# Shared Genetic Architecture Between Kidney Function and Alzheimer Disease Across Ancestries

**DOI:** 10.64898/2026.04.04.26350158

**Authors:** Diya Yang, Yihe Yang, Nicholas R. Ray, Mengxuan Li, Penelope Benchek, Dana C. Crawford, John F. O’Toole, John R. Sedor, Christiane Reitz, Audrey Lynn, Xiaofeng Zhu, Jonathan L. Haines, Alzheimer’s Disease Genetics Consortium (ADGC), William S. Bush

**Affiliations:** Department of Population and Quantitative Health Sciences, School of Medicine, Case Western Reserve University, Cleveland, OH, USA; Cleveland Institute for Computational Biology, Case Western Reserve University, Cleveland, OH, USA; Department of Genetics and Genome Sciences, School of Medicine, Case Western Reserve University, Cleveland, OH, USA; Departments of Molecular Medicine and Physiology and Biophysics, School of Medicine, Case Western Reserve University, Cleveland, OH, USA; Cleveland Clinic Medical Specialties Institute and Department of Heart, Blood and Kidney Research, Cleveland, OH, USA; Columbia University Irving Medical Center, New York, NY, USA; Gertrude H. Sergievsky Center, Taub Institute for Research on the Aging Brain, Departments of Neurology, Psychiatry, and Epidemiology, College of Physicians and Surgeons, Columbia University, New York, NY, USA

**Keywords:** Alzheimer disease, Estimated Glomerular Filtration Rate, multi-ancestry, genetic correlation, pleiotropy, Mendelian randomization

## Abstract

Epidemiological studies have consistently shown that chronic kidney disease is associated with increased Alzheimer disease risk. However, the underlying genetic architecture connecting these two conditions remains largely unexplored beyond genome-wide correlation analyses. Here, we conducted the first comprehensive, multi-ancestry, large-scale genetic investigation to identify shared genetic components between kidney function and Alzheimer disease.

We leveraged large-scale genome-wide association study summary statistics for estimated glomerular filtration rate (N≈1.5 million European, N≈145,000 African ancestry) and late-onset Alzheimer disease (N=63,926 and N=398,058 in two European cohorts; N=9,168 in African ancestry) corrected for competing risk bias. We deployed a novel analytical framework integrating linkage disequilibrium score regression and polygenic risk score analysis, local analysis of [co]variant association, conjunctional false discovery rate analysis with Bayesian colocalization and fine-mapping, and bidirectional cis-Mendelian randomization to identify vertical pleiotropy.

Despite the absence of genome-wide genetic correlation (r_g_ ≈ 0, p > 0.1), local genetic analysis uncovered striking regional heterogeneity. Sixteen pleiotropic loci were identified in individuals of European ancestry (conjunctional false discovery rate < 0.05), including *APOE*, *PICALM*, *SPI1*, and *EFTUD1*, alongside 15 loci with significant local genetic correlations. Fine-mapping revealed that most pleiotropic loci harbored distinct causal variants for kidney function and Alzheimer disease, indicating horizontal pleiotropy. An *APOE* ε4-defining allele (rs429358) was the sole variant with shared causality across both traits. We identified vertical pleiotropy using cis-Mendelian randomization at the *PICALM* and *EFTUD1* loci, providing evidence that kidney function-related genetic variants can causally affect Alzheimer disease risk at specific genomic loci. In contrast, loci such as *CD2AP*, *MAT1A*, and *SYMPK* demonstrated horizontal pleiotropy, reflecting shared upstream biological pathways rather than direct causal mediation. Notably, *APOE* was the only pleiotropic locus shared between European and African ancestry groups, underscoring marked ancestry-specific genetic architectures with critical implications for risk prediction and therapeutic translation.

Alzheimer disease and kidney function share genetic components at specific loci rather than genome-wide, with mixed directional effects and horizontal pleiotropy explaining the absent global correlation despite strong local signals. At a subset of loci, we identified directional effects linking kidney genetic determinants to Alzheimer disease risk using cis-Mendelian randomization, supporting a complex kidney–brain genetic axis. Most overlap reflects horizontal pleiotropy, with limited loci showing vertical pleiotropy. *APOE* was the only shared locus across ancestries, underscoring ancestry-specific architectures with implications for risk prediction. The multi-scale approach used here also provides a methodological framework for dissecting complex disease relationships missed by traditional genome-wide analyses.

## Introduction

Alzheimer disease (AD) and chronic kidney disease (CKD) represent two of the most pressing health challenges of aging populations.^1–4^ AD affects 50 million people worldwide and is projected to reach 200 million by 2050, with over one-third of individuals aged 80 and older affected globally.^5,6^ CKD presents a parallel burden, affecting 10-13% of adults and rising to nearly 40% among those 60 and older.^7,8^ Kidney function is clinically assessed through estimated glomerular filtration rate (eGFR), which has become a key quantitative outcome in genetic studies due to its scalability and statistical power.^9–12^ While traditionally studied as separate conditions, growing evidence suggests the two systems may be more closely connected than previously recognized.^13,14^

The repeated failure of clinical trials targeting amyloid-β and tau pathology has shifted research focus toward peripheral and systemic drivers of neurodegeneration.^15–18^ Growing evidence suggests that organ systems outside the brain, including the kidneys^19–22^, liver^23–25^, and gut microbiome^26,27^, contribute to AD pathology through impaired metabolic clearance^28,29^, systemic inflammation^30–32^, and compromised blood-brain barrier (BBB) integrity.^33,34^ This broader perspective recognizes that AD may arise not solely from intrinsic brain pathology but also from the brain’s vulnerability to peripheral organ dysfunction, particularly in aging populations where multi-organ comorbidity is common.^35,36^

Recent epidemiological evidence largely supports a connection between kidney function and brain health, though some findings remain inconsistent.^19,37–40^ Patients with moderate kidney impairment (eGFR <60 mL/min/1.73 m^2^) face 50% increased dementia risk^19^ and experience cognitive decline acceleration of approximately 1.5 years.^41,42^ This relationship extends beyond comorbidity: reduced kidney function correlates with elevated serum amyloid-β levels^43–46^, increased cerebrospinal fluid phosphorylated tau independent of cardiovascular factors^47^, and progressive BBB breakdown.^34,37,48,49^ However, it remains unclear whether these links are driven by common genetic factors or purely by environmental and comorbid conditions. The recent comprehensive review by Zuo *et al.* in *Brain* highlighted critical gaps in understanding the kidney-neurodegeneration link, specifically calling for genetic investigations to better understand the biological mechanisms linking these organ systems.^50^ Both conditions demonstrate substantial heritability (60-80% for AD with over 75 known risk loci identified^51–59^; 30-75% for CKD with 264 known genetic loci^60–62^), yet their genetic interconnection remains largely unexplored. Thus, the primary objective of our work is to define the genetic architecture linking kidney function and AD across ancestries.

Genome-wide association studies (GWAS) have now mapped hundreds of loci for late-onset Alzheimer disease and for kidney function traits such as eGFR.^56,63,64^ Intriguingly, several genes have emerged as candidates influencing both organ systems, suggesting potential shared biological mechanisms. A prominent example is *CD2AP*, which encodes an adaptor protein essential for podocyte integrity in the kidney and is also among the top loci influencing AD risk.^65^ Variants in *CD2AP* cause focal segmental glomerulosclerosis, and the same locus has been implicated in amyloid-β clearance and microglial function in the brain.^66^ Another example is *APOE*, the strongest genetic risk factor for AD, which plays a key role in lipid metabolism affecting renal and vascular health.^67–70^ Notably, the ε4 allele substantially increases AD risk^71–73^, whereas the ε2 allele, protective in AD^74^, has been associated with adverse lipid profiles and cardiovascular outcomes^75^ that may influence kidney health. These overlapping signals at well-characterized loci raise the question of whether a broader shared genetic architecture exists between kidney function and AD beyond these known genes.

Despite the clinical and mechanistic evidence, genetic studies have yielded inconsistent results. Mendelian randomization (MR) analyses have not provided robust evidence for causal relationships between AD and kidney function, with some studies suggesting effects in opposing directions, while genome-wide genetic correlation estimates have revealed weak or null signals.^76–79^ Most studies have also focused on European ancestry (EUR) populations, despite African ancestry (AFR) populations experiencing disproportionately higher CKD and AD burden. Genetic analyses offer unique advantages in addressing these limitations. Variants are assigned at meiosis, largely eliminating confounding, and methods now exist to distinguish horizontal pleiotropy from vertical pleiotropy. In horizontal pleiotropy, genetic variants influence both traits through shared upstream pathways, whereas in vertical pleiotropy, one trait causally mediates the effect on the other, which is a distinction with important clinical implications for intervention strategies.^80,81^

We hypothesize that shared genetic architecture between kidney function and AD is subtle and masked in genome-wide analyses due to a mixture of concordant and discordant effects across loci. Variants influencing shared pathways, such as inflammation, lipid metabolism, or vascular integrity, may exert opposing effects on each trait, leading to near-zero genome-wide genetic correlations despite substantial underlying biological overlap. Furthermore, competing risk bias may attenuate genetic signals if individuals with kidney disease or related comorbidities do not survive to the age of AD onset, thereby systematically excluding some shared risk variants from AD case populations. To address these gaps, our study leverages large-scale diverse genomic data and multiple complementary analytical approaches capable of detecting both concordant and discordant genetic sharing to comprehensively characterize the genetic mechanisms linking AD and kidney function.

In this study, we present the first comprehensive, multi-ancestry genome-wide investigation of shared genetic architecture between kidney function and AD. Our approach (**Fig. 1**) combined complementary analytical approaches: (1) Genome-wide correlation analyses to test for overall genetic correlation and polygenic score overlap between eGFR and AD; (2) Local genetic overlap mapping to pinpoint specific genomic loci with joint association signals, using methods like Local Analysis of [co]Variant Association (*LAVA*)^82^ and conjunctional false discovery rate (*conjFDR*)^83,84^ that increase power to detect pleiotropy; (3) Colocalization fine-mapping to distinguish whether overlapping signals at a locus likely arise from the same causal variant or independent variants in linkage disequilibrium (LD); and (4) cis-Mendelian randomization (cis-MR) to infer the magnitude and direction of effect between eGFR and AD at each pleiotropic locus accounting for horizontal pleiotropy by leveraging large GWAS datasets from EUR and AFR. We also implemented specific corrections to account for collider bias and competing risk. Using these adjusted models, we investigated whether a shared genetic basis links AD and eGFR and if this relationship persists across diverse ancestries. Ultimately, clarifying these connections can improve our understanding of AD pathogenesis and identify new genetic risk factors that span neurological and kidney function pathways.

**Figure 1.**
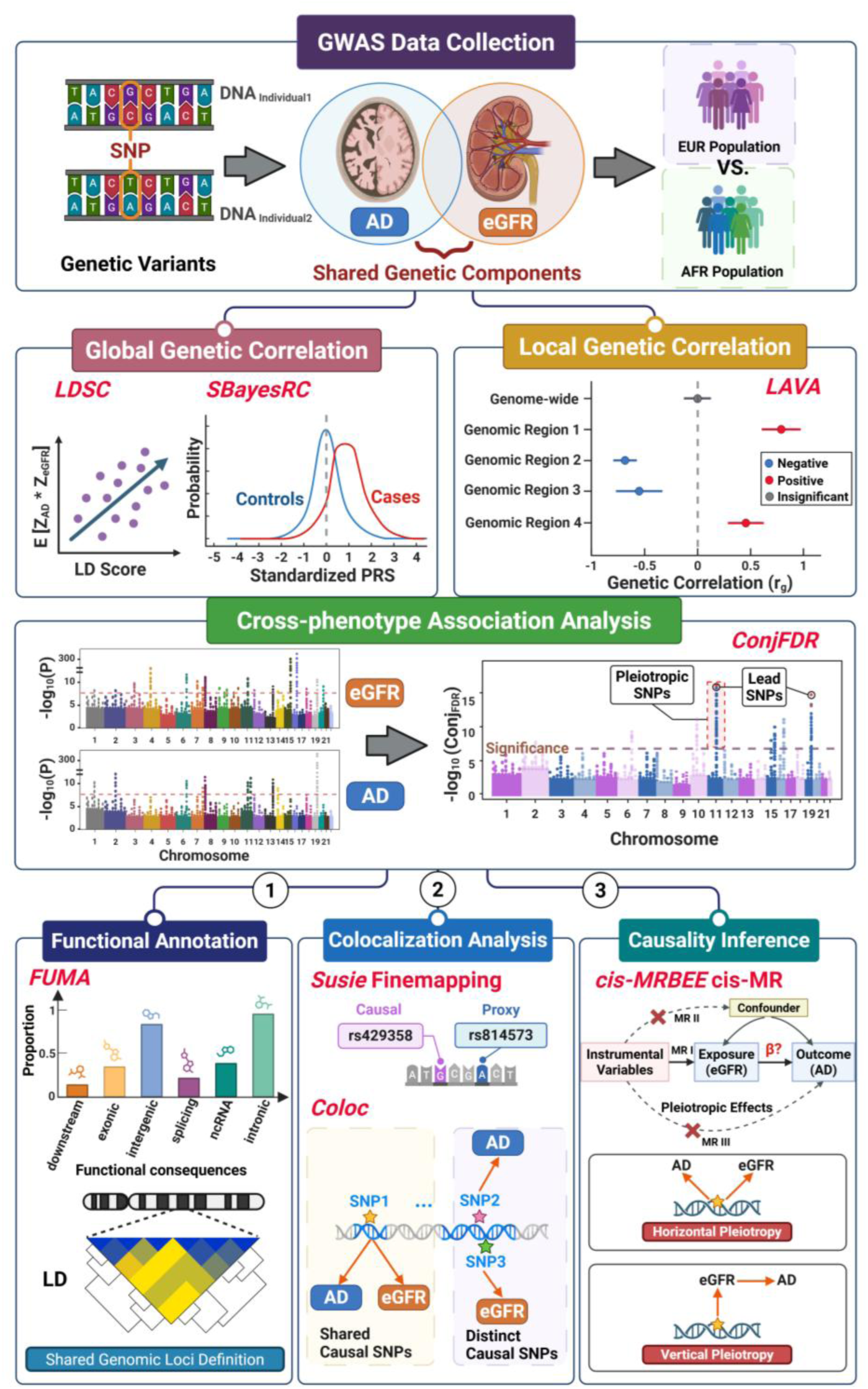
Comprehensive multi-level genetic analysis workflow investigating shared genetic architecture between eGFR and AD. Schematic overview of the systematic analytical pipeline employed to dissect genetic relationships between estimated glomerular filtration rate (eGFR) and Alzheimer’s disease (AD). The workflow begins with GWAS Data Collection (top), showing genetic variants (SNPs) from EUR and AFR populations analyzed for shared genetic components between brain (AD) and kidney (eGFR) phenotypes, comparing European versus African American ancestry groups. The analysis proceeds through four major analytical tiers: (1) Global Genetic Correlation using *LDSC* (Linkage Disequilibrium Score Regression) to estimate genome-wide genetic correlation, and *SBayesRC* for polygenic prediction with functional annotations, showing the relationship between LD scores and test statistics, and distribution of polygenic risk scores between cases and controls; (2) Local Genetic Correlation using *LAVA* (Local Analysis of [co]Variant Association) to identify specific genomic regions showing positive, negative, or insignificant genetic correlations (rg) across four genomic regions, with genome-wide significance testing; (3) Cross-phenotype Association Analysis using *ConjFDR* (Conjunctional False Discovery Rate) to identify pleiotropic SNPs (variants affecting both traits), showing Manhattan plots for both eGFR and AD with significance thresholds (FDR 0.001, 0.01, 0.05) and identification of lead SNPs across chromosomes. The bottom tier branches into three parallel downstream analyses: (1) Functional Annotation via *FUMA* (Functional Mapping and Annotation) examining functional consequences of identified variants across different functional genomic contexts, with linkage disequilibrium (LD) patterns visualized in a correlation matrix for shared genomic loci to help with the shared genomic loci definition; (2) Colocalization Analysis using *Susie* Fine-mapping to distinguish between shared causal SNPs versus distinct causal SNPs in the same locus, employing the *Coloc* method for proxy and causal variant identification in both eGFR and AD, with SNP-level resolution; and (3) Causality Inference through cis-MR (Mendelian Randomization) to evaluate locus-specific directional alignment of genetic effects between eGFR and AD. Forward (eGFR to AD) and reverse (AD to eGFR) cis-MR analyses distinguish loci showing directional effects compatible with kidney-to-brain causal pathways, reverse directionality, bidirectional effects, or patterns consistent with shared regulatory architecture without direct causal mediation. This integrated workflow enabled comprehensive characterization of genetic sharing patterns, from genome-wide correlations to locus-specific mechanisms and potential causal pathways, across two ancestral populations and analytical scales. The figure was created using *BioRender* and included with permission for publication.

## Materials and methods

### GWAS summary statistics and data sources

We used publicly available GWAS summary statistics for all analyses. For AD in EUR, we used clinically adjudicated data from Kunkle *et al.* (Stage 1, N=63,926)^52^ and Wightman *et al.* (N=398,058)^85^, prioritizing clinically adjudicated samples over proxy cases. For AD in AFR, we used summary statistics from Ray *et al.* (N=9,168).^86^

We implemented a correction for participation bias in AD GWAS following the collider bias correction approach described by Wang *et al.*^87^ Competing risk occurs when individuals with certain conditions die or become unable to participate before AD onset, creating bias that distorts genetic associations. Using multivariable *MRBEE*^88^, we adjusted AD effect sizes for competing mortality from five major diseases.

For kidney function, we analyzed eGFR. For EUR, we performed fixed-effects meta-analysis (N=1,482,631) combining data from the Million Veteran Program (MVP)^89^, CKDGen Consortium^90^, and UK Biobank^91^ using *Stouffer*’s method^92^ to harmonize effect scales. For AFR eGFR, we meta-analyzed data from Hughes *et al.*^93^, MVP African Americans^94^, and the COGENT-Kidney Consortium^95^. Individual-level genotype data for polygenic risk score (PRS) analysis were obtained from the Alzheimer’s Disease Genetics Consortium (ADGC).^96^ Genome-wide association results for all trait-ancestry combinations are visualized in **Supplementary Figs. S1–S3**.

All GWAS utilized were approved by relevant ethics committees, and ADGC participants provided written informed consent, as detailed in the **Supplementary Methods**.

### Genome-wide level genetic analyses

#### Genetic correlation and polygenic risk scores

We estimated genetic correlations using LD score regression (*LDSC*)^97^ with 1000 Genomes Phase 3 reference panels^98^, restricted to HapMap3^99^ Single Nucleotide Polymorphisms (SNPs). We calculated PRS using *SBayesRC*^100,101^, which integrates functional genomic annotations to improve SNP weight estimation, with final scores computed in *PLINK*.^102^ Associations between eGFR PRS and AD diagnosis were estimated using logistic regression adjusting for age, sex, and the first three principal components.

#### Conjunctional false discovery rate analysis

To identify pleiotropic variants, we performed conjunctional false discovery rate (*conjFDR*) analysis. This Bayesian method estimates the posterior probability that a SNP is null for either phenotype given observed p-values for both traits, enabling detection of shared variants regardless of effect direction. We defined significant pleiotropic loci at conjFDR<0.05 to be consistent with prior cross-trait genetic studies. Genomic loci were defined per *FUMA*^103^ protocols, merging independent significant SNPs (r² < 0.6) within 250 kb, and SNPs were annotated using *CADD*^104^ (impact on protein structure/function), *RegulomeDB*^105^ (regulatory roles), and *chromatin state* information (transcriptional/regulatory effects). Detailed mathematical definitions are in the Supplementary Methods.

*ConjFDR* identifies variants jointly associated with both traits regardless of effect direction, making it robust to opposing allelic effects within loci. In contrast, local genetic correlation methods quantify effect size correlations and may yield null results when opposing effects coexist. Accordingly, *conjFDR* served as the primary framework for pleiotropic locus discovery. Unless otherwise noted, primary results reflect bias-corrected analyses, with uncorrected findings reported in the Supplementary Materials.

### Locus-level genetic analyses

#### Local genetic correlations using LAVA

We assessed local genetic correlations using *LAVA* with ∼1 Mb semi-independent LD blocks (EUR: 2,495 blocks; AFR: 2,687 blocks) derived from 1000 Genomes Phase 3 reference panels. *LDSC* intercepts accounted for sample overlap. Genomic loci with significant local h²_SNP_ (p<1E-4) proceeded to bivariate analysis, with significant loci identified at FDR<0.05.

#### Cross-trait colocalization analysis

We performed cross-trait colocalization using *coloc* (v.5.2.3)^106^ with *SuSiE*^107^ fine-mapping to test whether pleiotropic loci share causal variants or harbor distinct variants in LD or proximity. Prior probabilities were set as P_1_=P_2_=1E-4, P_12_=5E-6. Five hypotheses were evaluated: H_0_ (no association), H_1_/H_2_ (single trait associations), H_3_ (distinct causal variants), H_4_ (shared causal variant). We selected loci with shared causal variants with PP.H_3_+PP.H_4_≥0.8 and PP.H_4_/PP.H_3_≥5.

#### Cis-Mendelian randomization analysis

We applied cis-Mendelian randomization bias correction estimating equation (*cis-MRBEE*)^108^ to evaluate potential causal effects of eGFR on AD risk at pleiotropic genomic loci identified through *conjFDR* analysis. This method addresses weak instrument bias and sparse genetic architecture through two-stage fine-mapping and sparse prediction. It distinguishes vertical pleiotropy (causal mediation) from horizontal pleiotropy (independent effects via shared upstream pathways). Findings were validated with *MAPLE*^109^ cis-MR. Analyses were restricted to EUR due to limited statistical power in AFR.

### Statistical analysis

Genome-wide genetic correlations were estimated using LD score regression. Polygenic risk scores were computed via *SBayesRC*, with associations tested by logistic regression adjusting for age, sex, and three principal components. Local genetic correlations were assessed using *LAVA*, with significance determined at Bonferroni-corrected p < 6.39E-5 or Benjamini-Hochberg FDR < 0.05. Pleiotropic variants were identified at conjFDR < 0.05 in Europeans and conjFDR < 0.2 in African ancestry. Bayesian colocalization with *SuSiE* fine-mapping distinguished shared from distinct causal variants. Bidirectional cis-Mendelian randomization used *cis-MRBEE*, validated with *MAPLE*, to distinguish vertical from horizontal pleiotropy. Two-sided tests were used throughout with α = 0.05 unless otherwise noted. Analyses were performed in R v4.3.1, Python v3.7 and PLINK v2.0.

## Results

Throughout this study, effect directions for variant-specific analyses are described relative to the eGFR-decreasing allele, while regional analyses report whether genetic effects on eGFR and AD are positively or negatively correlated.

### Genome-wide genetic correlation and polygenic overlap are minimal

*LDSC* revealed no significant genome-wide genetic correlation between eGFR and AD in European ancestry (r_g_ = −0.06, SE = 0.05, p ≈ 0.24) using bias-corrected effect sizes, confirmed in Wightman *et al.* for validation (r_g_ ∼ 0, p ≈ 0.97). AFR analysis revealed unique challenges: reliable genome-wide r_g_ could not be estimated due to heritability estimates falling outside biologically plausible bounds, attributable to smaller sample size and reduced statistical power. These findings indicate that in aggregate and on a genome-wide level the genetic architectures of kidney function and AD do not overlap (**Table 1** and **Supplementary Table S3**).

**Table 1.**
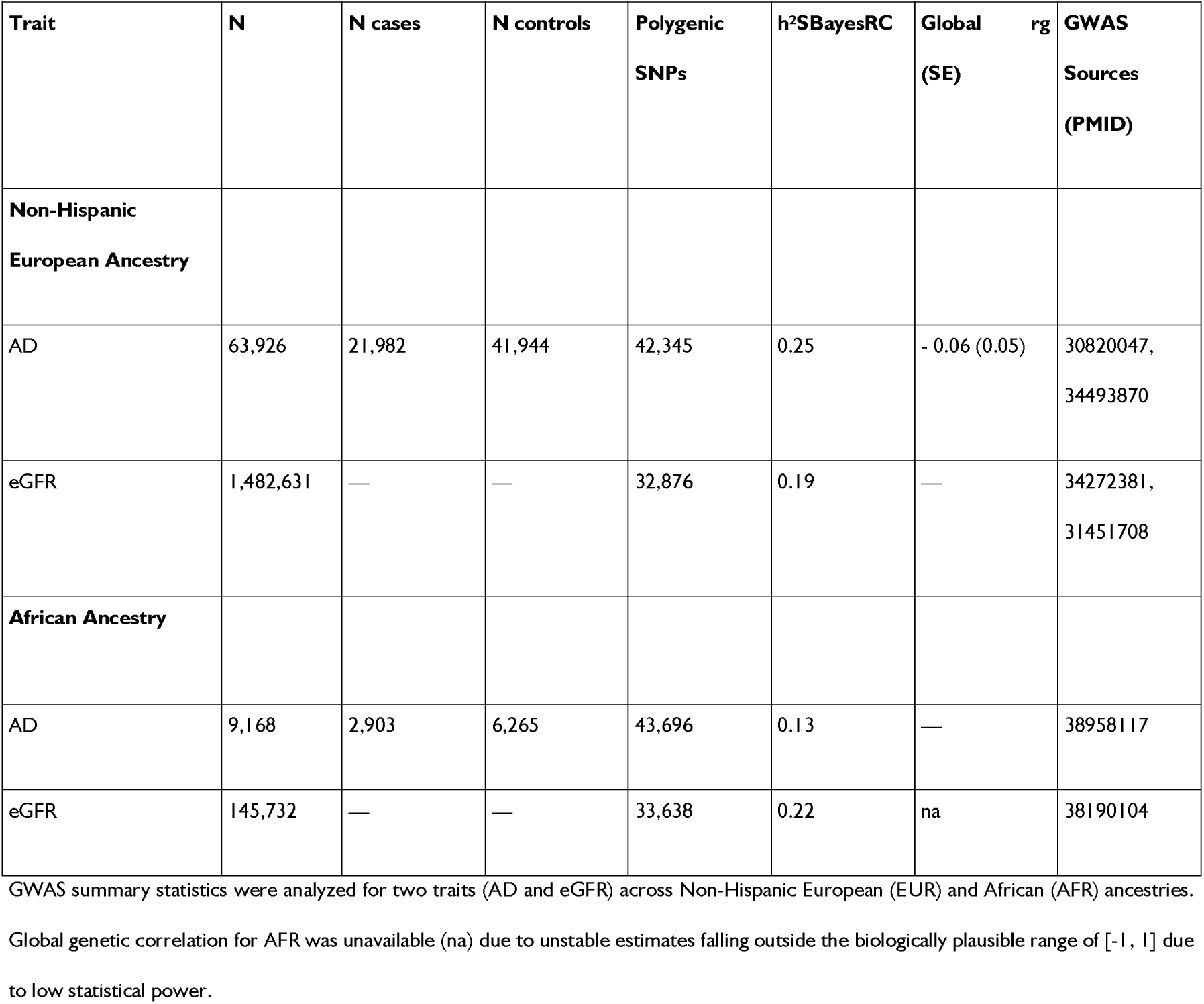
Overview of GWAS datasets used and genome-wide genetic correlation calculations between Alzheimer’s disease (AD) and estimated glomerular filtration rate (eGFR) across ancestries. GWAS Summary statistics used for the two traits analyzed in this study across Non-Hispanic European (EUR) and African (AFR) ancestries. For each trait and ancestry, the table provides: total sample size (N), number of cases and controls (for AD), number of polygenic SNPs used in analyses, SNP-based heritability estimates from both *LDSC* (h²_LDSC_) and *SBayesRC* (h²_SbayesRC_) methods, genome-wide genetic correlation (global r_g_) with standard error, and GWAS source PMIDs. In EUR populations, AD (N=63,926; 21,982 cases, 41,944 controls) showed h²_SbayesRC_=0.25, while eGFR (N=1,482,631) showed h²_SbayesRC_=0.19. The global genetic correlation between AD and eGFR in EUR was −0.06 (SE=0.05), not reaching statistical significance. In AFR populations, AD (N=9,168; 2,903 cases, 6,265 controls) showed lower h²_SbayesRC_=0.13, while eGFR (N=145,732) showed h²_SbayesRC_=0.22. Genome-wide genetic correlation estimates for AFR were not available (na) because *LDSC*-based rg estimates were unstable and fell outside the biologically plausible range of −1 to 1, reflecting limited precision associated with smaller sample size and reduced *LDSC* heritability estimates in this ancestry.

In EUR, AD showed SNP-based heritability (h^2^_SBayesRC_) of 0.25 via *SBayesRC*, indicating that common genetic variants explain roughly one-quarter of phenotypic variation in AD risk. eGFR showed similar heritability (h^2^_SBayesRC_ = 0.19), reflecting a moderately polygenic basis where genetic variants explain a substantial portion of trait variation. In AFR, AD showed lower heritability (h²_SBayesRC_ = 0.13), while eGFR showed h²_SBayesRC_ = 0.22. We report heritability estimates from *SBayesRC*, as this approach performs robustly under heterogeneous genetic architectures and complex LD structure, conditions under which *LDSC* may yield conservative estimates. These heritability estimates are consistent with prior reports from Liu *et al*.^59^, though likely represent underestimates of true heritability due to incomplete LD capture and common variant focus.

To further probe polygenic overlap, we constructed eGFR-PRS using *SBayesRC* in 31,315 European ancestry individuals from ADGC (15,063 cases, 16,252 controls) based on 32,876 SNPs. The eGFR-PRS showed no significant discriminative ability for AD status (OR = 0.98 per standard deviation increase, 95% CI = 0.96–1.01, SE = 0.012, p = 0.149; **Fig. 2B**), with PRS distributions between cases and controls overlapping almost completely (**Fig. 2A**).

**Figure 2.**
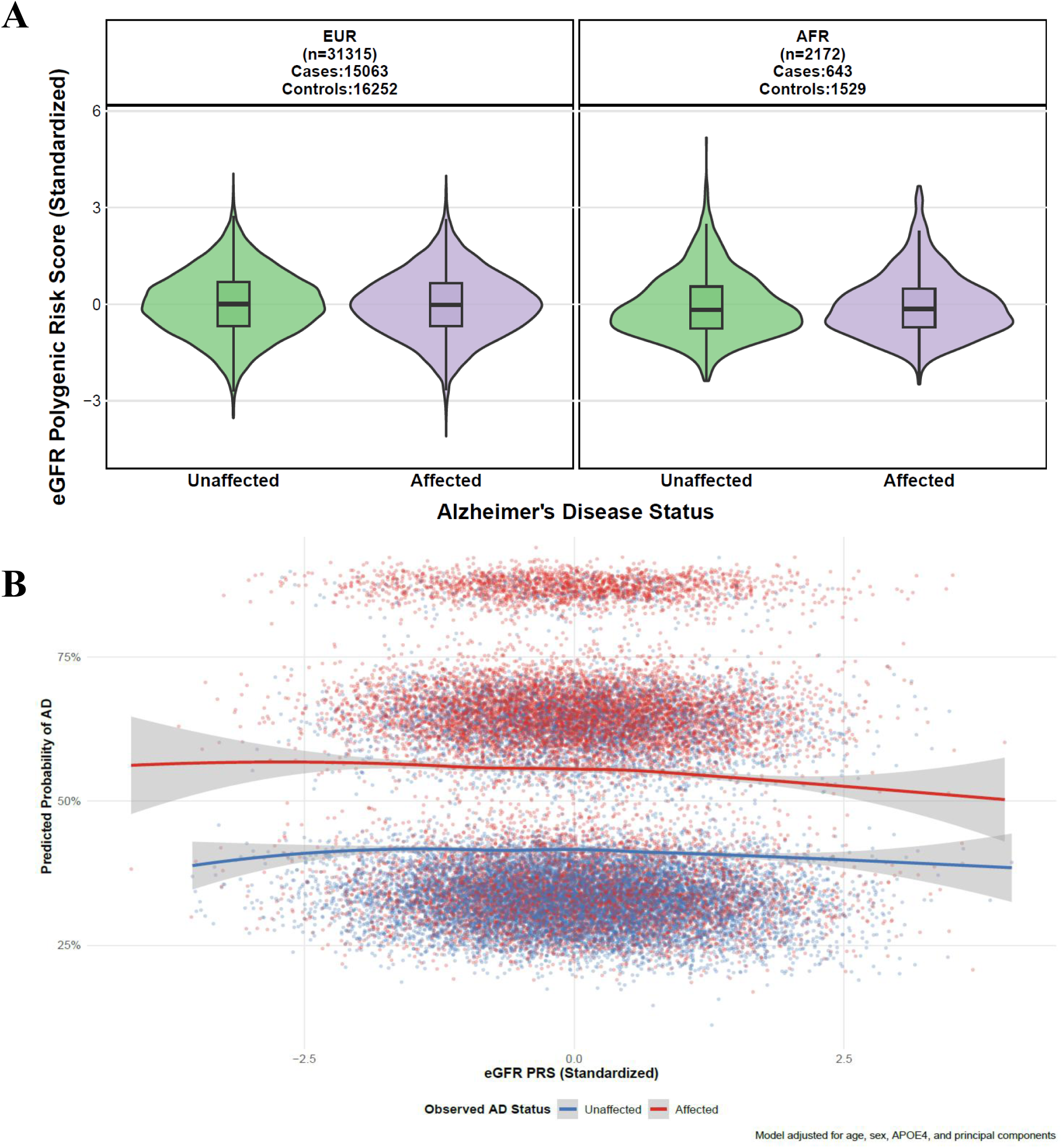
Distribution of eGFR polygenic risk scores and their association with AD risk across ancestries. **(A)** Violin plots with embedded box plots showing standardized eGFR-PRS distribution by AD status in EUR (n=31,315: 15,063 cases, 16,252 controls) and AFR (n=2,172: 643 cases, 1,529 controls). Unaffected individuals shown in green, affected individuals in purple. Box plots display median (center line), interquartile range (box), and whiskers extending to 1.5 times IQR. **(B)** Scatter plot showing relationship between eGFR-PRS (x-axis) and predicted AD probability (y-axis) in EUR. Red points and line represent affected individuals, blue points and line represent unaffected individuals, with gray shading showing 95% confidence intervals from *LOESS* curves. The modest association weakens after *APOE* ε4 adjustment, suggesting the relationship is largely driven by *APOE* genotype.

When stratified by tertiles, individuals in the highest eGFR-PRS tertile showed a nominal trend toward lower AD risk (OR = 0.94, 95% CI = 0.89–1.00, p = 0.033), but this signal weakened after adjusting for *APOE* ε4 genotype (adjusted OR = 0.95, 95% CI = 0.89–1.00, p = 0.064). Predicted AD probability remained essentially flat across the PRS spectrum, with any subtle gradient eliminated after *APOE* adjustment (**Fig. 2B; Supplementary Figs. S7–S10**).

In the African ancestry sample (N=2,172; 643 cases, 1,529 controls), neither EUR-derived nor AFR-specific PRS showed significant association with AD (all p > 0.25; **Supplementary Tables S6, S7**). These PRS analyses reinforce the hypothesis of locus-specific rather than genome-wide genetic sharing.

Because AD GWAS are susceptible to survival-induced collider bias, where competing terminal diseases reduce the probability of surviving to AD diagnosis and thereby create spurious genetic associations, we applied a multivariable Mendelian randomization (MVMR) correction.^88^ Specifically, survival to study participation acts as a collider: conditioning on it induces non-causal associations between genetic variants and competing diseases. Using MRBEE, we adjusted AD effect sizes for genetic pathways operating through five competing terminal diseases (coronary artery disease,^110^ heart failure,^111^ type 1 diabetes,^112^ kidney cancer,^113^ and lung cancer^114^) that showed negative effect estimates on AD risk consistent with selection bias rather than genuine protective effects, yielding debiased summary statistics for all downstream analyses (**Supplementary Methods** and **Supplementary Fig. S28**).

### Local genetic correlations reveal regional hotspots

To test whether localized genetic relationships exist despite null global correlation, we partitioned the genome into semi-independent LD blocks using *LAVA*. Complete bivariate local genetic correlation results for all tested loci are provided in **Supplementary Tables S8-S14**.

In EUR, 15 of 781 tested loci (1.9%; i.e., those passing the univariate heritability threshold of p < 1E-4 from 2,495 total blocks) showed significant local correlations after *Bonferroni* correction (p < 6.39E-5). Over half (53%) were negative, indicating that genetic effects on eGFR and AD were inversely related in these loci. The strongest negative correlation occurred at the *APOE* locus (chr19:45040933-45893307, r_g_ = −0.34, 95% CI [-0.43, −0.25], p = 2.01E-12; **Fig. 3A**), spanning *APOE*, *TOMM40*, and *APOC1*. Conversely, we identified a positive correlation at chromosome 11 (chr11:95589814-96629176, r_g_ = 0.57, 95% CI [0.25, 0.90], p = 6.01E-4), a locus containing *MADD* and *PACS1*, reflecting concordant regional effect directions for eGFR and AD. Additional significant EUR loci included a second region on chromosome 11 near 47.4 Mb (chr11:47257514-48317148, r_g_ = −0.46, 95% CI [–0.92, – 0.19], p = 9.49E-4), encompassing *SPI1*, *PTPMT1*, and *CELF1*, again demonstrating opposing regional genetic effects across the two traits.

**Figure 3.**
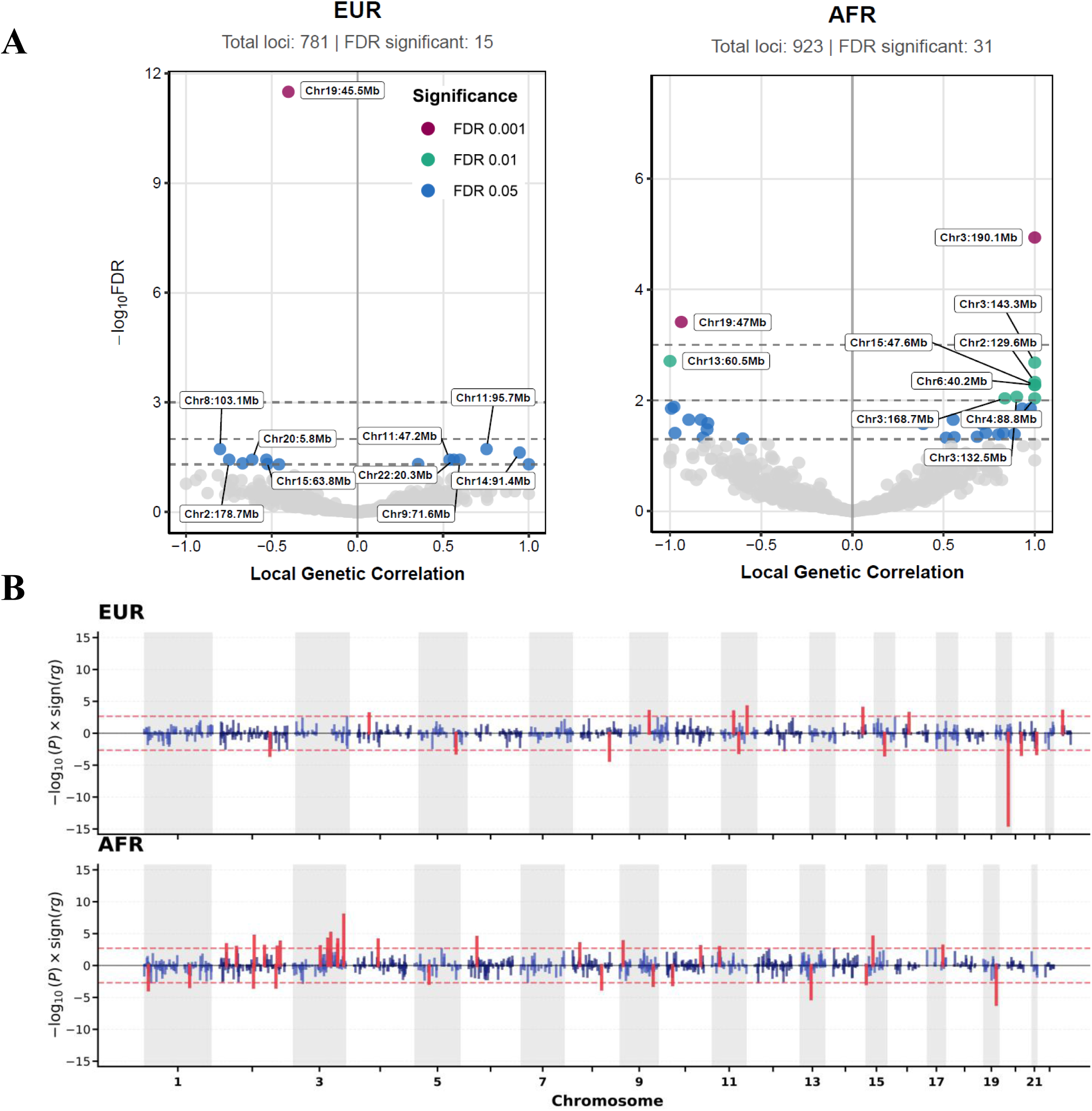
Local genetic correlations between AD and eGFR in European (EUR) and African American (AFR) ancestries using *LAVA*. **(A)** Genome-wide local genetic correlation (rg) analysis using *LAVA* (Local Analysis of [co]Variant Association) in European (EUR, left) and African American (AFR, right) ancestry populations. Volcano plots displaying local genetic correlation coefficients (x-axis) against −log10(FDR-adjusted P-value) values (y-axis) for all tested genomic loci. EUR analysis (left panel) identified 781 total testable loci with 15 FDR-significant regions, while AFR analysis (right panel) identified 923 total loci with 31 FDR-significant regions. Colored points indicate significance levels: pink (FDR < 0.001), teal (FDR < 0.01), and blue (FDR < 0.05), with gray points representing non-significant loci. Key regions are annotated with chromosomal positions (e.g., Chr19:45.5Mb in EUR, Chr3:190.1Mb in AFR). Dashed horizontal lines denote FDR significance thresholds. **(B)** Manhattan plots showing −log_10_(P) × sign(r_g_) values across all 22 autosomes for EUR (top) and AFR (bottom) populations. Positive correlations shown in red (above zero), negative correlations in blue (below zero), with alternating gray chromosomal bands for visualization. Dashed horizontal lines indicate Bonferroni-corrected significance threshold. AFR shows more uniform distribution of significant signals compared to EUR’s APOE-dominated pattern, demonstrating substantial ancestry-specific genetic architecture.

In African ancestry *LAVA* analysis, 31 of 923 tested loci (3.4%) showed significant local r_g_ at 5% FDR. The *APOE* region showed robust negative correlation in both ancestries (r_g_ = −0.34 in EUR, r_g_ = −0.94 in AFR). However, ancestry-specific patterns emerged (**Fig. 3A**): AFR analysis revealed additional significant loci on chromosomes 2, 3, 6, 8, 15, and 19 not detected in EUR. Many AFR signals had wide confidence intervals due to smaller sample size and very low SNP-based heritability (h^2^_SNP_) for both traits, and some significant r_g_ estimates likely reflect noise or false positive signals. These ancestry-specific patterns are visualized in **Fig. 3B**, showing the *APOE* region as the dominant signal in EUR while AFR exhibits more distributed signals across multiple chromosomes. Despite absent genome-wide r_g_, these regional analyses uncovered concentrated genetic correlations within loci of interest otherwise hidden by genome-wide averaging. Examples of null LAVA results with similar variance-covariance structure are shown in **Supplementary Figs. S12–S13**.

The direction of effects diverged between ancestries: positive local correlations (alleles raising eGFR associated with increased AD risk) predominated in AFR (64.5% of significant loci) versus negative correlations in EUR (53.3%). Only one significant locus (*APOE/TOMM40/APOC1* region) overlapped between ancestries, revealing substantial population-specific architecture. Notably, the *APOL1* locus on chromosome 22, known for strong kidney disease links in AFR, showed no significant local genetic correlation with AD. This may stem from GWAS models assuming additive effects while *APOL1* acts recessively.

### Cross-Phenotype association test identifies pleiotropic loci

To pinpoint specific genetic variants influencing both kidney function and AD, we applied conjunctional false discovery rate analysis. This approach leverages the observation that SNPs associated with one trait are often enriched for associations with related traits, thereby boosting power to detect shared signals that might not reach significance in single-trait analyses. Full results are provided in **Supplementary Tables S19-S23** and **Supplementary Figs. S4-S6**.

In European ancestry, we identified 16 distinct pleiotropic loci at conjFDR<0.05 (**Fig. 4A; Supplementary Table S19**). The strongest signals clustered around well-established AD genes: *SPI1* on chromosome 11 (lead SNP rs3824869, conjFDR=5.77E-6), *APOE* on chromosome 19 (lead SNP rs7412, conjFDR=1.30E-4), *PVRL2* (lead SNP rs11666329, conjFDR=6.05E-4), and *APOE/TOMM40* (lead SNP rs157580, conjFDR=2.21E-5; lead SNP rs115881343, conjFDR=5.27E-4). Beyond these known loci, we detected novel kidney-brain connections at *PICALM* (lead SNP rs648270, conjFDR=4.24E-3), *SCIMP* (lead SNP rs61182333, conjFDR=1.95E-2), *APOE/SYMPK* (lead SNP rs16980051, conjFDR=3.66E-2), and *EFTUD1* (also known as *EFL1*) (lead SNP rs905450, conjFDR=4.34E-2). A regional plot of the *SPI1* locus (**Fig. 4B**) illustrates the overlapping association peaks, with both eGFR and AD signals exceeding genome-wide significance across a shared genomic locus encompassing multiple genes including *SPI1*, *PTPMT1*, and *CELF1*. Notably, eGFR associations generally drove the conjFDR significance at most loci, reflecting the substantially larger sample size and greater statistical power of the kidney function GWAS.

**Figure 4.**
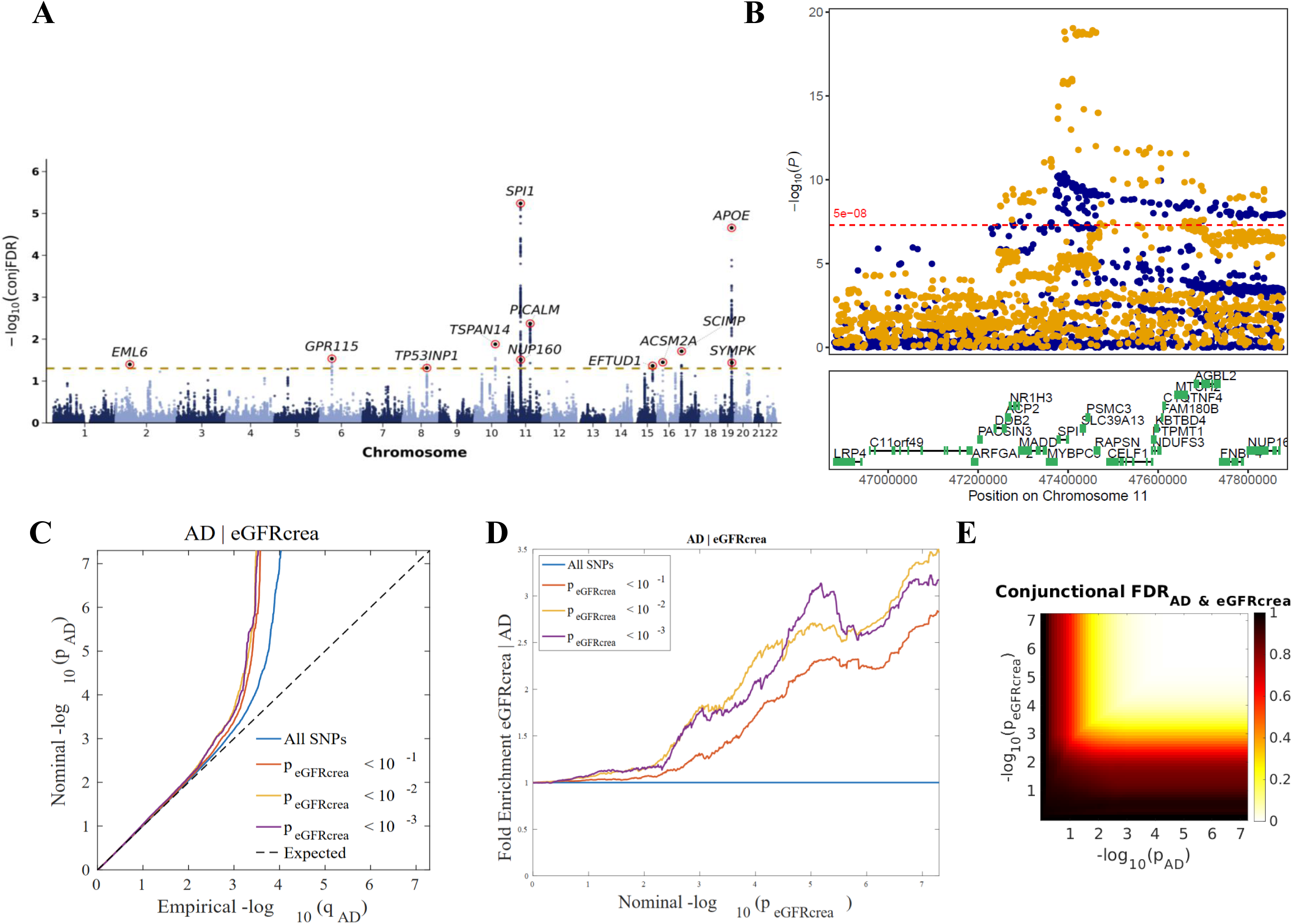
Pleiotropic signals from conjunctional false discovery rate (conjFDR) analysis for AD and eGFR in European ancestry. **(A)** Manhattan plot showing −log10(conjFDR) values (y-axis) across chromosomes (x-axis), with alternating blue and gray points by chromosome. Orange dashed line indicates conjFDR<0.05 threshold. Sixteen independent loci identified, with strongest signals at *SPI1* (Chr11) and *TOMM40/APOE* (Chr19), plus *PICALM*, *TSPAN14*, *GPR111*, *NDUFAF6*, *SCIMP*, and *EFTUD1*. **(B)** Regional plot of Chr11 *SPI1* locus (47.0-47.8 Mb) showing three panels: top panel displays −log10(P) values with orange and yellow points for variants exceeding genome-wide significance (red dashed line at −log[[(P)=7.3); middle panel shows gene annotations. **(C)** Q-Q plot for AD conditioned on eGFR association, with different thresholds: all SNPs (blue), P_eGFR<10E-3 (orange), P_eGFR<10E-2 (green), P_eGFR<10E-1 (red), versus expected null (dashed line). **(D)** Fold enrichment plot with color gradient from red (low enrichment) to yellow to blue (high enrichment). **(E)** ConjunctionalFDR heatmap with color scale from yellow (conjFDR=1) to dark red (conjFDR approaching 0), showing strongest evidence in upper right corner where both traits show strong associations.

Evidence for cross-trait enrichment is visualized in **Figs. 4C-E**. Conditional Q-Q plots (**Fig. 4C**) revealed progressive leftward deflection of AD p-values when stratified by eGFR association strength, where SNPs in the top eGFR stratum showed approximately 3-fold enrichment for AD associations compared to unstratified SNPs, providing quantitative evidence for pleiotropy between the two traits. Fold enrichment analysis (**Fig. 4D**) demonstrated increasing enrichment as the eGFR association threshold became more stringent, with distinct peaks visible at specific p-value combinations. The two-dimensional *conjFDR* heatmap (**Fig. 4E**) confirmed that joint significance concentrates in the upper-right quadrant where both traits show strong associations, with the color gradient transitioning from yellow (conjFDR=1) to dark red (conjFDR approaching 0).

To further characterize shared loci, we evaluated allelic effect directions using Z-score concordance for lead pleiotropic variants. Across EUR loci, effect directions showed locus-specific heterogeneity: at most loci, the eGFR-decreasing allele was associated with increased AD risk, while at others the eGFR-decreasing allele was associated with decreased AD risk (**Supplementary Fig. S11**; **Supplementary Tables S16-S22**). Functional annotation of pleiotropic SNPs indicated that the majority mapped to non-coding regions, with most variants located in intronic and intergenic regions across both discovery and validation datasets (**Supplementary Figs. S4-S6**; **Supplementary Tables S19-S21**), consistent with regulatory mechanisms underlying shared genetic effects.

African ancestry analysis was constrained by smaller sample sizes, yielding no loci at conjFDR<0.05. Using a relaxed threshold of conjFDR<0.2 due to reduced statistical power, we identified five suggestive loci (**Fig. 5A, C; Supplementary Table S23**): chromosome 6 near *RP1-292B18.4* (lead SNP rs140069490, conjFDR=0.179), chromosome 17 near *PITPNM3* (lead SNP rs12453781, conjFDR=0.143), chromosome 7 near *AUTS2* (lead SNP rs10242748, conjFDR=0.131), chromosome 11 near *RP11-64I17.1* (lead SNP rs35302065, conjFDR=0.184), and chromosome 19 near *ZNF254* (lead SNP rs3943791, conjFDR=0.183). Regional association plots for both AD and eGFR at the chromosome 7 *AUTS2* locus (**Fig. 5B**) display the lead SNP rs10242748 with surrounding variants colored by LD. The conjFDR heatmap for African ancestry (**Fig. 5D**) showed a similar pattern to EUR but with weaker overall signal intensity, reflecting reduced statistical power. Strikingly, *APOE* was again the only overlapping locus between ancestries, with the remaining signals appearing population-specific.

**Figure 5.**
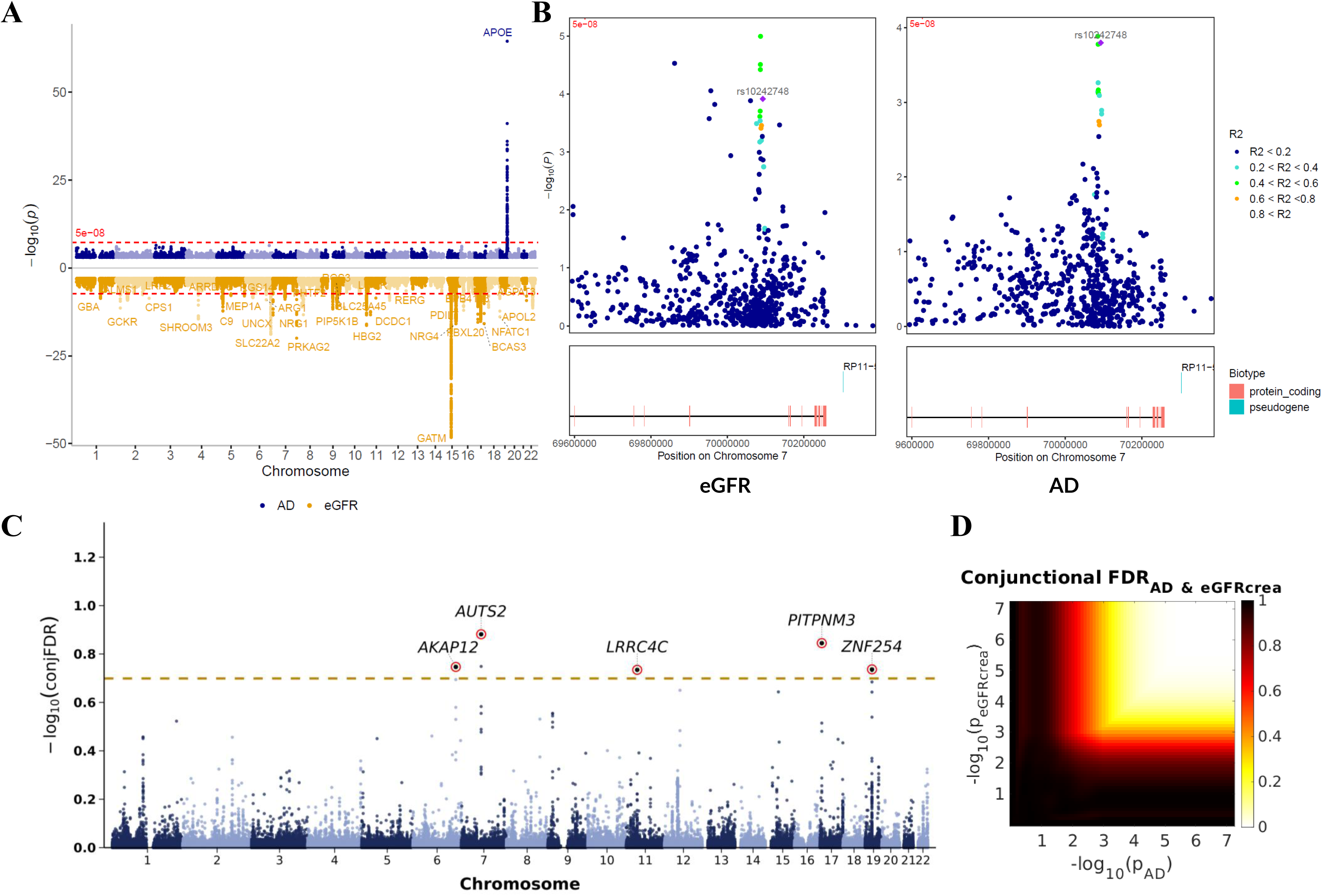
ConjFDR analysis for pleiotropic loci in African American ancestry. **(A)** Miami plot showing AD (top, blue points) and eGFR (bottom, orange/yellow points) GWAS results with −log10(P) on y-axis and chromosomes on x-axis. **(B)** Regional association plots for Chr7:70Mb region showing −log10(P) for both traits with gene annotations. Lead SNP is highlighted in green. Blue points indicate eGFR associations, black points indicate AD associations. **(C)** Manhattan plot of conjFDR identifying five pleiotropic loci (FDR<0.2): *AUTS2* (Chr7), *RP11-64I17.1* (Chr11), *PITPNM3* (Chr17), *ZNF254* (Chr19), and *RP1-292B18* (Chr6). Red points represent significant AD and eGFR associations, with gray chromosomal bands alternating for visualization. **(D)** *ConjFDR* heatmap with color scale from yellow (conjFDR=1) to dark red (conjFDR approaching 0), showing strongest conjunctional evidence in upper right where both traits show strong associations.

### Distinct but colocalized causal variants at most pleiotropic loci

A critical question is whether identified pleiotropic loci harbor the same causal variant affecting both traits or distinct variants in LD or near each other. We performed Bayesian colocalization integrated with *SuSiE* fine-mapping to distinguish these scenarios, with complete results in **Supplementary Tables S24-S28** and **Supplementary Fig. S14**.

Among EUR pleiotropic loci meeting our analysis criteria, most showed evidence for distinct causal variants driving eGFR and AD associations. At the *SPI1* locus (Chr11:47.4Mb), colocalization strongly supported hypothesis H_3_ (PP.H_3_ = 0.997), indicating that separate variants in proximity independently influence each trait. This pattern, termed “horizontal pleiotropy”, suggests the *SPI1* locus contains multiple functional elements with tissue-specific regulatory effects rather than a single variant with dual consequences. The *APOE* locus showed strong colocalization evidence (PP.H_4_ ≈ 0.85) and fine-mapping identifying the ε4 allele (rs429358, PIP ∼0.999) as the shared driver of both reduced eGFR and elevated AD risk. Unlike other pleiotropic loci where distinct colocalized variants drove associations, *APOE* represents true biological pleiotropy with a single SNP affecting both organ systems.

### Cis-Mendelian randomization provides evidence of vertical pleiotropy

Identifying shared loci does not establish whether kidney function causally influences AD or whether both traits simply share upstream genetic regulators. To distinguish these possibilities, we performed bidirectional cis-MR at each pleiotropic locus using *Cis-MRBEE*, with validation by *MAPLE*. Complete results appear in **Supplementary Tables S29-S32** and **Supplementary Figs. S15-S27**.

Two loci showed clear evidence of vertical pleiotropy, where genetic effects on AD are mediated through kidney function. At the *PICALM* locus (Chr11:85.7Mb), genetically predicted lower eGFR was associated with reduced AD risk (β = 0.711, SE = 0.206, P=5.65E-4), with consistent effect directions across instrumental variables and minimal evidence of horizontal pleiotropy (**Fig. 6B**). In contrast, at the *EFTUD1* locus (Chr15:81.4-83.4 Mb), genetically predicted lower eGFR was associated with higher AD risk (β = −0.97, SE = 0.36, P=6.9E-3; **Fig. 6A**). Notably, these two loci showed opposite effect directions: at *EFTUD1*, the eGFR-decreasing allele increased AD risk, whereas at *PICALM*, the eGFR-decreasing allele decreased AD risk. This directional heterogeneity demonstrates that eGFR-related genetic effects on AD risk vary by locus. At *EFTUD1*, the findings align with the hypothesis that impaired kidney clearance of neurotoxic metabolites may accelerate AD pathology, whereas the *PICALM* pattern suggests more complex relationships between kidney-related genetic variation and neurodegeneration.

**Figure 6.**
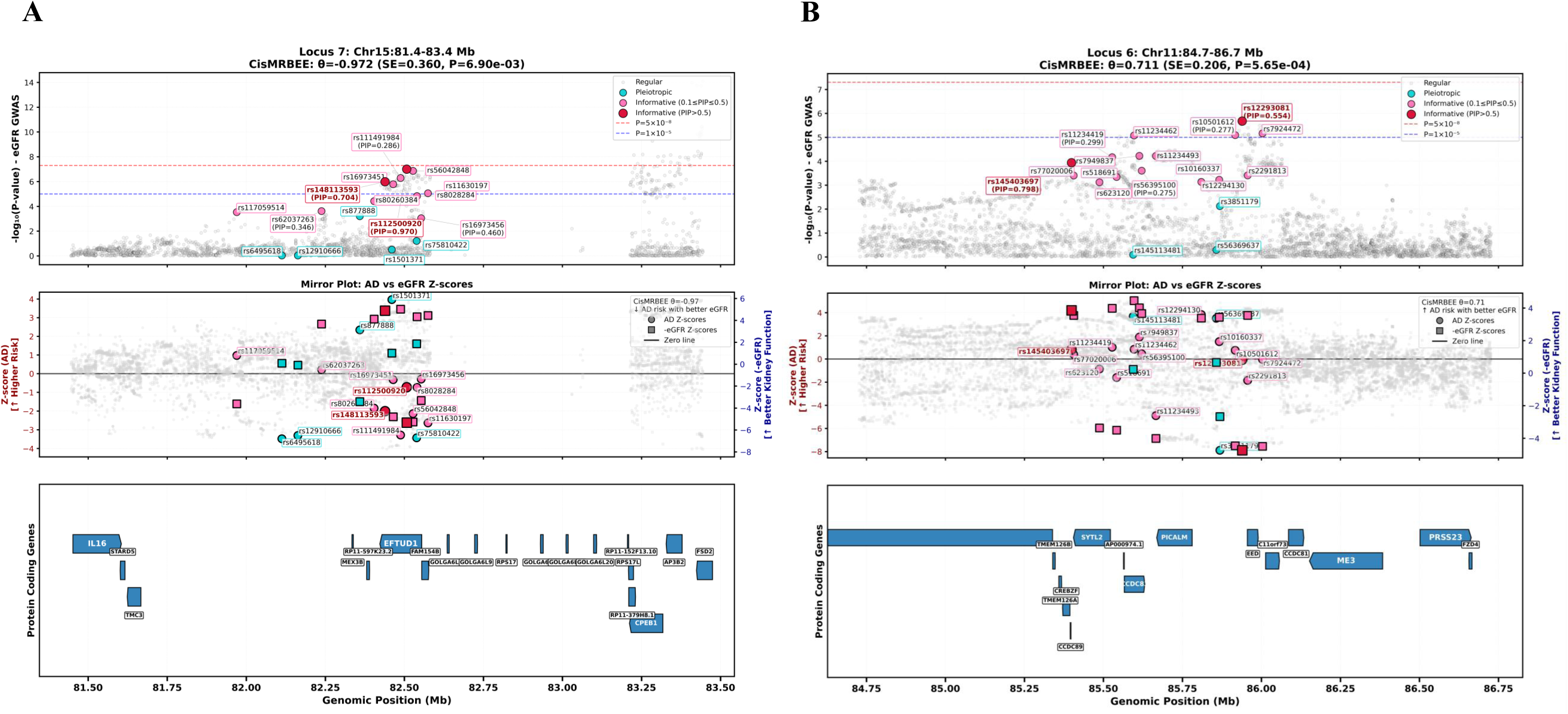
Cis-Mendelian randomization analysis of eGFR-to-AD causal effects at two genomic loci. Locus-specific cis-MR visualization showing SNP-level evidence for causal effects of kidney function (eGFR) on Alzheimer’s disease (AD) risk. Effect directions are reported relative to the eGFR-decreasing allele, consistent with the convention used throughout the main text. A negative θ indicates that eGFR-decreasing alleles increase AD risk; a positive θ indicates that eGFR-decreasing alleles decrease AD risk. Both panels display three integrated components: **(Top) eGFR GWAS Manhattan plot.** The y-axis shows −log_10_(P-value) for eGFR associations across genomic positions (x-axis, in megabases). SNPs are colored by category: light gray (regular), dark turquoise (pleiotropic), hot pink (informative with PIP 0.1–0.5), dark magenta (informative and pleiotropic), and crimson red (informative with PIP>0.5). Red dashed line indicates genome-wide significance (P=5E-8); blue dashed line indicates suggestive significance (P=1E-5). Lead variants are labeled with rsIDs and posterior inclusion probabilities (PIP). **(Middle) Mirror plot of AD vs eGFR Z-scores.** A dual-axis plot comparing effect sizes between traits. The left y-axis (dark red) shows AD Z-scores where higher values indicate increased AD risk. The right y-axis (dark blue) shows inverted eGFR Z-scores (-eGFR), where higher values indicate better kidney function. Circles represent AD Z-scores; squares represent -eGFR Z-scores. The black horizontal line marks zero. When a variant shows a positive AD Z-score (above zero) alongside a positive -eGFR Z-score (meaning the eGFR-increasing allele), this indicates that alleles associated with better kidney function are associated with higher AD risk, which is consistent with a positive θ (as in panel B). Conversely, concordant negative values support a negative θ (as in panel A). The legend displays the *CisMRBEE* θ estimate and indicates the direction of effect. **(Bottom) Protein-coding gene track.** Blue arrow-shaped boxes indicate protein-coding genes within the locus, with arrow direction indicating transcriptional strand orientation. Gene names are displayed within or adjacent to each gene body. **(A)** Locus 7 (Chr15:81.4–83.4 Mb): *CisMRBEE* θ=-0.972 (SE=0.36, P=6.9E-3), indicating that genetically predicted lower eGFR is associated with higher AD risk at this locus. **(B)** Locus 6 (Chr11:84.7–86.7 Mb): *CisMRBEE* θ=0.711 (SE=0.206, P=5.65E-4), indicating that genetically predicted lower eGFR is associated with lower AD risk at this locus.

In contrast, three loci, *CD2AP* (Chr6:47.7 Mb), *MAT1A* (Chr10:82.3 Mb), and *SYMPK* (Chr19:46.3 Mb), exhibited horizontal pleiotropy. Here, genetic variants influenced both traits but without evidence that one mediates the other. This pattern suggests these loci harbor genes affecting kidney and brain through independent biological pathways. The *SPI1* locus presented a more complex picture, with significant effects in both forward (eGFR to AD: β = −0.85, P=1.84E-2) and reverse directions. This bidirectional signal could reflect true reciprocal causation, tightly linked regulatory variants with independent effects, or limitations in distinguishing causality at loci with dense functional architecture. The *APOE* region similarly showed reverse causality, where genetic liability for AD influenced eGFR, consistent with APOE’s established pleiotropic effects across multiple organ systems. Methodological robustness was confirmed by strong agreement between *Cis-MRBEE* and *MAPLE* estimates across all loci (r = 0.92, 95% CI [0.78-0.97], P<0.001).

### Convergent Evidence from Multiple Approaches Validates and Characterizes Pleiotropic Loci

Synthesizing evidence across all analytical approaches, we categorized 11 genome-wide significant shared loci into mechanistic classes using a systematic decision framework (**Fig. 7**). Among these 11 loci, seven showed significant local r_g_ and six showed high colocalization probability (**Fig. 7A**). The systematic classification based on MR results and colocalization patterns (**Fig. 7B**) identified five mechanistic categories. First, ambiguous bidirectional pleiotropy characterized two loci (Chr11:47.4 Mb/SPI1, Chr17:5.1 Mb/SCIMP) showing significant bidirectional MR with shared causal variants, suggesting either reciprocal causation or tightly linked regulatory elements affecting both traits. Second, bidirectional effects with distinct variants were observed at the *APOE* locus only, where both forward and reverse MR were significant but mediated by different causal variants in linkage disequilibrium. Third, vertical pleiotropy with kidney-to-brain causation was evident at two loci (Chr11:85.7 Mb/*PICALM*, Chr15:82.4 Mb/*EFTUD1*), where improved kidney function causally reduces AD risk, potentially through enhanced protein clearance or reduced systemic inflammation. Fourth, horizontal pleiotropy characterized three loci (Chr6:47.7 Mb/*CD2AP*, Chr10:82.3 Mb/*MAT1A*, Chr19:46.3 Mb/*SYMPK*), where genetic variants affect both traits through independent mechanisms, possibly reflecting shared developmental programs or cellular processes. Fifth, three loci showed joint association without significant causal relationships in either direction, potentially due to complex polygenicity, insufficient instrument strength, or gene-environment interactions not captured by our analyses (**Supplementary Table S32**).

**Figure 7.**
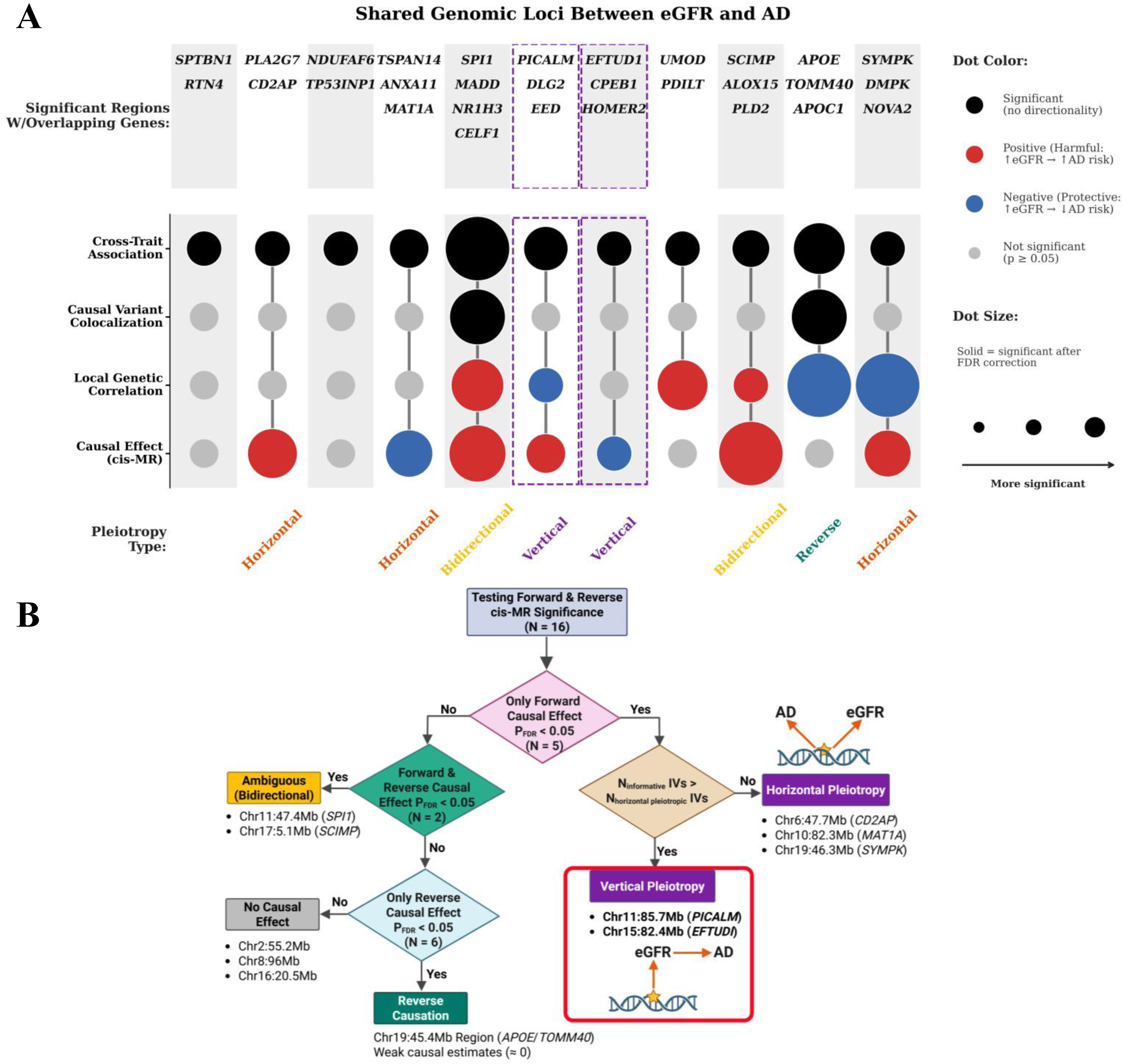
Integration of multi-level genetic evidence reveals distinct pleiotropic mechanisms. **(A) UpSet plot synthesizing results across 11 significant genomic regions shared between eGFR and AD**. They were consolidated from 16 loci, as five overlapping loci in the *APOE* region were merged into a single region, with four analytical tiers: (1) Cross-Trait Association (*conjFDR* analysis): dot size indicates conjFDR significance (larger means more significant; all regions conjFDR < 0.05); (2) Causal Variant Colocalization (*coloc.SuSiE* analysis): filled black or gray circles indicate shared/distinct causal variants (PP.H3 + PP.H4 >= 0.8); (3) Local Genetic Correlation (*LAVA* analysis): red dots indicate positive (harmful: increased eGFR associated with increased AD risk) genetic correlation, blue dots indicate negative (protective: increased eGFR associated with decreased AD risk) correlation, and gray dots indicate non-significant results (p >= 0.05); (4) Causal Effect (cis-MR analysis): dot color and size follow the same convention. Solid dots denote significance after FDR correction. Dashed boxes highlight loci classified as vertical pleiotropy (*PICALM* and *EFTUD1* regions). Pleiotropy type for each region is labeled below in colored text. Key overlapping genes are annotated above each region. **(B) Decision tree flowchart categorizing the loci into five mechanistic classes based on MR significance (FDR<0.05)**: (1) Ambiguous bidirectional pleiotropy in yellow box (Chr11:47.4Mb *SPI1*, Chr17:5.1Mb *SCIMP*); (2) Forward & reverse causal effect in green box; (3) Vertical pleiotropy in red box (Chr11:85.7Mb *PICALM*, Chr15:82.4Mb *EFTUD1*) showing eGFR to AD pathway; (4) Horizontal pleiotropy in purple box (Chr6:47.7Mb *CD2AP*, Chr10:82.3Mb *MAT1A*, Chr19:46.3Mb *SYMPK*); (5) No clear causal effect in gray box. Arrows and decision nodes (pink diamonds, green ovals) guide classification logic. The flowchart was created using *BioRender* and included with permission for publication.

These mechanistic categories segregate pleiotropic loci into those where eGFR causally influences AD, those where AD might influence kidney health, bidirectional loops, and those reflecting horizontal pleiotropy or unresolved complexity. Notably, these classifications were derived exclusively for EUR loci where adequate power and resolution were available. None of the suggestive AFR loci met criteria for detailed categorization, reinforcing that much of the identified shared genetic architecture remains population specific. Regional association plots and *LocusZoom* visualizations for key loci are provided in **Supplementary Figs. S18-S26**.

## Discussion

To our knowledge, this study represents the first comprehensive, large-scale, multi-ancestry genome-wide investigation systematically characterizing the shared genetic architecture between kidney function and AD. We hypothesized that genetic connections between these traits would concentrate at specific genomic loci rather than manifest as diffuse polygenic overlap, and that this architecture would differ across ancestries. Using an integrated analytical framework combining conjFDR analysis, local genetic correlation, fine-mapping with colocalization, and locus-specific cis-MR, we confirm this hypothesis and demonstrate that kidney–brain genetic relationships are regionally constrained, mechanistically heterogeneous, and highly ancestry dependent.

Despite near-zero genome-wide r_g_ between eGFR and AD in EUR, we identified 16 distinct pleiotropic loci at conjFDR < 0.05 and 15 loci showing significant local genetic correlation using *LAVA*. Differences between conjFDR and *LAVA* results reflect complementary rather than conflicting views of shared genetic architecture. *ConjFDR* detects loci with joint association to both traits, even when individual variants exert effects in different directions, whereas *LAVA* summarizes net correlation of effects across regions. Consequently, loci with complex regulatory structure may exhibit strong *conjFDR* signals but weak or null local rg, underscoring the value of integrating both approaches. The absence of significant PRS association further supports a model in which shared genetic architecture between eGFR and AD arises from specific genomic loci rather than diffuse genome-wide polygenic overlap. These findings explain prior weak or null global genetic correlations despite robust epidemiological associations between kidney dysfunction and dementia: when the eGFR-decreasing allele increases AD risk at some loci but decreases it at others, global estimates obscure meaningful signal through averaging. This pattern also contextualizes apparently conflicting MR findings: studies by Huang *et al*. (2024)^77^, Zhao *et al*. (2024)^76^ reported associations between genetically predicted chronic kidney disease liability and AD risk, whereas Kjaergaard *et al*. (2022)^79^ and Liu *et al*. (2023)^78^ observed weaker or null effects when focusing on estimated glomerular filtration rate. Traditional genome-wide MR approaches aggregate instruments across loci with opposing effect directions, attenuating signals when effects cancel at the global level.

Fine-mapping and colocalization analyses further revealed that most pleiotropic loci harbor distinct causal variants influencing kidney function and AD risk, consistent with horizontal pleiotropy. The *APOE* locus represents a notable exception, where the ε4-defining variant showed shared causality driving both reduced eGFR and increased AD risk. This locus also exhibited evidence of reverse causality in cis-MR, with genetic liability for AD influencing eGFR, consistent with *APOE’*s established pleiotropic effects across lipid metabolism, vascular biology, and multiple organ systems. In contrast, loci such as *SPI1* demonstrated overlapping association signals driven by distinct nearby variants, suggesting modular regulatory architecture with tissue- or cell-type-specific effects. The *APOE* locus served as a positive control across multiple analytical layers, as it was the only region demonstrating robust local genetic correlation, strong colocalization with a shared causal variant, and consistent effects across ancestries.

Cis-Mendelian randomization enabled mechanistic stratification of pleiotropic loci, revealing directional heterogeneity among regions with significant causal evidence. Importantly, these locus-specific findings do not imply systemic mediation of AD risk by eGFR; rather, they indicate that genetic variants regulating kidney-related biology at specific loci exert parallel or partially overlapping effects on neurodegenerative processes. At the *EFTUD1* locus, genetically predicted reductions in eGFR were causally associated with increased AD risk, whereas the *PICALM* locus exhibited an opposing relationship, where genetic liability for lower eGFR is associated with decreased AD risk. This directional heterogeneity likely explains why genome-wide MR studies yield attenuated estimates. Most pleiotropic loci identified through conjFDR showed no significant causal effects in cis-MR, suggesting horizontal pleiotropy or shared upstream regulation rather than direct mediation.

At both loci, causal effects were driven by non-coding regulatory variants. The lead *PICALM* instrument rs145403697 lies within an intergenic enhancer region between *PICALM* and *SYTL2*, while the *EFTUD1* intronic variant rs112500920 exhibits strong regulatory annotations and prior associations with blood pressure traits. Expression data further support a kidney-first model: *PICALM* shows higher expression in kidney tubular cells and immune populations than in most brain cell types, and *EFTUD1* is more robustly expressed in kidney than brain tissue.^115,116^

The mechanistic basis for vertical pleiotropy at these loci involves distinct biological pathways. PICALM (phosphatidylinositol-binding clathrin assembly protein) orchestrates clathrin-mediated endocytosis in both kidney tubular epithelium and brain endothelium.^116,117^ In the brain, PICALM mediates amyloid-β clearance through LRP1-dependent transcytosis at the BBB and regulates tau clearance via SNARE-dependent autophagosome-lysosome fusion.^118^ In the kidney, PICALM supports proximal tubular reabsorption of filtered proteins and uremic solutes. Our findings suggest that kidney function-associated *PICALM* locus variants affect AD risk through impaired kidney clearance or diminishing systemic endocytic activity^115,116^, thereby creating a pro-amyloidogenic environment consistent with evidence that kidney dysfunction elevates circulating amyloid-β levels.^43,44^

At the *EFTUD1* locus, vertical pleiotropy reveals an unexpected connection between ribosome biogenesis and the kidney-brain axis. *EFTUD1* encodes a GTPase essential for 60S ribosomal subunit maturation; mutations cause Shwachman-Diamond syndrome type 2, underscoring its fundamental role in protein synthesis.^119^ Proximal tubular cells exhibit extraordinarily high protein synthesis demands, making them sensitive to ribosomal stress, while in the brain, aberrant ribosome biogenesis represents an early feature of AD and tauopathies.^119,120^ Tau directly impairs 60S subunit biogenesis, creating a pathogenic feedforward loop.^121^ Genetic variants affecting *EFTUD1* function may thus simultaneously compromise kidney tubular function and neuronal proteostatic resilience.

In contrast, loci exhibiting horizontal pleiotropy, including *CD2AP*, *MAT1A*, and *SYMPK*, influence kidney and brain through shared upstream pathways rather than direct causal mediation. At the *CD2AP* locus, fine-mapping identified rs7763746 as a candidate causal variant with a GERP score of 10.4 indicating strong evolutionary conservation. *CD2AP* plays essential roles in podocyte integrity in the kidney and in endocytic and blood–brain barrier regulation in the brain, with established links to focal segmental glomerulosclerosis and AD risk.^66^ We identified novel genetic variants in this locus that may help elucidate how *CD2AP* functions at the junction of nephropathy and AD, warranting functional investigation. Recent experimental studies demonstrated that *CD2AP* loss in brain endothelial cells causes BBB breakdown and cognitive deficits, providing biological support for our finding.^66^ *MAT1A* regulates S-adenosylmethionine synthesis, a central node in methylation and lipid metabolism affecting oxidative stress in both renal and neuronal tissues.^122^ *SYMPK* participates in mRNA processing and cell-cycle regulation across energy-demanding tissues. Collectively, these horizontally pleiotropic loci converge on membrane trafficking, methylation balance, and cellular stress resilience, which are processes fundamental to both organ systems but operating through shared systemic biological mechanisms.

Similarly, the *SPI1* locus (Chr11:47.4Mb) showed bidirectional effects in cis-MR, with variant rs10838702 residing in a CAGE-defined enhancer overlapping multiple super-enhancers. *SPI1* expression is highly enriched in myeloid lineages, including microglia and peripheral macrophages, with minimal expression in brain neurons. The *SPI1* (PU.1) transcription factor controls both microglial activation in brain and macrophage function in kidney, with risk-lowering alleles associated with better function in both organs, suggesting that genetically mediated immune regulation may influence both kidney and brain function through shared inflammatory pathways.^123,124^

Marked ancestry-specific differences further highlight the complexity of kidney-brain genetic architecture. With *APOE* region being the only overlap in significant loci, genetic risk factors for kidney-brain dysfunction vary substantially across populations. Given the substantially smaller sample size and lower h^2^_SNP_ estimates in AFR, ancestry-specific signals beyond the *APOE* locus should be interpreted as hypothesis-generating rather than definitive evidence of biological divergence. Local correlations showed opposite predominant directions between ancestries alongside distinct polygenic risk score findings, suggesting fundamentally different genetic architecture. The African-specific signal near *AGTR1* may partly explain the disproportionate burden of both chronic kidney disease and vascular cognitive impairment in AFR, given its central role in both kidney function regulation and cerebrovascular biology.^125^ Such heterogeneity has important implications for precision medicine and cautions against extrapolating genetic risk models across ancestries without appropriate validation.

These findings have important implications for genetic risk prediction and clinical translation. Because kidney-related contributions to AD risk are concentrated at specific genomic loci with heterogeneous effect directions, polygenic risk scores that aggregate eGFR-associated variants genome-wide may fail to capture this relationship or even obscure it when opposing effects cancel. Our results suggest that genetic liability arising from vertically pleiotropic loci such as *PICALM* and *EFTUD1*, validated across multiple complementary analytical approaches, represents a distinct component of AD risk operating through kidney-related biological pathways. In contrast, horizontally pleiotropic loci primarily reflect shared systemic processes such as inflammation, endocytosis, and vascular integrity. Given that some loci show protective and others detrimental effects of eGFR on AD risk, partitioned or pathway-informed polygenic scores that separately model directionally consistent or vertically pleiotropic loci may enhance predictive performance and clarify biological subtypes of risk. Moreover, with *APOE* as the only cross-ancestry signal identified, developing ancestry-specific or pathway-weighted genetic risk models will be essential to ensure equitable prediction and translation across populations.

These findings also have implications for interpreting blood-based AD biomarkers, where kidney function is routinely treated as a covariate. Kidney function can influence circulating biomarker concentrations through at least two non-exclusive mechanisms: impaired renal clearance elevating plasma levels independent of central nervous system pathology, and pleiotropic genetic pathways contributing to both kidney dysfunction and neurodegeneration. Routine adjustment for kidney function may therefore attenuate biologically meaningful signal rather than isolate measurement noise alone. Reporting both kidney-adjusted and unadjusted biomarker associations may clarify whether kidney-related variation reflects clearance effects or shared disease biology.

We acknowledge several limitations. First, eGFR captures only one dimension of kidney physiology; other clinically relevant aspects such as albuminuria or tubular injury could not be comprehensively evaluated due to limited GWAS sample sizes. Second, although we applied a competing risk correction to mitigate survival bias in AD GWAS summary statistics, residual bias may persist in late-onset disease GWAS contexts. Third, cis-MR is restricted to local genetic effects and cannot capture trans-regulatory mechanisms; while these approaches reduce susceptibility to horizontal pleiotropy, they cannot fully exclude shared regulatory architecture within complex loci. Fourth, our analyses were limited to common autosomal variants in predominantly EUR samples, potentially missing contributions from rare variants, structural variation, or sex chromosome loci, and AFR findings beyond *APOE* should be interpreted cautiously given reduced statistical power.

In summary, this study extends beyond conventional genome-wide genetic correlation approaches by implementing a multi-scale framework that reveals localized, ancestry-specific, and mechanistically diverse genetic connections between kidney function and AD. Future research should prioritize functional validation of identified pleiotropic loci using cellular and animal models, and integration of multi-omic data to clarify how genetic variants translate into cross-organ dysfunction. Longitudinal studies tracking kidney function trajectories alongside cognitive performance would help establish temporal relationships and identify critical windows for intervention. Evaluating whether polygenic risk scores incorporating these pleiotropic loci improve prediction of cognitive decline could facilitate clinical translation. As AD biomarkers move toward broader clinical use, kidney function should be considered in biomarker interpretation to ensure accurate and equitable translation. By revealing specific genetic components connecting peripheral organ function to neurodegeneration, this work reshapes how we conceptualize the kidney-brain axis and provides a foundation for experimental validation and mechanistic exploration.

## Data availability

All GWAS summary statistics used in this study are publicly available. AD summary statistics are available from Kunkle *et al.* (2019)^52^, Wightman *et al.* (2021)^85^, and Ray *et al.* (2024)^86^. eGFR summary statistics are available from Stanzick *et al*. (2021)^126^, Hughes *et al*. (2023)^93^, and the Million Veteran Program (Hellwege *et al*., 2019)^94^. ADGC individual-level data is available through authorized access following dbGaP procedures.^96^ All additional scripts and summary level data are available upon request. Complete methodological details, including instrumental variable selection, parameter specifications, and sensitivity analyses, are provided in the Supplementary Methods.

## Acknowledgements

We sincerely thank the participants whose genetic data contributed to the GWAS studies used in this research, as well as the investigators who generated, curated, and shared these valuable resources. We are especially grateful to the Alzheimer’s Disease Genetics Consortium for their leadership and commitment to data sharing, which made this project possible.

## Funding

This study was supported in part by the National Institute on Aging (NIA) of the National Institutes of Health under grant numbers T32 AG071474 (Bush) and U01 AG032984 (Schellenberg). Y.Y, M.L. and X.Z. were supported by HG011052 (Zhu) and HG011052-03S1 (Zhu). We also acknowledge the use of computational resources provided by the Case Western Reserve University High Performance Computing Center (HPCC), which played a vital role in enabling the large-scale genomic analyses presented in this study.

## Competing interests

The authors report no competing interests.

## Supplementary material

Supplementary material is provided as a separate file.

## Notes

### Competing Interest Statement

The authors have declared no competing interest.

### Author Declarations

IRB of University Hospitals Cleveland Medical Center (IRB number EM-14-02, PI: Jonathan Haines) gave ethical approval for this work. All publicly available GWAS summary statistics utilized were approved by relevant ethics committees in their respective publications, and ADGC individual-level data were de-identified and accessed under approved protocols. ADGC participants provided written informed consent, as detailed in the Supplementary Methods.

