## Supplementary Material for "Shared Genetic Architecture Between Kidney Function and Alzheimer Disease Across Ancestries"

### Supplementary Methods

#### GWAS summary statistics

Single Nucleotide Polymorphisms (SNPs) on the X, Y, and within the MHC region (chr6:25,119,106-33,854,733, hg19) were excluded from all analyses to mitigate potential bias arising from the complex linkage disequilibrium (LD) patterns. All the GWAS utilized in this study were approved by relevant ethics committees, with detailed genotyping and phenotype data processing details documented in the respective publications. To accurately estimate statistical power, control Type I error rate, and maximize efficiency, effective sample size for each cohort was calculated as $n_{eff} = 4/(1/n_{cases} + 1/n_{controls})$ to account for imbalances in the case/control ratio. GWAS Manhattan plots were generated using the *topr* package in R v4.3.1.

##### AD GWAS datasets

For AD in EUR, we obtained GWAS summary statistics from Kunkle *et al.* stage 1 with clinically adjudicated AD (n = 63,926) and from Wightman *et al.* with clinically diagnosed AD (n = 398,058), excluding 23andMe and UK Biobank AD proxies, provided by the authors upon request. For AD in AFR, we used summary statistics from Ray *et al.* model 1 without APOE adjustment with clinically adjudicated AD (n = 9,168).

##### AD GWAS Corrected for Competing Risk

Late-onset AD genetic studies are susceptible to competing risk bias because individuals with genetic liability to conditions causing earlier mortality (cardiovascular disease, kidney failure, cancer) are underrepresented in elderly AD cohorts. If kidney disease genetic variants increase mortality risk, standard AD GWAS will underestimate or fail to detect true genetic overlap between kidney function and AD. We implemented a correction for participation bias in AD GWAS following the collider bias correction approach described by Wang *et al.* (2024). Competing risk occurs when individuals with certain conditions die or become unable to participate before AD onset, creating selection bias that distorts genetic associations. This is a form of collider bias where survival to study participation acts as a collider between genetic variants, competing risks, and AD. Correcting for this bias is critical because participation bias can significantly distort genetic associations and compromise downstream analyses, particularly genetic correlation and Mendelian randomization (MR) analyses that rely on accurate SNP effect size estimates.

This selection bias can be formally conceptualized within a causal framework. Let S denote survival to recruitment or study participation. If both genetic liability and competing diseases influence S, then conditioning on S, which occurs implicitly when analyzing GWAS data restricted to recruited and surviving individuals, induces a non-causal association between genetic variants and those competing diseases, even if they are independent in the underlying population. In a directed acyclic graph (DAG), S functions as a collider on the path from genetic variants to S and from competing disease to S. Conditioning on this collider opens a backdoor path and generates spurious associations (**Supplementary Figure S28**). In the context of AD, several conditions that compete with AD for survival may reduce the probability of surviving to the age at which AD is typically diagnosed. Individuals affected by these conditions are therefore less likely to be included among both AD cases and controls. As a result, genetic variants predisposing to these diseases may appear to have protective effects on AD risk due to differential survival rather than true biological mechanisms.

To mitigate this survival-induced collider bias in the AD GWAS summary statistics, we applied a multivariable Mendelian randomization (MVMR)-based correction strategy (Wang et al., 2024). We first conducted univariable Mendelian randomization (UVMR) using MRBEE to evaluate the apparent causal effects of candidate competing diseases on AD. Diseases showing negative effect estimates on AD risk, a pattern consistent with selection bias rather than genuine protective effects, were then incorporated as covariates in an MVMR model. We considered only terminal diseases as exposures and did not include common exposures such as BMI. We then applied the genome-wide pleiotropy test within the MRBEE framework to obtain debiased AD GWAS summary statistics, thereby removing the component of SNP-AD association attributable to genetic pathways operating through competing diseases that influence survival or study participation.

We selected five diseases with known epidemiological associations with mortality or reduced study participation as competing exposures: coronary artery disease (CAD; Aragam *et al.*, 2022), heart failure (HF; Rasooly *et al.*, 2023), kidney cancer (Purdue *et al.*, 2024), lung cancer (Gorman *et al.*, 2024), and type 1 diabetes (T1D; Chiou *et al.*, 2021), with AD as the outcome. These conditions may lead to death or study exclusion before AD onset, potentially biasing the observed genetic associations with AD. Notably, we did not apply this correction to the kidney function GWAS, as the study population for eGFR has a substantially younger age distribution compared to AD cohorts, making competing risk from these age-related conditions less relevant.

Instrumental variables (IVs) for each exposure were selected using PLINK with the following clumping parameters: --clump-kb 1000, --clump-p1 5E-8, --clump-p2 5E-8, and --clump-r2 0.01. The LD reference panel consisted of 9,680 individuals of genetically European ancestry from the UK Biobank (46% male, 54% female). We restricted our analyses to common variants in the European population (minor allele frequency > 0.01), as defined by variants passing quality control conducted by the Neale lab.

We applied *MRBEE* (Mendelian Randomization Bias and Efficiency Estimation) to estimate the causal effects of the five diseases on AD using the selected instrumental variables. For multivariable MR, each instrument was required to show strong association (F-statistic > 10) with at least one exposure to ensure instrument strength. To construct bias-corrected AD summary statistics, we performed a genome-wide residualization process. Specifically, for each variant across the entire genome, we modelled its reported AD effect size (beta coefficient) as the response variable, with the corresponding effect sizes for the five competing diseases as covariates in a linear model. Using the causal effect parameters (beta coefficients representing the causal impact of each disease on AD) estimated from the *MRBEE* analysis, we predicted the expected AD effect size for each variant based on its associations with the five competing diseases. The bias-corrected effect size was computed as the residual: the difference between the observed AD effect size and the predicted value (residual = observed AD beta - predicted AD beta). This residualization was performed genome-wide across all variants, not restricted to the instrumental variables used in the *MRBEE* analysis, thereby generating bias-corrected AD effect sizes that account for competing risk from the five selected diseases.

#### eGFR GWAS datasets

For eGFR in EUR, we conducted fixed-effects meta-analysis using *Stouffer's* method (implemented in *METAL*) combining data from three sources: (1) Million Veteran Program (MVP) from Hellwege et al., (2) Chronic Kidney Disease Genetics (CKDGen) Consortium data, and (3) UK Biobank (UKB) data from Stanzick et al. (n = 1,482,631). Stouffer's method was employed because different eGFR transformation approaches across resources produced effect estimates on different scales. SNVs not reported in at least one resource were excluded to ensure cross-study signal replication. Allelic effects were aligned to the eGFR-decreasing allele. For eGFR in AFR, we conducted fixed-effects meta-analysis by aggregating association summary statistics from the multi-ancestry GWAS meta-analysis by Hughes et al. (2023), which included individuals of African and admixed American (AMS) ancestry, with those from two additional resources: (i) an African American GWAS of 57,336 individuals from the MVP, and (ii) a meta-analysis of AMS GWAS comprising 5,010 individuals from a prior COGENT-Kidney Consortium publication. This approach was necessary due to limited sample sizes in African ancestry populations.

### Individual-level Data from ADGC

For the PRS analysis, we employed genotype and phenotype data from the Alzheimer's Disease Genetics Consortium (ADGC). This dataset comprised 17,781 healthy controls and 15,706 individuals with AD across both ancestry groups. The inclusion criteria for all individuals were (1) non-Hispanic European ancestry and African American ancestry (2) complete genetic and phenotype data, and (3) genetically unrelated to each other. Following these criteria, our final EUR sample included 15,063 individuals with AD (mean [SD] age 73.41 [8.49] years, 57.3% female) and 16,252 healthy controls (mean [SD] age 76.35 [8.25] years, 59.7% female). In the AFR sample, we included 643 individuals with AD (mean [SD] age 74.12 [7.79] years, 71.5% female) and 1,529 healthy controls (mean [SD] age 72.54 [7.87] years, 77.6% female). APOE ε4 dosage distributions differed significantly between cases and controls in both ancestry groups ($p < 0.001$), with AD cases showing higher frequencies of ε4 carriers as expected. All participants in the ADGC provided their written informed consent.

### Genome-wide level genetic analyses

#### Genetic correlation estimation using *LDSC*

To estimate the genetic correlation between eGFR and AD, we applied the LD score regression (*LDSC*) method. *LDSC* works by regressing GWAS summary statistics for variants across the genome on each variant's LD score, which reflects its ability to tag nearby variants through linkage disequilibrium. This approach infers the posterior mean effect size of each marker by conditioning on a specified prior genetic architecture and LD information derived from European and African ancestry reference panels in the 1000 Genomes Project (Phase 3). We restricted analyses to *HapMap3* SNPs to ensure data quality and comparability across studies. We used the GWAS summary-level data described above to estimate genetic correlations between eGFR and clinically adjudicated AD phenotypes in non-Hispanic and African ancestry populations. Manhattan plots and other GWAS association results were generated using the *topr* package in R 4.3.1. Genome-wide significant loci were then mapped to genes using the *FUMA* web platform. Importantly, LDSC relies solely on GWAS summary statistics and can provide estimates of genetic correlations (rg) while accounting for population stratification and cryptic relatedness. However, sample overlap between studies can introduce bias in genetic correlation estimates, which can be assessed using the cross-trait LDSC intercept.

#### Polygenic risk score analysis

Polygenic risk score analysis was conducted using *SBayesRC* toolbox (v0.2.6). After removing duplicate and ambiguous SNPs, we applied quality control excluding variants with MAF < 0.01 and related individuals (kinship coefficient > 0.0442). This yielded 4,992,903 and 4,580,818 SNPs for PRS calculation in 31,315 EUR and 2,172 AFR samples, respectively. We computed SNP weights and calculated PRS for ADGC individuals using eGFR GWAS summary statistics. This Bayesian method integrates functional genomic annotations with GWAS effect sizes to improve SNP effect estimation by distinguishing likely causal from non-causal SNPs in high LD regions. We analyzed imputed common SNPs using a hierarchical multicomponent mixture prior incorporating annotation information via a full Bayesian learning framework, utilizing functional genomic annotations for approximately 8 million SNPs provided by the authors. We used the Baseline LD v2.2 annotation model (96 annotations; Márquez-Luna *et al*., 2021) with LD reference panels from UK Biobank and LifeLines (~7M SNPs for EUR, ~6M for AFR overlapping with BaselineLD v2.2) to estimate genome-wide SNP effects. Final PRS was calculated using *PLINK* 2.0's --score command and standardized to mean 0, variance 1. Multivariate logistic regression models estimated associations between eGFR-based PRS and AD diagnosis, adjusting for sex, age, and three genetic principal components. Analyses were performed with and without APOE ε4 dosage adjustment to assess independent PRS contribution.

#### Conjunctional false discovery rate analysis

To assess local genetic overlap and identify shared genetic loci between AD and eGFR, we used the conjunctional false discovery rate (*conjFDR*) method from the *pleioFDR* toolbox. The *conjFDR* approach aims to boost statistical power for discovering shared genetic variants between two phenotypes by leveraging cross-trait enrichment patterns. This method is defined as the posterior probability that a SNP is null for either phenotype or both simultaneously, given that its p-values for association with both phenotypes are as small as or smaller than the observed p-values. The *conjFDR* approach performs two conditional FDR (*condFDR*) analyses, first conditioning the primary phenotype (e.g., eGFR) on SNP associations with the secondary phenotype (e.g., AD), and vice versa. The final *conjFDR* value is defined as the maximum of these two *condFDR* statistics, representing an estimate of the posterior probability that a given SNP is not associated with either trait under the condition that the p-values for both traits are as small or smaller than observed.

Cross-trait enrichment was visualized using conditional quantile-quantile (*Q-Q*) plots, which display the distribution of p-values for a primary phenotype across all SNPs and for SNP strata defined by their association strength with the secondary phenotype. Specifically, we computed the empirical cumulative distribution of nominal p-values for all SNPs and for SNPs meeting significance thresholds below indicated cut-offs ($p < 0.1$, $p < 0.01$, $p < 0.001$) for the conditional phenotypes. Nominal p-values -log10(p) values were plotted against the empirical cumulative distribution function. Enrichment was evidenced by leftward deflections in conditional Q-Q plots with increasing auxiliary phenotype association strength, directly interpretable as true discovery rate (1-FDR). We applied a conjFDR threshold of 0.05 following established methods to ensure adequate power.

#### Pleiotropic genomic loci definition and functional annotation

Independent genomic loci were defined following the *FUMA* protocol to identify pleiotropic regions associated with both phenotypes. Independent significant SNPs were first identified using stringent criteria of linkage disequilibrium $r^2 < 0.60$ and *conjFDR* < 0.05. Among these independent significant SNPs, those with $r^2 < 0.1$ were considered to be in approximate linkage equilibrium and designated as lead SNPs representing distinct genomic loci. Candidate SNPs were subsequently defined as variants with *conjFDR* values < 0.05 and linkage disequilibrium $r^2 > 0.60$ with any independent significant SNP, thereby capturing the full spectrum of potentially causal variants within each locus. Genomic loci were refined through a merging process where all independent loci located within 250 kb of each other were combined into single larger loci, with the SNP showing the most significant conjFDR value selected as the lead SNP for the merged region. The boundaries of each locus were precisely defined by identifying all candidate SNPs in linkage disequilibrium ($r^2 \geq 0.6$) with any of the independent significant SNPs within that locus. For downstream analyses, each final locus was further extended to include regions 500 kb upstream and downstream of the lead SNP, ensuring comprehensive coverage of regulatory elements and potential long-range effects. All linkage disequilibrium $r^2$ values were calculated using the 1000 Genomes Project European-ancestry and African-ancestry haplotype reference panels, ensuring population-appropriate linkage disequilibrium estimates for downstream functional interpretation and prioritization of potentially causal variants within identified pleiotropic loci.

### Locus-level genetic analyses

#### Local genetic correlations using LAVA

We used the R package *LAVA* to assess bivariate local genetic correlations between AD and eGFR. For this analysis, we implemented approximately equal-sized (~1 Mb), semi-independent genomic blocks as defined by *LAVA* authors. These blocks partition the European genome into 2,495 semi-independent LD blocks using parameters in the *LAVA* blocking algorithm, with a minimum block size of 2,500 base pairs. To account for any potential sample overlap in the input summary statistics, we supplied the intercepts from the bivariate cross-trait *LDSC* analyses to *LAVA* as these represent an estimate of the sampling correlation between the datasets. We used the 1000 Genomes Project Phase 3 European/African reference panels (GRCh37/hg19) as the LD references for these analyses. To quantify the amount of local genetic correlation for each pair of phenotypes and remove loci without associations, we first tested the local joint univariate association signal for each phenotype. Univariate analyses were conducted using *LAVA* to assess SNP-based heritability ($h^2_{SNP}$) at the level of each individual genomic region for each phenotype. We selected only those genomic regions exhibiting significant local $h^2_{SNP}$ values (univariate p-value < 1E-4) for the subsequent bivariate analyses. Significant bivariate loci were identified using a Benjamini-Hochberg (BH) false discovery rate (FDR) threshold of less than 0.05 to account for multiple testing. For the AFR genome, we partitioned it into 2,687 semi-independent LD blocks using a minimum block size of 4,000 base pairs, following a similar approach to the one used by the authors for the EUR genome. We then applied the same workflow to evaluate local genetic correlations between AD and eGFR in the AFR population.

#### Cross-trait colocalization analysis

To test whether AD and eGFR traits share causal genetic variants, we performed cross-trait colocalization analysis using the R *coloc* package (v.5.2.3) with Sum of Single Effects

(*SuSiE*) frameworks to accommodate loci with multiple causal variants, utilizing LD matrices from the 1000 Genomes Project Phase 3. The traditional *coloc.abf* method assumes each trait has at most one causal signal per locus, while the *SuSiE* extension uses fine-mapping to perform more accurate inference when multiple independent associations exist for the same trait. We set prior probabilities for SNP associations as follows: $P_1$=1E-4 (probability of association only with trait 1), $P_2$=1E-4 (probability of association only with trait 2), and $P_{12}$=5E-6 (probability of association with both traits, using a more conservative value than the default 1E-5 to reduce false positives). The Bayesian colocalization analysis evaluated support for five mutually exclusive hypotheses: $H_0$ (no association with either trait), $H_1$ (association only with trait 1), $H_2$ (association only with trait 2), $H_3$ (association with both traits but driven by distinct causal variants in linkage disequilibrium), and $H_4$ (association with both traits sharing the same causal variant). For pleiotropic genomic loci identified through *conjFDR* analysis (conjFDR <0.05), we used colocalization to determine whether associations were driven by shared or distinct genetic variants. We selected genetically correlated loci only if the cumulative posterior probabilities for hypotheses $H_3$ and $H_4$ exceeded 0.8, ensuring substantial confidence that identified loci host at least one causal variant for either or both traits. Additionally, we imposed a ratio constraint requiring $PP.H_4$ to be at least five-fold greater than $PP\_H_3$, enriching our selection for genomic loci where evidence supports shared causal variants influencing both traits rather than distinct causal variants for each trait. After identifying loci meeting our pre-established criteria ($PP\_H_3+PP\_H_4 \geq 0.8$ and $PP\_H_4/PP\_H_3 \geq 5$), we pinpointed the most likely causal variants within each locus by identifying variants with the highest posterior inclusion probabilities (PIP) from *SuSiE* fine-mapping results and examined their functional annotations to prioritize biologically relevant candidates shared between AD and eGFR. This analysis was only performed in EUR due to the limited statistical power in AFR datasets.

#### Cis-Mendelian randomization analysis

We employed the *cis-MRBEE* (cis-Mendelian randomization bias correction estimating equation) method to evaluate potential causal effects of eGFR on AD risk at pleiotropic genomic loci identified through conjFDR analysis. *Cis-MRBEE* advances traditional cis-MR approaches by addressing weak instrument bias from genetic effect size estimation errors and accounting for sparse local genetic architecture. It uses a two-stage approach: fine-mapping to identify informative variants, then sparse prediction to re-estimate genetic associations using only selected variants, substantially reducing estimation errors. *Cis-MRBEE* then employs double-penalized minimization to simultaneously estimate causal effects and identify horizontal pleiotropy, using *SuSiE* for exposure selection and MCP (Minimax Concave Penalty) for pleiotropy detection.

For each locus identified from conjFDR, we applied the *CisMRBEE_UV* function with the following key parameters: xQTL sample size set to the mean eGFR meta-analysis sample size, maximum number of credible sets (xQTL.max.L = 4), posterior inclusion probability threshold (xQTL.pip.thres = 0.5), minimum PIP for credible set purification (xQTL.pip.min = 0.2), tuning parameter vector for direct causal effect penalization (tauvec = seq(0.5, 2, by = 0.1)), and ADMM algorithm parameter (admm.rho = 1). Prior to analysis, genetic variants were clumped using PLINK with $r^2 < 0.64$ and $p < 1E-3$ to remove highly correlated variants while preserving informative proxies for causal variants. The correlation matrix between exposure and outcome estimation errors (Rxy) was set to account for potential sample overlap, with minimal correlation (0.01) used between eGFR and AD studies. We then validated findings by comparing *cis-MRBEE* estimates with those from the *MAPLE* cis-MR package, strengthening evidence where methods converged. Statistical significance was assessed using bias-corrected estimating equations. This approach provides accurate causal effect estimation, better type I error control with small effect sizes, and distinguishes direct causal pathways from horizontal pleiotropy. This analysis was only performed in EUR due to limited statistical power in AFR datasets.

### Supplementary Figures

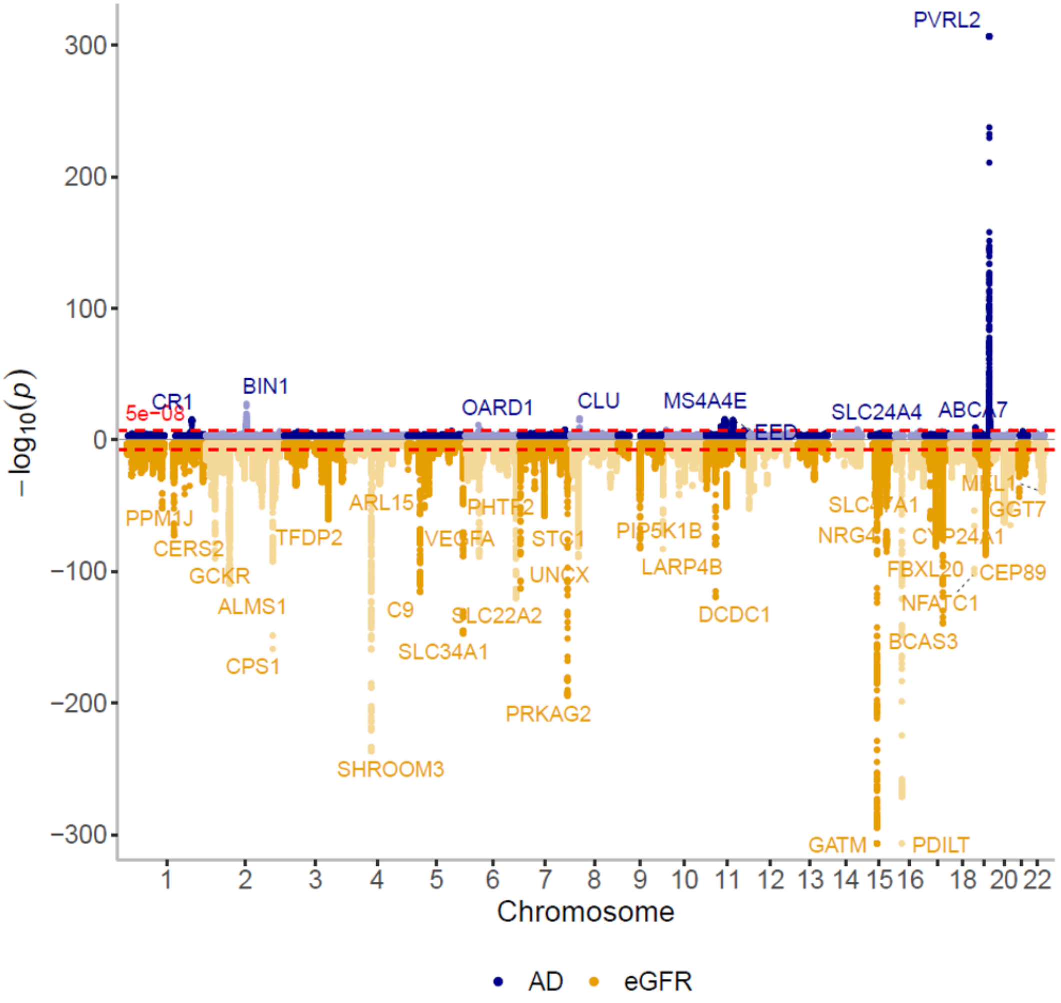

**Supplementary Figure S1. Miami Plot of Genome-Wide Association Results for Alzheimer's Disease (Discovery Dataset: Kunkle Corrected) and eGFR in European Ancestry.** Miami plot displaying genome-wide association study (GWAS) results for Alzheimer's disease (AD; discovery dataset using Kunkle data corrected for competing risks) and estimated glomerular filtration rate based on creatinine (eGFR) in individuals of European ancestry. The upper panel shows -log10(P-values) for AD associations across all chromosomes, while the lower panel displays -log10(P-values) for eGFR associations. Each point represents a single nucleotide polymorphism (SNP), with chromosomal positions indicated on the x-axis. The horizontal dashed line indicates the genome-wide significance threshold (P < 5E-8).

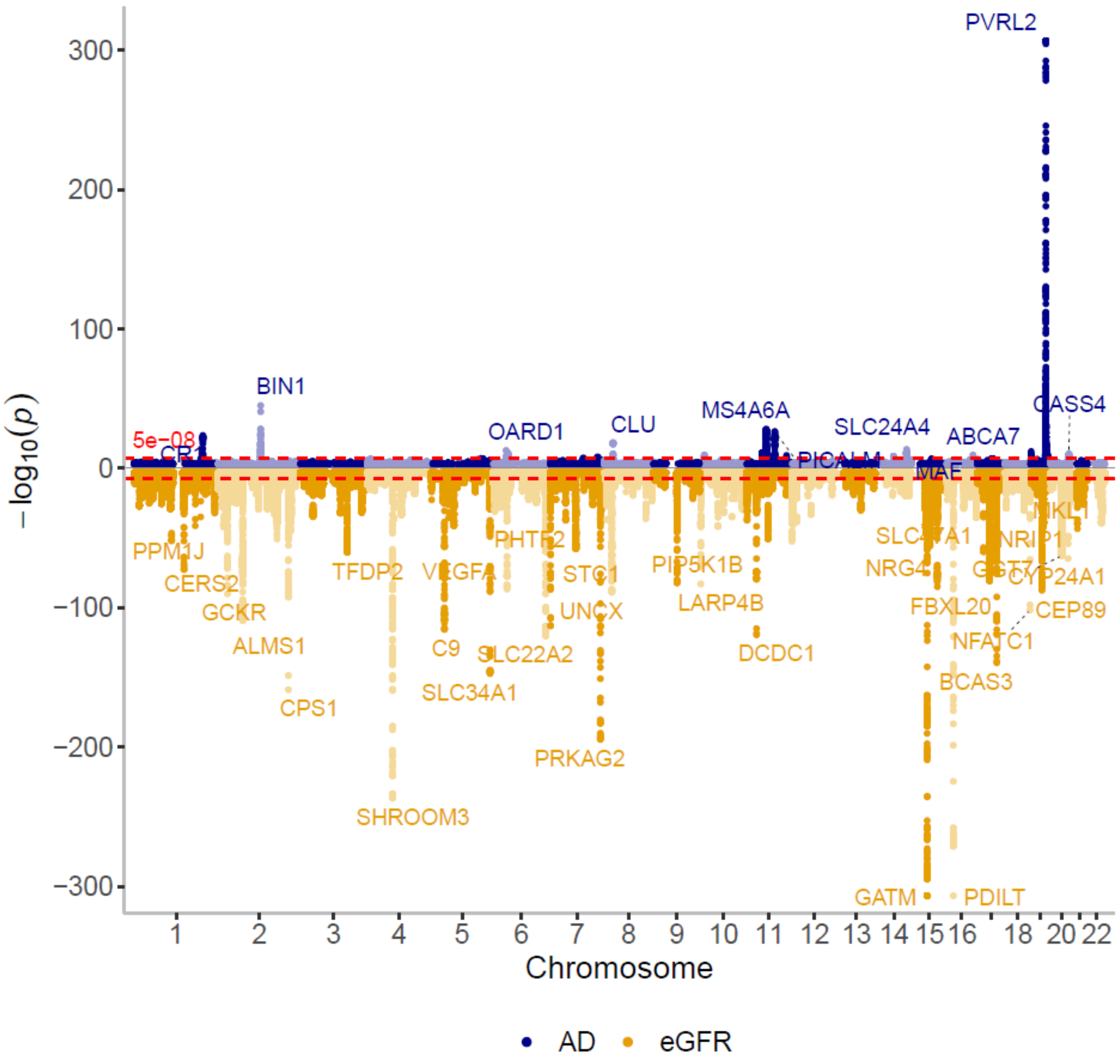

**Supplementary Figure S2. Miami Plot of Genome-Wide Association Results for Alzheimer's Disease (Validation Dataset: Wightman Corrected) and eGFR in European Ancestry**. This plot represents GWAS results for AD (validation dataset using Wightman data corrected for competing risk) and eGFR in European ancestry individuals. The plot displays -log10(P-values) for association with both traits across the genome, with each point representing a SNP colored by chromosome. The x-axis shows chromosomal positions (chromosomes 1-22), and the y-axis shows the strength of genetic association. The horizontal red line indicates the genome-wide significance threshold (P < 5E10-8).

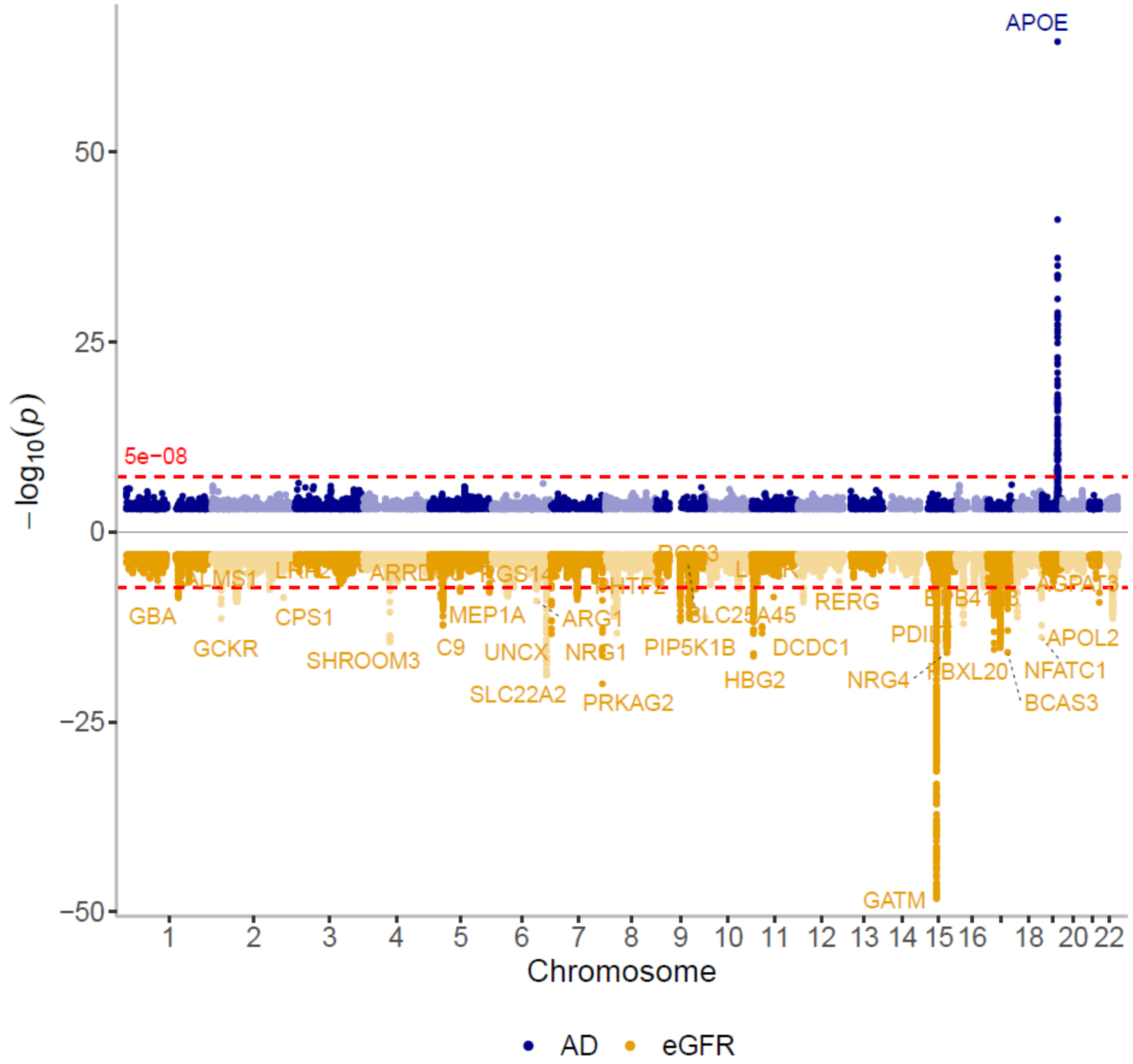

**Supplementary Figure S3. Miami Plot of Genome-Wide Association Results for Alzheimer's Disease and eGFR in African American Ancestry.** This plot displays GWAS results for AD and eGFR in African American ancestry individuals. The plot shows -log10(P-values) across all autosomes, with each point representing a SNP. Colors alternate by chromosome for visual clarity. The horizontal dashed line indicates genome-wide significance (P < 5E-8).

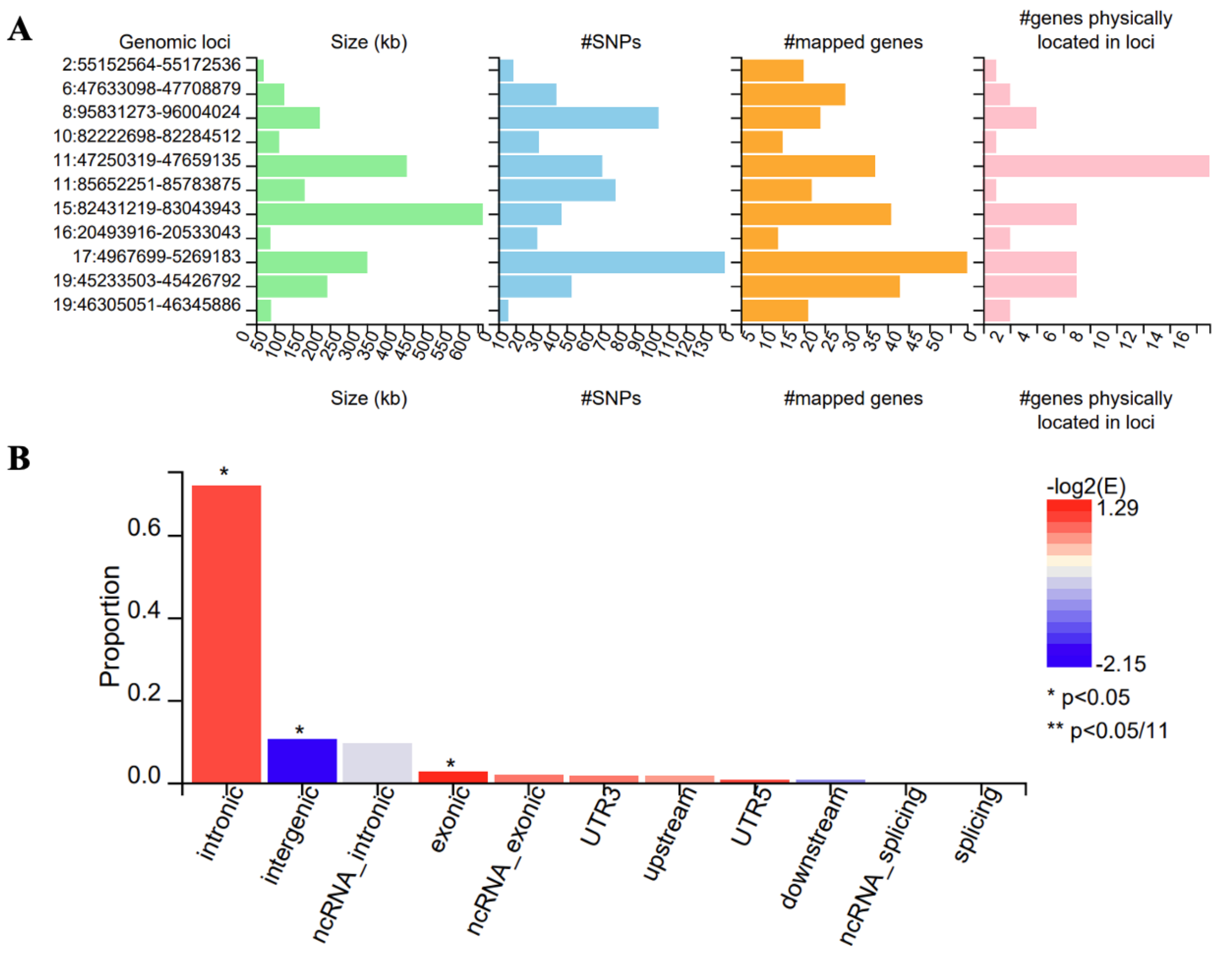

**Supplementary Figure S4. FUMA Annotation of Pleiotropic SNPs Identified by ConjFDR Analysis for Alzheimer's Disease (Discovery Dataset: Kunkle Corrected) in European Ancestry for Loci Definition**. This figure summarizes FUMA analysis of AD GWAS data from Kunkle et al. (corrected), including lead SNPs, genomic risk loci, gene mapping, and tissue expression enrichment. Results highlight prioritized genes and pathways related to AD. Complete pleiotropic loci from conjFDR analysis are in Supplementary Tables S19 and S20.

A

B

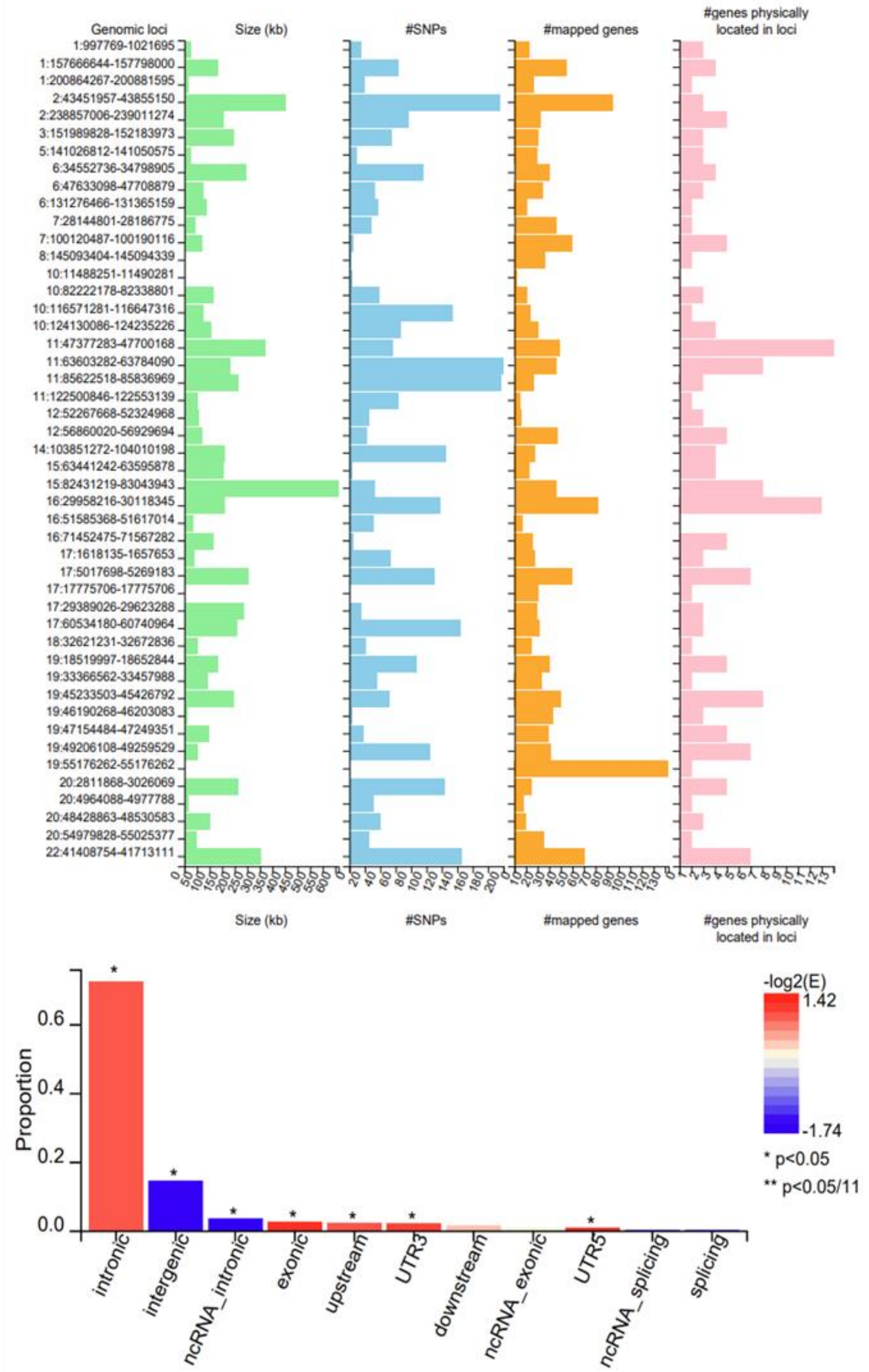

**Supplementary Figure S5. FUMA Annotation of Pleiotropic Loci Identified by ConjFDR Analysis for Alzheimer's Disease (Validation Dataset: Wightman Corrected) in European Ancestry for Loci Definition.**

A

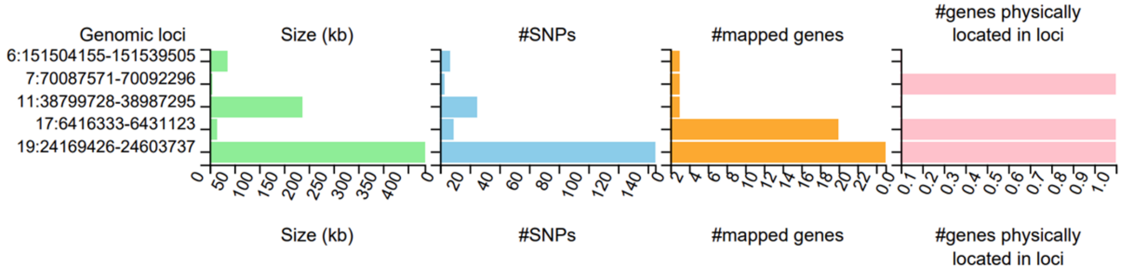

B

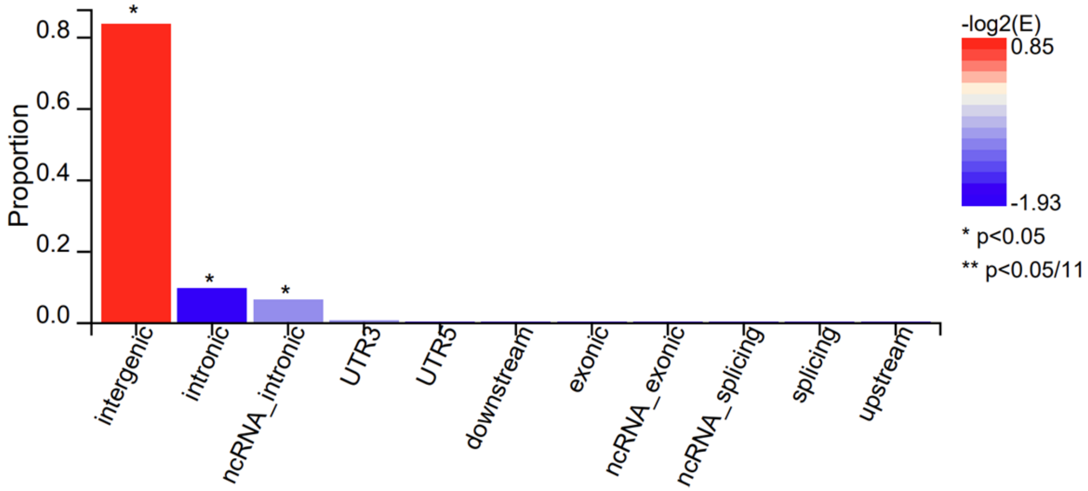

**Supplementary Figure S6. FUMA Annotation of Pleiotropic Loci Identified by ConjFDR Analysis for Alzheimer's Disease in African American Ancestry for Loci Definition.**

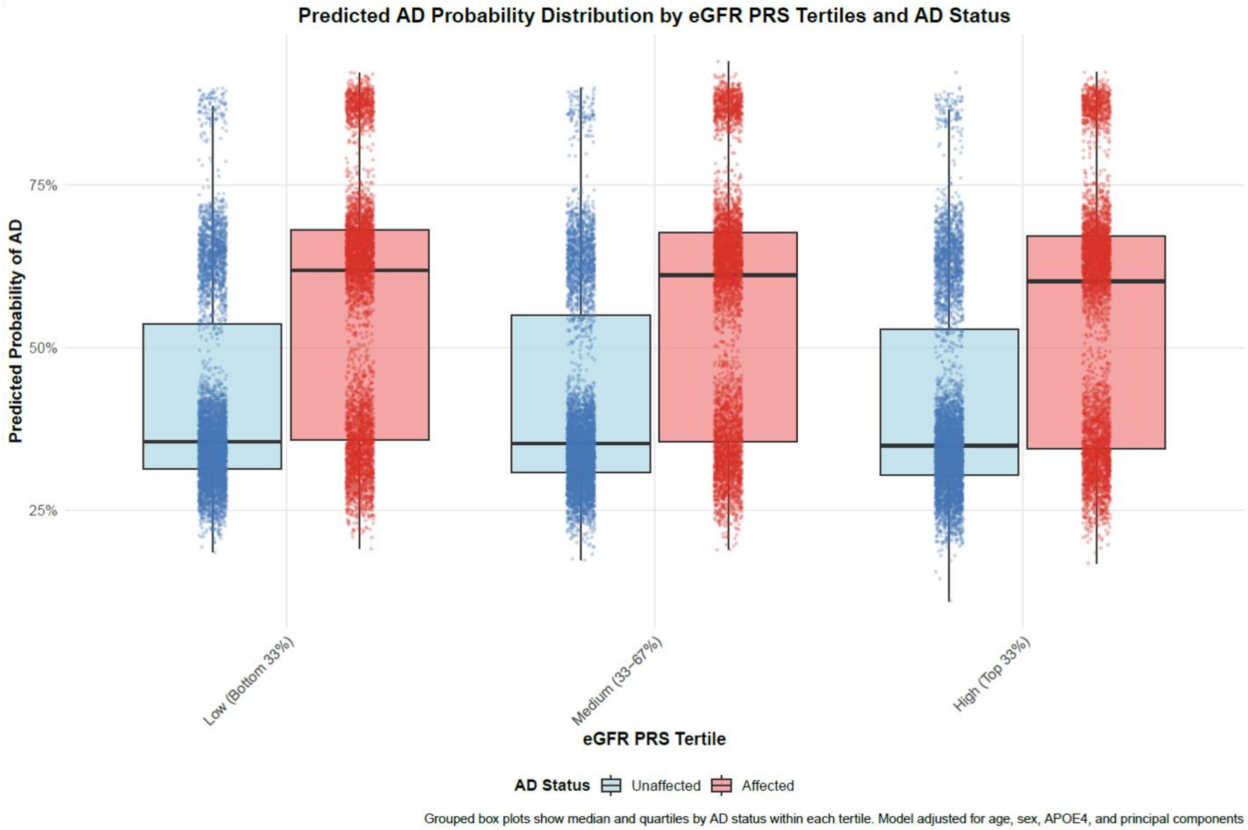

**Supplementary Figure S7. Predicted Probability of Alzheimer's Disease by eGFR Polygenic Risk Score Tertiles in European Ancestry (With APOE ε4 Dosage Adjustment).** Box plots displaying predicted probability of AD across eGFR polygenic risk score (PRS) tertiles in European ancestry individuals (N=31,315), adjusted for APOE ε4 dosage, age, sex, and the first three principal components of genetic ancestry. The x-axis shows eGFR-PRS tertiles (low, medium, high genetic predisposition for reduced kidney function), and the y-axis shows predicted probability of AD from logistic regression models. Each box represents the interquartile range (IQR), with the median shown as a horizontal line within each box. Whiskers extend to 1.5 times IQR. Individual data points may be overlaid. This figure illustrates whether genetic liability for reduced kidney function, as captured by eGFR-PRS, is associated with AD risk after accounting for the APOE ε4 allele.

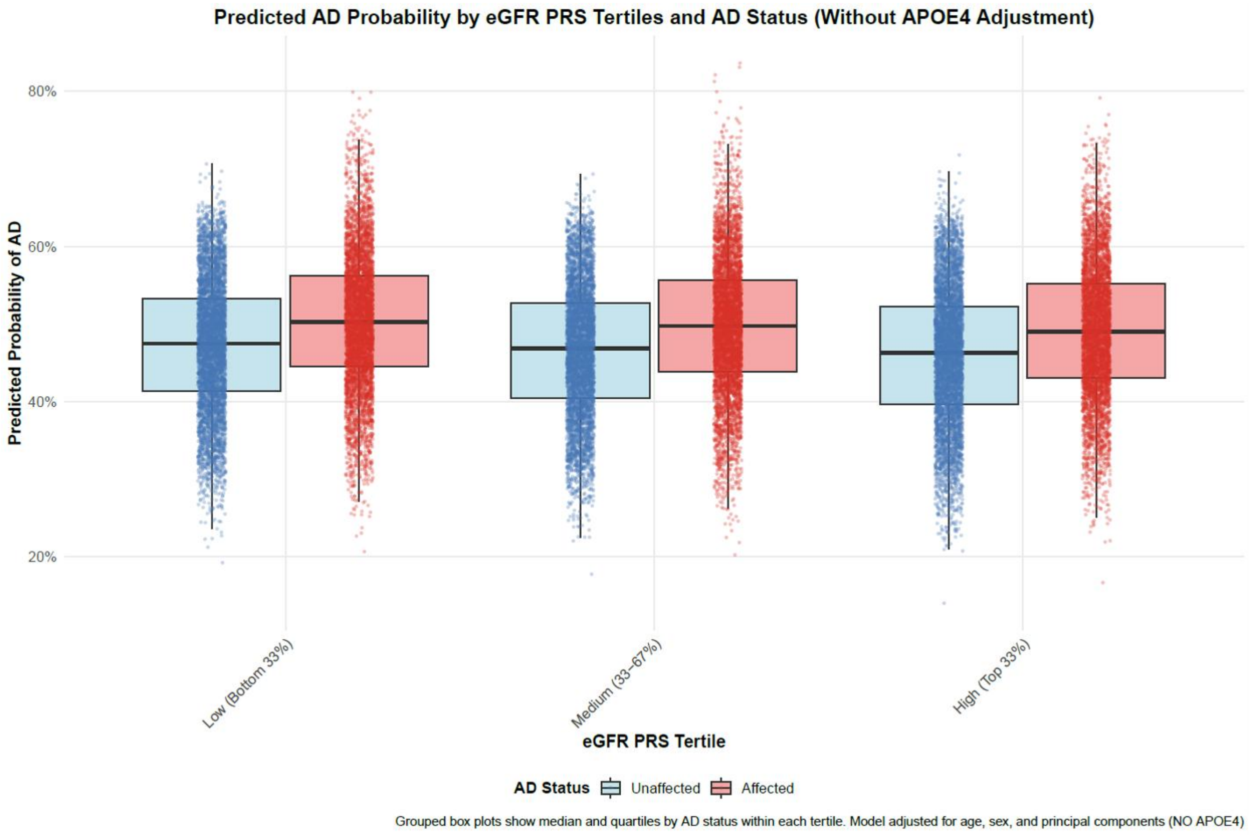

**Supplementary Figure S8. Predicted Probability of Alzheimer's Disease by eGFR Polygenic Risk Score Tertiles in European Ancestry (Without APOE ε4 Adjustment).** Box plots showing predicted probability of AD across eGFR-PRS tertiles in European ancestry individuals, without adjustment for APOE ε4 dosage. Models are adjusted for age, sex, and the first three principal components of genetic ancestry. The figure displays the distribution of predicted AD probability within each PRS tertile group.

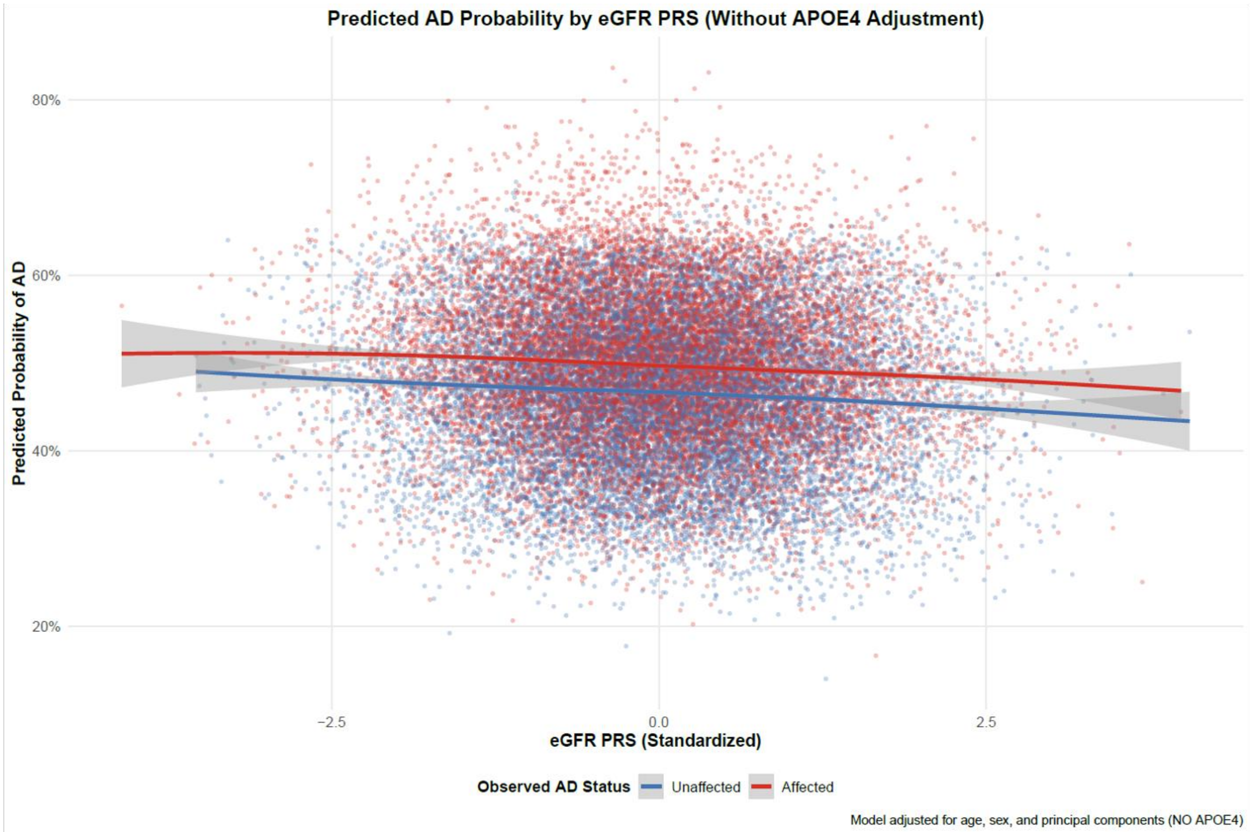

**Supplementary Figure S9. Predicted Probability of Alzheimer's Disease by eGFR Polygenic Risk Score in European Ancestry (Without APOE ε4 Adjustment).** This shows the predicted AD probability by eGFR-PRS in European ancestry, without APOE ε4 dosage adjustment.

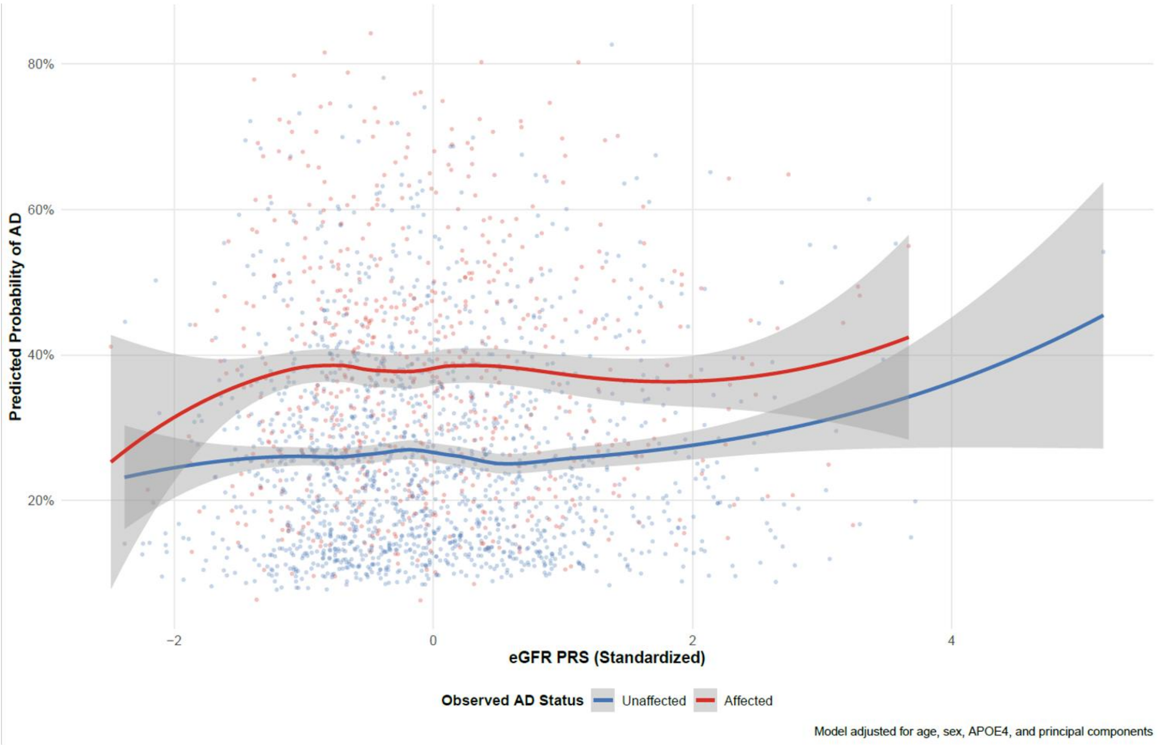

**Supplementary Figure S10. Predicted Probability of Alzheimer's Disease by eGFR Polygenic Risk Score in African American Ancestry (With APOE ε4 Adjustment).** Predicted AD probability across eGFR-PRS categories in African American ancestry individuals, adjusted for APOE ε4 dosage, age, sex, and principal components of genetic ancestry.

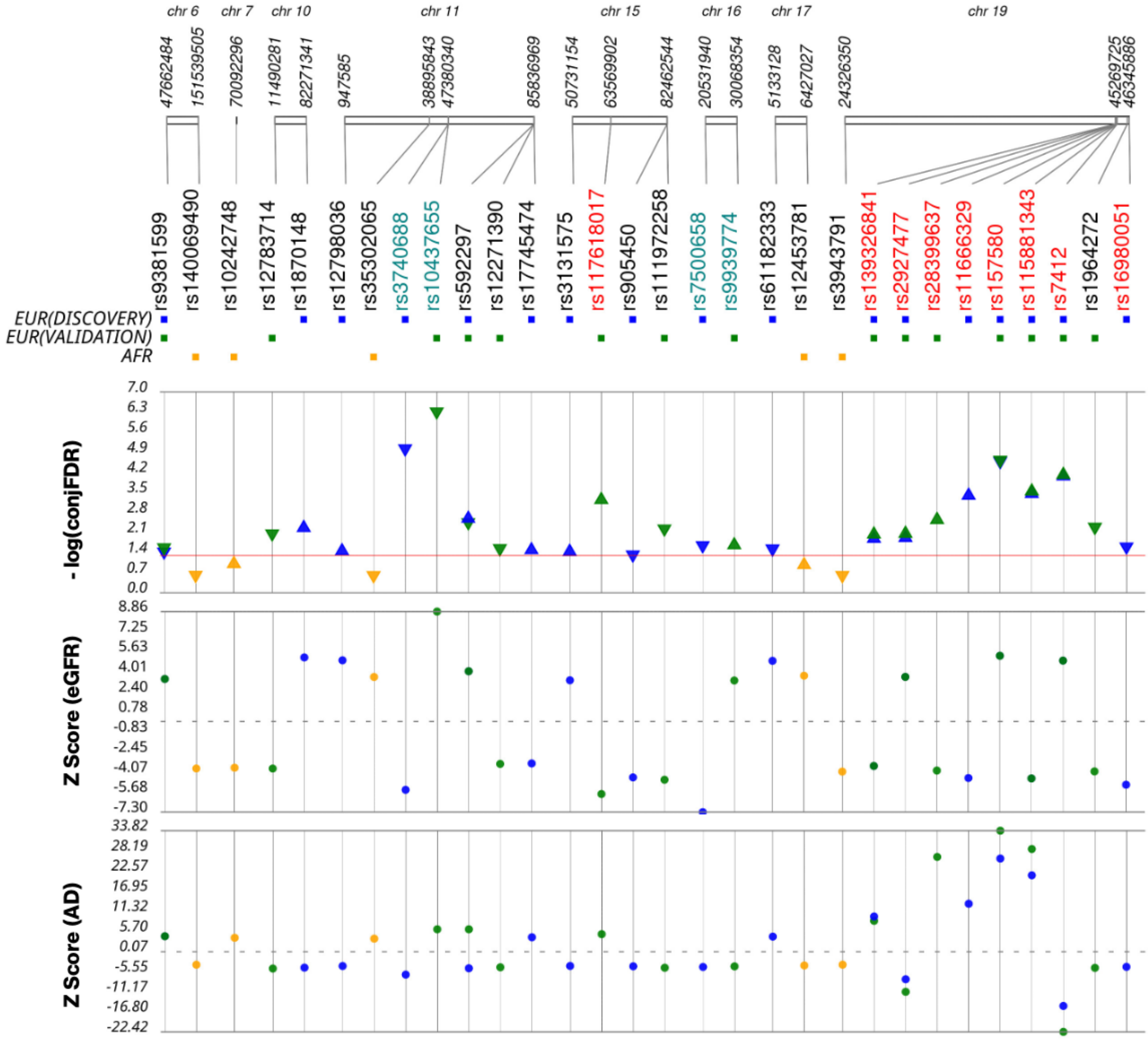

**Supplementary Figure S11. ConjFDR Lead Loci and Z-Score Concordance for Pleiotropic Variants Between Alzheimer's Disease and eGFR in European Ancestry.** This figure displays lead loci from conjFDR analysis in European ancestry, with Z-score concordance plots for pleiotropic SNPs. Panels show effect direction alignment between AD and eGFR. Full SNP lists and loci are in Supplementary Tables S16, S17, S19, S20, S21, and S22.

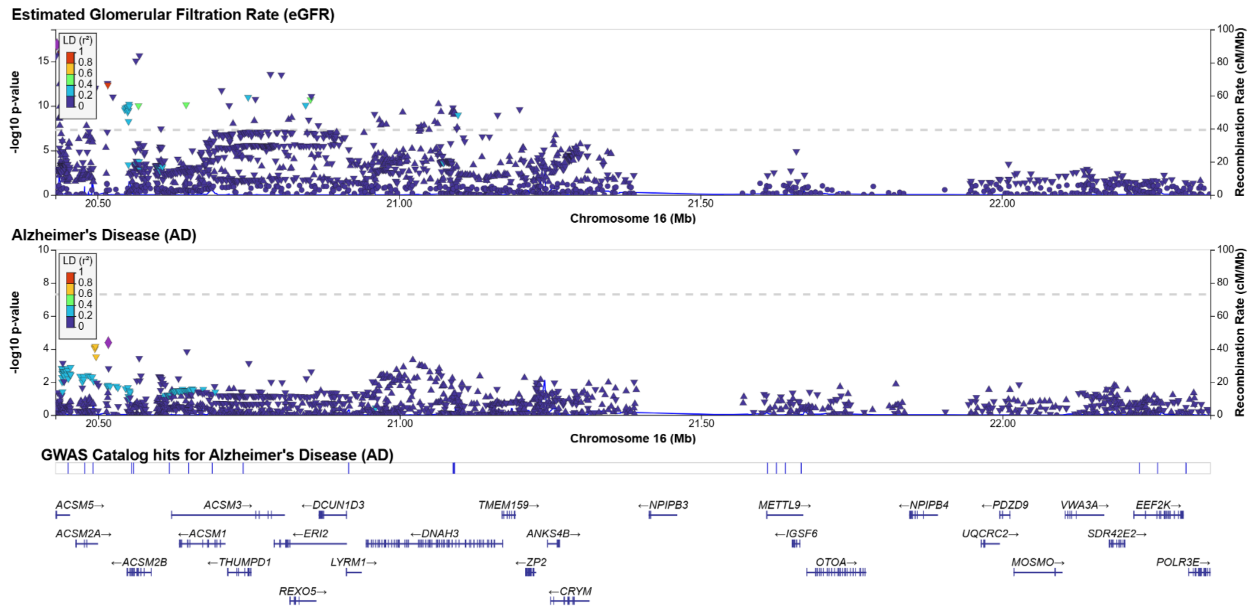

**Supplementary Figure S12. Example 1 of LAVA Null Results with Similar Variance-Covariance Structure Between AD and eGFR in European Ancestry.** This figure provides an example of a genomic locus with null local genetic correlation (Rg) from LAVA analysis between AD and eGFR in European ancestry. Signals are weak, but variance/covariance patterns suggest potential shared genetics. Complete LAVA results are in Supplementary Tables S8, S9, S13, and S14.

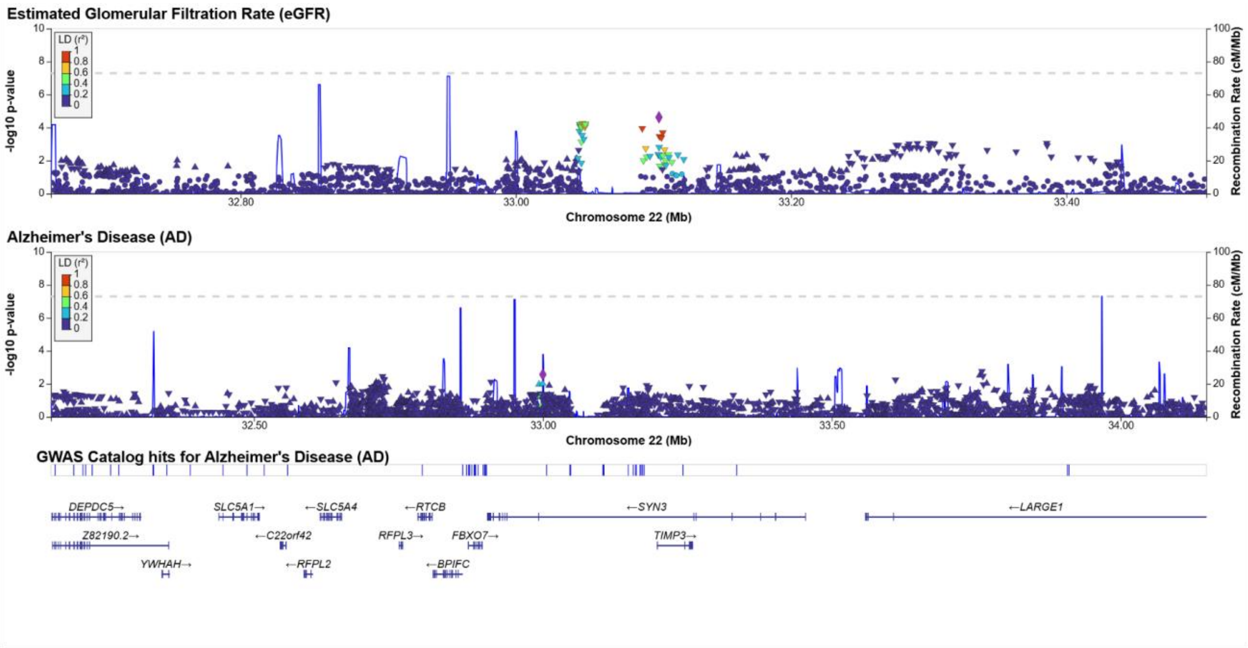

**Supplementary Figure S13. Example 2 of LAVA Null Results with Similar Variance-Covariance Structure Between AD and eGFR in European Ancestry.** This figure shows another example of a locus with null local Rg from LAVA, where weak signals and similar variance/covariance matrices indicate candidate shared genetics between AD and eGFR in European ancestry. Related data are in Supplementary Tables S8, S9, S13, and S14.

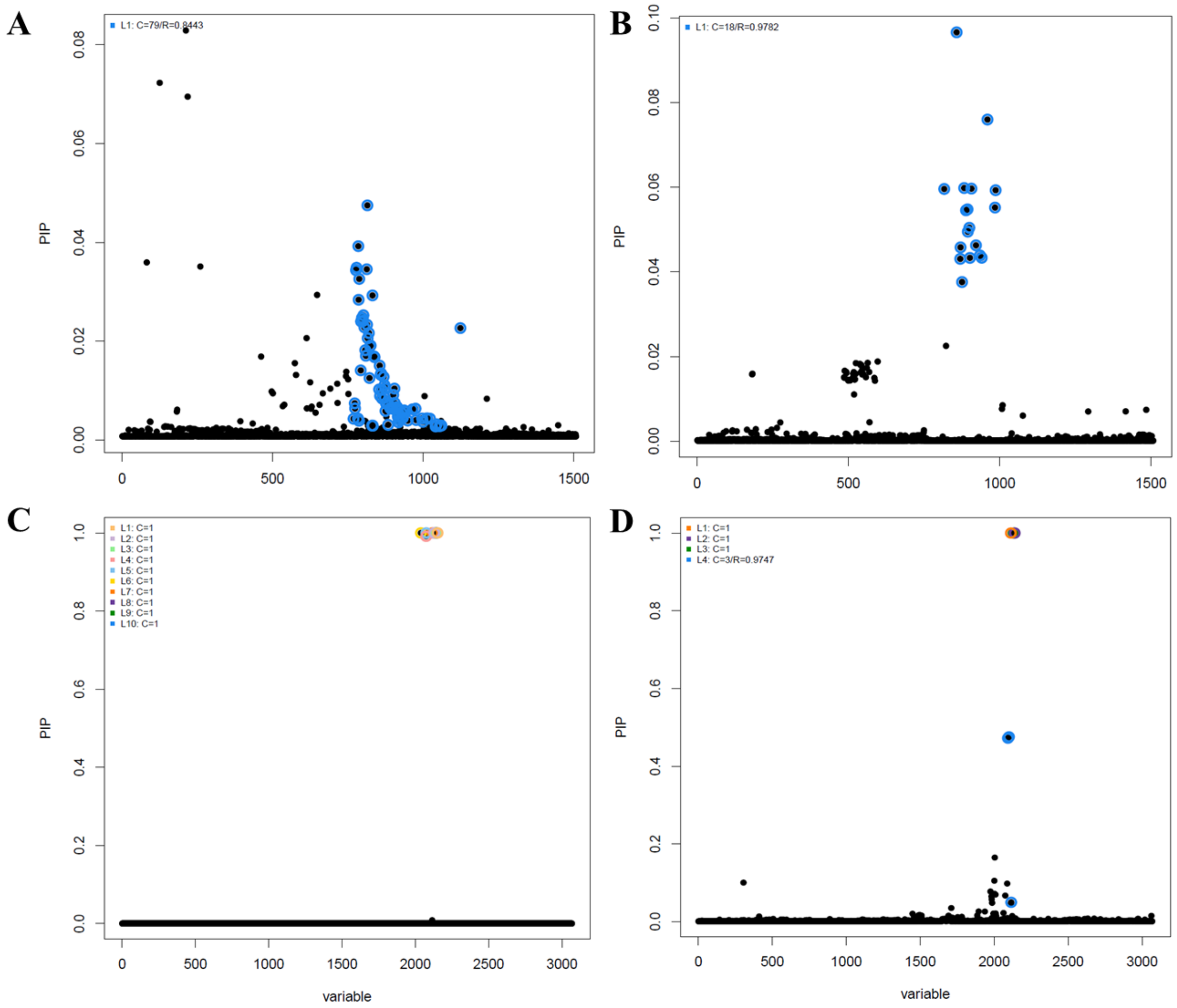

**Supplementary Figure S14. Colocalization Analysis Using SuSiE Credible Sets for Locus 5 (SPI1 Region) and Locus 10 (APOE Region) in European Ancestry.** Four-panel colocalization plot displaying results from Sum of Single Effects (SuSiE) fine-mapping analysis for two key pleiotropic loci. Panels (a) and (b) show locus 5 (chromosome 11, SPI1 region, approximately 46.4-48.4 Mb), while panels (c) and (d) show locus 10 (chromosome 19, APOE region, approximately 45.0-45.9 Mb). For each locus, one panel displays $-\log_{10}$(P-values) for AD associations and the other for eGFR associations across the region, with credible sets of potentially causal variants highlighted. Colored circles indicate SNPs with high posterior inclusion probability (PIP) in the SuSiE credible sets.

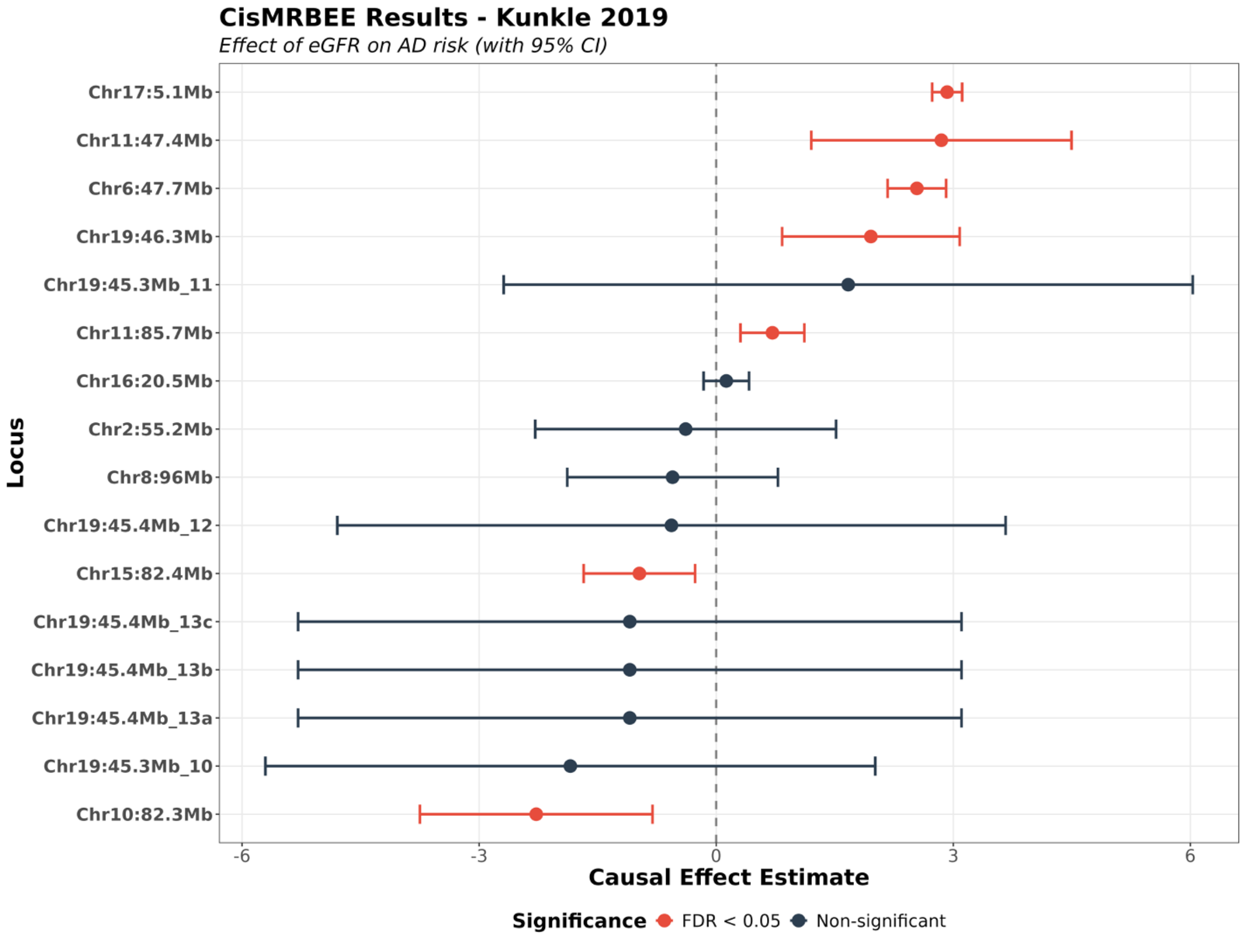

**Supplementary Figure S15. Forest Plot of Forward Cis-Mendelian Randomization Analysis (eGFR to AD) in European Ancestry (Discovery Dataset: Kunkle Corrected).** Forest plot displaying causal effect estimates from forward cis-Mendelian randomization (cis-MR) analysis in European ancestry using the discovery dataset (Kunkle corrected). Each row represents a distinct genomic locus identified through conjFDR analysis. The x-axis shows the causal effect estimate (theta) of genetically predicted eGFR on AD risk, with error bars indicating 95% confidence intervals. Effect estimates to the left of zero suggest that genetically predicted higher eGFR reduces AD risk, while estimates to the right suggest increased risk. Loci are ordered by effect size or chromosomal position. Statistical significance is indicated when confidence intervals do not cross zero.

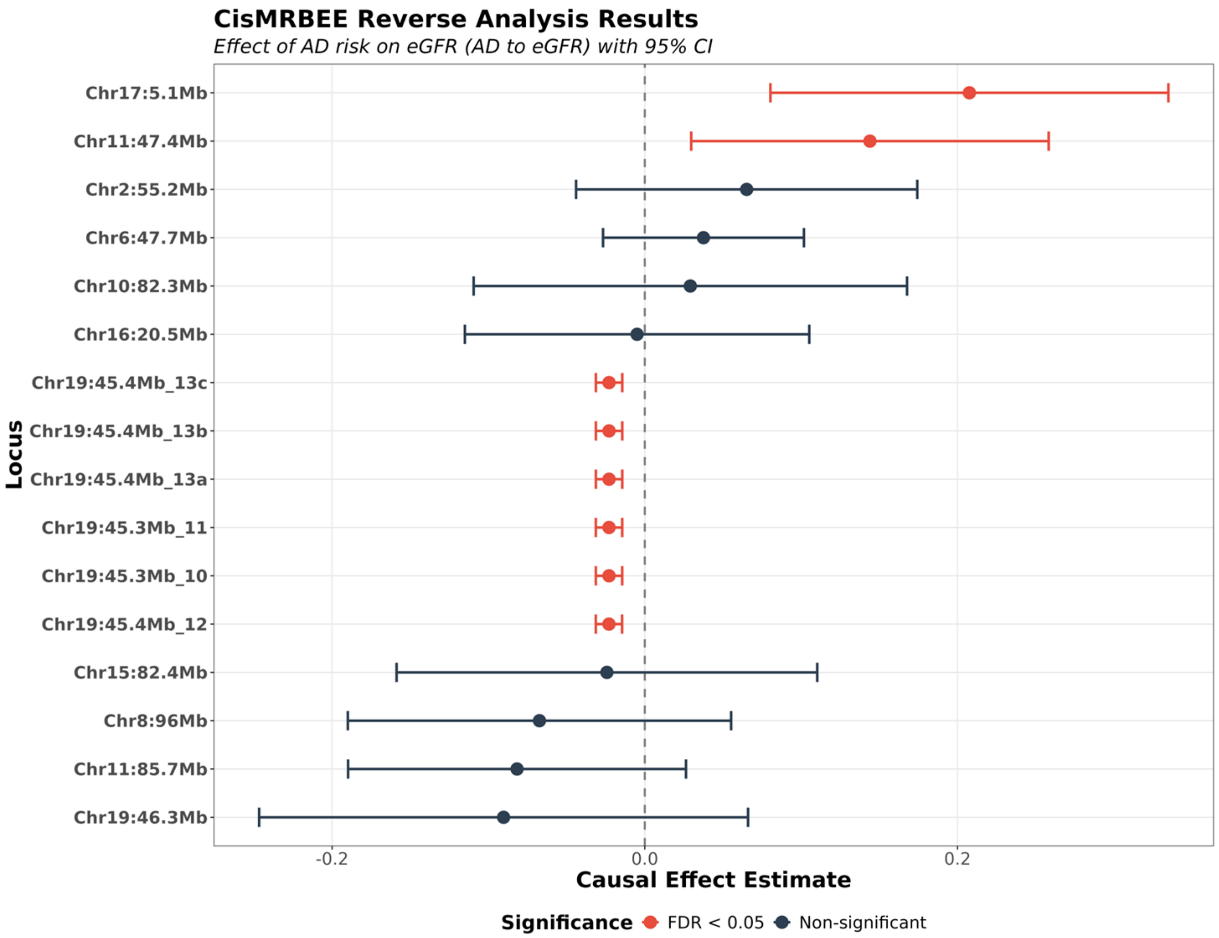

**Supplementary Figure S16. Forest Plot of Reverse Cis-Mendelian Randomization Analysis (AD to eGFR) in European Ancestry (Discovery Dataset: Kunkle Corrected).** Forest plot presenting causal effect estimates from reverse cis-MR analysis in European ancestry using the discovery dataset (Kunkle corrected). Each row represents a genomic locus, showing the estimated causal effect (theta) of genetically predicted AD liability on eGFR, with 95% confidence intervals. Effect estimates to the left of zero indicate that genetic liability for AD decreases eGFR, while estimates to the right indicate increased eGFR.

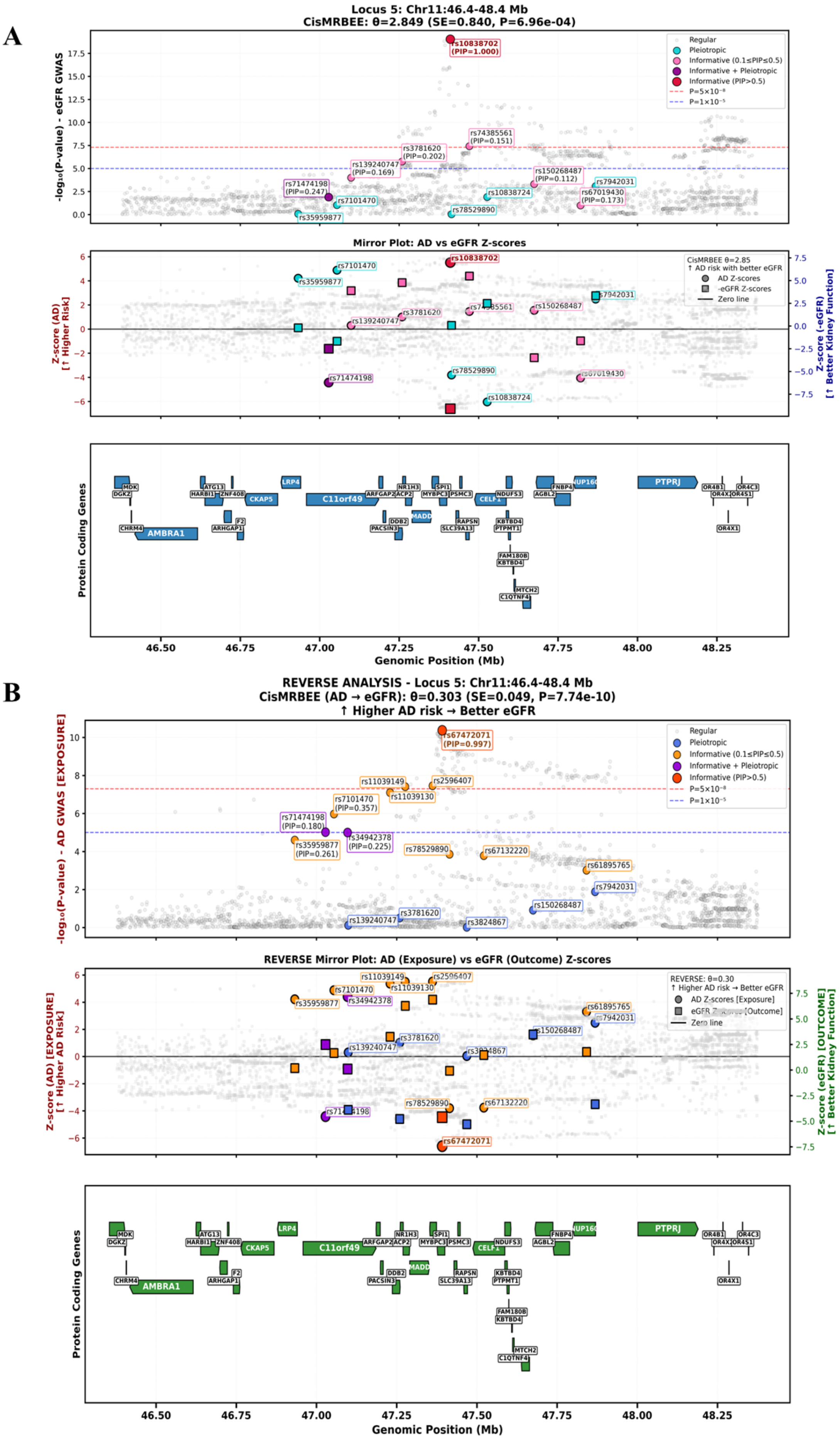
A
Locus 5: Chr11:46.4-48.4 Mb
CisMRBEE: θ=2.849 (SE=0.840, P=6.96e-04)
-log₁₀(P-value) - eGFR GWAS
Regular
Pleiotropic
Informative (0.1≤PIP≤0.5)
Informative + Pleiotropic
Informative (PIP>0.5)
P=5×10⁻⁸
P=1×10⁻⁵
rs10838702 (PIP=1.000)
rs74385561 (PIP=0.151)
rs3781620 (PIP=0.202)
rs139240747 (PIP=0.169)
rs71474198 (PIP=0.247)
rs150268487 (PIP=0.112)
rs67019430 (PIP=0.173)
rs7942031
rs10838724
rs7101470
rs78529890
rs35959877
Mirror Plot: AD vs eGFR Z-scores
Z-score (AD) [↑ Higher Risk]
Z-score (-eGFR) [↑ Better Kidney Function]
CisMRBEE θ=2.85
↑ AD risk with better eGFR
AD Z-scores
-eGFR Z-scores
Zero line
Protein Coding Genes
Genomic Position (Mb)
MDK
DGKZ
CHRM4
AMBRA1
ATG13
HARBI1
ZNF408
ARHGAP1
F2
CKAP5
LRP4
C11orf49
ARFGAP2
ACP2
NR1H3
DDB2
PACSIN3
MADD
SPI1
MYBPC3
PSMC3
RAPSN
SLC39A13
CELF1
NDUFS3
KBTBD4
PTPMT1
FAM180B
MTCH2
C1QTNF4
AGBL2
FNBP4
NUP160
PTPRJ
OR4B1
OR4X2
OR4C3
OR4S1
OR4X1
B
REVERSE ANALYSIS - Locus 5: Chr11:46.4-48.4 Mb
CisMRBEE (AD → eGFR): θ=0.303 (SE=0.049, P=7.74e-10)
↑ Higher AD risk → Better eGFR
-log₁₀(P-value) - AD GWAS [EXPOSURE]
rs67472071 (PIP=0.997)
rs11039149
rs11039130
rs2596407
rs7101470 (PIP=0.357)
rs71474198 (PIP=0.180)
rs34942378 (PIP=0.225)
rs35959877 (PIP=0.261)
rs78529890
rs67132220
rs61895765
rs7942031
rs150268487
rs3781620
rs139240747
rs3824867
REVERSE Mirror Plot: AD (Exposure) vs eGFR (Outcome) Z-scores
Z-score (AD) [EXPOSURE] [↑ Higher AD Risk]
Z-score (eGFR) [OUTCOME] [↑ Better Kidney Function]
REVERSE: θ=0.30
↑ Higher AD risk → Better eGFR
AD Z-scores [Exposure]
Zero line
Protein Coding Genes
Genomic Position (Mb)

**Supplementary Figure S17. Cis-Mendelian Randomization Reveals Bidirectional and Ambiguous Causal Relationship at Locus 5 (SPI1 Region).** Detailed three-panel visualization of bidirectional cis-MR analysis at locus 5 on chromosome 11 (46.4-48.4 Mb) encompassing PTPMT1, SPI1, and surrounding genes. (a) Forward analysis (eGFR to AD): Top panel displays -$\log_{10}$(P-values) for both eGFR (orange/yellow points) and AD (pink/red points) associations across the locus. Middle panel shows a mirror plot with concordant effect directions, where z-scores from both traits align in the same direction for most credible set variants. Credible set variants are marked by squares (AD) and circles (eGFR), with colors indicating effect direction (red for positive effects, blue for negative effects). The mirror plot demonstrates positive correlation between effect sizes. Bottom panel shows protein-coding gene annotations. Gene region spans chromosome 11 positions 46.5-48.25 Mb.

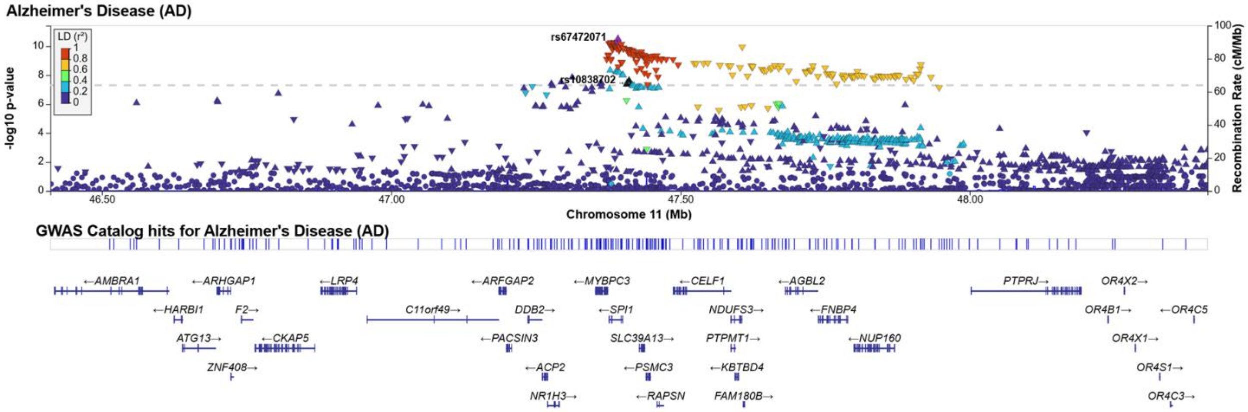

**Supplementary Figure S18. LocusZoom Plot of Locus 5 (SPI1 Region) Demonstrating Linkage Disequilibrium Structure and Recombination Rates**. The two labeled SNPs are the two putative SNPs for AD and eGFR respectively and that are colocalized with each other. Regional LD plot shows correlation ($R^2$) patterns between variants. Recombination rate is plotted as blue line showing recombination hotspots in the region.

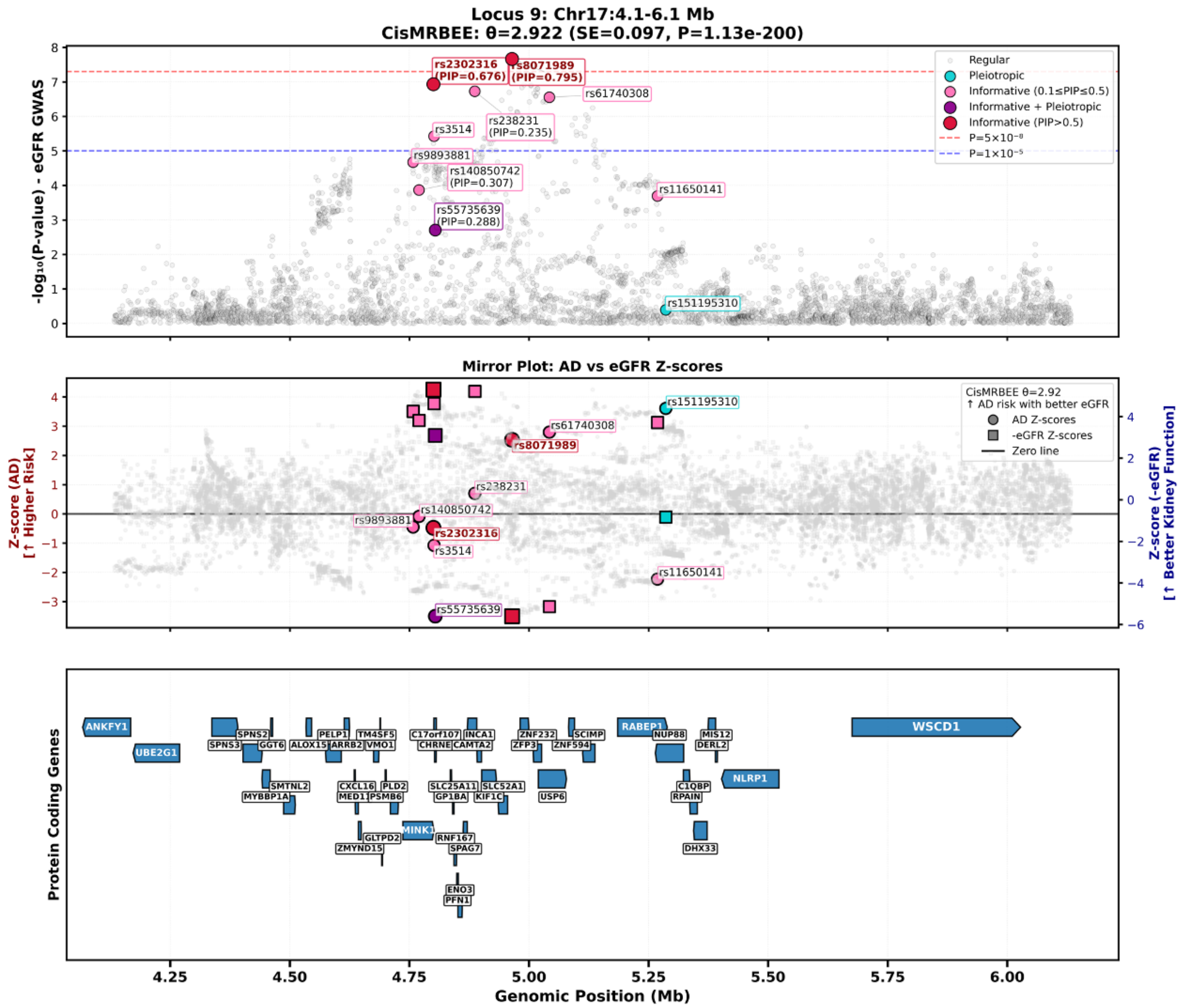

**Supplementary Figure S19. Forward Cis-Mendelian Randomization Analysis (eGFR to AD) for Locus 9 Showing Bidirectional Relationship in European Ancestry.** Detailed cis-MR analysis for locus 9 on chromosome 17, displaying forward analysis where eGFR serves as the exposure and AD as the outcome. The multi-panel figure includes: top panel showing -log$_{10}$(P-values) for both AD and eGFR associations across the locus; middle panel presenting a mirror or scatter plot of effect sizes demonstrating the relationship between eGFR and AD genetic effects; bottom panel displaying gene annotations for the region. This locus exhibits evidence of bidirectional causality between eGFR and AD, meaning genetic variants in this region suggest that kidney function causally influences AD risk and vice versa.

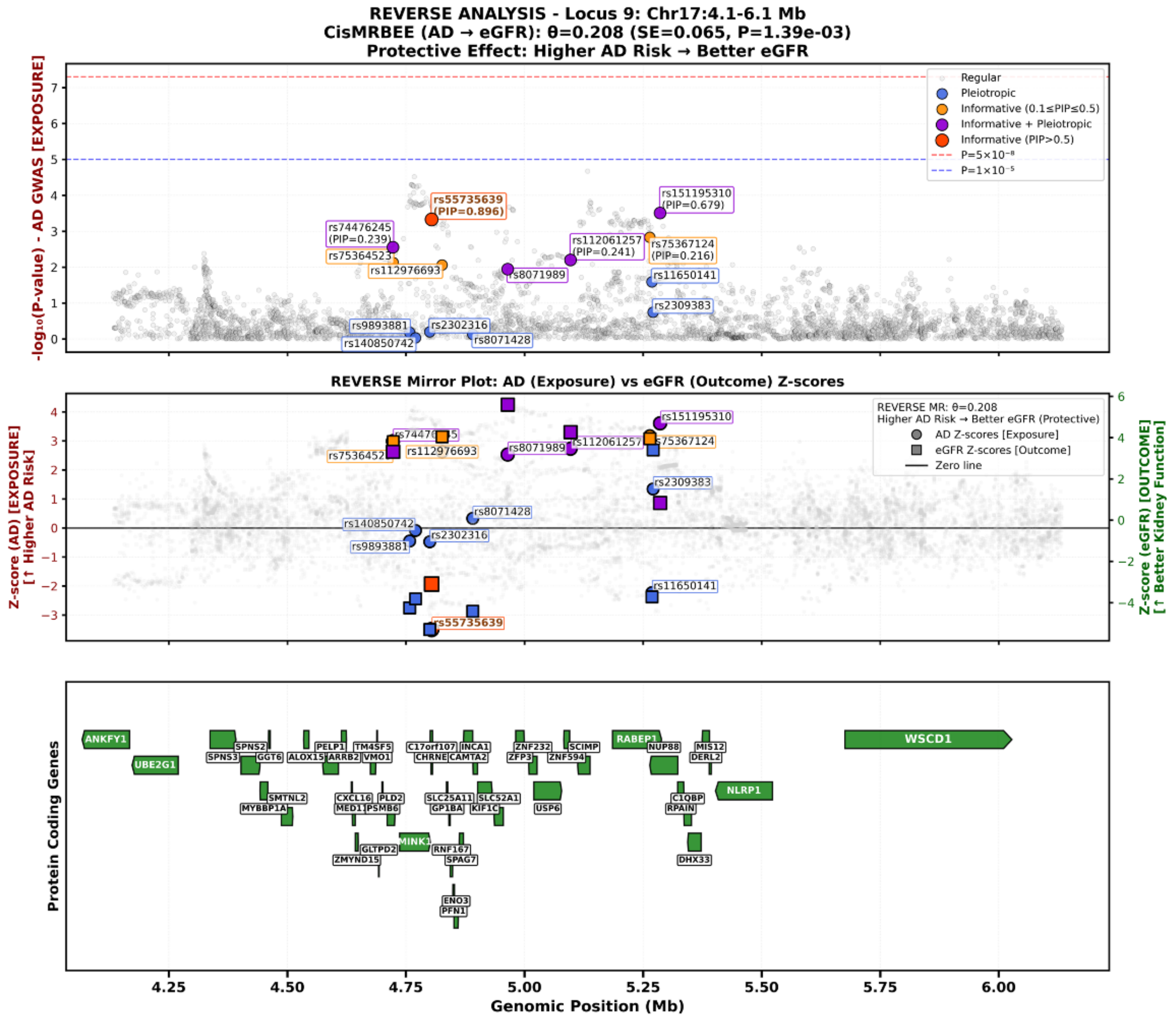

**Supplementary Figure S20. Reverse Cis-Mendelian Randomization Analysis (AD to eGFR) for Locus 9 Showing Bidirectional Relationship in European Ancestry.** Reverse cis-MR analysis for locus 9 on chromosome 17, where AD serves as the exposure and eGFR as the outcome. The figure layout parallels Figure 19, showing association patterns, effect size relationships, and gene annotations. The combination of significant forward (Figure 19) and reverse (Figure 20) cis-MR results at this locus provides strong evidence for bidirectional causality, suggesting complex biological pleiotropy where genetic variation in this region influences both AD risk and kidney function through interconnected mechanisms.

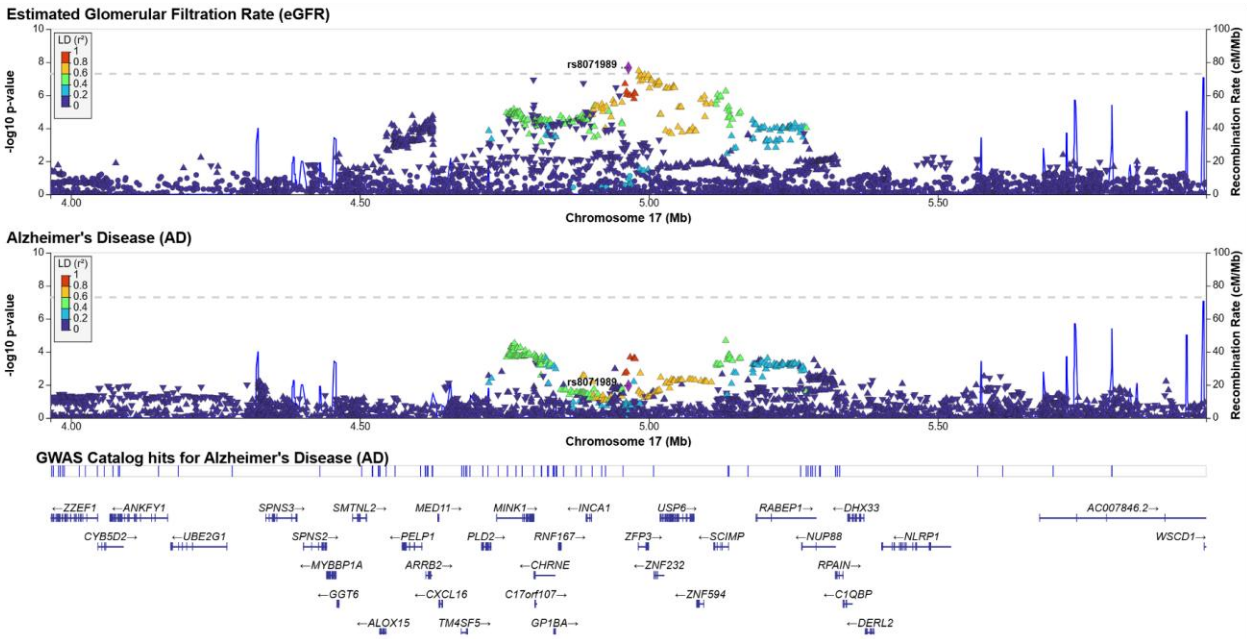

**Supplementary Figure S21. LocusZoom Plot of Locus 9 Region on Chromosome 17 with Bidirectional Pleiotropy in European Ancestry.**

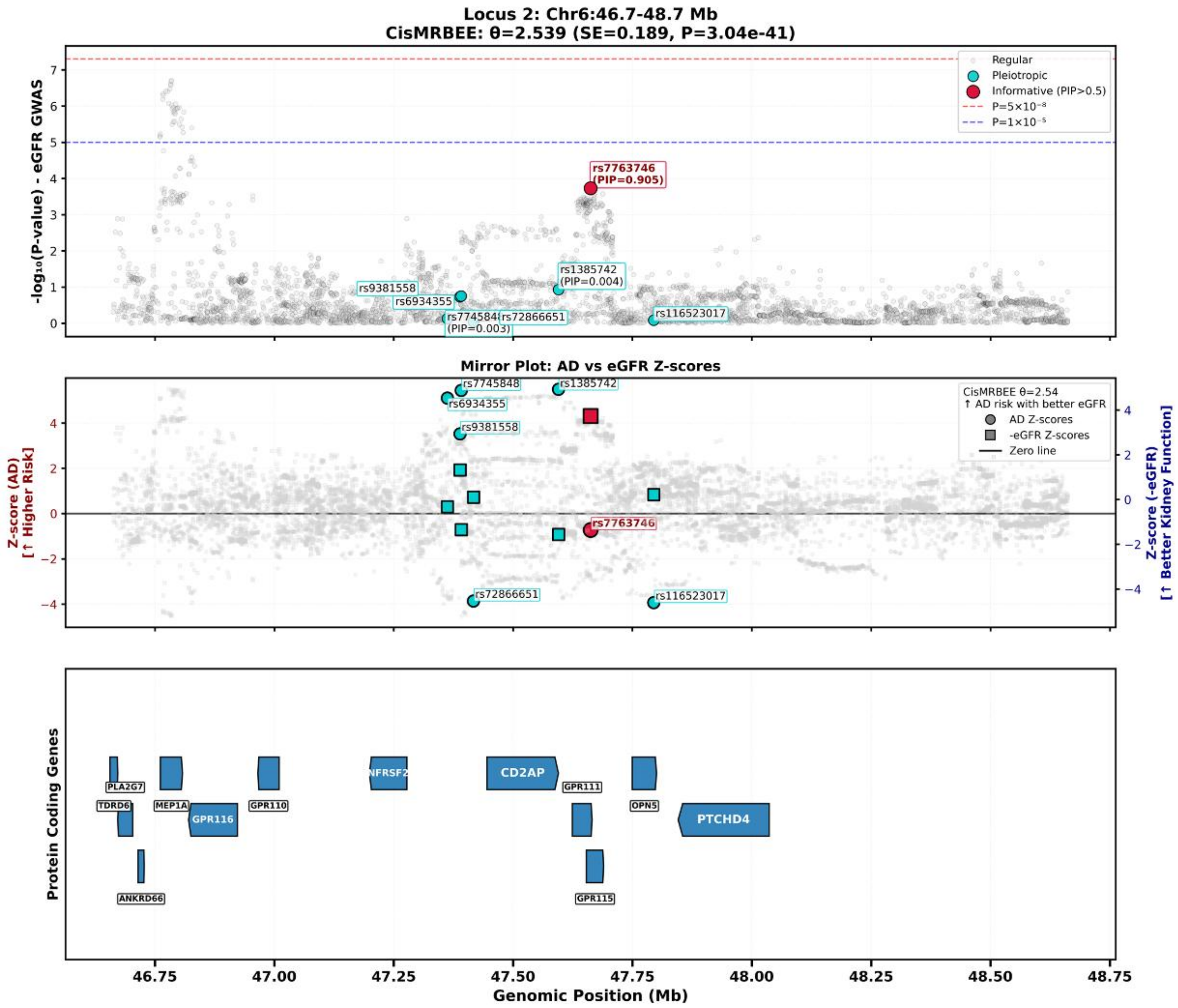

**Supplementary Figure S22. Forward Cis-Mendelian Randomization Analysis (eGFR to AD) for Locus 2 Showing Horizontal Pleiotropy in European Ancestry**. Cis-MR analysis for locus 2, displaying forward analysis (eGFR to AD). The figure shows association patterns, effect size relationships, and gene annotations for this locus. In contrast to bidirectional causality seen at other loci, this region exhibits horizontal pleiotropy, where genetic variants affect both AD and eGFR through independent biological pathways rather than through causal relationships between the traits.

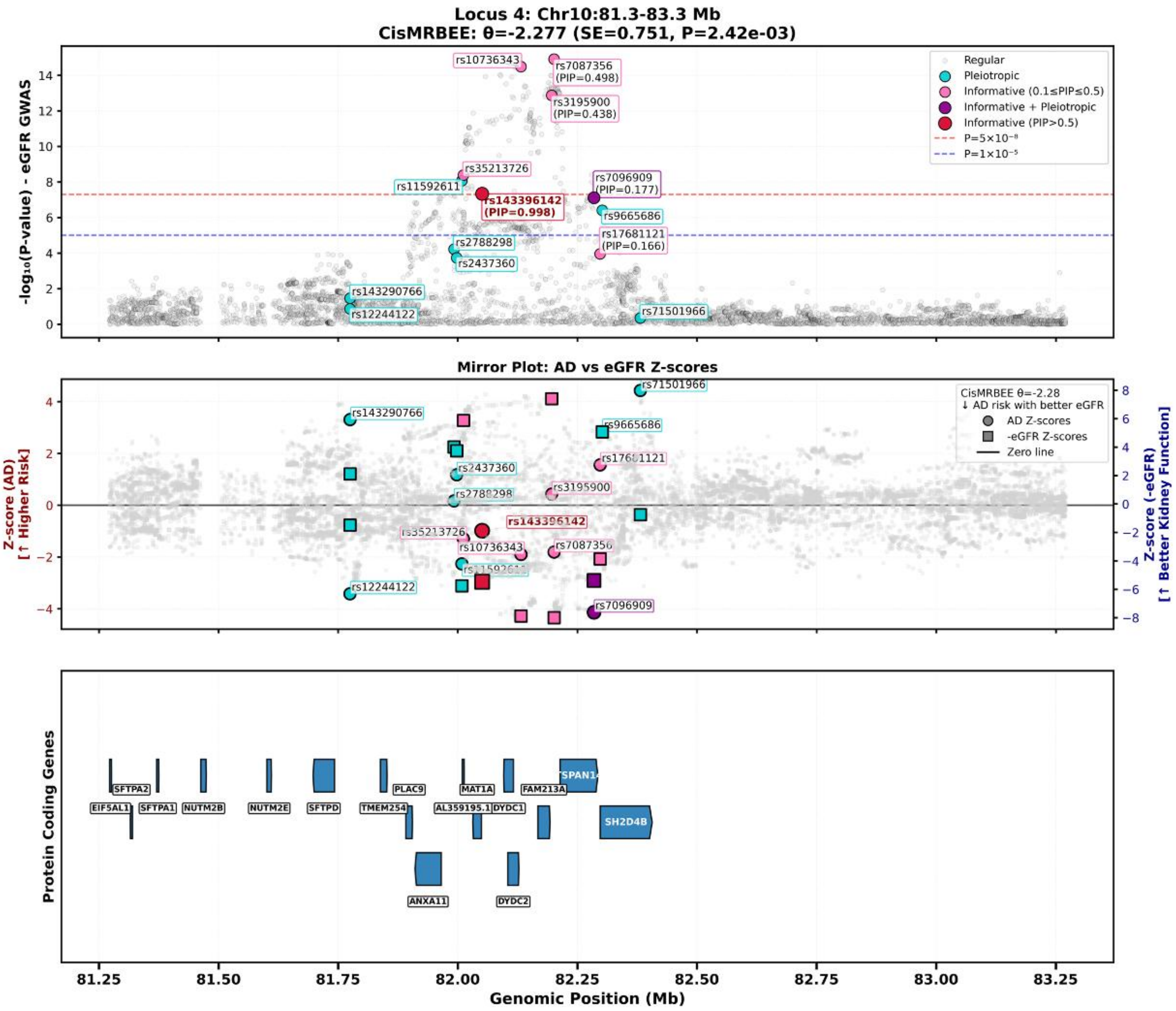

**Supplementary Figure S23. Forward Cis-Mendelian Randomization Analysis (eGFR to AD) for Locus 4 Showing Horizontal Pleiotropy in European Ancestry.** Cis-MR forward analysis for locus 4, showing genetic associations, effect relationships, and gene annotations. Similar to locus 2 (Figure 22), this region demonstrates horizontal pleiotropy, where genetic variants influence both AD and kidney function through separate biological mechanisms.

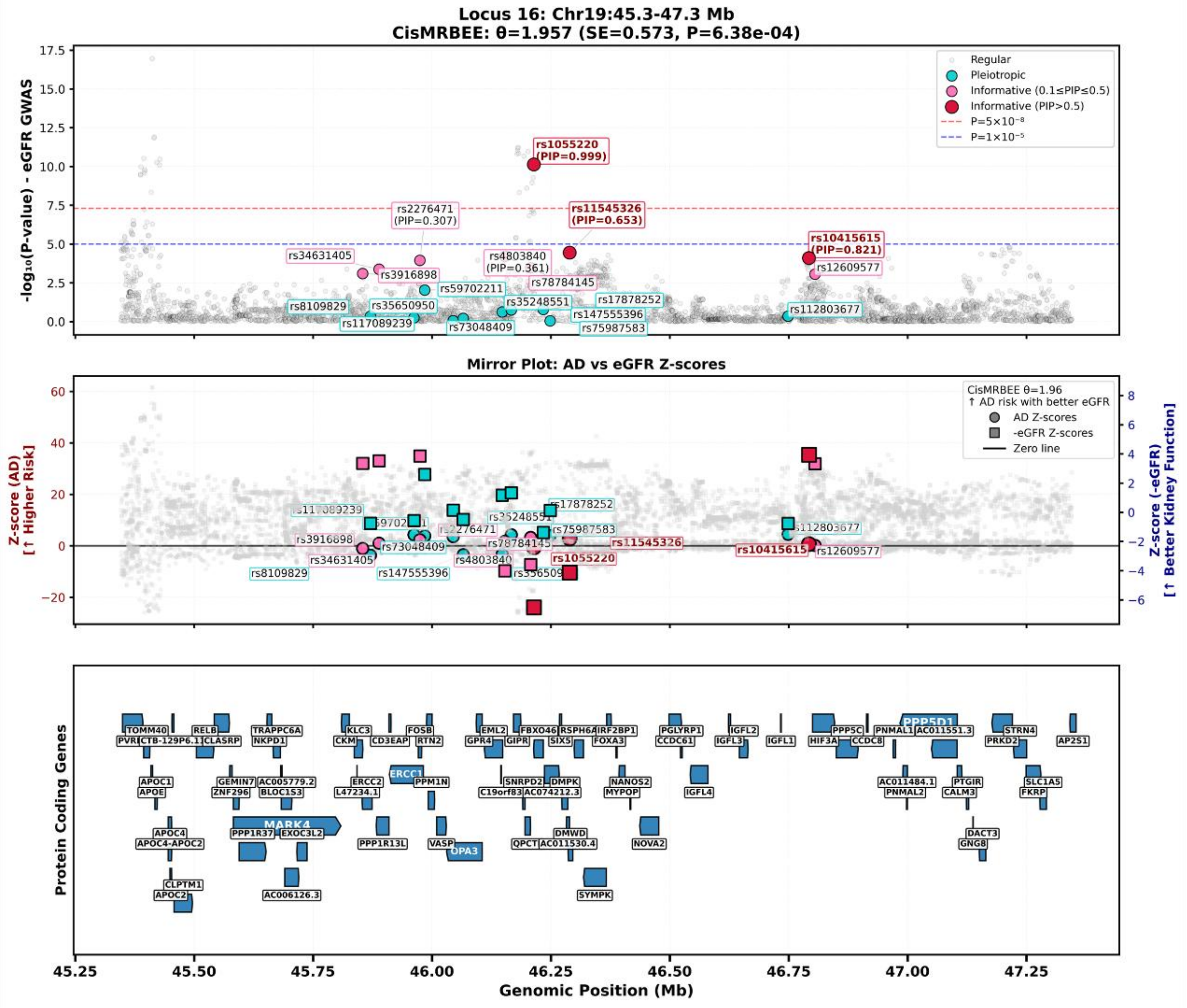

**Supplementary Figure S24. Forward Cis-Mendelian Randomization Analysis (eGFR to AD) for Locus 16 Showing Horizontal Pleiotropy in European Ancestry.** Cis-MR forward analysis for locus 16, displaying association patterns and effect relationships. This locus represents another example of horizontal pleiotropy, where genetic overlap between AD and eGFR does not reflect causal relationships but rather independent effects on both traits.

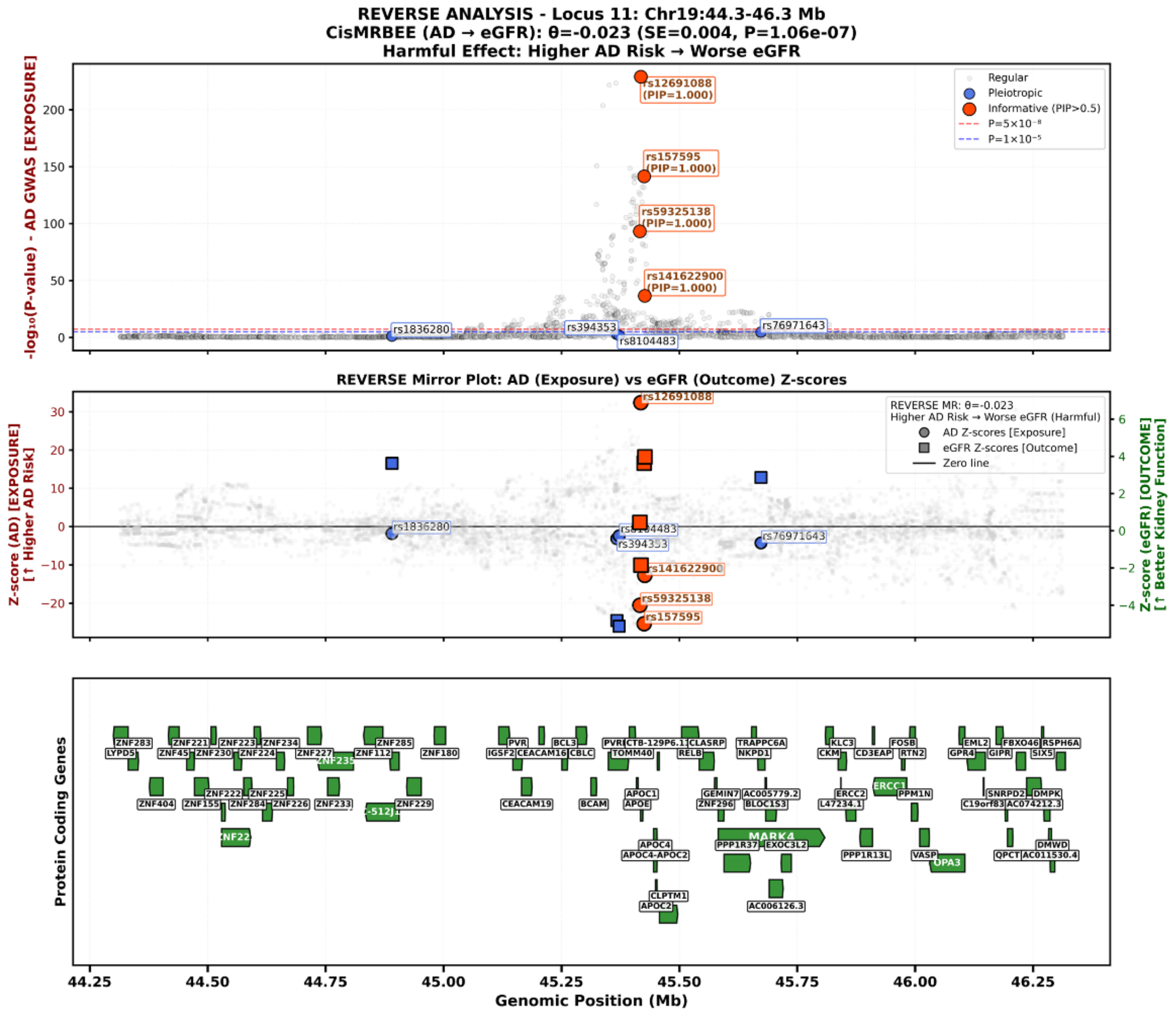

**Supplementary Figure S25. Reverse Cis-Mendelian Randomization Analysis (AD to eGFR) for Locus 11 Showing Reverse Causality in European Ancestry**. Reverse cis-MR analysis for locus 11, displaying AD as exposure and eGFR as outcome. The multi-panel figure shows association patterns, effect relationships, and gene annotations. This locus demonstrates reverse causality, where genetic liability for AD causally influences kidney function, but the forward direction (eGFR to AD) shows no significant causal effect.

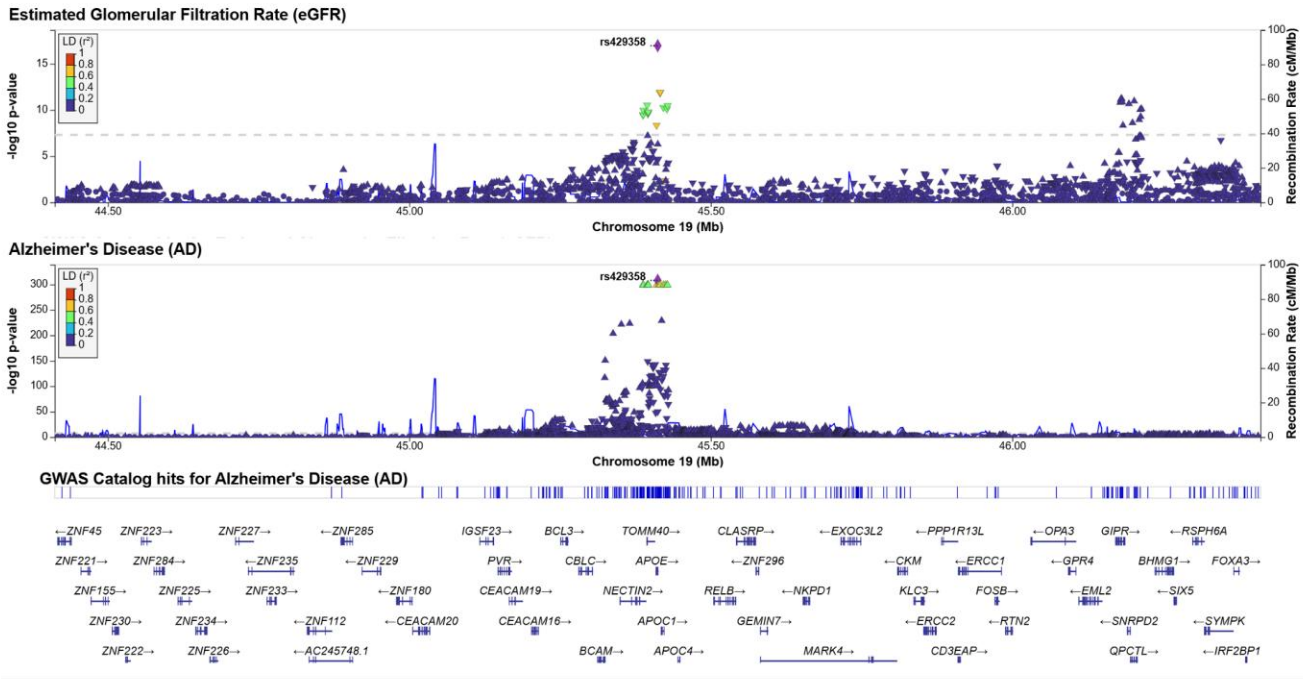

**Supplementary Figure S26. LocusZoom Plot of Chromosome 19 Region Spanning *APOE/TOMM40/APOC1* with Reverse Causality in European Ancestry.** Regional association plot for the APOE locus on chromosome 19, displaying LD structure and genetic associations. The plot shows -$\log_{10}$(P-values) for both AD and eGFR across the region encompassing APOE, TOMM40, and APOC1 genes. Points are colored by LD ($R^2$) with the lead SNP, and recombination rate is overlaid.

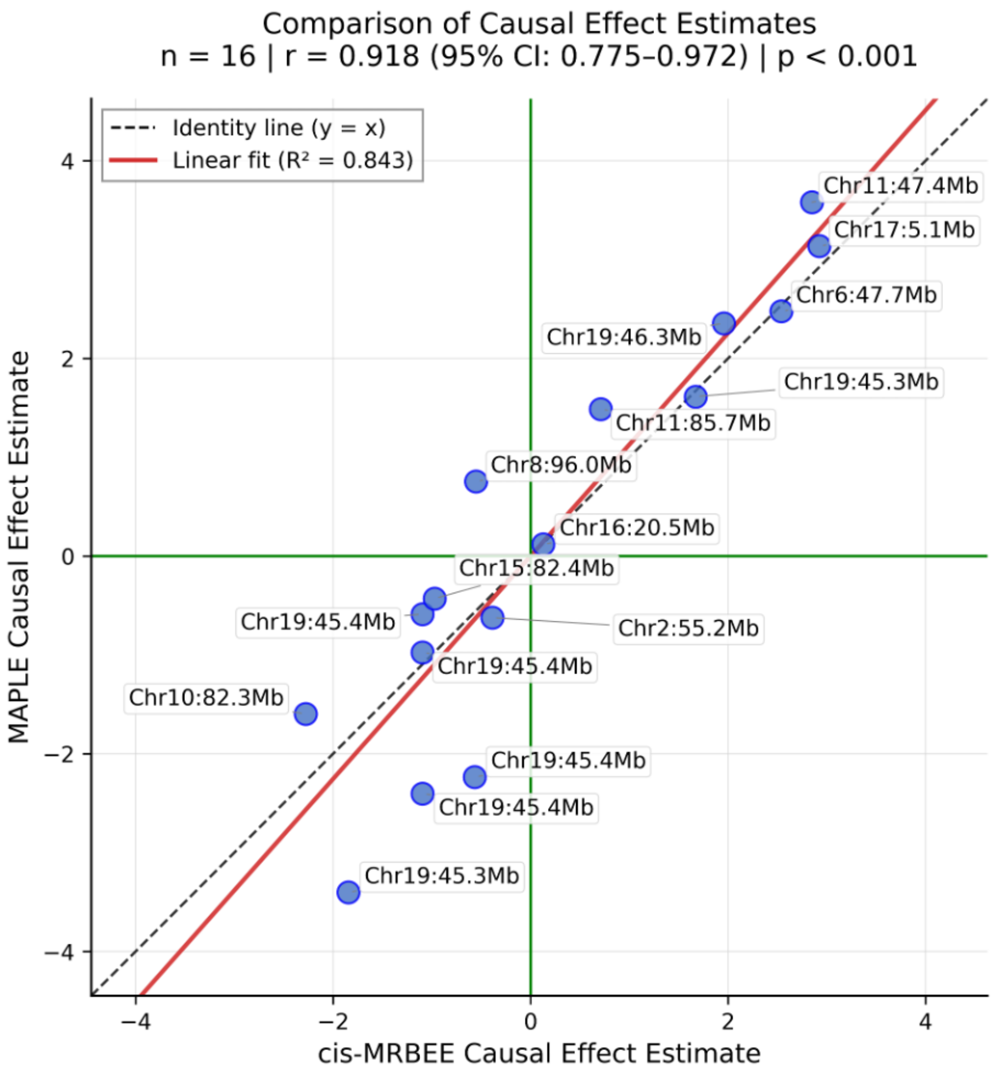

**Supplementary Figure S27. Comparison Between Cis-MR and MAPLE Methods for Robustness of Causal Estimation**. Comparison plot displaying causal effect estimates from two different methodological approaches: cis-Mendelian randomization (cis-MRBEE) and MAPLE (MR method with self-adaptive determination of sample structure and multiple pleiotropic effects). For each locus analyzed, the figure shows effect estimates from both methods with confidence intervals or standard errors.

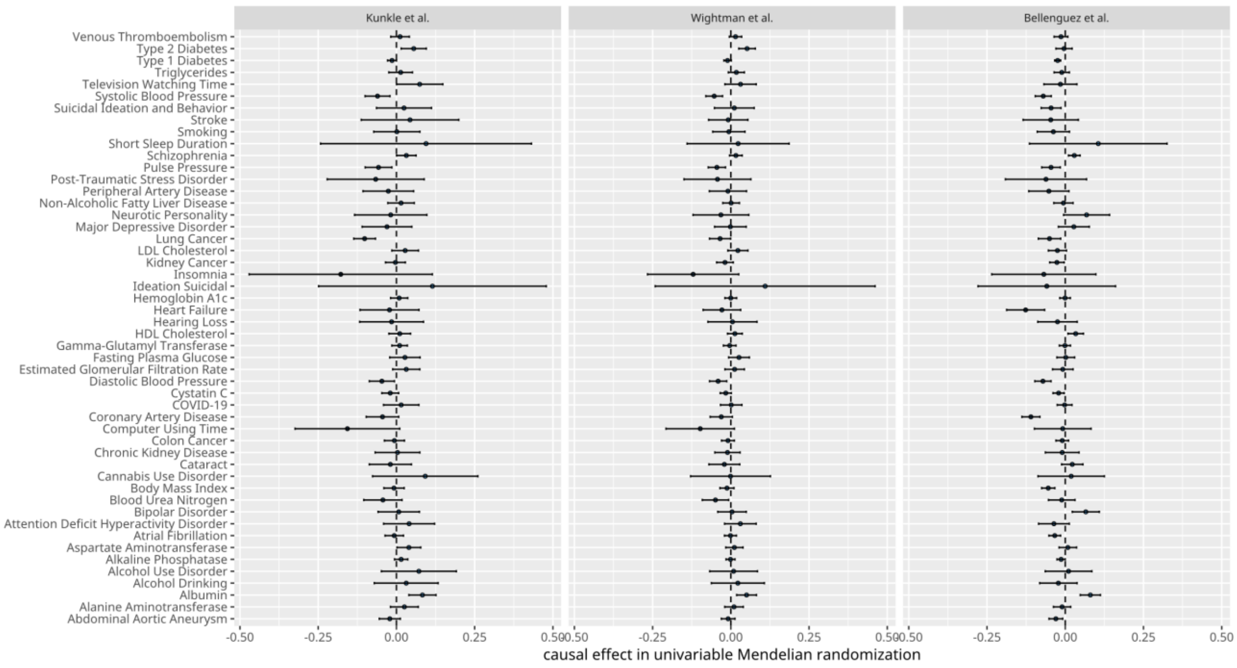

**Supplementary Figure S28. Forest Plot Illustrating Collider Bias Correction for Competing Risk in AD GWAS.** It shows the multivariable Mendelian randomization (MVMR)-based correction strategy used to mitigate survival-induced collider bias in AD GWAS summary statistics. Survival to study participation (S) functions as a collider between genetic variants and competing terminal diseases. Five competing diseases (coronary artery disease, heart failure, type 1 diabetes, kidney cancer, and lung cancer) showed negative effect estimates on AD risk consistent with selection bias. Using MRBEE, AD effect sizes were adjusted for genetic pathways operating through these competing diseases, yielding corrected summary statistics for downstream analyses.

### Supplementary Tables

#### Table of Contents

| Table number | Table ID | Table Title | Table Legend |
|---|---|---|---|
| 1 | S1_GWAS_Summary | Summary of Alzheimer's Disease and eGFR GWAS Data Sources | Overview of ancestry, traits, SNP counts, sample sizes, effective sample sizes, and sources for AD and eGFR GWAS datasets across European and African ancestries. |
| 2 | S2_Individual_Data_PRS | Individual-Level Genotype Data from ADGC MEA Dataset for PRS Analysis | Demographic and genetic characteristics of healthy controls and AD cases in European and African ancestry cohorts from ADGC, including age, sex, education, and APOE ε4 dosage distributions. |
| 3 | S3_LDSC_GlobalCorrelation | Genome-Wide Genetic Correlation Analysis Using LD Score Regression | Heritability estimates, genetic correlation, and covariance between AD and eGFR in European and African ancestries using LDSC, with standard errors and significance levels. |
| 4 | S4_PRS_QC_Metrics | Quality Control Metrics for Polygenic Risk Score Calculation | Quality control metrics for SNPs in PRS calculation, including totals, exclusions, and matched SNPs between GWAS weights and individual-level data for European and African ancestries. |
| 5 | S5_PRS_Categorization | Polygenic Risk Score Categorization Approaches and Cutoff Definitions | Categorization methods for eGFR-PRS (quantile, SD-based, tertile) with mean PRS values and risk classifications for low, moderate, high, and very high risk groups. |
| 6 | S6_PRS_EUR_Results | Association of eGFR Polygenic Risk Scores with Alzheimer's Disease Risk in European Ancestry | Logistic regression results for eGFR-PRS associations with AD in European ancestry, including beta coefficients, odds ratios, and p-values across quantile, SD-based, tertile, and continuous groupings, with and without APOE adjustment. |
| 7 | S7_PRS_AFR_Results | Association of eGFR | Logistic regression results |

| Table number | Table ID | Table Title | Table Legend |
|---|---|---|---|
| | | Polygenic Risk Scores with Alzheimer's Disease Risk in African American Ancestry | for eGFR-PRS associations with AD in African ancestry, including beta coefficients, odds ratios, and p-values across quantile and tertile groupings, with and without APOE adjustment |
| 8 | S8_LAVA_EUR_Discovery_Sig | Significant Local Genetic Correlations Between AD and eGFR in European Ancestry (Kunkle Discovery) | Local genetic correlations, heritabilities, p-values, and direction indicators for significant shared loci in European ancestry using Kunkle corrected AD data. |
| 9 | S9_LAVA_EUR_Validation_Sig | Significant Local Genetic Correlations Between AD and eGFR in European Ancestry (Wightman Validation) | Local genetic correlations, heritabilities, p-values, and direction indicators for significant shared loci in European ancestry using Wightman validation AD data. |
| 10 | S10_LAVA_AFR_Significant | Significant Local Genetic Correlations Between AD and eGFR in African American Ancestry | Local genetic correlations, heritabilities, p-values, and direction indicators for significant shared loci in African ancestry using Ray AD data. |
| 11 | S11_LAVA_Summary_Statistics | Summary Statistics of Local Genetic Correlation Analysis Across Ancestries | Counts of LAVA testable loci, significant loci, and percentages of positive/negative correlations across European (discovery and validation datasets) and African ancestries. |
| 12 | S12_LAVA_Ancestry_Comparison | Cross-Ancestry Comparison of Significant Local Genetic Correlations | Classification of significant LAVA loci as EUR-only, AFR-only, or shared, with local genetic correlations and p-values across ancestries. |
| 13 | S13_LAVA_EUR_Discovery_Complete | Complete Local Genetic Correlation Results in European Ancestry (Kunkle Discovery) | Comprehensive list of all tested loci in European ancestry (Kunkle corrected), including genetic correlations, heritabilities, p-values, FDR, nearest genes, and functional annotations. |
| 14 | S14_LAVA_EUR_Validation_Complete | Complete Local Genetic Correlation Results in European Ancestry (Wightman Validation) | Comprehensive list of all tested loci in European ancestry (Wightman corrected), including genetic correlations, |

| Table number | Table ID | Table Title | Table Legend |
|---|---|---|---|
| | | | heritabilities, p-values, FDR, and genes in the LAVA regions. |
| 15 | S15_LAVA_AFR_Complete | Complete Local Genetic Correlation Results in African American Ancestry | Comprehensive list of all tested loci in African ancestry, including genetic correlations, heritabilities, p-values, FDR, and genes in the LAVA regions. |
| 16 | S16_ConjFDR_EUR_Discovery_AllSNPs | Pleiotropic SNPs Identified by Conjunctional FDR in European Ancestry (Kunkle Discovery) | List of pleiotropic SNPs in European ancestry discovery dataset (Kunkle corrected), with chromosomal positions, Z-scores for AD and eGFR, conjFDR values, and rsIDs. |
| 17 | S17_ConjFDR_EUR_Validation_AllSNPs | Pleiotropic SNPs Identified by Conjunctional FDR in European Ancestry (Wightman Validation) | List of pleiotropic SNPs in European ancestry validation dataset (Wightman corrected), with chromosomal positions, Z-scores for AD and eGFR, conjFDR values, and rsIDs. |
| 18 | S18_ConjFDR_AFR_AllSNPs | Pleiotropic SNPs Identified by Conjunctional FDR in African American Ancestry | List of pleiotropic SNPs in African ancestry, with chromosomal positions, Z-scores for AD and eGFR, conjFDR values, and rsIDs. |
| 19 | S19_ConjFDR_EUR_Discovery_Loci | Distinct Pleiotropic Genomic Loci in European Ancestry (Kunkle Discovery, conjFDR<0.05) | Distinct pleiotropic loci in European ancestry discovery dataset (Kunkle corrected), including lead SNPs, positions, conjFDR, Z-scores, nearest genes, functions, and effect directions. |
| 20 | S20_ConjFDR_EUR_Discovery_Uncorr_Loci | Distinct Pleiotropic Genomic Loci in European Ancestry (Kunkle Uncorrected, conjFDR<0.05) | Distinct pleiotropic loci in European ancestry (Kunkle uncorrected), including lead SNPs, positions, conjFDR, Z-scores, nearest genes, functions, and effect directions. |
| 21 | S21_ConjFDR_EUR_Validation_Loci | Distinct Pleiotropic Genomic Loci in European Ancestry (Wightman Validation, conjFDR<0.05) | Distinct pleiotropic loci in European ancestry validation dataset (Wightman corrected), including lead SNPs, positions, conjFDR, Z-scores, nearest genes, functions, and effect directions. |
| 22 | S22_ConjFDR_EUR_Validation_Uncorr_Loci | Distinct Pleiotropic Genomic Loci in | Distinct pleiotropic loci in European ancestry |

| Table number | Table ID | Table Title | Table Legend |
|---|---|---|---|
| | | European Ancestry (Wightman Uncorrected, conjFDR<0.05) | (Wightman uncorrected), including lead SNPs, positions, conjFDR, Z-scores, nearest genes, functions, and effect directions. |
| 23 | S23_ConjFDR_AFR_Loci | Distinct Pleiotropic Genomic Loci in African American Ancestry (conjFDR<0.2) | Distinct pleiotropic loci in African ancestry (conjFDR<0.2), including lead SNPs, positions, conjFDR, Z-scores, nearest genes, functions, and effect directions. |
| 24 | S24_Coloc_EUR_Discovery | Colocalization Analysis Results Using SuSiE Framework (Kunkle Discovery) | Colocalization probabilities (PP.H0 to PP.H4) for loci in European ancestry using AD discovery data corrected for competing risks, with lead SNPs and indicators for shared causal variants. |
| 25 | S25_Coloc_EUR_Discovery_Uncorr | Colocalization Analysis Results Using SuSiE Framework (Kunkle Uncorrected) | Colocalization probabilities (PP.H0 to PP.H4) for loci in European ancestry using uncorrected AD discovery data, with lead SNPs and indicators for shared causal variants. |
| 26 | S26_Coloc_EUR_Validation | Colocalization Analysis Results Using SuSiE Framework (Wightman Validation) | Colocalization probabilities (PP.H0 to PP.H4) for loci in European ancestry using AD validation data corrected for competing risks, with lead SNPs and indicators for shared causal variants. |
| 27 | S27_Coloc_EUR_Validation_Uncorr | Colocalization Analysis Results Using SuSiE Framework (Wightman Uncorrected) | Colocalization probabilities (PP.H0 to PP.H4) for loci in European ancestry using Wightman uncorrected AD validation data, with lead SNPs and indicators for shared causal variants. |
| 28 | S28_Coloc_EUR_Summary | Summary of Colocalization Analysis in European Ancestry | Counts and percentages of colocalized loci across AD datasets (Kunkle and Wightman, corrected/uncorrected) in European ancestry, including single vs. multiple signal breakdowns. |
| 29 | S29_CisMR_EUR_Results | Cis-Mendelian Randomization Results | Cis-MR beta coefficients, standard errors, Z-scores, p- |

| Table number | Table ID | Table Title | Table Legend |
|---|---|---|---|
| | | in European Ancestry (Kunkle Discovery) | values, and FDR for forward (eGFR to AD) and reverse (AD to eGFR) analyses in European ancestry using AD discovery data corrected for competing risks. |
| 30 | S30_CisMR_EUR_Forward_Ivs | Instrumental Variables for Forward Cis-MR Analysis (eGFR to AD) | Instrumental variables (IVs) for forward cis-MR (eGFR to AD), including SNP counts, lead SNPs, pleiotropic variants, and LD r-values in European ancestry. |
| 31 | S31_CisMR_EUR_Reverse_Ivs | Instrumental Variables for Reverse Cis-MR Analysis (AD to eGFR) | Instrumental variables (IVs) for reverse cis-MR (AD to eGFR), including SNP counts, lead SNPs, pleiotropic variants, and LD r-values in European ancestry. |
| 32 | S32_CisMR_EUR_IV_Overlap | Instrumental Variable Overlap Between Forward and Reverse Cis-MR Analyses | Overlap counts of IVs between forward and reverse cis-MR analyses, including shared, distinct, and pleiotropic SNPs across loci in European ancestry. |

#### Appendix

| Abbreviation | Full Term |
|---|---|
| AD | Alzheimer's disease |
| ADGC | Alzheimer's Disease Genetics Consortium |
| AFR | African American ancestry |
| APOE | Apolipoprotein E |
| BP | Base pair position |
| CADD | Combined Annotation-Dependent Depletion |
| CHR | Chromosome |
| CI | Confidence interval |
| cis-MR | Cis-Mendelian randomization |
| CKD | Chronic kidney disease |
| conjFDR | Conjunctional false discovery rate |
| eGFR | Estimated glomerular filtration rate |
| eQTL | Expression quantitative trait locus |
| EUR | European ancestry |
| FDR | False discovery rate |
| GWAS | Genome-wide association study |
| $h^2$SNP | SNP-based heritability |
| IV | Instrumental variable |

| Abbreviation | Full Term |
|---|---|
| LAVA | Local Analysis of [co]Variant Association |
| LD | Linkage disequilibrium |
| LDSC | Linkage disequilibrium score regression |
| LOAD | Late-onset Alzheimer's disease |
| MR | Mendelian randomization |
| Neff | Effective sample size |
| OR | Odds ratio |
| PC | Principal component |
| PIP | Posterior inclusion probability |
| PMID | PubMed identifier |
| PP | Posterior probability |
| PRS | Polygenic risk score |
| QC | Quality control |
| Rg | Genetic correlation |
| SE | Standard error |
| SMD | Standardized mean difference |
| SNP | Single nucleotide polymorphism |
| SuSiE | Sum of Single Effects |
| λGC | Genomic inflation factor |

**Supplementary Table S1. Summary of Alzheimer's Disease and eGFR GWAS Data Sources.**

This table provides comprehensive metadata for the genome-wide association study (GWAS) summary statistics utilized in this study. Data are stratified by ancestry (European [EUR] and African American [AFR]) and trait (Alzheimer's disease [AD] and estimated glomerular filtration rate [eGFR]). For each GWAS dataset, the table reports the number of single nucleotide polymorphisms (SNPs) that passed quality control procedures, sample sizes including the number of cases and controls for AD or total sample size for eGFR, the effective sample size (Neff), and complete citation information including PubMed identifiers (PMIDs) and primary authors. The Kunkle dataset represents AD GWAS corrected for competing risk according to the method section, while the Wightman dataset serves as a validation cohort. This table establishes the foundational data sources for all subsequent genetic correlation, pleiotropy, and causal inference analyses.

| Ancestry | Traits | Trait Description | No. of SNPS passed QC | No. of Cases | No. of Controls | Neff | Total | Source (PMID) | Author(s) |
|---|---|---|---|---|---|---|---|---|---|
| EUR | AD | late-onset Alzheimer's disease (LOAD) | 9,649,357 | 21,982 | 41,944 | 57,692 | 63,926 | 30820047 | Kunkle et al. |
| | AD | late-onset Alzheimer's disease (LOAD) | 8,113,049 | 39,918 | 358,140 | 143,663 | 398,058 | 34493870 | Wightman et al. |
| | eGFR | creatinine-based estimated glomerular filtration rate per 2021 CKD-EPI formula | 8,097,136 | | | | 1,482,631 | 34272381, 31451708 | Stanzick et al. and Hellwege et al. |
| AFR | AD | late-onset Alzheimer's disease (LOAD) | 29,750,779 | 2,903 | 6,265 | 7,935 | 9,168 | 38958117 | Ray et. al. |
| *AFR + AMS(27.5%) | eGFR | creatinine-based estimated glomerular filtration rate per 2009 CKD-EPI formula | 14,755,831 | | | | 145,732 | 38190104 | Hughes et. al. |

**Supplementary Table S2. Individual-Level Genotype Data from ADGC MEA Dataset for PRS Analysis.** This table summarizes the individual-level genotype data obtained from the Alzheimer's Disease Genetics Consortium (ADGC) Multi-Ethnic GWAS Array (MEA) dataset, which was used to calculate and validate polygenic risk scores (PRS). The data are stratified by ancestry (EUR and AFR) and diagnostic status (healthy controls versus AD cases). For each ancestry group, demographic and genetic characteristics are reported, including sample sizes with proportions, mean age at AD onset/exam distribution (mean ± standard deviation), sex distribution, apolipoprotein E (APOE) ε4 allele dosage, and the first three principal components (PC1, PC2, PC3) of genetic ancestry. Statistical comparisons between AD cases and controls were performed using chi-square tests for categorical variables, t-tests for continuous variables with normal distributions, and non-parametric tests for variables that were not normally distributed. Standardized mean differences (SMD) are reported to quantify effect sizes. These data allow the evaluation of whether genetic liability for reduced kidney function, as captured by eGFR-PRS, is associated with AD risk after adjusting for demographic and genetic factors.

| | | **Healthy Controls** | **AD** | **P value** | **SMD** |
|---|---|---|---|---|---|
| EUR Ancestry | N (%) | 16,252 (51.9) | 15,063 (48.1) | | |
| | Age (mean (SD)) | 76.35 (8.25) | 73.41 (8.49) | <0.001 * | 0.351 |
| | Sex (%) | | | | |
| | Male | 6,551 (40.3) | 6,431 (42.7) | <0.001 * | 0.048 |
| | Female | 9,701 (59.7) | 8,632 (57.3) | | |
| | APOE ε4 dosage (%) | | | <0.001 * | 0.775 |
| | 0 copies | 12,021 (74.0) | 5,980 (39.7) | | |
| | 1 copy | 3,906 (24.0) | 7,084 (47.0) | | |
| | 2 copies | 325 ( 2.0) | 1,999 (13.3) | | |
| AFR Ancestry | N (%) | 1,529 (70.4) | 643 (29.6) | | |
| | Age (mean (SD)) | 72.54 (7.87) | 74.12 (7.79) | <0.001 * | 0.201 |
| | Sex (%) | | | | |
| | Male | 343 (22.4) | 183 (28.5) | 0.003 * | 0.139 |
| | Female | 1,186 (77.6) | 460 (71.5) | | |
| | APOE ε4 dosage (%) | | | <0.001 * | 0.637 |
| | 0 copies | 961 (62.9) | 227 (35.3) | | |
| | 1 copy | 527 (34.5) | 327 (50.9) | | |
| | 2 copies | 41 ( 2.7) | 89 (13.8) | | |

**Supplementary Table S3. Genome-Wide Genetic Correlation Analysis Using LD Score Regression.**
This table presents the results of linkage disequilibrium score regression (LDSC) analysis, which estimates genome-wide SNP-based heritability ($h^2SNP$) for each trait and calculates the global genetic correlation (rg) between AD and eGFR across both European and African American ancestries. For each trait-ancestry combination, the table reports heritability estimates on the liability scale with standard errors, mean chi-square statistics (Mean $\chi^2$), genomic inflation factors ($\lambda GC$), and LDSC intercepts that distinguish polygenic signal from population stratification. The genetic correlation (rg) between AD and eGFR is reported with its standard error, z-score, and p-value, along with the genetic covariance intercept.

| | | **Heritability of Phenotype** | | | | **Genetic Correlation** | | | **Genetic Covariance** |
|---|---|---|---|---|---|---|---|---|---|
| | **Phenotype** | **H2LDSC (SE)** | **Mean χ2** | **λGC** | **Intercept (SE)** | **Rg (SE)** | **Z-score** | **P** | **Intercept (SE)** |
| EUR Discovery | AD | 0.0536 (0.0446) | 1.1216 | 1.0932 | 1.0738 (0.0505) | -0.0707 (0.0481) | -1.4715 | 0.1411 | 0.0064 (0.0086) |
| | eGFR | 0.0765 (0.0053) | 2.969 | 1.6292 | 0.9797 (0.0559) | | | | |
| EUR Discovery Corrected | AD | 0.0507 (0.0464) | 1.1203 | 1.0911 | 1.075 (0.052) | -0.059 (0.0503) | -1.1709 | 0.2416 | 0.0054 (0.0089) |
| | eGFR | 0.0759 (0.0053) | 2.9703 | 1.6306 | 0.9852 (0.059) | | | | |
| EUR Validation | AD | 0.0145 (0.0423) | 1.1698 | 1.099 | 1.1336 (0.1211) | 0.0016 (0.0192) | 0.0809 | 0.9355 | 0.0008 (0.0036) |
| | eGFR | 0.0766 (0.0052) | 1.856 | 1.6292 | 0.9781 (0.0555) | | | | |
| EUR Validation Corrected | AD | 0.0115 (0.0441) | 1.1654 | 1.095 | 1.1359 (0.1258) | 0.003 (0.0836) | 0.036 | 0.9713 | -0.0091 (0.0158) |
| | eGFR | 0.0758 (0.0054) | 2.9711 | 1.6306 | 0.9867 (0.0598) | | | | |
| AFR | AD | -0.0014 (0.0296) | 0.9501 | 0.9518 | 0.9401 (0.0041) | na | na | na | 0.0051 (0.003) |
| | eGFR | 0.0157 (0.005) | 1.0832 | 1.0679 | 1.0487 (0.0051) | | | | |

**Supplementary Table S4. Quality Control Metrics for Polygenic Risk Score Calculation.**
This table documents the comprehensive quality control (QC) pipeline applied to GWAS summary statistics prior to polygenic risk score (PRS) calculation using the SBayesRC method. QC metrics are reported separately for African American (AFR) and European (EUR) ancestry groups. The table tracks the number of SNPs at each stage of the QC process, including total SNPs in the original eGFR GWAS summary statistics pre-SBayesRC, SNPs remaining after standard QC filters, SNPs in LD reference panel overlap, and final SNPs used in PRS calculation post-SBayesRC.

| QC Metric | AFR | EUR |
|---|---|---|
| Total SNPs in eGFR GWAS summary statistics (pre-SBayesRC) | 14,755,738 | 8,097,045 |
| SNPs with weights estimated by SBayesRC | 6,056,626 | 7,329,491 |
| Total SNPs in individual-level genotype data | 133,245,772 | 161,248,802 |
| SNPs excluded due to missing variant IDs | 128,664,954 | 156,255,899 |
| SNPs used for PRS calculation (matched between SNP weights and individual-level data) | **4,580,818** | **4,992,903** |

**Supplementary Table S5. Polygenic Risk Score Categorization Approaches and Cutoff Definitions.**
This table defines the various approaches used to categorize continuous polygenic risk scores (PRS) into discrete risk groups for association analysis with AD. Three complementary categorization strategies are presented: quartiles (dividing the PRS distribution into four equal groups, Q1-Q4), tertiles (dividing into three equal groups), and extreme groups (comparing the highest 20% versus the lowest 20% of the PRS distribution). For each approach, the table specifies the PRS cutoff values in the European ancestry sample that define group boundaries and provides interpretative labels for each category. Quartiles allow for assessment of dose-response relationships across the full PRS spectrum, tertiles provide increased statistical power with fewer comparisons, and extreme groups maximize contrast between high and low genetic risk individuals. These multiple categorization approaches enable sensitivity analyses to determine whether associations between eGFR-PRS and AD risk are consistent across different modeling strategies.

| Approach | PRS Category | EUR_Cutoff | Interpretation |
|---|---|---|---|
| Quartile | Q1 (Lowest 25%) | -4.102 | Lowest risk group |
| Quartile | Q2 (25-50%) | -0.677 | Low-moderate risk |
| Quartile | Q3 (50-75%) | -0.009 | Moderate-high risk |
| Quartile | Q4 (Highest 25%) | 0.668 | Highest risk group |
| SD-based | Low (<-0.5 SD) | | Below average risk |
| SD-based | Average (-0.5 to +0.5 SD) | | Average risk |
| SD-based | High (+0.5 to +1.5 SD) | | Above average risk |
| SD-based | Very High (>+1.5 SD) | | High risk |
| Tertile | Low (Bottom 33%) | -4.102 | Low risk |
| Tertile | Medium (33-67%) | -0.434 | Moderate risk |
| Tertile | High (Top 33%) | 0.427 | High risk |

**Supplementary Table S6. Association of eGFR Polygenic Risk Scores with Alzheimer's Disease Risk in European Ancestry.**
This table presents the results of logistic regression analyses testing the association between eGFR polygenic risk scores (eGFR-PRS) and AD risk in individuals of European ancestry (N=31,315). Analyses are stratified by PRS categorization approach (quartiles, tertiles, extreme groups, and continuous per standard deviation increase) and by adjustment for APOE ε4 dosage, the strongest genetic risk factor for AD. For each comparison, the table reports regression coefficients (Beta), odds ratios (OR) with 95% confidence intervals, and p-values. Results are

shown both with and without adjustment for APOE ε4 dosage, age, sex, and the first three principal components of genetic ancestry.

| eGFR-PRS Grouping | Comparison of eGFR-PRS | APOE Adjustment | Beta (SE) | OR (95% CI) | P-value |
|---|---|---|---|---|---|
| Quartiles | Q2 vs. Q1 | Yes | 0.021 (0.035) | 1.02 (0.95 - 1.09) | 0.549 |
| | Q3 vs. Q1 | | -0.006 (0.035) | 0.99 (0.93 - 1.06) | 0.861 |
| | Q4 vs. Q1 | | -0.046 (0.035) | 0.95 (0.89 - 1.02) | 0.182 |
| | Q2 vs. Q1 | No | 0.022 (0.032) | 1.02 (0.96 - 1.09) | 0.494 |
| | Q3 vs. Q1 | | 0.004 (0.032) | 1.00 (0.94 - 1.07) | 0.897 |
| | Q4 vs. Q1 | | -0.056 (0.033) | 0.95 (0.89 - 1.01) | 0.085 |
| SD-based | Average vs. Low | Yes | 0.005 (0.029) | 1.01 (0.95 - 1.06) | 0.859 |
| | High vs. Low | | -0.059 (0.033) | 0.94 (0.88 - 1.01) | 0.078 |
| | Very High vs. Low | | -0.040 (0.052) | 0.96 (0.87 - 1.06) | 0.441 |
| | Average vs. Low | No | 0.019 (0.028) | 1.02 (0.97 - 1.08) | 0.495 |
| | High vs. Low | | -0.054 (0.031) | 0.95 (0.89 - 1.01) | 0.084 |
| | Very High vs. Low | | -0.067 (0.049) | 0.94 (0.85 - 1.03) | 0.172 |
| Tertiles | Medium vs. Low | Yes | 0.019 (0.030) | 1.02 (0.96 - 1.08) | 0.519 |
| | High vs. Low | | -0.055 (0.030) | 0.95 (0.89 - 1.00) | 0.064 |
| | Medium vs. Low | No | 0.027 (0.028) | 1.03 (0.97 - 1.09) | 0.342 |
| | High vs. Low | | -0.060 (0.028) | 0.94 (0.89 - 1.00) | 0.033 * |
| Continuous | | Yes | -0.018 (0.012) | 0.98 (0.96 - 1.01) | 0.149 |
| | | No | -0.021 (0.012) | 0.98 (0.96 - 1.00) | 0.075 |

**Supplementary Table S7. Association of eGFR Polygenic Risk Scores with Alzheimer's Disease Risk in African American Ancestry.**

This table presents parallel analyses to Table S6, examining the association between eGFR polygenic risk scores and AD risk specifically in individuals of African American ancestry. Given the smaller sample size in the African American cohort and potential differences in linkage disequilibrium structure compared to European ancestry, these analyses are critical for assessing whether genetic relationships between kidney function and AD are ancestry-specific or generalizable across populations. Similar to the European ancestry results, associations are tested using multiple PRS categorization strategies and with or without APOE ε4 adjustment. All models adjust for age, sex, and the first three principal components of genetic ancestry.

| eGFR-PRS Grouping | Comparison of eGFR-PRS | APOE Adjustment | Beta | OR (95% CI) | P-value |
|---|---|---|---|---|---|
| Continuous (per 1 SD increase in eGFR-PRS) | Higher vs. lower | Yes | 0.014 | 1.01 (0.92 – 1.12) | 0.791 |
| | Higher vs. lower | No | -0.035 | 0.97 (0.88 – 1.06) | 0.472 |
| Tertiles (EUR-derived) | Medium vs. Low | Yes | 0.016 | 1.02 (0.81 – 1.28) | 0.891 |
| | High vs. Low | Yes | -0.002 | 1 (0.79 – 1.28) | 0.988 |
| | Medium vs. Low | No | 0.002 | 1.00 (0.80 – 1.25) | 0.989 |
| | High vs. Low | No | -0.135 | 0.87 (0.69 – 1.11) | 0.262 |
| Tertiles (AFR-specific) | Medium vs. Low | Yes | 0.006 | 1.01 (0.79 – 1.28) | 0.963 |
| | High vs. Low | Yes | 0.089 | 1.09 (0.86 – 1.4) | 0.473 |

**Supplementary Table S8. Significant Local Genetic Correlations Between AD and eGFR in European Ancestry (Kunkle Discovery).**
This table reports genomic loci exhibiting statistically significant local genetic correlations between AD and eGFR in the European ancestry discovery dataset (Kunkle corrected for competing risk). Local Analysis of [co]Variant Association (LAVA) was used to partition the genome into approximately 2,500 independent LD blocks and test for local genetic correlations within each block. Only loci meeting the Benjamini-Hochberg false discovery rate (FDR) threshold of 0.05 are included. For each significant locus, the table provides: locus identifier, traits analyzed, chromosomal location (chromosome, start and stop positions), number of SNPs in the locus, estimated local genetic correlation (rho) with 95% confidence interval, squared correlation coefficient ($R^2$), and p-value. The direction of correlation (positive or negative) indicates whether genetic variants increasing AD risk also increase or decrease eGFR within that specific genomic region. Note: 0 indicates "No", and 1 indicates "Yes".

| locus | Trait1 | Trait2 | chr | start | stop | n.snps | Rg (95% CI) | R2 (95% CI) | P | PFDR | Concordant with global Rg |
|---|---|---|---|---|---|---|---|---|---|---|---|
| 1 | AD Kunkle Corrected | eGFR | 19 | 45,040,933 | 45,893,307 | 2,065 | -0.403 (-0.500, -0.308) | 0.162 (0.095, 0.250) | 4.059E-15 | 3.178E-12 | 1 |
| 2 | | | 8 | 102,548,681 | 103,716,053 | 2,738 | -0.802 (-1.000, -0.442) | 0.643 (0.195, 1.000) | 5.756E-05 | 2.253E-02 | 1 |
| 3 | | | 11 | 95,327,211 | 96,150,134 | 2,019 | 0.753 (0.415, 1.000) | 0.567 (0.172, 1.000) | 7.278E-05 | 1.900E-02 | 0 |
| 4 | | | 14 | 90,662,837 | 92,101,228 | 3,299 | 0.946 (0.498, 1.000) | 0.895 (0.248, 1.000) | 1.206E-04 | 2.360E-02 | 0 |
| 5 | | | 2 | 178,149,982 | 179,224,870 | 2,621 | -0.749 (-1.000, -0.369) | 0.561 (0.136, 1.000) | 3.486E-04 | 4.549E-02 | 1 |
| 6 | | | 22 | 19,635,655 | 20,969,184 | 2,345 | 0.562 (0.272, 0.928) | 0.316 (0.074, 0.862) | 3.510E-04 | 3.926E-02 | 0 |
| 7 | | | 9 | 70,939,678 | 72,288,878 | 3,117 | 0.597 (0.275, 1.000) | 0.356 (0.075, 1.000) | 3.864E-04 | 3.782E-02 | 0 |
| 8 | | | 15 | 63,218,979 | 64,339,945 | 2,725 | -0.532 (-0.901, -0.256) | 0.283 (0.065, 0.812) | 3.983E-04 | 3.465E-02 | 1 |
| 9 | | | 11 | 46,316,006 | 48,066,366 | 1,965 | 0.538 (0.259, 0.863) | 0.289 (0.067, 0.746) | 4.575E-04 | 3.582E-02 | 0 |
| 10 | | | 20 | 5,379,451 | 6,128,358 | 2,236 | -0.614 (-1.000, -0.286) | 0.377 (0.082, 1.000) | 4.730E-04 | 3.367E-02 | 1 |
| 11 | | | 20 | 60,766,558 | 61,470,768 | 1,965 | -0.670 (-1.000, -0.293) | 0.449 (0.086, 1.000) | 6.531E-04 | 4.261E-02 | 1 |
| 12 | | | 16 | 20,351,262 | 22,667,048 | 2,501 | 0.354 (0.160, 0.603) | 0.125 (0.026, 0.364) | 7.687E-04 | 4.630E-02 | 0 |
| 13 | | | 5 | 136,949,854 | 139,408,116 | 2,941 | -0.525 (-0.947, -0.235) | 0.275 (0.055, 0.897) | 8.036E-04 | 4.494E-02 | 1 |
| 14 | | | 4 | 40,590,542 | 41,360,937 | 1,973 | 1.000 (0.399, 1.000) | 1.000 (0.159, 1.000) | 8.832E-04 | 4.610E-02 | 0 |
| 15 | | | 11 | 64,594,823 | 66,782,661 | 2,884 | -0.457 (-0.916, - | 0.209 (0.036, 0.839) | 9.492E-04 | 4.645E-02 | 1 |

| locus | Trait1 | Trait2 | chr | start | stop | n.snps | Rg (95% CI) | R2 (95% CI) | P | PFDR | Concordant with global Rg |
|---|---|---|---|---|---|---|---|---|---|---|---|
| | | | | | | | 0.190) | | | | |
| 1 | AD Kunkle | eGFR | 19 | 45,040,933 | 45,893,307 | 2,205 | -0.338 (-0.430, -0.247) | 0.115 (0.061, 0.185) | 2.010E-12 | 1.530E-09 | 1 |
| 2 | | | 5 | 136,949,854 | 139,408,116 | 3,343 | -0.621 (-1.000, -0.338) | 0.386 (0.114, 1.000) | 4.365E-05 | 1.661E-02 | 1 |
| 3 | | | 22 | 32,662,294 | 33,501,725 | 2,645 | 1.000 (0.466, 1.000) | 0.737 (0.217, 1.000) | 5.213E-05 | 1.322E-02 | 0 |
| 4 | | | 8 | 102,548,681 | 103,716,053 | 3,029 | 0.737 (-0.000, 1.000) | 0.543 (0.138, 1.000) | 1.384E-04 | 2.633E-02 | 0 |
| 5 | | | 15 | 63,218,979 | 64,339,945 | 2,841 | -0.527 (-0.845, -0.268) | 0.278 (0.072, 0.715) | 1.795E-04 | 2.732E-02 | 1 |
| 6 | | | 2 | 178,149,982 | 179,224,870 | 2,779 | -0.630 (-1.000, -0.324) | 0.397 (0.105, 1.000) | 2.736E-04 | 3.470E-02 | 1 |
| 7 | | | 22 | 19,635,655 | 20,969,184 | 2,595 | 0.609 (0.299, 1.000) | 0.371 (0.089, 1.000) | 3.553E-04 | 3.863E-02 | 0 |

**Supplementary Table S9. Significant Local Genetic Correlations Between AD and eGFR in European Ancestry (Wightman Validation).** This table presents results from local genetic correlation analysis using the Wightman AD GWAS (corrected for competing risk) as a validation dataset, parallel to the discovery analysis in Table S8. By analyzing an independent AD GWAS, this validation approach tests whether local genetic correlations identified in the discovery phase replicate in an independent sample, thereby establishing the robustness and generalizability of findings. For each significant locus (FDR < 0.05), the table reports locus identifiers, chromosomal coordinates, number of SNPs, number of principal components used to capture local LD structure, and the estimated local genetic correlation (rho) with lower and upper bounds of the 95% confidence interval. Note: 0 indicates "No", and 1 indicates "Yes".

| locus | Trait1 | Trait2 | chr | start | stop | n.snps | n.pcs | rho | rho.lower | rho.upper | r2 | r2.lower | r2.upper | P | PFDR | Concordant with global rg |
|---|---|---|---|---|---|---|---|---|---|---|---|---|---|---|---|---|
| 1 | AD Wightman Corrected | eGFR | 19 | 45040933 | 45893307 | 1925 | 244 | -0.348 | -0.448 | -0.246 | 0.121 | 0.061 | 0.201 | 7.035E-11 | 7.759E-08 | 0 |
| 2 | | | 14 | 90662837 | 92101228 | 3137 | 353 | 0.885 | 0.556 | 1.000 | 0.784 | 0.309 | 1.000 | 1.970E-06 | 1.087E-03 | 1 |
| 3 | | | 4 | 142807915 | 144401042 | 3222 | 268 | 0.720 | 0.434 | 1.000 | 0.519 | 0.188 | 1.000 | 9.635E-06 | 3.543E-03 | 1 |
| 4 | | | 16 | 80559686 | 81437861 | 2544 | | 0.831 | 0.499 | 1.000 | 0.690 | 0.249 | 1.000 | 1.086E-05 | 2.995E-03 | 1 |
| 5 | | | 11 | 46316006 | 48066366 | 1892 | | 0.640 | 0.357 | 0.967 | 0.410 | 0.127 | 0.934 | 4.338E-05 | 9.570E-03 | 1 |
| 6 | | | 5 | 136949854 | 139408116 | 2773 | | -0.604 | -0.953 | -0.316 | 0.364 | 0.100 | 0.909 | 9.727E-05 | 1.788E-02 | 0 |
| 7 | | | 15 | 63218979 | 64339945 | 2622 | | -0.558 | -0.899 | -0.291 | 0.311 | 0.085 | 0.807 | 1.054E-04 | 1.660E-02 | 0 |
| 8 | | | 5 | 132554691 | 134556570 | 3340 | | -0.698 | -1.000 | -0.364 | 0.487 | 0.133 | 1.000 | 1.119E- | 1.542E- | 0 |

| locus | Trait1 | Trait2 | chr | start | stop | n.snps | n.pcs | rho | rho.lower | rho.upper | r2 | r2.lower | r2.upper | P | PFDR | Concordant with global rg |
|---|---|---|---|---|---|---|---|---|---|---|---|---|---|---|---|---|
| | | | | | | | | | | | | | | 04 | 02 | |
| 9 | | | 12 | 53039988 | 54371448 | 2766 | | -0.580 | -0.914 | -0.306 | 0.337 | 0.093 | 0.835 | 1.159E-04 | 1.420E-02 | 0 |
| 10 | | | 4 | 32746821 | 33640822 | 1694 | | -0.626 | -1.000 | -0.317 | 0.391 | 0.100 | 1.000 | 3.617E-04 | 3.989E-02 | 0 |
| 1 | AD Wightman | eGFR | 17 | 43460501 | 44865832 | 2962 | 0.293381 | 0.293 | 0.172 | 0.410 | 0.086 | 0.030 | 0.168 | 3.123E-06 | 5.268E-03 | 1 |
| 2 | | | 6 | 148184845 | 149278798 | 3577 | -0.741949 | -0.742 | -1.000 | -0.417 | 0.550 | 0.174 | 1.000 | 2.347E-05 | 1.980E-02 | 0 |
| 3 | | | 16 | 29043178 | 31384210 | 2767 | 0.378548 | 0.379 | 0.203 | 0.560 | 0.143 | 0.041 | 0.314 | 5.373E-05 | 3.022E-02 | 1 |
| 4 | | | 4 | 23858351 | 24764724 | 2219 | 0.506822 | 0.507 | 0.280 | 0.772 | 0.257 | 0.078 | 0.596 | 5.671E-05 | 2.392E-02 | 1 |
| 5 | | | 2 | 240325601 | 241113279 | 2812 | -0.481551 | -0.482 | -0.780 | -0.243 | 0.232 | 0.059 | 0.608 | 1.126E-04 | 3.800E-02 | 0 |
| 6 | | | 12 | 43980532 | 44923825 | 2040 | 0.466734 | 0.467 | 0.235 | 0.713 | 0.218 | 0.055 | 0.508 | 1.394E-04 | 3.919E-02 | 1 |
| 7 | | | 18 | 38498382 | 39410165 | 2078 | -1 | -1.000 | -1.000 | -0.503 | 1.000 | 0.253 | 1.000 | 1.963E-04 | 4.730E-02 | 0 |
| 8 | | | 10 | 107877788 | 108725997 | 2321 | 0.53031 | 0.530 | 0.260 | 0.824 | 0.281 | 0.067 | 0.679 | 2.327E-04 | 4.906E-02 | 1 |

**Supplementary Table S10. Significant Local Genetic Correlations Between AD and eGFR in African American Ancestry.**
This table reports genomic loci with significant local genetic correlations between AD and eGFR specifically in African American ancestry (FDR < 0.05). Given the distinct linkage disequilibrium structure, allele frequencies, and potentially different genetic architectures in African ancestry populations compared to European ancestry, ancestry-specific analyses are essential for comprehensive understanding of genetic pleiotropy. The table includes locus identifiers, traits analyzed, chromosomal positions, number of SNPs, estimated local genetic correlations (rg) with 95% confidence intervals, p-values, and number of principal components used to model local LD structure. Note: 0 indicates "No", and 1 indicates "Yes".

| locus | Trait1 | Trait2 | chr | start | stop | n.snps | rho | rho.lower | rho.upper | r2 | r2.lower | r2.upper | P | PFDR |
|---|---|---|---|---|---|---|---|---|---|---|---|---|---|---|
| 1 | AD Ray | eGFR | 3 | 189757981 | 190357798 | 3382 | 1.000 | 0.642 | 1.000 | 1.000 | 0.412 | 1.000 | 1.247E-08 | 1.152E-05 |
| 2 | | | 19 | 46508672 | 47544622 | 4529 | -0.938 | -1.000 | -0.587 | 0.880 | 0.345 | 1.000 | 8.331E-07 | 3.849E-04 |
| 3 | | | 13 | 59706604 | 61268194 | 6321 | -1.000 | -1.000 | -0.522 | 1.000 | 0.272 | 1.000 | 6.338E-06 | 1.952E-03 |
| 4 | | | 3 | 142567286 | 143976384 | 6678 | 1.000 | 0.472 | 1.000 | 1.000 | 0.223 | 1.000 | 8.993E-06 | 2.077E-03 |
| 5 | | | 2 | 129313012 | 129915501 | 3452 | 1.000 | 0.530 | 1.000 | 1.000 | 0.281 | 1.000 | 2.516E-05 | 4.650E-03 |
| 6 | | | 15 | 46833894 | 48301725 | 6328 | 1.000 | 0.485 | 1.000 | 1.000 | 0.235 | 1.000 | 3.432E-05 | 5.286E-03 |
| 7 | | | 6 | 39814082 | 40550913 | 3748 | 1.000 | 0.496 | 1.000 | 1.000 | 0.246 | 1.000 | 4.022E-05 | 5.310E-03 |
| 8 | | | 3 | 132061684 | 132880614 | 3734 | 0.901 | 0.481 | 1.000 | 0.812 | 0.232 | 1.000 | 7.458E-05 | 8.614E-03 |
| 9 | | | 3 | 168302338 | 169190677 | 3439 | 0.836 | 0.432 | 1.000 | 0.699 | 0.187 | 1.000 | 9.319E-05 | 9.567E-03 |
| 10 | | | 4 | 88480371 | 89180351 | 3290 | 1.000 | 0.506 | 1.000 | 1.000 | 0.256 | 1.000 | 9.930E-05 | 9.176E-03 |
| 11 | | | 1 | 15102430 | 15747105 | 3820 | -0.978 | -1.000 | -0.485 | 0.957 | 0.236 | 1.000 | 1.558E-04 | 1.309E-02 |
| 12 | | | 9 | 12044381 | 12797991 | 4146 | 0.976 | 0.490 | 1.000 | 0.953 | 0.240 | 1.000 | 1.822E-04 | 1.403E-02 |
| 13 | | | 8 | 105982692 | 107379445 | 5592 | -0.992 | -1.000 | -0.487 | 0.985 | 0.237 | 1.000 | 2.065E-04 | 1.468E-02 |
| 14 | | | 2 | 223561337 | 224269422 | 3434 | 0.932 | 0.468 | 1.000 | 0.869 | 0.219 | 1.000 | 2.133E-04 | 1.407E-02 |
| 15 | | | 8 | 27425268 | 28158440 | 3949 | 0.553 | 0.264 | 0.909 | 0.305 | 0.070 | 0.827 | 3.969E-04 | 2.445E-02 |

| locus | Trait1 | Trait2 | chr | start | stop | n.snps | rho | rho.lower | rho.upper | r2 | r2.lower | r2.upper | P | PFDR |
|---|---|---|---|---|---|---|---|---|---|---|---|---|---|---|
| 16 | | | 2 | 126680360 | 127770280 | 5665 | -0.897 | -1.000 | -0.419 | 0.805 | 0.176 | 1.000 | 3.999E-04 | 2.309E-02 |
| 17 | | | 2 | 207726268 | 208825721 | 4160 | -0.830 | -1.000 | -0.390 | 0.689 | 0.152 | 1.000 | 4.113E-04 | 2.235E-02 |
| 18 | | | 1 | 164440353 | 165828905 | 6604 | -0.793 | -1.000 | -0.365 | 0.628 | 0.133 | 1.000 | 5.027E-04 | 2.581E-02 |
| 19 | | | 2 | 28802271 | 29633040 | 4064 | 0.712 | 0.326 | 1.000 | 0.507 | 0.106 | 1.000 | 5.544E-04 | 2.696E-02 |
| 20 | | | 3 | 167521068 | 168302337 | 3313 | 0.388 | 0.172 | 0.620 | 0.151 | 0.030 | 0.385 | 5.780E-04 | 2.671E-02 |
| 21 | | | 9 | 120600042 | 121473024 | 4013 | -0.797 | -1.000 | -0.359 | 0.635 | 0.129 | 1.000 | 7.592E-04 | 3.340E-02 |
| 22 | | | 17 | 63346710 | 64331974 | 4964 | 0.732 | 0.321 | 1.000 | 0.536 | 0.103 | 1.000 | 9.179E-04 | 3.855E-02 |
| 23 | | | 2 | 166395569 | 167373038 | 4156 | 0.828 | 0.367 | 1.000 | 0.686 | 0.135 | 1.000 | 9.674E-04 | 3.887E-02 |
| 24 | | | 10 | 22542648 | 23839537 | 5498 | -0.973 | -1.000 | -0.413 | 0.946 | 0.171 | 1.000 | 1.008E-03 | 3.880E-02 |
| 25 | | | 10 | 123853589 | 124868063 | 4785 | 0.888 | 0.367 | 1.000 | 0.789 | 0.135 | 1.000 | 1.099E-03 | 4.062E-02 |
| 26 | | | 3 | 104829898 | 105669644 | 3507 | 0.801 | 0.333 | 1.000 | 0.642 | 0.111 | 1.000 | 1.163E-03 | 4.135E-02 |
| 27 | | | 2 | 212383518 | 213059448 | 3506 | 0.684 | 0.283 | 1.000 | 0.468 | 0.080 | 1.000 | 1.325E-03 | 4.535E-02 |
| 28 | | | 2 | 64623202 | 65712249 | 4852 | 0.559 | 0.227 | 1.000 | 0.313 | 0.052 | 1.000 | 1.399E-03 | 4.615E-02 |
| 29 | | | 15 | 22881864 | 24199319 | 2764 | -0.819 | -1.000 | -0.329 | 0.670 | 0.108 | 1.000 | 1.473E-03 | 4.694E-02 |
| 30 | | | 11 | 30688687 | 32065472 | 4215 | 0.514 | 0.201 | 0.898 | 0.265 | 0.040 | 0.806 | 1.535E-03 | 4.726E-02 |
| 31 | | | 5 | 56326600 | 57441632 | 4873 | -0.601 | -1.000 | -0.238 | 0.361 | 0.057 | 1.000 | 1.633E-03 | 4.869E-02 |

**Supplementary Table S11. Summary Statistics of Local Genetic Correlation Analysis Across Ancestries.**
This table provides summary statistics comparing LAVA results across ancestries (EUR and AFR) and AD GWAS datasets (Kunkle discovery and Wightman validation). For each ancestry-dataset combination, the table reports: the number of genomic regions demonstrating significant univariate heritability for eGFR alone, for AD alone, the total number of regions tested in bivariate LAVA analysis (requiring significant heritability in both traits), the number of regions showing statistically significant local genetic correlations (FDR < 0.05), and among significant correlations, the number showing positive versus negative correlation directions.

| | **Genomic regions in univariate LAVA** | | **Genomic regions in eGFR & AD bivariate LAVA** | | | |
|---|---|---|---|---|---|---|
| **Ancestry** | **Trait 1 (eGFR)** | **Trait 2 (AD)** | **Total** | **Significant (PFDR <0.05)** | **Positive local Rg** | **Negative local Rg** |
| EUR | 2,224 | Kunkle: 1,975 | 761 | 7 | 2 | 5 (71.43%) |
| | 2,206 | Kunkle Corrected: 1,979 | 781 | 15 | 7 | 8 (53.33%) |
| | 2,342 | Wightman: 2,286 | 1,687 | 8 | 5 | 3 (37.50%) |
| | 1,933 | Wightman Corrected: 2,339 | 1,103 | 15 | 7 | 9 (60.00%) |
| AFR | 2,390 | Ray: 2,261 | 923 | 31 | 20 | 11 (35.48%) |

**Supplementary Table S12. Cross-Ancestry Comparison of Significant Local Genetic Correlations.**
This table systematically categorizes all genomic loci identified in LAVA analysis according to their ancestry-specific patterns: EUR-only significant, AFR-only significant, or significant in both ancestries. For loci appearing in multiple categories, the table reports the direction of correlation (positive or negative) in each ancestry, FDR-adjusted p-values, unadjusted p-values, and the estimated local genetic correlation (rg) in each ancestry.

| **Locus** | **category** | **Direction_EUR** | **Direction_AFR** | **FDR_EUR** | **FDR_AFR** | **P_value_EUR** | **P_value_AFR** | **rho_EUR** | **rho_AFR** |
|---|---|---|---|---|---|---|---|---|---|
| Chr10:124.4 Mb | AFR-only | | 1 | | 4.057E-02 | | 1.099E-03 | | 0.888 |
| Chr10:23.2Mb | AFR-only | | -1 | | 3.876E-02 | | 1.008E-03 | | -0.973 |
| Chr11:31.4Mb | AFR-only | | 1 | | 4.721E-02 | | 1.535E-03 | | 0.514 |
| Chr11:47.2Mb | EUR-only | 1 | | 3.699E-02 | | 4.575E-04 | | 0.538 | |
| Chr11:65.7Mb | EUR-only | -1 | | 4.948E-02 | | 9.492E-04 | | -0.457 | |
| Chr11:95.7Mb | EUR-only | 1 | | 1.897E-02 | | 7.278E-05 | | 0.753 | |
| Chr13:60.5Mb | AFR-only | | -1 | | 1.950E-03 | | 6.338E-06 | | -1.000 |
| Chr14:91.4Mb | EUR-only | 1 | | 2.357E-02 | | 1.206E-04 | | 0.946 | |

| Locus | category | Direction_EUR | Direction_AFR | FDR_EUR | FDR_AFR | P_value_EUR | P_value_AFR | rho_EUR | rho_AFR |
|---|---|---|---|---|---|---|---|---|---|
| Chr15:23.5Mb | AFR-only | | -1 | | 4.689E-02 | | 1.473E-03 | | -0.819 |
| Chr15:47.6Mb | AFR-only | | 1 | | 5.280E-03 | | 3.432E-05 | | 1.000 |
| Chr15:63.8Mb | EUR-only | -1 | | 3.699E-02 | | 3.983E-04 | | -0.532 | |
| Chr16:21.5Mb | EUR-only | 1 | | 4.834E-02 | | 7.687E-04 | | 0.354 | |
| Chr17:63.8Mb | AFR-only | | 1 | | 3.851E-02 | | 9.179E-04 | | 0.732 |
| Chr19:45.5Mb | EUR-only | -1 | | 3.174E-12 | | 4.059E-15 | | -0.403 | |
| Chr19:47Mb | AFR-only | | -1 | | 3.845E-04 | | 8.331E-07 | | -0.938 |
| Chr1:15.4Mb | AFR-only | | -1 | | 1.307E-02 | | 1.558E-04 | | -0.978 |
| Chr1:165.1Mb | AFR-only | | -1 | | 2.578E-02 | | 5.027E-04 | | -0.793 |
| Chr20:5.8Mb | EUR-only | -1 | | 3.699E-02 | | 4.730E-04 | | -0.614 | |
| Chr20:61.1Mb | EUR-only | -1 | | 4.643E-02 | | 6.531E-04 | | -0.670 | |
| Chr22:20.3Mb | EUR-only | 1 | | 3.699E-02 | | 3.510E-04 | | 0.562 | |
| Chr2:127.2Mb | AFR-only | | -1 | | 2.233E-02 | | 3.999E-04 | | -0.897 |
| Chr2:129.6Mb | AFR-only | | 1 | | 4.645E-03 | | 2.516E-05 | | 1.000 |
| Chr2:166.9Mb | AFR-only | | 1 | | 3.876E-02 | | 9.674E-04 | | 0.828 |
| Chr2:178.7Mb | EUR-only | -1 | | 3.699E-02 | | 3.486E-04 | | -0.749 | |
| Chr2:208.3Mb | AFR-only | | -1 | | 2.233E-02 | | 4.113E-04 | | -0.830 |
| Chr2:212.7Mb | AFR-only | | 1 | | 4.530E-02 | | 1.325E-03 | | 0.684 |
| Chr2:223.9Mb | AFR-only | | 1 | | 1.406E-02 | | 2.133E-04 | | 0.932 |
| Chr2:29.2Mb | AFR-only | | 1 | | 2.668E-02 | | 5.544E-04 | | 0.712 |
| Chr2:65.2Mb | AFR-only | | 1 | | 4.610E-02 | | 1.399E-03 | | 0.559 |
| Chr3:105.2Mb | AFR-only | | 1 | | 4.130E-02 | | 1.163E-03 | | 0.801 |
| Chr3:132.5Mb | AFR-only | | 1 | | 8.604E-03 | | 7.458E-05 | | 0.901 |
| Chr3:143.3Mb | AFR-only | | 1 | | 2.075E-03 | | 8.993E-06 | | 1.000 |
| Chr3:167.9Mb | AFR-only | | 1 | | 2.668E-02 | | 5.780E-04 | | 0.388 |
| Chr3:168.7Mb | AFR-only | | 1 | | 9.166E-03 | | 9.319E-05 | | 0.836 |
| Chr3:190.1Mb | AFR-only | | 1 | | 1.151E-05 | | 1.247E-08 | | 1.000 |
| Chr4:41Mb | EUR-only | 1 | | 4.933E-02 | | 8.832E-04 | | 1.000 | |
| Chr4:88.8Mb | AFR-only | | 1 | | 9.166E-03 | | 9.930E-05 | | 1.000 |

| Locus | category | Direction_EUR | Direction_AFR | FDR_EUR | FDR_AFR | P_value_EUR | P_value_AFR | rho_EUR | rho_AFR |
|---|---|---|---|---|---|---|---|---|---|
| Chr5:138.2Mb | EUR-only | -1 | | 4.834E-02 | | 8.036E-04 | | -0.525 | |
| Chr5:56.9Mb | AFR-only | | -1 | | 4.863E-02 | | 1.633E-03 | | -0.601 |
| Chr6:40.2Mb | AFR-only | | 1 | | 5.304E-03 | | 4.022E-05 | | 1.000 |
| Chr8:103.1Mb | EUR-only | -1 | | 1.897E-02 | | 5.756E-05 | | -0.802 | |
| Chr8:106.7Mb | AFR-only | | -1 | | 1.406E-02 | | 2.065E-04 | | -0.992 |
| Chr8:27.8Mb | AFR-only | | 1 | | 2.233E-02 | | 3.969E-04 | | 0.553 |
| Chr9:12.4Mb | AFR-only | | 1 | | 1.401E-02 | | 1.822E-04 | | 0.976 |
| Chr9:121Mb | AFR-only | | -1 | | 3.337E-02 | | 7.592E-04 | | -0.797 |
| Chr9:71.6Mb | EUR-only | 1 | | 3.699E-02 | | 3.864E-04 | | 0.597 | |

**Supplementary Table S13.** Complete Local Genetic Correlation Results in European Ancestry (Kunkle Discovery). This comprehensive table presents the complete set of local genetic correlation analysis results for all genomic loci tested in European ancestry using the Kunkle AD GWAS (corrected). Unlike Table S8 which includes only statistically significant loci, this table reports results for all approximately 2,500 genomic regions genome-wide, regardless of statistical significance. For each locus, the table provides: locus label and number, chromosomal coordinates, number of SNPs, number of principal components used, phenotypes analyzed, estimated local genetic correlation (rho) with standard error, test statistics, p-value, and false discovery rate.

| locus_label | locus | chr | start | stop | n.snps | n.pcs | phen1 | phen2 | rho | rho.lower | rho.upper | r2 | r2.lower | r2.upper | p | rank | FDR |
|---|---|---|---|---|---|---|---|---|---|---|---|---|---|---|---|---|---|
| Chr19:45.5Mb | 2351 | 19 | 45040933 | 45893307 | 2065 | 284 | AD | eGFR | -0.402723 | -0.50029 | -0.30754 | 0.162186 | 0.09458 | 0.25029 | 4.06E-15 | 1 | 3.17E-12 |
| Chr8:103.1Mb | 1331 | 8 | 102548681 | 103716053 | 2738 | 270 | AD | eGFR | -0.801759 | -1 | -0.44182 | 0.642818 | 0.1952 | 1 | 5.76E-05 | 2 | 0.0189712418 |
| Chr11:95.7Mb | 1702 | 11 | 95327211 | 96150134 | 2019 | 247 | AD | eGFR | 0.753041 | 0.41506 | 1 | 0.567071 | 0.17228 | 1 | 7.28E-05 | 3 | 0.0189712418 |
| Chr14:91.4Mb | 2018 | 14 | 90662837 | 92101228 | 3299 | 300 | AD | eGFR | 0.946164 | 0.49794 | 1 | 0.895225 | 0.24794 | 1 | 0.000120578 | 4 | 0.023572999 |
| Chr2:178.7Mb | 362 | 2 | 178149982 | 179224870 | 2621 | 153 | AD | eGFR | -0.748717 | -1 | -0.3694 | 0.560577 | 0.13646 | 1 | 0.000348571 | 5 | 0.036985472 |
| Chr22:20.3Mb | 2465 | 22 | 19635655 | 20969184 | 2345 | 299 | AD | eGFR | 0.562458 | 0.27167 | 0.92835 | 0.316359 | 0.07381 | 0.86183 | 0.000350992 | 6 | 0.036985472 |
| Chr9:71.6Mb | 1418 | 9 | 70939678 | 72288878 | 3117 | 266 | AD | eGFR | 0.596974 | 0.27461 | 1 | 0.356378 | 0.07541 | 1 | 0.000386415 | 7 | 0.036985472 |
| Chr15:63.8Mb | 2068 | 15 | 63218979 | 64339945 | 2725 | 235 | AD | eGFR | -0.531517 | -0.90088 | -0.25564 | 0.282511 | 0.06535 | 0.81159 | 0.000398331 | 8 | 0.036985472 |
| Chr11:47.2Mb | 1658 | 11 | 46316006 | 48066366 | 1965 | 168 | AD | eGFR | 0.537697 | 0.25875 | 0.86349 | 0.289118 | 0.06695 | 0.74561 | 0.000457453 | 9 | 0.036985472 |
| Chr20:5.8Mb | 2375 | 20 | 5379451 | 6128358 | 2236 | 287 | AD | eGFR | -0.613903 | -1 | -0.28599 | 0.376877 | 0.08179 | 1 | 0.00047296 | 10 | 0.036985472 |
| Chr20:61.1Mb | 2427 | 20 | 60766558 | 61470768 | 1965 | 293 | AD | eGFR | -0.670331 | -1 | -0.29258 | 0.449344 | 0.0856 | 1 | 0.000653057 | 11 | 0.0464264158181818 |
| Chr16:21.5Mb | 2122 | 16 | 20351262 | 22667048 | 2501 | 267 | AD | eGFR | 0.35401 | 0.16037 | 0.60302 | 0.125323 | 0.02572 | 0.36363 | 0.000768721 | 12 | 0.048337766 |
| Chr5:138.2Mb | 891 | 5 | 136949854 | 139408116 | 2941 | 279 | AD | eGFR | -0.524841 | -0.94692 | -0.23501 | 0.275458 | 0.05523 | 0.89665 | 0.000803569 | 13 | 0.048337766 |
| Chr4:41Mb | 641 | 4 | 40590542 | 41360937 | 1973 | 223 | AD | eGFR | 1 | 0.39924 | 1 | 1 | 0.1594 | 1 | 0.000883223 | 14 | 0.0493343132857143 |
| Chr11:65.7Mb | 1674 | 11 | 64594823 | 66782661 | 2884 | 268 | AD | eGFR | -0.457388 | -0.91603 | -0.19002 | 0.209204 | 0.03611 | 0.8391 | 0.000949182 | 15 | 0.0494840216 |
| Chr12:72.6Mb | 1807 | 12 | 71948186 | 73154515 | 2079 | 211 | AD | eGFR | -0.915809 | -1 | -0.36306 | 0.838707 | 0.13182 | 1 | 0.00219376 | 16 | 0.0985711 |
| Chr2:211.7Mb | 388 | 2 | 210842458 | 212520453 | 3254 | 275 | AD | eGFR | -0.367721 | -0.67567 | -0.13663 | 0.135219 | 0.01892 | 0.45653 | 0.00222985 | 17 | 0.0985711 |
| Chr17:34Mb | 2200 | 17 | 33614190 | 34474611 | 1991 | 216 | AD | eGFR | -0.81415 | -1 | -0.31963 | 0.662841 | 0.10216 | 1 | 0.0022689 | 18 | 0.0985711 |
| Chr3:187.3M | 584 | 3 | 18660204 | 18793919 | 3449 | 358 | AD | eGFR | 0.57983 | 0.21553 | 1 | 0.336203 | 0.04665 | 1 | 0.00262585 | 19 | 0.10807445789473 |

| locus_label | locus | chr | start | stop | n.snps | n.pcs | phen1 | phen2 | rho | rho.lower | rho.upper | r2 | r2.lower | r2.upper | p | rank | FDR |
|---|---|---|---|---|---|---|---|---|---|---|---|---|---|---|---|---|---|
| b | | | 6 | 9 | | | | | | | | | | | | | 7 |
| Chr9:136.4Mb | 1479 | 9 | 136042491 | 136770926 | 2255 | 278 | AD | eGFR | 0.553776 | 0.20186 | 1 | 0.306668 | 0.04075 | 1 | 0.00307174 | 20 | 0.120105034 |
| Chr21:28.2Mb | 2443 | 21 | 27774351 | 28616539 | 2504 | 264 | AD | eGFR | -0.677113 | -1 | -0.24478 | 0.458482 | 0.06024 | 1 | 0.00383599 | 21 | 0.133634718709677 |
| Chr2:240.7Mb | 418 | 2 | 240325601 | 241113279 | 2543 | 304 | AD | eGFR | -0.727055 | -1 | -0.2376 | 0.528609 | 0.05683 | 1 | 0.00405171 | 22 | 0.133634718709677 |
| Chr2:17.9Mb | 222 | 2 | 17241116 | 18656325 | 3783 | 272 | AD | eGFR | -0.943341 | -1 | -0.31966 | 0.889892 | 0.10254 | 1 | 0.00416996 | 23 | 0.133634718709677 |
| Chr15:101.1Mb | 2100 | 15 | 100742118 | 101550750 | 2503 | 279 | AD | eGFR | 0.627283 | 0.21791 | 1 | 0.393483 | 0.04748 | 1 | 0.00434182 | 24 | 0.133634718709677 |
| Chr9:102.4Mb | 1446 | 9 | 101596751 | 103165725 | 2205 | 262 | AD | eGFR | 0.59672 | 0.18628 | 1 | 0.356074 | 0.03479 | 1 | 0.00449598 | 25 | 0.133634718709677 |
| Chr5:52.5Mb | 819 | 5 | 52139774 | 52837225 | 2026 | 173 | AD | eGFR | -0.495796 | -1 | -0.15814 | 0.245814 | 0.02574 | 1 | 0.00496575 | 26 | 0.133634718709677 |
| Chr17:2.1Mb | 2175 | 17 | 1480057 | 2699135 | 2649 | 325 | AD | eGFR | -0.475675 | -0.95731 | -0.15713 | 0.226267 | 0.02469 | 0.91645 | 0.00517476 | 27 | 0.133634718709677 |
| Chr2:183.8Mb | 367 | 2 | 183352291 | 184148655 | 1885 | 176 | AD | eGFR | -0.734706 | -1 | -0.24742 | 0.539793 | 0.06122 | 1 | 0.00518504 | 28 | 0.133634718709677 |
| Chr12:102.4Mb | 1832 | 12 | 101825062 | 102966633 | 2257 | 250 | AD | eGFR | 0.66162 | 0.22361 | 1 | 0.437741 | 0.05049 | 1 | 0.00520263 | 29 | 0.133634718709677 |
| Chr3:26.2Mb | 446 | 3 | 25516194 | 26905340 | 2979 | 265 | AD | eGFR | 0.545034 | 0.17343 | 1 | 0.297062 | 0.03087 | 1 | 0.00520469 | 30 | 0.133634718709677 |
| Chr3:188.7Mb | 585 | 3 | 187939200 | 189451805 | 3343 | 342 | AD | eGFR | -0.488818 | -0.91222 | -0.14827 | 0.238943 | 0.02205 | 0.83214 | 0.00529754 | 31 | 0.133634718709677 |
| Chr20:55.1Mb | 2419 | 20 | 54743695 | 55448075 | 1938 | 296 | AD | eGFR | -0.575176 | -1 | -0.18318 | 0.330828 | 0.03358 | 1 | 0.00563628 | 32 | 0.1377365925 |
| Chr10:53.5Mb | 1535 | 10 | 53129610 | 53919161 | 2346 | 235 | AD | eGFR | 0.819062 | 0.26032 | 1 | 0.670863 | 0.06859 | 1 | 0.00587348 | 33 | 0.139183677575758 |
| Chr15:39.9Mb | 2049 | 15 | 39238841 | 40604780 | 2984 | 288 | AD | eGFR | 0.46515 | 0.14087 | 1 | 0.216365 | 0.02002 | 1 | 0.00630586 | 34 | 0.14503478 |
| Chr8:29Mb | 1274 | 8 | 28344177 | 29744540 | 2719 | 307 | AD | eGFR | 0.863859 | 0.2536 | 1 | 0.746253 | 0.06566 | 1 | 0.00692258 | 35 | 0.154670216 |
| Chr6:164Mb | 1087 | 6 | 163336362 | 164687462 | 3725 | 334 | AD | eGFR | -0.765942 | -1 | -0.2145 | 0.586667 | 0.04689 | 1 | 0.00747853 | 36 | 0.162450290555556 |
| Chr6:116.9Mb | 1045 | 6 | 116132148 | 117674076 | 3364 | 215 | AD | eGFR | -1 | -1 | -0.16407 | 1 | 0.04044 | 1 | 0.00795055 | 37 | 0.168035948648649 |
| Chr16:10.7Mb | 2114 | 16 | 10351809 | 11022542 | 1850 | 271 | AD | eGFR | -0.572724 | -1 | -0.16641 | 0.328013 | 0.028 | 1 | 0.00823051 | 38 | 0.169375232105263 |
| Chr7:127.3Mb | 1206 | 7 | 126640873 | 127903738 | 2360 | 209 | AD | eGFR | 0.649714 | 0.18423 | 1 | 0.422129 | 0.03427 | 1 | 0.00864295 | 39 | 0.173302228205128 |
| Chr5:133.6Mb | 888 | 5 | 132554691 | 134556570 | 3475 | 353 | AD | eGFR | -0.563168 | -1 | -0.14478 | 0.317158 | 0.02153 | 1 | 0.00933888 | 40 | 0.182006685365854 |
| Chr4:77.3Mb | 670 | 4 | 76497359 | 78045637 | 3789 | 268 | AD | eGFR | -0.28005 | -0.53331 | -0.07357 | 0.0784282 | 0.00561 | 0.28442 | 0.00954255 | 41 | 0.182006685365854 |
| Chr1:92.4Mb | 80 | 1 | 91870060 | 92904465 | 2079 | 213 | AD | eGFR | -0.610397 | -1 | -0.15249 | 0.372584 | 0.02457 | 1 | 0.0118904 | 42 | 0.221387923809524 |

| locus_label | locus | chr | start | stop | n.snps | n.pcs | phen1 | phen2 | rho | rho.lower | rho.upper | r2 | r2.lower | r2.upper | p | rank | FDR |
|---|---|---|---|---|---|---|---|---|---|---|---|---|---|---|---|---|---|
| Chr2:46.2Mb | 251 | 2 | 45850048 | 46499326 | 1940 | 316 | AD | eGFR | 0.426077 | 0.09729 | 0.8982 | 0.181541 | 0.01019 | 0.80676 | 0.0121895 | 43 | 0.221678813953488 |
| Chr10:86Mb | 1565 | 10 | 85463485 | 86459980 | 2745 | 227 | AD | eGFR | -0.729758 | -1 | -0.18255 | 0.532546 | 0.03544 | 1 | 0.0126906 | 44 | 0.225546572727273 |
| Chr1:205.5Mb | 158 | 1 | 205009624 | 205917548 | 2342 | 249 | AD | eGFR | 0.460768 | 0.09478 | 0.93458 | 0.212307 | 0.01075 | 0.87345 | 0.0135794 | 45 | 0.235979795555556 |
| Chr12:23.4Mb | 1763 | 12 | 22928056 | 23923798 | 1993 | 273 | AD | eGFR | 0.55324 | 0.12645 | 1 | 0.306075 | 0.01734 | 1 | 0.014822 | 46 | 0.251974 |
| Chr1:71.8Mb | 61 | 1 | 70992906 | 72513119 | 2798 | 217 | AD | eGFR | -0.712848 | -1 | -0.16587 | 0.508153 | 0.02956 | 1 | 0.015668 | 47 | 0.26068885106383 |
| Chr17:77Mb | 2233 | 17 | 76596288 | 77412786 | 2548 | 340 | AD | eGFR | -0.740992 | -1 | -0.16656 | 0.549068 | 0.0312 | 1 | 0.0166753 | 48 | 0.269923056716418 |
| Chr17:35.3Mb | 2201 | 17 | 34474612 | 36116884 | 2108 | 301 | AD | eGFR | 0.474408 | 0.0925 | 1 | 0.225063 | 0.0096 | 1 | 0.0169569 | 49 | 0.269923056716418 |
| Chr3:186.2Mb | 583 | 3 | 185709997 | 186602045 | 2023 | 307 | AD | eGFR | -0.356526 | -0.79964 | -0.06652 | 0.127111 | 0.0057 | 0.63942 | 0.0178926 | 50 | 0.269923056716418 |
| Chr20:42.6Mb | 2406 | 20 | 42185672 | 43008890 | 1887 | 263 | AD | eGFR | 0.563984 | 0.11687 | 1 | 0.318078 | 0.01536 | 1 | 0.0184129 | 51 | 0.269923056716418 |
| Chr12:53.7Mb | 1791 | 12 | 53039988 | 54371448 | 2914 | 263 | AD | eGFR | -0.496433 | -1 | -0.08627 | 0.246445 | 0.01014 | 1 | 0.0187011 | 52 | 0.269923056716418 |
| Chr7:116.1Mb | 1198 | 7 | 115321302 | 116835336 | 2722 | 252 | AD | eGFR | 0.749783 | 0.16443 | 1 | 0.562175 | 0.02966 | 1 | 0.0187635 | 53 | 0.269923056716418 |
| Chr12:97.3Mb | 1828 | 12 | 96828315 | 97814021 | 2516 | 213 | AD | eGFR | 0.918292 | 0.18054 | 1 | 0.843261 | 0.04341 | 1 | 0.0188622 | 54 | 0.269923056716418 |
| Chr8:118.9Mb | 1345 | 8 | 118300069 | 119423298 | 2141 | 252 | AD | eGFR | 0.779513 | 0.13654 | 1 | 0.607641 | 0.02604 | 1 | 0.0195537 | 55 | 0.269923056716418 |
| Chr5:67.6Mb | 833 | 5 | 67096192 | 68006993 | 2135 | 266 | AD | eGFR | 0.442141 | 0.07822 | 1 | 0.195488 | 0.00698 | 1 | 0.0198003 | 56 | 0.269923056716418 |
| Chr20:62.6Mb | 2429 | 20 | 62179242 | 62963627 | 1967 | 247 | AD | eGFR | -0.354164 | -0.79521 | -0.05547 | 0.125432 | 0.00428 | 0.63296 | 0.0204528 | 57 | 0.269923056716418 |
| Chr2:0.4Mb | 201 | 2 | 10181 | 789195 | 1851 | 210 | AD | eGFR | -0.662317 | -1 | -0.12065 | 0.438663 | 0.01771 | 1 | 0.0207951 | 58 | 0.269923056716418 |
| Chr1:15.2Mb | 16 | 1 | 14720132 | 15755230 | 3063 | 343 | AD | eGFR | -0.553518 | -1 | -0.09837 | 0.306382 | 0.01218 | 1 | 0.0211973 | 59 | 0.269923056716418 |
| Chr3:4.5Mb | 426 | 3 | 4109394 | 4835083 | 1757 | 276 | AD | eGFR | 0.693824 | 0.11913 | 1 | 0.481391 | 0.01821 | 1 | 0.0213572 | 60 | 0.269923056716418 |
| Chr2:2.3Mb | 204 | 2 | 1875061 | 2680063 | 2025 | 265 | AD | eGFR | 0.310846 | 0.05099 | 0.60196 | 0.096625 | 0.0032 | 0.36236 | 0.0219638 | 61 | 0.269923056716418 |
| Chr20:1.3Mb | 2370 | 20 | 884242 | 1771285 | 2348 | 305 | AD | eGFR | 0.349947 | 0.05301 | 0.68255 | 0.122463 | 0.00403 | 0.46588 | 0.0224421 | 62 | 0.269923056716418 |
| Chr10:5.8Mb | 1492 | 10 | 5396243 | 6234718 | 2657 | 312 | AD | eGFR | 0.509233 | 0.07597 | 1 | 0.259318 | 0.00907 | 1 | 0.022454 | 63 | 0.269923056716418 |
| Chr15:58.2Mb | 2063 | 15 | 57641165 | 58712954 | 2934 | 326 | AD | eGFR | -0.427388 | -1 | -0.05356 | 0.18266 | 0.00465 | 1 | 0.0227646 | 64 | 0.269923056716418 |
| Chr5:159.1Mb | 908 | 5 | 158484776 | 159663733 | 2762 | 279 | AD | eGFR | -0.620093 | -1 | -0.10707 | 0.384516 | 0.01478 | 1 | 0.0229209 | 65 | 0.269923056716418 |
| Chr22:49.3Mb | 2493 | 22 | 48977539 | 49559207 | 2964 | 303 | AD | eGFR | -0.53321 | -1 | -0.08476 | 0.284313 | 0.01014 | 1 | 0.0230786 | 66 | 0.269923056716418 |

| locus_label | locus | chr | start | stop | n.snps | n.pcs | phen1 | phen2 | rho | rho.lower | rho.upper | r2 | r2.lower | r2.upper | p | rank | FDR |
|---|---|---|---|---|---|---|---|---|---|---|---|---|---|---|---|---|---|
| Chr21:43.4Mb | 2457 | 21 | 43081435 | 43812791 | 2550 | 339 | AD | eGFR | 0.526086 | 0.08054 | 1 | 0.276767 | 0.00962 | 1 | 0.0231264 | 67 | 0.269923056716418 |
| Chr10:6.6Mb | 1493 | 10 | 6234719 | 6897037 | 1915 | 301 | AD | eGFR | -0.400416 | -0.81137 | -0.05908 | 0.160333 | 0.00476 | 0.65833 | 0.0242141 | 68 | 0.277791333333333 |
| Chr4:68.2Mb | 663 | 4 | 67616363 | 68742859 | 2520 | 207 | AD | eGFR | 0.867639 | 0.15714 | 1 | 0.752798 | 0.03192 | 1 | 0.024511 | 69 | 0.277791333333333 |
| Chr18:42.2Mb | 2271 | 18 | 41425455 | 42974165 | 3005 | 286 | AD | eGFR | -0.429303 | -0.9677 | -0.06059 | 0.184301 | 0.00572 | 0.93644 | 0.0261414 | 70 | 0.292036782857143 |
| Chr1:87.2Mb | 75 | 1 | 86761555 | 87667572 | 1844 | 156 | AD | eGFR | -0.874331 | -1 | -0.11994 | 0.764455 | 0.02425 | 1 | 0.0273727 | 71 | 0.301485230985916 |
| Chr1:94.6Mb | 82 | 1 | 94168577 | 95122541 | 1932 | 218 | AD | eGFR | -0.583986 | -1 | -0.08642 | 0.341039 | 0.01059 | 1 | 0.0278018 | 72 | 0.301958438888889 |
| Chr9:73.1Mb | 1419 | 9 | 72288879 | 73915494 | 3825 | 246 | AD | eGFR | 0.54917 | 0.07215 | 1 | 0.301588 | 0.00908 | 1 | 0.0287394 | 73 | 0.307865901369863 |
| Chr1:201.8Mb | 155 | 1 | 201067953 | 202583884 | 3259 | 355 | AD | eGFR | 0.327391 | 0.03514 | 0.68906 | 0.107185 | 0.00248 | 0.47481 | 0.0292429 | 74 | 0.309026321621622 |
| Chr15:35.9Mb | 2046 | 15 | 35118928 | 36583327 | 3133 | 295 | AD | eGFR | 0.549795 | 0.07726 | 1 | 0.302274 | 0.00937 | 1 | 0.0302002 | 75 | 0.314887418666667 |
| Chr9:5Mb | 1377 | 9 | 4596615 | 5437591 | 2062 | 255 | AD | eGFR | -0.610359 | -1 | -0.0784 | 0.372539 | 0.00972 | 1 | 0.0307954 | 76 | 0.316868457894737 |
| Chr9:131.5Mb | 1474 | 9 | 130902728 | 132193798 | 1779 | 250 | AD | eGFR | 0.401419 | 0.04254 | 0.95018 | 0.161137 | 0.00399 | 0.90474 | 0.0314144 | 77 | 0.317021869047619 |
| Chr12:89.5Mb | 1821 | 12 | 88344765 | 90567001 | 3081 | 254 | AD | eGFR | 0.403288 | 0.0464 | 0.89156 | 0.162641 | 0.00391 | 0.79617 | 0.0321113 | 78 | 0.317021869047619 |
| Chr13:113.9Mb | 1952 | 13 | 113573119 | 114302561 | 1831 | 251 | AD | eGFR | -0.417052 | -0.97885 | -0.03021 | 0.173932 | 0.00404 | 0.95814 | 0.0327479 | 79 | 0.317021869047619 |
| Chr14:25.2Mb | 1959 | 14 | 24906057 | 25585041 | 1874 | 161 | AD | eGFR | 1 | 0.10879 | 1 | 1 | 0.03241 | 1 | 0.0327997 | 80 | 0.317021869047619 |
| Chr11:68.4Mb | 1676 | 11 | 68000950 | 68887344 | 1792 | 192 | AD | eGFR | 0.452765 | 0.05825 | 1 | 0.204996 | 0.00554 | 1 | 0.0328876 | 81 | 0.317021869047619 |
| Chr11:86.2Mb | 1692 | 11 | 85869060 | 86629333 | 2111 | 201 | AD | eGFR | -0.631517 | -1 | -0.0608 | 0.398814 | 0.00898 | 1 | 0.033401 | 82 | 0.317021869047619 |
| Chr2:5.9Mb | 209 | 2 | 5568654 | 6306606 | 2126 | 248 | AD | eGFR | 0.577553 | 0.05956 | 1 | 0.333568 | 0.00822 | 1 | 0.0339912 | 83 | 0.317021869047619 |
| Chr12:127.2Mb | 1854 | 12 | 126871453 | 127545377 | 1947 | 222 | AD | eGFR | -0.678466 | -1 | -0.08432 | 0.460316 | 0.01455 | 1 | 0.0340535 | 84 | 0.317021869047619 |
| Chr10:89.2Mb | 1568 | 10 | 88443653 | 89971628 | 2672 | 262 | AD | eGFR | -0.3834 | -0.83307 | -0.03375 | 0.146996 | 0.00284 | 0.694 | 0.0347111 | 85 | 0.31934212 |
| Chr2:179.8Mb | 363 | 2 | 179224871 | 180336045 | 2260 | 231 | AD | eGFR | -0.42624 | -1 | -0.02825 | 0.181681 | 0.00314 | 1 | 0.035569 | 86 | 0.322169618390805 |
| Chr20:2.2Mb | 2371 | 20 | 1771286 | 2668108 | 2551 | 310 | AD | eGFR | -0.514436 | -1 | -0.04814 | 0.264645 | 0.00567 | 1 | 0.0358424 | 87 | 0.322169618390805 |
| Chr19:53.7Mb | 2361 | 19 | 53431218 | 54042240 | 2392 | 298 | AD | eGFR | -0.423849 | -1 | -0.04118 | 0.179648 | 0.00369 | 1 | 0.0375823 | 88 | 0.325789519148936 |
| Chr15:50.9Mb | 2057 | 15 | 50195877 | 51605477 | 3282 | 202 | AD | eGFR | 0.543015 | 0.04254 | 1 | 0.294865 | 0.00531 | 1 | 0.0377649 | 89 | 0.325789519148936 |
| Chr2:65.3Mb | 271 | 2 | 64696090 | 65938002 | 2705 | 309 | AD | eGFR | 0.436424 | 0.02937 | 1 | 0.190466 | 0.00369 | 1 | 0.0379262 | 90 | 0.32578951914893 |

| locus_label | locus | chr | start | stop | n.snps | n.pcs | phen1 | phen2 | rho | rho.lower | rho.upper | r2 | r2.lower | r2.upper | p | rank | FDR |
|---|---|---|---|---|---|---|---|---|---|---|---|---|---|---|---|---|---|
| | | | | | | | | | | | | | | | | | 6 |
| Chr9:111Mb | 1454 | 9 | 110630151 | 111368107 | 1903 | 209 | AD | eGFR | -0.608411 | -1 | -0.03722 | 0.370164 | 0.00835 | 1 | 0.0385503 | 91 | 0.325789519148936 |
| Chr21:16.4Mb | 2432 | 21 | 15840034 | 16873443 | 2338 | 311 | AD | eGFR | -0.231736 | -0.47675 | -0.0152 | 0.0537017 | 0.00106 | 0.22729 | 0.0387913 | 92 | 0.325789519148936 |
| Chr4:6.6Mb | 603 | 4 | 5972710 | 7135819 | 3332 | 379 | AD | eGFR | -0.571749 | -1 | -0.03295 | 0.326897 | 0.00632 | 1 | 0.0391522 | 93 | 0.325789519148936 |
| Chr7:32.1Mb | 1129 | 7 | 31730555 | 32460228 | 2234 | 224 | AD | eGFR | -0.566158 | -1 | -0.03728 | 0.320535 | 0.00624 | 1 | 0.0391614 | 94 | 0.325789519148936 |
| Chr3:76.8Mb | 488 | 3 | 76157471 | 77507832 | 3268 | 267 | AD | eGFR | 0.622994 | 0.02524 | 1 | 0.388121 | 0.00811 | 1 | 0.0404084 | 95 | 0.331712995833333 |
| Chr13:110.4Mb | 1948 | 13 | 109813577 | 110995432 | 3620 | 348 | AD | eGFR | -0.409544 | -1 | -0.01143 | 0.167726 | 0.00267 | 1 | 0.0407218 | 96 | 0.331712995833333 |
| Chr4:39.4Mb | 639 | 4 | 38984839 | 39820061 | 1908 | 188 | AD | eGFR | -0.771398 | -1 | -0.04508 | 0.595055 | 0.01162 | 1 | 0.0418713 | 97 | 0.336645414285714 |
| Chr15:25Mb | 2037 | 15 | 24531188 | 25384327 | 3062 | 205 | AD | eGFR | 0.330687 | 0.01877 | 0.67459 | 0.109354 | 0.00178 | 0.45508 | 0.0421883 | 98 | 0.336645414285714 |
| Chr2:200.5Mb | 379 | 2 | 199654970 | 201324132 | 2267 | 220 | AD | eGFR | -0.532417 | -1 | -0.03649 | 0.283468 | 0.00496 | 1 | 0.0435588 | 99 | 0.343052452 |
| Chr7:138.2Mb | 1214 | 7 | 137657460 | 138755107 | 2740 | 331 | AD | eGFR | -0.288599 | -0.59565 | -0.01042 | 0.0832892 | 0.00122 | 0.3548 | 0.0438686 | 100 | 0.343052452 |
| Chr3:169.6Mb | 570 | 3 | 168972663 | 170174578 | 2412 | 253 | AD | eGFR | -0.35027 | -0.8462 | -0.00419 | 0.122689 | 0.00169 | 0.71606 | 0.0456126 | 101 | 0.349654217475728 |
| Chr3:171.6Mb | 572 | 3 | 171134850 | 172004760 | 1998 | 247 | AD | eGFR | -0.366659 | -0.82127 | -0.02485 | 0.134439 | 0.00268 | 0.67448 | 0.0458437 | 102 | 0.349654217475728 |
| Chr19:52.3Mb | 2359 | 19 | 51903805 | 52678748 | 2523 | 270 | AD | eGFR | 0.559618 | 0.01007 | 1 | 0.313172 | 0.00544 | 1 | 0.0460542 | 103 | 0.349654217475728 |
| Chr2:12.4Mb | 217 | 2 | 11995766 | 12732506 | 1999 | 261 | AD | eGFR | -0.453562 | -1 | -0.01488 | 0.205719 | 0.00303 | 1 | 0.0466638 | 104 | 0.350875880769231 |
| Chr17:5Mb | 2178 | 17 | 4463972 | 5573784 | 2804 | 264 | AD | eGFR | 0.477665 | -0.00972 | 1 | 0.228164 | 0 | 1 | 0.0498265 | 105 | 0.369053062385321 |
| Chr10:63.3Mb | 1544 | 10 | 62626466 | 64069687 | 3053 | 268 | AD | eGFR | -0.474869 | -1 | 0.00238 | 0.2255 | 0 | 1 | 0.0501973 | 106 | 0.369053062385321 |
| Chr16:87.2Mb | 2169 | 16 | 86748868 | 87669168 | 2777 | 337 | AD | eGFR | -0.411128 | -1 | -0.00444 | 0.169026 | 0.0024 | 1 | 0.0512105 | 107 | 0.369053062385321 |
| Chr4:5.5Mb | 602 | 4 | 5051835 | 5972709 | 3101 | 320 | AD | eGFR | 0.420142 | 0.00413 | 0.99616 | 0.176519 | 0.00249 | 0.99234 | 0.0513704 | 108 | 0.369053062385321 |
| Chr16:11.5Mb | 2115 | 16 | 11022543 | 11916631 | 2513 | 280 | AD | eGFR | 0.479414 | 0.0017 | 1 | 0.229838 | 0.00353 | 1 | 0.0514409 | 109 | 0.369053062385321 |
| Chr19:42.5Mb | 2348 | 19 | 41733464 | 43317282 | 1794 | 229 | AD | eGFR | -0.396478 | -0.95876 | 0.00122 | 0.157195 | 0 | 0.91921 | 0.0527081 | 110 | 0.371906500854701 |
| Chr6:46.9Mb | 981 | 6 | 46430702 | 47313443 | 1891 | 227 | AD | eGFR | 0.538809 | -0.02116 | 1 | 0.290315 | 0 | 1 | 0.0537479 | 111 | 0.371906500854701 |
| Chr16:81Mb | 2161 | 16 | 80559686 | 81437861 | 2720 | 297 | AD | eGFR | 0.393173 | -0.01134 | 0.93064 | 0.154585 | 0 | 0.86609 | 0.0540395 | 112 | 0.371906500854701 |
| Chr2:235.3Mb | 411 | 2 | 234945578 | 235625789 | 2141 | 272 | AD | eGFR | -0.533746 | -1 | 0.00056 | 0.284885 | 0 | 1 | 0.0540634 | 113 | 0.371906500854701 |

| locus_label | locus | chr | start | stop | n.snps | n.pcs | phen1 | phen2 | rho | rho.lower | rho.upper | r2 | r2.lower | r2.upper | p | rank | FDR |
|---|---|---|---|---|---|---|---|---|---|---|---|---|---|---|---|---|---|
| Chr15:86.8Mb | 2086 | 15 | 86341599 | 87158587 | 2561 | 236 | AD | eGFR | -0.536873 | -1 | 0.00116 | 0.288233 | 0 | 1 | 0.0543343 | 114 | 0.371906500854701 |
| Chr4:107.3Mb | 695 | 4 | 106479156 | 108131206 | 2627 | 246 | AD | eGFR | 0.558389 | -0.00748 | 1 | 0.311798 | 0 | 1 | 0.055422 | 115 | 0.371906500854701 |
| Chr4:72.4Mb | 667 | 4 | 71468301 | 73258294 | 3113 | 228 | AD | eGFR | -0.480635 | -1 | 0.00194 | 0.23101 | 0 | 1 | 0.0554222 | 116 | 0.371906500854701 |
| Chr6:23.6Mb | 947 | 6 | 23192338 | 23939306 | 2253 | 252 | AD | eGFR | 0.407216 | 0.00013 | 0.93343 | 0.165825 | 0.00199 | 0.87128 | 0.0556433 | 117 | 0.371906500854701 |
| Chr1:99.8Mb | 86 | 1 | 99425001 | 100269207 | 2028 | 209 | AD | eGFR | 0.452316 | -0.02317 | 1 | 0.20459 | 0 | 1 | 0.0579972 | 118 | 0.373666427737226 |
| Chr13:30.8Mb | 1874 | 13 | 30068526 | 31433218 | 3649 | 326 | AD | eGFR | -0.564711 | -1 | 0.03094 | 0.318899 | 0 | 1 | 0.0583849 | 119 | 0.373666427737226 |
| Chr12:46.8Mb | 1785 | 12 | 46037745 | 47499603 | 2500 | 228 | AD | eGFR | 0.576565 | -0.01533 | 1 | 0.332428 | 0 | 1 | 0.0584771 | 120 | 0.373666427737226 |
| Chr3:141.5Mb | 544 | 3 | 141034638 | 141898978 | 1984 | 227 | AD | eGFR | 0.249598 | -0.01282 | 0.5653 | 0.062299 | 0 | 0.31956 | 0.0587327 | 121 | 0.373666427737226 |
| Chr16:81.9Mb | 2162 | 16 | 81437862 | 82310204 | 2968 | 337 | AD | eGFR | -0.321253 | -0.71484 | 0.01589 | 0.103203 | 0 | 0.51099 | 0.06051 | 122 | 0.373666427737226 |
| Chr9:3.6Mb | 1375 | 9 | 3079046 | 4025355 | 2186 | 311 | AD | eGFR | 0.353073 | -0.01771 | 0.80833 | 0.12466 | 0 | 0.65339 | 0.0605823 | 123 | 0.373666427737226 |
| Chr1:243.9Mb | 195 | 1 | 243048004 | 244723196 | 2670 | 313 | AD | eGFR | 0.223073 | -0.00699 | 0.47765 | 0.0497616 | 0 | 0.22815 | 0.0607146 | 124 | 0.373666427737226 |
| Chr14:100.1Mb | 2026 | 14 | 99474534 | 100786189 | 2910 | 338 | AD | eGFR | 0.356145 | -0.01298 | 0.97416 | 0.126839 | 0 | 0.95216 | 0.0607749 | 125 | 0.373666427737226 |
| Chr5:79.7Mb | 841 | 5 | 79005159 | 80448225 | 3599 | 308 | AD | eGFR | 0.538029 | -0.01832 | 1 | 0.289475 | 0 | 1 | 0.0611603 | 126 | 0.373666427737226 |
| Chr7:149.2Mb | 1222 | 7 | 148622914 | 149842999 | 2426 | 261 | AD | eGFR | -0.428971 | -1 | 0.01727 | 0.184016 | 0 | 1 | 0.0615515 | 127 | 0.373666427737226 |
| Chr5:152.4Mb | 901 | 5 | 151892618 | 152878627 | 2102 | 165 | AD | eGFR | -0.719243 | -1 | 0.01817 | 0.517311 | 0 | 1 | 0.0616264 | 128 | 0.373666427737226 |
| Chr19:43.6Mb | 2349 | 19 | 43317283 | 43960958 | 773 | 138 | AD | eGFR | 0.383123 | -0.01074 | 0.92319 | 0.146783 | 0 | 0.85227 | 0.0619525 | 129 | 0.373666427737226 |
| Chr2:102Mb | 298 | 2 | 101556800 | 102494487 | 1804 | 214 | AD | eGFR | -0.346948 | -0.8158 | 0.01993 | 0.120373 | 0 | 0.66553 | 0.0628329 | 130 | 0.373666427737226 |
| Chr17:66.2Mb | 2223 | 17 | 65582328 | 66757556 | 2552 | 271 | AD | eGFR | 0.290223 | -0.01308 | 0.6512 | 0.0842295 | 0 | 0.42425 | 0.0634205 | 131 | 0.373666427737226 |
| Chr9:114.9Mb | 1459 | 9 | 114443476 | 115390112 | 2340 | 194 | AD | eGFR | 0.578103 | -0.03597 | 1 | 0.334203 | 0 | 1 | 0.064168 | 132 | 0.373666427737226 |
| Chr8:10.2Mb | 1249 | 8 | 9835864 | 10478851 | 2118 | 256 | AD | eGFR | 0.40127 | -0.02124 | 0.99621 | 0.161017 | 0 | 0.99243 | 0.0647704 | 133 | 0.373666427737226 |
| Chr4:145.3Mb | 730 | 4 | 144401043 | 146174871 | 2999 | 220 | AD | eGFR | 0.278596 | -0.01933 | 0.6214 | 0.0776156 | 0 | 0.38613 | 0.0649355 | 134 | 0.373666427737226 |
| Chr18:13.2Mb | 2251 | 18 | 12735559 | 13641479 | 2068 | 270 | AD | eGFR | -0.34472 | -0.78422 | 0.02339 | 0.118832 | 0 | 0.615 | 0.0652083 | 135 | 0.373666427737226 |
| Chr9:37.2Mb | 1413 | 9 | 36749274 | 37687112 | 2018 | 253 | AD | eGFR | 0.550974 | -0.02916 | 1 | 0.303572 | 0 | 1 | 0.0654386 | 136 | 0.373666427737226 |
| Chr9:118.5M | 146 | 9 | 11798901 | 11900122 | 2176 | 206 | AD | eGFR | -0.417143 | -1 | 0.02422 | 0.174008 | 0 | 1 | 0.0654633 | 137 | 0.37366642773722 |

| locus_label | locus | chr | start | stop | n.snps | n.pcs | phen1 | phen2 | rho | rho.lower | rho.upper | r2 | r2.lower | r2.upper | p | rank | FDR |
|---|---|---|---|---|---|---|---|---|---|---|---|---|---|---|---|---|---|
| b | 3 | | 8 | 7 | | | | | | | | | | | | | 6 |
| Chr19:2.7Mb | 2309 | 19 | 2253175 | 3085446 | 2639 | 330 | AD | eGFR | 0.34714 | -0.01223 | 1 | 0.120506 | 0 | 1 | 0.0663603 | 138 | 0.3760417 |
| Chr6:126.5Mb | 1054 | 6 | 125365055 | 127545459 | 3050 | 286 | AD | eGFR | -0.430523 | -1 | 0.02942 | 0.18535 | 0 | 1 | 0.0674967 | 139 | 0.379729635971223 |
| Chr13:92.5Mb | 1932 | 13 | 91764205 | 93285142 | 3436 | 284 | AD | eGFR | -0.424997 | -1 | 0.0397 | 0.180622 | 0 | 1 | 0.0693574 | 140 | 0.382310289189189 |
| Chr1:186.6Mb | 141 | 1 | 186179448 | 187072605 | 1915 | 158 | AD | eGFR | -0.318701 | -0.74084 | 0.02273 | 0.10157 | 0 | 0.55039 | 0.0700675 | 141 | 0.382310289189189 |
| Chr19:50.9Mb | 2357 | 19 | 50451926 | 51259178 | 1808 | 284 | AD | eGFR | -0.281415 | -0.61143 | 0.02893 | 0.0791944 | 0 | 0.37385 | 0.0704277 | 142 | 0.382310289189189 |
| Chr18:11.3Mb | 2249 | 18 | 10675549 | 11872226 | 3011 | 316 | AD | eGFR | -0.342608 | -0.78607 | 0.02485 | 0.11738 | 0 | 0.6179 | 0.0710488 | 143 | 0.382310289189189 |
| Chr9:119.4Mb | 1464 | 9 | 119001228 | 119890464 | 2021 | 245 | AD | eGFR | 0.308424 | -0.03585 | 0.76954 | 0.0951256 | 0 | 0.59555 | 0.0710986 | 144 | 0.382310289189189 |
| Chr20:53.5Mb | 2417 | 20 | 53120335 | 53879803 | 1825 | 190 | AD | eGFR | -0.48256 | -1 | 0.04108 | 0.232864 | 0 | 1 | 0.0714052 | 145 | 0.382310289189189 |
| Chr1:55.2Mb | 47 | 1 | 54771294 | 55727134 | 2661 | 293 | AD | eGFR | -0.346014 | -0.9859 | 0.03459 | 0.119726 | 0 | 0.972 | 0.0715028 | 146 | 0.382310289189189 |
| Chr17:76.1Mb | 2232 | 17 | 75619257 | 76596287 | 2982 | 357 | AD | eGFR | 0.312935 | -0.02551 | 0.7181 | 0.0979282 | 0 | 0.51567 | 0.0720498 | 147 | 0.382310289189189 |
| Chr2:24.1Mb | 227 | 2 | 23538528 | 24669224 | 2693 | 225 | AD | eGFR | 0.755585 | -0.0637 | 1 | 0.570909 | 0 | 1 | 0.0723554 | 148 | 0.382310289189189 |
| Chr10:29.7Mb | 1518 | 10 | 29393750 | 30000223 | 1974 | 228 | AD | eGFR | 0.367393 | -0.0409 | 1 | 0.134977 | 0 | 1 | 0.0763321 | 149 | 0.3910107525 |
| Chr18:61.4Mb | 2289 | 18 | 60780195 | 62074993 | 3254 | 276 | AD | eGFR | -0.295503 | -0.69666 | 0.02276 | 0.0873218 | 0 | 0.48602 | 0.0763615 | 150 | 0.3910107525 |
| Chr11:15.8Mb | 1627 | 11 | 15126768 | 16383386 | 2271 | 243 | AD | eGFR | -0.324968 | -0.76265 | 0.02811 | 0.105604 | 0 | 0.58263 | 0.0769806 | 151 | 0.3910107525 |
| Chr19:15.3Mb | 2322 | 19 | 14891101 | 15650185 | 1916 | 246 | AD | eGFR | 0.368792 | -0.03689 | 0.88138 | 0.136008 | 0 | 0.77724 | 0.0770677 | 152 | 0.3910107525 |
| Chr8:31.8Mb | 1276 | 8 | 31134787 | 32454962 | 3481 | 234 | AD | eGFR | -0.386045 | -1 | 0.04587 | 0.149031 | 0 | 1 | 0.0777804 | 153 | 0.3910107525 |
| Chr6:20.9Mb | 944 | 6 | 20418021 | 21295864 | 2004 | 212 | AD | eGFR | -0.424228 | -1 | 0.03732 | 0.17997 | 0 | 1 | 0.0781644 | 154 | 0.3910107525 |
| Chr1:242.1Mb | 193 | 1 | 241813535 | 242417923 | 1897 | 240 | AD | eGFR | 0.309906 | -0.03569 | 0.68896 | 0.0960415 | 0 | 0.47466 | 0.0788154 | 155 | 0.3910107525 |
| Chr1:157.4Mb | 114 | 1 | 156813846 | 158081627 | 2881 | 290 | AD | eGFR | -0.492403 | -1 | 0.05395 | 0.242461 | 0 | 1 | 0.0788848 | 156 | 0.3910107525 |
| Chr21:45.4Mb | 2459 | 21 | 44815009 | 46045433 | 2948 | 310 | AD | eGFR | 0.484974 | -0.06699 | 1 | 0.2352 | 0 | 1 | 0.0789921 | 157 | 0.3910107525 |
| Chr1:191.9Mb | 146 | 1 | 191419276 | 192468373 | 1776 | 173 | AD | eGFR | 0.53084 | -0.06207 | 1 | 0.281791 | 0 | 1 | 0.079447 | 158 | 0.3910107525 |
| Chr4:190.5Mb | 774 | 4 | 189899574 | 191043880 | 2303 | 242 | AD | eGFR | 0.426234 | -0.04293 | 1 | 0.181675 | 0 | 1 | 0.0798632 | 159 | 0.3910107525 |
| Chr10:121.2Mb | 1594 | 10 | 120592840 | 121874445 | 3143 | 277 | AD | eGFR | -0.55676 | -1 | 0.04946 | 0.309981 | 0 | 1 | 0.0800022 | 160 | 0.3910107525 |
| Chr17:77.9M | 223 | 17 | 77412787 | 78438332 | 2437 | 328 | AD | eGFR | -0.363839 | -0.94707 | 0.05001 | 0.132379 | 0 | 0.89762 | 0.0812712 | 161 | 0.39317222222222 |

| locus_label | locus | chr | start | stop | n.snps | n.pcs | phen1 | phen2 | rho | rho.lower | rho.upper | r2 | r2.lower | r2.upper | p | rank | FDR |
|---|---|---|---|---|---|---|---|---|---|---|---|---|---|---|---|---|---|
| b | 4 | | | | | | | | | | | | | | | | 2 |
| Chr20:44.9Mb | 2408 | 20 | 44072211 | 45673603 | 3288 | 327 | AD | eGFR | -0.424058 | -1 | 0.06716 | 0.179826 | 0 | 1 | 0.08145 | 162 | 0.393172222222222 |
| Chr13:21Mb | 1865 | 13 | 20491182 | 21540733 | 2593 | 277 | AD | eGFR | 0.332373 | -0.04825 | 0.83813 | 0.110472 | 0 | 0.70336 | 0.0820012 | 163 | 0.39340453006135 |
| Chr1:248.8Mb | 200 | 1 | 248341595 | 249240538 | 1179 | 77 | AD | eGFR | 0.497832 | -0.05105 | 1 | 0.247836 | 0 | 1 | 0.0830048 | 164 | 0.395791180487805 |
| Chr22:25.7Mb | 2470 | 22 | 25282437 | 26166934 | 1840 | 261 | AD | eGFR | 0.244792 | -0.02891 | 0.53953 | 0.059923 | 0 | 0.29109 | 0.0845068 | 165 | 0.400511015757576 |
| Chr22:18.1Mb | 2463 | 22 | 17587475 | 18535225 | 2784 | 333 | AD | eGFR | 0.347948 | -0.04351 | 0.86284 | 0.121068 | 0 | 0.74449 | 0.0869423 | 166 | 0.409571557831325 |
| Chr10:31.7Mb | 1520 | 10 | 30919470 | 32457293 | 3050 | 245 | AD | eGFR | 0.383807 | -0.0699 | 1 | 0.147307 | 0 | 1 | 0.090292 | 167 | 0.42280445508982 |
| Chr1:68.2Mb | 58 | 1 | 67761891 | 68633860 | 1911 | 241 | AD | eGFR | -0.268246 | -0.62233 | 0.04814 | 0.0719561 | 0 | 0.38729 | 0.0908904 | 168 | 0.423073171428571 |
| Chr1:234Mb | 183 | 1 | 233639474 | 234365796 | 2152 | 245 | AD | eGFR | 0.341581 | -0.06127 | 0.82335 | 0.116678 | 0 | 0.67791 | 0.0915773 | 169 | 0.423748216568047 |
| Chr2:241.5Mb | 419 | 2 | 241113280 | 241910366 | 2644 | 272 | AD | eGFR | 0.618932 | -0.13113 | 1 | 0.383077 | 0 | 1 | 0.0929774 | 170 | 0.42769604 |
| Chr3:148Mb | 550 | 3 | 147358244 | 148638308 | 3039 | 284 | AD | eGFR | -0.459563 | -1 | 0.08022 | 0.211198 | 0 | 1 | 0.0944436 | 171 | 0.431899971929825 |
| Chr5:15.8Mb | 790 | 5 | 14910296 | 16592319 | 3042 | 290 | AD | eGFR | -0.540818 | -1 | 0.10493 | 0.292484 | 0 | 1 | 0.0965649 | 172 | 0.439033440697674 |
| Chr1:18.8Mb | 20 | 1 | 18427821 | 19238924 | 2365 | 325 | AD | eGFR | -0.24286 | -0.55545 | 0.04083 | 0.0589811 | 0 | 0.30853 | 0.0974167 | 173 | 0.440346008092486 |
| Chr10:91.5Mb | 1570 | 10 | 91021322 | 91960435 | 2489 | 209 | AD | eGFR | -0.3729 | -1 | 0.07515 | 0.139054 | 0 | 1 | 0.0992296 | 174 | 0.445962914942529 |
| Chr10:80.6Mb | 1559 | 10 | 79952997 | 81190573 | 3097 | 355 | AD | eGFR | 0.309622 | -0.06135 | 0.74355 | 0.095866 | 0 | 0.55287 | 0.100885 | 175 | 0.450811828571429 |
| Chr20:17.7Mb | 2388 | 20 | 17359291 | 18058524 | 2005 | 254 | AD | eGFR | -0.305015 | -0.74504 | 0.06094 | 0.093034 | 0 | 0.55508 | 0.102447 | 176 | 0.455190647727273 |
| Chr2:232.6Mb | 408 | 2 | 231847387 | 233352722 | 3327 | 287 | AD | eGFR | 0.40699 | -0.09496 | 1 | 0.165641 | 0 | 1 | 0.104422 | 177 | 0.458549072625698 |
| Chr2:133.9Mb | 325 | 2 | 133352951 | 134359059 | 2021 | 265 | AD | eGFR | 0.757007 | -0.17863 | 1 | 0.57306 | 0 | 1 | 0.104742 | 178 | 0.458549072625698 |
| Chr9:90.5Mb | 1434 | 9 | 90135155 | 90890979 | 2020 | 219 | AD | eGFR | -0.370621 | -1 | 0.07006 | 0.13736 | 0 | 1 | 0.104962 | 179 | 0.458549072625698 |
| Chr2:198.9Mb | 378 | 2 | 198140351 | 199654969 | 1857 | 198 | AD | eGFR | 0.50849 | -0.10346 | 1 | 0.258562 | 0 | 1 | 0.107644 | 180 | 0.46495 |
| Chr16:86.4Mb | 2168 | 16 | 86058598 | 86748867 | 2776 | 358 | AD | eGFR | -0.548839 | -1 | 0.14051 | 0.301224 | 0 | 1 | 0.108658 | 181 | 0.46495 |
| Chr19:17.4Mb | 2325 | 19 | 17045964 | 17750518 | 2072 | 265 | AD | eGFR | -0.288241 | -0.73677 | 0.07718 | 0.0830827 | 0 | 0.54283 | 0.108838 | 182 | 0.46495 |
| Chr6:41.4Mb | 976 | 6 | 40778402 | 42103738 | 2973 | 330 | AD | eGFR | 0.20587 | -0.04849 | 0.47201 | 0.0423824 | 0 | 0.22279 | 0.108999 | 183 | 0.46495 |
| Chr1:108.6Mb | 95 | 1 | 107874654 | 109271449 | 2860 | 267 | AD | eGFR | 0.527201 | -0.13118 | 1 | 0.27794 | 0 | 1 | 0.1094 | 184 | 0.46495 |
| Chr9:138.6M | 148 | 9 | 13820211 | 13899579 | 2193 | 327 | AD | eGFR | -0.341294 | -0.96269 | 0.08584 | 0.116481 | 0 | 0.92937 | 0.111545 | 185 | 0.47115079569892 |

| locus_label | locus | chr | start | stop | n.snps | n.pcs | phen1 | phen2 | rho | rho.lower | rho.upper | r2 | r2.lower | r2.upper | p | rank | FDR |
|---|---|---|---|---|---|---|---|---|---|---|---|---|---|---|---|---|---|
| b | 2 | | 3 | 1 | | | | | | | | | | | | | 5 |
| Chr1:200.6Mb | 154 | 1 | 200134006 | 201067952 | 2115 | 212 | AD | eGFR | 0.266434 | -0.06124 | 0.6915 | 0.0709872 | 0 | 0.47838 | 0.112064 | 186 | 0.471150795698925 |
| Chr11:78.3Mb | 1684 | 11 | 77719871 | 78912252 | 2310 | 280 | AD | eGFR | 0.363112 | -0.09376 | 1 | 0.131851 | 0 | 1 | 0.11441 | 187 | 0.478351063829787 |
| Chr17:62.8Mb | 2220 | 17 | 62112374 | 63548724 | 2299 | 233 | AD | eGFR | -0.296946 | -0.79561 | 0.07769 | 0.0881767 | 0 | 0.63374 | 0.115 | 188 | 0.478351063829787 |
| Chr4:58.5Mb | 654 | 4 | 58011086 | 58940256 | 2406 | 231 | AD | eGFR | -0.589548 | -1 | 0.15959 | 0.347567 | 0 | 1 | 0.117044 | 189 | 0.483181336842105 |
| Chr1:91.4Mb | 79 | 1 | 90941550 | 91870059 | 1892 | 203 | AD | eGFR | -0.482311 | -1 | 0.12687 | 0.232624 | 0 | 1 | 0.117397 | 190 | 0.483181336842105 |
| Chr10:114.9Mb | 1589 | 10 | 114255955 | 115588903 | 2518 | 316 | AD | eGFR | 0.443321 | -0.12806 | 1 | 0.196534 | 0 | 1 | 0.118488 | 191 | 0.485118408376963 |
| Chr14:92.7Mb | 2019 | 14 | 92101229 | 93386328 | 3413 | 321 | AD | eGFR | 0.226047 | -0.0699 | 0.5295 | 0.0510971 | 0 | 0.28037 | 0.120202 | 192 | 0.489572729166667 |
| Chr2:74.3Mb | 279 | 2 | 73544291 | 75148103 | 2368 | 253 | AD | eGFR | -0.277768 | -0.98711 | 0.084 | 0.0771548 | 0 | 0.97739 | 0.121037 | 193 | 0.490419347150259 |
| Chr15:97.4Mb | 2096 | 15 | 96864279 | 98025684 | 2524 | 329 | AD | eGFR | -0.309115 | -0.78623 | 0.09082 | 0.0955522 | 0 | 0.61816 | 0.122314 | 194 | 0.493038907216495 |
| Chr5:65Mb | 831 | 5 | 64438519 | 65606870 | 2215 | 200 | AD | eGFR | 0.63468 | -0.18034 | 1 | 0.402819 | 0 | 1 | 0.123754 | 195 | 0.494859126903553 |
| Chr1:117.6Mb | 103 | 1 | 117046312 | 118111667 | 1971 | 255 | AD | eGFR | 0.341411 | -0.09866 | 1 | 0.116562 | 0 | 1 | 0.124634 | 196 | 0.494859126903553 |
| Chr3:175.2Mb | 575 | 3 | 174684708 | 175671424 | 2652 | 261 | AD | eGFR | 0.50445 | -0.16007 | 1 | 0.25447 | 0 | 1 | 0.124664 | 197 | 0.494859126903553 |
| Chr19:38.9Mb | 2344 | 19 | 38472871 | 39370419 | 2134 | 224 | AD | eGFR | -0.232554 | -0.5984 | 0.06963 | 0.0540814 | 0 | 0.35809 | 0.125822 | 198 | 0.496933353535354 |
| Chr1:19.9Mb | 21 | 1 | 19238925 | 20556202 | 3291 | 304 | AD | eGFR | 0.246656 | -0.07963 | 0.59233 | 0.0608392 | 0 | 0.35086 | 0.126986 | 199 | 0.499010311557789 |
| Chr10:50.9Mb | 1533 | 10 | 50145674 | 51724039 | 2285 | 209 | AD | eGFR | 0.333087 | -0.09518 | 1 | 0.110947 | 0 | 1 | 0.1294 | 200 | 0.504977472636816 |
| Chr9:19.1Mb | 1395 | 9 | 18754197 | 19463292 | 2036 | 234 | AD | eGFR | 0.573902 | -0.16944 | 1 | 0.329363 | 0 | 1 | 0.129796 | 201 | 0.504977472636816 |
| Chr7:6.9Mb | 1102 | 7 | 6716906 | 7167610 | 1494 | 167 | AD | eGFR | 0.280445 | -0.07999 | 0.76509 | 0.0786491 | 0 | 0.58536 | 0.132712 | 202 | 0.510448601941748 |
| Chr13:95.5Mb | 1934 | 13 | 94925137 | 96132647 | 3653 | 322 | AD | eGFR | -0.347402 | -1 | 0.13206 | 0.120688 | 0 | 1 | 0.132932 | 203 | 0.510448601941748 |
| Chr7:40.8Mb | 1137 | 7 | 40261241 | 41415925 | 2026 | 245 | AD | eGFR | -0.28126 | -0.77779 | 0.08435 | 0.0791073 | 0 | 0.60496 | 0.134076 | 204 | 0.510448601941748 |
| Chr2:166.7Mb | 350 | 2 | 165917789 | 167488576 | 3235 | 240 | AD | eGFR | -0.598787 | -1 | 0.22045 | 0.358546 | 0 | 1 | 0.134392 | 205 | 0.510448601941748 |
| Chr5:14.3Mb | 789 | 5 | 13608148 | 14910295 | 2888 | 296 | AD | eGFR | -0.307266 | -0.91782 | 0.10461 | 0.0944122 | 0 | 0.84353 | 0.134466 | 206 | 0.510448601941748 |
| Chr12:66.6Mb | 1801 | 12 | 66114644 | 67183620 | 2221 | 256 | AD | eGFR | -0.38145 | -1 | 0.1136 | 0.145504 | 0 | 1 | 0.135267 | 207 | 0.511008666666667 |
| Chr1:24.9Mb | 26 | 1 | 23907046 | 25885138 | 3459 | 339 | AD | eGFR | -0.357418 | -1 | 0.11715 | 0.127747 | 0 | 1 | 0.136911 | 208 | 0.514732701923077 |

| locus_label | locus | chr | start | stop | n.snps | n.pcs | phen1 | phen2 | rho | rho.lower | rho.upper | r2 | r2.lower | r2.upper | p | rank | FDR |
|---|---|---|---|---|---|---|---|---|---|---|---|---|---|---|---|---|---|
| Chr14:87.8Mb | 2015 | 14 | 87350276 | 88208595 | 1948 | 161 | AD | eGFR | 0.475636 | -0.17872 | 1 | 0.22623 | 0 | 1 | 0.140195 | 209 | 0.524557368421053 |
| Chr11:77.1Mb | 1683 | 11 | 76518907 | 77719870 | 2367 | 196 | AD | eGFR | -0.282175 | -0.74299 | 0.09262 | 0.0796226 | 0 | 0.55204 | 0.142313 | 210 | 0.527580726415094 |
| Chr19:54.9Mb | 2363 | 19 | 54602371 | 55182973 | 1895 | 295 | AD | eGFR | 0.360807 | -0.12055 | 1 | 0.130182 | 0 | 1 | 0.14252 | 211 | 0.527580726415094 |
| Chr9:129.2Mb | 1472 | 9 | 128785783 | 129617771 | 1808 | 281 | AD | eGFR | 0.326102 | -0.11788 | 0.87974 | 0.106343 | 0 | 0.77488 | 0.143027 | 212 | 0.527580726415094 |
| Chr2:43.7Mb | 248 | 2 | 43295777 | 44104110 | 2033 | 225 | AD | eGFR | -0.249442 | -0.70844 | 0.08787 | 0.0622216 | 0 | 0.50231 | 0.146047 | 213 | 0.534350831775701 |
| Chr17:71Mb | 2227 | 17 | 70495120 | 71466953 | 2608 | 352 | AD | eGFR | -0.411302 | -1 | 0.1479 | 0.16917 | 0 | 1 | 0.146229 | 214 | 0.534350831775701 |
| Chr2:53.4Mb | 260 | 2 | 53032977 | 53852120 | 2496 | 234 | AD | eGFR | -0.559421 | -1 | 0.21348 | 0.312952 | 0 | 1 | 0.149164 | 215 | 0.542540688372093 |
| Chr8:6.3Mb | 1244 | 8 | 6019164 | 6663427 | 3026 | 315 | AD | eGFR | -0.390561 | -1 | 0.14918 | 0.152538 | 0 | 1 | 0.149907 | 216 | 0.542718861111111 |
| Chr4:140.3Mb | 727 | 4 | 139553761 | 141087047 | 2932 | 334 | AD | eGFR | 0.367677 | -0.14511 | 1 | 0.135187 | 0 | 1 | 0.151735 | 217 | 0.546805391705069 |
| Chr17:1.1Mb | 2174 | 17 | 815654 | 1480056 | 2181 | 331 | AD | eGFR | -0.23285 | -0.58568 | 0.09096 | 0.0542191 | 0 | 0.34302 | 0.152596 | 218 | 0.547385651376147 |
| Chr2:159.6Mb | 345 | 2 | 159040980 | 160200379 | 3009 | 227 | AD | eGFR | -0.370574 | -1 | 0.15536 | 0.137325 | 0 | 1 | 0.157319 | 219 | 0.559901336363636 |
| Chr2:162.5Mb | 347 | 2 | 161432090 | 163607635 | 3552 | 251 | AD | eGFR | -0.349958 | -1 | 0.14694 | 0.122471 | 0 | 1 | 0.157517 | 220 | 0.559901336363636 |
| Chr17:6.8Mb | 2180 | 17 | 6305079 | 7264458 | 2340 | 316 | AD | eGFR | -0.388462 | -1 | 0.16737 | 0.150902 | 0 | 1 | 0.158236 | 221 | 0.559912 |
| Chr10:1.6Mb | 1486 | 10 | 1329233 | 1872960 | 1788 | 262 | AD | eGFR | 0.289486 | -0.11749 | 0.77199 | 0.0838023 | 0 | 0.59597 | 0.159304 | 222 | 0.561151927927928 |
| Chr1:10.1Mb | 12 | 1 | 9475967 | 10753427 | 2039 | 241 | AD | eGFR | 0.260536 | -0.11056 | 0.88362 | 0.0678788 | 0 | 0.78449 | 0.161584 | 223 | 0.566630887892377 |
| Chr17:57.4Mb | 2217 | 17 | 56245228 | 58652809 | 2637 | 211 | AD | eGFR | -0.27597 | -0.91244 | 0.13257 | 0.0761595 | 0 | 0.83587 | 0.164627 | 224 | 0.573387236051502 |
| Chr7:98.8Mb | 1185 | 7 | 98173565 | 99465540 | 2345 | 209 | AD | eGFR | 0.364283 | -0.15837 | 1 | 0.132702 | 0 | 1 | 0.165746 | 225 | 0.573387236051502 |
| Chr19:1.1Mb | 2307 | 19 | 762047 | 1515920 | 2092 | 335 | AD | eGFR | -0.129237 | -0.31543 | 0.05164 | 0.0167022 | 0 | 0.0995 | 0.167079 | 226 | 0.573387236051502 |
| Chr11:120.6Mb | 1725 | 11 | 120064529 | 121208186 | 2420 | 302 | AD | eGFR | 0.281466 | -0.14063 | 0.76736 | 0.0792234 | 0 | 0.58884 | 0.168031 | 227 | 0.573387236051502 |
| Chr4:38.4Mb | 638 | 4 | 37880861 | 38984838 | 3109 | 301 | AD | eGFR | -0.190676 | -0.47371 | 0.08313 | 0.0363574 | 0 | 0.2244 | 0.168488 | 228 | 0.573387236051502 |
| Chr8:104.1Mb | 1332 | 8 | 103716054 | 104467845 | 1599 | 215 | AD | eGFR | 0.341936 | -0.161 | 1 | 0.11692 | 0 | 1 | 0.168704 | 229 | 0.573387236051502 |
| Chr13:106Mb | 1943 | 13 | 105672501 | 106313847 | 1737 | 220 | AD | eGFR | 0.44998 | -0.22952 | 1 | 0.202482 | 0 | 1 | 0.169394 | 230 | 0.573387236051502 |
| Chr21:46.5Mb | 2460 | 21 | 46045434 | 47014880 | 3131 | 311 | AD | eGFR | 0.226328 | -0.09143 | 0.5984 | 0.0512242 | 0 | 0.35809 | 0.170518 | 231 | 0.573387236051502 |
| Chr7:34.7Mb | 113 | 7 | 34114533 | 35354135 | 2861 | 212 | AD | eGFR | -0.359352 | -1 | 0.16797 | 0.129134 | 0 | 1 | 0.17067 | 232 | 0.57338723605150 |

| locus_label | locus | chr | start | stop | n.snps | n.pcs | phen1 | phen2 | rho | rho.lower | rho.upper | r2 | r2.lower | r2.upper | p | rank | FDR |
|---|---|---|---|---|---|---|---|---|---|---|---|---|---|---|---|---|---|
| | 2 | | | | | | | | | | | | | | | | 2 |
| Chr9:22.4Mb | 1398 | 9 | 21674033 | 23091193 | 3194 | 298 | AD | eGFR | 0.463984 | -0.23009 | 1 | 0.215281 | 0 | 1 | 0.170843 | 233 | 0.573387236051502 |
| Chr11:57.1Mb | 1667 | 11 | 56623495 | 57675986 | 1861 | 187 | AD | eGFR | 0.307619 | -0.15057 | 0.95333 | 0.0946296 | 0 | 0.91291 | 0.173014 | 234 | 0.578192085470085 |
| Chr7:20.8Mb | 1119 | 7 | 20122180 | 21390644 | 3152 | 318 | AD | eGFR | 0.252144 | -0.10833 | 0.67531 | 0.0635765 | 0 | 0.45605 | 0.176388 | 235 | 0.581840314049587 |
| Chr7:148Mb | 1221 | 7 | 147352172 | 148622913 | 3594 | 319 | AD | eGFR | 0.367294 | -0.17303 | 1 | 0.134905 | 0 | 1 | 0.176795 | 236 | 0.581840314049587 |
| Chr2:107Mb | 302 | 2 | 106546525 | 107412295 | 2196 | 220 | AD | eGFR | 0.384416 | -0.19451 | 1 | 0.147776 | 0 | 1 | 0.176896 | 237 | 0.581840314049587 |
| Chr1:150.4Mb | 109 | 1 | 149716038 | 151146407 | 1929 | 159 | AD | eGFR | 0.191113 | -0.09012 | 0.51583 | 0.036524 | 0 | 0.26608 | 0.178794 | 238 | 0.581840314049587 |
| Chr7:105.9Mb | 1191 | 7 | 105312564 | 106399739 | 2695 | 315 | AD | eGFR | 0.273728 | -0.13963 | 0.79701 | 0.0749271 | 0 | 0.63523 | 0.178915 | 239 | 0.581840314049587 |
| Chr15:73.9Mb | 2075 | 15 | 73375719 | 74458113 | 1983 | 220 | AD | eGFR | 0.258503 | -0.12696 | 0.79073 | 0.0668238 | 0 | 0.62818 | 0.179571 | 240 | 0.581840314049587 |
| Chr2:114.6Mb | 307 | 2 | 113930669 | 115203835 | 2401 | 219 | AD | eGFR | -0.281692 | -0.94077 | 0.14815 | 0.0793505 | 0 | 0.88793 | 0.179879 | 241 | 0.581840314049587 |
| Chr5:88.8Mb | 849 | 5 | 87943483 | 89584466 | 2054 | 272 | AD | eGFR | 0.303824 | -0.16463 | 0.89795 | 0.0923091 | 0 | 0.8099 | 0.180058 | 242 | 0.581840314049587 |
| Chr12:57.9Mb | 1794 | 12 | 56987106 | 58748139 | 2712 | 235 | AD | eGFR | -0.19948 | -0.57549 | 0.09759 | 0.0397924 | 0 | 0.33368 | 0.182609 | 243 | 0.587655300411523 |
| Chr11:125.8Mb | 1730 | 11 | 125266685 | 126304240 | 2682 | 244 | AD | eGFR | -0.359361 | -1 | 0.20099 | 0.12914 | 0 | 1 | 0.185764 | 244 | 0.593611412244898 |
| Chr15:25.9Mb | 2038 | 15 | 25384328 | 26392947 | 2915 | 335 | AD | eGFR | 0.486233 | -0.2943 | 1 | 0.236423 | 0 | 1 | 0.185978 | 245 | 0.593611412244898 |
| Chr11:49.1Mb | 1660 | 11 | 48765367 | 49386071 | 1424 | 34 | AD | eGFR | 0.382686 | -0.21332 | 1 | 0.146448 | 0 | 1 | 0.187108 | 246 | 0.594433975708502 |
| Chr10:42.7Mb | 1527 | 10 | 42392743 | 42984427 | 559 | 37 | AD | eGFR | -0.514595 | -1 | 0.31402 | 0.264808 | 0 | 1 | 0.187756 | 247 | 0.594433975708502 |
| Chr4:22.1Mb | 620 | 4 | 21416381 | 22835185 | 3428 | 264 | AD | eGFR | -0.446814 | -1 | 0.24421 | 0.199643 | 0 | 1 | 0.188526 | 248 | 0.594465048387097 |
| Chr14:77.1Mb | 2006 | 14 | 76694202 | 77425453 | 1857 | 274 | AD | eGFR | -0.281079 | -0.80934 | 0.14165 | 0.0790056 | 0 | 0.65504 | 0.19078 | 249 | 0.599156465863454 |
| Chr11:34.3Mb | 1646 | 11 | 33905328 | 34672697 | 1933 | 261 | AD | eGFR | -0.325095 | -1 | 0.1786 | 0.105686 | 0 | 1 | 0.191703 | 250 | 0.599646984 |
| Chr9:108.7Mb | 1452 | 9 | 108203096 | 109150717 | 1662 | 178 | AD | eGFR | -0.248192 | -0.66863 | 0.13152 | 0.0615995 | 0 | 0.44719 | 0.193713 | 251 | 0.603520183266932 |
| Chr8:122.1Mb | 1348 | 8 | 121554391 | 122622804 | 2596 | 253 | AD | eGFR | 0.340017 | -0.20749 | 1 | 0.115611 | 0 | 1 | 0.195216 | 252 | 0.605789333333333 |
| Chr1:173Mb | 130 | 1 | 172465154 | 173582511 | 1907 | 233 | AD | eGFR | 0.340658 | -0.20319 | 1 | 0.116048 | 0 | 1 | 0.197467 | 253 | 0.610215595330739 |
| Chr17:12.2Mb | 2185 | 17 | 11778165 | 12676353 | 1819 | 263 | AD | eGFR | -0.271732 | -0.86713 | 0.14637 | 0.0738383 | 0 | 0.76419 | 0.198447 | 254 | 0.610215595330739 |
| Chr5:82.2Mb | 843 | 5 | 81710195 | 82720248 | 1832 | 248 | AD | eGFR | 0.364662 | -0.2347 | 1 | 0.132978 | 0 | 1 | 0.199771 | 255 | 0.610215595330739 |

| locus_label | locus | chr | start | stop | n.snps | n.pcs | phen1 | phen2 | rho | rho.lower | rho.upper | r2 | r2.lower | r2.upper | p | rank | FDR |
|---|---|---|---|---|---|---|---|---|---|---|---|---|---|---|---|---|---|
| Chr12:20.5Mb | 1760 | 12 | 20074931 | 20866285 | 2034 | 257 | AD | eGFR | 0.263732 | -0.15612 | 0.74363 | 0.0695544 | 0 | 0.55448 | 0.200062 | 256 | 0.610215595330739 |
| Chr17:14.1Mb | 2187 | 17 | 13648447 | 14508610 | 2362 | 295 | AD | eGFR | -0.233422 | -0.64735 | 0.12863 | 0.0544856 | 0 | 0.41906 | 0.200544 | 257 | 0.610215595330739 |
| Chr8:95.6Mb | 1325 | 8 | 94999067 | 96175630 | 2987 | 226 | AD | eGFR | 0.281866 | -0.15228 | 0.81678 | 0.0794486 | 0 | 0.66714 | 0.205212 | 258 | 0.618628169230769 |
| Chr1:12.6Mb | 14 | 1 | 11709174 | 13481025 | 2446 | 298 | AD | eGFR | 0.189915 | -0.10567 | 0.50969 | 0.0360678 | 0 | 0.25982 | 0.205383 | 259 | 0.618628169230769 |
| Chr6:111.9Mb | 1041 | 6 | 111363444 | 112342873 | 1926 | 180 | AD | eGFR | -0.373589 | -1 | 0.23265 | 0.139568 | 0 | 1 | 0.205682 | 260 | 0.618628169230769 |
| Chr18:33.3Mb | 2264 | 18 | 32458908 | 34153297 | 3291 | 302 | AD | eGFR | -0.403755 | -1 | 0.26027 | 0.163018 | 0 | 1 | 0.208645 | 261 | 0.625135593869732 |
| Chr4:181.6Mb | 764 | 4 | 181122434 | 182065646 | 2910 | 311 | AD | eGFR | -0.357379 | -1 | 0.20669 | 0.12772 | 0 | 1 | 0.209491 | 262 | 0.625274664122138 |
| Chr2:37.2Mb | 241 | 2 | 36867390 | 37603360 | 1829 | 218 | AD | eGFR | 0.450651 | -0.32671 | 1 | 0.203086 | 0 | 1 | 0.210846 | 263 | 0.626926129277567 |
| Chr7:107Mb | 1192 | 7 | 106399740 | 107658603 | 2345 | 264 | AD | eGFR | -0.336971 | -1 | 0.21214 | 0.11355 | 0 | 1 | 0.212919 | 264 | 0.630691886363636 |
| Chr2:5.2Mb | 208 | 2 | 4766970 | 5568653 | 1774 | 215 | AD | eGFR | -0.340255 | -1 | 0.22665 | 0.115774 | 0 | 1 | 0.217524 | 265 | 0.641901011320755 |
| Chr10:76.4Mb | 1556 | 10 | 75694024 | 77144961 | 2067 | 186 | AD | eGFR | 0.308467 | -0.22333 | 1 | 0.0951521 | 0 | 1 | 0.220984 | 266 | 0.649659729323308 |
| Chr20:55.8Mb | 2420 | 20 | 55448076 | 56172843 | 2225 | 280 | AD | eGFR | 0.238649 | -0.16041 | 0.91116 | 0.0569533 | 0 | 0.85951 | 0.225641 | 267 | 0.657282416058394 |
| Chr1:82.8Mb | 71 | 1 | 82123369 | 83461215 | 3204 | 251 | AD | eGFR | -0.235779 | -0.73863 | 0.1554 | 0.0555916 | 0 | 0.55356 | 0.226254 | 268 | 0.657282416058394 |
| Chr18:9.3Mb | 2247 | 18 | 8788525 | 9816164 | 3153 | 303 | AD | eGFR | 0.369759 | -0.28705 | 1 | 0.136722 | 0 | 1 | 0.227966 | 269 | 0.657282416058394 |
| Chr5:129.7Mb | 886 | 5 | 128734906 | 130573817 | 3093 | 176 | AD | eGFR | 0.229104 | -0.17138 | 0.67488 | 0.0524885 | 0 | 0.45738 | 0.228273 | 270 | 0.657282416058394 |
| Chr16:2.7Mb | 2104 | 16 | 1996493 | 3379996 | 2326 | 334 | AD | eGFR | 0.151095 | -0.10533 | 0.43841 | 0.0228296 | 0 | 0.19274 | 0.229462 | 271 | 0.657282416058394 |
| Chr11:123Mb | 1727 | 11 | 122589225 | 123398882 | 2289 | 296 | AD | eGFR | -0.286939 | -0.91991 | 0.20372 | 0.0823337 | 0 | 0.8482 | 0.229909 | 272 | 0.657282416058394 |
| Chr21:35.1Mb | 2449 | 21 | 34383795 | 35729261 | 2767 | 311 | AD | eGFR | 0.212458 | -0.15056 | 0.66062 | 0.0451385 | 0 | 0.43694 | 0.23006 | 273 | 0.657282416058394 |
| Chr17:36.7Mb | 2202 | 17 | 36116885 | 37361178 | 2241 | 288 | AD | eGFR | 0.205641 | -0.1389 | 0.66091 | 0.0422884 | 0 | 0.44013 | 0.230301 | 274 | 0.657282416058394 |
| Chr6:151.1Mb | 1073 | 6 | 150635316 | 151629954 | 3214 | 316 | AD | eGFR | 0.263099 | -0.18012 | 0.91891 | 0.069221 | 0 | 0.85217 | 0.234129 | 275 | 0.662287446808511 |
| Chr22:23.5Mb | 2468 | 22 | 23143365 | 23950732 | 2033 | 243 | AD | eGFR | 0.220062 | -0.14958 | 0.63593 | 0.0484272 | 0 | 0.40587 | 0.234749 | 276 | 0.662287446808511 |
| Chr2:138.8Mb | 329 | 2 | 138073792 | 139505819 | 2979 | 220 | AD | eGFR | -0.279406 | -0.87461 | 0.20023 | 0.0780675 | 0 | 0.76662 | 0.235677 | 277 | 0.662287446808511 |
| Chr15:71.4Mb | 2073 | 15 | 70767984 | 72058129 | 2485 | 278 | AD | eGFR | 0.329743 | -0.24121 | 1 | 0.10873 | 0 | 1 | 0.235687 | 278 | 0.662287446808511 |
| Chr2:224.5M | 401 | 2 | 22412308 | 22493873 | 1864 | 226 | AD | eGFR | -0.281725 | -0.91902 | 0.21296 | 0.0793687 | 0 | 0.85326 | 0.236601 | 279 | 0.66228744680851 |

| locus_label | locus | chr | start | stop | n.snps | n.pcs | phen1 | phen2 | rho | rho.lower | rho.upper | r2 | r2.lower | r2.upper | p | rank | FDR |
|---|---|---|---|---|---|---|---|---|---|---|---|---|---|---|---|---|---|
| b | | | 0 | 1 | | | | | | | | | | | | | 1 |
| Chr4:183.3Mb | 766 | 4 | 182946259 | 183603892 | 1919 | 287 | AD | eGFR | 0.256192 | -0.19106 | 0.77966 | 0.0656345 | 0 | 0.60846 | 0.23814 | 280 | 0.662287446808511 |
| Chr16:14.9Mb | 2118 | 16 | 13893408 | 15921108 | 2560 | 291 | AD | eGFR | 0.294049 | -0.23443 | 0.97291 | 0.0864651 | 0 | 0.95286 | 0.238214 | 281 | 0.662287446808511 |
| Chr18:6.2Mb | 2244 | 18 | 5575813 | 6756496 | 2782 | 339 | AD | eGFR | -0.216477 | -0.65891 | 0.14932 | 0.0468622 | 0 | 0.4359 | 0.23883 | 282 | 0.662287446808511 |
| Chr2:128.7Mb | 320 | 2 | 127895645 | 129486897 | 3863 | 286 | AD | eGFR | -0.387026 | -1 | 0.31087 | 0.149789 | 0 | 1 | 0.240943 | 283 | 0.664533560810811 |
| Chr13:38.8Mb | 1881 | 13 | 38290690 | 39381743 | 2193 | 208 | AD | eGFR | 0.370412 | -0.28942 | 1 | 0.137205 | 0 | 1 | 0.24297 | 284 | 0.664533560810811 |
| Chr10:125.3Mb | 1598 | 10 | 124894143 | 125798319 | 2768 | 297 | AD | eGFR | 0.31422 | -0.25462 | 1 | 0.0987341 | 0 | 1 | 0.243637 | 285 | 0.664533560810811 |
| Chr11:12.9Mb | 1625 | 11 | 12470450 | 13364727 | 2247 | 255 | AD | eGFR | -0.38337 | -1 | 0.31684 | 0.146972 | 0 | 1 | 0.243741 | 286 | 0.664533560810811 |
| Chr8:13.2Mb | 1253 | 8 | 12947936 | 13395051 | 1738 | 169 | AD | eGFR | -0.364212 | -1 | 0.29643 | 0.132651 | 0 | 1 | 0.244932 | 287 | 0.664533560810811 |
| Chr18:23.4Mb | 2257 | 18 | 22746154 | 24088163 | 2519 | 283 | AD | eGFR | 0.292211 | -0.24585 | 0.98266 | 0.0853875 | 0 | 0.96651 | 0.245787 | 288 | 0.664533560810811 |
| Chr2:121.6Mb | 313 | 2 | 121073753 | 122026817 | 2262 | 310 | AD | eGFR | 0.136721 | -0.09508 | 0.3906 | 0.0186927 | 0 | 0.15257 | 0.246079 | 289 | 0.664533560810811 |
| Chr12:128.8Mb | 1856 | 12 | 128372589 | 129170296 | 2348 | 328 | AD | eGFR | -0.399658 | -1 | 0.32888 | 0.159727 | 0 | 1 | 0.247977 | 290 | 0.664533560810811 |
| Chr8:23.3Mb | 1268 | 8 | 22895019 | 23788494 | 2772 | 280 | AD | eGFR | -0.171367 | -0.56577 | 0.12699 | 0.0293666 | 0 | 0.32192 | 0.249398 | 291 | 0.664533560810811 |
| Chr20:5Mb | 2374 | 20 | 4591848 | 5379450 | 2348 | 277 | AD | eGFR | -0.314519 | -1 | 0.24813 | 0.0989224 | 0 | 1 | 0.249405 | 292 | 0.664533560810811 |
| Chr5:56.4Mb | 823 | 5 | 55968967 | 56896890 | 2062 | 226 | AD | eGFR | 0.357281 | -0.27861 | 1 | 0.12765 | 0 | 1 | 0.249924 | 293 | 0.664533560810811 |
| Chr6:3.4Mb | 928 | 6 | 2746532 | 3964072 | 3137 | 345 | AD | eGFR | 0.223473 | -0.16327 | 0.66265 | 0.0499404 | 0 | 0.43951 | 0.25052 | 294 | 0.664533560810811 |
| Chr8:61.8Mb | 1298 | 8 | 61324890 | 62330900 | 2009 | 229 | AD | eGFR | -0.349509 | -1 | 0.26357 | 0.122157 | 0 | 1 | 0.250955 | 295 | 0.664533560810811 |
| Chr5:34.6Mb | 806 | 5 | 33971057 | 35157146 | 3027 | 245 | AD | eGFR | -0.216726 | -0.75404 | 0.18325 | 0.0469702 | 0 | 0.58476 | 0.251537 | 296 | 0.664533560810811 |
| Chr11:11.4Mb | 1623 | 11 | 11081300 | 11632650 | 1924 | 287 | AD | eGFR | -0.412996 | -1 | 0.35444 | 0.170566 | 0 | 1 | 0.254695 | 297 | 0.670611077441078 |
| Chr14:88.8Mb | 2016 | 14 | 88208596 | 89482883 | 2950 | 226 | AD | eGFR | -0.18772 | -0.54161 | 0.1306 | 0.0352389 | 0 | 0.29334 | 0.256888 | 298 | 0.674115489932886 |
| Chr7:139.5Mb | 1215 | 7 | 138755108 | 140217630 | 3025 | 320 | AD | eGFR | 0.237684 | -0.19383 | 0.9059 | 0.0564936 | 0 | 0.82786 | 0.259798 | 299 | 0.678747980456026 |
| Chr19:41.4Mb | 2347 | 19 | 41004106 | 41733463 | 1723 | 233 | AD | eGFR | 0.166441 | -0.1346 | 0.51834 | 0.0277027 | 0 | 0.2691 | 0.264921 | 300 | 0.678747980456026 |
| Chr15:49.7Mb | 2056 | 15 | 49260007 | 50195876 | 1936 | 115 | AD | eGFR | -0.200356 | -0.58131 | 0.15449 | 0.0401426 | 0 | 0.33793 | 0.265575 | 301 | 0.678747980456026 |
| Chr10:98.7Mb | 1576 | 10 | 98005995 | 99367800 | 2730 | 290 | AD | eGFR | -0.280719 | -1 | 0.23435 | 0.0788032 | 0 | 1 | 0.265685 | 302 | 0.678747980456026 |

| locus_label | locus | chr | start | stop | n.snps | n.pcs | phen1 | phen2 | rho | rho.lower | rho.upper | r2 | r2.lower | r2.upper | p | rank | FDR |
|---|---|---|---|---|---|---|---|---|---|---|---|---|---|---|---|---|---|
| Chr17:64Mb | 2221 | 17 | 63548725 | 64438387 | 2089 | 190 | AD | eGFR | 0.316323 | -0.2712 | 1 | 0.10006 | 0 | 1 | 0.265906 | 303 | 0.678747980456026 |
| Chr8:27.9Mb | 1273 | 8 | 27406512 | 28344176 | 2371 | 258 | AD | eGFR | -0.188724 | -0.551 | 0.15364 | 0.0356168 | 0 | 0.30423 | 0.266194 | 304 | 0.678747980456026 |
| Chr12:25.5Mb | 1765 | 12 | 25058715 | 25990813 | 2524 | 254 | AD | eGFR | 0.296818 | -0.26553 | 1 | 0.0881006 | 0 | 1 | 0.26636 | 305 | 0.678747980456026 |
| Chr3:31.8Mb | 451 | 3 | 31269698 | 32378805 | 2568 | 309 | AD | eGFR | -0.166897 | -0.47938 | 0.13314 | 0.0278546 | 0 | 0.2298 | 0.266428 | 306 | 0.678747980456026 |
| Chr6:150Mb | 1072 | 6 | 149278799 | 150635315 | 3181 | 274 | AD | eGFR | -0.330854 | -1 | 0.3106 | 0.109464 | 0 | 1 | 0.266465 | 307 | 0.678747980456026 |
| Chr16:6.1Mb | 2107 | 16 | 5782969 | 6446081 | 2194 | 338 | AD | eGFR | -0.490436 | -1 | 0.48654 | 0.240527 | 0 | 1 | 0.26932 | 308 | 0.683792987012987 |
| Chr19:51.6Mb | 2358 | 19 | 51259179 | 51903804 | 1798 | 303 | AD | eGFR | -0.176003 | -0.53605 | 0.14987 | 0.0309771 | 0 | 0.28874 | 0.273854 | 309 | 0.691081949206349 |
| Chr3:154.2Mb | 555 | 3 | 153406719 | 155046423 | 3599 | 244 | AD | eGFR | -0.213887 | -0.6716 | 0.17889 | 0.0457476 | 0 | 0.45281 | 0.274682 | 310 | 0.691081949206349 |
| Chr1:211.7Mb | 162 | 1 | 211082893 | 212347582 | 3490 | 233 | AD | eGFR | -0.373247 | -1 | 0.32449 | 0.139313 | 0 | 1 | 0.27557 | 311 | 0.691081949206349 |
| Chr10:72.2Mb | 1553 | 10 | 71672998 | 72659223 | 2824 | 291 | AD | eGFR | 0.367908 | -0.36112 | 1 | 0.135356 | 0 | 1 | 0.276164 | 312 | 0.691081949206349 |
| Chr6:102.2Mb | 1032 | 6 | 101793543 | 102637706 | 1894 | 203 | AD | eGFR | 0.435648 | -0.41975 | 1 | 0.189789 | 0 | 1 | 0.276686 | 313 | 0.691081949206349 |
| Chr8:135.8Mb | 1360 | 8 | 135313674 | 136220342 | 2273 | 262 | AD | eGFR | 0.296627 | -0.27605 | 1 | 0.0879875 | 0 | 1 | 0.277598 | 314 | 0.691081949206349 |
| Chr1:116.6Mb | 102 | 1 | 116207298 | 117046311 | 1854 | 207 | AD | eGFR | 0.22789 | -0.20897 | 0.72895 | 0.0519341 | 0 | 0.53399 | 0.278377 | 315 | 0.691081949206349 |
| Chr5:103.3Mb | 861 | 5 | 102903986 | 103788460 | 1802 | 247 | AD | eGFR | -0.291198 | -1 | 0.29263 | 0.0847964 | 0 | 1 | 0.283794 | 316 | 0.698950616352201 |
| Chr4:78.5Mb | 671 | 4 | 78045638 | 78967715 | 2354 | 183 | AD | eGFR | -0.304592 | -1 | 0.29556 | 0.0927765 | 0 | 1 | 0.284088 | 317 | 0.698950616352201 |
| Chr1:88.2Mb | 76 | 1 | 87667573 | 88656689 | 1801 | 279 | AD | eGFR | 0.257095 | -0.24027 | 0.85096 | 0.0660978 | 0 | 0.7307 | 0.284228 | 318 | 0.698950616352201 |
| Chr13:46Mb | 1888 | 13 | 45476447 | 46493236 | 2052 | 233 | AD | eGFR | 0.290719 | -0.28729 | 1 | 0.0845173 | 0 | 1 | 0.286533 | 319 | 0.70241005015674 |
| Chr16:0.6Mb | 2102 | 16 | 60169 | 1049079 | 2709 | 307 | AD | eGFR | 0.220587 | -0.21561 | 0.86787 | 0.0486587 | 0 | 0.7712 | 0.289121 | 320 | 0.703374534954407 |
| Chr19:49.9Mb | 2356 | 19 | 49415190 | 50451925 | 2074 | 292 | AD | eGFR | 0.144976 | -0.12649 | 0.43771 | 0.0210179 | 0 | 0.1916 | 0.289373 | 321 | 0.703374534954407 |
| Chr13:71.5Mb | 1911 | 13 | 71010209 | 71976862 | 2643 | 262 | AD | eGFR | -0.27532 | -0.98804 | 0.28045 | 0.0758011 | 0 | 0.98546 | 0.290419 | 322 | 0.703374534954407 |
| Chr13:99Mb | 1937 | 13 | 98527336 | 99402294 | 2295 | 300 | AD | eGFR | -0.19945 | -0.63841 | 0.1762 | 0.0397802 | 0 | 0.40867 | 0.291507 | 323 | 0.703374534954407 |
| Chr8:145.6Mb | 1369 | 8 | 144934289 | 146303787 | 1587 | 247 | AD | eGFR | 0.159467 | -0.1419 | 0.51455 | 0.0254296 | 0 | 0.2657 | 0.292887 | 324 | 0.703374534954407 |
| Chr8:0.3Mb | 1232 | 8 | 11740 | 660414 | 1963 | 237 | AD | eGFR | -0.277512 | -1 | 0.27045 | 0.0770128 | 0 | 1 | 0.292897 | 325 | 0.703374534954407 |
| Chr11:17Mb | 162 | 11 | 16383387 | 17583948 | 2471 | 224 | AD | eGFR | 0.257392 | -0.25622 | 1 | 0.0662508 | 0 | 1 | 0.294621 | 326 | 0.70337453495440 |

| locus_label | locus | chr | start | stop | n.snps | n.pcs | phen1 | phen2 | rho | rho.lower | rho.upper | r2 | r2.lower | r2.upper | p | rank | FDR |
|---|---|---|---|---|---|---|---|---|---|---|---|---|---|---|---|---|---|
| | 8 | | | | | | | | | | | | | | | | 7 |
| Chr10:60.6Mb | 1542 | 10 | 59822947 | 61470383 | 3078 | 278 | AD | eGFR | 0.238932 | -0.23726 | 0.79306 | 0.0570885 | 0 | 0.63102 | 0.295585 | 327 | 0.703374534954407 |
| Chr6:167.3Mb | 1090 | 6 | 166853326 | 167714865 | 2589 | 271 | AD | eGFR | -0.405385 | -1 | 0.42958 | 0.164337 | 0 | 1 | 0.295817 | 328 | 0.703374534954407 |
| Chr10:23.9Mb | 1511 | 10 | 23365686 | 24489447 | 2251 | 211 | AD | eGFR | 0.244627 | -0.22861 | 0.83753 | 0.0598424 | 0 | 0.71608 | 0.295921 | 329 | 0.703374534954407 |
| Chr4:2Mb | 598 | 4 | 1529268 | 2468935 | 2090 | 250 | AD | eGFR | -0.177539 | -0.56704 | 0.16749 | 0.0315201 | 0 | 0.32403 | 0.299548 | 330 | 0.708919619335347 |
| Chr17:30.6Mb | 2197 | 17 | 29783142 | 31318753 | 2997 | 295 | AD | eGFR | 0.283776 | -0.30765 | 1 | 0.0805285 | 0 | 1 | 0.300067 | 331 | 0.708919619335347 |
| Chr21:27.1Mb | 2442 | 21 | 26467827 | 27774350 | 2926 | 256 | AD | eGFR | -0.336603 | -1 | 0.35099 | 0.113302 | 0 | 1 | 0.301528 | 332 | 0.710225590361446 |
| Chr1:31Mb | 30 | 1 | 30405183 | 31639217 | 3159 | 302 | AD | eGFR | -0.20035 | -0.64142 | 0.19092 | 0.04014 | 0 | 0.41172 | 0.303801 | 333 | 0.713430576576577 |
| Chr6:19.9Mb | 943 | 6 | 19449153 | 20418020 | 2417 | 300 | AD | eGFR | -0.152432 | -0.46608 | 0.14092 | 0.0232355 | 0 | 0.21766 | 0.307177 | 334 | 0.716246833333333 |
| Chr1:2.6Mb | 3 | 1 | 2215496 | 2983519 | 1724 | 262 | AD | eGFR | 0.166088 | -0.15902 | 0.52414 | 0.0275851 | 0 | 0.27479 | 0.307678 | 335 | 0.716246833333333 |
| Chr18:53.1Mb | 2281 | 18 | 52512524 | 53762996 | 2122 | 231 | AD | eGFR | -0.352602 | -1 | 0.37855 | 0.124328 | 0 | 1 | 0.307748 | 336 | 0.716246833333333 |
| Chr3:133.3Mb | 538 | 3 | 132925968 | 133718493 | 1931 | 238 | AD | eGFR | 0.315915 | -0.34242 | 1 | 0.0998025 | 0 | 1 | 0.309727 | 337 | 0.716853384615385 |
| Chr7:62.6Mb | 1156 | 7 | 61969718 | 63311297 | 1344 | 82 | AD | eGFR | -0.375422 | -1 | 0.44756 | 0.140941 | 0 | 1 | 0.309842 | 338 | 0.716853384615385 |
| Chr5:22.7Mb | 796 | 5 | 22149043 | 23246745 | 2067 | 198 | AD | eGFR | -0.317928 | -1 | 0.37948 | 0.101078 | 0 | 1 | 0.31177 | 339 | 0.71697704398827 |
| Chr6:124.6Mb | 1053 | 6 | 123854858 | 125365054 | 3336 | 335 | AD | eGFR | 0.416218 | -0.46768 | 1 | 0.173238 | 0 | 1 | 0.312113 | 340 | 0.71697704398827 |
| Chr2:172.9Mb | 357 | 2 | 172501316 | 173287857 | 1890 | 203 | AD | eGFR | -0.336387 | -1 | 0.40697 | 0.113156 | 0 | 1 | 0.312646 | 341 | 0.71697704398827 |
| Chr8:143.1Mb | 1367 | 8 | 142611890 | 143611461 | 2632 | 297 | AD | eGFR | -0.266331 | -1 | 0.29461 | 0.0709322 | 0 | 1 | 0.314604 | 342 | 0.719357684210526 |
| Chr7:131.1Mb | 1209 | 7 | 130418705 | 131856481 | 3031 | 348 | AD | eGFR | 0.294755 | -0.32969 | 1 | 0.0868806 | 0 | 1 | 0.315627 | 343 | 0.719592752186589 |
| Chr9:93.8Mb | 1438 | 9 | 93441051 | 94175374 | 1884 | 222 | AD | eGFR | 0.319612 | -0.36303 | 1 | 0.102152 | 0 | 1 | 0.318111 | 344 | 0.723147680232558 |
| Chr7:70.7Mb | 1164 | 7 | 70144615 | 71215741 | 3099 | 277 | AD | eGFR | -0.375608 | -1 | 0.4614 | 0.141082 | 0 | 1 | 0.322126 | 345 | 0.730152266666667 |
| Chr1:231.7Mb | 180 | 1 | 231287765 | 232177704 | 1854 | 235 | AD | eGFR | -0.261049 | -1 | 0.28402 | 0.0681468 | 0 | 1 | 0.324609 | 346 | 0.73181941954023 |
| Chr12:106.6Mb | 1836 | 12 | 106153788 | 106964977 | 1846 | 242 | AD | eGFR | -0.242079 | -0.95465 | 0.27692 | 0.0586024 | 0 | 0.92272 | 0.324832 | 347 | 0.73181941954023 |
| Chr3:24.9Mb | 445 | 3 | 24270190 | 25516193 | 3474 | 321 | AD | eGFR | 0.229622 | -0.25071 | 0.9098 | 0.0527264 | 0 | 0.83622 | 0.325669 | 348 | 0.73181941954023 |
| Chr15:37.3Mb | 2047 | 15 | 36583328 | 37962915 | 3210 | 326 | AD | eGFR | -0.183017 | -0.58922 | 0.19305 | 0.0334953 | 0 | 0.34762 | 0.328639 | 349 | 0.735274395480226 |
| Chr9:123.4Mb | 1467 | 9 | 122677720 | 124119883 | 1978 | 247 | AD | eGFR | -0.360917 | -1 | 0.47968 | 0.130261 | 0 | 1 | 0.329631 | 350 | 0.735274395480226 |

| locus_label | locus | chr | start | stop | n.snps | n.pcs | phen1 | phen2 | rho | rho.lower | rho.upper | r2 | r2.lower | r2.upper | p | rank | FDR |
|---|---|---|---|---|---|---|---|---|---|---|---|---|---|---|---|---|---|
| Chr5:75.6Mb | 838 | 5 | 75239303 | 75959515 | 1771 | 241 | AD | eGFR | 0.213938 | -0.23653 | 0.74011 | 0.0457696 | 0 | 0.54868 | 0.330357 | 351 | 0.735274395480226 |
| Chr12:12.2Mb | 1752 | 12 | 11742150 | 12721874 | 2332 | 307 | AD | eGFR | 0.17228 | -0.18985 | 0.62312 | 0.0296803 | 0 | 0.39096 | 0.332572 | 352 | 0.735274395480226 |
| Chr11:45.7Mb | 1657 | 11 | 45019560 | 46316005 | 2595 | 257 | AD | eGFR | -0.31981 | -1 | 0.40918 | 0.102278 | 0 | 1 | 0.33278 | 353 | 0.735274395480226 |
| Chr4:184Mb | 767 | 4 | 183603893 | 184338584 | 2057 | 271 | AD | eGFR | -0.269943 | -1 | 0.30778 | 0.0728693 | 0 | 1 | 0.332848 | 354 | 0.735274395480226 |
| Chr18:0.4Mb | 2238 | 18 | 10719 | 848350 | 2396 | 301 | AD | eGFR | 0.251488 | -0.30152 | 0.94193 | 0.0632463 | 0 | 0.91341 | 0.336678 | 355 | 0.740446365168539 |
| Chr7:150.4Mb | 1223 | 7 | 149843000 | 150897818 | 2666 | 244 | AD | eGFR | 0.185409 | -0.21883 | 0.6359 | 0.0343766 | 0 | 0.40683 | 0.337083 | 356 | 0.740446365168539 |
| Chr8:126.1Mb | 1351 | 8 | 125453323 | 126766827 | 2859 | 311 | AD | eGFR | 0.120442 | -0.13413 | 0.39179 | 0.0145063 | 0 | 0.15368 | 0.34048 | 357 | 0.745813333333333 |
| Chr4:0.4Mb | 596 | 4 | 10227 | 812415 | 1808 | 160 | AD | eGFR | -0.209894 | -0.83205 | 0.25451 | 0.0440557 | 0 | 0.72606 | 0.34261 | 358 | 0.748382737430168 |
| Chr12:54.9Mb | 1792 | 12 | 54371449 | 55416802 | 2238 | 255 | AD | eGFR | -0.322823 | -1 | 0.43018 | 0.104215 | 0 | 1 | 0.344376 | 359 | 0.750144935933148 |
| Chr6:15.8Mb | 938 | 6 | 15069422 | 16566882 | 3312 | 314 | AD | eGFR | -0.287327 | -1 | 0.36012 | 0.082557 | 0 | 1 | 0.346928 | 360 | 0.753604711111111 |
| Chr9:25.4Mb | 1401 | 9 | 24757730 | 25957606 | 3261 | 308 | AD | eGFR | -0.25305 | -1 | 0.30581 | 0.0640342 | 0 | 1 | 0.350894 | 361 | 0.757369136612022 |
| Chr7:101.4Mb | 1187 | 7 | 100849308 | 101950748 | 3028 | 261 | AD | eGFR | 0.182104 | -0.21953 | 0.70157 | 0.0331617 | 0 | 0.49681 | 0.352035 | 362 | 0.757369136612022 |
| Chr9:112.5Mb | 1456 | 9 | 112076382 | 112865520 | 2166 | 242 | AD | eGFR | 0.236732 | -0.30089 | 1 | 0.0560419 | 0 | 1 | 0.353119 | 363 | 0.757369136612022 |
| Chr5:160.1Mb | 909 | 5 | 159663734 | 160519049 | 2110 | 232 | AD | eGFR | -0.272149 | -1 | 0.36523 | 0.0740653 | 0 | 1 | 0.354345 | 364 | 0.757369136612022 |
| Chr5:174.1Mb | 920 | 5 | 173606996 | 174662885 | 2936 | 333 | AD | eGFR | -0.257687 | -1 | 0.3495 | 0.0664025 | 0 | 1 | 0.354395 | 365 | 0.757369136612022 |
| Chr8:3.2Mb | 1238 | 8 | 2970492 | 3386667 | 2443 | 264 | AD | eGFR | -0.310904 | -1 | 0.42221 | 0.0966614 | 0 | 1 | 0.354472 | 366 | 0.757369136612022 |
| Chr17:5.9Mb | 2179 | 17 | 5573785 | 6305078 | 2253 | 306 | AD | eGFR | -0.278539 | -1 | 0.39364 | 0.077584 | 0 | 1 | 0.359316 | 367 | 0.763878 |
| Chr16:19.7Mb | 2121 | 16 | 19104258 | 20351261 | 2995 | 284 | AD | eGFR | 0.153471 | -0.20177 | 0.68758 | 0.0235533 | 0 | 0.49035 | 0.359472 | 368 | 0.763878 |
| Chr9:101Mb | 1445 | 9 | 100310518 | 101596750 | 2994 | 292 | AD | eGFR | -0.151075 | -0.50632 | 0.18213 | 0.0228235 | 0 | 0.25888 | 0.361669 | 369 | 0.766463842818428 |
| Chr8:102Mb | 1330 | 8 | 101448440 | 102548680 | 2304 | 311 | AD | eGFR | -0.189882 | -0.68312 | 0.24714 | 0.0360552 | 0 | 0.47104 | 0.364732 | 370 | 0.767970247311828 |
| Chr20:16.3Mb | 2386 | 20 | 15962577 | 16661516 | 2184 | 220 | AD | eGFR | 0.360053 | -0.575 | 1 | 0.129638 | 0 | 1 | 0.365173 | 371 | 0.767970247311828 |
| Chr4:3.9Mb | 600 | 4 | 3549230 | 4266178 | 1600 | 237 | AD | eGFR | 0.163843 | -0.21276 | 0.67164 | 0.0268445 | 0 | 0.46147 | 0.365326 | 372 | 0.767970247311828 |
| Chr19:20.1Mb | 2328 | 19 | 19749167 | 20485499 | 1963 | 173 | AD | eGFR | 0.198185 | -0.24319 | 0.76082 | 0.0392773 | 0 | 0.58628 | 0.369251 | 373 | 0.774140166219839 |
| Chr2:1.6Mb | 203 | 2 | 1376546 | 1875060 | 1736 | 223 | AD | eGFR | -0.240256 | -1 | 0.33238 | 0.0577229 | 0 | 1 | 0.372248 | 374 | 0.77833672727272 |

| locus_label | locus | chr | start | stop | n.snps | n.pcs | phen1 | phen2 | rho | rho.lower | rho.upper | r2 | r2.lower | r2.upper | p | rank | FDR |
|---|---|---|---|---|---|---|---|---|---|---|---|---|---|---|---|---|---|
| | | | | | | | | | | | | | | | | | 7 |
| Chr19:13.6Mb | 2320 | 19 | 13213187 | 13968319 | 1717 | 284 | AD | eGFR | -0.130781 | -0.45228 | 0.1697 | 0.0171037 | 0 | 0.20491 | 0.375707 | 375 | 0.781722783068783 |
| Chr2:44.6Mb | 249 | 2 | 44104111 | 45189468 | 2277 | 250 | AD | eGFR | -0.20335 | -0.74769 | 0.27698 | 0.0413513 | 0 | 0.56552 | 0.37751 | 376 | 0.781722783068783 |
| Chr10:8.2Mb | 1495 | 10 | 7659873 | 8771920 | 3504 | 323 | AD | eGFR | 0.298825 | -0.42516 | 1 | 0.0892963 | 0 | 1 | 0.377762 | 377 | 0.781722783068783 |
| Chr12:78.6Mb | 1812 | 12 | 77800464 | 79315178 | 3363 | 302 | AD | eGFR | 0.199809 | -0.2697 | 0.75275 | 0.0399236 | 0 | 0.56829 | 0.377866 | 378 | 0.781722783068783 |
| Chr20:49.9Mb | 2413 | 20 | 49236419 | 50653620 | 3625 | 338 | AD | eGFR | -0.214162 | -0.84918 | 0.29518 | 0.0458654 | 0 | 0.72833 | 0.380306 | 379 | 0.784694701846966 |
| Chr18:47.8Mb | 2276 | 18 | 47455926 | 48228379 | 1811 | 205 | AD | eGFR | -0.257403 | -1 | 0.37931 | 0.0662563 | 0 | 1 | 0.383853 | 380 | 0.785217604166667 |
| Chr4:84.7Mb | 676 | 4 | 84003063 | 85437805 | 2880 | 273 | AD | eGFR | 0.323944 | -0.52404 | 1 | 0.104939 | 0 | 1 | 0.384392 | 381 | 0.785217604166667 |
| Chr4:79.4Mb | 672 | 4 | 78967716 | 79880101 | 1887 | 177 | AD | eGFR | 0.209851 | -0.30962 | 0.86083 | 0.0440375 | 0 | 0.75395 | 0.385077 | 382 | 0.785217604166667 |
| Chr4:110.9Mb | 698 | 4 | 109978983 | 111733579 | 3632 | 307 | AD | eGFR | 0.182095 | -0.25362 | 0.69111 | 0.0331587 | 0 | 0.48176 | 0.38519 | 383 | 0.785217604166667 |
| Chr1:159.4Mb | 116 | 1 | 158900370 | 159882515 | 2244 | 223 | AD | eGFR | 0.306023 | -0.4846 | 1 | 0.0936501 | 0 | 1 | 0.38558 | 384 | 0.785217604166667 |
| Chr10:13.6Mb | 1501 | 10 | 13208062 | 14008135 | 2200 | 317 | AD | eGFR | 0.189776 | -0.27608 | 0.76427 | 0.0360148 | 0 | 0.58771 | 0.392001 | 385 | 0.794159492227979 |
| Chr2:31Mb | 233 | 2 | 30575620 | 31457418 | 2614 | 252 | AD | eGFR | 0.310329 | -0.53078 | 1 | 0.0963038 | 0 | 1 | 0.392002 | 386 | 0.794159492227979 |
| Chr9:4.3Mb | 1376 | 9 | 4025356 | 4596614 | 1881 | 289 | AD | eGFR | 0.146491 | -0.19593 | 0.53205 | 0.0214596 | 0 | 0.28476 | 0.395035 | 387 | 0.797332360406091 |
| Chr3:168Mb | 569 | 3 | 167118894 | 168972662 | 3351 | 291 | AD | eGFR | -0.193348 | -0.78521 | 0.26889 | 0.0373836 | 0 | 0.62036 | 0.396786 | 388 | 0.797332360406091 |
| Chr9:13.1Mb | 1387 | 9 | 12664859 | 13512852 | 1674 | 269 | AD | eGFR | 0.270231 | -0.41245 | 1 | 0.0730249 | 0 | 1 | 0.39684 | 389 | 0.797332360406091 |
| Chr5:73.8Mb | 836 | 5 | 73314062 | 74245354 | 2137 | 266 | AD | eGFR | -0.283593 | -1 | 0.46115 | 0.0804248 | 0 | 1 | 0.400957 | 390 | 0.797332360406091 |
| Chr6:160.1Mb | 1083 | 6 | 159604053 | 160583918 | 2463 | 241 | AD | eGFR | -0.1456 | -0.68793 | 0.23134 | 0.0211994 | 0 | 0.47726 | 0.400995 | 391 | 0.797332360406091 |
| Chr7:79.4Mb | 1170 | 7 | 78685010 | 80055721 | 2713 | 227 | AD | eGFR | -0.303847 | -1 | 0.50905 | 0.0923232 | 0 | 1 | 0.401222 | 392 | 0.797332360406091 |
| Chr1:109.7Mb | 96 | 1 | 109271450 | 110224230 | 1889 | 223 | AD | eGFR | 0.13597 | -0.2096 | 0.51468 | 0.0184878 | 0 | 0.26694 | 0.401549 | 393 | 0.797332360406091 |
| Chr3:194.7Mb | 591 | 3 | 194217141 | 195196789 | 3420 | 305 | AD | eGFR | 0.202771 | -0.31539 | 0.87244 | 0.0411163 | 0 | 0.80647 | 0.401725 | 394 | 0.797332360406091 |
| Chr19:1.9Mb | 2308 | 19 | 1515921 | 2253174 | 1803 | 302 | AD | eGFR | 0.151917 | -0.23249 | 0.68654 | 0.0230786 | 0 | 0.48093 | 0.405335 | 395 | 0.799604824120603 |
| Chr2:105.8Mb | 301 | 2 | 105120579 | 106546524 | 3448 | 313 | AD | eGFR | 0.190564 | -0.29134 | 0.72372 | 0.0363145 | 0 | 0.52873 | 0.405761 | 396 | 0.799604824120603 |
| Chr2:111.4Mb | 305 | 2 | 110097869 | 112644828 | 2641 | 281 | AD | eGFR | -0.139622 | -0.51672 | 0.20567 | 0.0194943 | 0 | 0.27179 | 0.406031 | 397 | 0.799604824120603 |

| locus_label | locus | chr | start | stop | n.snps | n.pcs | phen1 | phen2 | rho | rho.lower | rho.upper | r2 | r2.lower | r2.upper | p | rank | FDR |
|---|---|---|---|---|---|---|---|---|---|---|---|---|---|---|---|---|---|
| Chr19:49Mb | 2355 | 19 | 48573066 | 49415189 | 2371 | 261 | AD | eGFR | -0.173442 | -0.81846 | 0.26862 | 0.030082 | 0 | 0.70089 | 0.40696 | 398 | 0.799604824120603 |
| Chr5:9.9Mb | 785 | 5 | 9203856 | 10549029 | 3276 | 346 | AD | eGFR | 0.27296 | -0.46398 | 1 | 0.0745071 | 0 | 1 | 0.408111 | 399 | 0.799856646616541 |
| Chr6:152Mb | 1074 | 6 | 151629955 | 152376499 | 1919 | 223 | AD | eGFR | -0.23807 | -1 | 0.38455 | 0.0566773 | 0 | 1 | 0.409862 | 400 | 0.80128021 |
| Chr19:47.3Mb | 2353 | 19 | 46765061 | 47752727 | 2100 | 284 | AD | eGFR | -0.129812 | -0.47545 | 0.19136 | 0.0168511 | 0 | 0.22693 | 0.411479 | 401 | 0.801404424821002 |
| Chr8:55.8Mb | 1292 | 8 | 55275355 | 56346878 | 2365 | 214 | AD | eGFR | 0.332905 | -0.60061 | 1 | 0.110826 | 0 | 1 | 0.416358 | 402 | 0.801404424821002 |
| Chr10:2.9Mb | 1488 | 10 | 2637066 | 3225336 | 2005 | 260 | AD | eGFR | -0.181517 | -0.70094 | 0.275 | 0.0329485 | 0 | 0.49425 | 0.416828 | 403 | 0.801404424821002 |
| Chr11:128.9Mb | 1733 | 11 | 128262551 | 129451784 | 2557 | 313 | AD | eGFR | 0.167254 | -0.27846 | 0.78984 | 0.0279738 | 0 | 0.64276 | 0.417826 | 404 | 0.801404424821002 |
| Chr15:38.6Mb | 2048 | 15 | 37962916 | 39238840 | 2717 | 311 | AD | eGFR | 0.148812 | -0.23776 | 0.55113 | 0.0221449 | 0 | 0.30729 | 0.419008 | 405 | 0.801404424821002 |
| Chr11:2.2Mb | 1612 | 11 | 1857846 | 2477449 | 1788 | 263 | AD | eGFR | 0.14613 | -0.2335 | 0.66543 | 0.0213538 | 0 | 0.46697 | 0.419088 | 406 | 0.801404424821002 |
| Chr19:19.1Mb | 2327 | 19 | 18504868 | 19749166 | 2219 | 236 | AD | eGFR | 0.17111 | -0.26422 | 0.85733 | 0.0292787 | 0 | 0.76663 | 0.419305 | 407 | 0.801404424821002 |
| Chr4:8.6Mb | 606 | 4 | 8309827 | 8882616 | 1316 | 198 | AD | eGFR | -0.138598 | -0.49396 | 0.20335 | 0.0192095 | 0 | 0.24552 | 0.420196 | 408 | 0.801404424821002 |
| Chr5:146.2Mb | 896 | 5 | 145319097 | 147180171 | 3572 | 302 | AD | eGFR | 0.202332 | -0.32669 | 0.85691 | 0.040938 | 0 | 0.7578 | 0.420555 | 409 | 0.801404424821002 |
| Chr17:26.6Mb | 2195 | 17 | 25872111 | 27344401 | 1913 | 265 | AD | eGFR | -0.188056 | -0.88004 | 0.32671 | 0.0353649 | 0 | 0.81033 | 0.420666 | 410 | 0.801404424821002 |
| Chr7:26.5Mb | 1125 | 7 | 25671576 | 27351286 | 3349 | 285 | AD | eGFR | -0.164002 | -0.77254 | 0.27317 | 0.0268968 | 0 | 0.62084 | 0.422119 | 411 | 0.801404424821002 |
| Chr8:108.2Mb | 1336 | 8 | 107787164 | 108647463 | 1838 | 181 | AD | eGFR | 0.219084 | -0.36869 | 0.99895 | 0.047998 | 0 | 1 | 0.422252 | 412 | 0.801404424821002 |
| Chr8:78Mb | 1312 | 8 | 77377990 | 78633659 | 2774 | 199 | AD | eGFR | 0.174466 | -0.27534 | 0.66813 | 0.0304384 | 0 | 0.44937 | 0.425185 | 413 | 0.801404424821002 |
| Chr12:22.3Mb | 1762 | 12 | 21572253 | 22928055 | 2664 | 257 | AD | eGFR | 0.240504 | -0.41248 | 1 | 0.0578422 | 0 | 1 | 0.425883 | 414 | 0.801404424821002 |
| Chr11:132Mb | 1736 | 11 | 131424362 | 132584401 | 2850 | 325 | AD | eGFR | 0.275185 | -0.51512 | 1 | 0.0757265 | 0 | 1 | 0.426231 | 415 | 0.801404424821002 |
| Chr5:147.9Mb | 897 | 5 | 147180172 | 148662214 | 3422 | 282 | AD | eGFR | 0.162997 | -0.27274 | 0.71295 | 0.0265681 | 0 | 0.52199 | 0.426648 | 416 | 0.801404424821002 |
| Chr3:61.9Mb | 474 | 3 | 61277588 | 62609818 | 2867 | 342 | AD | eGFR | 0.243077 | -0.42887 | 1 | 0.0590863 | 0 | 1 | 0.428236 | 417 | 0.801404424821002 |
| Chr15:95.1Mb | 2094 | 15 | 94565522 | 95662425 | 2777 | 347 | AD | eGFR | -0.163382 | -0.6308 | 0.25829 | 0.0266936 | 0 | 0.40238 | 0.428603 | 418 | 0.801404424821002 |
| Chr3:30.6Mb | 450 | 3 | 29877023 | 31269697 | 3560 | 303 | AD | eGFR | -0.205092 | -1 | 0.34925 | 0.0420627 | 0 | 1 | 0.429397 | 419 | 0.801404424821002 |
| Chr16:89.7Mb | 2172 | 16 | 89106305 | 90292787 | 3021 | 262 | AD | eGFR | 0.120704 | -0.19526 | 0.48363 | 0.0145695 | 0 | 0.23499 | 0.433212 | 420 | 0.806599485714286 |
| Chr17:49.3M | 221 | 17 | 48722591 | 49909298 | 2207 | 276 | AD | eGFR | -0.159645 | -0.62414 | 0.26679 | 0.0254866 | 0 | 0.39528 | 0.43676 | 421 | 0.80930899290780 |

| locus_label | locus | chr | start | stop | n.snps | n.pcs | phen1 | phen2 | rho | rho.lower | rho.upper | r2 | r2.lower | r2.upper | p | rank | FDR |
|---|---|---|---|---|---|---|---|---|---|---|---|---|---|---|---|---|---|
| b | 1 | | | | | | | | | | | | | | | | 1 |
| Chr3:196.8Mb | 594 | 3 | 196383153 | 197147990 | 1844 | 247 | AD | eGFR | 0.153368 | -0.26158 | 0.63078 | 0.0235218 | 0 | 0.40754 | 0.437448 | 422 | 0.809308992907801 |
| Chr17:67.5Mb | 2224 | 17 | 66757557 | 68176219 | 2462 | 251 | AD | eGFR | 0.170255 | -0.2858 | 0.66899 | 0.0289867 | 0 | 0.45322 | 0.437772 | 423 | 0.809308992907801 |
| Chr11:124.8Mb | 1729 | 11 | 124403478 | 125266684 | 1940 | 292 | AD | eGFR | 0.174334 | -0.30243 | 0.7606 | 0.0303923 | 0 | 0.59142 | 0.439083 | 424 | 0.809818174528302 |
| Chr14:76.2Mb | 2005 | 14 | 75699502 | 76694201 | 1973 | 260 | AD | eGFR | 0.306083 | -0.63008 | 1 | 0.0936866 | 0 | 1 | 0.441559 | 425 | 0.81246856 |
| Chr13:36.2Mb | 1878 | 13 | 35744569 | 36579413 | 2062 | 216 | AD | eGFR | -0.256555 | -1 | 0.50752 | 0.0658207 | 0 | 1 | 0.442764 | 426 | 0.812773352112676 |
| Chr7:0.3Mb | 1094 | 7 | 15064 | 617319 | 888 | 176 | AD | eGFR | -0.151816 | -0.67262 | 0.26563 | 0.023048 | 0 | 0.4645 | 0.446703 | 427 | 0.818083714285714 |
| Chr5:4.2Mb | 780 | 5 | 3677596 | 4636542 | 2163 | 268 | AD | eGFR | 0.138412 | -0.24798 | 0.59714 | 0.0191579 | 0 | 0.36111 | 0.451467 | 428 | 0.824876621495327 |
| Chr7:17.2Mb | 1116 | 7 | 16739014 | 17596578 | 2247 | 294 | AD | eGFR | -0.178021 | -0.84603 | 0.33846 | 0.0316914 | 0 | 0.75167 | 0.454489 | 429 | 0.827750637209302 |
| Chr5:149.3Mb | 898 | 5 | 148662215 | 149850203 | 2424 | 322 | AD | eGFR | 0.182274 | -0.3386 | 0.82446 | 0.0332238 | 0 | 0.69996 | 0.455157 | 430 | 0.827750637209302 |
| Chr5:33.3Mb | 805 | 5 | 32727915 | 33971056 | 3022 | 254 | AD | eGFR | 0.250677 | -0.51848 | 1 | 0.0628391 | 0 | 1 | 0.456262 | 431 | 0.827834997679814 |
| Chr3:160Mb | 561 | 3 | 159478752 | 160584961 | 2044 | 172 | AD | eGFR | 0.251135 | -0.52108 | 1 | 0.0630689 | 0 | 1 | 0.459543 | 432 | 0.830431625866051 |
| Chr11:74.9Mb | 1681 | 11 | 74399332 | 75445253 | 1955 | 235 | AD | eGFR | 0.195033 | -0.37949 | 0.94282 | 0.0380377 | 0 | 0.92471 | 0.459817 | 433 | 0.830431625866051 |
| Chr10:22.4Mb | 1510 | 10 | 21477693 | 23365685 | 2829 | 251 | AD | eGFR | 0.213855 | -0.41834 | 1 | 0.0457341 | 0 | 1 | 0.469477 | 434 | 0.843028871681416 |
| Chr8:134.9Mb | 1359 | 8 | 134509449 | 135313673 | 2675 | 285 | AD | eGFR | -0.17329 | -0.76729 | 0.33559 | 0.0300294 | 0 | 0.59949 | 0.470431 | 435 | 0.843028871681416 |
| Chr7:7.5Mb | 1103 | 7 | 7167611 | 7926500 | 2271 | 264 | AD | eGFR | -0.249073 | -1 | 0.53718 | 0.0620372 | 0 | 1 | 0.47256 | 436 | 0.843028871681416 |
| Chr8:142.1Mb | 1366 | 8 | 141527882 | 142611889 | 2539 | 289 | AD | eGFR | -0.201254 | -1 | 0.4172 | 0.040503 | 0 | 1 | 0.473819 | 437 | 0.843028871681416 |
| Chr2:8.5Mb | 212 | 2 | 7966710 | 8978798 | 2064 | 303 | AD | eGFR | -0.111985 | -0.44962 | 0.20428 | 0.0125407 | 0 | 0.20377 | 0.473961 | 438 | 0.843028871681416 |
| Chr7:109.8Mb | 1194 | 7 | 109095559 | 110479479 | 2964 | 272 | AD | eGFR | -0.149642 | -0.61936 | 0.29002 | 0.0223926 | 0 | 0.38967 | 0.475607 | 439 | 0.843028871681416 |
| Chr4:146.7Mb | 731 | 4 | 146174872 | 147161042 | 2119 | 257 | AD | eGFR | 0.112421 | -0.20445 | 0.44393 | 0.0126384 | 0 | 0.20022 | 0.475948 | 440 | 0.843028871681416 |
| Chr9:107.6Mb | 1451 | 9 | 107061020 | 108203095 | 3311 | 275 | AD | eGFR | 0.137606 | -0.27428 | 0.58175 | 0.0189355 | 0 | 0.34395 | 0.477484 | 441 | 0.843028871681416 |
| Chr13:51Mb | 1893 | 13 | 50419897 | 51554236 | 1988 | 219 | AD | eGFR | -0.144164 | -0.74617 | 0.29603 | 0.0207834 | 0 | 0.58674 | 0.478816 | 442 | 0.843028871681416 |
| Chr3:43.2Mb | 461 | 3 | 42301658 | 44141693 | 3395 | 306 | AD | eGFR | -0.251914 | -1 | 0.57622 | 0.0634605 | 0 | 1 | 0.479211 | 443 | 0.843028871681416 |
| Chr1:21.8Mb | 23 | 1 | 21416122 | 22145629 | 1987 | 224 | AD | eGFR | -0.165877 | -0.7233 | 0.32255 | 0.0275153 | 0 | 0.54075 | 0.480433 | 444 | 0.843028871681416 |

| locus_label | locus | chr | start | stop | n.snps | n.pcs | phen1 | phen2 | rho | rho.lower | rho.upper | r2 | r2.lower | r2.upper | p | rank | FDR |
|---|---|---|---|---|---|---|---|---|---|---|---|---|---|---|---|---|---|
| Chr2:143.9Mb | 334 | 2 | 143259781 | 144503688 | 2260 | 227 | AD | eGFR | -0.12797 | -0.52197 | 0.24125 | 0.0163764 | 0 | 0.27455 | 0.480536 | 445 | 0.843028871681416 |
| Chr1:167.7Mb | 125 | 1 | 167163955 | 168179086 | 1983 | 276 | AD | eGFR | 0.242961 | -0.56615 | 1 | 0.05903 | 0 | 1 | 0.482731 | 446 | 0.843028871681416 |
| Chr18:58.1Mb | 2286 | 18 | 57635822 | 58527236 | 2118 | 190 | AD | eGFR | 0.282139 | -0.70268 | 1 | 0.0796026 | 0 | 1 | 0.484652 | 447 | 0.843028871681416 |
| Chr6:143.9Mb | 1067 | 6 | 143083348 | 144791455 | 3119 | 327 | AD | eGFR | 0.182295 | -0.37708 | 0.85565 | 0.0332316 | 0 | 0.74677 | 0.48476 | 448 | 0.843028871681416 |
| Chr12:49.7Mb | 1788 | 12 | 48960266 | 50344346 | 2374 | 238 | AD | eGFR | 0.103078 | -0.1961 | 0.42178 | 0.0106251 | 0 | 0.17964 | 0.48494 | 449 | 0.843028871681416 |
| Chr4:154.3Mb | 737 | 4 | 153520884 | 155044409 | 3700 | 332 | AD | eGFR | -0.165531 | -0.78658 | 0.34065 | 0.0274003 | 0 | 0.63895 | 0.485129 | 450 | 0.843028871681416 |
| Chr2:87.7Mb | 292 | 2 | 86867033 | 88547312 | 967 | 151 | AD | eGFR | 0.185955 | -0.3999 | 0.99083 | 0.0345794 | 0 | 1 | 0.487233 | 451 | 0.843028871681416 |
| Chr10:90.5Mb | 1569 | 10 | 89971629 | 91021321 | 2238 | 253 | AD | eGFR | 0.166672 | -0.34302 | 0.75032 | 0.0277796 | 0 | 0.57467 | 0.487275 | 452 | 0.843028871681416 |
| Chr1:162.5Mb | 119 | 1 | 161945443 | 163141862 | 2924 | 288 | AD | eGFR | -0.128755 | -0.53943 | 0.25609 | 0.0165777 | 0 | 0.29884 | 0.490149 | 453 | 0.844568593406593 |
| Chr1:57.2Mb | 49 | 1 | 56799637 | 57529670 | 1967 | 238 | AD | eGFR | 0.163344 | -0.33804 | 0.77209 | 0.0266813 | 0 | 0.60471 | 0.490325 | 454 | 0.844568593406593 |
| Chr19:0.4Mb | 2306 | 19 | 64705 | 762046 | 1667 | 310 | AD | eGFR | -0.104417 | -0.46634 | 0.20617 | 0.0109029 | 0 | 0.22103 | 0.491405 | 455 | 0.844568593406593 |
| Chr11:69.3Mb | 1677 | 11 | 68887345 | 69713996 | 2067 | 265 | AD | eGFR | 0.10214 | -0.20382 | 0.44281 | 0.0104325 | 0 | 0.19904 | 0.49556 | 456 | 0.848881489082969 |
| Chr21:38.6Mb | 2452 | 21 | 38046450 | 39252059 | 2985 | 279 | AD | eGFR | 0.148125 | -0.32155 | 0.70399 | 0.0219409 | 0 | 0.51956 | 0.497133 | 457 | 0.848881489082969 |
| Chr10:103.4Mb | 1580 | 10 | 102509299 | 104206837 | 2486 | 263 | AD | eGFR | 0.135348 | -0.29903 | 0.65137 | 0.0183192 | 0 | 0.44254 | 0.497171 | 458 | 0.848881489082969 |
| Chr7:50.3Mb | 1146 | 7 | 49632427 | 50894508 | 3192 | 241 | AD | eGFR | -0.129789 | -0.56076 | 0.2538 | 0.0168453 | 0 | 0.32024 | 0.498269 | 459 | 0.848902740740741 |
| Chr8:133Mb | 1357 | 8 | 132534637 | 133461551 | 2443 | 242 | AD | eGFR | 0.227105 | -0.54755 | 1 | 0.0515766 | 0 | 1 | 0.501007 | 460 | 0.8517119 |
| Chr6:90.5Mb | 1022 | 6 | 89974268 | 91097024 | 2047 | 255 | AD | eGFR | -0.134222 | -0.63622 | 0.29001 | 0.0180157 | 0 | 0.42523 | 0.503925 | 461 | 0.852142726495727 |
| Chr13:29.3Mb | 1873 | 13 | 28587126 | 30068525 | 3391 | 311 | AD | eGFR | 0.202147 | -0.48403 | 1 | 0.0408635 | 0 | 1 | 0.503989 | 462 | 0.852142726495727 |
| Chr7:144.9Mb | 1218 | 7 | 144324465 | 145415775 | 2643 | 230 | AD | eGFR | 0.242186 | -0.62744 | 1 | 0.058654 | 0 | 1 | 0.505773 | 463 | 0.852142726495727 |
| Chr9:14.6Mb | 1389 | 9 | 14244748 | 14902924 | 1845 | 305 | AD | eGFR | -0.103763 | -0.4297 | 0.20927 | 0.0107669 | 0 | 0.18689 | 0.506407 | 464 | 0.852142726495727 |
| Chr9:113.3Mb | 1457 | 9 | 112865521 | 113646149 | 2293 | 238 | AD | eGFR | -0.186416 | -1 | 0.43855 | 0.0347508 | 0 | 1 | 0.507325 | 465 | 0.852142726495727 |
| Chr13:43.4Mb | 1886 | 13 | 42950158 | 43931931 | 2095 | 262 | AD | eGFR | -0.129391 | -0.5639 | 0.27484 | 0.0167419 | 0 | 0.32363 | 0.508656 | 466 | 0.852142726495727 |
| Chr5:32.2Mb | 804 | 5 | 31679465 | 32727914 | 2804 | 318 | AD | eGFR | 0.194258 | -0.47717 | 1 | 0.0377361 | 0 | 1 | 0.509445 | 467 | 0.852142726495727 |
| Chr1:133.1M | 106 | 1 | 12034506 | 14580384 | 282 | 56 | AD | eGFR | 0.194636 | -0.48077 | 1 | 0.0378832 | 0 | 1 | 0.509978 | 468 | 0.85214272649572 |

| locus_label | locus | chr | start | stop | n.snps | n.pcs | phen1 | phen2 | rho | rho.lower | rho.upper | r2 | r2.lower | r2.upper | p | rank | FDR |
|---|---|---|---|---|---|---|---|---|---|---|---|---|---|---|---|---|---|
| b | | | 7 | 8 | | | | | | | | | | | | | 7 |
| Chr21:37.7Mb | 2451 | 21 | 37303549 | 38046449 | 2261 | 207 | AD | eGFR | -0.157553 | -0.83608 | 0.37477 | 0.0248231 | 0 | 0.73993 | 0.512406 | 469 | 0.853910723404255 |
| Chr2:148.4Mb | 337 | 2 | 147252197 | 149568875 | 3620 | 268 | AD | eGFR | 0.143488 | -0.32853 | 0.74829 | 0.0205888 | 0 | 0.59943 | 0.51322 | 470 | 0.853910723404255 |
| Chr15:46.5Mb | 2053 | 15 | 46022643 | 46884071 | 1892 | 189 | AD | eGFR | -0.139127 | -0.77768 | 0.32356 | 0.0193562 | 0 | 0.63095 | 0.514508 | 471 | 0.854236212314225 |
| Chr6:105.5Mb | 1036 | 6 | 104951345 | 106053915 | 2046 | 220 | AD | eGFR | 0.150759 | -0.34916 | 0.71045 | 0.0227282 | 0 | 0.5213 | 0.516218 | 472 | 0.855251742071882 |
| Chr15:53.8Mb | 2059 | 15 | 53263127 | 54332799 | 2965 | 238 | AD | eGFR | -0.131596 | -0.72964 | 0.32184 | 0.0173175 | 0 | 0.58403 | 0.517307 | 473 | 0.855251742071882 |
| Chr18:4.2Mb | 2242 | 18 | 3722829 | 4603961 | 2294 | 328 | AD | eGFR | 0.157155 | -0.38091 | 0.85657 | 0.0246976 | 0 | 0.79917 | 0.520952 | 474 | 0.857561571428571 |
| Chr9:86Mb | 1430 | 9 | 85135752 | 86769886 | 3590 | 311 | AD | eGFR | 0.147059 | -0.33601 | 0.69136 | 0.0216262 | 0 | 0.48773 | 0.521446 | 475 | 0.857561571428571 |
| Chr10:101.9Mb | 1579 | 10 | 101271452 | 102509298 | 2199 | 225 | AD | eGFR | -0.145825 | -0.80346 | 0.35296 | 0.0212648 | 0 | 0.67933 | 0.521994 | 476 | 0.857561571428571 |
| Chr15:54.7Mb | 2060 | 15 | 54332800 | 55136717 | 2693 | 201 | AD | eGFR | 0.148997 | -0.33915 | 0.70234 | 0.0222 | 0 | 0.50966 | 0.523363 | 477 | 0.858008104821803 |
| Chr22:36.7Mb | 2481 | 22 | 36135969 | 37364004 | 2851 | 312 | AD | eGFR | 0.149168 | -0.3635 | 0.8476 | 0.0222511 | 0 | 0.75971 | 0.524593 | 478 | 0.858225368200837 |
| Chr3:5.2Mb | 427 | 3 | 4835084 | 5496182 | 1765 | 309 | AD | eGFR | -0.107813 | -0.51066 | 0.24117 | 0.0116235 | 0 | 0.26743 | 0.526639 | 479 | 0.859773899791232 |
| Chr7:151.6Mb | 1224 | 7 | 150897819 | 152253294 | 2586 | 343 | AD | eGFR | 0.0920195 | -0.22802 | 0.46356 | 0.00846759 | 0 | 0.23152 | 0.530426 | 480 | 0.863202814968815 |
| Chr1:242.7Mb | 194 | 1 | 242417924 | 243048003 | 1847 | 216 | AD | eGFR | -0.143317 | -0.68021 | 0.32567 | 0.0205399 | 0 | 0.47747 | 0.530947 | 481 | 0.863202814968815 |
| Chr11:12.1Mb | 1624 | 11 | 11632651 | 12470449 | 2430 | 315 | AD | eGFR | 0.164632 | -0.40981 | 0.93783 | 0.0271036 | 0 | 0.94858 | 0.532778 | 482 | 0.864382564315353 |
| Chr22:21.7Mb | 2466 | 22 | 20969185 | 22404422 | 2188 | 239 | AD | eGFR | -0.130258 | -0.60851 | 0.31321 | 0.0169672 | 0 | 0.38528 | 0.533937 | 483 | 0.864469428571428 |
| Chr22:46.8Mb | 2490 | 22 | 46351049 | 47299625 | 2527 | 303 | AD | eGFR | 0.100105 | -0.23033 | 0.44718 | 0.010021 | 0 | 0.20208 | 0.538001 | 484 | 0.869249549586777 |
| Chr22:47.8Mb | 2491 | 22 | 47299626 | 48269177 | 2704 | 306 | AD | eGFR | 0.13802 | -0.34311 | 0.68531 | 0.0190495 | 0 | 0.48659 | 0.539779 | 485 | 0.870324078350516 |
| Chr13:111.3Mb | 1949 | 13 | 110995433 | 111621244 | 1956 | 271 | AD | eGFR | -0.131051 | -0.71521 | 0.34031 | 0.0171743 | 0 | 0.53832 | 0.542008 | 486 | 0.87157031211499 |
| Chr20:20.8Mb | 2391 | 20 | 19957929 | 21682377 | 2971 | 287 | AD | eGFR | -0.144379 | -0.75452 | 0.36772 | 0.0208453 | 0 | 0.57631 | 0.542781 | 487 | 0.87157031211499 |
| Chr22:43.2Mb | 2486 | 22 | 42867928 | 43574017 | 1926 | 214 | AD | eGFR | 0.11812 | -0.31921 | 0.64195 | 0.0139523 | 0 | 0.43934 | 0.548328 | 488 | 0.877004204498978 |
| Chr16:70.3Mb | 2148 | 16 | 69549150 | 71062712 | 1862 | 194 | AD | eGFR | 0.0683462 | -0.16052 | 0.30288 | 0.00467121 | 0 | 0.09258 | 0.548408 | 489 | 0.877004204498978 |
| Chr17:78.9Mb | 2235 | 17 | 78438333 | 79333774 | 2875 | 276 | AD | eGFR | -0.159321 | -0.85826 | 0.42372 | 0.0253833 | 0 | 0.78036 | 0.551269 | 490 | 0.877494350597609 |
| Chr2:171.2Mb | 355 | 2 | 170865766 | 171566482 | 1981 | 270 | AD | eGFR | 0.189501 | -0.5363 | 1 | 0.0359106 | 0 | 1 | 0.554302 | 491 | 0.877494350597609 |

| locus_label | locus | chr | start | stop | n.snps | n.pcs | phen1 | phen2 | rho | rho.lower | rho.upper | r2 | r2.lower | r2.upper | p | rank | FDR |
|---|---|---|---|---|---|---|---|---|---|---|---|---|---|---|---|---|---|
| Chr18:56.3Mb | 2284 | 18 | 55809316 | 56832312 | 2595 | 283 | AD | eGFR | -0.196871 | -1 | 0.57691 | 0.0387582 | 0 | 1 | 0.554348 | 492 | 0.877494350597609 |
| Chr2:60Mb | 267 | 2 | 59251997 | 60775066 | 3338 | 315 | AD | eGFR | -0.135453 | -0.70359 | 0.34727 | 0.0183476 | 0 | 0.51525 | 0.555163 | 493 | 0.877494350597609 |
| Chr12:121.2Mb | 1849 | 12 | 120567741 | 121817509 | 3164 | 233 | AD | eGFR | 0.10556 | -0.27796 | 0.52608 | 0.011143 | 0 | 0.29486 | 0.557098 | 494 | 0.877494350597609 |
| Chr14:35.9Mb | 1968 | 14 | 35379989 | 36422077 | 2166 | 164 | AD | eGFR | 0.185462 | -0.53774 | 1 | 0.0343962 | 0 | 1 | 0.558385 | 495 | 0.877494350597609 |
| Chr13:67.1Mb | 1906 | 13 | 66382287 | 67718879 | 2656 | 235 | AD | eGFR | 0.169326 | -0.50214 | 0.95446 | 0.0286712 | 0 | 0.9592 | 0.560064 | 496 | 0.877494350597609 |
| Chr19:6.9Mb | 2314 | 19 | 6455429 | 7249359 | 2532 | 328 | AD | eGFR | 0.149952 | -0.43812 | 0.9168 | 0.0224855 | 0 | 0.93499 | 0.560519 | 497 | 0.877494350597609 |
| Chr3:46.7Mb | 463 | 3 | 45844192 | 47588461 | 3151 | 218 | AD | eGFR | 0.146609 | -0.40201 | 0.83372 | 0.0214941 | 0 | 0.73539 | 0.560767 | 498 | 0.877494350597609 |
| Chr16:72.6Mb | 2150 | 16 | 72089512 | 73140781 | 1713 | 239 | AD | eGFR | -0.128683 | -0.76695 | 0.35347 | 0.0165593 | 0 | 0.64394 | 0.561485 | 499 | 0.877494350597609 |
| Chr8:72.5Mb | 1307 | 8 | 72013185 | 72917489 | 2000 | 277 | AD | eGFR | -0.133081 | -0.67554 | 0.34944 | 0.0177105 | 0 | 0.47097 | 0.562051 | 500 | 0.877494350597609 |
| Chr13:37.9Mb | 1880 | 13 | 37499811 | 38290689 | 1922 | 160 | AD | eGFR | -0.186296 | -1 | 0.5635 | 0.034706 | 0 | 1 | 0.562592 | 501 | 0.877494350597609 |
| Chr2:233.7Mb | 409 | 2 | 233352723 | 234115092 | 1906 | 238 | AD | eGFR | -0.116095 | -0.58221 | 0.29882 | 0.0134781 | 0 | 0.35038 | 0.563302 | 502 | 0.877494350597609 |
| Chr1:203.3Mb | 156 | 1 | 202583885 | 204092537 | 3115 | 333 | AD | eGFR | 0.163228 | -0.46883 | 1 | 0.0266434 | 0 | 1 | 0.564885 | 503 | 0.878210874751491 |
| Chr14:101.4Mb | 2027 | 14 | 100786190 | 102004377 | 2818 | 358 | AD | eGFR | 0.0912562 | -0.25048 | 0.45774 | 0.0083277 | 0 | 0.22037 | 0.567915 | 504 | 0.878961818181818 |
| Chr6:155Mb | 1078 | 6 | 154462011 | 155523957 | 2834 | 323 | AD | eGFR | 0.116714 | -0.32643 | 0.69679 | 0.0136222 | 0 | 0.53679 | 0.568376 | 505 | 0.878961818181818 |
| Chr9:0.3Mb | 1370 | 9 | 10469 | 653661 | 1728 | 220 | AD | eGFR | -0.147009 | -0.83786 | 0.4123 | 0.0216115 | 0 | 0.76475 | 0.56874 | 506 | 0.878961818181818 |
| Chr21:42.6Mb | 2456 | 21 | 42187153 | 43081434 | 2479 | 329 | AD | eGFR | -0.103744 | -0.51448 | 0.27752 | 0.0107629 | 0 | 0.26979 | 0.575045 | 507 | 0.881388476190476 |
| Chr6:109.8Mb | 1039 | 6 | 109043245 | 110647842 | 3147 | 225 | AD | eGFR | -0.122839 | -0.7201 | 0.35603 | 0.0150895 | 0 | 0.55811 | 0.576183 | 508 | 0.881388476190476 |
| Chr13:106.7Mb | 1944 | 13 | 106313848 | 107037864 | 1970 | 257 | AD | eGFR | 0.121544 | -0.34708 | 0.64079 | 0.014773 | 0 | 0.4335 | 0.579274 | 509 | 0.881388476190476 |
| Chr11:70.5Mb | 1678 | 11 | 69713997 | 71242834 | 3630 | 326 | AD | eGFR | 0.150315 | -0.43825 | 0.83652 | 0.0225945 | 0 | 0.74772 | 0.581174 | 510 | 0.881388476190476 |
| Chr9:139.5Mb | 1483 | 9 | 138995792 | 140097759 | 1944 | 277 | AD | eGFR | -0.0998198 | -0.58259 | 0.29176 | 0.009964 | 0 | 0.35915 | 0.58159 | 511 | 0.881388476190476 |
| Chr14:21.4Mb | 1955 | 14 | 20876081 | 21982654 | 2735 | 374 | AD | eGFR | -0.102647 | -0.52901 | 0.28868 | 0.0105364 | 0 | 0.28954 | 0.584955 | 512 | 0.881388476190476 |
| Chr15:75.4Mb | 2076 | 15 | 74458114 | 76401952 | 2843 | 257 | AD | eGFR | 0.0922865 | -0.2894 | 0.54226 | 0.00851679 | 0 | 0.33293 | 0.5851 | 513 | 0.881388476190476 |
| Chr3:75.8Mb | 487 | 3 | 75374968 | 76157470 | 826 | 87 | AD | eGFR | -0.151669 | -0.84765 | 0.46576 | 0.0230036 | 0 | 0.79445 | 0.586352 | 514 | 0.881388476190476 |
| Chr6:168.2M | 109 | 6 | 16771486 | 16870733 | 2553 | 298 | AD | eGFR | 0.107172 | -0.31383 | 0.55701 | 0.0114859 | 0 | 0.32149 | 0.58801 | 515 | 0.88138847619047 |

| locus_label | locus | chr | start | stop | n.snps | n.pcs | phen1 | phen2 | rho | rho.lower | rho.upper | r2 | r2.lower | r2.upper | p | rank | FDR |
|---|---|---|---|---|---|---|---|---|---|---|---|---|---|---|---|---|---|
| b | 1 | | 6 | 8 | | | | | | | | | | | | | 6 |
| Chr1:9Mb | 11 | 1 | 8580988 | 9475966 | 1775 | 283 | AD | eGFR | 0.126149 | -0.383 | 0.69563 | 0.0159135 | 0 | 0.52115 | 0.588169 | 516 | 0.881388476190476 |
| Chr12:115.1Mb | 1843 | 12 | 114685671 | 115439451 | 2029 | 261 | AD | eGFR | -0.137638 | -0.74842 | 0.39999 | 0.0189441 | 0 | 0.58967 | 0.589225 | 517 | 0.881388476190476 |
| Chr11:121.9Mb | 1726 | 11 | 121208187 | 122589224 | 2741 | 300 | AD | eGFR | -0.116836 | -0.73303 | 0.36526 | 0.0136507 | 0 | 0.59023 | 0.589459 | 518 | 0.881388476190476 |
| Chr12:15.4Mb | 1755 | 12 | 14656850 | 16136742 | 2367 | 258 | AD | eGFR | -0.0966199 | -0.53989 | 0.28808 | 0.00933541 | 0 | 0.32011 | 0.590774 | 519 | 0.881388476190476 |
| Chr20:9.8Mb | 2379 | 20 | 9279551 | 10279603 | 2399 | 298 | AD | eGFR | -0.14753 | -0.91951 | 0.4715 | 0.021765 | 0 | 0.90948 | 0.591349 | 520 | 0.881388476190476 |
| Chr9:21.1Mb | 1397 | 9 | 20483881 | 21674032 | 2604 | 257 | AD | eGFR | 0.168775 | -0.56118 | 1 | 0.0284849 | 0 | 1 | 0.592437 | 521 | 0.881388476190476 |
| Chr20:0.5Mb | 2369 | 20 | 60795 | 884241 | 2545 | 358 | AD | eGFR | 0.119438 | -0.35954 | 0.63479 | 0.0142655 | 0 | 0.41851 | 0.592686 | 522 | 0.881388476190476 |
| Chr3:195.5Mb | 592 | 3 | 195196790 | 195710290 | 963 | 184 | AD | eGFR | -0.0938424 | -0.52321 | 0.28943 | 0.00880639 | 0 | 0.3011 | 0.595075 | 523 | 0.881388476190476 |
| Chr11:67.4Mb | 1675 | 11 | 66782662 | 68000949 | 2284 | 218 | AD | eGFR | -0.0968107 | -0.56201 | 0.30062 | 0.0093723 | 0 | 0.3308 | 0.596651 | 524 | 0.881388476190476 |
| Chr8:27Mb | 1272 | 8 | 26679136 | 27406511 | 2244 | 178 | AD | eGFR | 0.131948 | -0.41621 | 0.79187 | 0.0174103 | 0 | 0.70251 | 0.59677 | 525 | 0.881388476190476 |
| Chr3:29.3Mb | 449 | 3 | 28649811 | 29877022 | 3550 | 312 | AD | eGFR | 0.170362 | -0.57862 | 1 | 0.0290233 | 0 | 1 | 0.597133 | 526 | 0.881388476190476 |
| Chr10:123.3Mb | 1596 | 10 | 122803507 | 123856184 | 2491 | 295 | AD | eGFR | -0.150631 | -0.95922 | 0.51241 | 0.0226898 | 0 | 1 | 0.604064 | 527 | 0.881388476190476 |
| Chr8:26.3Mb | 1271 | 8 | 25859278 | 26679135 | 2705 | 241 | AD | eGFR | 0.114951 | -0.37376 | 0.63282 | 0.0132138 | 0 | 0.41593 | 0.604266 | 528 | 0.881388476190476 |
| Chr13:72.5Mb | 1912 | 13 | 71976863 | 73114509 | 2268 | 263 | AD | eGFR | 0.0946589 | -0.29729 | 0.54077 | 0.0089603 | 0 | 0.31764 | 0.604684 | 529 | 0.881388476190476 |
| Chr9:93Mb | 1437 | 9 | 92500278 | 93441050 | 2258 | 221 | AD | eGFR | 0.14336 | -0.48176 | 0.85991 | 0.020552 | 0 | 0.79855 | 0.605117 | 530 | 0.881388476190476 |
| Chr2:203.1Mb | 381 | 2 | 202504863 | 203639686 | 1911 | 224 | AD | eGFR | 0.159738 | -0.54455 | 1 | 0.0255161 | 0 | 1 | 0.6053 | 531 | 0.881388476190476 |
| Chr18:60.1Mb | 2288 | 18 | 59449423 | 60780194 | 3339 | 260 | AD | eGFR | -0.101542 | -0.52581 | 0.30404 | 0.0103109 | 0 | 0.28609 | 0.605505 | 532 | 0.881388476190476 |
| Chr20:36.6Mb | 2401 | 20 | 35990262 | 37253657 | 2393 | 280 | AD | eGFR | -0.130911 | -0.7916 | 0.42287 | 0.0171376 | 0 | 0.67048 | 0.606423 | 533 | 0.881388476190476 |
| Chr4:108.6Mb | 696 | 4 | 108131207 | 109122458 | 1941 | 209 | AD | eGFR | 0.243816 | -0.99012 | 1 | 0.0594461 | 0 | 1 | 0.607467 | 534 | 0.881388476190476 |
| Chr4:130.4Mb | 718 | 4 | 129625377 | 131098950 | 3246 | 240 | AD | eGFR | 0.200721 | -0.79477 | 1 | 0.0402889 | 0 | 1 | 0.607495 | 535 | 0.881388476190476 |
| Chr4:86.2Mb | 677 | 4 | 85437806 | 86931670 | 3003 | 201 | AD | eGFR | 0.149977 | -0.51028 | 0.96492 | 0.0224931 | 0 | 1 | 0.607768 | 536 | 0.881388476190476 |
| Chr20:14.7Mb | 2384 | 20 | 14203756 | 15238519 | 2399 | 218 | AD | eGFR | -0.15298 | -1 | 0.51715 | 0.0234029 | 0 | 1 | 0.607969 | 537 | 0.881388476190476 |
| Chr10:95Mb | 1573 | 10 | 94503300 | 95528160 | 2628 | 250 | AD | eGFR | 0.0840431 | -0.25813 | 0.44499 | 0.00706325 | 0 | 0.20786 | 0.609883 | 538 | 0.881388476190476 |

| locus_label | locus | chr | start | stop | n.snps | n.pcs | phen1 | phen2 | rho | rho.lower | rho.upper | r2 | r2.lower | r2.upper | p | rank | FDR |
|---|---|---|---|---|---|---|---|---|---|---|---|---|---|---|---|---|---|
| Chr11:53.5Mb | 1664 | 11 | 51581596 | 55477633 | 2043 | 24 | AD | eGFR | -0.150041 | -1 | 0.53122 | 0.0225122 | 0 | 1 | 0.609983 | 539 | 0.881388476190476 |
| Chr1:208.9Mb | 160 | 1 | 208162952 | 209711169 | 3327 | 346 | AD | eGFR | -0.123131 | -0.73227 | 0.39079 | 0.0151613 | 0 | 0.57187 | 0.610726 | 540 | 0.881388476190476 |
| Chr5:154.2Mb | 903 | 5 | 153734495 | 154599750 | 1966 | 238 | AD | eGFR | 0.157167 | -0.56542 | 1 | 0.0247016 | 0 | 1 | 0.612051 | 541 | 0.881388476190476 |
| Chr2:120.4Mb | 312 | 2 | 119635490 | 121073752 | 3027 | 268 | AD | eGFR | 0.0986789 | -0.313 | 0.54737 | 0.00973753 | 0 | 0.3198 | 0.613487 | 542 | 0.881388476190476 |
| Chr6:56.8Mb | 991 | 6 | 56200986 | 57316157 | 1723 | 172 | AD | eGFR | -0.15494 | -1 | 0.54076 | 0.0240063 | 0 | 1 | 0.613517 | 543 | 0.881388476190476 |
| Chr12:112.8Mb | 1841 | 12 | 111592382 | 113947983 | 3104 | 257 | AD | eGFR | 0.12041 | -0.41458 | 0.7259 | 0.0144985 | 0 | 0.55856 | 0.614283 | 544 | 0.881388476190476 |
| Chr2:55.2Mb | 262 | 2 | 54688235 | 55663336 | 2402 | 216 | AD | eGFR | -0.107521 | -0.64936 | 0.36699 | 0.0115608 | 0 | 0.4906 | 0.614468 | 545 | 0.881388476190476 |
| Chr6:161.8Mb | 1085 | 6 | 161371015 | 162254229 | 2134 | 299 | AD | eGFR | -0.086259 | -0.4665 | 0.27166 | 0.00744061 | 0 | 0.22415 | 0.615394 | 546 | 0.881388476190476 |
| Chr1:175.9Mb | 132 | 1 | 175385221 | 176449096 | 1961 | 213 | AD | eGFR | -0.156134 | -1 | 0.57811 | 0.0243778 | 0 | 1 | 0.620943 | 547 | 0.886788 |
| Chr9:137.9Mb | 1481 | 9 | 137593528 | 138202112 | 2230 | 328 | AD | eGFR | 0.13111 | -0.47799 | 0.82949 | 0.0171898 | 0 | 0.76278 | 0.621726 | 548 | 0.886788 |
| Chr19:57.6Mb | 2367 | 19 | 57143510 | 57959952 | 2188 | 293 | AD | eGFR | -0.0926855 | -0.50731 | 0.30304 | 0.0085906 | 0 | 0.2696 | 0.622566 | 549 | 0.886788 |
| Chr1:62.1Mb | 53 | 1 | 61377616 | 62778733 | 3247 | 335 | AD | eGFR | -0.107476 | -0.64643 | 0.36096 | 0.0115511 | 0 | 0.45988 | 0.624639 | 550 | 0.887927723404255 |
| Chr18:59Mb | 2287 | 18 | 58527237 | 59449422 | 1832 | 268 | AD | eGFR | -0.0848249 | -0.49007 | 0.28289 | 0.00719526 | 0 | 0.25175 | 0.627049 | 551 | 0.887927723404255 |
| Chr2:63.1Mb | 269 | 2 | 62429911 | 63672118 | 2032 | 230 | AD | eGFR | 0.191204 | -0.80389 | 1 | 0.036559 | 0 | 1 | 0.627063 | 552 | 0.887927723404255 |
| Chr17:10.1Mb | 2183 | 17 | 9619357 | 10572617 | 2707 | 318 | AD | eGFR | 0.109028 | -0.37184 | 0.63017 | 0.0118871 | 0 | 0.43403 | 0.629049 | 553 | 0.887927723404255 |
| Chr8:82.1Mb | 1315 | 8 | 81340232 | 82878769 | 3018 | 273 | AD | eGFR | -0.119126 | -0.77671 | 0.43887 | 0.0141911 | 0 | 0.66157 | 0.631928 | 554 | 0.887927723404255 |
| Chr3:9.3Mb | 431 | 3 | 8664893 | 9970731 | 3169 | 363 | AD | eGFR | 0.191046 | -0.87709 | 1 | 0.0364984 | 0 | 1 | 0.632057 | 555 | 0.887927723404255 |
| Chr2:194.2Mb | 375 | 2 | 193033983 | 195300250 | 2450 | 227 | AD | eGFR | -0.161641 | -1 | 0.63472 | 0.0261278 | 0 | 1 | 0.632115 | 556 | 0.887927723404255 |
| Chr6:130.2Mb | 1057 | 6 | 129850179 | 130550137 | 1764 | 243 | AD | eGFR | 0.102753 | -0.38324 | 0.67415 | 0.0105581 | 0 | 0.50923 | 0.633929 | 557 | 0.887927723404255 |
| Chr7:156.8Mb | 1229 | 7 | 156344387 | 157199331 | 2674 | 239 | AD | eGFR | -0.0720958 | -0.40004 | 0.24081 | 0.0051978 | 0 | 0.16827 | 0.634054 | 558 | 0.887927723404255 |
| Chr20:52.8Mb | 2416 | 20 | 52411533 | 53120334 | 2024 | 249 | AD | eGFR | 0.0850805 | -0.33164 | 0.56424 | 0.00723869 | 0 | 0.37336 | 0.635177 | 559 | 0.887927723404255 |
| Chr5:141.9Mb | 893 | 5 | 141253831 | 142532956 | 2905 | 315 | AD | eGFR | 0.109965 | -0.41129 | 0.7276 | 0.0120922 | 0 | 0.58691 | 0.636919 | 560 | 0.887927723404255 |
| Chr1:40.6Mb | 37 | 1 | 40200951 | 41093875 | 1855 | 211 | AD | eGFR | 0.0955822 | -0.32841 | 0.56607 | 0.00913595 | 0 | 0.33363 | 0.637018 | 561 | 0.887927723404255 |
| Chr14:23.4M | 195 | 14 | 22760701 | 23985936 | 2909 | 342 | AD | eGFR | - | -0.44835 | 0.27309 | 0.00609906 | 0 | 0.20983 | 0.638347 | 562 | 0.88792772340425 |

| locus_label | locus | chr | start | stop | n.snps | n.pcs | phen1 | phen2 | rho | rho.lower | rho.upper | r2 | r2.lower | r2.upper | p | rank | FDR |
|---|---|---|---|---|---|---|---|---|---|---|---|---|---|---|---|---|---|
| b | 7 | | | | | | | | 0.0780965 | | | | | | | | 5 |
| Chr4:80.5Mb | 673 | 4 | 79880102 | 81206182 | 2769 | 226 | AD | eGFR | 0.0676332 | -0.22859 | 0.37109 | 0.00457425 | 0 | 0.14218 | 0.639635 | 563 | 0.887927723404255 |
| Chr1:115.4Mb | 101 | 1 | 114664388 | 116207297 | 3684 | 286 | AD | eGFR | 0.0823424 | -0.28226 | 0.4702 | 0.00678028 | 0 | 0.22898 | 0.640398 | 564 | 0.887927723404255 |
| Chr6:7.2Mb | 932 | 6 | 6621938 | 7808323 | 3403 | 345 | AD | eGFR | -0.0829978 | -0.56948 | 0.30487 | 0.00688864 | 0 | 0.35146 | 0.642939 | 565 | 0.88987309380531 |
| Chr10:134.3Mb | 1608 | 10 | 133818037 | 134856054 | 3326 | 312 | AD | eGFR | -0.16086 | -1 | 0.6788 | 0.0258759 | 0 | 1 | 0.646614 | 566 | 0.892527959390863 |
| Chr1:48.6Mb | 43 | 1 | 48105234 | 49185414 | 2007 | 240 | AD | eGFR | -0.142598 | -1 | 0.58438 | 0.0203342 | 0 | 1 | 0.648685 | 567 | 0.892527959390863 |
| Chr3:32.7Mb | 452 | 3 | 32378806 | 33020868 | 1786 | 201 | AD | eGFR | -0.109433 | -0.8209 | 0.45819 | 0.0119756 | 0 | 0.77655 | 0.652609 | 568 | 0.892527959390863 |
| Chr2:215.5Mb | 392 | 2 | 215002485 | 215899570 | 2089 | 208 | AD | eGFR | -0.178281 | -1 | 0.80461 | 0.0317841 | 0 | 1 | 0.653002 | 569 | 0.892527959390863 |
| Chr6:91.6Mb | 1023 | 6 | 91097025 | 92015279 | 2130 | 233 | AD | eGFR | 0.123002 | -0.51614 | 0.8558 | 0.0151294 | 0 | 0.84267 | 0.656214 | 570 | 0.892527959390863 |
| Chr9:81.9Mb | 1427 | 9 | 81161598 | 82601465 | 2722 | 330 | AD | eGFR | 0.11912 | -0.49099 | 0.81314 | 0.0141896 | 0 | 0.72931 | 0.656902 | 571 | 0.892527959390863 |
| Chr1:160.5Mb | 117 | 1 | 159882516 | 161054076 | 2492 | 294 | AD | eGFR | 0.116118 | -0.44282 | 0.74575 | 0.0134835 | 0 | 0.61379 | 0.657479 | 572 | 0.892527959390863 |
| Chr13:48.8Mb | 1891 | 13 | 48137480 | 49381727 | 2642 | 188 | AD | eGFR | -0.113392 | -0.82635 | 0.45801 | 0.0128577 | 0 | 0.73426 | 0.658057 | 573 | 0.892527959390863 |
| Chr2:10.3Mb | 214 | 2 | 9960671 | 10611654 | 2063 | 261 | AD | eGFR | 0.106674 | -0.42819 | 0.69914 | 0.0113793 | 0 | 0.52364 | 0.659629 | 574 | 0.892527959390863 |
| Chr1:67.3Mb | 57 | 1 | 66778016 | 67761890 | 2274 | 200 | AD | eGFR | -0.123568 | -0.86274 | 0.52009 | 0.015269 | 0 | 0.87367 | 0.660094 | 575 | 0.892527959390863 |
| Chr12:13.1Mb | 1753 | 12 | 12721875 | 13559527 | 1754 | 283 | AD | eGFR | 0.0916169 | -0.36304 | 0.57925 | 0.00839365 | 0 | 0.38137 | 0.660564 | 576 | 0.892527959390863 |
| Chr3:16.6Mb | 438 | 3 | 16084006 | 17182494 | 2992 | 281 | AD | eGFR | -0.119373 | -0.79264 | 0.4815 | 0.0142499 | 0 | 0.69959 | 0.661221 | 577 | 0.892527959390863 |
| Chr7:155.8Mb | 1228 | 7 | 155280611 | 156344386 | 3205 | 357 | AD | eGFR | 0.0715299 | -0.28339 | 0.51262 | 0.00511652 | 0 | 0.29653 | 0.661638 | 578 | 0.892527959390863 |
| Chr11:27.8Mb | 1640 | 11 | 27018308 | 28577718 | 2360 | 217 | AD | eGFR | -0.0899872 | -0.57125 | 0.34956 | 0.0080977 | 0 | 0.35258 | 0.662252 | 579 | 0.892527959390863 |
| Chr13:23.3Mb | 1867 | 13 | 22795189 | 23729049 | 3270 | 323 | AD | eGFR | -0.146694 | -1 | 0.69829 | 0.021519 | 0 | 1 | 0.665416 | 580 | 0.892527959390863 |
| Chr6:100.3Mb | 1030 | 6 | 99678877 | 100878645 | 2721 | 238 | AD | eGFR | 0.0747422 | -0.28231 | 0.44218 | 0.00558639 | 0 | 0.20255 | 0.665456 | 581 | 0.892527959390863 |
| Chr9:78.2Mb | 1423 | 9 | 77862309 | 78630915 | 2203 | 243 | AD | eGFR | 0.150182 | -0.7238 | 1 | 0.0225546 | 0 | 1 | 0.666601 | 582 | 0.892527959390863 |
| Chr7:135.5Mb | 1212 | 7 | 134943222 | 136113467 | 2805 | 273 | AD | eGFR | -0.109533 | -0.72992 | 0.43659 | 0.0119975 | 0 | 0.57306 | 0.667503 | 583 | 0.892527959390863 |
| Chr3:193.7Mb | 590 | 3 | 193122091 | 194217140 | 2691 | 332 | AD | eGFR | -0.0838423 | -0.57406 | 0.33211 | 0.00702954 | 0 | 0.36386 | 0.667511 | 584 | 0.892527959390863 |
| Chr3:14.7Mb | 436 | 3 | 14312008 | 15150876 | 2116 | 279 | AD | eGFR | -0.0783949 | -0.47224 | 0.29976 | 0.00614576 | 0 | 0.23273 | 0.670022 | 585 | 0.892527959390863 |

| locus_label | locus | chr | start | stop | n.snps | n.pcs | phen1 | phen2 | rho | rho.lower | rho.upper | r2 | r2.lower | r2.upper | p | rank | FDR |
|---|---|---|---|---|---|---|---|---|---|---|---|---|---|---|---|---|---|
| Chr4:100.7Mb | 690 | 4 | 100333335 | 101132995 | 1879 | 161 | AD | eGFR | 0.119874 | -0.51408 | 0.83745 | 0.0143699 | 0 | 0.78579 | 0.670373 | 586 | 0.892527959390863 |
| Chr8:24.3Mb | 1269 | 8 | 23788495 | 24863243 | 2208 | 211 | AD | eGFR | -0.0691735 | -0.43597 | 0.26704 | 0.00478497 | 0 | 0.20238 | 0.6712 | 587 | 0.892527959390863 |
| Chr2:3Mb | 205 | 2 | 2680064 | 3314202 | 1930 | 263 | AD | eGFR | -0.113764 | -0.79094 | 0.47724 | 0.0129421 | 0 | 0.66593 | 0.672686 | 588 | 0.892527959390863 |
| Chr19:39.8Mb | 2345 | 19 | 39370420 | 40171413 | 1976 | 228 | AD | eGFR | 0.131461 | -0.60044 | 0.99541 | 0.0172821 | 0 | 1 | 0.673096 | 589 | 0.892527959390863 |
| Chr17:73.1Mb | 2229 | 17 | 72383473 | 73741321 | 2933 | 300 | AD | eGFR | 0.0822681 | -0.35291 | 0.52849 | 0.00676805 | 0 | 0.30454 | 0.674463 | 590 | 0.892527959390863 |
| Chr9:20Mb | 1396 | 9 | 19463293 | 20483880 | 2288 | 297 | AD | eGFR | -0.0960869 | -0.62429 | 0.39598 | 0.00923269 | 0 | 0.41647 | 0.674532 | 591 | 0.892527959390863 |
| Chr5:172.9Mb | 919 | 5 | 172285683 | 173606995 | 3382 | 328 | AD | eGFR | 0.07835 | -0.30919 | 0.49184 | 0.00613872 | 0 | 0.25054 | 0.6778 | 592 | 0.895337162162162 |
| Chr1:3.9Mb | 5 | 1 | 3582048 | 4281476 | 2037 | 239 | AD | eGFR | 0.0691341 | -0.28732 | 0.42743 | 0.00477952 | 0 | 0.19826 | 0.682599 | 593 | 0.900155848229342 |
| Chr14:53.6Mb | 1985 | 14 | 52760554 | 54359598 | 2743 | 291 | AD | eGFR | 0.102685 | -0.45693 | 0.74696 | 0.0105441 | 0 | 0.6273 | 0.687586 | 594 | 0.903557125412541 |
| Chr2:25.3Mb | 228 | 2 | 24669225 | 25895394 | 2010 | 217 | AD | eGFR | -0.0751993 | -0.49302 | 0.32884 | 0.00565493 | 0 | 0.26442 | 0.688357 | 595 | 0.903557125412541 |
| Chr2:64.2Mb | 270 | 2 | 63672119 | 64696089 | 1782 | 193 | AD | eGFR | 0.114672 | -0.56045 | 0.85161 | 0.0131496 | 0 | 0.79817 | 0.690252 | 596 | 0.903557125412541 |
| Chr3:138Mb | 541 | 3 | 137372142 | 138693846 | 2030 | 217 | AD | eGFR | -0.0946779 | -0.65457 | 0.41316 | 0.0089639 | 0 | 0.45806 | 0.690387 | 597 | 0.903557125412541 |
| Chr11:50.7Mb | 1662 | 11 | 50217009 | 51098366 | 1536 | 12 | AD | eGFR | -0.115357 | -0.79075 | 0.52008 | 0.0133071 | 0 | 0.68596 | 0.692256 | 598 | 0.903557125412541 |
| Chr8:8.9Mb | 1247 | 8 | 8589771 | 9167795 | 1883 | 241 | AD | eGFR | -0.0782226 | -0.57063 | 0.34914 | 0.00611878 | 0 | 0.35953 | 0.6926 | 599 | 0.903557125412541 |
| Chr2:9.5Mb | 213 | 2 | 8978799 | 9960670 | 1950 | 242 | AD | eGFR | -0.0825957 | -0.55595 | 0.3647 | 0.00682205 | 0 | 0.33308 | 0.694656 | 600 | 0.903557125412541 |
| Chr7:136.9Mb | 1213 | 7 | 136113468 | 137657459 | 2850 | 278 | AD | eGFR | 0.126437 | -0.65686 | 1 | 0.0159864 | 0 | 1 | 0.696446 | 601 | 0.903557125412541 |
| Chr19:36.1Mb | 2342 | 19 | 35649344 | 36638122 | 2386 | 297 | AD | eGFR | -0.0594291 | -0.3857 | 0.25835 | 0.00353182 | 0 | 0.16061 | 0.696998 | 602 | 0.903557125412541 |
| Chr13:107.8Mb | 1945 | 13 | 107037865 | 108521978 | 3607 | 366 | AD | eGFR | 0.0934195 | -0.43054 | 0.64387 | 0.00872719 | 0 | 0.44876 | 0.698416 | 603 | 0.903557125412541 |
| Chr18:3.3Mb | 2241 | 18 | 2839843 | 3722828 | 2498 | 340 | AD | eGFR | -0.0858589 | -0.59293 | 0.39241 | 0.00737175 | 0 | 0.38774 | 0.698975 | 604 | 0.903557125412541 |
| Chr5:175.4Mb | 921 | 5 | 174662886 | 176180301 | 2772 | 329 | AD | eGFR | -0.0555733 | -0.35453 | 0.23836 | 0.0030884 | 0 | 0.13212 | 0.699223 | 605 | 0.903557125412541 |
| Chr1:39.3Mb | 36 | 1 | 38474037 | 40200950 | 3625 | 322 | AD | eGFR | -0.0941849 | -0.66937 | 0.41742 | 0.0088708 | 0 | 0.48597 | 0.700199 | 606 | 0.903557125412541 |
| Chr1:11.2Mb | 13 | 1 | 10753428 | 11709173 | 2044 | 266 | AD | eGFR | 0.0674155 | -0.31402 | 0.45773 | 0.00454485 | 0 | 0.22946 | 0.702623 | 607 | 0.90453435483871 |
| Chr5:81.1Mb | 842 | 5 | 80448226 | 81710194 | 2024 | 250 | AD | eGFR | -0.104465 | -0.83565 | 0.52627 | 0.010913 | 0 | 0.81391 | 0.703587 | 608 | 0.90453435483871 |
| Chr9:117.5Mb | 1462 | 9 | 117054910 | 117989017 | 2045 | 245 | AD | eGFR | 0.130101 | -0.72945 | 1 | 0.0169264 | 0 | 1 | 0.707334 | 609 | 0.90453435483871 |
| Chr5:176.9M | 922 | 5 | 17618030 | 17765990 | 2592 | 265 | AD | eGFR | - | -0.30985 | 0.20802 | 0.00206877 | 0 | 0.10378 | 0.707778 | 610 | 0.90453435483871 |

| locus_label | locus | chr | start | stop | n.snps | n.pcs | phen1 | phen2 | rho | rho.lower | rho.upper | r2 | r2.lower | r2.upper | p | rank | FDR |
|---|---|---|---|---|---|---|---|---|---|---|---|---|---|---|---|---|---|
| b | | | 2 | 2 | | | | | 0.0454838 | | | | | | | | |
| Chr7:80.8Mb | 1171 | 7 | 80055722 | 81533566 | 3030 | 311 | AD | eGFR | -0.107299 | -0.8541 | 0.54564 | 0.011513 | 0 | 0.80114 | 0.707789 | 611 | 0.90453435483871 |
| Chr2:231.2Mb | 407 | 2 | 230541093 | 231847386 | 3295 | 296 | AD | eGFR | -0.0844426 | -0.63145 | 0.40987 | 0.00713055 | 0 | 0.43669 | 0.710302 | 612 | 0.90453435483871 |
| Chr7:154.8Mb | 1227 | 7 | 154407488 | 155280610 | 2876 | 334 | AD | eGFR | 0.118604 | -0.65263 | 1 | 0.0140669 | 0 | 1 | 0.710717 | 613 | 0.90453435483871 |
| Chr16:59.2Mb | 2139 | 16 | 58508979 | 59885499 | 3593 | 264 | AD | eGFR | 0.155379 | -1 | 1 | 0.0241425 | 0 | 1 | 0.711113 | 614 | 0.90453435483871 |
| Chr9:2.8Mb | 1374 | 9 | 2441121 | 3079045 | 1747 | 277 | AD | eGFR | 0.0917736 | -0.45294 | 0.68714 | 0.00842239 | 0 | 0.50913 | 0.712576 | 615 | 0.90453435483871 |
| Chr9:111.7Mb | 1455 | 9 | 111368108 | 112076381 | 1953 | 193 | AD | eGFR | -0.111118 | -0.95317 | 0.60948 | 0.0123472 | 0 | 1 | 0.71501 | 616 | 0.90453435483871 |
| Chr11:5.2Mb | 1616 | 11 | 4946052 | 5533057 | 2422 | 191 | AD | eGFR | -0.0916413 | -0.82226 | 0.50704 | 0.00839813 | 0 | 0.79835 | 0.715158 | 617 | 0.90453435483871 |
| Chr19:54.3Mb | 2362 | 19 | 54042241 | 54602370 | 1618 | 292 | AD | eGFR | 0.0417071 | -0.1913 | 0.27454 | 0.00173948 | 0 | 0.08036 | 0.715707 | 618 | 0.90453435483871 |
| Chr17:0.4Mb | 2173 | 17 | 52 | 815653 | 2305 | 302 | AD | eGFR | -0.0602848 | -0.42624 | 0.28728 | 0.00363425 | 0 | 0.19755 | 0.716668 | 619 | 0.90453435483871 |
| Chr17:3.1Mb | 2176 | 17 | 2699136 | 3430440 | 2085 | 268 | AD | eGFR | 0.0673333 | -0.32459 | 0.49969 | 0.00453377 | 0 | 0.26339 | 0.71715 | 620 | 0.90453435483871 |
| Chr8:75.7Mb | 1310 | 8 | 75011700 | 76295483 | 2919 | 249 | AD | eGFR | 0.0963654 | -0.53754 | 0.80564 | 0.00928629 | 0 | 0.80896 | 0.719615 | 621 | 0.906181851851852 |
| Chr21:41.8Mb | 2455 | 21 | 41344743 | 42187152 | 2506 | 282 | AD | eGFR | -0.0945527 | -0.85428 | 0.51272 | 0.00894021 | 0 | 0.81417 | 0.724387 | 622 | 0.908112862865948 |
| Chr3:149.3Mb | 551 | 3 | 148638309 | 149998411 | 3303 | 305 | AD | eGFR | 0.083074 | -0.43989 | 0.63198 | 0.0069013 | 0 | 0.44782 | 0.726602 | 623 | 0.908112862865948 |
| Chr12:131.7Mb | 1860 | 12 | 131404043 | 132012316 | 1828 | 232 | AD | eGFR | 0.0703561 | -0.38465 | 0.58084 | 0.00494998 | 0 | 0.37346 | 0.729094 | 624 | 0.908112862865948 |
| Chr13:75.2Mb | 1914 | 13 | 74437636 | 75882384 | 3549 | 304 | AD | eGFR | -0.0818894 | -0.71047 | 0.47796 | 0.00670588 | 0 | 0.61063 | 0.730301 | 625 | 0.908112862865948 |
| Chr12:129.5Mb | 1857 | 12 | 129170297 | 129887418 | 2436 | 321 | AD | eGFR | -0.118051 | -1 | 0.71914 | 0.0139361 | 0 | 1 | 0.733565 | 626 | 0.908112862865948 |
| Chr7:8.3Mb | 1104 | 7 | 7926501 | 8718351 | 2374 | 284 | AD | eGFR | -0.113652 | -1 | 0.72017 | 0.0129168 | 0 | 1 | 0.733683 | 627 | 0.908112862865948 |
| Chr13:28Mb | 1872 | 13 | 27392006 | 28587125 | 3497 | 311 | AD | eGFR | 0.0840683 | -0.48838 | 0.72869 | 0.00706749 | 0 | 0.61914 | 0.73378 | 628 | 0.908112862865948 |
| Chr7:28.1Mb | 1126 | 7 | 27351287 | 28890886 | 3569 | 328 | AD | eGFR | -0.138203 | -1 | 0.96329 | 0.0191 | 0 | 1 | 0.734609 | 629 | 0.908112862865948 |
| Chr2:168.8Mb | 352 | 2 | 168437446 | 169224445 | 1905 | 229 | AD | eGFR | 0.0984057 | -0.58991 | 0.85053 | 0.00968367 | 0 | 0.841 | 0.736016 | 630 | 0.908112862865948 |
| Chr6:162.8Mb | 1086 | 6 | 162254230 | 163336361 | 2930 | 252 | AD | eGFR | 0.120638 | -0.78078 | 1 | 0.0145536 | 0 | 1 | 0.73619 | 631 | 0.908112862865948 |
| Chr19:5.1Mb | 2312 | 19 | 4741719 | 5478684 | 2004 | 304 | AD | eGFR | 0.0678983 | -0.35272 | 0.51291 | 0.00461018 | 0 | 0.28143 | 0.736417 | 632 | 0.908112862865948 |
| Chr6:132.2Mb | 1059 | 6 | 131457165 | 132890694 | 2752 | 287 | AD | eGFR | 0.0521683 | -0.27686 | 0.39128 | 0.00272153 | 0 | 0.17095 | 0.736439 | 633 | 0.908112862865948 |

| locus_label | locus | chr | start | stop | n.snps | n.pcs | phen1 | phen2 | rho | rho.lower | rho.upper | r2 | r2.lower | r2.upper | p | rank | FDR |
|---|---|---|---|---|---|---|---|---|---|---|---|---|---|---|---|---|---|
| Chr17:75.3Mb | 2231 | 17 | 74908267 | 75619256 | 2324 | 321 | AD | eGFR | -0.0737472 | -0.5925 | 0.40643 | 0.00543865 | 0 | 0.39911 | 0.738075 | 634 | 0.908112862865948 |
| Chr22:38Mb | 2482 | 22 | 37364005 | 38718589 | 3042 | 329 | AD | eGFR | 0.074699 | -0.43518 | 0.63624 | 0.00557994 | 0 | 0.49505 | 0.738709 | 635 | 0.908112862865948 |
| Chr5:179.2Mb | 924 | 5 | 178595253 | 179794710 | 3233 | 353 | AD | eGFR | -0.0695304 | -0.55876 | 0.37155 | 0.00483448 | 0 | 0.33625 | 0.740888 | 636 | 0.908112862865948 |
| Chr10:81.5Mb | 1560 | 10 | 81190574 | 81763230 | 1657 | 89 | AD | eGFR | -0.0717973 | -0.54447 | 0.39299 | 0.00515486 | 0 | 0.32403 | 0.740947 | 637 | 0.908112862865948 |
| Chr6:106.7Mb | 1037 | 6 | 106053916 | 107309327 | 3361 | 306 | AD | eGFR | 0.0985508 | -0.63095 | 0.92714 | 0.00971226 | 0 | 1 | 0.74129 | 638 | 0.908112862865948 |
| Chr6:166.3Mb | 1089 | 6 | 165835954 | 166853325 | 3055 | 308 | AD | eGFR | -0.0710592 | -0.58717 | 0.40368 | 0.00504941 | 0 | 0.37338 | 0.745101 | 639 | 0.908112862865948 |
| Chr12:133.4Mb | 1862 | 12 | 132957946 | 133841814 | 2081 | 204 | AD | eGFR | 0.063159 | -0.37164 | 0.55228 | 0.00398906 | 0 | 0.33868 | 0.747857 | 640 | 0.908112862865948 |
| Chr5:94.5Mb | 853 | 5 | 93814605 | 95117259 | 2393 | 262 | AD | eGFR | 0.109639 | -0.73373 | 1 | 0.0120208 | 0 | 1 | 0.748433 | 641 | 0.908112862865948 |
| Chr7:129.6Mb | 1208 | 7 | 128779174 | 130418704 | 3352 | 295 | AD | eGFR | 0.0695521 | -0.4446 | 0.63098 | 0.00483749 | 0 | 0.50955 | 0.749241 | 642 | 0.908112862865948 |
| Chr1:65.1Mb | 55 | 1 | 64344227 | 65894184 | 3102 | 290 | AD | eGFR | 0.0846889 | -0.51223 | 0.72869 | 0.00717221 | 0 | 0.63233 | 0.750409 | 643 | 0.908112862865948 |
| Chr3:70.5Mb | 482 | 3 | 69784970 | 71223281 | 2368 | 279 | AD | eGFR | -0.0812876 | -0.79561 | 0.52169 | 0.00660767 | 0 | 0.75917 | 0.752018 | 644 | 0.908112862865948 |
| Chr17:48.1Mb | 2210 | 17 | 47516225 | 48722590 | 2168 | 350 | AD | eGFR | 0.0604774 | -0.34738 | 0.48987 | 0.00365751 | 0 | 0.2737 | 0.752388 | 645 | 0.908112862865948 |
| Chr19:32.4Mb | 2338 | 19 | 31763009 | 33028099 | 2498 | 291 | AD | eGFR | -0.0537994 | -0.41255 | 0.29789 | 0.00289437 | 0 | 0.18741 | 0.752596 | 646 | 0.908112862865948 |
| Chr11:116.8Mb | 1722 | 11 | 116247378 | 117295641 | 2851 | 264 | AD | eGFR | 0.0560855 | -0.3091 | 0.43534 | 0.00314558 | 0 | 0.20202 | 0.752978 | 647 | 0.908112862865948 |
| Chr10:82.1Mb | 1561 | 10 | 81763231 | 82378048 | 1970 | 165 | AD | eGFR | -0.0567258 | -0.442 | 0.31599 | 0.00321781 | 0 | 0.21317 | 0.753433 | 648 | 0.908112862865948 |
| Chr15:52.4Mb | 2058 | 15 | 51605478 | 53263126 | 3303 | 262 | AD | eGFR | -0.0674862 | -0.5553 | 0.38496 | 0.00455439 | 0 | 0.34969 | 0.753664 | 649 | 0.908112862865948 |
| Chr9:32.7Mb | 1410 | 9 | 32253284 | 33194892 | 2320 | 229 | AD | eGFR | 0.0683749 | -0.44442 | 0.63973 | 0.00467512 | 0 | 0.51601 | 0.756107 | 650 | 0.909654883076923 |
| Chr3:71.8Mb | 483 | 3 | 71223282 | 72334704 | 2571 | 348 | AD | eGFR | 0.104509 | -0.73104 | 1 | 0.0109222 | 0 | 1 | 0.757381 | 651 | 0.909787929339478 |
| Chr1:111.8Mb | 98 | 1 | 111134063 | 112489248 | 3436 | 298 | AD | eGFR | 0.106127 | -0.7865 | 1 | 0.0112629 | 0 | 1 | 0.75878 | 652 | 0.910070490797546 |
| Chr12:6.5Mb | 1746 | 12 | 6023701 | 6948954 | 2210 | 295 | AD | eGFR | 0.0513252 | -0.29893 | 0.39673 | 0.00263427 | 0 | 0.17312 | 0.760552 | 653 | 0.910798872894334 |
| Chr11:29.1Mb | 1641 | 11 | 28577719 | 29530460 | 1668 | 176 | AD | eGFR | -0.0782034 | -0.82224 | 0.52301 | 0.00611577 | 0 | 0.83056 | 0.764068 | 654 | 0.912616683969466 |
| Chr12:116.6Mb | 1845 | 12 | 116197676 | 117091843 | 2136 | 273 | AD | eGFR | 0.059297 | -0.35357 | 0.49726 | 0.00351613 | 0 | 0.26894 | 0.764404 | 655 | 0.912616683969466 |
| Chr8:124.7Mb | 1350 | 8 | 124001020 | 125453322 | 3134 | 305 | AD | eGFR | 0.068803 | -0.44188 | 0.61091 | 0.00473386 | 0 | 0.43267 | 0.767137 | 656 | 0.91448343597561 |
| Chr5:3.3Mb | 779 | 5 | 2960069 | 3677595 | 2916 | 290 | AD | eGFR | -0.101032 | -1 | 0.78701 | 0.0102074 | 0 | 1 | 0.769022 | 657 | 0.91533516590563 |

| locus_label | locus | chr | start | stop | n.snps | n.pcs | phen1 | phen2 | rho | rho.lower | rho.upper | r2 | r2.lower | r2.upper | p | rank | FDR |
|---|---|---|---|---|---|---|---|---|---|---|---|---|---|---|---|---|---|
| | | | | | | | | | | | | | | | | | 2 |
| Chr20:56.6Mb | 2421 | 20 | 56172844 | 56951773 | 2063 | 296 | AD | eGFR | -0.0812986 | -0.80746 | 0.55376 | 0.00660947 | 0 | 0.72383 | 0.771274 | 658 | 0.916502818731118 |
| Chr10:74.6Mb | 1555 | 10 | 73514215 | 75694023 | 2971 | 200 | AD | eGFR | -0.0796727 | -0.79018 | 0.55702 | 0.00634773 | 0 | 0.7509 | 0.774337 | 659 | 0.916502818731118 |
| Chr10:4.3Mb | 1490 | 10 | 3880323 | 4813182 | 2551 | 302 | AD | eGFR | 0.0852112 | -0.62555 | 0.88344 | 0.00726095 | 0 | 0.9859 | 0.774806 | 660 | 0.916502818731118 |
| Chr18:7.3Mb | 2245 | 18 | 6756497 | 7862479 | 2615 | 329 | AD | eGFR | 0.0647253 | -0.44301 | 0.62139 | 0.00418936 | 0 | 0.44741 | 0.775455 | 661 | 0.916502818731118 |
| Chr12:7.5Mb | 1747 | 12 | 6948955 | 8123317 | 2270 | 257 | AD | eGFR | 0.104187 | -0.86592 | 1 | 0.0108549 | 0 | 1 | 0.775863 | 662 | 0.916502818731118 |
| Chr16:28.2Mb | 2127 | 16 | 27443062 | 29043177 | 1982 | 276 | AD | eGFR | -0.0559066 | -0.54814 | 0.39252 | 0.00312554 | 0 | 0.35066 | 0.777236 | 663 | 0.916739897435898 |
| Chr4:174.9Mb | 756 | 4 | 174470207 | 175295842 | 1784 | 261 | AD | eGFR | -0.103076 | -1 | 0.80349 | 0.0106246 | 0 | 1 | 0.779175 | 664 | 0.917642846385542 |
| Chr11:56.3Mb | 1666 | 11 | 55947453 | 56623494 | 1933 | 59 | AD | eGFR | -0.0874331 | -0.92032 | 0.65423 | 0.00764454 | 0 | 1 | 0.782458 | 665 | 0.920116933933934 |
| Chr11:48.4Mb | 1659 | 11 | 48066367 | 48765366 | 1569 | 41 | AD | eGFR | -0.062115 | -0.54045 | 0.42476 | 0.00385828 | 0 | 0.33213 | 0.783629 | 666 | 0.920116933933934 |
| Chr13:32.2Mb | 1875 | 13 | 31433219 | 32986686 | 3586 | 324 | AD | eGFR | -0.0773952 | -0.91319 | 0.6017 | 0.00599002 | 0 | 1 | 0.787511 | 667 | 0.922553103139014 |
| Chr19:46.3Mb | 2352 | 19 | 45893308 | 46765060 | 2105 | 246 | AD | eGFR | 0.0391366 | -0.25258 | 0.34103 | 0.00153167 | 0 | 0.12803 | 0.788992 | 668 | 0.922553103139014 |
| Chr6:148.7Mb | 1071 | 6 | 148184845 | 149278798 | 3253 | 317 | AD | eGFR | -0.0801162 | -0.89973 | 0.60678 | 0.0064186 | 0 | 0.98776 | 0.789243 | 669 | 0.922553103139014 |
| Chr3:66Mb | 478 | 3 | 65326752 | 66702084 | 2916 | 322 | AD | eGFR | 0.0679175 | -0.5353 | 0.70054 | 0.00461279 | 0 | 0.60056 | 0.796608 | 670 | 0.929134696428571 |
| Chr1:44.3Mb | 40 | 1 | 43512670 | 45167235 | 3067 | 284 | AD | eGFR | 0.0652679 | -0.55555 | 0.75435 | 0.0042599 | 0 | 0.79041 | 0.797292 | 671 | 0.929134696428571 |
| Chr18:57.2Mb | 2285 | 18 | 56832313 | 57635821 | 2345 | 282 | AD | eGFR | -0.11433 | -1 | 1 | 0.0130713 | 0 | 1 | 0.798438 | 672 | 0.929134696428571 |
| Chr11:76Mb | 1682 | 11 | 75445254 | 76518906 | 2210 | 260 | AD | eGFR | -0.0591588 | -0.60931 | 0.45414 | 0.00349976 | 0 | 0.42962 | 0.806674 | 673 | 0.937307056379822 |
| Chr6:134.9Mb | 1061 | 6 | 134313644 | 135544083 | 2598 | 305 | AD | eGFR | -0.0554577 | -0.57989 | 0.4329 | 0.00307555 | 0 | 0.37863 | 0.807858 | 674 | 0.937307056379822 |
| Chr15:80.5Mb | 2081 | 15 | 79998207 | 81077025 | 2695 | 280 | AD | eGFR | -0.0676147 | -0.79887 | 0.58574 | 0.00457174 | 0 | 0.76965 | 0.810927 | 675 | 0.938216068047337 |
| Chr7:143.2Mb | 1217 | 7 | 142098980 | 144324464 | 2831 | 260 | AD | eGFR | -0.0674533 | -0.79049 | 0.59559 | 0.00454994 | 0 | 0.7453 | 0.811041 | 676 | 0.938216068047337 |
| Chr22:39.5Mb | 2483 | 22 | 38718590 | 40378783 | 3115 | 320 | AD | eGFR | -0.0463284 | -0.51361 | 0.39182 | 0.00214632 | 0 | 0.31638 | 0.81487 | 677 | 0.941142766961652 |
| Chr1:247.9Mb | 199 | 1 | 247531113 | 248341594 | 2563 | 229 | AD | eGFR | 0.0566219 | -0.48524 | 0.62727 | 0.00320604 | 0 | 0.46388 | 0.815978 | 678 | 0.941142766961652 |
| Chr15:48.5Mb | 2055 | 15 | 47693317 | 49260006 | 2379 | 233 | AD | eGFR | 0.066921 | -0.61738 | 0.85267 | 0.00447843 | 0 | 0.89788 | 0.817328 | 679 | 0.941311481590575 |
| Chr8:1.8Mb | 1235 | 8 | 1628040 | 2070902 | 1730 | 311 | AD | eGFR | -0.0596115 | -0.71066 | 0.54522 | 0.00355353 | 0 | 0.62699 | 0.821531 | 680 | 0.944420485380117 |

| locus_label | locus | chr | start | stop | n.snps | n.pcs | phen1 | phen2 | rho | rho.lower | rho.upper | r2 | r2.lower | r2.upper | p | rank | FDR |
|---|---|---|---|---|---|---|---|---|---|---|---|---|---|---|---|---|---|
| Chr21:39.9Mb | 2453 | 21 | 39252060 | 40480831 | 2783 | 324 | AD | eGFR | -0.0548596 | -0.64368 | 0.48808 | 0.00300957 | 0 | 0.48135 | 0.82341 | 681 | 0.944420485380117 |
| Chr11:111.9Mb | 1718 | 11 | 111117781 | 112755446 | 2638 | 263 | AD | eGFR | -0.0457887 | -0.52032 | 0.39926 | 0.00209661 | 0 | 0.31308 | 0.824963 | 682 | 0.944420485380117 |
| Chr4:40.2Mb | 640 | 4 | 39820062 | 40590541 | 1970 | 265 | AD | eGFR | 0.0427566 | -0.37542 | 0.4822 | 0.00182813 | 0 | 0.25879 | 0.825251 | 683 | 0.944420485380117 |
| Chr2:197.3Mb | 377 | 2 | 196446300 | 198140350 | 3219 | 224 | AD | eGFR | 0.0506823 | -0.45378 | 0.57369 | 0.0025687 | 0 | 0.3811 | 0.826066 | 684 | 0.944420485380117 |
| Chr2:46.9Mb | 252 | 2 | 46499327 | 47320114 | 2291 | 272 | AD | eGFR | 0.0634069 | -0.63295 | 0.82465 | 0.00402043 | 0 | 0.88532 | 0.827488 | 685 | 0.944665132846715 |
| Chr8:25.4Mb | 1270 | 8 | 24863244 | 25859277 | 2266 | 265 | AD | eGFR | -0.0497474 | -0.63697 | 0.48264 | 0.0024748 | 0 | 0.47686 | 0.834852 | 686 | 0.951591554585153 |
| Chr1:207Mb | 159 | 1 | 205917549 | 208162951 | 3081 | 290 | AD | eGFR | 0.0326112 | -0.29077 | 0.3634 | 0.00106349 | 0 | 0.15096 | 0.835989 | 687 | 0.951591554585153 |
| Chr15:69.9Mb | 2072 | 15 | 69089816 | 70767983 | 3436 | 345 | AD | eGFR | 0.0443432 | -0.44169 | 0.56733 | 0.00196632 | 0 | 0.39175 | 0.840288 | 688 | 0.955094790697674 |
| Chr3:144.8Mb | 547 | 3 | 144408822 | 145211907 | 2030 | 150 | AD | eGFR | 0.0856447 | -1 | 1 | 0.00733502 | 0 | 1 | 0.842548 | 689 | 0.956273637155297 |
| Chr15:87.6Mb | 2087 | 15 | 87158588 | 88035757 | 2325 | 248 | AD | eGFR | 0.063864 | -0.76019 | 0.97726 | 0.00407862 | 0 | 1 | 0.845431 | 690 | 0.957114810419681 |
| Chr19:44.5Mb | 2350 | 19 | 43960959 | 45040932 | 2548 | 237 | AD | eGFR | -0.0367591 | -0.45354 | 0.34725 | 0.00135123 | 0 | 0.22353 | 0.845737 | 691 | 0.957114810419681 |
| Chr8:117.2Mb | 1344 | 8 | 116094204 | 118300068 | 3512 | 319 | AD | eGFR | 0.052705 | -0.62432 | 0.77562 | 0.00277782 | 0 | 0.79539 | 0.847677 | 692 | 0.957492243902439 |
| Chr10:29.1Mb | 1517 | 10 | 28734423 | 29393749 | 1915 | 224 | AD | eGFR | -0.0570044 | -0.85521 | 0.64976 | 0.0032495 | 0 | 0.8802 | 0.849464 | 693 | 0.957492243902439 |
| Chr2:71.1Mb | 276 | 2 | 70756284 | 71385813 | 1933 | 232 | AD | eGFR | -0.0489183 | -0.68443 | 0.52774 | 0.002393 | 0 | 0.58309 | 0.850634 | 694 | 0.957492243902439 |
| Chr5:63.8Mb | 830 | 5 | 63214090 | 64438518 | 1716 | 195 | AD | eGFR | -0.0736442 | -1 | 0.9729 | 0.00542347 | 0 | 1 | 0.852008 | 695 | 0.957492243902439 |
| Chr16:55.4Mb | 2136 | 16 | 54866096 | 55907389 | 2801 | 293 | AD | eGFR | 0.0704321 | -0.95887 | 1 | 0.00496068 | 0 | 1 | 0.853239 | 696 | 0.957492243902439 |
| Chr7:152.7Mb | 1225 | 7 | 152253295 | 153241228 | 3224 | 306 | AD | eGFR | -0.0499763 | -0.72459 | 0.58455 | 0.00249764 | 0 | 0.64873 | 0.853417 | 697 | 0.957492243902439 |
| Chr14:97.7Mb | 2023 | 14 | 97174315 | 98268391 | 2489 | 301 | AD | eGFR | -0.0727436 | -1 | 1 | 0.00529164 | 0 | 1 | 0.859219 | 698 | 0.962620713467049 |
| Chr2:20.6Mb | 224 | 2 | 20064518 | 21084898 | 2432 | 290 | AD | eGFR | -0.0397325 | -0.54561 | 0.44894 | 0.00157867 | 0 | 0.35693 | 0.860506 | 699 | 0.96268339341917 |
| Chr12:122.6Mb | 1850 | 12 | 121817510 | 123396634 | 2659 | 227 | AD | eGFR | -0.0299134 | -0.40418 | 0.33098 | 0.00089481 | 0 | 0.19129 | 0.863855 | 700 | 0.964693495007133 |
| Chr15:79.6Mb | 2080 | 15 | 79292537 | 79998206 | 1853 | 229 | AD | eGFR | 0.0429379 | -0.52876 | 0.63411 | 0.00184366 | 0 | 0.50491 | 0.86477 | 701 | 0.964693495007133 |
| Chr7:103.7Mb | 1189 | 7 | 103191323 | 104109231 | 2735 | 229 | AD | eGFR | -0.0595403 | -1 | 0.95123 | 0.00354504 | 0 | 1 | 0.868406 | 702 | 0.964842692198582 |
| Chr12:98.5Mb | 1829 | 12 | 97814022 | 99244337 | 3080 | 282 | AD | eGFR | -0.0426092 | -0.67397 | 0.53799 | 0.00181555 | 0 | 0.5758 | 0.868472 | 703 | 0.964842692198582 |
| Chr5:143.1M | 894 | 5 | 14253295 | 14360590 | 1996 | 234 | AD | eGFR | - | -0.60859 | 0.51349 | 0.00162052 | 0 | 0.44132 | 0.869162 | 704 | 0.96484269219858 |

| locus_label | locus | chr | start | stop | n.snps | n.pcs | phen1 | phen2 | rho | rho.lower | rho.upper | r2 | r2.lower | r2.upper | p | rank | FDR |
|---|---|---|---|---|---|---|---|---|---|---|---|---|---|---|---|---|---|
| b | | | 7 | 8 | | | | | 0.0402557 | | | | | | | | 2 |
| Chr16:51.3Mb | 2133 | 16 | 50467034 | 52041336 | 2988 | 341 | AD | eGFR | 0.0281675 | -0.36502 | 0.47695 | 0.000793408 | 0 | 0.30311 | 0.869839 | 705 | 0.964842692198582 |
| Chr16:23.3Mb | 2123 | 16 | 22667049 | 24014705 | 3018 | 290 | AD | eGFR | 0.0308526 | -0.36755 | 0.42945 | 0.000951885 | 0 | 0.22555 | 0.871871 | 706 | 0.965726801699717 |
| Chr19:12.4Mb | 2319 | 19 | 11681979 | 13213186 | 2806 | 231 | AD | eGFR | -0.0343056 | -0.51585 | 0.43577 | 0.00117688 | 0 | 0.32228 | 0.873571 | 707 | 0.966241190947666 |
| Chr7:2.8Mb | 1097 | 7 | 2473750 | 3100211 | 1917 | 295 | AD | eGFR | 0.0326883 | -0.44427 | 0.52057 | 0.00106852 | 0 | 0.33854 | 0.875059 | 708 | 0.966519968926554 |
| Chr19:48.2Mb | 2354 | 19 | 47752728 | 48573065 | 2113 | 261 | AD | eGFR | 0.029871 | -0.37953 | 0.44428 | 0.000892276 | 0 | 0.23737 | 0.877142 | 709 | 0.967454222849083 |
| Chr1:90.3Mb | 78 | 1 | 89753965 | 90941549 | 2069 | 256 | AD | eGFR | -0.0301969 | -0.47638 | 0.39383 | 0.000911852 | 0 | 0.27391 | 0.88101 | 710 | 0.970351859154929 |
| Chr7:46.8Mb | 1142 | 7 | 46247336 | 47253454 | 2920 | 244 | AD | eGFR | 0.0294209 | -0.45314 | 0.54293 | 0.000865588 | 0 | 0.39669 | 0.882844 | 711 | 0.971004230661041 |
| Chr6:17Mb | 939 | 6 | 16566883 | 17391994 | 1911 | 264 | AD | eGFR | -0.0248011 | -0.4077 | 0.34145 | 0.000615093 | 0 | 0.19623 | 0.890746 | 712 | 0.976466022222222 |
| Chr7:94.4Mb | 1181 | 7 | 93691726 | 95174847 | 2389 | 249 | AD | eGFR | 0.0415599 | -0.74334 | 0.85754 | 0.00172722 | 0 | 1 | 0.893188 | 713 | 0.976466022222222 |
| Chr15:62.7Mb | 2067 | 15 | 62106139 | 63218978 | 2486 | 299 | AD | eGFR | 0.0241493 | -0.35631 | 0.42098 | 0.000583188 | 0 | 0.20936 | 0.894438 | 714 | 0.976466022222222 |
| Chr17:55.6Mb | 2216 | 17 | 54950108 | 56245227 | 2805 | 369 | AD | eGFR | 0.0220815 | -0.33528 | 0.37477 | 0.000487594 | 0 | 0.16863 | 0.895103 | 715 | 0.976466022222222 |
| Chr18:10.2Mb | 2248 | 18 | 9816165 | 10675548 | 2503 | 328 | AD | eGFR | -0.0331073 | -0.67234 | 0.5719 | 0.00109609 | 0 | 0.62805 | 0.896169 | 716 | 0.976466022222222 |
| Chr1:204.6Mb | 157 | 1 | 204092538 | 205009623 | 2079 | 263 | AD | eGFR | -0.0341362 | -0.69247 | 0.58186 | 0.00116528 | 0 | 0.60957 | 0.898242 | 717 | 0.976466022222222 |
| Chr9:137.2Mb | 1480 | 9 | 136770927 | 137593527 | 2210 | 293 | AD | eGFR | -0.0144656 | -0.24309 | 0.21326 | 0.000209252 | 0 | 0.06787 | 0.898945 | 718 | 0.976466022222222 |
| Chr11:44.5Mb | 1656 | 11 | 43949378 | 45019559 | 2758 | 340 | AD | eGFR | -0.037663 | -0.786 | 0.68209 | 0.0014185 | 0 | 0.85386 | 0.898969 | 719 | 0.976466022222222 |
| Chr11:59.9Mb | 1670 | 11 | 59256233 | 60515105 | 2437 | 202 | AD | eGFR | 0.0309843 | -0.5132 | 0.59662 | 0.000960024 | 0 | 0.44193 | 0.899048 | 720 | 0.976466022222222 |
| Chr1:155.7Mb | 113 | 1 | 154685546 | 156813845 | 2602 | 332 | AD | eGFR | 0.022249 | -0.39687 | 0.47708 | 0.000495019 | 0 | 0.3223 | 0.901477 | 721 | 0.977746205270458 |
| Chr17:65Mb | 2222 | 17 | 64438388 | 65582327 | 2647 | 318 | AD | eGFR | -0.0275539 | -0.56885 | 0.50067 | 0.000759218 | 0 | 0.41867 | 0.909056 | 722 | 0.980989997252747 |
| Chr7:74.2Mb | 1166 | 7 | 73128831 | 75243899 | 1808 | 270 | AD | eGFR | 0.0192926 | -0.35783 | 0.39461 | 0.000372205 | 0 | 0.19861 | 0.909145 | 723 | 0.980989997252747 |
| Chr8:56.9Mb | 1293 | 8 | 56346879 | 57436722 | 2575 | 218 | AD | eGFR | -0.0416014 | -1 | 0.96206 | 0.00173067 | 0 | 1 | 0.909351 | 724 | 0.980989997252747 |
| Chr15:94.1Mb | 2093 | 15 | 93707011 | 94565521 | 2958 | 331 | AD | eGFR | -0.024024 | -0.51553 | 0.45708 | 0.000577154 | 0 | 0.34592 | 0.911234 | 725 | 0.980989997252747 |
| Chr13:22.2Mb | 1866 | 13 | 21540734 | 22795188 | 3353 | 347 | AD | eGFR | -0.0243626 | -0.57208 | 0.48794 | 0.000593538 | 0 | 0.407 | 0.91216 | 726 | 0.980989997252747 |
| Chr19:56.1Mb | 2365 | 19 | 55714086 | 56411086 | 1924 | 324 | AD | eGFR | -0.0198514 | -0.41078 | 0.35528 | 0.000394077 | 0 | 0.19981 | 0.912381 | 727 | 0.980989997252747 |

| locus_label | locus | chr | start | stop | n.snps | n.pcs | phen1 | phen2 | rho | rho.lower | rho.upper | r2 | r2.lower | r2.upper | p | rank | FDR |
|---|---|---|---|---|---|---|---|---|---|---|---|---|---|---|---|---|---|
| Chr9:10.3Mb | 1383 | 9 | 9920259 | 10675474 | 2475 | 269 | AD | eGFR | -0.0247923 | -0.54263 | 0.47169 | 0.000614659 | 0 | 0.35767 | 0.913249 | 728 | 0.980989997252747 |
| Chr18:77.6Mb | 2305 | 18 | 77148138 | 78017072 | 2553 | 278 | AD | eGFR | -0.0143293 | -0.32673 | 0.299 | 0.00020533 | 0 | 0.14199 | 0.918642 | 729 | 0.984134145205479 |
| Chr1:163.5Mb | 120 | 1 | 163141863 | 163854567 | 1875 | 175 | AD | eGFR | -0.0235987 | -0.52397 | 0.46858 | 0.0005569 | 0 | 0.3423 | 0.918693 | 730 | 0.984134145205479 |
| Chr5:66.4Mb | 832 | 5 | 65606871 | 67096191 | 3094 | 285 | AD | eGFR | 0.0331357 | -0.8989 | 0.99407 | 0.00109798 | 0 | 1 | 0.922846 | 731 | 0.987230604651163 |
| Chr12:47.9Mb | 1786 | 12 | 47499604 | 48227035 | 2024 | 222 | AD | eGFR | 0.0253032 | -0.64325 | 0.71966 | 0.00064025 | 0 | 0.70686 | 0.927726 | 732 | 0.988734838018742 |
| Chr9:88.5Mb | 1432 | 9 | 87815450 | 89171796 | 2623 | 256 | AD | eGFR | 0.0158232 | -0.35267 | 0.38048 | 0.000250373 | 0 | 0.18404 | 0.929176 | 733 | 0.988734838018742 |
| Chr2:150.5Mb | 338 | 2 | 149568876 | 151428988 | 3352 | 309 | AD | eGFR | 0.0272774 | -0.74831 | 0.84019 | 0.000744055 | 0 | 1 | 0.930285 | 734 | 0.988734838018742 |
| Chr4:24.3Mb | 622 | 4 | 23858351 | 24764724 | 1921 | 268 | AD | eGFR | -0.0198079 | -0.56795 | 0.517 | 0.000392352 | 0 | 0.45993 | 0.931875 | 735 | 0.988734838018742 |
| Chr3:1.1Mb | 422 | 3 | 795747 | 1440048 | 1889 | 272 | AD | eGFR | -0.0221331 | -0.7045 | 0.62423 | 0.000489875 | 0 | 0.70526 | 0.932117 | 736 | 0.988734838018742 |
| Chr20:57.4Mb | 2422 | 20 | 56951774 | 57924037 | 1970 | 297 | AD | eGFR | -0.0139026 | -0.3649 | 0.32122 | 0.000193283 | 0 | 0.16889 | 0.932362 | 737 | 0.988734838018742 |
| Chr13:108.8Mb | 1946 | 13 | 108521979 | 109156765 | 1894 | 239 | AD | eGFR | 0.0284131 | -0.91579 | 0.9648 | 0.000807303 | 0 | 1 | 0.934103 | 738 | 0.988734838018742 |
| Chr2:228.2Mb | 404 | 2 | 227557842 | 228812485 | 3599 | 290 | AD | eGFR | -0.0146691 | -0.39204 | 0.34835 | 0.000215184 | 0 | 0.18914 | 0.934554 | 739 | 0.988734838018742 |
| Chr1:210.4Mb | 161 | 1 | 209711170 | 211082892 | 3142 | 284 | AD | eGFR | 0.0205424 | -0.58549 | 0.62909 | 0.000421989 | 0 | 0.55514 | 0.937436 | 740 | 0.988734838018742 |
| Chr11:131Mb | 1735 | 11 | 130500784 | 131424361 | 2232 | 326 | AD | eGFR | -0.0182562 | -0.56002 | 0.4863 | 0.00033329 | 0 | 0.39464 | 0.937626 | 741 | 0.988734838018742 |
| Chr16:84.8Mb | 2166 | 16 | 84440155 | 85146805 | 2881 | 340 | AD | eGFR | 0.0114928 | -0.30438 | 0.33515 | 0.000132085 | 0 | 0.13402 | 0.939569 | 742 | 0.988734838018742 |
| Chr8:80.5Mb | 1314 | 8 | 79756170 | 81340231 | 3222 | 281 | AD | eGFR | -0.0223334 | -0.82325 | 0.7464 | 0.00049878 | 0 | 1 | 0.94211 | 743 | 0.988734838018742 |
| Chr22:50Mb | 2494 | 22 | 49559208 | 50347996 | 2806 | 293 | AD | eGFR | 0.0125425 | -0.35738 | 0.37516 | 0.000157314 | 0 | 0.18068 | 0.942457 | 744 | 0.988734838018742 |
| Chr4:37.3Mb | 637 | 4 | 36671802 | 37880860 | 3597 | 285 | AD | eGFR | 0.0287621 | -1 | 1 | 0.000827258 | 0 | 1 | 0.942591 | 745 | 0.988734838018742 |
| Chr14:73Mb | 2002 | 14 | 72665320 | 73373373 | 1794 | 237 | AD | eGFR | 0.0128945 | -0.40508 | 0.41275 | 0.000166269 | 0 | 0.23685 | 0.944421 | 746 | 0.988734838018742 |
| Chr18:12.3Mb | 2250 | 18 | 11872227 | 12735558 | 2373 | 221 | AD | eGFR | -0.0215147 | -0.82633 | 0.74133 | 0.000462881 | 0 | 0.91239 | 0.944482 | 747 | 0.988734838018742 |
| Chr18:22.2Mb | 2256 | 18 | 21622717 | 22746153 | 2464 | 289 | AD | eGFR | 0.0176859 | -0.63232 | 0.65753 | 0.000312791 | 0 | 0.6062 | 0.947561 | 748 | 0.989225829333333 |
| Chr8:37.7Mb | 1280 | 8 | 36641175 | 38803980 | 3055 | 354 | AD | eGFR | 0.0213504 | -0.78624 | 0.88581 | 0.000455838 | 0 | 1 | 0.947619 | 749 | 0.989225829333333 |
| Chr3:10.6Mb | 432 | 3 | 9970732 | 11226367 | 3214 | 334 | AD | eGFR | -0.0155035 | -0.59818 | 0.5385 | 0.000240358 | 0 | 0.47866 | 0.948746 | 750 | 0.989225829333333 |
| Chr12:26.5M | 176 | 12 | 25990814 | 26958056 | 2537 | 247 | AD | eGFR | 0.0203672 | -0.80782 | 0.86181 | 0.00041482 | 0 | 1 | 0.950297 | 751 | 0.98942757978723 |

| locus_label | locus | chr | start | stop | n.snps | n.pcs | phen1 | phen2 | rho | rho.lower | rho.upper | r2 | r2.lower | r2.upper | p | rank | FDR |
|---|---|---|---|---|---|---|---|---|---|---|---|---|---|---|---|---|---|
| b | 6 | | | | | | | | | | | 2 | | | | | 4 |
| Chr17:33Mb | 2199 | 17 | 32356062 | 33614189 | 2750 | 292 | AD | eGFR | 0.0151067 | -0.54809 | 0.59481 | 0.000228212 | 0 | 0.46669 | 0.95147 | 752 | 0.989427579787234 |
| Chr10:93.7Mb | 1572 | 10 | 92911507 | 94503299 | 2739 | 194 | AD | eGFR | -0.0125583 | -0.57797 | 0.50474 | 0.000157711 | 0 | 0.47154 | 0.956233 | 753 | 0.992501590488772 |
| Chr2:75.5Mb | 280 | 2 | 75148104 | 75942397 | 2120 | 250 | AD | eGFR | 0.0198826 | -1 | 1 | 0.00039532 | 0 | 1 | 0.957932 | 754 | 0.992501590488772 |
| Chr19:3.5Mb | 2310 | 19 | 3085447 | 3893909 | 2167 | 332 | AD | eGFR | 0.00721387 | -0.28536 | 0.3076 | 5.20E-05 | 0 | 0.1239 | 0.959585 | 755 | 0.992501590488772 |
| Chr15:41.5Mb | 2050 | 15 | 40604781 | 42332685 | 2945 | 249 | AD | eGFR | -0.0103101 | -0.53085 | 0.47425 | 0.000106298 | 0 | 0.41996 | 0.959962 | 756 | 0.992501590488772 |
| Chr10:124.4Mb | 1597 | 10 | 123856185 | 124894142 | 2675 | 274 | AD | eGFR | 0.0136702 | -0.6521 | 0.68199 | 0.000186875 | 0 | 0.66099 | 0.960772 | 757 | 0.992501590488772 |
| Chr4:87.7Mb | 678 | 4 | 86931671 | 88502717 | 3231 | 204 | AD | eGFR | -0.0131308 | -0.69029 | 0.66072 | 0.000172419 | 0 | 0.68477 | 0.963134 | 758 | 0.992825181935484 |
| Chr4:182.5Mb | 765 | 4 | 182065647 | 182946258 | 2837 | 308 | AD | eGFR | -0.0194781 | -1 | 1 | 0.000379396 | 0 | 1 | 0.964285 | 759 | 0.992825181935484 |
| Chr17:61.3Mb | 2219 | 17 | 60453121 | 62112373 | 2228 | 237 | AD | eGFR | -0.00718861 | -0.34604 | 0.33033 | 5.17E-05 | 0 | 0.15521 | 0.96545 | 760 | 0.992825181935484 |
| Chr11:62.3Mb | 1672 | 11 | 61717118 | 62800368 | 2079 | 249 | AD | eGFR | -0.00689103 | -0.36035 | 0.3534 | 4.75E-05 | 0 | 0.1676 | 0.967544 | 761 | 0.992825181935484 |
| Chr7:134.4Mb | 1211 | 7 | 133807217 | 134943221 | 2722 | 212 | AD | eGFR | -0.0113487 | -0.81104 | 0.73491 | 0.000128793 | 0 | 0.96741 | 0.970164 | 762 | 0.992825181935484 |
| Chr4:112.3Mb | 699 | 4 | 111733580 | 112830229 | 2112 | 214 | AD | eGFR | 0.00872987 | -0.5031 | 0.53675 | 7.62E-05 | 0 | 0.39328 | 0.970442 | 763 | 0.992825181935484 |
| Chr14:62.2Mb | 1993 | 14 | 61652817 | 62717985 | 2342 | 274 | AD | eGFR | -0.00996235 | -0.77357 | 0.7084 | 9.92E-05 | 0 | 0.93959 | 0.972058 | 764 | 0.992825181935484 |
| Chr13:68.4Mb | 1907 | 13 | 67718880 | 69004702 | 2498 | 224 | AD | eGFR | 0.011889 | -0.94693 | 0.95721 | 0.000141349 | 0 | 1 | 0.972727 | 765 | 0.992825181935484 |
| Chr18:54.4Mb | 2282 | 18 | 53762997 | 54957867 | 2447 | 228 | AD | eGFR | -0.00869566 | -0.69624 | 0.67939 | 7.56E-05 | 0 | 0.73854 | 0.975072 | 766 | 0.992825181935484 |
| Chr2:177.4Mb | 361 | 2 | 176734751 | 178149981 | 2988 | 304 | AD | eGFR | 0.00501552 | -0.40609 | 0.40969 | 2.52E-05 | 0 | 0.26763 | 0.975344 | 767 | 0.992825181935484 |
| Chr12:3.5Mb | 1743 | 12 | 3069507 | 3871114 | 2346 | 319 | AD | eGFR | 0.00426954 | -0.32402 | 0.32673 | 1.82E-05 | 0 | 0.15455 | 0.976603 | 768 | 0.992825181935484 |
| Chr4:75.7Mb | 669 | 4 | 74885901 | 76497358 | 3173 | 262 | AD | eGFR | 0.00675588 | -0.57629 | 0.59397 | 4.56E-05 | 0 | 0.53091 | 0.977732 | 769 | 0.992825181935484 |
| Chr5:95.8Mb | 854 | 5 | 95117260 | 96467377 | 2737 | 257 | AD | eGFR | -0.00642786 | -0.61011 | 0.57965 | 4.13E-05 | 0 | 0.50185 | 0.980147 | 770 | 0.992825181935484 |
| Chr11:55.7Mb | 1665 | 11 | 55477634 | 55947452 | 2271 | 27 | AD | eGFR | 0.00715551 | -0.76834 | 0.77516 | 5.12E-05 | 0 | 0.89298 | 0.981611 | 771 | 0.992825181935484 |
| Chr16:79.3M | 215 | 16 | 78973581 | 79662634 | 2381 | 357 | AD | eGFR | - | -0.30489 | 0.298 | 1.13E-05 | 0 | 0.12053 | 0.981765 | 772 | 0.99282518193548 |

| locus_label | locus | chr | start | stop | n.snps | n.pcs | phen1 | phen2 | rho | rho.lower | rho.upper | r2 | r2.lower | r2.upper | p | rank | FDR |
|---|---|---|---|---|---|---|---|---|---|---|---|---|---|---|---|---|---|
| b | 9 | | | | | | | | 0.00336254 | | | | | | | | 4 |
| Chr13:81.3Mb | 1920 | 13 | 80778340 | 81724089 | 2307 | 177 | AD | eGFR | -0.00886219 | -1 | 1 | 7.85E-05 | 0 | 1 | 0.982444 | 773 | 0.992825181935484 |
| Chr1:239.8Mb | 190 | 1 | 239332149 | 240248508 | 2265 | 278 | AD | eGFR | -0.00608631 | -0.75718 | 0.73261 | 3.70E-05 | 0 | 0.86659 | 0.983583 | 774 | 0.992825181935484 |
| Chr7:153.8Mb | 1226 | 7 | 153241229 | 154407487 | 2491 | 299 | AD | eGFR | -0.00378248 | -0.41125 | 0.39129 | 1.43E-05 | 0 | 0.22264 | 0.983938 | 775 | 0.992825181935484 |
| Chr17:20.9Mb | 2193 | 17 | 20402495 | 21432873 | 1428 | 210 | AD | eGFR | -0.00241599 | -0.28363 | 0.2869 | 5.84E-06 | 0 | 0.1056 | 0.986129 | 776 | 0.993753708762887 |
| Chr4:119.5Mb | 706 | 4 | 119138098 | 119927262 | 1796 | 145 | AD | eGFR | 0.00585706 | -1 | 1 | 3.43E-05 | 0 | 1 | 0.989047 | 777 | 0.99469496534018 |
| Chr20:52Mb | 2415 | 20 | 51533751 | 52411532 | 1918 | 312 | AD | eGFR | -0.00221386 | -0.359 | 0.35451 | 4.90E-06 | 0 | 0.17225 | 0.989836 | 778 | 0.99469496534018 |
| Chr5:144.5Mb | 895 | 5 | 143605909 | 145319096 | 3366 | 224 | AD | eGFR | 0.00489232 | -1 | 1 | 2.39E-05 | 0 | 1 | 0.990879 | 779 | 0.99469496534018 |
| Chr2:30.1Mb | 232 | 2 | 29627933 | 30575619 | 2126 | 265 | AD | eGFR | -0.00158735 | -0.55999 | 0.57368 | 2.52E-06 | 0 | 0.45131 | 0.994886 | 780 | 0.99743698974359 |
| Chr14:34.1Mb | 1966 | 14 | 33591114 | 34695195 | 2869 | 374 | AD | eGFR | 0.00122171 | -0.81274 | 0.81598 | 1.49E-06 | 0 | 1 | 0.99691 | 781 | 0.998186453265045 |
| Chr14:75Mb | 2004 | 14 | 74245429 | 75699501 | 2869 | 226 | AD | eGFR | 7.87E-05 | -0.53903 | 0.54173 | 6.20E-09 | 0 | 0.45256 | 0.999726 | 782 | 0.999726 |

**Supplementary Table S14. Complete Local Genetic Correlation Results in European Ancestry (Wightman Validation).**
This table provides comprehensive local genetic correlation results using the Wightman AD GWAS (corrected) as validation data, paralleling Table S13 for the discovery analysis. All genomic loci tested genome-wide are included regardless of statistical significance, enabling direct comparison with discovery results and assessment of replication. For each locus, the table reports locus identifiers, genomic coordinates, SNP counts, number of principal components, phenotypes, estimated local genetic correlation with confidence bounds, test statistics, genes contained in the regions, p-values, and FDR.

| locus_label | locus | chr | start | stop | n.snps | n.pcs | phen1 | phen2 | rho | rho.lower | rho.upper | r2 | r2.lower | r2.upper | p | rank | FDR |
|---|---|---|---|---|---|---|---|---|---|---|---|---|---|---|---|---|---|
| Chr19:45.5Mb | 2351 | 19 | 45040933 | 45893307 | 1925 | 277 | AD | eGFR | -0.347578 | -0.44847 | -0.24601 | 0.12081 | 0.06052 | 0.20113 | 7.03E-11 | 1 | 7.75E-08 |
| Chr14:91.4Mb | 2018 | 14 | 90662837 | 92101228 | 3137 | 296 | AD | eGFR | 0.885286 | 0.55559 | 1 | 0.783732 | 0.30868 | 1 | 1.97E-06 | 2 | 0.00108468318 |
| Chr4:143.6Mb | 729 | 4 | 142807915 | 144401042 | 3222 | 205 | AD | eGFR | 0.72048 | 0.43382 | 1 | 0.519091 | 0.1882 | 1 | 9.64E-06 | 3 | 0.00298915995 |
| Chr16:81Mb | 2161 | 16 | 80559686 | 81437861 | 2544 | 291 | AD | eGFR | 0.830951 | 0.49934 | 1 | 0.690479 | 0.24934 | 1 | 1.09E-05 | 4 | 0.00298915995 |
| Chr11:47.2Mb | 1658 | 11 | 46316006 | 48066366 | 1892 | 160 | AD | eGFR | 0.640467 | 0.35672 | 0.96666 | 0.410198 | 0.12725 | 0.93444 | 4.34E-05 | 5 | 0.00955298064 |
| Chr5:138.2Mb | 891 | 5 | 136949854 | 139408116 | 2773 | 270 | AD | eGFR | -0.603575 | -0.95337 | -0.31643 | 0.364303 | 0.10013 | 0.90891 | 9.73E-05 | 6 | 0.014172806 |
| Chr15:63.8Mb | 2068 | 15 | 63218979 | 64339945 | 2622 | 228 | AD | eGFR | -0.557613 | -0.89851 | -0.29114 | 0.310933 | 0.08476 | 0.80733 | 0.000105361 | 7 | 0.014172806 |
| Chr5:133.6Mb | 888 | 5 | 132554691 | 134556570 | 3340 | 346 | AD | eGFR | -0.69781 | -1 | -0.36444 | 0.486939 | 0.13282 | 1 | 0.000111872 | 8 | 0.014172806 |
| Chr12:53.7Mb | 1791 | 12 | 53039988 | 54371448 | 2766 | 256 | AD | eGFR | -0.580225 | -0.91351 | -0.30555 | 0.336661 | 0.09336 | 0.8345 | 0.000115854 | 9 | 0.014172806 |
| Chr4:33.2Mb | 632 | 4 | 32746821 | 33640822 | 1694 | 109 | AD | eGFR | -0.625615 | -1 | -0.31663 | 0.391394 | 0.10025 | 1 | 0.000361666 | 10 | 0.0398194266 |
| Chr20:42.6Mb | 2406 | 20 | 42185672 | 43008890 | 1802 | 256 | AD | eGFR | 0.670498 | 0.31163 | 1 | 0.449567 | 0.09711 | 1 | 0.000559149 | 11 | 0.0530581075 |
| Chr15:96.3Mb | 2095 | 15 | 95662426 | 96864278 | 2535 | 326 | AD | eGFR | -0.780233 | -1 | -0.34775 | 0.608763 | 0.12093 | 1 | 0.00057829 | 12 | 0.0530581075 |
| Chr2:178.7Mb | 362 | 2 | 178149982 | 179224870 | 2505 | 145 | AD | eGFR | -0.744084 | -1 | -0.34108 | 0.553661 | 0.11634 | 1 | 0.000779 | 13 | 0.0620188162 |
| Chr2:179.8Mb | 363 | 2 | 179224871 | 180336045 | 2093 | 222 | AD | eGFR | -0.814033 | -1 | -0.36957 | 0.662649 | 0.13707 | 1 | 0.000812441 | 14 | 0.0620188162 |
| Chr3:119.3Mb | 524 | 3 | 118903054 | 119759983 | 1877 | 184 | AD | eGFR | 0.654597 | 0.29366 | 1 | 0.428498 | 0.08624 | 1 | 0.000844943 | 15 | 0.0620188162 |
| Chr19:18.1Mb | 2326 | 19 | 17750519 | 18504867 | 1909 | 275 | AD | eGFR | 0.694214 | 0.29867 | 1 | 0.481934 | 0.0892 | 1 | 0.00109349 | 16 | 0.075245780625 |
| Chr13:114.7Mb | 1953 | 13 | 114302562 | 115109851 | 1460 | 170 | AD | eGFR | 0.683634 | 0.29353 | 1 | 0.467356 | 0.08616 | 1 | 0.00116522 | 17 | 0.0754651305882353 |
| Chr22:20.3Mb | 2465 | 22 | 19635655 | 20969184 | 2239 | 293 | AD | eGFR | 0.54502 | 0.22186 | 0.97087 | 0.297047 | 0.04922 | 0.9426 | 0.00134169 | 18 | 0.0779918027586207 |
| Chr2:162.5Mb | 347 | 2 | 161432090 | 163607635 | 3352 | 244 | AD | eGFR | -0.691207 | -1 | -0.29001 | 0.477767 | 0.0841 | 1 | 0.00143397 | 19 | 0.0779918027586207 |
| Chr5:67.6Mb | 833 | 5 | 67096192 | 68006993 | 2007 | 261 | AD | eGFR | 0.491358 | 0.20207 | 1 | 0.241433 | 0.04083 | 1 | 0.00162034 | 20 | 0.0779918027586207 |
| Chr1:37.7Mb | 35 | 1 | 36855140 | 38474036 | 3119 | 319 | AD | eGFR | 0.635344 | 0.24766 | 1 | 0.403662 | 0.06133 | 1 | 0.00165522 | 21 | 0.0779918027586207 |
| Chr15:58.2Mb | 2063 | 15 | 57641165 | 58712954 | 2830 | 320 | AD | eGFR | -0.511305 | -1 | -0.19894 | 0.261433 | 0.03969 | 1 | 0.00166989 | 22 | 0.0779918027586207 |

| locus_label | locus | chr | start | stop | n.snps | n.pcs | phen1 | phen2 | rho | rho.lower | rho.upper | r2 | r2.lower | r2.upper | p | rank | FDR |
|---|---|---|---|---|---|---|---|---|---|---|---|---|---|---|---|---|---|
| Chr8:73.4Mb | 1308 | 8 | 72917490 | 73838027 | 2162 | 210 | AD | eGFR | -0.864502 | -1 | -0.35273 | 0.747364 | 0.12442 | 1 | 0.00173792 | 23 | 0.0779918027586207 |
| Chr8:7.4Mb | 1245 | 8 | 6663428 | 8064600 | 1209 | 232 | AD | eGFR | 0.368168 | 0.13878 | 0.61006 | 0.135547 | 0.01942 | 0.37217 | 0.00182542 | 24 | 0.0779918027586207 |
| Chr5:44.7Mb | 814 | 5 | 44049879 | 45305566 | 1721 | 141 | AD | eGFR | -0.640954 | -1 | -0.26628 | 0.410822 | 0.07118 | 1 | 0.00183036 | 25 | 0.0779918027586207 |
| Chr19:17.4Mb | 2325 | 19 | 17045964 | 17750518 | 1900 | 252 | AD | eGFR | -0.565819 | -1 | -0.23427 | 0.320151 | 0.05488 | 1 | 0.00195281 | 26 | 0.0779918027586207 |
| Chr12:69.5Mb | 1804 | 12 | 68839662 | 70097805 | 2970 | 280 | AD | eGFR | -0.703418 | -1 | -0.28014 | 0.494797 | 0.07848 | 1 | 0.00195823 | 27 | 0.0779918027586207 |
| Chr12:3.5Mb | 1743 | 12 | 3069507 | 3871114 | 2240 | 314 | AD | eGFR | -0.445088 | -0.93164 | -0.17339 | 0.198103 | 0.03007 | 0.86795 | 0.00198811 | 28 | 0.0779918027586207 |
| Chr19:10.9Mb | 2318 | 19 | 10028841 | 11681978 | 3153 | 326 | AD | eGFR | -0.493609 | -0.93909 | -0.18891 | 0.243649 | 0.03569 | 0.8819 | 0.00205428 | 29 | 0.0779918027586207 |
| Chr1:32.4Mb | 31 | 1 | 31639218 | 33066846 | 2022 | 218 | AD | eGFR | -0.748815 | -1 | -0.29306 | 0.560724 | 0.08588 | 1 | 0.00217943 | 30 | 0.079985081 |
| Chr4:103.5Mb | 692 | 4 | 102544804 | 104384534 | 3173 | 251 | AD | eGFR | 0.603787 | 0.22775 | 1 | 0.364558 | 0.05187 | 1 | 0.00234654 | 31 | 0.0817282621875 |
| Chr22:49.3Mb | 2493 | 22 | 48977539 | 49559207 | 2814 | 299 | AD | eGFR | -1 | -1 | -0.33685 | 1 | 0.11347 | 1 | 0.00237539 | 32 | 0.0817282621875 |
| Chr9:98.2Mb | 1443 | 9 | 97418634 | 98885862 | 2541 | 247 | AD | eGFR | -1 | -1 | -0.34612 | 1 | 0.12025 | 1 | 0.00289856 | 33 | 0.0967065018181818 |
| Chr22:50.8Mb | 2495 | 22 | 50347997 | 51244236 | 2214 | 263 | AD | eGFR | -0.513362 | -0.91862 | -0.18499 | 0.263541 | 0.03422 | 0.84386 | 0.00304949 | 34 | 0.0987496614705882 |
| Chr1:245.3Mb | 196 | 1 | 244723197 | 245970505 | 3037 | 354 | AD | eGFR | 0.54834 | 0.1847 | 1 | 0.300677 | 0.03423 | 1 | 0.00326906 | 35 | 0.102835287428571 |
| Chr13:21Mb | 1865 | 13 | 20491182 | 21540733 | 2457 | 270 | AD | eGFR | 0.578746 | 0.19895 | 1 | 0.334947 | 0.03975 | 1 | 0.00340109 | 36 | 0.104016669166667 |
| Chr1:191.9Mb | 146 | 1 | 191419276 | 192468373 | 1726 | 167 | AD | eGFR | 0.787805 | 0.29457 | 1 | 0.620636 | 0.08729 | 1 | 0.00408277 | 37 | 0.121489993783784 |
| Chr2:160.8Mb | 346 | 2 | 160200380 | 161432089 | 2779 | 184 | AD | eGFR | -0.686713 | -1 | -0.23726 | 0.471574 | 0.05691 | 1 | 0.0046088 | 38 | 0.133533915789474 |
| Chr2:242.5Mb | 420 | 2 | 241910367 | 243188366 | 1902 | 211 | AD | eGFR | -0.777227 | -1 | -0.27862 | 0.604082 | 0.07828 | 1 | 0.0047529 | 39 | 0.134178023076923 |
| Chr11:3.9Mb | 1614 | 11 | 3362509 | 4405707 | 1829 | 210 | AD | eGFR | -0.611196 | -1 | -0.19094 | 0.373561 | 0.03656 | 1 | 0.00543728 | 40 | 0.149661132 |
| Chr4:5.5Mb | 602 | 4 | 5051835 | 5972709 | 2942 | 316 | AD | eGFR | 0.603383 | 0.18822 | 1 | 0.364071 | 0.036 | 1 | 0.005921 | 41 | 0.159000512195122 |
| Chr1:57.2Mb | 49 | 1 | 56799637 | 57529670 | 1888 | 233 | AD | eGFR | 0.602172 | 0.18285 | 1 | 0.362611 | 0.03444 | 1 | 0.0062872 | 42 | 0.159642497727273 |
| Chr4:149.2Mb | 733 | 4 | 148462209 | 149998528 | 2783 | 268 | AD | eGFR | -0.529138 | -1 | -0.1688 | 0.279987 | 0.02874 | 1 | 0.00631699 | 43 | 0.159642497727273 |
| Chr2:122.6Mb | 314 | 2 | 122026818 | 123219243 | 2134 | 168 | AD | eGFR | -0.567006 | -1 | -0.1789 | 0.321496 | 0.03265 | 1 | 0.0063799 | 44 | 0.159642497727273 |
| Chr1:214.4Mb | 164 | 1 | 213958293 | 214783872 | 1881 | 230 | AD | eGFR | -0.458373 | -0.88854 | -0.13663 | 0.210106 | 0.01881 | 0.78951 | 0.00677524 | 45 | 0.1618837734 |
| Chr10:53.5Mb | 1535 | 10 | 53129610 | 53919161 | 2196 | 227 | AD | eGFR | 0.944332 | 0.28474 | 1 | 0.891762 | 0.08247 | 1 | 0.00693207 | 46 | 0.1618837734 |
| Chr7:105.9Mb | 1191 | 7 | 105312564 | 106399739 | 2574 | 312 | AD | eGFR | 0.476852 | 0.13612 | 0.90529 | 0.227388 | 0.01862 | 0.81954 | 0.00709189 | 47 | 0.1618837734 |
| Chr3:26.2Mb | 446 | 3 | 25516194 | 26905340 | 2756 | 257 | AD | eGFR | 0.818184 | 0.24116 | 1 | 0.669426 | 0.0593 | 1 | 0.0072976 | 48 | 0.1618837734 |
| Chr1:93.5Mb | 81 | 1 | 92904466 | 94168576 | 1557 | 168 | AD | eGFR | -0.660093 | -1 | -0.20506 | 0.435722 | 0.04321 | 1 | 0.00732267 | 49 | 0.1618837734 |
| Chr21:28.2Mb | 2443 | 21 | 27774351 | 28616539 | 2392 | 258 | AD | eGFR | -0.600343 | -1 | -0.17298 | 0.360412 | 0.03003 | 1 | 0.00735167 | 50 | 0.1618837734 |
| Chr15:78.9Mb | 2079 | 15 | 78514102 | 79292536 | 1672 | 192 | AD | eGFR | 0.641432 | 0.19486 | 1 | 0.411435 | 0.03938 | 1 | 0.00769549 | 51 | 0.165930016153846 |

| locus_label | locus | chr | start | stop | n.snps | n.pcs | phen1 | phen2 | rho | rho.lower | rho.upper | r2 | r2.lower | r2.upper | p | rank | FDR |
|---|---|---|---|---|---|---|---|---|---|---|---|---|---|---|---|---|---|
| Chr18:2.4Mb | 2240 | 18 | 1940212 | 2839842 | 2370 | 280 | AD | eGFR | 0.787034 | 0.23279 | 1 | 0.619423 | 0.05573 | 1 | 0.00783684 | 52 | 0.165930016153846 |
| Chr10:6.6Mb | 1493 | 10 | 6234719 | 6897037 | 1734 | 292 | AD | eGFR | -0.603683 | -1 | -0.17436 | 0.364433 | 0.03058 | 1 | 0.00802369 | 53 | 0.166149463333333 |
| Chr21:35.1Mb | 2449 | 21 | 34383795 | 35729261 | 2663 | 306 | AD | eGFR | 0.568038 | 0.16547 | 1 | 0.322667 | 0.02785 | 1 | 0.00814902 | 54 | 0.166149463333333 |
| Chr17:2.1Mb | 2175 | 17 | 1480057 | 2699135 | 2427 | 316 | AD | eGFR | -0.356337 | -0.64378 | -0.10059 | 0.126976 | 0.01018 | 0.41445 | 0.00836668 | 55 | 0.167485721454545 |
| Chr12:46.8Mb | 1785 | 12 | 46037745 | 47499603 | 2369 | 223 | AD | eGFR | 0.745984 | 0.21026 | 1 | 0.556492 | 0.04538 | 1 | 0.00906041 | 56 | 0.171149603076923 |
| Chr14:100.1Mb | 2026 | 14 | 99474534 | 100786189 | 2794 | 332 | AD | eGFR | 0.41717 | 0.11491 | 0.82228 | 0.174031 | 0.01349 | 0.67615 | 0.00908744 | 57 | 0.171149603076923 |
| Chr16:88Mb | 2170 | 16 | 87669169 | 88387228 | 2434 | 287 | AD | eGFR | 0.553382 | 0.1436 | 1 | 0.306232 | 0.02166 | 1 | 0.00942784 | 58 | 0.171149603076923 |
| Chr16:81.9Mb | 2162 | 16 | 81437862 | 82310204 | 2822 | 332 | AD | eGFR | -0.50682 | -1 | -0.14371 | 0.256866 | 0.02171 | 1 | 0.00953621 | 59 | 0.171149603076923 |
| Chr13:92.5Mb | 1932 | 13 | 91764205 | 93285142 | 3290 | 279 | AD | eGFR | -0.570108 | -1 | -0.15377 | 0.325023 | 0.02376 | 1 | 0.0095586 | 60 | 0.171149603076923 |
| Chr5:7.5Mb | 783 | 5 | 6936734 | 8079415 | 2970 | 256 | AD | eGFR | 0.924918 | 0.26523 | 1 | 0.855473 | 0.07139 | 1 | 0.0095749 | 61 | 0.171149603076923 |
| Chr2:146.6Mb | 336 | 2 | 145977921 | 147252196 | 1976 | 215 | AD | eGFR | -0.709762 | -1 | -0.20218 | 0.503762 | 0.04208 | 1 | 0.00973349 | 62 | 0.171149603076923 |
| Chr18:73.2Mb | 2300 | 18 | 72785207 | 73568260 | 2225 | 266 | AD | eGFR | -0.881215 | -1 | -0.2505 | 0.77654 | 0.06587 | 1 | 0.00992443 | 63 | 0.171149603076923 |
| Chr3:176.3Mb | 576 | 3 | 175671425 | 176931163 | 2663 | 249 | AD | eGFR | -0.685276 | -1 | -0.18386 | 0.469603 | 0.0346 | 1 | 0.0101032 | 64 | 0.171149603076923 |
| Chr4:150.5Mb | 734 | 4 | 149998529 | 151014322 | 1657 | 178 | AD | eGFR | 0.788674 | 0.21374 | 1 | 0.622007 | 0.04745 | 1 | 0.0101042 | 65 | 0.171149603076923 |
| Chr3:4.5Mb | 426 | 3 | 4109394 | 4835083 | 1684 | 273 | AD | eGFR | 0.608879 | 0.16116 | 1 | 0.370734 | 0.02662 | 1 | 0.0103272 | 66 | 0.172276472727273 |
| Chr2:98.2Mb | 295 | 2 | 97339302 | 98989360 | 1742 | 158 | AD | eGFR | 0.84235 | 0.24365 | 1 | 0.709554 | 0.06255 | 1 | 0.0111621 | 67 | 0.183424956716418 |
| Chr2:183.8Mb | 367 | 2 | 183352291 | 184148655 | 1791 | 169 | AD | eGFR | -0.58818 | -1 | -0.15181 | 0.345955 | 0.02394 | 1 | 0.0116322 | 68 | 0.188339002941176 |
| Chr10:63.3Mb | 1544 | 10 | 62626466 | 64069687 | 2968 | 261 | AD | eGFR | -0.563201 | -1 | -0.14387 | 0.317195 | 0.02147 | 1 | 0.0118153 | 69 | 0.188531091304348 |
| Chr1:154Mb | 112 | 1 | 153410810 | 154685545 | 1762 | 222 | AD | eGFR | 0.653684 | 0.16251 | 1 | 0.427303 | 0.02787 | 1 | 0.0121328 | 70 | 0.190831611428571 |
| Chr18:74.9Mb | 2302 | 18 | 74471746 | 75240760 | 2674 | 304 | AD | eGFR | 0.603442 | 0.14885 | 1 | 0.364142 | 0.02414 | 1 | 0.0125424 | 71 | 0.194495526760563 |
| Chr1:242.1Mb | 193 | 1 | 241813535 | 242417923 | 1755 | 235 | AD | eGFR | 0.608504 | 0.1456 | 1 | 0.370277 | 0.02208 | 1 | 0.01273 | 72 | 0.194662916666667 |
| Chr8:59.1Mb | 1295 | 8 | 58675118 | 59555977 | 1989 | 214 | AD | eGFR | -0.728878 | -1 | -0.1697 | 0.531263 | 0.03135 | 1 | 0.013158 | 73 | 0.197575937837838 |
| Chr6:2.1Mb | 927 | 6 | 1380680 | 2746531 | 3115 | 342 | AD | eGFR | 0.361799 | 0.07734 | 0.7201 | 0.130898 | 0.00641 | 0.51854 | 0.0132794 | 74 | 0.197575937837838 |
| Chr4:4.7Mb | 601 | 4 | 4266179 | 5051834 | 2272 | 272 | AD | eGFR | 0.567376 | 0.12129 | 1 | 0.321916 | 0.01633 | 1 | 0.0136958 | 75 | 0.200734598734177 |
| Chr7:8.3Mb | 1104 | 7 | 7926501 | 8718351 | 2257 | 278 | AD | eGFR | -0.562512 | -1 | -0.13105 | 0.31642 | 0.01751 | 1 | 0.0139129 | 76 | 0.200734598734177 |
| Chr16:10.7Mb | 2114 | 16 | 10351809 | 11022542 | 1772 | 267 | AD | eGFR | -0.479401 | -0.95255 | -0.1153 | 0.229825 | 0.0146 | 0.90735 | 0.0140532 | 77 | 0.200734598734177 |
| Chr13:108.8Mb | 1946 | 13 | 108521979 | 109156765 | 1822 | 232 | AD | eGFR | 0.696715 | 0.15753 | 1 | 0.485412 | 0.02687 | 1 | 0.0143024 | 78 | 0.200734598734177 |
| Chr17:28.6Mb | 2196 | 17 | 27344402 | 29783141 | 2488 | 227 | AD | eGFR | 0.593278 | 0.13604 | 1 | 0.351979 | 0.02002 | 1 | 0.0144033 | 79 | 0.200734598734177 |
| Chr12:23.4Mb | 1763 | 12 | 22928056 | 23923798 | 1891 | 267 | AD | eGFR | 0.524219 | 0.1261 | 1 | 0.274806 | 0.01713 | 1 | 0.0147805 | 80 | 0.20341663125 |
| Chr3:132.5Mb | 537 | 3 | 132124019 | 132925967 | 1774 | 151 | AD | eGFR | 0.549875 | 0.13027 | 1 | 0.302363 | 0.01874 | 1 | 0.0154395 | 81 | 0.207271872289157 |
| Chr15:45.5Mb | 2052 | 15 | 44987756 | 46022642 | 1791 | 179 | AD | eGFR | 0.340029 | 0.06859 | 0.78057 | 0.115619 | 0.00556 | 0.6093 | 0.01562 | 82 | 0.207271872289157 |
| Chr1:10.1Mb | 12 | 1 | 9475967 | 10753427 | 1904 | 232 | AD | eGFR | 0.364344 | 0.07735 | 0.74151 | 0.132747 | 0.00616 | 0.54984 | 0.0156254 | 83 | 0.207271872289157 |
| Chr3:197.6Mb | 595 | 3 | 197147991 | 197962380 | 1372 | 177 | AD | eGFR | -0.664161 | -1 | -0.13593 | 0.44111 | 0.0223 | 1 | 0.0161877 | 84 | 0.211601837647059 |
| Chr4:3.9Mb | 600 | 4 | 3549230 | 4266178 | 1324 | 223 | AD | eGFR | 0.264549 | 0.05066 | 0.49407 | 0.0699859 | 0.00295 | 0.24411 | 0.0163362 | 85 | 0.211601837647059 |
| Chr7:15.4Mb | 1114 | 7 | 15013695 | 15877564 | 2144 | 243 | AD | eGFR | -0.634715 | -1 | -0.13396 | 0.402863 | 0.01994 | 1 | 0.0170111 | 86 | 0.215387237931034 |
| Chr5:136.1Mb | 890 | 5 | 13534494 | 13694985 | 2957 | 260 | AD | eGFR | -0.666246 | -1 | -0.12729 | 0.443883 | 0.02007 | 1 | 0.0170197 | 87 | 0.215387237931034 |

| locus_label | locus | chr | start | stop | n.snps | n.pcs | phen1 | phen2 | rho | rho.lower | rho.upper | r2 | r2.lower | r2.upper | p | rank | FDR |
|---|---|---|---|---|---|---|---|---|---|---|---|---|---|---|---|---|---|
| | | | 4 | 3 | | | | | | | | | | | | | |
| Chr9:81.9Mb | 1427 | 9 | 81161598 | 82601465 | 2576 | 324 | AD | eGFR | 0.574793 | 0.11839 | 1 | 0.330387 | 0.01588 | 1 | 0.0175795 | 88 | 0.218621487640449 |
| Chr17:14.1Mb | 2187 | 17 | 13648447 | 14508610 | 2235 | 287 | AD | eGFR | -0.621446 | -1 | -0.12724 | 0.386195 | 0.01842 | 1 | 0.0176724 | 89 | 0.218621487640449 |
| Chr18:47Mb | 2275 | 18 | 46558307 | 47455925 | 2088 | 160 | AD | eGFR | -0.583625 | -1 | -0.12343 | 0.340619 | 0.0181 | 1 | 0.0178907 | 90 | 0.218862896666667 |
| Chr11:128.9Mb | 1733 | 11 | 128262551 | 129451784 | 2454 | 309 | AD | eGFR | -0.349271 | -0.68858 | -0.06276 | 0.12199 | 0.00496 | 0.47414 | 0.0185257 | 91 | 0.224140612087912 |
| Chr5:34.6Mb | 806 | 5 | 33971057 | 35157146 | 2721 | 239 | AD | eGFR | -0.380878 | -0.785 | -0.07107 | 0.145068 | 0.00646 | 0.61623 | 0.0195192 | 92 | 0.231500476595745 |
| Chr21:16.4Mb | 2432 | 21 | 15840034 | 16873443 | 2236 | 303 | AD | eGFR | -0.276172 | -0.53661 | -0.05231 | 0.076271 | 0.00317 | 0.28795 | 0.0195563 | 93 | 0.231500476595745 |
| Chr20:5.8Mb | 2375 | 20 | 5379451 | 6128358 | 2099 | 281 | AD | eGFR | -0.393538 | -0.7822 | -0.06546 | 0.154872 | 0.00548 | 0.61184 | 0.0197648 | 94 | 0.231500476595745 |
| Chr13:30.8Mb | 1874 | 13 | 30068526 | 31433218 | 3463 | 320 | AD | eGFR | -0.616396 | -1 | -0.10589 | 0.379944 | 0.01561 | 1 | 0.0209044 | 95 | 0.242270993684211 |
| Chr2:17.9Mb | 222 | 2 | 17241116 | 18656325 | 3618 | 264 | AD | eGFR | -0.713563 | -1 | -0.10921 | 0.509172 | 0.01768 | 1 | 0.0216989 | 96 | 0.248859259375 |
| Chr11:14.2Mb | 1626 | 11 | 13364728 | 15126767 | 2767 | 249 | AD | eGFR | -0.811796 | -1 | -0.15651 | 0.659012 | 0.02885 | 1 | 0.0221858 | 97 | 0.251820265979381 |
| Chr13:111.3Mb | 1949 | 13 | 110995433 | 111621244 | 1859 | 261 | AD | eGFR | -0.382501 | -0.75526 | -0.05203 | 0.146307 | 0.00405 | 0.57042 | 0.0235998 | 98 | 0.261598679411765 |
| Chr6:165.3Mb | 1088 | 6 | 164687463 | 165835953 | 3163 | 253 | AD | eGFR | 0.967153 | 0.17071 | 1 | 0.935386 | 0.0398 | 1 | 0.0238273 | 99 | 0.261598679411765 |
| Chr18:33.3Mb | 2264 | 18 | 32458908 | 34153297 | 3116 | 294 | AD | eGFR | -0.669586 | -1 | -0.09915 | 0.448345 | 0.01488 | 1 | 0.0239565 | 100 | 0.261598679411765 |
| Chr17:65Mb | 2222 | 17 | 64438388 | 65582327 | 2545 | 314 | AD | eGFR | 0.607873 | 0.09299 | 1 | 0.36951 | 0.012 | 1 | 0.0240982 | 101 | 0.261598679411765 |
| Chr2:152Mb | 339 | 2 | 151428989 | 152497375 | 2390 | 204 | AD | eGFR | 0.49223 | 0.06924 | 1 | 0.242291 | 0.00743 | 1 | 0.0242353 | 102 | 0.261598679411765 |
| Chr9:118.5Mb | 1463 | 9 | 117989018 | 119001227 | 2072 | 199 | AD | eGFR | -0.440389 | -0.94602 | -0.06427 | 0.193943 | 0.00632 | 0.89517 | 0.0251407 | 103 | 0.268736997087379 |
| Chr16:86.4Mb | 2168 | 16 | 86058598 | 86748867 | 2632 | 354 | AD | eGFR | -0.630052 | -1 | -0.0817 | 0.396965 | 0.01324 | 1 | 0.0256739 | 104 | 0.269783071962617 |
| Chr17:18.1Mb | 2191 | 17 | 17326541 | 18900978 | 2046 | 200 | AD | eGFR | 0.440448 | 0.05719 | 1 | 0.193995 | 0.00585 | 1 | 0.0258737 | 105 | 0.269783071962617 |
| Chr11:95.7Mb | 1702 | 11 | 95327211 | 96150134 | 1941 | 245 | AD | eGFR | 0.464863 | 0.06791 | 1 | 0.216097 | 0.00728 | 1 | 0.0260016 | 106 | 0.269783071962617 |
| Chr5:146.2Mb | 896 | 5 | 145319097 | 147180171 | 3442 | 295 | AD | eGFR | 0.489455 | 0.0715 | 1 | 0.239566 | 0.00744 | 1 | 0.0262187 | 107 | 0.269783071962617 |
| Chr9:3.6Mb | 1375 | 9 | 3079046 | 4025355 | 2060 | 304 | AD | eGFR | 0.407307 | 0.05583 | 0.83956 | 0.165899 | 0.005 | 0.70485 | 0.0270748 | 108 | 0.276012544444444 |
| Chr15:69.9Mb | 2072 | 15 | 69089816 | 70767983 | 3333 | 341 | AD | eGFR | -0.385033 | -0.79801 | -0.05238 | 0.14825 | 0.00454 | 0.63682 | 0.0274881 | 109 | 0.277267832727273 |
| Chr6:134.9Mb | 1061 | 6 | 134313644 | 135544083 | 2485 | 298 | AD | eGFR | -0.683476 | -1 | -0.08554 | 0.46714 | 0.01383 | 1 | 0.0277016 | 110 | 0.277267832727273 |
| Chr9:102.4Mb | 1446 | 9 | 101596751 | 103165725 | 2110 | 254 | AD | eGFR | 0.482225 | 0.05601 | 1 | 0.232541 | 0.00676 | 1 | 0.0279537 | 111 | 0.277270483783784 |
| Chr12:13.1Mb | 1753 | 12 | 12721875 | 13559527 | 1684 | 279 | AD | eGFR | 0.335445 | 0.04321 | 0.66487 | 0.112524 | 0.0032 | 0.44205 | 0.0283544 | 112 | 0.278733878571429 |
| Chr10:92.4Mb | 1571 | 10 | 91960436 | 92911506 | 2330 | 200 | AD | eGFR | 0.979562 | 0.14558 | 1 | 0.959541 | 0.03444 | 1 | 0.0290427 | 113 | 0.282973563716814 |
| Chr20:62.6Mb | 2429 | 20 | 62179242 | 62963627 | 1788 | 238 | AD | eGFR | -0.400203 | -1 | -0.05367 | 0.160162 | 0.0047 | 1 | 0.0295177 | 114 | 0.285078839473684 |
| Chr19:19.1Mb | 2327 | 19 | 18504868 | 19749166 | 2088 | 228 | AD | eGFR | 0.303564 | 0.0307 | 0.61396 | 0.0921512 | 0.00202 | 0.37694 | 0.0299215 | 115 | 0.286465839130435 |
| Chr13:25.4Mb | 1869 | 13 | 24818334 | 25977140 | 3137 | 274 | AD | eGFR | 0.56609 | 0.05895 | 1 | 0.320457 | 0.00719 | 1 | 0.0302235 | 116 | 0.286862702586207 |
| Chr7:100.2Mb | 1186 | 7 | 99465541 | 100849307 | 1941 | 228 | AD | eGFR | -0.506707 | -1 | -0.05339 | 0.256752 | 0.00508 | 1 | 0.0304853 | 117 | 0.28687448974359 |
| Chr11:68.4Mb | 1676 | 11 | 68000950 | 68887344 | 1649 | 180 | AD | eGFR | 0.391883 | 0.0503 | 0.87499 | 0.153572 | 0.00429 | 0.7656 | 0.0309073 | 118 | 0.287773643697479 |
| Chr10:5.8Mb | 1492 | 10 | 5396243 | 6234718 | 2511 | 308 | AD | eGFR | 0.454245 | 0.05455 | 1 | 0.206338 | 0.00532 | 1 | 0.0311036 | 119 | 0.287773643697479 |
| Chr1:151.8Mb | 110 | 1 | 151146408 | 152535630 | 1977 | 222 | AD | eGFR | -0.347559 | -0.70361 | -0.03398 | 0.120797 | 0.00269 | 0.49507 | 0.0317708 | 120 | 0.288392510655738 |
| Chr16:67.6Mb | 2146 | 16 | 66738844 | 68435728 | 1454 | 151 | AD | eGFR | -0.441963 | -0.98513 | -0.05093 | 0.195331 | 0.00472 | 0.97048 | 0.0318245 | 121 | 0.288392510655738 |
| Chr15:101.1M | 2100 | 15 | 10074211 | 10155075 | 2341 | 270 | AD | eGFR | 0.664424 | 0.07888 | 1 | 0.44146 | 0.01256 | 1 | 0.0319563 | 122 | 0.288392510655738 |

| locus_label | locus | chr | start | stop | n.snps | n.pcs | phen1 | phen2 | rho | rho.lower | rho.upper | r2 | r2.lower | r2.upper | p | rank | FDR |
|---|---|---|---|---|---|---|---|---|---|---|---|---|---|---|---|---|---|
| b | | | 8 | 0 | | | | | | | | | | | | | |
| Chr6:34.4Mb | 970 | 6 | 33864263 | 34979270 | 1804 | 175 | AD | eGFR | 0.258286 | 0.02592 | 0.50418 | 0.0667117 | 0.00147 | 0.2542 | 0.0326407 | 123 | 0.292174070731707 |
| Chr19:33.4Mb | 2339 | 19 | 33028100 | 33785835 | 2105 | 203 | AD | eGFR | 0.248396 | 0.02082 | 0.49285 | 0.0617007 | 0.00116 | 0.2429 | 0.0330093 | 124 | 0.2922600096 |
| Chr19:15.3Mb | 2322 | 19 | 14891101 | 15650185 | 1791 | 234 | AD | eGFR | 0.481927 | 0.04304 | 1 | 0.232254 | 0.00503 | 1 | 0.0331812 | 125 | 0.2922600096 |
| Chr8:103.1Mb | 1331 | 8 | 102548681 | 103716053 | 2589 | 264 | AD | eGFR | -0.454481 | -1 | -0.04057 | 0.206553 | 0.00415 | 1 | 0.0335948 | 126 | 0.293554561904762 |
| Chr7:127.3Mb | 1206 | 7 | 126640873 | 127903738 | 2273 | 204 | AD | eGFR | 0.444541 | 0.04219 | 0.9653 | 0.197617 | 0.00422 | 0.9318 | 0.0343716 | 127 | 0.29680809609375 |
| Chr3:57.8Mb | 470 | 3 | 56954879 | 58557718 | 3307 | 218 | AD | eGFR | -0.557278 | -1 | -0.04503 | 0.310559 | 0.00698 | 1 | 0.0345063 | 128 | 0.29680809609375 |
| Chr1:236.8Mb | 186 | 1 | 236199672 | 237409458 | 3371 | 304 | AD | eGFR | 0.473777 | 0.0458 | 1 | 0.224465 | 0.00452 | 1 | 0.0353107 | 129 | 0.300840627692308 |
| Chr7:116.1Mb | 1198 | 7 | 115321302 | 116835336 | 2561 | 246 | AD | eGFR | 0.487322 | 0.04253 | 1 | 0.237482 | 0.00597 | 1 | 0.0355216 | 130 | 0.300840627692308 |
| Chr10:98.7Mb | 1576 | 10 | 98005995 | 99367800 | 2624 | 284 | AD | eGFR | -0.425783 | -0.95808 | -0.03251 | 0.181291 | 0.00366 | 0.91791 | 0.0367495 | 131 | 0.307191952941177 |
| Chr4:140.3Mb | 727 | 4 | 139553761 | 141087047 | 2759 | 328 | AD | eGFR | 0.644949 | 0.04138 | 1 | 0.415959 | 0.00821 | 1 | 0.0369054 | 132 | 0.307191952941177 |
| Chr5:73.8Mb | 836 | 5 | 73314062 | 74245354 | 2051 | 258 | AD | eGFR | -0.61019 | -1 | -0.04804 | 0.372331 | 0.00656 | 1 | 0.0371097 | 133 | 0.307191952941177 |
| Chr20:5Mb | 2374 | 20 | 4591848 | 5379450 | 2220 | 274 | AD | eGFR | -0.747165 | -1 | -0.05832 | 0.558255 | 0.01223 | 1 | 0.0375303 | 134 | 0.307191952941177 |
| Chr2:211.7Mb | 388 | 2 | 210842458 | 212520453 | 3089 | 270 | AD | eGFR | -0.26252 | -0.58125 | -0.01814 | 0.068917 | 0.00128 | 0.33785 | 0.0377708 | 135 | 0.307191952941177 |
| Chr7:4.5Mb | 1099 | 7 | 4139432 | 4938442 | 2558 | 308 | AD | eGFR | -0.428959 | -0.96694 | -0.02961 | 0.184006 | 0.00323 | 0.93497 | 0.0379456 | 136 | 0.307191952941177 |
| Chr17:49.3Mb | 2211 | 17 | 48722591 | 49909298 | 2120 | 270 | AD | eGFR | -0.484971 | -1 | -0.02841 | 0.235197 | 0.0046 | 1 | 0.0386034 | 137 | 0.309494291304348 |
| Chr17:36.7Mb | 2202 | 17 | 36116885 | 37361178 | 2077 | 280 | AD | eGFR | 0.243829 | 0.01393 | 0.48717 | 0.0594527 | 0.00086 | 0.23733 | 0.0387922 | 138 | 0.309494291304348 |
| Chr11:15.8Mb | 1627 | 11 | 15126768 | 16383386 | 2214 | 239 | AD | eGFR | -0.318905 | -0.65447 | -0.01698 | 0.1017 | 0.00162 | 0.42833 | 0.0397301 | 139 | 0.314696691366906 |
| Chr10:82.1Mb | 1561 | 10 | 81763231 | 82378048 | 1820 | 157 | AD | eGFR | -0.450233 | -1 | -0.02801 | 0.20271 | 0.00432 | 1 | 0.0407798 | 140 | 0.319517227659574 |
| Chr20:58.4Mb | 2423 | 20 | 57924038 | 58780894 | 1919 | 299 | AD | eGFR | 0.567424 | 0.03952 | 1 | 0.32197 | 0.0058 | 1 | 0.0409191 | 141 | 0.319517227659574 |
| Chr8:13.2Mb | 1253 | 8 | 12947936 | 13395051 | 1637 | 161 | AD | eGFR | -0.702987 | -1 | -0.03851 | 0.49419 | 0.00894 | 1 | 0.0412985 | 142 | 0.320208792253521 |
| Chr7:51.4Mb | 1147 | 7 | 50894509 | 51951647 | 2358 | 237 | AD | eGFR | -0.716361 | -1 | -0.03874 | 0.513173 | 0.00956 | 1 | 0.0420424 | 143 | 0.323170260416667 |
| Chr20:56.6Mb | 2421 | 20 | 56172844 | 56951773 | 2002 | 291 | AD | eGFR | -0.540843 | -1 | -0.04154 | 0.292512 | 0.00596 | 1 | 0.0422675 | 144 | 0.323170260416667 |
| Chr14:58Mb | 1989 | 14 | 57460782 | 58447798 | 2075 | 221 | AD | eGFR | 0.603161 | 0.05153 | 1 | 0.363803 | 0.0083 | 1 | 0.0437311 | 145 | 0.331824809589041 |
| Chr16:89.7Mb | 2172 | 16 | 89106305 | 90292787 | 2733 | 253 | AD | eGFR | 0.234288 | 0.0077 | 0.46951 | 0.0548908 | 8.00E-04 | 0.22044 | 0.0440022 | 146 | 0.331824809589041 |
| Chr1:215.3Mb | 165 | 1 | 214783873 | 215825791 | 1773 | 169 | AD | eGFR | -0.418784 | -0.92347 | -0.01437 | 0.17538 | 0.00291 | 0.8528 | 0.0452225 | 147 | 0.333675184768212 |
| Chr11:65.7Mb | 1674 | 11 | 64594823 | 66782661 | 2717 | 259 | AD | eGFR | -0.225958 | -0.46772 | -0.00585 | 0.0510571 | 0.00069 | 0.21876 | 0.0452404 | 148 | 0.333675184768212 |
| Chr1:218Mb | 168 | 1 | 217352166 | 218563960 | 3122 | 255 | AD | eGFR | -0.513104 | -1 | -0.02023 | 0.263275 | 0.00365 | 1 | 0.0455922 | 149 | 0.333675184768212 |
| Chr18:23.4Mb | 2257 | 18 | 22746154 | 24088163 | 2413 | 277 | AD | eGFR | 0.468333 | 0.01736 | 1 | 0.219336 | 0.00359 | 1 | 0.0457225 | 150 | 0.333675184768212 |
| Chr20:10.8Mb | 2380 | 20 | 10279604 | 11257012 | 2420 | 273 | AD | eGFR | -0.321494 | -0.67731 | -0.00696 | 0.103359 | 0.00149 | 0.45875 | 0.0457629 | 151 | 0.333675184768212 |
| Chr3:160Mb | 561 | 3 | 159478752 | 160584961 | 1993 | 166 | AD | eGFR | 0.644137 | 0.02278 | 1 | 0.414913 | 0.00707 | 1 | 0.0461719 | 152 | 0.3344425125 |
| Chr7:31Mb | 1128 | 7 | 30175114 | 31730554 | 3298 | 324 | AD | eGFR | 0.557174 | 0.02475 | 1 | 0.310443 | 0.00578 | 1 | 0.0472654 | 153 | 0.340125525490196 |
| Chr20:40.8Mb | 2404 | 20 | 40272391 | 41351514 | 2664 | 282 | AD | eGFR | 0.588775 | 0.00684 | 1 | 0.346656 | 0.0048 | 1 | 0.0484433 | 154 | 0.344162655483871 |
| Chr12:49.7Mb | 1788 | 12 | 48960266 | 50344346 | 2228 | 227 | AD | eGFR | 0.28962 | 0.01198 | 0.61575 | 0.0838795 | 0.00133 | 0.37914 | 0.0484516 | 155 | 0.344162655483871 |
| Chr17:66.2Mb | 2223 | 17 | 65582328 | 66757556 | 2395 | 267 | AD | eGFR | 0.306264 | -0.0069 | 0.67188 | 0.0937978 | 0 | 0.45142 | 0.0504425 | 156 | 0.348337049090909 |
| Chr22:23.5Mb | 2468 | 22 | 23143365 | 23950732 | 1846 | 234 | AD | eGFR | 0.416851 | 0.00192 | 0.95281 | 0.173765 | 0.00227 | 0.90785 | 0.0506769 | 157 | 0.348337049090909 |

| locus_label | locus | chr | start | stop | n.snps | n.pcs | phen1 | phen2 | rho | rho.lower | rho.upper | r2 | r2.lower | r2.upper | p | rank | FDR |
|---|---|---|---|---|---|---|---|---|---|---|---|---|---|---|---|---|---|
| Chr13:28Mb | 1872 | 13 | 27392006 | 28587125 | 3368 | 309 | AD | eGFR | -0.385036 | -0.8671 | -0.00398 | 0.148253 | 0.00174 | 0.75187 | 0.0508302 | 158 | 0.348337049090909 |
| Chr5:66.4Mb | 832 | 5 | 65606871 | 67096191 | 2891 | 275 | AD | eGFR | 0.671017 | -0.00228 | 1 | 0.450263 | 0 | 1 | 0.0510172 | 159 | 0.348337049090909 |
| Chr3:180.4Mb | 579 | 3 | 179372166 | 181424323 | 3009 | 267 | AD | eGFR | -0.550349 | -1 | -0.00138 | 0.302884 | 0.00408 | 1 | 0.0511652 | 160 | 0.348337049090909 |
| Chr13:51Mb | 1893 | 13 | 50419897 | 51554236 | 1888 | 214 | AD | eGFR | 0.401301 | -0.00131 | 1 | 0.161042 | 0 | 1 | 0.0512993 | 161 | 0.348337049090909 |
| Chr11:1.5Mb | 1611 | 11 | 1061400 | 1857845 | 2392 | 240 | AD | eGFR | 0.403177 | -0.00177 | 0.89256 | 0.162552 | 0 | 0.79666 | 0.0515974 | 162 | 0.348337049090909 |
| Chr6:136.5Mb | 1062 | 6 | 135544084 | 137553212 | 2763 | 306 | AD | eGFR | 0.51333 | -0.00378 | 1 | 0.263507 | 0 | 1 | 0.0519773 | 163 | 0.348337049090909 |
| Chr19:49.9Mb | 2356 | 19 | 49415190 | 50451925 | 1857 | 284 | AD | eGFR | 0.257924 | -0.00339 | 0.53926 | 0.0665246 | 0 | 0.2908 | 0.0520391 | 164 | 0.348337049090909 |
| Chr6:19.9Mb | 943 | 6 | 19449153 | 20418020 | 2279 | 296 | AD | eGFR | -0.302963 | -0.64961 | 0.00376 | 0.0917867 | 0 | 0.422 | 0.0522031 | 165 | 0.348337049090909 |
| Chr6:46.9Mb | 981 | 6 | 46430702 | 47313443 | 1891 | 227 | AD | eGFR | 0.538809 | -0.02116 | 1 | 0.290315 | 0 | 1 | 0.0537479 | 166 | 0.354113057142857 |
| Chr10:126.3Mb | 1599 | 10 | 125798320 | 126875816 | 2610 | 294 | AD | eGFR | 0.345651 | 0.00797 | 1 | 0.119474 | 0.00171 | 1 | 0.0539278 | 167 | 0.354113057142857 |
| Chr2:240.7Mb | 418 | 2 | 240325601 | 241113279 | 2447 | 299 | AD | eGFR | -0.429794 | -1 | 0.00249 | 0.184723 | 0 | 1 | 0.0540336 | 168 | 0.354113057142857 |
| Chr1:108.6Mb | 95 | 1 | 107874654 | 109271449 | 2701 | 262 | AD | eGFR | 0.429263 | -0.0117 | 1 | 0.184267 | 0 | 1 | 0.0547824 | 169 | 0.356895990532544 |
| Chr13:106Mb | 1943 | 13 | 105672501 | 106313847 | 1650 | 213 | AD | eGFR | 0.625046 | -0.0234 | 1 | 0.390683 | 0 | 1 | 0.0564697 | 170 | 0.364399542857143 |
| Chr3:148Mb | 550 | 3 | 147358244 | 148638308 | 2889 | 279 | AD | eGFR | -0.487505 | -1 | 0.00733 | 0.237661 | 0 | 1 | 0.0569183 | 171 | 0.364399542857143 |
| Chr9:128Mb | 1471 | 9 | 127184583 | 128785782 | 1852 | 195 | AD | eGFR | -0.485949 | -1 | 0.00387 | 0.236146 | 0 | 1 | 0.0570618 | 172 | 0.364399542857143 |
| Chr2:102Mb | 298 | 2 | 101556800 | 102494487 | 1712 | 206 | AD | eGFR | -0.316477 | -0.67362 | 0.00938 | 0.100158 | 0 | 0.45377 | 0.0576991 | 173 | 0.364399542857143 |
| Chr14:35.9Mb | 1968 | 14 | 35379989 | 36422077 | 2012 | 155 | AD | eGFR | 0.5014 | -0.0129 | 1 | 0.251402 | 0 | 1 | 0.0578347 | 174 | 0.364399542857143 |
| Chr3:71.8Mb | 483 | 3 | 71223282 | 72334704 | 2476 | 343 | AD | eGFR | 0.377203 | -0.00249 | 0.95497 | 0.142282 | 0 | 0.91197 | 0.05792 | 175 | 0.364399542857143 |
| Chr5:17.2Mb | 791 | 5 | 16592320 | 17803477 | 2568 | 330 | AD | eGFR | -0.603044 | -1 | 0.02602 | 0.363662 | 0 | 1 | 0.0592734 | 176 | 0.370795530681818 |
| Chr17:54.2Mb | 2215 | 17 | 53468007 | 54950107 | 2793 | 273 | AD | eGFR | -0.348792 | -0.80118 | 0.00747 | 0.121656 | 0 | 0.64189 | 0.0599824 | 177 | 0.372864783707865 |
| Chr3:171.6Mb | 572 | 3 | 171134850 | 172004760 | 1897 | 242 | AD | eGFR | -0.367016 | -0.84219 | 0.01284 | 0.134701 | 0 | 0.70929 | 0.0602815 | 178 | 0.372864783707865 |
| Chr14:86.1Mb | 2013 | 14 | 85562937 | 86653343 | 2187 | 238 | AD | eGFR | -0.506068 | -1 | 0.02632 | 0.256105 | 0 | 1 | 0.0610826 | 179 | 0.37343973480663 |
| Chr2:55.2Mb | 262 | 2 | 54688235 | 55663336 | 2304 | 211 | AD | eGFR | -0.455892 | -1 | 0.01016 | 0.207837 | 0 | 1 | 0.0611776 | 180 | 0.37343973480663 |
| Chr12:15.4Mb | 1755 | 12 | 14656850 | 16136742 | 2249 | 252 | AD | eGFR | -0.207652 | -0.4332 | 0.00929 | 0.0431194 | 0 | 0.18767 | 0.061392 | 181 | 0.37343973480663 |
| Chr19:57.6Mb | 2367 | 19 | 57143510 | 57959952 | 2015 | 285 | AD | eGFR | -0.411545 | -0.97764 | 0.01675 | 0.169369 | 0 | 0.95578 | 0.0622827 | 182 | 0.37582635 |
| Chr1:22.4Mb | 24 | 1 | 22145630 | 22740287 | 1718 | 214 | AD | eGFR | -0.356844 | -0.80465 | 0.01494 | 0.127337 | 0 | 0.64746 | 0.0626852 | 183 | 0.37582635 |
| Chr4:176.5Mb | 758 | 4 | 175959698 | 177129678 | 2501 | 257 | AD | eGFR | 0.609738 | -0.0524 | 1 | 0.37178 | 0 | 1 | 0.0628084 | 184 | 0.37582635 |
| Chr3:134.5Mb | 539 | 3 | 133718494 | 135288394 | 3197 | 294 | AD | eGFR | -0.482399 | -1 | 0.02506 | 0.232708 | 0 | 1 | 0.0642494 | 185 | 0.382370753513513 |
| Chr9:139.5Mb | 1483 | 9 | 138995792 | 140097759 | 1791 | 268 | AD | eGFR | -0.269147 | -0.64385 | 0.0259 | 0.0724401 | 0 | 0.41455 | 0.0687228 | 186 | 0.406694680748663 |
| Chr15:80.5Mb | 2081 | 15 | 79998207 | 81077025 | 2575 | 277 | AD | eGFR | -0.419673 | -1 | 0.03681 | 0.176125 | 0 | 1 | 0.0690753 | 187 | 0.406694680748663 |
| Chr3:76.8Mb | 488 | 3 | 76157471 | 77507832 | 3112 | 262 | AD | eGFR | 0.541212 | -0.04326 | 1 | 0.29291 | 0 | 1 | 0.0708164 | 188 | 0.413324092105263 |
| Chr15:35.9Mb | 2046 | 15 | 35118928 | 36583327 | 2949 | 287 | AD | eGFR | 0.413663 | -0.02995 | 1 | 0.171117 | 0 | 1 | 0.0710355 | 189 | 0.413324092105263 |
| Chr19:6Mb | 2313 | 19 | 5478685 | 6455428 | 2300 | 264 | AD | eGFR | -0.54984 | -1 | 0.05531 | 0.302324 | 0 | 1 | 0.0713275 | 190 | 0.413324092105263 |

| locus_label | locus | chr | start | stop | n.snps | n.pcs | phen1 | phen2 | rho | rho.lower | rho.upper | r2 | r2.lower | r2.upper | p | rank | FDR |
|---|---|---|---|---|---|---|---|---|---|---|---|---|---|---|---|---|---|
| Chr12:63.4Mb | 1798 | 12 | 63055937 | 63764111 | 1718 | 217 | AD | eGFR | -0.710942 | -1 | 0.06716 | 0.505438 | 0 | 1 | 0.07232 | 191 | 0.414610929015544 |
| Chr1:170.6Mb | 128 | 1 | 170193208 | 171044255 | 1786 | 157 | AD | eGFR | 0.524646 | -0.03984 | 1 | 0.275254 | 0 | 1 | 0.0723962 | 192 | 0.414610929015544 |
| Chr9:119.4Mb | 1464 | 9 | 119001228 | 119890464 | 1925 | 241 | AD | eGFR | 0.244105 | -0.02272 | 0.53159 | 0.0595871 | 0 | 0.28259 | 0.0726793 | 193 | 0.414610929015544 |
| Chr4:24.3Mb | 622 | 4 | 23858351 | 24764724 | 1846 | 263 | AD | eGFR | 0.304138 | -0.02262 | 0.68336 | 0.0924997 | 0 | 0.46699 | 0.0732385 | 194 | 0.41506969744898 |
| Chr7:72.2Mb | 1165 | 7 | 71215742 | 73128830 | 2476 | 234 | AD | eGFR | -0.571555 | -1 | 0.04917 | 0.326676 | 0 | 1 | 0.0737588 | 195 | 0.41506969744898 |
| Chr16:14.9Mb | 2118 | 16 | 13893408 | 15921108 | 2372 | 282 | AD | eGFR | 0.304007 | -0.02834 | 0.69003 | 0.0924204 | 0 | 0.47613 | 0.0738907 | 196 | 0.41506969744898 |
| Chr3:37.8Mb | 457 | 3 | 36840137 | 38729767 | 3307 | 290 | AD | eGFR | -0.291297 | -0.7113 | 0.02965 | 0.084854 | 0 | 0.50594 | 0.0759602 | 197 | 0.424528833502538 |
| Chr1:67.3Mb | 57 | 1 | 66778016 | 67761890 | 2181 | 193 | AD | eGFR | -0.494774 | -1 | 0.04736 | 0.244802 | 0 | 1 | 0.0772875 | 198 | 0.429765340909091 |
| Chr2:192Mb | 374 | 2 | 191051955 | 193033982 | 3311 | 305 | AD | eGFR | -0.433667 | -1 | 0.04545 | 0.188067 | 0 | 1 | 0.0779343 | 199 | 0.431184242713568 |
| Chr18:60.1Mb | 2288 | 18 | 59449423 | 60780194 | 3191 | 251 | AD | eGFR | 0.399942 | -0.04579 | 1 | 0.159954 | 0 | 1 | 0.0790561 | 200 | 0.43412156119403 |
| Chr9:4.3Mb | 1376 | 9 | 4025356 | 4596614 | 1760 | 285 | AD | eGFR | -0.266705 | -0.59089 | 0.0338 | 0.0711313 | 0 | 0.34915 | 0.0792538 | 201 | 0.43412156119403 |
| Chr22:38Mb | 2482 | 22 | 37364005 | 38718589 | 2886 | 326 | AD | eGFR | 0.314022 | -0.0394 | 0.74825 | 0.0986098 | 0 | 0.55988 | 0.0802749 | 202 | 0.435841965865385 |
| Chr14:104.8Mb | 2030 | 14 | 104439107 | 105091340 | 1652 | 235 | AD | eGFR | 0.380327 | -0.05387 | 0.94317 | 0.144648 | 0 | 0.88956 | 0.0806598 | 203 | 0.435841965865385 |
| Chr6:24.4Mb | 948 | 6 | 23939307 | 24950379 | 2874 | 255 | AD | eGFR | -0.389712 | -1 | 0.0587 | 0.151876 | 0 | 1 | 0.0809671 | 204 | 0.435841965865385 |
| Chr12:25.5Mb | 1765 | 12 | 25058715 | 25990813 | 2398 | 246 | AD | eGFR | -0.497865 | -1 | 0.07634 | 0.247869 | 0 | 1 | 0.0817 | 205 | 0.435841965865385 |
| Chr6:37Mb | 972 | 6 | 36346354 | 37570051 | 2863 | 296 | AD | eGFR | 0.473173 | -0.06108 | 1 | 0.223892 | 0 | 1 | 0.081751 | 206 | 0.435841965865385 |
| Chr12:12.2Mb | 1752 | 12 | 11742150 | 12721874 | 2167 | 300 | AD | eGFR | 0.210878 | -0.02534 | 0.45858 | 0.0444696 | 0 | 0.21029 | 0.0820881 | 207 | 0.435841965865385 |
| Chr13:79.1Mb | 1918 | 13 | 78693311 | 79489045 | 1881 | 192 | AD | eGFR | -0.611552 | -1 | 0.06016 | 0.373996 | 0 | 1 | 0.0823389 | 208 | 0.435841965865385 |
| Chr2:43.7Mb | 248 | 2 | 43295777 | 44104110 | 1941 | 222 | AD | eGFR | -0.272168 | -0.62215 | 0.0337 | 0.0740755 | 0 | 0.38707 | 0.0832992 | 209 | 0.436594054929577 |
| Chr7:5.5Mb | 1100 | 7 | 4938443 | 6020653 | 3077 | 316 | AD | eGFR | 0.669724 | -0.09876 | 1 | 0.448531 | 0 | 1 | 0.0839411 | 210 | 0.436594054929577 |
| Chr10:2.9Mb | 1488 | 10 | 2637066 | 3225336 | 1880 | 252 | AD | eGFR | 0.368531 | -0.06039 | 0.99838 | 0.135815 | 0 | 0.99677 | 0.0841974 | 211 | 0.436594054929577 |
| Chr14:92.7Mb | 2019 | 14 | 92101229 | 93386328 | 3237 | 314 | AD | eGFR | 0.209568 | -0.02866 | 0.45463 | 0.0439185 | 0 | 0.20669 | 0.084243 | 212 | 0.436594054929577 |
| Chr3:120.2Mb | 525 | 3 | 119759984 | 120574477 | 1754 | 186 | AD | eGFR | -0.447079 | -1 | 0.06712 | 0.199879 | 0 | 1 | 0.0844637 | 213 | 0.436594054929577 |
| Chr7:156.8Mb | 1229 | 7 | 156344387 | 157199331 | 2492 | 231 | AD | eGFR | -0.196173 | -0.42771 | 0.02435 | 0.0384837 | 0 | 0.18294 | 0.0861402 | 214 | 0.440417934841629 |
| Chr4:2Mb | 598 | 4 | 1529268 | 2468935 | 1964 | 242 | AD | eGFR | -0.261058 | -0.57817 | 0.04219 | 0.0681512 | 0 | 0.33428 | 0.0869093 | 215 | 0.440417934841629 |
| Chr1:160.5Mb | 117 | 1 | 159882516 | 161054076 | 2362 | 288 | AD | eGFR | 0.44478 | -0.07638 | 1 | 0.19783 | 0 | 1 | 0.08732 | 216 | 0.440417934841629 |
| Chr17:62.8Mb | 2220 | 17 | 62112374 | 63548724 | 2204 | 226 | AD | eGFR | -0.329631 | -0.80004 | 0.05306 | 0.108657 | 0 | 0.64006 | 0.087498 | 217 | 0.440417934841629 |
| Chr8:8.9Mb | 1247 | 8 | 8589771 | 9167795 | 1791 | 235 | AD | eGFR | 0.386253 | -0.05782 | 1 | 0.149191 | 0 | 1 | 0.0876005 | 218 | 0.440417934841629 |
| Chr21:43.4Mb | 2457 | 21 | 43081435 | 43812791 | 2454 | 336 | AD | eGFR | 0.528893 | -0.07652 | 1 | 0.279728 | 0 | 1 | 0.0879133 | 219 | 0.440417934841629 |
| Chr21:42.6Mb | 2456 | 21 | 42187153 | 43081434 | 2383 | 325 | AD | eGFR | -0.310697 | -0.72792 | 0.0429 | 0.0965327 | 0 | 0.52987 | 0.0880402 | 220 | 0.440417934841629 |
| Chr1:241.4Mb | 192 | 1 | 241033585 | 241813534 | 1893 | 271 | AD | eGFR | 0.643492 | -0.10172 | 1 | 0.414081 | 0 | 1 | 0.0884036 | 221 | 0.440417934841629 |
| Chr10:70.5Mb | 1551 | 10 | 69904171 | 71050504 | 2861 | 198 | AD | eGFR | 0.350282 | -0.05189 | 1 | 0.122698 | 0 | 1 | 0.0889623 | 222 | 0.44120492027027 |
| Chr12:26.5Mb | 1766 | 12 | 25990814 | 26958056 | 2445 | 241 | AD | eGFR | 0.461206 | -0.06783 | 1 | 0.212711 | 0 | 1 | 0.0897433 | 223 | 0.443082391479821 |
| Chr8:144.3Mb | 1368 | 8 | 143611462 | 144934288 | 3306 | 281 | AD | eGFR | 0.226975 | -0.03088 | 0.49831 | 0.0515178 | 0 | 0.24831 | 0.0907976 | 224 | 0.443683684210526 |
| Chr6:45.9Mb | 980 | 6 | 45408422 | 46430701 | 1998 | 244 | AD | eGFR | 0.329407 | -0.05708 | 0.77274 | 0.108509 | 0 | 0.59713 | 0.0911381 | 225 | 0.443683684210526 |
| Chr2:228.2Mb | 404 | 2 | 227557842 | 228812485 | 3385 | 284 | AD | eGFR | -0.307206 | -0.72309 | 0.06078 | 0.0943755 | 0 | 0.52286 | 0.0912645 | 226 | 0.443683684210526 |

| locus_label | locus | chr | start | stop | n.snps | n.pcs | phen1 | phen2 | rho | rho.lower | rho.upper | r2 | r2.lower | r2.upper | p | rank | FDR |
|---|---|---|---|---|---|---|---|---|---|---|---|---|---|---|---|---|---|
| Chr8:86.9Mb | 1319 | 8 | 86251871 | 87563145 | 1744 | 154 | AD | eGFR | -0.265802 | -0.59007 | 0.0379 | 0.0706509 | 0 | 0.34818 | 0.091748 | 227 | 0.443683684210526 |
| Chr11:0.6Mb | 1610 | 11 | 87150 | 1061399 | 2220 | 275 | AD | eGFR | -0.300432 | -0.73108 | 0.04455 | 0.0902595 | 0 | 0.53448 | 0.09188 | 228 | 0.443683684210526 |
| Chr8:61.8Mb | 1298 | 8 | 61324890 | 62330900 | 1909 | 222 | AD | eGFR | -0.492966 | -1 | 0.08697 | 0.243016 | 0 | 1 | 0.09255 | 229 | 0.444021811304348 |
| Chr7:143.2Mb | 1217 | 7 | 142098980 | 144324464 | 2694 | 255 | AD | eGFR | 0.409933 | -0.07005 | 1 | 0.168045 | 0 | 1 | 0.0927566 | 230 | 0.444021811304348 |
| Chr11:91.6Mb | 1698 | 11 | 90967062 | 92209168 | 2669 | 201 | AD | eGFR | -0.704244 | -1 | 0.13098 | 0.49596 | 0 | 1 | 0.0936202 | 231 | 0.444383531896552 |
| Chr3:121.9Mb | 527 | 3 | 121362443 | 122340832 | 2014 | 160 | AD | eGFR | 0.391189 | -0.06936 | 1 | 0.153029 | 0 | 1 | 0.0936394 | 232 | 0.444383531896552 |
| Chr14:77.1Mb | 2006 | 14 | 76694202 | 77425453 | 1774 | 269 | AD | eGFR | -0.362121 | -0.9714 | 0.06726 | 0.131131 | 0 | 0.94411 | 0.0948059 | 233 | 0.447988394420601 |
| Chr16:77.5Mb | 2156 | 16 | 77270666 | 77739132 | 1740 | 186 | AD | eGFR | -0.527437 | -1 | 0.10256 | 0.27819 | 0 | 1 | 0.0956886 | 234 | 0.450227130769231 |
| Chr12:128.8Mb | 1856 | 12 | 128372589 | 129170296 | 2234 | 323 | AD | eGFR | -0.569612 | -1 | 0.12805 | 0.324458 | 0 | 1 | 0.0969847 | 235 | 0.454033272457627 |
| Chr15:66.9Mb | 2070 | 15 | 66355635 | 67396520 | 2760 | 290 | AD | eGFR | 0.336503 | -0.0744 | 0.87115 | 0.113234 | 0 | 0.75891 | 0.0973223 | 236 | 0.454033272457627 |
| Chr4:41.8Mb | 642 | 4 | 41360938 | 42189251 | 1733 | 167 | AD | eGFR | -0.367303 | -0.97873 | 0.07825 | 0.134912 | 0 | 0.95791 | 0.0986722 | 237 | 0.456985464 |
| Chr7:141.2Mb | 1216 | 7 | 140217631 | 142098979 | 2899 | 286 | AD | eGFR | -0.26205 | -0.60361 | 0.04201 | 0.0686704 | 0 | 0.36435 | 0.0993853 | 238 | 0.456985464 |
| Chr10:29.7Mb | 1518 | 10 | 29393750 | 30000223 | 1876 | 220 | AD | eGFR | 0.29072 | -0.05024 | 0.67227 | 0.084518 | 0 | 0.45195 | 0.0999785 | 239 | 0.456985464 |
| Chr17:69.9Mb | 2226 | 17 | 69245591 | 70495119 | 2578 | 318 | AD | eGFR | 0.361533 | -0.06903 | 1 | 0.130706 | 0 | 1 | 0.0999845 | 240 | 0.456985464 |
| Chr14:62.2Mb | 1993 | 14 | 61652817 | 62717985 | 2267 | 271 | AD | eGFR | 0.291564 | -0.06547 | 0.68823 | 0.0850097 | 0 | 0.47366 | 0.100445 | 241 | 0.456985464 |
| Chr16:24.6Mb | 2124 | 16 | 24014706 | 25235251 | 2556 | 321 | AD | eGFR | -0.516208 | -1 | 0.11684 | 0.26647 | 0 | 1 | 0.100804 | 242 | 0.456985464 |
| Chr1:235.6Mb | 185 | 1 | 235097114 | 236199671 | 1856 | 216 | AD | eGFR | 0.256297 | -0.04798 | 0.59589 | 0.0656883 | 0 | 0.35509 | 0.100954 | 243 | 0.456985464 |
| Chr2:221.9Mb | 398 | 2 | 221445192 | 222327713 | 1716 | 248 | AD | eGFR | -0.547794 | -1 | 0.11839 | 0.300078 | 0 | 1 | 0.101315 | 244 | 0.456985464 |
| Chr1:92.4Mb | 80 | 1 | 91870060 | 92904465 | 1967 | 208 | AD | eGFR | -0.33999 | -0.85615 | 0.07043 | 0.115593 | 0 | 0.73299 | 0.101784 | 245 | 0.456985464 |
| Chr6:139.3Mb | 1064 | 6 | 138825722 | 139716713 | 2124 | 233 | AD | eGFR | -0.435562 | -1 | 0.09947 | 0.189714 | 0 | 1 | 0.102286 | 246 | 0.456985464 |
| Chr2:133Mb | 324 | 2 | 132598286 | 133352950 | 804 | 106 | AD | eGFR | 0.676191 | -0.13413 | 1 | 0.457234 | 0 | 1 | 0.102737 | 247 | 0.456985464 |
| Chr4:145.3Mb | 730 | 4 | 144401043 | 146174871 | 2604 | 209 | AD | eGFR | 0.293253 | -0.05578 | 0.67892 | 0.0859974 | 0 | 0.46093 | 0.103279 | 248 | 0.456985464 |
| Chr9:138.6Mb | 1482 | 9 | 138202113 | 138995791 | 2044 | 318 | AD | eGFR | -0.444569 | -1 | 0.09884 | 0.197641 | 0 | 1 | 0.103521 | 249 | 0.456985464 |
| Chr10:131.2Mb | 1604 | 10 | 130694009 | 131694738 | 2596 | 296 | AD | eGFR | -0.319039 | -0.88128 | 0.07236 | 0.101786 | 0 | 0.7771 | 0.103766 | 250 | 0.456985464 |
| Chr3:190Mb | 586 | 3 | 189451806 | 190591293 | 3085 | 288 | AD | eGFR | 0.364959 | -0.082 | 0.9496 | 0.133195 | 0 | 0.90174 | 0.104222 | 251 | 0.457165027888446 |
| Chr8:74.4Mb | 1309 | 8 | 73838028 | 75011699 | 2436 | 257 | AD | eGFR | 0.59351 | -0.13376 | 1 | 0.352254 | 0 | 1 | 0.10598 | 252 | 0.46187380078125 |
| Chr18:24.7Mb | 2258 | 18 | 24088164 | 25305260 | 2510 | 287 | AD | eGFR | 0.228606 | -0.05012 | 0.51989 | 0.0522608 | 0 | 0.27028 | 0.107055 | 253 | 0.46187380078125 |
| Chr10:132.1Mb | 1605 | 10 | 131694739 | 132576793 | 2681 | 256 | AD | eGFR | -0.497538 | -1 | 0.11504 | 0.247544 | 0 | 1 | 0.10714 | 254 | 0.46187380078125 |
| Chr16:32.1Mb | 2129 | 16 | 31384211 | 32847928 | 867 | 92 | AD | eGFR | -0.344564 | -0.84589 | 0.0838 | 0.118724 | 0 | 0.71636 | 0.107369 | 255 | 0.46187380078125 |
| Chr1:198.6Mb | 152 | 1 | 197935822 | 199263389 | 2835 | 234 | AD | eGFR | 0.524033 | -0.11703 | 1 | 0.274611 | 0 | 1 | 0.107393 | 256 | 0.46187380078125 |
| Chr20:20.8Mb | 2391 | 20 | 19957929 | 21682377 | 2875 | 280 | AD | eGFR | 0.36556 | -0.08314 | 0.99116 | 0.133634 | 0 | 0.98239 | 0.108105 | 257 | 0.46312686770428 |
| Chr17:76.1Mb | 2232 | 17 | 75619257 | 76596287 | 2854 | 354 | AD | eGFR | 0.256395 | -0.05144 | 0.59915 | 0.0657382 | 0 | 0.35898 | 0.110261 | 258 | 0.470532406976744 |

| locus_label | locus | chr | start | stop | n.snps | n.pcs | phen1 | phen2 | rho | rho.lower | rho.upper | r2 | r2.lower | r2.upper | p | rank | FDR |
|---|---|---|---|---|---|---|---|---|---|---|---|---|---|---|---|---|---|
| Chr19:56.1Mb | 2365 | 19 | 55714086 | 56411086 | 1748 | 315 | AD | eGFR | -0.394572 | -1 | 0.09649 | 0.155687 | 0 | 1 | 0.111127 | 259 | 0.471261876923077 |
| Chr9:95.4Mb | 1440 | 9 | 94924483 | 95854925 | 1962 | 137 | AD | eGFR | -0.354506 | -0.96008 | 0.08693 | 0.125674 | 0 | 0.92175 | 0.111288 | 260 | 0.471261876923077 |
| Chr2:230.2Mb | 406 | 2 | 229764919 | 230541092 | 1878 | 252 | AD | eGFR | 0.523022 | -0.13957 | 1 | 0.273552 | 0 | 1 | 0.113154 | 261 | 0.473588477272727 |
| Chr2:42.8Mb | 247 | 2 | 42307419 | 43295776 | 2189 | 233 | AD | eGFR | 0.400264 | -0.10793 | 1 | 0.160211 | 0 | 1 | 0.113176 | 262 | 0.473588477272727 |
| Chr3:187.3Mb | 584 | 3 | 186602046 | 187939199 | 3267 | 353 | AD | eGFR | 0.282834 | -0.07058 | 0.68601 | 0.0799951 | 0 | 0.47062 | 0.11345 | 263 | 0.473588477272727 |
| Chr2:68.4Mb | 274 | 2 | 67979829 | 68877774 | 1975 | 208 | AD | eGFR | 0.429029 | -0.11729 | 1 | 0.184066 | 0 | 1 | 0.113558 | 264 | 0.473588477272727 |
| Chr20:61.1Mb | 2427 | 20 | 60766558 | 61470768 | 1857 | 284 | AD | eGFR | -0.264593 | -0.7012 | 0.06682 | 0.0700092 | 0 | 0.49168 | 0.115782 | 265 | 0.481041441509434 |
| Chr11:62.3Mb | 1672 | 11 | 61717118 | 62800368 | 1937 | 240 | AD | eGFR | 0.503325 | -0.13881 | 1 | 0.253336 | 0 | 1 | 0.117077 | 266 | 0.484593146616541 |
| Chr8:37.7Mb | 1280 | 8 | 36641175 | 38803980 | 2922 | 349 | AD | eGFR | 0.646713 | -0.15339 | 1 | 0.418238 | 0 | 1 | 0.117557 | 267 | 0.484757516853933 |
| Chr8:11Mb | 1250 | 8 | 10478852 | 11466761 | 2718 | 285 | AD | eGFR | 0.307887 | -0.08935 | 0.83582 | 0.0947942 | 0 | 0.69933 | 0.119524 | 268 | 0.491029567164179 |
| Chr3:15.6Mb | 437 | 3 | 15150877 | 16084005 | 1730 | 184 | AD | eGFR | -0.537217 | -1 | 0.16433 | 0.288602 | 0 | 1 | 0.120696 | 269 | 0.494001100371747 |
| Chr1:91.4Mb | 79 | 1 | 90941550 | 91870059 | 1774 | 197 | AD | eGFR | -0.388685 | -1 | 0.09878 | 0.151076 | 0 | 1 | 0.121962 | 270 | 0.497333933333333 |
| Chr20:0.5Mb | 2369 | 20 | 60795 | 884241 | 2360 | 348 | AD | eGFR | 0.400446 | -0.12776 | 1 | 0.160357 | 0 | 1 | 0.123879 | 271 | 0.502217911764706 |
| Chr8:78Mb | 1312 | 8 | 77377990 | 78633659 | 2648 | 191 | AD | eGFR | 0.303719 | -0.08062 | 0.7685 | 0.0922454 | 0 | 0.59059 | 0.124072 | 272 | 0.502217911764706 |
| Chr7:91.6Mb | 1179 | 7 | 90660832 | 92530165 | 2317 | 261 | AD | eGFR | -0.414104 | -1 | 0.13937 | 0.171482 | 0 | 1 | 0.128106 | 273 | 0.515240076086957 |
| Chr1:186.6Mb | 141 | 1 | 186179448 | 187072605 | 1824 | 152 | AD | eGFR | -0.222755 | -0.52518 | 0.06712 | 0.0496197 | 0 | 0.27582 | 0.128652 | 274 | 0.515240076086957 |
| Chr15:95.1Mb | 2094 | 15 | 94565522 | 95662425 | 2670 | 342 | AD | eGFR | -0.387559 | -1 | 0.10941 | 0.150202 | 0 | 1 | 0.129006 | 275 | 0.515240076086957 |
| Chr11:118.1Mb | 1723 | 11 | 117295642 | 118901213 | 3601 | 333 | AD | eGFR | -0.250821 | -0.6349 | 0.07464 | 0.062911 | 0 | 0.40309 | 0.129161 | 276 | 0.515240076086957 |
| Chr8:124.7Mb | 1350 | 8 | 124001020 | 125453322 | 3007 | 301 | AD | eGFR | 0.365126 | -0.10552 | 1 | 0.133317 | 0 | 1 | 0.1302 | 277 | 0.517509747292419 |
| Chr10:122.3Mb | 1595 | 10 | 121874446 | 122803506 | 2507 | 257 | AD | eGFR | 0.584362 | -0.20859 | 1 | 0.34148 | 0 | 1 | 0.131197 | 278 | 0.519233167259786 |
| Chr18:67.6Mb | 2295 | 18 | 67195981 | 68051035 | 2006 | 235 | AD | eGFR | -0.450097 | -1 | 0.14351 | 0.202587 | 0 | 1 | 0.131632 | 279 | 0.519233167259786 |
| Chr15:99.4Mb | 2098 | 15 | 98828262 | 99925972 | 2730 | 326 | AD | eGFR | 0.231003 | -0.07159 | 0.55246 | 0.0533622 | 0 | 0.30521 | 0.132241 | 280 | 0.519233167259786 |
| Chr16:72.6Mb | 2150 | 16 | 72089512 | 73140781 | 1654 | 234 | AD | eGFR | 0.188477 | -0.0595 | 0.4402 | 0.0355238 | 0 | 0.19378 | 0.13252 | 281 | 0.519233167259786 |
| Chr6:6Mb | 931 | 6 | 5378214 | 6621937 | 3421 | 352 | AD | eGFR | 0.486861 | -0.18106 | 1 | 0.237033 | 0 | 1 | 0.133381 | 282 | 0.520753478723404 |
| Chr11:114.3Mb | 1720 | 11 | 113889020 | 114742317 | 1793 | 253 | AD | eGFR | 0.332976 | -0.09725 | 0.94896 | 0.110873 | 0 | 0.90559 | 0.134387 | 283 | 0.522498158450704 |
| Chr11:92.9Mb | 1699 | 11 | 92209169 | 93576641 | 2339 | 243 | AD | eGFR | -0.421173 | -1 | 0.13823 | 0.177387 | 0 | 1 | 0.134777 | 284 | 0.522498158450704 |
| Chr3:128.3Mb | 533 | 3 | 127495174 | 129091975 | 2937 | 241 | AD | eGFR | -0.304843 | -0.78395 | 0.09927 | 0.0929291 | 0 | 0.61457 | 0.13536 | 285 | 0.522917052631579 |
| Chr1:193Mb | 147 | 1 | 192468374 | 193575516 | 1679 | 191 | AD | eGFR | -0.515924 | -1 | 0.18309 | 0.266178 | 0 | 1 | 0.135872 | 286 | 0.523059692307692 |
| Chr16:12.4Mb | 2116 | 16 | 11916632 | 12793149 | 2795 | 276 | AD | eGFR | -0.37119 | -1 | 0.14262 | 0.137782 | 0 | 1 | 0.138445 | 287 | 0.529948 |
| Chr16:28.2Mb | 2127 | 16 | 27443062 | 29043177 | 1883 | 269 | AD | eGFR | -0.256411 | -0.66862 | 0.09063 | 0.0657465 | 0 | 0.44705 | 0.138624 | 288 | 0.529948 |
| Chr14:104Mb | 2029 | 14 | 103617473 | 104439106 | 1874 | 172 | AD | eGFR | 0.41297 | -0.13347 | 1 | 0.170544 | 0 | 1 | 0.139855 | 289 | 0.532803996539792 |
| Chr8:9.5Mb | 1248 | 8 | 9167796 | 9835863 | 1645 | 180 | AD | eGFR | 0.239915 | -0.08144 | 0.61585 | 0.057559 | 0 | 0.37927 | 0.140564 | 290 | 0.533562698630137 |
| Chr9:112.5Mb | 1456 | 9 | 112076382 | 112865520 | 2070 | 235 | AD | eGFR | 0.322593 | -0.1121 | 0.8616 | 0.104066 | 0 | 0.74235 | 0.141184 | 291 | 0.533562698630137 |
| Chr8:72.5Mb | 1307 | 8 | 72013185 | 72917489 | 1916 | 270 | AD | eGFR | -0.315083 | -0.83773 | 0.09926 | 0.0992775 | 0 | 0.70179 | 0.141508 | 292 | 0.533562698630137 |
| Chr2:238.5Mb | 415 | 2 | 23818548 | 23881463 | 1763 | 218 | AD | eGFR | 0.444216 | -0.16056 | 1 | 0.197328 | 0 | 1 | 0.142344 | 293 | 0.534883085324232 |

| locus_label | locus | chr | start | stop | n.snps | n.pcs | phen1 | phen2 | rho | rho.lower | rho.upper | r2 | r2.lower | r2.upper | p | rank | FDR |
|---|---|---|---|---|---|---|---|---|---|---|---|---|---|---|---|---|---|
| | | | 3 | 0 | | | | | | | | | | | | | |
| Chr20:53.5Mb | 2417 | 20 | 53120335 | 53879803 | 1740 | 187 | AD | eGFR | -0.341712 | -0.94014 | 0.12339 | 0.116767 | 0 | 0.88469 | 0.142851 | 294 | 0.534962418367347 |
| Chr1:162.5Mb | 119 | 1 | 161945443 | 163141862 | 2776 | 285 | AD | eGFR | -0.296736 | -0.77769 | 0.11421 | 0.0880523 | 0 | 0.6048 | 0.144024 | 295 | 0.537526861016949 |
| Chr8:27.9Mb | 1273 | 8 | 27406512 | 28344176 | 2302 | 253 | AD | eGFR | -0.300288 | -0.93663 | 0.11954 | 0.0901731 | 0 | 0.87803 | 0.14517 | 296 | 0.538522454545455 |
| Chr1:117.6Mb | 103 | 1 | 117046312 | 118118667 | 1875 | 248 | AD | eGFR | 0.311585 | -0.11845 | 0.82115 | 0.0970853 | 0 | 0.67428 | 0.145269 | 297 | 0.538522454545455 |
| Chr4:100.7Mb | 690 | 4 | 100333335 | 101132995 | 1812 | 159 | AD | eGFR | 0.420691 | -0.16868 | 1 | 0.176981 | 0 | 1 | 0.147085 | 298 | 0.542343762541806 |
| Chr12:117.7Mb | 1846 | 12 | 117091844 | 118256124 | 2529 | 299 | AD | eGFR | -0.333951 | -0.90911 | 0.11877 | 0.111523 | 0 | 0.83011 | 0.147285 | 299 | 0.542343762541806 |
| Chr4:107.3Mb | 695 | 4 | 106479156 | 108131206 | 2470 | 240 | AD | eGFR | 0.43758 | -0.1895 | 1 | 0.191476 | 0 | 1 | 0.148804 | 300 | 0.54492550166113 |
| Chr6:9.7Mb | 934 | 6 | 9020711 | 10416550 | 3195 | 283 | AD | eGFR | 0.308281 | -0.12896 | 0.8538 | 0.0950369 | 0 | 0.72974 | 0.148976 | 301 | 0.54492550166113 |
| Chr5:131.6Mb | 887 | 5 | 130573818 | 132554690 | 2673 | 202 | AD | eGFR | 0.218588 | -0.08459 | 0.58183 | 0.0477806 | 0 | 0.33901 | 0.150773 | 302 | 0.547671444078947 |
| Chr15:75.4Mb | 2076 | 15 | 74458114 | 76401952 | 2661 | 248 | AD | eGFR | -0.175065 | -0.45599 | 0.06718 | 0.0306478 | 0 | 0.20793 | 0.151074 | 303 | 0.547671444078947 |
| Chr1:18Mb | 19 | 1 | 17557747 | 18427820 | 2180 | 273 | AD | eGFR | 0.470029 | -0.19869 | 1 | 0.220927 | 0 | 1 | 0.151219 | 304 | 0.547671444078947 |
| Chr19:53.1Mb | 2360 | 19 | 52678749 | 53431217 | 2380 | 281 | AD | eGFR | -0.223365 | -0.55513 | 0.08295 | 0.0498917 | 0 | 0.30817 | 0.151782 | 305 | 0.547808666666667 |
| Chr16:4.1Mb | 2105 | 16 | 3379997 | 4816145 | 2970 | 329 | AD | eGFR | -0.212203 | -0.56435 | 0.07429 | 0.04503 | 0 | 0.31965 | 0.152252 | 306 | 0.547808666666667 |
| Chr17:46.7Mb | 2209 | 17 | 45883902 | 47516224 | 3243 | 245 | AD | eGFR | -0.215376 | -0.55627 | 0.0787 | 0.046387 | 0 | 0.30944 | 0.153205 | 307 | 0.549442035830619 |
| Chr5:118Mb | 874 | 5 | 117348498 | 118605251 | 2782 | 242 | AD | eGFR | -0.505203 | -1 | 0.21005 | 0.25523 | 0 | 1 | 0.154141 | 308 | 0.550532067961165 |
| Chr16:0.6Mb | 2102 | 16 | 60169 | 1049079 | 2359 | 295 | AD | eGFR | -0.23326 | -0.63301 | 0.08942 | 0.05441 | 0 | 0.4007 | 0.154509 | 309 | 0.550532067961165 |
| Chr8:142.1Mb | 1366 | 8 | 141527882 | 142611889 | 2434 | 283 | AD | eGFR | -0.341297 | -1 | 0.15714 | 0.116483 | 0 | 1 | 0.156308 | 310 | 0.553494439490446 |
| Chr7:74.2Mb | 1166 | 7 | 73128831 | 75243899 | 1639 | 261 | AD | eGFR | 0.253487 | -0.1117 | 0.68443 | 0.0642556 | 0 | 0.46899 | 0.156636 | 311 | 0.553494439490446 |
| Chr1:207Mb | 159 | 1 | 205917549 | 208162951 | 2953 | 280 | AD | eGFR | 0.175899 | -0.06867 | 0.42871 | 0.0309405 | 0 | 0.1838 | 0.157318 | 312 | 0.553494439490446 |
| Chr18:9.3Mb | 2247 | 18 | 8788525 | 9816164 | 2964 | 298 | AD | eGFR | 0.431327 | -0.18842 | 1 | 0.186043 | 0 | 1 | 0.157687 | 313 | 0.553494439490446 |
| Chr1:222.3Mb | 172 | 1 | 221856865 | 222763597 | 1646 | 200 | AD | eGFR | -0.361114 | -1 | 0.14644 | 0.130404 | 0 | 1 | 0.157854 | 314 | 0.553494439490446 |
| Chr7:0.3Mb | 1094 | 7 | 15064 | 617319 | 801 | 165 | AD | eGFR | 0.267673 | -0.10744 | 0.68098 | 0.0716489 | 0 | 0.46373 | 0.158871 | 315 | 0.553826787037037 |
| Chr5:159.1Mb | 908 | 5 | 158484776 | 159663733 | 2632 | 275 | AD | eGFR | -0.333032 | -0.92728 | 0.1453 | 0.11091 | 0 | 0.85998 | 0.159027 | 316 | 0.553826787037037 |
| Chr17:30.6Mb | 2197 | 17 | 29783142 | 31318753 | 2782 | 287 | AD | eGFR | 0.363639 | -0.15897 | 1 | 0.132233 | 0 | 1 | 0.160634 | 317 | 0.553826787037037 |
| Chr10:76.4Mb | 1556 | 10 | 75694024 | 77144961 | 1973 | 179 | AD | eGFR | 0.257052 | -0.10323 | 0.65573 | 0.0660755 | 0 | 0.42998 | 0.161003 | 318 | 0.553826787037037 |
| Chr11:11.4Mb | 1623 | 11 | 11081300 | 11632650 | 1858 | 283 | AD | eGFR | -0.327584 | -0.9753 | 0.12757 | 0.107311 | 0 | 0.95122 | 0.161453 | 319 | 0.553826787037037 |
| Chr4:110.9Mb | 698 | 4 | 109978983 | 111733579 | 3341 | 301 | AD | eGFR | -0.293678 | -0.79019 | 0.13452 | 0.0862466 | 0 | 0.62441 | 0.16244 | 320 | 0.553826787037037 |
| Chr15:98.4Mb | 2097 | 15 | 98025685 | 98828261 | 2477 | 284 | AD | eGFR | 0.315268 | -0.13941 | 0.97273 | 0.099394 | 0 | 0.95074 | 0.162453 | 321 | 0.553826787037037 |
| Chr6:39.2Mb | 974 | 6 | 38688721 | 39785907 | 2993 | 263 | AD | eGFR | -0.349294 | -1 | 0.16392 | 0.122007 | 0 | 1 | 0.162484 | 322 | 0.553826787037037 |
| Chr15:102Mb | 2101 | 15 | 101550751 | 102520975 | 2145 | 310 | AD | eGFR | -0.404414 | -1 | 0.19023 | 0.163551 | 0 | 1 | 0.162712 | 323 | 0.553826787037037 |
| Chr11:18.2Mb | 1629 | 11 | 17583949 | 18740267 | 2452 | 247 | AD | eGFR | -0.248494 | -0.64629 | 0.10033 | 0.0617491 | 0 | 0.41946 | 0.162979 | 324 | 0.553826787037037 |
| Chr12:51.1Mb | 1789 | 12 | 50344347 | 51769419 | 2211 | 155 | AD | eGFR | -0.425173 | -1 | 0.20186 | 0.180772 | 0 | 1 | 0.163722 | 325 | 0.55463976 |

| locus_label | locus | chr | start | stop | n.snps | n.pcs | phen1 | phen2 | rho | rho.lower | rho.upper | r2 | r2.lower | r2.upper | p | rank | FDR |
|---|---|---|---|---|---|---|---|---|---|---|---|---|---|---|---|---|---|
| Chr12:89.5Mb | 1821 | 12 | 88344765 | 90567001 | 2946 | 248 | AD | eGFR | 0.226675 | -0.09215 | 0.5681 | 0.0513814 | 0 | 0.32273 | 0.164317 | 326 | 0.554947904907975 |
| Chr14:54.8Mb | 1986 | 14 | 54359599 | 55292832 | 1760 | 231 | AD | eGFR | 0.204669 | -0.08238 | 0.51706 | 0.0418893 | 0 | 0.26735 | 0.16556 | 327 | 0.557435963302752 |
| Chr20:49.9Mb | 2413 | 20 | 49236419 | 50653620 | 3474 | 334 | AD | eGFR | -0.345089 | -1 | 0.17523 | 0.119086 | 0 | 1 | 0.166318 | 328 | 0.558280847560976 |
| Chr6:131Mb | 1058 | 6 | 130550138 | 131457164 | 2594 | 169 | AD | eGFR | -0.376874 | -1 | 0.16904 | 0.142034 | 0 | 1 | 0.1685 | 329 | 0.563886018237082 |
| Chr10:69.3Mb | 1550 | 10 | 68623275 | 69904170 | 2413 | 168 | AD | eGFR | -0.318369 | -0.88748 | 0.14493 | 0.101359 | 0 | 0.78809 | 0.171772 | 330 | 0.569818441791045 |
| Chr6:3.4Mb | 928 | 6 | 2746532 | 3964072 | 2921 | 339 | AD | eGFR | 0.319197 | -0.15545 | 0.97859 | 0.101887 | 0 | 0.9642 | 0.171856 | 331 | 0.569818441791045 |
| Chr1:3.9Mb | 5 | 1 | 3582048 | 4281476 | 1903 | 231 | AD | eGFR | -0.244072 | -0.64141 | 0.10718 | 0.0595711 | 0 | 0.41141 | 0.171921 | 332 | 0.569818441791045 |
| Chr2:220.2Mb | 396 | 2 | 219678784 | 220771552 | 1806 | 255 | AD | eGFR | 0.178028 | -0.07684 | 0.46794 | 0.0316941 | 0 | 0.21897 | 0.172909 | 333 | 0.569818441791045 |
| Chr4:69.2Mb | 664 | 4 | 68742860 | 69603368 | 1465 | 129 | AD | eGFR | 0.463999 | -0.23636 | 1 | 0.215295 | 0 | 1 | 0.172981 | 334 | 0.569818441791045 |
| Chr2:182.8Mb | 366 | 2 | 182307878 | 183352290 | 2036 | 199 | AD | eGFR | -0.318354 | -1 | 0.14841 | 0.10135 | 0 | 1 | 0.173378 | 335 | 0.569818441791045 |
| Chr11:9.7Mb | 1621 | 11 | 9080940 | 10370100 | 2707 | 217 | AD | eGFR | -0.354929 | -1 | 0.16937 | 0.125975 | 0 | 1 | 0.175845 | 336 | 0.57620638392857 |
| Chr2:111.4Mb | 305 | 2 | 110097869 | 112644828 | 2441 | 269 | AD | eGFR | -0.243359 | -0.62873 | 0.12151 | 0.0592238 | 0 | 0.3953 | 0.177443 | 337 | 0.579717338278932 |
| Chr12:65.3Mb | 1800 | 12 | 64403858 | 66114643 | 2322 | 270 | AD | eGFR | 0.393192 | -0.1789 | 1 | 0.1546 | 0 | 1 | 0.178542 | 338 | 0.581582076923077 |
| Chr11:86.2Mb | 1692 | 11 | 85869060 | 86629333 | 1959 | 198 | AD | eGFR | -0.35564 | -1 | 0.17099 | 0.12648 | 0 | 1 | 0.179223 | 339 | 0.582078238938053 |
| Chr15:50.9Mb | 2057 | 15 | 50195877 | 51605477 | 3136 | 196 | AD | eGFR | 0.298614 | -0.15478 | 0.83311 | 0.0891705 | 0 | 0.69759 | 0.180555 | 340 | 0.58269298245614 |
| Chr17:57.4Mb | 2217 | 17 | 56245228 | 58652809 | 2478 | 201 | AD | eGFR | -0.228763 | -0.65436 | 0.10476 | 0.0523327 | 0 | 0.42819 | 0.180558 | 341 | 0.58269298245614 |
| Chr13:95.5Mb | 1934 | 13 | 94925137 | 96132647 | 3498 | 319 | AD | eGFR | 0.268798 | -0.13919 | 0.78931 | 0.0722526 | 0 | 0.62301 | 0.181 | 342 | 0.58269298245614 |
| Chr5:81.1Mb | 842 | 5 | 80448226 | 81710194 | 1929 | 244 | AD | eGFR | -0.298966 | -0.84493 | 0.1574 | 0.0893808 | 0 | 0.71594 | 0.18316 | 343 | 0.586272953757225 |
| Chr2:2.3Mb | 204 | 2 | 1875061 | 2680063 | 1955 | 258 | AD | eGFR | 0.249566 | -0.12002 | 0.65681 | 0.0622834 | 0 | 0.43139 | 0.183749 | 344 | 0.586272953757225 |
| Chr18:75.7Mb | 2303 | 18 | 75240761 | 76231685 | 3079 | 319 | AD | eGFR | 0.4906 | -0.26897 | 1 | 0.240688 | 0 | 1 | 0.184103 | 345 | 0.586272953757225 |
| Chr2:235.3Mb | 411 | 2 | 234945578 | 235625789 | 2066 | 269 | AD | eGFR | -0.340479 | -1 | 0.17379 | 0.115926 | 0 | 1 | 0.184242 | 346 | 0.586272953757225 |
| Chr7:107Mb | 1192 | 7 | 106399740 | 107658603 | 2243 | 258 | AD | eGFR | -0.335257 | -1 | 0.1727 | 0.112397 | 0 | 1 | 0.185635 | 347 | 0.587908834285714 |
| Chr6:53.9Mb | 988 | 6 | 53425493 | 54452767 | 2289 | 196 | AD | eGFR | -0.350587 | -1 | 0.17511 | 0.122911 | 0 | 1 | 0.186401 | 348 | 0.587908834285714 |
| Chr1:116.6Mb | 102 | 1 | 116207298 | 117046311 | 1761 | 203 | AD | eGFR | 0.272577 | -0.14554 | 0.74304 | 0.0742984 | 0 | 0.55211 | 0.186891 | 349 | 0.587908834285714 |
| Chr6:170.4Mb | 1093 | 6 | 169667887 | 171053405 | 3052 | 241 | AD | eGFR | 0.329275 | -0.17346 | 1 | 0.108422 | 0 | 1 | 0.186892 | 350 | 0.587908834285714 |
| Chr9:137.9Mb | 1481 | 9 | 137593528 | 138202112 | 2118 | 321 | AD | eGFR | 0.318606 | -0.17163 | 0.95405 | 0.10151 | 0 | 0.91354 | 0.189042 | 351 | 0.589815872282609 |
| Chr22:25.7Mb | 2470 | 22 | 25282437 | 26166934 | 1612 | 245 | AD | eGFR | 0.222692 | -0.11567 | 0.60191 | 0.0495919 | 0 | 0.3623 | 0.190409 | 352 | 0.589815872282609 |
| Chr7:139.5Mb | 1215 | 7 | 138755108 | 140217630 | 2885 | 315 | AD | eGFR | 0.247272 | -0.13569 | 0.70393 | 0.0611434 | 0 | 0.4956 | 0.190731 | 353 | 0.589815872282609 |
| Chr1:55.2Mb | 47 | 1 | 54771294 | 55727134 | 2505 | 288 | AD | eGFR | -0.197938 | -0.52636 | 0.09852 | 0.0391795 | 0 | 0.27706 | 0.190881 | 354 | 0.589815872282609 |
| Chr2:197.3Mb | 377 | 2 | 196446300 | 198140350 | 3095 | 217 | AD | eGFR | -0.272173 | -0.77197 | 0.13897 | 0.0740781 | 0 | 0.59651 | 0.191188 | 355 | 0.589815872282609 |
| Chr4:1.2Mb | 597 | 4 | 812416 | 1529267 | 1897 | 184 | AD | eGFR | -0.230809 | -0.60712 | 0.13155 | 0.0532729 | 0 | 0.36908 | 0.191595 | 356 | 0.589815872282609 |
| Chr1:2.6Mb | 3 | 1 | 2215496 | 2983519 | 1608 | 252 | AD | eGFR | -0.213989 | -0.55934 | 0.11416 | 0.0457914 | 0 | 0.31286 | 0.191689 | 357 | 0.589815872282609 |
| Chr15:62.7Mb | 2067 | 15 | 62106139 | 63218978 | 2388 | 293 | AD | eGFR | 0.211839 | -0.11473 | 0.55897 | 0.0448759 | 0 | 0.31245 | 0.193809 | 358 | 0.589815872282609 |
| Chr2:5.2Mb | 208 | 2 | 4766970 | 5568653 | 1685 | 209 | AD | eGFR | -0.475244 | -1 | 0.29783 | 0.225857 | 0 | 1 | 0.194216 | 359 | 0.589815872282609 |
| Chr1:94.6Mb | 82 | 1 | 94168577 | 95122541 | 1855 | 215 | AD | eGFR | -0.338896 | -1 | 0.19253 | 0.11485 | 0 | 1 | 0.194749 | 360 | 0.589815872282609 |

| locus_label | locus | chr | start | stop | n.snps | n.pcs | phen1 | phen2 | rho | rho.lower | rho.upper | r2 | r2.lower | r2.upper | p | rank | FDR |
|---|---|---|---|---|---|---|---|---|---|---|---|---|---|---|---|---|---|
| Chr8:31.8Mb | 1276 | 8 | 31134787 | 32454962 | 3340 | 230 | AD | eGFR | -0.227174 | -0.62705 | 0.11954 | 0.051608 | 0 | 0.39319 | 0.194795 | 361 | 0.589815872282609 |
| Chr12:22.3Mb | 1762 | 12 | 21572253 | 22928055 | 2536 | 252 | AD | eGFR | 0.390042 | -0.23772 | 1 | 0.152133 | 0 | 1 | 0.194894 | 362 | 0.589815872282609 |
| Chr21:38.6Mb | 2452 | 21 | 38046450 | 39252059 | 2856 | 274 | AD | eGFR | 0.23619 | -0.14443 | 0.64188 | 0.0557856 | 0 | 0.41201 | 0.195374 | 363 | 0.589815872282609 |
| Chr10:74.6Mb | 1555 | 10 | 73514215 | 75694023 | 2807 | 191 | AD | eGFR | -0.392886 | -1 | 0.22492 | 0.15436 | 0 | 1 | 0.195963 | 364 | 0.589815872282609 |
| Chr2:105.8Mb | 301 | 2 | 105120579 | 106546524 | 3324 | 307 | AD | eGFR | 0.221878 | -0.1144 | 0.61177 | 0.0492297 | 0 | 0.37426 | 0.196101 | 365 | 0.589815872282609 |
| Chr9:132.6Mb | 1475 | 9 | 132193799 | 132999452 | 2403 | 250 | AD | eGFR | -0.179265 | -0.4694 | 0.09225 | 0.0321358 | 0 | 0.22034 | 0.196848 | 366 | 0.589815872282609 |
| Chr14:35Mb | 1967 | 14 | 34695196 | 35379988 | 1759 | 189 | AD | eGFR | 0.328554 | -0.19018 | 1 | 0.107948 | 0 | 1 | 0.196941 | 367 | 0.589815872282609 |
| Chr1:213.2Mb | 163 | 1 | 212347583 | 213958292 | 3500 | 295 | AD | eGFR | 0.268288 | -0.14742 | 0.76741 | 0.0719786 | 0 | 0.59081 | 0.197141 | 368 | 0.589815872282609 |
| Chr8:145.6Mb | 1369 | 8 | 144934289 | 146303787 | 1443 | 232 | AD | eGFR | 0.139863 | -0.07584 | 0.35867 | 0.0195616 | 0 | 0.12865 | 0.198287 | 369 | 0.591011458221024 |
| Chr7:59.7Mb | 1155 | 7 | 57383658 | 61969717 | 598 | 49 | AD | eGFR | 0.372334 | -0.22825 | 1 | 0.138633 | 0 | 1 | 0.199087 | 370 | 0.591011458221024 |
| Chr7:131.1Mb | 1209 | 7 | 130418705 | 131856481 | 2918 | 344 | AD | eGFR | -0.242136 | -0.67387 | 0.12998 | 0.0586299 | 0 | 0.4541 | 0.199151 | 371 | 0.591011458221024 |
| Chr12:94.8Mb | 1825 | 12 | 94385885 | 95147845 | 2070 | 283 | AD | eGFR | 0.264306 | -0.15085 | 0.7528 | 0.0698578 | 0 | 0.56679 | 0.200582 | 372 | 0.59229372386059 |
| Chr12:81.4Mb | 1814 | 12 | 80952473 | 81843890 | 1760 | 150 | AD | eGFR | -0.568121 | -1 | 0.36697 | 0.322762 | 0 | 1 | 0.200659 | 373 | 0.59229372386059 |
| Chr18:56.3Mb | 2284 | 18 | 55809316 | 56832312 | 2485 | 279 | AD | eGFR | -0.481625 | -1 | 0.29944 | 0.231963 | 0 | 1 | 0.204221 | 374 | 0.599762468253968 |
| Chr1:1.8Mb | 2 | 1 | 1376205 | 2215495 | 1054 | 172 | AD | eGFR | -0.22714 | -0.73196 | 0.13385 | 0.0515926 | 0 | 0.53676 | 0.204618 | 375 | 0.599762468253968 |
| Chr11:7.6Mb | 1619 | 11 | 7191231 | 7915071 | 2070 | 240 | AD | eGFR | 0.579358 | -0.40528 | 1 | 0.335656 | 0 | 1 | 0.204992 | 376 | 0.599762468253968 |
| Chr2:177.4Mb | 361 | 2 | 176734751 | 178149981 | 2892 | 299 | AD | eGFR | -0.163279 | -0.48657 | 0.09642 | 0.02666 | 0 | 0.2373 | 0.205413 | 377 | 0.599762468253968 |
| Chr22:42.3Mb | 2485 | 22 | 41722788 | 42867927 | 1699 | 172 | AD | eGFR | -0.202432 | -0.57624 | 0.12446 | 0.0409786 | 0 | 0.33277 | 0.205913 | 378 | 0.599762468253968 |
| Chr11:48.4Mb | 1659 | 11 | 48066367 | 48765366 | 1346 | 35 | AD | eGFR | 0.309951 | -0.19116 | 0.91795 | 0.0960695 | 0 | 0.84468 | 0.206834 | 379 | 0.600855498680739 |
| Chr6:12.6Mb | 936 | 6 | 11790672 | 13352326 | 3363 | 314 | AD | eGFR | 0.246194 | -0.1295 | 0.6959 | 0.0606114 | 0 | 0.48434 | 0.207755 | 380 | 0.601336724409449 |
| Chr2:40.4Mb | 244 | 2 | 40087419 | 40786176 | 1794 | 233 | AD | eGFR | -0.259452 | -0.71855 | 0.16219 | 0.0673156 | 0 | 0.51632 | 0.208092 | 381 | 0.601336724409449 |
| Chr15:71.4Mb | 2073 | 15 | 70767984 | 72058129 | 2397 | 272 | AD | eGFR | 0.277769 | -0.16739 | 0.8208 | 0.0771554 | 0 | 0.67738 | 0.210545 | 382 | 0.60490775 |
| Chr17:73.1Mb | 2229 | 17 | 72383473 | 73741321 | 2768 | 293 | AD | eGFR | 0.198719 | -0.11286 | 0.54285 | 0.0394892 | 0 | 0.29468 | 0.210959 | 383 | 0.60490775 |
| Chr4:48.4Mb | 648 | 4 | 47408266 | 49325138 | 2945 | 222 | AD | eGFR | 0.337128 | -0.22521 | 1 | 0.113655 | 0 | 1 | 0.210976 | 384 | 0.60490775 |
| Chr18:3.3Mb | 2241 | 18 | 2839843 | 3722828 | 2303 | 332 | AD | eGFR | 0.210045 | -0.11757 | 0.58726 | 0.0441187 | 0 | 0.34487 | 0.212366 | 385 | 0.605860100775194 |
| Chr10:114.9Mb | 1589 | 10 | 114255955 | 115588903 | 2436 | 312 | AD | eGFR | 0.326467 | -0.21788 | 1 | 0.106581 | 0 | 1 | 0.212823 | 386 | 0.605860100775194 |
| Chr11:2.2Mb | 1612 | 11 | 1857846 | 2477449 | 1669 | 253 | AD | eGFR | 0.133776 | -0.08076 | 0.35294 | 0.0178961 | 0 | 0.12457 | 0.212959 | 387 | 0.605860100775194 |
| Chr1:133.1Mb | 106 | 1 | 120345067 | 145803848 | 257 | 51 | AD | eGFR | 0.309851 | -0.19907 | 0.92987 | 0.0960074 | 0 | 0.8681 | 0.213686 | 388 | 0.606053799485861 |
| Chr4:75.7Mb | 669 | 4 | 74885901 | 76497358 | 2990 | 256 | AD | eGFR | 0.306447 | -0.18433 | 1 | 0.0939096 | 0 | 1 | 0.214128 | 389 | 0.606053799485861 |
| Chr6:166.3Mb | 1089 | 6 | 165835954 | 166853325 | 2913 | 300 | AD | eGFR | -0.223446 | -0.61642 | 0.13297 | 0.0499281 | 0 | 0.37998 | 0.21554 | 390 | 0.608486 |
| Chr2:216.7Mb | 393 | 2 | 215899571 | 217566011 | 3328 | 340 | AD | eGFR | -0.219337 | -0.6231 | 0.13718 | 0.0481089 | 0 | 0.38852 | 0.216761 | 391 | 0.609191984732824 |
| Chr19:56.8Mb | 2366 | 19 | 56411087 | 57143509 | 1998 | 293 | AD | eGFR | -0.273484 | -0.95123 | 0.18481 | 0.0747936 | 0 | 0.90882 | 0.217213 | 392 | 0.609191984732824 |
| Chr1:78.5Mb | 67 | 1 | 77895395 | 79065286 | 2011 | 173 | AD | eGFR | 0.369632 | -0.24334 | 1 | 0.136627 | 0 | 1 | 0.21745 | 393 | 0.609191984732824 |
| Chr3:169.6Mb | 570 | 3 | 168972663 | 170174578 | 2282 | 242 | AD | eGFR | -0.204256 | -0.58878 | 0.13461 | 0.0417207 | 0 | 0.34682 | 0.218182 | 394 | 0.609585192118227 |
| Chr14:81.8Mb | 2010 | 14 | 81001921 | 82524785 | 3308 | 267 | AD | eGFR | -0.273557 | -0.83578 | 0.1765 | 0.0748334 | 0 | 0.69954 | 0.21957 | 395 | 0.609585192118227 |

| locus_label | locus | chr | start | stop | n.snps | n.pcs | phen1 | phen2 | rho | rho.lower | rho.upper | r2 | r2.lower | r2.upper | p | rank | FDR |
|---|---|---|---|---|---|---|---|---|---|---|---|---|---|---|---|---|---|
| Chr22:46.8Mb | 2490 | 22 | 46351049 | 47299625 | 2370 | 294 | AD | eGFR | 0.191722 | -0.11676 | 0.52528 | 0.0367575 | 0 | 0.27592 | 0.21983 | 396 | 0.609585192118227 |
| Chr11:125.8Mb | 1730 | 11 | 125266685 | 126304240 | 2541 | 237 | AD | eGFR | -0.287047 | -0.87062 | 0.18688 | 0.082396 | 0 | 0.75798 | 0.220025 | 397 | 0.609585192118227 |
| Chr18:76.7Mb | 2304 | 18 | 76231686 | 77148137 | 2656 | 248 | AD | eGFR | -0.238392 | -0.69029 | 0.15668 | 0.0568309 | 0 | 0.4765 | 0.22083 | 398 | 0.609585192118227 |
| Chr1:23.3Mb | 25 | 1 | 22740288 | 23907045 | 1739 | 251 | AD | eGFR | 0.165578 | -0.10129 | 0.45114 | 0.0274161 | 0 | 0.20353 | 0.221659 | 399 | 0.609585192118227 |
| Chr8:12.6Mb | 1252 | 8 | 12296850 | 12947935 | 1370 | 177 | AD | eGFR | 0.451319 | -0.3482 | 1 | 0.203689 | 0 | 1 | 0.223654 | 400 | 0.609585192118227 |
| Chr14:70.5Mb | 2000 | 14 | 69794543 | 71140426 | 2872 | 257 | AD | eGFR | 0.508041 | -0.38167 | 1 | 0.258105 | 0 | 1 | 0.223866 | 401 | 0.609585192118227 |
| Chr5:147.9Mb | 897 | 5 | 147180172 | 148662214 | 3181 | 274 | AD | eGFR | 0.192446 | -0.11086 | 0.53491 | 0.0370353 | 0 | 0.28613 | 0.224208 | 402 | 0.609585192118227 |
| Chr1:159.4Mb | 116 | 1 | 158900370 | 159882515 | 2067 | 215 | AD | eGFR | 0.441778 | -0.34777 | 1 | 0.195168 | 0 | 1 | 0.224227 | 403 | 0.609585192118227 |
| Chr4:86.2Mb | 677 | 4 | 85437806 | 86931670 | 2862 | 192 | AD | eGFR | 0.308324 | -0.19333 | 1 | 0.0950636 | 0 | 1 | 0.224271 | 404 | 0.609585192118227 |
| Chr8:143.1Mb | 1367 | 8 | 142611890 | 143611461 | 2504 | 288 | AD | eGFR | -0.404032 | -1 | 0.29331 | 0.163242 | 0 | 1 | 0.224745 | 405 | 0.609585192118227 |
| Chr4:26.6Mb | 624 | 4 | 25832019 | 27385580 | 2702 | 327 | AD | eGFR | -0.288523 | -0.91441 | 0.19985 | 0.0832457 | 0 | 0.83645 | 0.224788 | 406 | 0.609585192118227 |
| Chr19:47.3Mb | 2353 | 19 | 46765061 | 47752727 | 1932 | 275 | AD | eGFR | -0.128271 | -0.34216 | 0.07867 | 0.0164535 | 0 | 0.11708 | 0.2255 | 407 | 0.610013513513514 |
| Chr11:70.5Mb | 1678 | 11 | 69713997 | 71242834 | 3472 | 322 | AD | eGFR | 0.310132 | -0.20992 | 1 | 0.0961817 | 0 | 1 | 0.226549 | 408 | 0.611349139705882 |
| Chr4:158.4Mb | 740 | 4 | 157597310 | 159176073 | 3182 | 238 | AD | eGFR | 0.332775 | -0.24858 | 1 | 0.110739 | 0 | 1 | 0.227321 | 409 | 0.611932569682152 |
| Chr13:26.3Mb | 1870 | 13 | 25977141 | 26681709 | 1635 | 203 | AD | eGFR | 0.258227 | -0.17944 | 0.76806 | 0.0666809 | 0 | 0.59271 | 0.228316 | 410 | 0.613111990243902 |
| Chr5:140.3Mb | 892 | 5 | 139408117 | 141253830 | 2853 | 269 | AD | eGFR | -0.270019 | -0.83404 | 0.18446 | 0.0729102 | 0 | 0.696 | 0.228919 | 411 | 0.613158123786408 |
| Chr9:101Mb | 1445 | 9 | 100310518 | 101596750 | 2819 | 288 | AD | eGFR | -0.193975 | -0.55443 | 0.12625 | 0.0376263 | 0 | 0.30739 | 0.229447 | 412 | 0.613158123786408 |
| Chr18:14Mb | 2252 | 18 | 13641480 | 14439251 | 1447 | 153 | AD | eGFR | -0.371651 | -1 | 0.29129 | 0.138125 | 0 | 1 | 0.230225 | 413 | 0.613747518159806 |
| Chr1:178.3Mb | 134 | 1 | 177746473 | 178915113 | 2106 | 183 | AD | eGFR | -0.479465 | -1 | 0.39472 | 0.229887 | 0 | 1 | 0.231401 | 414 | 0.614434937349398 |
| Chr19:46.3Mb | 2352 | 19 | 45893308 | 46765060 | 1986 | 241 | AD | eGFR | -0.129236 | -0.34543 | 0.08435 | 0.016702 | 0 | 0.11932 | 0.231599 | 415 | 0.614434937349398 |
| Chr16:54.1Mb | 2135 | 16 | 53393883 | 54866095 | 3199 | 351 | AD | eGFR | -0.213379 | -0.60107 | 0.1389 | 0.0455305 | 0 | 0.36129 | 0.232213 | 416 | 0.614582963942308 |
| Chr16:11.5Mb | 2115 | 16 | 11022543 | 11916631 | 2416 | 275 | AD | eGFR | 0.253603 | -0.17756 | 0.76499 | 0.0643146 | 0 | 0.58803 | 0.232978 | 417 | 0.615128964028777 |
| Chr18:11.3Mb | 2249 | 18 | 10675549 | 11872226 | 2841 | 311 | AD | eGFR | -0.288297 | -0.99475 | 0.20325 | 0.0831154 | 0 | 0.99533 | 0.233555 | 418 | 0.61517716507177 |
| Chr9:111Mb | 1454 | 9 | 110630151 | 111368107 | 1840 | 207 | AD | eGFR | 0.346436 | -0.26196 | 1 | 0.120018 | 0 | 1 | 0.23437 | 419 | 0.615243474056604 |
| Chr19:42.5Mb | 2348 | 19 | 41733464 | 43317282 | 1677 | 219 | AD | eGFR | 0.206916 | -0.14483 | 0.58443 | 0.0428144 | 0 | 0.34242 | 0.235125 | 420 | 0.615243474056604 |
| Chr7:155.8Mb | 1228 | 7 | 155280611 | 156344386 | 3072 | 352 | AD | eGFR | 0.189995 | -0.13584 | 0.69653 | 0.0360982 | 0 | 0.49041 | 0.236071 | 421 | 0.615243474056604 |
| Chr9:13.1Mb | 1387 | 9 | 12664859 | 13512852 | 1595 | 263 | AD | eGFR | 0.234628 | -0.16526 | 0.69997 | 0.0550501 | 0 | 0.49403 | 0.236269 | 422 | 0.615243474056604 |
| Chr4:40.2Mb | 640 | 4 | 39820062 | 40590541 | 1861 | 257 | AD | eGFR | 0.177203 | -0.11997 | 0.49443 | 0.0314009 | 0 | 0.24446 | 0.23663 | 423 | 0.615243474056604 |
| Chr8:60Mb | 1296 | 8 | 59555978 | 60476248 | 1637 | 221 | AD | eGFR | 0.299142 | -0.20681 | 0.97168 | 0.0894859 | 0 | 0.94951 | 0.236933 | 424 | 0.615243474056604 |
| Chr19:2.7Mb | 2309 | 19 | 2253175 | 3085446 | 2370 | 322 | AD | eGFR | -0.158872 | -0.43116 | 0.10758 | 0.0252403 | 0 | 0.1859 | 0.239296 | 425 | 0.617168244755245 |
| Chr11:77.1Mb | 1683 | 11 | 76518907 | 77719870 | 2210 | 189 | AD | eGFR | -0.194191 | -0.53584 | 0.13226 | 0.0377103 | 0 | 0.28713 | 0.239822 | 426 | 0.617168244755245 |
| Chr15:91.1Mb | 2090 | 15 | 90632719 | 91593623 | 2212 | 240 | AD | eGFR | -0.281485 | -0.88675 | 0.20127 | 0.0792337 | 0 | 0.79689 | 0.240179 | 427 | 0.617168244755245 |
| Chr8:6.3Mb | 1244 | 8 | 6019164 | 6663427 | 2818 | 313 | AD | eGFR | -0.288405 | -1 | 0.20325 | 0.0831773 | 0 | 1 | 0.240427 | 428 | 0.617168244755245 |
| Chr17:77Mb | 2233 | 17 | 76596288 | 77412786 | 2426 | 336 | AD | eGFR | -0.257348 | -0.79369 | 0.19971 | 0.066228 | 0 | 0.63112 | 0.240477 | 429 | 0.617168244755245 |
| Chr2:164.2Mb | 348 | 2 | 163607636 | 164702312 | 2015 | 201 | AD | eGFR | -0.270523 | -0.86135 | 0.19868 | 0.0731828 | 0 | 0.74788 | 0.24271 | 430 | 0.621450488372093 |

| locus_label | locus | chr | start | stop | n.snps | n.pcs | phen1 | phen2 | rho | rho.lower | rho.upper | r2 | r2.lower | r2.upper | p | rank | FDR |
|---|---|---|---|---|---|---|---|---|---|---|---|---|---|---|---|---|---|
| Chr16:1.5Mb | 2103 | 16 | 1049080 | 1996492 | 2779 | 279 | AD | eGFR | -0.168088 | -0.48343 | 0.11671 | 0.0282537 | 0 | 0.2339 | 0.24374 | 431 | 0.622082840277778 |
| Chr5:69.7Mb | 834 | 5 | 68006994 | 71468651 | 2872 | 258 | AD | eGFR | 0.18587 | -0.13813 | 0.58067 | 0.0345478 | 0 | 0.33901 | 0.244087 | 432 | 0.622082840277778 |
| Chr7:95.9Mb | 1182 | 7 | 95174848 | 96564981 | 2598 | 238 | AD | eGFR | 0.384369 | -0.31564 | 1 | 0.147739 | 0 | 1 | 0.246336 | 433 | 0.623372123287671 |
| Chr19:54.9Mb | 2363 | 19 | 54602371 | 55182973 | 1682 | 282 | AD | eGFR | 0.2032 | -0.13844 | 0.58628 | 0.0412904 | 0 | 0.34373 | 0.246568 | 434 | 0.623372123287671 |
| Chr16:79.3Mb | 2159 | 16 | 78973581 | 79662634 | 2307 | 353 | AD | eGFR | -0.142723 | -0.39783 | 0.09851 | 0.0203698 | 0 | 0.15827 | 0.247206 | 435 | 0.623372123287671 |
| Chr2:136.1Mb | 327 | 2 | 135160198 | 137061003 | 2692 | 162 | AD | eGFR | 0.274864 | -0.21278 | 0.96054 | 0.07555 | 0 | 0.92684 | 0.247479 | 436 | 0.623372123287671 |
| Chr2:190.5Mb | 373 | 2 | 189882703 | 191051954 | 2074 | 170 | AD | eGFR | 0.224287 | -0.17219 | 0.65413 | 0.0503049 | 0 | 0.42973 | 0.247632 | 437 | 0.623372123287671 |
| Chr7:101.4Mb | 1187 | 7 | 100849308 | 101950748 | 2864 | 253 | AD | eGFR | 0.21977 | -0.17419 | 0.69962 | 0.0482987 | 0 | 0.49506 | 0.24799 | 438 | 0.623372123287671 |
| Chr6:151.1Mb | 1073 | 6 | 150635316 | 151629954 | 3059 | 311 | AD | eGFR | 0.266988 | -0.20919 | 0.92907 | 0.0712823 | 0 | 0.86838 | 0.250563 | 439 | 0.627417707482993 |
| Chr18:5.1Mb | 2243 | 18 | 4603962 | 5575812 | 2548 | 283 | AD | eGFR | 0.254106 | -0.19775 | 0.799 | 0.0645698 | 0 | 0.64399 | 0.251142 | 440 | 0.627417707482993 |
| Chr4:184.9Mb | 768 | 4 | 184338585 | 185483262 | 3298 | 357 | AD | eGFR | 0.287111 | -0.22926 | 1 | 0.0824326 | 0 | 1 | 0.251309 | 441 | 0.627417707482993 |
| Chr2:94.6Mb | 294 | 2 | 91860962 | 97339301 | 1113 | 132 | AD | eGFR | 0.265545 | -0.21545 | 0.85491 | 0.070514 | 0 | 0.73532 | 0.252003 | 442 | 0.627726929864253 |
| Chr8:108.2Mb | 1336 | 8 | 107787164 | 108647463 | 1766 | 176 | AD | eGFR | -0.290407 | -0.96351 | 0.23029 | 0.084336 | 0 | 0.93862 | 0.252892 | 443 | 0.62851939503386 |
| Chr7:150.4Mb | 1223 | 7 | 149843000 | 150897818 | 2445 | 240 | AD | eGFR | -0.153715 | -0.43027 | 0.11869 | 0.0236284 | 0 | 0.18538 | 0.254677 | 444 | 0.631530128378378 |
| Chr1:211.7Mb | 162 | 1 | 211082893 | 212347582 | 3327 | 227 | AD | eGFR | -0.384268 | -1 | 0.32764 | 0.147662 | 0 | 1 | 0.255593 | 445 | 0.632377287640449 |
| Chr8:98.8Mb | 1328 | 8 | 98132520 | 99388944 | 2660 | 278 | AD | eGFR | 0.283844 | -0.21931 | 0.95859 | 0.0805673 | 0 | 0.92076 | 0.256332 | 446 | 0.632783704035874 |
| Chr20:46.1Mb | 2409 | 20 | 45673604 | 46486132 | 1844 | 246 | AD | eGFR | 0.317376 | -0.27114 | 1 | 0.100727 | 0 | 1 | 0.258536 | 447 | 0.635414318485523 |
| Chr6:90.5Mb | 1022 | 6 | 89974268 | 91097024 | 1972 | 252 | AD | eGFR | -0.227866 | -0.75565 | 0.1754 | 0.051923 | 0 | 0.57188 | 0.259004 | 448 | 0.635414318485523 |
| Chr16:87.2Mb | 2169 | 16 | 86748868 | 87669168 | 2630 | 332 | AD | eGFR | -0.182318 | -0.52805 | 0.13446 | 0.03324 | 0 | 0.27891 | 0.259129 | 449 | 0.635414318485523 |
| Chr4:147.8Mb | 732 | 4 | 147161043 | 148462208 | 2463 | 193 | AD | eGFR | -0.372872 | -1 | 0.32344 | 0.139034 | 0 | 1 | 0.26062 | 450 | 0.63714359602649 |
| Chr2:104.2Mb | 300 | 2 | 103305258 | 105120578 | 2786 | 264 | AD | eGFR | 0.182903 | -0.13436 | 0.52917 | 0.0334535 | 0 | 0.28002 | 0.26133 | 451 | 0.63714359602649 |
| Chr12:56.2Mb | 1793 | 12 | 55416803 | 56987105 | 2167 | 237 | AD | eGFR | 0.242373 | -0.20494 | 0.88411 | 0.0587449 | 0 | 0.79016 | 0.261761 | 452 | 0.63714359602649 |
| Chr16:52.7Mb | 2134 | 16 | 52041337 | 53393882 | 2736 | 277 | AD | eGFR | -0.207381 | -0.61891 | 0.16451 | 0.0430069 | 0 | 0.38383 | 0.262149 | 453 | 0.63714359602649 |
| Chr15:52.4Mb | 2058 | 15 | 51605478 | 53263126 | 3146 | 259 | AD | eGFR | -0.212128 | -0.64397 | 0.17219 | 0.0449982 | 0 | 0.41609 | 0.263958 | 454 | 0.639095413186813 |
| Chr11:2.9Mb | 1613 | 11 | 2477450 | 3362508 | 1943 | 324 | AD | eGFR | 0.172657 | -0.13693 | 0.56851 | 0.0298103 | 0 | 0.32353 | 0.264113 | 455 | 0.639095413186813 |
| Chr2:66.6Mb | 272 | 2 | 65938003 | 67224027 | 2799 | 280 | AD | eGFR | -0.208884 | -0.62555 | 0.17489 | 0.0436325 | 0 | 0.39131 | 0.265835 | 456 | 0.639322700431034 |
| Chr9:1.5Mb | 1372 | 9 | 1103599 | 1874961 | 2496 | 278 | AD | eGFR | 0.387232 | -0.3562 | 1 | 0.149949 | 0 | 1 | 0.26655 | 457 | 0.639322700431034 |
| Chr7:1Mb | 1095 | 7 | 617320 | 1366972 | 2011 | 225 | AD | eGFR | -0.123314 | -0.3603 | 0.0964 | 0.0152062 | 0 | 0.13052 | 0.266858 | 458 | 0.639322700431034 |
| Chr6:160.1Mb | 1083 | 6 | 159604053 | 160583918 | 2325 | 234 | AD | eGFR | -0.173021 | -0.59789 | 0.14543 | 0.0299364 | 0 | 0.35951 | 0.268347 | 459 | 0.639322700431034 |
| Chr1:146.4Mb | 107 | 1 | 145803849 | 147083835 | 1572 | 126 | AD | eGFR | 0.281814 | -0.25049 | 0.92496 | 0.0794193 | 0 | 0.87097 | 0.268413 | 460 | 0.639322700431034 |
| Chr10:79.3Mb | 1558 | 10 | 78665482 | 79952996 | 2604 | 286 | AD | eGFR | 0.321828 | -0.2987 | 1 | 0.103574 | 0 | 1 | 0.268684 | 461 | 0.639322700431034 |
| Chr17:12.2Mb | 2185 | 17 | 11778165 | 12676353 | 1716 | 258 | AD | eGFR | -0.181474 | -0.53394 | 0.1434 | 0.0329327 | 0 | 0.28584 | 0.269015 | 462 | 0.639322700431034 |
| Chr17:5Mb | 2178 | 17 | 4463972 | 5573784 | 2619 | 256 | AD | eGFR | 0.219035 | -0.18235 | 0.66673 | 0.0479763 | 0 | 0.44453 | 0.269386 | 463 | 0.639322700431034 |
| Chr9:93.8Mb | 1438 | 9 | 93441051 | 94175374 | 1772 | 220 | AD | eGFR | 0.303514 | -0.26053 | 1 | 0.0921207 | 0 | 1 | 0.269433 | 464 | 0.639322700431034 |

| locus_label | locus | chr | start | stop | n.snps | n.pcs | phen1 | phen2 | rho | rho.lower | rho.upper | r2 | r2.lower | r2.upper | p | rank | FDR |
|---|---|---|---|---|---|---|---|---|---|---|---|---|---|---|---|---|---|
| Chr14:34.1Mb | 1966 | 14 | 33591114 | 34695195 | 2746 | 372 | AD | eGFR | -0.289575 | -1 | 0.24434 | 0.0838537 | 0 | 1 | 0.271555 | 465 | 0.641817128205128 |
| Chr20:39.3Mb | 2403 | 20 | 38427595 | 40272390 | 3356 | 304 | AD | eGFR | 0.213652 | -0.17758 | 0.67775 | 0.0456472 | 0 | 0.46041 | 0.271914 | 466 | 0.641817128205128 |
| Chr10:44.3Mb | 1529 | 10 | 43769843 | 44893846 | 3276 | 220 | AD | eGFR | -0.383174 | -1 | 0.38216 | 0.146822 | 0 | 1 | 0.272688 | 467 | 0.641817128205128 |
| Chr9:114.9Mb | 1459 | 9 | 114443476 | 115390112 | 2232 | 188 | AD | eGFR | 0.317197 | -0.28315 | 1 | 0.100614 | 0 | 1 | 0.272816 | 468 | 0.641817128205128 |
| Chr15:54.7Mb | 2060 | 15 | 54332800 | 55136717 | 2591 | 198 | AD | eGFR | 0.30117 | -0.28878 | 1 | 0.0907035 | 0 | 1 | 0.275157 | 469 | 0.645944257995736 |
| Chr3:196Mb | 593 | 3 | 195710291 | 196383152 | 1671 | 238 | AD | eGFR | -0.279103 | -1 | 0.26835 | 0.0778985 | 0 | 1 | 0.277074 | 470 | 0.648982229299363 |
| Chr3:125.7Mb | 531 | 3 | 125475723 | 125992650 | 1479 | 125 | AD | eGFR | -0.228931 | -0.76973 | 0.18559 | 0.0524092 | 0 | 0.60208 | 0.27763 | 471 | 0.648982229299363 |
| Chr18:13.2Mb | 2251 | 18 | 12735559 | 13641479 | 1978 | 266 | AD | eGFR | -0.214072 | -0.6743 | 0.18168 | 0.045827 | 0 | 0.45602 | 0.278672 | 472 | 0.65003786440678 |
| Chr19:20.1Mb | 2328 | 19 | 19749167 | 20485499 | 1832 | 161 | AD | eGFR | 0.198103 | -0.16862 | 0.5955 | 0.0392449 | 0 | 0.35774 | 0.280454 | 473 | 0.652811530655391 |
| Chr12:102.4Mb | 1832 | 12 | 101825062 | 102966633 | 2160 | 244 | AD | eGFR | 0.329281 | -0.32448 | 1 | 0.108426 | 0 | 1 | 0.28336 | 474 | 0.655555669456067 |
| Chr5:75.6Mb | 838 | 5 | 75239303 | 75959515 | 1675 | 237 | AD | eGFR | 0.330102 | -0.32851 | 1 | 0.108967 | 0 | 1 | 0.283747 | 475 | 0.655555669456067 |
| Chr1:114Mb | 100 | 1 | 113418038 | 114664387 | 2037 | 229 | AD | eGFR | 0.278292 | -0.26873 | 1 | 0.0774465 | 0 | 1 | 0.284321 | 476 | 0.655555669456067 |
| Chr6:100.3Mb | 1030 | 6 | 99678877 | 100878645 | 2559 | 231 | AD | eGFR | 0.242538 | -0.22171 | 0.79499 | 0.0588245 | 0 | 0.63652 | 0.284506 | 477 | 0.655555669456067 |
| Chr18:6.2Mb | 2244 | 18 | 5575813 | 6756496 | 2685 | 335 | AD | eGFR | -0.178667 | -0.54063 | 0.15114 | 0.031922 | 0 | 0.2931 | 0.28461 | 478 | 0.655555669456067 |
| Chr1:203.3Mb | 156 | 1 | 202583885 | 204092537 | 2965 | 327 | AD | eGFR | -0.25135 | -0.83269 | 0.22887 | 0.063177 | 0 | 0.69654 | 0.28546 | 479 | 0.656140835073069 |
| Chr17:26.6Mb | 2195 | 17 | 25872111 | 27344401 | 1834 | 258 | AD | eGFR | -0.227138 | -0.7227 | 0.21313 | 0.0515918 | 0 | 0.5223 | 0.286473 | 480 | 0.65709744375 |
| Chr11:123Mb | 1727 | 11 | 122589225 | 123398882 | 2175 | 293 | AD | eGFR | -0.207732 | -0.65291 | 0.18553 | 0.0431526 | 0 | 0.42725 | 0.288403 | 481 | 0.659058143153527 |
| Chr12:115.8Mb | 1844 | 12 | 115439452 | 116197675 | 1921 | 268 | AD | eGFR | -0.180457 | -0.56048 | 0.15586 | 0.0325646 | 0 | 0.31413 | 0.288525 | 482 | 0.659058143153527 |
| Chr6:55Mb | 989 | 6 | 54452768 | 55469137 | 3002 | 164 | AD | eGFR | 0.22526 | -0.20721 | 0.71869 | 0.0507422 | 0 | 0.51929 | 0.2899 | 483 | 0.659788975308642 |
| Chr1:166Mb | 123 | 1 | 165553813 | 166458031 | 1773 | 223 | AD | eGFR | -0.251225 | -0.83128 | 0.23496 | 0.0631138 | 0 | 0.69317 | 0.290605 | 484 | 0.659788975308642 |
| Chr7:151.6Mb | 1224 | 7 | 150897819 | 152253294 | 2445 | 337 | AD | eGFR | -0.0988845 | -0.29162 | 0.08506 | 0.00977815 | 0 | 0.08504 | 0.291034 | 485 | 0.659788975308642 |
| Chr5:121.5Mb | 878 | 5 | 120960738 | 122005291 | 1838 | 220 | AD | eGFR | -0.441898 | -1 | 0.50643 | 0.195274 | 0 | 1 | 0.291242 | 486 | 0.659788975308642 |
| Chr15:94.1Mb | 2093 | 15 | 93707011 | 94565521 | 2776 | 324 | AD | eGFR | 0.199606 | -0.18559 | 0.62971 | 0.0398426 | 0 | 0.397 | 0.294832 | 487 | 0.661825306451613 |
| Chr10:64.7Mb | 1545 | 10 | 64069688 | 65400431 | 2784 | 205 | AD | eGFR | -0.246431 | -0.83856 | 0.23403 | 0.0607284 | 0 | 0.70999 | 0.295051 | 488 | 0.661825306451613 |
| Chr11:25.3Mb | 1637 | 11 | 24892440 | 25662918 | 2111 | 137 | AD | eGFR | -0.373794 | -1 | 0.39698 | 0.139722 | 0 | 1 | 0.295175 | 489 | 0.661825306451613 |
| Chr14:105.7Mb | 2031 | 14 | 105091341 | 106347143 | 1609 | 197 | AD | eGFR | -0.13972 | -0.41424 | 0.13025 | 0.0195216 | 0 | 0.17276 | 0.295415 | 490 | 0.661825306451613 |
| Chr3:140.4Mb | 543 | 3 | 139831268 | 141034637 | 2516 | 270 | AD | eGFR | 0.327186 | -0.36098 | 1 | 0.107051 | 0 | 1 | 0.296458 | 491 | 0.661825306451613 |
| Chr3:138Mb | 541 | 3 | 137372142 | 138693846 | 1920 | 213 | AD | eGFR | -0.218365 | -0.69583 | 0.20539 | 0.0476831 | 0 | 0.48457 | 0.296512 | 492 | 0.661825306451613 |
| Chr7:134.4Mb | 1211 | 7 | 133807217 | 134943221 | 2574 | 208 | AD | eGFR | 0.273623 | -0.27741 | 1 | 0.0748693 | 0 | 1 | 0.296906 | 493 | 0.661825306451613 |
| Chr1:3.3Mb | 4 | 1 | 2983520 | 3582047 | 1640 | 292 | AD | eGFR | -0.233183 | -0.95463 | 0.23564 | 0.0543742 | 0 | 0.92626 | 0.297452 | 494 | 0.661825306451613 |
| Chr1:21Mb | 22 | 1 | 20556203 | 21416121 | 1845 | 191 | AD | eGFR | 0.272553 | -0.27029 | 0.95548 | 0.074285 | 0 | 0.92165 | 0.297843 | 495 | 0.661825306451613 |

| locus_label | locus | chr | start | stop | n.snps | n.pcs | phen1 | phen2 | rho | rho.lower | rho.upper | r2 | r2.lower | r2.upper | p | rank | FDR |
|---|---|---|---|---|---|---|---|---|---|---|---|---|---|---|---|---|---|
| Chr8:133Mb | 1357 | 8 | 132534637 | 133461551 | 2320 | 237 | AD | eGFR | -0.35305 | -1 | 0.38383 | 0.124645 | 0 | 1 | 0.298152 | 496 | 0.661825306451613 |
| Chr16:16.6Mb | 2119 | 16 | 15921109 | 17343277 | 2284 | 264 | AD | eGFR | 0.204457 | -0.1951 | 0.65397 | 0.0418027 | 0 | 0.42856 | 0.299536 | 497 | 0.663559629778672 |
| Chr4:22.1Mb | 620 | 4 | 21416381 | 22835185 | 3267 | 256 | AD | eGFR | -0.298076 | -1 | 0.32325 | 0.0888491 | 0 | 1 | 0.30042 | 498 | 0.66418156626506 |
| Chr3:136.3Mb | 540 | 3 | 135288395 | 137372141 | 2620 | 230 | AD | eGFR | -0.246997 | -0.95312 | 0.2634 | 0.0610073 | 0 | 0.93823 | 0.30165 | 499 | 0.664856466 |
| Chr14:24.4Mb | 1958 | 14 | 23985937 | 24906056 | 1727 | 255 | AD | eGFR | -0.233024 | -0.92355 | 0.22893 | 0.0543003 | 0 | 0.86184 | 0.301933 | 500 | 0.664856466 |
| Chr1:199.7Mb | 153 | 1 | 199263390 | 200134005 | 2343 | 169 | AD | eGFR | -0.26497 | -0.95822 | 0.27686 | 0.0702091 | 0 | 0.91818 | 0.303012 | 501 | 0.665359302788845 |
| Chr2:143.9Mb | 334 | 2 | 143259781 | 144503688 | 2156 | 221 | AD | eGFR | -0.129562 | -0.38807 | 0.12324 | 0.0167862 | 0 | 0.15085 | 0.30337 | 502 | 0.665359302788845 |
| Chr17:77.9Mb | 2234 | 17 | 77412787 | 78438332 | 2291 | 322 | AD | eGFR | 0.167395 | -0.16035 | 0.52528 | 0.0280209 | 0 | 0.27657 | 0.308574 | 503 | 0.675427383697813 |
| Chr4:8.6Mb | 606 | 4 | 8309827 | 8882616 | 1256 | 193 | AD | eGFR | 0.225038 | -0.22326 | 0.74721 | 0.0506421 | 0 | 0.55847 | 0.313315 | 504 | 0.680569117647059 |
| Chr19:13.6Mb | 2320 | 19 | 13213187 | 13968319 | 1610 | 274 | AD | eGFR | -0.152634 | -0.47857 | 0.14967 | 0.0232973 | 0 | 0.22903 | 0.313458 | 505 | 0.680569117647059 |
| Chr4:131.5Mb | 719 | 4 | 131098951 | 131873453 | 1906 | 162 | AD | eGFR | -0.380464 | -1 | 0.46939 | 0.144753 | 0 | 1 | 0.313774 | 506 | 0.680569117647059 |
| Chr15:97.4Mb | 2096 | 15 | 96864279 | 98025684 | 2363 | 321 | AD | eGFR | -0.272145 | -1 | 0.2974 | 0.0740628 | 0 | 1 | 0.314422 | 507 | 0.680569117647059 |
| Chr16:73.6Mb | 2151 | 16 | 73140782 | 74135275 | 2178 | 328 | AD | eGFR | 0.281695 | -0.29886 | 1 | 0.0793519 | 0 | 1 | 0.314559 | 508 | 0.680569117647059 |
| Chr10:50.9Mb | 1533 | 10 | 50145674 | 51724039 | 2139 | 199 | AD | eGFR | 0.251794 | -0.27352 | 1 | 0.0634001 | 0 | 1 | 0.314651 | 509 | 0.680569117647059 |
| Chr14:31.6Mb | 1964 | 14 | 30831155 | 32382245 | 2873 | 233 | AD | eGFR | 0.210065 | -0.21913 | 0.69994 | 0.0441273 | 0 | 0.49217 | 0.31525 | 510 | 0.680569117647059 |
| Chr9:130.3Mb | 1473 | 9 | 129617772 | 130902727 | 1730 | 210 | AD | eGFR | 0.143655 | -0.14321 | 0.44832 | 0.0206367 | 0 | 0.20174 | 0.31783 | 511 | 0.683770458984375 |
| Chr1:74.6Mb | 63 | 1 | 73992171 | 75132092 | 1878 | 184 | AD | eGFR | -0.210312 | -0.71139 | 0.21035 | 0.0442312 | 0 | 0.50902 | 0.317975 | 512 | 0.683770458984375 |
| Chr10:93.7Mb | 1572 | 10 | 92911507 | 94503299 | 2580 | 189 | AD | eGFR | 0.179591 | -0.18126 | 0.57803 | 0.0322531 | 0 | 0.33419 | 0.320117 | 513 | 0.685985118677043 |
| Chr5:43.5Mb | 813 | 5 | 42856333 | 44049878 | 2374 | 148 | AD | eGFR | 0.28695 | -0.32546 | 1 | 0.0823403 | 0 | 1 | 0.320251 | 514 | 0.685985118677043 |
| Chr11:126.8Mb | 1731 | 11 | 126304241 | 127306871 | 2581 | 292 | AD | eGFR | 0.413937 | -0.55144 | 1 | 0.171344 | 0 | 1 | 0.321019 | 515 | 0.686044386847195 |
| Chr19:51.6Mb | 2358 | 19 | 51259179 | 51903804 | 1700 | 295 | AD | eGFR | -0.142381 | -0.44046 | 0.14385 | 0.0202724 | 0 | 0.19412 | 0.321544 | 516 | 0.686044386847195 |
| Chr9:108.7Mb | 1452 | 9 | 108203096 | 109150717 | 1574 | 171 | AD | eGFR | -0.217711 | -0.72923 | 0.22717 | 0.0473981 | 0 | 0.53629 | 0.322148 | 517 | 0.686044386847195 |
| Chr1:200.6Mb | 154 | 1 | 200134006 | 201067952 | 2002 | 204 | AD | eGFR | -0.200528 | -0.78937 | 0.21722 | 0.0402116 | 0 | 0.63011 | 0.325595 | 518 | 0.691145855491329 |
| Chr12:66.6Mb | 1801 | 12 | 66114644 | 67183620 | 2089 | 250 | AD | eGFR | -0.175682 | -0.56495 | 0.17909 | 0.0308641 | 0 | 0.31975 | 0.325799 | 519 | 0.691145855491329 |
| Chr21:44.3Mb | 2458 | 21 | 43812792 | 44815008 | 2870 | 291 | AD | eGFR | 0.190499 | -0.18512 | 0.65061 | 0.0362897 | 0 | 0.42429 | 0.32867 | 520 | 0.693904195402299 |
| Chr12:72.6Mb | 1807 | 12 | 71948186 | 73154515 | 1952 | 198 | AD | eGFR | -0.28682 | -1 | 0.34032 | 0.0822659 | 0 | 1 | 0.32885 | 521 | 0.693904195402299 |
| Chr1:239Mb | 189 | 1 | 238704858 | 239332148 | 1776 | 183 | AD | eGFR | -0.270775 | -1 | 0.32954 | 0.0733193 | 0 | 1 | 0.32899 | 522 | 0.693904195402299 |
| Chr1:77.4Mb | 66 | 1 | 76977916 | 77895394 | 2071 | 198 | AD | eGFR | 0.410005 | -0.56691 | 1 | 0.168104 | 0 | 1 | 0.330015 | 523 | 0.694397554285714 |
| Chr7:149.2Mb | 1222 | 7 | 148622914 | 149842999 | 2226 | 250 | AD | eGFR | -0.182737 | -0.62456 | 0.194 | 0.033393 | 0 | 0.39215 | 0.330508 | 524 | 0.694397554285714 |
| Chr16:70.3Mb | 2148 | 16 | 69549150 | 71062712 | 1624 | 176 | AD | eGFR | 0.133431 | -0.13984 | 0.41147 | 0.0178037 | 0 | 0.16931 | 0.331116 | 525 | 0.694397554285714 |
| Chr13:80.1Mb | 1919 | 13 | 79489046 | 80778339 | 2287 | 238 | AD | eGFR | 0.244749 | -0.28456 | 0.91202 | 0.059902 | 0 | 0.8485 | 0.33532 | 526 | 0.701877034220532 |
| Chr8:123.3Mb | 1349 | 8 | 122622805 | 124001019 | 2513 | 293 | AD | eGFR | 0.23653 | -0.2746 | 0.86597 | 0.0559465 | 0 | 0.75919 | 0.339659 | 527 | 0.708434183364839 |
| Chr10:30.5Mb | 1519 | 10 | 30000224 | 30919469 | 2379 | 267 | AD | eGFR | -0.376905 | -1 | 0.51952 | 0.142057 | 0 | 1 | 0.340042 | 528 | 0.708434183364839 |
| Chr5:129.7Mb | 886 | 5 | 12873490 | 13057381 | 2881 | 167 | AD | eGFR | 0.142255 | -0.15594 | 0.45307 | 0.0202366 | 0 | 0.20528 | 0.340383 | 529 | 0.708434183364839 |

| locus_label | locus | chr | start | stop | n.snps | n.pcs | phen1 | phen2 | rho | rho.lower | rho.upper | r2 | r2.lower | r2.upper | p | rank | FDR |
|---|---|---|---|---|---|---|---|---|---|---|---|---|---|---|---|---|---|
| | | | 6 | 7 | | | | | | | | | | | | | |
| Chr19:37.6Mb | 2343 | 19 | 36638123 | 38472870 | 2958 | 139 | AD | eGFR | -0.211418 | -0.88711 | 0.26267 | 0.0446976 | 0 | 0.82285 | 0.342416 | 530 | 0.710537495429616 |
| Chr6:23.6Mb | 947 | 6 | 23192338 | 23939306 | 2085 | 242 | AD | eGFR | 0.25005 | -0.28659 | 0.94161 | 0.062525 | 0 | 0.90098 | 0.342871 | 531 | 0.710537495429616 |
| Chr7:93.1Mb | 1180 | 7 | 92530166 | 93691725 | 2138 | 245 | AD | eGFR | -0.236968 | -0.8839 | 0.2877 | 0.0561539 | 0 | 0.78859 | 0.343801 | 532 | 0.710537495429616 |
| Chr11:57.1Mb | 1667 | 11 | 56623495 | 57675986 | 1764 | 179 | AD | eGFR | 0.200552 | -0.24728 | 0.69997 | 0.0402211 | 0 | 0.4916 | 0.343996 | 533 | 0.710537495429616 |
| Chr7:42Mb | 1138 | 7 | 41415926 | 42494806 | 2221 | 261 | AD | eGFR | 0.182754 | -0.219 | 0.69775 | 0.0333989 | 0 | 0.49173 | 0.345535 | 534 | 0.710537495429616 |
| Chr9:129.2Mb | 1472 | 9 | 128785783 | 129617771 | 1726 | 277 | AD | eGFR | 0.197779 | -0.23696 | 0.67635 | 0.0391166 | 0 | 0.46062 | 0.347172 | 535 | 0.710537495429616 |
| Chr3:185.1Mb | 582 | 3 | 184524269 | 185709996 | 2064 | 184 | AD | eGFR | 0.193454 | -0.24774 | 0.78972 | 0.0374243 | 0 | 0.64126 | 0.34746 | 536 | 0.710537495429616 |
| Chr2:142.9Mb | 333 | 2 | 142494981 | 143259780 | 1835 | 179 | AD | eGFR | 0.284687 | -0.39065 | 1 | 0.0810466 | 0 | 1 | 0.348344 | 537 | 0.710537495429616 |
| Chr16:66.3Mb | 2145 | 16 | 65882821 | 66738843 | 1870 | 244 | AD | eGFR | -0.227097 | -0.8918 | 0.28545 | 0.0515729 | 0 | 0.80571 | 0.348438 | 538 | 0.710537495429616 |
| Chr15:68.2Mb | 2071 | 15 | 67396521 | 69089815 | 3059 | 294 | AD | eGFR | -0.218315 | -0.98253 | 0.27032 | 0.0476615 | 0 | 0.98846 | 0.348765 | 539 | 0.710537495429616 |
| Chr3:91.9Mb | 500 | 3 | 89612940 | 94250144 | 1854 | 133 | AD | eGFR | 0.256937 | -0.31316 | 0.97554 | 0.0660165 | 0 | 0.96814 | 0.349702 | 540 | 0.710537495429616 |
| Chr8:30.4Mb | 1275 | 8 | 29744541 | 31134786 | 2585 | 236 | AD | eGFR | 0.148555 | -0.16576 | 0.4983 | 0.0220686 | 0 | 0.24877 | 0.350579 | 541 | 0.710537495429616 |
| Chr10:120.1Mb | 1593 | 10 | 119528731 | 120592839 | 2441 | 265 | AD | eGFR | -0.210057 | -0.74839 | 0.25063 | 0.0441239 | 0 | 0.56208 | 0.351556 | 542 | 0.710537495429616 |
| Chr3:29.3Mb | 449 | 3 | 28649811 | 29877022 | 3410 | 308 | AD | eGFR | -0.247875 | -0.99723 | 0.3077 | 0.0614419 | 0 | 0.99591 | 0.351658 | 543 | 0.710537495429616 |
| Chr9:103.7Mb | 1447 | 9 | 103165726 | 104222848 | 2352 | 197 | AD | eGFR | 0.309126 | -0.42575 | 1 | 0.0955591 | 0 | 1 | 0.352472 | 544 | 0.710537495429616 |
| Chr11:59.9Mb | 1670 | 11 | 59256233 | 60515105 | 2220 | 198 | AD | eGFR | 0.171958 | -0.20088 | 0.60508 | 0.0295695 | 0 | 0.37412 | 0.352869 | 545 | 0.710537495429616 |
| Chr7:135.5Mb | 1212 | 7 | 134943222 | 136113467 | 2645 | 267 | AD | eGFR | -0.252578 | -1 | 0.33223 | 0.0637956 | 0 | 1 | 0.352911 | 546 | 0.710537495429616 |
| Chr10:99.9Mb | 1577 | 10 | 99367801 | 100337830 | 1900 | 252 | AD | eGFR | 0.214146 | -0.26435 | 0.78685 | 0.0458586 | 0 | 0.62161 | 0.35301 | 547 | 0.710537495429616 |
| Chr6:15.8Mb | 938 | 6 | 15069422 | 16566882 | 3174 | 309 | AD | eGFR | -0.251594 | -0.96605 | 0.32291 | 0.0632997 | 0 | 0.95719 | 0.355706 | 548 | 0.714210076502732 |
| Chr1:12.6Mb | 14 | 1 | 11709174 | 13481025 | 2299 | 289 | AD | eGFR | 0.142747 | -0.16204 | 0.47064 | 0.0203767 | 0 | 0.2215 | 0.356132 | 549 | 0.714210076502732 |
| Chr10:91.5Mb | 1570 | 10 | 91021322 | 91960435 | 2326 | 201 | AD | eGFR | -0.239182 | -1 | 0.31673 | 0.0572078 | 0 | 1 | 0.357067 | 550 | 0.714364508108108 |
| Chr1:50.5Mb | 44 | 1 | 49185415 | 51777577 | 2661 | 175 | AD | eGFR | -0.292971 | -1 | 0.39354 | 0.0858321 | 0 | 1 | 0.35753 | 551 | 0.714364508108108 |
| Chr16:85.6Mb | 2167 | 16 | 85146806 | 86058597 | 2315 | 376 | AD | eGFR | -0.178994 | -0.65786 | 0.21468 | 0.0320389 | 0 | 0.43299 | 0.35837 | 552 | 0.714364508108108 |
| Chr4:77.3Mb | 670 | 4 | 76497359 | 78045637 | 3584 | 264 | AD | eGFR | -0.0992879 | -0.33012 | 0.12327 | 0.00985809 | 0 | 0.10924 | 0.359529 | 553 | 0.714364508108108 |
| Chr1:0.7Mb | 1 | 1 | 10539 | 1376204 | 1027 | 138 | AD | eGFR | -0.172248 | -0.64161 | 0.22114 | 0.0296694 | 0 | 0.41958 | 0.359953 | 554 | 0.714364508108108 |
| Chr14:73.8Mb | 2003 | 14 | 73373374 | 74245428 | 1602 | 154 | AD | eGFR | -0.193623 | -0.76738 | 0.25935 | 0.03749 | 0 | 0.60306 | 0.360102 | 555 | 0.714364508108108 |
| Chr20:60.4Mb | 2426 | 20 | 60125334 | 60766557 | 1782 | 296 | AD | eGFR | 0.179336 | -0.23251 | 0.67856 | 0.0321614 | 0 | 0.46438 | 0.362335 | 556 | 0.717501501798561 |
| Chr19:38.9Mb | 2344 | 19 | 38472871 | 39370419 | 2008 | 218 | AD | eGFR | -0.111774 | -0.35855 | 0.14145 | 0.0124934 | 0 | 0.12856 | 0.363614 | 557 | 0.718741497307002 |
| Chr13:33.9Mb | 1876 | 13 | 32986687 | 34722391 | 3307 | 301 | AD | eGFR | -0.242847 | -1 | 0.31808 | 0.0589746 | 0 | 1 | 0.366437 | 558 | 0.723023543010753 |
| Chr14:64.6Mb | 1995 | 14 | 64033238 | 65229391 | 2189 | 177 | AD | eGFR | 0.391496 | -0.62823 | 1 | 0.153269 | 0 | 1 | 0.368633 | 559 | 0.724995788632327 |
| Chr6:161.8Mb | 1085 | 6 | 161371015 | 162254229 | 2014 | 289 | AD | eGFR | -0.160285 | -0.54794 | 0.20291 | 0.0256912 | 0 | 0.30078 | 0.369224 | 560 | 0.724995788632327 |
| Chr1:17.1Mb | 18 | 1 | 16732169 | 17557746 | 1008 | 166 | AD | eGFR | 0.240266 | -0.33844 | 1 | 0.0577278 | 0 | 1 | 0.369824 | 561 | 0.724995788632327 |
| Chr1:40.6Mb | 37 | 1 | 40200951 | 41093875 | 1747 | 202 | AD | eGFR | 0.188812 | -0.24935 | 0.69556 | 0.03565 | 0 | 0.4876 | 0.37053 | 562 | 0.724995788632327 |
| Chr6:91.6Mb | 1023 | 6 | 91097025 | 92015279 | 2037 | 223 | AD | eGFR | 0.327585 | -0.49131 | 1 | 0.107312 | 0 | 1 | 0.370729 | 563 | 0.724995788632327 |
| Chr12:7.5Mb | 1747 | 12 | 6948955 | 8123317 | 2212 | 257 | AD | eGFR | -0.26363 | -1 | 0.39042 | 0.0695009 | 0 | 1 | 0.371796 | 564 | 0.725793255319149 |
| Chr1:163.5Mb | 120 | 1 | 163141863 | 163854567 | 1788 | 170 | AD | eGFR | -0.165429 | -0.56012 | 0.20356 | 0.0273668 | 0 | 0.31692 | 0.373455 | 565 | 0.727741513274336 |

| locus_label | locus | chr | start | stop | n.snps | n.pcs | phen1 | phen2 | rho | rho.lower | rho.upper | r2 | r2.lower | r2.upper | p | rank | FDR |
|---|---|---|---|---|---|---|---|---|---|---|---|---|---|---|---|---|---|
| Chr17:75.3Mb | 2231 | 17 | 74908267 | 75619256 | 2219 | 317 | AD | eGFR | 0.172565 | -0.21306 | 0.61228 | 0.0297786 | 0 | 0.3752 | 0.374706 | 566 | 0.728061978835979 |
| Chr1:36Mb | 34 | 1 | 35201627 | 36855139 | 1363 | 212 | AD | eGFR | -0.127942 | -0.42877 | 0.15834 | 0.0163692 | 0 | 0.18481 | 0.374942 | 567 | 0.728061978835979 |
| Chr20:36.6Mb | 2401 | 20 | 35990262 | 37253657 | 2293 | 275 | AD | eGFR | 0.191743 | -0.24899 | 0.72254 | 0.0367654 | 0 | 0.52603 | 0.37648 | 568 | 0.729301239015817 |
| Chr1:243.9Mb | 195 | 1 | 243048004 | 244723196 | 2413 | 306 | AD | eGFR | -0.0971395 | -0.31515 | 0.11793 | 0.00943608 | 0 | 0.09953 | 0.376905 | 569 | 0.729301239015817 |
| Chr1:185.6Mb | 140 | 1 | 184953090 | 186179447 | 1482 | 208 | AD | eGFR | -0.11981 | -0.39989 | 0.15035 | 0.0143544 | 0 | 0.15991 | 0.377911 | 570 | 0.729964931578947 |
| Chr19:12.4Mb | 2319 | 19 | 11681979 | 13213186 | 2559 | 223 | AD | eGFR | -0.173677 | -0.60457 | 0.23004 | 0.0301637 | 0 | 0.3681 | 0.379213 | 571 | 0.731197045534151 |
| Chr15:25.9Mb | 2038 | 15 | 25384328 | 26392947 | 2769 | 330 | AD | eGFR | 0.265688 | -0.38519 | 1 | 0.0705901 | 0 | 1 | 0.38114 | 572 | 0.733622149565217 |
| Chr1:65.1Mb | 55 | 1 | 64344227 | 65894184 | 2909 | 282 | AD | eGFR | -0.334119 | -1 | 0.54675 | 0.111635 | 0 | 1 | 0.382668 | 573 | 0.733622149565217 |
| Chr14:107.1Mb | 2033 | 14 | 106876549 | 107289435 | 993 | 69 | AD | eGFR | -0.251258 | -1 | 0.37516 | 0.0631306 | 0 | 1 | 0.382971 | 574 | 0.733622149565217 |
| Chr2:0.4Mb | 201 | 2 | 10181 | 789195 | 1756 | 201 | AD | eGFR | -0.279017 | -1 | 0.40996 | 0.0778507 | 0 | 1 | 0.383136 | 575 | 0.733622149565217 |
| Chr1:173Mb | 130 | 1 | 172465154 | 173582511 | 1805 | 227 | AD | eGFR | 0.176855 | -0.2346 | 0.61919 | 0.0312777 | 0 | 0.38761 | 0.384879 | 576 | 0.733749830449827 |
| Chr6:35.7Mb | 971 | 6 | 34979271 | 36346353 | 2514 | 238 | AD | eGFR | -0.187029 | -0.71929 | 0.25774 | 0.0349799 | 0 | 0.5235 | 0.384931 | 577 | 0.733749830449827 |
| Chr13:37Mb | 1879 | 13 | 36579414 | 37499810 | 1955 | 212 | AD | eGFR | -0.282009 | -1 | 0.44128 | 0.0795293 | 0 | 1 | 0.385202 | 578 | 0.733749830449827 |
| Chr12:116.6Mb | 1845 | 12 | 116197676 | 117091843 | 2069 | 271 | AD | eGFR | 0.312692 | -0.48875 | 1 | 0.0977764 | 0 | 1 | 0.385903 | 579 | 0.733815549222798 |
| Chr7:79.4Mb | 1170 | 7 | 78685010 | 80055721 | 2608 | 220 | AD | eGFR | -0.278661 | -1 | 0.43058 | 0.077652 | 0 | 1 | 0.387843 | 580 | 0.736233005172414 |
| Chr10:11.4Mb | 1498 | 10 | 10969482 | 11856924 | 2117 | 282 | AD | eGFR | -0.20091 | -0.93397 | 0.29971 | 0.0403649 | 0 | 0.9104 | 0.389186 | 581 | 0.737510819277108 |
| Chr18:22.2Mb | 2256 | 18 | 21622717 | 22746153 | 2364 | 284 | AD | eGFR | -0.257392 | -1 | 0.406 | 0.0662508 | 0 | 1 | 0.391369 | 582 | 0.739006256410256 |
| Chr6:22.7Mb | 946 | 6 | 22199714 | 23192337 | 2455 | 226 | AD | eGFR | -0.321141 | -1 | 0.54201 | 0.103132 | 0 | 1 | 0.391481 | 583 | 0.739006256410256 |
| Chr5:180.3Mb | 925 | 5 | 179794711 | 180902886 | 2345 | 260 | AD | eGFR | 0.132867 | -0.17714 | 0.47128 | 0.0176536 | 0 | 0.2221 | 0.392436 | 584 | 0.739006256410256 |
| Chr12:20.5Mb | 1760 | 12 | 20074931 | 20866285 | 1955 | 253 | AD | eGFR | -0.159489 | -0.56713 | 0.21727 | 0.0254367 | 0 | 0.32216 | 0.39266 | 585 | 0.739006256410256 |
| Chr6:7.2Mb | 932 | 6 | 6621938 | 7808323 | 3195 | 343 | AD | eGFR | -0.0957285 | -0.33467 | 0.13313 | 0.00916395 | 0 | 0.11236 | 0.393897 | 586 | 0.739248474489796 |
| Chr1:18.8Mb | 20 | 1 | 18427821 | 19238924 | 2279 | 319 | AD | eGFR | -0.138104 | -0.4842 | 0.1804 | 0.0190728 | 0 | 0.23519 | 0.394785 | 587 | 0.739248474489796 |
| Chr4:74.1Mb | 668 | 4 | 73258295 | 74885900 | 2741 | 215 | AD | eGFR | 0.174312 | -0.24901 | 0.69226 | 0.0303846 | 0 | 0.50162 | 0.394803 | 588 | 0.739248474489796 |
| Chr16:74.5Mb | 2152 | 16 | 74135276 | 74889127 | 1601 | 195 | AD | eGFR | 0.275548 | -0.42734 | 1 | 0.0759269 | 0 | 1 | 0.395765 | 589 | 0.739250018549747 |
| Chr7:29.5Mb | 1127 | 7 | 28890887 | 30175113 | 3371 | 303 | AD | eGFR | -0.283917 | -1 | 0.4666 | 0.0806089 | 0 | 1 | 0.396481 | 590 | 0.739250018549747 |
| Chr12:114.3Mb | 1842 | 12 | 113947984 | 114685670 | 2143 | 218 | AD | eGFR | -0.333381 | -1 | 0.55793 | 0.111143 | 0 | 1 | 0.397482 | 591 | 0.739250018549747 |
| Chr16:23.3Mb | 2123 | 16 | 22667049 | 24014705 | 2892 | 283 | AD | eGFR | -0.135102 | -0.4678 | 0.18878 | 0.0182525 | 0 | 0.2208 | 0.398104 | 592 | 0.739250018549747 |
| Chr19:1.1Mb | 2307 | 19 | 762047 | 1515920 | 1890 | 327 | AD | eGFR | 0.0751038 | -0.10333 | 0.25385 | 0.00564058 | 0 | 0.06454 | 0.398161 | 593 | 0.739250018549747 |
| Chr3:5.2Mb | 427 | 3 | 4835084 | 5496182 | 1649 | 300 | AD | eGFR | 0.136822 | -0.19686 | 0.48749 | 0.0187203 | 0 | 0.23834 | 0.404746 | 594 | 0.75021102020202 |
| Chr3:193.7Mb | 590 | 3 | 193122091 | 194217140 | 2581 | 328 | AD | eGFR | 0.155392 | -0.2407 | 0.6262 | 0.0241468 | 0 | 0.40497 | 0.406976 | 595 | 0.753076598319328 |
| Chr2:232.6Mb | 408 | 2 | 231847387 | 233352722 | 3080 | 278 | AD | eGFR | -0.28441 | -1 | 0.48668 | 0.0808888 | 0 | 1 | 0.408039 | 596 | 0.753776743288591 |
| Chr11:87.2Mb | 1693 | 11 | 86629334 | 87676870 | 3192 | 224 | AD | eGFR | -0.312901 | -1 | 0.57367 | 0.097907 | 0 | 1 | 0.41068 | 597 | 0.757384723618091 |
| Chr18:47.8Mb | 2276 | 18 | 47455926 | 48228379 | 1742 | 201 | AD | eGFR | -0.212892 | -0.93279 | 0.35918 | 0.0453231 | 0 | 0.9072 | 0.417793 | 598 | 0.769214202341137 |
| Chr1:234.7Mb | 184 | 1 | 234365797 | 235097113 | 1958 | 293 | AD | eGFR | 0.126042 | -0.18899 | 0.45932 | 0.0158867 | 0 | 0.21183 | 0.419522 | 599 | 0.771108050083472 |
| Chr4:88.9Mb | 679 | 4 | 88502718 | 89244554 | 1817 | 187 | AD | eGFR | 0.215325 | -0.37561 | 0.91673 | 0.0463647 | 0 | 0.8485 | 0.421337 | 600 | 0.771537637873754 |
| Chr10:16.5Mb | 1504 | 10 | 15946678 | 17065860 | 2596 | 271 | AD | eGFR | -0.151363 | -0.56943 | 0.227 | 0.0229107 | 0 | 0.32517 | 0.421608 | 601 | 0.771537637873754 |

| locus_label | locus | chr | start | stop | n.snps | n.pcs | phen1 | phen2 | rho | rho.lower | rho.upper | r2 | r2.lower | r2.upper | p | rank | FDR |
|---|---|---|---|---|---|---|---|---|---|---|---|---|---|---|---|---|---|
| Chr14:75Mb | 2004 | 14 | 74245429 | 75699501 | 2733 | 220 | AD | eGFR | 0.160027 | -0.24863 | 0.62769 | 0.0256088 | 0 | 0.39883 | 0.421858 | 602 | 0.771537637873754 |
| Chr3:75.8Mb | 487 | 3 | 75374968 | 76157470 | 719 | 80 | AD | eGFR | -0.212031 | -0.95582 | 0.36171 | 0.044957 | 0 | 0.93741 | 0.423905 | 603 | 0.772956415156507 |
| Chr20:3.2Mb | 2372 | 20 | 2668109 | 3736551 | 2346 | 269 | AD | eGFR | 0.16728 | -0.26934 | 0.71682 | 0.0279826 | 0 | 0.54116 | 0.424481 | 604 | 0.772956415156507 |
| Chr9:14.6Mb | 1389 | 9 | 14244748 | 14902924 | 1762 | 303 | AD | eGFR | 0.153546 | -0.25202 | 0.57703 | 0.0235765 | 0 | 0.33473 | 0.425586 | 605 | 0.772956415156507 |
| Chr14:90.1Mb | 2017 | 14 | 89482884 | 90662836 | 2630 | 316 | AD | eGFR | 0.235214 | -0.40752 | 1 | 0.0553255 | 0 | 1 | 0.426055 | 606 | 0.772956415156507 |
| Chr7:2.8Mb | 1097 | 7 | 2473750 | 3100211 | 1792 | 287 | AD | eGFR | -0.15971 | -0.61539 | 0.24686 | 0.0255072 | 0 | 0.38004 | 0.426144 | 607 | 0.772956415156507 |
| Chr17:79.9Mb | 2236 | 17 | 79333775 | 80459149 | 2125 | 249 | AD | eGFR | 0.079704 | -0.11846 | 0.28166 | 0.00635272 | 0 | 0.07975 | 0.430086 | 608 | 0.777104838499184 |
| Chr15:55.7Mb | 2061 | 15 | 55136718 | 56322588 | 3263 | 231 | AD | eGFR | 0.289197 | -0.53846 | 1 | 0.083635 | 0 | 1 | 0.430682 | 609 | 0.777104838499184 |
| Chr8:126.1Mb | 1351 | 8 | 125453323 | 126766827 | 2760 | 306 | AD | eGFR | -0.0878713 | -0.31954 | 0.13137 | 0.00772136 | 0 | 0.1022 | 0.431566 | 610 | 0.777104838499184 |
| Chr2:65.3Mb | 271 | 2 | 64696090 | 65938002 | 2575 | 301 | AD | eGFR | 0.115663 | -0.17915 | 0.42006 | 0.0133779 | 0 | 0.17711 | 0.431968 | 611 | 0.777104838499184 |
| Chr16:56.5Mb | 2137 | 16 | 55907390 | 57119889 | 2697 | 288 | AD | eGFR | 0.21813 | -0.39091 | 1 | 0.0475808 | 0 | 1 | 0.432501 | 612 | 0.777104838499184 |
| Chr2:26.4Mb | 229 | 2 | 25895395 | 26894102 | 1625 | 225 | AD | eGFR | -0.128834 | -0.49096 | 0.2058 | 0.0165982 | 0 | 0.24176 | 0.432666 | 613 | 0.777104838499184 |
| Chr11:56.3Mb | 1666 | 11 | 55947453 | 56623494 | 1817 | 56 | AD | eGFR | 0.246935 | -0.46466 | 1 | 0.0609766 | 0 | 1 | 0.437179 | 614 | 0.780865959481362 |
| Chr19:32.4Mb | 2338 | 19 | 31763009 | 33028099 | 2417 | 283 | AD | eGFR | 0.121927 | -0.19644 | 0.45487 | 0.0148661 | 0 | 0.20758 | 0.437219 | 615 | 0.780865959481362 |
| Chr20:16.3Mb | 2386 | 20 | 15962577 | 16661516 | 2108 | 215 | AD | eGFR | -0.32543 | -1 | 0.63559 | 0.105905 | 0 | 1 | 0.437301 | 616 | 0.780865959481362 |
| Chr8:134.9Mb | 1359 | 8 | 134509449 | 135313673 | 2576 | 279 | AD | eGFR | -0.254152 | -1 | 0.45758 | 0.0645931 | 0 | 1 | 0.437597 | 617 | 0.780865959481362 |
| Chr4:10.7Mb | 610 | 4 | 10370667 | 11050119 | 1806 | 189 | AD | eGFR | 0.199082 | -0.35518 | 0.88002 | 0.0396336 | 0 | 0.80845 | 0.440558 | 618 | 0.783697473429952 |
| Chr20:52.8Mb | 2416 | 20 | 52411533 | 53120334 | 1951 | 246 | AD | eGFR | 0.0735314 | -0.11492 | 0.26438 | 0.00540687 | 0 | 0.07043 | 0.44062 | 619 | 0.783697473429952 |
| Chr6:143.9Mb | 1067 | 6 | 143083348 | 144791455 | 3004 | 323 | AD | eGFR | 0.191461 | -0.33131 | 0.82223 | 0.0366573 | 0 | 0.69526 | 0.441624 | 620 | 0.783697473429952 |
| Chr9:109.9Mb | 1453 | 9 | 109150718 | 110630150 | 3315 | 338 | AD | eGFR | 0.262286 | -0.53751 | 1 | 0.0687941 | 0 | 1 | 0.442031 | 621 | 0.783697473429952 |
| Chr20:44.9Mb | 2408 | 20 | 44072211 | 45673603 | 3138 | 322 | AD | eGFR | -0.21701 | -1 | 0.38301 | 0.0470934 | 0 | 1 | 0.443593 | 622 | 0.785202400321543 |
| Chr10:123.3Mb | 1596 | 10 | 122803507 | 123856184 | 2364 | 289 | AD | eGFR | -0.206491 | -0.92368 | 0.37397 | 0.0426386 | 0 | 0.86678 | 0.444678 | 623 | 0.785859515248796 |
| Chr1:87.2Mb | 75 | 1 | 86761555 | 87667572 | 1772 | 148 | AD | eGFR | -0.195215 | -0.92026 | 0.34321 | 0.0381089 | 0 | 0.87497 | 0.445551 | 624 | 0.786140466346154 |
| Chr5:4.2Mb | 780 | 5 | 3677596 | 4636542 | 2090 | 265 | AD | eGFR | 0.107694 | -0.16577 | 0.3971 | 0.011598 | 0 | 0.15775 | 0.446524 | 625 | 0.7865966784 |
| Chr3:31.8Mb | 451 | 3 | 31269698 | 32378805 | 2459 | 305 | AD | eGFR | -0.183112 | -0.82844 | 0.33936 | 0.0335299 | 0 | 0.71442 | 0.448302 | 626 | 0.786814795238095 |
| Chr3:24.9Mb | 445 | 3 | 24270190 | 25516193 | 3351 | 317 | AD | eGFR | -0.130262 | -0.50809 | 0.22518 | 0.0169681 | 0 | 0.25963 | 0.44889 | 627 | 0.786814795238095 |
| Chr7:138.2Mb | 1214 | 7 | 137657460 | 138755107 | 2600 | 325 | AD | eGFR | -0.119627 | -0.45473 | 0.20119 | 0.0143107 | 0 | 0.2079 | 0.449856 | 628 | 0.786814795238095 |
| Chr9:135.6Mb | 1478 | 9 | 135135888 | 136042490 | 1947 | 280 | AD | eGFR | 0.212231 | -0.39901 | 1 | 0.0450421 | 0 | 1 | 0.449887 | 629 | 0.786814795238095 |
| Chr9:92.1Mb | 1436 | 9 | 91643256 | 92500277 | 1893 | 217 | AD | eGFR | 0.137289 | -0.2323 | 0.54062 | 0.0188483 | 0 | 0.30141 | 0.450221 | 630 | 0.786814795238095 |
| Chr7:128.3Mb | 1207 | 7 | 127903739 | 128779173 | 2086 | 205 | AD | eGFR | 0.134191 | -0.24254 | 0.57952 | 0.0180073 | 0 | 0.34063 | 0.452083 | 631 | 0.788816771790808 |
| Chr2:219Mb | 395 | 2 | 218396259 | 219678783 | 2217 | 244 | AD | eGFR | -0.131139 | -0.53895 | 0.22017 | 0.0171974 | 0 | 0.29613 | 0.455008 | 632 | 0.791129089622642 |
| Chr3:11.6Mb | 433 | 3 | 11226368 | 11997658 | 1673 | 228 | AD | eGFR | 0.241494 | -0.48807 | 1 | 0.0583194 | 0 | 1 | 0.455061 | 633 | 0.791129089622642 |
| Chr10:8.2Mb | 1495 | 10 | 7659873 | 8771920 | 3359 | 316 | AD | eGFR | 0.205797 | -0.39758 | 0.99927 | 0.0423526 | 0 | 1 | 0.45655 | 634 | 0.791129089622642 |
| Chr18:72.1Mb | 2299 | 18 | 71491861 | 72785206 | 2966 | 320 | AD | eGFR | -0.147919 | -0.60639 | 0.2614 | 0.0218801 | 0 | 0.3726 | 0.456867 | 635 | 0.791129089622642 |
| Chr11:45.7Mb | 1657 | 11 | 45019560 | 46316005 | 2492 | 253 | AD | eGFR | 0.172291 | -0.32466 | 0.78616 | 0.0296841 | 0 | 0.63921 | 0.457001 | 636 | 0.791129089622642 |
| Chr8:129.8Mb | 1354 | 8 | 129291119 | 130383563 | 2408 | 203 | AD | eGFR | 0.196425 | -0.38244 | 0.87911 | 0.0385828 | 0 | 0.80003 | 0.458405 | 637 | 0.792313822605966 |

| locus_label | locus | chr | start | stop | n.snps | n.pcs | phen1 | phen2 | rho | rho.lower | rho.upper | r2 | r2.lower | r2.upper | p | rank | FDR |
|---|---|---|---|---|---|---|---|---|---|---|---|---|---|---|---|---|---|
| Chr10:105.2Mb | 1581 | 10 | 104206838 | 106142283 | 3358 | 250 | AD | eGFR | 0.122208 | -0.21692 | 0.48342 | 0.0149347 | 0 | 0.23681 | 0.459702 | 638 | 0.792721723004695 |
| Chr20:8.6Mb | 2378 | 20 | 7959829 | 9279550 | 2929 | 293 | AD | eGFR | 0.112701 | -0.19665 | 0.42627 | 0.0127015 | 0 | 0.18225 | 0.460081 | 639 | 0.792721723004695 |
| Chr1:19.9Mb | 21 | 1 | 19238925 | 20556202 | 3123 | 298 | AD | eGFR | 0.111552 | -0.19249 | 0.42304 | 0.0124438 | 0 | 0.17907 | 0.461393 | 640 | 0.7937401453125 |
| Chr15:49.7Mb | 2056 | 15 | 49260007 | 50195876 | 1809 | 109 | AD | eGFR | -0.167326 | -0.74474 | 0.31419 | 0.0279981 | 0 | 0.56662 | 0.462243 | 641 | 0.793961845553822 |
| Chr20:34.9Mb | 2400 | 20 | 33890061 | 35990261 | 2639 | 238 | AD | eGFR | -0.151546 | -0.78618 | 0.30706 | 0.0229662 | 0 | 0.68244 | 0.466745 | 642 | 0.796942183486239 |
| Chr2:9.5Mb | 213 | 2 | 8978799 | 9960670 | 1828 | 235 | AD | eGFR | 0.153851 | -0.27863 | 0.64228 | 0.0236702 | 0 | 0.41995 | 0.466915 | 643 | 0.796942183486239 |
| Chr1:98.6Mb | 85 | 1 | 97721187 | 99425000 | 3322 | 274 | AD | eGFR | -0.181426 | -0.81497 | 0.3357 | 0.0329156 | 0 | 0.68167 | 0.467109 | 644 | 0.796942183486239 |
| Chr18:31.4Mb | 2263 | 18 | 30401129 | 32458907 | 2985 | 251 | AD | eGFR | 0.203499 | -0.39547 | 0.98609 | 0.0414118 | 0 | 1 | 0.467557 | 645 | 0.796942183486239 |
| Chr3:66Mb | 478 | 3 | 65326752 | 66702084 | 2830 | 319 | AD | eGFR | -0.181403 | -0.81832 | 0.35403 | 0.0329072 | 0 | 0.68812 | 0.469297 | 646 | 0.796942183486239 |
| Chr4:190.5Mb | 774 | 4 | 189899574 | 191043880 | 2042 | 229 | AD | eGFR | -0.165898 | -0.72272 | 0.3257 | 0.0275221 | 0 | 0.54513 | 0.470801 | 647 | 0.796942183486239 |
| Chr2:15.2Mb | 220 | 2 | 14692121 | 15721703 | 2072 | 197 | AD | eGFR | -0.219329 | -1 | 0.44395 | 0.0481054 | 0 | 1 | 0.470909 | 648 | 0.796942183486239 |
| Chr2:46.9Mb | 252 | 2 | 46499327 | 47320114 | 2219 | 267 | AD | eGFR | -0.214114 | -1 | 0.44673 | 0.0458449 | 0 | 1 | 0.471958 | 649 | 0.796942183486239 |
| Chr2:44.6Mb | 249 | 2 | 44104111 | 45189468 | 2199 | 246 | AD | eGFR | 0.16562 | -0.31693 | 0.7111 | 0.02743 | 0 | 0.52196 | 0.471971 | 650 | 0.796942183486239 |
| Chr21:46.5Mb | 2460 | 21 | 46045434 | 47014880 | 2920 | 302 | AD | eGFR | 0.11689 | -0.21308 | 0.45472 | 0.0136634 | 0 | 0.20802 | 0.472636 | 651 | 0.796942183486239 |
| Chr17:7.9Mb | 2181 | 17 | 7264459 | 8554763 | 2573 | 288 | AD | eGFR | -0.127251 | -0.53437 | 0.22151 | 0.0161929 | 0 | 0.28975 | 0.472751 | 652 | 0.796942183486239 |
| Chr3:141.5Mb | 544 | 3 | 141034638 | 141898978 | 1881 | 220 | AD | eGFR | 0.0828585 | -0.14977 | 0.31745 | 0.00686552 | 0 | 0.10102 | 0.473127 | 653 | 0.796942183486239 |
| Chr17:11.2Mb | 2184 | 17 | 10572618 | 11778164 | 2537 | 312 | AD | eGFR | 0.199703 | -0.39483 | 0.97159 | 0.0398814 | 0 | 0.98493 | 0.473388 | 654 | 0.796942183486239 |
| Chr1:11.2Mb | 13 | 1 | 10753428 | 11709173 | 1941 | 258 | AD | eGFR | 0.110909 | -0.1976 | 0.42761 | 0.0123007 | 0 | 0.18588 | 0.475597 | 655 | 0.797345205479452 |
| Chr4:72.4Mb | 667 | 4 | 71468301 | 73258294 | 2960 | 222 | AD | eGFR | -0.185477 | -0.98275 | 0.38433 | 0.0344018 | 0 | 1 | 0.475689 | 656 | 0.797345205479452 |
| Chr2:130Mb | 321 | 2 | 129486898 | 130485398 | 2407 | 207 | AD | eGFR | -0.212538 | -1 | 0.44587 | 0.0451723 | 0 | 1 | 0.4758 | 657 | 0.797345205479452 |
| Chr17:71.9Mb | 2228 | 17 | 71466954 | 72383472 | 2394 | 349 | AD | eGFR | -0.207519 | -1 | 0.44131 | 0.043064 | 0 | 1 | 0.479951 | 658 | 0.802735763636364 |
| Chr1:53.9Mb | 46 | 1 | 53114187 | 54771293 | 3367 | 318 | AD | eGFR | 0.156531 | -0.31501 | 0.66938 | 0.0245018 | 0 | 0.45836 | 0.480732 | 659 | 0.802735763636364 |
| Chr3:19.9Mb | 440 | 3 | 19030989 | 20754842 | 3431 | 248 | AD | eGFR | 0.176945 | -0.3554 | 0.81584 | 0.0313095 | 0 | 0.6794 | 0.481204 | 660 | 0.802735763636364 |
| Chr14:73Mb | 2002 | 14 | 72665320 | 73373373 | 1734 | 236 | AD | eGFR | -0.109721 | -0.44342 | 0.20954 | 0.0120388 | 0 | 0.19835 | 0.482544 | 661 | 0.803753319213313 |
| Chr3:67.1Mb | 479 | 3 | 66702085 | 67454740 | 2071 | 198 | AD | eGFR | -0.264553 | -1 | 0.5955 | 0.0699882 | 0 | 1 | 0.484029 | 662 | 0.804106963800905 |
| Chr15:73.9Mb | 2075 | 15 | 73375719 | 74458113 | 1914 | 214 | AD | eGFR | 0.135722 | -0.26999 | 0.60861 | 0.0184206 | 0 | 0.37455 | 0.484217 | 663 | 0.804106963800905 |
| Chr11:108.9Mb | 1716 | 11 | 107846825 | 109891837 | 3058 | 268 | AD | eGFR | -0.211836 | -1 | 0.4622 | 0.0448745 | 0 | 1 | 0.486341 | 664 | 0.804255776946108 |
| Chr22:18.1Mb | 2463 | 22 | 17587475 | 18535225 | 2613 | 327 | AD | eGFR | 0.152978 | -0.3138 | 0.69659 | 0.0234024 | 0 | 0.49747 | 0.487175 | 665 | 0.804255776946108 |
| Chr10:112.6Mb | 1587 | 10 | 112164982 | 113117615 | 1935 | 260 | AD | eGFR | 0.234402 | -0.53679 | 1 | 0.0549445 | 0 | 1 | 0.487546 | 666 | 0.804255776946108 |
| Chr8:76.8Mb | 1311 | 8 | 76295484 | 77377989 | 2500 | 147 | AD | eGFR | 0.178835 | -0.39435 | 0.91559 | 0.0319821 | 0 | 0.89337 | 0.487767 | 667 | 0.804255776946108 |
| Chr1:228.6Mb | 177 | 1 | 228045555 | 229075156 | 1677 | 192 | AD | eGFR | 0.190611 | -0.41696 | 0.96397 | 0.0363324 | 0 | 0.99153 | 0.487959 | 668 | 0.804255776946108 |
| Chr1:204.6Mb | 157 | 1 | 204092538 | 205009623 | 2010 | 256 | AD | eGFR | -0.129871 | -0.53302 | 0.24891 | 0.0168665 | 0 | 0.28558 | 0.491591 | 669 | 0.806997008915305 |
| Chr9:113.3Mb | 1457 | 9 | 112865521 | 113646149 | 2153 | 232 | AD | eGFR | 0.155148 | -0.30784 | 0.67097 | 0.0240709 | 0 | 0.45222 | 0.49199 | 670 | 0.806997008915305 |
| Chr17:1.1Mb | 2174 | 17 | 815654 | 1480056 | 1925 | 318 | AD | eGFR | 0.103463 | -0.20008 | 0.418 | 0.0107047 | 0 | 0.17533 | 0.492814 | 671 | 0.806997008915305 |
| Chr17:19.7Mb | 2192 | 17 | 18900979 | 20402494 | 1400 | 153 | AD | eGFR | 0.0922123 | -0.17706 | 0.38155 | 0.00850311 | 0 | 0.14743 | 0.493105 | 672 | 0.806997008915305 |
| Chr8:23.3Mb | 1268 | 8 | 22895019 | 23788494 | 2604 | 273 | AD | eGFR | -0.119717 | -0.63387 | 0.26224 | 0.014332 | 0 | 0.42485 | 0.493287 | 673 | 0.806997008915305 |
| Chr1:109.7Mb | 96 | 1 | 10927145 | 11022423 | 1823 | 220 | AD | eGFR | 0.0897378 | -0.17766 | 0.36412 | 0.00805287 | 0 | 0.13449 | 0.494889 | 674 | 0.808416600890208 |

| locus_label | locus | chr | start | stop | n.snps | n.pcs | phen1 | phen2 | rho | rho.lower | rho.upper | r2 | r2.lower | r2.upper | p | rank | FDR |
|---|---|---|---|---|---|---|---|---|---|---|---|---|---|---|---|---|---|
| | | | 0 | 0 | | | | | | | | | | | | | |
| Chr5:52.5Mb | 819 | 5 | 52139774 | 52837225 | 1906 | 168 | AD | eGFR | -0.114544 | -0.4948 | 0.22518 | 0.0131203 | 0 | 0.246 | 0.496855 | 675 | 0.808686903930131 |
| Chr16:59.2Mb | 2139 | 16 | 58508979 | 59885499 | 3333 | 255 | AD | eGFR | 0.230394 | -0.56342 | 1 | 0.0530813 | 0 | 1 | 0.497823 | 676 | 0.808686903930131 |
| Chr1:28.6Mb | 28 | 1 | 27992666 | 29188426 | 1784 | 175 | AD | eGFR | 0.181624 | -0.39327 | 0.93787 | 0.0329872 | 0 | 0.92075 | 0.498396 | 677 | 0.808686903930131 |
| Chr19:50.9Mb | 2357 | 19 | 50451926 | 51259178 | 1663 | 277 | AD | eGFR | -0.108451 | -0.44174 | 0.21284 | 0.0117617 | 0 | 0.19693 | 0.500413 | 678 | 0.808686903930131 |
| Chr19:41.4Mb | 2347 | 19 | 41004106 | 41733463 | 1572 | 224 | AD | eGFR | 0.0915943 | -0.18073 | 0.36927 | 0.00838952 | 0 | 0.13736 | 0.500431 | 679 | 0.808686903930131 |
| Chr22:43.2Mb | 2486 | 22 | 42867928 | 43574017 | 1766 | 202 | AD | eGFR | -0.116032 | -0.52125 | 0.2449 | 0.0134635 | 0 | 0.2877 | 0.501264 | 680 | 0.808686903930131 |
| Chr1:66.3Mb | 56 | 1 | 65894185 | 66778015 | 1819 | 160 | AD | eGFR | -0.14302 | -0.61015 | 0.29936 | 0.0204548 | 0 | 0.3771 | 0.501313 | 681 | 0.808686903930131 |
| Chr17:0.4Mb | 2173 | 17 | 52 | 815653 | 2049 | 291 | AD | eGFR | -0.183461 | -0.94093 | 0.41743 | 0.033658 | 0 | 0.95317 | 0.502135 | 682 | 0.808686903930131 |
| Chr11:116.8Mb | 1722 | 11 | 116247378 | 117295641 | 2756 | 262 | AD | eGFR | -0.164974 | -0.80347 | 0.35152 | 0.0272165 | 0 | 0.68073 | 0.502508 | 683 | 0.808686903930131 |
| Chr2:168.8Mb | 352 | 2 | 168437446 | 169224445 | 1817 | 225 | AD | eGFR | -0.173599 | -0.83771 | 0.37889 | 0.0301367 | 0 | 0.73137 | 0.503684 | 684 | 0.808686903930131 |
| Chr15:38.6Mb | 2048 | 15 | 37962916 | 39238840 | 2586 | 306 | AD | eGFR | 0.107947 | -0.21087 | 0.44988 | 0.0116525 | 0 | 0.20512 | 0.504202 | 685 | 0.808686903930131 |
| Chr8:17.2Mb | 1260 | 8 | 16988031 | 17492672 | 1802 | 255 | AD | eGFR | 0.14785 | -0.32277 | 0.69172 | 0.0218597 | 0 | 0.49745 | 0.504311 | 686 | 0.808686903930131 |
| Chr10:116.2Mb | 1590 | 10 | 115588904 | 116845213 | 2298 | 272 | AD | eGFR | 0.133746 | -0.27749 | 0.57334 | 0.0178881 | 0 | 0.33357 | 0.504603 | 687 | 0.808686903930131 |
| Chr1:220.5Mb | 170 | 1 | 220073133 | 220978151 | 1764 | 175 | AD | eGFR | 0.147132 | -0.3316 | 0.7507 | 0.0216477 | 0 | 0.58574 | 0.506798 | 688 | 0.811024125 |
| Chr19:22.3Mb | 2331 | 19 | 21822654 | 22729408 | 2268 | 146 | AD | eGFR | -0.272868 | -1 | 0.73773 | 0.0744572 | 0 | 1 | 0.508116 | 689 | 0.811953143686502 |
| Chr11:12.9Mb | 1625 | 11 | 12470450 | 13364727 | 2152 | 249 | AD | eGFR | 0.144596 | -0.31325 | 0.6488 | 0.0209079 | 0 | 0.43077 | 0.509555 | 690 | 0.812730515895954 |
| Chr6:161Mb | 1084 | 6 | 160583919 | 161371014 | 1849 | 161 | AD | eGFR | 0.0964126 | -0.21051 | 0.44378 | 0.00929539 | 0 | 0.20447 | 0.510695 | 691 | 0.812730515895954 |
| Chr17:38.1Mb | 2203 | 17 | 37361179 | 38880481 | 2177 | 218 | AD | eGFR | 0.0731232 | -0.14897 | 0.30324 | 0.00534701 | 0 | 0.09226 | 0.510817 | 692 | 0.812730515895954 |
| Chr20:59.8Mb | 2425 | 20 | 59560458 | 60125333 | 1836 | 237 | AD | eGFR | 0.15263 | -0.34707 | 0.73331 | 0.0232958 | 0 | 0.54717 | 0.515669 | 693 | 0.818307556834532 |
| Chr2:10.9Mb | 215 | 2 | 10611655 | 11230349 | 1890 | 249 | AD | eGFR | 0.159969 | -0.36812 | 0.7586 | 0.0255901 | 0 | 0.59587 | 0.516236 | 694 | 0.818307556834532 |
| Chr12:115.1Mb | 1843 | 12 | 114685671 | 115439451 | 1950 | 256 | AD | eGFR | -0.168879 | -0.81999 | 0.40451 | 0.0285201 | 0 | 0.69792 | 0.516552 | 695 | 0.818307556834532 |
| Chr19:5.1Mb | 2312 | 19 | 4741719 | 5478684 | 1860 | 295 | AD | eGFR | 0.167631 | -0.39712 | 0.86946 | 0.0281 | 0 | 0.79533 | 0.520896 | 696 | 0.824003586206897 |
| Chr22:44.8Mb | 2488 | 22 | 44409534 | 45170649 | 2444 | 355 | AD | eGFR | 0.152944 | -0.37409 | 0.76793 | 0.0233918 | 0 | 0.63108 | 0.524562 | 697 | 0.828556130372493 |
| Chr1:99.8Mb | 86 | 1 | 99425001 | 100269207 | 1929 | 203 | AD | eGFR | 0.153712 | -0.38439 | 0.76072 | 0.0236273 | 0 | 0.6193 | 0.525279 | 698 | 0.828556130372493 |
| Chr10:83Mb | 1562 | 10 | 82378049 | 83596350 | 3349 | 252 | AD | eGFR | -0.22205 | -1 | 0.60223 | 0.0493062 | 0 | 1 | 0.527027 | 699 | 0.830124072961373 |
| Chr7:1.9Mb | 1096 | 7 | 1366973 | 2473749 | 2914 | 277 | AD | eGFR | 0.137579 | -0.35333 | 0.7986 | 0.0189281 | 0 | 0.69516 | 0.530039 | 700 | 0.832933223300971 |
| Chr8:105.9Mb | 1334 | 8 | 105370131 | 106500950 | 2182 | 222 | AD | eGFR | -0.162518 | -0.80425 | 0.38926 | 0.0264121 | 0 | 0.68418 | 0.531356 | 701 | 0.832933223300971 |
| Chr12:106.6Mb | 1836 | 12 | 106153788 | 106964977 | 1777 | 237 | AD | eGFR | 0.138697 | -0.32662 | 0.66082 | 0.0192369 | 0 | 0.45907 | 0.531646 | 702 | 0.832933223300971 |
| Chr11:121.9Mb | 1726 | 11 | 121208187 | 122589224 | 2622 | 296 | AD | eGFR | 0.0930749 | -0.21003 | 0.40363 | 0.00866293 | 0 | 0.16421 | 0.531875 | 703 | 0.832933223300971 |
| Chr16:84.8Mb | 2166 | 16 | 84440155 | 85146805 | 2718 | 335 | AD | eGFR | 0.0936404 | -0.21241 | 0.41133 | 0.00876852 | 0 | 0.17082 | 0.533437 | 704 | 0.832933223300971 |
| Chr2:121.6Mb | 313 | 2 | 121073753 | 122026817 | 2161 | 305 | AD | eGFR | -0.0787279 | -0.35133 | 0.17734 | 0.00619808 | 0 | 0.12592 | 0.5344 | 705 | 0.832933223300971 |
| Chr1:201.8Mb | 155 | 1 | 201067953 | 202583884 | 3143 | 352 | AD | eGFR | 0.0908774 | -0.20704 | 0.40755 | 0.00825869 | 0 | 0.16755 | 0.535742 | 706 | 0.832933223300971 |
| Chr6:42.9Mb | 977 | 6 | 42103739 | 43770626 | 2844 | 286 | AD | eGFR | -0.111204 | -0.58398 | 0.27868 | 0.0123664 | 0 | 0.37669 | 0.53647 | 707 | 0.832933223300971 |

| locus_label | locus | chr | start | stop | n.snps | n.pcs | phen1 | phen2 | rho | rho.lower | rho.upper | r2 | r2.lower | r2.upper | p | rank | FDR |
|---|---|---|---|---|---|---|---|---|---|---|---|---|---|---|---|---|---|
| Chr5:79.7Mb | 841 | 5 | 79005159 | 80448225 | 3388 | 303 | AD | eGFR | 0.223574 | -0.62996 | 1 | 0.0499855 | 0 | 1 | 0.537172 | 708 | 0.832933223300971 |
| Chr3:10.6Mb | 432 | 3 | 9970732 | 11226367 | 3054 | 330 | AD | eGFR | 0.16237 | -0.42787 | 0.85338 | 0.0263642 | 0 | 0.76984 | 0.537593 | 709 | 0.832933223300971 |
| Chr12:98.5Mb | 1829 | 12 | 97814022 | 99244337 | 2925 | 277 | AD | eGFR | 0.126406 | -0.29849 | 0.57723 | 0.0159785 | 0 | 0.34047 | 0.539164 | 710 | 0.832933223300971 |
| Chr8:25.4Mb | 1270 | 8 | 24863244 | 25859277 | 2113 | 260 | AD | eGFR | -0.134752 | -0.64491 | 0.33593 | 0.018158 | 0 | 0.44003 | 0.539496 | 711 | 0.832933223300971 |
| Chr3:105.5Mb | 511 | 3 | 104878978 | 106174685 | 3066 | 242 | AD | eGFR | 0.170352 | -0.42478 | 0.90899 | 0.0290199 | 0 | 0.87266 | 0.539756 | 712 | 0.832933223300971 |
| Chr1:44.3Mb | 40 | 1 | 43512670 | 45167235 | 2920 | 278 | AD | eGFR | -0.123763 | -0.59738 | 0.30986 | 0.0153172 | 0 | 0.36914 | 0.539913 | 713 | 0.832933223300971 |
| Chr12:6.5Mb | 1746 | 12 | 6023701 | 6948954 | 2044 | 287 | AD | eGFR | 0.126705 | -0.307 | 0.61655 | 0.0160541 | 0 | 0.38483 | 0.542102 | 714 | 0.832933223300971 |
| Chr1:180.8Mb | 136 | 1 | 180221609 | 181324205 | 2153 | 224 | AD | eGFR | 0.131368 | -0.32488 | 0.62276 | 0.0172577 | 0 | 0.40616 | 0.542429 | 715 | 0.832933223300971 |
| Chr11:127.8Mb | 1732 | 11 | 127306872 | 128262550 | 2241 | 229 | AD | eGFR | -0.120534 | -0.56532 | 0.298 | 0.0145285 | 0 | 0.33114 | 0.542602 | 716 | 0.832933223300971 |
| Chr12:57.9Mb | 1794 | 12 | 56987106 | 58748139 | 2519 | 226 | AD | eGFR | -0.081055 | -0.36636 | 0.187 | 0.00656991 | 0 | 0.13566 | 0.543208 | 717 | 0.832933223300971 |
| Chr8:27Mb | 1272 | 8 | 26679136 | 27406511 | 2150 | 175 | AD | eGFR | -0.164516 | -0.93292 | 0.42637 | 0.0270656 | 0 | 0.97089 | 0.544058 | 718 | 0.832933223300971 |
| Chr12:24.5Mb | 1764 | 12 | 23923799 | 25058714 | 2749 | 298 | AD | eGFR | -0.149569 | -0.7805 | 0.36174 | 0.0223709 | 0 | 0.6224 | 0.544731 | 719 | 0.832933223300971 |
| Chr2:236.7Mb | 413 | 2 | 236276288 | 237194297 | 1968 | 258 | AD | eGFR | 0.181463 | -0.52599 | 1 | 0.032929 | 0 | 1 | 0.545215 | 720 | 0.832933223300971 |
| Chr3:149.3Mb | 551 | 3 | 148638309 | 149998411 | 3169 | 299 | AD | eGFR | -0.141952 | -0.71011 | 0.36123 | 0.0201502 | 0 | 0.51938 | 0.545454 | 721 | 0.832933223300971 |
| Chr1:164.3Mb | 121 | 1 | 163854568 | 164726579 | 1722 | 226 | AD | eGFR | 0.182544 | -0.50041 | 1 | 0.0333222 | 0 | 1 | 0.547051 | 722 | 0.833977804979253 |
| Chr15:23.4Mb | 2035 | 15 | 22799927 | 24099946 | 861 | 191 | AD | eGFR | -0.146547 | -0.73975 | 0.38535 | 0.021476 | 0 | 0.56762 | 0.547653 | 723 | 0.833977804979253 |
| Chr10:108.3Mb | 1583 | 10 | 107877788 | 108725997 | 1946 | 202 | AD | eGFR | -0.196395 | -1 | 0.53806 | 0.038571 | 0 | 1 | 0.549741 | 724 | 0.835824074277854 |
| Chr10:29.1Mb | 1517 | 10 | 28734423 | 29393749 | 1831 | 217 | AD | eGFR | 0.202831 | -0.59192 | 1 | 0.0411403 | 0 | 1 | 0.551224 | 725 | 0.835824074277854 |
| Chr18:61.4Mb | 2289 | 18 | 60780195 | 62074993 | 3081 | 268 | AD | eGFR | -0.0879952 | -0.39563 | 0.21901 | 0.00774316 | 0 | 0.16002 | 0.551439 | 726 | 0.835824074277854 |
| Chr2:36.5Mb | 240 | 2 | 36120744 | 36867389 | 1898 | 233 | AD | eGFR | -0.185544 | -1 | 0.52307 | 0.0344267 | 0 | 1 | 0.551902 | 727 | 0.835824074277854 |
| Chr20:1.3Mb | 2370 | 20 | 884242 | 1771285 | 2232 | 299 | AD | eGFR | 0.0914212 | -0.21865 | 0.4073 | 0.00835784 | 0 | 0.1705 | 0.554875 | 728 | 0.838495091655267 |
| Chr13:81.3Mb | 1920 | 13 | 80778340 | 81724089 | 2205 | 169 | AD | eGFR | 0.236403 | -0.71186 | 1 | 0.0558865 | 0 | 1 | 0.555234 | 729 | 0.838495091655267 |
| Chr10:95Mb | 1573 | 10 | 94503300 | 95528160 | 2531 | 245 | AD | eGFR | -0.106158 | -0.53822 | 0.2817 | 0.0112694 | 0 | 0.29957 | 0.556062 | 730 | 0.838495091655267 |
| Chr16:71.6Mb | 2149 | 16 | 71062713 | 72089511 | 1654 | 155 | AD | eGFR | 0.132801 | -0.3386 | 0.64306 | 0.0176361 | 0 | 0.42484 | 0.556712 | 731 | 0.838495091655267 |
| Chr8:24.3Mb | 1269 | 8 | 23788495 | 24863243 | 2106 | 204 | AD | eGFR | -0.100844 | -0.49987 | 0.26703 | 0.0101695 | 0 | 0.26483 | 0.558417 | 732 | 0.839914094262295 |
| Chr18:46.1Mb | 2274 | 18 | 45718493 | 46558306 | 2155 | 321 | AD | eGFR | -0.1405 | -0.82197 | 0.38055 | 0.0197402 | 0 | 0.72953 | 0.561218 | 733 | 0.842392095302014 |
| Chr2:69.8Mb | 275 | 2 | 68877775 | 70756283 | 3304 | 297 | AD | eGFR | -0.127186 | -0.66258 | 0.33475 | 0.0161762 | 0 | 0.45415 | 0.563065 | 734 | 0.842392095302014 |
| Chr1:9Mb | 11 | 1 | 8580988 | 9475966 | 1680 | 279 | AD | eGFR | -0.127276 | -0.62347 | 0.33808 | 0.0161992 | 0 | 0.40177 | 0.563278 | 735 | 0.842392095302014 |
| Chr7:64.8Mb | 1158 | 7 | 64216136 | 65294277 | 1650 | 77 | AD | eGFR | 0.161225 | -0.46383 | 0.83361 | 0.0259936 | 0 | 0.73472 | 0.563385 | 736 | 0.842392095302014 |
| Chr2:10.3Mb | 214 | 2 | 9960671 | 10611654 | 1966 | 254 | AD | eGFR | -0.263093 | -1 | 0.95795 | 0.0692181 | 0 | 1 | 0.566979 | 737 | 0.842392095302014 |
| Chr2:29.2Mb | 231 | 2 | 28819511 | 29627932 | 1764 | 221 | AD | eGFR | -0.165271 | -0.96675 | 0.49875 | 0.0273145 | 0 | 1 | 0.567259 | 738 | 0.842392095302014 |
| Chr21:32.7Mb | 2447 | 21 | 32202240 | 33270272 | 2463 | 265 | AD | eGFR | -0.158452 | -0.97342 | 0.47826 | 0.0251071 | 0 | 1 | 0.567626 | 739 | 0.842392095302014 |
| Chr9:2.2Mb | 1373 | 9 | 1874962 | 2441120 | 1676 | 283 | AD | eGFR | -0.13107 | -0.67056 | 0.36622 | 0.0171794 | 0 | 0.4712 | 0.568702 | 740 | 0.842392095302014 |
| Chr10:13.6Mb | 1501 | 10 | 13208062 | 14008135 | 2085 | 311 | AD | eGFR | -0.153989 | -0.84376 | 0.45702 | 0.0237125 | 0 | 0.73741 | 0.568703 | 741 | 0.842392095302014 |
| Chr12:103.6Mb | 1833 | 12 | 102966634 | 104140831 | 2592 | 271 | AD | eGFR | 0.21551 | -0.73965 | 1 | 0.0464444 | 0 | 1 | 0.568786 | 742 | 0.842392095302014 |
| Chr12:121.2Mb | 1849 | 12 | 120567741 | 121817509 | 3006 | 227 | AD | eGFR | -0.129809 | -0.79334 | 0.36492 | 0.0168503 | 0 | 0.69708 | 0.569332 | 743 | 0.842392095302014 |
| Chr7:20.8Mb | 1119 | 7 | 20122180 | 21390644 | 2984 | 312 | AD | eGFR | 0.129012 | -0.35399 | 0.64981 | 0.0166442 | 0 | 0.43643 | 0.569854 | 744 | 0.842392095302014 |

| locus_label | locus | chr | start | stop | n.snps | n.pcs | phen1 | phen2 | rho | rho.lower | rho.upper | r2 | r2.lower | r2.upper | p | rank | FDR |
|---|---|---|---|---|---|---|---|---|---|---|---|---|---|---|---|---|---|
| Chr1:208.9Mb | 160 | 1 | 208162952 | 209711169 | 3186 | 342 | AD | eGFR | 0.173706 | -0.49698 | 1 | 0.0301738 | 0 | 1 | 0.570011 | 745 | 0.842392095302014 |
| Chr9:131.5Mb | 1474 | 9 | 130902728 | 132193798 | 1661 | 238 | AD | eGFR | 0.0917947 | -0.2356 | 0.43314 | 0.00842627 | 0 | 0.19037 | 0.571109 | 746 | 0.842883390080429 |
| Chr10:101.9Mb | 1579 | 10 | 101271452 | 102509298 | 2093 | 221 | AD | eGFR | 0.0999532 | -0.27351 | 0.4847 | 0.00999065 | 0 | 0.24334 | 0.5723 | 747 | 0.843510441767068 |
| Chr17:52.8Mb | 2214 | 17 | 52102650 | 53468006 | 2947 | 275 | AD | eGFR | -0.175571 | -1 | 0.52567 | 0.0308252 | 0 | 1 | 0.573883 | 748 | 0.843592373831776 |
| Chr9:20Mb | 1396 | 9 | 19463293 | 20483880 | 2154 | 292 | AD | eGFR | -0.158903 | -0.97944 | 0.47824 | 0.0252502 | 0 | 1 | 0.573888 | 749 | 0.843592373831776 |
| Chr10:60.6Mb | 1542 | 10 | 59822947 | 61470383 | 2899 | 270 | AD | eGFR | -0.128842 | -0.69172 | 0.35846 | 0.0166004 | 0 | 0.5068 | 0.574845 | 750 | 0.84387246 |
| Chr17:45.4Mb | 2208 | 17 | 44865833 | 45883901 | 1745 | 153 | AD | eGFR | -0.115685 | -0.6021 | 0.32488 | 0.0133831 | 0 | 0.38418 | 0.575926 | 751 | 0.844293003984064 |
| Chr11:134.6Mb | 1739 | 11 | 134351065 | 134946450 | 2307 | 244 | AD | eGFR | 0.242047 | -0.89895 | 1 | 0.058587 | 0 | 1 | 0.576716 | 752 | 0.844293003984064 |
| Chr9:89.7Mb | 1433 | 9 | 89171797 | 90135154 | 2412 | 260 | AD | eGFR | 0.159474 | -0.46973 | 0.94622 | 0.025432 | 0 | 0.94819 | 0.577432 | 753 | 0.844293003984064 |
| Chr14:76.2Mb | 2005 | 14 | 75699502 | 76694201 | 1914 | 256 | AD | eGFR | 0.166629 | -0.52429 | 1 | 0.0277651 | 0 | 1 | 0.579864 | 754 | 0.846724488063661 |
| Chr7:50.3Mb | 1146 | 7 | 49632427 | 50894508 | 3072 | 236 | AD | eGFR | 0.0893772 | -0.23428 | 0.42261 | 0.00798829 | 0 | 0.18424 | 0.580758 | 755 | 0.846906699337748 |
| Chr2:4.4Mb | 207 | 2 | 4016105 | 4766969 | 1960 | 215 | AD | eGFR | 0.135593 | -0.38257 | 0.7147 | 0.0183856 | 0 | 0.53155 | 0.582563 | 756 | 0.848370545454546 |
| Chr3:107Mb | 512 | 3 | 106174686 | 107736519 | 2969 | 282 | AD | eGFR | -0.153143 | -0.95083 | 0.45937 | 0.0234528 | 0 | 0.9973 | 0.583949 | 757 | 0.848370545454546 |
| Chr2:148.4Mb | 337 | 2 | 147252197 | 149568875 | 3436 | 260 | AD | eGFR | 0.107357 | -0.32045 | 0.5853 | 0.0115255 | 0 | 0.36646 | 0.584399 | 758 | 0.848370545454546 |
| Chr20:17.7Mb | 2388 | 20 | 17359291 | 18058524 | 1927 | 251 | AD | eGFR | -0.130121 | -0.76668 | 0.38209 | 0.0169315 | 0 | 0.6329 | 0.584844 | 759 | 0.848370545454546 |
| Chr13:110.4Mb | 1948 | 13 | 109813577 | 110995432 | 3439 | 344 | AD | eGFR | -0.11019 | -0.67829 | 0.33283 | 0.0121418 | 0 | 0.51399 | 0.585892 | 760 | 0.848772489473684 |
| Chr15:90Mb | 2089 | 15 | 89385188 | 90632718 | 2442 | 309 | AD | eGFR | 0.122009 | -0.36033 | 0.63487 | 0.0148862 | 0 | 0.41906 | 0.588742 | 761 | 0.84933978515625 |
| Chr9:86Mb | 1430 | 9 | 85135752 | 86769886 | 3385 | 308 | AD | eGFR | -0.151025 | -0.93439 | 0.47299 | 0.0228086 | 0 | 0.94875 | 0.589468 | 762 | 0.84933978515625 |
| Chr16:18.2Mb | 2120 | 16 | 17343278 | 19104257 | 2492 | 256 | AD | eGFR | -0.158942 | -0.95844 | 0.50185 | 0.0252625 | 0 | 0.96749 | 0.5895 | 763 | 0.84933978515625 |
| Chr4:3Mb | 599 | 4 | 2468936 | 3549229 | 2244 | 231 | AD | eGFR | -0.0816731 | -0.39304 | 0.22717 | 0.0066705 | 0 | 0.15757 | 0.591259 | 764 | 0.84933978515625 |
| Chr2:237.7Mb | 414 | 2 | 237194298 | 238185482 | 2051 | 245 | AD | eGFR | 0.104336 | -0.30102 | 0.53267 | 0.010886 | 0 | 0.29746 | 0.591706 | 765 | 0.84933978515625 |
| Chr6:20.9Mb | 944 | 6 | 20418021 | 21295864 | 1942 | 207 | AD | eGFR | -0.103621 | -0.53914 | 0.30521 | 0.0107374 | 0 | 0.30126 | 0.591973 | 766 | 0.84933978515625 |
| Chr12:54.9Mb | 1792 | 12 | 54371449 | 55416802 | 2080 | 253 | AD | eGFR | -0.188604 | -1 | 0.63897 | 0.0355713 | 0 | 1 | 0.592428 | 767 | 0.84933978515625 |
| Chr5:176.9Mb | 922 | 5 | 176180302 | 177659902 | 2321 | 258 | AD | eGFR | 0.047566 | -0.13123 | 0.22452 | 0.00226252 | 0 | 0.05128 | 0.592455 | 768 | 0.84933978515625 |
| Chr1:175.9Mb | 132 | 1 | 175385221 | 176449096 | 1850 | 211 | AD | eGFR | -0.139603 | -0.86034 | 0.44553 | 0.019489 | 0 | 0.80431 | 0.593605 | 769 | 0.849881801040312 |
| Chr3:186.2Mb | 583 | 3 | 185709997 | 186602045 | 1936 | 303 | AD | eGFR | -0.0530405 | -0.24989 | 0.14508 | 0.0028133 | 0 | 0.06434 | 0.595246 | 770 | 0.850691719844358 |
| Chr6:41.4Mb | 976 | 6 | 40778402 | 42103738 | 2858 | 325 | AD | eGFR | -0.0722035 | -0.35234 | 0.20448 | 0.00521334 | 0 | 0.12737 | 0.595716 | 771 | 0.850691719844358 |
| Chr6:105.5Mb | 1036 | 6 | 104951345 | 106053915 | 1932 | 212 | AD | eGFR | -0.122865 | -0.66424 | 0.37191 | 0.0150959 | 0 | 0.46078 | 0.597651 | 772 | 0.852349418393782 |
| Chr18:20.8Mb | 2255 | 18 | 20009697 | 21622716 | 2596 | 227 | AD | eGFR | 0.16357 | -0.5448 | 1 | 0.026755 | 0 | 1 | 0.601297 | 773 | 0.856439840879689 |
| Chr5:179.2Mb | 924 | 5 | 178595253 | 179794710 | 2927 | 345 | AD | eGFR | 0.137349 | -0.44144 | 0.81193 | 0.0188648 | 0 | 0.69151 | 0.60213 | 774 | 0.856518255813954 |
| Chr11:120.6Mb | 1725 | 11 | 120064529 | 121208186 | 2335 | 296 | AD | eGFR | 0.0959757 | -0.27794 | 0.50179 | 0.00921134 | 0 | 0.25719 | 0.603703 | 775 | 0.857647745806452 |
| Chr4:146.7Mb | 731 | 4 | 146174872 | 147161042 | 1991 | 251 | AD | eGFR | 0.0810388 | -0.24353 | 0.42106 | 0.00656728 | 0 | 0.18127 | 0.606742 | 776 | 0.858995201765448 |

| locus_label | locus | chr | start | stop | n.snps | n.pcs | phen1 | phen2 | rho | rho.lower | rho.upper | r2 | r2.lower | r2.upper | p | rank | FDR |
|---|---|---|---|---|---|---|---|---|---|---|---|---|---|---|---|---|---|
| Chr1:39.3Mb | 36 | 1 | 38474037 | 40200950 | 3347 | 315 | AD | eGFR | 0.123312 | -0.41512 | 0.7461 | 0.015206 | 0 | 0.59849 | 0.607207 | 777 | 0.858995201765448 |
| Chr4:89.7Mb | 680 | 4 | 89244555 | 90236971 | 1984 | 220 | AD | eGFR | -0.109086 | -0.619 | 0.37267 | 0.0118998 | 0 | 0.43832 | 0.608647 | 778 | 0.858995201765448 |
| Chr19:39.8Mb | 2345 | 19 | 39370420 | 40171413 | 1816 | 218 | AD | eGFR | 0.134317 | -0.43692 | 0.82313 | 0.0180411 | 0 | 0.7345 | 0.609538 | 779 | 0.858995201765448 |
| Chr18:10.2Mb | 2248 | 18 | 9816165 | 10675548 | 2305 | 325 | AD | eGFR | -0.0909031 | -0.47098 | 0.27083 | 0.00826338 | 0 | 0.22721 | 0.610143 | 780 | 0.858995201765448 |
| Chr1:88.2Mb | 76 | 1 | 87667573 | 88656689 | 1748 | 274 | AD | eGFR | 0.119157 | -0.39231 | 0.70734 | 0.0141983 | 0 | 0.5184 | 0.611179 | 781 | 0.858995201765448 |
| Chr2:8.5Mb | 212 | 2 | 7966710 | 8978798 | 1942 | 295 | AD | eGFR | -0.0684834 | -0.34061 | 0.19826 | 0.00468998 | 0 | 0.11796 | 0.611463 | 782 | 0.858995201765448 |
| Chr17:61.3Mb | 2219 | 17 | 60453121 | 62112373 | 2093 | 227 | AD | eGFR | -0.0819711 | -0.42149 | 0.24236 | 0.00671926 | 0 | 0.18146 | 0.613273 | 783 | 0.858995201765448 |
| Chr1:24.9Mb | 26 | 1 | 23907046 | 25885138 | 3277 | 335 | AD | eGFR | -0.096519 | -0.51458 | 0.28613 | 0.00931593 | 0 | 0.2683 | 0.613303 | 784 | 0.858995201765448 |
| Chr10:25Mb | 1512 | 10 | 24489448 | 25577761 | 1979 | 282 | AD | eGFR | -0.128295 | -0.75489 | 0.41992 | 0.0164597 | 0 | 0.60023 | 0.614401 | 785 | 0.858995201765448 |
| Chr3:182.3Mb | 580 | 3 | 181424324 | 183203155 | 2894 | 314 | AD | eGFR | -0.140137 | -0.93335 | 0.48303 | 0.0196384 | 0 | 0.99723 | 0.616056 | 786 | 0.858995201765448 |
| Chr7:136.9Mb | 1213 | 7 | 136113468 | 137657459 | 2707 | 274 | AD | eGFR | -0.1308 | -0.80699 | 0.4635 | 0.0171087 | 0 | 0.72538 | 0.616162 | 787 | 0.858995201765448 |
| Chr5:175.4Mb | 921 | 5 | 174662886 | 176180301 | 2602 | 325 | AD | eGFR | 0.0763115 | -0.22817 | 0.39317 | 0.00582344 | 0 | 0.1584 | 0.616276 | 788 | 0.858995201765448 |
| Chr2:226.9Mb | 403 | 2 | 226242843 | 227557841 | 2361 | 223 | AD | eGFR | -0.0707328 | -0.37286 | 0.22049 | 0.00500312 | 0 | 0.14246 | 0.616957 | 789 | 0.858995201765448 |
| Chr17:33Mb | 2199 | 17 | 32356062 | 33614189 | 2607 | 285 | AD | eGFR | -0.118748 | -0.68517 | 0.39416 | 0.0141011 | 0 | 0.49709 | 0.617479 | 790 | 0.858995201765448 |
| Chr1:150.4Mb | 109 | 1 | 149716038 | 151146407 | 1803 | 150 | AD | eGFR | 0.0671554 | -0.2059 | 0.36021 | 0.00450985 | 0 | 0.13434 | 0.617519 | 791 | 0.858995201765448 |
| Chr5:143.1Mb | 894 | 5 | 142532957 | 143605908 | 1929 | 229 | AD | eGFR | -0.104137 | -0.57206 | 0.33845 | 0.0108446 | 0 | 0.34178 | 0.618391 | 792 | 0.858995201765448 |
| Chr9:88.5Mb | 1432 | 9 | 87815450 | 89171796 | 2439 | 246 | AD | eGFR | 0.0729085 | -0.2252 | 0.37231 | 0.00531564 | 0 | 0.1421 | 0.618695 | 793 | 0.858995201765448 |
| Chr14:95.4Mb | 2021 | 14 | 94892241 | 95946792 | 3461 | 353 | AD | eGFR | -0.108542 | -0.62296 | 0.35171 | 0.0117814 | 0 | 0.41274 | 0.622408 | 794 | 0.862005542713568 |
| Chr12:112.8Mb | 1841 | 12 | 111592382 | 113947983 | 2955 | 248 | AD | eGFR | -0.097514 | -0.54346 | 0.31424 | 0.00950898 | 0 | 0.30536 | 0.62268 | 795 | 0.862005542713568 |
| Chr10:89.2Mb | 1568 | 10 | 88443653 | 89971628 | 2425 | 255 | AD | eGFR | -0.086371 | -0.45357 | 0.27501 | 0.00745995 | 0 | 0.21391 | 0.623212 | 796 | 0.862005542713568 |
| Chr13:19.3Mb | 1863 | 13 | 19020095 | 19648628 | 908 | 106 | AD | eGFR | 0.214215 | -0.90336 | 1 | 0.045888 | 0 | 1 | 0.625594 | 797 | 0.864214547051443 |
| Chr1:171.8Mb | 129 | 1 | 171044256 | 172465153 | 3283 | 233 | AD | eGFR | -0.0870893 | -0.49819 | 0.28112 | 0.00758455 | 0 | 0.26152 | 0.626993 | 798 | 0.864561972465582 |
| Chr1:231.7Mb | 180 | 1 | 231287765 | 232177704 | 1738 | 228 | AD | eGFR | 0.130609 | -0.48617 | 0.82323 | 0.0170588 | 0 | 0.75431 | 0.627416 | 799 | 0.864561972465582 |
| Chr4:155.8Mb | 738 | 4 | 155044410 | 156520288 | 2984 | 280 | AD | eGFR | 0.180512 | -0.74558 | 1 | 0.0325847 | 0 | 1 | 0.630119 | 800 | 0.86568939276808 |
| Chr4:101.8Mb | 691 | 4 | 101132996 | 102544803 | 3075 | 231 | AD | eGFR | 0.129497 | -0.48006 | 0.84923 | 0.0167695 | 0 | 0.77316 | 0.630134 | 801 | 0.86568939276808 |
| Chr2:153.2Mb | 340 | 2 | 152497376 | 153999309 | 2980 | 226 | AD | eGFR | 0.135403 | -0.48537 | 0.8702 | 0.0183341 | 0 | 0.80937 | 0.630593 | 802 | 0.86568939276808 |
| Chr8:66.5Mb | 1302 | 8 | 66018205 | 66960438 | 1909 | 164 | AD | eGFR | -0.118495 | -0.73901 | 0.42689 | 0.014041 | 0 | 0.59473 | 0.632636 | 803 | 0.867412498132005 |
| Chr13:43.4Mb | 1886 | 13 | 42950158 | 43931931 | 2014 | 255 | AD | eGFR | 0.0944487 | -0.32268 | 0.53239 | 0.00892055 | 0 | 0.29644 | 0.635737 | 804 | 0.870162919354839 |
| Chr13:75.2Mb | 1914 | 13 | 74437636 | 75882384 | 3402 | 299 | AD | eGFR | -0.0799501 | -0.43877 | 0.26285 | 0.00639201 | 0 | 0.19722 | 0.636888 | 805 | 0.870162919354839 |
| Chr13:40.5Mb | 1883 | 13 | 40091624 | 41003943 | 2003 | 269 | AD | eGFR | -0.141641 | -0.99718 | 0.54195 | 0.0200622 | 0 | 1 | 0.637013 | 806 | 0.870162919354839 |
| Chr10:124.4Mb | 1597 | 10 | 123856185 | 124894142 | 2549 | 268 | AD | eGFR | -0.113093 | -0.68269 | 0.42412 | 0.01279 | 0 | 0.51244 | 0.64201 | 807 | 0.875902118959108 |
| Chr4:112.3Mb | 699 | 4 | 111733580 | 112830229 | 1954 | 204 | AD | eGFR | 0.11867 | -0.45202 | 0.83981 | 0.0140827 | 0 | 0.79911 | 0.643332 | 808 | 0.87661947029703 |

| locus_label | locus | chr | start | stop | n.snps | n.pcs | phen1 | phen2 | rho | rho.lower | rho.upper | r2 | r2.lower | r2.upper | p | rank | FDR |
|---|---|---|---|---|---|---|---|---|---|---|---|---|---|---|---|---|---|
| Chr3:87Mb | 497 | 3 | 86660980 | 87411258 | 1721 | 128 | AD | eGFR | -0.160139 | -1 | 0.70113 | 0.0256444 | 0 | 1 | 0.64592 | 809 | 0.879057997527812 |
| Chr13:32.2Mb | 1875 | 13 | 31433219 | 32986686 | 3457 | 319 | AD | eGFR | -0.114828 | -0.73309 | 0.44496 | 0.0131854 | 0 | 0.5822 | 0.64821 | 810 | 0.880384579531443 |
| Chr16:21.5Mb | 2122 | 16 | 20351262 | 22667048 | 2324 | 258 | AD | eGFR | 0.0380622 | -0.1288 | 0.20393 | 0.00144873 | 0 | 0.04298 | 0.648494 | 811 | 0.880384579531443 |
| Chr2:71.1Mb | 276 | 2 | 70756284 | 71385813 | 1791 | 225 | AD | eGFR | 0.129383 | -0.5244 | 0.91281 | 0.0167401 | 0 | 0.93841 | 0.651474 | 812 | 0.880832145985401 |
| Chr4:39.4Mb | 639 | 4 | 38984839 | 39820061 | 1800 | 182 | AD | eGFR | -0.141809 | -1 | 0.60549 | 0.0201097 | 0 | 1 | 0.65187 | 813 | 0.880832145985401 |
| Chr9:90.5Mb | 1434 | 9 | 90135155 | 90890979 | 1916 | 215 | AD | eGFR | -0.0962499 | -0.60357 | 0.35909 | 0.00926404 | 0 | 0.39011 | 0.652698 | 814 | 0.880832145985401 |
| Chr7:56.8Mb | 1154 | 7 | 56303514 | 57383657 | 2216 | 128 | AD | eGFR | -0.165051 | -1 | 0.75143 | 0.0272418 | 0 | 1 | 0.65369 | 815 | 0.880832145985401 |
| Chr11:5.9Mb | 1617 | 11 | 5533058 | 6180840 | 2182 | 215 | AD | eGFR | 0.0944715 | -0.3483 | 0.59571 | 0.00892486 | 0 | 0.37438 | 0.653723 | 816 | 0.880832145985401 |
| Chr2:74.3Mb | 279 | 2 | 73544291 | 75148103 | 2220 | 247 | AD | eGFR | -0.0491378 | -0.27643 | 0.16751 | 0.00241452 | 0 | 0.07951 | 0.655096 | 817 | 0.880832145985401 |
| Chr1:119.9Mb | 105 | 1 | 119408889 | 120345066 | 1847 | 210 | AD | eGFR | 0.15455 | -0.70046 | 1 | 0.0238856 | 0 | 1 | 0.656042 | 818 | 0.880832145985401 |
| Chr10:135.2Mb | 1609 | 10 | 134856055 | 135524372 | 1355 | 188 | AD | eGFR | 0.0731981 | -0.26616 | 0.42881 | 0.00535796 | 0 | 0.19533 | 0.657056 | 819 | 0.880832145985401 |
| Chr3:195.5Mb | 592 | 3 | 195196790 | 195710290 | 794 | 159 | AD | eGFR | -0.101961 | -0.68523 | 0.40898 | 0.010396 | 0 | 0.51965 | 0.657175 | 820 | 0.880832145985401 |
| Chr7:153.8Mb | 1226 | 7 | 153241229 | 154407487 | 2333 | 294 | AD | eGFR | 0.0891015 | -0.33366 | 0.52784 | 0.00793909 | 0 | 0.29468 | 0.657487 | 821 | 0.880832145985401 |
| Chr22:36.7Mb | 2481 | 22 | 36135969 | 37364004 | 2698 | 308 | AD | eGFR | 0.0766746 | -0.27227 | 0.44804 | 0.005879 | 0 | 0.20881 | 0.657624 | 822 | 0.880832145985401 |
| Chr11:34.3Mb | 1646 | 11 | 33905328 | 34672697 | 1861 | 254 | AD | eGFR | -0.114121 | -0.73943 | 0.4392 | 0.0130236 | 0 | 0.58582 | 0.659053 | 823 | 0.881673575941677 |
| Chr5:33.3Mb | 805 | 5 | 32727915 | 33971056 | 2899 | 250 | AD | eGFR | 0.135374 | -0.60114 | 0.99744 | 0.0183262 | 0 | 1 | 0.660409 | 824 | 0.881791963768116 |
| Chr15:59.3Mb | 2064 | 15 | 58712955 | 59901116 | 2347 | 285 | AD | eGFR | 0.0631126 | -0.2314 | 0.35947 | 0.00398321 | 0 | 0.13331 | 0.661822 | 825 | 0.881791963768116 |
| Chr18:53.1Mb | 2281 | 18 | 52512524 | 53762996 | 2059 | 223 | AD | eGFR | -0.124478 | -0.88592 | 0.51088 | 0.0154947 | 0 | 0.85375 | 0.662634 | 826 | 0.881791963768116 |
| Chr9:134.6Mb | 1477 | 9 | 134141937 | 135135887 | 1878 | 264 | AD | eGFR | -0.105694 | -0.693 | 0.43227 | 0.0111711 | 0 | 0.52653 | 0.663067 | 827 | 0.881791963768116 |
| Chr18:59Mb | 2287 | 18 | 58527237 | 59449422 | 1747 | 260 | AD | eGFR | 0.077818 | -0.2959 | 0.48125 | 0.00605565 | 0 | 0.24889 | 0.663146 | 828 | 0.881791963768116 |
| Chr1:205.5Mb | 158 | 1 | 205009624 | 205917548 | 2202 | 245 | AD | eGFR | 0.0770858 | -0.28075 | 0.44926 | 0.00594222 | 0 | 0.21054 | 0.664541 | 829 | 0.882109261445783 |
| Chr12:131Mb | 1859 | 12 | 130629841 | 131404042 | 2428 | 309 | AD | eGFR | -0.151052 | -1 | 0.72733 | 0.0228168 | 0 | 1 | 0.664987 | 830 | 0.882109261445783 |
| Chr2:16.5Mb | 221 | 2 | 15721704 | 17241115 | 3510 | 342 | AD | eGFR | -0.0632986 | -0.41235 | 0.25028 | 0.00400671 | 0 | 0.18301 | 0.667007 | 831 | 0.883724075812274 |
| Chr1:16.2Mb | 17 | 1 | 15755231 | 16732168 | 1788 | 192 | AD | eGFR | -0.076252 | -0.49654 | 0.31489 | 0.00581437 | 0 | 0.26536 | 0.6702 | 832 | 0.886887259615385 |
| Chr15:81.6Mb | 2082 | 15 | 81077026 | 82191077 | 1791 | 259 | AD | eGFR | -0.0570394 | -0.32964 | 0.2081 | 0.00325349 | 0 | 0.11218 | 0.671643 | 833 | 0.887729823529412 |
| Chr2:30.1Mb | 232 | 2 | 29627933 | 30575619 | 2074 | 262 | AD | eGFR | 0.0854993 | -0.3548 | 0.56812 | 0.00731013 | 0 | 0.34932 | 0.673079 | 834 | 0.888561125899281 |
| Chr4:38.4Mb | 638 | 4 | 37880861 | 38984838 | 2933 | 298 | AD | eGFR | -0.0512779 | -0.29645 | 0.19134 | 0.00262942 | 0 | 0.09114 | 0.676161 | 835 | 0.888814651605232 |
| Chr12:43.5Mb | 1782 | 12 | 43072680 | 43980531 | 2382 | 175 | AD | eGFR | 0.168668 | -0.92068 | 1 | 0.0284488 | 0 | 1 | 0.676987 | 836 | 0.888814651605232 |
| Chr16:19.7Mb | 2121 | 16 | 19104258 | 20351261 | 2881 | 280 | AD | eGFR | -0.0528491 | -0.33217 | 0.21018 | 0.00279303 | 0 | 0.11775 | 0.6775 | 837 | 0.888814651605232 |
| Chr21:39.9Mb | 2453 | 21 | 39252060 | 40480831 | 2676 | 319 | AD | eGFR | -0.106475 | -0.77159 | 0.47396 | 0.011337 | 0 | 0.65955 | 0.678099 | 838 | 0.888814651605232 |
| Chr5:166.1Mb | 914 | 5 | 165289212 | 166863413 | 2973 | 328 | AD | eGFR | -0.0947641 | -0.61173 | 0.39153 | 0.00898024 | 0 | 0.40531 | 0.678104 | 839 | 0.888814651605232 |
| Chr8:102Mb | 1330 | 8 | 101448440 | 102548680 | 2195 | 308 | AD | eGFR | -0.0673287 | -0.40532 | 0.25989 | 0.00453316 | 0 | 0.17025 | 0.678525 | 840 | 0.888814651605232 |
| Chr7:94.4Mb | 1181 | 7 | 93691726 | 95174847 | 2241 | 239 | AD | eGFR | 0.16271 | -0.88822 | 1 | 0.0264747 | 0 | 1 | 0.678922 | 841 | 0.888814651605232 |
| Chr17:41.3Mb | 2205 | 17 | 40233546 | 42348003 | 2737 | 258 | AD | eGFR | 0.0621117 | -0.25517 | 0.38623 | 0.00385786 | 0 | 0.15858 | 0.68494 | 842 | 0.895231850533808 |
| Chr20:57.4Mb | 2422 | 20 | 56951774 | 57924037 | 1901 | 291 | AD | eGFR | -0.0611313 | -0.37796 | 0.23665 | 0.00373704 | 0 | 0.14771 | 0.68545 | 843 | 0.895231850533808 |
| Chr1:111.8Mb | 98 | 1 | 111134063 | 112489248 | 3251 | 294 | AD | eGFR | -0.120291 | -0.91927 | 0.56615 | 0.0144698 | 0 | 0.96829 | 0.68711 | 844 | 0.895940378698225 |

| locus_label | locus | chr | start | stop | n.snps | n.pcs | phen1 | phen2 | rho | rho.lower | rho.upper | r2 | r2.lower | r2.upper | p | rank | FDR |
|---|---|---|---|---|---|---|---|---|---|---|---|---|---|---|---|---|---|
| Chr9:96.3Mb | 1441 | 9 | 95854926 | 96670171 | 2130 | 173 | AD | eGFR | -0.133024 | -1 | 0.66648 | 0.0176955 | 0 | 1 | 0.68762 | 845 | 0.895940378698225 |
| Chr6:44.2Mb | 978 | 6 | 43770627 | 44596897 | 1978 | 311 | AD | eGFR | -0.058243 | -0.40497 | 0.25805 | 0.00339225 | 0 | 0.17997 | 0.689556 | 846 | 0.896622498233216 |
| Chr2:195.9Mb | 376 | 2 | 195300251 | 196446299 | 2470 | 168 | AD | eGFR | -0.113304 | -0.81446 | 0.53168 | 0.0128379 | 0 | 0.76602 | 0.690471 | 847 | 0.896622498233216 |
| Chr9:32.7Mb | 1410 | 9 | 32253284 | 33194892 | 2190 | 224 | AD | eGFR | -0.0762442 | -0.49946 | 0.32763 | 0.00581318 | 0 | 0.26293 | 0.690681 | 848 | 0.896622498233216 |
| Chr19:49Mb | 2355 | 19 | 48573066 | 49415189 | 2192 | 254 | AD | eGFR | 0.0561689 | -0.23237 | 0.34463 | 0.00315494 | 0 | 0.12437 | 0.691401 | 849 | 0.896622498233216 |
| Chr11:44.5Mb | 1656 | 11 | 43949378 | 45019559 | 2690 | 336 | AD | eGFR | 0.114987 | -0.56835 | 0.93741 | 0.013222 | 0 | 1 | 0.692411 | 850 | 0.896875895294118 |
| Chr12:2.5Mb | 1742 | 12 | 1942427 | 3069506 | 2449 | 302 | AD | eGFR | 0.135391 | -0.70932 | 1 | 0.0183308 | 0 | 1 | 0.695305 | 851 | 0.898772661971831 |
| Chr15:37.3Mb | 2047 | 15 | 36583328 | 37962915 | 3063 | 322 | AD | eGFR | -0.0729553 | -0.47267 | 0.31219 | 0.00532247 | 0 | 0.2366 | 0.695508 | 852 | 0.898772661971831 |
| Chr9:17Mb | 1392 | 9 | 16658378 | 17392382 | 1850 | 172 | AD | eGFR | 0.118442 | -0.6129 | 0.94369 | 0.0140285 | 0 | 1 | 0.698849 | 853 | 0.90203135873381 |
| Chr10:62Mb | 1543 | 10 | 61470384 | 62626465 | 2897 | 235 | AD | eGFR | 0.114414 | -0.59885 | 0.9374 | 0.0130906 | 0 | 1 | 0.702681 | 854 | 0.904140174621653 |
| Chr17:16.9Mb | 2190 | 17 | 16405208 | 17326540 | 1685 | 202 | AD | eGFR | 0.0926492 | -0.45602 | 0.72515 | 0.00858387 | 0 | 0.58655 | 0.703543 | 855 | 0.904140174621653 |
| Chr22:22.8Mb | 2467 | 22 | 22404423 | 23143364 | 2036 | 197 | AD | eGFR | 0.0800374 | -0.37866 | 0.59375 | 0.00640598 | 0 | 0.38426 | 0.704545 | 856 | 0.904140174621653 |
| Chr5:135Mb | 889 | 5 | 134556571 | 135344943 | 1931 | 232 | AD | eGFR | -0.110325 | -0.92661 | 0.55732 | 0.0121716 | 0 | 1 | 0.704741 | 857 | 0.904140174621653 |
| Chr3:16.6Mb | 438 | 3 | 16084006 | 17182494 | 2852 | 277 | AD | eGFR | -0.0797541 | -0.5595 | 0.36173 | 0.00636071 | 0 | 0.33744 | 0.704806 | 858 | 0.904140174621653 |
| Chr4:152.8Mb | 736 | 4 | 152162099 | 153520883 | 2753 | 229 | AD | eGFR | 0.118961 | -0.64003 | 0.99312 | 0.0141518 | 0 | 1 | 0.70541 | 859 | 0.904140174621653 |
| Chr2:60Mb | 267 | 2 | 59251997 | 60775066 | 3192 | 312 | AD | eGFR | -0.0934793 | -0.69361 | 0.47145 | 0.00873839 | 0 | 0.54076 | 0.707081 | 860 | 0.90522811744186 |
| Chr10:77.9Mb | 1557 | 10 | 77144962 | 78665481 | 2327 | 265 | AD | eGFR | -0.111984 | -0.91605 | 0.58611 | 0.0125405 | 0 | 0.94896 | 0.708847 | 861 | 0.905555972286374 |
| Chr11:74.9Mb | 1681 | 11 | 74399332 | 75445253 | 1882 | 232 | AD | eGFR | -0.096881 | -0.77176 | 0.48845 | 0.00938592 | 0 | 0.71333 | 0.709042 | 862 | 0.905555972286374 |
| Chr2:181.8Mb | 365 | 2 | 181309940 | 182307877 | 2145 | 192 | AD | eGFR | -0.0805452 | -0.55557 | 0.38633 | 0.00648753 | 0 | 0.33984 | 0.710171 | 863 | 0.905555972286374 |
| Chr11:50.7Mb | 1662 | 11 | 50217009 | 51098366 | 1452 | 12 | AD | eGFR | 0.134127 | -0.78323 | 1 | 0.0179901 | 0 | 1 | 0.711203 | 864 | 0.905555972286374 |
| Chr22:39.5Mb | 2483 | 22 | 38718590 | 40378783 | 2897 | 312 | AD | eGFR | -0.055176 | -0.35459 | 0.23994 | 0.00304439 | 0 | 0.13395 | 0.711557 | 865 | 0.905555972286374 |
| Chr22:19.1Mb | 2464 | 22 | 18535226 | 19635654 | 2130 | 228 | AD | eGFR | -0.0591643 | -0.39015 | 0.27251 | 0.00350042 | 0 | 0.16177 | 0.712272 | 866 | 0.905555972286374 |
| Chr10:130.3Mb | 1603 | 10 | 129831970 | 130694008 | 2447 | 325 | AD | eGFR | -0.0894928 | -0.67128 | 0.46016 | 0.00800897 | 0 | 0.48466 | 0.715319 | 867 | 0.906372943246311 |
| Chr7:40.8Mb | 1137 | 7 | 40261241 | 41415925 | 1932 | 235 | AD | eGFR | 0.0501212 | -0.22404 | 0.32967 | 0.00251213 | 0 | 0.11525 | 0.716614 | 868 | 0.906372943246311 |
| Chr7:6.9Mb | 1102 | 7 | 6716906 | 7167610 | 1237 | 154 | AD | eGFR | -0.0647864 | -0.44875 | 0.32397 | 0.00419728 | 0 | 0.22126 | 0.716808 | 869 | 0.906372943246311 |
| Chr1:85.4Mb | 73 | 1 | 84728348 | 86017267 | 2923 | 279 | AD | eGFR | -0.10359 | -0.88226 | 0.56966 | 0.0107308 | 0 | 0.93222 | 0.717874 | 870 | 0.906372943246311 |
| Chr10:17.8Mb | 1505 | 10 | 17065861 | 18529742 | 3230 | 279 | AD | eGFR | -0.0920522 | -0.74914 | 0.4939 | 0.00847362 | 0 | 0.65947 | 0.720246 | 871 | 0.906372943246311 |
| Chr11:8.5Mb | 1620 | 11 | 7915072 | 9080939 | 2769 | 288 | AD | eGFR | 0.0874749 | -0.47136 | 0.70843 | 0.00765186 | 0 | 0.56662 | 0.721148 | 872 | 0.906372943246311 |
| Chr17:3.1Mb | 2176 | 17 | 2699136 | 3430440 | 1855 | 260 | AD | eGFR | 0.0713361 | -0.34932 | 0.50464 | 0.00508884 | 0 | 0.28255 | 0.721209 | 873 | 0.906372943246311 |
| Chr17:48.1Mb | 2210 | 17 | 47516225 | 48722590 | 2059 | 343 | AD | eGFR | -0.0487752 | -0.32984 | 0.2277 | 0.00237902 | 0 | 0.11536 | 0.721282 | 874 | 0.906372943246311 |
| Chr19:40.6Mb | 2346 | 19 | 40171414 | 41004105 | 1509 | 190 | AD | eGFR | 0.0623832 | -0.30242 | 0.4247 | 0.00389166 | 0 | 0.1924 | 0.722399 | 875 | 0.906372943246311 |
| Chr19:7.7Mb | 2315 | 19 | 7249360 | 8199016 | 2453 | 303 | AD | eGFR | 0.0845891 | -0.44408 | 0.64903 | 0.00715531 | 0 | 0.46633 | 0.723279 | 876 | 0.906372943246311 |
| Chr19:36.1Mb | 2342 | 19 | 35649344 | 36638122 | 2232 | 288 | AD | eGFR | 0.0821102 | -0.45677 | 0.70769 | 0.00674208 | 0 | 0.6135 | 0.723953 | 877 | 0.906372943246311 |
| Chr17:64Mb | 2221 | 17 | 63548725 | 64438387 | 2006 | 182 | AD | eGFR | 0.117862 | -0.70537 | 1 | 0.0138914 | 0 | 1 | 0.724625 | 878 | 0.906372943246311 |
| Chr17:44.2Mb | 2207 | 17 | 43460501 | 44865832 | 2319 | 87 | AD | eGFR | 0.0837306 | -0.4491 | 0.66241 | 0.00701081 | 0 | 0.49469 | 0.724935 | 879 | 0.906372943246311 |
| Chr9:21.1Mb | 1397 | 9 | 20483881 | 21674032 | 2448 | 251 | AD | eGFR | -0.0804289 | -0.60898 | 0.40975 | 0.00646882 | 0 | 0.41617 | 0.725236 | 880 | 0.906372943246311 |
| Chr15:39.9Mb | 2049 | 15 | 39238841 | 40604780 | 2856 | 283 | AD | eGFR | -0.0511951 | -0.35252 | 0.24392 | 0.00262094 | 0 | 0.13137 | 0.725263 | 881 | 0.906372943246311 |
| Chr19:20.8Mb | 2329 | 19 | 20485500 | 21155862 | 1991 | 87 | AD | eGFR | -0.126079 | -1 | 0.74748 | 0.0158958 | 0 | 1 | 0.726663 | 882 | 0.907092928571428 |
| Chr10:86Mb | 1565 | 10 | 85463485 | 86459980 | 2624 | 224 | AD | eGFR | -0.109273 | -0.97136 | 0.63635 | 0.0119405 | 0 | 1 | 0.729076 | 883 | 0.909074378255946 |
| Chr5:37.2Mb | 808 | 5 | 36431968 | 37925487 | 2524 | 242 | AD | eGFR | -0.106957 | -0.94902 | 0.62416 | 0.0114399 | 0 | 1 | 0.729985 | 884 | 0.909178150452489 |
| Chr2:218Mb | 394 | 2 | 21756601 | 21839625 | 2083 | 231 | AD | eGFR | -0.0618832 | -0.50639 | 0.35447 | 0.00382953 | 0 | 0.30248 | 0.731761 | 885 | 0.910175326185101 |

| locus_label | locus | chr | start | stop | n.snps | n.pcs | phen1 | phen2 | rho | rho.lower | rho.upper | r2 | r2.lower | r2.upper | p | rank | FDR |
|---|---|---|---|---|---|---|---|---|---|---|---|---|---|---|---|---|---|
| | | | 2 | 8 | | | | | | | | | | | | | |
| Chr1:221.4Mb | 171 | 1 | 220978152 | 221856864 | 2036 | 166 | AD | eGFR | -0.0692089 | -0.51201 | 0.36264 | 0.00478988 | 0 | 0.28161 | 0.732439 | 886 | 0.910175326185101 |
| Chr13:38.8Mb | 1881 | 13 | 38290690 | 39381743 | 2094 | 203 | AD | eGFR | -0.0944831 | -0.83817 | 0.53916 | 0.00892705 | 0 | 0.80039 | 0.734763 | 887 | 0.912033892897407 |
| Chr20:61.8Mb | 2428 | 20 | 61470769 | 62179241 | 2232 | 281 | AD | eGFR | 0.0711007 | -0.41246 | 0.60507 | 0.00505531 | 0 | 0.42333 | 0.739049 | 888 | 0.916181686516854 |
| Chr3:30.6Mb | 450 | 3 | 29877023 | 31269697 | 3439 | 298 | AD | eGFR | 0.0665009 | -0.3624 | 0.51217 | 0.00442237 | 0 | 0.28882 | 0.740082 | 889 | 0.916181686516854 |
| Chr19:55.4Mb | 2364 | 19 | 55182974 | 55714085 | 1249 | 210 | AD | eGFR | -0.0894316 | -0.78067 | 0.53437 | 0.00799802 | 0 | 0.69888 | 0.740601 | 890 | 0.916181686516854 |
| Chr5:149.3Mb | 898 | 5 | 148662215 | 149850203 | 2342 | 317 | AD | eGFR | -0.0935055 | -0.8312 | 0.56017 | 0.00874328 | 0 | 0.80523 | 0.744451 | 891 | 0.918269754759239 |
| Chr7:46.8Mb | 1142 | 7 | 46247336 | 47253454 | 2822 | 236 | AD | eGFR | -0.0566507 | -0.47401 | 0.32086 | 0.00320931 | 0 | 0.25796 | 0.744614 | 892 | 0.918269754759239 |
| Chr17:55.6Mb | 2216 | 17 | 54950108 | 56245227 | 2676 | 365 | AD | eGFR | -0.0565282 | -0.42473 | 0.30213 | 0.00319544 | 0 | 0.19894 | 0.744791 | 893 | 0.918269754759239 |
| Chr3:56.3Mb | 469 | 3 | 55616626 | 56954878 | 3092 | 234 | AD | eGFR | -0.108717 | -1 | 0.70207 | 0.0118193 | 0 | 1 | 0.749194 | 894 | 0.920009331848552 |
| Chr3:154.2Mb | 555 | 3 | 153406719 | 155046423 | 3437 | 240 | AD | eGFR | -0.0497293 | -0.3626 | 0.25927 | 0.00247301 | 0 | 0.13964 | 0.749621 | 895 | 0.920009331848552 |
| Chr1:115.4Mb | 101 | 1 | 114664388 | 116207297 | 3487 | 280 | AD | eGFR | -0.0534392 | -0.40246 | 0.28676 | 0.00285575 | 0 | 0.17407 | 0.749777 | 896 | 0.920009331848552 |
| Chr8:82.1Mb | 1315 | 8 | 81340232 | 82878769 | 2892 | 266 | AD | eGFR | -0.0777825 | -0.67693 | 0.47155 | 0.00605012 | 0 | 0.53007 | 0.750037 | 897 | 0.920009331848552 |
| Chr1:82.8Mb | 71 | 1 | 82123369 | 83461215 | 3067 | 245 | AD | eGFR | -0.0647849 | -0.5525 | 0.39521 | 0.00419708 | 0 | 0.3538 | 0.75038 | 898 | 0.920009331848552 |
| Chr9:111.7Mb | 1455 | 9 | 111368108 | 112076381 | 1883 | 186 | AD | eGFR | 0.133632 | -1 | 1 | 0.0178576 | 0 | 1 | 0.751306 | 899 | 0.920120028921023 |
| Chr2:131.1Mb | 322 | 2 | 130485399 | 131760638 | 1289 | 176 | AD | eGFR | 0.0922826 | -0.59197 | 0.88231 | 0.00851608 | 0 | 0.91623 | 0.75415 | 900 | 0.92197349833887 |
| Chr20:31.5Mb | 2398 | 20 | 30569660 | 32484506 | 2793 | 274 | AD | eGFR | -0.048167 | -0.36386 | 0.26912 | 0.00232006 | 0 | 0.14582 | 0.75542 | 901 | 0.92197349833887 |
| Chr17:80.8Mb | 2237 | 17 | 80459150 | 81192460 | 1717 | 184 | AD | eGFR | 0.10189 | -0.72084 | 1 | 0.0103815 | 0 | 1 | 0.756046 | 902 | 0.92197349833887 |
| Chr12:122.6Mb | 1850 | 12 | 121817510 | 123396634 | 2453 | 217 | AD | eGFR | 0.0563733 | -0.32026 | 0.44087 | 0.00317795 | 0 | 0.20863 | 0.756169 | 903 | 0.92197349833887 |
| Chr10:103.4Mb | 1580 | 10 | 102509299 | 104206837 | 2405 | 259 | AD | eGFR | -0.0494718 | -0.37958 | 0.28351 | 0.00244746 | 0 | 0.15992 | 0.757203 | 904 | 0.92221294579646 |
| Chr11:33.4Mb | 1645 | 11 | 32830423 | 33905327 | 2231 | 208 | AD | eGFR | 0.066108 | -0.39854 | 0.572 | 0.00437027 | 0 | 0.38134 | 0.761638 | 905 | 0.925944645695364 |
| Chr8:95.6Mb | 1325 | 8 | 94999067 | 96175630 | 2866 | 221 | AD | eGFR | -0.0730233 | -0.63379 | 0.45521 | 0.0053324 | 0 | 0.45048 | 0.761949 | 906 | 0.925944645695364 |
| Chr10:1.6Mb | 1486 | 10 | 1329233 | 1872960 | 1672 | 252 | AD | eGFR | 0.0735107 | -0.47165 | 0.64894 | 0.00540383 | 0 | 0.46917 | 0.76283 | 907 | 0.925993197353914 |
| Chr11:63.7Mb | 1673 | 11 | 62800369 | 64594822 | 2651 | 263 | AD | eGFR | 0.0495058 | -0.30107 | 0.39122 | 0.00245082 | 0 | 0.16756 | 0.76559 | 908 | 0.926808491208791 |
| Chr11:133.1Mb | 1737 | 11 | 132584402 | 133521642 | 2338 | 325 | AD | eGFR | 0.10371 | -0.7807 | 1 | 0.0107557 | 0 | 1 | 0.765979 | 909 | 0.926808491208791 |
| Chr16:30.2Mb | 2128 | 16 | 29043178 | 31384210 | 2015 | 249 | AD | eGFR | -0.0527819 | -0.43061 | 0.30976 | 0.00278593 | 0 | 0.19812 | 0.766027 | 910 | 0.926808491208791 |
| Chr6:25.3Mb | 949 | 6 | 24950380 | 25684629 | 2105 | 203 | AD | eGFR | 0.0607836 | -0.38843 | 0.54375 | 0.00369464 | 0 | 0.33705 | 0.76925 | 911 | 0.929618243421053 |
| Chr5:128.4Mb | 885 | 5 | 128067415 | 128734905 | 1997 | 133 | AD | eGFR | -0.124797 | -1 | 1 | 0.0155742 | 0 | 1 | 0.770038 | 912 | 0.929618243421053 |
| Chr14:101.4Mb | 2027 | 14 | 100786190 | 102004377 | 2718 | 354 | AD | eGFR | -0.038629 | -0.31485 | 0.23422 | 0.0014922 | 0 | 0.10792 | 0.772077 | 913 | 0.930187198030635 |
| Chr4:165.3Mb | 747 | 4 | 164781848 | 165824650 | 2917 | 234 | AD | eGFR | -0.106699 | -1 | 0.82667 | 0.0113847 | 0 | 1 | 0.772199 | 914 | 0.930187198030635 |
| Chr19:16.6Mb | 2324 | 19 | 16079007 | 17045963 | 2029 | 230 | AD | eGFR | -0.0703688 | -0.64673 | 0.49248 | 0.00495177 | 0 | 0.48916 | 0.774339 | 915 | 0.931745616393443 |
| Chr3:168Mb | 569 | 3 | 167118894 | 168972662 | 3196 | 284 | AD | eGFR | -0.0509699 | -0.44584 | 0.31329 | 0.00259793 | 0 | 0.20974 | 0.778305 | 916 | 0.932954996777658 |
| Chr10:22.4Mb | 1510 | 10 | 21477693 | 23365685 | 2715 | 246 | AD | eGFR | -0.0656308 | -0.58777 | 0.45048 | 0.0043074 | 0 | 0.39456 | 0.779549 | 917 | 0.932954996777658 |

| locus_label | locus | chr | start | stop | n.snps | n.pcs | phen1 | phen2 | rho | rho.lower | rho.upper | r2 | r2.lower | r2.upper | p | rank | FDR |
|---|---|---|---|---|---|---|---|---|---|---|---|---|---|---|---|---|---|
| Chr15:46.5Mb | 2053 | 15 | 46022643 | 46884071 | 1796 | 182 | AD | eGFR | -0.0407647 | -0.34397 | 0.26171 | 0.00166176 | 0 | 0.13486 | 0.779727 | 918 | 0.932954996777658 |
| Chr3:196.8Mb | 594 | 3 | 196383153 | 197147990 | 1716 | 237 | AD | eGFR | -0.0684594 | -0.63022 | 0.48513 | 0.00468669 | 0 | 0.46579 | 0.780402 | 919 | 0.932954996777658 |
| Chr15:84.9Mb | 2084 | 15 | 84209761 | 85494083 | 2104 | 212 | AD | eGFR | 0.0478322 | -0.3044 | 0.40702 | 0.00228792 | 0 | 0.1815 | 0.780603 | 920 | 0.932954996777658 |
| Chr16:88.7Mb | 2171 | 16 | 88387229 | 89106304 | 2114 | 306 | AD | eGFR | 0.0405347 | -0.25735 | 0.33887 | 0.00164306 | 0 | 0.12805 | 0.781215 | 921 | 0.932954996777658 |
| Chr2:46.2Mb | 251 | 2 | 45850048 | 46499326 | 1886 | 311 | AD | eGFR | 0.038574 | -0.24393 | 0.32283 | 0.00148795 | 0 | 0.11516 | 0.781403 | 922 | 0.932954996777658 |
| Chr3:170.7Mb | 571 | 3 | 170174579 | 171134849 | 1846 | 238 | AD | eGFR | -0.0556627 | -0.50585 | 0.3672 | 0.00309833 | 0 | 0.2806 | 0.782836 | 923 | 0.932954996777658 |
| Chr7:129.6Mb | 1208 | 7 | 128779174 | 130418704 | 3198 | 289 | AD | eGFR | -0.0414508 | -0.36521 | 0.27189 | 0.00171817 | 0 | 0.15101 | 0.783559 | 924 | 0.932954996777658 |
| Chr6:152.7Mb | 1075 | 6 | 152376500 | 153082361 | 1769 | 219 | AD | eGFR | 0.0868503 | -0.68771 | 0.97353 | 0.00754298 | 0 | 1 | 0.7844 | 925 | 0.932954996777658 |
| Chr2:233.7Mb | 409 | 2 | 233352723 | 234115092 | 1809 | 231 | AD | eGFR | 0.0447207 | -0.28093 | 0.38974 | 0.00199995 | 0 | 0.16787 | 0.785212 | 926 | 0.932954996777658 |
| Chr16:2.7Mb | 2104 | 16 | 1996493 | 3379996 | 2110 | 325 | AD | eGFR | 0.0306107 | -0.19862 | 0.2603 | 0.000937014 | 0 | 0.07601 | 0.787157 | 927 | 0.932954996777658 |
| Chr2:145.2Mb | 335 | 2 | 144503689 | 145977920 | 1861 | 258 | AD | eGFR | -0.0428244 | -0.38839 | 0.29024 | 0.00183393 | 0 | 0.17031 | 0.78717 | 928 | 0.932954996777658 |
| Chr9:80.6Mb | 1426 | 9 | 80137025 | 81161597 | 1713 | 211 | AD | eGFR | 0.101412 | -0.94445 | 1 | 0.0102845 | 0 | 1 | 0.788253 | 929 | 0.932954996777658 |
| Chr8:135.8Mb | 1360 | 8 | 135313674 | 136220342 | 2179 | 254 | AD | eGFR | -0.0611647 | -0.5951 | 0.43733 | 0.00374112 | 0 | 0.39049 | 0.788364 | 930 | 0.932954996777658 |
| Chr17:10.1Mb | 2183 | 17 | 9619357 | 10572617 | 2559 | 311 | AD | eGFR | 0.0429743 | -0.29788 | 0.38108 | 0.00184679 | 0 | 0.16058 | 0.788902 | 931 | 0.932954996777658 |
| Chr6:145.3Mb | 1068 | 6 | 144791456 | 145778282 | 2067 | 172 | AD | eGFR | -0.0694863 | -0.70344 | 0.5267 | 0.00482835 | 0 | 0.59462 | 0.790144 | 932 | 0.932956482315113 |
| Chr4:55.2Mb | 651 | 4 | 54659779 | 55681701 | 1922 | 248 | AD | eGFR | -0.0549035 | -0.51412 | 0.39126 | 0.00301439 | 0 | 0.30544 | 0.790598 | 933 | 0.932956482315113 |
| Chr10:97.5Mb | 1575 | 10 | 97040173 | 98005994 | 2145 | 200 | AD | eGFR | -0.0750678 | -0.85345 | 0.61655 | 0.00563518 | 0 | 0.91789 | 0.793199 | 934 | 0.935023660599572 |
| Chr19:6.9Mb | 2314 | 19 | 6455429 | 7249359 | 2307 | 321 | AD | eGFR | -0.0485449 | -0.44471 | 0.33981 | 0.00235661 | 0 | 0.22179 | 0.796088 | 935 | 0.936285470149254 |
| Chr17:14.9Mb | 2188 | 17 | 14508611 | 15308722 | 2528 | 282 | AD | eGFR | -0.0709013 | -0.75855 | 0.57762 | 0.005027 | 0 | 0.69347 | 0.796751 | 936 | 0.936285470149254 |
| Chr8:122.1Mb | 1348 | 8 | 121554391 | 122622804 | 2440 | 246 | AD | eGFR | 0.0580398 | -0.43253 | 0.58566 | 0.00336862 | 0 | 0.39583 | 0.797536 | 937 | 0.936285470149254 |
| Chr13:22.2Mb | 1866 | 13 | 21540734 | 22795188 | 3128 | 344 | AD | eGFR | 0.0473676 | -0.34085 | 0.43219 | 0.00224369 | 0 | 0.20817 | 0.797671 | 938 | 0.936285470149254 |
| Chr4:109.6Mb | 697 | 4 | 109122459 | 109978982 | 1943 | 208 | AD | eGFR | -0.0562075 | -0.618 | 0.45058 | 0.00315928 | 0 | 0.44172 | 0.800397 | 939 | 0.938484661341853 |
| Chr1:8.1Mb | 10 | 1 | 7711795 | 8580987 | 1875 | 223 | AD | eGFR | -0.0992044 | -1 | 1 | 0.00984152 | 0 | 1 | 0.805293 | 940 | 0.942085982528263 |
| Chr5:1.7Mb | 777 | 5 | 1206610 | 2106885 | 2920 | 324 | AD | eGFR | 0.0676822 | -0.57568 | 0.77253 | 0.00458088 | 0 | 0.74854 | 0.806159 | 941 | 0.942085982528263 |
| Chr20:59.2Mb | 2424 | 20 | 58780895 | 59560457 | 1952 | 248 | AD | eGFR | -0.0851087 | -1 | 0.8504 | 0.00724349 | 0 | 1 | 0.80682 | 942 | 0.942085982528263 |
| Chr6:17Mb | 939 | 6 | 16566883 | 17391994 | 1802 | 253 | AD | eGFR | 0.055646 | -0.46121 | 0.57718 | 0.00309648 | 0 | 0.39138 | 0.807888 | 943 | 0.942085982528263 |
| Chr17:6.8Mb | 2180 | 17 | 6305079 | 7264458 | 2200 | 309 | AD | eGFR | 0.0510923 | -0.39969 | 0.5119 | 0.00261042 | 0 | 0.2986 | 0.808933 | 944 | 0.942085982528263 |
| Chr12:52.4Mb | 1790 | 12 | 51769420 | 53039987 | 3071 | 303 | AD | eGFR | -0.0524766 | -0.55256 | 0.41794 | 0.00275379 | 0 | 0.34271 | 0.809972 | 945 | 0.942085982528263 |
| Chr14:21.4Mb | 1955 | 14 | 20876081 | 21982654 | 2570 | 370 | AD | eGFR | -0.0499329 | -0.50276 | 0.39369 | 0.00249329 | 0 | 0.28892 | 0.810001 | 946 | 0.942085982528263 |
| Chr11:5.2Mb | 1616 | 11 | 4946052 | 5533057 | 2093 | 180 | AD | eGFR | 0.0627291 | -0.52634 | 0.71183 | 0.00393493 | 0 | 0.61412 | 0.811067 | 947 | 0.942085982528263 |
| Chr16:69Mb | 2147 | 16 | 68435729 | 69549149 | 1727 | 208 | AD | eGFR | -0.0393968 | -0.3896 | 0.31165 | 0.00155211 | 0 | 0.1694 | 0.813133 | 948 | 0.942085982528263 |
| Chr13:72.5Mb | 1912 | 13 | 71976863 | 73114509 | 2154 | 258 | AD | eGFR | 0.0337106 | -0.25985 | 0.3341 | 0.00113641 | 0 | 0.12223 | 0.814641 | 949 | 0.942085982528263 |
| Chr20:2.2Mb | 2371 | 20 | 1771286 | 2668108 | 2434 | 304 | AD | eGFR | -0.0553195 | -0.60494 | 0.46152 | 0.00306025 | 0 | 0.42732 | 0.815277 | 950 | 0.942085982528263 |
| Chr1:56.3Mb | 48 | 1 | 55727135 | 56799636 | 2663 | 209 | AD | eGFR | -0.0436734 | -0.44964 | 0.3559 | 0.00190736 | 0 | 0.23417 | 0.816154 | 951 | 0.942085982528263 |
| Chr4:115.8Mb | 702 | 4 | 11530615 | 11620470 | 1991 | 149 | AD | eGFR | -0.0593382 | -0.68427 | 0.52474 | 0.00352102 | 0 | 0.55456 | 0.816682 | 952 | 0.942085982528263 |

| locus_label | locus | chr | start | stop | n.snps | n.pcs | phen1 | phen2 | rho | rho.lower | rho.upper | r2 | r2.lower | r2.upper | p | rank | FDR |
|---|---|---|---|---|---|---|---|---|---|---|---|---|---|---|---|---|---|
| | | | 0 | 3 | | | | | | | | | | | | | |
| Chr15:83.2Mb | 2083 | 15 | 82191078 | 84209760 | 2411 | 235 | AD | eGFR | -0.0503699 | -0.57434 | 0.44419 | 0.00253712 | 0 | 0.38384 | 0.818568 | 953 | 0.942085982528263 |
| Chr4:154.3Mb | 737 | 4 | 153520884 | 155044409 | 3520 | 329 | AD | eGFR | -0.0494987 | -0.54071 | 0.41362 | 0.00245012 | 0 | 0.32843 | 0.818842 | 954 | 0.942085982528263 |
| Chr18:77.6Mb | 2305 | 18 | 77148138 | 78017072 | 2376 | 266 | AD | eGFR | 0.0261012 | -0.20226 | 0.26238 | 0.000681271 | 0 | 0.07512 | 0.81885 | 955 | 0.942085982528263 |
| Chr1:68.2Mb | 58 | 1 | 67761891 | 68633860 | 1839 | 233 | AD | eGFR | 0.0357943 | -0.2922 | 0.36633 | 0.00128123 | 0 | 0.14876 | 0.820225 | 956 | 0.942085982528263 |
| Chr17:5.9Mb | 2179 | 17 | 5573785 | 6305078 | 2139 | 299 | AD | eGFR | 0.0554512 | -0.50775 | 0.63167 | 0.00307483 | 0 | 0.47102 | 0.822236 | 957 | 0.942085982528263 |
| Chr17:42.9Mb | 2206 | 17 | 42348004 | 43460500 | 1889 | 208 | AD | eGFR | -0.0446244 | -0.4747 | 0.3665 | 0.00199134 | 0 | 0.25132 | 0.822485 | 958 | 0.942085982528263 |
| Chr13:107.8Mb | 1945 | 13 | 107037865 | 108521978 | 3461 | 362 | AD | eGFR | 0.0674382 | -0.70628 | 0.90309 | 0.00454792 | 0 | 1 | 0.824354 | 959 | 0.942085982528263 |
| Chr2:231.2Mb | 407 | 2 | 230541093 | 231847386 | 3126 | 289 | AD | eGFR | -0.0632413 | -0.7945 | 0.65331 | 0.00399946 | 0 | 0.86676 | 0.825362 | 960 | 0.942085982528263 |
| Chr20:52Mb | 2415 | 20 | 51533751 | 52411532 | 1818 | 306 | AD | eGFR | -0.0368888 | -0.38128 | 0.30799 | 0.00136079 | 0 | 0.16679 | 0.827142 | 961 | 0.942085982528263 |
| Chr3:150.6Mb | 552 | 3 | 149998412 | 151131307 | 2210 | 261 | AD | eGFR | 0.0511818 | -0.49508 | 0.61623 | 0.00261958 | 0 | 0.47978 | 0.827533 | 962 | 0.942085982528263 |
| Chr6:126.5Mb | 1054 | 6 | 125365055 | 127545459 | 2907 | 279 | AD | eGFR | 0.0499772 | -0.47598 | 0.58927 | 0.00249772 | 0 | 0.44411 | 0.828049 | 963 | 0.942085982528263 |
| Chr8:46.4Mb | 1285 | 8 | 43825634 | 49044813 | 2084 | 86 | AD | eGFR | 0.0637023 | -0.64752 | 0.82344 | 0.00405799 | 0 | 0.84543 | 0.828681 | 964 | 0.942085982528263 |
| Chr4:105.9Mb | 694 | 4 | 105319196 | 106479155 | 2267 | 190 | AD | eGFR | -0.0610079 | -0.78729 | 0.60843 | 0.00372196 | 0 | 0.79261 | 0.828786 | 965 | 0.942085982528263 |
| Chr11:133.9Mb | 1738 | 11 | 133521643 | 134351064 | 1888 | 299 | AD | eGFR | -0.0627841 | -0.86944 | 0.64954 | 0.00394185 | 0 | 1 | 0.828967 | 966 | 0.942085982528263 |
| Chr1:26.9Mb | 27 | 1 | 25885139 | 27992665 | 2841 | 286 | AD | eGFR | -0.0331965 | -0.35892 | 0.28584 | 0.00110201 | 0 | 0.14584 | 0.829659 | 967 | 0.942085982528263 |
| Chr11:27.8Mb | 1640 | 11 | 27018308 | 28577718 | 2261 | 211 | AD | eGFR | -0.0413849 | -0.45057 | 0.37738 | 0.00171271 | 0 | 0.23553 | 0.830292 | 968 | 0.942085982528263 |
| Chr6:8.4Mb | 933 | 6 | 7808324 | 9020710 | 3045 | 247 | AD | eGFR | -0.0582311 | -0.72893 | 0.59011 | 0.00339086 | 0 | 0.67449 | 0.830836 | 969 | 0.942085982528263 |
| Chr3:63.1Mb | 475 | 3 | 62609819 | 63670803 | 2979 | 277 | AD | eGFR | -0.0666136 | -0.91323 | 0.7202 | 0.00443737 | 0 | 1 | 0.831072 | 970 | 0.942085982528263 |
| Chr1:155.7Mb | 113 | 1 | 154685546 | 156813845 | 2452 | 325 | AD | eGFR | -0.0258533 | -0.27856 | 0.22448 | 0.000668393 | 0 | 0.08551 | 0.831902 | 971 | 0.942085982528263 |
| Chr11:119.5Mb | 1724 | 11 | 118901214 | 120064528 | 2362 | 283 | AD | eGFR | -0.0326017 | -0.35363 | 0.28509 | 0.00106287 | 0 | 0.1374 | 0.832356 | 972 | 0.942085982528263 |
| Chr2:171.2Mb | 355 | 2 | 170865766 | 171566482 | 1882 | 265 | AD | eGFR | 0.0624629 | -0.65805 | 0.89279 | 0.00390161 | 0 | 1 | 0.832561 | 973 | 0.942085982528263 |
| Chr22:24.6Mb | 2469 | 22 | 23950733 | 25282436 | 2567 | 250 | AD | eGFR | 0.0477791 | -0.44012 | 0.55546 | 0.00228284 | 0 | 0.35275 | 0.8335 | 974 | 0.942180184804928 |
| Chr10:34.4Mb | 1522 | 10 | 33655459 | 35178224 | 3080 | 308 | AD | eGFR | -0.0798524 | -1 | 0.94113 | 0.00637641 | 0 | 1 | 0.836305 | 975 | 0.944381338461538 |
| Chr17:3.9Mb | 2177 | 17 | 3430441 | 4463971 | 2521 | 320 | AD | eGFR | 0.0319661 | -0.29606 | 0.36521 | 0.00102183 | 0 | 0.15027 | 0.839412 | 976 | 0.946918659836066 |
| Chr14:53.6Mb | 1985 | 14 | 52760554 | 54359598 | 2602 | 283 | AD | eGFR | 0.0334631 | -0.32103 | 0.3993 | 0.00111978 | 0 | 0.18259 | 0.843891 | 977 | 0.950996920163767 |
| Chr6:132.2Mb | 1059 | 6 | 131457165 | 132890694 | 2519 | 276 | AD | eGFR | -0.0265358 | -0.31212 | 0.25122 | 0.00070415 | 0 | 0.10724 | 0.848418 | 978 | 0.953450367139959 |
| Chr4:129Mb | 717 | 4 | 128321877 | 129625376 | 1863 | 190 | AD | eGFR | -0.0284278 | -0.34015 | 0.27983 | 0.000808139 | 0 | 0.13255 | 0.849078 | 979 | 0.953450367139959 |
| Chr5:150.4Mb | 899 | 5 | 149850204 | 150981934 | 2566 | 253 | AD | eGFR | -0.0529101 | -0.75014 | 0.596 | 0.00279948 | 0 | 0.72028 | 0.849601 | 980 | 0.953450367139959 |
| Chr5:157.8Mb | 907 | 5 | 157191082 | 158484775 | 2769 | 248 | AD | eGFR | 0.0535422 | -0.60668 | 0.76704 | 0.00286677 | 0 | 0.72199 | 0.850665 | 981 | 0.953450367139959 |
| Chr11:110.5Mb | 1717 | 11 | 109891838 | 111117780 | 2517 | 241 | AD | eGFR | -0.055006 | -0.8443 | 0.65172 | 0.00302566 | 0 | 0.90679 | 0.852754 | 982 | 0.953450367139959 |

| locus_label | locus | chr | start | stop | n.snps | n.pcs | phen1 | phen2 | rho | rho.lower | rho.upper | r2 | r2.lower | r2.upper | p | rank | FDR |
|---|---|---|---|---|---|---|---|---|---|---|---|---|---|---|---|---|---|
| Chr3:126.7Mb | 532 | 3 | 125992651 | 127495173 | 3408 | 294 | AD | eGFR | -0.0385336 | -0.48945 | 0.39322 | 0.00148484 | 0 | 0.28126 | 0.853035 | 983 | 0.953450367139959 |
| Chr2:83.7Mb | 289 | 2 | 82941456 | 84395899 | 2958 | 160 | AD | eGFR | -0.0628503 | -1 | 0.8092 | 0.00395016 | 0 | 1 | 0.853601 | 984 | 0.953450367139959 |
| Chr2:34Mb | 236 | 2 | 33735319 | 34239776 | 1681 | 175 | AD | eGFR | 0.0628196 | -0.84388 | 1 | 0.0039463 | 0 | 1 | 0.853862 | 985 | 0.953450367139959 |
| Chr15:41.5Mb | 2050 | 15 | 40604781 | 42332685 | 2825 | 244 | AD | eGFR | -0.0245994 | -0.3033 | 0.24865 | 0.000605132 | 0 | 0.10608 | 0.853862 | 986 | 0.953450367139959 |
| Chr2:240Mb | 417 | 2 | 239634351 | 240325600 | 1749 | 268 | AD | eGFR | -0.0463888 | -0.72677 | 0.56117 | 0.00215192 | 0 | 0.7025 | 0.854877 | 987 | 0.953616592705167 |
| Chr9:107.6Mb | 1451 | 9 | 107061020 | 108203095 | 3139 | 268 | AD | eGFR | 0.0364487 | -0.40674 | 0.5126 | 0.00132851 | 0 | 0.31518 | 0.855872 | 988 | 0.953760194331984 |
| Chr5:45.8Mb | 815 | 5 | 45305567 | 46388848 | 1461 | 47 | AD | eGFR | -0.0511577 | -0.78473 | 0.64367 | 0.00261711 | 0 | 0.81556 | 0.860523 | 989 | 0.95627288407258 |
| Chr8:55.8Mb | 1292 | 8 | 55275355 | 56346878 | 2263 | 209 | AD | eGFR | 0.0592587 | -0.81185 | 1 | 0.00351159 | 0 | 1 | 0.861104 | 990 | 0.95627288407258 |
| Chr5:57.4Mb | 824 | 5 | 56896891 | 57895012 | 2254 | 194 | AD | eGFR | 0.0497632 | -0.63246 | 0.75177 | 0.00247638 | 0 | 0.74015 | 0.861185 | 991 | 0.95627288407258 |
| Chr7:47.8Mb | 1143 | 7 | 47253455 | 48363585 | 2681 | 285 | AD | eGFR | -0.0494779 | -0.78374 | 0.66597 | 0.00244807 | 0 | 0.85139 | 0.861601 | 992 | 0.95627288407258 |
| Chr19:0.4Mb | 2306 | 19 | 64705 | 762046 | 1451 | 297 | AD | eGFR | 0.0163689 | -0.17418 | 0.20692 | 0.000267942 | 0 | 0.04807 | 0.863139 | 993 | 0.95629683201581 |
| Chr6:109.8Mb | 1039 | 6 | 109043245 | 110647842 | 2979 | 221 | AD | eGFR | 0.0297757 | -0.32578 | 0.38978 | 0.000886594 | 0 | 0.17739 | 0.864105 | 994 | 0.95629683201581 |
| Chr2:119.1Mb | 311 | 2 | 118663654 | 119635489 | 2210 | 206 | AD | eGFR | 0.0437409 | -0.5277 | 0.6509 | 0.00191326 | 0 | 0.51092 | 0.865388 | 995 | 0.95629683201581 |
| Chr1:90.3Mb | 78 | 1 | 89753965 | 90941549 | 1921 | 248 | AD | eGFR | 0.0454316 | -0.58799 | 0.71374 | 0.00206403 | 0 | 0.6855 | 0.866196 | 996 | 0.95629683201581 |
| Chr8:16.5Mb | 1259 | 8 | 15999329 | 16988030 | 2713 | 272 | AD | eGFR | 0.0579545 | -0.84648 | 1 | 0.00335873 | 0 | 1 | 0.866484 | 997 | 0.95629683201581 |
| Chr9:137.2Mb | 1480 | 9 | 136770927 | 137593527 | 2083 | 289 | AD | eGFR | -0.0204934 | -0.26395 | 0.2297 | 0.00041998 | 0 | 0.08157 | 0.866855 | 998 | 0.95629683201581 |
| Chr20:28.4Mb | 2397 | 20 | 26318859 | 30569659 | 1295 | 96 | AD | eGFR | -0.0423611 | -0.66002 | 0.54548 | 0.00179446 | 0 | 0.55167 | 0.870414 | 999 | 0.95629683201581 |
| Chr18:40.9Mb | 2270 | 18 | 40471408 | 41425454 | 1964 | 181 | AD | eGFR | -0.0639679 | -1 | 1 | 0.0040919 | 0 | 1 | 0.870801 | 1000 | 0.95629683201581 |
| Chr12:76Mb | 1810 | 12 | 75563969 | 76524573 | 1952 | 250 | AD | eGFR | 0.0360499 | -0.47444 | 0.57701 | 0.0012996 | 0 | 0.42746 | 0.871403 | 1001 | 0.95629683201581 |
| Chr18:7.3Mb | 2245 | 18 | 6756497 | 7862479 | 2439 | 322 | AD | eGFR | -0.0513157 | -0.94313 | 0.74824 | 0.0026333 | 0 | 1 | 0.872243 | 1002 | 0.95629683201581 |
| Chr2:225.6Mb | 402 | 2 | 224938732 | 226242842 | 3131 | 221 | AD | eGFR | -0.0554331 | -1 | 0.85336 | 0.00307283 | 0 | 1 | 0.872416 | 1003 | 0.95629683201581 |
| Chr6:130.2Mb | 1057 | 6 | 129850179 | 130550137 | 1677 | 236 | AD | eGFR | -0.0350945 | -0.55175 | 0.45557 | 0.00123163 | 0 | 0.38893 | 0.872606 | 1004 | 0.95629683201581 |
| Chr4:80.5Mb | 673 | 4 | 79880102 | 81206182 | 2648 | 220 | AD | eGFR | -0.0249615 | -0.35375 | 0.3041 | 0.000623074 | 0 | 0.14807 | 0.873152 | 1005 | 0.95629683201581 |
| Chr6:150Mb | 1072 | 6 | 149278799 | 150635315 | 3065 | 269 | AD | eGFR | 0.0415436 | -0.55189 | 0.64338 | 0.00172587 | 0 | 0.55223 | 0.87428 | 1006 | 0.95629683201581 |
| Chr19:29.9Mb | 2336 | 19 | 29389091 | 30375792 | 2666 | 271 | AD | eGFR | -0.0493685 | -0.88975 | 0.72636 | 0.00243725 | 0 | 1 | 0.87662 | 1007 | 0.95629683201581 |
| Chr4:81.9Mb | 674 | 4 | 81206183 | 82655623 | 2463 | 220 | AD | eGFR | 0.0369595 | -0.53792 | 0.6294 | 0.00136601 | 0 | 0.57485 | 0.877353 | 1008 | 0.95629683201581 |
| Chr7:104.7Mb | 1190 | 7 | 104109232 | 105312563 | 1994 | 229 | AD | eGFR | 0.0506354 | -0.79652 | 0.93651 | 0.00256394 | 0 | 1 | 0.877612 | 1009 | 0.95629683201581 |
| Chr7:114.3Mb | 1197 | 7 | 113339387 | 115321301 | 2800 | 252 | AD | eGFR | -0.0499349 | -0.91717 | 0.77872 | 0.00249349 | 0 | 1 | 0.878302 | 1010 | 0.95629683201581 |

| locus_label | locus | chr | start | stop | n.snps | n.pcs | phen1 | phen2 | rho | rho.lower | rho.upper | r2 | r2.lower | r2.upper | p | rank | FDR |
|---|---|---|---|---|---|---|---|---|---|---|---|---|---|---|---|---|---|
| Chr17:35.3Mb | 2201 | 17 | 34474612 | 36116884 | 2017 | 293 | AD | eGFR | 0.0236861 | -0.29376 | 0.34704 | 0.000561033 | 0 | 0.14274 | 0.878583 | 1011 | 0.95629683201581 |
| Chr2:169.7Mb | 353 | 2 | 169224446 | 170155561 | 1920 | 246 | AD | eGFR | -0.0221484 | -0.33839 | 0.28814 | 0.000490552 | 0 | 0.14095 | 0.878994 | 1012 | 0.95629683201581 |
| Chr21:41.8Mb | 2455 | 21 | 41344743 | 42187152 | 2395 | 277 | AD | eGFR | -0.0368811 | -0.61749 | 0.5301 | 0.00136022 | 0 | 0.47515 | 0.880364 | 1013 | 0.956841820335637 |
| Chr8:139.9Mb | 1364 | 8 | 139404632 | 140355988 | 2464 | 277 | AD | eGFR | -0.0375951 | -0.67604 | 0.55596 | 0.00141339 | 0 | 0.57137 | 0.883977 | 1014 | 0.95694477179236 |
| Chr2:7.5Mb | 211 | 2 | 7108209 | 7966709 | 2012 | 211 | AD | eGFR | 0.0353865 | -0.53111 | 0.60199 | 0.00125221 | 0 | 0.47161 | 0.884583 | 1015 | 0.95694477179236 |
| Chr10:80.6Mb | 1559 | 10 | 79952997 | 81190573 | 3019 | 350 | AD | eGFR | 0.028397 | -0.37527 | 0.46685 | 0.000806387 | 0 | 0.25148 | 0.884939 | 1016 | 0.95694477179236 |
| Chr2:120.4Mb | 312 | 2 | 119635490 | 121073752 | 2872 | 259 | AD | eGFR | -0.0252391 | -0.40357 | 0.34009 | 0.000637012 | 0 | 0.1902 | 0.885008 | 1017 | 0.95694477179236 |
| Chr20:9.8Mb | 2379 | 20 | 9279551 | 10279603 | 2302 | 291 | AD | eGFR | -0.0361796 | -0.61314 | 0.54757 | 0.00130897 | 0 | 0.5108 | 0.886013 | 1018 | 0.95694477179236 |
| Chr1:21.8Mb | 23 | 1 | 21416122 | 22145629 | 1859 | 212 | AD | eGFR | 0.0322949 | -0.4638 | 0.53179 | 0.00104296 | 0 | 0.35252 | 0.886599 | 1019 | 0.95694477179236 |
| Chr11:17Mb | 1628 | 11 | 16383387 | 17583948 | 2331 | 217 | AD | eGFR | -0.0280327 | -0.44817 | 0.37912 | 0.00078583 | 0 | 0.24194 | 0.886703 | 1020 | 0.95694477179236 |
| Chr7:26.5Mb | 1125 | 7 | 25671576 | 27351286 | 3209 | 281 | AD | eGFR | -0.0207899 | -0.32244 | 0.28149 | 0.00043222 | 0 | 0.12051 | 0.887412 | 1021 | 0.95694477179236 |
| Chr3:188.7Mb | 585 | 3 | 187939200 | 189451805 | 3174 | 338 | AD | eGFR | -0.0222001 | -0.35187 | 0.30037 | 0.000492843 | 0 | 0.14408 | 0.889391 | 1022 | 0.957340445853659 |
| Chr19:54.3Mb | 2362 | 19 | 54042241 | 54602370 | 1444 | 281 | AD | eGFR | 0.021071 | -0.28812 | 0.33393 | 0.000443986 | 0 | 0.13266 | 0.890131 | 1023 | 0.957340445853659 |
| Chr15:92.1Mb | 2091 | 15 | 91593624 | 92675405 | 2475 | 318 | AD | eGFR | 0.0322424 | -0.4971 | 0.58255 | 0.00103957 | 0 | 0.43625 | 0.89103 | 1024 | 0.957340445853659 |
| Chr2:20.6Mb | 224 | 2 | 20064518 | 21084898 | 2354 | 286 | AD | eGFR | 0.035448 | -0.55784 | 0.64448 | 0.00125656 | 0 | 0.5376 | 0.891257 | 1025 | 0.957340445853659 |
| Chr6:52.2Mb | 986 | 6 | 51826424 | 52598879 | 1958 | 225 | AD | eGFR | -0.0292069 | -0.54483 | 0.45229 | 0.000853045 | 0 | 0.36585 | 0.895062 | 1026 | 0.959051325875486 |
| Chr17:67.5Mb | 2224 | 17 | 66757557 | 68176219 | 2351 | 247 | AD | eGFR | 0.0352596 | -0.60795 | 0.64012 | 0.00124324 | 0 | 0.56708 | 0.895245 | 1027 | 0.959051325875486 |
| Chr2:200.5Mb | 379 | 2 | 199654970 | 201324132 | 2190 | 215 | AD | eGFR | -0.0324719 | -0.61651 | 0.49756 | 0.00105443 | 0 | 0.43759 | 0.895463 | 1028 | 0.959051325875486 |
| Chr4:19.4Mb | 618 | 4 | 18753264 | 20086838 | 3460 | 199 | AD | eGFR | 0.0388283 | -0.70665 | 0.7916 | 0.00150764 | 0 | 0.84778 | 0.896731 | 1029 | 0.959136684466019 |
| Chr12:132.5Mb | 1861 | 12 | 132012317 | 132957945 | 1521 | 227 | AD | eGFR | 0.025461 | -0.39264 | 0.44013 | 0.000648262 | 0 | 0.23499 | 0.897285 | 1030 | 0.959136684466019 |
| Chr6:108.2Mb | 1038 | 6 | 107309328 | 109043244 | 2868 | 292 | AD | eGFR | -0.0179795 | -0.3158 | 0.26774 | 0.000323263 | 0 | 0.11655 | 0.899403 | 1031 | 0.96046818913676 |
| Chr21:37.7Mb | 2451 | 21 | 37303549 | 38046449 | 2143 | 203 | AD | eGFR | -0.0234895 | -0.44653 | 0.38548 | 0.000551755 | 0 | 0.23954 | 0.903152 | 1032 | 0.963537162790698 |
| Chr3:114.7Mb | 519 | 3 | 113657666 | 115649909 | 3320 | 321 | AD | eGFR | -0.030608 | -0.633 | 0.54161 | 0.000936852 | 0 | 0.50555 | 0.90442 | 1033 | 0.963955876089061 |
| Chr9:30Mb | 1407 | 9 | 29596502 | 30410078 | 1865 | 150 | AD | eGFR | -0.0503855 | -1 | 1 | 0.0025387 | 0 | 1 | 0.907555 | 103 | 0.964778589199614 |

| locus_label | locus | chr | start | stop | n.snps | n.pcs | phen1 | phen2 | rho | rho.lower | rho.upper | r2 | r2.lower | r2.upper | p | rank | FDR |
|---|---|---|---|---|---|---|---|---|---|---|---|---|---|---|---|---|---|
| | | | | | | | | | | | | | | | | 4 | |
| Chr19:1.9Mb | 2308 | 19 | 1515921 | 2253174 | 1589 | 288 | AD | eGFR | 0.0115296 | -0.18856 | 0.20688 | 0.000132931 | 0 | 0.05085 | 0.908278 | 1035 | 0.964778589199614 |
| Chr8:80.5Mb | 1314 | 8 | 79756170 | 81340231 | 3078 | 275 | AD | eGFR | -0.0425283 | -1 | 0.96103 | 0.00180866 | 0 | 1 | 0.908646 | 1036 | 0.964778589199614 |
| Chr6:84.3Mb | 1016 | 6 | 83571954 | 85038134 | 2184 | 204 | AD | eGFR | -0.0261816 | -0.54323 | 0.48093 | 0.000685478 | 0 | 0.36481 | 0.908697 | 1037 | 0.964778589199614 |
| Chr11:69.3Mb | 1677 | 11 | 68887345 | 69713996 | 1998 | 257 | AD | eGFR | -0.0127231 | -0.24148 | 0.22229 | 0.000161877 | 0 | 0.07019 | 0.911588 | 1038 | 0.965664699616123 |
| Chr4:56.4Mb | 652 | 4 | 55681702 | 57103100 | 3518 | 274 | AD | eGFR | -0.0341918 | -0.86545 | 0.73664 | 0.00116908 | 0 | 1 | 0.912155 | 1039 | 0.965664699616123 |
| Chr14:94.1Mb | 2020 | 14 | 93386329 | 94892240 | 3564 | 316 | AD | eGFR | -0.0199804 | -0.43269 | 0.37576 | 0.000399218 | 0 | 0.23188 | 0.913439 | 1040 | 0.965664699616123 |
| Chr7:88.6Mb | 1177 | 7 | 87836642 | 89330087 | 3177 | 237 | AD | eGFR | -0.0438878 | -1 | 1 | 0.00192614 | 0 | 1 | 0.913498 | 1041 | 0.965664699616123 |
| Chr9:133.6Mb | 1476 | 9 | 132999453 | 134141936 | 2280 | 317 | AD | eGFR | 0.0317759 | -0.69188 | 0.78643 | 0.00100971 | 0 | 0.89289 | 0.913917 | 1042 | 0.965664699616123 |
| Chr15:65.3Mb | 2069 | 15 | 64339946 | 66355634 | 3047 | 250 | AD | eGFR | 0.0204914 | -0.40642 | 0.42973 | 0.000419897 | 0 | 0.23849 | 0.916776 | 1043 | 0.967756832214765 |
| Chr5:174.1Mb | 920 | 5 | 173606996 | 174662885 | 2823 | 330 | AD | eGFR | 0.0320474 | -0.82698 | 0.96449 | 0.00102703 | 0 | 1 | 0.921142 | 1044 | 0.969974696564886 |
| Chr10:119Mb | 1592 | 10 | 118416050 | 119528730 | 1950 | 282 | AD | eGFR | -0.0237659 | -0.60381 | 0.52362 | 0.00056482 | 0 | 0.45556 | 0.921901 | 1045 | 0.969974696564886 |
| Chr3:14.7Mb | 436 | 3 | 14312008 | 15150876 | 2049 | 277 | AD | eGFR | -0.0172199 | -0.391 | 0.35025 | 0.000296525 | 0 | 0.18848 | 0.922861 | 1046 | 0.969974696564886 |
| Chr5:32.2Mb | 804 | 5 | 31679465 | 32727914 | 2649 | 312 | AD | eGFR | 0.0264315 | -0.62953 | 0.68605 | 0.000698624 | 0 | 0.64858 | 0.922935 | 1047 | 0.969974696564886 |
| Chr5:172.9Mb | 919 | 5 | 172285683 | 173606995 | 3235 | 323 | AD | eGFR | 0.0174608 | -0.35893 | 0.39064 | 0.000304879 | 0 | 0.19082 | 0.923282 | 1048 | 0.969974696564886 |
| Chr10:90.5Mb | 1569 | 10 | 89971629 | 91021321 | 2169 | 250 | AD | eGFR | -0.022981 | -0.60307 | 0.53136 | 0.000528126 | 0 | 0.47329 | 0.926964 | 1049 | 0.972914551000953 |
| Chr5:141.9Mb | 893 | 5 | 141253831 | 142532956 | 2791 | 312 | AD | eGFR | -0.0148646 | -0.36801 | 0.33112 | 0.000220956 | 0 | 0.16709 | 0.92972 | 1050 | 0.974877828571429 |
| Chr19:14.4Mb | 2321 | 19 | 13968320 | 14891100 | 1814 | 280 | AD | eGFR | 0.0164284 | -0.40742 | 0.45272 | 0.000269892 | 0 | 0.26317 | 0.931077 | 1051 | 0.974962061494797 |
| Chr10:23.9Mb | 1511 | 10 | 23365686 | 24489447 | 2110 | 205 | AD | eGFR | -0.0208098 | -0.62462 | 0.56896 | 0.000433046 | 0 | 0.5483 | 0.933302 | 1052 | 0.974962061494797 |
| Chr2:128.7Mb | 320 | 2 | 127895645 | 129486897 | 3711 | 281 | AD | eGFR | -0.018911 | -0.52534 | 0.49119 | 0.000357625 | 0 | 0.36245 | 0.934046 | 1053 | 0.974962061494797 |
| Chr10:81.5Mb | 1560 | 10 | 81190574 | 81763230 | 1445 | 81 | AD | eGFR | -0.0253582 | -0.781 | 0.73071 | 0.000643037 | 0 | 0.92108 | 0.934402 | 1054 | 0.974962061494797 |
| Chr5:53.3Mb | 820 | 5 | 52837226 | 53747856 | 2109 | 222 | AD | eGFR | 0.0131249 | -0.36626 | 0.38441 | 0.000172262 | 0 | 0.20895 | 0.935701 | 1055 | 0.974962061494797 |
| Chr3:144.8Mb | 547 | 3 | 144408822 | 145211907 | 1927 | 143 | AD | eGFR | -0.0276627 | -0.95978 | 0.86588 | 0.000765225 | 0 | 1 | 0.935998 | 1056 | 0.974962061494797 |
| Chr11:124.8Mb | 1729 | 11 | 124403478 | 125266684 | 1845 | 290 | AD | eGFR | 0.0204775 | -0.57688 | 0.60224 | 0.000419329 | 0 | 0.51068 | 0.935999 | 1057 | 0.974962061494797 |

| locus_label | locus | chr | start | stop | n.snps | n.pcs | phen1 | phen2 | rho | rho.lower | rho.upper | r2 | r2.lower | r2.upper | p | rank | FDR |
|---|---|---|---|---|---|---|---|---|---|---|---|---|---|---|---|---|---|
| Chr5:106.8Mb | 864 | 5 | 106389389 | 107265911 | 1990 | 248 | AD | eGFR | -0.0201578 | -0.72947 | 0.63717 | 0.000406335 | 0 | 0.83936 | 0.939038 | 1058 | 0.977203060491493 |
| Chr11:61.1Mb | 1671 | 11 | 60515106 | 61717117 | 1794 | 276 | AD | eGFR | -0.0169281 | -0.57043 | 0.5294 | 0.000286562 | 0 | 0.41786 | 0.943655 | 1059 | 0.979938771913289 |
| Chr4:135Mb | 723 | 4 | 134109979 | 135809542 | 3091 | 222 | AD | eGFR | 0.0228445 | -0.79924 | 0.88066 | 0.00052187 | 0 | 1 | 0.944216 | 1060 | 0.979938771913289 |
| Chr3:192.6Mb | 589 | 3 | 192049689 | 193122090 | 2262 | 264 | AD | eGFR | 0.0196226 | -0.65382 | 0.72774 | 0.000385045 | 0 | 0.73522 | 0.944337 | 1061 | 0.979938771913289 |
| Chr8:71.4Mb | 1306 | 8 | 70872089 | 72013184 | 2058 | 147 | AD | eGFR | 0.0200616 | -0.69233 | 0.75877 | 0.000402468 | 0 | 0.81316 | 0.946211 | 1062 | 0.980958861581921 |
| Chr10:72.2Mb | 1553 | 10 | 71672998 | 72659223 | 2726 | 287 | AD | eGFR | 0.0183452 | -0.72323 | 0.79186 | 0.000336548 | 0 | 0.89346 | 0.951175 | 1063 | 0.985177492944497 |
| Chr7:6.4Mb | 1101 | 7 | 6020654 | 6716905 | 1783 | 199 | AD | eGFR | -0.016386 | -0.67404 | 0.64302 | 0.0002685 | 0 | 0.6659 | 0.953092 | 1064 | 0.986059735211268 |
| Chr12:131.7Mb | 1860 | 12 | 131404043 | 132012316 | 1610 | 218 | AD | eGFR | 0.0132984 | -0.49368 | 0.52259 | 0.000176847 | 0 | 0.35811 | 0.953818 | 1065 | 0.986059735211268 |
| Chr6:168.2Mb | 1091 | 6 | 167714866 | 168707338 | 2392 | 288 | AD | eGFR | -0.0107052 | -0.42183 | 0.3989 | 0.000114602 | 0 | 0.22571 | 0.955475 | 1066 | 0.986151357075914 |
| Chr11:35.9Mb | 1648 | 11 | 35500368 | 36331189 | 1777 | 262 | AD | eGFR | -0.0192401 | -0.98998 | 0.89927 | 0.000370183 | 0 | 1 | 0.955698 | 1067 | 0.986151357075914 |
| Chr1:62.1Mb | 53 | 1 | 61377616 | 62778733 | 3068 | 328 | AD | eGFR | 0.0112455 | -0.46904 | 0.50846 | 0.000126462 | 0 | 0.34767 | 0.95852 | 1068 | 0.988137191011236 |
| Chr10:31.7Mb | 1520 | 10 | 30919470 | 32457293 | 2882 | 239 | AD | eGFR | -0.0106846 | -0.51319 | 0.48385 | 0.000114161 | 0 | 0.35961 | 0.9621 | 1069 | 0.989433028011204 |
| Chr8:107.1Mb | 1335 | 8 | 106500951 | 107787163 | 2523 | 218 | AD | eGFR | -0.0135603 | -0.75518 | 0.707 | 0.000183882 | 0 | 0.88908 | 0.962283 | 1070 | 0.989433028011204 |
| Chr13:99Mb | 1937 | 13 | 98527336 | 99402294 | 2202 | 295 | AD | eGFR | -0.00820665 | -0.37516 | 0.34818 | 6.73E-05 | 0 | 0.18082 | 0.962473 | 1071 | 0.989433028011204 |
| Chr22:21.7Mb | 2466 | 22 | 20969185 | 22404422 | 2032 | 226 | AD | eGFR | 0.0108515 | -0.5376 | 0.57855 | 0.000117754 | 0 | 0.46555 | 0.964617 | 1072 | 0.990448518173346 |
| Chr19:43.6Mb | 2349 | 19 | 43317283 | 43960958 | 576 | 112 | AD | eGFR | -0.00645966 | -0.30386 | 0.30035 | 4.17E-05 | 0 | 0.11915 | 0.96526 | 1073 | 0.990448518173346 |
| Chr11:72.1Mb | 1679 | 11 | 71242835 | 72875068 | 2865 | 219 | AD | eGFR | -0.0109821 | -0.6982 | 0.66529 | 0.000120607 | 0 | 0.69415 | 0.968729 | 1074 | 0.99257044744186 |
| Chr22:50Mb | 2494 | 22 | 49559208 | 50347996 | 2679 | 289 | AD | eGFR | 0.00737035 | -0.40771 | 0.41207 | 5.43E-05 | 0 | 0.22864 | 0.969131 | 1075 | 0.99257044744186 |
| Chr1:165.1Mb | 122 | 1 | 164726580 | 165553812 | 2098 | 252 | AD | eGFR | -0.00605939 | -0.40194 | 0.37759 | 3.67E-05 | 0 | 0.20275 | 0.973578 | 1076 | 0.994175777472527 |
| Chr19:48.2Mb | 2354 | 19 | 47752728 | 48573065 | 1919 | 248 | AD | eGFR | -0.0072379 | -0.52069 | 0.51402 | 5.24E-05 | 0 | 0.38624 | 0.973703 | 1077 | 0.994175777472527 |
| Chr17:20.9Mb | 2193 | 17 | 20402495 | 21432873 | 1284 | 200 | AD | eGFR | -0.00677046 | -0.46815 | 0.45606 | 4.58E-05 | 0 | 0.29272 | 0.975106 | 1078 | 0.994175777472527 |
| Chr2:11.6Mb | 216 | 2 | 11230350 | 11995765 | 1831 | 249 | AD | eGFR | -0.00552104 | -0.39008 | 0.36792 | 3.05E-05 | 0 | 0.19787 | 0.975906 | 1079 | 0.994175777472527 |
| Chr8:134Mb | 1358 | 8 | 133461552 | 134509448 | 2672 | 298 | AD | eGFR | 0.00608728 | -0.41398 | 0.4437 | 3.71E-05 | 0 | 0.2519 | 0.975975 | 1080 | 0.994175777472527 |
| Chr16:42.1Mb | 2131 | 16 | 35259454 | 49010761 | 2375 | 221 | AD | eGFR | -0.0103762 | -1 | 0.98365 | 0.00010766 | 0 | 1 | 0.976993 | 108 | 0.994175777472527 |

| locus_label | locus | chr | start | stop | n.snps | n.pcs | phen1 | phen2 | rho | rho.lower | rho.upper | r2 | r2.lower | r2.upper | p | rank | FDR |
|---|---|---|---|---|---|---|---|---|---|---|---|---|---|---|---|---|---|
| | | | | | | | | | | | | 5 | | | | 1 | |
| Chr1:110.7Mb | 97 | 1 | 110224231 | 111134062 | 1779 | 284 | AD | eGFR | -0.00516187 | -0.4125 | 0.39839 | 2.66E-05 | 0 | 0.22533 | 0.978029 | 1082 | 0.994175777472527 |
| Chr5:76.6Mb | 839 | 5 | 75959516 | 77290255 | 3076 | 312 | AD | eGFR | -0.0076221 | -0.70666 | 0.68382 | 5.81E-05 | 0 | 0.76077 | 0.978171 | 1083 | 0.994175777472527 |
| Chr8:26.3Mb | 1271 | 8 | 25859278 | 26679135 | 2604 | 236 | AD | eGFR | -0.00519602 | -0.44473 | 0.44689 | 2.70E-05 | 0 | 0.27065 | 0.979854 | 1084 | 0.994175777472527 |
| Chr13:47Mb | 1889 | 13 | 46493237 | 47433528 | 1750 | 219 | AD | eGFR | 0.00442164 | -0.45456 | 0.45834 | 1.96E-05 | 0 | 0.28731 | 0.983456 | 1085 | 0.994175777472527 |
| Chr4:78.5Mb | 671 | 4 | 78045638 | 78967715 | 2191 | 177 | AD | eGFR | -0.0056019 | -0.71244 | 0.71478 | 3.14E-05 | 0 | 0.76341 | 0.98447 | 1086 | 0.994175777472527 |
| Chr22:41.1Mb | 2484 | 22 | 40378784 | 41722787 | 1595 | 174 | AD | eGFR | -0.00371781 | -0.46341 | 0.43737 | 1.38E-05 | 0 | 0.30513 | 0.98453 | 1087 | 0.994175777472527 |
| Chr1:227.3Mb | 176 | 1 | 226534712 | 228045554 | 3557 | 228 | AD | eGFR | 0.00679223 | -1 | 1 | 4.61E-05 | 0 | 1 | 0.985292 | 1088 | 0.994175777472527 |
| Chr14:23.4Mb | 1957 | 14 | 22760701 | 23985936 | 2701 | 337 | AD | eGFR | -0.00305118 | -0.35018 | 0.34773 | 9.31E-06 | 0 | 0.16708 | 0.985405 | 1089 | 0.994175777472527 |
| Chr6:128.2Mb | 1055 | 6 | 127545460 | 128763380 | 2020 | 199 | AD | eGFR | -0.00635911 | -0.95845 | 1 | 4.04E-05 | 0 | 1 | 0.985855 | 1090 | 0.994175777472527 |
| Chr8:117.2Mb | 1344 | 8 | 116094204 | 118300068 | 3349 | 313 | AD | eGFR | -0.00322753 | -0.39559 | 0.37196 | 1.04E-05 | 0 | 0.20315 | 0.985981 | 1091 | 0.994175777472527 |
| Chr11:111.9Mb | 1718 | 11 | 111117781 | 112755446 | 2532 | 254 | AD | eGFR | -0.00315468 | -0.39488 | 0.395 | 9.95E-06 | 0 | 0.21292 | 0.986049 | 1092 | 0.994175777472527 |
| Chr2:208.2Mb | 385 | 2 | 207726595 | 208674588 | 1771 | 240 | AD | eGFR | 0.00480099 | -0.85686 | 0.85386 | 2.30E-05 | 0 | 1 | 0.988158 | 1093 | 0.994782156164384 |
| Chr19:3.5Mb | 2310 | 19 | 3085447 | 3893909 | 1987 | 323 | AD | eGFR | 0.00172634 | -0.26664 | 0.26991 | 2.98E-06 | 0 | 0.09432 | 0.989356 | 1094 | 0.994782156164384 |
| Chr11:67.4Mb | 1675 | 11 | 66782662 | 68000949 | 1870 | 206 | AD | eGFR | -0.00279181 | -0.48015 | 0.47149 | 7.79E-06 | 0 | 0.32516 | 0.989361 | 1095 | 0.994782156164384 |
| Chr14:88.8Mb | 2016 | 14 | 88208596 | 89482883 | 2847 | 222 | AD | eGFR | 0.00140859 | -0.33686 | 0.33424 | 1.98E-06 | 0 | 0.15061 | 0.993075 | 1096 | 0.996878721057429 |
| Chr12:0.6Mb | 1740 | 12 | 60317 | 1078397 | 2186 | 292 | AD | eGFR | -0.00131904 | -0.35128 | 0.34827 | 1.74E-06 | 0 | 0.17274 | 0.993257 | 1097 | 0.996878721057429 |
| Chr1:210.4Mb | 161 | 1 | 209711170 | 211082892 | 3018 | 277 | AD | eGFR | -0.00107468 | -0.50548 | 0.5129 | 1.15E-06 | 0 | 0.36017 | 0.996275 | 1098 | 0.99899706284153 |
| Chr11:76Mb | 1682 | 11 | 75445254 | 76518906 | 2109 | 257 | AD | eGFR | 0.000252326 | -0.45627 | 0.45782 | 6.37E-08 | 0 | 0.28663 | 0.999044 | 1099 | 0.999914 |
| Chr3:194.7Mb | 591 | 3 | 194217141 | 195196789 | 3247 | 297 | AD | eGFR | -0.000232588 | -0.44818 | 0.44222 | 5.41E-08 | 0 | 0.27394 | 0.999102 | 1100 | 0.999914 |
| Chr7:77.1Mb | 1168 | 7 | 76451929 | 77663964 | 2252 | 204 | AD | eGFR | 2.08E-05 | -0.45203 | 0.45272 | 4.31E-10 | 0 | 0.32079 | 0.999914 | 1101 | 0.999914 |

**Supplementary Table S15. Complete Local Genetic Correlation Results in African American Ancestry.**
This table presents the complete set of local genetic correlation analysis results for all genomic loci tested in African American ancestry individuals. Given the distinct linkage disequilibrium patterns and genetic architecture in African ancestry populations, this comprehensive dataset is essential for understanding ancestry-specific genetic pleiotropy between AD and eGFR. The table includes all tested loci regardless of statistical significance.

| locus_label | locus | chr | start | stop | n.snps | n.pcs | phen1 | phen2 | rho | rho.lower | rho.upper | r2 | r2.lower | r2.upper | p | rank | FDR |
|---|---|---|---|---|---|---|---|---|---|---|---|---|---|---|---|---|---|
| Chr3:190.1Mb | 676 | 3 | 189757981 | 190357798 | 3382 | 428 | AD | eGFR | 1 | 0.64181 | 1 | 1 | 0.41192 | 1 | 1.25E-08 | 1 | 1.15E-05 |
| Chr19:47Mb | 2542 | 19 | 46508672 | 47544622 | 4529 | 489 | AD | eGFR | -0.938095 | -1 | -0.58728 | 0.880022 | 0.3449 | 1 | 8.33E-07 | 2 | 0.0003844862645 |
| Chr13:60.5Mb | 2069 | 13 | 59706604 | 61268194 | 6321 | 452 | AD | eGFR | -1 | -1 | -0.52163 | 1 | 0.2721 | 1 | 6.34E-06 | 3 | 0.00194996672 |
| Chr3:143.3Mb | 627 | 3 | 142567286 | 143976384 | 6678 | 519 | AD | eGFR | 1 | 0.47181 | 1 | 1 | 0.22261 | 1 | 8.99E-06 | 4 | 0.0020752062825 |
| Chr2:129.6Mb | 372 | 2 | 129313012 | 129915501 | 3452 | 364 | AD | eGFR | 1 | 0.53041 | 1 | 1 | 0.28134 | 1 | 2.52E-05 | 5 | 0.00464505288 |
| Chr15:47.6Mb | 2238 | 15 | 46833894 | 48301725 | 6328 | 481 | AD | eGFR | 1 | 0.4851 | 1 | 1 | 0.23533 | 1 | 3.43E-05 | 6 | 0.0052800215 |
| Chr6:40.2Mb | 1106 | 6 | 39814082 | 40550913 | 3748 | 384 | AD | eGFR | 1 | 0.49563 | 1 | 1 | 0.24565 | 1 | 4.02E-05 | 7 | 0.0053038349 |
| Chr3:132.5Mb | 619 | 3 | 132061684 | 132880614 | 3734 | 317 | AD | eGFR | 0.900919 | 0.48149 | 1 | 0.811656 | 0.23184 | 1 | 7.46E-05 | 8 | 0.008604367525 |
| Chr3:168.7Mb | 655 | 3 | 168302338 | 169190677 | 3439 | 395 | AD | eGFR | 0.835951 | 0.43224 | 1 | 0.698814 | 0.18683 | 1 | 9.32E-05 | 9 | 0.00916572228 |
| Chr4:88.8Mb | 787 | 4 | 88480371 | 89180351 | 3290 | 384 | AD | eGFR | 1 | 0.50573 | 1 | 1 | 0.25577 | 1 | 9.93E-05 | 10 | 0.00916572228 |
| Chr1:15.4Mb | 20 | 1 | 15102430 | 15747105 | 3820 | 506 | AD | eGFR | -0.978381 | -1 | -0.48536 | 0.957229 | 0.23558 | 1 | 0.000155807 | 11 | 0.0130736237272727 |
| Chr9:12.4Mb | 1530 | 9 | 12044381 | 12797991 | 4146 | 374 | AD | eGFR | 0.976364 | 0.48958 | 1 | 0.953287 | 0.23969 | 1 | 0.000182199 | 12 | 0.01401413975 |
| Chr8:106.7Mb | 1475 | 8 | 105982692 | 107379445 | 5592 | 426 | AD | eGFR | -0.992374 | -1 | -0.48682 | 0.984807 | 0.23699 | 1 | 0.000206541 | 13 | 0.0140597293571429 |
| Chr2:223.9Mb | 462 | 2 | 223561337 | 224269422 | 3434 | 366 | AD | eGFR | 0.93202 | 0.46766 | 1 | 0.868662 | 0.2187 | 1 | 0.000213257 | 14 | 0.0140597293571429 |
| Chr8:27.8Mb | 1409 | 8 | 27425268 | 28158440 | 3949 | 426 | AD | eGFR | 0.552667 | 0.2638 | 0.9094 | 0.305441 | 0.06959 | 0.82701 | 0.000396867 | 15 | 0.0223297046470588 |
| Chr2:127.2Mb | 369 | 2 | 126680360 | 127770280 | 5665 | 403 | AD | eGFR | -0.89697 | -1 | -0.41925 | 0.804555 | 0.17577 | 1 | 0.000399865 | 16 | 0.0223297046470588 |
| Chr2:208.3Mb | 445 | 2 | 207726268 | 208825721 | 4160 | 447 | AD | eGFR | -0.829862 | -1 | -0.38971 | 0.688671 | 0.15187 | 1 | 0.000411273 | 17 | 0.0223297046470588 |
| Chr1:165.1Mb | 148 | 1 | 164440353 | 165828905 | 6604 | 533 | AD | eGFR | -0.792772 | -1 | -0.36457 | 0.628488 | 0.13291 | 1 | 0.000502718 | 18 | 0.0257782618888889 |
| Chr2:29.2Mb | 273 | 2 | 28802271 | 29633040 | 4064 | 424 | AD | eGFR | 0.712237 | 0.32621 | 1 | 0.507282 | 0.10641 | 1 | 0.000554439 | 19 | 0.0266770075 |
| Chr3:167.9Mb | 654 | 3 | 167521068 | 168302337 | 3313 | 348 | AD | eGFR | 0.388097 | 0.17215 | 0.62029 | 0.150619 | 0.02963 | 0.38476 | 0.00057805 | 20 | 0.0266770075 |
| Chr9:121Mb | 1617 | 9 | 120600042 | 121473024 | 4013 | 360 | AD | eGFR | -0.796781 | -1 | -0.35907 | 0.634861 | 0.12893 | 1 | 0.000759204 | 21 | 0.0333688234285714 |
| Chr17:63.8Mb | 2412 | 17 | 63346710 | 64331974 | 4964 | 427 | AD | eGFR | 0.731831 | 0.32146 | 1 | 0.535577 | 0.10333 | 1 | 0.000917873 | 22 | 0.0385089445 |
| Chr2:166.9Mb | 409 | 2 | 16639556 | 16737303 | 4156 | 365 | AD | eGFR | 0.828223 | 0.36692 | 1 | 0.685954 | 0.13463 | 1 | 0.00096742 | 23 | 0.03875561625 |

| locus_label | locus | chr | start | stop | n.snps | n.pcs | phen1 | phen2 | rho | rho.lower | rho.upper | r2 | r2.lower | r2.upper | p | rank | FDR |
|---|---|---|---|---|---|---|---|---|---|---|---|---|---|---|---|---|---|
| | | | 9 | 8 | | | | | | | | | | | 4 | | |
| Chr10:23.2Mb | 1664 | 10 | 22542648 | 23839537 | 5498 | 435 | AD | eGFR | -0.972627 | -1 | -0.41302 | 0.946004 | 0.17059 | 1 | 0.00100773 | 24 | 0.03875561625 |
| Chr10:124.4Mb | 1756 | 10 | 123853589 | 124868063 | 4785 | 469 | AD | eGFR | 0.888056 | 0.36744 | 1 | 0.788643 | 0.13539 | 1 | 0.00109899 | 25 | 0.0405747108 |
| Chr3:105.2Mb | 592 | 3 | 104829898 | 105669644 | 3507 | 357 | AD | eGFR | 0.80109 | 0.33271 | 1 | 0.641745 | 0.11069 | 1 | 0.0011634 | 26 | 0.0413007 |
| Chr2:212.7Mb | 450 | 2 | 212383518 | 213059448 | 3506 | 416 | AD | eGFR | 0.684288 | 0.28323 | 1 | 0.46825 | 0.08022 | 1 | 0.00132503 | 27 | 0.0452963959259259 |
| Chr2:65.2Mb | 313 | 2 | 64623202 | 65712249 | 4852 | 452 | AD | eGFR | 0.559073 | 0.22725 | 1 | 0.312563 | 0.05164 | 1 | 0.00139862 | 28 | 0.0461045092857143 |
| Chr15:23.5Mb | 2216 | 15 | 22881864 | 24199319 | 2764 | 464 | AD | eGFR | -0.818639 | -1 | -0.32926 | 0.67017 | 0.10841 | 1 | 0.0014733 | 29 | 0.0468915827586207 |
| Chr11:31.4Mb | 1801 | 11 | 30688687 | 32065472 | 4215 | 356 | AD | eGFR | 0.514492 | 0.20091 | 0.89762 | 0.264702 | 0.04037 | 0.80572 | 0.00153451 | 30 | 0.0472117576666667 |
| Chr5:56.9Mb | 953 | 5 | 56326600 | 57441632 | 4873 | 417 | AD | eGFR | -0.600877 | -1 | -0.23814 | 0.361053 | 0.05671 | 1 | 0.00163345 | 31 | 0.0486346564516129 |
| Chr3:42.3Mb | 538 | 3 | 41935928 | 42726675 | 3673 | 385 | AD | eGFR | -0.785911 | -1 | -0.30076 | 0.617656 | 0.09086 | 1 | 0.00197607 | 32 | 0.0569972690625 |
| Chr17:36.8Mb | 2392 | 17 | 36131866 | 37377590 | 4451 | 494 | AD | eGFR | 1 | 0.34497 | 1 | 1 | 0.12014 | 1 | 0.00220679 | 33 | 0.0617232475757576 |
| Chr12:13.2Mb | 1913 | 12 | 12712669 | 13652723 | 4065 | 515 | AD | eGFR | -0.460987 | -0.86107 | -0.16951 | 0.212509 | 0.02873 | 0.74144 | 0.00228604 | 34 | 0.0620592623529412 |
| Chr5:102.7Mb | 991 | 5 | 102307559 | 103184572 | 3437 | 397 | AD | eGFR | 0.936028 | 0.33533 | 1 | 0.876149 | 0.11245 | 1 | 0.00253401 | 35 | 0.0668254637142857 |
| Chr14:22.4Mb | 2134 | 14 | 22083534 | 22621154 | 2620 | 413 | AD | eGFR | 0.746501 | 0.26423 | 1 | 0.557264 | 0.06982 | 1 | 0.00266724 | 36 | 0.06701193 |
| Chr1:25.4Mb | 32 | 1 | 24881152 | 25864438 | 3747 | 426 | AD | eGFR | -0.810113 | -1 | -0.29576 | 0.656284 | 0.08747 | 1 | 0.00270596 | 37 | 0.06701193 |
| Chr3:141.7Mb | 626 | 3 | 140865160 | 142567285 | 7031 | 469 | AD | eGFR | 0.542351 | 0.19178 | 1 | 0.294145 | 0.03678 | 1 | 0.00282303 | 38 | 0.06701193 |
| Chr7:25.6Mb | 1254 | 7 | 25073338 | 26180381 | 5342 | 491 | AD | eGFR | -0.692288 | -1 | -0.26153 | 0.479263 | 0.0684 | 1 | 0.00283149 | 39 | 0.06701193 |
| Chr2:14.8Mb | 258 | 2 | 14334726 | 15230095 | 4235 | 422 | AD | eGFR | -0.557716 | -1 | -0.19305 | 0.311048 | 0.03729 | 1 | 0.00339557 | 40 | 0.0779140790697674 |
| Chr12:52.5Mb | 1952 | 12 | 51925777 | 53036280 | 5590 | 498 | AD | eGFR | 0.810255 | 0.29671 | 1 | 0.656513 | 0.08804 | 1 | 0.00348666 | 41 | 0.0779140790697674 |
| Chr3:130.1Mb | 616 | 3 | 129756877 | 130375086 | 2849 | 353 | AD | eGFR | 0.79925 | 0.28362 | 1 | 0.638801 | 0.08044 | 1 | 0.00358414 | 42 | 0.0779140790697674 |
| Chr2:72.5Mb | 321 | 2 | 71868054 | 73127122 | 4410 | 428 | AD | eGFR | -0.985038 | -1 | -0.35016 | 0.9703 | 0.12356 | 1 | 0.0036298 | 43 | 0.0779140790697674 |
| Chr12:104.6Mb | 1998 | 12 | 104141497 | 105143830 | 4842 | 451 | AD | eGFR | -0.820257 | -1 | -0.28284 | 0.672822 | 0.08127 | 1 | 0.00379201 | 44 | 0.0795460279545455 |
| Chr1:44.6Mb | 51 | 1 | 44129648 | 45085625 | 3609 | 432 | AD | eGFR | -0.743365 | -1 | -0.25683 | 0.552592 | 0.06596 | 1 | 0.00393139 | 45 | 0.0806371771111111 |
| Chr9:87.4Mb | 1582 | 9 | 86922196 | 87887525 | 4374 | 443 | AD | eGFR | 0.621901 | 0.20755 | 1 | 0.38676 | 0.04308 | 1 | 0.0044001 | 46 | 0.0878589953191489 |
| Chr2:68.1Mb | 316 | 2 | 67696942 | 68545962 | 4316 | 442 | AD | eGFR | 0.550759 | 0.1935 | 1 | 0.303336 | 0.03744 | 1 | 0.00447386 | 47 | 0.0878589953191489 |
| Chr1:226.3Mb | 212 | 1 | 225828363 | 226816212 | 3682 | 414 | AD | eGFR | -0.877909 | -1 | -0.29009 | 0.770724 | 0.08415 | 1 | 0.00487141 | 48 | 0.0934294506122449 |
| Chr2:17.4Mb | 261 | 2 | 16815315 | 17992213 | 5027 | 387 | AD | eGFR | -0.636663 | -1 | -0.20446 | 0.405339 | 0.04252 | 1 | 0.00495996 | 49 | 0.093429450612244 |

| locus_label | locus | chr | start | stop | n.snps | n.pcs | phen1 | phen2 | rho | rho.lower | rho.upper | r2 | r2.lower | r2.upper | p | rank | FDR |
|---|---|---|---|---|---|---|---|---|---|---|---|---|---|---|---|---|---|
| | | | | | | | | | | | | | | | | | 9 |
| Chr3:136.4Mb | 622 | 3 | 135469023 | 137366620 | 5830 | 348 | AD | eGFR | 0.578901 | 0.18068 | 1 | 0.335126 | 0.03304 | 1 | 0.00559144 | 50 | 0.101129452830189 |
| Chr17:6.4Mb | 2370 | 17 | 5942910 | 6893025 | 5160 | 545 | AD | eGFR | 0.509894 | 0.15669 | 0.97726 | 0.259991 | 0.02481 | 0.95504 | 0.00561344 | 51 | 0.101129452830189 |
| Chr8:2.5Mb | 1374 | 8 | 2206115 | 2802295 | 3038 | 429 | AD | eGFR | 0.477185 | 0.14732 | 0.89986 | 0.227706 | 0.0218 | 0.80975 | 0.00579363 | 52 | 0.101129452830189 |
| Chr9:22.4Mb | 1543 | 9 | 21944639 | 22920514 | 4385 | 418 | AD | eGFR | -0.443766 | -0.87979 | -0.13499 | 0.196928 | 0.01835 | 0.77404 | 0.005807 | 53 | 0.101129452830189 |
| Chr15:85.5Mb | 2270 | 15 | 84704361 | 86341459 | 5678 | 390 | AD | eGFR | 0.700336 | 0.20782 | 1 | 0.49047 | 0.04334 | 1 | 0.00599294 | 54 | 0.102434881851852 |
| Chr19:29.9Mb | 2526 | 19 | 29484293 | 30381693 | 5568 | 453 | AD | eGFR | -1 | -1 | -0.2689 | 1 | 0.07523 | 1 | 0.00630225 | 55 | 0.104319727017544 |
| Chr13:95.8Mb | 2107 | 13 | 95470651 | 96133543 | 3781 | 450 | AD | eGFR | 0.776689 | 0.22436 | 1 | 0.603245 | 0.051 | 1 | 0.00642626 | 56 | 0.104319727017544 |
| Chr5:41Mb | 941 | 5 | 40623661 | 41435701 | 4028 | 301 | AD | eGFR | -1 | -1 | -0.28991 | 1 | 0.08631 | 1 | 0.00644228 | 57 | 0.104319727017544 |
| Chr13:50.5Mb | 2060 | 13 | 49988960 | 51054676 | 3936 | 325 | AD | eGFR | -0.873601 | -1 | -0.26453 | 0.763179 | 0.07097 | 1 | 0.00694144 | 58 | 0.109104701186441 |
| Chr3:37Mb | 532 | 3 | 36488583 | 37535683 | 4117 | 352 | AD | eGFR | -0.441046 | -0.97267 | -0.13342 | 0.194521 | 0.01798 | 0.94609 | 0.00697419 | 59 | 0.109104701186441 |
| Chr7:155.9Mb | 1367 | 7 | 155382682 | 156354440 | 5648 | 539 | AD | eGFR | 0.694989 | 0.20184 | 1 | 0.48301 | 0.04218 | 1 | 0.00746612 | 60 | 0.11320580578125 |
| Chr6:72.2Mb | 1135 | 6 | 71819903 | 72654006 | 4199 | 369 | AD | eGFR | -0.771628 | -1 | -0.21948 | 0.59541 | 0.04909 | 1 | 0.00761117 | 61 | 0.11320580578125 |
| Chr6:79.3Mb | 1143 | 6 | 78719822 | 79859802 | 5398 | 284 | AD | eGFR | -1 | -1 | -0.25462 | 1 | 0.06662 | 1 | 0.00764275 | 62 | 0.11320580578125 |
| Chr1:202.7Mb | 187 | 1 | 202130356 | 203201027 | 3930 | 441 | AD | eGFR | -0.774879 | -1 | -0.23103 | 0.600437 | 0.05371 | 1 | 0.00775783 | 63 | 0.11320580578125 |
| Chr12:44.5Mb | 1944 | 12 | 44044408 | 44923230 | 3867 | 272 | AD | eGFR | 0.65082 | 0.19019 | 1 | 0.423567 | 0.03741 | 1 | 0.00784959 | 64 | 0.11320580578125 |
| Chr18:4.5Mb | 2433 | 18 | 4003482 | 4927955 | 4947 | 521 | AD | eGFR | 0.839596 | 0.23337 | 1 | 0.704922 | 0.05669 | 1 | 0.00807049 | 65 | 0.113346540289855 |
| Chr4:178.7Mb | 880 | 4 | 178336518 | 179013564 | 3369 | 377 | AD | eGFR | -0.558495 | -1 | -0.16469 | 0.311917 | 0.0278 | 1 | 0.0082415 | 66 | 0.113346540289855 |
| Chr5:155.1Mb | 1037 | 5 | 154445639 | 155730801 | 5611 | 427 | AD | eGFR | -0.625452 | -1 | -0.17615 | 0.39119 | 0.03126 | 1 | 0.00845261 | 67 | 0.113346540289855 |
| Chr6:116.9Mb | 1175 | 6 | 116127419 | 117675181 | 5721 | 394 | AD | eGFR | -0.713614 | -1 | -0.20011 | 0.509245 | 0.04129 | 1 | 0.0084669 | 68 | 0.113346540289855 |
| Chr11:34.3Mb | 1804 | 11 | 33972428 | 34654944 | 3464 | 425 | AD | eGFR | 0.41032 | 0.10198 | 0.75563 | 0.168362 | 0.01056 | 0.57097 | 0.00847336 | 69 | 0.113346540289855 |
| Chr1:242.5Mb | 231 | 1 | 242201565 | 242844236 | 3403 | 416 | AD | eGFR | -0.431195 | -0.80761 | -0.11582 | 0.18593 | 0.01372 | 0.65224 | 0.0087042 | 70 | 0.114771094285714 |
| Chr21:18.8Mb | 2626 | 21 | 18360544 | 19333169 | 5168 | 488 | AD | eGFR | 0.608563 | 0.16076 | 1 | 0.370348 | 0.02618 | 1 | 0.00903602 | 71 | 0.116733220138889 |
| Chr4:66.1Mb | 762 | 4 | 65564708 | 66597940 | 5015 | 376 | AD | eGFR | 0.529166 | 0.14112 | 1 | 0.280017 | 0.02093 | 1 | 0.00910595 | 72 | 0.116733220138889 |
| Chr15:72.7Mb | 2261 | 15 | 72050679 | 73349295 | 3766 | 311 | AD | eGFR | 1 | 0.27633 | 1 | 1 | 0.0782 | 1 | 0.00957284 | 73 | 0.121037415342466 |
| Chr10:35.2Mb | 1677 | 10 | 34533511 | 35901901 | 5675 | 334 | AD | eGFR | 0.61495 | 0.16462 | 1 | 0.378163 | 0.02844 | 1 | 0.0097824 | 74 | 0.122015610810811 |
| Chr2:55.2Mb | 304 | 2 | 54688914 | 55664322 | 4272 | 350 | AD | eGFR | -0.665867 | -1 | -0.18482 | 0.443379 | 0.0351 | 1 | 0.0100989 | 75 | 0.124283796 |
| Chr11:26.4Mb | 1797 | 11 | 26019496 | 26753511 | 4025 | 341 | AD | eGFR | 0.509053 | 0.12298 | 1 | 0.259135 | 0.01537 | 1 | 0.0105137 | 76 | 0.126388667532468 |
| Chr10:2.3Mb | 1641 | 10 | 1873171 | 2639820 | 4944 | 447 | AD | eGFR | 0.568974 | 0.13515 | 1 | 0.323732 | 0.0193 | 1 | 0.0105438 | 77 | 0.126388667532468 |
| Chr3:160.1Mb | 646 | 3 | 159700711 | 160588293 | 3697 | 320 | AD | eGFR | 0.525109 | 0.13005 | 1 | 0.275739 | 0.01737 | 1 | 0.0110475 | 78 | 0.127807359756098 |
| Chr8:136Mb | 1501 | 8 | 135410222 | 136666004 | 5744 | 456 | AD | eGFR | 0.643457 | 0.15854 | 1 | 0.414036 | 0.02724 | 1 | 0.0111611 | 79 | 0.127807359756098 |
| Chr6:68.3Mb | 1130 | 6 | 67906207 | 68594060 | 3638 | 266 | AD | eGFR | -0.620173 | -1 | -0.14912 | 0.384615 | 0.02397 | 1 | 0.0111696 | 80 | 0.127807359756098 |
| Chr2:30.1Mb | 274 | 2 | 29633041 | 30575911 | 5120 | 466 | AD | eGFR | 0.508335 | 0.11947 | 1 | 0.258404 | 0.01465 | 1 | 0.0112847 | 81 | 0.127807359756098 |
| Chr9:0.3Mb | 1511 | 9 | 10327 | 524546 | 2226 | 425 | AD | eGFR | -0.657035 | -1 | -0.15378 | 0.431695 | 0.02527 | 1 | 0.0113545 | 82 | 0.127807359756098 |
| Chr13:35.1Mb | 2046 | 13 | 34683829 | 35435560 | 3503 | 413 | AD | eGFR | 0.486148 | 0.12059 | 1 | 0.23634 | 0.01577 | 1 | 0.0115533 | 83 | 0.128478263855422 |
| Chr19:40Mb | 2535 | 19 | 39372330 | 40695320 | 6085 | 474 | AD | eGFR | -1 | -1 | -0.20251 | 1 | 0.04698 | 1 | 0.0120809 | 84 | 0.131744297674419 |
| Chr3:149.6Mb | 634 | 3 | 14914360 | 14999388 | 3743 | 411 | AD | eGFR | 0.477201 | 0.10857 | 0.95194 | 0.227721 | 0.01246 | 0.9062 | 0.0122706 | 85 | 0.131744297674419 |

| locus_label | locus | chr | start | stop | n.snps | n.pcs | phen1 | phen2 | rho | rho.lower | rho.upper | r2 | r2.lower | r2.upper | p | rank | FDR |
|---|---|---|---|---|---|---|---|---|---|---|---|---|---|---|---|---|---|
| | | | 8 | 1 | | | | | | | | | | | | | |
| Chr12:64.8Mb | 1963 | 12 | 64304578 | 65391079 | 4212 | 423 | AD | eGFR | -0.714336 | -1 | -0.16764 | 0.510276 | 0.03094 | 1 | 0.0122752 | 86 | 0.131744297674419 |
| Chr6:47.1Mb | 1112 | 6 | 46729086 | 47443603 | 3485 | 415 | AD | eGFR | -0.293765 | -0.52888 | -0.07355 | 0.0862979 | 0.00555 | 0.27972 | 0.0125687 | 87 | 0.133343794252874 |
| Chr1:66.1Mb | 73 | 1 | 65690454 | 66554286 | 3395 | 364 | AD | eGFR | -0.498506 | -1 | -0.12323 | 0.248508 | 0.01608 | 1 | 0.0132211 | 88 | 0.138671310227273 |
| Chr7:50.4Mb | 1277 | 7 | 49881606 | 50906575 | 4581 | 372 | AD | eGFR | 0.754081 | 0.16278 | 1 | 0.568639 | 0.03119 | 1 | 0.0147495 | 89 | 0.149867024210526 |
| Chr11:81.8Mb | 1847 | 11 | 81213006 | 82354010 | 6158 | 366 | AD | eGFR | 0.753766 | 0.17897 | 1 | 0.568162 | 0.03488 | 1 | 0.014957 | 90 | 0.149867024210526 |
| Chr13:28.6Mb | 2039 | 13 | 28271193 | 28905323 | 3251 | 427 | AD | eGFR | 0.726728 | 0.14511 | 1 | 0.528134 | 0.02348 | 1 | 0.0150218 | 91 | 0.149867024210526 |
| Chr14:72.8Mb | 2182 | 14 | 72488344 | 73156662 | 3511 | 432 | AD | eGFR | 0.581078 | 0.1236 | 1 | 0.337651 | 0.01677 | 1 | 0.0151567 | 92 | 0.149867024210526 |
| Chr10:27.9Mb | 1669 | 10 | 27561439 | 28300040 | 3363 | 358 | AD | eGFR | 0.758874 | 0.1549 | 1 | 0.575889 | 0.02738 | 1 | 0.0153395 | 93 | 0.149867024210526 |
| Chr14:87.3Mb | 2195 | 14 | 86654443 | 87996998 | 6241 | 434 | AD | eGFR | -0.620526 | -1 | -0.12927 | 0.385053 | 0.01855 | 1 | 0.0154059 | 94 | 0.149867024210526 |
| Chr6:33.2Mb | 1099 | 6 | 32898138 | 33464010 | 3479 | 280 | AD | eGFR | -0.535705 | -1 | -0.1152 | 0.286979 | 0.01491 | 1 | 0.0154251 | 95 | 0.149867024210526 |
| Chr9:88.5Mb | 1583 | 9 | 87887526 | 89019173 | 4777 | 401 | AD | eGFR | 0.687007 | 0.14794 | 1 | 0.471978 | 0.02383 | 1 | 0.0159028 | 96 | 0.152898795833333 |
| Chr10:21.9Mb | 1663 | 10 | 21317459 | 22542647 | 4245 | 385 | AD | eGFR | -0.594791 | -1 | -0.11672 | 0.353777 | 0.01651 | 1 | 0.0166146 | 97 | 0.158095626804124 |
| Chr1:31.6Mb | 38 | 1 | 31122464 | 32039316 | 4300 | 417 | AD | eGFR | 0.621728 | 0.11836 | 1 | 0.386545 | 0.01751 | 1 | 0.0171167 | 98 | 0.161211368367347 |
| Chr14:89.1Mb | 2197 | 14 | 88686161 | 89483323 | 3519 | 299 | AD | eGFR | -0.720592 | -1 | -0.14866 | 0.519253 | 0.02605 | 1 | 0.0176756 | 99 | 0.164793725252525 |
| Chr8:55Mb | 1431 | 8 | 54398263 | 55510108 | 5068 | 409 | AD | eGFR | -0.570456 | -1 | -0.11209 | 0.32542 | 0.014 | 1 | 0.0180506 | 100 | 0.166607038 |
| Chr21:38.6Mb | 2645 | 21 | 37994705 | 39224094 | 5755 | 483 | AD | eGFR | -0.612755 | -1 | -0.10143 | 0.375469 | 0.01268 | 1 | 0.0185493 | 101 | 0.16951489009901 |
| Chr7:128.3Mb | 1343 | 7 | 127901550 | 128777331 | 3605 | 362 | AD | eGFR | -0.530959 | -1 | -0.09499 | 0.281918 | 0.0109 | 1 | 0.0188292 | 102 | 0.170338305825243 |
| Chr2:70.6Mb | 319 | 2 | 70191440 | 71033479 | 4204 | 406 | AD | eGFR | 0.411404 | 0.07003 | 0.81528 | 0.169254 | 0.00608 | 0.66468 | 0.0190085 | 103 | 0.170338305825243 |
| Chr10:8.1Mb | 1648 | 10 | 7633505 | 8543257 | 4953 | 511 | AD | eGFR | -0.541927 | -1 | -0.09237 | 0.293684 | 0.01036 | 1 | 0.0192652 | 104 | 0.17097865 |
| Chr5:83.6Mb | 976 | 5 | 83172905 | 84089616 | 3487 | 416 | AD | eGFR | -0.57368 | -1 | -0.10814 | 0.329108 | 0.01341 | 1 | 0.019718 | 105 | 0.17333060952381 |
| Chr1:211.3Mb | 196 | 1 | 210792521 | 211884316 | 4842 | 391 | AD | eGFR | 0.769967 | 0.15763 | 1 | 0.592849 | 0.02806 | 1 | 0.0201285 | 106 | 0.175269863207547 |
| Chr5:112.3Mb | 1001 | 5 | 111987726 | 112661351 | 3546 | 315 | AD | eGFR | 0.364576 | 0.05991 | 0.77677 | 0.132916 | 0.00448 | 0.60337 | 0.0203587 | 107 | 0.175617571028037 |
| Chr22:16.9Mb | 2655 | 22 | 16050115 | 17670444 | 2436 | 415 | AD | eGFR | -0.643333 | -1 | -0.11369 | 0.413878 | 0.01511 | 1 | 0.0209172 | 108 | 0.177945085321101 |
| Chr13:98.4Mb | 2110 | 13 | 98068416 | 98829196 | 3454 | 401 | AD | eGFR | 0.652054 | 0.11865 | 1 | 0.425174 | 0.01726 | 1 | 0.0210141 | 109 | 0.177945085321101 |
| Chr5:25.1Mb | 925 | 5 | 24494035 | 25720601 | 5531 | 456 | AD | eGFR | 0.470417 | 0.07878 | 1 | 0.221292 | 0.00837 | 1 | 0.0213744 | 110 | 0.179350647272727 |
| Chr10:20.9Mb | 1662 | 10 | 20504211 | 21317458 | 4510 | 420 | AD | eGFR | -0.468343 | -1 | -0.07658 | 0.219345 | 0.00673 | 1 | 0.0222066 | 111 | 0.182874637719298 |
| Chr2:175.3Mb | 418 | 2 | 174714723 | 175906698 | 4707 | 446 | AD | eGFR | 0.744271 | 0.12483 | 1 | 0.553939 | 0.02207 | 1 | 0.0222265 | 112 | 0.182874637719298 |
| Chr8:40.6Mb | 1420 | 8 | 40176051 | 40978026 | 4172 | 395 | AD | eGFR | -0.684771 | -1 | -0.12778 | 0.468912 | 0.02136 | 1 | 0.0224384 | 113 | 0.182874637719298 |
| Chr12:33.5Mb | 1933 | 12 | 33076216 | 33839351 | 4184 | 251 | AD | eGFR | 0.55288 | 0.09513 | 1 | 0.305676 | 0.01179 | 1 | 0.0225869 | 114 | 0.182874637719298 |
| Chr6:19.8Mb | 1077 | 6 | 19448730 | 20157121 | 3667 | 450 | AD | eGFR | -0.737878 | -1 | -0.13845 | 0.544464 | 0.02319 | 1 | 0.0231646 | 115 | 0.185921093913043 |
| Chr8:61.8Mb | 1437 | 8 | 61288513 | 62217395 | 3689 | 390 | AD | eGFR | 0.548118 | 0.08324 | 1 | 0.300433 | 0.01001 | 1 | 0.0241358 | 116 | 0.191535911111111 |
| Chr3:120.6Mb | 607 | 3 | 120145746 | 120964887 | 3390 | 280 | AD | eGFR | 0.535134 | 0.06957 | 1 | 0.286368 | 0.00764 | 1 | 0.0242792 | 117 | 0.191535911111111 |
| Chr4:183.5Mb | 887 | 4 | 183112064 | 183807246 | 3770 | 503 | AD | eGFR | -0.40632 | -0.86752 | -0.05299 | 0.165096 | 0.0042 | 0.75259 | 0.0252512 | 118 | 0.194624226612903 |
| Chr12:128Mb | 2020 | 12 | 127547824 | 128373043 | 4628 | 530 | AD | eGFR | 0.438843 | 0.05991 | 0.99761 | 0.192583 | 0.0065 | 0.99523 | 0.0254103 | 119 | 0.194624226612903 |
| Chr13:45.9Mb | 2056 | 13 | 45479681 | 46349853 | 3471 | 374 | AD | eGFR | -0.39413 | -0.81546 | -0.05215 | 0.155339 | 0.00384 | 0.66497 | 0.0255005 | 120 | 0.194624226612903 |
| Chr12:80.4Mb | 1977 | 12 | 79756933 | 81134128 | 4801 | 381 | AD | eGFR | -0.637394 | -1 | -0.09233 | 0.406272 | 0.01327 | 1 | 0.0255969 | 121 | 0.194624226612903 |
| Chr3:87.5Mb | 578 | 3 | 86741867 | 88270959 | 6564 | 356 | AD | eGFR | -0.544533 | -1 | -0.0656 | 0.296516 | 0.00713 | 1 | 0.0260601 | 122 | 0.194624226612903 |
| Chr14:20Mb | 2132 | 14 | 19000060 | 20942037 | 2853 | 414 | AD | eGFR | -0.332995 | -0.66844 | -0.04412 | 0.110885 | 0.00309 | 0.44681 | 0.0261304 | 123 | 0.194624226612903 |

| locus_label | locus | chr | start | stop | n.snps | n.pcs | phen1 | phen2 | rho | rho.lower | rho.upper | r2 | r2.lower | r2.upper | p | rank | FDR |
|---|---|---|---|---|---|---|---|---|---|---|---|---|---|---|---|---|---|
| Chr4:184.8Mb | 889 | 4 | 184508773 | 185088918 | 3287 | 504 | AD | eGFR | 0.746515 | 0.08507 | 1 | 0.557285 | 0.01417 | 1 | 0.0261467 | 124 | 0.194624226612903 |
| Chr3:115.6Mb | 602 | 3 | 115236929 | 116048470 | 3446 | 441 | AD | eGFR | 0.484947 | 0.06141 | 1 | 0.235173 | 0.00607 | 1 | 0.0266181 | 125 | 0.1965480504 |
| Chr13:89.6Mb | 2100 | 13 | 89060617 | 90173070 | 4805 | 264 | AD | eGFR | 0.789399 | 0.10687 | 1 | 0.62315 | 0.02167 | 1 | 0.0271362 | 126 | 0.198783433333333 |
| Chr13:51.5Mb | 2061 | 13 | 51054677 | 51849614 | 3399 | 442 | AD | eGFR | 0.539117 | 0.0835 | 1 | 0.290648 | 0.00967 | 1 | 0.0280636 | 127 | 0.20395828976378 |
| Chr2:74.6Mb | 323 | 2 | 74129635 | 75122684 | 3660 | 409 | AD | eGFR | -0.557765 | -1 | -0.0658 | 0.311102 | 0.00772 | 1 | 0.028377 | 128 | 0.2046247734375 |
| Chr9:107.5Mb | 1603 | 9 | 107061377 | 107933334 | 5124 | 440 | AD | eGFR | -0.678684 | -1 | -0.07789 | 0.460612 | 0.01302 | 1 | 0.0286982 | 129 | 0.205336733333333 |
| Chr1:0.7Mb | 1 | 1 | 10505 | 1359454 | 2336 | 392 | AD | eGFR | -0.711693 | -1 | -0.07487 | 0.506507 | 0.01286 | 1 | 0.0290935 | 130 | 0.20656385 |
| Chr14:106.9Mb | 2214 | 14 | 106506452 | 107289451 | 2256 | 330 | AD | eGFR | 0.645659 | 0.07238 | 1 | 0.416876 | 0.01111 | 1 | 0.0298211 | 131 | 0.210113551908397 |
| Chr1:17.8Mb | 23 | 1 | 17496754 | 18190728 | 3757 | 464 | AD | eGFR | -0.416271 | -0.8987 | -0.03465 | 0.173282 | 0.00334 | 0.80766 | 0.0301851 | 132 | 0.211067025 |
| Chr12:115.9Mb | 2008 | 12 | 115438413 | 116380071 | 5054 | 534 | AD | eGFR | -0.560307 | -1 | -0.04361 | 0.313944 | 0.00646 | 1 | 0.0312715 | 133 | 0.215969601492537 |
| Chr15:31.4Mb | 2224 | 15 | 30638082 | 32177341 | 5350 | 473 | AD | eGFR | -0.608142 | -1 | -0.06987 | 0.369837 | 0.00905 | 1 | 0.0313542 | 134 | 0.215969601492537 |
| Chr7:144.6Mb | 1356 | 7 | 143925686 | 145174442 | 5454 | 434 | AD | eGFR | -0.908173 | -1 | -0.10086 | 0.824778 | 0.02257 | 1 | 0.0320283 | 135 | 0.217954659854015 |
| Chr3:118.6Mb | 605 | 3 | 118118783 | 119094221 | 4283 | 355 | AD | eGFR | -0.832264 | -1 | -0.09657 | 0.692664 | 0.01728 | 1 | 0.0321184 | 136 | 0.217954659854015 |
| Chr6:70.6Mb | 1133 | 6 | 70222512 | 71015024 | 3472 | 326 | AD | eGFR | -0.592553 | -1 | -0.06769 | 0.351119 | 0.00797 | 1 | 0.0323508 | 137 | 0.217954659854015 |
| Chr2:191.7Mb | 432 | 2 | 191036943 | 192331625 | 4953 | 388 | AD | eGFR | 0.405516 | 0.03618 | 0.89263 | 0.164443 | 0.00334 | 0.79678 | 0.0327293 | 138 | 0.217986038848921 |
| Chr18:44.5Mb | 2465 | 18 | 43876349 | 45128523 | 5646 | 451 | AD | eGFR | 0.786927 | 0.05448 | 1 | 0.619254 | 0.01241 | 1 | 0.0328278 | 139 | 0.217986038848921 |
| Chr8:49.7Mb | 1426 | 8 | 49045766 | 50288821 | 4600 | 312 | AD | eGFR | 0.643664 | 0.07075 | 1 | 0.414304 | 0.01007 | 1 | 0.0332866 | 140 | 0.219453798571429 |
| Chr5:69Mb | 963 | 5 | 67295903 | 70672360 | 5932 | 493 | AD | eGFR | -0.331771 | -0.67734 | -0.02212 | 0.110072 | 0.00188 | 0.45878 | 0.0335372 | 141 | 0.219537841134752 |
| Chr1:168.7Mb | 152 | 1 | 168379444 | 169088053 | 3635 | 440 | AD | eGFR | -0.633939 | -1 | -0.04959 | 0.401878 | 0.00792 | 1 | 0.0340102 | 142 | 0.2210663 |
| Chr7:48.1Mb | 1274 | 7 | 47720491 | 48456573 | 3717 | 349 | AD | eGFR | -0.544336 | -1 | -0.0442 | 0.296301 | 0.00661 | 1 | 0.0342566 | 143 | 0.221110781818182 |
| Chr3:148.8Mb | 633 | 3 | 148358957 | 149143607 | 3819 | 432 | AD | eGFR | 0.515144 | 0.03999 | 1 | 0.265374 | 0.00628 | 1 | 0.0345186 | 144 | 0.2212546375 |
| Chr9:30.6Mb | 1553 | 9 | 30077282 | 31115154 | 5254 | 272 | AD | eGFR | -0.883693 | -1 | -0.07911 | 0.780914 | 0.01697 | 1 | 0.0352649 | 145 | 0.224479328965517 |
| Chr4:139.8Mb | 839 | 4 | 139403464 | 140176426 | 3536 | 404 | AD | eGFR | 0.509576 | 0.03602 | 1 | 0.259668 | 0.00504 | 1 | 0.0357523 | 146 | 0.226023102054795 |
| Chr1:120Mb | 128 | 1 | 119581605 | 120347432 | 3367 | 374 | AD | eGFR | 0.608098 | 0.04767 | 1 | 0.369783 | 0.00828 | 1 | 0.0363123 | 147 | 0.228001720408163 |
| Chr3:159.1Mb | 645 | 3 | 158578925 | 159700710 | 4987 | 433 | AD | eGFR | 0.292716 | 0.01557 | 0.62954 | 0.0856824 | 0.00155 | 0.39633 | 0.0366647 | 148 | 0.228658906081081 |
| Chr15:38.9Mb | 2230 | 15 | 38377990 | 39517797 | 5497 | 508 | AD | eGFR | -0.401719 | -0.93109 | -0.02418 | 0.161378 | 0.00327 | 0.86694 | 0.0373051 | 149 | 0.231091324161074 |
| Chr9:120.1Mb | 1616 | 9 | 119557908 | 120600041 | 4886 | 458 | AD | eGFR | -0.37842 | -0.84632 | -0.02074 | 0.143202 | 0.0027 | 0.71626 | 0.0376875 | 150 | 0.23190375 |
| Chr6:71.4Mb | 1134 | 6 | 71015025 | 71819902 | 3523 | 318 | AD | eGFR | -0.648957 | -1 | -0.04339 | 0.421146 | 0.00957 | 1 | 0.0392344 | 151 | 0.239778397368421 |
| Chr10:131.9Mb | 1764 | 10 | 131463050 | 132346400 | 4845 | 491 | AD | eGFR | 0.607044 | 0.02097 | 1 | 0.368502 | 0.00567 | 1 | 0.0394868 | 152 | 0.239778397368421 |
| Chr3:11.2Mb | 503 | 3 | 10776480 | 11625632 | 3552 | 441 | AD | eGFR | -0.312118 | -0.65268 | -0.01274 | 0.0974177 | 0.00143 | 0.42599 | 0.0411326 | 153 | 0.248139802614379 |
| Chr11:6Mb | 1776 | 11 | 5611305 | 6477448 | 4430 | 472 | AD | eGFR | 0.477534 | 0.01812 | 1 | 0.228039 | 0.00347 | 1 | 0.0416153 | 154 | 0.249064701935484 |
| Chr6:20.8Mb | 1078 | 6 | 20157122 | 21484707 | 6435 | 502 | AD | eGFR | 0.471278 | 0.02984 | 1 | 0.222103 | 0.00422 | 1 | 0.0418256 | 155 | 0.249064701935484 |

| locus_label | locus | chr | start | stop | n.snps | n.pcs | phen1 | phen2 | rho | rho.lower | rho.upper | r2 | r2.lower | r2.upper | p | rank | FDR |
|---|---|---|---|---|---|---|---|---|---|---|---|---|---|---|---|---|---|
| Chr6:168.3Mb | 1223 | 6 | 167808368 | 168718581 | 5100 | 519 | AD | eGFR | 0.399465 | 0.0078 | 1 | 0.159572 | 0.0019 | 1 | 0.0433586 | 156 | 0.256538383333333 |
| Chr13:31.3Mb | 2042 | 13 | 30872647 | 31693457 | 3986 | 505 | AD | eGFR | 0.433741 | 0.01625 | 1 | 0.188131 | 0.0023 | 1 | 0.043979 | 157 | 0.2569632 |
| Chr6:163.3Mb | 1218 | 6 | 162929107 | 163615965 | 3328 | 325 | AD | eGFR | -0.534968 | -1 | -0.02324 | 0.286191 | 0.00503 | 1 | 0.0439872 | 158 | 0.2569632 |
| Chr14:98.4Mb | 2206 | 14 | 97967918 | 98751682 | 4056 | 439 | AD | eGFR | 0.692484 | 0.01138 | 1 | 0.479534 | 0.0087 | 1 | 0.0446383 | 159 | 0.258220645061728 |
| Chr10:3.8Mb | 1643 | 10 | 3357439 | 4300374 | 5042 | 503 | AD | eGFR | -0.463553 | -1 | 0.00335 | 0.214881 | 0 | 1 | 0.0451756 | 160 | 0.258220645061728 |
| Chr7:41.4Mb | 1268 | 7 | 40781543 | 42094599 | 5537 | 480 | AD | eGFR | 0.47281 | 0.01714 | 1 | 0.22355 | 0.0037 | 1 | 0.0452748 | 161 | 0.258220645061728 |
| Chr9:98.6Mb | 1593 | 9 | 98132586 | 99084179 | 4550 | 439 | AD | eGFR | 0.273131 | 0.0083 | 0.56058 | 0.0746003 | 0.00107 | 0.31425 | 0.0453215 | 162 | 0.258220645061728 |
| Chr8:98.6Mb | 1469 | 8 | 98130796 | 99109424 | 4623 | 450 | AD | eGFR | -0.483872 | -1 | 0.00587 | 0.234132 | 0 | 1 | 0.0463164 | 163 | 0.262270166871166 |
| Chr1:204.5Mb | 189 | 1 | 204071164 | 204922955 | 3912 | 443 | AD | eGFR | 0.479036 | 0.00564 | 1 | 0.229475 | 0.00301 | 1 | 0.0469488 | 164 | 0.264164268072289 |
| Chr10:4.7Mb | 1644 | 10 | 4300375 | 5114049 | 4754 | 474 | AD | eGFR | -0.608143 | -1 | -0.00131 | 0.369838 | 0.00569 | 1 | 0.0474438 | 165 | 0.264164268072289 |
| Chr18:65.1Mb | 2482 | 18 | 64708992 | 65490203 | 4476 | 420 | AD | eGFR | 0.475966 | -0.0094 | 1 | 0.226544 | 0 | 1 | 0.0475095 | 166 | 0.264164268072289 |
| Chr1:12.8Mb | 17 | 1 | 12266602 | 13382928 | 2335 | 385 | AD | eGFR | 0.533836 | -0.00869 | 1 | 0.28498 | 0 | 1 | 0.0486873 | 167 | 0.269092083233533 |
| Chr7:0.5Mb | 1226 | 7 | 14808 | 937564 | 3910 | 504 | AD | eGFR | -0.575016 | -1 | -0.0062 | 0.330643 | 0.00425 | 1 | 0.0493585 | 168 | 0.269694122807018 |
| Chr15:32.9Mb | 2225 | 15 | 32177342 | 33540410 | 4975 | 507 | AD | eGFR | 0.450287 | 0 | 1 | 0.202758 | 0 | 1 | 0.0498714 | 169 | 0.269694122807018 |
| Chr3:186.6Mb | 672 | 3 | 186226757 | 186894704 | 3309 | 473 | AD | eGFR | 0.426957 | 0.00593 | 1 | 0.182292 | 0.00262 | 1 | 0.0498803 | 170 | 0.269694122807018 |
| Chr21:37.4Mb | 2644 | 21 | 36818143 | 37994704 | 5716 | 459 | AD | eGFR | -0.385802 | -0.89856 | 0.00154 | 0.148843 | 0 | 0.80741 | 0.049965 | 171 | 0.269694122807018 |
| Chr12:131.7Mb | 2025 | 12 | 131404541 | 132011601 | 3610 | 459 | AD | eGFR | 0.726102 | 0.01596 | 1 | 0.527224 | 0.00751 | 1 | 0.0506041 | 172 | 0.271555722674419 |
| Chr1:162Mb | 145 | 1 | 161567518 | 162445320 | 3643 | 425 | AD | eGFR | -0.38161 | -0.91244 | -0.00079 | 0.145626 | 0.00177 | 0.83255 | 0.0511274 | 173 | 0.271754009770115 |
| Chr4:184.2Mb | 888 | 4 | 183807247 | 184508772 | 3660 | 423 | AD | eGFR | -0.644003 | -1 | 0.00199 | 0.414739 | 0 | 1 | 0.0512299 | 174 | 0.271754009770115 |
| Chr11:80.8Mb | 1846 | 11 | 80385741 | 81213005 | 4556 | 394 | AD | eGFR | 0.819173 | 0.00208 | 1 | 0.671044 | 0.0095 | 1 | 0.0518669 | 175 | 0.272322945555556 |
| Chr19:35.2Mb | 2531 | 19 | 34637027 | 35692489 | 4399 | 434 | AD | eGFR | 0.553966 | 0.00929 | 1 | 0.306878 | 0.00407 | 1 | 0.0521596 | 176 | 0.272322945555556 |
| Chr12:28.1Mb | 1927 | 12 | 27529206 | 28739039 | 4961 | 437 | AD | eGFR | 0.78998 | 0.0013 | 1 | 0.624068 | 0.00937 | 1 | 0.0522602 | 177 | 0.272322945555556 |
| Chr4:108.4Mb | 807 | 4 | 107500911 | 109375128 | 6883 | 477 | AD | eGFR | -0.855212 | -1 | 0.02384 | 0.731387 | 0 | 1 | 0.0528395 | 178 | 0.272322945555556 |
| Chr4:38.4Mb | 737 | 4 | 37911854 | 38986235 | 5750 | 519 | AD | eGFR | 0.581467 | 0.01027 | 1 | 0.338104 | 0.00431 | 1 | 0.0529605 | 179 | 0.272322945555556 |
| Chr11:69.3Mb | 1835 | 11 | 68801334 | 69793989 | 4855 | 500 | AD | eGFR | -0.623066 | -1 | 0.00352 | 0.388211 | 0 | 1 | 0.0531074 | 180 | 0.272322945555556 |
| Chr9:26.8Mb | 1548 | 9 | 26449871 | 27159584 | 4000 | 351 | AD | eGFR | -0.480029 | -1 | 0.01585 | 0.230427 | 0 | 1 | 0.0561186 | 181 | 0.286173855248619 |
| Chr13:24.2Mb | 2033 | 13 | 23842814 | 24534812 | 4138 | 418 | AD | eGFR | -0.486875 | -1 | 0.00141 | 0.237047 | 0 | 1 | 0.0578387 | 182 | 0.2920487375 |
| Chr8:5.6Mb | 1381 | 8 | 5410536 | 5853366 | 4028 | 435 | AD | eGFR | 0.530091 | -0.01452 | 1 | 0.280997 | 0 | 1 | 0.058152 | 183 | 0.2920487375 |
| Chr3:18.9Mb | 512 | 3 | 18415431 | 19338054 | 3567 | 367 | AD | eGFR | -0.486668 | -1 | 0.0237 | 0.236845 | 0 | 1 | 0.0582199 | 184 | 0.2920487375 |
| Chr1:76.4Mb | 84 | 1 | 75991177 | 76775178 | 3597 | 322 | AD | eGFR | -0.540877 | -1 | 0.01982 | 0.292548 | 0 | 1 | 0.0589396 | 185 | 0.294060815135135 |
| Chr10:115.8Mb | 1748 | 10 | 115330651 | 116364396 | 4971 | 474 | AD | eGFR | -0.450879 | -1 | 0.01533 | 0.203292 | 0 | 1 | 0.0598708 | 186 | 0.297100797849462 |
| Chr2:165.8Mb | 408 | 2 | 165205274 | 166395568 | 4649 | 409 | AD | eGFR | 0.598538 | -0.01673 | 1 | 0.358248 | 0 | 1 | 0.0611552 | 187 | 0.301851602139037 |
| Chr2:220.2Mb | 458 | 2 | 219684223 | 220693042 | 3944 | 439 | AD | eGFR | -0.403872 | -1 | -0.00023 | 0.163112 | 0.00219 | 1 | 0.0622596 | 188 | 0.305668142553191 |
| Chr16:10.2Mb | 2300 | 16 | 9877644 | 10434117 | 3411 | 399 | AD | eGFR | -0.376231 | -0.89312 | 0.01792 | 0.14155 | 0 | 0.79841 | 0.0643988 | 189 | 0.314497843386243 |
| Chr4:22.1Mb | 716 | 4 | 21696551 | 22525039 | 4624 | 365 | AD | eGFR | -0.48597 | -1 | 0.01832 | 0.236167 | 0 | 1 | 0.0653395 | 190 | 0.31717914611399 |

| locus_label | locus | chr | start | stop | n.snps | n.pcs | phen1 | phen2 | rho | rho.lower | rho.upper | r2 | r2.lower | r2.upper | p | rank | FDR |
|---|---|---|---|---|---|---|---|---|---|---|---|---|---|---|---|---|---|
| Chr6:157.9Mb | 1211 | 6 | 157421696 | 158392668 | 4019 | 434 | AD | eGFR | 0.592418 | -0.04573 | 1 | 0.35096 | 0 | 1 | 0.0659952 | 191 | 0.31717914611399 |
| Chr16:21.5Mb | 2309 | 16 | 20264005 | 22698695 | 5394 | 474 | AD | eGFR | -0.174077 | -0.36323 | 0.00871 | 0.0303029 | 0 | 0.13194 | 0.0661313 | 192 | 0.31717914611399 |
| Chr9:128.5Mb | 1624 | 9 | 127985943 | 129061376 | 4402 | 408 | AD | eGFR | 0.564267 | -0.02861 | 1 | 0.318398 | 0 | 1 | 0.0663224 | 193 | 0.31717914611399 |
| Chr4:187.3Mb | 893 | 4 | 186970654 | 187563492 | 3275 | 445 | AD | eGFR | -0.383587 | -1 | 0.02391 | 0.147139 | 0 | 1 | 0.0672467 | 194 | 0.31994177371134 |
| Chr4:84.8Mb | 783 | 4 | 84465634 | 85145677 | 3310 | 394 | AD | eGFR | 0.431629 | -0.02918 | 1 | 0.186303 | 0 | 1 | 0.0676008 | 195 | 0.31997712 |
| Chr5:111.4Mb | 1000 | 5 | 110843368 | 111987725 | 5873 | 437 | AD | eGFR | 0.450601 | -0.02745 | 1 | 0.203041 | 0 | 1 | 0.0679683 | 196 | 0.320075208673469 |
| Chr7:9.8Mb | 1237 | 7 | 9381316 | 10136829 | 4838 | 423 | AD | eGFR | 0.348588 | -0.02015 | 0.82466 | 0.121514 | 0 | 0.68007 | 0.068333 | 197 | 0.320159182741117 |
| Chr6:44.7Mb | 1110 | 6 | 44019199 | 45401733 | 5898 | 453 | AD | eGFR | 0.294531 | -0.02338 | 0.66923 | 0.0867487 | 0 | 0.44787 | 0.0689544 | 198 | 0.321438945454545 |
| Chr20:16.2Mb | 2576 | 20 | 15796810 | 16660088 | 4808 | 448 | AD | eGFR | 0.550617 | -0.05403 | 1 | 0.303179 | 0 | 1 | 0.0696427 | 199 | 0.32301614120603 |
| Chr11:36.5Mb | 1807 | 11 | 36125684 | 36900413 | 4009 | 418 | AD | eGFR | 0.411342 | -0.03491 | 1 | 0.169202 | 0 | 1 | 0.0701003 | 200 | 0.3235128845 |
| Chr1:159.4Mb | 142 | 1 | 158899795 | 159822095 | 3615 | 401 | AD | eGFR | -0.354239 | -0.83905 | 0.03355 | 0.125485 | 0 | 0.704 | 0.071317 | 201 | 0.327490502487562 |
| Chr14:58.9Mb | 2169 | 14 | 58300003 | 59449546 | 4256 | 427 | AD | eGFR | -0.468943 | -1 | 0.05284 | 0.219908 | 0 | 1 | 0.0717187 | 202 | 0.327704752970297 |
| Chr2:139.3Mb | 383 | 2 | 138878413 | 139810112 | 3829 | 348 | AD | eGFR | 0.532618 | -0.0497 | 1 | 0.283682 | 0 | 1 | 0.0730906 | 203 | 0.332222588516746 |
| Chr1:67.7Mb | 75 | 1 | 67359902 | 68125319 | 3356 | 404 | AD | eGFR | -0.478456 | -1 | 0.05995 | 0.22892 | 0 | 1 | 0.0738518 | 204 | 0.332222588516746 |
| Chr11:128.9Mb | 1892 | 11 | 128375176 | 129494348 | 4968 | 512 | AD | eGFR | -0.595356 | -1 | 0.07158 | 0.354449 | 0 | 1 | 0.0741279 | 205 | 0.332222588516746 |
| Chr7:150.9Mb | 1362 | 7 | 150453552 | 151443100 | 5092 | 533 | AD | eGFR | 0.190839 | -0.0178 | 0.41103 | 0.0364194 | 0 | 0.16895 | 0.0744534 | 206 | 0.332222588516746 |
| Chr2:115.8Mb | 357 | 2 | 115204695 | 116411514 | 4972 | 402 | AD | eGFR | -0.610692 | -1 | 0.06015 | 0.372945 | 0 | 1 | 0.0748527 | 207 | 0.332222588516746 |
| Chr12:15.2Mb | 1915 | 12 | 14661771 | 15816042 | 4787 | 391 | AD | eGFR | 0.290374 | -0.02947 | 0.64768 | 0.084317 | 0 | 0.41949 | 0.0748802 | 208 | 0.332222588516746 |
| Chr15:62.4Mb | 2252 | 15 | 61805518 | 63018961 | 5489 | 480 | AD | eGFR | 0.466988 | -0.05356 | 1 | 0.218078 | 0 | 1 | 0.075227 | 209 | 0.332222588516746 |
| Chr17:74.3Mb | 2421 | 17 | 73738588 | 74848710 | 5323 | 488 | AD | eGFR | 0.376641 | -0.0317 | 1 | 0.141859 | 0 | 1 | 0.0758931 | 210 | 0.333568244285714 |
| Chr8:26.1Mb | 1407 | 8 | 25571730 | 26678701 | 5563 | 463 | AD | eGFR | 0.297048 | -0.02617 | 0.66787 | 0.0882376 | 0 | 0.44605 | 0.0768668 | 211 | 0.336246712796209 |
| Chr14:57.8Mb | 2168 | 14 | 57351066 | 58300002 | 4256 | 400 | AD | eGFR | -0.379922 | -0.92624 | 0.04869 | 0.144341 | 0 | 0.8627 | 0.0775539 | 212 | 0.337652121226415 |
| Chr1:116.5Mb | 124 | 1 | 116026338 | 117021533 | 4759 | 394 | AD | eGFR | -0.404292 | -1 | 0.06049 | 0.163452 | 0 | 1 | 0.0790196 | 213 | 0.342367872429907 |
| Chr4:166.6Mb | 866 | 4 | 166255334 | 167016476 | 4228 | 420 | AD | eGFR | -0.451086 | -1 | 0.05644 | 0.203479 | 0 | 1 | 0.0793789 | 214 | 0.342367872429907 |
| Chr7:103.7Mb | 1322 | 7 | 103186233 | 104157174 | 4995 | 450 | AD | eGFR | -0.541207 | -1 | 0.06223 | 0.292905 | 0 | 1 | 0.0819534 | 215 | 0.351827852093023 |
| Chr2:10.8Mb | 253 | 2 | 10468904 | 11113631 | 3937 | 486 | AD | eGFR | 0.516681 | -0.07818 | 1 | 0.266959 | 0 | 1 | 0.087094 | 216 | 0.366260536036036 |
| Chr9:80.7Mb | 1575 | 9 | 80226639 | 81241434 | 4393 | 416 | AD | eGFR | -0.477533 | -1 | 0.06892 | 0.228037 | 0 | 1 | 0.0872074 | 217 | 0.366260536036036 |
| Chr4:85.7Mb | 784 | 4 | 85145678 | 86182800 | 3827 | 318 | AD | eGFR | 0.396647 | -0.06673 | 1 | 0.157329 | 0 | 1 | 0.0872329 | 218 | 0.366260536036036 |
| Chr4:132.3Mb | 831 | 4 | 131623098 | 132958221 | 5077 | 350 | AD | eGFR | 0.519492 | -0.09895 | 1 | 0.269871 | 0 | 1 | 0.0878079 | 219 | 0.366260536036036 |
| Chr1:10.3Mb | 14 | 1 | 9890598 | 10807358 | 3512 | 376 | AD | eGFR | 0.510542 | -0.0711 | 1 | 0.260653 | 0 | 1 | 0.0879121 | 220 | 0.366260536036036 |
| Chr11:20.5Mb | 1790 | 11 | 20015987 | 20944092 | 5088 | 497 | AD | eGFR | -0.581016 | -1 | 0.10185 | 0.337579 | 0 | 1 | 0.0880356 | 221 | 0.366260536036036 |
| Chr14:91.4Mb | 2199 | 14 | 90665759 | 92141002 | 6653 | 513 | AD | eGFR | 0.372039 | -0.05291 | 0.97771 | 0.138413 | 0 | 0.95591 | 0.088093 | 222 | 0.366260536036036 |
| Chr4:15.8Mb | 709 | 4 | 15342994 | 16244635 | 3897 | 441 | AD | eGFR | 0.492197 | -0.11051 | 1 | 0.242258 | 0 | 1 | 0.0887917 | 223 | 0.367510040807175 |

| locus_label | locus | chr | start | stop | n.snps | n.pcs | phen1 | phen2 | rho | rho.lower | rho.upper | r2 | r2.lower | r2.upper | p | rank | FDR |
|---|---|---|---|---|---|---|---|---|---|---|---|---|---|---|---|---|---|
| Chr12:10.4Mb | 1910 | 12 | 10002351 | 10822910 | 4381 | 417 | AD | eGFR | -0.462196 | -1 | 0.05808 | 0.213625 | 0 | 1 | 0.0894738 | 224 | 0.368233899559471 |
| Chr1:16.1Mb | 21 | 1 | 15747106 | 16517143 | 3615 | 371 | AD | eGFR | -0.416513 | -1 | 0.06898 | 0.173483 | 0 | 1 | 0.0903754 | 225 | 0.368233899559471 |
| Chr5:76.7Mb | 969 | 5 | 76252855 | 77124514 | 3743 | 447 | AD | eGFR | 0.345669 | -0.05418 | 0.93951 | 0.119487 | 0 | 0.88408 | 0.0905527 | 226 | 0.368233899559471 |
| Chr8:13.2Mb | 1391 | 8 | 12988092 | 13425611 | 3213 | 351 | AD | eGFR | -0.420153 | -1 | 0.07409 | 0.176529 | 0 | 1 | 0.0905624 | 227 | 0.368233899559471 |
| Chr10:135.1Mb | 1768 | 10 | 134726849 | 135524372 | 3755 | 450 | AD | eGFR | -0.417984 | -1 | 0.06803 | 0.174711 | 0 | 1 | 0.092459 | 228 | 0.37276478652173 9 |
| Chr13:71.9Mb | 2081 | 13 | 71427785 | 72331636 | 3730 | 387 | AD | eGFR | -0.183697 | -0.43618 | 0.03408 | 0.0337444 | 0 | 0.19026 | 0.0927261 | 229 | 0.372764786521739 |
| Chr2:34.6Mb | 279 | 2 | 34235016 | 34890734 | 3613 | 370 | AD | eGFR | -0.381786 | -1 | 0.0693 | 0.145761 | 0 | 1 | 0.0928883 | 230 | 0.372764786521739 |
| Chr9:97.4Mb | 1592 | 9 | 96679610 | 98132585 | 5493 | 360 | AD | eGFR | -0.307599 | -0.83683 | 0.05288 | 0.094617 | 0 | 0.70029 | 0.0961796 | 231 | 0.384302038095238 |
| Chr11:75.7Mb | 1841 | 11 | 75144190 | 76353165 | 5139 | 451 | AD | eGFR | -0.479998 | -1 | 0.0923 | 0.230398 | 0 | 1 | 0.0984507 | 232 | 0.389700855555556 |
| Chr2:20Mb | 264 | 2 | 19629636 | 20410561 | 3916 | 411 | AD | eGFR | 0.290047 | -0.05472 | 0.69586 | 0.0841271 | 0 | 0.48422 | 0.0985977 | 233 | 0.389700855555556 |
| Chr6:74Mb | 1137 | 6 | 73539517 | 74395508 | 3559 | 380 | AD | eGFR | -0.260185 | -0.60143 | 0.04521 | 0.0676963 | 0 | 0.36172 | 0.0987974 | 234 | 0.389700855555556 |
| Chr2:180.2Mb | 422 | 2 | 179573439 | 180776608 | 5175 | 424 | AD | eGFR | -0.292586 | -0.70423 | 0.06956 | 0.0856065 | 0 | 0.49594 | 0.0998044 | 235 | 0.391715838174274 |
| Chr14:60.1Mb | 2170 | 14 | 59449547 | 60663448 | 4903 | 321 | AD | eGFR | 0.250079 | -0.04547 | 0.68357 | 0.0625397 | 0 | 0.46864 | 0.101047 | 236 | 0.391715838174274 |
| Chr4:37Mb | 735 | 4 | 36734793 | 37318906 | 3472 | 309 | AD | eGFR | 0.627792 | -0.15602 | 1 | 0.394123 | 0 | 1 | 0.101326 | 237 | 0.391715838174274 |
| Chr1:150.2Mb | 133 | 1 | 149421695 | 150976251 | 3803 | 256 | AD | eGFR | -0.560479 | -1 | 0.10664 | 0.314137 | 0 | 1 | 0.101698 | 238 | 0.391715838174274 |
| Chr6:82.8Mb | 1147 | 6 | 82390065 | 83297550 | 3694 | 349 | AD | eGFR | -0.351895 | -0.88036 | 0.06646 | 0.12383 | 0 | 0.77556 | 0.101809 | 239 | 0.391715838174274 |
| Chr7:52.7Mb | 1280 | 7 | 52499355 | 52989121 | 3726 | 257 | AD | eGFR | -0.457988 | -1 | 0.09975 | 0.209753 | 0 | 1 | 0.102002 | 240 | 0.391715838174274 |
| Chr17:73.1Mb | 2420 | 17 | 72385919 | 73738587 | 6050 | 496 | AD | eGFR | 0.412371 | -0.0966 | 1 | 0.17005 | 0 | 1 | 0.102279 | 241 | 0.391715838174274 |
| Chr3:112.9Mb | 600 | 3 | 112519270 | 113309390 | 4030 | 319 | AD | eGFR | 0.31605 | -0.06649 | 0.77348 | 0.0998875 | 0 | 0.59828 | 0.10432 | 242 | 0.397881652892562 |
| Chr6:153.1Mb | 1207 | 6 | 152417752 | 153694693 | 6209 | 491 | AD | eGFR | 0.375773 | -0.09048 | 1 | 0.141205 | 0 | 1 | 0.105192 | 243 | 0.398558209016393 |
| Chr3:104.5Mb | 591 | 3 | 104116589 | 104829897 | 3681 | 302 | AD | eGFR | -0.434128 | -1 | 0.10192 | 0.188467 | 0 | 1 | 0.105361 | 244 | 0.398558209016393 |
| Chr13:24.8Mb | 2034 | 13 | 24534813 | 25092419 | 3575 | 480 | AD | eGFR | -0.401052 | -1 | 0.09713 | 0.160843 | 0 | 1 | 0.106434 | 245 | 0.400973804081633 |
| Chr15:53.8Mb | 2244 | 15 | 53381240 | 54165792 | 4169 | 403 | AD | eGFR | -0.35527 | -1 | 0.0815 | 0.126217 | 0 | 1 | 0.107818 | 246 | 0.404536642276423 |
| Chr3:34Mb | 529 | 3 | 33195529 | 34816866 | 6137 | 405 | AD | eGFR | -0.348294 | -0.89626 | 0.08137 | 0.121309 | 0 | 0.80395 | 0.108994 | 247 | 0.405423116465863 |
| Chr7:101.8Mb | 1320 | 7 | 101200585 | 102332526 | 3513 | 413 | AD | eGFR | 0.417431 | -0.09659 | 1 | 0.174248 | 0 | 1 | 0.109183 | 248 | 0.405423116465863 |
| Chr8:81.7Mb | 1454 | 8 | 81089813 | 82267431 | 5151 | 460 | AD | eGFR | 0.360922 | -0.07696 | 1 | 0.130264 | 0 | 1 | 0.109372 | 249 | 0.405423116465863 |
| Chr11:35.7Mb | 1806 | 11 | 35328321 | 36125683 | 3746 | 474 | AD | eGFR | 0.396652 | -0.08766 | 1 | 0.157332 | 0 | 1 | 0.110456 | 250 | 0.407675494023904 |
| Chr12:49.7Mb | 1950 | 12 | 49151269 | 50339059 | 3593 | 416 | AD | eGFR | 0.264351 | -0.05332 | 0.61955 | 0.0698814 | 0 | 0.38384 | 0.110863 | 251 | 0.407675494023904 |
| Chr4:165.9Mb | 865 | 4 | 165557683 | 166255333 | 3920 | 399 | AD | eGFR | 0.426758 | -0.10715 | 1 | 0.182123 | 0 | 1 | 0.111613 | 252 | 0.408609192913386 |
| Chr13:53.2Mb | 2063 | 13 | 52880747 | 53606081 | 2939 | 332 | AD | eGFR | 0.464043 | -0.11783 | 1 | 0.215335 | 0 | 1 | 0.112019 | 253 | 0.408609192913386 |
| Chr8:129.8Mb | 1495 | 8 | 129306459 | 130378156 | 5151 | 407 | AD | eGFR | -0.646751 | -1 | 0.17919 | 0.418287 | 0 | 1 | 0.112445 | 254 | 0.408609192913386 |
| Chr4:6.5Mb | 696 | 4 | 6265235 | 6767782 | 3295 | 510 | AD | eGFR | -0.379705 | -1 | 0.10003 | 0.144176 | 0 | 1 | 0.113297 | 255 | 0.410090709803922 |
| Chr12:30Mb | 1929 | 12 | 29648751 | 30359017 | 4201 | 420 | AD | eGFR | -0.343301 | -0.95365 | 0.08917 | 0.117856 | 0 | 0.91027 | 0.113939 | 256 | 0.410242972762646 |
| Chr6:54.1Mb | 1119 | 6 | 53559343 | 54633142 | 4979 | 361 | AD | eGFR | -0.288952 | -0.71977 | 0.07304 | 0.0834933 | 0 | 0.51854 | 0.114228 | 257 | 0.410242972762646 |
| Chr16:23.4Mb | 2310 | 16 | 22698696 | 24096055 | 5712 | 499 | AD | eGFR | 0.497713 | -0.10707 | 1 | 0.247718 | 0 | 1 | 0.114978 | 258 | 0.411336023255814 |
| Chr10:29.9Mb | 1671 | 10 | 29531856 | 30222721 | 3883 | 443 | AD | eGFR | 0.33473 | -0.09665 | 0.89363 | 0.112044 | 0 | 0.79857 | 0.116245 | 259 | 0.41426306949807 |
| Chr5:53.2Mb | 949 | 5 | 52665456 | 53703869 | 4652 | 399 | AD | eGFR | -0.346021 | -0.99826 | 0.09451 | 0.11973 | 0 | 0.99653 | 0.11716 | 260 | 0.415918 |

| locus_label | locus | chr | start | stop | n.snps | n.pcs | phen1 | phen2 | rho | rho.lower | rho.upper | r2 | r2.lower | r2.upper | p | rank | FDR |
|---|---|---|---|---|---|---|---|---|---|---|---|---|---|---|---|---|---|
| Chr2:38.3Mb | 284 | 2 | 37959619 | 38731387 | 4094 | 436 | AD | eGFR | 0.347707 | -0.0988 | 1 | 0.1209 | 0 | 1 | 0.117638 | 261 | 0.416014842911877 |
| Chr20:59.4Mb | 2617 | 20 | 58962376 | 59777918 | 4330 | 501 | AD | eGFR | -0.234985 | -0.564 | 0.05935 | 0.0552178 | 0 | 0.31809 | 0.118546 | 262 | 0.41762579389313 |
| Chr9:75.2Mb | 1569 | 9 | 74623888 | 75785593 | 4700 | 400 | AD | eGFR | 0.471201 | -0.14562 | 1 | 0.22203 | 0 | 1 | 0.119309 | 263 | 0.418715615969582 |
| Chr3:126.6Mb | 613 | 3 | 126216594 | 127082016 | 3839 | 370 | AD | eGFR | -0.295923 | -0.74926 | 0.08057 | 0.0875702 | 0 | 0.56138 | 0.120799 | 264 | 0.422338928030303 |
| Chr18:45.7Mb | 2466 | 18 | 45128524 | 46363231 | 5572 | 523 | AD | eGFR | 0.267759 | -0.07779 | 0.7293 | 0.0716946 | 0 | 0.53431 | 0.121917 | 265 | 0.424639211320755 |
| Chr4:110.6Mb | 809 | 4 | 110185542 | 110987607 | 3282 | 348 | AD | eGFR | 0.436038 | -0.12459 | 1 | 0.190129 | 0 | 1 | 0.123116 | 266 | 0.426246239700375 |
| Chr9:90.3Mb | 1585 | 9 | 89865702 | 90689576 | 4348 | 483 | AD | eGFR | -0.380677 | -1 | 0.09876 | 0.144915 | 0 | 1 | 0.123302 | 267 | 0.426246239700375 |
| Chr18:61.7Mb | 2479 | 18 | 61178036 | 62286818 | 5507 | 437 | AD | eGFR | -0.38032 | -1 | 0.11884 | 0.144643 | 0 | 1 | 0.124637 | 268 | 0.429253548507463 |
| Chr1:222.3Mb | 208 | 1 | 221856635 | 222730493 | 3321 | 408 | AD | eGFR | -0.274265 | -0.71184 | 0.08085 | 0.0752214 | 0 | 0.50671 | 0.125822 | 269 | 0.431723814126394 |
| Chr12:53.6Mb | 1953 | 12 | 53036281 | 54149495 | 4661 | 449 | AD | eGFR | 0.351606 | -0.10397 | 1 | 0.123627 | 0 | 1 | 0.127334 | 270 | 0.435293637037037 |
| Chr1:23.5Mb | 30 | 1 | 22969974 | 23973227 | 3665 | 432 | AD | eGFR | -0.293547 | -0.76367 | 0.09035 | 0.0861697 | 0 | 0.58319 | 0.129866 | 271 | 0.442311136531365 |
| Chr10:97.4Mb | 1732 | 10 | 96756083 | 97986657 | 4864 | 369 | AD | eGFR | -0.437446 | -1 | 0.14727 | 0.191359 | 0 | 1 | 0.130862 | 272 | 0.444064801470588 |
| Chr6:137.4Mb | 1193 | 6 | 136997275 | 137875770 | 3835 | 432 | AD | eGFR | 0.388379 | -0.1272 | 1 | 0.150838 | 0 | 1 | 0.132134 | 273 | 0.446132386861314 |
| Chr6:46.1Mb | 1111 | 6 | 45401734 | 46729085 | 5429 | 476 | AD | eGFR | 0.362432 | -0.10584 | 1 | 0.131357 | 0 | 1 | 0.132438 | 274 | 0.446132386861314 |
| Chr6:43.2Mb | 1109 | 6 | 42458360 | 44019198 | 6243 | 507 | AD | eGFR | 0.248464 | -0.07073 | 0.62327 | 0.0617345 | 0 | 0.38925 | 0.13298 | 275 | 0.446329236363636 |
| Chr10:106.6Mb | 1739 | 10 | 106134194 | 107043496 | 3857 | 352 | AD | eGFR | 0.316047 | -0.09293 | 0.84958 | 0.0998859 | 0 | 0.72179 | 0.133843 | 276 | 0.447598148550725 |
| Chr19:0.4Mb | 2495 | 19 | 64705 | 825194 | 3194 | 538 | AD | eGFR | 0.256478 | -0.08029 | 0.64759 | 0.0657811 | 0 | 0.41937 | 0.135328 | 277 | 0.448224017921147 |
| Chr19:4.3Mb | 2500 | 19 | 3899294 | 4743472 | 3899 | 499 | AD | eGFR | -0.259194 | -0.66339 | 0.07676 | 0.0671815 | 0 | 0.44107 | 0.13545 | 278 | 0.448224017921147 |
| Chr19:36.2Mb | 2532 | 19 | 35692490 | 36643012 | 4170 | 473 | AD | eGFR | 0.277237 | -0.10103 | 0.75387 | 0.0768605 | 0 | 0.56931 | 0.135487 | 279 | 0.448224017921147 |
| Chr13:109.8Mb | 2126 | 13 | 109464153 | 110141685 | 3671 | 416 | AD | eGFR | 0.381694 | -0.14533 | 1 | 0.14569 | 0 | 1 | 0.139747 | 280 | 0.460666003571429 |
| Chr10:103.4Mb | 1737 | 10 | 102618666 | 104223057 | 4260 | 413 | AD | eGFR | -0.34083 | -0.97612 | 0.12344 | 0.116165 | 0 | 0.9528 | 0.140691 | 281 | 0.462127377224199 |
| Chr2:238.8Mb | 479 | 2 | 238449460 | 239219396 | 4227 | 472 | AD | eGFR | -0.505626 | -1 | 0.17695 | 0.255658 | 0 | 1 | 0.141283 | 282 | 0.462426273049645 |
| Chr19:10.4Mb | 2507 | 19 | 10026873 | 10831357 | 3376 | 455 | AD | eGFR | -0.388749 | -1 | 0.14645 | 0.151126 | 0 | 1 | 0.143035 | 283 | 0.466506378091873 |
| Chr2:69.8Mb | 318 | 2 | 69336308 | 70191439 | 3727 | 376 | AD | eGFR | -0.446295 | -1 | 0.18148 | 0.199179 | 0 | 1 | 0.145026 | 284 | 0.4713345 |
| Chr9:36.5Mb | 1559 | 9 | 36138840 | 36919931 | 3424 | 422 | AD | eGFR | 0.236838 | -0.08869 | 0.63618 | 0.0560921 | 0 | 0.40518 | 0.147377 | 285 | 0.477294635087719 |
| Chr9:103.8Mb | 1598 | 9 | 103364050 | 104229070 | 3917 | 364 | AD | eGFR | -0.306524 | -0.89369 | 0.10918 | 0.093957 | 0 | 0.79868 | 0.148518 | 286 | 0.479308090909091 |
| Chr3:52.9Mb | 545 | 3 | 52210786 | 53593195 | 4447 | 354 | AD | eGFR | 0.278498 | -0.09733 | 0.70922 | 0.0775609 | 0 | 0.50299 | 0.149053 | 287 | 0.479358602787456 |
| Chr7:89Mb | 1310 | 7 | 88731393 | 89328433 | 3190 | 296 | AD | eGFR | -0.330902 | -0.9722 | 0.12628 | 0.109496 | 0 | 0.95365 | 0.150498 | 288 | 0.480306125 |
| Chr11:114.3Mb | 1878 | 11 | 113923802 | 114744691 | 3858 | 453 | AD | eGFR | 0.593945 | -0.24433 | 1 | 0.352771 | 0 | 1 | 0.150933 | 289 | 0.480306125 |
| Chr6:83.8Mb | 1148 | 6 | 83297551 | 84217916 | 3632 | 314 | AD | eGFR | 0.244362 | -0.08746 | 0.65711 | 0.0597129 | 0 | 0.43253 | 0.151816 | 290 | 0.480306125 |
| Chr14:29.8Mb | 2142 | 14 | 29022162 | 30543661 | 5762 | 500 | AD | eGFR | 0.342884 | -0.15001 | 1 | 0.117569 | 0 | 1 | 0.151903 | 291 | 0.480306125 |
| Chr2:10.1Mb | 252 | 2 | 9788424 | 10468903 | 4053 | 487 | AD | eGFR | 0.331068 | -0.13083 | 0.99284 | 0.109606 | 0 | 0.98573 | 0.152945 | 292 | 0.480306125 |
| Chr6:90.5Mb | 1155 | 6 | 89749336 | 91211005 | 5715 | 495 | AD | eGFR | 0.27346 | -0.10899 | 0.81217 | 0.0747806 | 0 | 0.65962 | 0.152998 | 293 | 0.480306125 |
| Chr2:177.9Mb | 420 | 2 | 177322587 | 178553262 | 5518 | 453 | AD | eGFR | 0.401894 | -0.16653 | 1 | 0.161518 | 0 | 1 | 0.153039 | 294 | 0.480306125 |
| Chr5:36.2Mb | 937 | 5 | 35823298 | 36645311 | 4031 | 409 | AD | eGFR | -0.388216 | -1 | 0.15373 | 0.150712 | 0 | 1 | 0.153827 | 295 | 0.480306125 |

| locus_label | locus | chr | start | stop | n.snps | n.pcs | phen1 | phen2 | rho | rho.lower | rho.upper | r2 | r2.lower | r2.upper | p | rank | FDR |
|---|---|---|---|---|---|---|---|---|---|---|---|---|---|---|---|---|---|
| Chr21:26Mb | 2633 | 21 | 25504502 | 26468313 | 5009 | 419 | AD | eGFR | -0.520453 | -1 | 0.21696 | 0.270871 | 0 | 1 | 0.154031 | 296 | 0.480306125 |
| Chr16:13.3Mb | 2304 | 16 | 12792107 | 13891462 | 6334 | 490 | AD | eGFR | 0.39029 | -0.1635 | 1 | 0.152326 | 0 | 1 | 0.155301 | 297 | 0.482264402684564 |
| Chr14:62.1Mb | 2172 | 14 | 61651879 | 62502252 | 3851 | 454 | AD | eGFR | 0.308552 | -0.12746 | 0.86709 | 0.0952042 | 0 | 0.7526 | 0.155704 | 298 | 0.482264402684564 |
| Chr8:10.5Mb | 1387 | 8 | 10211810 | 10707264 | 3258 | 419 | AD | eGFR | -0.398544 | -1 | 0.1763 | 0.158837 | 0 | 1 | 0.158923 | 299 | 0.485991275409836 |
| Chr14:55.5Mb | 2166 | 14 | 54799380 | 56173644 | 6007 | 442 | AD | eGFR | 0.379109 | -0.14618 | 1 | 0.143724 | 0 | 1 | 0.159285 | 300 | 0.485991275409836 |
| Chr13:96.6Mb | 2108 | 13 | 96133544 | 97125240 | 3665 | 253 | AD | eGFR | 0.433339 | -0.19363 | 1 | 0.187783 | 0 | 1 | 0.160144 | 301 | 0.485991275409836 |
| Chr3:110.5Mb | 597 | 3 | 110099173 | 110940758 | 3468 | 252 | AD | eGFR | -0.277255 | -0.72331 | 0.12226 | 0.0768703 | 0 | 0.52499 | 0.160207 | 302 | 0.485991275409836 |
| Chr1:91.8Mb | 101 | 1 | 91341822 | 92213244 | 3783 | 415 | AD | eGFR | 0.322571 | -0.12914 | 0.96998 | 0.104052 | 0 | 0.94252 | 0.160413 | 303 | 0.485991275409836 |
| Chr7:47.2Mb | 1273 | 7 | 46607993 | 47720490 | 5850 | 501 | AD | eGFR | 0.318166 | -0.14288 | 0.92177 | 0.10123 | 0 | 0.8509 | 0.16048 | 304 | 0.485991275409836 |
| Chr8:87.5Mb | 1460 | 8 | 86987926 | 88055045 | 5108 | 366 | AD | eGFR | 0.358703 | -0.16867 | 1 | 0.128668 | 0 | 1 | 0.160593 | 305 | 0.485991275409836 |
| Chr1:243.5Mb | 232 | 1 | 242844237 | 244080110 | 4092 | 406 | AD | eGFR | -0.206265 | -0.56736 | 0.08615 | 0.0425451 | 0 | 0.3219 | 0.162123 | 306 | 0.487412260450161 |
| Chr9:124.2Mb | 1620 | 9 | 123640379 | 124679659 | 4343 | 399 | AD | eGFR | -0.38812 | -1 | 0.16766 | 0.150637 | 0 | 1 | 0.162503 | 307 | 0.487412260450161 |
| Chr9:113.2Mb | 1609 | 9 | 112776854 | 113649462 | 4230 | 434 | AD | eGFR | 0.357668 | -0.18359 | 1 | 0.127926 | 0 | 1 | 0.162733 | 308 | 0.487412260450161 |
| Chr2:184.9Mb | 426 | 2 | 184447091 | 185277367 | 4054 | 256 | AD | eGFR | 0.424347 | -0.20042 | 1 | 0.18007 | 0 | 1 | 0.164181 | 309 | 0.487412260450161 |
| Chr12:75.1Mb | 1972 | 12 | 74577926 | 75563059 | 4293 | 344 | AD | eGFR | 0.465423 | -0.2261 | 1 | 0.216619 | 0 | 1 | 0.164229 | 310 | 0.487412260450161 |
| Chr3:180.9Mb | 667 | 3 | 180298525 | 181426564 | 3494 | 341 | AD | eGFR | 0.455622 | -0.19812 | 1 | 0.207592 | 0 | 1 | 0.164231 | 311 | 0.487412260450161 |
| Chr13:27.3Mb | 2037 | 13 | 26952970 | 27562471 | 3588 | 448 | AD | eGFR | -0.232969 | -0.60354 | 0.09695 | 0.0542744 | 0 | 0.36479 | 0.164988 | 312 | 0.4880895 |
| Chr6:68.9Mb | 1131 | 6 | 68594061 | 69298685 | 3653 | 312 | AD | eGFR | -0.227238 | -0.59509 | 0.09917 | 0.051637 | 0 | 0.35413 | 0.16717 | 313 | 0.492258828025478 |
| Chr2:119.6Mb | 361 | 2 | 119261516 | 120029001 | 3791 | 417 | AD | eGFR | 0.305683 | -0.13479 | 0.88865 | 0.0934421 | 0 | 0.79117 | 0.167464 | 314 | 0.492258828025478 |
| Chr3:162.1Mb | 648 | 3 | 161659377 | 162449018 | 4445 | 279 | AD | eGFR | 0.247406 | -0.11282 | 0.64395 | 0.0612098 | 0 | 0.41467 | 0.168872 | 315 | 0.493715584375 |
| Chr4:126Mb | 825 | 4 | 125495904 | 126425045 | 4455 | 375 | AD | eGFR | -0.58222 | -1 | 0.28708 | 0.33898 | 0 | 1 | 0.169416 | 316 | 0.493715584375 |
| Chr1:63.4Mb | 70 | 1 | 62901691 | 63799928 | 3804 | 410 | AD | eGFR | 0.373693 | -0.17305 | 1 | 0.139646 | 0 | 1 | 0.170431 | 317 | 0.493715584375 |
| Chr9:117.2Mb | 1613 | 9 | 116739984 | 117697061 | 4531 | 484 | AD | eGFR | -0.298514 | -0.85782 | 0.13442 | 0.0891108 | 0 | 0.73687 | 0.17054 | 318 | 0.493715584375 |
| Chr4:140.6Mb | 840 | 4 | 140176427 | 141086487 | 3717 | 485 | AD | eGFR | 0.419019 | -0.20278 | 1 | 0.175577 | 0 | 1 | 0.170992 | 319 | 0.493715584375 |
| Chr5:58.1Mb | 954 | 5 | 57441633 | 58835549 | 6302 | 448 | AD | eGFR | 0.302287 | -0.1459 | 0.8694 | 0.0913772 | 0 | 0.75586 | 0.171169 | 320 | 0.493715584375 |
| Chr10:85.9Mb | 1722 | 10 | 85423416 | 86459024 | 4826 | 423 | AD | eGFR | 0.229253 | -0.09544 | 0.61598 | 0.0525569 | 0 | 0.37943 | 0.173117 | 321 | 0.497098562111801 |
| Chr9:5.1Mb | 1519 | 9 | 4762810 | 5524508 | 4173 | 431 | AD | eGFR | -0.302241 | -0.87259 | 0.13705 | 0.0913494 | 0 | 0.76141 | 0.173419 | 322 | 0.497098562111801 |
| Chr2:153.4Mb | 397 | 2 | 152735766 | 153999228 | 5293 | 402 | AD | eGFR | -0.281317 | -0.99747 | 0.14402 | 0.0791393 | 0 | 1 | 0.173979 | 323 | 0.49715980495356 |
| Chr10:36.4Mb | 1678 | 10 | 35901902 | 36887609 | 4607 | 442 | AD | eGFR | -0.154802 | -0.39967 | 0.06958 | 0.0239635 | 0 | 0.15996 | 0.1752 | 324 | 0.499103703703704 |
| Chr2:133Mb | 376 | 2 | 132690445 | 133386614 | 2309 | 396 | AD | eGFR | -0.263559 | -0.71172 | 0.1244 | 0.0694634 | 0 | 0.50654 | 0.177397 | 325 | 0.502173533333333 |
| Chr4:121.9Mb | 821 | 4 | 121388190 | 122460107 | 5811 | 405 | AD | eGFR | -0.146871 | -0.37484 | 0.06597 | 0.0215711 | 0 | 0.1405 | 0.177942 | 326 | 0.502173533333333 |
| Chr9:101.5Mb | 1596 | 9 | 101078128 | 101860663 | 3724 | 446 | AD | eGFR | -0.223346 | -0.60199 | 0.11089 | 0.0498835 | 0 | 0.36239 | 0.178787 | 327 | 0.502173533333333 |

| locus_label | locus | chr | start | stop | n.snps | n.pcs | phen1 | phen2 | rho | rho.lower | rho.upper | r2 | r2.lower | r2.upper | p | rank | FDR |
|---|---|---|---|---|---|---|---|---|---|---|---|---|---|---|---|---|---|
| Chr12:101.5Mb | 1995 | 12 | 100936112 | 101976787 | 4523 | 453 | AD | eGFR | -0.337962 | -1 | 0.16273 | 0.114218 | 0 | 1 | 0.179358 | 328 | 0.502173533333333 |
| Chr6:75.4Mb | 1139 | 6 | 75052177 | 75834629 | 3418 | 300 | AD | eGFR | -0.491258 | -1 | 0.27043 | 0.241334 | 0 | 1 | 0.179487 | 329 | 0.502173533333333 |
| Chr1:112.1Mb | 120 | 1 | 111732156 | 112496017 | 3814 | 429 | AD | eGFR | -0.262768 | -0.73162 | 0.13332 | 0.0690469 | 0 | 0.53527 | 0.179542 | 330 | 0.502173533333333 |
| Chr6:56.7Mb | 1122 | 6 | 56106488 | 57316570 | 4293 | 342 | AD | eGFR | 0.250612 | -0.11957 | 0.68885 | 0.0628063 | 0 | 0.47452 | 0.180851 | 331 | 0.504306564954683 |
| Chr4:175.1Mb | 876 | 4 | 174689458 | 175593531 | 4167 | 475 | AD | eGFR | -0.469365 | -1 | 0.23954 | 0.220303 | 0 | 1 | 0.182173 | 332 | 0.506462888554217 |
| Chr2:171.4Mb | 414 | 2 | 170973618 | 171748742 | 3980 | 494 | AD | eGFR | 0.317184 | -0.15734 | 0.9653 | 0.100606 | 0 | 0.93427 | 0.182898 | 333 | 0.506951513513514 |
| Chr8:25.1Mb | 1406 | 8 | 24670169 | 25571729 | 4045 | 431 | AD | eGFR | 0.351307 | -0.18935 | 1 | 0.123417 | 0 | 1 | 0.184016 | 334 | 0.50852325748503 |
| Chr2:88.5Mb | 338 | 2 | 87808602 | 89145572 | 3014 | 346 | AD | eGFR | -0.164844 | -0.41533 | 0.0772 | 0.0271736 | 0 | 0.1725 | 0.18524 | 335 | 0.510377671641791 |
| Chr10:90.2Mb | 1726 | 10 | 89590337 | 90905621 | 4952 | 460 | AD | eGFR | 0.455396 | -0.29813 | 1 | 0.207385 | 0 | 1 | 0.189466 | 336 | 0.520467613095238 |
| Chr6:169.2Mb | 1224 | 6 | 168718582 | 169717655 | 6208 | 484 | AD | eGFR | 0.285948 | -0.15136 | 0.86121 | 0.0817664 | 0 | 0.74228 | 0.190285 | 337 | 0.521166335311573 |
| Chr2:152.3Mb | 396 | 2 | 151773672 | 152735765 | 4024 | 341 | AD | eGFR | -0.492849 | -1 | 0.29371 | 0.2429 | 0 | 1 | 0.191591 | 338 | 0.523190807692308 |
| Chr10:60.7Mb | 1698 | 10 | 60136913 | 61309677 | 4506 | 433 | AD | eGFR | 0.331999 | -0.18131 | 1 | 0.110223 | 0 | 1 | 0.192336 | 339 | 0.52367589380531 |
| Chr19:6.9Mb | 2503 | 19 | 6554210 | 7310557 | 4249 | 533 | AD | eGFR | 0.274769 | -0.15669 | 0.87681 | 0.0754979 | 0 | 0.77144 | 0.194714 | 340 | 0.528591241176471 |
| Chr6:11.5Mb | 1070 | 6 | 10898991 | 12178501 | 6149 | 504 | AD | eGFR | 0.276109 | -0.15776 | 0.8063 | 0.0762363 | 0 | 0.65012 | 0.196566 | 341 | 0.531558938596491 |
| Chr6:51.1Mb | 1116 | 6 | 50425358 | 51705697 | 5338 | 340 | AD | eGFR | 0.345141 | -0.19452 | 1 | 0.119122 | 0 | 1 | 0.196959 | 342 | 0.531558938596491 |
| Chr4:143.2Mb | 843 | 4 | 142807867 | 143504537 | 3350 | 333 | AD | eGFR | 0.37586 | -0.21916 | 1 | 0.141271 | 0 | 1 | 0.19784 | 343 | 0.532379941690962 |
| Chr13:95.1Mb | 2106 | 13 | 94722546 | 95470650 | 3551 | 447 | AD | eGFR | 0.262654 | -0.14807 | 0.74198 | 0.0689871 | 0 | 0.552 | 0.198823 | 344 | 0.533469851744186 |
| Chr7:64.5Mb | 1290 | 7 | 64102826 | 64833290 | 2928 | 199 | AD | eGFR | -0.49344 | -1 | 0.32893 | 0.243483 | 0 | 1 | 0.199874 | 345 | 0.534735368115942 |
| Chr1:206.5Mb | 191 | 1 | 205811878 | 207251465 | 3105 | 444 | AD | eGFR | -0.323505 | -1 | 0.19501 | 0.104656 | 0 | 1 | 0.204995 | 346 | 0.546850823699422 |
| Chr5:140.8Mb | 1025 | 5 | 140352430 | 141337297 | 3683 | 400 | AD | eGFR | -0.458855 | -1 | 0.30646 | 0.210548 | 0 | 1 | 0.206755 | 347 | 0.549956383285303 |
| Chr12:103.6Mb | 1997 | 12 | 103150650 | 104141496 | 4225 | 444 | AD | eGFR | -0.360811 | -1 | 0.25314 | 0.130185 | 0 | 1 | 0.210461 | 348 | 0.558205468390805 |
| Chr8:91.6Mb | 1464 | 8 | 90886634 | 92407388 | 5958 | 366 | AD | eGFR | -0.51933 | -1 | 0.33772 | 0.269704 | 0 | 1 | 0.212234 | 349 | 0.561295077363897 |
| Chr1:58.3Mb | 64 | 1 | 57954900 | 58712789 | 3345 | 402 | AD | eGFR | -0.260621 | -0.76664 | 0.15851 | 0.0679233 | 0 | 0.58899 | 0.214747 | 350 | 0.566318517142857 |
| Chr15:99.2Mb | 2283 | 15 | 98724596 | 99636933 | 5188 | 537 | AD | eGFR | 0.255995 | -0.16839 | 0.75046 | 0.0655332 | 0 | 0.56336 | 0.215965 | 351 | 0.56784690625 |
| Chr10:9Mb | 1649 | 10 | 8543258 | 9434190 | 4408 | 435 | AD | eGFR | -0.146534 | -0.39318 | 0.07966 | 0.0214723 | 0 | 0.1551 | 0.216557 | 352 | 0.56784690625 |
| Chr1:61.8Mb | 68 | 1 | 61258170 | 62243662 | 4081 | 490 | AD | eGFR | -0.380931 | -1 | 0.26748 | 0.145108 | 0 | 1 | 0.219543 | 353 | 0.574045861189802 |
| Chr3:161.1Mb | 647 | 3 | 160588294 | 161659376 | 4993 | 312 | AD | eGFR | 0.280772 | -0.18498 | 0.84393 | 0.0788329 | 0 | 0.71797 | 0.221291 | 354 | 0.576981901129943 |
| Chr2:103.2Mb | 346 | 2 | 102689084 | 103781210 | 4649 | 418 | AD | eGFR | -0.160225 | -0.45648 | 0.09547 | 0.025672 | 0 | 0.20846 | 0.223306 | 355 | 0.5805956 |
| Chr4:95.2Mb | 794 | 4 | 94755265 | 95638487 | 4177 | 342 | AD | eGFR | -0.267814 | -0.81975 | 0.18931 | 0.0717241 | 0 | 0.6737 | 0.224672 | 356 | 0.582472464985994 |
| Chr11:102.9Mb | 1868 | 11 | 102383680 | 103449312 | 5639 | 384 | AD | eGFR | -0.419031 | -1 | 0.29906 | 0.175587 | 0 | 1 | 0.22529 | 357 | 0.582472464985994 |
| Chr11:79.9Mb | 1845 | 11 | 79480999 | 80385740 | 4425 | 477 | AD | eGFR | 0.319575 | -0.22429 | 1 | 0.102128 | 0 | 1 | 0.227878 | 358 | 0.586256690807799 |
| Chr16:67.4Mb | 2335 | 16 | 66396822 | 68427119 | 5464 | 314 | AD | eGFR | -0.334051 | -1 | 0.24726 | 0.11159 | 0 | 1 | 0.228024 | 359 | 0.586256690807799 |
| Chr10:64.2Mb | 1701 | 10 | 63553783 | 64758500 | 5146 | 478 | AD | eGFR | -0.267333 | -0.83777 | 0.18157 | 0.0714672 | 0 | 0.70271 | 0.22961 | 360 | 0.58833451800554 |

| locus_label | locus | chr | start | stop | n.snps | n.pcs | phen1 | phen2 | rho | rho.lower | rho.upper | r2 | r2.lower | r2.upper | p | rank | FDR |
|---|---|---|---|---|---|---|---|---|---|---|---|---|---|---|---|---|---|
| Chr3:182.3Mb | 668 | 3 | 181426565 | 183111760 | 6470 | 514 | AD | eGFR | 0.436664 | -0.32086 | 1 | 0.190675 | 0 | 1 | 0.230107 | 361 | 0.58833451800554 |
| Chr8:60.9Mb | 1436 | 8 | 60478318 | 61288512 | 3816 | 305 | AD | eGFR | 0.276291 | -0.20673 | 1 | 0.0763367 | 0 | 1 | 0.231733 | 362 | 0.589362198347107 |
| Chr10:74.9Mb | 1712 | 10 | 74069133 | 75693329 | 4744 | 279 | AD | eGFR | -0.312913 | -1 | 0.21619 | 0.0979147 | 0 | 1 | 0.231786 | 363 | 0.589362198347107 |
| Chr9:19.8Mb | 1540 | 9 | 19452714 | 20187320 | 3322 | 440 | AD | eGFR | 0.368724 | -0.25711 | 1 | 0.135957 | 0 | 1 | 0.233864 | 364 | 0.592037487671233 |
| Chr13:100.8Mb | 2113 | 13 | 100400375 | 101264876 | 3336 | 422 | AD | eGFR | 0.186008 | -0.13029 | 0.52661 | 0.0345989 | 0 | 0.27731 | 0.234121 | 365 | 0.592037487671233 |
| Chr13:40.1Mb | 2051 | 13 | 39403027 | 40706371 | 6681 | 480 | AD | eGFR | -0.4361 | -1 | 0.33581 | 0.190183 | 0 | 1 | 0.235875 | 366 | 0.592947344736842 |
| Chr20:23.4Mb | 2584 | 20 | 23024048 | 23748117 | 4036 | 365 | AD | eGFR | 0.365559 | -0.27757 | 1 | 0.133633 | 0 | 1 | 0.237431 | 367 | 0.592947344736842 |
| Chr5:27.7Mb | 928 | 5 | 27076165 | 28315408 | 5808 | 428 | AD | eGFR | -0.297849 | -1 | 0.21882 | 0.088714 | 0 | 1 | 0.237989 | 368 | 0.592947344736842 |
| Chr16:14.9Mb | 2305 | 16 | 13891463 | 15919297 | 4805 | 463 | AD | eGFR | -0.378011 | -1 | 0.26488 | 0.142893 | 0 | 1 | 0.238137 | 369 | 0.592947344736842 |
| Chr1:231.6Mb | 217 | 1 | 231205932 | 232074277 | 3665 | 421 | AD | eGFR | -0.386382 | -1 | 0.30583 | 0.149291 | 0 | 1 | 0.239378 | 370 | 0.592947344736842 |
| Chr14:81.7Mb | 2190 | 14 | 80998273 | 82456810 | 6727 | 434 | AD | eGFR | 0.293818 | -0.23127 | 0.97646 | 0.086329 | 0 | 0.95659 | 0.239674 | 371 | 0.592947344736842 |
| Chr7:44.7Mb | 1271 | 7 | 44221299 | 45231981 | 4040 | 379 | AD | eGFR | 0.398504 | -0.33657 | 1 | 0.158805 | 0 | 1 | 0.239942 | 372 | 0.592947344736842 |
| Chr3:107.4Mb | 594 | 3 | 106985790 | 107876312 | 3423 | 400 | AD | eGFR | 0.339443 | -0.26168 | 1 | 0.115222 | 0 | 1 | 0.240278 | 373 | 0.592947344736842 |
| Chr9:81.8Mb | 1576 | 9 | 81241435 | 82345830 | 4861 | 468 | AD | eGFR | 0.214411 | -0.1459 | 0.65076 | 0.0459721 | 0 | 0.42714 | 0.241853 | 374 | 0.592947344736842 |
| Chr21:30.3Mb | 2637 | 21 | 29723404 | 30845782 | 5206 | 344 | AD | eGFR | -0.269154 | -0.84968 | 0.19185 | 0.0724439 | 0 | 0.72348 | 0.242936 | 375 | 0.592947344736842 |
| Chr3:109.4Mb | 596 | 3 | 108639230 | 110099172 | 6467 | 388 | AD | eGFR | 0.311472 | -0.24701 | 1 | 0.0970148 | 0 | 1 | 0.243188 | 376 | 0.592947344736842 |
| Chr6:149.7Mb | 1204 | 6 | 149107630 | 150253141 | 5657 | 435 | AD | eGFR | 0.204784 | -0.15405 | 0.6122 | 0.0419364 | 0 | 0.37479 | 0.243506 | 377 | 0.592947344736842 |
| Chr3:125.9Mb | 612 | 3 | 125539207 | 126216593 | 4027 | 374 | AD | eGFR | 0.239478 | -0.16664 | 0.76388 | 0.0573496 | 0 | 0.59041 | 0.243535 | 378 | 0.592947344736842 |
| Chr1:107.9Mb | 116 | 1 | 107488167 | 108407186 | 4053 | 445 | AD | eGFR | -0.326087 | -1 | 0.24357 | 0.106333 | 0 | 1 | 0.243671 | 379 | 0.592947344736842 |
| Chr12:125.1Mb | 2016 | 12 | 124708655 | 125465022 | 4252 | 529 | AD | eGFR | -0.298143 | -1 | 0.22133 | 0.0888891 | 0 | 1 | 0.244117 | 380 | 0.592947344736842 |
| Chr12:68.3Mb | 1966 | 12 | 67817776 | 68739921 | 4194 | 475 | AD | eGFR | -0.238429 | -0.75562 | 0.1744 | 0.0568484 | 0 | 0.57511 | 0.246022 | 381 | 0.595180361256544 |
| Chr8:71.4Mb | 1445 | 8 | 70798885 | 72094739 | 5143 | 387 | AD | eGFR | -0.347629 | -1 | 0.28415 | 0.120846 | 0 | 1 | 0.246326 | 382 | 0.595180361256544 |
| Chr12:77.2Mb | 1974 | 12 | 76526221 | 77797978 | 5557 | 426 | AD | eGFR | 0.243014 | -0.1863 | 0.9156 | 0.0590559 | 0 | 0.85233 | 0.250331 | 383 | 0.602853254545455 |
| Chr8:69.1Mb | 1443 | 8 | 68568192 | 69725316 | 5040 | 484 | AD | eGFR | 0.373469 | -0.30316 | 1 | 0.139479 | 0 | 1 | 0.251267 | 384 | 0.602853254545455 |
| Chr5:151.1Mb | 1034 | 5 | 150558437 | 151659329 | 5461 | 441 | AD | eGFR | 0.281699 | -0.22032 | 1 | 0.0793541 | 0 | 1 | 0.251461 | 385 | 0.602853254545455 |
| Chr3:100Mb | 587 | 3 | 99373270 | 100557770 | 4366 | 298 | AD | eGFR | -0.430711 | -1 | 0.38135 | 0.185512 | 0 | 1 | 0.254575 | 386 | 0.608737629533679 |
| Chr15:42Mb | 2233 | 15 | 41563960 | 42442110 | 3423 | 378 | AD | eGFR | -0.218846 | -0.68565 | 0.15916 | 0.0478937 | 0 | 0.47099 | 0.255263 | 387 | 0.608805552971576 |
| Chr10:56.6Mb | 1694 | 10 | 56203309 | 57038323 | 4323 | 428 | AD | eGFR | 0.326627 | -0.27932 | 1 | 0.106685 | 0 | 1 | 0.256614 | 388 | 0.609901020356234 |
| Chr1:107.1Mb | 115 | 1 | 106769893 | 107488166 | 3425 | 287 | AD | eGFR | -0.271213 | -0.9072 | 0.20878 | 0.0735564 | 0 | 0.82511 | 0.258918 | 389 | 0.609901020356234 |
| Chr6:25.9Mb | 1085 | 6 | 25485854 | 26346790 | 3714 | 298 | AD | eGFR | 0.240742 | -0.19432 | 0.75363 | 0.0579567 | 0 | 0.56989 | 0.258971 | 390 | 0.609901020356234 |
| Chr7:13.9Mb | 1242 | 7 | 13577007 | 14138484 | 3155 | 441 | AD | eGFR | -0.324802 | -1 | 0.25531 | 0.105496 | 0 | 1 | 0.25946 | 391 | 0.609901020356234 |
| Chr11:65.3Mb | 1832 | 11 | 64576162 | 66057666 | 5217 | 410 | AD | eGFR | 0.228687 | -0.18766 | 0.71772 | 0.0522976 | 0 | 0.51579 | 0.259645 | 392 | 0.609901020356234 |
| Chr1:163Mb | 146 | 1 | 162445321 | 163504124 | 5051 | 445 | AD | eGFR | -0.210024 | -0.65944 | 0.16785 | 0.04411 | 0 | 0.43487 | 0.259687 | 393 | 0.609901020356234 |

| locus_label | locus | chr | start | stop | n.snps | n.pcs | phen1 | phen2 | rho | rho.lower | rho.upper | r2 | r2.lower | r2.upper | p | rank | FDR |
|---|---|---|---|---|---|---|---|---|---|---|---|---|---|---|---|---|---|
| Chr12:130.2Mb | 2023 | 12 | 129782736 | 130581203 | 4776 | 527 | AD | eGFR | 0.206195 | -0.16894 | 0.70116 | 0.0425163 | 0 | 0.49827 | 0.26065 | 394 | 0.610609010152284 |
| Chr17:58.3Mb | 2408 | 17 | 57387919 | 59288226 | 5259 | 365 | AD | eGFR | -0.301332 | -1 | 0.2588 | 0.0908011 | 0 | 1 | 0.262156 | 395 | 0.612506063131313 |
| Chr20:17.1Mb | 2577 | 20 | 16660089 | 17484756 | 4238 | 424 | AD | eGFR | -0.191135 | -0.56929 | 0.15145 | 0.0365324 | 0 | 0.3244 | 0.262787 | 396 | 0.612506063131313 |
| Chr3:122.7Mb | 609 | 3 | 121971907 | 123412340 | 6287 | 471 | AD | eGFR | -0.307313 | -1 | 0.26442 | 0.0944414 | 0 | 1 | 0.264068 | 397 | 0.613414668341709 |
| Chr5:84.7Mb | 977 | 5 | 84089617 | 85315276 | 6413 | 346 | AD | eGFR | -0.438712 | -1 | 0.40028 | 0.192468 | 0 | 1 | 0.264506 | 398 | 0.613414668341709 |
| Chr10:80.2Mb | 1716 | 10 | 79713338 | 80687808 | 4738 | 484 | AD | eGFR | 0.274104 | -0.24033 | 0.95236 | 0.0751329 | 0 | 0.90925 | 0.267342 | 399 | 0.6179138875 |
| Chr5:26Mb | 926 | 5 | 25720602 | 26358164 | 3846 | 350 | AD | eGFR | 0.345531 | -0.31602 | 1 | 0.119392 | 0 | 1 | 0.267785 | 400 | 0.6179138875 |
| Chr13:65.4Mb | 2074 | 13 | 64617610 | 66213201 | 6362 | 396 | AD | eGFR | -0.31956 | -1 | 0.29987 | 0.102119 | 0 | 1 | 0.269025 | 401 | 0.619227119700748 |
| Chr4:86.6Mb | 785 | 4 | 86182801 | 86928785 | 3815 | 360 | AD | eGFR | 0.355638 | -0.3418 | 1 | 0.126478 | 0 | 1 | 0.270045 | 402 | 0.619953677419355 |
| Chr12:99.2Mb | 1993 | 12 | 98578209 | 99815375 | 4926 | 441 | AD | eGFR | 0.213803 | -0.17361 | 0.65741 | 0.0457119 | 0 | 0.4335 | 0.270684 | 403 | 0.619953677419355 |
| Chr14:27.9Mb | 2140 | 14 | 27530683 | 28282153 | 3334 | 335 | AD | eGFR | 0.268903 | -0.23972 | 0.90885 | 0.072309 | 0 | 0.82769 | 0.272508 | 404 | 0.621520203791469 |
| Chr15:70.4Mb | 2259 | 15 | 69965289 | 70765763 | 3910 | 509 | AD | eGFR | -0.337358 | -1 | 0.32304 | 0.11381 | 0 | 1 | 0.27353 | 405 | 0.621520203791469 |
| Chr1:43.7Mb | 50 | 1 | 43299581 | 44129647 | 3517 | 406 | AD | eGFR | 0.209855 | -0.18681 | 0.65312 | 0.0440391 | 0 | 0.42657 | 0.273652 | 406 | 0.621520203791469 |
| Chr8:72.6Mb | 1446 | 8 | 72094740 | 73169868 | 5034 | 481 | AD | eGFR | 0.209016 | -0.17827 | 0.65495 | 0.0436877 | 0 | 0.43025 | 0.274983 | 407 | 0.621520203791469 |
| Chr2:110.9Mb | 353 | 2 | 109959776 | 111849815 | 3886 | 419 | AD | eGFR | -0.289291 | -1 | 0.27032 | 0.0836893 | 0 | 1 | 0.275754 | 408 | 0.621520203791469 |
| Chr22:34.3Mb | 2670 | 22 | 33752428 | 34863126 | 5993 | 468 | AD | eGFR | -0.34949 | -1 | 0.33386 | 0.122143 | 0 | 1 | 0.277916 | 409 | 0.621520203791469 |
| Chr20:47.7Mb | 2604 | 20 | 47197529 | 48212707 | 4114 | 383 | AD | eGFR | 0.273319 | -0.25573 | 0.94675 | 0.0747031 | 0 | 0.89829 | 0.278634 | 410 | 0.621520203791469 |
| Chr12:31.6Mb | 1931 | 12 | 31140248 | 32117204 | 4842 | 413 | AD | eGFR | 0.241149 | -0.20678 | 0.79685 | 0.0581531 | 0 | 0.64044 | 0.280699 | 411 | 0.621520203791469 |
| Chr5:19.2Mb | 919 | 5 | 18790682 | 19543381 | 3786 | 331 | AD | eGFR | -0.30875 | -1 | 0.27659 | 0.0953263 | 0 | 1 | 0.280787 | 412 | 0.621520203791469 |
| Chr1:223.2Mb | 209 | 1 | 222730494 | 223634500 | 3684 | 372 | AD | eGFR | 0.191388 | -0.16029 | 0.57218 | 0.0366292 | 0 | 0.3294 | 0.280925 | 413 | 0.621520203791469 |
| Chr8:38Mb | 1418 | 8 | 37166048 | 38815774 | 5595 | 490 | AD | eGFR | -0.332088 | -1 | 0.30268 | 0.110282 | 0 | 1 | 0.28142 | 414 | 0.621520203791469 |
| Chr5:161.7Mb | 1044 | 5 | 161361943 | 162106453 | 3269 | 372 | AD | eGFR | -0.359374 | -1 | 0.35112 | 0.129149 | 0 | 1 | 0.281431 | 415 | 0.621520203791469 |
| Chr2:67.2Mb | 315 | 2 | 66720836 | 67696941 | 4213 | 448 | AD | eGFR | 0.187547 | -0.16202 | 0.68271 | 0.0351737 | 0 | 0.49206 | 0.281491 | 416 | 0.621520203791469 |
| Chr1:228.5Mb | 214 | 1 | 227983745 | 229114216 | 5037 | 391 | AD | eGFR | -0.314509 | -1 | 0.30418 | 0.0989157 | 0 | 1 | 0.282103 | 417 | 0.621520203791469 |
| Chr13:43.4Mb | 2054 | 13 | 43045031 | 43773841 | 3514 | 415 | AD | eGFR | 0.180092 | -0.15276 | 0.53329 | 0.0324331 | 0 | 0.28453 | 0.28288 | 418 | 0.621520203791469 |
| Chr2:219.2Mb | 457 | 2 | 218756740 | 219684222 | 3785 | 385 | AD | eGFR | -0.386857 | -1 | 0.39243 | 0.149658 | 0 | 1 | 0.283143 | 419 | 0.621520203791469 |
| Chr2:25.2Mb | 269 | 2 | 24670283 | 25816250 | 3645 | 383 | AD | eGFR | -0.274499 | -1 | 0.26434 | 0.0753499 | 0 | 1 | 0.283814 | 420 | 0.621520203791469 |
| Chr3:195.5Mb | 683 | 3 | 195198579 | 195779322 | 1744 | 354 | AD | eGFR | 0.253488 | -0.22397 | 0.90601 | 0.064256 | 0 | 0.82992 | 0.283964 | 421 | 0.621520203791469 |
| Chr7:38.6Mb | 1265 | 7 | 38270124 | 38964435 | 3637 | 367 | AD | eGFR | -0.33285 | -1 | 0.34523 | 0.110789 | 0 | 1 | 0.284162 | 422 | 0.621520203791469 |
| Chr1:55.9Mb | 61 | 1 | 55523105 | 56284175 | 3749 | 374 | AD | eGFR | 0.352054 | -0.3503 | 1 | 0.123942 | 0 | 1 | 0.285797 | 423 | 0.623618513002364 |
| Chr3:32.2Mb | 527 | 3 | 31854998 | 32507705 | 3396 | 449 | AD | eGFR | -0.263718 | -0.92388 | 0.2415 | 0.0695469 | 0 | 0.8542 | 0.289933 | 424 | 0.629720550588235 |
| Chr3:40Mb | 535 | 3 | 39607202 | 40356575 | 3151 | 306 | AD | eGFR | -0.293357 | -1 | 0.28977 | 0.0860585 | 0 | 1 | 0.289958 | 425 | 0.629720550588235 |
| Chr22:36.4Mb | 2672 | 22 | 35855903 | 37007880 | 5276 | 488 | AD | eGFR | -0.167429 | -0.51341 | 0.14986 | 0.0280323 | 0 | 0.26383 | 0.293513 | 426 | 0.633430315420561 |
| Chr19:7.8Mb | 2504 | 19 | 7310558 | 8201240 | 4399 | 499 | AD | eGFR | -0.234434 | -0.86346 | 0.24333 | 0.0549592 | 0 | 0.75152 | 0.293711 | 427 | 0.633430315420561 |
| Chr6:159.5Mb | 1213 | 6 | 159082296 | 159985061 | 5082 | 442 | AD | eGFR | 0.219575 | -0.20834 | 0.72835 | 0.0482133 | 0 | 0.53251 | 0.293725 | 428 | 0.633430315420561 |
| Chr16:53.2Mb | 2323 | 16 | 52507565 | 53852141 | 5736 | 477 | AD | eGFR | -0.356098 | -1 | 0.38336 | 0.126806 | 0 | 1 | 0.296286 | 429 | 0.637463818181818 |
| Chr15:81.8Mb | 2268 | 15 | 81106582 | 82405384 | 5392 | 504 | AD | eGFR | -0.214297 | -0.77583 | 0.21016 | 0.0459233 | 0 | 0.61444 | 0.297893 | 430 | 0.638204174013921 |

| locus_label | locus | chr | start | stop | n.snps | n.pcs | phen1 | phen2 | rho | rho.lower | rho.upper | r2 | r2.lower | r2.upper | p | rank | FDR |
|---|---|---|---|---|---|---|---|---|---|---|---|---|---|---|---|---|---|
| Chr6:129Mb | 1185 | 6 | 128335095 | 129604950 | 5416 | 404 | AD | eGFR | 0.205731 | -0.20623 | 0.70733 | 0.0423252 | 0 | 0.50346 | 0.298013 | 431 | 0.638204174013921 |
| Chr6:67.5Mb | 1129 | 6 | 67028075 | 67906206 | 5760 | 362 | AD | eGFR | 0.229992 | -0.23385 | 0.8649 | 0.0528964 | 0 | 0.75134 | 0.299819 | 432 | 0.640585502314815 |
| Chr14:54.1Mb | 2165 | 14 | 53472709 | 54799379 | 5549 | 446 | AD | eGFR | -0.325013 | -1 | 0.34248 | 0.105633 | 0 | 1 | 0.302596 | 433 | 0.644381310344828 |
| Chr3:153Mb | 638 | 3 | 152644859 | 153406787 | 3521 | 368 | AD | eGFR | 0.381123 | -0.40927 | 1 | 0.145255 | 0 | 1 | 0.303457 | 434 | 0.644381310344828 |
| Chr6:80.3Mb | 1144 | 6 | 79859803 | 80695438 | 4045 | 386 | AD | eGFR | -0.27325 | -1 | 0.27611 | 0.0746656 | 0 | 1 | 0.30369 | 435 | 0.644381310344828 |
| Chr19:49.8Mb | 2545 | 19 | 49361068 | 50231619 | 3981 | 455 | AD | eGFR | -0.279212 | -1 | 0.30737 | 0.0779593 | 0 | 1 | 0.305777 | 436 | 0.647321493119266 |
| Chr10:105.2Mb | 1738 | 10 | 104223058 | 106134193 | 6362 | 418 | AD | eGFR | -0.228186 | -0.81934 | 0.23423 | 0.0520688 | 0 | 0.67648 | 0.307031 | 437 | 0.648488816933638 |
| Chr5:157.6Mb | 1040 | 5 | 157203560 | 158053484 | 4236 | 350 | AD | eGFR | -0.242635 | -0.84652 | 0.25023 | 0.0588716 | 0 | 0.71951 | 0.308932 | 438 | 0.651014237442922 |
| Chr10:89Mb | 1725 | 10 | 88447920 | 89590336 | 3719 | 378 | AD | eGFR | -0.175018 | -0.55148 | 0.16204 | 0.0306313 | 0 | 0.3052 | 0.31168 | 439 | 0.652127501123596 |
| Chr12:66Mb | 1964 | 12 | 65391080 | 66632168 | 4088 | 431 | AD | eGFR | -0.219223 | -0.76866 | 0.22741 | 0.0480586 | 0 | 0.59441 | 0.312005 | 440 | 0.652127501123596 |
| Chr1:70.5Mb | 78 | 1 | 69940038 | 70994447 | 4076 | 370 | AD | eGFR | 0.266025 | -0.28563 | 1 | 0.0707691 | 0 | 1 | 0.312297 | 441 | 0.652127501123596 |
| Chr3:32.9Mb | 528 | 3 | 32507706 | 33195528 | 3265 | 380 | AD | eGFR | -0.198595 | -0.6441 | 0.199 | 0.0394398 | 0 | 0.41659 | 0.312752 | 442 | 0.652127501123596 |
| Chr9:37.3Mb | 1560 | 9 | 36919932 | 37732939 | 3467 | 384 | AD | eGFR | 0.226048 | -0.23282 | 0.79892 | 0.0510979 | 0 | 0.64097 | 0.313181 | 443 | 0.652127501123596 |
| Chr13:109.1Mb | 2125 | 13 | 108805638 | 109464152 | 3588 | 419 | AD | eGFR | 0.237435 | -0.25162 | 0.8249 | 0.0563753 | 0 | 0.68823 | 0.313794 | 444 | 0.652127501123596 |
| Chr8:24.1Mb | 1405 | 8 | 23547241 | 24670168 | 5662 | 418 | AD | eGFR | 0.218942 | -0.24753 | 0.88704 | 0.0479356 | 0 | 0.79918 | 0.314406 | 445 | 0.652127501123596 |
| Chr12:93.9Mb | 1988 | 12 | 93451081 | 94319629 | 3905 | 481 | AD | eGFR | -0.135688 | -0.47039 | 0.13422 | 0.0184113 | 0 | 0.22306 | 0.31652 | 446 | 0.655040269058296 |
| Chr9:17.8Mb | 1537 | 9 | 17454989 | 18123170 | 3888 | 428 | AD | eGFR | 0.274337 | -0.31 | 1 | 0.075261 | 0 | 1 | 0.322912 | 447 | 0.659577784946237 |
| Chr9:112.2Mb | 1608 | 9 | 111624009 | 112776853 | 5228 | 439 | AD | eGFR | -0.27969 | -1 | 0.32547 | 0.0782264 | 0 | 1 | 0.323419 | 448 | 0.659577784946237 |
| Chr10:81.2Mb | 1717 | 10 | 80687809 | 81629519 | 3840 | 479 | AD | eGFR | -0.273235 | -1 | 0.30489 | 0.0746573 | 0 | 1 | 0.323815 | 449 | 0.659577784946237 |
| Chr19:8.7Mb | 2505 | 19 | 8201241 | 9124135 | 4056 | 495 | AD | eGFR | -0.314606 | -1 | 0.35584 | 0.0989767 | 0 | 1 | 0.324865 | 450 | 0.659577784946237 |
| Chr11:42.9Mb | 1814 | 11 | 42311483 | 43567706 | 6351 | 383 | AD | eGFR | 0.315517 | -0.38605 | 1 | 0.0995512 | 0 | 1 | 0.326028 | 451 | 0.659577784946237 |
| Chr13:46.7Mb | 2057 | 13 | 46349854 | 47059423 | 3309 | 423 | AD | eGFR | 0.255663 | -0.26789 | 0.98372 | 0.0653636 | 0 | 0.97348 | 0.32604 | 452 | 0.659577784946237 |
| Chr15:76.6Mb | 2264 | 15 | 75958825 | 77253638 | 4916 | 292 | AD | eGFR | 0.143225 | -0.1491 | 0.45563 | 0.0205134 | 0 | 0.20791 | 0.327506 | 453 | 0.659577784946237 |
| Chr5:156.8Mb | 1039 | 5 | 156463091 | 157203559 | 3599 | 350 | AD | eGFR | -0.361744 | -1 | 0.45 | 0.130858 | 0 | 1 | 0.327551 | 454 | 0.659577784946237 |
| Chr19:16.7Mb | 2514 | 19 | 16253961 | 17230838 | 4173 | 466 | AD | eGFR | 0.177933 | -0.19679 | 0.58795 | 0.03166 | 0 | 0.34677 | 0.327745 | 455 | 0.659577784946237 |
| Chr20:15.4Mb | 2575 | 20 | 14919028 | 15796809 | 4406 | 495 | AD | eGFR | 0.25796 | -0.29685 | 1 | 0.0665434 | 0 | 1 | 0.32775 | 456 | 0.659577784946237 |
| Chr3:54Mb | 546 | 3 | 53593196 | 54488511 | 4163 | 464 | AD | eGFR | -0.249653 | -0.97816 | 0.2747 | 0.0623266 | 0 | 0.96742 | 0.328556 | 457 | 0.659577784946237 |
| Chr1:13.9Mb | 18 | 1 | 13382929 | 14395688 | 3562 | 455 | AD | eGFR | 0.309573 | -0.36676 | 1 | 0.0958355 | 0 | 1 | 0.329909 | 458 | 0.659577784946237 |
| Chr1:50.5Mb | 56 | 1 | 49202325 | 51776644 | 6277 | 337 | AD | eGFR | -0.194881 | -0.65259 | 0.2145 | 0.0379784 | 0 | 0.42683 | 0.330046 | 459 | 0.659577784946237 |
| Chr6:131.6Mb | 1187 | 6 | 130940046 | 132277919 | 6253 | 460 | AD | eGFR | -0.124732 | -0.38581 | 0.12694 | 0.015558 | 0 | 0.14885 | 0.330204 | 460 | 0.659577784946237 |
| Chr18:60.4Mb | 2478 | 18 | 59695907 | 61178035 | 5919 | 479 | AD | eGFR | -0.284463 | -1 | 0.33894 | 0.0809195 | 0 | 1 | 0.331346 | 461 | 0.659577784946237 |
| Chr7:7.3Mb | 1234 | 7 | 6975015 | 7674381 | 4658 | 432 | AD | eGFR | -0.322556 | -1 | 0.40067 | 0.104042 | 0 | 1 | 0.331883 | 462 | 0.659577784946237 |
| Chr9:84.4Mb | 1579 | 9 | 83712031 | 85125779 | 5044 | 482 | AD | eGFR | 0.311083 | -0.38063 | 1 | 0.0967723 | 0 | 1 | 0.332085 | 463 | 0.659577784946237 |
| Chr16:54.4Mb | 2324 | 16 | 53852142 | 54991694 | 5901 | 544 | AD | eGFR | 0.30617 | -0.36081 | 1 | 0.0937401 | 0 | 1 | 0.332251 | 464 | 0.659577784946237 |
| Chr20:21.6Mb | 2582 | 20 | 20965010 | 22290924 | 4794 | 371 | AD | eGFR | 0.237948 | -0.2746 | 0.9324 | 0.0566192 | 0 | 0.88857 | 0.33229 | 465 | 0.659577784946237 |
| Chr18:29.5Mb | 2453 | 18 | 29001460 | 30081048 | 4781 | 455 | AD | eGFR | 0.28045 | -0.33303 | 1 | 0.0786522 | 0 | 1 | 0.333029 | 466 | 0.65962610944206 |
| Chr13:27.9Mb | 2038 | 13 | 27562472 | 28271192 | 3725 | 414 | AD | eGFR | 0.229638 | -0.27393 | 0.84802 | 0.0527335 | 0 | 0.7224 | 0.334499 | 467 | 0.660783734607219 |
| Chr8:4.9Mb | 1379 | 8 | 4759959 | 5060433 | 2942 | 351 | AD | eGFR | -0.249547 | -0.97538 | 0.28471 | 0.0622737 | 0 | 0.95249 | 0.335686 | 468 | 0.660783734607219 |

| locus_label | locus | chr | start | stop | n.snps | n.pcs | phen1 | phen2 | rho | rho.lower | rho.upper | r2 | r2.lower | r2.upper | p | rank | FDR |
|---|---|---|---|---|---|---|---|---|---|---|---|---|---|---|---|---|---|
| Chr12:51.1Mb | 1951 | 12 | 50339060 | 51925776 | 5609 | 407 | AD | eGFR | -0.218036 | -0.78068 | 0.24928 | 0.0475396 | 0 | 0.6104 | 0.335865 | 469 | 0.660783734607219 |
| Chr10:13.7Mb | 1654 | 10 | 13272266 | 14138949 | 5071 | 518 | AD | eGFR | -0.270403 | -1 | 0.335 | 0.073118 | 0 | 1 | 0.336799 | 470 | 0.660783734607219 |
| Chr3:98.8Mb | 586 | 3 | 98256686 | 99373269 | 5667 | 352 | AD | eGFR | -0.283336 | -1 | 0.35923 | 0.0802794 | 0 | 1 | 0.337193 | 471 | 0.660783734607219 |
| Chr4:182.8Mb | 886 | 4 | 182422631 | 183112063 | 3818 | 484 | AD | eGFR | 0.312502 | -0.38735 | 1 | 0.0976576 | 0 | 1 | 0.338586 | 472 | 0.662107792372881 |
| Chr1:81.1Mb | 90 | 1 | 80787154 | 81495917 | 3751 | 380 | AD | eGFR | -0.237643 | -0.9023 | 0.28267 | 0.056474 | 0 | 0.82108 | 0.339523 | 473 | 0.662536424947146 |
| Chr3:117.5Mb | 604 | 3 | 116827914 | 118118782 | 5913 | 461 | AD | eGFR | -0.379642 | -1 | 0.49392 | 0.144128 | 0 | 1 | 0.341126 | 474 | 0.663011303157895 |
| Chr16:5.2Mb | 2292 | 16 | 5008305 | 5460499 | 2946 | 453 | AD | eGFR | -0.235454 | -0.97707 | 0.28956 | 0.0554386 | 0 | 0.99892 | 0.341203 | 475 | 0.663011303157895 |
| Chr4:96Mb | 795 | 4 | 95638488 | 96347018 | 3318 | 373 | AD | eGFR | 0.227437 | -0.27195 | 0.90786 | 0.0517277 | 0 | 0.83863 | 0.345569 | 476 | 0.670084426470588 |
| Chr1:106.4Mb | 114 | 1 | 106089281 | 106769892 | 3235 | 319 | AD | eGFR | 0.206855 | -0.24807 | 0.72621 | 0.0427892 | 0 | 0.53253 | 0.347043 | 477 | 0.67132029916318 |
| Chr14:77.1Mb | 2186 | 14 | 76780226 | 77430026 | 3452 | 467 | AD | eGFR | 0.267921 | -0.35212 | 1 | 0.0717815 | 0 | 1 | 0.347661 | 478 | 0.67132029916318 |
| Chr18:62.9Mb | 2480 | 18 | 62286819 | 63458747 | 5924 | 433 | AD | eGFR | 0.251686 | -0.31485 | 0.98764 | 0.0633457 | 0 | 0.99303 | 0.3491 | 479 | 0.671842297297297 |
| Chr9:132.9Mb | 1628 | 9 | 132441849 | 133394685 | 4068 | 481 | AD | eGFR | -0.162544 | -0.55293 | 0.19074 | 0.0264205 | 0 | 0.30668 | 0.349782 | 480 | 0.671842297297297 |
| Chr3:45.9Mb | 541 | 3 | 45176511 | 46660634 | 6873 | 427 | AD | eGFR | -0.377416 | -1 | 0.50342 | 0.142443 | 0 | 1 | 0.350115 | 481 | 0.671842297297297 |
| Chr12:121.4Mb | 2013 | 12 | 120833677 | 122003558 | 5440 | 416 | AD | eGFR | 0.20875 | -0.25727 | 0.76131 | 0.0435766 | 0 | 0.58263 | 0.351263 | 482 | 0.672601374741201 |
| Chr3:183.8Mb | 669 | 3 | 183111761 | 184524290 | 6617 | 530 | AD | eGFR | -0.266001 | -1 | 0.34941 | 0.0707565 | 0 | 1 | 0.351968 | 483 | 0.672601374741201 |
| Chr7:108.8Mb | 1327 | 7 | 108265780 | 109295963 | 5238 | 354 | AD | eGFR | 0.399829 | -0.57756 | 1 | 0.159863 | 0 | 1 | 0.352935 | 484 | 0.673055795454545 |
| Chr4:25.5Mb | 720 | 4 | 25110198 | 25873928 | 3884 | 457 | AD | eGFR | -0.215782 | -0.92326 | 0.27195 | 0.0465621 | 0 | 0.87393 | 0.354543 | 485 | 0.674728224742268 |
| Chr17:60Mb | 2409 | 17 | 59288227 | 60735733 | 5111 | 488 | AD | eGFR | 0.158298 | -0.20143 | 0.53945 | 0.0250583 | 0 | 0.29122 | 0.359903 | 486 | 0.683519483539095 |
| Chr1:185.9Mb | 170 | 1 | 185515602 | 186267150 | 3143 | 283 | AD | eGFR | 0.32832 | -0.50802 | 1 | 0.107794 | 0 | 1 | 0.363045 | 487 | 0.6880709137577 |
| Chr2:112.4Mb | 354 | 2 | 111849816 | 112918649 | 2799 | 377 | AD | eGFR | -0.183158 | -0.67388 | 0.23223 | 0.033547 | 0 | 0.45776 | 0.368126 | 488 | 0.695219077709611 |
| Chr1:98.4Mb | 106 | 1 | 97800990 | 98964964 | 4331 | 406 | AD | eGFR | -0.191317 | -0.71589 | 0.24215 | 0.0366021 | 0 | 0.51723 | 0.368323 | 489 | 0.695219077709611 |
| Chr12:76Mb | 1973 | 12 | 75563060 | 76526220 | 4116 | 455 | AD | eGFR | 0.13333 | -0.16837 | 0.44139 | 0.0177769 | 0 | 0.19507 | 0.369796 | 490 | 0.695797252545825 |
| Chr3:116.4Mb | 603 | 3 | 116048471 | 116827913 | 3672 | 383 | AD | eGFR | -0.274106 | -1 | 0.40133 | 0.0751339 | 0 | 1 | 0.370137 | 491 | 0.695797252545825 |
| Chr12:63.8Mb | 1962 | 12 | 63237669 | 64304577 | 4923 | 399 | AD | eGFR | 0.279497 | -0.40342 | 1 | 0.0781186 | 0 | 1 | 0.371657 | 492 | 0.695927052631579 |
| Chr10:130.1Mb | 1762 | 10 | 129667642 | 130481895 | 4515 | 495 | AD | eGFR | -0.310926 | -1 | 0.42773 | 0.096675 | 0 | 1 | 0.372181 | 493 | 0.695927052631579 |
| Chr4:31.8Mb | 728 | 4 | 31312684 | 32320369 | 5182 | 392 | AD | eGFR | 0.240822 | -0.32692 | 1 | 0.057995 | 0 | 1 | 0.372468 | 494 | 0.695927052631579 |
| Chr9:79.1Mb | 1573 | 9 | 78726503 | 79457934 | 3897 | 471 | AD | eGFR | -0.194447 | -0.72096 | 0.26425 | 0.0378095 | 0 | 0.5219 | 0.377641 | 495 | 0.703121302419355 |
| Chr13:49.2Mb | 2059 | 13 | 48382404 | 49988959 | 6815 | 409 | AD | eGFR | 0.20553 | -0.27997 | 0.77185 | 0.0422424 | 0 | 0.60314 | 0.377842 | 496 | 0.703121302419355 |
| Chr8:63.7Mb | 1439 | 8 | 63157950 | 64198821 | 4992 | 316 | AD | eGFR | -0.227359 | -0.9249 | 0.32156 | 0.051692 | 0 | 0.86036 | 0.380416 | 497 | 0.706486857142857 |
| Chr20:19.5Mb | 2580 | 20 | 19110515 | 19948089 | 4218 | 460 | AD | eGFR | 0.13752 | -0.17773 | 0.48341 | 0.0189118 | 0 | 0.23383 | 0.384995 | 498 | 0.713554989959839 |
| Chr4:111.4Mb | 810 | 4 | 110987608 | 111731728 | 3385 | 421 | AD | eGFR | 0.260599 | -0.40374 | 1 | 0.067912 | 0 | 1 | 0.391499 | 499 | 0.72415546492986 |
| Chr20:2Mb | 2560 | 20 | 1652525 | 2331544 | 3650 | 495 | AD | eGFR | 0.198058 | -0.27255 | 0.78734 | 0.039227 | 0 | 0.62511 | 0.393017 | 500 | 0.725509382 |
| Chr9:18.4Mb | 1538 | 9 | 18123171 | 18751638 | 3364 | 399 | AD | eGFR | 0.247821 | -0.3901 | 1 | 0.0614151 | 0 | 1 | 0.395941 | 501 | 0.729448189620758 |
| Chr3:39.1Mb | 534 | 3 | 38604837 | 39607201 | 4814 | 411 | AD | eGFR | 0.232429 | -0.38167 | 1 | 0.0540232 | 0 | 1 | 0.397598 | 502 | 0.730026940357853 |

| locus_label | locus | chr | start | stop | n.snps | n.pcs | phen1 | phen2 | rho | rho.lower | rho.upper | r2 | r2.lower | r2.upper | p | rank | FDR |
|---|---|---|---|---|---|---|---|---|---|---|---|---|---|---|---|---|---|
| Chr8:28.7Mb | 1410 | 8 | 28158441 | 29250773 | 4352 | 441 | AD | eGFR | 0.181883 | -0.27256 | 0.74226 | 0.0330815 | 0 | 0.56138 | 0.397837 | 503 | 0.730026940357853 |
| Chr5:109.1Mb | 998 | 5 | 108632394 | 109613714 | 4610 | 422 | AD | eGFR | 0.139716 | -0.19452 | 0.50346 | 0.0195205 | 0 | 0.25348 | 0.398631 | 504 | 0.730032565476191 |
| Chr2:37.5Mb | 283 | 2 | 37047209 | 37959618 | 4581 | 431 | AD | eGFR | -0.306417 | -1 | 0.52441 | 0.0938916 | 0 | 1 | 0.39969 | 505 | 0.730522514851485 |
| Chr1:146.5Mb | 131 | 1 | 145830857 | 147164393 | 2587 | 297 | AD | eGFR | -0.378697 | -1 | 0.68987 | 0.143412 | 0 | 1 | 0.401984 | 506 | 0.731173762278978 |
| Chr16:77.9Mb | 2344 | 16 | 77445654 | 78275341 | 5915 | 493 | AD | eGFR | -0.128073 | -0.45633 | 0.17466 | 0.0164027 | 0 | 0.20824 | 0.402151 | 507 | 0.731173762278978 |
| Chr22:45.8Mb | 2681 | 22 | 45264357 | 46273866 | 5291 | 516 | AD | eGFR | -0.213584 | -0.94893 | 0.3291 | 0.045618 | 0 | 0.93525 | 0.403041 | 508 | 0.731173762278978 |
| Chr14:22.9Mb | 2135 | 14 | 22621155 | 23239528 | 3040 | 428 | AD | eGFR | -0.208371 | -0.88938 | 0.31161 | 0.0434184 | 0 | 0.79347 | 0.403215 | 509 | 0.731173762278978 |
| Chr10:55.8Mb | 1693 | 10 | 55336213 | 56203308 | 5459 | 359 | AD | eGFR | 0.204011 | -0.33168 | 0.92307 | 0.0416204 | 0 | 0.89937 | 0.407012 | 510 | 0.73661191372549 |
| Chr1:40Mb | 46 | 1 | 39544840 | 40538447 | 3782 | 376 | AD | eGFR | -0.2552 | -1 | 0.41212 | 0.065127 | 0 | 1 | 0.408117 | 511 | 0.737166322896282 |
| Chr9:24.2Mb | 1545 | 9 | 23681385 | 24750125 | 4731 | 405 | AD | eGFR | 0.177505 | -0.26711 | 0.82113 | 0.0315081 | 0 | 0.69445 | 0.40894 | 512 | 0.7372101953125 |
| Chr9:85.5Mb | 1580 | 9 | 85125780 | 85940303 | 3533 | 428 | AD | eGFR | 0.196714 | -0.31729 | 0.84811 | 0.0386965 | 0 | 0.74479 | 0.412702 | 513 | 0.740226323643411 |
| Chr8:19.7Mb | 1401 | 8 | 19267303 | 20100434 | 5141 | 481 | AD | eGFR | -0.192698 | -0.79675 | 0.30215 | 0.0371324 | 0 | 0.64549 | 0.41303 | 514 | 0.740226323643411 |
| Chr13:38.8Mb | 2050 | 13 | 38289967 | 39403026 | 5336 | 455 | AD | eGFR | 0.220716 | -0.35015 | 0.96491 | 0.0487157 | 0 | 0.96685 | 0.413048 | 515 | 0.740226323643411 |
| Chr20:49.7Mb | 2606 | 20 | 49238194 | 50182144 | 4538 | 518 | AD | eGFR | -0.264145 | -1 | 0.45861 | 0.0697724 | 0 | 1 | 0.413821 | 516 | 0.740226323643411 |
| Chr3:7.1Mb | 498 | 3 | 6757829 | 7372532 | 3235 | 452 | AD | eGFR | -0.331336 | -1 | 0.58806 | 0.109784 | 0 | 1 | 0.415926 | 517 | 0.740535430210325 |
| Chr6:10.6Mb | 1069 | 6 | 10257684 | 10898990 | 3401 | 447 | AD | eGFR | -0.146658 | -0.54311 | 0.22086 | 0.0215086 | 0 | 0.29575 | 0.417236 | 518 | 0.740535430210325 |
| Chr5:170.5Mb | 1051 | 5 | 170051309 | 170888950 | 3406 | 355 | AD | eGFR | -0.191315 | -0.76998 | 0.30948 | 0.0366013 | 0 | 0.60763 | 0.418193 | 519 | 0.740535430210325 |
| Chr6:63.3Mb | 1125 | 6 | 62703091 | 63987530 | 5012 | 254 | AD | eGFR | 0.186903 | -0.29657 | 0.79931 | 0.0349326 | 0 | 0.67325 | 0.418493 | 520 | 0.740535430210325 |
| Chr9:110.4Mb | 1606 | 9 | 109994245 | 110759561 | 4267 | 496 | AD | eGFR | -0.152773 | -0.57622 | 0.23478 | 0.0233397 | 0 | 0.33393 | 0.418827 | 521 | 0.740535430210325 |
| Chr2:19.1Mb | 263 | 2 | 18644945 | 19629635 | 3825 | 387 | AD | eGFR | 0.22165 | -0.38293 | 0.99383 | 0.0491289 | 0 | 1 | 0.419182 | 522 | 0.740535430210325 |
| Chr8:23Mb | 1404 | 8 | 22414044 | 23547240 | 5734 | 532 | AD | eGFR | 0.20657 | -0.34904 | 0.98824 | 0.0426713 | 0 | 1 | 0.41961 | 523 | 0.740535430210325 |
| Chr11:47.2Mb | 1817 | 11 | 46260159 | 48112449 | 5149 | 285 | AD | eGFR | -0.250179 | -1 | 0.45984 | 0.0625896 | 0 | 1 | 0.422775 | 524 | 0.74328172243346 |
| Chr18:53.3Mb | 2472 | 18 | 52515306 | 54049083 | 4955 | 460 | AD | eGFR | -0.157209 | -0.61853 | 0.25037 | 0.0247147 | 0 | 0.38973 | 0.42343 | 525 | 0.74328172243346 |
| Chr1:173.3Mb | 157 | 1 | 172659528 | 173883516 | 4298 | 364 | AD | eGFR | 0.238372 | -0.41556 | 1 | 0.0568211 | 0 | 1 | 0.423582 | 526 | 0.74328172243346 |
| Chr10:131Mb | 1763 | 10 | 130481896 | 131463049 | 5293 | 529 | AD | eGFR | 0.202886 | -0.33592 | 0.88137 | 0.0411628 | 0 | 0.79539 | 0.424973 | 527 | 0.74430755028463 |
| Chr20:54.1Mb | 2611 | 20 | 53623184 | 54533715 | 5349 | 442 | AD | eGFR | -0.281323 | -1 | 0.51191 | 0.0791425 | 0 | 1 | 0.427337 | 528 | 0.74673302443609 |
| Chr22:30.2Mb | 2667 | 22 | 29460033 | 30971247 | 6213 | 406 | AD | eGFR | 0.181265 | -0.28904 | 0.7264 | 0.032857 | 0 | 0.53429 | 0.429069 | 529 | 0.74673302443609 |
| Chr2:195.4Mb | 435 | 2 | 194750209 | 195952215 | 4482 | 342 | AD | eGFR | -0.225091 | -1 | 0.39954 | 0.0506661 | 0 | 1 | 0.429637 | 530 | 0.74673302443609 |
| Chr4:168.2Mb | 868 | 4 | 167852837 | 168635745 | 3435 | 328 | AD | eGFR | -0.174325 | -0.69749 | 0.27976 | 0.0303894 | 0 | 0.48879 | 0.429743 | 531 | 0.74673302443609 |
| Chr2:224.6Mb | 463 | 2 | 224269423 | 225028695 | 3699 | 405 | AD | eGFR | -0.183424 | -0.79521 | 0.30304 | 0.0336445 | 0 | 0.6361 | 0.430403 | 532 | 0.74673302443609 |
| Chr9:28.2Mb | 1550 | 9 | 27834392 | 28520307 | 3767 | 442 | AD | eGFR | 0.204306 | -0.35573 | 0.89769 | 0.041741 | 0 | 0.82461 | 0.431932 | 533 | 0.747979804878049 |
| Chr20:40Mb | 2596 | 20 | 39439793 | 40597919 | 4230 | 379 | AD | eGFR | -0.134131 | -0.50656 | 0.21182 | 0.0179913 | 0 | 0.25776 | 0.435912 | 534 | 0.752198678438662 |
| Chr17:67.4Mb | 2415 | 17 | 66752528 | 68107835 | 5288 | 471 | AD | eGFR | -0.0900398 | -0.32868 | 0.14265 | 0.00810717 | 0 | 0.10847 | 0.436765 | 535 | 0.752198678438662 |
| Chr1:17Mb | 22 | 1 | 16517144 | 17496753 | 2456 | 302 | AD | eGFR | 0.227981 | -0.4214 | 1 | 0.0519754 | 0 | 1 | 0.437398 | 536 | 0.752198678438662 |
| Chr18:59.3Mb | 2477 | 18 | 58816179 | 59695906 | 4339 | 464 | AD | eGFR | -0.168559 | -0.84094 | 0.29899 | 0.0284122 | 0 | 0.7458 | 0.438268 | 537 | 0.752198678438662 |
| Chr10:76.4Mb | 1713 | 10 | 75693330 | 77144930 | 4626 | 346 | AD | eGFR | -0.253669 | -1 | 0.4766 | 0.0643479 | 0 | 1 | 0.438443 | 538 | 0.752198678438662 |
| Chr3:150.5Mb | 635 | 3 | 149993888 | 150948644 | 3644 | 461 | AD | eGFR | 0.150782 | -0.25914 | 0.67696 | 0.0227353 | 0 | 0.46374 | 0.441195 | 539 | 0.75453635174954 |

| locus_label | locus | chr | start | stop | n.snps | n.pcs | phen1 | phen2 | rho | rho.lower | rho.upper | r2 | r2.lower | r2.upper | p | rank | FDR |
|---|---|---|---|---|---|---|---|---|---|---|---|---|---|---|---|---|---|
| | | | 2 | 2 | | | | | | | | | | | | | |
| Chr11:88Mb | 1853 | 11 | 87665008 | 88406993 | 3768 | 370 | AD | eGFR | -0.174076 | -0.74535 | 0.30663 | 0.0303023 | 0 | 0.56468 | 0.442652 | 540 | 0.75453635174954 |
| Chr17:33.7Mb | 2390 | 17 | 32992143 | 34441644 | 6101 | 475 | AD | eGFR | 0.183697 | -0.31852 | 0.78499 | 0.0337446 | 0 | 0.63336 | 0.442747 | 541 | 0.75453635174954 |
| Chr13:103.9Mb | 2117 | 13 | 103556028 | 104217311 | 3590 | 438 | AD | eGFR | 0.132463 | -0.21074 | 0.50032 | 0.0175464 | 0 | 0.25159 | 0.443505 | 542 | 0.75453635174954 |
| Chr14:105Mb | 2212 | 14 | 104545284 | 105416021 | 4991 | 514 | AD | eGFR | 0.263631 | -0.51945 | 1 | 0.0695012 | 0 | 1 | 0.443893 | 543 | 0.75453635174954 |
| Chr13:78.5Mb | 2087 | 13 | 78035060 | 78975435 | 4160 | 326 | AD | eGFR | 0.208023 | -0.40246 | 0.96328 | 0.0432735 | 0 | 0.96662 | 0.446448 | 544 | 0.755130642857143 |
| Chr20:3.6Mb | 2562 | 20 | 3148307 | 4073061 | 4500 | 435 | AD | eGFR | -0.246559 | -1 | 0.4813 | 0.0607913 | 0 | 1 | 0.446674 | 545 | 0.755130642857143 |
| Chr3:29.9Mb | 524 | 3 | 29603547 | 30284145 | 3892 | 415 | AD | eGFR | -0.157833 | -0.78082 | 0.28222 | 0.0249114 | 0 | 0.6373 | 0.446697 | 546 | 0.755130642857143 |
| Chr12:133.4Mb | 2027 | 12 | 132950440 | 133841814 | 3655 | 375 | AD | eGFR | -0.186561 | -0.79982 | 0.33428 | 0.0348052 | 0 | 0.66316 | 0.447648 | 547 | 0.755354851919561 |
| Chr12:56.8Mb | 1956 | 12 | 56052414 | 57544979 | 4209 | 437 | AD | eGFR | 0.130236 | -0.22687 | 0.50188 | 0.0169613 | 0 | 0.25327 | 0.45272 | 548 | 0.76070140647482 |
| Chr19:55.4Mb | 2553 | 19 | 55169699 | 55729890 | 2619 | 461 | AD | eGFR | -0.134522 | -0.52893 | 0.23102 | 0.018096 | 0 | 0.28202 | 0.453717 | 549 | 0.76070140647482 |
| Chr10:73.7Mb | 1711 | 10 | 73233629 | 74069132 | 4233 | 466 | AD | eGFR | 0.168745 | -0.30604 | 0.70066 | 0.0284747 | 0 | 0.50104 | 0.455239 | 550 | 0.76070140647482 |
| Chr4:174.2Mb | 875 | 4 | 173805574 | 174689457 | 3880 | 411 | AD | eGFR | -0.283191 | -1 | 0.57807 | 0.080197 | 0 | 1 | 0.456046 | 551 | 0.76070140647482 |
| Chr8:133.1Mb | 1498 | 8 | 132538724 | 133574211 | 5350 | 452 | AD | eGFR | -0.145652 | -0.72518 | 0.26679 | 0.0212145 | 0 | 0.56868 | 0.456348 | 552 | 0.76070140647482 |
| Chr12:102.6Mb | 1996 | 12 | 101976788 | 103150649 | 4642 | 404 | AD | eGFR | 0.141533 | -0.2499 | 0.56588 | 0.0200316 | 0 | 0.32314 | 0.456938 | 553 | 0.76070140647482 |
| Chr6:118.1Mb | 1176 | 6 | 117675182 | 118529268 | 3713 | 319 | AD | eGFR | -0.214475 | -1 | 0.41818 | 0.0459995 | 0 | 1 | 0.457381 | 554 | 0.76070140647482 |
| Chr19:49Mb | 2544 | 19 | 48556497 | 49361067 | 4324 | 470 | AD | eGFR | -0.12654 | -0.49179 | 0.21234 | 0.0160125 | 0 | 0.24386 | 0.457501 | 555 | 0.76070140647482 |
| Chr3:196.1Mb | 684 | 3 | 195779323 | 196404922 | 3271 | 454 | AD | eGFR | -0.253683 | -1 | 0.5341 | 0.0643553 | 0 | 1 | 0.458234 | 556 | 0.76070140647482 |
| Chr2:4.6Mb | 245 | 2 | 4146631 | 4973956 | 4351 | 446 | AD | eGFR | -0.167797 | -0.86381 | 0.31509 | 0.028156 | 0 | 0.78509 | 0.464247 | 557 | 0.768337951612903 |
| Chr4:73.3Mb | 771 | 4 | 72979558 | 73675587 | 3471 | 275 | AD | eGFR | -0.332238 | -1 | 0.83758 | 0.110382 | 0 | 1 | 0.464499 | 558 | 0.768337951612903 |
| Chr22:25.7Mb | 2663 | 22 | 25285954 | 26165003 | 4149 | 475 | AD | eGFR | 0.176109 | -0.33876 | 0.78776 | 0.0310144 | 0 | 0.6417 | 0.465359 | 559 | 0.768383465116279 |
| Chr3:106.3Mb | 593 | 3 | 105669645 | 106985789 | 6483 | 446 | AD | eGFR | 0.243959 | -0.54686 | 1 | 0.059516 | 0 | 1 | 0.466882 | 560 | 0.769521582142857 |
| Chr16:75.5Mb | 2341 | 16 | 74985938 | 75971114 | 4833 | 369 | AD | eGFR | 0.309688 | -0.72707 | 1 | 0.0959067 | 0 | 1 | 0.46838 | 561 | 0.76970809430605 |
| Chr6:161.8Mb | 1216 | 6 | 161365186 | 162319341 | 5335 | 530 | AD | eGFR | 0.166649 | -0.33106 | 0.71879 | 0.0277717 | 0 | 0.53008 | 0.468663 | 562 | 0.76970809430605 |
| Chr1:160.3Mb | 143 | 1 | 159822096 | 160687163 | 3881 | 466 | AD | eGFR | 0.244639 | -0.5282 | 1 | 0.0598482 | 0 | 1 | 0.470608 | 563 | 0.77152963410302 |
| Chr3:174.2Mb | 660 | 3 | 173489075 | 174854628 | 6365 | 474 | AD | eGFR | -0.140004 | -0.60844 | 0.28 | 0.0196012 | 0 | 0.37744 | 0.471524 | 564 | 0.771660730496454 |
| Chr5:12.3Mb | 913 | 5 | 11914800 | 12784341 | 3685 | 266 | AD | eGFR | 0.179297 | -0.36869 | 0.80924 | 0.0321474 | 0 | 0.66416 | 0.474027 | 565 | 0.772494324561404 |
| Chr2:196.6Mb | 436 | 2 | 195952216 | 197310902 | 5895 | 390 | AD | eGFR | -0.140596 | -0.59254 | 0.25288 | 0.0197672 | 0 | 0.35344 | 0.474345 | 566 | 0.772494324561404 |
| Chr3:97.8Mb | 585 | 3 | 97343667 | 98256685 | 5042 | 328 | AD | eGFR | -0.22202 | -1 | 0.46029 | 0.0492929 | 0 | 1 | 0.474547 | 567 | 0.772494324561404 |
| Chr14:26.3Mb | 2138 | 14 | 25927290 | 26673619 | 3309 | 436 | AD | eGFR | 0.185249 | -0.39774 | 0.93061 | 0.0343172 | 0 | 0.91077 | 0.476229 | 568 | 0.772494324561404 |
| Chr1:178.1Mb | 161 | 1 | 177652309 | 178633069 | 3866 | 373 | AD | eGFR | -0.19409 | -0.98697 | 0.41922 | 0.0376709 | 0 | 1 | 0.476788 | 569 | 0.772494324561404 |
| Chr4:190.2Mb | 898 | 4 | 189893889 | 190419640 | 3025 | 388 | AD | eGFR | -0.250011 | -1 | 0.5795 | 0.0625053 | 0 | 1 | 0.477055 | 570 | 0.772494324561404 |

| locus_label | locus | chr | start | stop | n.snps | n.pcs | phen1 | phen2 | rho | rho.lower | rho.upper | r2 | r2.lower | r2.upper | p | rank | FDR |
|---|---|---|---|---|---|---|---|---|---|---|---|---|---|---|---|---|---|
| Chr9:100.6Mb | 1595 | 9 | 100163464 | 101078127 | 3584 | 396 | AD | eGFR | -0.117371 | -0.47654 | 0.21627 | 0.013776 | 0 | 0.23345 | 0.47819 | 571 | 0.772976129597198 |
| Chr6:25.2Mb | 1084 | 6 | 24848404 | 25485853 | 3554 | 388 | AD | eGFR | 0.18428 | -0.37499 | 0.88203 | 0.0339591 | 0 | 0.78673 | 0.481748 | 572 | 0.774510970434783 |
| Chr2:20.8Mb | 265 | 2 | 20410562 | 21274992 | 4085 | 457 | AD | eGFR | 0.123044 | -0.241 | 0.50897 | 0.0151399 | 0 | 0.26789 | 0.481985 | 573 | 0.774510970434783 |
| Chr9:105.3Mb | 1600 | 9 | 105001961 | 105622333 | 3864 | 295 | AD | eGFR | 0.152626 | -0.29764 | 0.64844 | 0.0232946 | 0 | 0.4264 | 0.482481 | 574 | 0.774510970434783 |
| Chr22:46.7Mb | 2682 | 22 | 46273867 | 47098260 | 5226 | 536 | AD | eGFR | -0.171541 | -0.79657 | 0.3596 | 0.0294264 | 0 | 0.6436 | 0.482496 | 575 | 0.774510970434783 |
| Chr8:131Mb | 1496 | 8 | 130378157 | 131544172 | 5388 | 431 | AD | eGFR | 0.235196 | -0.55189 | 1 | 0.055317 | 0 | 1 | 0.486361 | 576 | 0.778189084337349 |
| Chr6:84.9Mb | 1149 | 6 | 84217917 | 85583640 | 6058 | 404 | AD | eGFR | 0.196876 | -0.39872 | 0.98614 | 0.0387602 | 0 | 0.9884 | 0.486852 | 577 | 0.778189084337349 |
| Chr4:187.9Mb | 894 | 4 | 187563493 | 188222775 | 3621 | 480 | AD | eGFR | 0.192476 | -0.40447 | 0.95011 | 0.0370471 | 0 | 0.9419 | 0.489168 | 578 | 0.778189084337349 |
| Chr16:12.4Mb | 2303 | 16 | 11916769 | 12792106 | 5806 | 455 | AD | eGFR | -0.222432 | -1 | 0.51947 | 0.0494759 | 0 | 1 | 0.489494 | 579 | 0.778189084337349 |
| Chr19:41.9Mb | 2537 | 19 | 41540892 | 42260894 | 3028 | 402 | AD | eGFR | -0.144543 | -0.65683 | 0.30142 | 0.0208927 | 0 | 0.43649 | 0.489839 | 580 | 0.778189084337349 |
| Chr8:120.9Mb | 1487 | 8 | 120216588 | 121550897 | 5738 | 383 | AD | eGFR | -0.106466 | -0.45265 | 0.21939 | 0.0113351 | 0 | 0.2131 | 0.489846 | 581 | 0.778189084337349 |
| Chr1:11.2Mb | 15 | 1 | 10807359 | 11509989 | 3326 | 447 | AD | eGFR | -0.133872 | -0.58154 | 0.26195 | 0.0179216 | 0 | 0.34151 | 0.49127 | 582 | 0.779110326460481 |
| Chr6:40.9Mb | 1107 | 6 | 40550914 | 41299800 | 3552 | 377 | AD | eGFR | 0.127598 | -0.26744 | 0.52583 | 0.0162812 | 0 | 0.28004 | 0.492788 | 583 | 0.78017722813036 |
| Chr10:16.3Mb | 1657 | 10 | 15938240 | 16721533 | 4359 | 448 | AD | eGFR | -0.173625 | -0.89703 | 0.38206 | 0.0301455 | 0 | 0.82378 | 0.496408 | 584 | 0.783808444444444 |
| Chr4:164.3Mb | 863 | 4 | 163798852 | 164770678 | 4374 | 450 | AD | eGFR | 0.236911 | -0.59447 | 1 | 0.0561266 | 0 | 1 | 0.49678 | 585 | 0.783808444444444 |
| Chr7:74.8Mb | 1298 | 7 | 73979428 | 75606530 | 2561 | 394 | AD | eGFR | -0.184666 | -0.97538 | 0.409 | 0.0341016 | 0 | 1 | 0.498692 | 586 | 0.784378608177172 |
| Chr9:83.4Mb | 1578 | 9 | 83076044 | 83712030 | 3754 | 277 | AD | eGFR | -0.150142 | -0.67308 | 0.31507 | 0.0225425 | 0 | 0.45902 | 0.498841 | 587 | 0.784378608177172 |
| Chr13:70.4Mb | 2079 | 13 | 70079765 | 70725873 | 3543 | 335 | AD | eGFR | -0.186307 | -0.95075 | 0.42929 | 0.0347102 | 0 | 0.93617 | 0.502241 | 588 | 0.786294313559322 |
| Chr3:111.3Mb | 598 | 3 | 110940759 | 111724651 | 3796 | 360 | AD | eGFR | 0.122039 | -0.25464 | 0.51443 | 0.0148935 | 0 | 0.26805 | 0.502528 | 589 | 0.786294313559322 |
| Chr14:36.7Mb | 2147 | 14 | 35881123 | 37449106 | 6152 | 500 | AD | eGFR | -0.265799 | -1 | 0.67375 | 0.0706494 | 0 | 1 | 0.502615 | 590 | 0.786294313559322 |
| Chr21:35Mb | 2642 | 21 | 34384933 | 35601770 | 5049 | 483 | AD | eGFR | -0.17777 | -0.9233 | 0.40899 | 0.0316023 | 0 | 0.88077 | 0.505532 | 591 | 0.789519519458545 |
| Chr7:158.7Mb | 1370 | 7 | 158173404 | 159128566 | 5340 | 422 | AD | eGFR | -0.227763 | -1 | 0.5665 | 0.0518759 | 0 | 1 | 0.508042 | 592 | 0.791771207986689 |
| Chr4:152.6Mb | 852 | 4 | 152166814 | 153095134 | 3968 | 328 | AD | eGFR | 0.199954 | -0.5001 | 1 | 0.0399816 | 0 | 1 | 0.509914 | 593 | 0.791771207986689 |
| Chr18:36.9Mb | 2459 | 18 | 36061344 | 37681534 | 5756 | 400 | AD | eGFR | 0.194956 | -0.48297 | 1 | 0.0380079 | 0 | 1 | 0.512957 | 594 | 0.791771207986689 |
| Chr6:98.8Mb | 1161 | 6 | 98213894 | 99301915 | 3420 | 375 | AD | eGFR | -0.135638 | -0.60863 | 0.29919 | 0.0183977 | 0 | 0.38366 | 0.513267 | 595 | 0.791771207986689 |
| Chr3:177.4Mb | 664 | 3 | 176968046 | 177741266 | 3378 | 430 | AD | eGFR | -0.107796 | -0.45535 | 0.22322 | 0.01162 | 0 | 0.20883 | 0.513504 | 596 | 0.791771207986689 |
| Chr2:18.3Mb | 262 | 2 | 17992214 | 18644944 | 3410 | 374 | AD | eGFR | 0.198485 | -0.47753 | 1 | 0.0393963 | 0 | 1 | 0.514257 | 597 | 0.791771207986689 |
| Chr15:50.4Mb | 2240 | 15 | 50004139 | 50810602 | 3427 | 383 | AD | eGFR | 0.139851 | -0.3287 | 0.71594 | 0.0195582 | 0 | 0.53871 | 0.514761 | 598 | 0.791771207986689 |
| Chr8:119.2Mb | 1485 | 8 | 118828658 | 119533403 | 3479 | 397 | AD | eGFR | 0.234697 | -0.61068 | 1 | 0.0550825 | 0 | 1 | 0.514773 | 599 | 0.791771207986689 |
| Chr13:47.7Mb | 2058 | 13 | 47059424 | 48382403 | 6423 | 490 | AD | eGFR | -0.151684 | -0.7432 | 0.34787 | 0.023008 | 0 | 0.57068 | 0.514975 | 600 | 0.791771207986689 |
| Chr16:74.5Mb | 2340 | 16 | 73990487 | 74985937 | 3993 | 454 | AD | eGFR | -0.139616 | -0.63641 | 0.30038 | 0.0194926 | 0 | 0.41087 | 0.515552 | 601 | 0.791771207986689 |
| Chr17:66.2Mb | 2414 | 17 | 65630909 | 66752527 | 5425 | 480 | AD | eGFR | 0.190929 | -0.47139 | 1 | 0.0364539 | 0 | 1 | 0.517085 | 602 | 0.792806403654485 |
| Chr2:216.5Mb | 454 | 2 | 216138383 | 216873959 | 3361 | 458 | AD | eGFR | -0.102678 | -0.43798 | 0.2202 | 0.0105427 | 0 | 0.19586 | 0.521343 | 603 | 0.795056265472313 |
| Chr21:33.8Mb | 2641 | 21 | 33231681 | 34384932 | 5000 | 505 | AD | eGFR | -0.13262 | -0.59754 | 0.30052 | 0.0175879 | 0 | 0.36706 | 0.521409 | 604 | 0.795056265472313 |

| locus_label | locus | chr | start | stop | n.snps | n.pcs | phen1 | phen2 | rho | rho.lower | rho.upper | r2 | r2.lower | r2.upper | p | rank | FDR |
|---|---|---|---|---|---|---|---|---|---|---|---|---|---|---|---|---|---|
| Chr9:99.6Mb | 1594 | 9 | 99084180 | 100163463 | 3930 | 283 | AD | eGFR | -0.14965 | -0.81012 | 0.35164 | 0.0223951 | 0 | 0.69456 | 0.523508 | 605 | 0.795056265472313 |
| Chr3:40.8Mb | 536 | 3 | 40356576 | 41295931 | 3467 | 384 | AD | eGFR | 0.164971 | -0.40545 | 0.84035 | 0.0272155 | 0 | 0.73311 | 0.523755 | 606 | 0.795056265472313 |
| Chr2:47.4Mb | 295 | 2 | 47026594 | 47765702 | 3893 | 446 | AD | eGFR | -0.104937 | -0.45098 | 0.22242 | 0.0110119 | 0 | 0.2065 | 0.523987 | 607 | 0.795056265472313 |
| Chr13:92.8Mb | 2103 | 13 | 92381083 | 93131001 | 3767 | 378 | AD | eGFR | 0.230752 | -0.65065 | 1 | 0.0532464 | 0 | 1 | 0.525728 | 608 | 0.795056265472313 |
| Chr9:1.3Mb | 1513 | 9 | 1075174 | 1617712 | 3623 | 464 | AD | eGFR | -0.167832 | -0.89992 | 0.41709 | 0.0281675 | 0 | 0.8364 | 0.527189 | 609 | 0.795056265472313 |
| Chr3:14.1Mb | 507 | 3 | 13739920 | 14410060 | 3539 | 398 | AD | eGFR | 0.140305 | -0.32407 | 0.65456 | 0.0196854 | 0 | 0.43849 | 0.527491 | 610 | 0.795056265472313 |
| Chr20:45Mb | 2601 | 20 | 44533135 | 45559112 | 4536 | 489 | AD | eGFR | -0.132223 | -0.62114 | 0.29939 | 0.0174828 | 0 | 0.39157 | 0.528146 | 611 | 0.795056265472313 |
| Chr2:240.5Mb | 481 | 2 | 240060325 | 240968745 | 4996 | 519 | AD | eGFR | 0.125285 | -0.2991 | 0.63252 | 0.0156963 | 0 | 0.41326 | 0.528158 | 612 | 0.795056265472313 |
| Chr16:73.5Mb | 2339 | 16 | 72932710 | 73990486 | 5262 | 524 | AD | eGFR | -0.199079 | -1 | 0.52255 | 0.0396324 | 0 | 1 | 0.528356 | 613 | 0.795056265472313 |
| Chr2:199.1Mb | 438 | 2 | 198496670 | 199661930 | 4270 | 389 | AD | eGFR | -0.142176 | -0.6693 | 0.32675 | 0.0202139 | 0 | 0.46292 | 0.528889 | 614 | 0.795056265472313 |
| Chr6:112.2Mb | 1171 | 6 | 111406304 | 112976488 | 5806 | 449 | AD | eGFR | 0.118194 | -0.25834 | 0.54379 | 0.0139698 | 0 | 0.30068 | 0.529762 | 615 | 0.795073700813008 |
| Chr22:41Mb | 2676 | 22 | 40378883 | 41630262 | 3767 | 290 | AD | eGFR | -0.147043 | -0.76709 | 0.36259 | 0.0216216 | 0 | 0.60464 | 0.531093 | 616 | 0.795777336038961 |
| Chr1:102.6Mb | 110 | 1 | 102180315 | 103101623 | 4866 | 380 | AD | eGFR | -0.183734 | -1 | 0.47305 | 0.0337582 | 0 | 1 | 0.533184 | 617 | 0.796786470873786 |
| Chr15:77.9Mb | 2265 | 15 | 77253639 | 78635159 | 6084 | 437 | AD | eGFR | -0.195811 | -1 | 0.5234 | 0.0383419 | 0 | 1 | 0.533493 | 618 | 0.796786470873786 |
| Chr2:79.5Mb | 329 | 2 | 79013336 | 79900021 | 4341 | 442 | AD | eGFR | -0.154461 | -0.78285 | 0.35619 | 0.023858 | 0 | 0.62664 | 0.534987 | 619 | 0.7970149375 |
| Chr3:144.3Mb | 628 | 3 | 143976385 | 144703587 | 3688 | 319 | AD | eGFR | -0.19334 | -1 | 0.51959 | 0.0373804 | 0 | 1 | 0.53711 | 620 | 0.7970149375 |
| Chr6:109.3Mb | 1169 | 6 | 108486470 | 110160229 | 6796 | 410 | AD | eGFR | 0.164167 | -0.42671 | 0.87847 | 0.0269507 | 0 | 0.80894 | 0.537528 | 621 | 0.7970149375 |
| Chr20:9.4Mb | 2569 | 20 | 8922498 | 9932817 | 4395 | 489 | AD | eGFR | -0.16698 | -0.96624 | 0.45176 | 0.0278825 | 0 | 1 | 0.538113 | 622 | 0.7970149375 |
| Chr4:40.5Mb | 739 | 4 | 40173280 | 40876504 | 3592 | 504 | AD | eGFR | -0.138031 | -0.70577 | 0.34132 | 0.0190527 | 0 | 0.5113 | 0.538499 | 623 | 0.7970149375 |
| Chr1:240.4Mb | 228 | 1 | 240113519 | 240708621 | 3521 | 448 | AD | eGFR | -0.183482 | -1 | 0.49816 | 0.0336657 | 0 | 1 | 0.538827 | 624 | 0.7970149375 |
| Chr11:48.5Mb | 1818 | 11 | 48112450 | 48867505 | 4362 | 125 | AD | eGFR | 0.168763 | -0.44373 | 0.95895 | 0.0284809 | 0 | 0.99875 | 0.540413 | 625 | 0.7980819184 |
| Chr8:57.3Mb | 1433 | 8 | 56603271 | 57906779 | 6337 | 408 | AD | eGFR | 0.166438 | -0.44557 | 0.93761 | 0.0277015 | 0 | 0.98159 | 0.543265 | 626 | 0.801012132587859 |
| Chr10:83.8Mb | 1720 | 10 | 83304172 | 84387256 | 5091 | 431 | AD | eGFR | -0.206221 | -1 | 0.57515 | 0.0425271 | 0 | 1 | 0.545656 | 627 | 0.801382296825397 |
| Chr4:92.1Mb | 791 | 4 | 91560822 | 92645779 | 5334 | 382 | AD | eGFR | -0.236019 | -1 | 0.69378 | 0.055705 | 0 | 1 | 0.546641 | 628 | 0.801382296825397 |
| Chr9:109.5Mb | 1605 | 9 | 109062084 | 109994244 | 3716 | 443 | AD | eGFR | -0.0965004 | -0.44448 | 0.22435 | 0.00931233 | 0 | 0.19973 | 0.546864 | 629 | 0.801382296825397 |
| Chr12:46.2Mb | 1946 | 12 | 45716268 | 46624746 | 3466 | 390 | AD | eGFR | -0.133932 | -0.67799 | 0.33923 | 0.0179378 | 0 | 0.48265 | 0.546989 | 630 | 0.801382296825397 |
| Chr1:227.4Mb | 213 | 1 | 226816213 | 227983744 | 5535 | 305 | AD | eGFR | -0.132958 | -0.68367 | 0.33854 | 0.0176779 | 0 | 0.49199 | 0.548654 | 631 | 0.802547768621236 |
| Chr9:70.3Mb | 1565 | 9 | 68993319 | 71590806 | 2302 | 315 | AD | eGFR | -0.103635 | -0.49103 | 0.25621 | 0.0107402 | 0 | 0.2473 | 0.550128 | 632 | 0.802810235387046 |
| Chr4:163.5Mb | 862 | 4 | 163174931 | 163798851 | 3546 | 325 | AD | eGFR | -0.231311 | -1 | 0.68343 | 0.0535049 | 0 | 1 | 0.550573 | 633 | 0.802810235387046 |
| Chr12:43.6Mb | 1943 | 12 | 43210526 | 44044407 | 3845 | 349 | AD | eGFR | 0.121753 | -0.29639 | 0.59284 | 0.0148237 | 0 | 0.3618 | 0.552128 | 634 | 0.803807798107255 |
| Chr12:71.5Mb | 1969 | 12 | 71012752 | 71948455 | 4298 | 408 | AD | eGFR | -0.128132 | -0.65591 | 0.33115 | 0.0164177 | 0 | 0.44478 | 0.555327 | 635 | 0.806276 |
| Chr13:81.2Mb | 2090 | 13 | 80783432 | 81657573 | 3992 | 294 | AD | eGFR | -0.210464 | -1 | 0.65567 | 0.0442949 | 0 | 1 | 0.556383 | 636 | 0.806276 |
| Chr1:208.6Mb | 193 | 1 | 208157326 | 208973257 | 3923 | 477 | AD | eGFR | -0.199439 | -1 | 0.62208 | 0.039776 | 0 | 1 | 0.556444 | 637 | 0.806276 |
| Chr4:144Mb | 844 | 4 | 14350453 | 14440978 | 3654 | 279 | AD | eGFR | 0.118538 | -0.31319 | 0.58722 | 0.0140512 | 0 | 0.35535 | 0.558449 | 638 | 0.80651768798752 |

| locus_label | locus | chr | start | stop | n.snps | n.pcs | phen1 | phen2 | rho | rho.lower | rho.upper | r2 | r2.lower | r2.upper | p | rank | FDR |
|---|---|---|---|---|---|---|---|---|---|---|---|---|---|---|---|---|---|
| | | | 8 | 7 | | | | | | | | | | | | | |
| Chr3:146Mb | 630 | 3 | 145353051 | 146643562 | 6735 | 439 | AD | eGFR | 0.196178 | -0.58792 | 1 | 0.0384859 | 0 | 1 | 0.559423 | 639 | 0.80651768798752 |
| Chr19:34.1Mb | 2530 | 19 | 33640646 | 34637026 | 5133 | 511 | AD | eGFR | 0.158168 | -0.46057 | 0.8807 | 0.0250172 | 0 | 0.82766 | 0.55962 | 640 | 0.80651768798752 |
| Chr4:175.9Mb | 877 | 4 | 175593532 | 176305193 | 3807 | 340 | AD | eGFR | -0.16106 | -0.91067 | 0.46507 | 0.0259402 | 0 | 0.86559 | 0.560106 | 641 | 0.80651768798752 |
| Chr1:229.7Mb | 215 | 1 | 229114217 | 230207176 | 4927 | 459 | AD | eGFR | 0.120268 | -0.31575 | 0.57669 | 0.0144644 | 0 | 0.34277 | 0.561016 | 642 | 0.806569732087227 |
| Chr3:12Mb | 504 | 3 | 11625633 | 12304562 | 3278 | 408 | AD | eGFR | 0.103212 | -0.28755 | 0.50968 | 0.0106527 | 0 | 0.27714 | 0.567232 | 643 | 0.812022245053272 |
| Chr1:234.8Mb | 221 | 1 | 234426276 | 235246448 | 3581 | 511 | AD | eGFR | 0.116111 | -0.33102 | 0.61092 | 0.0134818 | 0 | 0.39519 | 0.567626 | 644 | 0.812022245053272 |
| Chr6:24.5Mb | 1083 | 6 | 24099964 | 24848403 | 3855 | 384 | AD | eGFR | -0.120153 | -0.61532 | 0.3411 | 0.0144367 | 0 | 0.40536 | 0.56793 | 645 | 0.812022245053272 |
| Chr8:73.7Mb | 1447 | 8 | 73169869 | 74285426 | 5119 | 476 | AD | eGFR | -0.218494 | -1 | 0.69098 | 0.0477397 | 0 | 1 | 0.568473 | 646 | 0.812022245053272 |
| Chr11:99.9Mb | 1865 | 11 | 99295649 | 100410650 | 6043 | 449 | AD | eGFR | 0.169422 | -0.51419 | 1 | 0.0287038 | 0 | 1 | 0.570248 | 647 | 0.812022245053272 |
| Chr4:135.2Mb | 834 | 4 | 134629989 | 135790700 | 4957 | 323 | AD | eGFR | 0.149166 | -0.43615 | 0.80039 | 0.0222506 | 0 | 0.6775 | 0.570968 | 648 | 0.812022245053272 |
| Chr2:122.7Mb | 364 | 2 | 122036712 | 123305677 | 4712 | 362 | AD | eGFR | -0.158291 | -0.93603 | 0.46294 | 0.025056 | 0 | 0.94554 | 0.572384 | 649 | 0.812022245053272 |
| Chr2:123.7Mb | 365 | 2 | 123305678 | 124151942 | 4646 | 337 | AD | eGFR | 0.169772 | -0.51637 | 1 | 0.0288225 | 0 | 1 | 0.573658 | 650 | 0.812022245053272 |
| Chr9:134.6Mb | 1630 | 9 | 134170122 | 135039243 | 3693 | 448 | AD | eGFR | -0.229239 | -1 | 0.81134 | 0.0525504 | 0 | 1 | 0.575087 | 651 | 0.812022245053272 |
| Chr1:90.9Mb | 100 | 1 | 90378148 | 91341821 | 3619 | 419 | AD | eGFR | 0.156225 | -0.47692 | 0.91342 | 0.0244062 | 0 | 0.91218 | 0.575216 | 652 | 0.812022245053272 |
| Chr18:14.7Mb | 2443 | 18 | 14192411 | 15199439 | 3152 | 371 | AD | eGFR | 0.14367 | -0.43681 | 0.8522 | 0.0206409 | 0 | 0.81629 | 0.575247 | 653 | 0.812022245053272 |
| Chr5:40Mb | 940 | 5 | 39441429 | 40623660 | 5560 | 357 | AD | eGFR | -0.147438 | -0.84803 | 0.4361 | 0.0217379 | 0 | 0.78634 | 0.575399 | 654 | 0.812022245053272 |
| Chr20:46.8Mb | 2603 | 20 | 46394091 | 47197528 | 4481 | 491 | AD | eGFR | -0.113055 | -0.58221 | 0.30713 | 0.0127814 | 0 | 0.34745 | 0.57747 | 655 | 0.812022245053272 |
| Chr5:127.8Mb | 1016 | 5 | 126992655 | 128509205 | 6072 | 405 | AD | eGFR | 0.18742 | -0.59978 | 1 | 0.0351263 | 0 | 1 | 0.577823 | 656 | 0.812022245053272 |
| Chr15:55.5Mb | 2246 | 15 | 55134740 | 55830521 | 3717 | 334 | AD | eGFR | 0.107533 | -0.28856 | 0.52141 | 0.0115634 | 0 | 0.2816 | 0.578005 | 657 | 0.812022245053272 |
| Chr3:127.6Mb | 614 | 3 | 127082017 | 128183428 | 4683 | 424 | AD | eGFR | -0.181765 | -1 | 0.5969 | 0.0330387 | 0 | 1 | 0.579613 | 658 | 0.813043767477204 |
| Chr4:186.1Mb | 891 | 4 | 185740157 | 186381950 | 3493 | 456 | AD | eGFR | 0.193054 | -0.65754 | 1 | 0.0372697 | 0 | 1 | 0.581587 | 659 | 0.813165792738275 |
| Chr11:17.6Mb | 1787 | 11 | 16999441 | 18166577 | 4429 | 457 | AD | eGFR | 0.0631947 | -0.16616 | 0.30045 | 0.00399357 | 0 | 0.09252 | 0.582208 | 660 | 0.813165792738275 |
| Chr16:56.5Mb | 2326 | 16 | 55913026 | 57170193 | 6043 | 501 | AD | eGFR | 0.160812 | -0.5015 | 0.96903 | 0.0258605 | 0 | 0.99724 | 0.582343 | 661 | 0.813165792738275 |
| Chr12:113.9Mb | 2006 | 12 | 113349778 | 114456894 | 5324 | 470 | AD | eGFR | 0.139202 | -0.43882 | 0.84944 | 0.0193772 | 0 | 0.80352 | 0.585341 | 662 | 0.816117436555891 |
| Chr3:197.6Mb | 686 | 3 | 197150044 | 197962380 | 2834 | 374 | AD | eGFR | 0.191115 | -0.67825 | 1 | 0.0365249 | 0 | 1 | 0.586281 | 663 | 0.816195117647059 |
| Chr2:102.3Mb | 345 | 2 | 101827857 | 102689083 | 3799 | 352 | AD | eGFR | -0.145674 | -0.86209 | 0.45893 | 0.0212209 | 0 | 0.80432 | 0.594511 | 664 | 0.825156464767616 |
| Chr18:75.6Mb | 2492 | 18 | 75111248 | 76065048 | 5591 | 525 | AD | eGFR | -0.160691 | -1 | 0.52983 | 0.0258216 | 0 | 1 | 0.595279 | 665 | 0.825156464767616 |
| Chr9:74Mb | 1568 | 9 | 73411393 | 74623887 | 5130 | 447 | AD | eGFR | -0.142232 | -0.8662 | 0.42439 | 0.0202299 | 0 | 0.79067 | 0.595621 | 666 | 0.825156464767616 |
| Chr3:176.6Mb | 663 | 3 | 176266861 | 176968045 | 3433 | 373 | AD | eGFR | 0.162663 | -0.56457 | 1 | 0.0264591 | 0 | 1 | 0.596294 | 667 | 0.825156464767616 |
| Chr11:24.8Mb | 1795 | 11 | 24413370 | 25270470 | 5180 | 451 | AD | eGFR | 0.105658 | -0.31197 | 0.55739 | 0.0111636 | 0 | 0.32036 | 0.599125 | 668 | 0.827832896706587 |

| locus_label | locus | chr | start | stop | n.snps | n.pcs | phen1 | phen2 | rho | rho.lower | rho.upper | r2 | r2.lower | r2.upper | p | rank | FDR |
|---|---|---|---|---|---|---|---|---|---|---|---|---|---|---|---|---|---|
| Chr3:167.1Mb | 653 | 3 | 166597489 | 167521067 | 3778 | 317 | AD | eGFR | -0.111853 | -0.60273 | 0.34633 | 0.0125111 | 0 | 0.37712 | 0.602482 | 669 | 0.831014095522388 |
| Chr8:142.5Mb | 1507 | 8 | 142029622 | 142884238 | 4741 | 525 | AD | eGFR | 0.138442 | -0.44991 | 0.8178 | 0.0191663 | 0 | 0.71976 | 0.603228 | 670 | 0.831014095522388 |
| Chr2:66.2Mb | 314 | 2 | 65712250 | 66720835 | 4334 | 455 | AD | eGFR | 0.128229 | -0.44941 | 0.80762 | 0.0164428 | 0 | 0.72156 | 0.605872 | 671 | 0.833412602086438 |
| Chr9:23.3Mb | 1544 | 9 | 22920515 | 23681384 | 3999 | 464 | AD | eGFR | 0.132852 | -0.47402 | 0.85986 | 0.0176496 | 0 | 0.84765 | 0.609511 | 672 | 0.83594861961367 |
| Chr11:16.4Mb | 1786 | 11 | 15744947 | 16999440 | 4606 | 402 | AD | eGFR | 0.0997601 | -0.30235 | 0.53355 | 0.00995208 | 0 | 0.3025 | 0.609527 | 673 | 0.83594861961367 |
| Chr1:53.6Mb | 58 | 1 | 53112695 | 54015368 | 4323 | 392 | AD | eGFR | -0.177948 | -1 | 0.66287 | 0.0316655 | 0 | 1 | 0.611605 | 674 | 0.837554028189911 |
| Chr9:78.3Mb | 1572 | 9 | 77862458 | 78726502 | 4420 | 471 | AD | eGFR | -0.107171 | -0.61724 | 0.34905 | 0.0114857 | 0 | 0.40476 | 0.615305 | 675 | 0.841372614814815 |
| Chr1:59.1Mb | 65 | 1 | 58712790 | 59484886 | 3781 | 406 | AD | eGFR | 0.176621 | -0.67588 | 1 | 0.031195 | 0 | 1 | 0.617249 | 676 | 0.842782288461539 |
| Chr13:93.5Mb | 2104 | 13 | 93131002 | 93922801 | 3561 | 356 | AD | eGFR | -0.106124 | -0.6071 | 0.35859 | 0.0112623 | 0 | 0.39103 | 0.619169 | 677 | 0.844155076809453 |
| Chr3:7.7Mb | 499 | 3 | 7372533 | 7958599 | 3410 | 416 | AD | eGFR | 0.111933 | -0.38316 | 0.64223 | 0.0125291 | 0 | 0.44079 | 0.62051 | 678 | 0.844735589970501 |
| Chr15:79.3Mb | 2266 | 15 | 78635160 | 79991196 | 6598 | 509 | AD | eGFR | -0.174458 | -1 | 0.67931 | 0.0304357 | 0 | 1 | 0.622098 | 679 | 0.845650153166421 |
| Chr12:30.7Mb | 1930 | 12 | 30359018 | 31140247 | 4777 | 379 | AD | eGFR | 0.120199 | -0.40491 | 0.70965 | 0.0144479 | 0 | 0.53751 | 0.623337 | 680 | 0.846088310294118 |
| Chr9:111.2Mb | 1607 | 9 | 110759562 | 111624008 | 4709 | 429 | AD | eGFR | -0.0861146 | -0.46737 | 0.278 | 0.00741572 | 0 | 0.23128 | 0.628771 | 681 | 0.851745210834553 |
| Chr13:110.5Mb | 2127 | 13 | 110141686 | 110867691 | 4039 | 469 | AD | eGFR | 0.0899124 | -0.28568 | 0.48677 | 0.00808424 | 0 | 0.24604 | 0.62943 | 682 | 0.851745210834553 |
| Chr6:41.9Mb | 1108 | 6 | 41299801 | 42458359 | 5496 | 540 | AD | eGFR | 0.10523 | -0.37483 | 0.61345 | 0.0110733 | 0 | 0.40019 | 0.630273 | 683 | 0.851745210834553 |
| Chr6:132.8Mb | 1188 | 6 | 132277920 | 133282303 | 5342 | 440 | AD | eGFR | 0.0868195 | -0.28231 | 0.47318 | 0.00753762 | 0 | 0.23746 | 0.632291 | 684 | 0.852054312408759 |
| Chr6:160.3Mb | 1214 | 6 | 159985062 | 160698175 | 3397 | 387 | AD | eGFR | -0.0870567 | -0.56395 | 0.31301 | 0.00757886 | 0 | 0.35109 | 0.632348 | 685 | 0.852054312408759 |
| Chr3:114.3Mb | 601 | 3 | 113309391 | 115236928 | 6611 | 482 | AD | eGFR | 0.13092 | -0.49657 | 0.8958 | 0.0171401 | 0 | 0.87089 | 0.634147 | 686 | 0.853232771137026 |
| Chr12:69.3Mb | 1967 | 12 | 68739922 | 69841685 | 4785 | 485 | AD | eGFR | 0.120399 | -0.46247 | 0.80529 | 0.0144959 | 0 | 0.74212 | 0.636579 | 687 | 0.855258248908297 |
| Chr4:106.1Mb | 805 | 4 | 105707616 | 106585579 | 3238 | 335 | AD | eGFR | -0.141842 | -1 | 0.54568 | 0.0201192 | 0 | 1 | 0.639892 | 688 | 0.855420638205499 |
| Chr11:123.7Mb | 1887 | 11 | 123333325 | 124000844 | 3837 | 413 | AD | eGFR | -0.146813 | -1 | 0.59663 | 0.021554 | 0 | 1 | 0.639939 | 689 | 0.855420638205499 |
| Chr1:19.3Mb | 25 | 1 | 18906907 | 19631520 | 3628 | 438 | AD | eGFR | 0.0966521 | -0.33757 | 0.55789 | 0.00934163 | 0 | 0.32626 | 0.640352 | 690 | 0.855420638205499 |
| Chr9:6Mb | 1520 | 9 | 5524509 | 6552037 | 4139 | 380 | AD | eGFR | 0.148514 | -0.62156 | 1 | 0.0220563 | 0 | 1 | 0.640407 | 691 | 0.855420638205499 |
| Chr18:77.6Mb | 2494 | 18 | 77147775 | 78017155 | 4331 | 453 | AD | eGFR | 0.126449 | -0.5044 | 0.84979 | 0.0159894 | 0 | 0.78632 | 0.643042 | 692 | 0.857699083815029 |
| Chr14:100.8Mb | 2208 | 14 | 100051311 | 101550334 | 6868 | 550 | AD | eGFR | -0.103107 | -0.63688 | 0.38222 | 0.010631 | 0 | 0.43581 | 0.64553 | 693 | 0.859775165945166 |
| Chr10:31.2Mb | 1673 | 10 | 30882856 | 31484983 | 3450 | 359 | AD | eGFR | 0.101779 | -0.37665 | 0.62908 | 0.010359 | 0 | 0.42318 | 0.646906 | 694 | 0.860366337175793 |
| Chr10:132.7Mb | 1765 | 10 | 132346401 | 133038470 | 4782 | 500 | AD | eGFR | -0.135368 | -0.95165 | 0.55765 | 0.0183245 | 0 | 1 | 0.650523 | 695 | 0.861549093123209 |
| Chr9:76.4Mb | 1570 | 9 | 75785594 | 76970155 | 4680 | 371 | AD | eGFR | -0.159678 | -1 | 0.66882 | 0.0254971 | 0 | 1 | 0.650773 | 696 | 0.861549093123209 |
| Chr14:31.3Mb | 2143 | 14 | 30543662 | 32023462 | 6151 | 465 | AD | eGFR | -0.157422 | -1 | 0.69514 | 0.0247817 | 0 | 1 | 0.650969 | 697 | 0.861549093123209 |
| Chr5:4.8Mb | 905 | 5 | 4445879 | 5125444 | 3571 | 455 | AD | eGFR | 0.110603 | -0.43421 | 0.69461 | 0.0122331 | 0 | 0.52095 | 0.651529 | 698 | 0.861549093123209 |
| Chr1:55.2Mb | 60 | 1 | 54839340 | 55523104 | 3762 | 467 | AD | eGFR | -0.137545 | -1 | 0.56312 | 0.0189186 | 0 | 1 | 0.652906 | 699 | 0.862134818311874 |
| Chr1:7.8Mb | 11 | 1 | 7486181 | 8205854 | 3419 | 445 | AD | eGFR | 0.0750022 | -0.27459 | 0.42721 | 0.00562533 | 0 | 0.19048 | 0.654003 | 700 | 0.86234967 |
| Chr10:10Mb | 1650 | 10 | 9434191 | 10538234 | 4936 | 409 | AD | eGFR | -0.160108 | -1 | 0.70867 | 0.0256346 | 0 | 1 | 0.655946 | 701 | 0.863677828815977 |
| Chr13:102.3Mb | 2115 | 13 | 101942986 | 102735077 | 3804 | 412 | AD | eGFR | -0.115845 | -0.83029 | 0.4775 | 0.01342 | 0 | 0.78246 | 0.659673 | 702 | 0.867347833333333 |

| locus_label | locus | chr | start | stop | n.snps | n.pcs | phen1 | phen2 | rho | rho.lower | rho.upper | r2 | r2.lower | r2.upper | p | rank | FDR |
|---|---|---|---|---|---|---|---|---|---|---|---|---|---|---|---|---|---|
| Chr19:14.4Mb | 2511 | 19 | 13971487 | 14873771 | 3990 | 482 | AD | eGFR | -0.115677 | -0.89226 | 0.50299 | 0.0133812 | 0 | 0.90764 | 0.662872 | 703 | 0.870314162162162 |
| Chr4:138.3Mb | 837 | 4 | 137967770 | 138646228 | 3468 | 371 | AD | eGFR | 0.111591 | -0.4624 | 0.75578 | 0.0124527 | 0 | 0.61423 | 0.665075 | 704 | 0.871966228693182 |
| Chr16:78.5Mb | 2345 | 16 | 78275342 | 78646693 | 3253 | 388 | AD | eGFR | -0.110336 | -0.75897 | 0.46135 | 0.0121741 | 0 | 0.62022 | 0.667094 | 705 | 0.873372712056738 |
| Chr11:40.2Mb | 1811 | 11 | 39700365 | 40717555 | 5198 | 421 | AD | eGFR | -0.119629 | -0.88831 | 0.51749 | 0.0143112 | 0 | 0.88928 | 0.672551 | 706 | 0.877462725988701 |
| Chr9:125.1Mb | 1621 | 9 | 124679660 | 125557549 | 4064 | 451 | AD | eGFR | -0.109107 | -0.74611 | 0.46554 | 0.0119042 | 0 | 0.6188 | 0.672808 | 707 | 0.877462725988701 |
| Chr10:79.2Mb | 1715 | 10 | 78702552 | 79713337 | 4201 | 436 | AD | eGFR | 0.0797361 | -0.32066 | 0.48768 | 0.00635784 | 0 | 0.25544 | 0.67307 | 708 | 0.877462725988701 |
| Chr18:24.6Mb | 2448 | 18 | 24045642 | 25148997 | 4613 | 494 | AD | eGFR | -0.0833162 | -0.52636 | 0.34177 | 0.0069416 | 0 | 0.29226 | 0.677432 | 709 | 0.878925131837307 |
| Chr9:140.6Mb | 1638 | 9 | 140100310 | 141146682 | 3695 | 460 | AD | eGFR | 0.0747248 | -0.31478 | 0.48935 | 0.00558379 | 0 | 0.26289 | 0.677435 | 710 | 0.878925131837307 |
| Chr15:75.2Mb | 2263 | 15 | 74457454 | 75958824 | 4534 | 406 | AD | eGFR | 0.0548411 | -0.22759 | 0.35247 | 0.00300754 | 0 | 0.13416 | 0.677838 | 711 | 0.878925131837307 |
| Chr13:52.4Mb | 2062 | 13 | 51849615 | 52880746 | 3434 | 342 | AD | eGFR | 0.111246 | -0.47698 | 0.79185 | 0.0123756 | 0 | 0.68854 | 0.678646 | 712 | 0.878925131837307 |
| Chr3:29.2Mb | 523 | 3 | 28885831 | 29603546 | 3736 | 464 | AD | eGFR | -0.0727324 | -0.44775 | 0.27935 | 0.00529 | 0 | 0.20974 | 0.678953 | 713 | 0.878925131837307 |
| Chr3:192.4Mb | 679 | 3 | 192057039 | 192760241 | 3486 | 449 | AD | eGFR | -0.0839605 | -0.58167 | 0.3706 | 0.00704936 | 0 | 0.36712 | 0.683586 | 714 | 0.883683302521008 |
| Chr20:46Mb | 2602 | 20 | 45559113 | 46394090 | 4012 | 463 | AD | eGFR | 0.0745034 | -0.30205 | 0.46848 | 0.00555076 | 0 | 0.23043 | 0.685518 | 715 | 0.884941418181818 |
| Chr1:186.9Mb | 171 | 1 | 186267151 | 187522194 | 5356 | 390 | AD | eGFR | -0.109571 | -0.86199 | 0.54143 | 0.0120058 | 0 | 0.83795 | 0.691519 | 716 | 0.891441392458101 |
| Chr15:28.6Mb | 2222 | 15 | 27882172 | 29336984 | 4252 | 437 | AD | eGFR | -0.110502 | -0.87128 | 0.51769 | 0.0122108 | 0 | 0.88273 | 0.693617 | 717 | 0.891566160273973 |
| Chr9:136.2Mb | 1632 | 9 | 135882062 | 136616736 | 4022 | 501 | AD | eGFR | 0.063427 | -0.26631 | 0.40284 | 0.00402298 | 0 | 0.17646 | 0.695224 | 718 | 0.891566160273973 |
| Chr1:115.5Mb | 123 | 1 | 114914644 | 116026337 | 4394 | 366 | AD | eGFR | 0.110125 | -0.5484 | 0.87045 | 0.0121275 | 0 | 0.95764 | 0.697024 | 719 | 0.891566160273973 |
| Chr3:157.2Mb | 643 | 3 | 156791148 | 157636089 | 3871 | 333 | AD | eGFR | 0.0717654 | -0.32019 | 0.48069 | 0.00515027 | 0 | 0.2484 | 0.697377 | 720 | 0.891566160273973 |
| Chr14:27.1Mb | 2139 | 14 | 26673620 | 27530682 | 3395 | 388 | AD | eGFR | 0.070964 | -0.29746 | 0.46296 | 0.00503589 | 0 | 0.22674 | 0.700835 | 721 | 0.891566160273973 |
| Chr13:91.8Mb | 2102 | 13 | 91225981 | 92381082 | 5511 | 454 | AD | eGFR | 0.088643 | -0.41334 | 0.63433 | 0.00785758 | 0 | 0.42865 | 0.701086 | 722 | 0.891566160273973 |
| Chr2:145.7Mb | 390 | 2 | 145124173 | 146305887 | 3518 | 392 | AD | eGFR | 0.0831633 | -0.38729 | 0.61842 | 0.00691613 | 0 | 0.42489 | 0.701137 | 723 | 0.891566160273973 |
| Chr2:68.9Mb | 317 | 2 | 68545963 | 69336307 | 4189 | 409 | AD | eGFR | 0.130265 | -0.71518 | 1 | 0.016969 | 0 | 1 | 0.701193 | 724 | 0.891566160273973 |
| Chr4:26.3Mb | 721 | 4 | 25873929 | 26813071 | 3929 | 460 | AD | eGFR | -0.109552 | -0.87385 | 0.53178 | 0.0120016 | 0 | 0.89987 | 0.701569 | 725 | 0.891566160273973 |
| Chr7:91.4Mb | 1312 | 7 | 90660754 | 92044485 | 4251 | 393 | AD | eGFR | -0.130675 | -1 | 0.6999 | 0.0170759 | 0 | 1 | 0.70238 | 726 | 0.891566160273973 |
| Chr14:42Mb | 2152 | 14 | 41614107 | 42423193 | 3230 | 270 | AD | eGFR | 0.134144 | -0.80703 | 1 | 0.0179946 | 0 | 1 | 0.704436 | 727 | 0.891566160273973 |
| Chr19:48.1Mb | 2543 | 19 | 47544623 | 48556496 | 4410 | 469 | AD | eGFR | 0.0606759 | -0.27333 | 0.3986 | 0.00368157 | 0 | 0.17219 | 0.704561 | 728 | 0.891566160273973 |
| Chr13:80.3Mb | 2089 | 13 | 79809744 | 80783431 | 3462 | 388 | AD | eGFR | 0.0822156 | -0.38719 | 0.58633 | 0.00675941 | 0 | 0.38529 | 0.704806 | 729 | 0.891566160273973 |
| Chr12:106.5Mb | 2000 | 12 | 106098952 | 106981130 | 4229 | 444 | AD | eGFR | -0.0763041 | -0.54021 | 0.34682 | 0.00582232 | 0 | 0.31256 | 0.705139 | 730 | 0.891566160273973 |
| Chr17:68.7Mb | 2416 | 17 | 68107836 | 69244794 | 5491 | 460 | AD | eGFR | 0.115348 | -0.64294 | 0.97539 | 0.0133052 | 0 | 1 | 0.70643 | 731 | 0.89197659370725 |
| Chr9:38.2Mb | 1561 | 9 | 37732940 | 38577096 | 4220 | 449 | AD | eGFR | -0.12678 | -1 | 0.70712 | 0.0160731 | 0 | 1 | 0.708807 | 732 | 0.892610286863271 |
| Chr2:174.2Mb | 417 | 2 | 173775872 | 174714722 | 4092 | 480 | AD | eGFR | -0.100106 | -0.78912 | 0.51193 | 0.0100213 | 0 | 0.70994 | 0.709056 | 733 | 0.892610286863271 |
| Chr8:119.9Mb | 1486 | 8 | 119533404 | 120216587 | 3262 | 339 | AD | eGFR | -0.0962376 | -0.80997 | 0.52716 | 0.00926168 | 0 | 0.80843 | 0.711876 | 734 | 0.892610286863271 |
| Chr3:112.1Mb | 599 | 3 | 111724652 | 112519269 | 3943 | 359 | AD | eGFR | 0.0867459 | -0.43465 | 0.68523 | 0.00752486 | 0 | 0.53953 | 0.711891 | 735 | 0.892610286863271 |

| locus_label | locus | chr | start | stop | n.snps | n.pcs | phen1 | phen2 | rho | rho.lower | rho.upper | r2 | r2.lower | r2.upper | p | rank | FDR |
|---|---|---|---|---|---|---|---|---|---|---|---|---|---|---|---|---|---|
| Chr1:209.4Mb | 194 | 1 | 208973258 | 209781893 | 3619 | 423 | AD | eGFR | -0.0755118 | -0.53647 | 0.35831 | 0.00570203 | 0 | 0.31697 | 0.712182 | 736 | 0.892610286863271 |
| Chr20:52.6Mb | 2609 | 20 | 52170568 | 52973682 | 4224 | 499 | AD | eGFR | -0.059828 | -0.40545 | 0.27702 | 0.00357938 | 0 | 0.17554 | 0.714596 | 737 | 0.892610286863271 |
| Chr3:19.8Mb | 513 | 3 | 19338055 | 20263243 | 3843 | 330 | AD | eGFR | -0.0786689 | -0.56703 | 0.38486 | 0.00618879 | 0 | 0.34695 | 0.714655 | 738 | 0.892610286863271 |
| Chr2:169.7Mb | 412 | 2 | 169177539 | 170137500 | 4662 | 469 | AD | eGFR | -0.0778058 | -0.627 | 0.40349 | 0.00605374 | 0 | 0.43249 | 0.715743 | 739 | 0.892610286863271 |
| Chr2:164.7Mb | 407 | 2 | 164105607 | 165205273 | 4154 | 432 | AD | eGFR | 0.110593 | -0.62066 | 0.95856 | 0.0122308 | 0 | 1 | 0.716956 | 740 | 0.892610286863271 |
| Chr2:238.1Mb | 478 | 2 | 237728427 | 238449459 | 4192 | 449 | AD | eGFR | 0.0815009 | -0.42858 | 0.63892 | 0.0066424 | 0 | 0.468 | 0.717453 | 741 | 0.892610286863271 |
| Chr14:65Mb | 2175 | 14 | 64566625 | 65478132 | 3619 | 361 | AD | eGFR | 0.0676347 | -0.34891 | 0.53904 | 0.00457446 | 0 | 0.31935 | 0.717819 | 742 | 0.892610286863271 |
| Chr8:30.7Mb | 1412 | 8 | 30170367 | 31135129 | 4013 | 377 | AD | eGFR | 0.0657822 | -0.31584 | 0.46693 | 0.00432729 | 0 | 0.22995 | 0.719986 | 743 | 0.892610286863271 |
| Chr2:222Mb | 460 | 2 | 221543870 | 222517963 | 4368 | 479 | AD | eGFR | -0.0824738 | -0.71581 | 0.42868 | 0.00680192 | 0 | 0.59566 | 0.720656 | 744 | 0.892610286863271 |
| Chr14:97.6Mb | 2205 | 14 | 97174236 | 97967917 | 3691 | 463 | AD | eGFR | -0.0538937 | -0.39933 | 0.25288 | 0.00290453 | 0 | 0.16526 | 0.720745 | 745 | 0.892610286863271 |
| Chr8:31.6Mb | 1413 | 8 | 31135130 | 32122456 | 4475 | 362 | AD | eGFR | 0.096834 | -0.53383 | 0.80368 | 0.00937683 | 0 | 0.77454 | 0.721438 | 746 | 0.892610286863271 |
| Chr2:22.9Mb | 267 | 2 | 22427210 | 23361023 | 4017 | 449 | AD | eGFR | 0.0747067 | -0.40811 | 0.59543 | 0.00558109 | 0 | 0.40033 | 0.724707 | 747 | 0.89545456626506 |
| Chr12:105.6Mb | 1999 | 12 | 105143831 | 106098951 | 4258 | 368 | AD | eGFR | 0.100626 | -0.59423 | 0.87909 | 0.0101255 | 0 | 0.92991 | 0.728028 | 748 | 0.896455293193717 |
| Chr6:95.8Mb | 1159 | 6 | 95063172 | 96492580 | 6105 | 399 | AD | eGFR | 0.0980957 | -0.54512 | 0.82063 | 0.00962277 | 0 | 0.76647 | 0.729295 | 749 | 0.896455293193717 |
| Chr8:80.3Mb | 1453 | 8 | 79554455 | 81089812 | 6571 | 448 | AD | eGFR | -0.121129 | -1 | 0.80562 | 0.0146722 | 0 | 1 | 0.731074 | 750 | 0.896455293193717 |
| Chr9:82.7Mb | 1577 | 9 | 82345831 | 83076043 | 3771 | 416 | AD | eGFR | 0.0501583 | -0.25781 | 0.36586 | 0.00251585 | 0 | 0.14345 | 0.732437 | 751 | 0.896455293193717 |
| Chr10:71Mb | 1708 | 10 | 70646651 | 71354385 | 3735 | 418 | AD | eGFR | -0.0757586 | -0.57413 | 0.39721 | 0.00573937 | 0 | 0.36283 | 0.732639 | 752 | 0.896455293193717 |
| Chr13:111.1Mb | 2128 | 13 | 110867692 | 111429289 | 3514 | 491 | AD | eGFR | -0.0750696 | -0.57698 | 0.39585 | 0.00563545 | 0 | 0.36196 | 0.735064 | 753 | 0.896455293193717 |
| Chr2:35.3Mb | 280 | 2 | 34890735 | 35613187 | 3773 | 335 | AD | eGFR | 0.0780457 | -0.42038 | 0.59558 | 0.00609114 | 0 | 0.39257 | 0.735926 | 754 | 0.896455293193717 |
| Chr14:38.2Mb | 2148 | 14 | 37449107 | 38993688 | 6264 | 429 | AD | eGFR | -0.105508 | -0.99255 | 0.63707 | 0.0111318 | 0 | 1 | 0.736465 | 755 | 0.896455293193717 |
| Chr1:220.5Mb | 206 | 1 | 220075136 | 220984131 | 3758 | 349 | AD | eGFR | -0.0661084 | -0.49634 | 0.34753 | 0.00437032 | 0 | 0.26222 | 0.737436 | 756 | 0.896455293193717 |
| Chr4:112.1Mb | 811 | 4 | 111731729 | 112515178 | 3361 | 335 | AD | eGFR | -0.106297 | -1 | 0.65409 | 0.0112991 | 0 | 1 | 0.73745 | 757 | 0.896455293193717 |
| Chr12:132.5Mb | 2026 | 12 | 132011602 | 132950439 | 4008 | 446 | AD | eGFR | 0.0920802 | -0.53476 | 0.80254 | 0.00847877 | 0 | 0.76203 | 0.738691 | 758 | 0.896455293193717 |
| Chr9:77.4Mb | 1571 | 9 | 76970156 | 77862457 | 3529 | 423 | AD | eGFR | -0.103292 | -0.98272 | 0.63728 | 0.0106693 | 0 | 1 | 0.738978 | 759 | 0.896455293193717 |
| Chr2:226.9Mb | 465 | 2 | 226242042 | 227528404 | 5118 | 421 | AD | eGFR | 0.0841623 | -0.48945 | 0.77603 | 0.00708329 | 0 | 0.71228 | 0.740029 | 760 | 0.896455293193717 |
| Chr10:18.2Mb | 1659 | 10 | 17585172 | 18785734 | 4408 | 468 | AD | eGFR | -0.11813 | -1 | 0.79492 | 0.0139546 | 0 | 1 | 0.740707 | 761 | 0.896455293193717 |
| Chr14:77.8Mb | 2187 | 14 | 77430027 | 78122292 | 3625 | 382 | AD | eGFR | 0.126375 | -0.89819 | 1 | 0.0159707 | 0 | 1 | 0.741414 | 762 | 0.896455293193717 |
| Chr10:98.5Mb | 1733 | 10 | 97986658 | 98958794 | 4025 | 473 | AD | eGFR | -0.0478097 | -0.34614 | 0.25008 | 0.00228577 | 0 | 0.12948 | 0.741527 | 763 | 0.896455293193717 |
| Chr5:153.7Mb | 1036 | 5 | 153053340 | 154445638 | 5721 | 417 | AD | eGFR | -0.0955027 | -0.8701 | 0.57194 | 0.00912076 | 0 | 0.90727 | 0.742028 | 764 | 0.896455293193717 |
| Chr9:3.9Mb | 1517 | 9 | 3565582 | 4307955 | 4035 | 513 | AD | eGFR | 0.0460451 | -0.24183 | 0.34016 | 0.00212015 | 0 | 0.12537 | 0.744211 | 765 | 0.897760101694915 |
| Chr19:54.3Mb | 2551 | 19 | 54042397 | 54533107 | 2965 | 527 | AD | eGFR | 0.0843089 | -0.58058 | 0.7773 | 0.00710799 | 0 | 0.77544 | 0.745776 | 766 | 0.897760101694915 |
| Chr3:65.8Mb | 558 | 3 | 65403327 | 66162873 | 3970 | 453 | AD | eGFR | -0.0850574 | -0.7524 | 0.50652 | 0.00723475 | 0 | 0.62568 | 0.746026 | 767 | 0.897760101694915 |
| Chr13:69.7Mb | 2078 | 13 | 69404685 | 70079764 | 3708 | 272 | AD | eGFR | 0.100869 | -0.68966 | 0.92146 | 0.0101745 | 0 | 1 | 0.747156 | 768 | 0.897949203125 |
| Chr15:21.4Mb | 2215 | 15 | 20000075 | 22881863 | 135 | 36 | AD | eGFR | 0.0935981 | -0.56167 | 0.79132 | 0.00876061 | 0 | 0.71037 | 0.748968 | 769 | 0.898956390117035 |

| locus_label | locus | chr | start | stop | n.snps | n.pcs | phen1 | phen2 | rho | rho.lower | rho.upper | r2 | r2.lower | r2.upper | p | rank | FDR |
|---|---|---|---|---|---|---|---|---|---|---|---|---|---|---|---|---|---|
| Chr10:24.3Mb | 1665 | 10 | 23839538 | 24846617 | 4966 | 467 | AD | eGFR | -0.100502 | -1 | 0.64052 | 0.0101006 | 0 | 1 | 0.750402 | 770 | 0.899507851948052 |
| Chr10:44.5Mb | 1683 | 10 | 44116079 | 44852810 | 4130 | 288 | AD | eGFR | -0.110981 | -1 | 0.79837 | 0.0123168 | 0 | 1 | 0.754449 | 771 | 0.903038331606218 |
| Chr12:7Mb | 1906 | 12 | 6420422 | 7525799 | 2154 | 405 | AD | eGFR | 0.0761346 | -0.4544 | 0.67865 | 0.00579647 | 0 | 0.51254 | 0.755304 | 772 | 0.903038331606218 |
| Chr4:146Mb | 846 | 4 | 145233056 | 146820830 | 5394 | 411 | AD | eGFR | 0.0982866 | -0.69174 | 0.99206 | 0.00966025 | 0 | 1 | 0.756734 | 773 | 0.903577596377749 |
| Chr11:73.5Mb | 1839 | 11 | 72908824 | 74156571 | 4998 | 357 | AD | eGFR | 0.0546389 | -0.32117 | 0.43311 | 0.00298541 | 0 | 0.21114 | 0.759695 | 774 | 0.905941195090439 |
| Chr16:89.3Mb | 2362 | 16 | 89045299 | 89642476 | 3229 | 374 | AD | eGFR | -0.085475 | -0.83527 | 0.5899 | 0.00730598 | 0 | 0.86613 | 0.761101 | 775 | 0.906446739354839 |
| Chr7:92.7Mb | 1313 | 7 | 92044486 | 93338416 | 4186 | 405 | AD | eGFR | -0.106071 | -1 | 0.79798 | 0.0112511 | 0 | 1 | 0.762937 | 776 | 0.90746243685567 |
| Chr1:148.3Mb | 132 | 1 | 147164394 | 149421694 | 1278 | 247 | AD | eGFR | -0.126121 | -1 | 1 | 0.0159064 | 0 | 1 | 0.76554 | 777 | 0.909386640926641 |
| Chr2:40.7Mb | 287 | 2 | 40281047 | 41199635 | 4496 | 416 | AD | eGFR | 0.0881448 | -0.63662 | 0.91941 | 0.00776951 | 0 | 1 | 0.768191 | 778 | 0.911221208121827 |
| Chr11:4Mb | 1773 | 11 | 3579508 | 4402586 | 3472 | 403 | AD | eGFR | -0.0549064 | -0.47277 | 0.34206 | 0.00301472 | 0 | 0.24479 | 0.769226 | 779 | 0.911221208121827 |
| Chr1:42.8Mb | 49 | 1 | 42358761 | 43299580 | 3969 | 300 | AD | eGFR | -0.10048 | -1 | 0.78783 | 0.0100963 | 0 | 1 | 0.770223 | 780 | 0.911221208121827 |
| Chr13:73.5Mb | 2083 | 13 | 73075521 | 73933729 | 3745 | 386 | AD | eGFR | 0.10906 | -0.90981 | 1 | 0.0118941 | 0 | 1 | 0.771881 | 781 | 0.911221208121827 |
| Chr3:187.3Mb | 673 | 3 | 186894705 | 187626850 | 3331 | 419 | AD | eGFR | 0.0727656 | -0.50189 | 0.70251 | 0.00529484 | 0 | 0.59713 | 0.77318 | 782 | 0.911221208121827 |
| Chr12:74Mb | 1971 | 12 | 73446368 | 74577925 | 4971 | 245 | AD | eGFR | 0.117839 | -1 | 1 | 0.0138861 | 0 | 1 | 0.77394 | 783 | 0.911221208121827 |
| Chr13:104.5Mb | 2118 | 13 | 104217312 | 104842810 | 3530 | 386 | AD | eGFR | 0.0548055 | -0.37221 | 0.49692 | 0.00300364 | 0 | 0.29234 | 0.776256 | 784 | 0.911221208121827 |
| Chr3:13.4Mb | 506 | 3 | 13074428 | 13739919 | 3626 | 451 | AD | eGFR | 0.106629 | -0.86805 | 1 | 0.0113696 | 0 | 1 | 0.776438 | 785 | 0.911221208121827 |
| Chr4:84.1Mb | 782 | 4 | 83680840 | 84465633 | 3315 | 397 | AD | eGFR | -0.100468 | -1 | 0.80859 | 0.0100938 | 0 | 1 | 0.776899 | 786 | 0.911221208121827 |
| Chr10:69.2Mb | 1706 | 10 | 68571604 | 69892269 | 5644 | 387 | AD | eGFR | 0.0703804 | -0.47829 | 0.6687 | 0.0049534 | 0 | 0.51869 | 0.777445 | 787 | 0.911221208121827 |
| Chr6:36.8Mb | 1103 | 6 | 36230132 | 37363762 | 5661 | 491 | AD | eGFR | 0.0665619 | -0.45066 | 0.62002 | 0.00443048 | 0 | 0.43544 | 0.777944 | 788 | 0.911221208121827 |
| Chr12:14.2Mb | 1914 | 12 | 13652724 | 14661770 | 4705 | 423 | AD | eGFR | 0.0429156 | -0.26808 | 0.35309 | 0.00184175 | 0 | 0.13899 | 0.779861 | 789 | 0.912308875792142 |
| Chr19:33.1Mb | 2529 | 19 | 32576922 | 33640645 | 5221 | 391 | AD | eGFR | 0.0790474 | -0.60686 | 0.83564 | 0.00624848 | 0 | 0.86297 | 0.781399 | 790 | 0.912950983544304 |
| Chr3:124.2Mb | 610 | 3 | 123412341 | 124921712 | 6633 | 484 | AD | eGFR | 0.0648667 | -0.46706 | 0.65076 | 0.00420769 | 0 | 0.49791 | 0.784478 | 791 | 0.914798122653317 |
| Chr5:131.7Mb | 1019 | 5 | 131312792 | 132135840 | 3329 | 328 | AD | eGFR | 0.0562533 | -0.40366 | 0.52073 | 0.00316444 | 0 | 0.30829 | 0.787289 | 792 | 0.914798122653317 |
| Chr13:34.1Mb | 2045 | 13 | 33508764 | 34683828 | 5884 | 496 | AD | eGFR | 0.0814758 | -0.6503 | 0.89277 | 0.00663831 | 0 | 0.96433 | 0.787792 | 793 | 0.914798122653317 |
| Chr3:145Mb | 629 | 3 | 144703588 | 145353050 | 3209 | 262 | AD | eGFR | 0.0914446 | -0.76742 | 1 | 0.00836211 | 0 | 1 | 0.789626 | 794 | 0.914798122653317 |
| Chr2:130.3Mb | 373 | 2 | 129915502 | 130662352 | 3939 | 392 | AD | eGFR | 0.0768114 | -0.60291 | 0.81937 | 0.00589999 | 0 | 0.84114 | 0.789643 | 795 | 0.914798122653317 |
| Chr15:34.1Mb | 2226 | 15 | 33540411 | 34647815 | 6224 | 518 | AD | eGFR | -0.0478661 | -0.42517 | 0.31838 | 0.00229117 | 0 | 0.20197 | 0.79006 | 796 | 0.914798122653317 |
| Chr1:181.4Mb | 165 | 1 | 181016516 | 181765046 | 3381 | 435 | AD | eGFR | -0.0792722 | -0.8726 | 0.62488 | 0.00628408 | 0 | 0.93475 | 0.791067 | 797 | 0.914798122653317 |
| Chr1:39.1Mb | 45 | 1 | 38607054 | 39544839 | 4088 | 479 | AD | eGFR | -0.0462315 | -0.42935 | 0.3271 | 0.00213736 | 0 | 0.20717 | 0.791361 | 798 | 0.914798122653317 |
| Chr7:51.4Mb | 1278 | 7 | 50906576 | 51953911 | 5122 | 437 | AD | eGFR | -0.0618764 | -0.65249 | 0.47365 | 0.00382869 | 0 | 0.54058 | 0.7919 | 799 | 0.914798122653317 |
| Chr6:59.6Mb | 1123 | 6 | 57316571 | 61883554 | 4059 | 143 | AD | eGFR | 0.107035 | -0.99429 | 1 | 0.0114564 | 0 | 1 | 0.793282 | 800 | 0.9152491075 |
| Chr9:86.4Mb | 1581 | 9 | 85940304 | 86922195 | 4267 | 405 | AD | eGFR | -0.0722825 | -0.81361 | 0.59532 | 0.00522476 | 0 | 0.79712 | 0.798548 | 801 | 0.917463142326733 |
| Chr9:72.1Mb | 1566 | 9 | 71590807 | 72513621 | 4011 | 396 | AD | eGFR | -0.0716097 | -0.80095 | 0.57149 | 0.00512795 | 0 | 0.76487 | 0.799135 | 802 | 0.917463142326733 |
| Chr10:112.8Mb | 1745 | 10 | 112378763 | 113304188 | 4424 | 411 | AD | eGFR | -0.0880792 | -1 | 0.7841 | 0.00775795 | 0 | 1 | 0.799378 | 803 | 0.917463142326733 |
| Chr2:125.3Mb | 367 | 2 | 124903614 | 125653808 | 3434 | 385 | AD | eGFR | -0.0520539 | -0.52119 | 0.38212 | 0.0027096 | 0 | 0.3074 | 0.800052 | 804 | 0.917463142326733 |

| locus_label | locus | chr | start | stop | n.snps | n.pcs | phen1 | phen2 | rho | rho.lower | rho.upper | r2 | r2.lower | r2.upper | p | rank | FDR |
|---|---|---|---|---|---|---|---|---|---|---|---|---|---|---|---|---|---|
| Chr8:27.1Mb | 1408 | 8 | 26678702 | 27425267 | 3903 | 336 | AD | eGFR | 0.0540651 | -0.43563 | 0.58387 | 0.00292303 | 0 | 0.43111 | 0.801769 | 805 | 0.917463142326733 |
| Chr11:118.4Mb | 1882 | 11 | 117904061 | 118958129 | 4230 | 411 | AD | eGFR | 0.0607332 | -0.48411 | 0.63809 | 0.00368852 | 0 | 0.46533 | 0.802198 | 806 | 0.917463142326733 |
| Chr2:173.4Mb | 416 | 2 | 172931483 | 173775871 | 4046 | 431 | AD | eGFR | -0.0624564 | -0.69101 | 0.50096 | 0.00390081 | 0 | 0.57476 | 0.802997 | 807 | 0.917463142326733 |
| Chr10:96.1Mb | 1731 | 10 | 95515829 | 96756082 | 5474 | 309 | AD | eGFR | 0.0802689 | -0.67851 | 0.98248 | 0.00644309 | 0 | 1 | 0.803153 | 808 | 0.917463142326733 |
| Chr10:118Mb | 1750 | 10 | 117641749 | 118415956 | 3622 | 428 | AD | eGFR | 0.0603694 | -0.49905 | 0.65585 | 0.00364447 | 0 | 0.51731 | 0.806965 | 809 | 0.920678238566131 |
| Chr2:53.5Mb | 302 | 2 | 53143665 | 53831584 | 4171 | 401 | AD | eGFR | 0.0930004 | -0.97049 | 1 | 0.00864908 | 0 | 1 | 0.810831 | 810 | 0.921326850673195 |
| Chr20:48.7Mb | 2605 | 20 | 48212708 | 49238193 | 4631 | 468 | AD | eGFR | 0.0400446 | -0.30774 | 0.38829 | 0.00160357 | 0 | 0.17406 | 0.81147 | 811 | 0.921326850673195 |
| Chr2:104.4Mb | 347 | 2 | 103781211 | 105116176 | 5112 | 403 | AD | eGFR | -0.0711281 | -0.84956 | 0.66065 | 0.0050592 | 0 | 0.9212 | 0.81245 | 812 | 0.921326850673195 |
| Chr4:36.4Mb | 734 | 4 | 36025562 | 36734792 | 3363 | 331 | AD | eGFR | -0.0568105 | -0.61809 | 0.47095 | 0.00322743 | 0 | 0.45418 | 0.812855 | 813 | 0.921326850673195 |
| Chr2:205.3Mb | 442 | 2 | 204817774 | 205764869 | 4129 | 416 | AD | eGFR | 0.084959 | -0.88658 | 1 | 0.00721804 | 0 | 1 | 0.813483 | 814 | 0.921326850673195 |
| Chr6:164.1Mb | 1219 | 6 | 163615966 | 164591970 | 5323 | 503 | AD | eGFR | -0.0707868 | -0.90846 | 0.68234 | 0.00501077 | 0 | 1 | 0.814806 | 815 | 0.921326850673195 |
| Chr1:3.1Mb | 4 | 1 | 2772098 | 3352199 | 3417 | 497 | AD | eGFR | 0.0517018 | -0.48255 | 0.58572 | 0.00267307 | 0 | 0.44004 | 0.814921 | 816 | 0.921326850673195 |
| Chr1:1.7Mb | 2 | 1 | 1359455 | 1966855 | 1699 | 366 | AD | eGFR | 0.0628994 | -0.54677 | 0.72782 | 0.00395634 | 0 | 0.6209 | 0.815519 | 817 | 0.921326850673195 |
| Chr4:141.5Mb | 841 | 4 | 141086488 | 141996946 | 3566 | 433 | AD | eGFR | -0.0483613 | -0.50891 | 0.38051 | 0.00233881 | 0 | 0.28677 | 0.81655 | 818 | 0.921363875305623 |
| Chr2:114.5Mb | 356 | 2 | 113872057 | 115204694 | 4864 | 427 | AD | eGFR | -0.0449841 | -0.47344 | 0.36168 | 0.00202357 | 0 | 0.25641 | 0.818234 | 819 | 0.92213673015873 |
| Chr12:81.8Mb | 1978 | 12 | 81134129 | 82502068 | 5266 | 371 | AD | eGFR | 0.0417008 | -0.41103 | 0.53772 | 0.00173895 | 0 | 0.38159 | 0.822183 | 820 | 0.925146171741778 |
| Chr16:51.9Mb | 2322 | 16 | 51277677 | 52507564 | 5778 | 496 | AD | eGFR | -0.0499666 | -0.55429 | 0.43402 | 0.00249666 | 0 | 0.35298 | 0.822909 | 821 | 0.925146171741778 |
| Chr19:50.7Mb | 2546 | 19 | 50231620 | 51113407 | 4196 | 469 | AD | eGFR | -0.0493386 | -0.62571 | 0.47608 | 0.0024343 | 0 | 0.47879 | 0.830466 | 822 | 0.93248203276699 |
| Chr1:47.7Mb | 54 | 1 | 47257677 | 48200179 | 3827 | 398 | AD | eGFR | -0.0466746 | -0.556 | 0.43803 | 0.00217851 | 0 | 0.3582 | 0.831846 | 823 | 0.93248203276699 |
| Chr10:20Mb | 1661 | 10 | 19574983 | 20504210 | 4914 | 406 | AD | eGFR | -0.0651366 | -0.90872 | 0.70584 | 0.00424278 | 0 | 1 | 0.832465 | 824 | 0.93248203276699 |
| Chr14:75.9Mb | 2185 | 14 | 75021275 | 76780225 | 6820 | 511 | AD | eGFR | -0.0466687 | -0.63036 | 0.46825 | 0.00217797 | 0 | 0.46891 | 0.836051 | 825 | 0.935363724848485 |
| Chr12:16.3Mb | 1916 | 12 | 15816043 | 16864533 | 3770 | 432 | AD | eGFR | 0.0336158 | -0.31723 | 0.38632 | 0.00113002 | 0 | 0.17078 | 0.841029 | 826 | 0.937213978391357 |
| Chr1:161.1Mb | 144 | 1 | 160687164 | 161567517 | 3287 | 373 | AD | eGFR | 0.0742266 | -0.95374 | 1 | 0.00550958 | 0 | 1 | 0.841197 | 827 | 0.937213978391357 |
| Chr12:72.7Mb | 1970 | 12 | 71948456 | 73446367 | 6090 | 413 | AD | eGFR | -0.0408463 | -0.51624 | 0.41176 | 0.00166842 | 0 | 0.31085 | 0.841878 | 828 | 0.937213978391357 |
| Chr10:34Mb | 1676 | 10 | 33452058 | 34533510 | 5446 | 507 | AD | eGFR | -0.0426511 | -0.5727 | 0.44113 | 0.00181912 | 0 | 0.41198 | 0.843656 | 829 | 0.937213978391357 |
| Chr1:22.6Mb | 29 | 1 | 22147134 | 22969973 | 4113 | 437 | AD | eGFR | 0.0653077 | -0.80707 | 0.99282 | 0.0042651 | 0 | 1 | 0.844338 | 830 | 0.937213978391357 |
| Chr3:38.1Mb | 533 | 3 | 37535684 | 38604836 | 4082 | 394 | AD | eGFR | 0.0653392 | -0.74653 | 0.99629 | 0.00426921 | 0 | 1 | 0.844738 | 831 | 0.937213978391357 |
| Chr12:47.9Mb | 1948 | 12 | 47508148 | 48287540 | 3848 | 434 | AD | eGFR | 0.0552533 | -0.66987 | 0.79594 | 0.00305292 | 0 | 0.94871 | 0.845198 | 832 | 0.937213978391357 |
| Chr10:17.2Mb | 1658 | 10 | 16721534 | 17585171 | 4990 | 452 | AD | eGFR | 0.0338196 | -0.32923 | 0.41123 | 0.00114376 | 0 | 0.1909 | 0.845828 | 833 | 0.937213978391357 |
| Chr10:65.2Mb | 1702 | 10 | 64758501 | 65737423 | 3894 | 318 | AD | eGFR | 0.0621683 | -0.77613 | 0.95528 | 0.0038649 | 0 | 1 | 0.848813 | 834 | 0.939352022754491 |
| Chr3:121.5Mb | 608 | 3 | 120964888 | 121971906 | 3562 | 302 | AD | eGFR | 0.0523885 | -0.62015 | 0.7354 | 0.00274456 | 0 | 0.70751 | 0.849793 | 835 | 0.939352022754491 |
| Chr15:73.9Mb | 2262 | 15 | 73349296 | 74457453 | 4245 | 376 | AD | eGFR | 0.0603166 | -0.80578 | 0.99396 | 0.00363809 | 0 | 1 | 0.856066 | 836 | 0.943832175771971 |
| Chr2:106.5Mb | 349 | 2 | 106078267 | 107001332 | 4684 | 437 | AD | eGFR | -0.0482766 | -0.74624 | 0.58327 | 0.00233063 | 0 | 0.69251 | 0.857299 | 837 | 0.943832175771971 |
| Chr8:59.8Mb | 1435 | 8 | 59077704 | 60478317 | 6131 | 462 | AD | eGFR | 0.032606 | -0.35361 | 0.42855 | 0.00106315 | 0 | 0.21666 | 0.857864 | 838 | 0.943832175771971 |
| Chr4:185.4Mb | 890 | 4 | 18508891 | 18574015 | 3449 | 464 | AD | eGFR | 0.0504356 | -0.63195 | 0.76372 | 0.00254375 | 0 | 0.73933 | 0.858143 | 839 | 0.943832175771971 |

| locus_label | locus | chr | start | stop | n.snps | n.pcs | phen1 | phen2 | rho | rho.lower | rho.upper | r2 | r2.lower | r2.upper | p | rank | FDR |
|---|---|---|---|---|---|---|---|---|---|---|---|---|---|---|---|---|---|
| | | | 9 | 6 | | | | | | | | | | | | | |
| Chr8:41.4Mb | 1421 | 8 | 40978027 | 41726919 | 3964 | 405 | AD | eGFR | 0.0486124 | -0.60311 | 0.73359 | 0.00236317 | 0 | 0.66695 | 0.859035 | 840 | 0.943832175771971 |
| Chr7:12.6Mb | 1240 | 7 | 12173114 | 13033825 | 5854 | 405 | AD | eGFR | -0.0567598 | -1 | 0.74494 | 0.00322168 | 0 | 1 | 0.860998 | 841 | 0.943832175771971 |
| Chr4:23Mb | 717 | 4 | 22525040 | 23487858 | 4407 | 401 | AD | eGFR | 0.0385173 | -0.44856 | 0.5672 | 0.00148358 | 0 | 0.39652 | 0.861004 | 842 | 0.943832175771971 |
| Chr9:91.8Mb | 1587 | 9 | 91332984 | 92288101 | 4797 | 440 | AD | eGFR | -0.0597739 | -1 | 0.82161 | 0.00357291 | 0 | 1 | 0.863963 | 843 | 0.945952371293001 |
| Chr13:74.6Mb | 2084 | 13 | 73933730 | 75304979 | 6666 | 533 | AD | eGFR | -0.0451717 | -0.75672 | 0.58801 | 0.00204048 | 0 | 0.72192 | 0.866914 | 844 | 0.947683158392435 |
| Chr9:102.6Mb | 1597 | 9 | 101860664 | 103364049 | 4118 | 335 | AD | eGFR | -0.0493812 | -0.8395 | 0.67856 | 0.0024385 | 0 | 0.97705 | 0.867605 | 845 | 0.947683158392435 |
| Chr2:192.8Mb | 433 | 2 | 192331626 | 193234273 | 3472 | 474 | AD | eGFR | -0.0442418 | -0.72416 | 0.57267 | 0.00195734 | 0 | 0.69541 | 0.868624 | 846 | 0.947683158392435 |
| Chr18:25.6Mb | 2449 | 18 | 25148998 | 26014629 | 3292 | 406 | AD | eGFR | 0.033442 | -0.42297 | 0.48523 | 0.00111837 | 0 | 0.29639 | 0.872301 | 847 | 0.950571219598583 |
| Chr1:94Mb | 103 | 1 | 93466126 | 94479821 | 3794 | 325 | AD | eGFR | -0.0321054 | -0.48408 | 0.40495 | 0.00103076 | 0 | 0.2828 | 0.875959 | 848 | 0.953431788915094 |
| Chr5:126.3Mb | 1015 | 5 | 125677793 | 126992654 | 6001 | 472 | AD | eGFR | 0.0301856 | -0.41498 | 0.50004 | 0.000911172 | 0 | 0.31294 | 0.877495 | 849 | 0.953978663133098 |
| Chr4:189.6Mb | 897 | 4 | 189397151 | 189893888 | 3437 | 372 | AD | eGFR | -0.0525568 | -1 | 0.86697 | 0.00276221 | 0 | 1 | 0.880459 | 850 | 0.954352784290739 |
| Chr15:45.5Mb | 2236 | 15 | 45002687 | 46002546 | 3973 | 371 | AD | eGFR | 0.0174835 | -0.23062 | 0.26487 | 0.000305674 | 0 | 0.08484 | 0.881115 | 851 | 0.954352784290739 |
| Chr16:10.7Mb | 2301 | 16 | 10434118 | 10987261 | 3413 | 473 | AD | eGFR | -0.0581382 | -1 | 1 | 0.00338005 | 0 | 1 | 0.881891 | 852 | 0.954352784290739 |
| Chr1:238.9Mb | 226 | 1 | 238440737 | 239327076 | 4871 | 445 | AD | eGFR | 0.0349785 | -0.49911 | 0.62905 | 0.00122349 | 0 | 0.52661 | 0.881975 | 853 | 0.954352784290739 |
| Chr2:77Mb | 326 | 2 | 76650969 | 77373927 | 4148 | 364 | AD | eGFR | 0.0290961 | -0.38985 | 0.4646 | 0.000846583 | 0 | 0.25948 | 0.885879 | 854 | 0.957079750877193 |
| Chr14:92.8Mb | 2200 | 14 | 92141003 | 93388064 | 6694 | 512 | AD | eGFR | -0.0386903 | -0.74553 | 0.61348 | 0.00149694 | 0 | 0.73725 | 0.886569 | 855 | 0.957079750877193 |
| Chr1:214.8Mb | 199 | 1 | 214406341 | 215126936 | 3628 | 376 | AD | eGFR | -0.0359951 | -0.69244 | 0.61035 | 0.00129565 | 0 | 0.65629 | 0.89188 | 856 | 0.960656676779463 |
| Chr16:7.9Mb | 2297 | 16 | 7527654 | 8309201 | 5781 | 495 | AD | eGFR | 0.0469464 | -0.85426 | 1 | 0.00220396 | 0 | 1 | 0.891964 | 857 | 0.960656676779463 |
| Chr12:0.6Mb | 1899 | 12 | 60181 | 1073030 | 3684 | 486 | AD | eGFR | -0.0306431 | -0.56794 | 0.4785 | 0.000939002 | 0 | 0.40168 | 0.894439 | 858 | 0.96219953030303 |
| Chr19:13.3Mb | 2510 | 19 | 12638785 | 13971486 | 5473 | 524 | AD | eGFR | -0.0326066 | -0.62133 | 0.53468 | 0.00106319 | 0 | 0.4839 | 0.896608 | 859 | 0.962494740697674 |
| Chr7:95.4Mb | 1315 | 7 | 94741237 | 96077028 | 5393 | 460 | AD | eGFR | -0.0299744 | -0.60568 | 0.52343 | 0.000898463 | 0 | 0.51339 | 0.896799 | 860 | 0.962494740697674 |
| Chr1:205.4Mb | 190 | 1 | 204922956 | 205811877 | 3882 | 463 | AD | eGFR | -0.0355523 | -0.74423 | 0.64659 | 0.00126397 | 0 | 0.75861 | 0.897915 | 861 | 0.962573222996516 |
| Chr13:33Mb | 2044 | 13 | 32542671 | 33508763 | 4052 | 403 | AD | eGFR | -0.0461727 | -1 | 0.967 | 0.00213191 | 0 | 1 | 0.900441 | 862 | 0.963676097222222 |
| Chr6:33.8Mb | 1100 | 6 | 33464011 | 34111032 | 4111 | 429 | AD | eGFR | -0.0142679 | -0.24879 | 0.2142 | 0.000203572 | 0 | 0.07143 | 0.901587 | 863 | 0.963676097222222 |
| Chr5:62.9Mb | 958 | 5 | 62539894 | 63229491 | 3259 | 268 | AD | eGFR | -0.0354892 | -0.76736 | 0.66417 | 0.00125948 | 0 | 0.79886 | 0.902076 | 864 | 0.963676097222222 |
| Chr2:77.7Mb | 327 | 2 | 77373928 | 78080661 | 3508 | 429 | AD | eGFR | 0.0239594 | -0.3934 | 0.46936 | 0.000574053 | 0 | 0.26401 | 0.904577 | 865 | 0.964206041522491 |
| Chr12:94.7Mb | 1989 | 12 | 94319630 | 95152978 | 3987 | 485 | AD | eGFR | 0.0228442 | -0.40684 | 0.44956 | 0.000521855 | 0 | 0.26281 | 0.904805 | 866 | 0.964206041522491 |
| Chr19:57.6Mb | 2556 | 19 | 57263494 | 57959397 | 3454 | 480 | AD | eGFR | 0.033554 | -0.70289 | 0.75673 | 0.00112587 | 0 | 0.87674 | 0.905706 | 867 | 0.964206041522491 |
| Chr6:78.3Mb | 1142 | 6 | 77941856 | 78719821 | 3852 | 257 | AD | eGFR | -0.0270374 | -0.55726 | 0.48522 | 0.00073102 | 0 | 0.40371 | 0.908166 | 868 | 0.965711080645161 |
| Chr22:23.7Mb | 2661 | 22 | 23216593 | 24188351 | 5309 | 506 | AD | eGFR | 0.027538 | -0.54773 | 0.61075 | 0.000758343 | 0 | 0.51057 | 0.91047 | 869 | 0.967046962025316 |
| Chr12:42Mb | 1941 | 12 | 41694309 | 42354236 | 3421 | 304 | AD | eGFR | -0.0278293 | -0.64548 | 0.56022 | 0.00077446 | 0 | 0.5325 | 0.913134 | 870 | 0.968252403669725 |

| locus_label | locus | chr | start | stop | n.snps | n.pcs | phen1 | phen2 | rho | rho.lower | rho.upper | r2 | r2.lower | r2.upper | p | rank | FDR |
|---|---|---|---|---|---|---|---|---|---|---|---|---|---|---|---|---|---|
| | | | | | | | | | | | | 9 | | | | | |
| Chr1:18.5Mb | 24 | 1 | 18190729 | 18906906 | 3918 | 508 | AD | eGFR | -0.0240311 | -0.58935 | 0.46903 | 0.000577494 | 0 | 0.44973 | 0.913891 | 871 | 0.968252403669725 |
| Chr3:131.7Mb | 618 | 3 | 131372701 | 132061683 | 3776 | 357 | AD | eGFR | -0.0341502 | -0.93067 | 0.7768 | 0.00116623 | 0 | 1 | 0.914752 | 872 | 0.968252403669725 |
| Chr5:64.7Mb | 960 | 5 | 64235496 | 65134948 | 3537 | 336 | AD | eGFR | -0.0272325 | -0.64459 | 0.60477 | 0.000741611 | 0 | 0.57038 | 0.918043 | 873 | 0.970622782359679 |
| Chr1:79.1Mb | 87 | 1 | 78719335 | 79452644 | 3497 | 334 | AD | eGFR | -0.0312326 | -0.8415 | 0.75248 | 0.000975472 | 0 | 1 | 0.919115 | 874 | 0.970644330663616 |
| Chr6:127.3Mb | 1184 | 6 | 126258608 | 128335094 | 6525 | 430 | AD | eGFR | 0.0209988 | -0.46916 | 0.53845 | 0.00044095 | 0 | 0.40621 | 0.920615 | 875 | 0.971117308571429 |
| Chr4:42.6Mb | 741 | 4 | 42225151 | 42926067 | 3788 | 366 | AD | eGFR | 0.0359187 | -0.99043 | 1 | 0.00129015 | 0 | 1 | 0.922187 | 876 | 0.971665069634703 |
| Chr9:138.8Mb | 1636 | 9 | 138451427 | 139222877 | 3975 | 480 | AD | eGFR | -0.0218007 | -0.54026 | 0.47874 | 0.000475269 | 0 | 0.37937 | 0.923934 | 877 | 0.971694797956867 |
| Chr10:100.8Mb | 1735 | 10 | 100245232 | 101355648 | 4248 | 344 | AD | eGFR | 0.0199963 | -0.43924 | 0.48693 | 0.000399852 | 0 | 0.29892 | 0.92519 | 878 | 0.971694797956867 |
| Chr7:6.4Mb | 1233 | 7 | 5846044 | 6975014 | 4961 | 447 | AD | eGFR | 0.0329058 | -0.93833 | 1 | 0.00108279 | 0 | 1 | 0.925713 | 879 | 0.971694797956867 |
| Chr11:127.8Mb | 1891 | 11 | 127312463 | 128375175 | 4814 | 451 | AD | eGFR | -0.0170989 | -0.41955 | 0.35785 | 0.000292371 | 0 | 0.20313 | 0.926536 | 880 | 0.971694797956867 |
| Chr6:22.5Mb | 1080 | 6 | 22193378 | 22882787 | 3548 | 384 | AD | eGFR | -0.0173125 | -0.44866 | 0.41247 | 0.000299722 | 0 | 0.26517 | 0.927479 | 881 | 0.971694797956867 |
| Chr4:80.8Mb | 779 | 4 | 80395885 | 81212223 | 3585 | 378 | AD | eGFR | -0.0255737 | -0.76852 | 0.70393 | 0.000654016 | 0 | 0.8504 | 0.930123 | 882 | 0.973360010204082 |
| Chr10:109.8Mb | 1742 | 10 | 109355908 | 110322098 | 4934 | 378 | AD | eGFR | -0.0201832 | -0.56496 | 0.50765 | 0.000407362 | 0 | 0.40713 | 0.931444 | 883 | 0.973638518686297 |
| Chr3:57.2Mb | 549 | 3 | 56433532 | 58056480 | 6671 | 429 | AD | eGFR | -0.0239046 | -0.79029 | 0.69062 | 0.000571432 | 0 | 0.90288 | 0.934747 | 884 | 0.975985838235294 |
| Chr1:65.2Mb | 72 | 1 | 64619458 | 65690453 | 4321 | 438 | AD | eGFR | 0.0165564 | -0.44387 | 0.48929 | 0.000274115 | 0 | 0.3026 | 0.937183 | 885 | 0.977423625988701 |
| Chr12:126.6Mb | 2018 | 12 | 126289544 | 126947707 | 4011 | 442 | AD | eGFR | -0.0136376 | -0.44888 | 0.40015 | 0.000185984 | 0 | 0.2553 | 0.945702 | 886 | 0.983222901234568 |
| Chr7:109.9Mb | 1328 | 7 | 109295964 | 110485679 | 5303 | 444 | AD | eGFR | -0.0213508 | -0.87226 | 0.77878 | 0.000455859 | 0 | 1 | 0.945887 | 887 | 0.983222901234568 |
| Chr9:29.7Mb | 1552 | 9 | 29331379 | 30077281 | 4359 | 337 | AD | eGFR | -0.0170396 | -0.61767 | 0.57697 | 0.000290349 | 0 | 0.5065 | 0.946623 | 888 | 0.983222901234568 |
| Chr1:95.3Mb | 104 | 1 | 94479822 | 96153036 | 6668 | 498 | AD | eGFR | 0.0193042 | -0.70592 | 0.80835 | 0.000372652 | 0 | 0.8941 | 0.948146 | 889 | 0.983222901234568 |
| Chr8:58.5Mb | 1434 | 8 | 57906780 | 59077703 | 6368 | 443 | AD | eGFR | -0.0145738 | -0.58086 | 0.51501 | 0.000212396 | 0 | 0.48251 | 0.948593 | 890 | 0.983222901234568 |
| Chr10:91.4Mb | 1727 | 10 | 90905622 | 91959168 | 4675 | 415 | AD | eGFR | 0.0229119 | -1 | 1 | 0.000524956 | 0 | 1 | 0.949135 | 891 | 0.983222901234568 |
| Chr3:76.3Mb | 568 | 3 | 75676787 | 76921061 | 4775 | 422 | AD | eGFR | -0.0217645 | -1 | 1 | 0.000473694 | 0 | 1 | 0.95208 | 892 | 0.98516798206278 |
| Chr10:84.9Mb | 1721 | 10 | 84387257 | 85423415 | 5641 | 349 | AD | eGFR | -0.0213297 | -1 | 1 | 0.000454954 | 0 | 1 | 0.95407 | 893 | 0.986121623740202 |
| Chr1:14.7Mb | 19 | 1 | 14395689 | 15102429 | 3718 | 467 | AD | eGFR | -0.0156318 | -0.80481 | 0.72597 | 0.000244353 | 0 | 0.9581 | 0.958063 | 894 | 0.988300449275362 |

| locus_label | locus | chr | start | stop | n.snps | n.pcs | phen1 | phen2 | rho | rho.lower | rho.upper | r2 | r2.lower | r2.upper | p | rank | FDR |
|---|---|---|---|---|---|---|---|---|---|---|---|---|---|---|---|---|---|
| Chr8:125.7Mb | 1491 | 8 | 125022220 | 126395685 | 6603 | 481 | AD | eGFR | -0.0109916 | -0.51391 | 0.47632 | 0.000120815 | 0 | 0.34753 | 0.959676 | 895 | 0.988300449275362 |
| Chr2:16.4Mb | 260 | 2 | 15979650 | 16815314 | 3667 | 502 | AD | eGFR | 0.012986 | -0.59333 | 0.61626 | 0.000168635 | 0 | 0.54491 | 0.960287 | 896 | 0.988300449275362 |
| Chr4:57.1Mb | 751 | 4 | 56700327 | 57458590 | 3298 | 402 | AD | eGFR | -0.0177688 | -1 | 0.92829 | 0.000315729 | 0 | 1 | 0.960461 | 897 | 0.988300449275362 |
| Chr18:1.5Mb | 2430 | 18 | 1043159 | 2033113 | 5196 | 467 | AD | eGFR | 0.0145785 | -0.78441 | 0.84217 | 0.000212534 | 0 | 1 | 0.963173 | 898 | 0.989457024417314 |
| Chr9:94Mb | 1589 | 9 | 93444865 | 94645815 | 5400 | 463 | AD | eGFR | 0.00790332 | -0.37364 | 0.39276 | 6.25E-05 | 0 | 0.2053 | 0.964565 | 899 | 0.989457024417314 |
| Chr13:19.3Mb | 2028 | 13 | 19020047 | 19649408 | 2318 | 331 | AD | eGFR | 0.0103856 | -0.57701 | 0.61305 | 0.00010786 | 0 | 0.57265 | 0.965175 | 900 | 0.989457024417314 |
| Chr1:3.7Mb | 5 | 1 | 3352200 | 4105408 | 3591 | 490 | AD | eGFR | 0.0109748 | -0.63275 | 0.67391 | 0.000120446 | 0 | 0.66274 | 0.965873 | 901 | 0.989457024417314 |
| Chr4:3.5Mb | 690 | 4 | 3242345 | 3825841 | 3312 | 458 | AD | eGFR | -0.00616746 | -0.33391 | 0.33006 | 3.80E-05 | 0 | 0.1492 | 0.967902 | 902 | 0.990436303769401 |
| Chr5:129Mb | 1017 | 5 | 128509206 | 129590200 | 5132 | 365 | AD | eGFR | 0.00661095 | -0.38586 | 0.38898 | 4.37E-05 | 0 | 0.20797 | 0.970899 | 903 | 0.992402853820598 |
| Chr12:92.9Mb | 1987 | 12 | 92377552 | 93451080 | 4705 | 459 | AD | eGFR | -0.00809939 | -0.65426 | 0.63364 | 6.56E-05 | 0 | 0.60556 | 0.975765 | 904 | 0.994180422612514 |
| Chr4:170.1Mb | 870 | 4 | 169581769 | 170530560 | 3831 | 416 | AD | eGFR | 0.00839188 | -0.69091 | 0.72552 | 7.04E-05 | 0 | 0.80942 | 0.976399 | 905 | 0.994180422612514 |
| Chr20:32.3Mb | 2590 | 20 | 31702601 | 32904273 | 4159 | 356 | AD | eGFR | -0.00696252 | -0.58705 | 0.56367 | 4.85E-05 | 0 | 0.47619 | 0.977772 | 906 | 0.994180422612514 |
| Chr6:133.7Mb | 1189 | 6 | 133282304 | 134145640 | 3991 | 386 | AD | eGFR | -0.00456117 | -0.36199 | 0.34953 | 2.08E-05 | 0 | 0.16627 | 0.978646 | 907 | 0.994180422612514 |
| Chr1:85.3Mb | 94 | 1 | 84848250 | 85710245 | 3740 | 414 | AD | eGFR | 0.00464051 | -0.39681 | 0.40958 | 2.15E-05 | 0 | 0.2186 | 0.980612 | 908 | 0.994180422612514 |
| Chr7:29.5Mb | 1257 | 7 | 28898718 | 30174980 | 5853 | 511 | AD | eGFR | 0.00759928 | -0.83529 | 0.8392 | 5.77E-05 | 0 | 1 | 0.981203 | 909 | 0.994180422612514 |
| Chr4:190.7Mb | 899 | 4 | 190419641 | 191043880 | 1652 | 307 | AD | eGFR | -0.00645846 | -0.68264 | 0.66978 | 4.17E-05 | 0 | 0.7162 | 0.981255 | 910 | 0.994180422612514 |
| Chr5:169.7Mb | 1050 | 5 | 169354257 | 170051308 | 3477 | 496 | AD | eGFR | -0.00675684 | -0.74396 | 0.70143 | 4.57E-05 | 0 | 0.80318 | 0.981255 | 911 | 0.994180422612514 |
| Chr1:69.4Mb | 77 | 1 | 68920419 | 69940037 | 3955 | 347 | AD | eGFR | 0.00430822 | -0.49018 | 0.49645 | 1.86E-05 | 0 | 0.33583 | 0.984586 | 912 | 0.996461489035088 |
| Chr4:1.7Mb | 688 | 4 | 993212 | 2308570 | 5399 | 477 | AD | eGFR | 0.00558204 | -1 | 1 | 3.12E-05 | 0 | 1 | 0.987748 | 913 | 0.996902134636265 |
| Chr21:29.2Mb | 2636 | 21 | 28715012 | 29723403 | 5229 | 440 | AD | eGFR | -0.00281307 | -0.5246 | 0.50664 | 7.91E-06 | 0 | 0.38476 | 0.990047 | 914 | 0.996902134636265 |
| Chr4:67.1Mb | 763 | 4 | 66597941 | 67596264 | 5025 | 370 | AD | eGFR | -0.00293124 | -0.56655 | 0.54723 | 8.59E-06 | 0 | 0.42412 | 0.990409 | 915 | 0.996902134636265 |
| Chr4:37.6Mb | 736 | 4 | 37318907 | 37911853 | 3537 | 442 | AD | eGFR | 0.00291278 | -0.60251 | 0.60676 | 8.48E-06 | 0 | 0.52384 | 0.991076 | 916 | 0.996902134636265 |
| Chr16:59.2Mb | 2328 | 16 | 58531866 | 59883527 | 6206 | 446 | AD | eGFR | 0.00263242 | -0.58788 | 0.58136 | 6.93E-06 | 0 | 0.50754 | 0.991432 | 917 | 0.996902134636265 |
| Chr15:44.3Mb | 2235 | 15 | 43544904 | 45002686 | 3732 | 258 | AD | eGFR | -0.00215158 | -0.56809 | 0.5713 | 4.63E-06 | 0 | 0.5066 | 0.992458 | 918 | 0.996902134636265 |
| Chr13:106.4Mb | 2121 | 13 | 106065364 | 106651014 | 3527 | 455 | AD | eGFR | 0.00206401 | -0.52363 | 0.5332 | 4.26E-06 | 0 | 0.38202 | 0.992882 | 919 | 0.996902134636265 |
| Chr6:55Mb | 1120 | 6 | 54633143 | 55306955 | 3984 | 310 | AD | eGFR | 0.000784428 | -0.20431 | 0.20642 | 6.15E-07 | 0 | 0.05437 | 0.993909 | 920 | 0.996902134636265 |
| Chr12:45.3Mb | 1945 | 12 | 44923231 | 45716267 | 3680 | 288 | AD | eGFR | 0.0012098 | -0.38624 | 0.40321 | 1.46E-06 | 0 | 0.21682 | 0.994742 | 921 | 0.996902134636265 |
| Chr2:98.1Mb | 341 | 2 | 97331746 | 98791542 | 2703 | 306 | AD | eGFR | 0.00106713 | -0.65504 | 0.64292 | 1.14E-06 | 0 | 0.60946 | 0.996828 | 922 | 0.99790915835141 |
| Chr7:100.7Mb | 1319 | 7 | 10019371 | 10120058 | 4527 | 411 | AD | eGFR | 0.00037906 | -0.88855 | 0.88226 | 1.44E-07 | 0 | 1 | 0.999092 | 923 | 0.999092 |

| locus_label | locus | chr | start | stop | n.snps | n.pcs | phen1 | phen2 | rho | rho.lower | rho.upper | r2 | r2.lower | r2.upper | p | rank | FDR |
|---|---|---|---|---|---|---|---|---|---|---|---|---|---|---|---|---|---|
| | | | 9 | 4 | | | | | 8 | | | | | | | | |

**Supplementary Table S16. Pleiotropic SNPs Identified by Conjunctional FDR in European Ancestry (Kunkle Discovery).**

This table catalogs all individual single nucleotide polymorphisms (SNPs) identified as pleiotropic for both AD and eGFR using the conjunctional false discovery rate (conjFDR) method in European ancestry individuals. The conjFDR approach leverages the full distribution of test statistics from both GWAS to increase power for detecting variants associated with both traits. Each row represents a single SNP, with information including: conjFDR locus number grouping nearby SNPs into genomic regions, chromosome, base pair position, z-scores for AD and eGFR associations, the conjunctional FDR value, a pruning indicator denoting independent lead SNPs within each region, the minimum conjFDR value in the region, and individual p-values for AD and eGFR associations. SNPs with conjFDR < 0.05 are considered statistically significant pleiotropic variants.

| cFDRlocusnum | CHR | BP | zscore_AD | zscore_eGFR | conjfdr_AD_eGFR | prune_AD_eGFR | min_conjfdr | pval_AD | pval_eGFR | rsID |
|---|---|---|---|---|---|---|---|---|---|---|
| 1 | 2 | 55159284 | 4.080 | -3.309 | 4.010E-02 | 1 | 4.010E-02 | 4.510E-05 | 9.350E-04 | rs13422342 |
| 2 | 6 | 47647221 | 3.996 | 3.499 | 4.650E-02 | 0 | 4.650E-02 | 6.450E-05 | 4.670E-04 | rs10948374 |
| 2 | 6 | 47660735 | 4.005 | 3.275 | 4.510E-02 | 0 | 4.510E-02 | 6.190E-05 | 1.050E-03 | rs1890666 |
| 2 | 6 | 47660777 | 4.009 | 3.281 | 4.450E-02 | 0 | 4.450E-02 | 6.100E-05 | 1.040E-03 | rs1890667 |
| 2 | 6 | 47662085 | 4.159 | 3.325 | 3.840E-02 | 0 | 3.840E-02 | 3.190E-05 | 8.840E-04 | rs7763444 |
| 2 | 6 | 47662484 | 4.156 | 3.418 | 2.930E-02 | 1 | 2.930E-02 | 3.240E-05 | 6.300E-04 | rs9381599 |
| 3 | 8 | 95969322 | -3.983 | -3.684 | 4.890E-02 | 1 | 4.890E-02 | 6.810E-05 | 2.300E-04 | rs10429294 |
| 4 | 10 | 82250831 | -3.983 | 4.875 | 4.890E-02 | 0 | 4.890E-02 | 6.810E-05 | 1.090E-06 | rs7097656 |
| 4 | 10 | 82269461 | -4.008 | 5.160 | 4.450E-02 | 0 | 4.450E-02 | 6.130E-05 | 2.470E-07 | rs7080009 |
| 4 | 10 | 82269611 | -3.993 | 5.217 | 4.700E-02 | 0 | 4.700E-02 | 6.520E-05 | 1.820E-07 | rs1870138 |
| 4 | 10 | 82271341 | -4.310 | 5.144 | 1.320E-02 | 1 | 1.320E-02 | 1.640E-05 | 2.690E-07 | rs1870148 |
| 4 | 10 | 82273079 | -4.099 | 5.228 | 3.140E-02 | 0 | 3.140E-02 | 4.150E-05 | 1.710E-07 | rs10748526 |
| 4 | 10 | 82280137 | -4.126 | 5.172 | 2.830E-02 | 0 | 2.830E-02 | 3.700E-05 | 2.320E-07 | rs1878036 |
| 5 | 11 | 47249223 | 4.795 | 5.756 | 1.430E-03 | 0 | 1.430E-03 | 1.630E-06 | 8.630E-09 | rs11039138 |
| 5 | 11 | 47250319 | 4.784 | 5.743 | 1.510E-03 | 0 | 1.510E-03 | 1.720E-06 | 9.280E-09 | rs11039139 |
| 5 | 11 | 47261260 | 4.764 | 5.974 | 1.660E-03 | 0 | 1.660E-03 | 1.900E-06 | 2.310E-09 | rs11988 |
| 5 | 11 | 47262840 | 4.786 | 5.870 | 1.500E-03 | 0 | 1.500E-03 | 1.700E-06 | 4.350E-09 | rs2167080 |
| 5 | 11 | 47276675 | 5.451 | 6.113 | 4.580E-05 | 0 | 4.580E-05 | 5.020E-08 | 9.800E-10 | rs11039149 |
| 5 | 11 | 47278502 | 5.395 | 5.992 | 6.290E-05 | 0 | 6.290E-05 | 6.860E-08 | 2.070E-09 | rs11039154 |
| 5 | 11 | 47294021 | 4.760 | 5.831 | 1.690E-03 | 0 | 1.690E-03 | 1.930E-06 | 5.500E-09 | rs11039159 |
| 5 | 11 | 47301225 | 5.351 | 5.978 | 7.980E-05 | 0 | 7.980E-05 | 8.720E-08 | 2.260E-09 | rs7944419 |
| 5 | 11 | 473027 | 4.755 | 5.834 | 1.730E-03 | 0 | 1.730E-03 | 1.990E | 5.420E- | rs116066 |

| cFDRlocusnum | CHR | BP | zscore_AD | zscore_eGFR | conjfdr_AD_eGFR | prune_AD_eGFR | min_conjfdr | pval_AD | pval_eGFR | rsID |
|---|---|---|---|---|---|---|---|---|---|---|
| | | 33 | | | | | | -06 | 09 | 83 |
| 5 | 11 | 47303612 | 4.783 | 5.876 | 1.520E-03 | 0 | 1.520E-03 | 1.720E-06 | 4.210E-09 | rs11039162 |
| 5 | 11 | 47309373 | 4.698 | 5.860 | 2.270E-03 | 0 | 2.270E-03 | 2.620E-06 | 4.640E-09 | rs12802374 |
| 5 | 11 | 47312689 | 5.625 | 6.023 | 1.720E-05 | 0 | 1.720E-05 | 1.860E-08 | 1.720E-09 | rs11039165 |
| 5 | 11 | 47318157 | 4.810 | 5.844 | 1.330E-03 | 0 | 1.330E-03 | 1.510E-06 | 5.090E-09 | rs749067 |
| 5 | 11 | 47322327 | 4.920 | 5.827 | 7.720E-04 | 0 | 7.720E-04 | 8.650E-07 | 5.630E-09 | rs7116451 |
| 5 | 11 | 47346723 | 5.341 | 6.085 | 8.460E-05 | 0 | 8.460E-05 | 9.260E-08 | 1.160E-09 | rs11039182 |
| 5 | 11 | 47346940 | 5.224 | 6.644 | 1.590E-04 | 0 | 1.590E-04 | 1.750E-07 | 3.060E-11 | rs11039183 |
| 5 | 11 | 47361084 | 5.446 | 6.670 | 4.720E-05 | 0 | 4.720E-05 | 5.160E-08 | 2.550E-11 | rs2856653 |
| 5 | 11 | 47361838 | 5.470 | 6.707 | 4.140E-05 | 0 | 4.140E-05 | 4.500E-08 | 1.980E-11 | rs2596407 |
| 5 | 11 | 47363215 | 5.471 | 6.559 | 4.120E-05 | 0 | 4.120E-05 | 4.480E-08 | 5.410E-11 | rs2697923 |
| 5 | 11 | 47363285 | 5.370 | 6.659 | 7.180E-05 | 0 | 7.180E-05 | 7.890E-08 | 2.750E-11 | rs10838696 |
| 5 | 11 | 47374998 | -6.205 | -3.856 | 7.180E-03 | 0 | 7.180E-03 | 5.450E-10 | 1.150E-04 | rs2856661 |
| 5 | 11 | 47375193 | -6.179 | -4.037 | 3.870E-03 | 0 | 3.870E-03 | 6.470E-10 | 5.420E-05 | rs11039195 |
| 5 | 11 | 47376363 | -6.480 | -4.178 | 2.350E-03 | 0 | 2.350E-03 | 9.190E-11 | 2.940E-05 | rs3740686 |
| 5 | 11 | 47376448 | -6.482 | -4.126 | 2.830E-03 | 0 | 2.830E-03 | 9.050E-11 | 3.690E-05 | rs1057233 |
| 5 | 11 | 47377283 | 5.300 | 7.437 | 1.060E-04 | 0 | 1.060E-04 | 1.160E-07 | 1.030E-13 | rs71475909 |
| 5 | 11 | 47378245 | 5.812 | 7.644 | 5.770E-06 | 1 | 5.770E-06 | 6.190E-09 | 2.110E-14 | rs3824869 |
| 5 | 11 | 47379407 | -6.504 | -4.155 | 2.550E-03 | 0 | 2.550E-03 | 7.830E-11 | 3.250E-05 | rs2084946 |
| 5 | 11 | 47379615 | -6.447 | -4.266 | 1.700E-03 | 0 | 1.700E-03 | 1.140E-10 | 1.990E-05 | rs1542321 |
| 5 | 11 | 47379646 | -6.076 | -4.130 | 2.790E-03 | 0 | 2.790E-03 | 1.230E-09 | 3.630E-05 | rs11039200 |
| 5 | 11 | 47380061 | -6.471 | -4.221 | 2.010E-03 | 0 | 2.010E-03 | 9.750E-11 | 2.440E-05 | rs10734557 |
| 5 | 11 | 47383275 | -6.427 | -4.444 | 8.600E-04 | 0 | 8.600E-04 | 1.300E-10 | 8.840E-06 | rs7105851 |
| 5 | 11 | 47384597 | -6.434 | -4.460 | 8.040E-04 | 0 | 8.040E-04 | 1.250E-10 | 8.200E-06 | rs11039203 |
| 5 | 11 | 47385194 | -6.434 | -4.474 | 7.590E-04 | 0 | 7.590E-04 | 1.250E-10 | 7.670E-06 | rs67116452 |
| 5 | 11 | 47385923 | -6.428 | -4.425 | 9.230E-04 | 0 | 9.230E-04 | 1.290E-10 | 9.640E-06 | rs10838698 |
| 5 | 11 | 47386073 | -6.437 | -4.394 | 1.050E-03 | 0 | 1.050E-03 | 1.220E-10 | 1.110E-05 | rs10838699 |
| 5 | 11 | 47387599 | -6.418 | -4.433 | 8.980E-04 | 0 | 8.980E-04 | 1.380E-10 | 9.310E-06 | rs935914 |
| 5 | 11 | 47388214 | 5.779 | 8.018 | 7.020E-06 | 0 | 7.020E-06 | 7.520E-09 | 1.070E-15 | rs12801188 |
| 5 | 11 | 47389223 | -6.381 | -4.518 | 6.360E-04 | 0 | 6.360E-04 | 1.760E-10 | 6.250E-06 | rs35996350 |

| cFDRlocusnum | CHR | BP | zscore_AD | zscore_eGFR | conjfdr_AD_eGFR | prune_AD_eGFR | min_conjfdr | pval_AD | pval_eGFR | rsID |
|---|---|---|---|---|---|---|---|---|---|---|
| 5 | 11 | 47389638 | 5.770 | 8.061 | 7.350E-06 | 0 | 7.350E-06 | 7.910E-09 | 7.580E-16 | rs34937994 |
| 5 | 11 | 47390692 | -6.488 | -4.381 | 1.100E-03 | 0 | 1.100E-03 | 8.680E-11 | 1.180E-05 | rs4752987 |
| 5 | 11 | 47391039 | -6.421 | -4.402 | 1.010E-03 | 0 | 1.010E-03 | 1.350E-10 | 1.070E-05 | rs10769258 |
| 5 | 11 | 47391745 | -6.545 | -4.489 | 7.170E-04 | 0 | 7.170E-04 | 5.970E-11 | 7.160E-06 | rs67472071 |
| 5 | 11 | 47391879 | -6.400 | -4.364 | 1.170E-03 | 0 | 1.170E-03 | 1.560E-10 | 1.280E-05 | rs7928163 |
| 5 | 11 | 47391948 | 5.775 | 8.813 | 7.190E-06 | 0 | 7.190E-06 | 7.710E-09 | 1.220E-18 | rs10437655 |
| 5 | 11 | 47392114 | -5.909 | -4.180 | 2.330E-03 | 0 | 2.330E-03 | 3.440E-09 | 2.920E-05 | rs7928419 |
| 5 | 11 | 47393638 | -6.314 | -4.410 | 9.780E-04 | 0 | 9.780E-04 | 2.720E-10 | 1.030E-05 | rs4992357 |
| 5 | 11 | 47394305 | 5.752 | 8.701 | 8.230E-06 | 0 | 8.230E-06 | 8.810E-09 | 3.290E-18 | rs896817 |
| 5 | 11 | 47394338 | -6.387 | -4.362 | 1.180E-03 | 0 | 1.180E-03 | 1.700E-10 | 1.290E-05 | rs896816 |
| 5 | 11 | 47395240 | -6.388 | -4.356 | 1.210E-03 | 0 | 1.210E-03 | 1.680E-10 | 1.320E-05 | rs7940536 |
| 5 | 11 | 47397353 | -6.020 | -4.216 | 2.040E-03 | 0 | 2.040E-03 | 1.740E-09 | 2.490E-05 | rs56030824 |
| 5 | 11 | 47398963 | -6.019 | -4.133 | 2.760E-03 | 0 | 2.760E-03 | 1.750E-09 | 3.590E-05 | rs12146565 |
| 5 | 11 | 47399602 | 5.691 | 8.009 | 1.170E-05 | 0 | 1.170E-05 | 1.260E-08 | 1.160E-15 | rs2278890 |
| 5 | 11 | 47400602 | 5.685 | 8.040 | 1.210E-05 | 0 | 1.210E-05 | 1.300E-08 | 8.980E-16 | rs3758675 |
| 5 | 11 | 47401058 | -6.367 | -4.521 | 6.280E-04 | 0 | 6.280E-04 | 1.920E-10 | 6.160E-06 | rs11601173 |
| 5 | 11 | 47401060 | -6.367 | -4.562 | 5.300E-04 | 0 | 5.300E-04 | 1.920E-10 | 5.060E-06 | rs11607981 |
| 5 | 11 | 47401171 | -6.366 | -4.510 | 6.580E-04 | 0 | 6.580E-04 | 1.940E-10 | 6.500E-06 | rs11601603 |
| 5 | 11 | 47406592 | -4.966 | -7.248 | 6.110E-04 | 0 | 6.110E-04 | 6.830E-07 | 4.240E-13 | rs11039216 |
| 5 | 11 | 47407439 | 5.481 | 8.081 | 3.900E-05 | 0 | 3.900E-05 | 4.240E-08 | 6.400E-16 | rs11606287 |
| 5 | 11 | 47408592 | 5.468 | 8.094 | 4.190E-05 | 0 | 4.190E-05 | 4.560E-08 | 5.770E-16 | rs35032070 |
| 5 | 11 | 47409212 | -6.275 | -4.372 | 1.140E-03 | 0 | 1.140E-03 | 3.490E-10 | 1.230E-05 | rs7111957 |
| 5 | 11 | 47410200 | -6.345 | -4.284 | 1.590E-03 | 0 | 1.590E-03 | 2.220E-10 | 1.830E-05 | rs6485753 |
| 5 | 11 | 47410393 | -6.252 | -4.456 | 8.180E-04 | 0 | 8.180E-04 | 4.050E-10 | 8.360E-06 | rs4752990 |
| 5 | 11 | 47410888 | 5.466 | 8.864 | 4.220E-05 | 0 | 4.220E-05 | 4.590E-08 | 7.690E-19 | rs10838702 |
| 5 | 11 | 47410951 | -6.322 | -4.341 | 1.280E-03 | 0 | 1.280E-03 | 2.590E-10 | 1.420E-05 | rs4752993 |
| 5 | 11 | 47411581 | -6.325 | -4.310 | 1.440E-03 | 0 | 1.440E-03 | 2.530E-10 | 1.630E-05 | rs10769262 |
| 5 | 11 | 47413475 | -6.276 | -4.454 | 8.250E-04 | 0 | 8.250E-04 | 3.470E-10 | 8.430E-06 | rs12803525 |
| 5 | 11 | 47413733 | -6.240 | -4.482 | 7.370E-04 | 0 | 7.370E-04 | 4.380E-10 | 7.390E-06 | rs11605672 |
| 5 | 11 | 474166 | -6.318 | -4.468 | 7.780E-04 | 0 | 7.780E-04 | 2.650E | 7.910E- | rs558761 |

| cFDRlocusnum | CHR | BP | zscore_AD | zscore_eGFR | conjfdr_AD_eGFR | prune_AD_eGFR | min_conjfdr | pval_AD | pval_eGFR | rsID |
|---|---|---|---|---|---|---|---|---|---|---|
| | | 36 | | | | | | -10 | 06 | 53 |
| 5 | 11 | 47416746 | 5.413 | 8.774 | 5.670E-05 | 0 | 5.670E-05 | 6.210E-08 | 1.730E-18 | rs1377416 |
| 5 | 11 | 47417183 | 5.397 | 8.780 | 6.200E-05 | 0 | 6.200E-05 | 6.770E-08 | 1.630E-18 | rs10769263 |
| 5 | 11 | 47418306 | -6.288 | -4.343 | 1.270E-03 | 0 | 1.270E-03 | 3.220E-10 | 1.410E-05 | rs12802273 |
| 5 | 11 | 47419129 | -6.277 | -4.377 | 1.120E-03 | 0 | 1.120E-03 | 3.440E-10 | 1.200E-05 | rs7924485 |
| 5 | 11 | 47419301 | -6.167 | -4.300 | 1.500E-03 | 0 | 1.500E-03 | 6.940E-10 | 1.710E-05 | rs896815 |
| 5 | 11 | 47419663 | 5.390 | 8.757 | 6.440E-05 | 0 | 6.440E-05 | 7.030E-08 | 2.010E-18 | rs1534576 |
| 5 | 11 | 47419757 | -6.214 | -4.477 | 7.510E-04 | 0 | 7.510E-04 | 5.160E-10 | 7.570E-06 | rs1317164 |
| 5 | 11 | 47423340 | 5.313 | 8.810 | 9.840E-05 | 0 | 9.840E-05 | 1.080E-07 | 1.250E-18 | rs10769264 |
| 5 | 11 | 47423553 | -6.031 | -4.203 | 2.140E-03 | 0 | 2.140E-03 | 1.630E-09 | 2.630E-05 | rs4752832 |
| 5 | 11 | 47424400 | -6.211 | -4.464 | 7.900E-04 | 0 | 7.900E-04 | 5.250E-10 | 8.050E-06 | rs11039219 |
| 5 | 11 | 47424717 | -5.806 | -4.257 | 1.760E-03 | 0 | 1.760E-03 | 6.390E-09 | 2.080E-05 | rs10838704 |
| 5 | 11 | 47424931 | 5.350 | 8.799 | 8.050E-05 | 0 | 8.050E-05 | 8.800E-08 | 1.380E-18 | rs10838705 |
| 5 | 11 | 47426956 | -6.187 | -4.474 | 7.590E-04 | 0 | 7.590E-04 | 6.110E-10 | 7.670E-06 | rs112060627 |
| 5 | 11 | 47427667 | 4.758 | 8.798 | 1.710E-03 | 0 | 1.710E-03 | 1.960E-06 | 1.390E-18 | rs61897431 |
| 5 | 11 | 47427739 | 5.355 | 8.787 | 7.840E-05 | 0 | 7.840E-05 | 8.580E-08 | 1.540E-18 | rs11039221 |
| 5 | 11 | 47428565 | -6.192 | -4.532 | 6.020E-04 | 0 | 6.020E-04 | 5.930E-10 | 5.840E-06 | rs4752999 |
| 5 | 11 | 47429904 | 5.347 | 8.789 | 8.170E-05 | 0 | 8.170E-05 | 8.930E-08 | 1.510E-18 | rs7947450 |
| 5 | 11 | 47430458 | -6.256 | -4.267 | 1.690E-03 | 0 | 1.690E-03 | 3.940E-10 | 1.980E-05 | rs7937331 |
| 5 | 11 | 47430599 | 5.341 | 8.771 | 8.460E-05 | 0 | 8.460E-05 | 9.260E-08 | 1.760E-18 | rs11039225 |
| 5 | 11 | 47431966 | -5.865 | -4.258 | 1.750E-03 | 0 | 1.750E-03 | 4.500E-09 | 2.070E-05 | rs755555 |
| 5 | 11 | 47432303 | -6.219 | -4.323 | 1.370E-03 | 0 | 1.370E-03 | 5.010E-10 | 1.540E-05 | rs755553 |
| 5 | 11 | 47432725 | 5.342 | 8.809 | 8.410E-05 | 0 | 8.410E-05 | 9.210E-08 | 1.260E-18 | rs10742802 |
| 5 | 11 | 47434777 | -6.183 | -4.251 | 1.800E-03 | 0 | 1.800E-03 | 6.280E-10 | 2.130E-05 | rs6485756 |
| 5 | 11 | 47434986 | 4.025 | 7.598 | 4.180E-02 | 0 | 4.180E-02 | 5.690E-05 | 3.000E-14 | rs2293576 |
| 5 | 11 | 47437202 | -6.197 | -4.302 | 1.490E-03 | 0 | 1.490E-03 | 5.770E-10 | 1.700E-05 | rs2293577 |
| 5 | 11 | 47437403 | -5.743 | -4.293 | 1.540E-03 | 0 | 1.540E-03 | 9.300E-09 | 1.760E-05 | rs2293578 |
| 5 | 11 | 47438282 | -6.127 | -4.105 | 3.040E-03 | 0 | 3.040E-03 | 8.930E-10 | 4.050E-05 | rs2053980 |
| 5 | 11 | 47439444 | -6.073 | -4.466 | 7.840E-04 | 0 | 7.840E-04 | 1.260E-09 | 7.980E-06 | rs2053979 |
| 5 | 11 | 47439938 | -6.152 | -4.274 | 1.650E-03 | 0 | 1.650E-03 | 7.640E-10 | 1.920E-05 | rs10742803 |

| cFDRlocusnum | CHR | BP | zscore_AD | zscore_eGFR | conjfdr_AD_eGFR | prune_AD_eGFR | min_conjfdr | pval_AD | pval_eGFR | rsID |
|---|---|---|---|---|---|---|---|---|---|---|
| 5 | 11 | 47440758 | 5.378 | 8.776 | 6.870E-05 | 0 | 6.870E-05 | 7.510E-08 | 1.690E-18 | rs2293579 |
| 5 | 11 | 47443080 | -6.112 | -4.451 | 8.350E-04 | 0 | 8.350E-04 | 9.840E-10 | 8.550E-06 | rs3781625 |
| 5 | 11 | 47443251 | -5.427 | -4.378 | 1.110E-03 | 0 | 1.110E-03 | 5.750E-08 | 1.200E-05 | rs3781624 |
| 5 | 11 | 47447346 | 5.330 | 8.772 | 8.930E-05 | 0 | 8.930E-05 | 9.820E-08 | 1.760E-18 | rs10838709 |
| 5 | 11 | 47448497 | -6.172 | -4.418 | 9.490E-04 | 0 | 9.490E-04 | 6.750E-10 | 9.980E-06 | rs11600581 |
| 5 | 11 | 47449072 | 5.343 | 8.770 | 8.370E-05 | 0 | 8.370E-05 | 9.160E-08 | 1.780E-18 | rs12292911 |
| 5 | 11 | 47449101 | -6.165 | -4.318 | 1.390E-03 | 0 | 1.390E-03 | 7.060E-10 | 1.570E-05 | rs60075622 |
| 5 | 11 | 47450338 | -6.089 | -4.446 | 8.510E-04 | 0 | 8.510E-04 | 1.140E-09 | 8.750E-06 | rs35496532 |
| 5 | 11 | 47457539 | -5.820 | -4.162 | 2.480E-03 | 0 | 2.480E-03 | 5.890E-09 | 3.150E-05 | rs11604680 |
| 5 | 11 | 47458765 | -6.185 | -4.283 | 1.600E-03 | 0 | 1.600E-03 | 6.190E-10 | 1.850E-05 | rs7106956 |
| 5 | 11 | 47460901 | -5.635 | -4.186 | 2.280E-03 | 0 | 2.280E-03 | 1.750E-08 | 2.840E-05 | rs11039238 |
| 5 | 11 | 47461693 | 4.117 | 6.950 | 2.930E-02 | 0 | 2.930E-02 | 3.850E-05 | 3.660E-12 | rs7103835 |
| 5 | 11 | 47461783 | 5.346 | 8.796 | 8.230E-05 | 0 | 8.230E-05 | 9.000E-08 | 1.420E-18 | rs7103648 |
| 5 | 11 | 47462140 | 5.344 | 8.804 | 8.310E-05 | 0 | 8.310E-05 | 9.090E-08 | 1.320E-18 | rs7104036 |
| 5 | 11 | 47466006 | -6.109 | -4.251 | 1.800E-03 | 0 | 1.800E-03 | 1.000E-09 | 2.120E-05 | rs4752838 |
| 5 | 11 | 47474146 | 4.429 | 5.793 | 7.840E-03 | 0 | 7.840E-03 | 9.450E-06 | 6.910E-09 | rs12361415 |
| 5 | 11 | 47475189 | -6.097 | -3.327 | 3.820E-02 | 0 | 3.820E-02 | 1.080E-09 | 8.780E-04 | rs35184771 |
| 5 | 11 | 47486885 | -6.091 | -3.392 | 3.180E-02 | 0 | 3.180E-02 | 1.120E-09 | 6.940E-04 | rs1317149 |
| 5 | 11 | 47495746 | -5.954 | -3.320 | 3.900E-02 | 0 | 3.900E-02 | 2.610E-09 | 9.020E-04 | rs11039255 |
| 5 | 11 | 47500400 | 4.405 | 6.305 | 8.720E-03 | 0 | 8.720E-03 | 1.060E-05 | 2.890E-10 | rs12224672 |
| 5 | 11 | 47523214 | -5.995 | -3.512 | 2.210E-02 | 0 | 2.210E-02 | 2.040E-09 | 4.450E-04 | rs11039265 |
| 5 | 11 | 47530024 | 4.284 | 6.829 | 1.480E-02 | 0 | 1.480E-02 | 1.840E-05 | 8.560E-12 | rs6485758 |
| 5 | 11 | 47532395 | 4.385 | 6.221 | 9.560E-03 | 0 | 9.560E-03 | 1.160E-05 | 4.930E-10 | rs11039266 |
| 5 | 11 | 47539697 | -5.994 | -3.354 | 3.530E-02 | 0 | 3.530E-02 | 2.050E-09 | 7.970E-04 | rs4752845 |
| 5 | 11 | 47547046 | -6.003 | -3.318 | 3.920E-02 | 0 | 3.920E-02 | 1.930E-09 | 9.070E-04 | rs34958982 |
| 5 | 11 | 47557871 | 4.277 | 6.874 | 1.520E-02 | 0 | 1.520E-02 | 1.900E-05 | 6.250E-12 | rs10838725 |
| 5 | 11 | 47560433 | 4.422 | 6.304 | 8.070E-03 | 0 | 8.070E-03 | 9.770E-06 | 2.890E-10 | rs61895112 |
| 5 | 11 | 47564785 | 4.274 | 6.187 | 1.530E-02 | 0 | 1.530E-02 | 1.920E-05 | 6.140E-10 | rs11039284 |
| 5 | 11 | 47568074 | -5.942 | -3.302 | 4.090E-02 | 0 | 4.090E-02 | 2.820E-09 | 9.580E-04 | rs66749409 |
| 5 | 11 | 475719 | 4.366 | 6.232 | 1.030E-02 | 0 | 1.030E-02 | 1.270E | 4.620E- | rs122250 |

| cFDRlocusnum | CHR | BP | zscore_AD | zscore_eGFR | conjfdr_AD_eGFR | prune_AD_eGFR | min_conjfdr | pval_AD | pval_eGFR | rsID |
|---|---|---|---|---|---|---|---|---|---|---|
| | | 67 | | | | | | -05 | 10 | 51 |
| 5 | 11 | 47572279 | 4.255 | 6.841 | 1.670E-02 | 0 | 1.670E-02 | 2.090E-05 | 7.850E-12 | rs11039290 |
| 5 | 11 | 47573342 | 4.212 | 6.822 | 1.990E-02 | 0 | 1.990E-02 | 2.530E-05 | 8.980E-12 | rs71475921 |
| 5 | 11 | 47581443 | 4.158 | 6.918 | 2.480E-02 | 0 | 2.480E-02 | 3.220E-05 | 4.570E-12 | rs11039297 |
| 5 | 11 | 47583121 | -5.929 | -3.311 | 3.990E-02 | 0 | 3.990E-02 | 3.050E-09 | 9.300E-04 | rs12798346 |
| 5 | 11 | 47586376 | -5.930 | -3.293 | 4.180E-02 | 0 | 4.180E-02 | 3.030E-09 | 9.900E-04 | rs56400411 |
| 5 | 11 | 47589707 | -5.690 | -3.302 | 4.090E-02 | 0 | 4.090E-02 | 1.270E-08 | 9.580E-04 | rs7945473 |
| 5 | 11 | 47600438 | 4.232 | 6.280 | 1.840E-02 | 0 | 1.840E-02 | 2.320E-05 | 3.390E-10 | rs2280231 |
| 5 | 11 | 47602729 | -5.886 | -3.318 | 3.910E-02 | 0 | 3.910E-02 | 3.950E-09 | 9.050E-04 | rs2030166 |
| 5 | 11 | 47606483 | -6.398 | -3.564 | 1.880E-02 | 0 | 1.880E-02 | 1.570E-10 | 3.650E-04 | rs11605348 |
| 5 | 11 | 47607135 | 4.159 | 6.818 | 2.470E-02 | 0 | 2.470E-02 | 3.200E-05 | 9.260E-12 | rs10838731 |
| 5 | 11 | 47622139 | -5.931 | -3.297 | 4.150E-02 | 0 | 4.150E-02 | 3.010E-09 | 9.790E-04 | rs12799623 |
| 5 | 11 | 47623890 | -5.890 | -3.430 | 2.830E-02 | 0 | 2.830E-02 | 3.850E-09 | 6.030E-04 | rs12363232 |
| 5 | 11 | 47624714 | -5.913 | -3.298 | 4.130E-02 | 0 | 4.130E-02 | 3.350E-09 | 9.720E-04 | rs12419692 |
| 5 | 11 | 47626492 | 4.184 | 6.818 | 2.240E-02 | 0 | 2.240E-02 | 2.870E-05 | 9.250E-12 | rs10769282 |
| 5 | 11 | 47648042 | -5.802 | -3.326 | 3.840E-02 | 0 | 3.840E-02 | 6.530E-09 | 8.820E-04 | rs4752856 |
| 5 | 11 | 47655752 | -5.853 | -3.312 | 3.980E-02 | 0 | 3.980E-02 | 4.840E-09 | 9.260E-04 | rs4752857 |
| 5 | 11 | 47658314 | -5.884 | -3.278 | 4.370E-02 | 0 | 4.370E-02 | 3.990E-09 | 1.050E-03 | rs12794570 |
| 5 | 11 | 47659135 | 4.281 | 6.251 | 1.490E-02 | 0 | 1.490E-02 | 1.860E-05 | 4.070E-10 | rs7118178 |
| 5 | 11 | 47662932 | 4.313 | 6.806 | 1.300E-02 | 0 | 1.300E-02 | 1.610E-05 | 1.000E-11 | rs7120548 |
| 5 | 11 | 47663049 | -5.936 | -3.337 | 3.720E-02 | 0 | 3.720E-02 | 2.930E-09 | 8.480E-04 | rs10838738 |
| 5 | 11 | 47676170 | 4.785 | 6.273 | 1.500E-03 | 0 | 1.500E-03 | 1.710E-06 | 3.540E-10 | rs7107356 |
| 5 | 11 | 47695840 | 4.078 | 6.175 | 3.420E-02 | 0 | 3.420E-02 | 4.550E-05 | 6.610E-10 | rs11039332 |
| 5 | 11 | 47733494 | 4.468 | 4.814 | 6.560E-03 | 0 | 6.560E-03 | 7.880E-06 | 1.480E-06 | rs1017730 |
| 5 | 11 | 47760078 | 4.021 | 5.917 | 4.240E-02 | 0 | 4.240E-02 | 5.790E-05 | 3.280E-09 | rs12807014 |
| 5 | 11 | 47769620 | 4.103 | 5.346 | 3.080E-02 | 0 | 3.080E-02 | 4.070E-05 | 8.990E-08 | rs11039364 |
| 5 | 11 | 47774238 | 4.030 | 5.911 | 4.110E-02 | 0 | 4.110E-02 | 5.580E-05 | 3.400E-09 | rs12577383 |
| 5 | 11 | 47780754 | 4.025 | 5.955 | 4.180E-02 | 0 | 4.180E-02 | 5.690E-05 | 2.610E-09 | rs7927445 |
| 5 | 11 | 47789082 | 4.030 | 5.932 | 4.100E-02 | 0 | 4.100E-02 | 5.570E-05 | 2.990E-09 | rs12223593 |
| 5 | 11 | 47820241 | 4.295 | 4.307 | 1.410E-02 | 0 | 1.410E-02 | 1.750E-05 | 1.660E-05 | rs34228231 |

| cFDRlocusnum | CHR | BP | zscore_AD | zscore_eGFR | conjfdr_AD_eGFR | prune_AD_eGFR | min_conjfdr | pval_AD | pval_eGFR | rsID |
|---|---|---|---|---|---|---|---|---|---|---|
| 5 | 11 | 47907641 | -5.717 | -3.374 | 3.340E-02 | 0 | 3.340E-02 | 1.080E-08 | 7.420E-04 | rs4752801 |
| 5 | 11 | 47910823 | -5.708 | -3.400 | 3.100E-02 | 0 | 3.100E-02 | 1.150E-08 | 6.740E-04 | rs12361256 |
| 5 | 11 | 47915299 | -5.961 | -3.364 | 3.440E-02 | 0 | 3.440E-02 | 2.510E-09 | 7.690E-04 | rs34467936 |
| 6 | 11 | 85655105 | -4.720 | 3.915 | 5.880E-03 | 0 | 5.880E-03 | 2.360E-06 | 9.040E-05 | rs1237230 |
| 6 | 11 | 85659847 | -4.623 | 3.912 | 5.940E-03 | 0 | 5.940E-03 | 3.780E-06 | 9.150E-05 | rs2458499 |
| 6 | 11 | 85666138 | -4.862 | 3.909 | 6.010E-03 | 0 | 6.010E-03 | 1.160E-06 | 9.290E-05 | rs11234493 |
| 6 | 11 | 85666440 | -4.644 | 3.904 | 6.090E-03 | 0 | 6.090E-03 | 3.420E-06 | 9.440E-05 | rs556562 |
| 6 | 11 | 85671702 | -4.564 | 3.962 | 5.000E-03 | 0 | 5.000E-03 | 5.010E-06 | 7.420E-05 | rs618679 |
| 6 | 11 | 85672383 | -4.374 | 3.878 | 9.980E-03 | 0 | 9.980E-03 | 1.220E-05 | 1.050E-04 | rs2077815 |
| 6 | 11 | 85673124 | -4.511 | 4.019 | 5.410E-03 | 0 | 5.410E-03 | 6.440E-06 | 5.850E-05 | rs592314 |
| 6 | 11 | 85675533 | -4.585 | 3.931 | 5.570E-03 | 0 | 5.570E-03 | 4.540E-06 | 8.460E-05 | rs548121 |
| 6 | 11 | 85682120 | -4.359 | 3.902 | 1.070E-02 | 0 | 1.070E-02 | 1.310E-05 | 9.550E-05 | rs626542 |
| 6 | 11 | 85685166 | -4.589 | 3.896 | 6.260E-03 | 0 | 6.260E-03 | 4.440E-06 | 9.780E-05 | rs637304 |
| 6 | 11 | 85689435 | -4.354 | 3.810 | 1.090E-02 | 0 | 1.090E-02 | 1.330E-05 | 1.390E-04 | rs618629 |
| 6 | 11 | 85692342 | -4.361 | 3.817 | 1.060E-02 | 0 | 1.060E-02 | 1.300E-05 | 1.350E-04 | rs2508688 |
| 6 | 11 | 85696670 | -4.482 | 3.903 | 6.210E-03 | 0 | 6.210E-03 | 7.410E-06 | 9.520E-05 | rs585820 |
| 6 | 11 | 85698598 | -4.505 | 3.947 | 5.580E-03 | 0 | 5.580E-03 | 6.640E-06 | 7.900E-05 | rs670606 |
| 6 | 11 | 85700000 | -4.535 | 3.907 | 6.050E-03 | 0 | 6.050E-03 | 5.760E-06 | 9.360E-05 | rs654194 |
| 6 | 11 | 85701622 | -4.549 | 3.973 | 4.820E-03 | 0 | 4.820E-03 | 5.380E-06 | 7.090E-05 | rs488134 |
| 6 | 11 | 85701741 | -4.567 | 3.998 | 4.410E-03 | 0 | 4.410E-03 | 4.960E-06 | 6.380E-05 | rs682366 |
| 6 | 11 | 85702967 | -4.054 | 3.929 | 3.730E-02 | 0 | 3.730E-02 | 5.040E-05 | 8.530E-05 | rs659801 |
| 6 | 11 | 85705885 | -4.444 | 4.027 | 7.360E-03 | 0 | 7.360E-03 | 8.840E-06 | 5.660E-05 | rs595972 |
| 6 | 11 | 85707228 | -4.357 | 3.838 | 1.080E-02 | 0 | 1.080E-02 | 1.320E-05 | 1.240E-04 | rs542167 |
| 6 | 11 | 85709559 | -4.358 | 3.876 | 1.070E-02 | 0 | 1.070E-02 | 1.310E-05 | 1.060E-04 | rs603615 |
| 6 | 11 | 85710169 | -4.351 | 3.847 | 1.110E-02 | 0 | 1.110E-02 | 1.360E-05 | 1.190E-04 | rs600258 |
| 6 | 11 | 85710180 | -4.400 | 4.028 | 8.930E-03 | 0 | 8.930E-03 | 1.080E-05 | 5.630E-05 | rs616555 |
| 6 | 11 | 85713565 | -4.376 | 3.798 | 9.920E-03 | 0 | 9.920E-03 | 1.210E-05 | 1.460E-04 | rs2155051 |
| 6 | 11 | 85715736 | -4.523 | 4.008 | 5.150E-03 | 0 | 5.150E-03 | 6.110E-06 | 6.130E-05 | rs527162 |
| 6 | 11 | 85715853 | -4.354 | 3.885 | 1.090E-02 | 0 | 1.090E-02 | 1.340E-05 | 1.020E-04 | rs528109 |
| 6 | 11 | 857256 | -4.274 | 3.871 | 1.540E-02 | 0 | 1.540E-02 | 1.920E | 1.090E- | rs622220 |

| cFDRlocusnum | CHR | BP | zscore_AD | zscore_eGFR | conjfdr_AD_eGFR | prune_AD_eGFR | min_conjfdr | pval_AD | pval_eGFR | rsID |
|---|---|---|---|---|---|---|---|---|---|---|
| | | 21 | | | | | | -05 | 04 | |
| 6 | 11 | 85725937 | -4.481 | 4.038 | 6.220E-03 | 0 | 6.220E-03 | 7.430E-06 | 5.400E-05 | rs592297 |
| 6 | 11 | 85726818 | -4.564 | 4.017 | 4.240E-03 | 1 | 4.240E-03 | 5.010E-06 | 5.890E-05 | rs648270 |
| 6 | 11 | 85728030 | -4.278 | 3.918 | 1.510E-02 | 0 | 1.510E-02 | 1.890E-05 | 8.930E-05 | rs1996108 |
| 6 | 11 | 85743449 | -4.405 | 3.982 | 8.730E-03 | 0 | 8.730E-03 | 1.060E-05 | 6.830E-05 | rs547767 |
| 6 | 11 | 85764875 | -4.509 | 3.861 | 7.060E-03 | 0 | 7.060E-03 | 6.520E-06 | 1.130E-04 | rs497711 |
| 6 | 11 | 85765737 | -4.661 | 3.974 | 4.810E-03 | 0 | 4.810E-03 | 3.150E-06 | 7.070E-05 | rs489486 |
| 6 | 11 | 85766582 | -4.494 | 3.851 | 7.300E-03 | 0 | 7.300E-03 | 6.990E-06 | 1.180E-04 | rs9804630 |
| 6 | 11 | 85771065 | -4.498 | 3.824 | 8.000E-03 | 0 | 8.000E-03 | 6.870E-06 | 1.310E-04 | rs634719 |
| 6 | 11 | 85771170 | -4.661 | 3.963 | 4.980E-03 | 0 | 4.980E-03 | 3.150E-06 | 7.390E-05 | rs635168 |
| 6 | 11 | 85778924 | -4.517 | 3.744 | 1.050E-02 | 0 | 1.050E-02 | 6.280E-06 | 1.810E-04 | rs544458 |
| 6 | 11 | 85779297 | -4.699 | 3.921 | 5.760E-03 | 0 | 5.760E-03 | 2.620E-06 | 8.820E-05 | rs2509608 |
| 6 | 11 | 85779310 | -4.694 | 3.901 | 6.150E-03 | 0 | 6.150E-03 | 2.690E-06 | 9.570E-05 | rs2458500 |
| 6 | 11 | 85780448 | -4.335 | 3.808 | 1.190E-02 | 0 | 1.190E-02 | 1.460E-05 | 1.400E-04 | rs3016327 |
| 6 | 11 | 85781322 | -4.614 | 3.698 | 1.220E-02 | 0 | 1.220E-02 | 3.940E-06 | 2.180E-04 | rs669556 |
| 6 | 11 | 85783875 | -4.780 | 3.768 | 9.650E-03 | 0 | 9.650E-03 | 1.760E-06 | 1.650E-04 | rs2458493 |
| 7 | 15 | 82444437 | -4.015 | -4.484 | 4.340E-02 | 1 | 4.340E-02 | 5.940E-05 | 7.330E-06 | rs905450 |
| 8 | 16 | 20516594 | -4.066 | -7.115 | 3.570E-02 | 1 | 3.570E-02 | 4.780E-05 | 1.120E-12 | rs2158031 |
| 9 | 17 | 4761912 | 4.024 | 4.220 | 4.190E-02 | 0 | 4.190E-02 | 5.720E-05 | 2.440E-05 | rs8067447 |
| 9 | 17 | 4761960 | 4.020 | 4.230 | 4.260E-02 | 0 | 4.260E-02 | 5.820E-05 | 2.340E-05 | rs111985639 |
| 9 | 17 | 4762035 | 4.021 | 4.281 | 4.240E-02 | 0 | 4.240E-02 | 5.790E-05 | 1.860E-05 | rs16954223 |
| 9 | 17 | 4763551 | 4.016 | 4.295 | 4.330E-02 | 0 | 4.330E-02 | 5.930E-05 | 1.750E-05 | rs8078173 |
| 9 | 17 | 4765043 | 4.004 | 4.226 | 4.510E-02 | 0 | 4.510E-02 | 6.220E-05 | 2.380E-05 | rs75604955 |
| 9 | 17 | 4765765 | 3.998 | 4.196 | 4.620E-02 | 0 | 4.620E-02 | 6.400E-05 | 2.710E-05 | rs8078887 |
| 9 | 17 | 4766144 | 4.000 | 4.212 | 4.580E-02 | 0 | 4.580E-02 | 6.350E-05 | 2.530E-05 | rs76031775 |
| 9 | 17 | 4772761 | 3.996 | 4.280 | 4.640E-02 | 0 | 4.640E-02 | 6.430E-05 | 1.870E-05 | rs8077447 |
| 9 | 17 | 4772762 | 3.980 | 4.286 | 4.940E-02 | 0 | 4.940E-02 | 6.880E-05 | 1.820E-05 | rs8077448 |
| 9 | 17 | 4777634 | 4.003 | 4.275 | 4.530E-02 | 0 | 4.530E-02 | 6.260E-05 | 1.910E-05 | rs12150370 |
| 9 | 17 | 5133128 | 4.217 | 4.874 | 1.950E-02 | 1 | 1.950E-02 | 2.480E-05 | 1.090E-06 | rs61182333 |
| 10 | 19 | 45269725 | 9.907 | -3.566 | 1.860E-02 | 1 | 1.860E-02 | 3.900E-23 | 3.620E-04 | rs139326841 |

| cFDRlocusnum | CHR | BP | zscore_AD | zscore_eGFR | conjfdr_AD_eGFR | prune_AD_eGFR | min_conjfdr | pval_AD | pval_eGFR | rsID |
|---|---|---|---|---|---|---|---|---|---|---|
| 10 | 19 | 45296364 | -8.929 | 3.227 | 5.000E-02 | 0 | 5.000E-02 | 4.310E-19 | 1.250E-03 | rs112450640 |
| 10 | 19 | 45301924 | -7.952 | 3.365 | 3.430E-02 | 0 | 3.430E-02 | 1.840E-15 | 7.670E-04 | rs2965113 |
| 10 | 19 | 45301942 | -7.956 | 3.380 | 3.280E-02 | 0 | 3.280E-02 | 1.780E-15 | 7.240E-04 | rs2967669 |
| 10 | 19 | 45302513 | -8.193 | 3.566 | 1.870E-02 | 0 | 1.870E-02 | 2.540E-16 | 3.630E-04 | rs2965112 |
| 10 | 19 | 45302951 | -11.721 | 3.291 | 4.210E-02 | 0 | 4.210E-02 | 9.910E-32 | 9.980E-04 | rs2967668 |
| 10 | 19 | 45311084 | 11.223 | -3.539 | 2.030E-02 | 0 | 2.030E-02 | 3.160E-29 | 4.020E-04 | rs140824606 |
| 11 | 19 | 45313715 | -7.843 | 3.584 | 1.760E-02 | 1 | 1.760E-02 | 4.400E-15 | 3.380E-04 | rs2927477 |
| 12 | 19 | 45324138 | 22.851 | -3.926 | 5.660E-03 | 0 | 5.660E-03 | 1.410E-115 | 8.630E-05 | rs28399637 |
| 12 | 19 | 45325309 | -7.405 | 3.456 | 2.620E-02 | 0 | 2.620E-02 | 1.310E-13 | 5.480E-04 | rs8112196 |
| 12 | 19 | 45325391 | -7.014 | 3.500 | 2.300E-02 | 0 | 2.300E-02 | 2.320E-12 | 4.660E-04 | rs6509172 |
| 12 | 19 | 45326217 | 18.051 | -4.028 | 3.990E-03 | 0 | 3.990E-03 | 7.680E-73 | 5.620E-05 | rs10406338 |
| 12 | 19 | 45326664 | 18.001 | -4.038 | 3.850E-03 | 0 | 3.850E-03 | 1.910E-72 | 5.390E-05 | rs10405693 |
| 12 | 19 | 45326768 | -6.883 | 3.401 | 3.090E-02 | 0 | 3.090E-02 | 5.880E-12 | 6.710E-04 | rs1871045 |
| 12 | 19 | 45327309 | 17.950 | -4.073 | 3.410E-03 | 0 | 3.410E-03 | 4.770E-72 | 4.630E-05 | rs10412413 |
| 12 | 19 | 45328380 | 17.414 | -4.082 | 3.310E-03 | 0 | 3.310E-03 | 6.450E-68 | 4.470E-05 | rs58446550 |
| 12 | 19 | 45329214 | 17.786 | -4.132 | 2.770E-03 | 0 | 2.770E-03 | 9.030E-71 | 3.600E-05 | rs10402271 |
| 12 | 19 | 45329344 | 17.875 | -4.244 | 1.850E-03 | 0 | 1.850E-03 | 1.850E-71 | 2.200E-05 | rs10402524 |
| 12 | 19 | 45331725 | 18.399 | -4.073 | 3.410E-03 | 0 | 3.410E-03 | 1.330E-75 | 4.640E-05 | rs4802238 |
| 12 | 19 | 45336035 | 17.021 | -4.106 | 3.030E-03 | 0 | 3.030E-03 | 5.760E-65 | 4.030E-05 | rs7359852 |
| 12 | 19 | 45336443 | 16.914 | -4.211 | 2.080E-03 | 0 | 2.080E-03 | 3.560E-64 | 2.540E-05 | rs3021439 |
| 12 | 19 | 45337249 | 16.211 | -4.124 | 2.850E-03 | 0 | 2.850E-03 | 4.210E-59 | 3.720E-05 | rs2927481 |
| 12 | 19 | 45340728 | -7.850 | 3.482 | 2.410E-02 | 0 | 2.410E-02 | 4.160E-15 | 4.970E-04 | rs10409208 |
| 12 | 19 | 45340736 | 16.977 | -4.356 | 1.210E-03 | 0 | 1.210E-03 | 1.210E-64 | 1.330E-05 | rs73048293 |
| 12 | 19 | 45341540 | 16.928 | -4.296 | 1.520E-03 | 0 | 1.520E-03 | 2.810E-64 | 1.740E-05 | rs12462274 |
| 12 | 19 | 45341904 | 17.004 | -4.284 | 1.590E-03 | 0 | 1.590E-03 | 7.690E-65 | 1.830E-05 | rs12459575 |
| 12 | 19 | 45342114 | 17.060 | -4.221 | 2.000E-03 | 0 | 2.000E-03 | 2.940E-65 | 2.430E-05 | rs4803761 |
| 12 | 19 | 45342161 | 14.942 | -4.298 | 1.510E-03 | 0 | 1.510E-03 | 1.740E-50 | 1.730E-05 | rs4803762 |
| 12 | 19 | 45346768 | -9.245 | 4.119 | 2.890E-03 | 0 | 2.890E-03 | 2.350E-20 | 3.800E-05 | rs1001611 |
| 12 | 19 | 45347911 | 11.660 | -3.810 | 8.380E-03 | 0 | 8.380E-03 | 2.040E-31 | 1.390E-04 | rs4452060 |
| 12 | 19 | 453482 | -9.239 | 4.090 | 3.220E-03 | 0 | 3.220E-03 | 2.490E | 4.320E- | rs104264 |

| cFDRlocusnum | CHR | BP | zscore_AD | zscore_eGFR | conjfdr_AD_eGFR | prune_AD_eGFR | min_conjfdr | pval_AD | pval_eGFR | rsID |
|---|---|---|---|---|---|---|---|---|---|---|
| | | 53 | | | | | | -20 | 05 | 23 |
| 12 | 19 | 45348522 | 17.770 | -4.364 | 1.170E-03 | 0 | 1.170E-03 | 1.210E-70 | 1.280E-05 | rs12162222 |
| 12 | 19 | 45349402 | 12.689 | -3.830 | 7.830E-03 | 0 | 7.830E-03 | 6.770E-37 | 1.280E-04 | rs3810143 |
| 12 | 19 | 45349963 | -9.406 | 4.159 | 2.500E-03 | 0 | 2.500E-03 | 5.160E-21 | 3.190E-05 | rs2306149 |
| 12 | 19 | 45351746 | -9.367 | 4.180 | 2.330E-03 | 0 | 2.330E-03 | 7.480E-21 | 2.910E-05 | rs1871047 |
| 12 | 19 | 45351937 | -11.142 | 4.140 | 2.700E-03 | 0 | 2.700E-03 | 7.850E-29 | 3.480E-05 | rs1871046 |
| 12 | 19 | 45352804 | -9.614 | 4.083 | 3.290E-03 | 0 | 3.290E-03 | 7.010E-22 | 4.440E-05 | rs4802240 |
| 12 | 19 | 45354044 | 13.331 | -4.498 | 6.910E-04 | 0 | 6.910E-04 | 1.520E-40 | 6.860E-06 | rs57537848 |
| 12 | 19 | 45354296 | 13.142 | -4.531 | 6.050E-04 | 1 | 6.050E-04 | 1.900E-39 | 5.880E-06 | rs11666329 |
| 12 | 19 | 45355743 | -6.633 | 3.275 | 4.400E-02 | 0 | 4.400E-02 | 3.300E-11 | 1.060E-03 | rs4802241 |
| 12 | 19 | 45357377 | 19.809 | -4.343 | 1.270E-03 | 0 | 1.270E-03 | 2.480E-87 | 1.400E-05 | rs4803764 |
| 12 | 19 | 45359586 | 20.007 | -4.129 | 2.800E-03 | 0 | 2.800E-03 | 4.800E-89 | 3.640E-05 | rs56317818 |
| 12 | 19 | 45359706 | 19.898 | -4.087 | 3.250E-03 | 0 | 3.250E-03 | 4.250E-88 | 4.380E-05 | rs12462573 |
| 12 | 19 | 45361224 | -10.527 | 3.587 | 1.750E-02 | 0 | 1.750E-02 | 6.470E-26 | 3.350E-04 | rs440277 |
| 12 | 19 | 45369203 | -12.448 | 3.892 | 6.340E-03 | 0 | 6.340E-03 | 1.430E-35 | 9.950E-05 | rs112422902 |
| 12 | 19 | 45370335 | -4.104 | -4.584 | 3.080E-02 | 0 | 3.080E-02 | 4.060E-05 | 4.570E-06 | rs384653 |
| 13 | 19 | 45386634 | 21.708 | -3.941 | 5.380E-03 | 0 | 5.380E-03 | 1.720E-104 | 8.130E-05 | rs147636938 |
| 13 | 19 | 45387057 | -7.032 | 3.251 | 4.700E-02 | 0 | 4.700E-02 | 2.030E-12 | 1.150E-03 | rs283809 |
| 13 | 19 | 45389596 | -8.606 | 4.358 | 1.200E-03 | 0 | 1.200E-03 | 7.580E-18 | 1.310E-05 | rs7254892 |
| 13 | 19 | 45395266 | -25.803 | 5.279 | 2.210E-05 | 1 | 2.210E-05 | 8.130E-147 | 1.300E-07 | rs157580 |
| 13 | 19 | 45396973 | 21.890 | -4.092 | 3.190E-03 | 0 | 3.190E-03 | 3.200E-106 | 4.280E-05 | rs77301115 |
| 13 | 19 | 45397229 | -8.811 | 4.814 | 1.820E-04 | 0 | 1.820E-04 | 1.240E-18 | 1.480E-06 | rs1160983 |
| 13 | 19 | 45397307 | 21.116 | -4.113 | 2.960E-03 | 0 | 2.960E-03 | 5.730E-99 | 3.910E-05 | rs112849259 |
| 13 | 19 | 45397952 | 19.563 | -4.136 | 2.730E-03 | 0 | 2.730E-03 | 3.200E-85 | 3.530E-05 | rs116881820 |
| 13 | 19 | 45398785 | 21.143 | -4.061 | 3.550E-03 | 0 | 3.550E-03 | 3.190E-99 | 4.890E-05 | rs79398853 |
| 13 | 19 | 45399344 | 21.181 | -4.078 | 3.360E-03 | 0 | 3.360E-03 | 1.440E-99 | 4.550E-05 | rs75687619 |
| 13 | 19 | 45399896 | 21.153 | -4.232 | 1.930E-03 | 0 | 1.930E-03 | 2.590E-99 | 2.320E-05 | rs76366838 |
| 13 | 19 | 45400725 | 21.204 | -4.106 | 3.020E-03 | 0 | 3.020E-03 | 8.800E-100 | 4.020E-05 | rs114536010 |
| 13 | 19 | 45403216 | 21.190 | -4.563 | 5.270E-04 | 1 | 5.270E-04 | 1.180E-99 | 5.030E-06 | rs115881343 |
| 13 | 19 | 45404691 | -17.430 | 4.204 | 2.140E-03 | 0 | 2.140E-03 | 4.870E-68 | 2.630E-05 | rs405697 |

| cFDRlocusnum | CHR | BP | zscore_AD | zscore_eGFR | conjfdr_AD_eGFR | prune_AD_eGFR | min_conjfdr | pval_AD | pval_eGFR | rsID |
|---|---|---|---|---|---|---|---|---|---|---|
| 13 | 19 | 45404857 | 21.192 | -4.556 | 5.440E-04 | 0 | 5.440E-04 | 1.140E-99 | 5.210E-06 | rs112019714 |
| 13 | 19 | 45412079 | -14.839 | 4.891 | 1.300E-04 | 1 | 1.300E-04 | 8.140E-50 | 1.000E-06 | rs7412 |
| 13 | 19 | 45412955 | 21.261 | -4.151 | 2.590E-03 | 0 | 2.590E-03 | 2.630E-100 | 3.320E-05 | rs1081105 |
| 14 | 19 | 45425460 | -25.179 | 3.540 | 2.020E-02 | 0 | 2.020E-02 | 6.840E-140 | 4.000E-04 | rs157595 |
| 15 | 19 | 45426792 | -12.650 | 3.868 | 6.910E-03 | 0 | 6.910E-03 | 1.120E-36 | 1.100E-04 | rs141622900 |
| 16 | 19 | 46345886 | -4.059 | -5.073 | 3.660E-02 | 1 | 3.660E-02 | 4.920E-05 | 3.920E-07 | rs16980051 |

**Supplementary Table S17. Pleiotropic SNPs Identified by Conjunctional FDR in European Ancestry (Wightman Validation).**

This table presents all pleiotropic SNPs identified using the conjFDR method in the validation dataset, employing the Wightman AD GWAS. The validation analysis serves to confirm findings from the discovery phase and assess the robustness of pleiotropic signals across independent AD samples. The table structure parallels Table S16, reporting conjFDR locus numbers, chromosomal positions, z-scores for both traits, conjFDR values, pruning indicators, minimum conjFDR values, and individual trait p-values.

| cFDRlocusnum | CHR | BP | zscore_AD | zscore_eGFR | conjfdr_AD_eGFR | prune_AD_eGFR | min_conjfdr | pval_AD | pval_eGFR | rsID |
|---|---|---|---|---|---|---|---|---|---|---|
| 1 | 1 | 1003629 | -3.805 | 3.541 | 4.730E-02 | 0 | 4.730E-02 | 1.420E-04 | 3.990E-04 | rs4075116 |
| 1 | 1 | 1021695 | -3.907 | 3.719 | 3.440E-02 | 1 | 3.440E-02 | 9.330E-05 | 2.000E-04 | rs9442398 |
| 2 | 1 | 157674997 | -3.907 | 3.291 | 3.450E-02 | 0 | 3.450E-02 | 9.330E-05 | 1.000E-03 | rs2317230 |
| 2 | 1 | 157686337 | -4.079 | 3.340 | 2.280E-02 | 0 | 2.280E-02 | 4.510E-05 | 8.370E-04 | rs2317231 |
| 2 | 1 | 157705725 | -4.061 | 3.414 | 2.060E-02 | 1 | 2.060E-02 | 4.880E-05 | 6.410E-04 | rs6427397 |
| 3 | 1 | 200868862 | 3.985 | 3.259 | 2.900E-02 | 0 | 2.900E-02 | 6.740E-05 | 1.120E-03 | rs296526 |
| 3 | 1 | 200871876 | 4.315 | 3.346 | 2.230E-02 | 0 | 2.230E-02 | 1.600E-05 | 8.190E-04 | rs296529 |
| 3 | 1 | 200872484 | 4.393 | 3.355 | 2.170E-02 | 0 | 2.170E-02 | 1.120E-05 | 7.930E-04 | rs296530 |
| 3 | 1 | 200873170 | 4.373 | 3.366 | 2.100E-02 | 1 | 2.100E-02 | 1.230E-05 | 7.630E-04 | rs511104 |
| 3 | 1 | 200874728 | 4.030 | 3.882 | 2.300E-02 | 0 | 2.300E-02 | 5.580E-05 | 1.040E-04 | rs55838263 |
| 3 | 1 | 200875095 | 4.019 | 3.942 | 2.380E-02 | 0 | 2.380E-02 | 5.830E-05 | 8.070E-05 | rs12132298 |
| 3 | 1 | 200875897 | 4.023 | 3.954 | 2.360E-02 | 0 | 2.360E-02 | 5.750E-05 | 7.680E-05 | rs59655222 |
| 3 | 1 | 200877562 | 3.998 | 3.981 | 2.560E-02 | 0 | 2.560E-02 | 6.390E-05 | 6.870E-05 | rs7554511 |
| 3 | 1 | 200877850 | 4.044 | 3.967 | 2.190E-02 | 0 | 2.190E-02 | 5.260E-05 | 7.280E-05 | rs41299637 |
| 3 | 1 | 200878727 | 4.013 | 3.978 | 2.440E-02 | 0 | 2.440E-02 | 6.010E-05 | 6.940E-05 | rs12131796 |
| 3 | 1 | 200881595 | 3.946 | 4.101 | 3.030E-02 | 0 | 3.030E-02 | 7.940E-05 | 4.110E-05 | rs7522462 |

| cFDRlocusnum | CHR | BP | zscore_AD | zscore_eGFR | conjfdr_AD_eGFR | prune_AD_eGFR | min_conjfdr | pval_AD | pval_eGFR | rsID |
|---|---|---|---|---|---|---|---|---|---|---|
| 4 | 2 | 43762959 | -3.914 | 8.995 | 3.380E-02 | 1 | 3.380E-02 | 9.080E-05 | 2.350E-19 | rs78572881 |
| 5 | 2 | 238857362 | -3.906 | 3.722 | 3.460E-02 | 0 | 3.460E-02 | 9.380E-05 | 1.970E-04 | rs72979251 |
| 5 | 2 | 238858015 | -4.201 | 3.864 | 1.250E-02 | 0 | 1.250E-02 | 2.650E-05 | 1.120E-04 | rs59894893 |
| 5 | 2 | 238859288 | -4.226 | 3.863 | 1.140E-02 | 0 | 1.140E-02 | 2.380E-05 | 1.120E-04 | rs7607448 |
| 5 | 2 | 238859988 | -4.075 | 3.857 | 1.970E-02 | 0 | 1.970E-02 | 4.590E-05 | 1.150E-04 | rs76950125 |
| 5 | 2 | 238860370 | -4.161 | 3.831 | 1.440E-02 | 0 | 1.440E-02 | 3.160E-05 | 1.270E-04 | rs62194502 |
| 5 | 2 | 238860554 | -4.190 | 3.861 | 1.300E-02 | 0 | 1.300E-02 | 2.780E-05 | 1.130E-04 | rs11887040 |
| 5 | 2 | 238862031 | -4.129 | 3.773 | 1.630E-02 | 0 | 1.630E-02 | 3.650E-05 | 1.620E-04 | rs58075114 |
| 5 | 2 | 238863486 | -4.231 | 3.876 | 1.120E-02 | 1 | 1.120E-02 | 2.320E-05 | 1.060E-04 | rs7558958 |
| 5 | 2 | 238863518 | -4.208 | 3.869 | 1.220E-02 | 0 | 1.220E-02 | 2.570E-05 | 1.090E-04 | rs7602700 |
| 5 | 2 | 238864939 | -3.950 | 3.911 | 3.000E-02 | 0 | 3.000E-02 | 7.820E-05 | 9.200E-05 | rs62194505 |
| 5 | 2 | 238939370 | -4.063 | 3.867 | 2.050E-02 | 0 | 2.050E-02 | 4.840E-05 | 1.100E-04 | rs11679080 |
| 5 | 2 | 238946396 | -4.105 | 3.804 | 1.770E-02 | 0 | 1.770E-02 | 4.050E-05 | 1.430E-04 | rs57508757 |
| 5 | 2 | 238947982 | -3.975 | 3.919 | 2.770E-02 | 0 | 2.770E-02 | 7.040E-05 | 8.870E-05 | rs11695668 |
| 6 | 3 | 152008390 | -3.896 | -4.642 | 3.570E-02 | 0 | 3.570E-02 | 9.800E-05 | 3.450E-06 | rs12492498 |
| 6 | 3 | 152033705 | -3.992 | -4.859 | 2.610E-02 | 0 | 2.610E-02 | 6.540E-05 | 1.180E-06 | rs12486405 |
| 6 | 3 | 152054222 | -4.171 | -4.651 | 1.400E-02 | 0 | 1.400E-02 | 3.030E-05 | 3.310E-06 | rs6440796 |
| 6 | 3 | 152062241 | -3.988 | -4.770 | 2.650E-02 | 0 | 2.650E-02 | 6.660E-05 | 1.840E-06 | rs13059175 |
| 6 | 3 | 152073055 | -4.222 | -4.579 | 1.160E-02 | 1 | 1.160E-02 | 2.430E-05 | 4.660E-06 | rs3920075 |
| 6 | 3 | 152077625 | -4.193 | -4.607 | 1.290E-02 | 0 | 1.290E-02 | 2.750E-05 | 4.080E-06 | rs2055355 |
| 6 | 3 | 152085277 | -4.169 | -4.648 | 1.410E-02 | 0 | 1.410E-02 | 3.060E-05 | 3.350E-06 | rs13082076 |
| 6 | 3 | 152094201 | -3.802 | -4.592 | 4.770E-02 | 0 | 4.770E-02 | 1.440E-04 | 4.390E-06 | rs6768807 |
| 6 | 3 | 152095576 | -3.799 | -4.680 | 4.810E-02 | 0 | 4.810E-02 | 1.450E-04 | 2.870E-06 | rs12632591 |
| 6 | 3 | 152101725 | -3.835 | -4.585 | 4.310E-02 | 0 | 4.310E-02 | 1.260E-04 | 4.530E-06 | rs1553218 |
| 6 | 3 | 152183512 | -3.817 | -5.075 | 4.550E-02 | 0 | 4.550E-02 | 1.350E-04 | 3.880E-07 | rs7221 |
| 6 | 3 | 152183973 | -3.806 | -4.983 | 4.710E-02 | 0 | 4.710E-02 | 1.410E-04 | 6.270E-07 | rs13082695 |
| 7 | 5 | 140984454 | -3.839 | 3.170 | 4.520E-02 | 0 | 4.520E-02 | 1.240E-04 | 1.530E-03 | rs62380760 |
| 7 | 5 | 141043299 | -3.881 | 3.635 | 3.740E-02 | 0 | 3.740E-02 | 1.040E-04 | 2.780E-04 | rs17208502 |
| 7 | 5 | 141044179 | -3.857 | 3.657 | 4.040E-02 | 0 | 4.040E-02 | 1.150E-04 | 2.550E-04 | rs62380786 |
| 7 | 5 | 1410459 | -3.926 | 3.657 | 3.240E-02 | 1 | 3.240E-02 | 8.620E | 2.550E- | rs623807 |

| cFDRlocusnum | CHR | BP | zscore_AD | zscore_eGFR | conjfdr_AD_eGFR | prune_AD_eGFR | min_conjfdr | pval_AD | pval_eGFR | rsID |
|---|---|---|---|---|---|---|---|---|---|---|
| | | 76 | | | | | | -05 | 04 | 87 |
| 7 | 5 | 141050575 | -3.788 | 3.667 | 4.960E-02 | 0 | 4.960E-02 | 1.520E-04 | 2.460E-04 | rs62380789 |
| 8 | 6 | 34592731 | -4.078 | 3.097 | 4.570E-02 | 0 | 4.570E-02 | 4.550E-05 | 1.950E-03 | rs2744963 |
| 8 | 6 | 34608640 | -4.092 | 3.151 | 3.950E-02 | 0 | 3.950E-02 | 4.270E-05 | 1.630E-03 | rs2814989 |
| 8 | 6 | 34611000 | -4.044 | 3.226 | 3.190E-02 | 1 | 3.190E-02 | 5.260E-05 | 1.260E-03 | rs60917185 |
| 8 | 6 | 34612379 | -4.111 | 3.182 | 3.630E-02 | 0 | 3.630E-02 | 3.930E-05 | 1.460E-03 | rs2814990 |
| 8 | 6 | 34622991 | -4.290 | 3.084 | 4.740E-02 | 0 | 4.740E-02 | 1.790E-05 | 2.040E-03 | rs2814997 |
| 8 | 6 | 34625211 | -4.133 | 3.147 | 4.000E-02 | 0 | 4.000E-02 | 3.580E-05 | 1.650E-03 | rs2814998 |
| 8 | 6 | 34634263 | -4.051 | 3.195 | 3.500E-02 | 0 | 3.500E-02 | 5.110E-05 | 1.400E-03 | rs2815000 |
| 8 | 6 | 34634953 | -4.153 | 3.121 | 4.310E-02 | 0 | 4.310E-02 | 3.280E-05 | 1.800E-03 | rs2744948 |
| 8 | 6 | 34635809 | -4.118 | 3.131 | 4.190E-02 | 0 | 4.190E-02 | 3.820E-05 | 1.740E-03 | rs2815003 |
| 8 | 6 | 34645322 | -4.008 | 3.199 | 3.460E-02 | 0 | 3.460E-02 | 6.130E-05 | 1.380E-03 | rs2064253 |
| 8 | 6 | 34656462 | -4.000 | 3.138 | 4.110E-02 | 0 | 4.110E-02 | 6.330E-05 | 1.700E-03 | rs2744953 |
| 8 | 6 | 34658363 | -4.111 | 3.125 | 4.260E-02 | 0 | 4.260E-02 | 3.940E-05 | 1.780E-03 | rs2814950 |
| 8 | 6 | 34664207 | -4.054 | 3.146 | 4.020E-02 | 0 | 4.020E-02 | 5.040E-05 | 1.660E-03 | rs3800456 |
| 8 | 6 | 34668442 | -3.927 | 3.161 | 3.840E-02 | 0 | 3.840E-02 | 8.610E-05 | 1.570E-03 | rs9368829 |
| 8 | 6 | 34668694 | -4.048 | 3.127 | 4.240E-02 | 0 | 4.240E-02 | 5.170E-05 | 1.770E-03 | rs9380451 |
| 9 | 6 | 47640097 | 3.919 | 3.276 | 3.350E-02 | 0 | 3.350E-02 | 8.890E-05 | 1.050E-03 | rs869002 |
| 9 | 6 | 47647221 | 3.912 | 3.501 | 3.390E-02 | 0 | 3.390E-02 | 9.140E-05 | 4.640E-04 | rs10948374 |
| 9 | 6 | 47652510 | 3.867 | 3.169 | 4.150E-02 | 0 | 4.150E-02 | 1.100E-04 | 1.530E-03 | rs6900120 |
| 9 | 6 | 47653629 | 3.950 | 3.224 | 3.210E-02 | 0 | 3.210E-02 | 7.820E-05 | 1.260E-03 | rs10948375 |
| 9 | 6 | 47655633 | 4.494 | 3.087 | 4.700E-02 | 0 | 4.700E-02 | 6.990E-06 | 2.020E-03 | rs12212406 |
| 9 | 6 | 47656139 | 3.963 | 3.237 | 3.090E-02 | 0 | 3.090E-02 | 7.410E-05 | 1.210E-03 | rs9473157 |
| 9 | 6 | 47656852 | 3.949 | 3.246 | 3.080E-02 | 0 | 3.080E-02 | 7.860E-05 | 1.170E-03 | rs7770492 |
| 9 | 6 | 47660735 | 3.997 | 3.277 | 2.750E-02 | 0 | 2.750E-02 | 6.420E-05 | 1.050E-03 | rs1890666 |
| 9 | 6 | 47660777 | 4.014 | 3.283 | 2.700E-02 | 0 | 2.700E-02 | 5.970E-05 | 1.030E-03 | rs1890667 |
| 9 | 6 | 47662085 | 4.205 | 3.327 | 2.370E-02 | 0 | 2.370E-02 | 2.610E-05 | 8.770E-04 | rs7763444 |
| 9 | 6 | 47662484 | 4.190 | 3.420 | 1.770E-02 | 1 | 1.770E-02 | 2.780E-05 | 6.250E-04 | rs9381599 |
| 10 | 6 | 131311108 | 3.852 | -5.364 | 4.100E-02 | 1 | 4.100E-02 | 1.170E-04 | 8.130E-08 | rs7761311 |
| 11 | 7 | 28149761 | -3.814 | 3.649 | 4.590E-02 | 0 | 4.590E-02 | 1.370E-04 | 2.630E-04 | rs11771411 |

| cFDRlocusnum | CHR | BP | zscore_AD | zscore_eGFR | conjfdr_AD_eGFR | prune_AD_eGFR | min_conjfdr | pval_AD | pval_eGFR | rsID |
|---|---|---|---|---|---|---|---|---|---|---|
| 11 | 7 | 28150689 | -3.861 | 3.617 | 3.980E-02 | 0 | 3.980E-02 | 1.130E-04 | 2.980E-04 | rs10276070 |
| 11 | 7 | 28159696 | -3.885 | 3.645 | 3.700E-02 | 1 | 3.700E-02 | 1.020E-04 | 2.670E-04 | rs10275989 |
| 12 | 7 | 100159567 | -3.942 | 4.012 | 3.080E-02 | 1 | 3.080E-02 | 8.090E-05 | 6.020E-05 | rs2734897 |
| 12 | 7 | 100175473 | -3.898 | 3.619 | 3.540E-02 | 0 | 3.540E-02 | 9.680E-05 | 2.960E-04 | rs909152 |
| 13 | 8 | 145093404 | 3.844 | 3.602 | 4.190E-02 | 1 | 4.190E-02 | 1.210E-04 | 3.160E-04 | rs7835415 |
| 13 | 8 | 145108151 | 3.831 | 8.819 | 4.360E-02 | 0 | 4.360E-02 | 1.270E-04 | 1.160E-18 | rs61732533 |
| 13 | 8 | 145154222 | 3.802 | 8.485 | 4.770E-02 | 0 | 4.770E-02 | 1.440E-04 | 2.160E-17 | rs34674752 |
| 14 | 10 | 11489713 | -4.317 | -3.649 | 8.550E-03 | 0 | 8.550E-03 | 1.580E-05 | 2.630E-04 | rs7903896 |
| 14 | 10 | 11490281 | -4.658 | -3.769 | 5.800E-03 | 1 | 5.800E-03 | 3.190E-06 | 1.640E-04 | rs12783714 |
| 14 | 10 | 11492581 | -3.943 | -3.555 | 3.060E-02 | 0 | 3.060E-02 | 8.040E-05 | 3.780E-04 | rs11257096 |
| 14 | 10 | 11524026 | -4.577 | -3.172 | 3.730E-02 | 0 | 3.730E-02 | 4.730E-06 | 1.510E-03 | rs7921833 |
| 14 | 10 | 11524693 | -4.459 | -3.104 | 4.490E-02 | 0 | 4.490E-02 | 8.220E-06 | 1.910E-03 | rs11257121 |
| 14 | 10 | 11526277 | -4.404 | -3.085 | 4.730E-02 | 0 | 4.730E-02 | 1.060E-05 | 2.040E-03 | rs12360091 |
| 14 | 10 | 11529868 | -4.550 | -3.132 | 4.170E-02 | 0 | 4.170E-02 | 5.370E-06 | 1.730E-03 | rs12354448 |
| 15 | 10 | 82246749 | -4.505 | 4.394 | 3.830E-03 | 0 | 3.830E-03 | 6.630E-06 | 1.110E-05 | rs1870140 |
| 15 | 10 | 82249752 | -4.469 | 5.036 | 4.430E-03 | 0 | 4.430E-03 | 7.850E-06 | 4.750E-07 | rs10749609 |
| 15 | 10 | 82250831 | -4.576 | 4.878 | 2.860E-03 | 0 | 2.860E-03 | 4.740E-06 | 1.070E-06 | rs7097656 |
| 15 | 10 | 82251544 | -4.526 | 4.996 | 3.520E-03 | 0 | 3.520E-03 | 6.010E-06 | 5.850E-07 | rs7922621 |
| 15 | 10 | 82251769 | -4.541 | 4.993 | 3.310E-03 | 0 | 3.310E-03 | 5.590E-06 | 5.930E-07 | rs7910643 |
| 15 | 10 | 82253701 | -4.543 | 4.995 | 3.290E-03 | 0 | 3.290E-03 | 5.560E-06 | 5.890E-07 | rs7900536 |
| 15 | 10 | 82253984 | -4.585 | 4.895 | 2.760E-03 | 0 | 2.760E-03 | 4.540E-06 | 9.850E-07 | rs6586028 |
| 15 | 10 | 82254047 | -4.569 | 4.533 | 2.940E-03 | 0 | 2.940E-03 | 4.910E-06 | 5.830E-06 | rs6586030 |
| 15 | 10 | 82257570 | -4.539 | 4.993 | 3.340E-03 | 0 | 3.340E-03 | 5.650E-06 | 5.950E-07 | rs7896832 |
| 15 | 10 | 82265271 | -4.515 | 5.043 | 3.670E-03 | 0 | 3.670E-03 | 6.330E-06 | 4.580E-07 | rs9633740 |
| 15 | 10 | 82269461 | -4.531 | 5.164 | 3.450E-03 | 0 | 3.450E-03 | 5.860E-06 | 2.420E-07 | rs7080009 |
| 15 | 10 | 82269611 | -4.495 | 5.220 | 4.000E-03 | 0 | 4.000E-03 | 6.950E-06 | 1.790E-07 | rs1870138 |
| 15 | 10 | 82271341 | -4.283 | 5.147 | 9.220E-03 | 0 | 9.220E-03 | 1.850E-05 | 2.640E-07 | rs1870148 |
| 15 | 10 | 82273079 | -4.678 | 5.231 | 1.850E-03 | 1 | 1.850E-03 | 2.890E-06 | 1.680E-07 | rs10748526 |
| 15 | 10 | 82280137 | -4.596 | 5.175 | 2.630E-03 | 0 | 2.630E-03 | 4.310E-06 | 2.280E-07 | rs1878036 |
| 16 | 10 | 8231529 | 3.927 | -4.479 | 3.230E-02 | 0 | 3.230E-02 | 8.590E | 7.510E- | rs111860 |

| cFDRlocusnum | CHR | BP | zscore_AD | zscore_eGFR | conjfdr_AD_eGFR | prune_AD_eGFR | min_conjfdr | pval_AD | pval_eGFR | rsID |
|---|---|---|---|---|---|---|---|---|---|---|
| | | 0 | | | | | | -05 | 06 | 29 |
| 16 | 10 | 82317158 | 4.009 | -4.456 | 2.470E-02 | 1 | 2.470E-02 | 6.110E-05 | 8.340E-06 | rs12267227 |
| 16 | 10 | 82318802 | 3.913 | -4.438 | 3.390E-02 | 0 | 3.390E-02 | 9.130E-05 | 9.060E-06 | rs11186047 |
| 17 | 10 | 116577161 | -3.832 | 3.290 | 4.340E-02 | 1 | 4.340E-02 | 1.270E-04 | 1.000E-03 | rs4752256 |
| 18 | 10 | 124130086 | 3.849 | 3.720 | 4.140E-02 | 0 | 4.140E-02 | 1.190E-04 | 1.990E-04 | rs72830775 |
| 18 | 10 | 124131714 | 3.851 | 3.709 | 4.110E-02 | 0 | 4.110E-02 | 1.180E-04 | 2.080E-04 | rs72830777 |
| 18 | 10 | 124135862 | 3.871 | 3.703 | 3.870E-02 | 0 | 3.870E-02 | 1.080E-04 | 2.130E-04 | rs72830780 |
| 18 | 10 | 124139893 | 3.878 | 3.749 | 3.780E-02 | 0 | 3.780E-02 | 1.050E-04 | 1.770E-04 | rs9887977 |
| 18 | 10 | 124141387 | 3.907 | 3.709 | 3.450E-02 | 0 | 3.450E-02 | 9.330E-05 | 2.080E-04 | rs7914912 |
| 18 | 10 | 124143406 | 3.943 | 3.914 | 3.070E-02 | 0 | 3.070E-02 | 8.060E-05 | 9.070E-05 | rs72830784 |
| 18 | 10 | 124145772 | 3.844 | 3.879 | 4.190E-02 | 0 | 4.190E-02 | 1.210E-04 | 1.050E-04 | rs72830788 |
| 18 | 10 | 124148234 | 3.976 | 3.862 | 2.760E-02 | 0 | 2.760E-02 | 7.010E-05 | 1.130E-04 | rs72830792 |
| 18 | 10 | 124151184 | 3.979 | 3.904 | 2.730E-02 | 0 | 2.730E-02 | 6.920E-05 | 9.480E-05 | rs72830795 |
| 18 | 10 | 124155845 | 3.981 | 3.855 | 2.710E-02 | 0 | 2.710E-02 | 6.860E-05 | 1.160E-04 | rs10399971 |
| 18 | 10 | 124156852 | 4.014 | 3.769 | 2.430E-02 | 0 | 2.430E-02 | 5.970E-05 | 1.640E-04 | rs4752692 |
| 18 | 10 | 124158516 | 3.995 | 3.850 | 2.580E-02 | 0 | 2.580E-02 | 6.460E-05 | 1.180E-04 | rs72830799 |
| 18 | 10 | 124160106 | 4.000 | 3.883 | 2.530E-02 | 0 | 2.530E-02 | 6.320E-05 | 1.030E-04 | rs72830800 |
| 18 | 10 | 124161144 | 3.978 | 3.839 | 2.740E-02 | 0 | 2.740E-02 | 6.950E-05 | 1.240E-04 | rs17649042 |
| 18 | 10 | 124163611 | 4.067 | 3.865 | 2.020E-02 | 0 | 2.020E-02 | 4.760E-05 | 1.110E-04 | rs4752693 |
| 18 | 10 | 124164988 | 4.000 | 3.849 | 2.540E-02 | 0 | 2.540E-02 | 6.330E-05 | 1.190E-04 | rs72830801 |
| 18 | 10 | 124166835 | 4.184 | 3.768 | 1.330E-02 | 0 | 1.330E-02 | 2.860E-05 | 1.640E-04 | rs76935275 |
| 18 | 10 | 124177898 | 4.037 | 3.838 | 2.250E-02 | 0 | 2.250E-02 | 5.420E-05 | 1.240E-04 | rs72832707 |
| 18 | 10 | 124179096 | 4.023 | 3.850 | 2.360E-02 | 0 | 2.360E-02 | 5.760E-05 | 1.180E-04 | rs41307060 |
| 18 | 10 | 124179856 | 4.023 | 3.859 | 2.360E-02 | 0 | 2.360E-02 | 5.760E-05 | 1.140E-04 | rs72832710 |
| 18 | 10 | 124183181 | 4.043 | 3.840 | 2.200E-02 | 0 | 2.200E-02 | 5.280E-05 | 1.230E-04 | rs41317040 |
| 18 | 10 | 124184567 | 4.039 | 3.861 | 2.230E-02 | 0 | 2.230E-02 | 5.370E-05 | 1.130E-04 | rs9783213 |
| 18 | 10 | 124186127 | 3.811 | 3.763 | 4.640E-02 | 0 | 4.640E-02 | 1.380E-04 | 1.680E-04 | rs2292623 |
| 18 | 10 | 124187107 | 4.067 | 3.825 | 2.020E-02 | 0 | 2.020E-02 | 4.760E-05 | 1.310E-04 | rs952508 |
| 18 | 10 | 124188743 | 4.039 | 3.862 | 2.230E-02 | 0 | 2.230E-02 | 5.380E-05 | 1.130E-04 | rs72832715 |
| 18 | 10 | 124189113 | 4.040 | 3.881 | 2.220E-02 | 0 | 2.220E-02 | 5.350E-05 | 1.040E-04 | rs72832716 |

| cFDRlocusnum | CHR | BP | zscore_AD | zscore_eGFR | conjfdr_AD_eGFR | prune_AD_eGFR | min_conjfdr | pval_AD | pval_eGFR | rsID |
|---|---|---|---|---|---|---|---|---|---|---|
| 18 | 10 | 124190121 | 4.043 | 3.800 | 2.200E-02 | 0 | 2.200E-02 | 5.280E-05 | 1.450E-04 | rs17103541 |
| 18 | 10 | 124192245 | 4.046 | 3.852 | 2.180E-02 | 0 | 2.180E-02 | 5.220E-05 | 1.170E-04 | rs72832718 |
| 18 | 10 | 124192474 | 4.042 | 3.869 | 2.200E-02 | 0 | 2.200E-02 | 5.290E-05 | 1.090E-04 | rs10510111 |
| 18 | 10 | 124197243 | 4.066 | 3.774 | 2.030E-02 | 0 | 2.030E-02 | 4.780E-05 | 1.610E-04 | rs12098648 |
| 18 | 10 | 124197641 | 4.095 | 3.816 | 1.840E-02 | 0 | 1.840E-02 | 4.230E-05 | 1.360E-04 | rs111348404 |
| 18 | 10 | 124203648 | 4.269 | 3.338 | 2.300E-02 | 0 | 2.300E-02 | 1.960E-05 | 8.450E-04 | rs2421027 |
| 18 | 10 | 124205304 | 4.271 | 3.398 | 1.900E-02 | 0 | 1.900E-02 | 1.950E-05 | 6.780E-04 | rs72834437 |
| 18 | 10 | 124213016 | 4.467 | 3.360 | 2.140E-02 | 0 | 2.140E-02 | 7.930E-06 | 7.810E-04 | rs72834439 |
| 18 | 10 | 124214251 | 4.400 | 3.139 | 4.090E-02 | 0 | 4.090E-02 | 1.080E-05 | 1.690E-03 | rs10490923 |
| 18 | 10 | 124214921 | 4.387 | 3.191 | 3.540E-02 | 0 | 3.540E-02 | 1.150E-05 | 1.420E-03 | rs17623531 |
| 18 | 10 | 124216397 | 4.300 | 3.292 | 2.630E-02 | 0 | 2.630E-02 | 1.710E-05 | 9.940E-04 | rs36213074 |
| 18 | 10 | 124216410 | 4.310 | 3.234 | 3.120E-02 | 0 | 3.120E-02 | 1.630E-05 | 1.220E-03 | rs7088128 |
| 18 | 10 | 124224904 | 4.634 | 3.286 | 2.680E-02 | 0 | 2.680E-02 | 3.580E-06 | 1.020E-03 | rs72834444 |
| 18 | 10 | 124230397 | 4.488 | 3.383 | 2.000E-02 | 0 | 2.000E-02 | 7.180E-06 | 7.180E-04 | rs74895474 |
| 18 | 10 | 124232420 | 4.627 | 3.433 | 1.710E-02 | 0 | 1.710E-02 | 3.700E-06 | 5.980E-04 | rs72834448 |
| 18 | 10 | 124233587 | 4.591 | 3.412 | 1.820E-02 | 0 | 1.820E-02 | 4.400E-06 | 6.440E-04 | rs77802229 |
| 18 | 10 | 124234320 | 4.012 | 3.512 | 2.440E-02 | 0 | 2.440E-02 | 6.030E-05 | 4.450E-04 | rs4752699 |
| 18 | 10 | 124235059 | 4.588 | 3.304 | 2.540E-02 | 0 | 2.540E-02 | 4.470E-06 | 9.520E-04 | rs17696741 |
| 18 | 10 | 124235187 | 4.496 | 3.562 | 1.140E-02 | 1 | 1.140E-02 | 6.920E-06 | 3.690E-04 | rs79057333 |
| 18 | 10 | 124235226 | 4.491 | 3.504 | 1.360E-02 | 0 | 1.360E-02 | 7.080E-06 | 4.580E-04 | rs72834453 |
| 19 | 11 | 47229316 | 4.359 | 3.171 | 3.740E-02 | 0 | 3.740E-02 | 1.310E-05 | 1.520E-03 | rs11039130 |
| 19 | 11 | 47249223 | 4.297 | 5.759 | 8.720E-03 | 0 | 8.720E-03 | 1.730E-05 | 8.450E-09 | rs11039138 |
| 19 | 11 | 47250319 | 4.288 | 5.747 | 9.030E-03 | 0 | 9.030E-03 | 1.800E-05 | 9.090E-09 | rs11039139 |
| 19 | 11 | 47261260 | 4.179 | 5.978 | 1.360E-02 | 0 | 1.360E-02 | 2.930E-05 | 2.260E-09 | rs11988 |
| 19 | 11 | 47262840 | 4.195 | 5.874 | 1.280E-02 | 0 | 1.280E-02 | 2.730E-05 | 4.250E-09 | rs2167080 |
| 19 | 11 | 47276675 | 5.244 | 6.116 | 1.200E-04 | 0 | 1.200E-04 | 1.570E-07 | 9.570E-10 | rs11039149 |
| 19 | 11 | 47278502 | 5.242 | 5.996 | 1.210E-04 | 0 | 1.210E-04 | 1.590E-07 | 2.020E-09 | rs11039154 |
| 19 | 11 | 47294021 | 4.146 | 5.835 | 1.530E-02 | 0 | 1.530E-02 | 3.380E-05 | 5.380E-09 | rs11039159 |
| 19 | 11 | 47301225 | 5.203 | 5.982 | 1.500E-04 | 0 | 1.500E-04 | 1.970E-07 | 2.210E-09 | rs7944419 |
| 19 | 11 | 4730273 | 4.122 | 5.837 | 1.670E-02 | 0 | 1.670E-02 | 3.760E | 5.300E- | rs116066 |

| cFDRlocusnum | CHR | BP | zscore_AD | zscore_eGFR | conjfdr_AD_eGFR | prune_AD_eGFR | min_conjfdr | pval_AD | pval_eGFR | rsID |
|---|---|---|---|---|---|---|---|---|---|---|
| | | 3 | | | | | | -05 | 09 | 83 |
| 19 | 11 | 47303612 | 4.157 | 5.879 | 1.470E-02 | 0 | 1.470E-02 | 3.230E-05 | 4.120E-09 | rs11039162 |
| 19 | 11 | 47309373 | 4.257 | 5.863 | 1.010E-02 | 0 | 1.010E-02 | 2.070E-05 | 4.540E-09 | rs12802374 |
| 19 | 11 | 47312689 | 5.220 | 6.026 | 1.370E-04 | 0 | 1.370E-04 | 1.790E-07 | 1.680E-09 | rs11039165 |
| 19 | 11 | 47318157 | 4.147 | 5.848 | 1.530E-02 | 0 | 1.530E-02 | 3.370E-05 | 4.980E-09 | rs749067 |
| 19 | 11 | 47322327 | 4.238 | 5.831 | 1.090E-02 | 0 | 1.090E-02 | 2.250E-05 | 5.510E-09 | rs7116451 |
| 19 | 11 | 47346723 | 5.183 | 6.089 | 1.670E-04 | 0 | 1.670E-04 | 2.190E-07 | 1.140E-09 | rs11039182 |
| 19 | 11 | 47346940 | 4.514 | 6.648 | 3.680E-03 | 0 | 3.680E-03 | 6.350E-06 | 2.970E-11 | rs11039183 |
| 19 | 11 | 47361084 | 4.596 | 6.675 | 2.630E-03 | 0 | 2.630E-03 | 4.300E-06 | 2.480E-11 | rs2856653 |
| 19 | 11 | 47361838 | 4.878 | 6.711 | 7.560E-04 | 0 | 7.560E-04 | 1.070E-06 | 1.930E-11 | rs2596407 |
| 19 | 11 | 47363215 | 4.698 | 6.563 | 1.700E-03 | 0 | 1.700E-03 | 2.630E-06 | 5.270E-11 | rs2697923 |
| 19 | 11 | 47363285 | 4.545 | 6.664 | 3.250E-03 | 0 | 3.250E-03 | 5.480E-06 | 2.670E-11 | rs10838696 |
| 19 | 11 | 47374998 | -6.418 | -3.859 | 4.360E-03 | 0 | 4.360E-03 | 1.380E-10 | 1.140E-04 | rs2856661 |
| 19 | 11 | 47375193 | -6.532 | -4.039 | 2.420E-03 | 0 | 2.420E-03 | 6.480E-11 | 5.360E-05 | rs11039195 |
| 19 | 11 | 47376363 | -6.643 | -4.181 | 1.500E-03 | 0 | 1.500E-03 | 3.080E-11 | 2.910E-05 | rs3740686 |
| 19 | 11 | 47376448 | -6.659 | -4.129 | 1.800E-03 | 0 | 1.800E-03 | 2.750E-11 | 3.650E-05 | rs1057233 |
| 19 | 11 | 47378245 | 6.024 | 7.648 | 1.420E-06 | 0 | 1.420E-06 | 1.700E-09 | 2.040E-14 | rs3824869 |
| 19 | 11 | 47379407 | -6.637 | -4.158 | 1.620E-03 | 0 | 1.620E-03 | 3.210E-11 | 3.220E-05 | rs2084946 |
| 19 | 11 | 47379615 | -6.595 | -4.268 | 1.100E-03 | 0 | 1.100E-03 | 4.250E-11 | 1.970E-05 | rs1542321 |
| 19 | 11 | 47379646 | -6.391 | -4.132 | 1.780E-03 | 0 | 1.780E-03 | 1.650E-10 | 3.590E-05 | rs11039200 |
| 19 | 11 | 47380061 | -6.594 | -4.223 | 1.290E-03 | 0 | 1.290E-03 | 4.270E-11 | 2.410E-05 | rs10734557 |
| 19 | 11 | 47383275 | -6.550 | -4.446 | 5.690E-04 | 0 | 5.690E-04 | 5.760E-11 | 8.730E-06 | rs7105851 |
| 19 | 11 | 47384597 | -6.785 | -4.463 | 5.330E-04 | 0 | 5.330E-04 | 1.160E-11 | 8.090E-06 | rs11039203 |
| 19 | 11 | 47385194 | -6.800 | -4.477 | 5.050E-04 | 0 | 5.050E-04 | 1.050E-11 | 7.570E-06 | rs67116452 |
| 19 | 11 | 47385923 | -6.763 | -4.428 | 6.100E-04 | 0 | 6.100E-04 | 1.350E-11 | 9.520E-06 | rs10838698 |
| 19 | 11 | 47386073 | -6.768 | -4.397 | 6.870E-04 | 0 | 6.870E-04 | 1.300E-11 | 1.100E-05 | rs10838699 |
| 19 | 11 | 47387599 | -6.831 | -4.435 | 5.930E-04 | 0 | 5.930E-04 | 8.430E-12 | 9.200E-06 | rs935914 |
| 19 | 11 | 47388214 | 6.158 | 8.023 | 6.310E-07 | 0 | 6.310E-07 | 7.360E-10 | 1.030E-15 | rs12801188 |
| 19 | 11 | 47389223 | -6.749 | -4.520 | 4.270E-04 | 0 | 4.270E-04 | 1.490E-11 | 6.170E-06 | rs35996350 |
| 19 | 11 | 47389638 | 6.011 | 8.066 | 1.530E-06 | 0 | 1.530E-06 | 1.840E-09 | 7.280E-16 | rs34937994 |

| cFDRlocusnum | CHR | BP | zscore_AD | zscore_eGFR | conjfdr_AD_eGFR | prune_AD_eGFR | min_conjfdr | pval_AD | pval_eGFR | rsID |
|---|---|---|---|---|---|---|---|---|---|---|
| 19 | 11 | 47390692 | -6.812 | -4.384 | 7.200E-04 | 0 | 7.200E-04 | 9.620E-12 | 1.160E-05 | rs4752987 |
| 19 | 11 | 47391039 | -6.766 | -4.405 | 6.660E-04 | 0 | 6.660E-04 | 1.320E-11 | 1.060E-05 | rs10769258 |
| 19 | 11 | 47391745 | -6.830 | -4.492 | 4.780E-04 | 0 | 4.780E-04 | 8.520E-12 | 7.070E-06 | rs67472071 |
| 19 | 11 | 47391879 | -6.715 | -4.367 | 7.660E-04 | 0 | 7.660E-04 | 1.880E-11 | 1.260E-05 | rs7928163 |
| 19 | 11 | 47391948 | 6.292 | 8.818 | 2.740E-07 | 1 | 2.740E-07 | 3.130E-10 | 1.160E-18 | rs10437655 |
| 19 | 11 | 47392114 | -6.257 | -4.182 | 1.490E-03 | 0 | 1.490E-03 | 3.930E-10 | 2.880E-05 | rs7928419 |
| 19 | 11 | 47393638 | -6.699 | -4.413 | 6.430E-04 | 0 | 6.430E-04 | 2.100E-11 | 1.020E-05 | rs4992357 |
| 19 | 11 | 47394305 | 6.262 | 8.707 | 3.310E-07 | 0 | 3.310E-07 | 3.790E-10 | 3.130E-18 | rs896817 |
| 19 | 11 | 47394338 | -6.761 | -4.364 | 7.730E-04 | 0 | 7.730E-04 | 1.370E-11 | 1.280E-05 | rs896816 |
| 19 | 11 | 47395240 | -6.689 | -4.359 | 7.900E-04 | 0 | 7.900E-04 | 2.240E-11 | 1.310E-05 | rs7940536 |
| 19 | 11 | 47397353 | -6.453 | -4.219 | 1.310E-03 | 0 | 1.310E-03 | 1.100E-10 | 2.460E-05 | rs56030824 |
| 19 | 11 | 47398963 | -6.464 | -4.135 | 1.760E-03 | 0 | 1.760E-03 | 1.020E-10 | 3.550E-05 | rs12146565 |
| 19 | 11 | 47399602 | 5.923 | 8.014 | 2.570E-06 | 0 | 2.570E-06 | 3.170E-09 | 1.110E-15 | rs2278890 |
| 19 | 11 | 47400602 | 5.921 | 8.045 | 2.590E-06 | 0 | 2.590E-06 | 3.190E-09 | 8.620E-16 | rs3758675 |
| 19 | 11 | 47401058 | -6.506 | -4.524 | 4.220E-04 | 0 | 4.220E-04 | 7.730E-11 | 6.070E-06 | rs11601173 |
| 19 | 11 | 47401060 | -6.502 | -4.565 | 3.580E-04 | 0 | 3.580E-04 | 7.940E-11 | 4.990E-06 | rs11607981 |
| 19 | 11 | 47401171 | -6.505 | -4.512 | 4.400E-04 | 0 | 4.400E-04 | 7.770E-11 | 6.410E-06 | rs11601603 |
| 19 | 11 | 47406592 | -5.587 | -7.252 | 1.810E-05 | 0 | 1.810E-05 | 2.310E-08 | 4.100E-13 | rs11039216 |
| 19 | 11 | 47407439 | 5.847 | 8.086 | 4.010E-06 | 0 | 4.010E-06 | 5.000E-09 | 6.140E-16 | rs11606287 |
| 19 | 11 | 47408592 | 5.822 | 8.099 | 4.660E-06 | 0 | 4.660E-06 | 5.800E-09 | 5.540E-16 | rs35032070 |
| 19 | 11 | 47409212 | -6.457 | -4.374 | 7.450E-04 | 0 | 7.450E-04 | 1.060E-10 | 1.220E-05 | rs7111957 |
| 19 | 11 | 47410200 | -5.793 | -4.287 | 1.030E-03 | 0 | 1.030E-03 | 6.920E-09 | 1.810E-05 | rs6485753 |
| 19 | 11 | 47410393 | -6.488 | -4.459 | 5.420E-04 | 0 | 5.420E-04 | 8.680E-11 | 8.250E-06 | rs4752990 |
| 19 | 11 | 47410888 | 6.019 | 8.870 | 1.460E-06 | 0 | 1.460E-06 | 1.760E-09 | 7.320E-19 | rs10838702 |
| 19 | 11 | 47410951 | -6.508 | -4.344 | 8.380E-04 | 0 | 8.380E-04 | 7.630E-11 | 1.400E-05 | rs4752993 |
| 19 | 11 | 47411581 | -6.533 | -4.313 | 9.350E-04 | 0 | 9.350E-04 | 6.450E-11 | 1.610E-05 | rs10769262 |
| 19 | 11 | 47413475 | -6.444 | -4.457 | 5.460E-04 | 0 | 5.460E-04 | 1.160E-10 | 8.320E-06 | rs12803525 |
| 19 | 11 | 47413733 | -6.447 | -4.485 | 4.910E-04 | 0 | 4.910E-04 | 1.140E-10 | 7.290E-06 | rs11605672 |
| 19 | 11 | 47416636 | -5.786 | -4.470 | 5.170E-04 | 0 | 5.170E-04 | 7.210E-09 | 7.810E-06 | rs55876153 |
| 19 | 11 | 4741674 | 6.000 | 8.779 | 1.630E-06 | 0 | 1.630E-06 | 1.970E | 1.650E- | rs137741 |

| cFDRlocusnum | CHR | BP | zscore_AD | zscore_eGFR | conjfdr_AD_eGFR | prune_AD_eGFR | min_conjfdr | pval_AD | pval_eGFR | rsID |
|---|---|---|---|---|---|---|---|---|---|---|
| | | 6 | | | | | | -09 | 18 | 6 |
| 19 | 11 | 47417183 | 5.980 | 8.786 | 1.840E-06 | 0 | 1.840E-06 | 2.230E-09 | 1.550E-18 | rs10769263 |
| 19 | 11 | 47418306 | -6.507 | -4.345 | 8.320E-04 | 0 | 8.320E-04 | 7.690E-11 | 1.390E-05 | rs12802273 |
| 19 | 11 | 47419129 | -6.523 | -4.380 | 7.320E-04 | 0 | 7.320E-04 | 6.900E-11 | 1.190E-05 | rs7924485 |
| 19 | 11 | 47419301 | -6.530 | -4.303 | 9.730E-04 | 0 | 9.730E-04 | 6.560E-11 | 1.690E-05 | rs896815 |
| 19 | 11 | 47419663 | 5.991 | 8.762 | 1.720E-06 | 0 | 1.720E-06 | 2.080E-09 | 1.910E-18 | rs1534576 |
| 19 | 11 | 47419757 | -6.478 | -4.480 | 5.000E-04 | 0 | 5.000E-04 | 9.290E-11 | 7.470E-06 | rs1317164 |
| 19 | 11 | 47423340 | 5.989 | 8.815 | 1.740E-06 | 0 | 1.740E-06 | 2.110E-09 | 1.190E-18 | rs10769264 |
| 19 | 11 | 47423553 | -6.313 | -4.206 | 1.370E-03 | 0 | 1.370E-03 | 2.730E-10 | 2.600E-05 | rs4752832 |
| 19 | 11 | 47424400 | -6.466 | -4.467 | 5.240E-04 | 0 | 5.240E-04 | 1.010E-10 | 7.950E-06 | rs11039219 |
| 19 | 11 | 47424717 | -6.181 | -4.259 | 1.140E-03 | 0 | 1.140E-03 | 6.370E-10 | 2.050E-05 | rs10838704 |
| 19 | 11 | 47424931 | 6.007 | 8.805 | 1.560E-06 | 0 | 1.560E-06 | 1.890E-09 | 1.310E-18 | rs10838705 |
| 19 | 11 | 47426956 | -6.446 | -4.477 | 5.050E-04 | 0 | 5.050E-04 | 1.150E-10 | 7.570E-06 | rs112060627 |
| 19 | 11 | 47427739 | 6.005 | 8.792 | 1.590E-06 | 0 | 1.590E-06 | 1.920E-09 | 1.460E-18 | rs11039221 |
| 19 | 11 | 47428565 | -6.442 | -4.535 | 4.050E-04 | 0 | 4.050E-04 | 1.180E-10 | 5.770E-06 | rs4752999 |
| 19 | 11 | 47429904 | 6.007 | 8.794 | 1.560E-06 | 0 | 1.560E-06 | 1.880E-09 | 1.440E-18 | rs7947450 |
| 19 | 11 | 47430458 | -6.485 | -4.270 | 1.090E-03 | 0 | 1.090E-03 | 8.900E-11 | 1.950E-05 | rs7937331 |
| 19 | 11 | 47430599 | 5.995 | 8.777 | 1.670E-06 | 0 | 1.670E-06 | 2.030E-09 | 1.680E-18 | rs11039225 |
| 19 | 11 | 47431966 | -6.175 | -4.260 | 1.130E-03 | 0 | 1.130E-03 | 6.620E-10 | 2.040E-05 | rs755555 |
| 19 | 11 | 47432303 | -6.471 | -4.326 | 8.920E-04 | 0 | 8.920E-04 | 9.760E-11 | 1.520E-05 | rs755553 |
| 19 | 11 | 47432725 | 5.961 | 8.814 | 2.050E-06 | 0 | 2.050E-06 | 2.510E-09 | 1.200E-18 | rs10742802 |
| 19 | 11 | 47434777 | -6.454 | -4.254 | 1.160E-03 | 0 | 1.160E-03 | 1.090E-10 | 2.100E-05 | rs6485756 |
| 19 | 11 | 47434986 | 4.511 | 7.603 | 3.740E-03 | 0 | 3.740E-03 | 6.450E-06 | 2.890E-14 | rs2293576 |
| 19 | 11 | 47437202 | -6.426 | -4.304 | 9.670E-04 | 0 | 9.670E-04 | 1.310E-10 | 1.680E-05 | rs2293577 |
| 19 | 11 | 47437403 | -6.140 | -4.295 | 9.990E-04 | 0 | 9.990E-04 | 8.230E-10 | 1.740E-05 | rs2293578 |
| 19 | 11 | 47439444 | -6.473 | -4.469 | 5.210E-04 | 0 | 5.210E-04 | 9.610E-11 | 7.870E-06 | rs2053979 |
| 19 | 11 | 47439938 | -6.412 | -4.277 | 1.070E-03 | 0 | 1.070E-03 | 1.430E-10 | 1.890E-05 | rs10742803 |
| 19 | 11 | 47440758 | 6.054 | 8.782 | 1.190E-06 | 0 | 1.190E-06 | 1.410E-09 | 1.610E-18 | rs2293579 |
| 19 | 11 | 47443080 | -5.574 | -4.454 | 5.530E-04 | 0 | 5.530E-04 | 2.490E-08 | 8.440E-06 | rs3781625 |
| 19 | 11 | 47447346 | 6.028 | 8.777 | 1.390E-06 | 0 | 1.390E-06 | 1.660E-09 | 1.670E-18 | rs10838709 |

| cFDRlocusnum | CHR | BP | zscore_AD | zscore_eGFR | conjfdr_AD_eGFR | prune_AD_eGFR | min_conjfdr | pval_AD | pval_eGFR | rsID |
|---|---|---|---|---|---|---|---|---|---|---|
| 19 | 11 | 47448497 | -6.439 | -4.420 | 6.260E-04 | 0 | 6.260E-04 | 1.200E-10 | 9.850E-06 | rs11600581 |
| 19 | 11 | 47449072 | 6.041 | 8.776 | 1.280E-06 | 0 | 1.280E-06 | 1.530E-09 | 1.700E-18 | rs12292911 |
| 19 | 11 | 47449101 | -6.431 | -4.321 | 9.070E-04 | 0 | 9.070E-04 | 1.270E-10 | 1.550E-05 | rs60075622 |
| 19 | 11 | 47450338 | -6.391 | -4.449 | 5.640E-04 | 0 | 5.640E-04 | 1.650E-10 | 8.630E-06 | rs35496532 |
| 19 | 11 | 47457539 | -6.192 | -4.165 | 1.580E-03 | 0 | 1.580E-03 | 5.950E-10 | 3.110E-05 | rs11604680 |
| 19 | 11 | 47458765 | -6.413 | -4.285 | 1.040E-03 | 0 | 1.040E-03 | 1.430E-10 | 1.830E-05 | rs7106956 |
| 19 | 11 | 47460901 | -6.041 | -4.189 | 1.460E-03 | 0 | 1.460E-03 | 1.540E-09 | 2.810E-05 | rs11039238 |
| 19 | 11 | 47461693 | 4.420 | 6.954 | 5.390E-03 | 0 | 5.390E-03 | 9.860E-06 | 3.550E-12 | rs7103835 |
| 19 | 11 | 47461783 | 6.018 | 8.802 | 1.470E-06 | 0 | 1.470E-06 | 1.760E-09 | 1.350E-18 | rs7103648 |
| 19 | 11 | 47462140 | 6.034 | 8.810 | 1.330E-06 | 0 | 1.330E-06 | 1.600E-09 | 1.250E-18 | rs7104036 |
| 19 | 11 | 47466006 | -6.341 | -4.254 | 1.160E-03 | 0 | 1.160E-03 | 2.280E-10 | 2.100E-05 | rs4752838 |
| 19 | 11 | 47466442 | 4.063 | 7.544 | 2.050E-02 | 0 | 2.050E-02 | 4.840E-05 | 4.560E-14 | rs11039244 |
| 19 | 11 | 47474146 | 5.130 | 5.797 | 2.190E-04 | 0 | 2.190E-04 | 2.890E-07 | 6.760E-09 | rs12361415 |
| 19 | 11 | 47475189 | -6.825 | -3.329 | 2.360E-02 | 0 | 2.360E-02 | 8.780E-12 | 8.720E-04 | rs35184771 |
| 19 | 11 | 47481533 | 4.712 | 5.300 | 1.600E-03 | 0 | 1.600E-03 | 2.460E-06 | 1.160E-07 | rs7950696 |
| 19 | 11 | 47486885 | -6.826 | -3.394 | 1.930E-02 | 0 | 1.930E-02 | 8.710E-12 | 6.890E-04 | rs1317149 |
| 19 | 11 | 47495746 | -6.627 | -3.322 | 2.410E-02 | 0 | 2.410E-02 | 3.430E-11 | 8.950E-04 | rs11039255 |
| 19 | 11 | 47500267 | 4.620 | 5.383 | 2.380E-03 | 0 | 2.380E-03 | 3.840E-06 | 7.330E-08 | rs2242081 |
| 19 | 11 | 47500400 | 5.250 | 6.308 | 1.170E-04 | 0 | 1.170E-04 | 1.520E-07 | 2.820E-10 | rs12224672 |
| 19 | 11 | 47504075 | 4.611 | 5.167 | 2.460E-03 | 0 | 2.460E-03 | 4.000E-06 | 2.380E-07 | rs3740699 |
| 19 | 11 | 47530024 | 5.304 | 6.833 | 8.740E-05 | 0 | 8.740E-05 | 1.130E-07 | 8.310E-12 | rs6485758 |
| 19 | 11 | 47532395 | 5.142 | 6.225 | 2.060E-04 | 0 | 2.060E-04 | 2.720E-07 | 4.810E-10 | rs11039266 |
| 19 | 11 | 47539697 | -6.677 | -3.356 | 2.160E-02 | 0 | 2.160E-02 | 2.440E-11 | 7.910E-04 | rs4752845 |
| 19 | 11 | 47547046 | -6.689 | -3.320 | 2.420E-02 | 0 | 2.420E-02 | 2.240E-11 | 9.000E-04 | rs34958982 |
| 19 | 11 | 47554337 | 4.593 | 5.318 | 2.660E-03 | 0 | 2.660E-03 | 4.360E-06 | 1.050E-07 | rs10742814 |
| 19 | 11 | 47557871 | 5.299 | 6.878 | 9.010E-05 | 0 | 9.010E-05 | 1.170E-07 | 6.070E-12 | rs10838725 |
| 19 | 11 | 47558858 | 4.632 | 5.287 | 2.260E-03 | 0 | 2.260E-03 | 3.620E-06 | 1.250E-07 | rs11039280 |
| 19 | 11 | 47560433 | 5.218 | 6.308 | 1.390E-04 | 0 | 1.390E-04 | 1.810E-07 | 2.820E-10 | rs61895112 |
| 19 | 11 | 47564785 | 5.115 | 6.191 | 2.350E-04 | 0 | 2.350E-04 | 3.130E-07 | 5.990E-10 | rs11039284 |
| 19 | 11 | 4756807 | -6.695 | -3.305 | 2.540E-02 | 0 | 2.540E-02 | 2.160E | 9.510E- | rs667494 |

| cFDRlocusnum | CHR | BP | zscore_AD | zscore_eGFR | conjfdr_AD_eGFR | prune_AD_eGFR | min_conjfdr | pval_AD | pval_eGFR | rsID |
|---|---|---|---|---|---|---|---|---|---|---|
| | | 4 | | | | | | -11 | 04 | 09 |
| 19 | 11 | 47571808 | 4.351 | 5.066 | 7.110E-03 | 0 | 7.110E-03 | 1.360E-05 | 4.060E-07 | rs12226590 |
| 19 | 11 | 47571967 | 5.208 | 6.235 | 1.460E-04 | 0 | 1.460E-04 | 1.910E-07 | 4.510E-10 | rs12225051 |
| 19 | 11 | 47572279 | 5.265 | 6.846 | 1.080E-04 | 0 | 1.080E-04 | 1.400E-07 | 7.620E-12 | rs11039290 |
| 19 | 11 | 47573342 | 5.249 | 6.826 | 1.170E-04 | 0 | 1.170E-04 | 1.530E-07 | 8.720E-12 | rs71475921 |
| 19 | 11 | 47581443 | 5.257 | 6.923 | 1.130E-04 | 0 | 1.130E-04 | 1.460E-07 | 4.440E-12 | rs11039297 |
| 19 | 11 | 47583121 | -6.669 | -3.313 | 2.470E-02 | 0 | 2.470E-02 | 2.580E-11 | 9.230E-04 | rs12798346 |
| 19 | 11 | 47585348 | 4.067 | 4.753 | 2.020E-02 | 0 | 2.020E-02 | 4.760E-05 | 2.010E-06 | rs7126378 |
| 19 | 11 | 47586376 | -6.661 | -3.295 | 2.600E-02 | 0 | 2.600E-02 | 2.730E-11 | 9.830E-04 | rs56400411 |
| 19 | 11 | 47589707 | -6.465 | -3.305 | 2.540E-02 | 0 | 2.540E-02 | 1.010E-10 | 9.510E-04 | rs7945473 |
| 19 | 11 | 47600438 | 5.103 | 6.284 | 2.510E-04 | 0 | 2.510E-04 | 3.340E-07 | 3.300E-10 | rs2280231 |
| 19 | 11 | 47602729 | -6.596 | -3.321 | 2.420E-02 | 0 | 2.420E-02 | 4.220E-11 | 8.990E-04 | rs2030166 |
| 19 | 11 | 47606483 | -6.811 | -3.566 | 1.120E-02 | 0 | 1.120E-02 | 9.670E-12 | 3.620E-04 | rs11605348 |
| 19 | 11 | 47607135 | 5.328 | 6.822 | 7.660E-05 | 0 | 7.660E-05 | 9.940E-08 | 8.990E-12 | rs10838731 |
| 19 | 11 | 47622139 | -6.671 | -3.299 | 2.580E-02 | 0 | 2.580E-02 | 2.540E-11 | 9.720E-04 | rs12799623 |
| 19 | 11 | 47623890 | -6.482 | -3.432 | 1.710E-02 | 0 | 1.710E-02 | 9.030E-11 | 5.980E-04 | rs12363232 |
| 19 | 11 | 47624714 | -6.656 | -3.300 | 2.570E-02 | 0 | 2.570E-02 | 2.810E-11 | 9.650E-04 | rs12419692 |
| 19 | 11 | 47626492 | 4.626 | 6.822 | 2.320E-03 | 0 | 2.320E-03 | 3.730E-06 | 8.980E-12 | rs10769282 |
| 19 | 11 | 47627302 | 4.618 | 5.234 | 2.400E-03 | 0 | 2.400E-03 | 3.880E-06 | 1.660E-07 | rs6485763 |
| 19 | 11 | 47648042 | -6.547 | -3.328 | 2.370E-02 | 0 | 2.370E-02 | 5.870E-11 | 8.760E-04 | rs4752856 |
| 19 | 11 | 47655380 | 4.626 | 5.288 | 2.320E-03 | 0 | 2.320E-03 | 3.740E-06 | 1.240E-07 | rs7945312 |
| 19 | 11 | 47655752 | -6.585 | -3.314 | 2.470E-02 | 0 | 2.470E-02 | 4.530E-11 | 9.190E-04 | rs4752857 |
| 19 | 11 | 47658314 | -5.897 | -3.280 | 2.730E-02 | 0 | 2.730E-02 | 3.690E-09 | 1.040E-03 | rs12794570 |
| 19 | 11 | 47659135 | 5.145 | 6.255 | 2.030E-04 | 0 | 2.030E-04 | 2.670E-07 | 3.970E-10 | rs7118178 |
| 19 | 11 | 47662932 | 5.304 | 6.811 | 8.750E-05 | 0 | 8.750E-05 | 1.130E-07 | 9.720E-12 | rs7120548 |
| 19 | 11 | 47663049 | -6.662 | -3.339 | 2.290E-02 | 0 | 2.290E-02 | 2.700E-11 | 8.420E-04 | rs10838738 |
| 19 | 11 | 47669249 | 4.530 | 5.383 | 3.460E-03 | 0 | 3.460E-03 | 5.890E-06 | 7.310E-08 | rs7107792 |
| 19 | 11 | 47669289 | 4.588 | 5.252 | 2.730E-03 | 0 | 2.730E-03 | 4.480E-06 | 1.500E-07 | rs7107922 |
| 19 | 11 | 47669665 | -6.483 | -3.155 | 3.910E-02 | 0 | 3.910E-02 | 8.990E-11 | 1.610E-03 | rs12787646 |
| 19 | 11 | 47672743 | 4.519 | 5.410 | 3.620E-03 | 0 | 3.620E-03 | 6.220E-06 | 6.290E-08 | rs10838739 |

| cFDRlocusnum | CHR | BP | zscore_AD | zscore_eGFR | conjfdr_AD_eGFR | prune_AD_eGFR | min_conjfdr | pval_AD | pval_eGFR | rsID |
|---|---|---|---|---|---|---|---|---|---|---|
| 19 | 11 | 47674101 | 4.585 | 5.410 | 2.760E-03 | 0 | 2.760E-03 | 4.530E-06 | 6.300E-08 | rs2869024 |
| 19 | 11 | 47674934 | 4.691 | 5.253 | 1.760E-03 | 0 | 1.760E-03 | 2.720E-06 | 1.490E-07 | rs7103013 |
| 19 | 11 | 47675316 | 4.621 | 5.329 | 2.370E-03 | 0 | 2.370E-03 | 3.830E-06 | 9.860E-08 | rs7119517 |
| 19 | 11 | 47676085 | 4.411 | 5.195 | 5.600E-03 | 0 | 5.600E-03 | 1.030E-05 | 2.040E-07 | rs7107535 |
| 19 | 11 | 47676170 | 6.006 | 6.277 | 1.580E-06 | 0 | 1.580E-06 | 1.900E-09 | 3.450E-10 | rs7107356 |
| 19 | 11 | 47679980 | 4.616 | 5.239 | 2.410E-03 | 0 | 2.410E-03 | 3.910E-06 | 1.620E-07 | rs10838740 |
| 19 | 11 | 47681370 | 4.486 | 5.393 | 4.140E-03 | 0 | 4.140E-03 | 7.240E-06 | 6.940E-08 | rs1056387 |
| 19 | 11 | 47682402 | 4.705 | 5.164 | 1.650E-03 | 0 | 1.650E-03 | 2.540E-06 | 2.420E-07 | rs7933184 |
| 19 | 11 | 47682671 | 4.600 | 5.266 | 2.580E-03 | 0 | 2.580E-03 | 4.220E-06 | 1.390E-07 | rs12362318 |
| 19 | 11 | 47683765 | 4.661 | 5.236 | 1.990E-03 | 0 | 1.990E-03 | 3.150E-06 | 1.640E-07 | rs10838741 |
| 19 | 11 | 47684908 | 4.655 | 5.180 | 2.050E-03 | 0 | 2.050E-03 | 3.240E-06 | 2.210E-07 | rs11039329 |
| 19 | 11 | 47685354 | 4.786 | 5.312 | 1.150E-03 | 0 | 1.150E-03 | 1.710E-06 | 1.080E-07 | rs4543938 |
| 19 | 11 | 47685829 | 4.586 | 5.381 | 2.750E-03 | 0 | 2.750E-03 | 4.520E-06 | 7.410E-08 | rs4752858 |
| 19 | 11 | 47689343 | 4.597 | 5.349 | 2.620E-03 | 0 | 2.620E-03 | 4.280E-06 | 8.860E-08 | rs10742817 |
| 19 | 11 | 47694699 | 4.577 | 5.432 | 2.850E-03 | 0 | 2.850E-03 | 4.710E-06 | 5.580E-08 | rs4539273 |
| 19 | 11 | 47695508 | 4.210 | 5.148 | 1.210E-02 | 0 | 1.210E-02 | 2.560E-05 | 2.640E-07 | rs7116970 |
| 19 | 11 | 47695840 | 5.345 | 6.179 | 7.020E-05 | 0 | 7.020E-05 | 9.060E-08 | 6.450E-10 | rs11039332 |
| 19 | 11 | 47700168 | 5.270 | 6.078 | 1.050E-04 | 0 | 1.050E-04 | 1.360E-07 | 1.220E-09 | rs147311748 |
| 19 | 11 | 47701528 | 4.569 | 5.443 | 2.940E-03 | 0 | 2.940E-03 | 4.900E-06 | 5.250E-08 | rs12286721 |
| 19 | 11 | 47701819 | 4.576 | 5.243 | 2.860E-03 | 0 | 2.860E-03 | 4.740E-06 | 1.580E-07 | rs7105791 |
| 19 | 11 | 47703943 | 4.597 | 5.022 | 2.620E-03 | 0 | 2.620E-03 | 4.290E-06 | 5.100E-07 | rs3758655 |
| 19 | 11 | 47704200 | 4.607 | 4.988 | 2.510E-03 | 0 | 2.510E-03 | 4.090E-06 | 6.090E-07 | rs7936948 |
| 19 | 11 | 47705015 | 4.497 | 5.183 | 3.970E-03 | 0 | 3.970E-03 | 6.900E-06 | 2.180E-07 | rs4752860 |
| 19 | 11 | 47706086 | 4.539 | 5.055 | 3.340E-03 | 0 | 3.340E-03 | 5.650E-06 | 4.300E-07 | rs10838745 |
| 19 | 11 | 47706141 | 4.633 | 5.205 | 2.250E-03 | 0 | 2.250E-03 | 3.600E-06 | 1.940E-07 | rs11039336 |
| 19 | 11 | 47706291 | 4.652 | 4.955 | 2.070E-03 | 0 | 2.070E-03 | 3.290E-06 | 7.250E-07 | rs10838746 |
| 19 | 11 | 47707156 | 4.653 | 4.988 | 2.060E-03 | 0 | 2.060E-03 | 3.270E-06 | 6.100E-07 | rs11039337 |
| 19 | 11 | 47709212 | 4.681 | 5.008 | 1.840E-03 | 0 | 1.840E-03 | 2.860E-06 | 5.500E-07 | rs10769285 |
| 19 | 11 | 47715524 | 4.556 | 4.904 | 3.110E-03 | 0 | 3.110E-03 | 5.220E-06 | 9.370E-07 | rs7943213 |
| 19 | 11 | 4772022 | 4.574 | 5.049 | 2.890E-03 | 0 | 2.890E-03 | 4.800E | 4.440E- | rs110393 |

| cFDRlocusnum | CHR | BP | zscore_AD | zscore_eGFR | conjfdr_AD_eGFR | prune_AD_eGFR | min_conjfdr | pval_AD | pval_eGFR | rsID |
|---|---|---|---|---|---|---|---|---|---|---|
| | | 8 | | | | | | -06 | 07 | 45 |
| 19 | 11 | 47720509 | 4.746 | 4.840 | 1.380E-03 | 0 | 1.380E-03 | 2.080E-06 | 1.300E-06 | rs7942074 |
| 19 | 11 | 47722798 | 4.688 | 4.838 | 1.780E-03 | 0 | 1.780E-03 | 2.760E-06 | 1.310E-06 | rs7935521 |
| 19 | 11 | 47722819 | 4.764 | 4.682 | 1.270E-03 | 0 | 1.270E-03 | 1.900E-06 | 2.840E-06 | rs7935528 |
| 19 | 11 | 47723938 | 4.618 | 5.049 | 2.400E-03 | 0 | 2.400E-03 | 3.880E-06 | 4.430E-07 | rs7939420 |
| 19 | 11 | 47729770 | 4.741 | 4.914 | 1.400E-03 | 0 | 1.400E-03 | 2.120E-06 | 8.910E-07 | rs12785949 |
| 19 | 11 | 47733494 | 5.289 | 4.817 | 1.310E-04 | 0 | 1.310E-04 | 1.230E-07 | 1.450E-06 | rs1017730 |
| 19 | 11 | 47734444 | 4.649 | 4.904 | 2.100E-03 | 0 | 2.100E-03 | 3.340E-06 | 9.410E-07 | rs7111509 |
| 19 | 11 | 47740662 | 4.811 | 4.920 | 1.030E-03 | 0 | 1.030E-03 | 1.500E-06 | 8.660E-07 | rs2869534 |
| 19 | 11 | 47744394 | 4.712 | 4.925 | 1.600E-03 | 0 | 1.600E-03 | 2.450E-06 | 8.420E-07 | rs10838751 |
| 19 | 11 | 47745374 | 4.703 | 4.962 | 1.660E-03 | 0 | 1.660E-03 | 2.560E-06 | 6.990E-07 | rs7102205 |
| 19 | 11 | 47753370 | 4.640 | 5.126 | 2.190E-03 | 0 | 2.190E-03 | 3.490E-06 | 2.960E-07 | rs2290851 |
| 19 | 11 | 47756568 | 4.753 | 4.924 | 1.330E-03 | 0 | 1.330E-03 | 2.010E-06 | 8.480E-07 | rs1563821 |
| 19 | 11 | 47758531 | 4.619 | 5.006 | 2.380E-03 | 0 | 2.380E-03 | 3.850E-06 | 5.560E-07 | rs10838753 |
| 19 | 11 | 47758532 | 4.618 | 5.021 | 2.400E-03 | 0 | 2.400E-03 | 3.880E-06 | 5.140E-07 | rs10838754 |
| 19 | 11 | 47758675 | 4.620 | 5.137 | 2.380E-03 | 0 | 2.380E-03 | 3.850E-06 | 2.790E-07 | rs7116320 |
| 19 | 11 | 47759841 | 4.653 | 5.142 | 2.060E-03 | 0 | 2.060E-03 | 3.270E-06 | 2.720E-07 | rs4752865 |
| 19 | 11 | 47760078 | 5.241 | 5.920 | 1.220E-04 | 0 | 1.220E-04 | 1.600E-07 | 3.210E-09 | rs12807014 |
| 19 | 11 | 47763509 | 4.669 | 4.919 | 1.920E-03 | 0 | 1.920E-03 | 3.020E-06 | 8.680E-07 | rs6485773 |
| 19 | 11 | 47769620 | 5.146 | 5.349 | 2.020E-04 | 0 | 2.020E-04 | 2.670E-07 | 8.830E-08 | rs11039364 |
| 19 | 11 | 47769936 | 4.688 | 4.930 | 1.780E-03 | 0 | 1.780E-03 | 2.760E-06 | 8.240E-07 | rs7932703 |
| 19 | 11 | 47771476 | 4.758 | 4.783 | 1.300E-03 | 0 | 1.300E-03 | 1.950E-06 | 1.730E-06 | rs10838758 |
| 19 | 11 | 47772208 | 4.612 | 5.143 | 2.450E-03 | 0 | 2.450E-03 | 3.990E-06 | 2.700E-07 | rs60833259 |
| 19 | 11 | 47773518 | 4.618 | 5.132 | 2.390E-03 | 0 | 2.390E-03 | 3.870E-06 | 2.870E-07 | rs6485775 |
| 19 | 11 | 47774238 | 5.263 | 5.915 | 1.090E-04 | 0 | 1.090E-04 | 1.410E-07 | 3.320E-09 | rs12577383 |
| 19 | 11 | 47776283 | 4.685 | 4.935 | 1.800E-03 | 0 | 1.800E-03 | 2.810E-06 | 8.020E-07 | rs7929014 |
| 19 | 11 | 47779045 | 4.527 | 5.023 | 3.510E-03 | 0 | 3.510E-03 | 5.990E-06 | 5.100E-07 | rs12360643 |
| 19 | 11 | 47780222 | 4.627 | 5.148 | 2.310E-03 | 0 | 2.310E-03 | 3.710E-06 | 2.640E-07 | rs12421563 |
| 19 | 11 | 47780754 | 5.064 | 5.958 | 3.060E-04 | 0 | 3.060E-04 | 4.110E-07 | 2.550E-09 | rs7927445 |
| 19 | 11 | 47781057 | 4.628 | 5.143 | 2.300E-03 | 0 | 2.300E-03 | 3.690E-06 | 2.700E-07 | rs10742820 |

| cFDRlocusnum | CHR | BP | zscore_AD | zscore_eGFR | conjfdr_AD_eGFR | prune_AD_eGFR | min_conjfdr | pval_AD | pval_eGFR | rsID |
|---|---|---|---|---|---|---|---|---|---|---|
| 19 | 11 | 47781460 | 4.702 | 4.952 | 1.670E-03 | 0 | 1.670E-03 | 2.580E-06 | 7.360E-07 | rs7108479 |
| 19 | 11 | 47782076 | 4.692 | 4.943 | 1.740E-03 | 0 | 1.740E-03 | 2.700E-06 | 7.680E-07 | rs6485778 |
| 19 | 11 | 47785347 | 3.932 | 4.858 | 3.180E-02 | 0 | 3.180E-02 | 8.410E-05 | 1.190E-06 | rs10838759 |
| 19 | 11 | 47786414 | 4.679 | 4.933 | 1.850E-03 | 0 | 1.850E-03 | 2.890E-06 | 8.090E-07 | rs10838760 |
| 19 | 11 | 47787445 | 4.759 | 4.944 | 1.300E-03 | 0 | 1.300E-03 | 1.950E-06 | 7.660E-07 | rs7111576 |
| 19 | 11 | 47787538 | 4.702 | 4.940 | 1.660E-03 | 0 | 1.660E-03 | 2.570E-06 | 7.830E-07 | rs7111606 |
| 19 | 11 | 47789082 | 5.232 | 5.936 | 1.280E-04 | 0 | 1.280E-04 | 1.670E-07 | 2.930E-09 | rs12223593 |
| 19 | 11 | 47789549 | 4.700 | 4.932 | 1.690E-03 | 0 | 1.690E-03 | 2.610E-06 | 8.140E-07 | rs11039376 |
| 19 | 11 | 47789776 | 4.654 | 5.054 | 2.050E-03 | 0 | 2.050E-03 | 3.250E-06 | 4.320E-07 | rs11039377 |
| 19 | 11 | 47791003 | 4.693 | 4.956 | 1.740E-03 | 0 | 1.740E-03 | 2.690E-06 | 7.210E-07 | rs11039379 |
| 19 | 11 | 47791429 | 4.616 | 5.126 | 2.410E-03 | 0 | 2.410E-03 | 3.920E-06 | 2.960E-07 | rs2869541 |
| 19 | 11 | 47791870 | 4.799 | 4.893 | 1.080E-03 | 0 | 1.080E-03 | 1.590E-06 | 9.920E-07 | rs7129237 |
| 19 | 11 | 47794802 | 4.699 | 4.949 | 1.690E-03 | 0 | 1.690E-03 | 2.610E-06 | 7.450E-07 | rs10838763 |
| 19 | 11 | 47798753 | 4.603 | 5.058 | 2.560E-03 | 0 | 2.560E-03 | 4.170E-06 | 4.230E-07 | rs11039392 |
| 19 | 11 | 47801101 | 4.669 | 4.955 | 1.920E-03 | 0 | 1.920E-03 | 3.020E-06 | 7.240E-07 | rs7114813 |
| 19 | 11 | 47805856 | 4.699 | 4.964 | 1.690E-03 | 0 | 1.690E-03 | 2.620E-06 | 6.910E-07 | rs12226431 |
| 19 | 11 | 47810617 | 4.622 | 4.899 | 2.360E-03 | 0 | 2.360E-03 | 3.810E-06 | 9.650E-07 | rs10838767 |
| 19 | 11 | 47811309 | 5.189 | 5.994 | 1.620E-04 | 0 | 1.620E-04 | 2.120E-07 | 2.040E-09 | rs7114011 |
| 19 | 11 | 47811933 | 4.639 | 5.066 | 2.200E-03 | 0 | 2.200E-03 | 3.510E-06 | 4.070E-07 | rs2125838 |
| 19 | 11 | 47814280 | 4.677 | 4.930 | 1.860E-03 | 0 | 1.860E-03 | 2.910E-06 | 8.240E-07 | rs754434 |
| 19 | 11 | 47815702 | 4.544 | 5.111 | 3.270E-03 | 0 | 3.270E-03 | 5.510E-06 | 3.200E-07 | rs4752792 |
| 19 | 11 | 47816052 | 4.681 | 4.936 | 1.830E-03 | 0 | 1.830E-03 | 2.850E-06 | 7.960E-07 | rs10838768 |
| 19 | 11 | 47817010 | 4.615 | 4.932 | 2.420E-03 | 0 | 2.420E-03 | 3.920E-06 | 8.120E-07 | rs3886262 |
| 19 | 11 | 47817290 | 4.607 | 4.935 | 2.510E-03 | 0 | 2.510E-03 | 4.090E-06 | 8.020E-07 | rs2904127 |
| 19 | 11 | 47820241 | 4.810 | 4.310 | 1.030E-03 | 0 | 1.030E-03 | 1.510E-06 | 1.640E-05 | rs34228231 |
| 19 | 11 | 47822217 | 5.218 | 5.970 | 1.390E-04 | 0 | 1.390E-04 | 1.810E-07 | 2.370E-09 | rs11039398 |
| 19 | 11 | 47823056 | 4.717 | 4.942 | 1.560E-03 | 0 | 1.560E-03 | 2.400E-06 | 7.720E-07 | rs7112248 |
| 19 | 11 | 47825333 | 4.537 | 5.152 | 3.370E-03 | 0 | 3.370E-03 | 5.710E-06 | 2.580E-07 | rs10838769 |
| 19 | 11 | 47825590 | 4.608 | 4.920 | 2.500E-03 | 0 | 2.500E-03 | 4.070E-06 | 8.670E-07 | rs11039401 |
| 19 | 11 | 4782609 | 4.627 | 4.941 | 2.310E-03 | 0 | 2.310E-03 | 3.710E | 7.790E- | rs110394 |

| cFDRlocusnum | CHR | BP | zscore_AD | zscore_eGFR | conjfdr_AD_eGFR | prune_AD_eGFR | min_conjfdr | pval_AD | pval_eGFR | rsID |
|---|---|---|---|---|---|---|---|---|---|---|
| | | 0 | | | | | | -06 | 07 | 02 |
| 19 | 11 | 47828988 | 4.557 | 5.153 | 3.100E-03 | 0 | 3.100E-03 | 5.200E-06 | 2.560E-07 | rs7115228 |
| 19 | 11 | 47830590 | 4.570 | 4.932 | 2.930E-03 | 0 | 2.930E-03 | 4.890E-06 | 8.130E-07 | rs12225854 |
| 19 | 11 | 47831332 | 4.604 | 5.118 | 2.550E-03 | 0 | 2.550E-03 | 4.150E-06 | 3.090E-07 | rs10838771 |
| 19 | 11 | 47834346 | 4.620 | 4.965 | 2.380E-03 | 0 | 2.380E-03 | 3.840E-06 | 6.880E-07 | rs10838773 |
| 19 | 11 | 47835721 | 4.225 | 5.887 | 1.140E-02 | 0 | 1.140E-02 | 2.380E-05 | 3.940E-09 | rs11039409 |
| 19 | 11 | 47836026 | 5.187 | 6.111 | 1.630E-04 | 0 | 1.630E-04 | 2.140E-07 | 9.900E-10 | rs10458914 |
| 19 | 11 | 47840622 | 4.538 | 5.152 | 3.360E-03 | 0 | 3.360E-03 | 5.680E-06 | 2.580E-07 | rs28617307 |
| 19 | 11 | 47841862 | 4.540 | 5.149 | 3.330E-03 | 0 | 3.330E-03 | 5.630E-06 | 2.610E-07 | rs4752796 |
| 19 | 11 | 47843133 | 4.550 | 5.151 | 3.190E-03 | 0 | 3.190E-03 | 5.370E-06 | 2.590E-07 | rs7122182 |
| 19 | 11 | 47844368 | 4.605 | 5.123 | 2.530E-03 | 0 | 2.530E-03 | 4.120E-06 | 3.010E-07 | rs7948468 |
| 19 | 11 | 47845814 | 4.619 | 4.938 | 2.380E-03 | 0 | 2.380E-03 | 3.860E-06 | 7.900E-07 | rs11039415 |
| 19 | 11 | 47848442 | 4.600 | 4.946 | 2.580E-03 | 0 | 2.580E-03 | 4.220E-06 | 7.580E-07 | rs7110300 |
| 19 | 11 | 47849470 | 4.610 | 4.941 | 2.470E-03 | 0 | 2.470E-03 | 4.030E-06 | 7.750E-07 | rs9633830 |
| 19 | 11 | 47851354 | 4.636 | 4.921 | 2.220E-03 | 0 | 2.220E-03 | 3.550E-06 | 8.590E-07 | rs11039419 |
| 19 | 11 | 47852057 | 4.603 | 5.027 | 2.560E-03 | 0 | 2.560E-03 | 4.170E-06 | 4.990E-07 | rs7123952 |
| 19 | 11 | 47856114 | 4.617 | 5.036 | 2.400E-03 | 0 | 2.400E-03 | 3.890E-06 | 4.750E-07 | rs10838774 |
| 19 | 11 | 47860193 | 4.049 | 4.917 | 2.150E-02 | 0 | 2.150E-02 | 5.150E-05 | 8.810E-07 | rs10838776 |
| 19 | 11 | 47860954 | 4.624 | 4.919 | 2.340E-03 | 0 | 2.340E-03 | 3.760E-06 | 8.700E-07 | rs10838777 |
| 19 | 11 | 47864842 | 4.668 | 4.930 | 1.940E-03 | 0 | 1.940E-03 | 3.050E-06 | 8.220E-07 | rs10742824 |
| 19 | 11 | 47865160 | 4.606 | 5.021 | 2.520E-03 | 0 | 2.520E-03 | 4.100E-06 | 5.150E-07 | rs10769299 |
| 19 | 11 | 47865560 | 4.668 | 4.932 | 1.940E-03 | 0 | 1.940E-03 | 3.050E-06 | 8.120E-07 | rs10769300 |
| 19 | 11 | 47867059 | 4.654 | 4.956 | 2.050E-03 | 0 | 2.050E-03 | 3.250E-06 | 7.200E-07 | rs11039429 |
| 19 | 11 | 47871017 | 4.557 | 5.013 | 3.090E-03 | 0 | 3.090E-03 | 5.190E-06 | 5.370E-07 | rs7950674 |
| 19 | 11 | 47872012 | 4.627 | 4.945 | 2.310E-03 | 0 | 2.310E-03 | 3.720E-06 | 7.610E-07 | rs11039431 |
| 19 | 11 | 47873327 | 4.628 | 4.941 | 2.300E-03 | 0 | 2.300E-03 | 3.690E-06 | 7.770E-07 | rs11039432 |
| 19 | 11 | 47875910 | 4.634 | 4.864 | 2.250E-03 | 0 | 2.250E-03 | 3.590E-06 | 1.150E-06 | rs2170815 |
| 19 | 11 | 47877800 | 4.545 | 5.019 | 3.260E-03 | 0 | 3.260E-03 | 5.490E-06 | 5.190E-07 | rs10838782 |
| 19 | 11 | 47878264 | 4.650 | 4.897 | 2.090E-03 | 0 | 2.090E-03 | 3.320E-06 | 9.710E-07 | rs12576933 |
| 19 | 11 | 47881740 | 4.637 | 4.892 | 2.220E-03 | 0 | 2.220E-03 | 3.540E-06 | 9.980E-07 | rs7929725 |

| cFDRlocusnum | CHR | BP | zscore_AD | zscore_eGFR | conjfdr_AD_eGFR | prune_AD_eGFR | min_conjfdr | pval_AD | pval_eGFR | rsID |
|---|---|---|---|---|---|---|---|---|---|---|
| 19 | 11 | 47882130 | 4.726 | 4.901 | 1.500E-03 | 0 | 1.500E-03 | 2.290E-06 | 9.560E-07 | rs12366244 |
| 19 | 11 | 47882341 | 4.675 | 4.893 | 1.880E-03 | 0 | 1.880E-03 | 2.940E-06 | 9.930E-07 | rs10838784 |
| 19 | 11 | 47882782 | 4.084 | 4.901 | 1.910E-02 | 0 | 1.910E-02 | 4.430E-05 | 9.530E-07 | rs7933896 |
| 19 | 11 | 47883337 | 5.144 | 5.961 | 2.030E-04 | 0 | 2.030E-04 | 2.680E-07 | 2.510E-09 | rs7934481 |
| 19 | 11 | 47883880 | 4.591 | 5.056 | 2.690E-03 | 0 | 2.690E-03 | 4.420E-06 | 4.270E-07 | rs7116346 |
| 19 | 11 | 47884260 | 4.656 | 4.844 | 2.040E-03 | 0 | 2.040E-03 | 3.230E-06 | 1.280E-06 | rs11039434 |
| 19 | 11 | 47886364 | 4.530 | 5.048 | 3.470E-03 | 0 | 3.470E-03 | 5.900E-06 | 4.470E-07 | rs4752798 |
| 19 | 11 | 47887087 | 4.555 | 4.942 | 3.120E-03 | 0 | 3.120E-03 | 5.240E-06 | 7.730E-07 | rs7944084 |
| 19 | 11 | 47889545 | 4.580 | 5.101 | 2.820E-03 | 0 | 2.820E-03 | 4.650E-06 | 3.380E-07 | rs7118747 |
| 19 | 11 | 47890172 | 4.745 | 5.282 | 1.380E-03 | 0 | 1.380E-03 | 2.080E-06 | 1.280E-07 | rs12803857 |
| 19 | 11 | 47899030 | 4.707 | 4.839 | 1.630E-03 | 0 | 1.630E-03 | 2.520E-06 | 1.310E-06 | rs1483988 |
| 19 | 11 | 47904086 | 4.549 | 4.710 | 3.200E-03 | 0 | 3.200E-03 | 5.390E-06 | 2.480E-06 | rs10838786 |
| 19 | 11 | 47907641 | -6.216 | -3.376 | 2.040E-02 | 0 | 2.040E-02 | 5.090E-10 | 7.360E-04 | rs4752801 |
| 19 | 11 | 47909313 | 4.780 | 5.546 | 1.180E-03 | 0 | 1.180E-03 | 1.750E-06 | 2.930E-08 | rs2046377 |
| 19 | 11 | 47910823 | -6.193 | -3.402 | 1.880E-02 | 0 | 1.880E-02 | 5.890E-10 | 6.680E-04 | rs12361256 |
| 19 | 11 | 47912150 | 4.533 | 5.339 | 3.430E-03 | 0 | 3.430E-03 | 5.830E-06 | 9.350E-08 | rs1471712 |
| 19 | 11 | 47913097 | 4.791 | 5.417 | 1.120E-03 | 0 | 1.120E-03 | 1.660E-06 | 6.080E-08 | rs7121038 |
| 19 | 11 | 47915299 | -5.797 | -3.366 | 2.100E-02 | 0 | 2.100E-02 | 6.750E-09 | 7.630E-04 | rs34467936 |
| 19 | 11 | 47917631 | 4.578 | 5.238 | 2.840E-03 | 0 | 2.840E-03 | 4.700E-06 | 1.630E-07 | rs11039447 |
| 19 | 11 | 47918416 | 4.531 | 5.332 | 3.450E-03 | 0 | 3.450E-03 | 5.870E-06 | 9.710E-08 | rs11039448 |
| 19 | 11 | 47929846 | 4.741 | 5.971 | 1.410E-03 | 0 | 1.410E-03 | 2.120E-06 | 2.350E-09 | rs6485795 |
| 19 | 11 | 47932666 | -6.077 | -3.205 | 3.400E-02 | 0 | 3.400E-02 | 1.220E-09 | 1.350E-03 | rs12802244 |
| 19 | 11 | 47943419 | 4.730 | 5.286 | 1.480E-03 | 0 | 1.480E-03 | 2.240E-06 | 1.250E-07 | rs10838791 |
| 19 | 11 | 47946836 | -6.036 | -3.193 | 3.520E-02 | 0 | 3.520E-02 | 1.580E-09 | 1.410E-03 | rs2930191 |
| 20 | 11 | 48913546 | 3.811 | 4.800 | 4.650E-02 | 0 | 4.650E-02 | 1.390E-04 | 1.590E-06 | rs12792807 |
| 20 | 11 | 48929342 | 3.946 | 4.696 | 3.030E-02 | 0 | 3.030E-02 | 7.930E-05 | 2.650E-06 | rs7946341 |
| 20 | 11 | 49003362 | 3.942 | 4.820 | 3.080E-02 | 0 | 3.080E-02 | 8.090E-05 | 1.440E-06 | rs11608048 |
| 20 | 11 | 49003921 | 3.787 | 4.945 | 4.980E-02 | 0 | 4.980E-02 | 1.530E-04 | 7.600E-07 | rs1847638 |
| 20 | 11 | 49003941 | 3.797 | 4.931 | 4.840E-02 | 0 | 4.840E-02 | 1.460E-04 | 8.190E-07 | rs1847639 |
| 20 | 11 | 4900987 | 4.053 | 3.830 | 2.120E-02 | 0 | 2.120E-02 | 5.050E | 1.280E- | rs286590 |

| cFDRlocusnum | CHR | BP | zscore_AD | zscore_eGFR | conjfdr_AD_eGFR | prune_AD_eGFR | min_conjfdr | pval_AD | pval_eGFR | rsID |
|---|---|---|---|---|---|---|---|---|---|---|
| | | 7 | | | | | | -05 | 04 | 8 |
| 20 | 11 | 49012833 | 3.822 | 4.907 | 4.490E-02 | 0 | 4.490E-02 | 1.330E-04 | 9.260E-07 | rs1827724 |
| 21 | 11 | 63745500 | 3.828 | 3.144 | 4.730E-02 | 0 | 4.730E-02 | 1.290E-04 | 1.670E-03 | rs4980501 |
| 21 | 11 | 63745677 | 3.839 | 3.113 | 4.640E-02 | 1 | 4.640E-02 | 1.240E-04 | 1.850E-03 | rs4980517 |
| 21 | 11 | 63761061 | 3.808 | 3.189 | 4.920E-02 | 0 | 4.920E-02 | 1.400E-04 | 1.430E-03 | rs34595065 |
| 21 | 11 | 63774597 | 3.822 | 3.205 | 4.690E-02 | 0 | 4.690E-02 | 1.330E-04 | 1.350E-03 | rs509701 |
| 22 | 11 | 85609073 | 4.036 | -3.198 | 3.470E-02 | 0 | 3.470E-02 | 5.430E-05 | 1.390E-03 | rs78563874 |
| 22 | 11 | 85615549 | 3.853 | -3.233 | 4.210E-02 | 0 | 4.210E-02 | 1.170E-04 | 1.230E-03 | rs7106460 |
| 22 | 11 | 85655105 | -6.073 | 3.918 | 3.610E-03 | 0 | 3.610E-03 | 1.250E-09 | 8.940E-05 | rs1237230 |
| 22 | 11 | 85659847 | -6.002 | 3.915 | 3.640E-03 | 0 | 3.640E-03 | 1.950E-09 | 9.060E-05 | rs2458499 |
| 22 | 11 | 85666138 | -6.212 | 3.911 | 3.680E-03 | 0 | 3.680E-03 | 5.220E-10 | 9.190E-05 | rs11234493 |
| 22 | 11 | 85666440 | -6.041 | 3.907 | 3.730E-03 | 0 | 3.730E-03 | 1.530E-09 | 9.350E-05 | rs556562 |
| 22 | 11 | 85671702 | -6.175 | 3.965 | 3.090E-03 | 0 | 3.090E-03 | 6.620E-10 | 7.340E-05 | rs618679 |
| 22 | 11 | 85672383 | -5.904 | 3.880 | 4.060E-03 | 0 | 4.060E-03 | 3.540E-09 | 1.040E-04 | rs2077815 |
| 22 | 11 | 85673124 | -6.141 | 4.021 | 2.570E-03 | 0 | 2.570E-03 | 8.200E-10 | 5.790E-05 | rs592314 |
| 22 | 11 | 85675533 | -5.948 | 3.933 | 3.420E-03 | 0 | 3.420E-03 | 2.720E-09 | 8.380E-05 | rs548121 |
| 22 | 11 | 85682120 | -5.914 | 3.904 | 3.760E-03 | 0 | 3.760E-03 | 3.340E-09 | 9.450E-05 | rs626542 |
| 22 | 11 | 85685166 | -5.972 | 3.899 | 3.830E-03 | 0 | 3.830E-03 | 2.340E-09 | 9.680E-05 | rs637304 |
| 22 | 11 | 85689435 | -5.774 | 3.812 | 5.060E-03 | 0 | 5.060E-03 | 7.750E-09 | 1.380E-04 | rs618629 |
| 22 | 11 | 85692342 | -5.784 | 3.820 | 4.940E-03 | 0 | 4.940E-03 | 7.270E-09 | 1.340E-04 | rs2508688 |
| 22 | 11 | 85696670 | -6.150 | 3.905 | 3.750E-03 | 0 | 3.750E-03 | 7.760E-10 | 9.420E-05 | rs585820 |
| 22 | 11 | 85698598 | -6.174 | 3.950 | 3.240E-03 | 0 | 3.240E-03 | 6.650E-10 | 7.820E-05 | rs670606 |
| 22 | 11 | 85700000 | -6.018 | 3.909 | 3.700E-03 | 0 | 3.700E-03 | 1.760E-09 | 9.270E-05 | rs654194 |
| 22 | 11 | 85701622 | -6.087 | 3.976 | 2.990E-03 | 0 | 2.990E-03 | 1.150E-09 | 7.020E-05 | rs488134 |
| 22 | 11 | 85701741 | -6.090 | 4.001 | 2.740E-03 | 0 | 2.740E-03 | 1.130E-09 | 6.310E-05 | rs682366 |
| 22 | 11 | 85702967 | -5.923 | 3.932 | 3.440E-03 | 0 | 3.440E-03 | 3.160E-09 | 8.440E-05 | rs659801 |
| 22 | 11 | 85705885 | -6.157 | 4.029 | 2.510E-03 | 0 | 2.510E-03 | 7.400E-10 | 5.600E-05 | rs595972 |
| 22 | 11 | 85707228 | -5.802 | 3.840 | 4.610E-03 | 0 | 4.610E-03 | 6.540E-09 | 1.230E-04 | rs542167 |
| 22 | 11 | 85709559 | -5.781 | 3.879 | 4.080E-03 | 0 | 4.080E-03 | 7.410E-09 | 1.050E-04 | rs603615 |
| 22 | 11 | 85710169 | -5.819 | 3.850 | 4.480E-03 | 0 | 4.480E-03 | 5.930E-09 | 1.180E-04 | rs600258 |

| cFDRlocusnum | CHR | BP | zscore_AD | zscore_eGFR | conjfdr_AD_eGFR | prune_AD_eGFR | min_conjfdr | pval_AD | pval_eGFR | rsID |
|---|---|---|---|---|---|---|---|---|---|---|
| 22 | 11 | 85710180 | -5.934 | 4.030 | 2.500E-03 | 0 | 2.500E-03 | 2.950E-09 | 5.570E-05 | rs616555 |
| 22 | 11 | 85713565 | -5.855 | 3.801 | 5.250E-03 | 0 | 5.250E-03 | 4.760E-09 | 1.440E-04 | rs2155051 |
| 22 | 11 | 85715736 | -6.187 | 4.010 | 2.660E-03 | 0 | 2.660E-03 | 6.150E-10 | 6.060E-05 | rs527162 |
| 22 | 11 | 85715853 | -5.989 | 3.887 | 3.960E-03 | 0 | 3.960E-03 | 2.110E-09 | 1.010E-04 | rs528109 |
| 22 | 11 | 85725621 | -5.918 | 3.873 | 4.160E-03 | 0 | 4.160E-03 | 3.260E-09 | 1.070E-04 | rs622220 |
| 22 | 11 | 85725937 | -6.148 | 4.040 | 2.420E-03 | 1 | 2.420E-03 | 7.870E-10 | 5.340E-05 | rs592297 |
| 22 | 11 | 85726818 | -6.038 | 4.020 | 2.590E-03 | 0 | 2.590E-03 | 1.560E-09 | 5.830E-05 | rs648270 |
| 22 | 11 | 85728030 | -5.722 | 3.920 | 3.570E-03 | 0 | 3.570E-03 | 1.050E-08 | 8.840E-05 | rs1996108 |
| 22 | 11 | 85743449 | -5.987 | 3.985 | 2.900E-03 | 0 | 2.900E-03 | 2.130E-09 | 6.760E-05 | rs547767 |
| 22 | 11 | 85764875 | -5.893 | 3.864 | 4.290E-03 | 0 | 4.290E-03 | 3.800E-09 | 1.120E-04 | rs497711 |
| 22 | 11 | 85765737 | -6.124 | 3.976 | 2.980E-03 | 0 | 2.980E-03 | 9.110E-10 | 7.000E-05 | rs489486 |
| 22 | 11 | 85766582 | -5.883 | 3.853 | 4.430E-03 | 0 | 4.430E-03 | 4.030E-09 | 1.170E-04 | rs9804630 |
| 22 | 11 | 85771065 | -5.895 | 3.826 | 4.830E-03 | 0 | 4.830E-03 | 3.750E-09 | 1.300E-04 | rs634719 |
| 22 | 11 | 85771170 | -6.107 | 3.966 | 3.080E-03 | 0 | 3.080E-03 | 1.020E-09 | 7.310E-05 | rs635168 |
| 22 | 11 | 85778924 | -5.873 | 3.746 | 6.270E-03 | 0 | 6.270E-03 | 4.280E-09 | 1.800E-04 | rs544458 |
| 22 | 11 | 85779297 | -6.109 | 3.923 | 3.540E-03 | 0 | 3.540E-03 | 1.000E-09 | 8.730E-05 | rs2509608 |
| 22 | 11 | 85779310 | -6.103 | 3.904 | 3.770E-03 | 0 | 3.770E-03 | 1.040E-09 | 9.480E-05 | rs2458500 |
| 22 | 11 | 85780448 | -5.283 | 3.810 | 5.100E-03 | 0 | 5.100E-03 | 1.270E-07 | 1.390E-04 | rs3016327 |
| 22 | 11 | 85781322 | -6.296 | 3.700 | 7.270E-03 | 0 | 7.270E-03 | 3.060E-10 | 2.160E-04 | rs669556 |
| 22 | 11 | 85783875 | -6.255 | 3.770 | 5.790E-03 | 0 | 5.790E-03 | 3.980E-10 | 1.630E-04 | rs2458493 |
| 22 | 11 | 85804432 | 4.120 | -3.347 | 2.230E-02 | 1 | 2.230E-02 | 3.800E-05 | 8.180E-04 | rs12284629 |
| 22 | 11 | 85809879 | 4.181 | -3.292 | 2.630E-02 | 0 | 2.630E-02 | 2.910E-05 | 9.950E-04 | rs12294130 |
| 23 | 11 | 122545146 | 3.986 | -5.195 | 2.670E-02 | 1 | 2.670E-02 | 6.720E-05 | 2.050E-07 | rs35483388 |
| 24 | 12 | 52272699 | 3.832 | 5.656 | 4.350E-02 | 0 | 4.350E-02 | 1.270E-04 | 1.540E-08 | rs11169942 |
| 24 | 12 | 52273248 | 3.859 | 5.649 | 4.010E-02 | 1 | 4.010E-02 | 1.140E-04 | 1.610E-08 | rs11169943 |
| 25 | 12 | 56860020 | 3.966 | 7.309 | 2.850E-02 | 0 | 2.850E-02 | 7.300E-05 | 2.690E-13 | rs2939302 |
| 25 | 12 | 56860073 | 3.999 | 6.751 | 2.540E-02 | 0 | 2.540E-02 | 6.350E-05 | 1.470E-11 | rs7298123 |
| 25 | 12 | 56860577 | 3.961 | 7.316 | 2.900E-02 | 0 | 2.900E-02 | 7.480E-05 | 2.550E-13 | rs2933243 |
| 25 | 12 | 56861034 | 3.950 | 7.300 | 3.000E-02 | 0 | 3.000E-02 | 7.810E-05 | 2.880E-13 | rs73114872 |
| 25 | 12 | 5686145 | 3.859 | 6.701 | 4.010E-02 | 0 | 4.010E-02 | 1.140E | 2.070E- | rs730292 |

| cFDRlocusnum | CHR | BP | zscore_AD | zscore_eGFR | conjfdr_AD_eGFR | prune_AD_eGFR | min_conjfdr | pval_AD | pval_eGFR | rsID |
|---|---|---|---|---|---|---|---|---|---|---|
| | | 8 | | | | | | -04 | 11 | 5 |
| 25 | 12 | 56865040 | 3.969 | 7.268 | 2.830E-02 | 0 | 2.830E-02 | 7.230E-05 | 3.640E-13 | rs1043011 |
| 25 | 12 | 56865338 | 3.965 | 7.250 | 2.860E-02 | 0 | 2.860E-02 | 7.340E-05 | 4.160E-13 | rs2657879 |
| 25 | 12 | 56866962 | 4.270 | 6.500 | 9.670E-03 | 1 | 9.670E-03 | 1.960E-05 | 8.040E-11 | rs2657878 |
| 26 | 14 | 103853367 | -4.050 | -3.167 | 3.780E-02 | 0 | 3.780E-02 | 5.130E-05 | 1.540E-03 | rs1951392 |
| 26 | 14 | 103858029 | -4.116 | -3.185 | 3.600E-02 | 0 | 3.600E-02 | 3.850E-05 | 1.450E-03 | rs113135977 |
| 26 | 14 | 103858673 | -4.077 | -3.130 | 4.190E-02 | 0 | 4.190E-02 | 4.570E-05 | 1.750E-03 | rs3783394 |
| 26 | 14 | 103860206 | -3.961 | -3.193 | 3.520E-02 | 0 | 3.520E-02 | 7.460E-05 | 1.410E-03 | rs12889639 |
| 26 | 14 | 103860309 | -3.890 | -3.187 | 3.820E-02 | 0 | 3.820E-02 | 1.000E-04 | 1.440E-03 | rs12890750 |
| 26 | 14 | 103862144 | -3.989 | -3.186 | 3.590E-02 | 0 | 3.590E-02 | 6.640E-05 | 1.440E-03 | rs1961600 |
| 26 | 14 | 103862193 | -3.820 | -3.212 | 4.700E-02 | 0 | 4.700E-02 | 1.340E-04 | 1.320E-03 | rs1989565 |
| 26 | 14 | 103862322 | -3.991 | -3.160 | 3.850E-02 | 0 | 3.850E-02 | 6.570E-05 | 1.580E-03 | rs2010281 |
| 26 | 14 | 103864885 | -4.000 | -3.167 | 3.790E-02 | 0 | 3.790E-02 | 6.340E-05 | 1.540E-03 | rs72708854 |
| 26 | 14 | 103865785 | -3.801 | -3.223 | 4.940E-02 | 0 | 4.940E-02 | 1.440E-04 | 1.270E-03 | rs7140564 |
| 26 | 14 | 103868851 | -3.830 | -3.231 | 4.500E-02 | 0 | 4.500E-02 | 1.280E-04 | 1.230E-03 | rs4900572 |
| 26 | 14 | 103869620 | -3.991 | -3.177 | 3.680E-02 | 0 | 3.680E-02 | 6.590E-05 | 1.490E-03 | rs11621510 |
| 26 | 14 | 103874420 | -3.813 | -3.207 | 4.810E-02 | 0 | 4.810E-02 | 1.380E-04 | 1.340E-03 | rs12879085 |
| 26 | 14 | 103876794 | -3.799 | -3.216 | 4.980E-02 | 0 | 4.980E-02 | 1.450E-04 | 1.300E-03 | rs7154246 |
| 26 | 14 | 103883633 | -3.962 | -3.260 | 2.930E-02 | 0 | 2.930E-02 | 7.430E-05 | 1.120E-03 | rs11623869 |
| 26 | 14 | 103884651 | -3.889 | -3.211 | 3.800E-02 | 0 | 3.800E-02 | 1.010E-04 | 1.320E-03 | rs62007681 |
| 26 | 14 | 103886757 | -3.816 | -3.198 | 4.790E-02 | 0 | 4.790E-02 | 1.360E-04 | 1.380E-03 | rs7152202 |
| 26 | 14 | 103887615 | -4.022 | -3.226 | 3.190E-02 | 0 | 3.190E-02 | 5.780E-05 | 1.260E-03 | rs7158144 |
| 26 | 14 | 103887868 | -3.937 | -3.275 | 3.160E-02 | 0 | 3.160E-02 | 8.250E-05 | 1.060E-03 | rs7158822 |
| 26 | 14 | 103888324 | -3.979 | -3.205 | 3.400E-02 | 0 | 3.400E-02 | 6.920E-05 | 1.350E-03 | rs1028613 |
| 26 | 14 | 103891346 | -3.975 | -3.260 | 2.900E-02 | 0 | 2.900E-02 | 7.040E-05 | 1.110E-03 | rs10145793 |
| 26 | 14 | 103894071 | -3.997 | -3.220 | 3.250E-02 | 0 | 3.250E-02 | 6.400E-05 | 1.280E-03 | rs62007683 |
| 26 | 14 | 103894223 | -4.043 | -3.196 | 3.500E-02 | 0 | 3.500E-02 | 5.270E-05 | 1.400E-03 | rs62007684 |
| 26 | 14 | 103895020 | -3.860 | -3.262 | 4.060E-02 | 0 | 4.060E-02 | 1.140E-04 | 1.100E-03 | rs3783395 |
| 26 | 14 | 103897899 | -3.932 | -3.226 | 3.300E-02 | 0 | 3.300E-02 | 8.440E-05 | 1.260E-03 | rs28496120 |
| 26 | 14 | 103899251 | -4.001 | -3.200 | 3.450E-02 | 0 | 3.450E-02 | 6.310E-05 | 1.370E-03 | rs72708874 |

| cFDRlocusnum | CHR | BP | zscore_AD | zscore_eGFR | conjfdr_AD_eGFR | prune_AD_eGFR | min_conjfdr | pval_AD | pval_eGFR | rsID |
|---|---|---|---|---|---|---|---|---|---|---|
| 26 | 14 | 103899344 | -4.000 | -3.199 | 3.460E-02 | 0 | 3.460E-02 | 6.330E-05 | 1.380E-03 | rs62007686 |
| 26 | 14 | 103903763 | -3.979 | -3.189 | 3.560E-02 | 0 | 3.560E-02 | 6.930E-05 | 1.430E-03 | rs62007687 |
| 26 | 14 | 103909203 | -3.914 | -3.224 | 3.500E-02 | 0 | 3.500E-02 | 9.090E-05 | 1.270E-03 | rs1013650 |
| 26 | 14 | 103916958 | -4.023 | -3.221 | 3.240E-02 | 0 | 3.240E-02 | 5.760E-05 | 1.280E-03 | rs17679475 |
| 26 | 14 | 103920927 | -3.962 | -3.219 | 3.260E-02 | 0 | 3.260E-02 | 7.430E-05 | 1.290E-03 | rs10143247 |
| 26 | 14 | 103923008 | -3.969 | -3.213 | 3.320E-02 | 0 | 3.320E-02 | 7.210E-05 | 1.310E-03 | rs10141120 |
| 26 | 14 | 103923414 | -3.971 | -3.181 | 3.640E-02 | 0 | 3.640E-02 | 7.160E-05 | 1.470E-03 | rs2273700 |
| 26 | 14 | 103923475 | -3.933 | -3.211 | 3.340E-02 | 0 | 3.340E-02 | 8.400E-05 | 1.320E-03 | rs2273699 |
| 26 | 14 | 103924477 | -3.969 | -3.201 | 3.440E-02 | 0 | 3.440E-02 | 7.210E-05 | 1.370E-03 | rs7146506 |
| 26 | 14 | 103928251 | -3.973 | -3.230 | 3.150E-02 | 0 | 3.150E-02 | 7.100E-05 | 1.240E-03 | rs10134580 |
| 26 | 14 | 103929535 | -4.011 | -3.175 | 3.710E-02 | 0 | 3.710E-02 | 6.040E-05 | 1.500E-03 | rs72708893 |
| 26 | 14 | 103930564 | -3.969 | -3.205 | 3.400E-02 | 0 | 3.400E-02 | 7.210E-05 | 1.350E-03 | rs28444993 |
| 26 | 14 | 103933158 | -3.912 | -3.194 | 3.560E-02 | 0 | 3.560E-02 | 9.140E-05 | 1.400E-03 | rs11626483 |
| 26 | 14 | 103938664 | -3.839 | -3.175 | 4.510E-02 | 0 | 4.510E-02 | 1.240E-04 | 1.500E-03 | rs28583452 |
| 26 | 14 | 103939275 | -3.905 | -3.137 | 4.120E-02 | 0 | 4.120E-02 | 9.420E-05 | 1.710E-03 | rs34216351 |
| 26 | 14 | 103939679 | -3.866 | -3.176 | 4.150E-02 | 0 | 4.150E-02 | 1.110E-04 | 1.490E-03 | rs10141388 |
| 26 | 14 | 103940436 | -3.923 | -3.140 | 4.080E-02 | 0 | 4.080E-02 | 8.750E-05 | 1.690E-03 | rs62007718 |
| 26 | 14 | 103940911 | -3.867 | -3.175 | 4.150E-02 | 0 | 4.150E-02 | 1.100E-04 | 1.500E-03 | rs11628969 |
| 26 | 14 | 103943130 | -3.926 | -3.127 | 4.230E-02 | 0 | 4.230E-02 | 8.630E-05 | 1.770E-03 | rs62007719 |
| 26 | 14 | 103944137 | -3.897 | -3.173 | 3.770E-02 | 0 | 3.770E-02 | 9.730E-05 | 1.510E-03 | rs7160366 |
| 26 | 14 | 103945414 | -3.923 | -3.158 | 3.870E-02 | 0 | 3.870E-02 | 8.740E-05 | 1.590E-03 | rs7145113 |
| 26 | 14 | 103948626 | -3.921 | -3.138 | 4.100E-02 | 0 | 4.100E-02 | 8.800E-05 | 1.700E-03 | rs2403180 |
| 26 | 14 | 103950368 | -3.906 | -3.180 | 3.660E-02 | 0 | 3.660E-02 | 9.380E-05 | 1.470E-03 | rs3742458 |
| 26 | 14 | 103952453 | -3.914 | -3.168 | 3.770E-02 | 0 | 3.770E-02 | 9.070E-05 | 1.530E-03 | rs11628213 |
| 26 | 14 | 103952621 | -3.845 | -3.176 | 4.430E-02 | 0 | 4.430E-02 | 1.210E-04 | 1.490E-03 | rs7156150 |
| 26 | 14 | 103953032 | -3.920 | -3.162 | 3.830E-02 | 0 | 3.830E-02 | 8.840E-05 | 1.570E-03 | rs17617941 |
| 26 | 14 | 103960026 | -3.939 | -3.158 | 3.880E-02 | 0 | 3.880E-02 | 8.170E-05 | 1.590E-03 | rs56365443 |
| 26 | 14 | 103962559 | -3.872 | -3.174 | 4.080E-02 | 0 | 4.080E-02 | 1.080E-04 | 1.510E-03 | rs10141433 |
| 26 | 14 | 103965528 | -3.859 | -3.175 | 4.250E-02 | 0 | 4.250E-02 | 1.140E-04 | 1.500E-03 | rs7160208 |
| 26 | 14 | 1039673 | -3.946 | -3.171 | 3.740E-02 | 0 | 3.740E-02 | 7.930E | 1.520E- | rs967191 |

| cFDRlocusnum | CHR | BP | zscore_AD | zscore_eGFR | conjfdr_AD_eGFR | prune_AD_eGFR | min_conjfdr | pval_AD | pval_eGFR | rsID |
|---|---|---|---|---|---|---|---|---|---|---|
| | | 43 | | | | | | -05 | 03 | 1 |
| 26 | 14 | 103977605 | -3.924 | -3.178 | 3.670E-02 | 0 | 3.670E-02 | 8.710E-05 | 1.480E-03 | rs10150072 |
| 26 | 14 | 103977724 | -3.937 | -3.181 | 3.640E-02 | 0 | 3.640E-02 | 8.260E-05 | 1.470E-03 | rs8019578 |
| 26 | 14 | 103979127 | -3.863 | -3.188 | 4.170E-02 | 0 | 4.170E-02 | 1.120E-04 | 1.430E-03 | rs10133035 |
| 26 | 14 | 103981849 | -4.065 | -3.219 | 3.260E-02 | 0 | 3.260E-02 | 4.800E-05 | 1.290E-03 | rs72710714 |
| 26 | 14 | 103986255 | -3.934 | -3.120 | 4.310E-02 | 0 | 4.310E-02 | 8.360E-05 | 1.810E-03 | rs1803283 |
| 26 | 14 | 103990799 | -3.866 | -3.526 | 3.930E-02 | 0 | 3.930E-02 | 1.110E-04 | 4.220E-04 | rs3759584 |
| 26 | 14 | 103991401 | -4.044 | -3.342 | 2.260E-02 | 0 | 2.260E-02 | 5.250E-05 | 8.320E-04 | rs55723020 |
| 26 | 14 | 103991749 | -4.051 | -3.357 | 2.160E-02 | 1 | 2.160E-02 | 5.110E-05 | 7.880E-04 | rs11626787 |
| 26 | 14 | 103992473 | 3.790 | 4.799 | 4.940E-02 | 0 | 4.940E-02 | 1.510E-04 | 1.590E-06 | rs2756118 |
| 26 | 14 | 103994961 | -4.021 | -3.258 | 2.920E-02 | 0 | 2.920E-02 | 5.790E-05 | 1.120E-03 | rs1467561 |
| 26 | 14 | 104000183 | 3.916 | 5.027 | 3.360E-02 | 0 | 3.360E-02 | 9.010E-05 | 4.990E-07 | rs2756127 |
| 26 | 14 | 104002611 | -4.261 | -3.297 | 2.590E-02 | 0 | 2.590E-02 | 2.040E-05 | 9.770E-04 | rs45441198 |
| 26 | 14 | 104003070 | -4.037 | -3.312 | 2.480E-02 | 0 | 2.480E-02 | 5.420E-05 | 9.250E-04 | rs2296486 |
| 26 | 14 | 104003094 | -4.030 | -3.289 | 2.650E-02 | 0 | 2.650E-02 | 5.570E-05 | 1.010E-03 | rs2296484 |
| 26 | 14 | 104004147 | -4.167 | -3.262 | 2.880E-02 | 0 | 2.880E-02 | 3.090E-05 | 1.110E-03 | rs10135868 |
| 26 | 14 | 104006160 | -4.175 | -3.237 | 3.090E-02 | 0 | 3.090E-02 | 2.980E-05 | 1.210E-03 | rs11621037 |
| 27 | 15 | 63569902 | 4.911 | -5.812 | 6.460E-04 | 1 | 6.460E-04 | 9.050E-07 | 6.160E-09 | rs117618017 |
| 27 | 15 | 63571820 | 4.510 | -5.824 | 3.760E-03 | 0 | 3.760E-03 | 6.490E-06 | 5.760E-09 | rs75763893 |
| 27 | 15 | 63595878 | 4.695 | -5.717 | 1.720E-03 | 0 | 1.720E-03 | 2.670E-06 | 1.080E-08 | rs76855300 |
| 28 | 15 | 82431219 | -4.506 | -3.906 | 3.820E-03 | 1 | 3.820E-03 | 6.600E-06 | 9.390E-05 | rs2457497 |
| 29 | 16 | 30068354 | -4.043 | 3.291 | 2.630E-02 | 1 | 2.630E-02 | 5.270E-05 | 9.980E-04 | rs9939774 |
| 30 | 16 | 51602710 | -3.811 | -3.262 | 4.710E-02 | 0 | 4.710E-02 | 1.390E-04 | 1.110E-03 | rs34841413 |
| 30 | 16 | 51603899 | -3.816 | -3.295 | 4.560E-02 | 1 | 4.560E-02 | 1.350E-04 | 9.850E-04 | rs1111316 |
| 31 | 16 | 71452475 | 4.287 | 4.110 | 9.070E-03 | 1 | 9.070E-03 | 1.810E-05 | 3.960E-05 | rs7192890 |
| 31 | 16 | 71458638 | 3.945 | 4.879 | 3.040E-02 | 0 | 3.040E-02 | 7.980E-05 | 1.060E-06 | rs7202340 |
| 32 | 17 | 1618135 | -4.435 | 5.594 | 5.090E-03 | 1 | 5.090E-03 | 9.190E-06 | 2.210E-08 | rs4790812 |
| 32 | 17 | 1618363 | 4.222 | -3.378 | 2.030E-02 | 0 | 2.030E-02 | 2.420E-05 | 7.310E-04 | rs11078597 |
| 32 | 17 | 1623675 | 5.032 | -4.149 | 1.670E-03 | 0 | 1.670E-03 | 4.840E-07 | 3.330E-05 | rs1574525 |
| 32 | 17 | 1625544 | -4.318 | 5.929 | 8.040E-03 | 0 | 8.040E-03 | 1.580E-05 | 3.050E-09 | rs62090040 |

| cFDRlocusnum | CHR | BP | zscore_AD | zscore_eGFR | conjfdr_AD_eGFR | prune_AD_eGFR | min_conjfdr | pval_AD | pval_eGFR | rsID |
|---|---|---|---|---|---|---|---|---|---|---|
| 32 | 17 | 1625892 | -4.158 | 5.985 | 1.460E-02 | 0 | 1.460E-02 | 3.200E-05 | 2.160E-09 | rs57937546 |
| 33 | 17 | 1630992 | 5.053 | -4.456 | 5.480E-04 | 0 | 5.480E-04 | 4.350E-07 | 8.360E-06 | rs9912287 |
| 33 | 17 | 1637458 | 4.885 | -4.656 | 7.290E-04 | 0 | 7.290E-04 | 1.040E-06 | 3.220E-06 | rs8065251 |
| 33 | 17 | 1639719 | 4.753 | -4.656 | 1.330E-03 | 0 | 1.330E-03 | 2.000E-06 | 3.230E-06 | rs4989024 |
| 33 | 17 | 1639965 | 4.757 | -4.647 | 1.310E-03 | 0 | 1.310E-03 | 1.970E-06 | 3.360E-06 | rs1317708 |
| 33 | 17 | 1640034 | 4.758 | -4.668 | 1.300E-03 | 0 | 1.300E-03 | 1.960E-06 | 3.050E-06 | rs874424 |
| 33 | 17 | 1640529 | 4.662 | -4.709 | 1.980E-03 | 0 | 1.980E-03 | 3.130E-06 | 2.490E-06 | rs34962442 |
| 33 | 17 | 1640793 | 4.850 | -4.730 | 8.580E-04 | 0 | 8.580E-04 | 1.240E-06 | 2.250E-06 | rs8077638 |
| 33 | 17 | 1641035 | 4.550 | -4.447 | 3.190E-03 | 0 | 3.190E-03 | 5.360E-06 | 8.690E-06 | rs2287322 |
| 33 | 17 | 1641715 | 4.725 | -4.723 | 1.510E-03 | 0 | 1.510E-03 | 2.300E-06 | 2.320E-06 | rs1310 |
| 33 | 17 | 1641806 | 4.712 | -4.698 | 1.590E-03 | 0 | 1.590E-03 | 2.450E-06 | 2.630E-06 | rs1045806 |
| 33 | 17 | 1645410 | 4.154 | -4.761 | 1.480E-02 | 0 | 1.480E-02 | 3.260E-05 | 1.930E-06 | rs62090056 |
| 33 | 17 | 1645526 | 4.737 | -4.781 | 1.440E-03 | 0 | 1.440E-03 | 2.170E-06 | 1.740E-06 | rs62090057 |
| 33 | 17 | 1645747 | 4.722 | -4.773 | 1.530E-03 | 0 | 1.530E-03 | 2.330E-06 | 1.810E-06 | rs34599286 |
| 33 | 17 | 1646317 | 4.969 | -4.817 | 4.890E-04 | 1 | 4.890E-04 | 6.720E-07 | 1.460E-06 | rs12937244 |
| 33 | 17 | 1648502 | 4.259 | -4.827 | 1.010E-02 | 0 | 1.010E-02 | 2.060E-05 | 1.390E-06 | rs2070863 |
| 33 | 17 | 1649365 | 4.883 | -4.797 | 7.360E-04 | 0 | 7.360E-04 | 1.040E-06 | 1.610E-06 | rs7501750 |
| 33 | 17 | 1650125 | 4.855 | -4.803 | 8.390E-04 | 0 | 8.390E-04 | 1.210E-06 | 1.560E-06 | rs4525526 |
| 33 | 17 | 1651516 | 4.738 | -4.830 | 1.430E-03 | 0 | 1.430E-03 | 2.160E-06 | 1.370E-06 | rs4790284 |
| 33 | 17 | 1651697 | 4.829 | -4.827 | 9.480E-04 | 0 | 9.480E-04 | 1.370E-06 | 1.380E-06 | rs4790285 |
| 33 | 17 | 1653091 | 4.494 | -4.729 | 4.020E-03 | 0 | 4.020E-03 | 6.990E-06 | 2.250E-06 | rs3976 |
| 33 | 17 | 1653504 | 4.949 | -4.545 | 5.400E-04 | 0 | 5.400E-04 | 7.480E-07 | 5.500E-06 | rs35050485 |
| 33 | 17 | 1653580 | 4.941 | -4.575 | 5.580E-04 | 0 | 5.580E-04 | 7.770E-07 | 4.770E-06 | rs12936190 |
| 33 | 17 | 1653732 | 4.460 | -4.436 | 4.600E-03 | 0 | 4.600E-03 | 8.200E-06 | 9.160E-06 | rs4790820 |
| 33 | 17 | 1654048 | 4.460 | -4.525 | 4.600E-03 | 0 | 4.600E-03 | 8.200E-06 | 6.040E-06 | rs4790821 |
| 33 | 17 | 1655605 | 4.953 | -4.572 | 5.300E-04 | 0 | 5.300E-04 | 7.320E-07 | 4.840E-06 | rs58147322 |
| 34 | 17 | 4761960 | 3.792 | 4.232 | 4.910E-02 | 0 | 4.910E-02 | 1.490E-04 | 2.310E-05 | rs111985639 |
| 34 | 17 | 4762035 | 3.792 | 4.283 | 4.900E-02 | 0 | 4.900E-02 | 1.490E-04 | 1.840E-05 | rs16954223 |
| 34 | 17 | 4763551 | 3.790 | 4.298 | 4.940E-02 | 0 | 4.940E-02 | 1.510E-04 | 1.730E-05 | rs8078173 |
| 34 | 17 | 4777634 | 3.846 | 4.278 | 4.170E-02 | 0 | 4.170E-02 | 1.200E | 1.890E- | rs121503 |

| cFDRlocusnum | CHR | BP | zscore_AD | zscore_eGFR | conjfdr_AD_eGFR | prune_AD_eGFR | min_conjfdr | pval_AD | pval_eGFR | rsID |
|---|---|---|---|---|---|---|---|---|---|---|
| | | | | | | | | -04 | 05 | 70 |
| 34 | 17 | 4819887 | 4.025 | 4.051 | 2.340E-02 | 0 | 2.340E-02 | 5.690E-05 | 5.100E-05 | rs72835074 |
| 34 | 17 | 4823657 | 3.961 | 4.034 | 2.900E-02 | 0 | 2.900E-02 | 7.460E-05 | 5.480E-05 | rs113426708 |
| 34 | 17 | 4829817 | 3.902 | 4.042 | 3.500E-02 | 0 | 3.500E-02 | 9.540E-05 | 5.310E-05 | rs72835079 |
| 34 | 17 | 4902473 | 3.997 | 4.223 | 2.570E-02 | 0 | 2.570E-02 | 6.420E-05 | 2.410E-05 | rs76101349 |
| 34 | 17 | 4958757 | 3.929 | 5.067 | 3.220E-02 | 0 | 3.220E-02 | 8.530E-05 | 4.040E-07 | rs56343966 |
| 34 | 17 | 4961538 | 4.094 | 4.864 | 1.840E-02 | 0 | 1.840E-02 | 4.230E-05 | 1.150E-06 | rs112466593 |
| 34 | 17 | 4967699 | 4.164 | 4.762 | 1.430E-02 | 0 | 1.430E-02 | 3.130E-05 | 1.920E-06 | rs113371019 |
| 34 | 17 | 4975231 | 4.167 | 4.827 | 1.420E-02 | 0 | 1.420E-02 | 3.090E-05 | 1.390E-06 | rs111971401 |
| 34 | 17 | 4975542 | 4.234 | 4.826 | 1.110E-02 | 0 | 1.110E-02 | 2.290E-05 | 1.390E-06 | rs8077508 |
| 34 | 17 | 4976401 | 4.203 | 4.811 | 1.240E-02 | 0 | 1.240E-02 | 2.630E-05 | 1.500E-06 | rs111566368 |
| 34 | 17 | 4985233 | 3.814 | 5.125 | 4.600E-02 | 0 | 4.600E-02 | 1.370E-04 | 2.970E-07 | rs111261071 |
| 34 | 17 | 5117806 | 4.152 | 4.633 | 1.500E-02 | 0 | 1.500E-02 | 3.300E-05 | 3.610E-06 | rs7502862 |
| 34 | 17 | 5118951 | 4.112 | 4.787 | 1.720E-02 | 0 | 1.720E-02 | 3.920E-05 | 1.690E-06 | rs77493189 |
| 34 | 17 | 5123477 | 4.141 | 4.714 | 1.560E-02 | 0 | 1.560E-02 | 3.460E-05 | 2.430E-06 | rs73976325 |
| 34 | 17 | 5123861 | 4.165 | 4.751 | 1.420E-02 | 0 | 1.420E-02 | 3.110E-05 | 2.030E-06 | rs113762960 |
| 34 | 17 | 5133128 | 4.241 | 4.877 | 1.080E-02 | 0 | 1.080E-02 | 2.230E-05 | 1.080E-06 | rs61182333 |
| 34 | 17 | 5137047 | 4.208 | 4.119 | 1.220E-02 | 0 | 1.220E-02 | 2.570E-05 | 3.810E-05 | rs7225151 |
| 34 | 17 | 5138304 | 4.285 | 4.450 | 9.140E-03 | 1 | 9.140E-03 | 1.830E-05 | 8.590E-06 | rs75511804 |
| 34 | 17 | 5138980 | 4.166 | 4.324 | 1.420E-02 | 0 | 1.420E-02 | 3.100E-05 | 1.530E-05 | rs113260531 |
| 34 | 17 | 5139808 | 4.275 | 4.213 | 9.490E-03 | 0 | 9.490E-03 | 1.910E-05 | 2.520E-05 | rs58124010 |
| 34 | 17 | 5140399 | 4.119 | 4.158 | 1.680E-02 | 0 | 1.680E-02 | 3.800E-05 | 3.210E-05 | rs4456560 |
| 34 | 17 | 5144696 | 4.057 | 4.074 | 2.090E-02 | 0 | 2.090E-02 | 4.970E-05 | 4.630E-05 | rs72479703 |
| 34 | 17 | 5155919 | 4.157 | 4.246 | 1.470E-02 | 0 | 1.470E-02 | 3.230E-05 | 2.180E-05 | rs113806568 |
| 34 | 17 | 5158714 | 4.216 | 4.293 | 1.180E-02 | 0 | 1.180E-02 | 2.490E-05 | 1.760E-05 | rs61481506 |
| 34 | 17 | 5179846 | 4.053 | 4.048 | 2.120E-02 | 0 | 2.120E-02 | 5.060E-05 | 5.160E-05 | rs78531165 |
| 34 | 17 | 5182430 | 4.162 | 3.926 | 1.440E-02 | 0 | 1.440E-02 | 3.160E-05 | 8.630E-05 | rs56117162 |
| 34 | 17 | 5184467 | 3.897 | 3.900 | 3.550E-02 | 0 | 3.550E-02 | 9.730E-05 | 9.600E-05 | rs77318561 |
| 34 | 17 | 5189416 | 3.883 | 3.734 | 3.710E-02 | 0 | 3.710E-02 | 1.030E-04 | 1.890E-04 | rs8077567 |
| 34 | 17 | 5194249 | 3.963 | 3.815 | 2.870E-02 | 0 | 2.870E-02 | 7.390E-05 | 1.360E-04 | rs80172399 |

| cFDRlocusnum | CHR | BP | zscore_AD | zscore_eGFR | conjfdr_AD_eGFR | prune_AD_eGFR | min_conjfdr | pval_AD | pval_eGFR | rsID |
|---|---|---|---|---|---|---|---|---|---|---|
| 34 | 17 | 5196409 | 4.001 | 3.787 | 2.530E-02 | 0 | 2.530E-02 | 6.310E-05 | 1.530E-04 | rs112065650 |
| 34 | 17 | 5200232 | 3.945 | 3.836 | 3.050E-02 | 0 | 3.050E-02 | 7.990E-05 | 1.250E-04 | rs79970806 |
| 34 | 17 | 5209510 | 3.980 | 3.737 | 2.730E-02 | 0 | 2.730E-02 | 6.900E-05 | 1.860E-04 | rs76253980 |
| 34 | 17 | 5216998 | 3.959 | 3.802 | 2.910E-02 | 0 | 2.910E-02 | 7.520E-05 | 1.440E-04 | rs16954523 |
| 34 | 17 | 5222071 | 3.982 | 3.809 | 2.700E-02 | 0 | 2.700E-02 | 6.830E-05 | 1.390E-04 | rs77752862 |
| 34 | 17 | 5230117 | 4.005 | 3.794 | 2.500E-02 | 0 | 2.500E-02 | 6.210E-05 | 1.480E-04 | rs59587437 |
| 34 | 17 | 5230867 | 4.059 | 3.795 | 2.070E-02 | 0 | 2.070E-02 | 4.920E-05 | 1.480E-04 | rs56125028 |
| 34 | 17 | 5231412 | 3.987 | 3.811 | 2.650E-02 | 0 | 2.650E-02 | 6.680E-05 | 1.380E-04 | rs55993740 |
| 34 | 17 | 5231618 | 4.005 | 3.795 | 2.490E-02 | 0 | 2.490E-02 | 6.190E-05 | 1.470E-04 | rs55947703 |
| 34 | 17 | 5232202 | 3.982 | 3.812 | 2.700E-02 | 0 | 2.700E-02 | 6.820E-05 | 1.380E-04 | rs112572092 |
| 34 | 17 | 5233817 | 3.954 | 3.795 | 2.960E-02 | 0 | 2.960E-02 | 7.680E-05 | 1.480E-04 | rs12600505 |
| 34 | 17 | 5241850 | 3.918 | 3.835 | 3.330E-02 | 0 | 3.330E-02 | 8.930E-05 | 1.260E-04 | rs112421661 |
| 34 | 17 | 5243201 | 3.920 | 3.838 | 3.310E-02 | 0 | 3.310E-02 | 8.840E-05 | 1.240E-04 | rs75262874 |
| 34 | 17 | 5243462 | 3.926 | 3.810 | 3.240E-02 | 0 | 3.240E-02 | 8.630E-05 | 1.390E-04 | rs78650284 |
| 34 | 17 | 5244853 | 3.919 | 3.813 | 3.320E-02 | 0 | 3.320E-02 | 8.880E-05 | 1.370E-04 | rs57638638 |
| 34 | 17 | 5246150 | 3.936 | 3.790 | 3.140E-02 | 0 | 3.140E-02 | 8.270E-05 | 1.510E-04 | rs55913010 |
| 34 | 17 | 5247732 | 3.938 | 3.800 | 3.120E-02 | 0 | 3.120E-02 | 8.230E-05 | 1.450E-04 | rs112888134 |
| 34 | 17 | 5248682 | 4.032 | 3.759 | 2.280E-02 | 0 | 2.280E-02 | 5.520E-05 | 1.710E-04 | rs12601720 |
| 34 | 17 | 5251671 | 3.960 | 3.869 | 2.910E-02 | 0 | 2.910E-02 | 7.500E-05 | 1.090E-04 | rs16954548 |
| 34 | 17 | 5252514 | 3.993 | 3.853 | 2.600E-02 | 0 | 2.600E-02 | 6.520E-05 | 1.170E-04 | rs16954552 |
| 34 | 17 | 5261743 | 3.851 | 3.643 | 4.110E-02 | 0 | 4.110E-02 | 1.180E-04 | 2.690E-04 | rs112918260 |
| 34 | 17 | 5264285 | 3.828 | 3.850 | 4.400E-02 | 0 | 4.400E-02 | 1.290E-04 | 1.180E-04 | rs75367124 |
| 34 | 17 | 5269183 | 3.918 | 3.844 | 3.340E-02 | 0 | 3.340E-02 | 8.940E-05 | 1.210E-04 | rs76418084 |
| 35 | 17 | 17775706 | -4.007 | -4.554 | 2.480E-02 | 1 | 2.480E-02 | 6.150E-05 | 5.260E-06 | rs61217632 |
| 36 | 17 | 29390671 | 4.330 | 3.549 | 1.180E-02 | 0 | 1.180E-02 | 1.490E-05 | 3.870E-04 | rs60642411 |
| 36 | 17 | 29395746 | 4.344 | 3.697 | 7.330E-03 | 1 | 7.330E-03 | 1.400E-05 | 2.180E-04 | rs11657989 |
| 36 | 17 | 29411573 | 3.910 | 4.127 | 3.420E-02 | 0 | 3.420E-02 | 9.240E-05 | 3.670E-05 | rs12449302 |
| 36 | 17 | 29461525 | 4.163 | 3.786 | 1.440E-02 | 0 | 1.440E-02 | 3.150E-05 | 1.530E-04 | rs72813607 |
| 36 | 17 | 29509946 | 4.162 | 4.061 | 1.440E-02 | 0 | 1.440E-02 | 3.150E-05 | 4.890E-05 | rs17885309 |
| 36 | 17 | 2952711 | 4.056 | 4.087 | 2.100E-02 | 0 | 2.100E-02 | 4.990E | 4.370E- | rs116502 |

| cFDRlocusnum | CHR | BP | zscore_AD | zscore_eGFR | conjfdr_AD_eGFR | prune_AD_eGFR | min_conjfdr | pval_AD | pval_eGFR | rsID |
|---|---|---|---|---|---|---|---|---|---|---|
| | | 7 | | | | | | -05 | 05 | 14 |
| 36 | 17 | 29530876 | 4.068 | 4.061 | 2.010E-02 | 0 | 2.010E-02 | 4.730E-05 | 4.880E-05 | rs72813627 |
| 36 | 17 | 29546765 | 4.177 | 3.930 | 1.370E-02 | 0 | 1.370E-02 | 2.960E-05 | 8.490E-05 | rs17884466 |
| 36 | 17 | 29573051 | 4.175 | 4.120 | 1.380E-02 | 0 | 1.380E-02 | 2.980E-05 | 3.790E-05 | rs17881943 |
| 36 | 17 | 29589988 | 4.080 | 4.214 | 1.930E-02 | 0 | 1.930E-02 | 4.500E-05 | 2.510E-05 | rs17886160 |
| 36 | 17 | 29599724 | 4.160 | 4.222 | 1.450E-02 | 0 | 1.450E-02 | 3.180E-05 | 2.430E-05 | rs72815614 |
| 36 | 17 | 29616516 | 4.150 | 4.198 | 1.510E-02 | 0 | 1.510E-02 | 3.320E-05 | 2.690E-05 | rs72815624 |
| 36 | 17 | 29623288 | 4.273 | 4.316 | 9.540E-03 | 0 | 9.540E-03 | 1.930E-05 | 1.590E-05 | rs11080149 |
| 37 | 17 | 60538932 | -3.925 | 5.091 | 3.260E-02 | 0 | 3.260E-02 | 8.670E-05 | 3.570E-07 | rs112781672 |
| 37 | 17 | 60554275 | -3.939 | 5.212 | 3.110E-02 | 1 | 3.110E-02 | 8.200E-05 | 1.870E-07 | rs9898829 |
| 37 | 17 | 60555375 | -3.824 | 5.144 | 4.460E-02 | 0 | 4.460E-02 | 1.310E-04 | 2.690E-07 | rs113301871 |
| 38 | 18 | 32621231 | 3.985 | -5.076 | 2.680E-02 | 0 | 2.680E-02 | 6.750E-05 | 3.840E-07 | rs530081 |
| 38 | 18 | 32623344 | 4.472 | -3.722 | 6.760E-03 | 1 | 6.760E-03 | 7.760E-06 | 1.980E-04 | rs686836 |
| 38 | 18 | 32624667 | 3.844 | -5.133 | 4.190E-02 | 0 | 4.190E-02 | 1.210E-04 | 2.850E-07 | rs595086 |
| 38 | 18 | 32631716 | 4.232 | -4.120 | 1.120E-02 | 0 | 1.120E-02 | 2.320E-05 | 3.790E-05 | rs668477 |
| 38 | 18 | 32631855 | 4.283 | -3.965 | 9.230E-03 | 0 | 9.230E-03 | 1.850E-05 | 7.330E-05 | rs549147 |
| 38 | 18 | 32631873 | 4.274 | -4.161 | 9.510E-03 | 0 | 9.510E-03 | 1.920E-05 | 3.170E-05 | rs502989 |
| 38 | 18 | 32632021 | 4.285 | -4.181 | 9.140E-03 | 0 | 9.140E-03 | 1.820E-05 | 2.910E-05 | rs501295 |
| 38 | 18 | 32634634 | 4.101 | -4.813 | 1.790E-02 | 0 | 1.790E-02 | 4.110E-05 | 1.490E-06 | rs7228414 |
| 38 | 18 | 32637037 | 3.854 | -4.816 | 4.080E-02 | 0 | 4.080E-02 | 1.160E-04 | 1.460E-06 | rs481492 |
| 38 | 18 | 32637135 | 3.942 | -4.868 | 3.070E-02 | 0 | 3.070E-02 | 8.070E-05 | 1.120E-06 | rs482405 |
| 38 | 18 | 32637939 | 3.881 | -4.655 | 3.740E-02 | 0 | 3.740E-02 | 1.040E-04 | 3.240E-06 | rs507549 |
| 38 | 18 | 32638528 | 4.057 | -4.748 | 2.090E-02 | 0 | 2.090E-02 | 4.980E-05 | 2.060E-06 | rs516412 |
| 38 | 18 | 32638852 | 3.851 | -4.717 | 4.110E-02 | 0 | 4.110E-02 | 1.180E-04 | 2.390E-06 | rs603493 |
| 38 | 18 | 32670021 | 3.793 | -4.728 | 4.890E-02 | 0 | 4.890E-02 | 1.490E-04 | 2.270E-06 | rs476348 |
| 38 | 18 | 32672836 | 3.793 | -4.611 | 4.890E-02 | 0 | 4.890E-02 | 1.490E-04 | 4.010E-06 | rs497999 |
| 39 | 19 | 18633755 | 3.815 | 3.154 | 4.900E-02 | 1 | 4.900E-02 | 1.360E-04 | 1.610E-03 | rs172032 |
| 40 | 19 | 33370070 | -4.388 | -5.957 | 6.130E-03 | 0 | 6.130E-03 | 1.140E-05 | 2.570E-09 | rs745961 |
| 40 | 19 | 33370161 | -4.105 | -5.951 | 1.770E-02 | 0 | 1.770E-02 | 4.050E-05 | 2.660E-09 | rs745959 |
| 40 | 19 | 33370566 | -4.400 | -5.918 | 5.850E-03 | 0 | 5.850E-03 | 1.080E-05 | 3.270E-09 | rs17206282 |

| cFDRlocusnum | CHR | BP | zscore_AD | zscore_eGFR | conjfdr_AD_eGFR | prune_AD_eGFR | min_conjfdr | pval_AD | pval_eGFR | rsID |
|---|---|---|---|---|---|---|---|---|---|---|
| 40 | 19 | 33370702 | -4.395 | -5.943 | 5.970E-03 | 0 | 5.970E-03 | 1.110E-05 | 2.790E-09 | rs55970832 |
| 40 | 19 | 33370952 | -4.387 | -5.992 | 6.170E-03 | 0 | 6.170E-03 | 1.150E-05 | 2.070E-09 | rs11665689 |
| 40 | 19 | 33371145 | -4.375 | -5.932 | 6.440E-03 | 0 | 6.440E-03 | 1.210E-05 | 2.990E-09 | rs17206303 |
| 40 | 19 | 33371234 | -4.385 | -5.930 | 6.210E-03 | 0 | 6.210E-03 | 1.160E-05 | 3.030E-09 | rs76375203 |
| 40 | 19 | 33372243 | -4.432 | -5.898 | 5.160E-03 | 1 | 5.160E-03 | 9.360E-06 | 3.680E-09 | rs73041776 |
| 40 | 19 | 33372601 | -4.349 | -5.932 | 7.150E-03 | 0 | 7.150E-03 | 1.370E-05 | 3.000E-09 | rs11666273 |
| 40 | 19 | 33373922 | -4.341 | -5.917 | 7.380E-03 | 0 | 7.380E-03 | 1.420E-05 | 3.280E-09 | rs113479494 |
| 40 | 19 | 33374940 | -4.253 | -5.934 | 1.030E-02 | 0 | 1.030E-02 | 2.110E-05 | 2.950E-09 | rs11084679 |
| 40 | 19 | 33376519 | -4.252 | -6.011 | 1.040E-02 | 0 | 1.040E-02 | 2.120E-05 | 1.850E-09 | rs60858882 |
| 40 | 19 | 33377053 | -4.248 | -5.905 | 1.050E-02 | 0 | 1.050E-02 | 2.150E-05 | 3.530E-09 | rs2868197 |
| 40 | 19 | 33379462 | -4.230 | -5.903 | 1.120E-02 | 0 | 1.120E-02 | 2.330E-05 | 3.580E-09 | rs59084925 |
| 40 | 19 | 33381610 | -4.331 | -5.941 | 7.680E-03 | 0 | 7.680E-03 | 1.490E-05 | 2.830E-09 | rs2098700 |
| 40 | 19 | 33382696 | -4.169 | -5.810 | 1.410E-02 | 0 | 1.410E-02 | 3.060E-05 | 6.230E-09 | rs73049192 |
| 40 | 19 | 33383409 | -4.283 | -5.905 | 9.220E-03 | 0 | 9.220E-03 | 1.840E-05 | 3.520E-09 | rs12150993 |
| 40 | 19 | 33384753 | -4.295 | -5.829 | 8.790E-03 | 0 | 8.790E-03 | 1.740E-05 | 5.590E-09 | rs78622628 |
| 40 | 19 | 33385808 | -4.188 | -5.731 | 1.320E-02 | 0 | 1.320E-02 | 2.820E-05 | 1.000E-08 | rs11084681 |
| 40 | 19 | 33387267 | -4.162 | -5.830 | 1.440E-02 | 0 | 1.440E-02 | 3.150E-05 | 5.540E-09 | rs59563992 |
| 40 | 19 | 33387931 | -4.162 | -5.808 | 1.440E-02 | 0 | 1.440E-02 | 3.150E-05 | 6.320E-09 | rs8111376 |
| 41 | 19 | 45269725 | 9.713 | -3.569 | 1.110E-02 | 1 | 1.110E-02 | 2.650E-22 | 3.590E-04 | rs139326841 |
| 41 | 19 | 45296364 | -11.053 | 3.229 | 3.170E-02 | 0 | 3.170E-02 | 2.110E-28 | 1.240E-03 | rs112450640 |
| 41 | 19 | 45301924 | -11.873 | 3.367 | 2.100E-02 | 0 | 2.100E-02 | 1.630E-32 | 7.610E-04 | rs2965113 |
| 41 | 19 | 45301942 | -11.573 | 3.382 | 2.000E-02 | 0 | 2.000E-02 | 5.650E-31 | 7.180E-04 | rs2967669 |
| 41 | 19 | 45302513 | -11.736 | 3.568 | 1.110E-02 | 0 | 1.110E-02 | 8.360E-32 | 3.590E-04 | rs2965112 |
| 41 | 19 | 45302951 | -14.582 | 3.293 | 2.620E-02 | 0 | 2.620E-02 | 3.670E-48 | 9.910E-04 | rs2967668 |
| 41 | 19 | 45311084 | 12.309 | -3.541 | 1.210E-02 | 0 | 1.210E-02 | 8.090E-35 | 3.990E-04 | rs140824606 |
| 42 | 19 | 45313715 | -11.414 | 3.587 | 1.050E-02 | 1 | 1.050E-02 | 3.540E-30 | 3.350E-04 | rs2927477 |
| 43 | 19 | 45324138 | 26.324 | -3.929 | 3.480E-03 | 1 | 3.480E-03 | 1.030E-152 | 8.550E-05 | rs28399637 |
| 43 | 19 | 45325309 | -10.876 | 3.459 | 1.580E-02 | 0 | 1.580E-02 | 1.510E-27 | 5.430E-04 | rs8112196 |
| 43 | 19 | 45325391 | -10.849 | 3.502 | 1.380E-02 | 0 | 1.380E-02 | 2.020E-27 | 4.620E-04 | rs6509172 |
| 43 | 19 | 4532621 | 24.365 | -4.031 | 2.490E-03 | 0 | 2.490E-03 | 3.980E | 5.560E- | rs104063 |

| cFDRlocusnum | CHR | BP | zscore_AD | zscore_eGFR | conjfdr_AD_eGFR | prune_AD_eGFR | min_conjfdr | pval_AD | pval_eGFR | rsID |
|---|---|---|---|---|---|---|---|---|---|---|
| | | 7 | | | | | | -131 | 05 | 38 |
| 43 | 19 | 45326664 | 24.376 | -4.040 | 2.410E-03 | 0 | 2.410E-03 | 3.090E-131 | 5.330E-05 | rs10405693 |
| 43 | 19 | 45326768 | -10.465 | 3.403 | 1.870E-02 | 0 | 1.870E-02 | 1.260E-25 | 6.660E-04 | rs1871045 |
| 43 | 19 | 45327309 | 24.231 | -4.076 | 2.150E-03 | 0 | 2.150E-03 | 1.040E-129 | 4.580E-05 | rs10412413 |
| 43 | 19 | 45328380 | 23.835 | -4.084 | 2.090E-03 | 0 | 2.090E-03 | 1.450E-125 | 4.420E-05 | rs58446550 |
| 43 | 19 | 45329214 | 24.092 | -4.134 | 1.760E-03 | 0 | 1.760E-03 | 3.040E-128 | 3.560E-05 | rs10402271 |
| 43 | 19 | 45329344 | 24.166 | -4.246 | 1.190E-03 | 0 | 1.190E-03 | 5.020E-129 | 2.170E-05 | rs10402524 |
| 43 | 19 | 45336035 | 22.079 | -4.108 | 1.920E-03 | 0 | 1.920E-03 | 5.050E-108 | 3.990E-05 | rs7359852 |
| 43 | 19 | 45336443 | 19.445 | -4.214 | 1.330E-03 | 0 | 1.330E-03 | 3.240E-84 | 2.510E-05 | rs3021439 |
| 43 | 19 | 45337249 | 21.404 | -4.127 | 1.810E-03 | 0 | 1.810E-03 | 1.220E-101 | 3.680E-05 | rs2927481 |
| 43 | 19 | 45340728 | -9.924 | 3.485 | 1.450E-02 | 0 | 1.450E-02 | 3.270E-23 | 4.930E-04 | rs10409208 |
| 43 | 19 | 45340736 | 22.653 | -4.358 | 7.920E-04 | 0 | 7.920E-04 | 1.320E-113 | 1.310E-05 | rs73048293 |
| 43 | 19 | 45341540 | 19.325 | -4.299 | 9.880E-04 | 0 | 9.880E-04 | 3.300E-83 | 1.720E-05 | rs12462274 |
| 43 | 19 | 45341904 | 22.650 | -4.287 | 1.030E-03 | 0 | 1.030E-03 | 1.380E-113 | 1.810E-05 | rs12459575 |
| 43 | 19 | 45342114 | 22.084 | -4.224 | 1.290E-03 | 0 | 1.290E-03 | 4.540E-108 | 2.400E-05 | rs4803761 |
| 43 | 19 | 45342161 | 21.030 | -4.300 | 9.810E-04 | 0 | 9.810E-04 | 3.470E-98 | 1.710E-05 | rs4803762 |
| 43 | 19 | 45345623 | 16.133 | -4.354 | 8.040E-04 | 0 | 8.040E-04 | 1.490E-58 | 1.330E-05 | rs2199575 |
| 43 | 19 | 45346768 | -11.595 | 4.122 | 1.840E-03 | 0 | 1.840E-03 | 4.360E-31 | 3.760E-05 | rs1001611 |
| 43 | 19 | 45347911 | 15.684 | -3.813 | 5.060E-03 | 0 | 5.060E-03 | 1.930E-55 | 1.370E-04 | rs4452060 |
| 43 | 19 | 45348253 | -9.228 | 4.092 | 2.030E-03 | 0 | 2.030E-03 | 2.750E-20 | 4.270E-05 | rs10426423 |
| 43 | 19 | 45348522 | 23.808 | -4.367 | 7.650E-04 | 0 | 7.650E-04 | 2.750E-125 | 1.260E-05 | rs12162222 |
| 43 | 19 | 45349402 | 17.257 | -3.832 | 4.730E-03 | 0 | 4.730E-03 | 9.890E-67 | 1.270E-04 | rs3810143 |
| 43 | 19 | 45349963 | -10.763 | 4.162 | 1.600E-03 | 0 | 1.600E-03 | 5.120E-27 | 3.150E-05 | rs2306149 |
| 43 | 19 | 45351746 | -11.733 | 4.183 | 1.490E-03 | 0 | 1.490E-03 | 8.630E-32 | 2.880E-05 | rs1871047 |
| 43 | 19 | 45351937 | -13.740 | 4.142 | 1.720E-03 | 0 | 1.720E-03 | 5.830E-43 | 3.440E-05 | rs1871046 |
| 43 | 19 | 45352804 | -12.002 | 4.086 | 2.080E-03 | 0 | 2.080E-03 | 3.460E-33 | 4.390E-05 | rs4802240 |
| 43 | 19 | 45354044 | 17.921 | -4.501 | 4.610E-04 | 0 | 4.610E-04 | 8.110E-72 | 6.770E-06 | rs57537848 |
| 43 | 19 | 45354296 | 17.882 | -4.534 | 4.070E-04 | 0 | 4.070E-04 | 1.640E-71 | 5.800E-06 | rs11666329 |
| 43 | 19 | 45355743 | -8.663 | 3.277 | 2.750E-02 | 0 | 2.750E-02 | 4.580E-18 | 1.050E-03 | rs4802241 |
| 43 | 19 | 45357377 | 27.202 | -4.346 | 8.300E-04 | 0 | 8.300E-04 | 6.150E-163 | 1.390E-05 | rs4803764 |

| cFDRlocusnum | CHR | BP | zscore_AD | zscore_eGFR | conjfdr_AD_eGFR | prune_AD_eGFR | min_conjfdr | pval_AD | pval_eGFR | rsID |
|---|---|---|---|---|---|---|---|---|---|---|
| 43 | 19 | 45358353 | -19.047 | 3.160 | 3.850E-02 | 1 | 3.850E-02 | 6.920E-81 | 1.580E-03 | rs10410835 |
| 43 | 19 | 45359586 | 27.280 | -4.132 | 1.780E-03 | 0 | 1.780E-03 | 7.270E-164 | 3.600E-05 | rs56317818 |
| 43 | 19 | 45359706 | 27.234 | -4.089 | 2.050E-03 | 0 | 2.050E-03 | 2.580E-163 | 4.330E-05 | rs12462573 |
| 43 | 19 | 45361224 | -15.896 | 3.589 | 1.040E-02 | 0 | 1.040E-02 | 6.800E-57 | 3.320E-04 | rs440277 |
| 43 | 19 | 45367473 | -6.399 | -4.698 | 2.130E-04 | 1 | 2.130E-04 | 1.570E-10 | 2.630E-06 | rs394353 |
| 43 | 19 | 45368320 | -6.191 | -4.653 | 2.540E-04 | 0 | 2.540E-04 | 5.980E-10 | 3.270E-06 | rs419010 |
| 43 | 19 | 45368424 | -6.187 | -4.620 | 2.890E-04 | 0 | 2.890E-04 | 6.130E-10 | 3.830E-06 | rs394221 |
| 43 | 19 | 45369203 | -15.323 | 3.894 | 3.880E-03 | 0 | 3.880E-03 | 5.400E-53 | 9.850E-05 | rs112422902 |
| 43 | 19 | 45370335 | -7.097 | -4.586 | 3.310E-04 | 0 | 3.310E-04 | 1.280E-12 | 4.510E-06 | rs384653 |
| 43 | 19 | 45371168 | -5.664 | -4.603 | 3.090E-04 | 0 | 3.090E-04 | 1.480E-08 | 4.160E-06 | rs4803766 |
| 43 | 19 | 45372354 | -4.087 | -4.998 | 1.890E-02 | 0 | 1.890E-02 | 4.380E-05 | 5.800E-07 | rs8104483 |
| 43 | 19 | 45377467 | -3.795 | -3.510 | 4.860E-02 | 0 | 4.860E-02 | 1.470E-04 | 4.480E-04 | rs2075642 |
| 43 | 19 | 45379309 | -4.305 | -3.375 | 2.050E-02 | 0 | 2.050E-02 | 1.670E-05 | 7.370E-04 | rs3852859 |
| 43 | 19 | 45381917 | -4.705 | -3.157 | 3.880E-02 | 0 | 3.880E-02 | 2.540E-06 | 1.590E-03 | rs3729640 |
| 44 | 19 | 45386634 | 31.210 | -3.943 | 3.310E-03 | 0 | 3.310E-03 | 7.910E-214 | 8.050E-05 | rs147636938 |
| 44 | 19 | 45387034 | -11.003 | 3.225 | 3.200E-02 | 0 | 3.200E-02 | 3.700E-28 | 1.260E-03 | rs283808 |
| 44 | 19 | 45387057 | -11.029 | 3.253 | 2.960E-02 | 0 | 2.960E-02 | 2.780E-28 | 1.140E-03 | rs283809 |
| 44 | 19 | 45389596 | -13.328 | 4.361 | 7.830E-04 | 0 | 7.830E-04 | 1.600E-40 | 1.290E-05 | rs7254892 |
| 44 | 19 | 45395266 | -33.669 | 5.282 | 1.660E-05 | 1 | 1.660E-05 | 1.630E-248 | 1.280E-07 | rs157580 |
| 44 | 19 | 45396973 | 31.501 | -4.094 | 2.020E-03 | 0 | 2.020E-03 | 8.420E-218 | 4.240E-05 | rs77301115 |
| 44 | 19 | 45397229 | -13.719 | 4.817 | 1.310E-04 | 0 | 1.310E-04 | 7.820E-43 | 1.450E-06 | rs1160983 |
| 44 | 19 | 45397307 | 31.120 | -4.115 | 1.880E-03 | 0 | 1.880E-03 | 1.280E-212 | 3.870E-05 | rs112849259 |
| 44 | 19 | 45397952 | 30.031 | -4.139 | 1.740E-03 | 0 | 1.740E-03 | 3.840E-198 | 3.500E-05 | rs116881820 |
| 44 | 19 | 45398785 | 31.136 | -4.064 | 2.230E-03 | 0 | 2.230E-03 | 7.890E-213 | 4.830E-05 | rs79398853 |
| 44 | 19 | 45399344 | 31.133 | -4.080 | 2.120E-03 | 0 | 2.120E-03 | 8.640E-213 | 4.500E-05 | rs75687619 |
| 44 | 19 | 45399896 | 31.045 | -4.234 | 1.250E-03 | 0 | 1.250E-03 | 1.340E-211 | 2.290E-05 | rs76366838 |
| 44 | 19 | 45400725 | 31.134 | -4.109 | 1.910E-03 | 0 | 1.910E-03 | 8.270E-213 | 3.970E-05 | rs114536010 |
| 44 | 19 | 45403216 | 28.617 | -4.566 | 3.570E-04 | 1 | 3.570E-04 | 4.180E-180 | 4.970E-06 | rs115881343 |
| 44 | 19 | 45404691 | -22.304 | 4.206 | 1.370E-03 | 0 | 1.370E-03 | 3.390E-110 | 2.600E-05 | rs405697 |
| 44 | 19 | 4540485 | 28.607 | -4.559 | 3.670E-04 | 0 | 3.670E-04 | 5.440E | 5.140E- | rs112019 |

| cFDRlocusnum | CHR | BP | zscore_AD | zscore_eGFR | conjfdr_AD_eGFR | prune_AD_eGFR | min_conjfdr | pval_AD | pval_eGFR | rsID |
|---|---|---|---|---|---|---|---|---|---|---|
| | | 7 | | | | | | -180 | 06 | 714 |
| 44 | 19 | 45406673 | 37.501 | -5.276 | 1.710E-05 | 1 | 1.710E-05 | 8.800E-308 | 1.320E-07 | rs10119 |
| 44 | 19 | 45412079 | -21.938 | 4.894 | 9.490E-05 | 1 | 9.490E-05 | 1.120E-106 | 9.860E-07 | rs7412 |
| 44 | 19 | 45412955 | 32.368 | -4.153 | 1.650E-03 | 0 | 1.650E-03 | 7.630E-230 | 3.280E-05 | rs1081105 |
| 45 | 19 | 45425460 | -33.342 | 3.542 | 1.200E-02 | 0 | 1.200E-02 | 9.650E-244 | 3.970E-04 | rs157595 |
| 46 | 19 | 45426792 | -18.197 | 3.870 | 4.200E-03 | 0 | 4.200E-03 | 5.410E-74 | 1.090E-04 | rs141622900 |
| 47 | 19 | 46190268 | -4.321 | -3.998 | 7.940E-03 | 1 | 7.940E-03 | 1.550E-05 | 6.380E-05 | rs1964272 |
| 47 | 19 | 46203083 | -3.845 | -3.992 | 4.180E-02 | 0 | 4.180E-02 | 1.210E-04 | 6.540E-05 | rs7256920 |
| 47 | 19 | 46239598 | -4.038 | -3.302 | 2.550E-02 | 0 | 2.550E-02 | 5.400E-05 | 9.590E-04 | rs9807819 |
| 47 | 19 | 46262286 | -4.134 | -3.337 | 2.300E-02 | 0 | 2.300E-02 | 3.560E-05 | 8.470E-04 | rs725660 |
| 47 | 19 | 46289503 | -4.022 | -3.182 | 3.630E-02 | 0 | 3.630E-02 | 5.760E-05 | 1.460E-03 | rs8109951 |
| 47 | 19 | 46290822 | -4.019 | -3.178 | 3.670E-02 | 0 | 3.670E-02 | 5.830E-05 | 1.480E-03 | rs17571725 |
| 47 | 19 | 46292060 | -3.971 | -3.199 | 3.470E-02 | 0 | 3.470E-02 | 7.160E-05 | 1.380E-03 | rs9676288 |
| 47 | 19 | 46304585 | 3.799 | 3.260 | 4.880E-02 | 0 | 4.880E-02 | 1.450E-04 | 1.120E-03 | rs12463359 |
| 48 | 19 | 47154484 | -5.751 | 3.987 | 2.870E-03 | 1 | 2.870E-03 | 8.870E-09 | 6.690E-05 | rs146335962 |
| 48 | 19 | 47176752 | -4.796 | 3.966 | 3.080E-03 | 0 | 3.080E-03 | 1.620E-06 | 7.310E-05 | rs874460 |
| 48 | 19 | 47179454 | -4.512 | 3.960 | 3.720E-03 | 0 | 3.720E-03 | 6.420E-06 | 7.480E-05 | rs141409639 |
| 48 | 19 | 47182842 | -4.541 | 3.948 | 3.320E-03 | 0 | 3.320E-03 | 5.600E-06 | 7.880E-05 | rs78320072 |
| 48 | 19 | 47182942 | -4.547 | 3.951 | 3.230E-03 | 0 | 3.230E-03 | 5.430E-06 | 7.780E-05 | rs137882482 |
| 48 | 19 | 47196079 | -4.524 | 3.925 | 3.550E-03 | 0 | 3.550E-03 | 6.080E-06 | 8.670E-05 | rs314675 |
| 48 | 19 | 47198932 | -4.481 | 4.123 | 4.230E-03 | 0 | 4.230E-03 | 7.440E-06 | 3.740E-05 | rs56010181 |
| 48 | 19 | 47203259 | -4.468 | 4.010 | 4.450E-03 | 0 | 4.450E-03 | 7.910E-06 | 6.080E-05 | rs314664 |
| 48 | 19 | 47205707 | -3.908 | 3.548 | 3.440E-02 | 0 | 3.440E-02 | 9.310E-05 | 3.880E-04 | rs60652743 |
| 48 | 19 | 47208481 | -3.866 | 3.561 | 3.930E-02 | 0 | 3.930E-02 | 1.110E-04 | 3.690E-04 | rs425105 |
| 48 | 19 | 47212343 | -3.992 | 4.225 | 2.610E-02 | 0 | 2.610E-02 | 6.560E-05 | 2.390E-05 | rs443081 |
| 48 | 19 | 47214411 | -4.190 | 4.181 | 1.300E-02 | 0 | 1.300E-02 | 2.790E-05 | 2.910E-05 | rs432157 |
| 48 | 19 | 47215153 | -4.216 | 4.105 | 1.180E-02 | 0 | 1.180E-02 | 2.490E-05 | 4.040E-05 | rs694273 |
| 48 | 19 | 47219122 | -3.841 | 3.745 | 4.230E-02 | 0 | 4.230E-02 | 1.220E-04 | 1.810E-04 | rs402072 |
| 48 | 19 | 47223949 | -4.106 | 4.003 | 1.760E-02 | 0 | 1.760E-02 | 4.030E-05 | 6.250E-05 | rs313841 |
| 48 | 19 | 47234885 | -4.249 | 4.003 | 1.050E-02 | 0 | 1.050E-02 | 2.140E-05 | 6.260E-05 | rs160544 |

| cFDRlocusnum | CHR | BP | zscore_AD | zscore_eGFR | conjfdr_AD_eGFR | prune_AD_eGFR | min_conjfdr | pval_AD | pval_eGFR | rsID |
|---|---|---|---|---|---|---|---|---|---|---|
| 49 | 19 | 49206985 | 4.035 | 4.444 | 2.260E-02 | 0 | 2.260E-02 | 5.470E-05 | 8.850E-06 | rs602662 |
| 49 | 19 | 49207206 | 4.085 | 4.395 | 1.900E-02 | 0 | 1.900E-02 | 4.400E-05 | 1.110E-05 | rs485186 |
| 49 | 19 | 49207255 | 4.140 | 4.422 | 1.560E-02 | 0 | 1.560E-02 | 3.470E-05 | 9.770E-06 | rs485073 |
| 49 | 19 | 49207257 | 4.123 | 4.422 | 1.660E-02 | 0 | 1.660E-02 | 3.740E-05 | 9.780E-06 | rs603985 |
| 49 | 19 | 49207554 | 4.043 | 4.540 | 2.200E-02 | 0 | 2.200E-02 | 5.280E-05 | 5.630E-06 | rs571689 |
| 49 | 19 | 49207651 | 4.219 | 4.538 | 1.170E-02 | 0 | 1.170E-02 | 2.450E-05 | 5.670E-06 | rs570794 |
| 49 | 19 | 49207792 | 4.071 | 4.533 | 2.000E-02 | 0 | 2.000E-02 | 4.690E-05 | 5.800E-06 | rs2251034 |
| 49 | 19 | 49208501 | 3.828 | 4.563 | 4.400E-02 | 0 | 4.400E-02 | 1.290E-04 | 5.040E-06 | rs507855 |
| 49 | 19 | 49208543 | 4.125 | 4.509 | 1.650E-02 | 0 | 1.650E-02 | 3.710E-05 | 6.520E-06 | rs507766 |
| 49 | 19 | 49208564 | 4.094 | 4.539 | 1.840E-02 | 0 | 1.840E-02 | 4.250E-05 | 5.650E-06 | rs507711 |
| 49 | 19 | 49208865 | 4.081 | 4.503 | 1.930E-02 | 0 | 1.930E-02 | 4.480E-05 | 6.710E-06 | rs504963 |
| 49 | 19 | 49208978 | 4.088 | 4.599 | 1.880E-02 | 0 | 1.880E-02 | 4.350E-05 | 4.250E-06 | rs632111 |
| 49 | 19 | 49209010 | 4.103 | 4.539 | 1.780E-02 | 0 | 1.780E-02 | 4.080E-05 | 5.650E-06 | rs503279 |
| 49 | 19 | 49209226 | 4.068 | 4.590 | 2.020E-02 | 0 | 2.020E-02 | 4.750E-05 | 4.430E-06 | rs633372 |
| 49 | 19 | 49209339 | 4.152 | 4.516 | 1.490E-02 | 0 | 1.490E-02 | 3.290E-05 | 6.300E-06 | rs2548459 |
| 49 | 19 | 49209464 | 4.142 | 4.626 | 1.550E-02 | 0 | 1.550E-02 | 3.450E-05 | 3.720E-06 | rs692854 |
| 49 | 19 | 49209566 | 4.260 | 4.680 | 1.000E-02 | 0 | 1.000E-02 | 2.040E-05 | 2.870E-06 | rs1704773 |
| 49 | 19 | 49209851 | 4.341 | 4.588 | 7.370E-03 | 0 | 7.370E-03 | 1.420E-05 | 4.480E-06 | rs646327 |
| 49 | 19 | 49210869 | -3.893 | -6.176 | 3.600E-02 | 0 | 3.600E-02 | 9.890E-05 | 6.590E-10 | rs2638281 |
| 49 | 19 | 49211969 | 4.304 | 4.240 | 8.490E-03 | 0 | 8.490E-03 | 1.670E-05 | 2.230E-05 | rs676388 |
| 49 | 19 | 49213284 | 4.382 | 4.333 | 6.290E-03 | 0 | 6.290E-03 | 1.180E-05 | 1.470E-05 | rs584768 |
| 49 | 19 | 49213504 | 4.451 | 4.327 | 4.780E-03 | 0 | 4.780E-03 | 8.560E-06 | 1.510E-05 | rs2452170 |
| 49 | 19 | 49213833 | 4.433 | 4.378 | 5.140E-03 | 0 | 5.140E-03 | 9.310E-06 | 1.200E-05 | rs2638282 |
| 49 | 19 | 49214274 | 4.388 | 4.611 | 6.140E-03 | 0 | 6.140E-03 | 1.140E-05 | 4.000E-06 | rs281379 |
| 49 | 19 | 49215095 | 4.021 | 3.734 | 2.370E-02 | 0 | 2.370E-02 | 5.800E-05 | 1.890E-04 | rs4002471 |
| 49 | 19 | 49218111 | 3.973 | 3.766 | 2.780E-02 | 0 | 2.780E-02 | 7.090E-05 | 1.660E-04 | rs33988101 |
| 49 | 19 | 49218430 | 3.952 | 3.812 | 2.980E-02 | 0 | 2.980E-02 | 7.760E-05 | 1.380E-04 | rs12978750 |
| 49 | 19 | 49218437 | 3.903 | 3.795 | 3.490E-02 | 0 | 3.490E-02 | 9.500E-05 | 1.470E-04 | rs12978752 |
| 49 | 19 | 49218602 | 3.931 | 3.773 | 3.200E-02 | 0 | 3.200E-02 | 8.460E-05 | 1.610E-04 | rs12979278 |
| 49 | 19 | 4921945 | 4.215 | 4.250 | 1.180E-02 | 0 | 1.180E-02 | 2.490E | 2.130E- | rs660745 |

| cFDRlocusnum | CHR | BP | zscore_AD | zscore_eGFR | conjfdr_AD_eGFR | prune_AD_eGFR | min_conjfdr | pval_AD | pval_eGFR | rsID |
|---|---|---|---|---|---|---|---|---|---|---|
| | | 9 | | | | | | -05 | 05 | |
| 49 | 19 | 49220323 | 4.163 | 4.006 | 1.440E-02 | 0 | 1.440E-02 | 3.150E-05 | 6.170E-05 | rs11672900 |
| 49 | 19 | 49224389 | -3.849 | -6.066 | 4.140E-02 | 0 | 4.140E-02 | 1.190E-04 | 1.310E-09 | rs3859439 |
| 49 | 19 | 49224966 | -3.805 | -6.063 | 4.720E-02 | 0 | 4.720E-02 | 1.420E-04 | 1.340E-09 | rs519757 |
| 49 | 19 | 49227043 | 4.292 | 3.948 | 8.910E-03 | 0 | 8.910E-03 | 1.770E-05 | 7.880E-05 | rs11666792 |
| 49 | 19 | 49227256 | 4.243 | 3.942 | 1.070E-02 | 0 | 1.070E-02 | 2.200E-05 | 8.070E-05 | rs12979891 |
| 49 | 19 | 49227443 | 4.226 | 3.968 | 1.140E-02 | 0 | 1.140E-02 | 2.380E-05 | 7.260E-05 | rs11878908 |
| 49 | 19 | 49228272 | 4.386 | 3.978 | 6.190E-03 | 0 | 6.190E-03 | 1.150E-05 | 6.950E-05 | rs2287921 |
| 49 | 19 | 49229323 | 4.580 | 4.233 | 2.820E-03 | 1 | 2.820E-03 | 4.660E-06 | 2.310E-05 | rs479486 |
| 49 | 19 | 49232226 | 4.411 | 3.975 | 5.600E-03 | 0 | 5.600E-03 | 1.030E-05 | 7.020E-05 | rs2287922 |
| 49 | 19 | 49236102 | 4.386 | 4.262 | 6.190E-03 | 0 | 6.190E-03 | 1.150E-05 | 2.030E-05 | rs439523 |
| 49 | 19 | 49246866 | 3.909 | 4.031 | 3.430E-02 | 0 | 3.430E-02 | 9.270E-05 | 5.560E-05 | rs838147 |
| 49 | 19 | 49250239 | 3.916 | 4.075 | 3.350E-02 | 0 | 3.350E-02 | 9.000E-05 | 4.600E-05 | rs838144 |
| 50 | 19 | 55176262 | 4.281 | 4.665 | 9.290E-03 | 1 | 9.290E-03 | 1.860E-05 | 3.090E-06 | rs731170 |
| 51 | 20 | 2811868 | 3.887 | 4.800 | 3.670E-02 | 1 | 3.670E-02 | 1.010E-04 | 1.590E-06 | rs6084183 |
| 52 | 20 | 4966941 | -4.609 | 3.157 | 3.890E-02 | 1 | 3.890E-02 | 4.050E-06 | 1.600E-03 | rs4813726 |
| 52 | 20 | 4967199 | -4.579 | 3.089 | 4.670E-02 | 0 | 4.670E-02 | 4.680E-06 | 2.010E-03 | rs1105838 |
| 52 | 20 | 4967355 | -4.606 | 3.125 | 4.250E-02 | 0 | 4.250E-02 | 4.110E-06 | 1.780E-03 | rs4076098 |
| 52 | 20 | 4967393 | -4.664 | 3.085 | 4.730E-02 | 0 | 4.730E-02 | 3.100E-06 | 2.040E-03 | rs2326576 |
| 52 | 20 | 4968220 | -4.681 | 3.085 | 4.730E-02 | 0 | 4.730E-02 | 2.850E-06 | 2.040E-03 | rs13044890 |
| 53 | 20 | 48438572 | -3.795 | 3.778 | 4.860E-02 | 0 | 4.860E-02 | 1.480E-04 | 1.580E-04 | rs6020069 |
| 53 | 20 | 48501606 | -3.955 | 4.106 | 2.950E-02 | 1 | 2.950E-02 | 7.640E-05 | 4.030E-05 | rs645544 |
| 54 | 20 | 54983075 | -6.381 | 3.089 | 4.670E-02 | 1 | 4.670E-02 | 1.760E-10 | 2.010E-03 | rs6069736 |
| 55 | 22 | 41418297 | 3.874 | -3.361 | 3.830E-02 | 1 | 3.830E-02 | 1.070E-04 | 7.770E-04 | rs9611460 |
| 55 | 22 | 41427446 | 3.866 | -3.114 | 4.380E-02 | 0 | 4.380E-02 | 1.100E-04 | 1.850E-03 | rs9607768 |

**Supplementary Table S18. Pleiotropic SNPs Identified by Conjunctional FDR in African American Ancestry.**

This table catalogs pleiotropic SNPs identified by conjFDR analysis specifically in African American ancestry individuals. Due to smaller sample sizes in the AFR AD GWAS, the conjFDR threshold was relaxed to 0.2 rather than the standard 0.05, allowing identification of suggestive pleiotropic signals that may warrant follow-up in larger studies. The table includes conjFDR locus numbers, chromosomal positions, z-scores for AD and eGFR, conjFDR values, pruning indicators, minimum conjFDR values, and individual trait p-values.

| cFDRlocusnum | CHR | BP | zscore_AD | zscore_eGFR | conjfdr_AD_eGFR | prune_AD_eGFR | min_conjfdr | pval_AD | pval_eGFR | rsID |
|---|---|---|---|---|---|---|---|---|---|---|
| 1 | 6 | 151539505 | -3.643 | -3.784 | 1.790E-01 | 1 | 1.790E-01 | 2.690E-04 | 1.540E-04 | rs140069490 |
| 2 | 7 | 70092296 | 3.888 | -3.723 | 1.310E-01 | 1 | 1.310E-01 | 1.010E-04 | 1.970E-04 | rs10242748 |
| 2 | 7 | 70085453 | -3.876 | 3.607 | 1.780E-01 | 0 | 1.780E-01 | 1.060E-04 | 3.100E-04 | rs11762238 |
| 3 | 11 | 38895843 | 3.640 | 3.593 | 1.840E-01 | 1 | 1.840E-01 | 2.730E-04 | 3.270E-04 | rs35302065 |
| 4 | 17 | 6427027 | -3.857 | 3.693 | 1.430E-01 | 1 | 1.430E-01 | 1.150E-04 | 2.210E-04 | rs12453781 |
| 5 | 19 | 24326350 | -3.635 | -4.034 | 1.830E-01 | 1 | 1.830E-01 | 2.780E-04 | 5.490E-05 | rs3943791 |
| 5 | 19 | 24275821 | -3.630 | -4.233 | 1.860E-01 | 0 | 1.860E-01 | 2.830E-04 | 2.310E-05 | rs55736631 |

**Supplementary Table S19. Distinct Pleiotropic Genomic Loci in European Ancestry (Kunkle Discovery, conjFDR<0.05)**

This table presents conjFDR analysis results using AD GWAS summary statistics that were not corrected for competing risk of mortality (Kunkle uncorrected). Competing risk correction accounts for the fact that individuals who die from other causes before developing AD are censored, which can bias genetic effect estimates if genetic variants influence both AD risk and mortality from other causes. By comparing results from corrected versus uncorrected analyses, this table enables assessment of whether competing risk adjustment influences the identification and characterization of pleiotropic loci. The most strongly associated lead SNPs in independent genomic loci are shown after including regions up and down-stream 500 KB of each lead SNP. Function = functional variant classification based on position in or outside of a gene; CADD = Combined Annotation-Dependent depletion score, which predict how deleterious the SNP effect is on protein structure/function (higher scores indicate more deleterious); RegulomeDB scores predict likelihood of regulatory functionality (lower scores indicate higher likelihood); NA = not annotated; minChrState = minimum chromatin state across 127 tissue types (lower scores indicate more open chromatin); commonChrState = most common chromatin state in 127 tissue types. Also shown are p-values and effect sizes (z-scores) from the original summary statistics. We also display results from gene mapping analyses, including mapping based on 1) SNPs' physical position (postMapFilt), 2) eQTL functionality (eqtlMapFilt) and 3) chromatin interactions (ciMapFilt). The following columns result from the LAVA analysis: LAVA.rho LAVA.p_rho - local genetic correlation between AD and eGFR and corresponding p-value are also shown, and the values is set to NA if LAVA algorithm failed to converge. Note: 0 indicates "No", and 1 indicates "Yes".

| cFDRlocusnum | CHR | Lead_SNP | Lead_BP | MinBP | MaxBP | conjFDR | AD_Z | AD_PVAL | eGFR_Z | eGFR_PVAL | nearestGene | dist | func | CADD | RDB | minChrState | commonChrState | posMapFilt | eqtlMapFilt | ciMapFilt | LAVA.rho | LAVA.rho.lower | LAVA.rho.upper | LAVA.p_rho |
|---|---|---|---|---|---|---|---|---|---|---|---|---|---|---|---|---|---|---|---|---|---|---|---|---|
| 1 | 2 | rs13422342 | 55159284 | 54659284 | 55659284 | 4.01E-02 | 4.08 | 4.51E-05 | -3.31 | 9.35E-04 | EML6 | 0 | intronic | 0.43 | 5 | 5 | 15 | 1 | 1 | 1 | -0.11 | -0.65 | 0.37 | 6.14E-01 |
| 2 | 6 | rs9381599 | 47662484 | 47162484 | 48162484 | 2.93E-02 | 4.16 | 3.24E-05 | 3.42 | 6.30E-04 | GPR111:GPR115 | 0 | intronic | 4.57 | 5 | 4 | 15 | 1 | 1 | 1 | 0.54 | -0.02 | 1.00 | 5.37E-02 |
| 3 | 8 | rs10429294 | 95969322 | 95469322 | 96469322 | 4.89E-02 | -3.98 | 6.81E-05 | -3.68 | 2.30E-04 | NDUFAF6 | 0 | intronic | 1.56 | 7 | 5 | 5 | 1 | 1 | 1 | 0.28 | -0.15 | 0.82 | 2.05E-01 |
| 4 | 10 | rs1870148 | 82271341 | 81771341 | 82771341 | 1.32E-02 | -4.31 | 1.64E-05 | 5.14 | 2.69E-07 | TSPAN14 | 0 | intronic | 0.18 | 4 | 4 | 4 | 1 | 1 | 1 | -0.06 | -0.44 | 0.32 | 7.53E-01 |
| 5 | 11 | rs3824869 | 47378245 | 46878245 | 47878245 | 5.77E-06 | 5.81 | 6.19E-09 | 7.64 | 2.11E-14 | SPI1 | 0 | intronic | 3.02 | 2b | 3 | 15 | 1 | 1 | 1 | 0.54 | 0.26 | 0.86 | 4.57E-04 |
| 6 | 11 | rs648270 | 85726818 | 85226818 | 86226818 | 4.24E-03 | -4.56 | 5.01E-06 | 4.02 | 5.89E-05 | PICALM | 0 | intronic | 3.40 | 5 | 4 | 5 | 1 | 1 | 1 | -0.63 | -1.00 | -0.06 | 3.34E-02 |
| 7 | 15 | rs905450 | 82444437 | 81944437 | 82944437 | 4.34E-02 | -4.02 | 5.94E-05 | -4.48 | 7.33E-06 | EFTUD1 | 0 | exonic | 0.65 | 7 | 4 | 4 | 1 | 1 | 1 | NA | NA | NA | NA |
| 8 | 16 | rs2158031 | 20516594 | 20016594 | 21016594 | 3.57E-02 | -4.07 | 4.78E-05 | -7.11 | 1.12E-12 | AC137056.1 | 15612 | intergenic | 1.19 | 5 | 9 | 15 | 0 | 1 | 1 | 0.35 | 0.16 | 0.60 | 7.69E-04 |
| 9 | 17 | rs61182333 | 5133128 | 4633128 | 5633128 | 1.95E-02 | 4.22 | 2.48E-05 | 4.87 | 1.09E-06 | RP11-333E1.1:SCIMP | 0 | ncRNA_intronic | 3.45 | 5 | 2 | 5 | 1 | 1 | 1 | 0.48 | -0.01 | 1.00 | 4.98E-02 |
| 10 | 19 | rs139326841 | 45269725 | 44769725 | 45769725 | 1.86E-02 | 9.91 | 3.90E-23 | -3.57 | 3.62E-04 | AC092066.6 | 1003 | intergenic | 1.29 | 7 | 2 | 5 | 1 | 1 | 0 | -0.40 | -0.50 | -0.31 | 4.06E-15 |
| 11 | 19 | rs2927477 | 45313715 | 44813715 | 45813715 | 1.76E-02 | -7.84 | 4.40E-15 | 3.58 | 3.38E-04 | BCAM | 0 | intronic | 15.16 | | 1 | 15 | 1 | 1 | 1 | -0.40 | -0.50 | -0.31 | 4.06E-15 |
| 12 | 19 | rs11666329 | 45354296 | 44854296 | 45854296 | 6.05E-04 | 13.14 | 1.90E-39 | -4.53 | 5.88E-06 | PVRL2 | 0 | intronic | 3.70 | 5 | 4 | 5 | 1 | 1 | 1 | -0.40 | -0.50 | -0.31 | 4.06E-15 |
| 13 | 19 | rs157580 | 45395266 | 44895266 | 45895266 | 2.21E-05 | -25.80 | 8.13E-147 | 5.28 | 1.30E-07 | TOMM40 | 0 | intronic | 6.84 | 1f | 1 | 2 | 1 | 1 | 1 | -0.40 | -0.50 | -0.31 | 4.06E-15 |
| 13 | 19 | rs115881343 | 45403216 | 44903216 | 45903216 | 5.27E-04 | 21.19 | 1.18E-99 | -4.56 | 5.03E-06 | TOMM40 | 0 | intronic | 4.05 | 6 | 3 | 4 | 1 | 1 | 1 | -0.40 | -0.50 | -0.31 | 4.06E-15 |

| cFDRlocusnum | CHR | Lead_SNP | Lead_BP | MinBP | MaxBP | conjFDR | AD_Z | AD_PVAL | eGFR_Z | eGFR_PVAL | nearestGene | dist | func | CADD | RDB | minChrState | commonChrState | posMapFilt | eqtlMapFilt | ciMapFilt | LAVA.rho | LAVA.rho.lower | LAVA.rho.upper | LAVA.p_rho |
|---|---|---|---|---|---|---|---|---|---|---|---|---|---|---|---|---|---|---|---|---|---|---|---|---|
| 13 | 19 | rs7412 | 45412079 | 44912079 | 45912079 | 1.30E-04 | -14.84 | 8.14E-50 | 4.89 | 1.00E-06 | APOE | 0 | exonic | 25.10 | 4 | 1 | 14 | 1 | 1 | 1 | -0.40 | -0.50 | -0.31 | 4.06E-15 |
| 16 | 19 | rs16980051 | 46345886 | 45845886 | 46845886 | 3.66E-02 | -4.06 | 4.92E-05 | -5.07 | 3.92E-07 | SYMPK | 0 | intronic | 2.25 | 5 | 4 | 4 | 1 | 1 | 1 | -0.40 | -0.50 | -0.31 | 4.06E-15 |

**Supplementary Table S20. Distinct Pleiotropic Genomic Loci in European Ancestry (Kunkle Uncorrected, conjFDR<0.05)**

This table reports distinct genomic loci identified as pleiotropic using the Kunkle AD GWAS without competing risk correction. The most strongly associated lead SNPs in independent genomic loci are shown after including regions up and down-stream 500 KB of each lead SNP. Function = functional variant classification based on position in or outside of a gene; CADD = Combined Annotation-Dependent depletion score, which predict how deleterious the SNP effect is on protein structure/function (higher scores indicate more deleterious); RegulomeDB scores predict likelihood of regulatory functionality (lower scores indicate higher likelihood); NA = not annotated; minChrState = minimum chromatin state across 127 tissue types (lower scores indicate more open chromatin); commonChrState = most common chromatin state in 127 tissue types. Also shown are p-values and effect sizes (z-scores) from the original summary statistics. We also display results from gene mapping analyses, including mapping based on 1) SNPs' physical position (postMapFilt), 2) eQTL functionality (eqtlMapFilt) and 3) chromatin interactions (ciMapFilt). The following columns result from the LAVA analysis: LAVA.rho LAVA.p_rho - local genetic correlation between AD and eGFR and corresponding p-value are also shown, and the values is set to NA if LAVA algorithm failed to converge. Note: 0 indicates "No", and 1 indicates "Yes".

| cFDRlocusnum | CHR | Lead_SNP | Lead_BP | MinBP | MaxBP | conjFDR | AD_Z | AD_PVAL | eGFR_Z | eGFR_PVAL | nearestGene | dist | func | CADD | RDB | minChrState | commonChrState | posMapFilt | eqtlMapFilt | ciMapFilt | LAVA.rho | LAVA.rho.lower | LAVA.rho.upper | LAVA.p_rho |
|---|---|---|---|---|---|---|---|---|---|---|---|---|---|---|---|---|---|---|---|---|---|---|---|---|
| 1 | 6 | rs9381599 | 47662484 | 47162484 | 48162484 | 2.75E-02 | 4.31 | 1.62E-05 | 3.43 | 6.00E-04 | GPR111:GPR115 | 0 | intronic | 4.57 | 5 | 4 | 15 | 1 | 1 | 1 | 0.23 | -0.49 | 1.00 | 4.71E-01 |
| 2 | 10 | rs1870148 | 82271341 | 81771341 | 82771341 | 7.26E-03 | -4.47 | 8.00E-06 | 5.16 | 2.41E-07 | TSPAN14 | 0 | intronic | 0.18 | 4 | 4 | 4 | 1 | 1 | 1 | -0.15 | -0.56 | 0.22 | 4.29E-01 |
| 3 | 11 | rs12798036 | 947585 | 447585 | 1447585 | 4.68E-02 | -4.01 | 6.07E-05 | 4.94 | 7.97E-07 | AP2A2 | 0 | intronic | 4.45 | 7 | 4 | 5 | 1 | 1 | 0 | NA | NA | NA | NA |
| 4 | 11 | rs3740688 | 47380340 | 46880340 | 47880340 | 7.02E-06 | -6.41 | 1.41E-10 | -5.50 | 3.70E-08 | SPI1 | 0 | exonic | 0.37 | 3a | 1 | 15 | 1 | 1 | 1 | 0.45456 | 0.18 | 0.78 | 2.64E-03 |
| 5 | 11 | rs592297 | 85725937 | 85225937 | 86225937 | 3.47E-03 | -4.64 | 3.47E-06 | 4.05 | 5.04E-05 | PICALM | 0 | exonic | 9.96 | 6 | 4 | 5 | 1 | 1 | 1 | NA | NA | NA | NA |
| 6 | 11 | rs17745474 | 85836969 | 85336969 | 86336969 | 4.33E-02 | 4.03 | 5.54E-05 | -3.37 | 7.39E-04 | RNU6-560P | 27299 | intergenic | 5.63 | 4 | 5 | 15 | 0 | 1 | 1 | NA | NA | NA | NA |
| 7 | 15 | rs3131575 | 50731154 | 50231154 | 51231154 | 4.93E-02 | -4.00 | 6.45E-05 | 3.33 | 8.82E-04 | USP8 | 0 | intronic | 6.15 |  | 4 | 5 | 1 | 1 | 1 | 0.49 | 0.07 | 1.00 | 2.50E-02 |
| 8 | 15 | rs905450 | 82444437 | 81944437 | 82944437 | 3.50E-02 | -4.09 | 4.34E-05 | -4.50 | 6.74E-06 | EFTUD1 | 0 | exonic | 0.65 | 7 | 4 | 4 | 1 | 1 | 1 | NA | NA | NA | NA |
| 9 | 16 | rs7500658 | 20531940 | 20031940 | 21031940 | 1.66E-02 | -4.27 | 1.91E-05 | -7.30 | 2.92E-13 | ACSM2B | 15607 | intergenic | 0.35 | 6 | 5 | 15 | 0 | 1 | 1 | 0.23 | 0.00 | 0.51 | 4.13E-02 |
| 10 | 17 | rs61182333 | 5133128 | 4633128 | 5633128 | 2.17E-02 | 4.21 | 2.57E-05 | 4.89 | 9.92E-07 | RP11-333E1.1:SCIMP | 0 | ncRNA_intronic | 3.45 | 5 | 2 | 5 | 1 | 1 | 1 | 0.33 | -0.08 | 0.88 | 1.05E-01 |
| 11 | 19 | rs139326841 | 45269725 | 44769725 | 45769725 | 1.73E-02 | 9.82 | 9.35E-23 | -3.58 | 3.43E-04 | AC092066.6 | 1003 | intergenic | 1.29 | 7 | 2 | 5 | 1 | 1 | 0 | -0.34 | -0.43 | -0.25 | 2.01E-12 |

| cFDRlocusnum | CHR | Lead_SNP | Lead_BP | MinBP | MaxBP | conjFDR | AD_Z | AD_PVAL | eGFR_Z | eGFR_PVAL | nearestGene | dist | func | CADD | RDB | minChrState | commonChrState | posMapFilt | eqtlMapFilt | ciMapFilt | LAVA.rho | LAVA.rho.lower | LAVA.rho.upper | LAVA.p_rho |
|---|---|---|---|---|---|---|---|---|---|---|---|---|---|---|---|---|---|---|---|---|---|---|---|---|
| 12 | 19 | rs2927477 | 45313715 | 44813715 | 45813715 | 1.62E-02 | -7.72 | 1.18E-14 | 3.60 | 3.20E-04 | BCAM | 0 | intronic | 15.16 | | 1 | 15 | 1 | 1 | 1 | -0.34 | -0.43 | -0.25 | 2.01E-12 |
| 13 | 19 | rs11666329 | 45354296 | 44854296 | 45854296 | 5.45E-04 | 13.40 | 6.38E-41 | -4.55 | 5.40E-06 | PVRL2 | 0 | intronic | 3.70 | 5 | 4 | 5 | 1 | 1 | 1 | -0.34 | -0.43 | -0.25 | 2.01E-12 |
| 14 | 19 | rs157580 | 45395266 | 44895266 | 45895266 | 1.95E-05 | 26.03 | 2.06E-149 | 5.30 | 1.16E-07 | TOMM40 | 0 | intronic | 6.84 | 1f | 1 | 2 | 1 | 1 | 1 | -0.34 | -0.43 | -0.25 | 2.01E-12 |
| 14 | 19 | rs115881343 | 45403216 | 44903216 | 45903216 | 4.77E-04 | 21.34 | 4.51E-101 | -4.58 | 4.62E-06 | TOMM40 | 0 | intronic | 4.05 | 6 | 3 | 4 | 1 | 1 | 1 | -0.34 | -0.43 | -0.25 | 2.01E-12 |
| 14 | 19 | rs7412 | 45412079 | 44912079 | 45912079 | 1.17E-04 | -15.18 | 5.05E-52 | 4.91 | 9.07E-07 | APOE | 0 | exonic | 25.10 | 4 | 1 | 14 | 1 | 1 | 1 | -0.34 | -0.43 | -0.25 | 2.01E-12 |
| 17 | 19 | rs16980051 | 46345886 | 45845886 | 46845886 | 1.85E-02 | -4.25 | 2.14E-05 | -5.09 | 3.53E-07 | SYMPK | 0 | intronic | 2.25 | 5 | 4 | 4 | 1 | 1 | 1 | -0.34 | -0.43 | -0.25 | 2.01E-12 |

**Supplementary Table S21. Distinct Pleiotropic Genomic Loci in European Ancestry (Wightman Validation, conjFDR<0.05)**

This table reports distinct genomic loci identified as pleiotropic in the validation dataset using the Wightman AD GWAS (corrected for competing risk). The table includes comprehensive annotation for each locus: identifiers, chromosomal coordinates, lead SNP information, conjFDR values, z-scores and p-values for both traits, functional annotations (CADD scores, RegulomeDB scores, chromatin states), gene mapping results from multiple approaches, and LAVA-derived local genetic correlations and significance. The most strongly associated lead SNPs in independent genomic loci are shown after including regions up and down-stream 500 KB of each lead SNP. Function = functional variant classification based on position in or outside of a gene; CADD = Combined Annotation-Dependent depletion score, which predict how deleterious the SNP effect is on protein structure/function (higher scores indicate more deleterious); RegulomeDB scores predict likelihood of regulatory functionality (lower scores indicate higher likelihood); NA = not annotated; minChrState = minimum chromatin state across 127 tissue types (lower scores indicate more open chromatin); commonChrState = most common chromatin state in 127 tissue types. Also shown are p-values and effect sizes (z-scores) from the original summary statistics. We also display results from gene mapping analyses, including mapping based on 1) SNPs' physical position (postMapFilt), 2) eQTL functionality (eqtlMapFilt) and 3) chromatin interactions (ciMapFilt). The following columns result from the LAVA analysis: LAVA.rho LAVA.p_rho - local genetic correlation between AD and eGFR and corresponding p-value are also shown, and the values is set to NA if LAVA algorithm failed to converge. Note: 0 indicates "No", and 1 indicates "Yes".

| cFDRlocusnum | CHR | Lead_SNP | Lead_BP | MinBP | MaxBP | conjFDR | AD_Z | AD_PVAL | eGFR_Z | eGFR_PVAL | nearestGene | dist | func | CADD | RDB | minChrState | commonChrState | posMapFilt | eqtlMapFilt | ciMapFilt | LAVA.rho | LAVA.rho.lower | LAVA.rho.upper | LAVA.p_rho |
|---|---|---|---|---|---|---|---|---|---|---|---|---|---|---|---|---|---|---|---|---|---|---|---|---|
| 1 | 1 | rs9442398 | 1021695 | 521695 | 1521695 | 3.44E-02 | -3.91 | 9.33E-05 | 3.72 | 2.00E-04 | C1orf159 | 0 | intronic | 0.13 | 3a | 3 | 15 | 1 | 1 | 1 | -0.23 | -0.73 | 0.13 | 2.05E-01 |
| 2 | 1 | rs6427397 | 157705725 | 157205725 | 158205725 | 2.06E-02 | -4.06 | 4.88E-05 | 3.41 | 6.41E-04 | FCRL2 | 9797 | intergenic | 0.93 | 6 | 5 | 15 | 1 | 1 | 1 | NA | NA | NA | NA |
| 3 | 1 | rs511104 | 200873170 | 200373170 | 201373170 | 2.10E-02 | 4.37 | 1.23E-05 | 3.37 | 7.63E-04 | C1orf106 | 0 | intronic | 0.34 | 5 | 4 | 15 | 1 | 1 | 1 | -0.20 | -0.79 | 0.22 | 3.26E-01 |
| 4 | 2 | rs78572881 | 43762959 | 43262959 | 44262959 | 3.38E-02 | -3.91 | 9.08E-05 | 9.00 | 2.35E-19 | THADA | 0 | intronic | 2.95 | 6 | 4 | 5 | 1 | 1 | 1 | -0.27 | -0.62 | 0.03 | 8.33E-02 |
| 5 | 2 | rs7558958 | 238863486 | 238363486 | 239363486 | 1.12E-02 | -4.23 | 2.32E-05 | 3.88 | 1.06E-04 | UBE2F | 11982 | intergenic | 2.96 | 5 | 5 | 14 | 0 | 1 | 1 | 0.44 | -0.16 | 1.00 | 1.42E-01 |
| 6 | 3 | rs3920075 | 152073055 | 151573055 | 152573055 | 1.16E-02 | -4.22 | 2.43E-05 | -4.58 | 4.66E-06 | MBNL1 | 0 | intronic | 4.24 | 2c | 4 | 5 | 1 | 1 | 1 | NA | NA | NA | NA |

| cFDRlocusnum | CHR | Lead_SNP | Lead_BP | MinBP | MaxBP | conjFDR | AD_Z | AD_PVAL | eGFR_Z | eGFR_PVAL | nearestGene | dist | func | CADD | RDB | minChrState | commonChrState | posMapFilt | eqtlMapFilt | ciMapFilt | LAVA.rho | LAVA.rho.lower | LAVA.rho.upper | LAVA.p_rho |
|---|---|---|---|---|---|---|---|---|---|---|---|---|---|---|---|---|---|---|---|---|---|---|---|---|
| 7 | 5 | rs62380787 | 141045976 | 140545976 | 141545976 | 3.24E-02 | -3.93 | 8.62E-05 | 3.66 | 2.55E-04 | ARAP3 | 0 | intronic | 2.84 | 5 | 4 | 5 | 1 | 1 | 1 | -0.27 | -0.83 | 0.18 | 2.29E-01 |
| 8 | 6 | rs60917185 | 34611000 | 34111000 | 35111000 | 3.19E-02 | -4.04 | 5.26E-05 | 3.23 | 1.26E-03 | C6orf106 | 0 | intronic | 0.03 | 6 | 4 | 5 | 1 | 1 | 1 | 0.26 | 0.03 | 0.50 | 3.26E-02 |
| 9 | 6 | rs9381599 | 47662484 | 47162484 | 48162484 | 1.77E-02 | 4.19 | 2.78E-05 | 3.42 | 6.25E-04 | GPR111:GPR115 | 0 | intronic | 4.57 | 5 | 4 | 15 | 1 | 1 | 1 | 0.54 | -0.02 | 1.00 | 5.37E-02 |
| 10 | 6 | rs7761311 | 131311108 | 130811108 | 131811108 | 4.10E-02 | 3.85 | 1.17E-04 | -5.36 | 8.13E-08 | EPB41L2 | 0 | intronic | 1.03 | 6 | 5 | 15 | 1 | 1 | 1 | -0.38 | -1.00 | 0.17 | 1.69E-01 |
| 11 | 7 | rs10275989 | 28159696 | 27659696 | 28659696 | 3.70E-02 | -3.88 | 1.02E-04 | 3.65 | 2.67E-04 | JAZF1 | 0 | intronic | 0.33 | 7 | 5 | 15 | 1 | 1 | 1 | NA | NA | NA | NA |
| 12 | 7 | rs2734897 | 100159567 | 99659567 | 100659567 | 3.08E-02 | -3.94 | 8.09E-05 | 4.01 | 6.02E-05 | AGFG2 | 0 | intronic | 7.90 |  | 3 | 5 | 1 | 1 | 1 | -0.51 | -1.00 | -0.05 | 3.05E-02 |
| 13 | 8 | rs7835415 | 145093404 | 144593404 | 145593404 | 4.19E-02 | 3.84 | 1.21E-04 | 3.60 | 3.16E-04 | SPATC1 | 0 | intronic | 0.03 | 7 | 5 | 14 | 1 | 1 | 1 | 0.23 | -0.03 | 0.50 | 9.08E-02 |
| 14 | 10 | rs12783714 | 11490281 | 10990281 | 11990281 | 5.80E-03 | -4.66 | 3.19E-06 | -3.77 | 1.64E-04 | USP6NL | 5663 | intergenic | 2.81 | 4 | 2 | 15 | 1 | 1 | 0 | -0.20 | -0.93 | 0.30 | 3.89E-01 |
| 15 | 10 | rs10748526 | 82273079 | 81773079 | 82773079 | 1.85E-03 | -4.68 | 2.89E-06 | 5.23 | 1.68E-07 | TSPAN14 | 0 | intronic | 7.41 | 1f | 4 | 4 | 1 | 1 | 1 | -0.45 | -1.00 | -0.03 | 4.08E-02 |
| 16 | 10 | rs12267227 | 82317158 | 81817158 | 82817158 | 2.47E-02 | 4.01 | 6.11E-05 | -4.46 | 8.34E-06 | SH2D4B | 0 | intronic | 3.71 | 7 | 13 | 15 | 1 | 1 | 1 | -0.45 | -1.00 | -0.03 | 4.08E-02 |
| 17 | 10 | rs4752256 | 116577161 | 116077161 | 117077161 | 4.34E-02 | -3.83 | 1.27E-04 | 3.29 | 1.00E-03 | FAM160B1 | 4341 | intergenic | 3.95 | 7 | 5 | 15 | 1 | 1 | 1 | 0.13 | -0.28 | 0.57 | 5.05E-01 |
| 18 | 10 | rs79057333 | 124235187 | 123735187 | 124735187 | 1.14E-02 | 4.50 | 6.92E-06 | 3.56 | 3.69E-04 | HTRA1 | 0 | intronic | 6.49 | 2b | 2 | 7 | 1 | 1 | 1 | -0.21 | -0.92 | 0.37 | 4.45E-01 |
| 19 | 11 | rs10437655 | 47391948 | 46891948 | 47891948 | 2.74E-07 | 6.29 | 3.13E-10 | 8.82 | 1.16E-18 | SPI1 | 0 | intronic | 0.04 | 7 | 3 | 15 | 1 | 1 | 1 | 0.64 | 0.36 | 0.97 | 4.34E-05 |
| 21 | 11 | rs4980517 | 63745677 | 63245677 | 64245677 | 4.64E-02 | 3.84 | 1.24E-04 | 3.11 | 1.85E-03 | AP000721.4 | 0 | intronic | 4.01 | 7 | 4 | 5 | 1 | 1 | 1 | 0.05 | -0.30 | 0.39 | 7.66E-01 |
| 22 | 11 | rs592297 | 85725937 | 85225937 | 86225937 | 2.42E-03 | -6.15 | 7.87E-10 | 4.04 | 5.34E-05 | PICALM | 0 | exonic | 9.96 | 6 | 4 | 5 | 1 | 1 | 1 | -0.36 | -1.00 | 0.17 | 1.79E-01 |
| 22 | 11 | rs12284629 | 85804432 | 85304432 | 86304432 | 2.23E-02 | 4.12 | 3.80E-05 | -3.35 | 8.18E-04 | snoU13 | 801 | upstream | 0.54 | 5 | 5 | 15 | 0 | 1 | 1 | -0.36 | -1.00 | 0.17 | 1.79E-01 |
| 23 | 11 | rs35483388 | 122545146 | 122045146 | 123045146 | 2.67E-02 | 3.99 | 6.72E-05 | -5.20 | 2.05E-07 | UBASH3B | 0 | intronic | 0.44 | 7 | 5 | 15 | 1 | 1 | 1 | -0.21 | -0.65 | 0.19 | 2.88E-01 |
| 24 | 12 | rs11169943 | 52273248 | 51773248 | 52773248 | 4.01E-02 | 3.86 | 1.14E-04 | 5.65 | 1.61E-08 | ANKRD33 | 8495 | intergenic | 0.18 | 7 | 5 | 15 | 1 | 1 | 1 | -0.05 | -0.55 | 0.42 | 8.10E-01 |
| 25 | 12 | rs2657878 | 56866962 | 56366962 | 57366962 | 9.67E-03 | 4.27 | 1.96E-05 | 6.50 | 8.04E-11 | GLS2 | 0 | intronic | 5.62 |  | 4 | 5 | 1 | 1 | 1 | 0.24 | -0.20 | 0.88 | 2.62E-01 |
| 26 | 14 | rs11626787 | 103991749 | 103491749 | 104491749 | 2.16E-02 | -4.05 | 5.11E-05 | -3.36 | 7.88E-04 | RP11-600F24.7 | 0 | ncRNA_exonic | 2.67 | 4 | 1 | 14 | 1 | 1 | 1 | 0.38 | -0.05 | 0.94 | 8.07E-02 |
| 27 | 15 | rs117618017 | 63569902 | 63069902 | 64069902 | 6.46E-04 | 4.91 | 9.05E-07 | -5.81 | 6.16E-09 | APH1B | 0 | exonic | 10.57 | 4 | 1 | 1 | 1 | 1 | 1 | -0.56 | -0.90 | -0.29 | 1.05E-04 |
| 28 | 15 | rs2457497 | 82431219 | 81931219 | 82931219 | 3.82E-03 | -4.51 | 6.60E-06 | -3.91 | 9.39E-05 | EFTUD1 | 0 | intronic | 0.64 |  | 4 | 5 | 1 | 1 | 1 | -0.06 | -0.33 | 0.21 | 6.72E-01 |
| 29 | 16 | rs9939774 | 30068354 | 29568354 | 30568354 | 2.63E-02 | -4.04 | 5.27E-05 | 3.29 | 9.98E-04 | ALDOA | 0 | intronic | 12.11 | 6 | 4 | 5 | 1 | 1 | 1 | -0.05 | -0.43 | 0.31 | 7.66E-01 |
| 30 | 16 | rs11113 | 516038 | 511038 | 521038 | 4.56E- | - | 1.35E- | -3.29 | 9.85E-04 | HNRNPA1 | 757 | intergenic | 4.25 |  | 7 | 15 | 0 | 0 | 1 | -0.21 | -0.62 | 0.16 | 2.62E-01 |

| cFDRlocusnum | CHR | Lead_SNP | Lead_BP | MinBP | MaxBP | conjFDR | AD_Z | AD_PVAL | eGFR_Z | eGFR_PVAL | nearestGene | dist | func | CADD | RDB | minChrState | commonChrState | posMapFilt | eqtlMapFilt | ciMapFilt | LAVA.rho | LAVA.rho.lower | LAVA.rho.upper | LAVA.p_rho |
|---|---|---|---|---|---|---|---|---|---|---|---|---|---|---|---|---|---|---|---|---|---|---|---|---|
| | | 16 | 99 | 99 | 99 | 02 | 3.82 | 04 | | | P48 | 67 | | | | | | | | | | | | |
| 31 | 16 | rs7192890 | 71452475 | 70952475 | 71952475 | 9.07E-03 | 4.29 | 1.81E-05 | 4.11 | 3.96E-05 | RP11-510M2.1 | 0 | ncRNA_intronic | 0.34 | 7 | 5 | 14 | 0 | 1 | 1 | 0.13 | -0.14 | 0.41 | 3.31E-01 |
| 32 | 17 | rs4790812 | 1618135 | 1118135 | 2118135 | 5.09E-03 | -4.44 | 9.19E-06 | 5.59 | 2.21E-08 | MIR22HG | 0 | ncRNA_intronic | 23.00 | 2a | 1 | 1 | 1 | 1 | 1 | -0.36 | -0.64 | -0.10 | 8.37E-03 |
| 33 | 17 | rs12937244 | 1646317 | 1146317 | 2146317 | 4.89E-04 | 4.97 | 6.72E-07 | -4.82 | 1.46E-06 | SERPINF2 | 0 | intronic | 4.93 | 2b | 1 | 15 | 1 | 1 | 1 | -0.36 | -0.64 | -0.10 | 8.37E-03 |
| 34 | 17 | rs75511804 | 5138304 | 4638304 | 5638304 | 9.14E-03 | 4.29 | 1.83E-05 | 4.45 | 8.59E-06 | RP11-333E1.1 | 0 | ncRNA_exonic | 3.25 | 4 | 1 | 15 | 1 | 1 | 1 | 0.22 | -0.18 | 0.67 | 2.69E-01 |
| 35 | 17 | rs61217632 | 17775706 | 17275706 | 18275706 | 2.48E-02 | -4.01 | 6.15E-05 | -4.55 | 5.26E-06 | TOM1L2 | 0 | intronic | 1.74 | 6 | 4 | 5 | 1 | 1 | 1 | 0.44 | 0.06 | 1.00 | 2.59E-02 |
| 36 | 17 | rs11657989 | 29395746 | 28895746 | 29895746 | 7.33E-03 | 4.34 | 1.40E-05 | 3.70 | 2.18E-04 | RP11-848P1.9 | 0 | ncRNA_intronic | 1.59 | 1f | 5 | 15 | 0 | 1 | 1 | 0.59 | 0.14 | 1.00 | 1.44E-02 |
| 37 | 17 | rs9898829 | 60554275 | 60054275 | 61054275 | 3.11E-02 | -3.94 | 8.20E-05 | 5.21 | 1.87E-07 | TLK2 | 0 | intronic | 1.76 | 7 | 5 | 15 | 1 | 1 | 0 | -0.08 | -0.42 | 0.24 | 6.13E-01 |
| 38 | 18 | rs686836 | 32623344 | 32123344 | 33123344 | 6.76E-03 | 4.47 | 7.76E-06 | -3.72 | 1.98E-04 | MAPRE2 | 0 | intronic | 6.54 | 4 | 1 | 7 | 1 | 1 | 1 | -0.67 | -1.00 | -0.10 | 2.40E-02 |
| 39 | 19 | rs172032 | 18633755 | 18133755 | 19133755 | 4.90E-02 | 3.82 | 1.36E-04 | 3.15 | 1.61E-03 | ELL | 817 | upstream | 2.06 | 4 | 1 | 5 | 1 | 1 | 1 | 0.69 | 0.30 | 1.00 | 1.09E-03 |
| 40 | 19 | rs73041776 | 33372243 | 32872243 | 33872243 | 5.16E-03 | -4.43 | 9.36E-06 | -5.90 | 3.68E-09 | CEP89 | 0 | intronic | 0.44 | 6 | 4 | 5 | 1 | 1 | 1 | 0.25 | 0.02 | 0.49 | 3.30E-02 |
| 41 | 19 | rs139326841 | 45269725 | 44769725 | 45769725 | 1.11E-02 | 9.71 | 2.65E-22 | -3.57 | 3.59E-04 | AC092066.6 | 1003 | intergenic | 1.29 | 7 | 2 | 5 | 1 | 1 | 0 | -0.35 | -0.45 | -0.25 | 7.03E-11 |
| 42 | 19 | rs2927477 | 45313715 | 44813715 | 45813715 | 1.05E-02 | -11.41 | 3.54E-30 | 3.59 | 3.35E-04 | BCAM | 0 | intronic | 15.16 | | 1 | 15 | 1 | 1 | 1 | -0.35 | -0.45 | -0.25 | 7.03E-11 |
| 43 | 19 | rs28399637 | 45324138 | 44824138 | 45824138 | 3.48E-03 | 26.32 | 1.03E-152 | -3.93 | 8.55E-05 | BCAM | 0 | intronic | 0.11 | 3a | 1 | 15 | 1 | 1 | 1 | -0.35 | -0.45 | -0.25 | 7.03E-11 |
| 43 | 19 | rs10410835 | 45358353 | 44858353 | 45858353 | 3.85E-02 | -19.05 | 6.92E-81 | 3.16 | 1.58E-03 | PVRL2 | 0 | intronic | 0.34 | 6 | 4 | 5 | 1 | 1 | 1 | -0.35 | -0.45 | -0.25 | 7.03E-11 |
| 43 | 19 | rs394353 | 45367473 | 44867473 | 45867473 | 2.13E-04 | -6.40 | 1.57E-10 | -4.70 | 2.63E-06 | PVRL2 | 0 | intronic | 3.54 | | 4 | 5 | 1 | 1 | 1 | -0.35 | -0.45 | -0.25 | 7.03E-11 |
| 44 | 19 | rs157580 | 45395266 | 44895266 | 45895266 | 1.66E-05 | -33.67 | 1.63E-248 | 5.28 | 1.28E-07 | TOMM40 | 0 | intronic | 6.84 | 1f | 1 | 2 | 1 | 1 | 1 | -0.35 | -0.45 | -0.25 | 7.03E-11 |
| 44 | 19 | rs115881343 | 45403216 | 44903216 | 45903216 | 3.57E-04 | 28.62 | 4.18E-180 | -4.57 | 4.97E-06 | TOMM40 | 0 | intronic | 4.05 | 6 | 3 | 4 | 1 | 1 | 1 | -0.35 | -0.45 | -0.25 | 7.03E-11 |
| 44 | 19 | rs10119 | 45406673 | 44906673 | 45906673 | 1.71E-05 | 37.50 | 8.80E-308 | -5.28 | 1.32E-07 | TOMM40 | 0 | UTR3 | 3.09 | | 2 | 5 | 1 | 1 | 1 | -0.35 | -0.45 | -0.25 | 7.03E-11 |
| 44 | 19 | rs7412 | 45412079 | 44912079 | 45912079 | 9.49E-05 | -21.94 | 1.12E-106 | 4.89 | 9.86E-07 | APOE | 0 | exonic | 25.10 | 4 | 1 | 14 | 1 | 1 | 1 | -0.35 | -0.45 | -0.25 | 7.03E-11 |
| 47 | 19 | rs1964272 | 46190268 | 45690268 | 46690268 | 7.94E-03 | -4.32 | 1.55E-05 | -4.00 | 6.38E-05 | SNRPD2 | 443 | downstream | 2.14 | 7 | 4 | 4 | 1 | 1 | 1 | -0.35 | -0.45 | -0.25 | 7.03E-11 |
| 48 | 19 | rs146335962 | 47154484 | 46654484 | 47654484 | 2.87E-03 | -5.75 | 8.87E-09 | 3.99 | 6.69E-05 | DACT3 | 0 | UTR3 | 1.54 | 6 | 4 | 14 | 1 | 1 | 0 | -0.13 | -0.34 | 0.08 | 2.26E-01 |

| cFDRlocusnum | CHR | Lead_SNP | Lead_BP | MinBP | MaxBP | conjFDR | AD_Z | AD_PVAL | eGFR_Z | eGFR_PVAL | nearestGene | dist | func | CADD | RDB | minChrState | commonChrState | posMapFilt | eqtlMapFilt | ciMapFilt | LAVA.rho | LAVA.rho.lower | LAVA.rho.upper | LAVA.p_rho |
|---|---|---|---|---|---|---|---|---|---|---|---|---|---|---|---|---|---|---|---|---|---|---|---|---|
| 49 | 19 | rs479486 | 49229323 | 48729323 | 49729323 | 2.82E-03 | 4.58 | 4.66E-06 | 4.23 | 2.31E-05 | RASIP1 | 0 | intronic | 1.27 | 6 | 5 | 5 | 1 | 1 | 1 | 0.26 | -0.00 | 0.54 | 5.20E-02 |
| 50 | 19 | rs731170 | 55176262 | 54676262 | 55676262 | 9.29E-03 | 4.28 | 1.86E-05 | 4.66 | 3.09E-06 | LILRB4 | 0 | exonic | 0.80 | 5 | 5 | 15 | 1 | 1 | 1 | 0.20 | -0.14 | 0.59 | 2.47E-01 |
| 51 | 20 | rs6084183 | 2811868 | 2311868 | 3311868 | 3.67E-02 | 3.89 | 1.01E-04 | 4.80 | 1.59E-06 | PCED1A | 4091 | intergenic | 1.81 | 6 | 5 | 14 | 1 | 1 | 1 | 0.17 | -0.27 | 0.72 | 4.24E-01 |
| 52 | 20 | rs4813726 | 4966941 | 4466941 | 5466941 | 3.89E-02 | -4.61 | 4.05E-06 | 3.16 | 1.60E-03 | SLC23A2 | 0 | intronic | 0.05 | 7 | 4 | 15 | 1 | 1 | 1 | -0.39 | -0.78 | -0.07 | 1.98E-02 |
| 53 | 20 | rs645544 | 48501606 | 48001606 | 49001606 | 2.95E-02 | -3.96 | 7.64E-05 | 4.11 | 4.03E-05 | SLC9A8 | 0 | intronic | 0.16 | 5 | 4 | 4 | 1 | 1 | 1 | NA | NA | NA | NA |
| 54 | 20 | rs6069736 | 54983075 | 54483075 | 55483075 | 4.67E-02 | -6.38 | 1.76E-10 | 3.09 | 2.01E-03 | CSTF1 | 3556 | intergenic | 2.41 | 6 | 4 | 4 | 1 | 1 | 1 | NA | NA | NA | NA |
| 55 | 22 | rs9611460 | 41418297 | 40918297 | 41918297 | 3.83E-02 | 3.87 | 1.07E-04 | -3.36 | 7.77E-04 | Y_RNA | 23203 | intergenic | 11.71 | 4 | 1 | 1 | 0 | 1 | 0 | -0.20 | -0.58 | 0.12 | 2.06E-01 |

**Supplementary Table S22. Distinct Pleiotropic Genomic Loci in European Ancestry (Wightman Uncorrected, conjFDR<0.05)**

This table presents conjFDR results using the Wightman AD GWAS summary statistics without competing risk correction. The most strongly associated lead SNPs in independent genomic loci are shown after including regions up and down-stream 500 KB of each lead SNP. Function = functional variant classification based on position in or outside of a gene; CADD = Combined Annotation-Dependent depletion score, which predict how deleterious the SNP effect is on protein structure/function (higher scores indicate more deleterious); RegulomeDB scores predict likelihood of regulatory functionality (lower scores indicate higher likelihood); NA = not annotated; minChrState = minimum chromatin state across 127 tissue types (lower scores indicate more open chromatin); commonChrState = most common chromatin state in 127 tissue types. Also shown are p-values and effect sizes (z-scores) from the original summary statistics. We also display results from gene mapping analyses, including mapping based on 1) SNPs' physical position (postMapFilt), 2) eQTL functionality (eqtlMapFilt) and 3) chromatin interactions (ciMapFilt). The following columns result from the LAVA analysis: LAVA.rho LAVA.p_rho - local genetic correlation between AD and eGFR and corresponding p-value are also shown, and the values is set to NA if LAVA algorithm failed to converge. Note: 0 indicates "No", and 1 indicates "Yes".

| cFDRlocusnum | CHR | Lead_SNP | Lead_BP | MinBP | MaxBP | conjFDR | AD_Z | AD_PVAL | eGFR_Z | eGFR_PVAL | nearestGene | dist | func | CADD | RDB | minChrState | commonChrState | posMapFilt | eqtlMapFilt | ciMapFilt | LAVA.rho | LAVA.rho.lower | LAVA.rho.upper | LAVA.p_rho |
|---|---|---|---|---|---|---|---|---|---|---|---|---|---|---|---|---|---|---|---|---|---|---|---|---|
| 1 | 1 | rs4075116 | 1003629 | 503629 | 1503629 | 3.15E-02 | 3.96 | 7.53E-05 | 3.56 | 3.73E-04 | RP11-465B22.3 | 0 | ncRNA_intronic | 4.14 |  | 1 | 14 | 1 | 1 | 0 | -0.13 | -0.25 | -0.00 | 4.32E-02 |
| 2 | 1 | rs6427397 | 157705725 | 157205725 | 158205725 | 2.44E-02 | -4.03 | 5.48E-05 | 3.43 | 6.02E-04 | FCRL2 | 9797 | intergenic | 0.93 | 6 | 5 | 15 | 1 | 1 | 1 | 0.49 | -0.12 | 1.00 | 1.06E-01 |
| 3 | 1 | rs511104 | 200873170 | 200373170 | 201373170 | 2.33E-02 | -4.44 | 8.81E-06 | 3.38 | 7.19E-04 | C1orf106 | 0 | intronic | 0.34 | 5 | 4 | 15 | 1 | 1 | 1 | 0.29 | 0.11 | 0.47 | 1.71E-03 |
| 4 | 2 | rs268134 | 65608363 | 65108363 | 66108363 | 2.76E-02 | 4.00 | 6.41E-05 | 4.80 | 1.55E-06 | SPRED2 | 0 | intronic | 1.32 |  | 4 | 5 | 1 | 1 | 1 | 0.39 | 0.08 | 0.79 | 1.61E-02 |
| 5 | 2 | rs7607448 | 238859288 | 238359288 | 239359288 | 2.61E-02 | 4.01 | 5.95E-05 | 3.88 | 1.03E-04 | UBE2F | 16180 | intergenic | 0.42 | 7 | 5 | 15 | 0 | 1 | 1 | 0.02 | -0.21 | 0.27 | 8.38E-01 |
| 6 | 3 | rs3920075 | 152073055 | 151573055 | 152573055 | 1.68E-02 | 4.14 | 3.47E-05 | -4.60 | 4.18E-06 | MBNL1 | 0 | intronic | 4.24 | 2c | 4 | 5 | 1 | 1 | 1 | -0.13 | -0.45 | 0.16 | 3.68E-01 |
| 7 | 5 | rs62380787 | 141045976 | 140545976 | 141545976 | 1.96E-02 | 4.10 | 4.18E-05 | 3.68 | 2.38E-04 | ARAP3 | 0 | intronic | 2.84 | 5 | 4 | 5 | 1 | 1 | 1 | 0.20 | -0.06 | 0.48 | 1.26E-01 |

| cFDRlocusnum | CHR | Lead_SNP | Lead_BP | MinBP | MaxBP | conjFDR | AD_Z | AD_PVAL | eGFR_Z | eGFR_PVAL | nearestGene | dist | func | CADD | RDB | minChrState | commonChrState | posMapFilt | eqtlMapFilt | ciMapFilt | LAVA.rho | LAVA.rho.lower | LAVA.rho.upper | LAVA.p_rho |
|---|---|---|---|---|---|---|---|---|---|---|---|---|---|---|---|---|---|---|---|---|---|---|---|---|
| 8 | 6 | rs60917185 | 34611000 | 34111000 | 35111000 | 3.52E-02 | -4.37 | 1.27E-05 | 3.24 | 1.19E-03 | C6orf106 | 0 | intronic | 0.03 | 6 | 4 | 5 | 1 | 1 | 1 | 0.06 | -0.11 | 0.22 | 5.05E-01 |
| 9 | 6 | rs9381599 | 47662484 | 47162484 | 48162484 | 1.97E-02 | 4.31 | 1.62E-05 | 3.44 | 5.88E-04 | GPR111:GPR115 | 0 | intronic | 4.57 | 5 | 4 | 15 | 1 | 1 | 1 | 0.11 | -0.29 | 0.52 | 5.81E-01 |
| 10 | 7 | rs10275989 | 28159696 | 27659696 | 28659696 | 3.17E-02 | 3.96 | 7.62E-05 | 3.66 | 2.49E-04 | JAZF1 | 0 | intronic | 0.33 | 7 | 5 | 15 | 1 | 1 | 1 | NA | NA | NA | NA |
| 11 | 7 | rs117791102 | 99531026 | 99031026 | 100031026 | 4.96E-02 | 3.82 | 1.36E-04 | -3.30 | 9.74E-04 | RP4-604G5.1 | 0 | ncRNA_intronic | 0.29 | 5 | 5 | 14 | 1 | 1 | 1 | 0.19 | 0.02 | 0.37 | 3.41E-02 |
| 12 | 7 | rs2734897 | 100159567 | 99659567 | 100659567 | 1.68E-02 | -4.14 | 3.47E-05 | 4.03 | 5.53E-05 | AGFG2 | 0 | intronic | 7.90 | | 3 | 5 | 1 | 1 | 1 | 0.19 | 0.02 | 0.37 | 3.41E-02 |
| 13 | 8 | rs61732533 | 145108151 | 144608151 | 145608151 | 2.78E-02 | 4.00 | 6.46E-05 | 8.86 | 7.82E-19 | OPLAH | 0 | exonic | 15.58 | 4 | 3 | 13 | 1 | 1 | 1 | 0.07 | -0.04 | 0.19 | 2.00E-01 |
| 14 | 10 | rs12783714 | 11490281 | 10990281 | 11990281 | 6.44E-03 | -4.72 | 2.39E-06 | -3.79 | 1.52E-04 | USP6NL | 5663 | intergenic | 2.81 | 4 | 2 | 15 | 1 | 1 | 0 | 0.39 | 0.13 | 0.70 | 3.10E-03 |
| 15 | 10 | rs10748526 | 82273079 | 81773079 | 82773079 | 1.83E-03 | -4.70 | 2.58E-06 | 5.26 | 1.46E-07 | TSPAN14 | 0 | intronic | 7.41 | 1f | 4 | 4 | 1 | 1 | 1 | -0.28 | -0.50 | -0.07 | 1.04E-02 |
| 16 | 10 | rs12267227 | 82317158 | 81817158 | 82817158 | 2.89E-02 | -3.98 | 6.77E-05 | -4.48 | 7.52E-06 | SH2D4B | 0 | intronic | 3.71 | 7 | 13 | 15 | 1 | 1 | 1 | -0.28 | -0.50 | -0.07 | 1.04E-02 |
| 17 | 10 | rs4752256 | 116577161 | 116077161 | 117077161 | 4.47E-02 | -3.85 | 1.18E-04 | 3.31 | 9.45E-04 | FAM160B1 | 4341 | intergenic | 3.95 | 7 | 5 | 15 | 1 | 1 | 1 | 0.30 | -0.02 | 0.67 | 6.94E-02 |
| 18 | 10 | rs76935275 | 124166835 | 123666835 | 124666835 | 9.54E-03 | -4.29 | 1.76E-05 | 3.79 | 1.53E-04 | PLEKHA1 | 0 | intronic | 3.39 | 7 | 4 | 5 | 1 | 1 | 1 | -0.33 | -0.97 | 0.14 | 1.68E-01 |
| 19 | 11 | rs10437655 | 47391948 | 46891948 | 47891948 | 3.64E-07 | 6.26 | 3.94E-10 | 8.86 | 7.88E-19 | SPI1 | 0 | intronic | 0.04 | 7 | 3 | 15 | 1 | 1 | 1 | -0.03 | -0.20 | 0.14 | 7.45E-01 |
| 20 | 11 | rs509701 | 63774597 | 63274597 | 64274597 | 3.83E-02 | -3.91 | 9.32E-05 | 3.22 | 1.28E-03 | MACROD1 | 0 | intronic | 7.12 | 3a | 1 | 7 | 1 | 1 | 1 | 0.28 | 0.08 | 0.48 | 6.75E-03 |
| 21 | 11 | rs592297 | 85725937 | 85225937 | 86225937 | 2.63E-03 | 6.26 | 3.97E-10 | 4.06 | 4.91E-05 | PICALM | 0 | exonic | 9.96 | 6 | 4 | 5 | 1 | 1 | 1 | -0.32 | -1.00 | 0.08 | 1.12E-01 |
| 21 | 11 | rs12271390 | 85804696 | 85304696 | 86304696 | 2.11E-02 | -4.37 | 1.25E-05 | -3.41 | 6.39E-04 | snoU13 | 537 | upstream | 5.74 | 5 | 5 | 15 | 0 | 1 | 1 | -0.32 | -1.00 | 0.08 | 1.12E-01 |
| 22 | 11 | rs35483388 | 122545146 | 122045146 | 123045146 | 2.81E-02 | 3.99 | 6.53E-05 | -5.22 | 1.78E-07 | UBASH3B | 0 | intronic | 0.44 | 7 | 5 | 15 | 1 | 1 | 1 | -0.10 | -0.30 | 0.11 | 3.49E-01 |
| 23 | 12 | rs11169943 | 52273248 | 51773248 | 52773248 | 4.54E-02 | 3.84 | 1.21E-04 | 5.68 | 1.37E-08 | ANKRD33 | 8495 | intergenic | 0.18 | 7 | 5 | 15 | 1 | 1 | 1 | -0.24 | -0.99 | 0.28 | 3.39E-01 |
| 24 | 12 | rs2657878 | 56866962 | 56366962 | 57366962 | 1.01E-02 | 4.28 | 1.89E-05 | 6.53 | 6.49E-11 | GLS2 | 0 | intronic | 5.62 | | 4 | 5 | 1 | 1 | 1 | -0.17 | -0.39 | 0.05 | 1.21E-01 |
| 25 | 14 | rs45441198 | 104002611 | 103502611 | 104502611 | 2.86E-02 | 4.04 | 5.42E-05 | -3.31 | 9.22E-04 | TRMT61A | 0 | UTR3 | 6.54 | 5 | 4 | 5 | 1 | 1 | 1 | 0.38 | 0.15 | 0.62 | 1.46E-03 |
| 26 | 15 | rs117618017 | 63569902 | 63069902 | 64069902 | 7.69E-04 | 4.89 | 9.99E-07 | -5.84 | 5.19E-09 | APH1B | 0 | exonic | 10.57 | 4 | 1 | 1 | 1 | 1 | 1 | -0.18 | -0.47 | 0.10 | 1.95E-01 |
| 27 | 15 | rs12909668 | 79043949 | 78543949 | 79543949 | 4.62E-02 | -3.84 | 1.24E-04 | -6.54 | 5.97E-11 | RP11-160C18.4 | 499 | upstream | 5.00 | 5 | 1 | 15 | 1 | 1 | 1 | 0.07 | -0.18 | 0.32 | 5.67E-01 |
| 28 | 15 | rs8025381 | 81330552 | 80830552 | 81830552 | 4.18E-02 | -3.87 | 1.08E-04 | -3.57 | 3.57E-04 | C15orf26 | 0 | intronic | 0.48 | 5 | 5 | 15 | 1 | 1 | 1 | -0.19 | -0.34 | -0.03 | 1.83E-02 |
| 29 | 15 | rs111972258 | 82462544 | 81962544 | 82962544 | 4.35E-03 | -4.50 | 6.96E-06 | -4.69 | 2.73E-06 | EFTUD1 | 0 | intronic | 1.56 | 6 | 4 | 5 | 1 | 1 | 1 | -0.19 | -0.34 | -0.03 | 1.83E-02 |
| 30 | 16 | rs99397 | 300683 | 295683 | 305683 | 2.91E- | - | 3.95E- | 3.31 | 9.42E-04 | ALDOA | 0 | intronic | 12.1 | 6 | 4 | 5 | 1 | 1 | 1 | 0.38 | 0.20 | 0.56 | 5.37E-05 |

| cFDRlocusnum | CHR | Lead_SNP | Lead_BP | MinBP | MaxBP | conjFDR | AD_Z | AD_PVAL | eGFR_Z | eGFR_PVAL | nearestGene | dist | func | CADD | RDB | minChrState | commonChrState | posMapFilt | eqtlMapFilt | ciMapFilt | LAVA.rho | LAVA.rho.lower | LAVA.rho.upper | LAVA.p_rho |
|---|---|---|---|---|---|---|---|---|---|---|---|---|---|---|---|---|---|---|---|---|---|---|---|---|
| | | 74 | 54 | 54 | 54 | 02 | 4.11 | 05 | | | | | | 1 | | | | | | | | | | |
| 31 | 16 | rs1111316 | 51603899 | 51103899 | 52103899 | 3.66E-02 | 3.91 | 9.12E-05 | -3.31 | 9.30E-04 | HNRNPA1P48 | 75767 | intergenic | 4.25 | | 7 | 15 | 0 | 0 | 1 | -0.13 | -0.39 | 0.12 | 2.94E-01 |
| 32 | 16 | rs7192890 | 71452475 | 70952475 | 71952475 | 1.40E-02 | -4.19 | 2.78E-05 | 4.13 | 3.63E-05 | RP11-510M2.1 | 0 | ncRNA_intronic | 0.34 | 7 | 5 | 14 | 0 | 1 | 1 | 0.19 | 0.01 | 0.38 | 3.35E-02 |
| 33 | 17 | rs4790812 | 1618135 | 1118135 | 2118135 | 1.24E-02 | -4.22 | 2.41E-05 | 5.62 | 1.89E-08 | MIR22HG | 0 | ncRNA_intronic | 23.00 | 2a | 1 | 1 | 1 | 1 | 1 | -0.18 | -0.49 | 0.11 | 2.12E-01 |
| 34 | 17 | rs9912287 | 1630992 | 1130992 | 2130992 | 5.92E-04 | 4.95 | 7.36E-07 | -4.48 | 7.54E-06 | WDR81:RP11-961A15.1 | 0 | exonic | 11.63 | 5 | 3 | 4 | 1 | 1 | 1 | -0.18 | -0.49 | 0.11 | 2.12E-01 |
| 35 | 17 | rs75511804 | 5138304 | 4638304 | 5638304 | 3.56E-03 | 4.54 | 5.51E-06 | 4.47 | 7.75E-06 | RP11-333E1.1 | 0 | ncRNA_exonic | 3.25 | 4 | 1 | 15 | 1 | 1 | 1 | 0.19 | -0.07 | 0.46 | 1.53E-01 |
| 36 | 17 | rs61217632 | 17775706 | 17275706 | 18275706 | 6.62E-03 | -4.39 | 1.14E-05 | -4.58 | 4.72E-06 | TOM1L2 | 0 | intronic | 1.74 | 6 | 4 | 5 | 1 | 1 | 1 | 0.13 | -0.03 | 0.29 | 1.16E-01 |
| 37 | 17 | rs11657989 | 29395746 | 28895746 | 29895746 | 8.12E-03 | 4.35 | 1.34E-05 | 3.72 | 2.03E-04 | RP11-848P1.9 | 0 | ncRNA_intronic | 1.59 | 1f | 5 | 15 | 0 | 1 | 1 | -0.02 | -0.26 | 0.22 | 8.81E-01 |
| 38 | 17 | rs112781672 | 60538932 | 60038932 | 61038932 | 2.66E-02 | 4.01 | 6.12E-05 | 5.12 | 3.13E-07 | TLK2 | 0 | intronic | 1.81 | 6 | 5 | 15 | 1 | 1 | 1 | 0.27 | 0.07 | 0.47 | 8.89E-03 |
| 39 | 18 | rs686836 | 32623344 | 32123344 | 33123344 | 7.53E-03 | -4.47 | 7.89E-06 | -3.74 | 1.84E-04 | MAPRE2 | 0 | intronic | 6.54 | 4 | 1 | 7 | 1 | 1 | 1 | -0.61 | -1.00 | -0.10 | 2.33E-02 |
| 40 | 19 | rs73041776 | 33372243 | 32872243 | 33872243 | 7.23E-03 | -4.37 | 1.27E-05 | -5.93 | 3.08E-09 | CEP89 | 0 | intronic | 0.44 | 6 | 4 | 5 | 1 | 1 | 1 | -0.31 | -0.59 | -0.05 | 2.14E-02 |
| 41 | 19 | rs139326841 | 45269725 | 44769725 | 45769725 | 1.23E-02 | -9.62 | 6.53E-22 | -3.59 | 3.36E-04 | AC092066.6 | 1003 | intergenic | 1.29 | 7 | 2 | 5 | 1 | 1 | 0 | -0.05 | -0.15 | 0.04 | 2.87E-01 |
| 42 | 19 | rs2927477 | 45313715 | 44813715 | 45813715 | 1.16E-02 | 11.22 | 3.23E-29 | 3.60 | 3.13E-04 | BCAM | 0 | intronic | 15.16 | | 1 | 15 | 1 | 1 | 1 | -0.05 | -0.15 | 0.04 | 2.87E-01 |
| 43 | 19 | rs28399637 | 45324138 | 44824138 | 45824138 | 3.81E-03 | 26.48 | 1.66E-154 | -3.95 | 7.88E-05 | BCAM | 0 | intronic | 0.11 | 3a | 1 | 15 | 1 | 1 | 1 | -0.05 | -0.15 | 0.04 | 2.87E-01 |
| 43 | 19 | rs10410835 | 45358353 | 44858353 | 45858353 | 4.23E-02 | -19.11 | 1.91E-81 | 3.18 | 1.49E-03 | PVRL2 | 0 | intronic | 0.34 | 6 | 4 | 5 | 1 | 1 | 1 | -0.05 | -0.15 | 0.04 | 2.87E-01 |
| 43 | 19 | rs394353 | 45367473 | 44867473 | 45867473 | 2.31E-04 | -6.36 | 1.98E-10 | -4.72 | 2.35E-06 | PVRL2 | 0 | intronic | 3.54 | | 4 | 5 | 1 | 1 | 1 | -0.05 | -0.15 | 0.04 | 2.87E-01 |
| 44 | 19 | rs157580 | 45395266 | 44895266 | 45895266 | 1.77E-05 | 33.82 | 1.15E-250 | 5.31 | 1.11E-07 | TOMM40 | 0 | intronic | 6.84 | 1f | 1 | 2 | 1 | 1 | 1 | 0.24 | 0.06 | 0.42 | 8.97E-03 |
| 44 | 19 | rs115881343 | 45403216 | 44903216 | 45903216 | 3.88E-04 | 28.70 | 4.40E-181 | -4.59 | 4.46E-06 | TOMM40 | 0 | intronic | 4.05 | 6 | 3 | 4 | 1 | 1 | 1 | 0.24 | 0.06 | 0.42 | 8.97E-03 |
| 44 | 19 | rs10119 | 45406673 | 44906673 | 45906673 | 1.82E-05 | 37.69 | 8.0702e-311 | -5.30 | 1.14E-07 | TOMM40 | 0 | UTR3 | 3.09 | | 2 | 5 | 1 | 1 | 1 | 0.24 | 0.06 | 0.42 | 8.97E-03 |
| 44 | 19 | rs7412 | 45412079 | 44912079 | 45912079 | 1.03E-04 | -22.42 | 2.49E-111 | 4.92 | 8.71E-07 | APOE | 0 | exonic | 25.10 | 4 | 1 | 14 | 1 | 1 | 1 | 0.24 | 0.06 | 0.42 | 8.97E-03 |
| 47 | 19 | rs1964272 | 46190268 | 45690268 | 46690268 | 3.82E-03 | -4.53 | 6.00E-06 | -4.02 | 5.87E-05 | SNRPD2 | 443 | downstream | 2.14 | 7 | 4 | 4 | 1 | 1 | 1 | 0.24 | 0.06 | 0.42 | 8.97E-03 |
| 48 | 19 | rs56010181 | 47198932 | 46698932 | 47698932 | 3.06E-03 | 4.58 | 4.64E-06 | 4.14 | 3.42E-05 | PRKD2 | 0 | intronic | 10.06 | 5 | 4 | 4 | 1 | 1 | 1 | 0.24 | 0.06 | 0.42 | 8.97E-03 |
| 49 | 19 | rs47948 | 492293 | 487293 | 497293 | 5.27E- | - | 8.72E- | 4.25 | 2.11E-05 | RASIP1 | 0 | intronic | 1.27 | 6 | 5 | 5 | 1 | 1 | 1 | -0.11 | -0.28 | 0.07 | 2.28E-01 |

| cFDRlocusnum | CHR | Lead_SNP | Lead_BP | MinBP | MaxBP | conjFDR | AD_Z | AD_PVAL | eGFR_Z | eGFR_PVAL | nearestGene | dist | func | CADD | RDB | minChrState | commonChrState | posMapFilt | eqtlMapFilt | ciMapFilt | LAVA.rho | LAVA.rho.lower | LAVA.rho.upper | LAVA.p_rho |
|---|---|---|---|---|---|---|---|---|---|---|---|---|---|---|---|---|---|---|---|---|---|---|---|---|
| | | 6 | 23 | 23 | 23 | 03 | 4.45 | 06 | | | | | | | | | | | | | | | | |
| 50 | 19 | rs731170 | 55176262 | 54676262 | 55676262 | 6.43E-03 | 4.40 | 1.10E-05 | 4.69 | 2.76E-06 | LILRB4 | 0 | exonic | 0.80 | 5 | 5 | 15 | 1 | 1 | 1 | -0.08 | -0.25 | 0.09 | 3.53E-01 |
| 51 | 20 | rs4813726 | 4966941 | 4466941 | 5466941 | 4.27E-02 | -4.51 | 6.46E-06 | 3.17 | 1.51E-03 | SLC23A2 | 0 | intronic | 0.05 | 7 | 4 | 15 | 1 | 1 | 1 | 0.22 | -0.14 | 0.62 | 2.15E-01 |
| 52 | 20 | rs645544 | 48501606 | 48001606 | 49001606 | 3.66E-02 | 3.91 | 9.11E-05 | 4.13 | 3.69E-05 | SLC9A8 | 0 | intronic | 0.16 | 5 | 4 | 4 | 1 | 1 | 1 | -0.26 | -0.58 | 0.05 | 9.75E-02 |
| 53 | 22 | rs9611460 | 41418297 | 40918297 | 41918297 | 2.36E-02 | -4.08 | 4.59E-05 | -3.38 | 7.31E-04 | Y_RNA | 23203 | intergenic | 11.71 | 4 | 1 | 1 | 0 | 1 | 0 | -0.15 | -0.36 | 0.05 | 1.52E-01 |

**Supplementary Table S23. Distinct Pleiotropic Genomic Loci in African American Ancestry (conjFDR<0.2)**

This table characterizes distinct genomic loci showing pleiotropy between AD and eGFR in African American ancestry individuals. Due to limited sample size in the African American AD GWAS, the significance threshold was set at conjFDR < 0.2 rather than < 0.05, identifying suggestive associations that warrant replication in larger studies. The most strongly associated lead SNPs in independent genomic loci are shown after including regions up and down-stream 500 KB of each lead SNP. Function = functional variant classification based on position in or outside of a gene; CADD = Combined Annotation-Dependent depletion score, which predict how deleterious the SNP effect is on protein structure/function (higher scores indicate more deleterious); RegulomeDB scores predict likelihood of regulatory functionality (lower scores indicate higher likelihood); NA = not annotated; minChrState = minimum chromatin state across 127 tissue types (lower scores indicate more open chromatin); commonChrState = most common chromatin state in 127 tissue types. Also shown are p-values and effect sizes (z-scores) from the original summary statistics. We also display results from gene mapping analyses, including mapping based on 1) SNPs' physical position (postMapFilt), 2) eQTL functionality (eqtlMapFilt) and 3) chromatin interactions (ciMapFilt). The following columns result from the LAVA analysis: LAVA.rho LAVA.p_rho - local genetic correlation between AD and eGFR and corresponding p-value are also shown, and the values is set to NA if LAVA algorithm failed to converge. Note: 0 indicates "No", and 1 indicates "Yes".

| cFDRlocusnum | CHR | Lead_SNP | Lead_BP | MinBP | MaxBP | conjFDR | AD_Z | AD_PVAL | eGFR_Z | eGFR_PVAL | nearestGene | dist | func | CADD | RDB | minChrState | commonChrState | posMapFilt | eqtlMapFilt | ciMapFilt | LAVA.rho | LAVA.rho.lower | LAVA.rho.upper | LAVA.p_rho |
|---|---|---|---|---|---|---|---|---|---|---|---|---|---|---|---|---|---|---|---|---|---|---|---|---|
| 1 | 6 | rs140069490 | 151539505 | 151039505 | 152039505 | 1.79E-01 | -3.64 | 2.69E-04 | -3.78 | 1.54E-04 | RP1-292B18.4 | 0 | ncRNA_intronic | 6.16 | 5 | 14 | 15 | 0 | 0 | 1 | NA | NA | NA | NA |
| 2 | 7 | rs10242748 | 70092296 | 69592296 | 70592296 | 1.31E-01 | 3.89 | 1.01E-04 | -3.72 | 1.97E-04 | AUTS2 | 0 | intronic | 4.66 | 6 | 5 | 15 | 1 | 0 | 0 | NA | NA | NA | NA |
| 3 | 11 | rs35302065 | 38895843 | 38395843 | 39395843 | 1.84E-01 | 3.64 | 2.73E-04 | 3.59 | 3.27E-04 | RP11-64I17.1 | 200051 | intergenic | 2.00 | 7 | 9 | 15 | 0 | 0 | 1 | NA | NA | NA | NA |
| 4 | 17 | rs12453781 | 6427027 | 5927027 | 6927027 | 1.43E-01 | -3.86 | 1.15E-04 | 3.69 | 2.21E-04 | PITPNM3 | 0 | intronic | 1.98 | 4 | 2 | 15 | 1 | 1 | 1 | 0.509894 | 0.15669 | 0.97726 | 0.00561344 |
| 5 | 19 | rs3943791 | 24326350 | 23826350 | 24826350 | 1.83E-01 | -3.63 | 2.78E-04 | -4.03 | 5.49E-05 | ZNF254 | 13695 | intergenic | 2.58 | 7 | 5 | 9 | 0 | 1 | 1 | NA | NA | NA | NA |

**Supplementary Table S24. Colocalization Analysis Results Using SuSiE Framework (Kunkle Discovery).**
This table presents results from colocalization analysis using the Sum of Single Effects (SuSiE) fine-mapping framework to assess whether AD and eGFR association signals at each conjFDR locus are driven by shared versus distinct causal variants. SuSiE accounts for linkage disequilibrium and identifies credible sets of potentially causal variants. For each locus tested, the table reports: traits analyzed (AD and eGFR), conjFDR locus number, number of SNPs analyzed, hit indices for credible sets in each trait, and posterior probabilities for five mutually exclusive hypotheses: H0 (no association with either trait), H1 (association with trait 1 only), H2 (association with trait 2 only), H3 (association with both traits but distinct causal variants), and H4 (association with both traits with shared causal variant). High posterior probability for H4 (PP.H4.abf) indicates strong evidence for a shared causal variant underlying both associations, supporting biological pleiotropy, while high PP.H3.abf suggests linkage of distinct causal variants. Note: 0 indicates "No", and 1 indicates "Yes".

| | | | coloc SUSIE method | | | | | | | | | | |
|---|---|---|---|---|---|---|---|---|---|---|---|---|---|
| **Trait1** | **Trait2** | **cFDRlocusnum** | **nsnps2** | **hit1** | **hit2** | **PP.H0.abf** | **PP.H1.abf** | **PP.H2.abf** | **PP.H3.abf** | **PP.H4.abf** | **PP.H3+PP.H4≥0.8** | **PP.H3/PP.H4≥3** | **PP.H4/PP.H3≥5** |
| AD Kunkle Corrected | eGFR | 5 | 1508 | rs67472071 | rs10838702 | 4.04E-18 | 2.09E-13 | 1.93E-05 | 9.97E-01 | 2.51E-03 | 1 | 0 | 0 |
| AD Kunkle Corrected | eGFR | 10 | 3068 | rs60049679 | rs429358 | 3.67E-60 | 5.34E-29 | 6.87E-32 | 1.00E+00 | 1.27E-26 | 1 | 0 | 0 |
| AD Kunkle Corrected | eGFR | | | rs12691088 | rs429358 | 2.62E-84 | 5.34E-29 | 4.91E-56 | 1.00E+00 | 7.74E-29 | 1 | 0 | 0 |
| AD Kunkle Corrected | eGFR | | | rs144311893 | rs429358 | 9.20E-217 | 5.34E-29 | 1.72E-188 | 1.00E+00 | 3.15E-31 | 1 | 0 | 0 |
| AD Kunkle Corrected | eGFR | | | rs77301115 | rs429358 | 1.06E-243 | 5.34E-29 | 1.99E-215 | 1.00E+00 | 8.59E-27 | 1 | 0 | 0 |
| AD Kunkle Corrected | eGFR | | | rs1160983 | rs429358 | 0.00E+00 | 5.34E-29 | 0.00E+00 | 1.00E+00 | 1.20E-27 | 1 | 0 | 0 |
| AD Kunkle | eGFR | | | rs41290120 | rs429358 | 0.00E+00 | 5.34E-29 | 0.00E+00 | 1.00E+00 | 6.21E-30 | 1 | 0 | 0 |

| | | | coloc SUSIE method | | | | | | | | | | |
|---|---|---|---|---|---|---|---|---|---|---|---|---|---|
| **Trait1** | **Trait 2** | **cFDRlocusnum** | **nsnps 2** | **hit1** | **hit2** | **PP.H0.abf** | **PP.H1.abf** | **PP.H2.abf** | **PP.H3.abf** | **PP.H4.abf** | **PP.H3+PP.H4≥0.8** | **PP.H3/PP.H4≥3** | **PP.H4/PP.H3≥5** |
| Corrected | | | | | | | | | | | | | |
| AD Kunkle Corrected | eGFR | | | rs75627662 | rs429358 | 0.00E+00 | 5.34E-29 | 0.00E+00 | 1.00E+00 | 3.98E-24 | 1 | 0 | 0 |
| AD Kunkle Corrected | eGFR | | | rs814573 | rs429358 | 0.00E+00 | 5.34E-29 | 0.00E+00 | 1.00E+00 | 3.56E-08 | 1 | 0 | 0 |
| AD Kunkle Corrected | eGFR | | | rs157595 | rs429358 | 0.00E+00 | 5.34E-29 | 0.00E+00 | 1.00E+00 | 3.73E-26 | 1 | 0 | 0 |
| AD Kunkle Corrected | eGFR | | | rs4803770 | rs429358 | 0.00E+00 | 5.34E-29 | 0.00E+00 | 1.00E+00 | 1.33E-29 | 1 | 0 | 0 |
| AD Kunkle Corrected | eGFR | | | rs60049679 | rs814573 | 4.81E-242 | 7.01E-211 | 6.87E-32 | 1.00E+00 | 1.07E-28 | 1 | 0 | 0 |
| AD Kunkle Corrected | eGFR | | | rs12691088 | rs814573 | 3.44E-266 | 7.01E-211 | 4.91E-56 | 1.00E+00 | 9.28E-55 | 1 | 0 | 0 |
| AD Kunkle Corrected | eGFR | | | rs144311893 | rs814573 | 0.00E+00 | 7.01E-211 | 1.72E-188 | 1.00E+00 | 4.92E-190 | 1 | 0 | 0 |
| AD Kunkle Corrected | eGFR | | | rs77301115 | rs814573 | 0.00E+00 | 7.01E-211 | 1.99E-215 | 1.00E+00 | 2.22E-191 | 1 | 0 | 0 |
| AD Kunkle Corrected | eGFR | | | rs1160983 | rs814573 | 0.00E+00 | 7.01E-211 | 0.00E+00 | 1.00E+00 | 1.96E-213 | 1 | 0 | 0 |
| AD Kunkle | eGFR | | | rs41290120 | rs814573 | 0.00E+00 | 7.01E-211 | 0.00E+00 | 1.00E+00 | 8.28E-213 | 1 | 0 | 0 |

| | | | coloc SUSIE method | | | | | | | | | | |
|---|---|---|---|---|---|---|---|---|---|---|---|---|---|
| **Trait1** | **Trait 2** | **cFDRlocusnum** | **nsnps 2** | **hit1** | **hit2** | **PP.H0.abf** | **PP.H1.abf** | **PP.H2.abf** | **PP.H3.abf** | **PP.H4.abf** | **PP.H3+PP.H4≥0.8** | **PP.H3/PP.H4≥3** | **PP.H4/PP.H3≥5** |
| Corrected | | | | | | | | | | | | | |
| AD Kunkle Corrected | eGFR | | | rs75627662 | rs814573 | 0.00E+00 | 7.01E-211 | 0.00E+00 | 1.00E+00 | 1.09E-141 | 1 | 0 | 0 |
| AD Kunkle Corrected | eGFR | | | rs814573 | rs814573 | 0.00E+00 | 1.40E-213 | 0.00E+00 | 0.00E+00 | 1.00E+00 | 1 | 1 | 1 |
| AD Kunkle Corrected | eGFR | | | rs157595 | rs814573 | 0.00E+00 | 7.01E-211 | 0.00E+00 | 1.00E+00 | 1.72E-193 | 1 | 0 | 0 |
| AD Kunkle Corrected | eGFR | | | rs4803770 | rs814573 | 0.00E+00 | 7.01E-211 | 0.00E+00 | 1.00E+00 | 2.34E-186 | 1 | 0 | 0 |
| AD Kunkle Corrected | eGFR | | | rs60049679 | rs12721046 | 4.49E-156 | 6.54E-125 | 6.87E-32 | 1.00E+00 | 8.64E-31 | 1 | 0 | 0 |
| AD Kunkle Corrected | eGFR | | | rs12691088 | rs12721046 | 3.21E-180 | 6.54E-125 | 4.91E-56 | 1.00E+00 | 3.97E-55 | 1 | 0 | 0 |
| AD Kunkle Corrected | eGFR | | | rs144311893 | rs12721046 | 1.13E-312 | 6.54E-125 | 1.72E-188 | 1.00E+00 | 6.42E-127 | 1 | 0 | 0 |
| AD Kunkle Corrected | eGFR | | | rs77301115 | rs12721046 | 0.00E+00 | 6.54E-125 | 1.99E-215 | 1.00E+00 | 5.00E-127 | 1 | 0 | 0 |
| AD Kunkle Corrected | eGFR | | | rs1160983 | rs12721046 | 0.00E+00 | 6.54E-125 | 0.00E+00 | 1.00E+00 | 2.78E-122 | 1 | 0 | 0 |
| AD Kunkle | eGFR | | | rs41290120 | rs12721046 | 0.00E+00 | 6.54E-125 | 0.00E+00 | 1.00E+00 | 1.24E-124 | 1 | 0 | 0 |

| | | | coloc SUSIE method | | | | | | | | | | |
|---|---|---|---|---|---|---|---|---|---|---|---|---|---|
| **Trait1** | **Trait 2** | **cFDRlocusnum** | **nsnps 2** | **hit1** | **hit2** | **PP.H0.abf** | **PP.H1.abf** | **PP.H2.abf** | **PP.H3.abf** | **PP.H4.abf** | **PP.H3+PP.H4≥0.8** | **PP.H3/PP.H4≥3** | **PP.H4/PP.H3≥5** |
| Corrected | | | | | | | | | | | | | |
| AD Kunkle Corrected | eGFR | | | rs75627662 | rs12721046 | 0.00E+00 | 6.54E-125 | 0.00E+00 | 1.00E+00 | 1.19E-90 | 1 | 0 | 0 |
| AD Kunkle Corrected | eGFR | | | rs814573 | rs12721046 | 0.00E+00 | 6.54E-125 | 0.00E+00 | 1.00E+00 | 3.28E-38 | 1 | 0 | 0 |
| AD Kunkle Corrected | eGFR | | | rs157595 | rs12721046 | 0.00E+00 | 6.54E-125 | 0.00E+00 | 1.00E+00 | 2.15E-110 | 1 | 0 | 0 |
| AD Kunkle Corrected | eGFR | | | rs4803770 | rs12721046 | 0.00E+00 | 6.54E-125 | 0.00E+00 | 1.00E+00 | 1.96E-117 | 1 | 0 | 0 |
| AD Kunkle Corrected | eGFR | | | rs60049679 | rs112019714 | 7.62E-55 | 1.11E-23 | 6.87E-32 | 1.00E+00 | 6.15E-22 | 1 | 0 | 0 |
| AD Kunkle Corrected | eGFR | | | rs12691088 | rs112019714 | 5.44E-79 | 1.11E-23 | 4.91E-56 | 1.00E+00 | 5.20E-26 | 1 | 0 | 0 |
| AD Kunkle Corrected | eGFR | | | rs144311893 | rs112019714 | 1.91E-211 | 1.11E-23 | 1.72E-188 | 1.00E+00 | 5.10E-26 | 1 | 0 | 0 |
| AD Kunkle Corrected | eGFR | | | rs77301115 | rs112019714 | 1.60E-238 | 8.07E-24 | 1.45E-215 | 7.27E-01 | 2.73E-01 | 1 | 0 | 0 |
| AD Kunkle Corrected | eGFR | | | rs1160983 | rs112019714 | 0.00E+00 | 1.11E-23 | 0.00E+00 | 1.00E+00 | 4.30E-23 | 1 | 0 | 0 |
| AD Kunkle | eGFR | | | rs41290120 | rs112019714 | 0.00E+00 | 1.11E-23 | 0.00E+00 | 1.00E+00 | 2.70E-25 | 1 | 0 | 0 |

| | | | coloc SUSIE method | | | | | | | | | | |
|---|---|---|---|---|---|---|---|---|---|---|---|---|---|
| **Trait1** | **Trait 2** | **cFDRlocusnum** | **nsnps 2** | **hit1** | **hit2** | **PP.H0.abf** | **PP.H1.abf** | **PP.H2.abf** | **PP.H3.abf** | **PP.H4.abf** | **PP.H3+PP.H4≥0.8** | **PP.H3/PP.H4≥3** | **PP.H4/PP.H3≥5** |
| Corrected | | | | | | | | | | | | | |
| AD Kunkle Corrected | eGFR | | | rs75627662 | rs112019714 | 0.00E+00 | 1.11E-23 | 0.00E+00 | 1.00E+00 | 2.78E-24 | 1 | 0 | 0 |
| AD Kunkle Corrected | eGFR | | | rs814573 | rs112019714 | 0.00E+00 | 1.11E-23 | 0.00E+00 | 1.00E+00 | 3.84E-23 | 1 | 0 | 0 |
| AD Kunkle Corrected | eGFR | | | rs157595 | rs112019714 | 0.00E+00 | 1.11E-23 | 0.00E+00 | 1.00E+00 | 1.11E-24 | 1 | 0 | 0 |
| AD Kunkle Corrected | eGFR | | | rs4803770 | rs112019714 | 0.00E+00 | 1.11E-23 | 0.00E+00 | 1.00E+00 | 5.52E-26 | 1 | 0 | 0 |
| AD Kunkle Corrected | eGFR | 11 | 3111 | rs60049679 | rs429358 | 3.99E-60 | 5.80E-29 | 6.87E-32 | 1.00E+00 | 1.37E-26 | 1 | 0 | 0 |
| AD Kunkle Corrected | eGFR | | | rs12691088 | rs429358 | 2.85E-84 | 5.80E-29 | 4.91E-56 | 1.00E+00 | 8.39E-29 | 1 | 0 | 0 |
| AD Kunkle Corrected | eGFR | | | rs144311893 | rs429358 | 9.99E-217 | 5.80E-29 | 1.72E-188 | 1.00E+00 | 3.42E-31 | 1 | 0 | 0 |
| AD Kunkle Corrected | eGFR | | | rs77301115 | rs429358 | 1.15E-243 | 5.80E-29 | 1.99E-215 | 1.00E+00 | 9.23E-27 | 1 | 0 | 0 |
| AD Kunkle Corrected | eGFR | | | rs1160983 | rs429358 | 0.00E+00 | 5.80E-29 | 0.00E+00 | 1.00E+00 | 1.31E-27 | 1 | 0 | 0 |
| AD | eGFR | | | rs41290120 | rs429358 | 0.00E+00 | 5.80E-29 | 0.00E+00 | 1.00E+00 | 6.72E-30 | 1 | 0 | 0 |

| | | | coloc SUSIE method | | | | | | | | | | |
|---|---|---|---|---|---|---|---|---|---|---|---|---|---|
| **Trait1** | **Trait 2** | **cFDRlocusnu m** | **nsnps 2** | **hit1** | **hit2** | **PP.H0.ab f** | **PP.H1.ab f** | **PP.H2.ab f** | **PP.H3.ab f** | **PP.H4.ab f** | **PP.H3+PP.H4≥0. 8** | **PP.H3/PP.H4≥ 3** | **PP.H4/PP.H3≥ 5** |
| Kunkle Corrected | | | | | | | | | | | | | |
| AD Kunkle Corrected | eGFR | | | rs75627662 | rs429358 | 0.00E+00 | 5.80E-29 | 0.00E+00 | 1.00E+00 | 4.22E-24 | 1 | 0 | 0 |
| AD Kunkle Corrected | eGFR | | | rs814573 | rs429358 | 0.00E+00 | 5.80E-29 | 0.00E+00 | 1.00E+00 | 3.60E-08 | 1 | 0 | 0 |
| AD Kunkle Corrected | eGFR | | | rs157595 | rs429358 | 0.00E+00 | 5.80E-29 | 0.00E+00 | 1.00E+00 | 4.04E-26 | 1 | 0 | 0 |
| AD Kunkle Corrected | eGFR | | | rs4803770 | rs429358 | 0.00E+00 | 5.80E-29 | 0.00E+00 | 1.00E+00 | 1.42E-29 | 1 | 0 | 0 |
| AD Kunkle Corrected | eGFR | | | rs60049679 | rs814573 | 5.83E-242 | 8.48E-211 | 6.87E-32 | 1.00E+00 | 1.07E-28 | 1 | 0 | 0 |
| AD Kunkle Corrected | eGFR | | | rs12691088 | rs814573 | 4.16E-266 | 8.48E-211 | 4.91E-56 | 1.00E+00 | 9.28E-55 | 1 | 0 | 0 |
| AD Kunkle Corrected | eGFR | | | rs144311893 | rs814573 | 0.00E+00 | 8.48E-211 | 1.72E-188 | 1.00E+00 | 4.92E-190 | 1 | 0 | 0 |
| AD Kunkle Corrected | eGFR | | | rs77301115 | rs814573 | 0.00E+00 | 8.48E-211 | 1.99E-215 | 1.00E+00 | 2.62E-191 | 1 | 0 | 0 |
| AD Kunkle Corrected | eGFR | | | rs1160983 | rs814573 | 0.00E+00 | 8.48E-211 | 0.00E+00 | 1.00E+00 | 2.38E-213 | 1 | 0 | 0 |
| AD | eGFR | | | rs41290120 | rs814573 | 0.00E+00 | 8.48E- | 0.00E+00 | 1.00E+00 | 1.00E- | 1 | 0 | 0 |

| | | | coloc SUSIE method | | | | | | | | | | |
|---|---|---|---|---|---|---|---|---|---|---|---|---|---|
| Trait1 | Trait 2 | cFDRlocusnum | nsnps 2 | hit1 | hit2 | PP.H0.abf | PP.H1.abf | PP.H2.abf | PP.H3.abf | PP.H4.abf | PP.H3+PP.H4≥0.8 | PP.H3/PP.H4≥3 | PP.H4/PP.H3≥5 |
| Kunkle Corrected | | | | | | | 211 | | | 212 | | | |
| AD Kunkle Corrected | eGFR | | | rs75627662 | rs814573 | 0.00E+00 | 8.48E-211 | 0.00E+00 | 1.00E+00 | 1.24E-141 | 1 | 0 | 0 |
| AD Kunkle Corrected | eGFR | | | rs814573 | rs814573 | 0.00E+00 | 1.70E-213 | 0.00E+00 | 0.00E+00 | 1.00E+00 | 1 | 1 | 1 |
| AD Kunkle Corrected | eGFR | | | rs157595 | rs814573 | 0.00E+00 | 8.48E-211 | 0.00E+00 | 1.00E+00 | 2.01E-193 | 1 | 0 | 0 |
| AD Kunkle Corrected | eGFR | | | rs4803770 | rs814573 | 0.00E+00 | 8.48E-211 | 0.00E+00 | 1.00E+00 | 2.84E-186 | 1 | 0 | 0 |
| AD Kunkle Corrected | eGFR | | | rs60049679 | rs12721046 | 4.47E-156 | 6.50E-125 | 6.87E-32 | 1.00E+00 | 8.64E-31 | 1 | 0 | 0 |
| AD Kunkle Corrected | eGFR | | | rs12691088 | rs12721046 | 3.19E-180 | 6.50E-125 | 4.91E-56 | 1.00E+00 | 3.97E-55 | 1 | 0 | 0 |
| AD Kunkle Corrected | eGFR | | | rs144311893 | rs12721046 | 1.12E-312 | 6.50E-125 | 1.72E-188 | 1.00E+00 | 6.40E-127 | 1 | 0 | 0 |
| AD Kunkle Corrected | eGFR | | | rs77301115 | rs12721046 | 0.00E+00 | 6.50E-125 | 1.99E-215 | 1.00E+00 | 4.97E-127 | 1 | 0 | 0 |
| AD Kunkle Corrected | eGFR | | | rs1160983 | rs12721046 | 0.00E+00 | 6.50E-125 | 0.00E+00 | 1.00E+00 | 2.79E-122 | 1 | 0 | 0 |
| AD | eGFR | | | rs41290120 | rs12721046 | 0.00E+00 | 6.50E- | 0.00E+00 | 1.00E+00 | 1.22E- | 1 | 0 | 0 |

| | | | coloc SUSIE method | | | | | | | | | | |
|---|---|---|---|---|---|---|---|---|---|---|---|---|---|
| **Trait1** | **Trait2** | **cFDRlocusnum** | **nsnps2** | **hit1** | **hit2** | **PP.H0.abf** | **PP.H1.abf** | **PP.H2.abf** | **PP.H3.abf** | **PP.H4.abf** | **PP.H3+PP.H4≥0.8** | **PP.H3/PP.H4≥3** | **PP.H4/PP.H3≥5** |
| Kunkle Corrected | | | | | | | 125 | | | 124 | | | |
| AD Kunkle Corrected | eGFR | | | rs75627662 | rs12721046 | 0.00E+00 | 6.50E-125 | 0.00E+00 | 1.00E+00 | 1.18E-90 | 1 | 0 | 0 |
| AD Kunkle Corrected | eGFR | | | rs814573 | rs12721046 | 0.00E+00 | 6.50E-125 | 0.00E+00 | 1.00E+00 | 3.23E-38 | 1 | 0 | 0 |
| AD Kunkle Corrected | eGFR | | | rs157595 | rs12721046 | 0.00E+00 | 6.50E-125 | 0.00E+00 | 1.00E+00 | 2.16E-110 | 1 | 0 | 0 |
| AD Kunkle Corrected | eGFR | | | rs4803770 | rs12721046 | 0.00E+00 | 6.50E-125 | 0.00E+00 | 1.00E+00 | 1.92E-117 | 1 | 0 | 0 |
| AD Kunkle Corrected | eGFR | | | rs60049679 | rs112019714 | 7.43E-55 | 1.08E-23 | 6.87E-32 | 1.00E+00 | 6.03E-22 | 1 | 0 | 0 |
| AD Kunkle Corrected | eGFR | | | rs12691088 | rs112019714 | 5.31E-79 | 1.08E-23 | 4.91E-56 | 1.00E+00 | 5.07E-26 | 1 | 0 | 0 |
| AD Kunkle Corrected | eGFR | | | rs144311893 | rs112019714 | 1.86E-211 | 1.08E-23 | 1.72E-188 | 1.00E+00 | 4.97E-26 | 1 | 0 | 0 |
| AD Kunkle Corrected | eGFR | | | rs77301115 | rs112019714 | 1.57E-238 | 7.87E-24 | 1.45E-215 | 7.27E-01 | 2.73E-01 | 1 | 0 | 0 |
| AD Kunkle Corrected | eGFR | | | rs1160983 | rs112019714 | 0.00E+00 | 1.08E-23 | 0.00E+00 | 1.00E+00 | 4.22E-23 | 1 | 0 | 0 |
| AD | eGFR | | | rs41290120 | rs11201971 | 0.00E+00 | 1.08E-23 | 0.00E+00 | 1.00E+00 | 2.63E-25 | 1 | 0 | 0 |

| | | | coloc SUSIE method | | | | | | | | | | |
|---|---|---|---|---|---|---|---|---|---|---|---|---|---|
| **Trait1** | **Trait 2** | **cFDRlocusnum** | **nsnps 2** | **hit1** | **hit2** | **PP.H0.abf** | **PP.H1.abf** | **PP.H2.abf** | **PP.H3.abf** | **PP.H4.abf** | **PP.H3+PP.H4≥0.8** | **PP.H3/PP.H4≥3** | **PP.H4/PP.H3≥5** |
| Kunkle Corrected | | | | | 4 | | | | | | | | |
| AD Kunkle Corrected | eGFR | | | rs75627662 | rs112019714 | 0.00E+00 | 1.08E-23 | 0.00E+00 | 1.00E+00 | 2.71E-24 | 1 | 0 | 0 |
| AD Kunkle Corrected | eGFR | | | rs814573 | rs112019714 | 0.00E+00 | 1.08E-23 | 0.00E+00 | 1.00E+00 | 3.75E-23 | 1 | 0 | 0 |
| AD Kunkle Corrected | eGFR | | | rs157595 | rs112019714 | 0.00E+00 | 1.08E-23 | 0.00E+00 | 1.00E+00 | 1.08E-24 | 1 | 0 | 0 |
| AD Kunkle Corrected | eGFR | | | rs4803770 | rs112019714 | 0.00E+00 | 1.08E-23 | 0.00E+00 | 1.00E+00 | 5.38E-26 | 1 | 0 | 0 |
| AD Kunkle Corrected | eGFR | 12 | 3122 | rs60049679 | rs429358 | 3.90E-60 | 5.67E-29 | 6.87E-32 | 1.00E+00 | 1.38E-26 | 1 | 0 | 0 |
| AD Kunkle Corrected | eGFR | | | rs12691088 | rs429358 | 2.78E-84 | 5.67E-29 | 4.91E-56 | 1.00E+00 | 8.05E-29 | 1 | 0 | 0 |
| AD Kunkle Corrected | eGFR | | | rs144311893 | rs429358 | 9.77E-217 | 5.67E-29 | 1.72E-188 | 1.00E+00 | 3.32E-31 | 1 | 0 | 0 |
| AD Kunkle Corrected | eGFR | | | rs77301115 | rs429358 | 1.13E-243 | 5.67E-29 | 1.99E-215 | 1.00E+00 | 9.07E-27 | 1 | 0 | 0 |
| AD Kunkle Corrected | eGFR | | | rs1160983 | rs429358 | 0.00E+00 | 5.67E-29 | 0.00E+00 | 1.00E+00 | 1.26E-27 | 1 | 0 | 0 |

| | | | coloc SUSIE method | | | | | | | | | | |
|---|---|---|---|---|---|---|---|---|---|---|---|---|---|
| **Trait1** | **Trait 2** | **cFDRlocusnum** | **nsnps 2** | **hit1** | **hit2** | **PP.H0.abf** | **PP.H1.abf** | **PP.H2.abf** | **PP.H3.abf** | **PP.H4.abf** | **PP.H3+PP.H4≥0.8** | **PP.H3/PP.H4≥3** | **PP.H4/PP.H3≥5** |
| AD Kunkle Corrected | eGFR | | | rs41290120 | rs429358 | 0.00E+00 | 5.67E-29 | 0.00E+00 | 1.00E+00 | 6.46E-30 | 1 | 0 | 0 |
| AD Kunkle Corrected | eGFR | | | rs75627662 | rs429358 | 0.00E+00 | 5.67E-29 | 0.00E+00 | 1.00E+00 | 4.29E-24 | 1 | 0 | 0 |
| AD Kunkle Corrected | eGFR | | | rs814573 | rs429358 | 0.00E+00 | 5.67E-29 | 0.00E+00 | 1.00E+00 | 3.58E-08 | 1 | 0 | 0 |
| AD Kunkle Corrected | eGFR | | | rs157595 | rs429358 | 0.00E+00 | 5.67E-29 | 0.00E+00 | 1.00E+00 | 3.97E-26 | 1 | 0 | 0 |
| AD Kunkle Corrected | eGFR | | | rs4803770 | rs429358 | 0.00E+00 | 5.67E-29 | 0.00E+00 | 1.00E+00 | 1.41E-29 | 1 | 0 | 0 |
| AD Kunkle Corrected | eGFR | | | rs60049679 | rs814573 | 4.38E-242 | 6.37E-211 | 6.87E-32 | 1.00E+00 | 1.07E-28 | 1 | 0 | 0 |
| AD Kunkle Corrected | eGFR | | | rs12691088 | rs814573 | 3.13E-266 | 6.37E-211 | 4.91E-56 | 1.00E+00 | 9.28E-55 | 1 | 0 | 0 |
| AD Kunkle Corrected | eGFR | | | rs144311893 | rs814573 | 0.00E+00 | 6.37E-211 | 1.72E-188 | 1.00E+00 | 4.92E-190 | 1 | 0 | 0 |
| AD Kunkle Corrected | eGFR | | | rs77301115 | rs814573 | 0.00E+00 | 6.37E-211 | 1.99E-215 | 1.00E+00 | 2.02E-191 | 1 | 0 | 0 |
| AD Kunkle Corrected | eGFR | | | rs1160983 | rs814573 | 0.00E+00 | 6.37E-211 | 0.00E+00 | 1.00E+00 | 1.78E-213 | 1 | 0 | 0 |

| | | | coloc SUSIE method | | | | | | | | | | |
|---|---|---|---|---|---|---|---|---|---|---|---|---|---|
| Trait1 | Trait 2 | cFDRlocusnum | nsnps 2 | hit1 | hit2 | PP.H0.abf | PP.H1.abf | PP.H2.abf | PP.H3.abf | PP.H4.abf | PP.H3+PP.H4≥0.8 | PP.H3/PP.H4≥3 | PP.H4/PP.H3≥5 |
| AD Kunkle Corrected | eGFR | | | rs41290120 | rs814573 | 0.00E+00 | 6.37E-211 | 0.00E+00 | 1.00E+00 | 7.64E-213 | 1 | 0 | 0 |
| AD Kunkle Corrected | eGFR | | | rs75627662 | rs814573 | 0.00E+00 | 6.37E-211 | 0.00E+00 | 1.00E+00 | 9.17E-142 | 1 | 0 | 0 |
| AD Kunkle Corrected | eGFR | | | rs814573 | rs814573 | 0.00E+00 | 1.27E-213 | 0.00E+00 | 0.00E+00 | 1.00E+00 | 1 | 1 | 1 |
| AD Kunkle Corrected | eGFR | | | rs157595 | rs814573 | 0.00E+00 | 6.37E-211 | 0.00E+00 | 1.00E+00 | 1.55E-193 | 1 | 0 | 0 |
| AD Kunkle Corrected | eGFR | | | rs4803770 | rs814573 | 0.00E+00 | 6.37E-211 | 0.00E+00 | 1.00E+00 | 2.13E-186 | 1 | 0 | 0 |
| AD Kunkle Corrected | eGFR | | | rs60049679 | rs12721046 | 3.95E-156 | 5.74E-125 | 6.87E-32 | 1.00E+00 | 8.64E-31 | 1 | 0 | 0 |
| AD Kunkle Corrected | eGFR | | | rs12691088 | rs12721046 | 2.82E-180 | 5.74E-125 | 4.91E-56 | 1.00E+00 | 3.97E-55 | 1 | 0 | 0 |
| AD Kunkle Corrected | eGFR | | | rs144311893 | rs12721046 | 9.89E-313 | 5.74E-125 | 1.72E-188 | 1.00E+00 | 5.58E-127 | 1 | 0 | 0 |
| AD Kunkle Corrected | eGFR | | | rs77301115 | rs12721046 | 0.00E+00 | 5.74E-125 | 1.99E-215 | 1.00E+00 | 4.39E-127 | 1 | 0 | 0 |
| AD Kunkle Corrected | eGFR | | | rs1160983 | rs12721046 | 0.00E+00 | 5.74E-125 | 0.00E+00 | 1.00E+00 | 2.42E-122 | 1 | 0 | 0 |

| | | | coloc SUSIE method | | | | | | | | | | |
|---|---|---|---|---|---|---|---|---|---|---|---|---|---|
| **Trait1** | **Trait 2** | **cFDRlocusnum** | **nsnps 2** | **hit1** | **hit2** | **PP.H0.abf** | **PP.H1.abf** | **PP.H2.abf** | **PP.H3.abf** | **PP.H4.abf** | **PP.H3+PP.H4≥0.8** | **PP.H3/PP.H4≥3** | **PP.H4/PP.H3≥5** |
| AD Kunkle Corrected | eGFR | | | rs41290120 | rs12721046 | 0.00E+00 | 5.74E-125 | 0.00E+00 | 1.00E+00 | 1.06E-124 | 1 | 0 | 0 |
| AD Kunkle Corrected | eGFR | | | rs75627662 | rs12721046 | 0.00E+00 | 5.74E-125 | 0.00E+00 | 1.00E+00 | 1.17E-90 | 1 | 0 | 0 |
| AD Kunkle Corrected | eGFR | | | rs814573 | rs12721046 | 0.00E+00 | 5.74E-125 | 0.00E+00 | 1.00E+00 | 3.10E-38 | 1 | 0 | 0 |
| AD Kunkle Corrected | eGFR | | | rs157595 | rs12721046 | 0.00E+00 | 5.74E-125 | 0.00E+00 | 1.00E+00 | 1.94E-110 | 1 | 0 | 0 |
| AD Kunkle Corrected | eGFR | | | rs4803770 | rs12721046 | 0.00E+00 | 5.74E-125 | 0.00E+00 | 1.00E+00 | 1.75E-117 | 1 | 0 | 0 |
| AD Kunkle Corrected | eGFR | | | rs60049679 | rs112019714 | 7.08E-55 | 1.03E-23 | 6.87E-32 | 1.00E+00 | 5.88E-22 | 1 | 0 | 0 |
| AD Kunkle Corrected | eGFR | | | rs12691088 | rs112019714 | 5.06E-79 | 1.03E-23 | 4.91E-56 | 1.00E+00 | 4.83E-26 | 1 | 0 | 0 |
| AD Kunkle Corrected | eGFR | | | rs144311893 | rs112019714 | 1.77E-211 | 1.03E-23 | 1.72E-188 | 1.00E+00 | 4.72E-26 | 1 | 0 | 0 |
| AD Kunkle Corrected | eGFR | | | rs77301115 | rs112019714 | 1.49E-238 | 7.50E-24 | 1.45E-215 | 7.27E-01 | 2.73E-01 | 1 | 0 | 0 |
| AD Kunkle Corrected | eGFR | | | rs1160983 | rs112019714 | 0.00E+00 | 1.03E-23 | 0.00E+00 | 1.00E+00 | 3.95E-23 | 1 | 0 | 0 |

| | | | coloc SUSIE method | | | | | | | | | | |
|---|---|---|---|---|---|---|---|---|---|---|---|---|---|
| **Trait1** | **Trait 2** | **cFDRlocusnum** | **nsnps 2** | **hit1** | **hit2** | **PP.H0.abf** | **PP.H1.abf** | **PP.H2.abf** | **PP.H3.abf** | **PP.H4.abf** | **PP.H3+PP.H4≥0.8** | **PP.H3/PP.H4≥3** | **PP.H4/PP.H3≥5** |
| AD Kunkle Corrected | eGFR | | | rs41290120 | rs112019714 | 0.00E+00 | 1.03E-23 | 0.00E+00 | 1.00E+00 | 2.48E-25 | 1 | 0 | 0 |
| AD Kunkle Corrected | eGFR | | | rs75627662 | rs112019714 | 0.00E+00 | 1.03E-23 | 0.00E+00 | 1.00E+00 | 2.55E-24 | 1 | 0 | 0 |
| AD Kunkle Corrected | eGFR | | | rs814573 | rs112019714 | 0.00E+00 | 1.03E-23 | 0.00E+00 | 1.00E+00 | 3.58E-23 | 1 | 0 | 0 |
| AD Kunkle Corrected | eGFR | | | rs157595 | rs112019714 | 0.00E+00 | 1.03E-23 | 0.00E+00 | 1.00E+00 | 1.03E-24 | 1 | 0 | 0 |
| AD Kunkle Corrected | eGFR | | | rs4803770 | rs112019714 | 0.00E+00 | 1.03E-23 | 0.00E+00 | 1.00E+00 | 5.13E-26 | 1 | 0 | 0 |
| AD Kunkle Corrected | eGFR | 13a | 3092 | rs60049679 | rs429358 | 3.98E-60 | 5.79E-29 | 6.87E-32 | 1.00E+00 | 1.47E-26 | 1 | 0 | 0 |
| AD Kunkle Corrected | eGFR | | | rs12691088 | rs429358 | 2.84E-84 | 5.79E-29 | 4.91E-56 | 1.00E+00 | 7.64E-29 | 1 | 0 | 0 |
| AD Kunkle Corrected | eGFR | | | rs144311893 | rs429358 | 9.98E-217 | 5.79E-29 | 1.72E-188 | 1.00E+00 | 3.44E-31 | 1 | 0 | 0 |
| AD Kunkle Corrected | eGFR | | | rs77301115 | rs429358 | 1.15E-243 | 5.79E-29 | 1.99E-215 | 1.00E+00 | 9.27E-27 | 1 | 0 | 0 |
| AD Kunkle Correcte | eGFR | | | rs1160983 | rs429358 | 0.00E+00 | 5.79E-29 | 0.00E+00 | 1.00E+00 | 1.32E-27 | 1 | 0 | 0 |

| | | | coloc SUSIE method | | | | | | | | | | |
|---|---|---|---|---|---|---|---|---|---|---|---|---|---|
| Trait1 | Trait 2 | cFDRlocusnum | nsnps 2 | hit1 | hit2 | PP.H0.abf | PP.H1.abf | PP.H2.abf | PP.H3.abf | PP.H4.abf | PP.H3+PP.H4≥0.8 | PP.H3/PP.H4≥3 | PP.H4/PP.H3≥5 |
| d | | | | | | | | | | | | | |
| AD Kunkle Corrected | eGFR | | | rs41290120 | rs429358 | 0.00E+00 | 5.79E-29 | 0.00E+00 | 1.00E+00 | 6.92E-30 | 1 | 0 | 0 |
| AD Kunkle Corrected | eGFR | | | rs75627662 | rs429358 | 0.00E+00 | 5.79E-29 | 0.00E+00 | 1.00E+00 | 4.05E-24 | 1 | 0 | 0 |
| AD Kunkle Corrected | eGFR | | | rs814573 | rs429358 | 0.00E+00 | 5.79E-29 | 0.00E+00 | 1.00E+00 | 3.51E-08 | 1 | 0 | 0 |
| AD Kunkle Corrected | eGFR | | | rs157595 | rs429358 | 0.00E+00 | 5.79E-29 | 0.00E+00 | 1.00E+00 | 4.12E-26 | 1 | 0 | 0 |
| AD Kunkle Corrected | eGFR | | | rs4803770 | rs429358 | 0.00E+00 | 5.79E-29 | 0.00E+00 | 1.00E+00 | 1.41E-29 | 1 | 0 | 0 |
| AD Kunkle Corrected | eGFR | | | rs60049679 | rs814573 | 1.12E-241 | 1.64E-210 | 6.87E-32 | 1.00E+00 | 1.07E-28 | 1 | 0 | 0 |
| AD Kunkle Corrected | eGFR | | | rs12691088 | rs814573 | 8.03E-266 | 1.64E-210 | 4.91E-56 | 1.00E+00 | 9.28E-55 | 1 | 0 | 0 |
| AD Kunkle Corrected | eGFR | | | rs144311893 | rs814573 | 0.00E+00 | 1.64E-210 | 1.72E-188 | 1.00E+00 | 4.92E-190 | 1 | 0 | 0 |
| AD Kunkle Corrected | eGFR | | | rs77301115 | rs814573 | 0.00E+00 | 1.64E-210 | 1.99E-215 | 1.00E+00 | 4.61E-191 | 1 | 0 | 0 |
| AD Kunkle Correcte | eGFR | | | rs1160983 | rs814573 | 0.00E+00 | 1.64E-210 | 0.00E+00 | 1.00E+00 | 4.61E-213 | 1 | 0 | 0 |

| | | | coloc SUSIE method | | | | | | | | | | |
|---|---|---|---|---|---|---|---|---|---|---|---|---|---|
| Trait1 | Trait 2 | cFDRlocusnum | nsnps 2 | hit1 | hit2 | PP.H0.abf | PP.H1.abf | PP.H2.abf | PP.H3.abf | PP.H4.abf | PP.H3+PP.H4≥0.8 | PP.H3/PP.H4≥3 | PP.H4/PP.H3≥5 |
| d | | | | | | | | | | | | | |
| AD Kunkle Corrected | eGFR | | | rs41290120 | rs814573 | 0.00E+00 | 1.64E-210 | 0.00E+00 | 1.00E+00 | 1.88E-212 | 1 | 0 | 0 |
| AD Kunkle Corrected | eGFR | | | rs75627662 | rs814573 | 0.00E+00 | 1.64E-210 | 0.00E+00 | 1.00E+00 | 2.17E-141 | 1 | 0 | 0 |
| AD Kunkle Corrected | eGFR | | | rs814573 | rs814573 | 0.00E+00 | 3.27E-213 | 0.00E+00 | 0.00E+00 | 1.00E+00 | 1 | 1 | 1 |
| AD Kunkle Corrected | eGFR | | | rs157595 | rs814573 | 0.00E+00 | 1.64E-210 | 0.00E+00 | 1.00E+00 | 3.44E-193 | 1 | 0 | 0 |
| AD Kunkle Corrected | eGFR | | | rs4803770 | rs814573 | 0.00E+00 | 1.64E-210 | 0.00E+00 | 1.00E+00 | 5.28E-186 | 1 | 0 | 0 |
| AD Kunkle Corrected | eGFR | | | rs60049679 | rs12721046 | 9.75E-156 | 1.42E-124 | 6.87E-32 | 1.00E+00 | 8.65E-31 | 1 | 0 | 0 |
| AD Kunkle Corrected | eGFR | | | rs12691088 | rs12721046 | 6.96E-180 | 1.42E-124 | 4.91E-56 | 1.00E+00 | 3.97E-55 | 1 | 0 | 0 |
| AD Kunkle Corrected | eGFR | | | rs144311893 | rs12721046 | 2.44E-312 | 1.42E-124 | 1.72E-188 | 1.00E+00 | 1.41E-126 | 1 | 0 | 0 |
| AD Kunkle Corrected | eGFR | | | rs77301115 | rs12721046 | 0.00E+00 | 1.42E-124 | 1.99E-215 | 1.00E+00 | 1.08E-126 | 1 | 0 | 0 |
| AD Kunkle Correcte | eGFR | | | rs1160983 | rs12721046 | 0.00E+00 | 1.42E-124 | 0.00E+00 | 1.00E+00 | 6.06E-122 | 1 | 0 | 0 |

| | | | coloc SUSIE method | | | | | | | | | | |
|---|---|---|---|---|---|---|---|---|---|---|---|---|---|
| **Trait1** | **Trait2** | **cFDRlocusnum** | **nsnps2** | **hit1** | **hit2** | **PP.H0.abf** | **PP.H1.abf** | **PP.H2.abf** | **PP.H3.abf** | **PP.H4.abf** | **PP.H3+PP.H4≥0.8** | **PP.H3/PP.H4≥3** | **PP.H4/PP.H3≥5** |
| d | | | | | | | | | | | | | |
| AD Kunkle Corrected | eGFR | | | rs41290120 | rs12721046 | 0.00E+00 | 1.42E-124 | 0.00E+00 | 1.00E+00 | 2.76E-124 | 1 | 0 | 0 |
| AD Kunkle Corrected | eGFR | | | rs75627662 | rs12721046 | 0.00E+00 | 1.42E-124 | 0.00E+00 | 1.00E+00 | 1.83E-90 | 1 | 0 | 0 |
| AD Kunkle Corrected | eGFR | | | rs814573 | rs12721046 | 0.00E+00 | 1.42E-124 | 0.00E+00 | 1.00E+00 | 3.96E-38 | 1 | 0 | 0 |
| AD Kunkle Corrected | eGFR | | | rs157595 | rs12721046 | 0.00E+00 | 1.42E-124 | 0.00E+00 | 1.00E+00 | 4.49E-110 | 1 | 0 | 0 |
| AD Kunkle Corrected | eGFR | | | rs4803770 | rs12721046 | 0.00E+00 | 1.42E-124 | 0.00E+00 | 1.00E+00 | 3.80E-117 | 1 | 0 | 0 |
| AD Kunkle Corrected | eGFR | | | rs60049679 | rs112019714 | 7.39E-55 | 1.07E-23 | 6.87E-32 | 1.00E+00 | 6.40E-22 | 1 | 0 | 0 |
| AD Kunkle Corrected | eGFR | | | rs12691088 | rs112019714 | 5.28E-79 | 1.07E-23 | 4.91E-56 | 1.00E+00 | 5.08E-26 | 1 | 0 | 0 |
| AD Kunkle Corrected | eGFR | | | rs144311893 | rs112019714 | 1.85E-211 | 1.07E-23 | 1.72E-188 | 1.00E+00 | 4.95E-26 | 1 | 0 | 0 |
| AD Kunkle Corrected | eGFR | | | rs77301115 | rs112019714 | 1.55E-238 | 7.80E-24 | 1.44E-215 | 7.25E-01 | 2.75E-01 | 1 | 0 | 0 |
| AD Kunkle Correcte | eGFR | | | rs1160983 | rs112019714 | 0.00E+00 | 1.07E-23 | 0.00E+00 | 1.00E+00 | 4.21E-23 | 1 | 0 | 0 |

| | | | coloc SUSIE method | | | | | | | | | | |
|---|---|---|---|---|---|---|---|---|---|---|---|---|---|
| **Trait1** | **Trait 2** | **cFDRlocusnum** | **nsnps 2** | **hit1** | **hit2** | **PP.H0.abf** | **PP.H1.abf** | **PP.H2.abf** | **PP.H3.abf** | **PP.H4.abf** | **PP.H3+PP.H4≥0.8** | **PP.H3/PP.H4≥3** | **PP.H4/PP.H3≥5** |
| d | | | | | | | | | | | | | |
| AD Kunkle Corrected | eGFR | | | rs41290120 | rs112019714 | 0.00E+00 | 1.07E-23 | 0.00E+00 | 1.00E+00 | 2.67E-25 | 1 | 0 | 0 |
| AD Kunkle Corrected | eGFR | | | rs75627662 | rs112019714 | 0.00E+00 | 1.07E-23 | 0.00E+00 | 1.00E+00 | 2.75E-24 | 1 | 0 | 0 |
| AD Kunkle Corrected | eGFR | | | rs814573 | rs112019714 | 0.00E+00 | 1.07E-23 | 0.00E+00 | 1.00E+00 | 3.70E-23 | 1 | 0 | 0 |
| AD Kunkle Corrected | eGFR | | | rs157595 | rs112019714 | 0.00E+00 | 1.07E-23 | 0.00E+00 | 1.00E+00 | 1.09E-24 | 1 | 0 | 0 |
| AD Kunkle Corrected | eGFR | | | rs4803770 | rs112019714 | 0.00E+00 | 1.07E-23 | 0.00E+00 | 1.00E+00 | 5.34E-26 | 1 | 0 | 0 |
| AD Kunkle Corrected | eGFR | 13b | 3039 | rs60049679 | rs429358 | 3.65E-60 | 5.32E-29 | 6.87E-32 | 1.00E+00 | 1.36E-26 | 1 | 0 | 0 |
| AD Kunkle Corrected | eGFR | | | rs12691088 | rs429358 | 2.61E-84 | 5.32E-29 | 4.91E-56 | 1.00E+00 | 7.07E-29 | 1 | 0 | 0 |
| AD Kunkle Corrected | eGFR | | | rs144311893 | rs429358 | 9.16E-217 | 5.32E-29 | 1.72E-188 | 1.00E+00 | 3.16E-31 | 1 | 0 | 0 |
| AD Kunkle Corrected | eGFR | | | rs77301115 | rs429358 | 1.06E-243 | 5.32E-29 | 1.99E-215 | 1.00E+00 | 8.61E-27 | 1 | 0 | 0 |
| AD Kunkle | eGFR | | | rs1160983 | rs429358 | 0.00E+00 | 5.32E-29 | 0.00E+00 | 1.00E+00 | 1.19E-27 | 1 | 0 | 0 |

| | | | coloc SUSIE method | | | | | | | | | | |
|---|---|---|---|---|---|---|---|---|---|---|---|---|---|
| **Trait1** | **Trait 2** | **cFDRlocusnum** | **nsnps 2** | **hit1** | **hit2** | **PP.H0.abf** | **PP.H1.abf** | **PP.H2.abf** | **PP.H3.abf** | **PP.H4.abf** | **PP.H3+PP.H4≥0.8** | **PP.H3/PP.H4≥3** | **PP.H4/PP.H3≥5** |
| Corrected | | | | | | | | | | | | | |
| AD Kunkle Corrected | eGFR | | | rs41290120 | rs429358 | 0.00E+00 | 5.32E-29 | 0.00E+00 | 1.00E+00 | 6.33E-30 | 1 | 0 | 0 |
| AD Kunkle Corrected | eGFR | | | rs75627662 | rs429358 | 0.00E+00 | 5.32E-29 | 0.00E+00 | 1.00E+00 | 3.88E-24 | 1 | 0 | 0 |
| AD Kunkle Corrected | eGFR | | | rs814573 | rs429358 | 0.00E+00 | 5.32E-29 | 0.00E+00 | 1.00E+00 | 3.45E-08 | 1 | 0 | 0 |
| AD Kunkle Corrected | eGFR | | | rs157595 | rs429358 | 0.00E+00 | 5.32E-29 | 0.00E+00 | 1.00E+00 | 3.79E-26 | 1 | 0 | 0 |
| AD Kunkle Corrected | eGFR | | | rs4803770 | rs429358 | 0.00E+00 | 5.32E-29 | 0.00E+00 | 1.00E+00 | 1.31E-29 | 1 | 0 | 0 |
| AD Kunkle Corrected | eGFR | | | rs60049679 | rs814573 | 9.71E-242 | 1.41E-210 | 6.87E-32 | 1.00E+00 | 1.07E-28 | 1 | 0 | 0 |
| AD Kunkle Corrected | eGFR | | | rs12691088 | rs814573 | 6.94E-266 | 1.41E-210 | 4.91E-56 | 1.00E+00 | 9.28E-55 | 1 | 0 | 0 |
| AD Kunkle Corrected | eGFR | | | rs144311893 | rs814573 | 0.00E+00 | 1.41E-210 | 1.72E-188 | 1.00E+00 | 4.92E-190 | 1 | 0 | 0 |
| AD Kunkle Corrected | eGFR | | | rs77301115 | rs814573 | 0.00E+00 | 1.41E-210 | 1.99E-215 | 1.00E+00 | 4.05E-191 | 1 | 0 | 0 |
| AD Kunkle | eGFR | | | rs1160983 | rs814573 | 0.00E+00 | 1.41E-210 | 0.00E+00 | 1.00E+00 | 3.96E-213 | 1 | 0 | 0 |

| | | | coloc SUSIE method | | | | | | | | | | |
|---|---|---|---|---|---|---|---|---|---|---|---|---|---|
| **Trait1** | **Trait 2** | **cFDRlocusnum** | **nsnps 2** | **hit1** | **hit2** | **PP.H0.abf** | **PP.H1.abf** | **PP.H2.abf** | **PP.H3.abf** | **PP.H4.abf** | **PP.H3+PP.H4≥0.8** | **PP.H3/PP.H4≥3** | **PP.H4/PP.H3≥5** |
| Corrected | | | | | | | | | | | | | |
| AD Kunkle Corrected | eGFR | | | rs41290120 | rs814573 | 0.00E+00 | 1.41E-210 | 0.00E+00 | 1.00E+00 | 1.63E-212 | 1 | 0 | 0 |
| AD Kunkle Corrected | eGFR | | | rs75627662 | rs814573 | 0.00E+00 | 1.41E-210 | 0.00E+00 | 1.00E+00 | 1.87E-141 | 1 | 0 | 0 |
| AD Kunkle Corrected | eGFR | | | rs814573 | rs814573 | 0.00E+00 | 2.83E-213 | 0.00E+00 | 0.00E+00 | 1.00E+00 | 1 | 1 | 1 |
| AD Kunkle Corrected | eGFR | | | rs157595 | rs814573 | 0.00E+00 | 1.41E-210 | 0.00E+00 | 1.00E+00 | 3.08E-193 | 1 | 0 | 0 |
| AD Kunkle Corrected | eGFR | | | rs4803770 | rs814573 | 0.00E+00 | 1.41E-210 | 0.00E+00 | 1.00E+00 | 4.47E-186 | 1 | 0 | 0 |
| AD Kunkle Corrected | eGFR | | | rs60049679 | rs12721046 | 9.69E-156 | 1.41E-124 | 6.87E-32 | 1.00E+00 | 8.65E-31 | 1 | 0 | 0 |
| AD Kunkle Corrected | eGFR | | | rs12691088 | rs12721046 | 6.93E-180 | 1.41E-124 | 4.91E-56 | 1.00E+00 | 3.97E-55 | 1 | 0 | 0 |
| AD Kunkle Corrected | eGFR | | | rs144311893 | rs12721046 | 2.43E-312 | 1.41E-124 | 1.72E-188 | 1.00E+00 | 1.40E-126 | 1 | 0 | 0 |
| AD Kunkle Corrected | eGFR | | | rs77301115 | rs12721046 | 0.00E+00 | 1.41E-124 | 1.99E-215 | 1.00E+00 | 1.07E-126 | 1 | 0 | 0 |
| AD Kunkle | eGFR | | | rs1160983 | rs12721046 | 0.00E+00 | 1.41E-124 | 0.00E+00 | 1.00E+00 | 5.90E-122 | 1 | 0 | 0 |

| | | | coloc SUSIE method | | | | | | | | | | |
|---|---|---|---|---|---|---|---|---|---|---|---|---|---|
| **Trait1** | **Trait 2** | **cFDRlocusnum** | **nsnps 2** | **hit1** | **hit2** | **PP.H0.abf** | **PP.H1.abf** | **PP.H2.abf** | **PP.H3.abf** | **PP.H4.abf** | **PP.H3+PP.H4≥0.8** | **PP.H3/PP.H4≥3** | **PP.H4/PP.H3≥5** |
| Corrected | | | | | | | | | | | | | |
| AD Kunkle Corrected | eGFR | | | rs41290120 | rs12721046 | 0.00E+00 | 1.41E-124 | 0.00E+00 | 1.00E+00 | 2.73E-124 | 1 | 0 | 0 |
| AD Kunkle Corrected | eGFR | | | rs75627662 | rs12721046 | 0.00E+00 | 1.41E-124 | 0.00E+00 | 1.00E+00 | 1.89E-90 | 1 | 0 | 0 |
| AD Kunkle Corrected | eGFR | | | rs814573 | rs12721046 | 0.00E+00 | 1.41E-124 | 0.00E+00 | 1.00E+00 | 3.99E-38 | 1 | 0 | 0 |
| AD Kunkle Corrected | eGFR | | | rs157595 | rs12721046 | 0.00E+00 | 1.41E-124 | 0.00E+00 | 1.00E+00 | 4.39E-110 | 1 | 0 | 0 |
| AD Kunkle Corrected | eGFR | | | rs4803770 | rs12721046 | 0.00E+00 | 1.41E-124 | 0.00E+00 | 1.00E+00 | 3.88E-117 | 1 | 0 | 0 |
| AD Kunkle Corrected | eGFR | | | rs60049679 | rs112019714 | 7.66E-55 | 1.11E-23 | 6.87E-32 | 1.00E+00 | 6.59E-22 | 1 | 0 | 0 |
| AD Kunkle Corrected | eGFR | | | rs12691088 | rs112019714 | 5.47E-79 | 1.11E-23 | 4.91E-56 | 1.00E+00 | 5.27E-26 | 1 | 0 | 0 |
| AD Kunkle Corrected | eGFR | | | rs144311893 | rs112019714 | 1.92E-211 | 1.11E-23 | 1.72E-188 | 1.00E+00 | 5.14E-26 | 1 | 0 | 0 |
| AD Kunkle Corrected | eGFR | | | rs77301115 | rs112019714 | 1.61E-238 | 8.09E-24 | 1.44E-215 | 7.25E-01 | 2.75E-01 | 1 | 0 | 0 |
| AD Kunkle | eGFR | | | rs1160983 | rs112019714 | 0.00E+00 | 1.11E-23 | 0.00E+00 | 1.00E+00 | 4.29E-23 | 1 | 0 | 0 |

| | | | coloc SUSIE method | | | | | | | | | | |
|---|---|---|---|---|---|---|---|---|---|---|---|---|---|
| **Trait1** | **Trait 2** | **cFDRlocusnum** | **nsnps 2** | **hit1** | **hit2** | **PP.H0.abf** | **PP.H1.abf** | **PP.H2.abf** | **PP.H3.abf** | **PP.H4.abf** | **PP.H3+PP.H4≥0.8** | **PP.H3/PP.H4≥3** | **PP.H4/PP.H3≥5** |
| Corrected | | | | | | | | | | | | | |
| AD Kunkle Corrected | eGFR | | | rs41290120 | rs112019714 | 0.00E+00 | 1.11E-23 | 0.00E+00 | 1.00E+00 | 2.76E-25 | 1 | 0 | 0 |
| AD Kunkle Corrected | eGFR | | | rs75627662 | rs112019714 | 0.00E+00 | 1.11E-23 | 0.00E+00 | 1.00E+00 | 2.83E-24 | 1 | 0 | 0 |
| AD Kunkle Corrected | eGFR | | | rs814573 | rs112019714 | 0.00E+00 | 1.11E-23 | 0.00E+00 | 1.00E+00 | 3.83E-23 | 1 | 0 | 0 |
| AD Kunkle Corrected | eGFR | | | rs157595 | rs112019714 | 0.00E+00 | 1.11E-23 | 0.00E+00 | 1.00E+00 | 1.12E-24 | 1 | 0 | 0 |
| AD Kunkle Corrected | eGFR | | | rs4803770 | rs112019714 | 0.00E+00 | 1.11E-23 | 0.00E+00 | 1.00E+00 | 5.55E-26 | 1 | 0 | 0 |
| AD Kunkle Corrected | eGFR | 13c | 3017 | rs60049679 | rs429358 | 3.53E-60 | 5.14E-29 | 6.87E-32 | 1.00E+00 | 1.31E-26 | 1 | 0 | 0 |
| AD Kunkle Corrected | eGFR | | | rs12691088 | rs429358 | 2.52E-84 | 5.14E-29 | 4.91E-56 | 1.00E+00 | 6.81E-29 | 1 | 0 | 0 |
| AD Kunkle Corrected | eGFR | | | rs144311893 | rs429358 | 8.85E-217 | 5.14E-29 | 1.72E-188 | 1.00E+00 | 3.05E-31 | 1 | 0 | 0 |
| AD Kunkle Corrected | eGFR | | | rs77301115 | rs429358 | 1.02E-243 | 5.14E-29 | 1.99E-215 | 1.00E+00 | 8.36E-27 | 1 | 0 | 0 |
| AD | eGFR | | | rs1160983 | rs429358 | 0.00E+00 | 5.14E-29 | 0.00E+00 | 1.00E+00 | 1.15E-27 | 1 | 0 | 0 |

| | | | coloc SUSIE method | | | | | | | | | | |
|---|---|---|---|---|---|---|---|---|---|---|---|---|---|
| **Trait1** | **Trait 2** | **cFDRlocusnum** | **nsnps 2** | **hit1** | **hit2** | **PP.H0.abf** | **PP.H1.abf** | **PP.H2.abf** | **PP.H3.abf** | **PP.H4.abf** | **PP.H3+PP.H4≥0.8** | **PP.H3/PP.H4≥3** | **PP.H4/PP.H3≥5** |
| Kunkle Corrected | | | | | | | | | | | | | |
| AD Kunkle Corrected | eGFR | | | rs41290120 | rs429358 | 0.00E+00 | 5.14E-29 | 0.00E+00 | 1.00E+00 | 6.13E-30 | 1 | 0 | 0 |
| AD Kunkle Corrected | eGFR | | | rs75627662 | rs429358 | 0.00E+00 | 5.14E-29 | 0.00E+00 | 1.00E+00 | 3.78E-24 | 1 | 0 | 0 |
| AD Kunkle Corrected | eGFR | | | rs814573 | rs429358 | 0.00E+00 | 5.14E-29 | 0.00E+00 | 1.00E+00 | 3.45E-08 | 1 | 0 | 0 |
| AD Kunkle Corrected | eGFR | | | rs157595 | rs429358 | 0.00E+00 | 5.14E-29 | 0.00E+00 | 1.00E+00 | 3.65E-26 | 1 | 0 | 0 |
| AD Kunkle Corrected | eGFR | | | rs4803770 | rs429358 | 0.00E+00 | 5.14E-29 | 0.00E+00 | 1.00E+00 | 1.28E-29 | 1 | 0 | 0 |
| AD Kunkle Corrected | eGFR | | | rs60049679 | rs814573 | 9.26E-242 | 1.35E-210 | 6.87E-32 | 1.00E+00 | 1.07E-28 | 1 | 0 | 0 |
| AD Kunkle Corrected | eGFR | | | rs12691088 | rs814573 | 6.61E-266 | 1.35E-210 | 4.91E-56 | 1.00E+00 | 9.28E-55 | 1 | 0 | 0 |
| AD Kunkle Corrected | eGFR | | | rs144311893 | rs814573 | 0.00E+00 | 1.35E-210 | 1.72E-188 | 1.00E+00 | 4.92E-190 | 1 | 0 | 0 |
| AD Kunkle Corrected | eGFR | | | rs77301115 | rs814573 | 0.00E+00 | 1.35E-210 | 1.99E-215 | 1.00E+00 | 3.90E-191 | 1 | 0 | 0 |
| AD | eGFR | | | rs1160983 | rs814573 | 0.00E+00 | 1.35E- | 0.00E+00 | 1.00E+00 | 3.77E- | 1 | 0 | 0 |

| | | | coloc SUSIE method | | | | | | | | | | |
|---|---|---|---|---|---|---|---|---|---|---|---|---|---|
| **Trait1** | **Trait 2** | **cFDRlocusnum** | **nsnps 2** | **hit1** | **hit2** | **PP.H0.abf** | **PP.H1.abf** | **PP.H2.abf** | **PP.H3.abf** | **PP.H4.abf** | **PP.H3+PP.H4≥0.8** | **PP.H3/PP.H4≥3** | **PP.H4/PP.H3≥5** |
| Kunkle Corrected | | | | | | | 210 | | | 213 | | | |
| AD Kunkle Corrected | eGFR | | | rs41290120 | rs814573 | 0.00E+00 | 1.35E-210 | 0.00E+00 | 1.00E+00 | 1.56E-212 | 1 | 0 | 0 |
| AD Kunkle Corrected | eGFR | | | rs75627662 | rs814573 | 0.00E+00 | 1.35E-210 | 0.00E+00 | 1.00E+00 | 1.83E-141 | 1 | 0 | 0 |
| AD Kunkle Corrected | eGFR | | | rs814573 | rs814573 | 0.00E+00 | 2.69E-213 | 0.00E+00 | 0.00E+00 | 1.00E+00 | 1 | 1 | 1 |
| AD Kunkle Corrected | eGFR | | | rs157595 | rs814573 | 0.00E+00 | 1.35E-210 | 0.00E+00 | 1.00E+00 | 3.00E-193 | 1 | 0 | 0 |
| AD Kunkle Corrected | eGFR | | | rs4803770 | rs814573 | 0.00E+00 | 1.35E-210 | 0.00E+00 | 1.00E+00 | 4.23E-186 | 1 | 0 | 0 |
| AD Kunkle Corrected | eGFR | | | rs60049679 | rs12721046 | 9.99E-156 | 1.45E-124 | 6.87E-32 | 1.00E+00 | 8.65E-31 | 1 | 0 | 0 |
| AD Kunkle Corrected | eGFR | | | rs12691088 | rs12721046 | 7.13E-180 | 1.45E-124 | 4.91E-56 | 1.00E+00 | 3.97E-55 | 1 | 0 | 0 |
| AD Kunkle Corrected | eGFR | | | rs144311893 | rs12721046 | 2.50E-312 | 1.45E-124 | 1.72E-188 | 1.00E+00 | 1.45E-126 | 1 | 0 | 0 |
| AD Kunkle Corrected | eGFR | | | rs77301115 | rs12721046 | 0.00E+00 | 1.45E-124 | 1.99E-215 | 1.00E+00 | 1.11E-126 | 1 | 0 | 0 |
| AD | eGFR | | | rs1160983 | rs12721046 | 0.00E+00 | 1.45E- | 0.00E+00 | 1.00E+00 | 6.06E- | 1 | 0 | 0 |

| | | | coloc SUSIE method | | | | | | | | | | |
|---|---|---|---|---|---|---|---|---|---|---|---|---|---|
| **Trait1** | **Trait 2** | **cFDRlocusnum** | **nsnps 2** | **hit1** | **hit2** | **PP.H0.abf** | **PP.H1.abf** | **PP.H2.abf** | **PP.H3.abf** | **PP.H4.abf** | **PP.H3+PP.H4≥0.8** | **PP.H3/PP.H4≥3** | **PP.H4/PP.H3≥5** |
| Kunkle Corrected | | | | | | | 124 | | | 122 | | | |
| AD Kunkle Corrected | eGFR | | | rs41290120 | rs12721046 | 0.00E+00 | 1.45E-124 | 0.00E+00 | 1.00E+00 | 2.81E-124 | 1 | 0 | 0 |
| AD Kunkle Corrected | eGFR | | | rs75627662 | rs12721046 | 0.00E+00 | 1.45E-124 | 0.00E+00 | 1.00E+00 | 1.92E-90 | 1 | 0 | 0 |
| AD Kunkle Corrected | eGFR | | | rs814573 | rs12721046 | 0.00E+00 | 1.45E-124 | 0.00E+00 | 1.00E+00 | 4.08E-38 | 1 | 0 | 0 |
| AD Kunkle Corrected | eGFR | | | rs157595 | rs12721046 | 0.00E+00 | 1.45E-124 | 0.00E+00 | 1.00E+00 | 4.45E-110 | 1 | 0 | 0 |
| AD Kunkle Corrected | eGFR | | | rs4803770 | rs12721046 | 0.00E+00 | 1.45E-124 | 0.00E+00 | 1.00E+00 | 4.02E-117 | 1 | 0 | 0 |
| AD Kunkle Corrected | eGFR | | | rs60049679 | rs112019714 | 7.80E-55 | 1.14E-23 | 6.87E-32 | 1.00E+00 | 6.69E-22 | 1 | 0 | 0 |
| AD Kunkle Corrected | eGFR | | | rs12691088 | rs112019714 | 5.57E-79 | 1.14E-23 | 4.91E-56 | 1.00E+00 | 5.37E-26 | 1 | 0 | 0 |
| AD Kunkle Corrected | eGFR | | | rs144311893 | rs112019714 | 1.96E-211 | 1.14E-23 | 1.72E-188 | 1.00E+00 | 5.23E-26 | 1 | 0 | 0 |
| AD Kunkle Corrected | eGFR | | | rs77301115 | rs112019714 | 1.64E-238 | 8.23E-24 | 1.44E-215 | 7.25E-01 | 2.75E-01 | 1 | 0 | 0 |
| AD | eGFR | | | rs1160983 | rs11201971 | 0.00E+00 | 1.14E-23 | 0.00E+00 | 1.00E+00 | 4.36E-23 | 1 | 0 | 0 |

| | | | coloc SUSIE method | | | | | | | | | | |
|---|---|---|---|---|---|---|---|---|---|---|---|---|---|
| **Trait1** | **Trait 2** | **cFDRlocusnum** | **nsnps 2** | **hit1** | **hit2** | **PP.H0.abf** | **PP.H1.abf** | **PP.H2.abf** | **PP.H3.abf** | **PP.H4.abf** | **PP.H3+PP.H4≥0.8** | **PP.H3/PP.H4≥3** | **PP.H4/PP.H3≥5** |
| Kunkle Corrected | | | | | 4 | | | | | | | | |
| AD Kunkle Corrected | eGFR | | | rs41290120 | rs112019714 | 0.00E+00 | 1.14E-23 | 0.00E+00 | 1.00E+00 | 2.82E-25 | 1 | 0 | 0 |
| AD Kunkle Corrected | eGFR | | | rs75627662 | rs112019714 | 0.00E+00 | 1.14E-23 | 0.00E+00 | 1.00E+00 | 2.88E-24 | 1 | 0 | 0 |
| AD Kunkle Corrected | eGFR | | | rs814573 | rs112019714 | 0.00E+00 | 1.14E-23 | 0.00E+00 | 1.00E+00 | 3.90E-23 | 1 | 0 | 0 |
| AD Kunkle Corrected | eGFR | | | rs157595 | rs112019714 | 0.00E+00 | 1.14E-23 | 0.00E+00 | 1.00E+00 | 1.14E-24 | 1 | 0 | 0 |
| AD Kunkle Corrected | eGFR | | | rs4803770 | rs112019714 | 0.00E+00 | 1.14E-23 | 0.00E+00 | 1.00E+00 | 5.65E-26 | 1 | 0 | 0 |

**Supplementary Table S25. Colocalization Analysis Results Using SuSiE Framework (Kunkle Uncorrected).**
This table presents colocalization results using AD GWAS data not corrected for competing risk (Kunkle uncorrected). The analysis structure is identical to Table S24, testing the same five hypotheses about causal variant architecture at each pleiotropic locus. Note: 0 indicates "No", and 1 indicates "Yes".

| | | | coloc SUSIE method | | | | | | | | | | |
|---|---|---|---|---|---|---|---|---|---|---|---|---|---|
| **Trait1** | **Trait 2** | **cFDRlocusnum** | **nsnps 2** | **hit1** | **hit2** | **PP.H0.abf** | **PP.H1.abf** | **PP.H2.abf** | **PP.H3.abf** | **PP.H4.abf** | **PP.H3+PP.H4≥0.8** | **PP.H3/PP.H4≥3** | **PP.H4/PP.H3≥5** |
| AD Kunkle | eGFR | 4 | 1673 | rs67472071 | rs10838702 | 4.93E-18 | 2.38E-13 | 2.07E-05 | 9.98E-01 | 2.32E-03 | 1 | 0 | 0 |

| | | | coloc SUSIE method | | | | | | | | | | |
|---|---|---|---|---|---|---|---|---|---|---|---|---|---|
| **Trait1** | **Trait 2** | **cFDRlocusnum** | **nsnps 2** | **hit1** | **hit2** | **PP.H0.abf** | **PP.H1.abf** | **PP.H2.abf** | **PP.H3.abf** | **PP.H4.abf** | **PP.H3+PP.H4≥0.8** | **PP.H3/PP.H4≥3** | **PP.H4/PP.H3≥5** |
| AD Kunkle | eGFR | 11 | 3276 | rs429358 | rs429358 | 0.00E+00 | 2.58E-29 | 0.00E+00 | 2.83E-09 | 1.00E+00 | 1 | 1 | 1 |
| AD Kunkle | eGFR | | | rs12691088 | rs429358 | 1.66E-94 | 1.29E-26 | 1.29E-68 | 1.00E+00 | 1.31E-26 | 1 | 0 | 0 |
| AD Kunkle | eGFR | | | rs814573 | rs429358 | 0.00E+00 | 1.29E-26 | 0.00E+00 | 1.00E+00 | 2.36E-07 | 1 | 0 | 0 |
| AD Kunkle | eGFR | | | rs60049679 | rs429358 | 4.38E-100 | 1.29E-26 | 3.40E-74 | 1.00E+00 | 4.62E-25 | 1 | 0 | 0 |
| AD Kunkle | eGFR | | | rs1160983 | rs429358 | 0.00E+00 | 1.29E-26 | 0.00E+00 | 1.00E+00 | 5.14E-26 | 1 | 0 | 0 |
| AD Kunkle | eGFR | | | rs41290120 | rs429358 | 0.00E+00 | 1.29E-26 | 0.00E+00 | 1.00E+00 | 1.37E-27 | 1 | 0 | 0 |
| AD Kunkle | eGFR | | | rs769449 | rs429358 | 0.00E+00 | 1.29E-26 | 0.00E+00 | 1.00E+00 | 3.25E-10 | 1 | 0 | 0 |
| AD Kunkle | eGFR | | | rs75627662 | rs429358 | 0.00E+00 | 1.29E-26 | 0.00E+00 | 1.00E+00 | 1.57E-21 | 1 | 0 | 0 |
| AD Kunkle | eGFR | | | rs10775543 | rs429358 | 0.00E+00 | 1.29E-26 | 0.00E+00 | 1.00E+00 | 1.18E-28 | 1 | 0 | 0 |
| AD Kunkle | eGFR | | | rs204907 | rs429358 | 0.00E+00 | 1.29E-26 | 0.00E+00 | 1.00E+00 | 7.29E-29 | 1 | 0 | 0 |
| AD Kunkle | eGFR | | | rs429358 | rs814573 | 0.00E+00 | 2.94E-253 | 0.00E+00 | 1.00E+00 | 2.10E-73 | 1 | 0 | 0 |
| AD Kunkle | eGFR | | | rs12691088 | rs814573 | 3.79E-321 | 2.94E-253 | 1.29E-68 | 1.00E+00 | 1.47E-66 | 1 | 0 | 0 |
| AD Kunkle | eGFR | | | rs814573 | rs814573 | 0.00E+00 | 5.87E-256 | 0.00E+00 | 0.00E+00 | 1.00E+00 | 1 | 1 | 1 |

| | | | coloc SUSIE method | | | | | | | | | | |
|---|---|---|---|---|---|---|---|---|---|---|---|---|---|
| Trait1 | Trait 2 | cFDRlocusnum | nsnps 2 | hit1 | hit2 | PP.H0.abf | PP.H1.abf | PP.H2.abf | PP.H3.abf | PP.H4.abf | PP.H3+PP.H4≥0.8 | PP.H3/PP.H4≥3 | PP.H4/PP.H3≥5 |
| AD Kunkle | eGFR | | | rs60049679 | rs814573 | 0.00E+00 | 2.94E-253 | 3.40E-74 | 1.00E+00 | 1.88E-64 | 1 | 0 | 0 |
| AD Kunkle | eGFR | | | rs1160983 | rs814573 | 0.00E+00 | 2.94E-253 | 0.00E+00 | 1.00E+00 | 4.62E-256 | 1 | 0 | 0 |
| AD Kunkle | eGFR | | | rs41290120 | rs814573 | 0.00E+00 | 2.94E-253 | 0.00E+00 | 1.00E+00 | 6.38E-255 | 1 | 0 | 0 |
| AD Kunkle | eGFR | | | rs769449 | rs814573 | 0.00E+00 | 2.94E-253 | 0.00E+00 | 1.00E+00 | 1.80E-99 | 1 | 0 | 0 |
| AD Kunkle | eGFR | | | rs75627662 | rs814573 | 0.00E+00 | 2.94E-253 | 0.00E+00 | 1.00E+00 | 1.59E-174 | 1 | 0 | 0 |
| AD Kunkle | eGFR | | | rs10775543 | rs814573 | 0.00E+00 | 2.94E-253 | 0.00E+00 | 1.00E+00 | 1.10E-255 | 1 | 0 | 0 |
| AD Kunkle | eGFR | | | rs204907 | rs814573 | 0.00E+00 | 2.94E-253 | 0.00E+00 | 1.00E+00 | 2.51E-255 | 1 | 0 | 0 |
| AD Kunkle | eGFR | | | rs429358 | rs12721046 | 0.00E+00 | 9.10E-156 | 0.00E+00 | 1.00E+00 | 7.29E-67 | 1 | 0 | 0 |
| AD Kunkle | eGFR | | | rs12691088 | rs12721046 | 1.18E-223 | 9.10E-156 | 1.29E-68 | 1.00E+00 | 2.70E-49 | 1 | 0 | 0 |
| AD Kunkle | eGFR | | | rs814573 | rs12721046 | 0.00E+00 | 9.10E-156 | 0.00E+00 | 1.00E+00 | 1.52E-47 | 1 | 0 | 0 |
| AD Kunkle | eGFR | | | rs60049679 | rs12721046 | 3.10E-229 | 9.10E-156 | 3.40E-74 | 1.00E+00 | 4.54E-46 | 1 | 0 | 0 |
| AD Kunkle | eGFR | | | rs1160983 | rs12721046 | 0.00E+00 | 9.10E-156 | 0.00E+00 | 1.00E+00 | 9.66E-154 | 1 | 0 | 0 |
| AD Kunkle | eGFR | | | rs41290120 | rs12721046 | 0.00E+00 | 9.10E-156 | 0.00E+00 | 1.00E+00 | 4.19E-155 | 1 | 0 | 0 |
| AD | eGFR | | | rs769449 | rs12721046 | 0.00E+00 | 9.10E-156 | 0.00E+00 | 1.00E+00 | 5.98E-53 | 1 | 0 | 0 |

| | | | coloc SUSIE method | | | | | | | | | | |
|---|---|---|---|---|---|---|---|---|---|---|---|---|---|
| **Trait1** | **Trait 2** | **cFDRlocusnum** | **nsnps 2** | **hit1** | **hit2** | **PP.H0.abf** | **PP.H1.abf** | **PP.H2.abf** | **PP.H3.abf** | **PP.H4.abf** | **PP.H3+PP.H4≥0.8** | **PP.H3/PP.H4≥3** | **PP.H4/PP.H3≥5** |
| Kunkle | | | | | | | | | | | | | |
| AD Kunkle | eGFR | | | rs75627662 | rs12721046 | 0.00E+00 | 9.10E-156 | 0.00E+00 | 1.00E+00 | 5.92E-109 | 1 | 0 | 0 |
| AD Kunkle | eGFR | | | rs10775543 | rs12721046 | 0.00E+00 | 9.10E-156 | 0.00E+00 | 1.00E+00 | 1.76E-157 | 1 | 0 | 0 |
| AD Kunkle | eGFR | | | rs204907 | rs12721046 | 0.00E+00 | 9.10E-156 | 0.00E+00 | 1.00E+00 | 6.76E-158 | 1 | 0 | 0 |
| AD Kunkle | eGFR | | | rs429358 | rs150966173 | 0.00E+00 | 3.50E-34 | 0.00E+00 | 1.00E+00 | 1.89E-30 | 1 | 0 | 0 |
| AD Kunkle | eGFR | | | rs12691088 | rs150966173 | 4.53E-102 | 3.50E-34 | 1.29E-68 | 1.00E+00 | 1.41E-36 | 1 | 0 | 0 |
| AD Kunkle | eGFR | | | rs814573 | rs150966173 | 0.00E+00 | 3.50E-34 | 0.00E+00 | 1.00E+00 | 4.71E-32 | 1 | 0 | 0 |
| AD Kunkle | eGFR | | | rs60049679 | rs150966173 | 1.19E-107 | 3.50E-34 | 3.40E-74 | 1.00E+00 | 7.65E-32 | 1 | 0 | 0 |
| AD Kunkle | eGFR | | | rs1160983 | rs150966173 | 0.00E+00 | 3.50E-34 | 0.00E+00 | 1.00E+00 | 3.07E-34 | 1 | 0 | 0 |
| AD Kunkle | eGFR | | | rs41290120 | rs150966173 | 0.00E+00 | 3.50E-34 | 0.00E+00 | 1.00E+00 | 8.94E-36 | 1 | 0 | 0 |
| AD Kunkle | eGFR | | | rs769449 | rs150966173 | 0.00E+00 | 3.50E-34 | 0.00E+00 | 1.00E+00 | 7.90E-36 | 1 | 0 | 0 |
| AD Kunkle | eGFR | | | rs75627662 | rs150966173 | 0.00E+00 | 3.50E-34 | 0.00E+00 | 1.00E+00 | 4.87E-35 | 1 | 0 | 0 |
| AD Kunkle | eGFR | | | rs10775543 | rs150966173 | 0.00E+00 | 3.50E-34 | 0.00E+00 | 1.00E+00 | 1.93E-36 | 1 | 0 | 0 |
| AD Kunkl | eGFR | | | rs204907 | rs150966173 | 0.00E+00 | 3.50E-34 | 0.00E+00 | 1.00E+00 | 1.47E-36 | 1 | 0 | 0 |

| | | | coloc SUSIE method | | | | | | | | | | |
|---|---|---|---|---|---|---|---|---|---|---|---|---|---|
| **Trait1** | **Trait 2** | **cFDRlocusnum** | **nsnps 2** | **hit1** | **hit2** | **PP.H0.abf** | **PP.H1.abf** | **PP.H2.abf** | **PP.H3.abf** | **PP.H4.abf** | **PP.H3+PP.H4≥0.8** | **PP.H3/PP.H4≥3** | **PP.H4/PP.H3≥5** |
| e | | | | | | | | | | | | | |
| AD Kunkle | eGFR | 12 | 3317 | rs429358 | rs429358 | 0.00E+00 | 2.44E-29 | 0.00E+00 | 2.81E-09 | 1.00E+00 | 1 | 1 | 1 |
| AD Kunkle | eGFR | | | rs12691088 | rs429358 | 1.58E-94 | 1.22E-26 | 1.29E-68 | 1.00E+00 | 1.26E-26 | 1 | 0 | 0 |
| AD Kunkle | eGFR | | | rs814573 | rs429358 | 0.00E+00 | 1.22E-26 | 0.00E+00 | 1.00E+00 | 2.29E-07 | 1 | 0 | 0 |
| AD Kunkle | eGFR | | | rs60049679 | rs429358 | 4.15E-100 | 1.22E-26 | 3.40E-74 | 1.00E+00 | 4.37E-25 | 1 | 0 | 0 |
| AD Kunkle | eGFR | | | rs1160983 | rs429358 | 0.00E+00 | 1.22E-26 | 0.00E+00 | 1.00E+00 | 8.43E-26 | 1 | 0 | 0 |
| AD Kunkle | eGFR | | | rs41290120 | rs429358 | 0.00E+00 | 1.22E-26 | 0.00E+00 | 1.00E+00 | 1.24E-27 | 1 | 0 | 0 |
| AD Kunkle | eGFR | | | rs769449 | rs429358 | 0.00E+00 | 1.22E-26 | 0.00E+00 | 1.00E+00 | 3.21E-10 | 1 | 0 | 0 |
| AD Kunkle | eGFR | | | rs75627662 | rs429358 | 0.00E+00 | 1.22E-26 | 0.00E+00 | 1.00E+00 | 1.12E-21 | 1 | 0 | 0 |
| AD Kunkle | eGFR | | | rs10775543 | rs429358 | 0.00E+00 | 1.22E-26 | 0.00E+00 | 1.00E+00 | 1.11E-28 | 1 | 0 | 0 |
| AD Kunkle | eGFR | | | rs204907 | rs429358 | 0.00E+00 | 1.22E-26 | 0.00E+00 | 1.00E+00 | 6.87E-29 | 1 | 0 | 0 |
| AD Kunkle | eGFR | | | rs429358 | rs814573 | 0.00E+00 | 3.02E-253 | 0.00E+00 | 1.00E+00 | 2.01E-73 | 1 | 0 | 0 |
| AD Kunkle | eGFR | | | rs12691088 | rs814573 | 3.90E-321 | 3.02E-253 | 1.29E-68 | 1.00E+00 | 1.47E-66 | 1 | 0 | 0 |
| AD Kunkl | eGFR | | | rs814573 | rs814573 | 0.00E+00 | 6.04E-256 | 0.00E+00 | 0.00E+00 | 1.00E+00 | 1 | | 1 |

| | | | coloc SUSIE method | | | | | | | | | | |
|---|---|---|---|---|---|---|---|---|---|---|---|---|---|
| Trait1 | Trait2 | cFDRlocusnum | nsnps2 | hit1 | hit2 | PP.H0.abf | PP.H1.abf | PP.H2.abf | PP.H3.abf | PP.H4.abf | PP.H3+PP.H4≥0.8 | PP.H3/PP.H4≥3 | PP.H4/PP.H3≥5 |
| e | | | | | | | | | | | | | |
| AD Kunkle | eGFR | | | rs60049679 | rs814573 | 0.00E+00 | 3.02E-253 | 3.40E-74 | 1.00E+00 | 1.88E-64 | 1 | 0 | 0 |
| AD Kunkle | eGFR | | | rs1160983 | rs814573 | 0.00E+00 | 3.02E-253 | 0.00E+00 | 1.00E+00 | 5.18E-256 | 1 | 0 | 0 |
| AD Kunkle | eGFR | | | rs41290120 | rs814573 | 0.00E+00 | 3.02E-253 | 0.00E+00 | 1.00E+00 | 6.84E-255 | 1 | 0 | 0 |
| AD Kunkle | eGFR | | | rs769449 | rs814573 | 0.00E+00 | 3.02E-253 | 0.00E+00 | 1.00E+00 | 1.74E-99 | 1 | 0 | 0 |
| AD Kunkle | eGFR | | | rs75627662 | rs814573 | 0.00E+00 | 3.02E-253 | 0.00E+00 | 1.00E+00 | 5.77E-174 | 1 | 0 | 0 |
| AD Kunkle | eGFR | | | rs10775543 | rs814573 | 0.00E+00 | 3.02E-253 | 0.00E+00 | 1.00E+00 | 1.14E-255 | 1 | 0 | 0 |
| AD Kunkle | eGFR | | | rs204907 | rs814573 | 0.00E+00 | 3.02E-253 | 0.00E+00 | 1.00E+00 | 2.62E-255 | 1 | 0 | 0 |
| AD Kunkle | eGFR | | | rs429358 | rs12721046 | 0.00E+00 | 6.64E-156 | 0.00E+00 | 1.00E+00 | 6.51E-67 | 1 | 0 | 0 |
| AD Kunkle | eGFR | | | rs12691088 | rs12721046 | 8.58E-224 | 6.64E-156 | 1.29E-68 | 1.00E+00 | 2.70E-49 | 1 | 0 | 0 |
| AD Kunkle | eGFR | | | rs814573 | rs12721046 | 0.00E+00 | 6.64E-156 | 0.00E+00 | 1.00E+00 | 1.35E-47 | 1 | 0 | 0 |
| AD Kunkle | eGFR | | | rs60049679 | rs12721046 | 2.26E-229 | 6.64E-156 | 3.40E-74 | 1.00E+00 | 4.53E-46 | 1 | 0 | 0 |
| AD Kunkle | eGFR | | | rs1160983 | rs12721046 | 0.00E+00 | 6.64E-156 | 0.00E+00 | 1.00E+00 | 1.65E-153 | 1 | 0 | 0 |
| AD Kunkle | eGFR | | | rs41290120 | rs12721046 | 0.00E+00 | 6.64E-156 | 0.00E+00 | 1.00E+00 | 2.85E-155 | 1 | 0 | 0 |

| | | | coloc SUSIE method | | | | | | | | | | |
|---|---|---|---|---|---|---|---|---|---|---|---|---|---|
| **Trait1** | **Trait 2** | **cFDRlocusnum** | **nsnps 2** | **hit1** | **hit2** | **PP.H0.abf** | **PP.H1.abf** | **PP.H2.abf** | **PP.H3.abf** | **PP.H4.abf** | **PP.H3+PP.H4≥0.8** | **PP.H3/PP.H4≥3** | **PP.H4/PP.H3≥5** |
| AD Kunkle | eGFR | | | rs769449 | rs12721046 | 0.00E+00 | 6.64E-156 | 0.00E+00 | 1.00E+00 | 5.54E-53 | 1 | 0 | 0 |
| AD Kunkle | eGFR | | | rs75627662 | rs12721046 | 0.00E+00 | 6.64E-156 | 0.00E+00 | 1.00E+00 | 1.78E-109 | 1 | 0 | 0 |
| AD Kunkle | eGFR | | | rs10775543 | rs12721046 | 0.00E+00 | 6.64E-156 | 0.00E+00 | 1.00E+00 | 1.26E-157 | 1 | 0 | 0 |
| AD Kunkle | eGFR | | | rs204907 | rs12721046 | 0.00E+00 | 6.64E-156 | 0.00E+00 | 1.00E+00 | 4.87E-158 | 1 | 0 | 0 |
| AD Kunkle | eGFR | | | rs429358 | rs150966173 | 0.00E+00 | 3.03E-34 | 0.00E+00 | 1.00E+00 | 1.68E-30 | 1 | 0 | 0 |
| AD Kunkle | eGFR | | | rs12691088 | rs150966173 | 3.92E-102 | 3.03E-34 | 1.29E-68 | 1.00E+00 | 1.22E-36 | 1 | 0 | 0 |
| AD Kunkle | eGFR | | | rs814573 | rs150966173 | 0.00E+00 | 3.03E-34 | 0.00E+00 | 1.00E+00 | 4.13E-32 | 1 | 0 | 0 |
| AD Kunkle | eGFR | | | rs60049679 | rs150966173 | 1.03E-107 | 3.03E-34 | 3.40E-74 | 1.00E+00 | 6.67E-32 | 1 | 0 | 0 |
| AD Kunkle | eGFR | | | rs1160983 | rs150966173 | 0.00E+00 | 3.03E-34 | 0.00E+00 | 1.00E+00 | 4.86E-34 | 1 | 0 | 0 |
| AD Kunkle | eGFR | | | rs41290120 | rs150966173 | 0.00E+00 | 3.03E-34 | 0.00E+00 | 1.00E+00 | 7.47E-36 | 1 | 0 | 0 |
| AD Kunkle | eGFR | | | rs769449 | rs150966173 | 0.00E+00 | 3.03E-34 | 0.00E+00 | 1.00E+00 | 6.83E-36 | 1 | 0 | 0 |
| AD Kunkle | eGFR | | | rs75627662 | rs150966173 | 0.00E+00 | 3.03E-34 | 0.00E+00 | 1.00E+00 | 5.06E-35 | 1 | 0 | 0 |
| AD Kunkle | eGFR | | | rs10775543 | rs150966173 | 0.00E+00 | 3.03E-34 | 0.00E+00 | 1.00E+00 | 1.66E-36 | 1 | 0 | 0 |
| AD | eGFR | | | rs204907 | rs15096617 | 0.00E+00 | 3.03E-34 | 0.00E+00 | 1.00E+00 | 1.27E-36 | 1 | 0 | 0 |

| | | | coloc SUSIE method | | | | | | | | | | |
|---|---|---|---|---|---|---|---|---|---|---|---|---|---|
| **Trait1** | **Trait 2** | **cFDRlocusnu m** | **nsnps 2** | **hit1** | **hit2** | **PP.H0.ab f** | **PP.H1.ab f** | **PP.H2.ab f** | **PP.H3.ab f** | **PP.H4.ab f** | **PP.H3+PP.H4≥0. 8** | **PP.H3/PP.H4≥ 3** | **PP.H4/PP.H3≥ 5** |
| Kunkl e | | | | | 3 | | | | | | | | |
| AD Kunkl e | eGFR | 13 | 3333 | rs429358 | rs429358 | 0.00E+00 | 2.08E-29 | 0.00E+00 | 2.68E-09 | 1.00E+00 | 1 | 1 | 1 |
| AD Kunkl e | eGFR | | | rs1269108 8 | rs429358 | 1.34E-94 | 1.04E-26 | 1.29E-68 | 1.00E+00 | 1.05E-26 | 1 | 0 | 0 |
| AD Kunkl e | eGFR | | | rs814573 | rs429358 | 0.00E+00 | 1.04E-26 | 0.00E+00 | 1.00E+00 | 2.11E-07 | 1 | 0 | 0 |
| AD Kunkl e | eGFR | | | rs6004967 9 | rs429358 | 3.54E-100 | 1.04E-26 | 3.40E-74 | 1.00E+00 | 3.78E-25 | 1 | 0 | 0 |
| AD Kunkl e | eGFR | | | rs1160983 | rs429358 | 0.00E+00 | 1.04E-26 | 0.00E+00 | 1.00E+00 | 1.89E-25 | 1 | 0 | 0 |
| AD Kunkl e | eGFR | | | rs4129012 0 | rs429358 | 0.00E+00 | 1.04E-26 | 0.00E+00 | 1.00E+00 | 9.54E-28 | 1 | 0 | 0 |
| AD Kunkl e | eGFR | | | rs769449 | rs429358 | 0.00E+00 | 1.04E-26 | 0.00E+00 | 1.00E+00 | 3.03E-10 | 1 | 0 | 0 |
| AD Kunkl e | eGFR | | | rs7562766 2 | rs429358 | 0.00E+00 | 1.04E-26 | 0.00E+00 | 1.00E+00 | 6.19E-22 | 1 | 0 | 0 |
| AD Kunkl e | eGFR | | | rs1077554 3 | rs429358 | 0.00E+00 | 1.04E-26 | 0.00E+00 | 1.00E+00 | 9.29E-29 | 1 | 0 | 0 |
| AD Kunkl e | eGFR | | | rs204907 | rs429358 | 0.00E+00 | 1.04E-26 | 0.00E+00 | 1.00E+00 | 5.79E-29 | 1 | 0 | 0 |
| AD Kunkl e | eGFR | | | rs429358 | rs814573 | 0.00E+00 | 9.99E-254 | 0.00E+00 | 1.00E+00 | 1.38E-73 | 1 | 0 | 0 |
| AD Kunkl e | eGFR | | | rs1269108 8 | rs814573 | 1.29E-321 | 9.99E-254 | 1.29E-68 | 1.00E+00 | 1.47E-66 | 1 | 0 | 0 |
| AD | eGFR | | | rs814573 | rs814573 | 0.00E+00 | 2.00E-256 | 0.00E+00 | 0.00E+00 | 1.00E+00 | 1 | 1 | 1 |

| | | | coloc SUSIE method | | | | | | | | | | |
|---|---|---|---|---|---|---|---|---|---|---|---|---|---|
| Trait1 | Trait 2 | cFDRlocusnum | nsnps 2 | hit1 | hit2 | PP.H0.abf | PP.H1.abf | PP.H2.abf | PP.H3.abf | PP.H4.abf | PP.H3+PP.H4≥0.8 | PP.H3/PP.H4≥3 | PP.H4/PP.H3≥5 |
| Kunkle | | | | | | | | | | | | | |
| AD Kunkle | eGFR | | | rs60049679 | rs814573 | 0.00E+00 | 9.99E-254 | 3.40E-74 | 1.00E+00 | 1.88E-64 | 1 | 0 | 0 |
| AD Kunkle | eGFR | | | rs1160983 | rs814573 | 0.00E+00 | 9.99E-254 | 0.00E+00 | 1.00E+00 | 2.04E-256 | 1 | 0 | 0 |
| AD Kunkle | eGFR | | | rs41290120 | rs814573 | 0.00E+00 | 9.99E-254 | 0.00E+00 | 1.00E+00 | 2.49E-255 | 1 | 0 | 0 |
| AD Kunkle | eGFR | | | rs769449 | rs814573 | 0.00E+00 | 9.99E-254 | 0.00E+00 | 1.00E+00 | 9.86E-100 | 1 | 0 | 0 |
| AD Kunkle | eGFR | | | rs75627662 | rs814573 | 0.00E+00 | 9.99E-254 | 0.00E+00 | 1.00E+00 | 1.35E-173 | 1 | 0 | 0 |
| AD Kunkle | eGFR | | | rs10775543 | rs814573 | 0.00E+00 | 9.99E-254 | 0.00E+00 | 1.00E+00 | 3.86E-256 | 1 | 0 | 0 |
| AD Kunkle | eGFR | | | rs204907 | rs814573 | 0.00E+00 | 9.99E-254 | 0.00E+00 | 1.00E+00 | 8.96E-256 | 1 | 0 | 0 |
| AD Kunkle | eGFR | | | rs429358 | rs12721046 | 0.00E+00 | 2.39E-156 | 0.00E+00 | 1.00E+00 | 4.33E-67 | 1 | 0 | 0 |
| AD Kunkle | eGFR | | | rs12691088 | rs12721046 | 3.09E-224 | 2.39E-156 | 1.29E-68 | 1.00E+00 | 2.70E-49 | 1 | 0 | 0 |
| AD Kunkle | eGFR | | | rs814573 | rs12721046 | 0.00E+00 | 2.39E-156 | 0.00E+00 | 1.00E+00 | 9.80E-48 | 1 | 0 | 0 |
| AD Kunkle | eGFR | | | rs60049679 | rs12721046 | 8.13E-230 | 2.39E-156 | 3.40E-74 | 1.00E+00 | 4.50E-46 | 1 | 0 | 0 |
| AD Kunkle | eGFR | | | rs1160983 | rs12721046 | 0.00E+00 | 2.39E-156 | 0.00E+00 | 1.00E+00 | 2.08E-153 | 1 | 0 | 0 |
| AD Kunkl | eGFR | | | rs41290120 | rs12721046 | 0.00E+00 | 2.39E-156 | 0.00E+00 | 1.00E+00 | 9.02E-156 | 1 | 0 | 0 |

| | | | coloc SUSIE method | | | | | | | | | | |
|---|---|---|---|---|---|---|---|---|---|---|---|---|---|
| **Trait1** | **Trait2** | **cFDRlocusnum** | **nsnps2** | **hit1** | **hit2** | **PP.H0.abf** | **PP.H1.abf** | **PP.H2.abf** | **PP.H3.abf** | **PP.H4.abf** | **PP.H3+PP.H4≥0.8** | **PP.H3/PP.H4≥3** | **PP.H4/PP.H3≥5** |
| e | | | | | | | | | | | | | |
| AD Kunkle | eGFR | | | rs769449 | rs12721046 | 0.00E+00 | 2.39E-156 | 0.00E+00 | 1.00E+00 | 4.36E-53 | 1 | 0 | 0 |
| AD Kunkle | eGFR | | | rs75627662 | rs12721046 | 0.00E+00 | 2.39E-156 | 0.00E+00 | 1.00E+00 | 2.56E-110 | 1 | 0 | 0 |
| AD Kunkle | eGFR | | | rs10775543 | rs12721046 | 0.00E+00 | 2.39E-156 | 0.00E+00 | 1.00E+00 | 4.44E-158 | 1 | 0 | 0 |
| AD Kunkle | eGFR | | | rs204907 | rs12721046 | 0.00E+00 | 2.39E-156 | 0.00E+00 | 1.00E+00 | 1.71E-158 | 1 | 0 | 0 |
| AD Kunkle | eGFR | | | rs429358 | rs150966173 | 0.00E+00 | 1.98E-34 | 0.00E+00 | 1.00E+00 | 1.19E-30 | 1 | 0 | 0 |
| AD Kunkle | eGFR | | | rs12691088 | rs150966173 | 2.56E-102 | 1.98E-34 | 1.29E-68 | 1.00E+00 | 7.96E-37 | 1 | 0 | 0 |
| AD Kunkle | eGFR | | | rs814573 | rs150966173 | 0.00E+00 | 1.98E-34 | 0.00E+00 | 1.00E+00 | 2.84E-32 | 1 | 0 | 0 |
| AD Kunkle | eGFR | | | rs60049679 | rs150966173 | 6.73E-108 | 1.98E-34 | 3.40E-74 | 1.00E+00 | 4.51E-32 | 1 | 0 | 0 |
| AD Kunkle | eGFR | | | rs1160983 | rs150966173 | 0.00E+00 | 1.98E-34 | 0.00E+00 | 1.00E+00 | 7.77E-34 | 1 | 0 | 0 |
| AD Kunkle | eGFR | | | rs41290120 | rs150966173 | 0.00E+00 | 1.98E-34 | 0.00E+00 | 1.00E+00 | 4.55E-36 | 1 | 0 | 0 |
| AD Kunkle | eGFR | | | rs769449 | rs150966173 | 0.00E+00 | 1.98E-34 | 0.00E+00 | 1.00E+00 | 4.44E-36 | 1 | 0 | 0 |
| AD Kunkle | eGFR | | | rs75627662 | rs150966173 | 0.00E+00 | 1.98E-34 | 0.00E+00 | 1.00E+00 | 4.20E-35 | 1 | 0 | 0 |
| AD Kunkle | eGFR | | | rs10775543 | rs150966173 | 0.00E+00 | 1.98E-34 | 0.00E+00 | 1.00E+00 | 1.07E-36 | 1 | 0 | 0 |

| | | | coloc SUSIE method | | | | | | | | | | |
|---|---|---|---|---|---|---|---|---|---|---|---|---|---|
| **Trait1** | **Trait 2** | **cFDRlocusnum** | **nsnps 2** | **hit1** | **hit2** | **PP.H0.abf** | **PP.H1.abf** | **PP.H2.abf** | **PP.H3.abf** | **PP.H4.abf** | **PP.H3+PP.H4≥0.8** | **PP.H3/PP.H4≥3** | **PP.H4/PP.H3≥5** |
| AD Kunkle | eGFR | | | rs204907 | rs150966173 | 0.00E+00 | 1.98E-34 | 0.00E+00 | 1.00E+00 | 8.19E-37 | 1 | 0 | 0 |
| AD Kunkle | eGFR | 14a | 3290 | rs429358 | rs429358 | 0.00E+00 | 1.89E-29 | 0.00E+00 | 2.60E-09 | 1.00E+00 | 1 | 1 | 1 |
| AD Kunkle | eGFR | | | rs12691088 | rs429358 | 1.22E-94 | 9.46E-27 | 1.29E-68 | 1.00E+00 | 8.99E-27 | 1 | 0 | 0 |
| AD Kunkle | eGFR | | | rs814573 | rs429358 | 0.00E+00 | 9.46E-27 | 0.00E+00 | 1.00E+00 | 2.06E-07 | 1 | 0 | 0 |
| AD Kunkle | eGFR | | | rs60049679 | rs429358 | 3.22E-100 | 9.46E-27 | 3.40E-74 | 1.00E+00 | 3.61E-25 | 1 | 0 | 0 |
| AD Kunkle | eGFR | | | rs1160983 | rs429358 | 0.00E+00 | 9.46E-27 | 0.00E+00 | 1.00E+00 | 1.75E-25 | 1 | 0 | 0 |
| AD Kunkle | eGFR | | | rs41290120 | rs429358 | 0.00E+00 | 9.46E-27 | 0.00E+00 | 1.00E+00 | 9.17E-28 | 1 | 0 | 0 |
| AD Kunkle | eGFR | | | rs769449 | rs429358 | 0.00E+00 | 9.46E-27 | 0.00E+00 | 1.00E+00 | 2.96E-10 | 1 | 0 | 0 |
| AD Kunkle | eGFR | | | rs75627662 | rs429358 | 0.00E+00 | 9.46E-27 | 0.00E+00 | 1.00E+00 | 5.39E-22 | 1 | 0 | 0 |
| AD Kunkle | eGFR | | | rs10775543 | rs429358 | 0.00E+00 | 9.46E-27 | 0.00E+00 | 1.00E+00 | 8.35E-29 | 1 | 0 | 0 |
| AD Kunkle | eGFR | | | rs204907 | rs429358 | 0.00E+00 | 9.46E-27 | 0.00E+00 | 1.00E+00 | 5.23E-29 | 1 | 0 | 0 |
| AD Kunkle | eGFR | | | rs429358 | rs814573 | 0.00E+00 | 1.61E-253 | 0.00E+00 | 1.00E+00 | 1.61E-73 | 1 | 0 | 0 |
| AD Kunkle | eGFR | | | rs12691088 | rs814573 | 2.08E-321 | 1.61E-253 | 1.29E-68 | 1.00E+00 | 1.47E-66 | 1 | 0 | 0 |

| | | | coloc SUSIE method | | | | | | | | | | |
|---|---|---|---|---|---|---|---|---|---|---|---|---|---|
| Trait1 | Trait 2 | cFDRlocusnum | nsnps 2 | hit1 | hit2 | PP.H0.abf | PP.H1.abf | PP.H2.abf | PP.H3.abf | PP.H4.abf | PP.H3+PP.H4≥0.8 | PP.H3/PP.H4≥3 | PP.H4/PP.H3≥5 |
| AD Kunkle | eGFR | | | rs814573 | rs814573 | 0.00E+00 | 3.22E-256 | 0.00E+00 | 0.00E+00 | 1.00E+00 | 1 | 1 | 1 |
| AD Kunkle | eGFR | | | rs60049679 | rs814573 | 0.00E+00 | 1.61E-253 | 3.40E-74 | 1.00E+00 | 1.88E-64 | 1 | 0 | 0 |
| AD Kunkle | eGFR | | | rs1160983 | rs814573 | 0.00E+00 | 1.61E-253 | 0.00E+00 | 1.00E+00 | 3.30E-256 | 1 | 0 | 0 |
| AD Kunkle | eGFR | | | rs41290120 | rs814573 | 0.00E+00 | 1.61E-253 | 0.00E+00 | 1.00E+00 | 3.79E-255 | 1 | 0 | 0 |
| AD Kunkle | eGFR | | | rs769449 | rs814573 | 0.00E+00 | 1.61E-253 | 0.00E+00 | 1.00E+00 | 1.21E-99 | 1 | 0 | 0 |
| AD Kunkle | eGFR | | | rs75627662 | rs814573 | 0.00E+00 | 1.61E-253 | 0.00E+00 | 1.00E+00 | 2.48E-173 | 1 | 0 | 0 |
| AD Kunkle | eGFR | | | rs10775543 | rs814573 | 0.00E+00 | 1.61E-253 | 0.00E+00 | 1.00E+00 | 6.30E-256 | 1 | 0 | 0 |
| AD Kunkle | eGFR | | | rs204907 | rs814573 | 0.00E+00 | 1.61E-253 | 0.00E+00 | 1.00E+00 | 1.47E-255 | 1 | 0 | 0 |
| AD Kunkle | eGFR | | | rs429358 | rs12721046 | 0.00E+00 | 5.43E-156 | 0.00E+00 | 1.00E+00 | 6.17E-67 | 1 | 0 | 0 |
| AD Kunkle | eGFR | | | rs12691088 | rs12721046 | 7.01E-224 | 5.43E-156 | 1.29E-68 | 1.00E+00 | 2.70E-49 | 1 | 0 | 0 |
| AD Kunkle | eGFR | | | rs814573 | rs12721046 | 0.00E+00 | 5.43E-156 | 0.00E+00 | 1.00E+00 | 1.28E-47 | 1 | 0 | 0 |
| AD Kunkle | eGFR | | | rs60049679 | rs12721046 | 1.85E-229 | 5.43E-156 | 3.40E-74 | 1.00E+00 | 4.53E-46 | 1 | 0 | 0 |
| AD Kunkle | eGFR | | | rs1160983 | rs12721046 | 0.00E+00 | 5.43E-156 | 0.00E+00 | 1.00E+00 | 4.79E-153 | 1 | 0 | 0 |
| AD | eGFR | | | rs4129012 | rs12721046 | 0.00E+00 | 5.43E-156 | 0.00E+00 | 1.00E+00 | 2.20E-155 | 1 | 0 | 0 |

| | | | coloc SUSIE method | | | | | | | | | | |
|---|---|---|---|---|---|---|---|---|---|---|---|---|---|
| Trait1 | Trait 2 | cFDRlocusnum | nsnps 2 | hit1 | hit2 | PP.H0.abf | PP.H1.abf | PP.H2.abf | PP.H3.abf | PP.H4.abf | PP.H3+PP.H4≥0.8 | PP.H3/PP.H4≥3 | PP.H4/PP.H3≥5 |
| Kunkle | | | | 0 | | | | | | | | | |
| AD Kunkle | eGFR | | | rs769449 | rs12721046 | 0.00E+00 | 5.43E-156 | 0.00E+00 | 1.00E+00 | 5.69E-53 | 1 | 0 | 0 |
| AD Kunkle | eGFR | | | rs75627662 | rs12721046 | 0.00E+00 | 5.43E-156 | 0.00E+00 | 1.00E+00 | 3.62E-110 | 1 | 0 | 0 |
| AD Kunkle | eGFR | | | rs10775543 | rs12721046 | 0.00E+00 | 5.43E-156 | 0.00E+00 | 1.00E+00 | 9.85E-158 | 1 | 0 | 0 |
| AD Kunkle | eGFR | | | rs204907 | rs12721046 | 0.00E+00 | 5.43E-156 | 0.00E+00 | 1.00E+00 | 3.82E-158 | 1 | 0 | 0 |
| AD Kunkle | eGFR | | | rs429358 | rs150966173 | 0.00E+00 | 2.03E-34 | 0.00E+00 | 1.00E+00 | 1.22E-30 | 1 | 0 | 0 |
| AD Kunkle | eGFR | | | rs12691088 | rs150966173 | 2.62E-102 | 2.03E-34 | 1.29E-68 | 1.00E+00 | 8.24E-37 | 1 | 0 | 0 |
| AD Kunkle | eGFR | | | rs814573 | rs150966173 | 0.00E+00 | 2.03E-34 | 0.00E+00 | 1.00E+00 | 2.92E-32 | 1 | 0 | 0 |
| AD Kunkle | eGFR | | | rs60049679 | rs150966173 | 6.91E-108 | 2.03E-34 | 3.40E-74 | 1.00E+00 | 4.84E-32 | 1 | 0 | 0 |
| AD Kunkle | eGFR | | | rs1160983 | rs150966173 | 0.00E+00 | 2.03E-34 | 0.00E+00 | 1.00E+00 | 8.11E-34 | 1 | 0 | 0 |
| AD Kunkle | eGFR | | | rs41290120 | rs150966173 | 0.00E+00 | 2.03E-34 | 0.00E+00 | 1.00E+00 | 4.86E-36 | 1 | 0 | 0 |
| AD Kunkle | eGFR | | | rs769449 | rs150966173 | 0.00E+00 | 2.03E-34 | 0.00E+00 | 1.00E+00 | 4.54E-36 | 1 | 0 | 0 |
| AD Kunkle | eGFR | | | rs75627662 | rs150966173 | 0.00E+00 | 2.03E-34 | 0.00E+00 | 1.00E+00 | 4.47E-35 | 1 | 0 | 0 |
| AD Kunkl | eGFR | | | rs10775543 | rs150966173 | 0.00E+00 | 2.03E-34 | 0.00E+00 | 1.00E+00 | 1.09E-36 | 1 | 0 | 0 |

| | | | coloc SUSIE method | | | | | | | | | | |
|---|---|---|---|---|---|---|---|---|---|---|---|---|---|
| Trait1 | Trait 2 | cFDRlocusnum | nsnps 2 | hit1 | hit2 | PP.H0.abf | PP.H1.abf | PP.H2.abf | PP.H3.abf | PP.H4.abf | PP.H3+PP.H4≥0.8 | PP.H3/PP.H4≥3 | PP.H4/PP.H3≥5 |
| e | | | | | | | | | | | | | |
| AD Kunkle | eGFR | | | rs204907 | rs150966173 | 0.00E+00 | 2.03E-34 | 0.00E+00 | 1.00E+00 | 8.36E-37 | 1 | 0 | 0 |
| AD Kunkle | eGFR | 14b | 3229 | rs429358 | rs429358 | 0.00E+00 | 1.84E-29 | 0.00E+00 | 2.59E-09 | 1.00E+00 | 1 | 1 | 1 |
| AD Kunkle | eGFR | | | rs12691088 | rs429358 | 1.19E-94 | 9.22E-27 | 1.29E-68 | 1.00E+00 | 8.79E-27 | 1 | 0 | 0 |
| AD Kunkle | eGFR | | | rs814573 | rs429358 | 0.00E+00 | 9.22E-27 | 0.00E+00 | 1.00E+00 | 2.03E-07 | 1 | 0 | 0 |
| AD Kunkle | eGFR | | | rs60049679 | rs429358 | 3.14E-100 | 9.22E-27 | 3.40E-74 | 1.00E+00 | 3.51E-25 | 1 | 0 | 0 |
| AD Kunkle | eGFR | | | rs1160983 | rs429358 | 0.00E+00 | 9.22E-27 | 0.00E+00 | 1.00E+00 | 1.64E-25 | 1 | 0 | 0 |
| AD Kunkle | eGFR | | | rs41290120 | rs429358 | 0.00E+00 | 9.22E-27 | 0.00E+00 | 1.00E+00 | 8.90E-28 | 1 | 0 | 0 |
| AD Kunkle | eGFR | | | rs769449 | rs429358 | 0.00E+00 | 9.22E-27 | 0.00E+00 | 1.00E+00 | 2.94E-10 | 1 | 0 | 0 |
| AD Kunkle | eGFR | | | rs75627662 | rs429358 | 0.00E+00 | 9.22E-27 | 0.00E+00 | 1.00E+00 | 5.46E-22 | 1 | 0 | 0 |
| AD Kunkle | eGFR | | | rs10775543 | rs429358 | 0.00E+00 | 9.22E-27 | 0.00E+00 | 1.00E+00 | 8.14E-29 | 1 | 0 | 0 |
| AD Kunkle | eGFR | | | rs204907 | rs429358 | 0.00E+00 | 9.22E-27 | 0.00E+00 | 1.00E+00 | 5.10E-29 | 1 | 0 | 0 |
| AD Kunkle | eGFR | | | rs429358 | rs814573 | 0.00E+00 | 1.95E-253 | 0.00E+00 | 1.00E+00 | 1.66E-73 | 1 | 0 | 0 |
| AD Kunkl | eGFR | | | rs12691088 | rs814573 | 2.51E-321 | 1.95E-253 | 1.29E-68 | 1.00E+00 | 1.47E-66 | 1 | 0 | 0 |

| | | | coloc SUSIE method | | | | | | | | | | |
|---|---|---|---|---|---|---|---|---|---|---|---|---|---|
| Trait1 | Trait 2 | cFDRlocusnum | nsnps2 | hit1 | hit2 | PP.H0.abf | PP.H1.abf | PP.H2.abf | PP.H3.abf | PP.H4.abf | PP.H3+PP.H4≥0.8 | PP.H3/PP.H4≥3 | PP.H4/PP.H3≥5 |
| e | | | | | | | | | | | | | |
| AD Kunkle | eGFR | | | rs814573 | rs814573 | 0.00E+00 | 3.89E-256 | 0.00E+00 | 0.00E+00 | 1.00E+00 | 1 | 1 | 1 |
| AD Kunkle | eGFR | | | rs60049679 | rs814573 | 0.00E+00 | 1.95E-253 | 3.40E-74 | 1.00E+00 | 1.88E-64 | 1 | 0 | 0 |
| AD Kunkle | eGFR | | | rs1160983 | rs814573 | 0.00E+00 | 1.95E-253 | 0.00E+00 | 1.00E+00 | 3.97E-256 | 1 | 0 | 0 |
| AD Kunkle | eGFR | | | rs41290120 | rs814573 | 0.00E+00 | 1.95E-253 | 0.00E+00 | 1.00E+00 | 4.60E-255 | 1 | 0 | 0 |
| AD Kunkle | eGFR | | | rs769449 | rs814573 | 0.00E+00 | 1.95E-253 | 0.00E+00 | 1.00E+00 | 1.28E-99 | 1 | 0 | 0 |
| AD Kunkle | eGFR | | | rs75627662 | rs814573 | 0.00E+00 | 1.95E-253 | 0.00E+00 | 1.00E+00 | 2.63E-173 | 1 | 0 | 0 |
| AD Kunkle | eGFR | | | rs10775543 | rs814573 | 0.00E+00 | 1.95E-253 | 0.00E+00 | 1.00E+00 | 7.62E-256 | 1 | 0 | 0 |
| AD Kunkle | eGFR | | | rs204907 | rs814573 | 0.00E+00 | 1.95E-253 | 0.00E+00 | 1.00E+00 | 1.77E-255 | 1 | 0 | 0 |
| AD Kunkle | eGFR | | | rs429358 | rs12721046 | 0.00E+00 | 6.29E-156 | 0.00E+00 | 1.00E+00 | 6.59E-67 | 1 | 0 | 0 |
| AD Kunkle | eGFR | | | rs12691088 | rs12721046 | 8.12E-224 | 6.29E-156 | 1.29E-68 | 1.00E+00 | 2.70E-49 | 1 | 0 | 0 |
| AD Kunkle | eGFR | | | rs814573 | rs12721046 | 0.00E+00 | 6.29E-156 | 0.00E+00 | 1.00E+00 | 1.32E-47 | 1 | 0 | 0 |
| AD Kunkle | eGFR | | | rs60049679 | rs12721046 | 2.14E-229 | 6.29E-156 | 3.40E-74 | 1.00E+00 | 4.53E-46 | 1 | 0 | 0 |
| AD Kunkle | eGFR | | | rs1160983 | rs12721046 | 0.00E+00 | 6.29E-156 | 0.00E+00 | 1.00E+00 | 5.39E-153 | 1 | 0 | 0 |

| | | | coloc SUSIE method | | | | | | | | | | |
|---|---|---|---|---|---|---|---|---|---|---|---|---|---|
| **Trait1** | **Trait2** | **cFDRlocusnum** | **nsnps2** | **hit1** | **hit2** | **PP.H0.abf** | **PP.H1.abf** | **PP.H2.abf** | **PP.H3.abf** | **PP.H4.abf** | **PP.H3+PP.H4≥0.8** | **PP.H3/PP.H4≥3** | **PP.H4/PP.H3≥5** |
| AD Kunkle | eGFR | | | rs41290120 | rs12721046 | 0.00E+00 | 6.29E-156 | 0.00E+00 | 1.00E+00 | 2.53E-155 | 1 | 0 | 0 |
| AD Kunkle | eGFR | | | rs769449 | rs12721046 | 0.00E+00 | 6.29E-156 | 0.00E+00 | 1.00E+00 | 5.96E-53 | 1 | 0 | 0 |
| AD Kunkle | eGFR | | | rs75627662 | rs12721046 | 0.00E+00 | 6.29E-156 | 0.00E+00 | 1.00E+00 | 4.19E-110 | 1 | 0 | 0 |
| AD Kunkle | eGFR | | | rs10775543 | rs12721046 | 0.00E+00 | 6.29E-156 | 0.00E+00 | 1.00E+00 | 1.14E-157 | 1 | 0 | 0 |
| AD Kunkle | eGFR | | | rs204907 | rs12721046 | 0.00E+00 | 6.29E-156 | 0.00E+00 | 1.00E+00 | 4.43E-158 | 1 | 0 | 0 |
| AD Kunkle | eGFR | | | rs429358 | rs150966173 | 0.00E+00 | 2.13E-34 | 0.00E+00 | 1.00E+00 | 1.27E-30 | 1 | 0 | 0 |
| AD Kunkle | eGFR | | | rs12691088 | rs150966173 | 2.76E-102 | 2.13E-34 | 1.29E-68 | 1.00E+00 | 8.66E-37 | 1 | 0 | 0 |
| AD Kunkle | eGFR | | | rs814573 | rs150966173 | 0.00E+00 | 2.13E-34 | 0.00E+00 | 1.00E+00 | 3.04E-32 | 1 | 0 | 0 |
| AD Kunkle | eGFR | | | rs60049679 | rs150966173 | 7.26E-108 | 2.13E-34 | 3.40E-74 | 1.00E+00 | 5.05E-32 | 1 | 0 | 0 |
| AD Kunkle | eGFR | | | rs1160983 | rs150966173 | 0.00E+00 | 2.13E-34 | 0.00E+00 | 1.00E+00 | 8.35E-34 | 1 | 0 | 0 |
| AD Kunkle | eGFR | | | rs41290120 | rs150966173 | 0.00E+00 | 2.13E-34 | 0.00E+00 | 1.00E+00 | 5.09E-36 | 1 | 0 | 0 |
| AD Kunkle | eGFR | | | rs769449 | rs150966173 | 0.00E+00 | 2.13E-34 | 0.00E+00 | 1.00E+00 | 4.77E-36 | 1 | 0 | 0 |
| AD Kunkle | eGFR | | | rs75627662 | rs150966173 | 0.00E+00 | 2.13E-34 | 0.00E+00 | 1.00E+00 | 4.65E-35 | 1 | 0 | 0 |
| AD | eGFR | | | rs1077554 | rs15096617 | 0.00E+00 | 2.13E-34 | 0.00E+00 | 1.00E+00 | 1.14E-36 | 1 | 0 | 0 |

| | | | coloc SUSIE method | | | | | | | | | | |
|---|---|---|---|---|---|---|---|---|---|---|---|---|---|
| **Trait1** | **Trait2** | **cFDRlocusnum** | **nsnps2** | **hit1** | **hit2** | **PP.H0.abf** | **PP.H1.abf** | **PP.H2.abf** | **PP.H3.abf** | **PP.H4.abf** | **PP.H3+PP.H4≥0.8** | **PP.H3/PP.H4≥3** | **PP.H4/PP.H3≥5** |
| Kunkle | | | | 3 | 3 | | | | | | | | |
| AD Kunkle | eGFR | | | rs204907 | rs150966173 | 0.00E+00 | 2.13E-34 | 0.00E+00 | 1.00E+00 | 8.79E-37 | 1 | 0 | 0 |
| AD Kunkle | eGFR | 14c | 3206 | rs429358 | rs429358 | 0.00E+00 | 1.82E-29 | 0.00E+00 | 2.60E-09 | 1.00E+00 | 1 | 1 | 1 |
| AD Kunkle | eGFR | | | rs12691088 | rs429358 | 1.18E-94 | 9.11E-27 | 1.29E-68 | 1.00E+00 | 8.64E-27 | 1 | 0 | 0 |
| AD Kunkle | eGFR | | | rs814573 | rs429358 | 0.00E+00 | 9.11E-27 | 0.00E+00 | 1.00E+00 | 2.03E-07 | 1 | 0 | 0 |
| AD Kunkle | eGFR | | | rs60049679 | rs429358 | 3.10E-100 | 9.11E-27 | 3.40E-74 | 1.00E+00 | 3.46E-25 | 1 | 0 | 0 |
| AD Kunkle | eGFR | | | rs1160983 | rs429358 | 0.00E+00 | 9.11E-27 | 0.00E+00 | 1.00E+00 | 1.60E-25 | 1 | 0 | 0 |
| AD Kunkle | eGFR | | | rs41290120 | rs429358 | 0.00E+00 | 9.11E-27 | 0.00E+00 | 1.00E+00 | 8.80E-28 | 1 | 0 | 0 |
| AD Kunkle | eGFR | | | rs769449 | rs429358 | 0.00E+00 | 9.11E-27 | 0.00E+00 | 1.00E+00 | 2.94E-10 | 1 | 0 | 0 |
| AD Kunkle | eGFR | | | rs75627662 | rs429358 | 0.00E+00 | 9.11E-27 | 0.00E+00 | 1.00E+00 | 5.46E-22 | 1 | 0 | 0 |
| AD Kunkle | eGFR | | | rs10775543 | rs429358 | 0.00E+00 | 9.11E-27 | 0.00E+00 | 1.00E+00 | 8.04E-29 | 1 | 0 | 0 |
| AD Kunkle | eGFR | | | rs204907 | rs429358 | 0.00E+00 | 9.11E-27 | 0.00E+00 | 1.00E+00 | 5.03E-29 | 1 | 0 | 0 |
| AD Kunkle | eGFR | | | rs429358 | rs814573 | 0.00E+00 | 1.93E-253 | 0.00E+00 | 1.00E+00 | 1.68E-73 | 1 | 0 | 0 |
| AD | eGFR | | | rs1269108 | rs814573 | 2.49E-321 | 1.93E-253 | 1.29E-68 | 1.00E+00 | 1.47E-66 | 1 | 0 | 0 |

| | | | coloc SUSIE method | | | | | | | | | | |
|---|---|---|---|---|---|---|---|---|---|---|---|---|---|
| **Trait1** | **Trait2** | **cFDRlocusnum** | **nsnps2** | **hit1** | **hit2** | **PP.H0.abf** | **PP.H1.abf** | **PP.H2.abf** | **PP.H3.abf** | **PP.H4.abf** | **PP.H3+PP.H4≥0.8** | **PP.H3/PP.H4≥3** | **PP.H4/PP.H3≥5** |
| Kunkle | | | | 8 | | | | | | | | | |
| AD Kunkle | eGFR | | | rs814573 | rs814573 | 0.00E+00 | 3.85E-256 | 0.00E+00 | 0.00E+00 | 1.00E+00 | 1 | 1 | 1 |
| AD Kunkle | eGFR | | | rs60049679 | rs814573 | 0.00E+00 | 1.93E-253 | 3.40E-74 | 1.00E+00 | 1.88E-64 | 1 | 0 | 0 |
| AD Kunkle | eGFR | | | rs1160983 | rs814573 | 0.00E+00 | 1.93E-253 | 0.00E+00 | 1.00E+00 | 3.94E-256 | 1 | 0 | 0 |
| AD Kunkle | eGFR | | | rs41290120 | rs814573 | 0.00E+00 | 1.93E-253 | 0.00E+00 | 1.00E+00 | 4.55E-255 | 1 | 0 | 0 |
| AD Kunkle | eGFR | | | rs769449 | rs814573 | 0.00E+00 | 1.93E-253 | 0.00E+00 | 1.00E+00 | 1.28E-99 | 1 | 0 | 0 |
| AD Kunkle | eGFR | | | rs75627662 | rs814573 | 0.00E+00 | 1.93E-253 | 0.00E+00 | 1.00E+00 | 2.65E-173 | 1 | 0 | 0 |
| AD Kunkle | eGFR | | | rs10775543 | rs814573 | 0.00E+00 | 1.93E-253 | 0.00E+00 | 1.00E+00 | 7.55E-256 | 1 | 0 | 0 |
| AD Kunkle | eGFR | | | rs204907 | rs814573 | 0.00E+00 | 1.93E-253 | 0.00E+00 | 1.00E+00 | 1.76E-255 | 1 | 0 | 0 |
| AD Kunkle | eGFR | | | rs429358 | rs12721046 | 0.00E+00 | 6.60E-156 | 0.00E+00 | 1.00E+00 | 6.71E-67 | 1 | 0 | 0 |
| AD Kunkle | eGFR | | | rs12691088 | rs12721046 | 8.53E-224 | 6.60E-156 | 1.29E-68 | 1.00E+00 | 2.70E-49 | 1 | 0 | 0 |
| AD Kunkle | eGFR | | | rs814573 | rs12721046 | 0.00E+00 | 6.60E-156 | 0.00E+00 | 1.00E+00 | 1.35E-47 | 1 | 0 | 0 |
| AD Kunkle | eGFR | | | rs60049679 | rs12721046 | 2.25E-229 | 6.60E-156 | 3.40E-74 | 1.00E+00 | 4.53E-46 | 1 | 0 | 0 |
| AD Kunkl | eGFR | | | rs1160983 | rs12721046 | 0.00E+00 | 6.60E-156 | 0.00E+00 | 1.00E+00 | 5.69E-153 | 1 | 0 | 0 |

| | | | coloc SUSIE method | | | | | | | | | | |
|---|---|---|---|---|---|---|---|---|---|---|---|---|---|
| **Trait1** | **Trait2** | **cFDRlocusnum** | **nsnps2** | **hit1** | **hit2** | **PP.H0.abf** | **PP.H1.abf** | **PP.H2.abf** | **PP.H3.abf** | **PP.H4.abf** | **PP.H3+PP.H4≥0.8** | **PP.H3/PP.H4≥3** | **PP.H4/PP.H3≥5** |
| e | | | | | | | | | | | | | |
| AD Kunkle | eGFR | | | rs41290120 | rs12721046 | 0.00E+00 | 6.60E-156 | 0.00E+00 | 1.00E+00 | 2.66E-155 | 1 | 0 | 0 |
| AD Kunkle | eGFR | | | rs769449 | rs12721046 | 0.00E+00 | 6.60E-156 | 0.00E+00 | 1.00E+00 | 6.06E-53 | 1 | 0 | 0 |
| AD Kunkle | eGFR | | | rs75627662 | rs12721046 | 0.00E+00 | 6.60E-156 | 0.00E+00 | 1.00E+00 | 4.30E-110 | 1 | 0 | 0 |
| AD Kunkle | eGFR | | | rs10775543 | rs12721046 | 0.00E+00 | 6.60E-156 | 0.00E+00 | 1.00E+00 | 1.20E-157 | 1 | 0 | 0 |
| AD Kunkle | eGFR | | | rs204907 | rs12721046 | 0.00E+00 | 6.60E-156 | 0.00E+00 | 1.00E+00 | 4.64E-158 | 1 | 0 | 0 |
| AD Kunkle | eGFR | | | rs429358 | rs150966173 | 0.00E+00 | 2.13E-34 | 0.00E+00 | 1.00E+00 | 1.27E-30 | 1 | 0 | 0 |
| AD Kunkle | eGFR | | | rs12691088 | rs150966173 | 2.75E-102 | 2.13E-34 | 1.29E-68 | 1.00E+00 | 8.66E-37 | 1 | 0 | 0 |
| AD Kunkle | eGFR | | | rs814573 | rs150966173 | 0.00E+00 | 2.13E-34 | 0.00E+00 | 1.00E+00 | 3.04E-32 | 1 | 0 | 0 |
| AD Kunkle | eGFR | | | rs60049679 | rs150966173 | 7.25E-108 | 2.13E-34 | 3.40E-74 | 1.00E+00 | 5.03E-32 | 1 | 0 | 0 |
| AD Kunkle | eGFR | | | rs1160983 | rs150966173 | 0.00E+00 | 2.13E-34 | 0.00E+00 | 1.00E+00 | 8.37E-34 | 1 | 0 | 0 |
| AD Kunkle | eGFR | | | rs41290120 | rs150966173 | 0.00E+00 | 2.13E-34 | 0.00E+00 | 1.00E+00 | 5.09E-36 | 1 | 0 | 0 |
| AD Kunkle | eGFR | | | rs769449 | rs150966173 | 0.00E+00 | 2.13E-34 | 0.00E+00 | 1.00E+00 | 4.77E-36 | 1 | 0 | 0 |
| AD Kunkle | eGFR | | | rs75627662 | rs150966173 | 0.00E+00 | 2.13E-34 | 0.00E+00 | 1.00E+00 | 4.66E-35 | 1 | 0 | 0 |

| | | | coloc SUSIE method | | | | | | | | | | |
|---|---|---|---|---|---|---|---|---|---|---|---|---|---|
| **Trait1** | **Trait 2** | **cFDRlocusnum** | **nsnps 2** | **hit1** | **hit2** | **PP.H0.abf** | **PP.H1.abf** | **PP.H2.abf** | **PP.H3.abf** | **PP.H4.abf** | **PP.H3+PP.H4≥0.8** | **PP.H3/PP.H4≥3** | **PP.H4/PP.H3≥5** |
| AD Kunkle | eGFR | | | rs10775543 | rs150966173 | 0.00E+00 | 2.13E-34 | 0.00E+00 | 1.00E+00 | 1.14E-36 | 1 | 0 | 0 |
| AD Kunkle | eGFR | | | rs204907 | rs150966173 | 0.00E+00 | 2.13E-34 | 0.00E+00 | 1.00E+00 | 8.78E-37 | 1 | 0 | 0 |

**Supplementary Table S26. Colocalization Analysis Results Using SuSiE Framework (Wightman Validation).**
This table presents colocalization analysis results in the validation dataset using the Wightman AD GWAS (corrected for competing risk). Validation of colocalization findings in an independent AD sample is critical for establishing that conclusions about shared versus distinct causal variants generalize beyond a single study. The table reports posterior probabilities for all five colocalization hypotheses at each locus, enabling comparison with discovery findings. Note: 0 indicates "No", and 1 indicates "Yes".

| | | | coloc SUSIE method | | | | | | | | | | |
|---|---|---|---|---|---|---|---|---|---|---|---|---|---|
| **Trait1** | **Trait 2** | **cFDRlocusnum** | **nsnps 2** | **hit1** | **hit2** | **PP.H0.abf** | **PP.H1.abf** | **PP.H2.abf** | **PP.H3.abf** | **PP.H4.abf** | **PP.H3+PP.H4≥0.8** | **PP.H3/PP.H4≥3** | **PP.H4/PP.H3≥5** |
| AD Wightman Corrected | eGFR | 13 | 1437 | rs6985003 | rs11786896 | 7.89E-17 | 6.57E-15 | 1.08E-02 | 8.94E-01 | 9.48E-02 | 1 | 0 | 0 |
| AD Wightman Corrected | eGFR | | | rs6985003 | rs6985555 | 8.76E-07 | 7.29E-05 | 1.19E-02 | 9.88E-01 | 1.42E-04 | 1 | 0 | 0 |
| AD Wightman Corrected | eGFR | 19 | 1453 | rs11039348 | rs10838702 | 3.92E-19 | 2.31E-13 | 1.69E-06 | 9.99E-01 | 1.12E-03 | 1 | 0 | 0 |
| AD Wightman Corrected | eGFR | 32 | 2912 | rs9912287 | rs57937546 | 1.29E-05 | 4.41E-04 | 1.55E-02 | 5.30E-01 | 4.54E-01 | 1 | 0 | 0 |
| AD Wightman Corrected | eGFR | | | rs9912287 | rs2131704 | 7.68E-08 | 2.63E-06 | 2.83E-02 | 9.71E-01 | 9.69E-04 | 1 | 0 | 0 |

| | | | coloc SUSIE method | | | | | | | | | | |
|---|---|---|---|---|---|---|---|---|---|---|---|---|---|
| **Trait1** | **Trait 2** | **cFDRlocusnum** | **nsnps 2** | **hit1** | **hit2** | **PP.H0.abf** | **PP.H1.abf** | **PP.H2.abf** | **PP.H3.abf** | **PP.H4.abf** | **PP.H3+PP.H4≥0.8** | **PP.H3/PP.H4≥3** | **PP.H4/PP.H3≥5** |
| AD Wightman Corrected | eGFR | 33 | 2813 | rs9912287 | rs57937546 | 1.29E-05 | 4.41E-04 | 1.55E-02 | 5.30E-01 | 4.54E-01 | 1 | 0 | 0 |
| AD Wightman Corrected | eGFR | | | rs9912287 | rs2131704 | 7.24E-08 | 2.48E-06 | 2.83E-02 | 9.71E-01 | 9.72E-04 | 1 | 0 | 0 |
| AD Wightman Corrected | eGFR | 41 | 2838 | rs429358 | rs429358 | 0.00E+00 | 5.17E-15 | 0.00E+00 | 5.81E-05 | 1.00E+00 | 1 | 1 | 1 |
| AD Wightman Corrected | eGFR | | | rs6857 | rs429358 | 0.00E+00 | 2.38E-13 | 0.00E+00 | 9.30E-02 | 9.07E-01 | 1 | 1 | 1 |
| AD Wightman Corrected | eGFR | | | rs2075650 | rs429358 | 0.00E+00 | 2.51E-12 | 0.00E+00 | 9.98E-01 | 1.79E-03 | 1 | 0 | 0 |
| AD Wightman Corrected | eGFR | | | rs71352238 | rs429358 | 0.00E+00 | 2.49E-12 | 0.00E+00 | 9.92E-01 | 8.26E-03 | 1 | 0 | 0 |
| AD Wightman Corrected | eGFR | | | rs5117 | rs429358 | 0.00E+00 | 2.51E-12 | 0.00E+00 | 1.00E+00 | 1.86E-09 | 1 | 0 | 0 |
| AD Wightman Corrected | eGFR | | | rs77301115 | rs429358 | 0.00E+00 | 2.51E-12 | 0.00E+00 | 1.00E+00 | 5.60E-12 | 1 | 0 | 0 |
| AD Wightman Corrected | eGFR | | | rs75627662 | rs429358 | 0.00E+00 | 2.51E-12 | 0.00E+00 | 1.00E+00 | 1.58E-11 | 1 | 0 | 0 |
| AD Wightma | eGFR | | | rs12972970 | rs429358 | 0.00E+00 | 2.51E-12 | 0.00E+00 | 9.99E-01 | 1.16E-03 | 1 | 0 | 0 |

| | | | coloc SUSIE method | | | | | | | | | | |
|---|---|---|---|---|---|---|---|---|---|---|---|---|---|
| Trait1 | Trait 2 | cFDRlocusnum | nsnps 2 | hit1 | hit2 | PP.H0.abf | PP.H1.abf | PP.H2.abf | PP.H3.abf | PP.H4.abf | PP.H3+PP.H4≥0.8 | PP.H3/PP.H4≥3 | PP.H4/PP.H3≥5 |
| n Corrected | | | | | | | | | | | | | |
| AD Wightman Corrected | eGFR | | | rs429358 | rs8104483 | 0.00E+00 | 1.36E-02 | 0.00E+00 | 9.86E-01 | 1.29E-04 | 1 | 0 | 0 |
| AD Wightman Corrected | eGFR | | | rs6857 | rs8104483 | 0.00E+00 | 1.36E-02 | 0.00E+00 | 9.86E-01 | 1.50E-04 | 1 | 0 | 0 |
| AD Wightman Corrected | eGFR | | | rs2075650 | rs8104483 | 0.00E+00 | 1.36E-02 | 0.00E+00 | 9.86E-01 | 1.29E-04 | 1 | 0 | 0 |
| AD Wightman Corrected | eGFR | | | rs71352238 | rs8104483 | 0.00E+00 | 1.36E-02 | 0.00E+00 | 9.86E-01 | 1.43E-04 | 1 | 0 | 0 |
| AD Wightman Corrected | eGFR | | | rs5117 | rs8104483 | 0.00E+00 | 1.36E-02 | 0.00E+00 | 9.85E-01 | 9.64E-04 | 1 | 0 | 0 |
| AD Wightman Corrected | eGFR | | | rs77301115 | rs8104483 | 0.00E+00 | 1.36E-02 | 0.00E+00 | 9.86E-01 | 1.93E-04 | 1 | 0 | 0 |
| AD Wightman Corrected | eGFR | | | rs75627662 | rs8104483 | 0.00E+00 | 1.35E-02 | 0.00E+00 | 9.84E-01 | 2.16E-03 | 1 | 0 | 0 |
| AD Wightman Corrected | eGFR | | | rs12972970 | rs8104483 | 0.00E+00 | 1.36E-02 | 0.00E+00 | 9.86E-01 | 1.30E-04 | 1 | 0 | 0 |
| AD Wightman Corrected | eGFR | 42 | 2877 | rs429358 | rs429358 | 0.00E+00 | 5.12E-15 | 0.00E+00 | 5.77E-05 | 1.00E+00 | 1 | 1 | 1 |
| AD | eGFR | | | rs6857 | rs429358 | 0.00E+00 | 2.39E-13 | 0.00E+00 | 9.40E-02 | 9.06E-01 | 1 | 1 | 1 |

| | | | coloc SUSIE method | | | | | | | | | | |
|---|---|---|---|---|---|---|---|---|---|---|---|---|---|
| **Trait1** | **Trait 2** | **cFDRlocusnum** | **nsnps 2** | **hit1** | **hit2** | **PP.H0.abf** | **PP.H1.abf** | **PP.H2.abf** | **PP.H3.abf** | **PP.H4.abf** | **PP.H3+PP.H4≥0.8** | **PP.H3/PP.H4≥3** | **PP.H4/PP.H3≥5** |
| Wightman Corrected | | | | | | | | | | | | | |
| AD Wightman Corrected | eGFR | | | rs2075650 | rs429358 | 0.00E+00 | 2.49E-12 | 0.00E+00 | 9.98E-01 | 1.77E-03 | 1 | 0 | 0 |
| AD Wightman Corrected | eGFR | | | rs71352238 | rs429358 | 0.00E+00 | 2.47E-12 | 0.00E+00 | 9.92E-01 | 8.18E-03 | 1 | 0 | 0 |
| AD Wightman Corrected | eGFR | | | rs5117 | rs429358 | 0.00E+00 | 2.49E-12 | 0.00E+00 | 1.00E+00 | 1.91E-09 | 1 | 0 | 0 |
| AD Wightman Corrected | eGFR | | | rs77301115 | rs429358 | 0.00E+00 | 2.49E-12 | 0.00E+00 | 1.00E+00 | 5.55E-12 | 1 | 0 | 0 |
| AD Wightman Corrected | eGFR | | | rs75627662 | rs429358 | 0.00E+00 | 2.49E-12 | 0.00E+00 | 1.00E+00 | 1.61E-11 | 1 | 0 | 0 |
| AD Wightman Corrected | eGFR | | | rs12972970 | rs429358 | 0.00E+00 | 2.49E-12 | 0.00E+00 | 9.99E-01 | 1.15E-03 | 1 | 0 | 0 |
| AD Wightman Corrected | eGFR | | | rs429358 | rs8104483 | 0.00E+00 | 1.35E-02 | 0.00E+00 | 9.86E-01 | 1.27E-04 | 1 | 0 | 0 |
| AD Wightman Corrected | eGFR | | | rs6857 | rs8104483 | 0.00E+00 | 1.35E-02 | 0.00E+00 | 9.86E-01 | 1.48E-04 | 1 | 0 | 0 |
| AD Wightman Corrected | eGFR | | | rs2075650 | rs8104483 | 0.00E+00 | 1.35E-02 | 0.00E+00 | 9.86E-01 | 1.27E-04 | 1 | 0 | 0 |
| AD | eGFR | | | rs71352238 | rs8104483 | 0.00E+00 | 1.35E-02 | 0.00E+00 | 9.86E-01 | 1.42E-04 | 1 | 0 | 0 |

| | | | coloc SUSIE method | | | | | | | | | | |
|---|---|---|---|---|---|---|---|---|---|---|---|---|---|
| **Trait1** | **Trait 2** | **cFDRlocusnum** | **nsnps 2** | **hit1** | **hit2** | **PP.H0.abf** | **PP.H1.abf** | **PP.H2.abf** | **PP.H3.abf** | **PP.H4.abf** | **PP.H3+PP.H4≥0.8** | **PP.H3/PP.H4≥3** | **PP.H4/PP.H3≥5** |
| Wightman Corrected | | | | | | | | | | | | | |
| AD Wightman Corrected | eGFR | | | rs5117 | rs8104483 | 0.00E+00 | 1.35E-02 | 0.00E+00 | 9.86E-01 | 9.43E-04 | 1 | 0 | 0 |
| AD Wightman Corrected | eGFR | | | rs77301115 | rs8104483 | 0.00E+00 | 1.35E-02 | 0.00E+00 | 9.86E-01 | 1.92E-04 | 1 | 0 | 0 |
| AD Wightman Corrected | eGFR | | | rs75627662 | rs8104483 | 0.00E+00 | 1.34E-02 | 0.00E+00 | 9.84E-01 | 2.11E-03 | 1 | 0 | 0 |
| AD Wightman Corrected | eGFR | | | rs12972970 | rs8104483 | 0.00E+00 | 1.35E-02 | 0.00E+00 | 9.86E-01 | 1.29E-04 | 1 | 0 | 0 |
| AD Wightman Corrected | eGFR | 43a | 2885 | rs429358 | rs429358 | 0.00E+00 | 5.21E-15 | 0.00E+00 | 5.75E-05 | 1.00E+00 | 1 | 1 | 1 |
| AD Wightman Corrected | eGFR | | | rs6857 | rs429358 | 0.00E+00 | 2.42E-13 | 0.00E+00 | 9.38E-02 | 9.06E-01 | 1 | 1 | 1 |
| AD Wightman Corrected | eGFR | | | rs2075650 | rs429358 | 0.00E+00 | 2.53E-12 | 0.00E+00 | 9.98E-01 | 1.78E-03 | 1 | 0 | 0 |
| AD Wightman Corrected | eGFR | | | rs71352238 | rs429358 | 0.00E+00 | 2.51E-12 | 0.00E+00 | 9.92E-01 | 8.24E-03 | 1 | 0 | 0 |
| AD Wightman Corrected | eGFR | | | rs5117 | rs429358 | 0.00E+00 | 2.53E-12 | 0.00E+00 | 1.00E+00 | 2.07E-09 | 1 | 0 | 0 |

| | | | coloc SUSIE method | | | | | | | | | | |
|---|---|---|---|---|---|---|---|---|---|---|---|---|---|
| **Trait1** | **Trait 2** | **cFDRlocusnum** | **nsnps 2** | **hit1** | **hit2** | **PP.H0.abf** | **PP.H1.abf** | **PP.H2.abf** | **PP.H3.abf** | **PP.H4.abf** | **PP.H3+PP.H4≥0.8** | **PP.H3/PP.H4≥3** | **PP.H4/PP.H3≥5** |
| AD Wightman Corrected | eGFR | | | rs77301115 | rs429358 | 0.00E+00 | 2.53E-12 | 0.00E+00 | 1.00E+00 | 5.63E-12 | 1 | 0 | 0 |
| AD Wightman Corrected | eGFR | | | rs75627662 | rs429358 | 0.00E+00 | 2.53E-12 | 0.00E+00 | 1.00E+00 | 1.73E-11 | 1 | 0 | 0 |
| AD Wightman Corrected | eGFR | | | rs12972970 | rs429358 | 0.00E+00 | 2.53E-12 | 0.00E+00 | 9.99E-01 | 1.16E-03 | 1 | 0 | 0 |
| AD Wightman Corrected | eGFR | | | rs429358 | rs8104483 | 0.00E+00 | 1.34E-02 | 0.00E+00 | 9.86E-01 | 1.27E-04 | 1 | 0 | 0 |
| AD Wightman Corrected | eGFR | | | rs6857 | rs8104483 | 0.00E+00 | 1.34E-02 | 0.00E+00 | 9.86E-01 | 1.48E-04 | 1 | 0 | 0 |
| AD Wightman Corrected | eGFR | | | rs2075650 | rs8104483 | 0.00E+00 | 1.34E-02 | 0.00E+00 | 9.86E-01 | 1.27E-04 | 1 | 0 | 0 |
| AD Wightman Corrected | eGFR | | | rs71352238 | rs8104483 | 0.00E+00 | 1.34E-02 | 0.00E+00 | 9.86E-01 | 1.41E-04 | 1 | 0 | 0 |
| AD Wightman Corrected | eGFR | | | rs5117 | rs8104483 | 0.00E+00 | 1.34E-02 | 0.00E+00 | 9.86E-01 | 9.17E-04 | 1 | 0 | 0 |
| AD Wightman Corrected | eGFR | | | rs77301115 | rs8104483 | 0.00E+00 | 1.34E-02 | 0.00E+00 | 9.86E-01 | 1.91E-04 | 1 | 0 | 0 |
| AD Wightman Corrected | eGFR | | | rs75627662 | rs8104483 | 0.00E+00 | 1.34E-02 | 0.00E+00 | 9.85E-01 | 2.04E-03 | 1 | 0 | 0 |

| | | | coloc SUSIE method | | | | | | | | | | |
|---|---|---|---|---|---|---|---|---|---|---|---|---|---|
| Trait1 | Trait 2 | cFDRlocusnum | nsnps 2 | hit1 | hit2 | PP.H0.abf | PP.H1.abf | PP.H2.abf | PP.H3.abf | PP.H4.abf | PP.H3+PP.H4≥0.8 | PP.H3/PP.H4≥3 | PP.H4/PP.H3≥5 |
| AD Wightman Corrected | eGFR | | | rs12972970 | rs8104483 | 0.00E+00 | 1.34E-02 | 0.00E+00 | 9.86E-01 | 1.28E-04 | 1 | 0 | 0 |
| AD Wightman Corrected | eGFR | 43b | 2890 | rs429358 | rs429358 | 0.00E+00 | 5.01E-15 | 0.00E+00 | 5.67E-05 | 1.00E+00 | 1 | 1 | 1 |
| AD Wightman Corrected | eGFR | | | rs6857 | rs429358 | 0.00E+00 | 2.35E-13 | 0.00E+00 | 9.44E-02 | 9.06E-01 | 1 | 1 | 1 |
| AD Wightman Corrected | eGFR | | | rs2075650 | rs429358 | 0.00E+00 | 2.43E-12 | 0.00E+00 | 9.98E-01 | 1.77E-03 | 1 | 0 | 0 |
| AD Wightman Corrected | eGFR | | | rs71352238 | rs429358 | 0.00E+00 | 2.42E-12 | 0.00E+00 | 9.92E-01 | 8.24E-03 | 1 | 0 | 0 |
| AD Wightman Corrected | eGFR | | | rs5117 | rs429358 | 0.00E+00 | 2.44E-12 | 0.00E+00 | 1.00E+00 | 2.22E-09 | 1 | 0 | 0 |
| AD Wightman Corrected | eGFR | | | rs77301115 | rs429358 | 0.00E+00 | 2.44E-12 | 0.00E+00 | 1.00E+00 | 5.39E-12 | 1 | 0 | 0 |
| AD Wightman Corrected | eGFR | | | rs75627662 | rs429358 | 0.00E+00 | 2.44E-12 | 0.00E+00 | 1.00E+00 | 1.82E-11 | 1 | 0 | 0 |
| AD Wightman Corrected | eGFR | | | rs12972970 | rs429358 | 0.00E+00 | 2.44E-12 | 0.00E+00 | 9.99E-01 | 1.16E-03 | 1 | 0 | 0 |
| AD Wightman | eGFR | | | rs429358 | rs8104483 | 0.00E+00 | 1.35E-02 | 0.00E+00 | 9.86E-01 | 1.28E-04 | 1 | 0 | 0 |

| | | | coloc SUSIE method | | | | | | | | | | |
|---|---|---|---|---|---|---|---|---|---|---|---|---|---|
| **Trait1** | **Trait 2** | **cFDRlocusnum** | **nsnps 2** | **hit1** | **hit2** | **PP.H0.abf** | **PP.H1.abf** | **PP.H2.abf** | **PP.H3.abf** | **PP.H4.abf** | **PP.H3+PP.H4≥0.8** | **PP.H3/PP.H4≥3** | **PP.H4/PP.H3≥5** |
| Corrected | | | | | | | | | | | | | |
| AD Wightman Corrected | eGFR | | | rs6857 | rs8104483 | 0.00E+00 | 1.35E-02 | 0.00E+00 | 9.86E-01 | 1.49E-04 | 1 | 0 | 0 |
| AD Wightman Corrected | eGFR | | | rs2075650 | rs8104483 | 0.00E+00 | 1.35E-02 | 0.00E+00 | 9.86E-01 | 1.28E-04 | 1 | 0 | 0 |
| AD Wightman Corrected | eGFR | | | rs71352238 | rs8104483 | 0.00E+00 | 1.35E-02 | 0.00E+00 | 9.86E-01 | 1.42E-04 | 1 | 0 | 0 |
| AD Wightman Corrected | eGFR | | | rs5117 | rs8104483 | 0.00E+00 | 1.35E-02 | 0.00E+00 | 9.86E-01 | 8.99E-04 | 1 | 0 | 0 |
| AD Wightman Corrected | eGFR | | | rs77301115 | rs8104483 | 0.00E+00 | 1.35E-02 | 0.00E+00 | 9.86E-01 | 1.92E-04 | 1 | 0 | 0 |
| AD Wightman Corrected | eGFR | | | rs75627662 | rs8104483 | 0.00E+00 | 1.35E-02 | 0.00E+00 | 9.85E-01 | 1.98E-03 | 1 | 0 | 0 |
| AD Wightman Corrected | eGFR | | | rs12972970 | rs8104483 | 0.00E+00 | 1.35E-02 | 0.00E+00 | 9.86E-01 | 1.29E-04 | 1 | 0 | 0 |
| AD Wightman Corrected | eGFR | 43c | 2881 | rs429358 | rs429358 | 0.00E+00 | 4.99E-15 | 0.00E+00 | 5.66E-05 | 1.00E+00 | 1 | 1 | 1 |
| AD Wightman Corrected | eGFR | | | rs6857 | rs429358 | 0.00E+00 | 2.34E-13 | 0.00E+00 | 9.45E-02 | 9.05E-01 | 1 | 1 | 1 |
| AD Wightma | eGFR | | | rs2075650 | rs429358 | 0.00E+00 | 2.42E-12 | 0.00E+00 | 9.98E-01 | 1.76E-03 | 1 | 0 | 0 |

| | | | coloc SUSIE method | | | | | | | | | | |
|---|---|---|---|---|---|---|---|---|---|---|---|---|---|
| Trait1 | Trait 2 | cFDRlocusnum | nsnps 2 | hit1 | hit2 | PP.H0.abf | PP.H1.abf | PP.H2.abf | PP.H3.abf | PP.H4.abf | PP.H3+PP.H4≥0.8 | PP.H3/PP.H4≥3 | PP.H4/PP.H3≥5 |
| n Corrected | | | | | | | | | | | | | |
| AD Wightman Corrected | eGFR | | | rs71352238 | rs429358 | 0.00E+00 | 2.41E-12 | 0.00E+00 | 9.92E-01 | 8.23E-03 | 1 | 0 | 0 |
| AD Wightman Corrected | eGFR | | | rs5117 | rs429358 | 0.00E+00 | 2.43E-12 | 0.00E+00 | 1.00E+00 | 2.22E-09 | 1 | 0 | 0 |
| AD Wightman Corrected | eGFR | | | rs77301115 | rs429358 | 0.00E+00 | 2.43E-12 | 0.00E+00 | 1.00E+00 | 5.37E-12 | 1 | 0 | 0 |
| AD Wightman Corrected | eGFR | | | rs75627662 | rs429358 | 0.00E+00 | 2.43E-12 | 0.00E+00 | 1.00E+00 | 1.83E-11 | 1 | 0 | 0 |
| AD Wightman Corrected | eGFR | | | rs12972970 | rs429358 | 0.00E+00 | 2.43E-12 | 0.00E+00 | 9.99E-01 | 1.15E-03 | 1 | 0 | 0 |
| AD Wightman Corrected | eGFR | | | rs429358 | rs8104483 | 0.00E+00 | 1.35E-02 | 0.00E+00 | 9.86E-01 | 1.28E-04 | 1 | 0 | 0 |
| AD Wightman Corrected | eGFR | | | rs6857 | rs8104483 | 0.00E+00 | 1.35E-02 | 0.00E+00 | 9.86E-01 | 1.49E-04 | 1 | 0 | 0 |
| AD Wightman Corrected | eGFR | | | rs2075650 | rs8104483 | 0.00E+00 | 1.35E-02 | 0.00E+00 | 9.86E-01 | 1.28E-04 | 1 | 0 | 0 |
| AD Wightman Corrected | eGFR | | | rs71352238 | rs8104483 | 0.00E+00 | 1.35E-02 | 0.00E+00 | 9.86E-01 | 1.42E-04 | 1 | 0 | 0 |
| AD Wightma | eGFR | | | rs5117 | rs8104483 | 0.00E+00 | 1.35E-02 | 0.00E+00 | 9.86E-01 | 8.98E-04 | 1 | 0 | 0 |

| | | | coloc SUSIE method | | | | | | | | | | |
|---|---|---|---|---|---|---|---|---|---|---|---|---|---|
| **Trait1** | **Trait 2** | **cFDRlocusnum** | **nsnps 2** | **hit1** | **hit2** | **PP.H0.abf** | **PP.H1.abf** | **PP.H2.abf** | **PP.H3.abf** | **PP.H4.abf** | **PP.H3+PP.H4≥0.8** | **PP.H3/PP.H4≥3** | **PP.H4/PP.H3≥5** |
| n Corrected | | | | | | | | | | | | | |
| AD Wightman Corrected | eGFR | | | rs77301115 | rs8104483 | 0.00E+00 | 1.35E-02 | 0.00E+00 | 9.86E-01 | 1.93E-04 | 1 | 0 | 0 |
| AD Wightman Corrected | eGFR | | | rs75627662 | rs8104483 | 0.00E+00 | 1.35E-02 | 0.00E+00 | 9.85E-01 | 1.98E-03 | 1 | 0 | 0 |
| AD Wightman Corrected | eGFR | | | rs12972970 | rs8104483 | 0.00E+00 | 1.35E-02 | 0.00E+00 | 9.86E-01 | 1.29E-04 | 1 | 0 | 0 |
| AD Wightman Corrected | eGFR | 44a | 2859 | rs429358 | rs429358 | 0.00E+00 | 5.01E-15 | 0.00E+00 | 5.65E-05 | 1.00E+00 | 1 | 1 | 1 |
| AD Wightman Corrected | eGFR | | | rs6857 | rs429358 | 0.00E+00 | 2.34E-13 | 0.00E+00 | 9.43E-02 | 9.06E-01 | 1 | 1 | 1 |
| AD Wightman Corrected | eGFR | | | rs2075650 | rs429358 | 0.00E+00 | 2.43E-12 | 0.00E+00 | 9.98E-01 | 1.76E-03 | 1 | 0 | 0 |
| AD Wightman Corrected | eGFR | | | rs71352238 | rs429358 | 0.00E+00 | 2.41E-12 | 0.00E+00 | 9.92E-01 | 8.23E-03 | 1 | 0 | 0 |
| AD Wightman Corrected | eGFR | | | rs5117 | rs429358 | 0.00E+00 | 2.43E-12 | 0.00E+00 | 1.00E+00 | 2.30E-09 | 1 | 0 | 0 |
| AD Wightman Corrected | eGFR | | | rs77301115 | rs429358 | 0.00E+00 | 2.43E-12 | 0.00E+00 | 1.00E+00 | 5.40E-12 | 1 | 0 | 0 |
| AD | eGFR | | | rs75627662 | rs429358 | 0.00E+00 | 2.43E-12 | 0.00E+00 | 1.00E+00 | 1.88E-11 | 1 | 0 | 0 |

| | | | coloc SUSIE method | | | | | | | | | | |
|---|---|---|---|---|---|---|---|---|---|---|---|---|---|
| **Trait1** | **Trait 2** | **cFDRlocusnum** | **nsnps 2** | **hit1** | **hit2** | **PP.H0.abf** | **PP.H1.abf** | **PP.H2.abf** | **PP.H3.abf** | **PP.H4.abf** | **PP.H3+PP.H4≥0.8** | **PP.H3/PP.H4≥3** | **PP.H4/PP.H3≥5** |
| Wightman Corrected | | | | | | | | | | | | | |
| AD Wightman Corrected | eGFR | | | rs12972970 | rs429358 | 0.00E+00 | 2.43E-12 | 0.00E+00 | 9.99E-01 | 1.16E-03 | 1 | 0 | 0 |
| AD Wightman Corrected | eGFR | | | rs429358 | rs8104483 | 0.00E+00 | 1.35E-02 | 0.00E+00 | 9.86E-01 | 1.28E-04 | 1 | 0 | 0 |
| AD Wightman Corrected | eGFR | | | rs6857 | rs8104483 | 0.00E+00 | 1.35E-02 | 0.00E+00 | 9.86E-01 | 1.49E-04 | 1 | 0 | 0 |
| AD Wightman Corrected | eGFR | | | rs2075650 | rs8104483 | 0.00E+00 | 1.35E-02 | 0.00E+00 | 9.86E-01 | 1.28E-04 | 1 | 0 | 0 |
| AD Wightman Corrected | eGFR | | | rs71352238 | rs8104483 | 0.00E+00 | 1.35E-02 | 0.00E+00 | 9.86E-01 | 1.42E-04 | 1 | 0 | 0 |
| AD Wightman Corrected | eGFR | | | rs5117 | rs8104483 | 0.00E+00 | 1.35E-02 | 0.00E+00 | 9.86E-01 | 8.85E-04 | 1 | 0 | 0 |
| AD Wightman Corrected | eGFR | | | rs77301115 | rs8104483 | 0.00E+00 | 1.35E-02 | 0.00E+00 | 9.86E-01 | 1.92E-04 | 1 | 0 | 0 |
| AD Wightman Corrected | eGFR | | | rs75627662 | rs8104483 | 0.00E+00 | 1.35E-02 | 0.00E+00 | 9.85E-01 | 1.95E-03 | 1 | 0 | 0 |
| AD Wightman Corrected | eGFR | | | rs12972970 | rs8104483 | 0.00E+00 | 1.35E-02 | 0.00E+00 | 9.86E-01 | 1.29E-04 | 1 | 0 | 0 |

| | | | coloc SUSIE method | | | | | | | | | | |
|---|---|---|---|---|---|---|---|---|---|---|---|---|---|
| Trait1 | Trait 2 | cFDRlocusnum | nsnps 2 | hit1 | hit2 | PP.H0.abf | PP.H1.abf | PP.H2.abf | PP.H3.abf | PP.H4.abf | PP.H3+PP.H4≥0.8 | PP.H3/PP.H4≥3 | PP.H4/PP.H3≥5 |
| AD Wightman Corrected | eGFR | 44b | 2815 | rs429358 | rs429358 | 0.00E+00 | 4.98E-15 | 0.00E+00 | 5.62E-05 | 1.00E+00 | 1 | 1 | 1 |
| AD Wightman Corrected | eGFR | | | rs6857 | rs429358 | 0.00E+00 | 2.33E-13 | 0.00E+00 | 9.46E-02 | 9.05E-01 | 1 | 1 | 1 |
| AD Wightman Corrected | eGFR | | | rs2075650 | rs429358 | 0.00E+00 | 2.42E-12 | 0.00E+00 | 9.98E-01 | 1.76E-03 | 1 | 0 | 0 |
| AD Wightman Corrected | eGFR | | | rs71352238 | rs429358 | 0.00E+00 | 2.40E-12 | 0.00E+00 | 9.92E-01 | 8.22E-03 | 1 | 0 | 0 |
| AD Wightman Corrected | eGFR | | | rs5117 | rs429358 | 0.00E+00 | 2.42E-12 | 0.00E+00 | 1.00E+00 | 2.34E-09 | 1 | 0 | 0 |
| AD Wightman Corrected | eGFR | | | rs77301115 | rs429358 | 0.00E+00 | 2.42E-12 | 0.00E+00 | 1.00E+00 | 5.36E-12 | 1 | 0 | 0 |
| AD Wightman Corrected | eGFR | | | rs75627662 | rs429358 | 0.00E+00 | 2.42E-12 | 0.00E+00 | 1.00E+00 | 1.90E-11 | 1 | 0 | 0 |
| AD Wightman Corrected | eGFR | | | rs12972970 | rs429358 | 0.00E+00 | 2.42E-12 | 0.00E+00 | 9.99E-01 | 1.16E-03 | 1 | 0 | 0 |
| AD Wightman Corrected | eGFR | | | rs429358 | rs8104483 | 0.00E+00 | 1.35E-02 | 0.00E+00 | 9.86E-01 | 1.28E-04 | 1 | 0 | 0 |
| AD Wightman Corrected | eGFR | | | rs6857 | rs8104483 | 0.00E+00 | 1.35E-02 | 0.00E+00 | 9.86E-01 | 1.49E-04 | 1 | 0 | 0 |

| | | | coloc SUSIE method | | | | | | | | | | |
|---|---|---|---|---|---|---|---|---|---|---|---|---|---|
| **Trait1** | **Trait 2** | **cFDRlocusnum** | **nsnps 2** | **hit1** | **hit2** | **PP.H0.abf** | **PP.H1.abf** | **PP.H2.abf** | **PP.H3.abf** | **PP.H4.abf** | **PP.H3+PP.H4≥0.8** | **PP.H3/PP.H4≥3** | **PP.H4/PP.H3≥5** |
| AD Wightman Corrected | eGFR | | | rs2075650 | rs8104483 | 0.00E+00 | 1.35E-02 | 0.00E+00 | 9.86E-01 | 1.28E-04 | 1 | 0 | 0 |
| AD Wightman Corrected | eGFR | | | rs71352238 | rs8104483 | 0.00E+00 | 1.35E-02 | 0.00E+00 | 9.86E-01 | 1.42E-04 | 1 | 0 | 0 |
| AD Wightman Corrected | eGFR | | | rs5117 | rs8104483 | 0.00E+00 | 1.35E-02 | 0.00E+00 | 9.86E-01 | 8.82E-04 | 1 | 0 | 0 |
| AD Wightman Corrected | eGFR | | | rs77301115 | rs8104483 | 0.00E+00 | 1.35E-02 | 0.00E+00 | 9.86E-01 | 1.93E-04 | 1 | 0 | 0 |
| AD Wightman Corrected | eGFR | | | rs75627662 | rs8104483 | 0.00E+00 | 1.35E-02 | 0.00E+00 | 9.85E-01 | 1.94E-03 | 1 | 0 | 0 |
| AD Wightman Corrected | eGFR | | | rs12972970 | rs8104483 | 0.00E+00 | 1.35E-02 | 0.00E+00 | 9.86E-01 | 1.29E-04 | 1 | 0 | 0 |
| AD Wightman Corrected | eGFR | 44c | 2805 | rs429358 | rs429358 | 0.00E+00 | 4.99E-15 | 0.00E+00 | 5.62E-05 | 1.00E+00 | 1 | 1 | 1 |
| AD Wightman Corrected | eGFR | | | rs6857 | rs429358 | 0.00E+00 | 2.34E-13 | 0.00E+00 | 9.44E-02 | 9.06E-01 | 1 | 1 | 1 |
| AD Wightman Corrected | eGFR | | | rs2075650 | rs429358 | 0.00E+00 | 2.42E-12 | 0.00E+00 | 9.98E-01 | 1.76E-03 | 1 | 0 | 0 |
| AD Wightman | eGFR | | | rs71352238 | rs429358 | 0.00E+00 | 2.41E-12 | 0.00E+00 | 9.92E-01 | 8.23E-03 | 1 | 0 | 0 |

| | | | coloc SUSIE method | | | | | | | | | | |
|---|---|---|---|---|---|---|---|---|---|---|---|---|---|
| **Trait1** | **Trait 2** | **cFDRlocusnum** | **nsnps 2** | **hit1** | **hit2** | **PP.H0.abf** | **PP.H1.abf** | **PP.H2.abf** | **PP.H3.abf** | **PP.H4.abf** | **PP.H3+PP.H4≥0.8** | **PP.H3/PP.H4≥3** | **PP.H4/PP.H3≥5** |
| Corrected | | | | | | | | | | | | | |
| AD Wightman Corrected | eGFR | | | rs5117 | rs429358 | 0.00E+00 | 2.43E-12 | 0.00E+00 | 1.00E+00 | 2.35E-09 | 1 | 0 | 0 |
| AD Wightman Corrected | eGFR | | | rs77301115 | rs429358 | 0.00E+00 | 2.43E-12 | 0.00E+00 | 1.00E+00 | 5.38E-12 | 1 | 0 | 0 |
| AD Wightman Corrected | eGFR | | | rs75627662 | rs429358 | 0.00E+00 | 2.43E-12 | 0.00E+00 | 1.00E+00 | 1.91E-11 | 1 | 0 | 0 |
| AD Wightman Corrected | eGFR | | | rs12972970 | rs429358 | 0.00E+00 | 2.43E-12 | 0.00E+00 | 9.99E-01 | 1.16E-03 | 1 | 0 | 0 |
| AD Wightman Corrected | eGFR | | | rs429358 | rs8104483 | 0.00E+00 | 1.35E-02 | 0.00E+00 | 9.86E-01 | 1.28E-04 | 1 | 0 | 0 |
| AD Wightman Corrected | eGFR | | | rs6857 | rs8104483 | 0.00E+00 | 1.35E-02 | 0.00E+00 | 9.86E-01 | 1.49E-04 | 1 | 0 | 0 |
| AD Wightman Corrected | eGFR | | | rs2075650 | rs8104483 | 0.00E+00 | 1.35E-02 | 0.00E+00 | 9.86E-01 | 1.28E-04 | 1 | 0 | 0 |
| AD Wightman Corrected | eGFR | | | rs71352238 | rs8104483 | 0.00E+00 | 1.35E-02 | 0.00E+00 | 9.86E-01 | 1.42E-04 | 1 | 0 | 0 |
| AD Wightman Corrected | eGFR | | | rs5117 | rs8104483 | 0.00E+00 | 1.35E-02 | 0.00E+00 | 9.86E-01 | 8.81E-04 | 1 | 0 | 0 |
| AD Wightman | eGFR | | | rs77301115 | rs8104483 | 0.00E+00 | 1.35E-02 | 0.00E+00 | 9.86E-01 | 1.93E-04 | 1 | 0 | 0 |

| | | | coloc SUSIE method | | | | | | | | | | |
|---|---|---|---|---|---|---|---|---|---|---|---|---|---|
| **Trait1** | **Trait 2** | **cFDRlocusnum** | **nsnps 2** | **hit1** | **hit2** | **PP.H0.abf** | **PP.H1.abf** | **PP.H2.abf** | **PP.H3.abf** | **PP.H4.abf** | **PP.H3+PP.H4≥0.8** | **PP.H3/PP.H4≥3** | **PP.H4/PP.H3≥5** |
| Corrected | | | | | | | | | | | | | |
| AD Wightman Corrected | eGFR | | | rs75627662 | rs8104483 | 0.00E+00 | 1.35E-02 | 0.00E+00 | 9.85E-01 | 1.93E-03 | 1 | 0 | 0 |
| AD Wightman Corrected | eGFR | | | rs12972970 | rs8104483 | 0.00E+00 | 1.35E-02 | 0.00E+00 | 9.86E-01 | 1.29E-04 | 1 | 0 | 0 |
| AD Wightman Corrected | eGFR | 44d | 2798 | rs429358 | rs429358 | 0.00E+00 | 5.00E-15 | 0.00E+00 | 5.63E-05 | 1.00E+00 | 1 | 1 | 1 |
| AD Wightman Corrected | eGFR | | | rs6857 | rs429358 | 0.00E+00 | 2.34E-13 | 0.00E+00 | 9.44E-02 | 9.06E-01 | 1 | 1 | 1 |
| AD Wightman Corrected | eGFR | | | rs2075650 | rs429358 | 0.00E+00 | 2.43E-12 | 0.00E+00 | 9.98E-01 | 1.77E-03 | 1 | 0 | 0 |
| AD Wightman Corrected | eGFR | | | rs71352238 | rs429358 | 0.00E+00 | 2.41E-12 | 0.00E+00 | 9.92E-01 | 8.23E-03 | 1 | 0 | 0 |
| AD Wightman Corrected | eGFR | | | rs5117 | rs429358 | 0.00E+00 | 2.43E-12 | 0.00E+00 | 1.00E+00 | 2.35E-09 | 1 | 0 | 0 |
| AD Wightman Corrected | eGFR | | | rs77301115 | rs429358 | 0.00E+00 | 2.43E-12 | 0.00E+00 | 1.00E+00 | 5.38E-12 | 1 | 0 | 0 |
| AD Wightman Corrected | eGFR | | | rs75627662 | rs429358 | 0.00E+00 | 2.43E-12 | 0.00E+00 | 1.00E+00 | 1.91E-11 | 1 | 0 | 0 |
| AD Wightma | eGFR | | | rs12972970 | rs429358 | 0.00E+00 | 2.43E-12 | 0.00E+00 | 9.99E-01 | 1.16E-03 | 1 | 0 | 0 |

| | | | coloc SUSIE method | | | | | | | | | | |
|---|---|---|---|---|---|---|---|---|---|---|---|---|---|
| **Trait1** | **Trait 2** | **cFDRlocusnum** | **nsnps 2** | **hit1** | **hit2** | **PP.H0.abf** | **PP.H1.abf** | **PP.H2.abf** | **PP.H3.abf** | **PP.H4.abf** | **PP.H3+PP.H4≥0.8** | **PP.H3/PP.H4≥3** | **PP.H4/PP.H3≥5** |
| n Corrected | | | | | | | | | | | | | |
| AD Wightman Corrected | eGFR | | | rs429358 | rs8104483 | 0.00E+00 | 1.36E-02 | 0.00E+00 | 9.86E-01 | 1.28E-04 | 1 | 0 | 0 |
| AD Wightman Corrected | eGFR | | | rs6857 | rs8104483 | 0.00E+00 | 1.36E-02 | 0.00E+00 | 9.86E-01 | 1.49E-04 | 1 | 0 | 0 |
| AD Wightman Corrected | eGFR | | | rs2075650 | rs8104483 | 0.00E+00 | 1.36E-02 | 0.00E+00 | 9.86E-01 | 1.28E-04 | 1 | 0 | 0 |
| AD Wightman Corrected | eGFR | | | rs71352238 | rs8104483 | 0.00E+00 | 1.36E-02 | 0.00E+00 | 9.86E-01 | 1.42E-04 | 1 | 0 | 0 |
| AD Wightman Corrected | eGFR | | | rs5117 | rs8104483 | 0.00E+00 | 1.35E-02 | 0.00E+00 | 9.86E-01 | 8.83E-04 | 1 | 0 | 0 |
| AD Wightman Corrected | eGFR | | | rs77301115 | rs8104483 | 0.00E+00 | 1.35E-02 | 0.00E+00 | 9.86E-01 | 1.93E-04 | 1 | 0 | 0 |
| AD Wightman Corrected | eGFR | | | rs75627662 | rs8104483 | 0.00E+00 | 1.35E-02 | 0.00E+00 | 9.85E-01 | 1.94E-03 | 1 | 0 | 0 |
| AD Wightman Corrected | eGFR | | | rs12972970 | rs8104483 | 0.00E+00 | 1.36E-02 | 0.00E+00 | 9.86E-01 | 1.29E-04 | 1 | 0 | 0 |
| AD Wightman Corrected | eGFR | 47 | 2674 | rs346758 | rs1800437 | 2.60E-21 | 4.83E-06 | 5.37E-16 | 1.00E+00 | 8.78E-08 | 1 | 0 | 0 |
| AD | eGFR | | | rs11695875 | rs1800437 | 1.57E-18 | 4.83E-06 | 3.25E-13 | 1.00E+00 | 4.93E-08 | 1 | 0 | 0 |

| | | | coloc SUSIE method | | | | | | | | | | |
|---|---|---|---|---|---|---|---|---|---|---|---|---|---|
| **Trait1** | **Trait 2** | **cFDRlocusnum** | **nsnps 2** | **hit1** | **hit2** | **PP.H0.abf** | **PP.H1.abf** | **PP.H2.abf** | **PP.H3.abf** | **PP.H4.abf** | **PP.H3+PP.H4≥0.8** | **PP.H3/PP.H4≥3** | **PP.H4/PP.H3≥5** |
| Wightman Corrected | | | | 5 | | | | | | | | | |
| AD Wightman Corrected | eGFR | | | rs344819 | rs1800437 | 1.92E-13 | 4.83E-06 | 3.98E-08 | 1.00E+00 | 4.93E-08 | 1 | 0 | 0 |
| AD Wightman Corrected | eGFR | | | rs111435864 | rs1800437 | 6.07E-12 | 4.83E-06 | 1.25E-06 | 1.00E+00 | 1.78E-06 | 1 | 0 | 0 |
| AD Wightman Corrected | eGFR | | | rs2072561 | rs1800437 | 6.44E-08 | 4.77E-06 | 1.33E-02 | 9.87E-01 | 1.25E-04 | 1 | 0 | 0 |
| AD Wightman Corrected | eGFR | | | rs8113514 | rs1800437 | 7.10E-09 | 4.83E-06 | 1.47E-03 | 9.99E-01 | 1.25E-05 | 1 | 0 | 0 |
| AD Wightman Corrected | eGFR | | | rs8112710 | rs1800437 | 3.18E-16 | 4.83E-06 | 6.57E-11 | 1.00E+00 | 3.76E-08 | 1 | 0 | 0 |
| AD Wightman Corrected | eGFR | | | rs12611268 | rs1800437 | 2.65E-09 | 4.83E-06 | 5.49E-04 | 9.99E-01 | 5.11E-06 | 1 | 0 | 0 |
| AD Wightman Corrected | eGFR | | | rs10404220 | rs1800437 | 9.82E-08 | 4.73E-06 | 2.03E-02 | 9.79E-01 | 2.22E-04 | 1 | 0 | 0 |

**Supplementary Table S27. Colocalization Analysis Results Using SuSiE Framework (Wightman Uncorrected).**
This table presents colocalization results using the Wightman AD GWAS without competing risk correction. The analysis provides a comprehensive assessment of how competing risk adjustment influences fine-mapping and colocalization conclusions in the validation dataset. Note: 0 indicates "No", and 1 indicates "Yes".

| | | | coloc SUSIE method | | | | | | | | | | |
|---|---|---|---|---|---|---|---|---|---|---|---|---|---|
| **Trait1** | **Trait 2** | **cFDRlocusnum** | **nsnps 2** | **hit1** | **hit2** | **PP.H0.abf** | **PP.H1.abf** | **PP.H2.abf** | **PP.H3.abf** | **PP.H4.abf** | **PP.H3+PP.H4≥0.8** | **PP.H3/PP.H4≥3** | **PP.H4/PP.H3≥5** |
| AD Wightman | eGFR | 1 | 2231 | rs9442367 | rs2710889 | 0.00E+00 | 2.42E-02 | 0.00E+00 | 9.37E-01 | 3.93E-02 | 1 | 0 | 0 |
| AD Wightman | eGFR | | | rs2298215 | rs2710889 | 0.00E+00 | 2.39E-02 | 0.00E+00 | 9.26E-01 | 5.02E-02 | 1 | 0 | 0 |
| AD Wightman | eGFR | | | rs1133647 | rs2710889 | 0.00E+00 | 2.39E-02 | 0.00E+00 | 9.23E-01 | 5.27E-02 | 1 | 0 | 0 |
| AD Wightman | eGFR | | | rs4246502 | rs2710889 | 0.00E+00 | 2.33E-02 | 0.00E+00 | 9.01E-01 | 7.62E-02 | 1 | 0 | 0 |
| AD Wightman | eGFR | | | rs4970401 | rs2710889 | 0.00E+00 | 2.33E-02 | 0.00E+00 | 9.03E-01 | 7.38E-02 | 1 | 0 | 0 |
| AD Wightman | eGFR | | | rs4075116 | rs2710889 | 0.00E+00 | 2.33E-02 | 0.00E+00 | 9.02E-01 | 7.50E-02 | 1 | 0 | 0 |
| AD Wightman | eGFR | 3 | 3261 | rs11806994 | rs3850625 | 0.00E+00 | 3.00E-43 | 0.00E+00 | 1.00E+00 | 1.13E-45 | 1 | 0 | 0 |
| AD Wightman | eGFR | | | rs6656481 | rs3850625 | 0.00E+00 | 3.00E-43 | 0.00E+00 | 1.00E+00 | 1.13E-44 | 1 | 0 | 0 |
| AD Wightman | eGFR | | | rs6692427 | rs3850625 | 0.00E+00 | 3.00E-43 | 0.00E+00 | 1.00E+00 | 4.40E-45 | 1 | 0 | 0 |
| AD Wightman | eGFR | | | rs12143520 | rs3850625 | 0.00E+00 | 3.00E-43 | 0.00E+00 | 1.00E+00 | 1.06E-45 | 1 | 0 | 0 |
| AD Wightma | eGFR | | | rs2297912 | rs3850625 | 0.00E+00 | 3.00E-43 | 0.00E+00 | 1.00E+00 | 1.19E-45 | 1 | 0 | 0 |

| | | | coloc SUSIE method | | | | | | | | | | |
|---|---|---|---|---|---|---|---|---|---|---|---|---|---|
| **Trait1** | **Trait 2** | **cFDRlocusnum** | **nsnps 2** | **hit1** | **hit2** | **PP.H0.abf** | **PP.H1.abf** | **PP.H2.abf** | **PP.H3.abf** | **PP.H4.abf** | **PP.H3+PP.H4≥0.8** | **PP.H3/PP.H4≥3** | **PP.H4/PP.H3≥5** |
| n | | | | | | | | | | | | | |
| AD Wightman | eGFR | | | rs57758014 | rs3850625 | 0.00E+00 | 3.00E-43 | 0.00E+00 | 1.00E+00 | 1.05E-45 | 1 | 0 | 0 |
| AD Wightman | eGFR | | | rs925059 | rs3850625 | 0.00E+00 | 3.00E-43 | 0.00E+00 | 1.00E+00 | 1.12E-45 | 1 | 0 | 0 |
| AD Wightman | eGFR | | | rs72749145 | rs3850625 | 0.00E+00 | 3.00E-43 | 0.00E+00 | 1.00E+00 | 1.07E-45 | 1 | 0 | 0 |
| AD Wightman | eGFR | | | rs2275485 | rs3850625 | 0.00E+00 | 3.00E-43 | 0.00E+00 | 1.00E+00 | 1.05E-45 | 1 | 0 | 0 |
| AD Wightman | eGFR | | | rs11806994 | rs12038184 | 0.00E+00 | 1.46E-122 | 0.00E+00 | 1.00E+00 | 3.85E-115 | 1 | 0 | 0 |
| AD Wightman | eGFR | | | rs6656481 | rs12038184 | 0.00E+00 | 1.46E-122 | 0.00E+00 | 1.00E+00 | 9.20E-117 | 1 | 0 | 0 |
| AD Wightman | eGFR | | | rs6692427 | rs12038184 | 0.00E+00 | 1.46E-122 | 0.00E+00 | 1.00E+00 | 2.46E-117 | 1 | 0 | 0 |
| AD Wightman | eGFR | | | rs12143520 | rs12038184 | 0.00E+00 | 1.46E-122 | 0.00E+00 | 1.00E+00 | 1.34E-115 | 1 | 0 | 0 |
| AD Wightman | eGFR | | | rs2297912 | rs12038184 | 0.00E+00 | 1.46E-122 | 0.00E+00 | 1.00E+00 | 3.48E-116 | 1 | 0 | 0 |
| AD Wightman | eGFR | | | rs57758014 | rs12038184 | 0.00E+00 | 1.46E-122 | 0.00E+00 | 1.00E+00 | 1.09E-115 | 1 | 0 | 0 |
| AD Wightman | eGFR | | | rs925059 | rs12038184 | 0.00E+00 | 1.46E-122 | 0.00E+00 | 1.00E+00 | 4.85E-115 | 1 | 0 | 0 |
| AD Wightman | eGFR | | | rs72749145 | rs12038184 | 0.00E+00 | 1.46E-122 | 0.00E+00 | 1.00E+00 | 1.54E-115 | 1 | 0 | 0 |
| AD Wightman | eGFR | | | rs2275485 | rs12038184 | 0.00E+00 | 1.46E-122 | 0.00E+00 | 1.00E+00 | 9.76E-116 | 1 | 0 | 0 |

| | | | coloc SUSIE method | | | | | | | | | | |
|---|---|---|---|---|---|---|---|---|---|---|---|---|---|
| Trait1 | Trait 2 | cFDRlocusnum | nsnps 2 | hit1 | hit2 | PP.H0.abf | PP.H1.abf | PP.H2.abf | PP.H3.abf | PP.H4.abf | PP.H3+PP.H4≥0.8 | PP.H3/PP.H4≥3 | PP.H4/PP.H3≥5 |
| AD Wightman | eGFR | | | rs11806994 | rs4915468 | 0.00E+00 | 2.69E-84 | 0.00E+00 | 1.00E+00 | 4.02E-82 | 1 | 0 | 0 |
| AD Wightman | eGFR | | | rs6656481 | rs4915468 | 0.00E+00 | 2.69E-84 | 0.00E+00 | 1.00E+00 | 2.25E-81 | 1 | 0 | 0 |
| AD Wightman | eGFR | | | rs6692427 | rs4915468 | 0.00E+00 | 2.69E-84 | 0.00E+00 | 1.00E+00 | 5.90E-83 | 1 | 0 | 0 |
| AD Wightman | eGFR | | | rs12143520 | rs4915468 | 0.00E+00 | 2.69E-84 | 0.00E+00 | 1.00E+00 | 8.49E-82 | 1 | 0 | 0 |
| AD Wightman | eGFR | | | rs2297912 | rs4915468 | 0.00E+00 | 2.69E-84 | 0.00E+00 | 1.00E+00 | 2.60E-79 | 1 | 0 | 0 |
| AD Wightman | eGFR | | | rs57758014 | rs4915468 | 0.00E+00 | 2.69E-84 | 0.00E+00 | 1.00E+00 | 9.51E-82 | 1 | 0 | 0 |
| AD Wightman | eGFR | | | rs925059 | rs4915468 | 0.00E+00 | 2.69E-84 | 0.00E+00 | 1.00E+00 | 5.76E-82 | 1 | 0 | 0 |
| AD Wightman | eGFR | | | rs72749145 | rs4915468 | 0.00E+00 | 2.69E-84 | 0.00E+00 | 1.00E+00 | 7.68E-82 | 1 | 0 | 0 |
| AD Wightman | eGFR | | | rs2275485 | rs4915468 | 0.00E+00 | 2.69E-84 | 0.00E+00 | 1.00E+00 | 1.05E-81 | 1 | 0 | 0 |
| AD Wightman | eGFR | 6 | 2465 | rs6440796 | rs4365603 | 0.00E+00 | 1.56E-03 | 0.00E+00 | 1.31E-01 | 8.67E-01 | 1 | 1 | 1 |
| AD Wightman | eGFR | | | rs12486405 | rs4365603 | 0.00E+00 | 6.26E-04 | 0.00E+00 | 5.15E-02 | 9.48E-01 | 1 | 1 | 1 |
| AD Wightman | eGFR | | | rs12492498 | rs4365603 | 0.00E+00 | 1.62E-03 | 0.00E+00 | 1.36E-01 | 8.62E-01 | 1 | 1 | 1 |
| AD Wightman | eGFR | | | rs1426384 | rs4365603 | 0.00E+00 | 7.53E-04 | 0.00E+00 | 6.23E-02 | 9.37E-01 | 1 | 1 | 1 |

| | | | coloc SUSIE method | | | | | | | | | | |
|---|---|---|---|---|---|---|---|---|---|---|---|---|---|
| **Trait1** | **Trait2** | **cFDRlocusnum** | **nsnps2** | **hit1** | **hit2** | **PP.H0.abf** | **PP.H1.abf** | **PP.H2.abf** | **PP.H3.abf** | **PP.H4.abf** | **PP.H3+PP.H4≥0.8** | **PP.H3/PP.H4≥3** | **PP.H4/PP.H3≥5** |
| AD Wightman | eGFR | | | rs12488207 | rs4365603 | 0.00E+00 | 5.36E-04 | 0.00E+00 | 4.38E-02 | 9.56E-01 | 1 | 1 | 1 |
| AD Wightman | eGFR | | | rs35650976 | rs4365603 | 0.00E+00 | 5.98E-04 | 0.00E+00 | 4.90E-02 | 9.50E-01 | 1 | 1 | 1 |
| AD Wightman | eGFR | | | rs13059175 | rs4365603 | 0.00E+00 | 9.41E-04 | 0.00E+00 | 7.83E-02 | 9.21E-01 | 1 | 1 | 1 |
| AD Wightman | eGFR | | | | | | | | | | | | |
| AD Wightman | eGFR | 8 | 2624 | rs3800456 | rs141704702 | 0.00E+00 | 0.00E+00 | 0.00E+00 | 1.00E+00 | 0.00E+00 | 1 | 0 | 0 |
| AD Wightman | eGFR | | | rs4713823 | rs141704702 | 0.00E+00 | 0.00E+00 | 0.00E+00 | 1.00E+00 | 0.00E+00 | 1 | 0 | 0 |
| AD Wightman | eGFR | | | rs2814997 | rs141704702 | 0.00E+00 | 0.00E+00 | 0.00E+00 | 1.00E+00 | 0.00E+00 | 1 | 0 | 0 |
| AD Wightman | eGFR | | | rs2252551 | rs141704702 | 0.00E+00 | 0.00E+00 | 0.00E+00 | 1.00E+00 | 0.00E+00 | 1 | 0 | 0 |
| AD Wightman | eGFR | | | rs3800456 | rs569521865 | 0.00E+00 | 0.00E+00 | 0.00E+00 | 1.00E+00 | 0.00E+00 | 1 | 0 | 0 |
| AD Wightman | eGFR | | | rs4713823 | rs569521865 | 0.00E+00 | 0.00E+00 | 0.00E+00 | 1.00E+00 | 0.00E+00 | 1 | 0 | 0 |
| AD Wightman | eGFR | | | rs2814997 | rs569521865 | 0.00E+00 | 0.00E+00 | 0.00E+00 | 1.00E+00 | 0.00E+00 | 1 | 0 | 0 |
| AD Wightman | eGFR | | | rs2252551 | rs569521865 | 0.00E+00 | 0.00E+00 | 0.00E+00 | 1.00E+00 | 0.00E+00 | 1 | 0 | 0 |
| AD Wightma | eGFR | 13 | 1849 | rs57957974 | rs11786896 | 0.00E+00 | 7.59E-15 | 0.00E+00 | 1.00E+00 | 8.41E-16 | 1 | 0 | 0 |

| | | | coloc SUSIE method | | | | | | | | | | |
|---|---|---|---|---|---|---|---|---|---|---|---|---|---|
| **Trait1** | **Trait 2** | **cFDRlocusnum** | **nsnps 2** | **hit1** | **hit2** | **PP.H0.abf** | **PP.H1.abf** | **PP.H2.abf** | **PP.H3.abf** | **PP.H4.abf** | **PP.H3+PP.H4≥0.8** | **PP.H3/PP.H4≥3** | **PP.H4/PP.H3≥5** |
| n | | | | | | | | | | | | | |
| AD Wightman | eGFR | | | rs62530297 | rs11786896 | 0.00E+00 | 7.59E-15 | 0.00E+00 | 1.00E+00 | 7.95E-16 | 1 | 0 | 0 |
| AD Wightman | eGFR | | | rs57957974 | rs6985555 | 0.00E+00 | 2.90E-05 | 0.00E+00 | 1.00E+00 | 2.63E-07 | 1 | 0 | 0 |
| AD Wightman | eGFR | | | rs62530297 | rs6985555 | 0.00E+00 | 2.90E-05 | 0.00E+00 | 1.00E+00 | 2.66E-07 | 1 | 0 | 0 |
| AD Wightman | eGFR | 15 | 4000 | rs7896832 | rs143396142 | 0.00E+00 | 8.21E-04 | 0.00E+00 | 9.99E-01 | 1.57E-05 | 1 | 0 | 0 |
| AD Wightman | eGFR | | | rs7910643 | rs143396142 | 0.00E+00 | 8.21E-04 | 0.00E+00 | 9.99E-01 | 1.57E-05 | 1 | 0 | 0 |
| AD Wightman | eGFR | | | rs7097656 | rs143396142 | 0.00E+00 | 8.21E-04 | 0.00E+00 | 9.99E-01 | 1.36E-05 | 1 | 0 | 0 |
| AD Wightman | eGFR | | | rs7896832 | rs10788630 | 0.00E+00 | 8.07E-11 | 0.00E+00 | 1.00E+00 | 1.30E-06 | 1 | 0 | 0 |
| AD Wightman | eGFR | | | rs7910643 | rs10788630 | 0.00E+00 | 8.07E-11 | 0.00E+00 | 1.00E+00 | 1.31E-06 | 1 | 0 | 0 |
| AD Wightman | eGFR | | | rs7097656 | rs10788630 | 0.00E+00 | 8.07E-11 | 0.00E+00 | 1.00E+00 | 6.94E-07 | 1 | 0 | 0 |
| AD Wightman | eGFR | 16 | 3788 | rs7896832 | rs143396142 | 0.00E+00 | 8.31E-04 | 0.00E+00 | 9.99E-01 | 1.63E-05 | 1 | 0 | 0 |
| AD Wightman | eGFR | | | rs7910643 | rs143396142 | 0.00E+00 | 8.31E-04 | 0.00E+00 | 9.99E-01 | 1.63E-05 | 1 | 0 | 0 |
| AD Wightman | eGFR | | | rs7097656 | rs143396142 | 0.00E+00 | 8.31E-04 | 0.00E+00 | 9.99E-01 | 1.41E-05 | 1 | 0 | 0 |

| | | | coloc SUSIE method | | | | | | | | | | |
|---|---|---|---|---|---|---|---|---|---|---|---|---|---|
| **Trait1** | **Trait 2** | **cFDRlocusnum** | **nsnps 2** | **hit1** | **hit2** | **PP.H0.abf** | **PP.H1.abf** | **PP.H2.abf** | **PP.H3.abf** | **PP.H4.abf** | **PP.H3+PP.H4≥0.8** | **PP.H3/PP.H4≥3** | **PP.H4/PP.H3≥5** |
| AD Wightman | eGFR | | | rs7896832 | rs10788630 | 0.00E+00 | 7.18E-11 | 0.00E+00 | 1.00E+00 | 1.33E-06 | 1 | 0 | 0 |
| AD Wightman | eGFR | | | rs7910643 | rs10788630 | 0.00E+00 | 7.18E-11 | 0.00E+00 | 1.00E+00 | 1.34E-06 | 1 | 0 | 0 |
| AD Wightman | eGFR | | | rs7097656 | rs10788630 | 0.00E+00 | 7.18E-11 | 0.00E+00 | 1.00E+00 | 7.09E-07 | 1 | 0 | 0 |
| AD Wightman | eGFR | | | | | | | | | | | | |
| AD Wightman | eGFR | 17 | 2411 | rs7081975 | rs6585310 | 0.00E+00 | 6.59E-03 | 0.00E+00 | 9.80E-01 | 1.33E-02 | 1 | 0 | 0 |
| AD Wightman | eGFR | | | rs4752255 | rs6585310 | 0.00E+00 | 6.56E-03 | 0.00E+00 | 9.76E-01 | 1.76E-02 | 1 | 0 | 0 |
| AD Wightman | eGFR | 18 | 3555 | rs79057333 | rs7089398 | 0.00E+00 | 1.36E-03 | 0.00E+00 | 9.96E-01 | 2.20E-03 | 1 | 0 | 0 |
| AD Wightman | eGFR | | | rs72834453 | rs7089398 | 0.00E+00 | 1.36E-03 | 0.00E+00 | 9.97E-01 | 1.84E-03 | 1 | 0 | 0 |
| AD Wightman | eGFR | | | rs17623531 | rs7089398 | 0.00E+00 | 1.36E-03 | 0.00E+00 | 9.98E-01 | 6.64E-04 | 1 | 0 | 0 |
| AD Wightman | eGFR | | | rs10490922 | rs7089398 | 0.00E+00 | 1.36E-03 | 0.00E+00 | 9.98E-01 | 8.84E-04 | 1 | 0 | 0 |
| AD Wightman | eGFR | | | rs10490923 | rs7089398 | 0.00E+00 | 1.36E-03 | 0.00E+00 | 9.98E-01 | 5.76E-04 | 1 | 0 | 0 |
| AD Wightman | eGFR | | | rs72834439 | rs7089398 | 0.00E+00 | 1.36E-03 | 0.00E+00 | 9.98E-01 | 1.09E-03 | 1 | 0 | 0 |
| AD Wightma | eGFR | 19 | 1763 | rs10769258 | rs10838702 | 0.00E+00 | 2.15E-13 | 0.00E+00 | 1.00E+00 | 2.25E-10 | 1 | 0 | 0 |

| | | | coloc SUSIE method | | | | | | | | | | |
|---|---|---|---|---|---|---|---|---|---|---|---|---|---|
| **Trait1** | **Trait 2** | **cFDRlocusnum** | **nsnps 2** | **hit1** | **hit2** | **PP.H0.abf** | **PP.H1.abf** | **PP.H2.abf** | **PP.H3.abf** | **PP.H4.abf** | **PP.H3+PP.H4≥0.8** | **PP.H3/PP.H4≥3** | **PP.H4/PP.H3≥5** |
| n | | | | | | | | | | | | | |
| AD Wightman | eGFR | | | rs4752987 | rs10838702 | 0.00E+00 | 2.15E-13 | 0.00E+00 | 1.00E+00 | 2.03E-10 | 1 | 0 | 0 |
| AD Wightman | eGFR | 22 | 3017 | rs7101940 | rs4935805 | 0.00E+00 | 1.94E-04 | 0.00E+00 | 1.00E+00 | 1.96E-06 | 1 | 0 | 0 |
| AD Wightman | eGFR | | | rs7118212 | rs4935805 | 0.00E+00 | 1.94E-04 | 0.00E+00 | 1.00E+00 | 2.02E-06 | 1 | 0 | 0 |
| AD Wightman | eGFR | | | rs7101940 | rs11218721 | 0.00E+00 | 2.02E-04 | 0.00E+00 | 2.92E-01 | 7.08E-01 | 1 | 0 | 0 |
| AD Wightman | eGFR | | | rs7118212 | rs11218721 | 0.00E+00 | 1.70E-04 | 0.00E+00 | 2.45E-01 | 7.55E-01 | 1 | 1 | 0 |
| AD Wightman | eGFR | 23 | 2724 | rs74626145 | rs2277383 | 0.00E+00 | 1.44E-06 | 0.00E+00 | 9.04E-01 | 9.56E-02 | 1 | 0 | 0 |
| AD Wightman | eGFR | | | rs76195628 | rs2277383 | 0.00E+00 | 1.48E-06 | 0.00E+00 | 9.33E-01 | 6.74E-02 | 1 | 0 | 0 |
| AD Wightman | eGFR | | | rs11169939 | rs2277383 | 0.00E+00 | 1.47E-06 | 0.00E+00 | 9.25E-01 | 7.48E-02 | 1 | 0 | 0 |
| AD Wightman | eGFR | | | rs12230130 | rs2277383 | 0.00E+00 | 1.48E-06 | 0.00E+00 | 9.31E-01 | 6.89E-02 | 1 | 0 | 0 |
| AD Wightman | eGFR | 24 | 1807 | rs1043011 | rs2277339 | 0.00E+00 | 1.54E-03 | 0.00E+00 | 9.98E-01 | 1.30E-05 | 1 | 0 | 0 |
| AD Wightman | eGFR | | | rs2657879 | rs2277339 | 0.00E+00 | 1.54E-03 | 0.00E+00 | 9.98E-01 | 1.30E-05 | 1 | 0 | 0 |
| AD Wightma | eGFR | | | rs2638314 | rs2277339 | 0.00E+00 | 1.54E-03 | 0.00E+00 | 9.98E-01 | 1.31E-05 | 1 | 0 | 0 |

| | | | coloc SUSIE method | | | | | | | | | | |
|---|---|---|---|---|---|---|---|---|---|---|---|---|---|
| **Trait1** | **Trait 2** | **cFDRlocusnum** | **nsnps 2** | **hit1** | **hit2** | **PP.H0.abf** | **PP.H1.abf** | **PP.H2.abf** | **PP.H3.abf** | **PP.H4.abf** | **PP.H3+PP.H4≥0.8** | **PP.H3/PP.H4≥3** | **PP.H4/PP.H3≥5** |
| n | | | | | | | | | | | | | |
| AD Wightman | eGFR | | | rs73114872 | rs2277339 | 0.00E+00 | 1.54E-03 | 0.00E+00 | 9.98E-01 | 1.31E-05 | 1 | 0 | 0 |
| AD Wightman | eGFR | | | rs1043011 | rs2657880 | 0.00E+00 | 6.95E-12 | 0.00E+00 | 2.20E-02 | 9.78E-01 | 1 | 1 | 1 |
| AD Wightman | eGFR | | | rs2657879 | rs2657880 | 0.00E+00 | 8.07E-12 | 0.00E+00 | 2.59E-02 | 9.74E-01 | 1 | 1 | 1 |
| AD Wightman | eGFR | | | rs2638314 | rs2657880 | 0.00E+00 | 5.86E-12 | 0.00E+00 | 1.82E-02 | 9.82E-01 | 1 | 1 | 1 |
| AD Wightman | eGFR | | | rs73114872 | rs2657880 | 0.00E+00 | 5.34E-12 | 0.00E+00 | 1.64E-02 | 9.84E-01 | 1 | 1 | 1 |
| AD Wightman | eGFR | | | rs1043011 | rs73129102 | 0.00E+00 | 5.43E-05 | 0.00E+00 | 1.00E+00 | 5.26E-07 | 1 | 0 | 0 |
| AD Wightman | eGFR | | | rs2657879 | rs73129102 | 0.00E+00 | 5.43E-05 | 0.00E+00 | 1.00E+00 | 5.32E-07 | 1 | 0 | 0 |
| AD Wightman | eGFR | | | rs2638314 | rs73129102 | 0.00E+00 | 5.43E-05 | 0.00E+00 | 1.00E+00 | 5.18E-07 | 1 | 0 | 0 |
| AD Wightman | eGFR | | | rs73114872 | rs73129102 | 0.00E+00 | 5.43E-05 | 0.00E+00 | 1.00E+00 | 5.14E-07 | 1 | 0 | 0 |
| AD Wightman | eGFR | 26 | 3075 | rs77612485 | rs7176597 | 0.00E+00 | 0.00E+00 | 0.00E+00 | 1.00E+00 | 0.00E+00 | 1 | 0 | 0 |
| AD Wightman | eGFR | | | rs190855337 | rs7176597 | 0.00E+00 | 0.00E+00 | 0.00E+00 | 1.00E+00 | 0.00E+00 | 1 | 0 | 0 |
| AD Wightman | eGFR | | | rs76855300 | rs7176597 | 0.00E+00 | 0.00E+00 | 1.89E-318 | 1.00E+00 | 1.79E-302 | 1 | 0 | 0 |
| AD Wightma | eGFR | | | rs117618017 | rs7176597 | 0.00E+00 | 0.00E+00 | 0.00E+00 | 1.00E+00 | 0.00E+00 | 1 | 0 | 0 |

| | | | coloc SUSIE method | | | | | | | | | | |
|---|---|---|---|---|---|---|---|---|---|---|---|---|---|
| **Trait1** | **Trait2** | **cFDRlocusnum** | **nsnps2** | **hit1** | **hit2** | **PP.H0.abf** | **PP.H1.abf** | **PP.H2.abf** | **PP.H3.abf** | **PP.H4.abf** | **PP.H3+PP.H4≥0.8** | **PP.H3/PP.H4≥3** | **PP.H4/PP.H3≥5** |
| n | | | | | | | | | | | | | |
| AD Wightman | eGFR | | | rs79474735 | rs7176597 | 0.00E+00 | 0.00E+00 | 0.00E+00 | 1.00E+00 | 0.00E+00 | 1 | 0 | 0 |
| AD Wightman | eGFR | | | rs117034065 | rs7176597 | 0.00E+00 | 0.00E+00 | 0.00E+00 | 1.00E+00 | 0.00E+00 | 1 | 0 | 0 |
| AD Wightman | eGFR | | | rs117618017 | rs7176597 | 0.00E+00 | 0.00E+00 | 1.12E-25 | 1.00E+00 | 1.06E-27 | 1 | 0 | 0 |
| AD Wightman | eGFR | | | rs77612485 | rs12595312 | 0.00E+00 | 0.00E+00 | 0.00E+00 | 1.00E+00 | 0.00E+00 | 1 | 0 | 0 |
| AD Wightman | eGFR | | | rs190855337 | rs12595312 | 0.00E+00 | 0.00E+00 | 0.00E+00 | 1.00E+00 | 0.00E+00 | 1 | 0 | 0 |
| AD Wightman | eGFR | | | rs76855300 | rs12595312 | 0.00E+00 | 0.00E+00 | 1.89E-318 | 1.00E+00 | 8.12E-312 | 1 | 0 | 0 |
| AD Wightman | eGFR | | | rs117618017 | rs12595312 | 0.00E+00 | 0.00E+00 | 0.00E+00 | 1.00E+00 | 0.00E+00 | 1 | 0 | 0 |
| AD Wightman | eGFR | | | rs79474735 | rs12595312 | 0.00E+00 | 0.00E+00 | 0.00E+00 | 1.00E+00 | 0.00E+00 | 1 | 0 | 0 |
| AD Wightman | eGFR | | | rs117034065 | rs12595312 | 0.00E+00 | 0.00E+00 | 0.00E+00 | 1.00E+00 | 0.00E+00 | 1 | 0 | 0 |
| AD Wightman | eGFR | | | rs117618017 | rs12595312 | 0.00E+00 | 0.00E+00 | 1.12E-25 | 1.00E+00 | 1.53E-24 | 1 | 0 | 0 |
| AD Wightman | eGFR | | | rs77612485 | rs2937855 | 0.00E+00 | 0.00E+00 | 0.00E+00 | 1.00E+00 | 0.00E+00 | 1 | 0 | 0 |
| AD Wightman | eGFR | | | rs190855337 | rs2937855 | 0.00E+00 | 0.00E+00 | 0.00E+00 | 1.00E+00 | 0.00E+00 | 1 | 0 | 0 |
| AD Wightman | eGFR | | | rs76855300 | rs2937855 | 0.00E+00 | 0.00E+00 | 1.89E-318 | 1.00E+00 | 4.15E-306 | 1 | 0 | 0 |

| | | | coloc SUSIE method | | | | | | | | | | |
|---|---|---|---|---|---|---|---|---|---|---|---|---|---|
| Trait1 | Trait 2 | cFDRlocusnum | nsnps 2 | hit1 | hit2 | PP.H0.abf | PP.H1.abf | PP.H2.abf | PP.H3.abf | PP.H4.abf | PP.H3+PP.H4≥0.8 | PP.H3/PP.H4≥3 | PP.H4/PP.H3≥5 |
| AD Wightman | eGFR | | | rs117618017 | rs2937855 | 0.00E+00 | 0.00E+00 | 0.00E+00 | 1.00E+00 | 0.00E+00 | 1 | 0 | 0 |
| AD Wightman | eGFR | | | rs79474735 | rs2937855 | 0.00E+00 | 0.00E+00 | 0.00E+00 | 1.00E+00 | 0.00E+00 | 1 | 0 | 0 |
| AD Wightman | eGFR | | | rs117034065 | rs2937855 | 0.00E+00 | 0.00E+00 | 0.00E+00 | 1.00E+00 | 0.00E+00 | 1 | 0 | 0 |
| AD Wightman | eGFR | | | rs117618017 | rs2937855 | 0.00E+00 | 0.00E+00 | 1.12E-25 | 1.00E+00 | 1.62E-27 | 1 | 0 | 0 |
| AD Wightman | eGFR | | | rs77612485 | rs2652804 | 0.00E+00 | 0.00E+00 | 0.00E+00 | 1.00E+00 | 0.00E+00 | 1 | 0 | 0 |
| AD Wightman | eGFR | | | rs190855337 | rs2652804 | 0.00E+00 | 0.00E+00 | 0.00E+00 | 1.00E+00 | 0.00E+00 | 1 | 0 | 0 |
| AD Wightman | eGFR | | | rs76855300 | rs2652804 | 0.00E+00 | 0.00E+00 | 1.89E-318 | 1.00E+00 | 1.50E-304 | 1 | 0 | 0 |
| AD Wightman | eGFR | | | rs117618017 | rs2652804 | 0.00E+00 | 0.00E+00 | 0.00E+00 | 1.00E+00 | 0.00E+00 | 1 | 0 | 0 |
| AD Wightman | eGFR | | | rs79474735 | rs2652804 | 0.00E+00 | 0.00E+00 | 0.00E+00 | 1.00E+00 | 0.00E+00 | 1 | 0 | 0 |
| AD Wightman | eGFR | | | rs117034065 | rs2652804 | 0.00E+00 | 0.00E+00 | 0.00E+00 | 1.00E+00 | 0.00E+00 | 1 | 0 | 0 |
| AD Wightman | eGFR | | | rs117618017 | rs2652804 | 0.00E+00 | 0.00E+00 | 1.12E-25 | 1.00E+00 | 6.87E-25 | 1 | 0 | 0 |
| AD Wightman | eGFR | | | rs77612485 | rs2729833 | 0.00E+00 | 0.00E+00 | 0.00E+00 | 1.00E+00 | 0.00E+00 | 1 | 0 | 0 |
| AD Wightman | eGFR | | | rs190855337 | rs2729833 | 0.00E+00 | 0.00E+00 | 0.00E+00 | 1.00E+00 | 0.00E+00 | 1 | 0 | 0 |
| AD | eGFR | | | rs76855300 | rs2729833 | 0.00E+00 | 0.00E+00 | 1.89E- | 1.00E+00 | 4.21E- | 1 | 0 | 0 |

| | | | coloc SUSIE method | | | | | | | | | | |
|---|---|---|---|---|---|---|---|---|---|---|---|---|---|
| **Trait1** | **Trait2** | **cFDRlocusnum** | **nsnps2** | **hit1** | **hit2** | **PP.H0.abf** | **PP.H1.abf** | **PP.H2.abf** | **PP.H3.abf** | **PP.H4.abf** | **PP.H3+PP.H4≥0.8** | **PP.H3/PP.H4≥3** | **PP.H4/PP.H3≥5** |
| Wightman | | | | | | | | 318 | | 317 | | | |
| AD Wightman | eGFR | | | rs117618017 | rs2729833 | 0.00E+00 | 0.00E+00 | 0.00E+00 | 1.00E+00 | 0.00E+00 | 1 | 0 | 0 |
| AD Wightman | eGFR | | | rs79474735 | rs2729833 | 0.00E+00 | 0.00E+00 | 0.00E+00 | 1.00E+00 | 0.00E+00 | 1 | 0 | 0 |
| AD Wightman | eGFR | | | rs117034065 | rs2729833 | 0.00E+00 | 0.00E+00 | 0.00E+00 | 1.00E+00 | 0.00E+00 | 1 | 0 | 0 |
| AD Wightman | eGFR | | | rs117618017 | rs2729833 | 0.00E+00 | 0.00E+00 | 1.12E-25 | 1.00E+00 | 1.20E-27 | 1 | 0 | 0 |
| AD Wightman | eGFR | | | rs77612485 | rs2652835 | 0.00E+00 | 0.00E+00 | 0.00E+00 | 1.00E+00 | 0.00E+00 | 1 | 0 | 0 |
| AD Wightman | eGFR | | | rs190855337 | rs2652835 | 0.00E+00 | 0.00E+00 | 0.00E+00 | 1.00E+00 | 0.00E+00 | 1 | 0 | 0 |
| AD Wightman | eGFR | | | rs76855300 | rs2652835 | 0.00E+00 | 0.00E+00 | 1.89E-318 | 1.00E+00 | 4.93E-315 | 1 | 0 | 0 |
| AD Wightman | eGFR | | | rs117618017 | rs2652835 | 0.00E+00 | 0.00E+00 | 0.00E+00 | 1.00E+00 | 0.00E+00 | 1 | 0 | 0 |
| AD Wightman | eGFR | | | rs79474735 | rs2652835 | 0.00E+00 | 0.00E+00 | 0.00E+00 | 1.00E+00 | 0.00E+00 | 1 | 0 | 0 |
| AD Wightman | eGFR | | | rs117034065 | rs2652835 | 0.00E+00 | 0.00E+00 | 0.00E+00 | 1.00E+00 | 0.00E+00 | 1 | 0 | 0 |
| AD Wightman | eGFR | | | rs117618017 | rs2652835 | 0.00E+00 | 0.00E+00 | 1.12E-25 | 1.00E+00 | 2.62E-27 | 1 | 0 | 0 |
| AD Wightman | eGFR | | | rs77612485 | rs2652833 | 0.00E+00 | 0.00E+00 | 0.00E+00 | 1.00E+00 | 0.00E+00 | 1 | 0 | 0 |
| AD Wightma | eGFR | | | rs190855337 | rs2652833 | 0.00E+00 | 0.00E+00 | 0.00E+00 | 1.00E+00 | 0.00E+00 | 1 | 0 | 0 |

| | | | coloc SUSIE method | | | | | | | | | | |
|---|---|---|---|---|---|---|---|---|---|---|---|---|---|
| **Trait1** | **Trait 2** | **cFDRlocusnum** | **nsnps 2** | **hit1** | **hit2** | **PP.H0.abf** | **PP.H1.abf** | **PP.H2.abf** | **PP.H3.abf** | **PP.H4.abf** | **PP.H3+PP.H4≥0.8** | **PP.H3/PP.H4≥3** | **PP.H4/PP.H3≥5** |
| n | | | | | | | | | | | | | |
| AD Wightman | eGFR | | | rs76855300 | rs2652833 | 0.00E+00 | 0.00E+00 | 1.89E-318 | 1.00E+00 | 1.71E-313 | 1 | 0 | 0 |
| AD Wightman | eGFR | | | rs117618017 | rs2652833 | 0.00E+00 | 0.00E+00 | 0.00E+00 | 1.00E+00 | 0.00E+00 | 1 | 0 | 0 |
| AD Wightman | eGFR | | | rs79474735 | rs2652833 | 0.00E+00 | 0.00E+00 | 0.00E+00 | 1.00E+00 | 0.00E+00 | 1 | 0 | 0 |
| AD Wightman | eGFR | | | rs117034065 | rs2652833 | 0.00E+00 | 0.00E+00 | 0.00E+00 | 1.00E+00 | 0.00E+00 | 1 | 0 | 0 |
| AD Wightman | eGFR | | | rs117618017 | rs2652833 | 0.00E+00 | 0.00E+00 | 1.12E-25 | 1.00E+00 | 1.39E-27 | 1 | 0 | 0 |
| AD Wightman | eGFR | | | rs77612485 | rs4774474 | 0.00E+00 | 1.26E-20 | 0.00E+00 | 1.00E+00 | 1.11E-21 | 1 | 0 | 0 |
| AD Wightman | eGFR | | | rs190855337 | rs4774474 | 0.00E+00 | 1.26E-20 | 0.00E+00 | 1.00E+00 | 1.50E-22 | 1 | 0 | 0 |
| AD Wightman | eGFR | | | rs76855300 | rs4774474 | 0.00E+00 | 1.26E-20 | 1.89E-318 | 1.00E+00 | 4.80E-21 | 1 | 0 | 0 |
| AD Wightman | eGFR | | | rs117618017 | rs4774474 | 0.00E+00 | 1.26E-20 | 0.00E+00 | 1.00E+00 | 1.96E-20 | 1 | 0 | 0 |
| AD Wightman | eGFR | | | rs79474735 | rs4774474 | 0.00E+00 | 1.26E-20 | 0.00E+00 | 1.00E+00 | 2.09E-22 | 1 | 0 | 0 |
| AD Wightman | eGFR | | | rs117034065 | rs4774474 | 0.00E+00 | 1.26E-20 | 0.00E+00 | 1.00E+00 | 1.81E-21 | 1 | 0 | 0 |
| AD Wightman | eGFR | | | rs117618017 | rs4774474 | 1.41E-45 | 1.26E-20 | 1.12E-25 | 1.00E+00 | 2.06E-20 | 1 | 0 | 0 |
| AD Wightman | eGFR | | | rs77612485 | rs11632401 | 0.00E+00 | 2.84E-10 | 0.00E+00 | 1.00E+00 | 8.32E-11 | 1 | 0 | 0 |

| | | | coloc SUSIE method | | | | | | | | | | |
|---|---|---|---|---|---|---|---|---|---|---|---|---|---|
| **Trait1** | **Trait 2** | **cFDRlocusnum** | **nsnps 2** | **hit1** | **hit2** | **PP.H0.abf** | **PP.H1.abf** | **PP.H2.abf** | **PP.H3.abf** | **PP.H4.abf** | **PP.H3+PP.H4≥0.8** | **PP.H3/PP.H4≥3** | **PP.H4/PP.H3≥5** |
| AD Wightman | eGFR | | | rs190855337 | rs11632401 | 0.00E+00 | 2.84E-10 | 0.00E+00 | 1.00E+00 | 6.98E-12 | 1 | 0 | 0 |
| AD Wightman | eGFR | | | rs76855300 | rs11632401 | 0.00E+00 | 2.84E-10 | 1.89E-318 | 1.00E+00 | 8.03E-08 | 1 | 0 | 0 |
| AD Wightman | eGFR | | | rs117618017 | rs11632401 | 0.00E+00 | 2.84E-10 | 0.00E+00 | 1.00E+00 | 1.68E-07 | 1 | 0 | 0 |
| AD Wightman | eGFR | | | rs79474735 | rs11632401 | 0.00E+00 | 2.84E-10 | 0.00E+00 | 1.00E+00 | 9.76E-12 | 1 | 0 | 0 |
| AD Wightman | eGFR | | | rs117034065 | rs11632401 | 0.00E+00 | 2.84E-10 | 0.00E+00 | 1.00E+00 | 1.16E-10 | 1 | 0 | 0 |
| AD Wightman | eGFR | | | rs117618017 | rs11632401 | 3.16E-35 | 2.84E-10 | 1.12E-25 | 1.00E+00 | 1.63E-07 | 1 | 0 | 0 |
| AD Wightman | eGFR | 27 | 2942 | rs12909668 | rs7176070 | 0.00E+00 | 0.00E+00 | 0.00E+00 | 1.00E+00 | 1.59E-30 | 1 | 0 | 0 |
| AD Wightman | eGFR | | | rs7182694 | rs7176070 | 0.00E+00 | 0.00E+00 | 0.00E+00 | 1.00E+00 | 7.10E-26 | 1 | 0 | 0 |
| AD Wightman | eGFR | | | rs7182567 | rs7176070 | 0.00E+00 | 0.00E+00 | 0.00E+00 | 1.00E+00 | 3.23E-26 | 1 | 0 | 0 |
| AD Wightman | eGFR | | | rs12909668 | rs78555828 | 0.00E+00 | 1.47E-06 | 0.00E+00 | 1.00E+00 | 1.74E-08 | 1 | 0 | 0 |
| AD Wightman | eGFR | | | rs7182694 | rs78555828 | 0.00E+00 | 1.47E-06 | 0.00E+00 | 1.00E+00 | 2.01E-08 | 1 | 0 | 0 |
| AD Wightman | eGFR | | | rs7182567 | rs78555828 | 0.00E+00 | 1.47E-06 | 0.00E+00 | 1.00E+00 | 2.01E-08 | 1 | 0 | 0 |
| AD Wightman | eGFR | | | rs12909668 | rs12916717 | 0.00E+00 | 0.00E+00 | 0.00E+00 | 1.00E+00 | 0.00E+00 | 1 | 0 | 0 |

| | | | coloc SUSIE method | | | | | | | | | | |
|---|---|---|---|---|---|---|---|---|---|---|---|---|---|
| **Trait1** | **Trait 2** | **cFDRlocusnum** | **nsnps 2** | **hit1** | **hit2** | **PP.H0.abf** | **PP.H1.abf** | **PP.H2.abf** | **PP.H3.abf** | **PP.H4.abf** | **PP.H3+PP.H4≥0.8** | **PP.H3/PP.H4≥3** | **PP.H4/PP.H3≥5** |
| AD Wightman | eGFR | | | rs7182694 | rs12916717 | 0.00E+00 | 0.00E+00 | 0.00E+00 | 1.00E+00 | 0.00E+00 | 1 | 0 | 0 |
| AD Wightman | eGFR | | | rs7182567 | rs12916717 | 0.00E+00 | 0.00E+00 | 0.00E+00 | 1.00E+00 | 0.00E+00 | 1 | 0 | 0 |
| AD Wightman | eGFR | | | rs12909668 | rs9888691 | 0.00E+00 | 0.00E+00 | 0.00E+00 | 1.00E+00 | 0.00E+00 | 1 | 0 | 0 |
| AD Wightman | eGFR | | | rs7182694 | rs9888691 | 0.00E+00 | 0.00E+00 | 0.00E+00 | 1.00E+00 | 0.00E+00 | 1 | 0 | 0 |
| AD Wightman | eGFR | | | rs7182567 | rs9888691 | 0.00E+00 | 0.00E+00 | 0.00E+00 | 1.00E+00 | 0.00E+00 | 1 | 0 | 0 |
| AD Wightman | eGFR | | | rs12909668 | rs11633152 | 0.00E+00 | 0.00E+00 | 0.00E+00 | 1.00E+00 | 0.00E+00 | 1 | 0 | 0 |
| AD Wightman | eGFR | | | rs7182694 | rs11633152 | 0.00E+00 | 0.00E+00 | 0.00E+00 | 1.00E+00 | 0.00E+00 | 1 | 0 | 0 |
| AD Wightman | eGFR | | | rs7182567 | rs11633152 | 0.00E+00 | 0.00E+00 | 0.00E+00 | 1.00E+00 | 0.00E+00 | 1 | 0 | 0 |
| AD Wightman | eGFR | | | rs12909668 | rs12913779 | 0.00E+00 | 0.00E+00 | 0.00E+00 | 1.00E+00 | 0.00E+00 | 1 | 0 | 0 |
| AD Wightman | eGFR | | | rs7182694 | rs12913779 | 0.00E+00 | 0.00E+00 | 0.00E+00 | 1.00E+00 | 0.00E+00 | 1 | 0 | 0 |
| AD Wightman | eGFR | | | rs7182567 | rs12913779 | 0.00E+00 | 0.00E+00 | 0.00E+00 | 1.00E+00 | 0.00E+00 | 1 | 0 | 0 |
| AD Wightman | eGFR | | | rs12909668 | rs12917502 | 0.00E+00 | 0.00E+00 | 0.00E+00 | 1.00E+00 | 0.00E+00 | 1 | 0 | 0 |
| AD Wightman | eGFR | | | rs7182694 | rs12917502 | 0.00E+00 | 0.00E+00 | 0.00E+00 | 1.00E+00 | 0.00E+00 | 1 | 0 | 0 |
| AD | eGFR | | | rs7182567 | rs12917502 | 0.00E+00 | 0.00E+00 | 0.00E+00 | 1.00E+00 | 0.00E+00 | 1 | 0 | 0 |

| | | | coloc SUSIE method | | | | | | | | | | |
|---|---|---|---|---|---|---|---|---|---|---|---|---|---|
| **Trait1** | **Trait 2** | **cFDRlocusnum** | **nsnps 2** | **hit1** | **hit2** | **PP.H0.abf** | **PP.H1.abf** | **PP.H2.abf** | **PP.H3.abf** | **PP.H4.abf** | **PP.H3+PP.H4≥0.8** | **PP.H3/PP.H4≥3** | **PP.H4/PP.H3≥5** |
| Wightman | | | | | | | | | | | | | |
| AD Wightman | eGFR | | | rs12909668 | rs2622821 | 0.00E+00 | 0.00E+00 | 0.00E+00 | 1.00E+00 | 0.00E+00 | 1 | 0 | 0 |
| AD Wightman | eGFR | | | rs7182694 | rs2622821 | 0.00E+00 | 0.00E+00 | 0.00E+00 | 1.00E+00 | 0.00E+00 | 1 | 0 | 0 |
| AD Wightman | eGFR | | | rs7182567 | rs2622821 | 0.00E+00 | 0.00E+00 | 0.00E+00 | 1.00E+00 | 0.00E+00 | 1 | 0 | 0 |
| AD Wightman | eGFR | 31 | 2775 | rs60913890 | rs142849507 | 0.00E+00 | 2.72E-02 | 0.00E+00 | 9.50E-01 | 2.31E-02 | 1 | 0 | 0 |
| AD Wightman | eGFR | | | rs1111316 | rs142849507 | 0.00E+00 | 2.72E-02 | 0.00E+00 | 9.49E-01 | 2.36E-02 | 1 | 0 | 0 |
| AD Wightman | eGFR | | | rs60913890 | rs74018944 | 0.00E+00 | 4.24E-14 | 0.00E+00 | 1.00E+00 | 3.56E-15 | 1 | 0 | 0 |
| AD Wightman | eGFR | | | rs1111316 | rs74018944 | 0.00E+00 | 4.24E-14 | 0.00E+00 | 1.00E+00 | 3.72E-15 | 1 | 0 | 0 |
| AD Wightman | eGFR | | | rs60913890 | rs9932625 | 0.00E+00 | 7.65E-34 | 0.00E+00 | 1.00E+00 | 1.60E-35 | 1 | 0 | 0 |
| AD Wightman | eGFR | | | rs1111316 | rs9932625 | 0.00E+00 | 7.65E-34 | 0.00E+00 | 1.00E+00 | 1.62E-35 | 1 | 0 | 0 |
| AD Wightman | eGFR | | | rs60913890 | rs117986012 | 0.00E+00 | 1.19E-03 | 0.00E+00 | 9.99E-01 | 3.37E-05 | 1 | 0 | 0 |
| AD Wightman | eGFR | | | rs1111316 | rs117986012 | 0.00E+00 | 1.19E-03 | 0.00E+00 | 9.99E-01 | 3.47E-05 | 1 | 0 | 0 |
| AD Wightman | eGFR | | | rs60913890 | rs12445948 | 0.00E+00 | 1.86E-03 | 0.00E+00 | 9.98E-01 | 2.30E-05 | 1 | 0 | 0 |
| AD | eGFR | | | rs1111316 | rs12445948 | 0.00E+00 | 1.86E-03 | 0.00E+00 | 9.98E-01 | 2.33E-05 | 1 | 0 | 0 |

| | | | coloc SUSIE method | | | | | | | | | | |
|---|---|---|---|---|---|---|---|---|---|---|---|---|---|
| **Trait1** | **Trait 2** | **cFDRlocusnum** | **nsnps 2** | **hit1** | **hit2** | **PP.H0.abf** | **PP.H1.abf** | **PP.H2.abf** | **PP.H3.abf** | **PP.H4.abf** | **PP.H3+PP.H4≥0.8** | **PP.H3/PP.H4≥3** | **PP.H4/PP.H3≥5** |
| Wightman | | | | | | | | | | | | | |
| AD Wightman | eGFR | 32 | 1960 | rs78815193 | rs7203216 | 0.00E+00 | 0.00E+00 | 0.00E+00 | 1.00E+00 | 0.00E+00 | 1 | 0 | 0 |
| AD Wightman | eGFR | | | rs76612312 | rs7203216 | 0.00E+00 | 0.00E+00 | 0.00E+00 | 1.00E+00 | 0.00E+00 | 1 | 0 | 0 |
| AD Wightman | eGFR | | | rs78815193 | rs8058222 | 0.00E+00 | 0.00E+00 | 0.00E+00 | 1.00E+00 | 0.00E+00 | 1 | 0 | 0 |
| AD Wightman | eGFR | | | rs76612312 | rs8058222 | 0.00E+00 | 0.00E+00 | 0.00E+00 | 1.00E+00 | 0.00E+00 | 1 | 0 | 0 |
| AD Wightman | eGFR | | | rs78815193 | rs11075890 | 0.00E+00 | 0.00E+00 | 0.00E+00 | 1.00E+00 | 0.00E+00 | 1 | 0 | 0 |
| AD Wightman | eGFR | | | rs76612312 | rs11075890 | 0.00E+00 | 0.00E+00 | 0.00E+00 | 1.00E+00 | 0.00E+00 | 1 | 0 | 0 |
| AD Wightman | eGFR | | | rs78815193 | rs61451008 | 0.00E+00 | 0.00E+00 | 0.00E+00 | 1.00E+00 | 0.00E+00 | 1 | 0 | 0 |
| AD Wightman | eGFR | | | rs76612312 | rs61451008 | 0.00E+00 | 0.00E+00 | 0.00E+00 | 1.00E+00 | 0.00E+00 | 1 | 0 | 0 |
| AD Wightman | eGFR | | | rs78815193 | rs140535806 | 0.00E+00 | 1.85E-50 | 0.00E+00 | 1.00E+00 | 1.64E-52 | 1 | 0 | 0 |
| AD Wightman | eGFR | | | rs76612312 | rs140535806 | 0.00E+00 | 1.85E-50 | 0.00E+00 | 1.00E+00 | 1.13E-52 | 1 | 0 | 0 |
| AD Wightman | eGFR | | | rs78815193 | rs6499539 | 0.00E+00 | 0.00E+00 | 0.00E+00 | 1.00E+00 | 0.00E+00 | 1 | 0 | 0 |
| AD Wightman | eGFR | | | rs76612312 | rs6499539 | 0.00E+00 | 0.00E+00 | 0.00E+00 | 1.00E+00 | 0.00E+00 | 1 | 0 | 0 |

| | | | coloc SUSIE method | | | | | | | | | | |
|---|---|---|---|---|---|---|---|---|---|---|---|---|---|
| **Trait1** | **Trait 2** | **cFDRlocusnum** | **nsnps 2** | **hit1** | **hit2** | **PP.H0.abf** | **PP.H1.abf** | **PP.H2.abf** | **PP.H3.abf** | **PP.H4.abf** | **PP.H3+PP.H4≥0.8** | **PP.H3/PP.H4≥3** | **PP.H4/PP.H3≥5** |
| AD Wightman | eGFR | 33 | 3627 | rs7503952 | rs9911200 | 0.00E+00 | 3.13E-216 | 0.00E+00 | 1.00E+00 | 4.97E-217 | 1 | 0 | 0 |
| AD Wightman | eGFR | | | rs12937244 | rs9911200 | 0.00E+00 | 3.13E-216 | 0.00E+00 | 1.00E+00 | 5.42E-217 | 1 | 0 | 0 |
| AD Wightman | eGFR | | | rs4790818 | rs9911200 | 0.00E+00 | 3.13E-216 | 0.00E+00 | 1.00E+00 | 5.04E-217 | 1 | 0 | 0 |
| AD Wightman | eGFR | | | rs12944946 | rs9911200 | 0.00E+00 | 3.13E-216 | 0.00E+00 | 1.00E+00 | 4.72E-217 | 1 | 0 | 0 |
| AD Wightman | eGFR | | | rs62090058 | rs9911200 | 0.00E+00 | 3.13E-216 | 0.00E+00 | 1.00E+00 | 4.23E-217 | 1 | 0 | 0 |
| AD Wightman | eGFR | | | rs7501750 | rs9911200 | 0.00E+00 | 3.13E-216 | 0.00E+00 | 1.00E+00 | 5.04E-217 | 1 | 0 | 0 |
| AD Wightman | eGFR | | | rs7503952 | rs9906546 | 0.00E+00 | 3.03E-42 | 0.00E+00 | 1.00E+00 | 1.09E-43 | 1 | 0 | 0 |
| AD Wightman | eGFR | | | rs12937244 | rs9906546 | 0.00E+00 | 3.03E-42 | 0.00E+00 | 1.00E+00 | 1.13E-43 | 1 | 0 | 0 |
| AD Wightman | eGFR | | | rs4790818 | rs9906546 | 0.00E+00 | 3.03E-42 | 0.00E+00 | 1.00E+00 | 1.10E-43 | 1 | 0 | 0 |
| AD Wightman | eGFR | | | rs12944946 | rs9906546 | 0.00E+00 | 3.03E-42 | 0.00E+00 | 1.00E+00 | 1.06E-43 | 1 | 0 | 0 |
| AD Wightman | eGFR | | | rs62090058 | rs9906546 | 0.00E+00 | 3.03E-42 | 0.00E+00 | 1.00E+00 | 9.76E-44 | 1 | 0 | 0 |
| AD Wightman | eGFR | | | rs7501750 | rs9906546 | 0.00E+00 | 3.03E-42 | 0.00E+00 | 1.00E+00 | 1.08E-43 | 1 | 0 | 0 |
| AD Wightman | eGFR | | | rs7503952 | rs9894634 | 0.00E+00 | 4.33E-101 | 0.00E+00 | 1.00E+00 | 1.20E-102 | 1 | 0 | 0 |
| AD | eGFR | | | rs12937244 | rs9894634 | 0.00E+00 | 4.33E- | 0.00E+00 | 1.00E+00 | 1.25E- | 1 | 0 | 0 |

| | | | coloc SUSIE method | | | | | | | | | | |
|---|---|---|---|---|---|---|---|---|---|---|---|---|---|
| **Trait1** | **Trait 2** | **cFDRlocusnum** | **nsnps 2** | **hit1** | **hit2** | **PP.H0.abf** | **PP.H1.abf** | **PP.H2.abf** | **PP.H3.abf** | **PP.H4.abf** | **PP.H3+PP.H4≥0.8** | **PP.H3/PP.H4≥3** | **PP.H4/PP.H3≥5** |
| Wightman | | | | | | | 101 | | | 102 | | | |
| AD Wightman | eGFR | | | rs4790818 | rs9894634 | 0.00E+00 | 4.33E-101 | 0.00E+00 | 1.00E+00 | 1.21E-102 | 1 | 0 | 0 |
| AD Wightman | eGFR | | | rs12944946 | rs9894634 | 0.00E+00 | 4.33E-101 | 0.00E+00 | 1.00E+00 | 1.16E-102 | 1 | 0 | 0 |
| AD Wightman | eGFR | | | rs62090058 | rs9894634 | 0.00E+00 | 4.33E-101 | 0.00E+00 | 1.00E+00 | 1.08E-102 | 1 | 0 | 0 |
| AD Wightman | eGFR | | | rs7501750 | rs9894634 | 0.00E+00 | 4.33E-101 | 0.00E+00 | 1.00E+00 | 1.19E-102 | 1 | 0 | 0 |
| AD Wightman | eGFR | | | rs7503952 | rs57937546 | 0.00E+00 | 1.43E-03 | 0.00E+00 | 8.92E-01 | 1.07E-01 | 1 | 0 | 0 |
| AD Wightman | eGFR | | | rs12937244 | rs57937546 | 0.00E+00 | 1.42E-03 | 0.00E+00 | 8.84E-01 | 1.14E-01 | 1 | 0 | 0 |
| AD Wightman | eGFR | | | rs4790818 | rs57937546 | 0.00E+00 | 1.43E-03 | 0.00E+00 | 8.92E-01 | 1.07E-01 | 1 | 0 | 0 |
| AD Wightman | eGFR | | | rs12944946 | rs57937546 | 0.00E+00 | 1.44E-03 | 0.00E+00 | 8.98E-01 | 1.00E-01 | 1 | 0 | 0 |
| AD Wightman | eGFR | | | rs62090058 | rs57937546 | 0.00E+00 | 1.46E-03 | 0.00E+00 | 9.11E-01 | 8.75E-02 | 1 | 0 | 0 |
| AD Wightman | eGFR | | | rs7501750 | rs57937546 | 0.00E+00 | 1.43E-03 | 0.00E+00 | 8.92E-01 | 1.06E-01 | 1 | 0 | 0 |
| AD Wightman | eGFR | | | rs7503952 | rs80314051 | 0.00E+00 | 1.17E-10 | 0.00E+00 | 1.00E+00 | 1.32E-11 | 1 | 0 | 0 |
| AD Wightman | eGFR | | | rs12937244 | rs80314051 | 0.00E+00 | 1.17E-10 | 0.00E+00 | 1.00E+00 | 1.38E-11 | 1 | 0 | 0 |
| AD Wightma | eGFR | | | rs4790818 | rs80314051 | 0.00E+00 | 1.17E-10 | 0.00E+00 | 1.00E+00 | 1.34E-11 | 1 | 0 | 0 |

| | | | coloc SUSIE method | | | | | | | | | | |
|---|---|---|---|---|---|---|---|---|---|---|---|---|---|
| Trait1 | Trait 2 | cFDRlocusnum | nsnps 2 | hit1 | hit2 | PP.H0.abf | PP.H1.abf | PP.H2.abf | PP.H3.abf | PP.H4.abf | PP.H3+PP.H4≥0.8 | PP.H3/PP.H4≥3 | PP.H4/PP.H3≥5 |
| n | | | | | | | | | | | | | |
| AD Wightman | eGFR | | | rs12944946 | rs80314051 | 0.00E+00 | 1.17E-10 | 0.00E+00 | 1.00E+00 | 1.27E-11 | 1 | 0 | 0 |
| AD Wightman | eGFR | | | rs62090058 | rs80314051 | 0.00E+00 | 1.17E-10 | 0.00E+00 | 1.00E+00 | 1.16E-11 | 1 | 0 | 0 |
| AD Wightman | eGFR | | | rs7501750 | rs80314051 | 0.00E+00 | 1.17E-10 | 0.00E+00 | 1.00E+00 | 1.31E-11 | 1 | 0 | 0 |
| AD Wightman | eGFR | | | rs7503952 | rs117026430 | 0.00E+00 | 8.05E-03 | 0.00E+00 | 9.90E-01 | 1.94E-03 | 1 | 0 | 0 |
| AD Wightman | eGFR | | | rs12937244 | rs117026430 | 0.00E+00 | 8.05E-03 | 0.00E+00 | 9.90E-01 | 2.04E-03 | 1 | 0 | 0 |
| AD Wightman | eGFR | | | rs4790818 | rs117026430 | 0.00E+00 | 8.05E-03 | 0.00E+00 | 9.90E-01 | 1.95E-03 | 1 | 0 | 0 |
| AD Wightman | eGFR | | | rs12944946 | rs117026430 | 0.00E+00 | 8.05E-03 | 0.00E+00 | 9.90E-01 | 1.87E-03 | 1 | 0 | 0 |
| AD Wightman | eGFR | | | rs62090058 | rs117026430 | 0.00E+00 | 8.05E-03 | 0.00E+00 | 9.90E-01 | 1.70E-03 | 1 | 0 | 0 |
| AD Wightman | eGFR | | | rs7501750 | rs117026430 | 0.00E+00 | 8.05E-03 | 0.00E+00 | 9.90E-01 | 1.94E-03 | 1 | 0 | 0 |
| AD Wightman | eGFR | 34 | 3559 | rs7503952 | rs9911200 | 0.00E+00 | 3.34E-216 | 0.00E+00 | 1.00E+00 | 5.27E-217 | 1 | 0 | 0 |
| AD Wightman | eGFR | | | rs12937244 | rs9911200 | 0.00E+00 | 3.34E-216 | 0.00E+00 | 1.00E+00 | 5.75E-217 | 1 | 0 | 0 |
| AD Wightman | eGFR | | | rs4790818 | rs9911200 | 0.00E+00 | 3.34E-216 | 0.00E+00 | 1.00E+00 | 5.34E-217 | 1 | 0 | 0 |
| AD Wightma | eGFR | | | rs12944946 | rs9911200 | 0.00E+00 | 3.34E-216 | 0.00E+00 | 1.00E+00 | 5.00E-217 | 1 | 0 | 0 |

| | | | coloc SUSIE method | | | | | | | | | | |
|---|---|---|---|---|---|---|---|---|---|---|---|---|---|
| **Trait1** | **Trait2** | **cFDRlocusnum** | **nsnps2** | **hit1** | **hit2** | **PP.H0.abf** | **PP.H1.abf** | **PP.H2.abf** | **PP.H3.abf** | **PP.H4.abf** | **PP.H3+PP.H4≥0.8** | **PP.H3/PP.H4≥3** | **PP.H4/PP.H3≥5** |
| n | | | | | | | | | | | | | |
| AD Wightman | eGFR | | | rs62090058 | rs9911200 | 0.00E+00 | 3.34E-216 | 0.00E+00 | 1.00E+00 | 4.49E-217 | 1 | 0 | 0 |
| AD Wightman | eGFR | | | rs7501750 | rs9911200 | 0.00E+00 | 3.34E-216 | 0.00E+00 | 1.00E+00 | 5.34E-217 | 1 | 0 | 0 |
| AD Wightman | eGFR | | | rs7503952 | rs9906546 | 0.00E+00 | 3.12E-42 | 0.00E+00 | 1.00E+00 | 1.11E-43 | 1 | 0 | 0 |
| AD Wightman | eGFR | | | rs12937244 | rs9906546 | 0.00E+00 | 3.12E-42 | 0.00E+00 | 1.00E+00 | 1.16E-43 | 1 | 0 | 0 |
| AD Wightman | eGFR | | | rs4790818 | rs9906546 | 0.00E+00 | 3.12E-42 | 0.00E+00 | 1.00E+00 | 1.12E-43 | 1 | 0 | 0 |
| AD Wightman | eGFR | | | rs12944946 | rs9906546 | 0.00E+00 | 3.12E-42 | 0.00E+00 | 1.00E+00 | 1.08E-43 | 1 | 0 | 0 |
| AD Wightman | eGFR | | | rs62090058 | rs9906546 | 0.00E+00 | 3.12E-42 | 0.00E+00 | 1.00E+00 | 9.99E-44 | 1 | 0 | 0 |
| AD Wightman | eGFR | | | rs7501750 | rs9906546 | 0.00E+00 | 3.12E-42 | 0.00E+00 | 1.00E+00 | 1.11E-43 | 1 | 0 | 0 |
| AD Wightman | eGFR | | | rs7503952 | rs9894634 | 0.00E+00 | 4.28E-101 | 0.00E+00 | 1.00E+00 | 1.18E-102 | 1 | 0 | 0 |
| AD Wightman | eGFR | | | rs12937244 | rs9894634 | 0.00E+00 | 4.28E-101 | 0.00E+00 | 1.00E+00 | 1.23E-102 | 1 | 0 | 0 |
| AD Wightman | eGFR | | | rs4790818 | rs9894634 | 0.00E+00 | 4.28E-101 | 0.00E+00 | 1.00E+00 | 1.19E-102 | 1 | 0 | 0 |
| AD Wightman | eGFR | | | rs12944946 | rs9894634 | 0.00E+00 | 4.28E-101 | 0.00E+00 | 1.00E+00 | 1.15E-102 | 1 | 0 | 0 |
| AD Wightman | eGFR | | | rs62090058 | rs9894634 | 0.00E+00 | 4.28E-101 | 0.00E+00 | 1.00E+00 | 1.06E-102 | 1 | 0 | 0 |

| | | | coloc SUSIE method | | | | | | | | | | |
|---|---|---|---|---|---|---|---|---|---|---|---|---|---|
| **Trait1** | **Trait 2** | **cFDRlocusnum** | **nsnps 2** | **hit1** | **hit2** | **PP.H0.abf** | **PP.H1.abf** | **PP.H2.abf** | **PP.H3.abf** | **PP.H4.abf** | **PP.H3+PP.H4≥0.8** | **PP.H3/PP.H4≥3** | **PP.H4/PP.H3≥5** |
| AD Wightman | eGFR | | | rs7501750 | rs9894634 | 0.00E+00 | 4.28E-101 | 0.00E+00 | 1.00E+00 | 1.17E-102 | 1 | 0 | 0 |
| AD Wightman | eGFR | | | rs7503952 | rs57937546 | 0.00E+00 | 1.43E-03 | 0.00E+00 | 8.92E-01 | 1.06E-01 | 1 | 0 | 0 |
| AD Wightman | eGFR | | | rs12937244 | rs57937546 | 0.00E+00 | 1.42E-03 | 0.00E+00 | 8.85E-01 | 1.13E-01 | 1 | 0 | 0 |
| AD Wightman | eGFR | | | rs4790818 | rs57937546 | 0.00E+00 | 1.43E-03 | 0.00E+00 | 8.92E-01 | 1.06E-01 | 1 | 0 | 0 |
| AD Wightman | eGFR | | | rs12944946 | rs57937546 | 0.00E+00 | 1.44E-03 | 0.00E+00 | 8.99E-01 | 9.98E-02 | 1 | 0 | 0 |
| AD Wightman | eGFR | | | rs62090058 | rs57937546 | 0.00E+00 | 1.47E-03 | 0.00E+00 | 9.12E-01 | 8.69E-02 | 1 | 0 | 0 |
| AD Wightman | eGFR | | | rs7501750 | rs57937546 | 0.00E+00 | 1.44E-03 | 0.00E+00 | 8.93E-01 | 1.06E-01 | 1 | 0 | 0 |
| AD Wightman | eGFR | | | rs7503952 | rs80314051 | 0.00E+00 | 1.17E-10 | 0.00E+00 | 1.00E+00 | 1.32E-11 | 1 | 0 | 0 |
| AD Wightman | eGFR | | | rs12937244 | rs80314051 | 0.00E+00 | 1.17E-10 | 0.00E+00 | 1.00E+00 | 1.37E-11 | 1 | 0 | 0 |
| AD Wightman | eGFR | | | rs4790818 | rs80314051 | 0.00E+00 | 1.17E-10 | 0.00E+00 | 1.00E+00 | 1.34E-11 | 1 | 0 | 0 |
| AD Wightman | eGFR | | | rs12944946 | rs80314051 | 0.00E+00 | 1.17E-10 | 0.00E+00 | 1.00E+00 | 1.27E-11 | 1 | 0 | 0 |
| AD Wightman | eGFR | | | rs62090058 | rs80314051 | 0.00E+00 | 1.17E-10 | 0.00E+00 | 1.00E+00 | 1.16E-11 | 1 | 0 | 0 |
| AD Wightman | eGFR | | | rs7501750 | rs80314051 | 0.00E+00 | 1.17E-10 | 0.00E+00 | 1.00E+00 | 1.31E-11 | 1 | 0 | 0 |
| AD | eGFR | | | rs7503952 | rs11702643 | 0.00E+00 | 8.08E-03 | 0.00E+00 | 9.90E-01 | 1.94E-03 | 1 | 0 | 0 |

| | | | coloc SUSIE method | | | | | | | | | | |
|---|---|---|---|---|---|---|---|---|---|---|---|---|---|
| **Trait1** | **Trait 2** | **cFDRlocusnum** | **nsnps 2** | **hit1** | **hit2** | **PP.H0.abf** | **PP.H1.abf** | **PP.H2.abf** | **PP.H3.abf** | **PP.H4.abf** | **PP.H3+PP.H4≥0.8** | **PP.H3/PP.H4≥3** | **PP.H4/PP.H3≥5** |
| Wightman | | | | | 0 | | | | | | | | |
| AD Wightman | eGFR | | | rs12937244 | rs117026430 | 0.00E+00 | 8.08E-03 | 0.00E+00 | 9.90E-01 | 2.04E-03 | 1 | 0 | 0 |
| AD Wightman | eGFR | | | rs4790818 | rs117026430 | 0.00E+00 | 8.08E-03 | 0.00E+00 | 9.90E-01 | 1.95E-03 | 1 | 0 | 0 |
| AD Wightman | eGFR | | | rs12944946 | rs117026430 | 0.00E+00 | 8.08E-03 | 0.00E+00 | 9.90E-01 | 1.87E-03 | 1 | 0 | 0 |
| AD Wightman | eGFR | | | rs62090058 | rs117026430 | 0.00E+00 | 8.08E-03 | 0.00E+00 | 9.90E-01 | 1.70E-03 | 1 | 0 | 0 |
| AD Wightman | eGFR | | | rs7501750 | rs117026430 | 0.00E+00 | 8.08E-03 | 0.00E+00 | 9.90E-01 | 1.94E-03 | 1 | 0 | 0 |
| AD Wightman | eGFR | 36 | 2076 | rs7212167 | rs1269402 | 0.00E+00 | 0.00E+00 | 0.00E+00 | 1.00E+00 | 0.00E+00 | 1 | 0 | 0 |
| AD Wightman | eGFR | | | rs6502632 | rs1269402 | 0.00E+00 | 0.00E+00 | 0.00E+00 | 1.00E+00 | 0.00E+00 | 1 | 0 | 0 |
| AD Wightman | eGFR | | | rs7212167 | rs9894492 | 0.00E+00 | 0.00E+00 | 0.00E+00 | 1.00E+00 | 0.00E+00 | 1 | 0 | 0 |
| AD Wightman | eGFR | | | rs6502632 | rs9894492 | 0.00E+00 | 0.00E+00 | 0.00E+00 | 1.00E+00 | 0.00E+00 | 1 | 0 | 0 |
| AD Wightman | eGFR | | | rs7212167 | rs11653424 | 0.00E+00 | 0.00E+00 | 0.00E+00 | 1.00E+00 | 0.00E+00 | 1 | 0 | 0 |
| AD Wightman | eGFR | | | rs6502632 | rs11653424 | 0.00E+00 | 0.00E+00 | 0.00E+00 | 1.00E+00 | 0.00E+00 | 1 | 0 | 0 |
| AD Wightman | eGFR | | | rs7212167 | rs76489162 | 0.00E+00 | 0.00E+00 | 0.00E+00 | 1.00E+00 | 0.00E+00 | 1 | 0 | 0 |
| AD | eGFR | | | rs6502632 | rs76489162 | 0.00E+00 | 0.00E+00 | 0.00E+00 | 1.00E+00 | 0.00E+00 | 1 | 0 | 0 |

| | | | coloc SUSIE method | | | | | | | | | | |
|---|---|---|---|---|---|---|---|---|---|---|---|---|---|
| Trait1 | Trait 2 | cFDRlocusnum | nsnps 2 | hit1 | hit2 | PP.H0.abf | PP.H1.abf | PP.H2.abf | PP.H3.abf | PP.H4.abf | PP.H3+PP.H4≥0.8 | PP.H3/PP.H4≥3 | PP.H4/PP.H3≥5 |
| Wightman | | | | | | | | | | | | | |
| AD Wightman | eGFR | | | rs7212167 | rs2660830 | 0.00E+00 | 0.00E+00 | 0.00E+00 | 1.00E+00 | 0.00E+00 | 1 | 0 | 0 |
| AD Wightman | eGFR | | | rs6502632 | rs2660830 | 0.00E+00 | 0.00E+00 | 0.00E+00 | 1.00E+00 | 0.00E+00 | 1 | 0 | 0 |
| AD Wightman | eGFR | | | rs7212167 | rs7212010 | 0.00E+00 | 0.00E+00 | 0.00E+00 | 1.00E+00 | 0.00E+00 | 1 | 0 | 0 |
| AD Wightman | eGFR | | | rs6502632 | rs7212010 | 0.00E+00 | 0.00E+00 | 0.00E+00 | 1.00E+00 | 0.00E+00 | 1 | 0 | 0 |
| AD Wightman | eGFR | | | rs7212167 | rs8073975 | 0.00E+00 | 0.00E+00 | 0.00E+00 | 1.00E+00 | 0.00E+00 | 1 | 0 | 0 |
| AD Wightman | eGFR | | | rs6502632 | rs8073975 | 0.00E+00 | 0.00E+00 | 0.00E+00 | 1.00E+00 | 0.00E+00 | 1 | 0 | 0 |
| AD Wightman | eGFR | | | rs7212167 | rs9910411 | 0.00E+00 | 3.73E-38 | 0.00E+00 | 1.00E+00 | 4.93E-32 | 1 | 0 | 0 |
| AD Wightman | eGFR | | | rs6502632 | rs9910411 | 0.00E+00 | 3.73E-38 | 0.00E+00 | 1.00E+00 | 4.54E-32 | 1 | 0 | 0 |
| AD Wightman | eGFR | 38 | 2019 | rs7223515 | rs4468695 | 0.00E+00 | 4.65E-06 | 0.00E+00 | 1.42E-01 | 8.58E-01 | 1 | 1 | 1 |
| AD Wightman | eGFR | | | rs7406820 | rs4468695 | 0.00E+00 | 7.11E-06 | 0.00E+00 | 2.18E-01 | 7.82E-01 | 1 | 1 | 0 |
| AD Wightman | eGFR | | | rs9898829 | rs4468695 | 0.00E+00 | 5.78E-06 | 0.00E+00 | 1.77E-01 | 8.23E-01 | 1 | 1 | 0 |
| AD Wightman | eGFR | | | rs4731478 | rs4468695 | 0.00E+00 | 6.30E-06 | 0.00E+00 | 1.93E-01 | 8.07E-01 | 1 | 1 | 0 |

| | | | coloc SUSIE method | | | | | | | | | | |
|---|---|---|---|---|---|---|---|---|---|---|---|---|---|
| **Trait1** | **Trait 2** | **cFDRlocusnum** | **nsnps 2** | **hit1** | **hit2** | **PP.H0.abf** | **PP.H1.abf** | **PP.H2.abf** | **PP.H3.abf** | **PP.H4.abf** | **PP.H3+PP.H4≥0.8** | **PP.H3/PP.H4≥3** | **PP.H4/PP.H3≥5** |
| AD Wightman | eGFR | 40 | 3498 | rs76375203 | rs8101667 | 0.00E+00 | 0.00E+00 | 0.00E+00 | 1.00E+00 | 0.00E+00 | 1 | 0 | 0 |
| AD Wightman | eGFR | | | rs17206303 | rs8101667 | 0.00E+00 | 0.00E+00 | 0.00E+00 | 1.00E+00 | 0.00E+00 | 1 | 0 | 0 |
| AD Wightman | eGFR | | | rs76375203 | rs2278402 | 0.00E+00 | 0.00E+00 | 0.00E+00 | 1.00E+00 | 0.00E+00 | 1 | 0 | 0 |
| AD Wightman | eGFR | | | rs17206303 | rs2278402 | 0.00E+00 | 0.00E+00 | 0.00E+00 | 1.00E+00 | 0.00E+00 | 1 | 0 | 0 |
| AD Wightman | eGFR | | | rs76375203 | rs8101546 | 0.00E+00 | 0.00E+00 | 0.00E+00 | 1.00E+00 | 0.00E+00 | 1 | 0 | 0 |
| AD Wightman | eGFR | | | rs17206303 | rs8101546 | 0.00E+00 | 0.00E+00 | 0.00E+00 | 1.00E+00 | 0.00E+00 | 1 | 0 | 0 |
| AD Wightman | eGFR | | | rs76375203 | rs11671862 | 0.00E+00 | 0.00E+00 | 0.00E+00 | 1.00E+00 | 0.00E+00 | 1 | 0 | 0 |
| AD Wightman | eGFR | | | rs17206303 | rs11671862 | 0.00E+00 | 0.00E+00 | 0.00E+00 | 1.00E+00 | 0.00E+00 | 1 | 0 | 0 |
| AD Wightman | eGFR | | | rs76375203 | rs113786673 | 0.00E+00 | 0.00E+00 | 0.00E+00 | 1.00E+00 | 0.00E+00 | 1 | 0 | 0 |
| AD Wightman | eGFR | | | rs17206303 | rs113786673 | 0.00E+00 | 0.00E+00 | 0.00E+00 | 1.00E+00 | 0.00E+00 | 1 | 0 | 0 |
| AD Wightman | eGFR | | | rs76375203 | rs111818441 | 0.00E+00 | 0.00E+00 | 0.00E+00 | 1.00E+00 | 0.00E+00 | 1 | 0 | 0 |
| AD Wightman | eGFR | | | rs17206303 | rs111818441 | 0.00E+00 | 0.00E+00 | 0.00E+00 | 1.00E+00 | 0.00E+00 | 1 | 0 | 0 |
| AD Wightman | eGFR | 41 | 3485 | rs429358 | rs814573 | 0.00E+00 | 4.94E-324 | 0.00E+00 | 1.00E+00 | 3.25E-98 | 1 | 0 | 0 |

| | | | coloc SUSIE method | | | | | | | | | | |
|---|---|---|---|---|---|---|---|---|---|---|---|---|---|
| Trait1 | Trait 2 | cFDRlocusnum | nsnps 2 | hit1 | hit2 | PP.H0.abf | PP.H1.abf | PP.H2.abf | PP.H3.abf | PP.H4.abf | PP.H3+PP.H4≥0.8 | PP.H3/PP.H4≥3 | PP.H4/PP.H3≥5 |
| AD Wightman | eGFR | | | rs769449 | rs814573 | 0.00E+00 | 4.94E-324 | 0.00E+00 | 1.00E+00 | 1.85E-134 | 1 | 0 | 0 |
| AD Wightman | eGFR | | | rs34404554 | rs814573 | 0.00E+00 | 4.94E-324 | 0.00E+00 | 1.00E+00 | 1.58E-197 | 1 | 0 | 0 |
| AD Wightman | eGFR | | | rs6857 | rs814573 | 0.00E+00 | 4.94E-324 | 0.00E+00 | 1.00E+00 | 8.51E-160 | 1 | 0 | 0 |
| AD Wightman | eGFR | | | rs2075650 | rs814573 | 0.00E+00 | 4.94E-324 | 0.00E+00 | 1.00E+00 | 2.18E-197 | 1 | 0 | 0 |
| AD Wightman | eGFR | | | rs11556505 | rs814573 | 0.00E+00 | 4.94E-324 | 0.00E+00 | 1.00E+00 | 6.92E-198 | 1 | 0 | 0 |
| AD Wightman | eGFR | | | rs429358 | rs4420638 | 0.00E+00 | 0.00E+00 | 0.00E+00 | 1.00E+00 | 3.42E-256 | 1 | 0 | 0 |
| AD Wightman | eGFR | | | rs769449 | rs4420638 | 0.00E+00 | 0.00E+00 | 0.00E+00 | 1.00E+00 | 0.00E+00 | 1 | 0 | 0 |
| AD Wightman | eGFR | | | rs34404554 | rs4420638 | 0.00E+00 | 0.00E+00 | 0.00E+00 | 1.00E+00 | 0.00E+00 | 1 | 0 | 0 |
| AD Wightman | eGFR | | | rs6857 | rs4420638 | 0.00E+00 | 0.00E+00 | 0.00E+00 | 1.00E+00 | 0.00E+00 | 1 | 0 | 0 |
| AD Wightman | eGFR | | | rs2075650 | rs4420638 | 0.00E+00 | 0.00E+00 | 0.00E+00 | 1.00E+00 | 0.00E+00 | 1 | 0 | 0 |
| AD Wightman | eGFR | | | rs11556505 | rs4420638 | 0.00E+00 | 0.00E+00 | 0.00E+00 | 1.00E+00 | 0.00E+00 | 1 | 0 | 0 |
| AD Wightman | eGFR | | | rs429358 | rs283811 | 0.00E+00 | 0.00E+00 | 0.00E+00 | 1.00E+00 | 0.00E+00 | 1 | 0 | 0 |
| AD Wightman | eGFR | | | rs769449 | rs283811 | 0.00E+00 | 0.00E+00 | 0.00E+00 | 1.00E+00 | 0.00E+00 | 1 | 0 | 0 |
| AD | eGFR | | | rs34404554 | rs283811 | 0.00E+00 | 0.00E+00 | 0.00E+00 | 1.00E+00 | 0.00E+00 | 1 | 0 | 0 |

| | | | coloc SUSIE method | | | | | | | | | | |
|---|---|---|---|---|---|---|---|---|---|---|---|---|---|
| Trait1 | Trait 2 | cFDRlocusnum | nsnps 2 | hit1 | hit2 | PP.H0.abf | PP.H1.abf | PP.H2.abf | PP.H3.abf | PP.H4.abf | PP.H3+PP.H4≥0.8 | PP.H3/PP.H4≥3 | PP.H4/PP.H3≥5 |
| Wightman | | | | | | | | | | | | | |
| AD Wightman | eGFR | | | rs6857 | rs283811 | 0.00E+00 | 0.00E+00 | 0.00E+00 | 1.00E+00 | 0.00E+00 | 1 | 0 | 0 |
| AD Wightman | eGFR | | | rs2075650 | rs283811 | 0.00E+00 | 0.00E+00 | 0.00E+00 | 1.00E+00 | 0.00E+00 | 1 | 0 | 0 |
| AD Wightman | eGFR | | | rs11556505 | rs283811 | 0.00E+00 | 0.00E+00 | 0.00E+00 | 1.00E+00 | 0.00E+00 | 1 | 0 | 0 |
| AD Wightman | eGFR | | | rs429358 | rs283815 | 0.00E+00 | 0.00E+00 | 0.00E+00 | 1.00E+00 | 0.00E+00 | 1 | 0 | 0 |
| AD Wightman | eGFR | | | rs769449 | rs283815 | 0.00E+00 | 0.00E+00 | 0.00E+00 | 1.00E+00 | 0.00E+00 | 1 | 0 | 0 |
| AD Wightman | eGFR | | | rs34404554 | rs283815 | 0.00E+00 | 0.00E+00 | 0.00E+00 | 1.00E+00 | 0.00E+00 | 1 | 0 | 0 |
| AD Wightman | eGFR | | | rs6857 | rs283815 | 0.00E+00 | 0.00E+00 | 0.00E+00 | 1.00E+00 | 0.00E+00 | 1 | 0 | 0 |
| AD Wightman | eGFR | | | rs2075650 | rs283815 | 0.00E+00 | 0.00E+00 | 0.00E+00 | 1.00E+00 | 0.00E+00 | 1 | 0 | 0 |
| AD Wightman | eGFR | | | rs11556505 | rs283815 | 0.00E+00 | 0.00E+00 | 0.00E+00 | 1.00E+00 | 0.00E+00 | 1 | 0 | 0 |
| AD Wightman | eGFR | | | rs429358 | rs157592 | 0.00E+00 | 1.01E-152 | 0.00E+00 | 1.00E+00 | 5.68E-60 | 1 | 0 | 0 |
| AD Wightman | eGFR | | | rs769449 | rs157592 | 0.00E+00 | 1.01E-152 | 0.00E+00 | 1.00E+00 | 1.69E-72 | 1 | 0 | 0 |
| AD Wightman | eGFR | | | rs34404554 | rs157592 | 0.00E+00 | 1.01E-152 | 0.00E+00 | 1.00E+00 | 1.57E-101 | 1 | 0 | 0 |
| AD Wightma | eGFR | | | rs6857 | rs157592 | 0.00E+00 | 1.01E-152 | 0.00E+00 | 1.00E+00 | 8.08E-84 | 1 | 0 | 0 |

| | | | coloc SUSIE method | | | | | | | | | | |
|---|---|---|---|---|---|---|---|---|---|---|---|---|---|
| **Trait1** | **Trait 2** | **cFDRlocusnum** | **nsnps 2** | **hit1** | **hit2** | **PP.H0.abf** | **PP.H1.abf** | **PP.H2.abf** | **PP.H3.abf** | **PP.H4.abf** | **PP.H3+PP.H4≥0.8** | **PP.H3/PP.H4≥3** | **PP.H4/PP.H3≥5** |
| n | | | | | | | | | | | | | |
| AD Wightman | eGFR | | | rs2075650 | rs157592 | 0.00E+00 | 1.01E-152 | 0.00E+00 | 1.00E+00 | 2.24E-101 | 1 | 0 | 0 |
| AD Wightman | eGFR | | | rs11556505 | rs157592 | 0.00E+00 | 1.01E-152 | 0.00E+00 | 1.00E+00 | 9.27E-102 | 1 | 0 | 0 |
| AD Wightman | eGFR | | | rs429358 | rs66807996 | 0.00E+00 | 1.20E-05 | 0.00E+00 | 1.00E+00 | 1.12E-07 | 1 | 0 | 0 |
| AD Wightman | eGFR | | | rs769449 | rs66807996 | 0.00E+00 | 1.20E-05 | 0.00E+00 | 1.00E+00 | 9.68E-08 | 1 | 0 | 0 |
| AD Wightman | eGFR | | | rs34404554 | rs66807996 | 0.00E+00 | 1.20E-05 | 0.00E+00 | 1.00E+00 | 8.94E-08 | 1 | 0 | 0 |
| AD Wightman | eGFR | | | rs6857 | rs66807996 | 0.00E+00 | 1.20E-05 | 0.00E+00 | 1.00E+00 | 8.91E-08 | 1 | 0 | 0 |
| AD Wightman | eGFR | | | rs2075650 | rs66807996 | 0.00E+00 | 1.20E-05 | 0.00E+00 | 1.00E+00 | 9.00E-08 | 1 | 0 | 0 |
| AD Wightman | eGFR | | | rs11556505 | rs66807996 | 0.00E+00 | 1.20E-05 | 0.00E+00 | 1.00E+00 | 8.90E-08 | 1 | 0 | 0 |
| AD Wightman | eGFR | | | rs429358 | rs34342646 | 0.00E+00 | 2.61E-21 | 0.00E+00 | 1.00E+00 | 4.59E-10 | 1 | 0 | 0 |
| AD Wightman | eGFR | | | rs769449 | rs34342646 | 0.00E+00 | 2.61E-21 | 0.00E+00 | 1.00E+00 | 3.27E-10 | 1 | 0 | 0 |
| AD Wightman | eGFR | | | rs34404554 | rs34342646 | 0.00E+00 | 2.57E-21 | 0.00E+00 | 9.86E-01 | 1.43E-02 | 1 | 0 | 0 |
| AD Wightman | eGFR | | | rs6857 | rs34342646 | 0.00E+00 | 2.61E-21 | 0.00E+00 | 1.00E+00 | 2.46E-06 | 1 | 0 | 0 |
| AD Wightman | eGFR | | | rs2075650 | rs34342646 | 0.00E+00 | 2.58E-21 | 0.00E+00 | 9.90E-01 | 1.01E-02 | 1 | 0 | 0 |

| | | | coloc SUSIE method | | | | | | | | | | |
|---|---|---|---|---|---|---|---|---|---|---|---|---|---|
| **Trait1** | **Trait 2** | **cFDRlocusnum** | **nsnps 2** | **hit1** | **hit2** | **PP.H0.abf** | **PP.H1.abf** | **PP.H2.abf** | **PP.H3.abf** | **PP.H4.abf** | **PP.H3+PP.H4≥0.8** | **PP.H3/PP.H4≥3** | **PP.H4/PP.H3≥5** |
| AD Wightman | eGFR | | | rs11556505 | rs34342646 | 0.00E+00 | 2.56E-21 | 0.00E+00 | 9.80E-01 | 1.95E-02 | 1 | 0 | 0 |
| AD Wightman | eGFR | | | rs429358 | rs375972689 | 0.00E+00 | 5.93E-02 | 0.00E+00 | 9.39E-01 | 1.23E-03 | 1 | 0 | 0 |
| AD Wightman | eGFR | | | rs769449 | rs375972689 | 0.00E+00 | 5.94E-02 | 0.00E+00 | 9.40E-01 | 8.44E-04 | 1 | 0 | 0 |
| AD Wightman | eGFR | | | rs34404554 | rs375972689 | 0.00E+00 | 5.94E-02 | 0.00E+00 | 9.40E-01 | 6.41E-04 | 1 | 0 | 0 |
| AD Wightman | eGFR | | | rs6857 | rs375972689 | 0.00E+00 | 5.94E-02 | 0.00E+00 | 9.40E-01 | 6.48E-04 | 1 | 0 | 0 |
| AD Wightman | eGFR | | | rs2075650 | rs375972689 | 0.00E+00 | 5.94E-02 | 0.00E+00 | 9.40E-01 | 6.56E-04 | 1 | 0 | 0 |
| AD Wightman | eGFR | | | rs11556505 | rs375972689 | 0.00E+00 | 5.94E-02 | 0.00E+00 | 9.40E-01 | 6.31E-04 | 1 | 0 | 0 |
| AD Wightman | eGFR | 42 | 3527 | rs429358 | rs814573 | 0.00E+00 | 4.94E-324 | 0.00E+00 | 1.00E+00 | 3.21E-98 | 1 | 0 | 0 |
| AD Wightman | eGFR | | | rs769449 | rs814573 | 0.00E+00 | 4.94E-324 | 0.00E+00 | 1.00E+00 | 1.84E-134 | 1 | 0 | 0 |
| AD Wightman | eGFR | | | rs34404554 | rs814573 | 0.00E+00 | 4.94E-324 | 0.00E+00 | 1.00E+00 | 1.56E-197 | 1 | 0 | 0 |
| AD Wightman | eGFR | | | rs6857 | rs814573 | 0.00E+00 | 4.94E-324 | 0.00E+00 | 1.00E+00 | 8.40E-160 | 1 | 0 | 0 |
| AD Wightman | eGFR | | | rs2075650 | rs814573 | 0.00E+00 | 4.94E-324 | 0.00E+00 | 1.00E+00 | 2.15E-197 | 1 | 0 | 0 |
| AD Wightman | eGFR | | | rs11556505 | rs814573 | 0.00E+00 | 4.94E-324 | 0.00E+00 | 1.00E+00 | 6.84E-198 | 1 | 0 | 0 |

| | | | coloc SUSIE method | | | | | | | | | | |
|---|---|---|---|---|---|---|---|---|---|---|---|---|---|
| **Trait1** | **Trait 2** | **cFDRlocusnum** | **nsnps 2** | **hit1** | **hit2** | **PP.H0.abf** | **PP.H1.abf** | **PP.H2.abf** | **PP.H3.abf** | **PP.H4.abf** | **PP.H3+PP.H4≥0.8** | **PP.H3/PP.H4≥3** | **PP.H4/PP.H3≥5** |
| AD Wightman | eGFR | | | rs429358 | rs4420638 | 0.00E+00 | 0.00E+00 | 0.00E+00 | 1.00E+00 | 3.39E-256 | 1 | 0 | 0 |
| AD Wightman | eGFR | | | rs769449 | rs4420638 | 0.00E+00 | 0.00E+00 | 0.00E+00 | 1.00E+00 | 0.00E+00 | 1 | 0 | 0 |
| AD Wightman | eGFR | | | rs34404554 | rs4420638 | 0.00E+00 | 0.00E+00 | 0.00E+00 | 1.00E+00 | 0.00E+00 | 1 | 0 | 0 |
| AD Wightman | eGFR | | | rs6857 | rs4420638 | 0.00E+00 | 0.00E+00 | 0.00E+00 | 1.00E+00 | 0.00E+00 | 1 | 0 | 0 |
| AD Wightman | eGFR | | | rs2075650 | rs4420638 | 0.00E+00 | 0.00E+00 | 0.00E+00 | 1.00E+00 | 0.00E+00 | 1 | 0 | 0 |
| AD Wightman | eGFR | | | rs11556505 | rs4420638 | 0.00E+00 | 0.00E+00 | 0.00E+00 | 1.00E+00 | 0.00E+00 | 1 | 0 | 0 |
| AD Wightman | eGFR | | | rs429358 | rs283811 | 0.00E+00 | 0.00E+00 | 0.00E+00 | 1.00E+00 | 0.00E+00 | 1 | 0 | 0 |
| AD Wightman | eGFR | | | rs769449 | rs283811 | 0.00E+00 | 0.00E+00 | 0.00E+00 | 1.00E+00 | 0.00E+00 | 1 | 0 | 0 |
| AD Wightman | eGFR | | | rs34404554 | rs283811 | 0.00E+00 | 0.00E+00 | 0.00E+00 | 1.00E+00 | 0.00E+00 | 1 | 0 | 0 |
| AD Wightman | eGFR | | | rs6857 | rs283811 | 0.00E+00 | 0.00E+00 | 0.00E+00 | 1.00E+00 | 0.00E+00 | 1 | 0 | 0 |
| AD Wightman | eGFR | | | rs2075650 | rs283811 | 0.00E+00 | 0.00E+00 | 0.00E+00 | 1.00E+00 | 0.00E+00 | 1 | 0 | 0 |
| AD Wightman | eGFR | | | rs11556505 | rs283811 | 0.00E+00 | 0.00E+00 | 0.00E+00 | 1.00E+00 | 0.00E+00 | 1 | 0 | 0 |
| AD Wightman | eGFR | | | rs429358 | rs283815 | 0.00E+00 | 0.00E+00 | 0.00E+00 | 1.00E+00 | 0.00E+00 | 1 | 0 | 0 |
| AD | eGFR | | | rs769449 | rs283815 | 0.00E+00 | 0.00E+00 | 0.00E+00 | 1.00E+00 | 0.00E+00 | 1 | 0 | 0 |

| | | | coloc SUSIE method | | | | | | | | | | |
|---|---|---|---|---|---|---|---|---|---|---|---|---|---|
| Trait1 | Trait 2 | cFDRlocusnum | nsnps 2 | hit1 | hit2 | PP.H0.abf | PP.H1.abf | PP.H2.abf | PP.H3.abf | PP.H4.abf | PP.H3+PP.H4≥0.8 | PP.H3/PP.H4≥3 | PP.H4/PP.H3≥5 |
| Wightman | | | | | | | | | | | | | |
| AD Wightman | eGFR | | | rs34404554 | rs283815 | 0.00E+00 | 0.00E+00 | 0.00E+00 | 1.00E+00 | 0.00E+00 | 1 | 0 | 0 |
| AD Wightman | eGFR | | | rs6857 | rs283815 | 0.00E+00 | 0.00E+00 | 0.00E+00 | 1.00E+00 | 0.00E+00 | 1 | 0 | 0 |
| AD Wightman | eGFR | | | rs2075650 | rs283815 | 0.00E+00 | 0.00E+00 | 0.00E+00 | 1.00E+00 | 0.00E+00 | 1 | 0 | 0 |
| AD Wightman | eGFR | | | rs11556505 | rs283815 | 0.00E+00 | 0.00E+00 | 0.00E+00 | 1.00E+00 | 0.00E+00 | 1 | 0 | 0 |
| AD Wightman | eGFR | | | rs429358 | rs157592 | 0.00E+00 | 1.05E-152 | 0.00E+00 | 1.00E+00 | 5.75E-60 | 1 | 0 | 0 |
| AD Wightman | eGFR | | | rs769449 | rs157592 | 0.00E+00 | 1.05E-152 | 0.00E+00 | 1.00E+00 | 1.72E-72 | 1 | 0 | 0 |
| AD Wightman | eGFR | | | rs34404554 | rs157592 | 0.00E+00 | 1.05E-152 | 0.00E+00 | 1.00E+00 | 1.61E-101 | 1 | 0 | 0 |
| AD Wightman | eGFR | | | rs6857 | rs157592 | 0.00E+00 | 1.05E-152 | 0.00E+00 | 1.00E+00 | 8.24E-84 | 1 | 0 | 0 |
| AD Wightman | eGFR | | | rs2075650 | rs157592 | 0.00E+00 | 1.05E-152 | 0.00E+00 | 1.00E+00 | 2.30E-101 | 1 | 0 | 0 |
| AD Wightman | eGFR | | | rs11556505 | rs157592 | 0.00E+00 | 1.05E-152 | 0.00E+00 | 1.00E+00 | 9.53E-102 | 1 | 0 | 0 |
| AD Wightman | eGFR | | | rs429358 | rs66807996 | 0.00E+00 | 1.20E-05 | 0.00E+00 | 1.00E+00 | 1.12E-07 | 1 | 0 | 0 |
| AD Wightman | eGFR | | | rs769449 | rs66807996 | 0.00E+00 | 1.20E-05 | 0.00E+00 | 1.00E+00 | 9.74E-08 | 1 | 0 | 0 |
| AD Wightma | eGFR | | | rs34404554 | rs66807996 | 0.00E+00 | 1.20E-05 | 0.00E+00 | 1.00E+00 | 8.99E-08 | 1 | 0 | 0 |

| | | | coloc SUSIE method | | | | | | | | | | |
|---|---|---|---|---|---|---|---|---|---|---|---|---|---|
| **Trait1** | **Trait2** | **cFDRlocusnum** | **nsnps2** | **hit1** | **hit2** | **PP.H0.abf** | **PP.H1.abf** | **PP.H2.abf** | **PP.H3.abf** | **PP.H4.abf** | **PP.H3+PP.H4≥0.8** | **PP.H3/PP.H4≥3** | **PP.H4/PP.H3≥5** |
| n | | | | | | | | | | | | | |
| AD Wightman | eGFR | | | rs6857 | rs66807996 | 0.00E+00 | 1.20E-05 | 0.00E+00 | 1.00E+00 | 8.96E-08 | 1 | 0 | 0 |
| AD Wightman | eGFR | | | rs2075650 | rs66807996 | 0.00E+00 | 1.20E-05 | 0.00E+00 | 1.00E+00 | 9.05E-08 | 1 | 0 | 0 |
| AD Wightman | eGFR | | | rs11556505 | rs66807996 | 0.00E+00 | 1.20E-05 | 0.00E+00 | 1.00E+00 | 8.95E-08 | 1 | 0 | 0 |
| AD Wightman | eGFR | | | rs429358 | rs34342646 | 0.00E+00 | 2.67E-21 | 0.00E+00 | 1.00E+00 | 4.65E-10 | 1 | 0 | 0 |
| AD Wightman | eGFR | | | rs769449 | rs34342646 | 0.00E+00 | 2.67E-21 | 0.00E+00 | 1.00E+00 | 3.30E-10 | 1 | 0 | 0 |
| AD Wightman | eGFR | | | rs34404554 | rs34342646 | 0.00E+00 | 2.63E-21 | 0.00E+00 | 9.86E-01 | 1.43E-02 | 1 | 0 | 0 |
| AD Wightman | eGFR | | | rs6857 | rs34342646 | 0.00E+00 | 2.67E-21 | 0.00E+00 | 1.00E+00 | 2.47E-06 | 1 | 0 | 0 |
| AD Wightman | eGFR | | | rs2075650 | rs34342646 | 0.00E+00 | 2.64E-21 | 0.00E+00 | 9.90E-01 | 1.01E-02 | 1 | 0 | 0 |
| AD Wightman | eGFR | | | rs11556505 | rs34342646 | 0.00E+00 | 2.62E-21 | 0.00E+00 | 9.80E-01 | 1.95E-02 | 1 | 0 | 0 |
| AD Wightman | eGFR | | | rs429358 | rs375972689 | 0.00E+00 | 5.91E-02 | 0.00E+00 | 9.40E-01 | 1.23E-03 | 1 | 0 | 0 |
| AD Wightman | eGFR | | | rs769449 | rs375972689 | 0.00E+00 | 5.91E-02 | 0.00E+00 | 9.40E-01 | 8.40E-04 | 1 | 0 | 0 |
| AD Wightman | eGFR | | | rs34404554 | rs375972689 | 0.00E+00 | 5.91E-02 | 0.00E+00 | 9.40E-01 | 6.38E-04 | 1 | 0 | 0 |
| AD Wightman | eGFR | | | rs6857 | rs375972689 | 0.00E+00 | 5.91E-02 | 0.00E+00 | 9.40E-01 | 6.45E-04 | 1 | 0 | 0 |

| | | | coloc SUSIE method | | | | | | | | | | |
|---|---|---|---|---|---|---|---|---|---|---|---|---|---|
| **Trait1** | **Trait 2** | **cFDRlocusnum** | **nsnps 2** | **hit1** | **hit2** | **PP.H0.abf** | **PP.H1.abf** | **PP.H2.abf** | **PP.H3.abf** | **PP.H4.abf** | **PP.H3+PP.H4≥0.8** | **PP.H3/PP.H4≥3** | **PP.H4/PP.H3≥5** |
| AD Wightman | eGFR | | | rs2075650 | rs375972689 | 0.00E+00 | 5.91E-02 | 0.00E+00 | 9.40E-01 | 6.53E-04 | 1 | 0 | 0 |
| AD Wightman | eGFR | | | rs11556505 | rs375972689 | 0.00E+00 | 5.91E-02 | 0.00E+00 | 9.40E-01 | 6.29E-04 | 1 | 0 | 0 |
| AD Wightman | eGFR | 43a | 3543 | rs429358 | rs814573 | 0.00E+00 | 4.94E-324 | 0.00E+00 | 1.00E+00 | 3.22E-98 | 1 | 0 | 0 |
| AD Wightman | eGFR | | | rs769449 | rs814573 | 0.00E+00 | 4.94E-324 | 0.00E+00 | 1.00E+00 | 1.86E-134 | 1 | 0 | 0 |
| AD Wightman | eGFR | | | rs34404554 | rs814573 | 0.00E+00 | 4.94E-324 | 0.00E+00 | 1.00E+00 | 1.57E-197 | 1 | 0 | 0 |
| AD Wightman | eGFR | | | rs6857 | rs814573 | 0.00E+00 | 4.94E-324 | 0.00E+00 | 1.00E+00 | 8.44E-160 | 1 | 0 | 0 |
| AD Wightman | eGFR | | | rs2075650 | rs814573 | 0.00E+00 | 4.94E-324 | 0.00E+00 | 1.00E+00 | 2.18E-197 | 1 | 0 | 0 |
| AD Wightman | eGFR | | | rs11556505 | rs814573 | 0.00E+00 | 4.94E-324 | 0.00E+00 | 1.00E+00 | 6.91E-198 | 1 | 0 | 0 |
| AD Wightman | eGFR | | | rs429358 | rs4420638 | 0.00E+00 | 0.00E+00 | 0.00E+00 | 1.00E+00 | 3.41E-256 | 1 | 0 | 0 |
| AD Wightman | eGFR | | | rs769449 | rs4420638 | 0.00E+00 | 0.00E+00 | 0.00E+00 | 1.00E+00 | 0.00E+00 | 1 | 0 | 0 |
| AD Wightman | eGFR | | | rs34404554 | rs4420638 | 0.00E+00 | 0.00E+00 | 0.00E+00 | 1.00E+00 | 0.00E+00 | 1 | 0 | 0 |
| AD Wightman | eGFR | | | rs6857 | rs4420638 | 0.00E+00 | 0.00E+00 | 0.00E+00 | 1.00E+00 | 0.00E+00 | 1 | 0 | 0 |
| AD Wightman | eGFR | | | rs2075650 | rs4420638 | 0.00E+00 | 0.00E+00 | 0.00E+00 | 1.00E+00 | 0.00E+00 | 1 | 0 | 0 |

| | | | coloc SUSIE method | | | | | | | | | | |
|---|---|---|---|---|---|---|---|---|---|---|---|---|---|
| **Trait1** | **Trait 2** | **cFDRlocusnum** | **nsnps 2** | **hit1** | **hit2** | **PP.H0.abf** | **PP.H1.abf** | **PP.H2.abf** | **PP.H3.abf** | **PP.H4.abf** | **PP.H3+PP.H4≥0.8** | **PP.H3/PP.H4≥3** | **PP.H4/PP.H3≥5** |
| AD Wightman | eGFR | | | rs11556505 | rs4420638 | 0.00E+00 | 0.00E+00 | 0.00E+00 | 1.00E+00 | 0.00E+00 | 1 | 0 | 0 |
| AD Wightman | eGFR | | | rs429358 | rs283811 | 0.00E+00 | 0.00E+00 | 0.00E+00 | 1.00E+00 | 0.00E+00 | 1 | 0 | 0 |
| AD Wightman | eGFR | | | rs769449 | rs283811 | 0.00E+00 | 0.00E+00 | 0.00E+00 | 1.00E+00 | 0.00E+00 | 1 | 0 | 0 |
| AD Wightman | eGFR | | | rs34404554 | rs283811 | 0.00E+00 | 0.00E+00 | 0.00E+00 | 1.00E+00 | 0.00E+00 | 1 | 0 | 0 |
| AD Wightman | eGFR | | | rs6857 | rs283811 | 0.00E+00 | 0.00E+00 | 0.00E+00 | 1.00E+00 | 0.00E+00 | 1 | 0 | 0 |
| AD Wightman | eGFR | | | rs2075650 | rs283811 | 0.00E+00 | 0.00E+00 | 0.00E+00 | 1.00E+00 | 0.00E+00 | 1 | 0 | 0 |
| AD Wightman | eGFR | | | rs11556505 | rs283811 | 0.00E+00 | 0.00E+00 | 0.00E+00 | 1.00E+00 | 0.00E+00 | 1 | 0 | 0 |
| AD Wightman | eGFR | | | rs429358 | rs283815 | 0.00E+00 | 0.00E+00 | 0.00E+00 | 1.00E+00 | 0.00E+00 | 1 | 0 | 0 |
| AD Wightman | eGFR | | | rs769449 | rs283815 | 0.00E+00 | 0.00E+00 | 0.00E+00 | 1.00E+00 | 0.00E+00 | 1 | 0 | 0 |
| AD Wightman | eGFR | | | rs34404554 | rs283815 | 0.00E+00 | 0.00E+00 | 0.00E+00 | 1.00E+00 | 0.00E+00 | 1 | 0 | 0 |
| AD Wightman | eGFR | | | rs6857 | rs283815 | 0.00E+00 | 0.00E+00 | 0.00E+00 | 1.00E+00 | 0.00E+00 | 1 | 0 | 0 |
| AD Wightman | eGFR | | | rs2075650 | rs283815 | 0.00E+00 | 0.00E+00 | 0.00E+00 | 1.00E+00 | 0.00E+00 | 1 | 0 | 0 |
| AD Wightman | eGFR | | | rs11556505 | rs283815 | 0.00E+00 | 0.00E+00 | 0.00E+00 | 1.00E+00 | 0.00E+00 | 1 | 0 | 0 |
| AD | eGFR | | | rs429358 | rs157592 | 0.00E+00 | 1.04E- | 0.00E+00 | 1.00E+00 | 5.74E-60 | 1 | 0 | 0 |

| | | | coloc SUSIE method | | | | | | | | | | |
|---|---|---|---|---|---|---|---|---|---|---|---|---|---|
| **Trait1** | **Trait 2** | **cFDRlocusnum** | **nsnps 2** | **hit1** | **hit2** | **PP.H0.abf** | **PP.H1.abf** | **PP.H2.abf** | **PP.H3.abf** | **PP.H4.abf** | **PP.H3+PP.H4≥0.8** | **PP.H3/PP.H4≥3** | **PP.H4/PP.H3≥5** |
| Wightman | | | | | | | 152 | | | | | | |
| AD Wightman | eGFR | | | rs769449 | rs157592 | 0.00E+00 | 1.04E-152 | 0.00E+00 | 1.00E+00 | 1.73E-72 | 1 | 0 | 0 |
| AD Wightman | eGFR | | | rs34404554 | rs157592 | 0.00E+00 | 1.04E-152 | 0.00E+00 | 1.00E+00 | 1.61E-101 | 1 | 0 | 0 |
| AD Wightman | eGFR | | | rs6857 | rs157592 | 0.00E+00 | 1.04E-152 | 0.00E+00 | 1.00E+00 | 8.23E-84 | 1 | 0 | 0 |
| AD Wightman | eGFR | | | rs2075650 | rs157592 | 0.00E+00 | 1.04E-152 | 0.00E+00 | 1.00E+00 | 2.31E-101 | 1 | 0 | 0 |
| AD Wightman | eGFR | | | rs11556505 | rs157592 | 0.00E+00 | 1.04E-152 | 0.00E+00 | 1.00E+00 | 9.53E-102 | 1 | 0 | 0 |
| AD Wightman | eGFR | | | rs429358 | rs66807996 | 0.00E+00 | 1.21E-05 | 0.00E+00 | 1.00E+00 | 1.13E-07 | 1 | 0 | 0 |
| AD Wightman | eGFR | | | rs769449 | rs66807996 | 0.00E+00 | 1.21E-05 | 0.00E+00 | 1.00E+00 | 9.78E-08 | 1 | 0 | 0 |
| AD Wightman | eGFR | | | rs34404554 | rs66807996 | 0.00E+00 | 1.21E-05 | 0.00E+00 | 1.00E+00 | 9.02E-08 | 1 | 0 | 0 |
| AD Wightman | eGFR | | | rs6857 | rs66807996 | 0.00E+00 | 1.21E-05 | 0.00E+00 | 1.00E+00 | 8.99E-08 | 1 | 0 | 0 |
| AD Wightman | eGFR | | | rs2075650 | rs66807996 | 0.00E+00 | 1.21E-05 | 0.00E+00 | 1.00E+00 | 9.09E-08 | 1 | 0 | 0 |
| AD Wightman | eGFR | | | rs11556505 | rs66807996 | 0.00E+00 | 1.21E-05 | 0.00E+00 | 1.00E+00 | 8.98E-08 | 1 | 0 | 0 |
| AD Wightman | eGFR | | | rs429358 | rs34342646 | 0.00E+00 | 2.72E-21 | 0.00E+00 | 1.00E+00 | 4.69E-10 | 1 | 0 | 0 |
| AD Wightma | eGFR | | | rs769449 | rs34342646 | 0.00E+00 | 2.72E-21 | 0.00E+00 | 1.00E+00 | 3.32E-10 | 1 | 0 | 0 |

| | | | coloc SUSIE method | | | | | | | | | | |
|---|---|---|---|---|---|---|---|---|---|---|---|---|---|
| **Trait1** | **Trait 2** | **cFDRlocusnum** | **nsnps 2** | **hit1** | **hit2** | **PP.H0.abf** | **PP.H1.abf** | **PP.H2.abf** | **PP.H3.abf** | **PP.H4.abf** | **PP.H3+PP.H4≥0.8** | **PP.H3/PP.H4≥3** | **PP.H4/PP.H3≥5** |
| n | | | | | | | | | | | | | |
| AD Wightman | eGFR | | | rs34404554 | rs34342646 | 0.00E+00 | 2.68E-21 | 0.00E+00 | 9.86E-01 | 1.43E-02 | 1 | 0 | 0 |
| AD Wightman | eGFR | | | rs6857 | rs34342646 | 0.00E+00 | 2.72E-21 | 0.00E+00 | 1.00E+00 | 2.48E-06 | 1 | 0 | 0 |
| AD Wightman | eGFR | | | rs2075650 | rs34342646 | 0.00E+00 | 2.69E-21 | 0.00E+00 | 9.90E-01 | 1.01E-02 | 1 | 0 | 0 |
| AD Wightman | eGFR | | | rs11556505 | rs34342646 | 0.00E+00 | 2.67E-21 | 0.00E+00 | 9.80E-01 | 1.95E-02 | 1 | 0 | 0 |
| AD Wightman | eGFR | | | rs429358 | rs375972689 | 0.00E+00 | 5.89E-02 | 0.00E+00 | 9.40E-01 | 1.22E-03 | 1 | 0 | 0 |
| AD Wightman | eGFR | | | rs769449 | rs375972689 | 0.00E+00 | 5.89E-02 | 0.00E+00 | 9.40E-01 | 8.38E-04 | 1 | 0 | 0 |
| AD Wightman | eGFR | | | rs34404554 | rs375972689 | 0.00E+00 | 5.89E-02 | 0.00E+00 | 9.40E-01 | 6.36E-04 | 1 | 0 | 0 |
| AD Wightman | eGFR | | | rs6857 | rs375972689 | 0.00E+00 | 5.89E-02 | 0.00E+00 | 9.40E-01 | 6.43E-04 | 1 | 0 | 0 |
| AD Wightman | eGFR | | | rs2075650 | rs375972689 | 0.00E+00 | 5.89E-02 | 0.00E+00 | 9.40E-01 | 6.51E-04 | 1 | 0 | 0 |
| AD Wightman | eGFR | | | rs11556505 | rs375972689 | 0.00E+00 | 5.89E-02 | 0.00E+00 | 9.40E-01 | 6.27E-04 | 1 | 0 | 0 |
| AD Wightman | eGFR | 43b | 3554 | rs429358 | rs814573 | 0.00E+00 | 4.94E-324 | 0.00E+00 | 1.00E+00 | 3.14E-98 | 1 | 0 | 0 |
| AD Wightman | eGFR | | | rs769449 | rs814573 | 0.00E+00 | 4.94E-324 | 0.00E+00 | 1.00E+00 | 1.83E-134 | 1 | 0 | 0 |
| AD Wightma | eGFR | | | rs34404554 | rs814573 | 0.00E+00 | 4.94E-324 | 0.00E+00 | 1.00E+00 | 1.53E-197 | 1 | 0 | 0 |

| | | | coloc SUSIE method | | | | | | | | | | |
|---|---|---|---|---|---|---|---|---|---|---|---|---|---|
| **Trait1** | **Trait 2** | **cFDRlocusnum** | **nsnps 2** | **hit1** | **hit2** | **PP.H0.abf** | **PP.H1.abf** | **PP.H2.abf** | **PP.H3.abf** | **PP.H4.abf** | **PP.H3+PP.H4≥0.8** | **PP.H3/PP.H4≥3** | **PP.H4/PP.H3≥5** |
| n | | | | | | | | | | | | | |
| AD Wightman | eGFR | | | rs6857 | rs814573 | 0.00E+00 | 4.94E-324 | 0.00E+00 | 1.00E+00 | 8.10E-160 | 1 | 0 | 0 |
| AD Wightman | eGFR | | | rs2075650 | rs814573 | 0.00E+00 | 4.94E-324 | 0.00E+00 | 1.00E+00 | 2.11E-197 | 1 | 0 | 0 |
| AD Wightman | eGFR | | | rs11556505 | rs814573 | 0.00E+00 | 4.94E-324 | 0.00E+00 | 1.00E+00 | 6.70E-198 | 1 | 0 | 0 |
| AD Wightman | eGFR | | | rs429358 | rs4420638 | 0.00E+00 | 0.00E+00 | 0.00E+00 | 1.00E+00 | 3.38E-256 | 1 | 0 | 0 |
| AD Wightman | eGFR | | | rs769449 | rs4420638 | 0.00E+00 | 0.00E+00 | 0.00E+00 | 1.00E+00 | 0.00E+00 | 1 | 0 | 0 |
| AD Wightman | eGFR | | | rs34404554 | rs4420638 | 0.00E+00 | 0.00E+00 | 0.00E+00 | 1.00E+00 | 0.00E+00 | 1 | 0 | 0 |
| AD Wightman | eGFR | | | rs6857 | rs4420638 | 0.00E+00 | 0.00E+00 | 0.00E+00 | 1.00E+00 | 0.00E+00 | 1 | 0 | 0 |
| AD Wightman | eGFR | | | rs2075650 | rs4420638 | 0.00E+00 | 0.00E+00 | 0.00E+00 | 1.00E+00 | 0.00E+00 | 1 | 0 | 0 |
| AD Wightman | eGFR | | | rs11556505 | rs4420638 | 0.00E+00 | 0.00E+00 | 0.00E+00 | 1.00E+00 | 0.00E+00 | 1 | 0 | 0 |
| AD Wightman | eGFR | | | rs429358 | rs283811 | 0.00E+00 | 0.00E+00 | 0.00E+00 | 1.00E+00 | 0.00E+00 | 1 | 0 | 0 |
| AD Wightman | eGFR | | | rs769449 | rs283811 | 0.00E+00 | 0.00E+00 | 0.00E+00 | 1.00E+00 | 0.00E+00 | 1 | 0 | 0 |
| AD Wightman | eGFR | | | rs34404554 | rs283811 | 0.00E+00 | 0.00E+00 | 0.00E+00 | 1.00E+00 | 0.00E+00 | 1 | 0 | 0 |
| AD Wightman | eGFR | | | rs6857 | rs283811 | 0.00E+00 | 0.00E+00 | 0.00E+00 | 1.00E+00 | 0.00E+00 | 1 | 0 | 0 |

| | | | coloc SUSIE method | | | | | | | | | | |
|---|---|---|---|---|---|---|---|---|---|---|---|---|---|
| **Trait1** | **Trait 2** | **cFDRlocusnum** | **nsnps 2** | **hit1** | **hit2** | **PP.H0.abf** | **PP.H1.abf** | **PP.H2.abf** | **PP.H3.abf** | **PP.H4.abf** | **PP.H3+PP.H4≥0.8** | **PP.H3/PP.H4≥3** | **PP.H4/PP.H3≥5** |
| AD Wightman | eGFR | | | rs2075650 | rs283811 | 0.00E+00 | 0.00E+00 | 0.00E+00 | 1.00E+00 | 0.00E+00 | 1 | 0 | 0 |
| AD Wightman | eGFR | | | rs11556505 | rs283811 | 0.00E+00 | 0.00E+00 | 0.00E+00 | 1.00E+00 | 0.00E+00 | 1 | 0 | 0 |
| AD Wightman | eGFR | | | rs429358 | rs283815 | 0.00E+00 | 0.00E+00 | 0.00E+00 | 1.00E+00 | 0.00E+00 | 1 | 0 | 0 |
| AD Wightman | eGFR | | | rs769449 | rs283815 | 0.00E+00 | 0.00E+00 | 0.00E+00 | 1.00E+00 | 0.00E+00 | 1 | 0 | 0 |
| AD Wightman | eGFR | | | rs34404554 | rs283815 | 0.00E+00 | 0.00E+00 | 0.00E+00 | 1.00E+00 | 0.00E+00 | 1 | 0 | 0 |
| AD Wightman | eGFR | | | rs6857 | rs283815 | 0.00E+00 | 0.00E+00 | 0.00E+00 | 1.00E+00 | 0.00E+00 | 1 | 0 | 0 |
| AD Wightman | eGFR | | | rs2075650 | rs283815 | 0.00E+00 | 0.00E+00 | 0.00E+00 | 1.00E+00 | 0.00E+00 | 1 | 0 | 0 |
| AD Wightman | eGFR | | | rs11556505 | rs283815 | 0.00E+00 | 0.00E+00 | 0.00E+00 | 1.00E+00 | 0.00E+00 | 1 | 0 | 0 |
| AD Wightman | eGFR | | | rs429358 | rs157592 | 0.00E+00 | 1.10E-152 | 0.00E+00 | 1.00E+00 | 5.85E-60 | 1 | 0 | 0 |
| AD Wightman | eGFR | | | rs769449 | rs157592 | 0.00E+00 | 1.10E-152 | 0.00E+00 | 1.00E+00 | 1.79E-72 | 1 | 0 | 0 |
| AD Wightman | eGFR | | | rs34404554 | rs157592 | 0.00E+00 | 1.10E-152 | 0.00E+00 | 1.00E+00 | 1.69E-101 | 1 | 0 | 0 |
| AD Wightman | eGFR | | | rs6857 | rs157592 | 0.00E+00 | 1.10E-152 | 0.00E+00 | 1.00E+00 | 8.46E-84 | 1 | 0 | 0 |
| AD Wightman | eGFR | | | rs2075650 | rs157592 | 0.00E+00 | 1.10E-152 | 0.00E+00 | 1.00E+00 | 2.41E-101 | 1 | 0 | 0 |
| AD | eGFR | | | rs11556505 | rs157592 | 0.00E+00 | 1.10E- | 0.00E+00 | 1.00E+00 | 9.97E- | 1 | 0 | 0 |

| | | | coloc SUSIE method | | | | | | | | | | |
|---|---|---|---|---|---|---|---|---|---|---|---|---|---|
| **Trait1** | **Trait 2** | **cFDRlocusnu m** | **nsnps 2** | **hit1** | **hit2** | **PP.H0.ab f** | **PP.H1.ab f** | **PP.H2.ab f** | **PP.H3.ab f** | **PP.H4.ab f** | **PP.H3+PP.H4≥0. 8** | **PP.H3/PP.H4≥ 3** | **PP.H4/PP.H3≥ 5** |
| Wightma n | | | | | | | 152 | | | 102 | | | |
| AD Wightma n | eGFR | | | rs429358 | rs66807996 | 0.00E+00 | 1.21E-05 | 0.00E+00 | 1.00E+00 | 1.13E-07 | 1 | 0 | 0 |
| AD Wightma n | eGFR | | | rs769449 | rs66807996 | 0.00E+00 | 1.21E-05 | 0.00E+00 | 1.00E+00 | 9.76E-08 | 1 | 0 | 0 |
| AD Wightma n | eGFR | | | rs34404554 | rs66807996 | 0.00E+00 | 1.21E-05 | 0.00E+00 | 1.00E+00 | 9.00E-08 | 1 | 0 | 0 |
| AD Wightma n | eGFR | | | rs6857 | rs66807996 | 0.00E+00 | 1.21E-05 | 0.00E+00 | 1.00E+00 | 8.97E-08 | 1 | 0 | 0 |
| AD Wightma n | eGFR | | | rs2075650 | rs66807996 | 0.00E+00 | 1.21E-05 | 0.00E+00 | 1.00E+00 | 9.07E-08 | 1 | 0 | 0 |
| AD Wightma n | eGFR | | | rs11556505 | rs66807996 | 0.00E+00 | 1.21E-05 | 0.00E+00 | 1.00E+00 | 8.96E-08 | 1 | 0 | 0 |
| AD Wightma n | eGFR | | | rs429358 | rs34342646 | 0.00E+00 | 2.82E-21 | 0.00E+00 | 1.00E+00 | 4.78E-10 | 1 | 0 | 0 |
| AD Wightma n | eGFR | | | rs769449 | rs34342646 | 0.00E+00 | 2.82E-21 | 0.00E+00 | 1.00E+00 | 3.36E-10 | 1 | 0 | 0 |
| AD Wightma n | eGFR | | | rs34404554 | rs34342646 | 0.00E+00 | 2.78E-21 | 0.00E+00 | 9.86E-01 | 1.43E-02 | 1 | 0 | 0 |
| AD Wightma n | eGFR | | | rs6857 | rs34342646 | 0.00E+00 | 2.82E-21 | 0.00E+00 | 1.00E+00 | 2.51E-06 | 1 | 0 | 0 |
| AD Wightma n | eGFR | | | rs2075650 | rs34342646 | 0.00E+00 | 2.80E-21 | 0.00E+00 | 9.90E-01 | 1.01E-02 | 1 | 0 | 0 |
| AD Wightma n | eGFR | | | rs11556505 | rs34342646 | 0.00E+00 | 2.77E-21 | 0.00E+00 | 9.81E-01 | 1.95E-02 | 1 | 0 | 0 |
| AD Wightma | eGFR | | | rs429358 | rs37597268 9 | 0.00E+00 | 5.89E-02 | 0.00E+00 | 9.40E-01 | 1.22E-03 | 1 | 0 | 0 |

| | | | coloc SUSIE method | | | | | | | | | | |
|---|---|---|---|---|---|---|---|---|---|---|---|---|---|
| **Trait1** | **Trait 2** | **cFDRlocusnum** | **nsnps 2** | **hit1** | **hit2** | **PP.H0.abf** | **PP.H1.abf** | **PP.H2.abf** | **PP.H3.abf** | **PP.H4.abf** | **PP.H3+PP.H4≥0.8** | **PP.H3/PP.H4≥3** | **PP.H4/PP.H3≥5** |
| n | | | | | | | | | | | | | |
| AD Wightman | eGFR | | | rs769449 | rs375972689 | 0.00E+00 | 5.89E-02 | 0.00E+00 | 9.40E-01 | 8.39E-04 | 1 | 0 | 0 |
| AD Wightman | eGFR | | | rs34404554 | rs375972689 | 0.00E+00 | 5.90E-02 | 0.00E+00 | 9.40E-01 | 6.37E-04 | 1 | 0 | 0 |
| AD Wightman | eGFR | | | rs6857 | rs375972689 | 0.00E+00 | 5.90E-02 | 0.00E+00 | 9.40E-01 | 6.43E-04 | 1 | 0 | 0 |
| AD Wightman | eGFR | | | rs2075650 | rs375972689 | 0.00E+00 | 5.90E-02 | 0.00E+00 | 9.40E-01 | 6.52E-04 | 1 | 0 | 0 |
| AD Wightman | eGFR | | | rs11556505 | rs375972689 | 0.00E+00 | 5.90E-02 | 0.00E+00 | 9.40E-01 | 6.27E-04 | 1 | 0 | 0 |
| AD Wightman | eGFR | 43c | 3543 | rs429358 | rs814573 | 0.00E+00 | 4.94E-324 | 0.00E+00 | 1.00E+00 | 3.13E-98 | 1 | 0 | 0 |
| AD Wightman | eGFR | | | rs769449 | rs814573 | 0.00E+00 | 4.94E-324 | 0.00E+00 | 1.00E+00 | 1.83E-134 | 1 | 0 | 0 |
| AD Wightman | eGFR | | | rs34404554 | rs814573 | 0.00E+00 | 4.94E-324 | 0.00E+00 | 1.00E+00 | 1.53E-197 | 1 | 0 | 0 |
| AD Wightman | eGFR | | | rs6857 | rs814573 | 0.00E+00 | 4.94E-324 | 0.00E+00 | 1.00E+00 | 8.09E-160 | 1 | 0 | 0 |
| AD Wightman | eGFR | | | rs2075650 | rs814573 | 0.00E+00 | 4.94E-324 | 0.00E+00 | 1.00E+00 | 2.11E-197 | 1 | 0 | 0 |
| AD Wightman | eGFR | | | rs11556505 | rs814573 | 0.00E+00 | 4.94E-324 | 0.00E+00 | 1.00E+00 | 6.70E-198 | 1 | 0 | 0 |
| AD Wightman | eGFR | | | rs429358 | rs4420638 | 0.00E+00 | 0.00E+00 | 0.00E+00 | 1.00E+00 | 3.38E-256 | 1 | 0 | 0 |
| AD Wightma | eGFR | | | rs769449 | rs4420638 | 0.00E+00 | 0.00E+00 | 0.00E+00 | 1.00E+00 | 0.00E+00 | 1 | 0 | 0 |

| | | | coloc SUSIE method | | | | | | | | | | |
|---|---|---|---|---|---|---|---|---|---|---|---|---|---|
| Trait1 | Trait 2 | cFDRlocusnum | nsnps 2 | hit1 | hit2 | PP.H0.abf | PP.H1.abf | PP.H2.abf | PP.H3.abf | PP.H4.abf | PP.H3+PP.H4≥0.8 | PP.H3/PP.H4≥3 | PP.H4/PP.H3≥5 |
| n | | | | | | | | | | | | | |
| AD Wightman | eGFR | | | rs34404554 | rs4420638 | 0.00E+00 | 0.00E+00 | 0.00E+00 | 1.00E+00 | 0.00E+00 | 1 | 0 | 0 |
| AD Wightman | eGFR | | | rs6857 | rs4420638 | 0.00E+00 | 0.00E+00 | 0.00E+00 | 1.00E+00 | 0.00E+00 | 1 | 0 | 0 |
| AD Wightman | eGFR | | | rs2075650 | rs4420638 | 0.00E+00 | 0.00E+00 | 0.00E+00 | 1.00E+00 | 0.00E+00 | 1 | 0 | 0 |
| AD Wightman | eGFR | | | rs11556505 | rs4420638 | 0.00E+00 | 0.00E+00 | 0.00E+00 | 1.00E+00 | 0.00E+00 | 1 | 0 | 0 |
| AD Wightman | eGFR | | | rs429358 | rs283811 | 0.00E+00 | 0.00E+00 | 0.00E+00 | 1.00E+00 | 0.00E+00 | 1 | 0 | 0 |
| AD Wightman | eGFR | | | rs769449 | rs283811 | 0.00E+00 | 0.00E+00 | 0.00E+00 | 1.00E+00 | 0.00E+00 | 1 | 0 | 0 |
| AD Wightman | eGFR | | | rs34404554 | rs283811 | 0.00E+00 | 0.00E+00 | 0.00E+00 | 1.00E+00 | 0.00E+00 | 1 | 0 | 0 |
| AD Wightman | eGFR | | | rs6857 | rs283811 | 0.00E+00 | 0.00E+00 | 0.00E+00 | 1.00E+00 | 0.00E+00 | 1 | 0 | 0 |
| AD Wightman | eGFR | | | rs2075650 | rs283811 | 0.00E+00 | 0.00E+00 | 0.00E+00 | 1.00E+00 | 0.00E+00 | 1 | 0 | 0 |
| AD Wightman | eGFR | | | rs11556505 | rs283811 | 0.00E+00 | 0.00E+00 | 0.00E+00 | 1.00E+00 | 0.00E+00 | 1 | 0 | 0 |
| AD Wightman | eGFR | | | rs429358 | rs283815 | 0.00E+00 | 0.00E+00 | 0.00E+00 | 1.00E+00 | 0.00E+00 | 1 | 0 | 0 |
| AD Wightman | eGFR | | | rs769449 | rs283815 | 0.00E+00 | 0.00E+00 | 0.00E+00 | 1.00E+00 | 0.00E+00 | 1 | 0 | 0 |
| AD Wightman | eGFR | | | rs34404554 | rs283815 | 0.00E+00 | 0.00E+00 | 0.00E+00 | 1.00E+00 | 0.00E+00 | 1 | 0 | 0 |

| | | | coloc SUSIE method | | | | | | | | | | |
|---|---|---|---|---|---|---|---|---|---|---|---|---|---|
| Trait1 | Trait 2 | cFDRlocusnum | nsnps 2 | hit1 | hit2 | PP.H0.abf | PP.H1.abf | PP.H2.abf | PP.H3.abf | PP.H4.abf | PP.H3+PP.H4≥0.8 | PP.H3/PP.H4≥3 | PP.H4/PP.H3≥5 |
| AD Wightman | eGFR | | | rs6857 | rs283815 | 0.00E+00 | 0.00E+00 | 0.00E+00 | 1.00E+00 | 0.00E+00 | 1 | 0 | 0 |
| AD Wightman | eGFR | | | rs2075650 | rs283815 | 0.00E+00 | 0.00E+00 | 0.00E+00 | 1.00E+00 | 0.00E+00 | 1 | 0 | 0 |
| AD Wightman | eGFR | | | rs11556505 | rs283815 | 0.00E+00 | 0.00E+00 | 0.00E+00 | 1.00E+00 | 0.00E+00 | 1 | 0 | 0 |
| AD Wightman | eGFR | | | rs429358 | rs157592 | 0.00E+00 | 1.10E-152 | 0.00E+00 | 1.00E+00 | 5.85E-60 | 1 | 0 | 0 |
| AD Wightman | eGFR | | | rs769449 | rs157592 | 0.00E+00 | 1.10E-152 | 0.00E+00 | 1.00E+00 | 1.79E-72 | 1 | 0 | 0 |
| AD Wightman | eGFR | | | rs34404554 | rs157592 | 0.00E+00 | 1.10E-152 | 0.00E+00 | 1.00E+00 | 1.69E-101 | 1 | 0 | 0 |
| AD Wightman | eGFR | | | rs6857 | rs157592 | 0.00E+00 | 1.10E-152 | 0.00E+00 | 1.00E+00 | 8.46E-84 | 1 | 0 | 0 |
| AD Wightman | eGFR | | | rs2075650 | rs157592 | 0.00E+00 | 1.10E-152 | 0.00E+00 | 1.00E+00 | 2.42E-101 | 1 | 0 | 0 |
| AD Wightman | eGFR | | | rs11556505 | rs157592 | 0.00E+00 | 1.10E-152 | 0.00E+00 | 1.00E+00 | 9.99E-102 | 1 | 0 | 0 |
| AD Wightman | eGFR | | | rs429358 | rs66807996 | 0.00E+00 | 1.21E-05 | 0.00E+00 | 1.00E+00 | 1.13E-07 | 1 | 0 | 0 |
| AD Wightman | eGFR | | | rs769449 | rs66807996 | 0.00E+00 | 1.21E-05 | 0.00E+00 | 1.00E+00 | 9.76E-08 | 1 | 0 | 0 |
| AD Wightman | eGFR | | | rs34404554 | rs66807996 | 0.00E+00 | 1.21E-05 | 0.00E+00 | 1.00E+00 | 9.00E-08 | 1 | 0 | 0 |
| AD Wightman | eGFR | | | rs6857 | rs66807996 | 0.00E+00 | 1.21E-05 | 0.00E+00 | 1.00E+00 | 8.97E-08 | 1 | 0 | 0 |
| AD | eGFR | | | rs2075650 | rs66807996 | 0.00E+00 | 1.21E-05 | 0.00E+00 | 1.00E+00 | 9.07E-08 | 1 | 0 | 0 |

| | | | coloc SUSIE method | | | | | | | | | | |
|---|---|---|---|---|---|---|---|---|---|---|---|---|---|
| Trait1 | Trait 2 | cFDRlocusnum | nsnps 2 | hit1 | hit2 | PP.H0.abf | PP.H1.abf | PP.H2.abf | PP.H3.abf | PP.H4.abf | PP.H3+PP.H4≥0.8 | PP.H3/PP.H4≥3 | PP.H4/PP.H3≥5 |
| Wightman | | | | | | | | | | | | | |
| AD Wightman | eGFR | | | rs11556505 | rs66807996 | 0.00E+00 | 1.21E-05 | 0.00E+00 | 1.00E+00 | 8.96E-08 | 1 | 0 | 0 |
| AD Wightman | eGFR | | | rs429358 | rs34342646 | 0.00E+00 | 2.83E-21 | 0.00E+00 | 1.00E+00 | 4.78E-10 | 1 | 0 | 0 |
| AD Wightman | eGFR | | | rs769449 | rs34342646 | 0.00E+00 | 2.83E-21 | 0.00E+00 | 1.00E+00 | 3.36E-10 | 1 | 0 | 0 |
| AD Wightman | eGFR | | | rs34404554 | rs34342646 | 0.00E+00 | 2.79E-21 | 0.00E+00 | 9.86E-01 | 1.42E-02 | 1 | 0 | 0 |
| AD Wightman | eGFR | | | rs6857 | rs34342646 | 0.00E+00 | 2.83E-21 | 0.00E+00 | 1.00E+00 | 2.51E-06 | 1 | 0 | 0 |
| AD Wightman | eGFR | | | rs2075650 | rs34342646 | 0.00E+00 | 2.80E-21 | 0.00E+00 | 9.90E-01 | 1.01E-02 | 1 | 0 | 0 |
| AD Wightman | eGFR | | | rs11556505 | rs34342646 | 0.00E+00 | 2.77E-21 | 0.00E+00 | 9.81E-01 | 1.95E-02 | 1 | 0 | 0 |
| AD Wightman | eGFR | | | rs429358 | rs375972689 | 0.00E+00 | 5.89E-02 | 0.00E+00 | 9.40E-01 | 1.22E-03 | 1 | 0 | 0 |
| AD Wightman | eGFR | | | rs769449 | rs375972689 | 0.00E+00 | 5.90E-02 | 0.00E+00 | 9.40E-01 | 8.39E-04 | 1 | 0 | 0 |
| AD Wightman | eGFR | | | rs34404554 | rs375972689 | 0.00E+00 | 5.90E-02 | 0.00E+00 | 9.40E-01 | 6.37E-04 | 1 | 0 | 0 |
| AD Wightman | eGFR | | | rs6857 | rs375972689 | 0.00E+00 | 5.90E-02 | 0.00E+00 | 9.40E-01 | 6.44E-04 | 1 | 0 | 0 |
| AD Wightman | eGFR | | | rs2075650 | rs375972689 | 0.00E+00 | 5.90E-02 | 0.00E+00 | 9.40E-01 | 6.52E-04 | 1 | 0 | 0 |
| AD Wightman | eGFR | | | rs11556505 | rs375972689 | 0.00E+00 | 5.90E-02 | 0.00E+00 | 9.40E-01 | 6.28E-04 | 1 | 0 | 0 |

| | | | coloc SUSIE method | | | | | | | | | | |
|---|---|---|---|---|---|---|---|---|---|---|---|---|---|
| Trait1 | Trait 2 | cFDRlocusnum | nsnps 2 | hit1 | hit2 | PP.H0.abf | PP.H1.abf | PP.H2.abf | PP.H3.abf | PP.H4.abf | PP.H3+PP.H4≥0.8 | PP.H3/PP.H4≥3 | PP.H4/PP.H3≥5 |
| n | | | | | | | | | | | | | |
| AD Wightman | eGFR | 44a | 3489 | rs429358 | rs814573 | 0.00E+00 | 4.94E-324 | 0.00E+00 | 1.00E+00 | 3.21E-98 | 1 | 0 | 0 |
| AD Wightman | eGFR | | | rs769449 | rs814573 | 0.00E+00 | 4.94E-324 | 0.00E+00 | 1.00E+00 | 1.86E-134 | 1 | 0 | 0 |
| AD Wightman | eGFR | | | rs34404554 | rs814573 | 0.00E+00 | 4.94E-324 | 0.00E+00 | 1.00E+00 | 1.57E-197 | 1 | 0 | 0 |
| AD Wightman | eGFR | | | rs6857 | rs814573 | 0.00E+00 | 4.94E-324 | 0.00E+00 | 1.00E+00 | 8.42E-160 | 1 | 0 | 0 |
| AD Wightman | eGFR | | | rs2075650 | rs814573 | 0.00E+00 | 4.94E-324 | 0.00E+00 | 1.00E+00 | 2.17E-197 | 1 | 0 | 0 |
| AD Wightman | eGFR | | | rs11556505 | rs814573 | 0.00E+00 | 4.94E-324 | 0.00E+00 | 1.00E+00 | 6.90E-198 | 1 | 0 | 0 |
| AD Wightman | eGFR | | | rs429358 | rs4420638 | 0.00E+00 | 0.00E+00 | 0.00E+00 | 1.00E+00 | 3.47E-256 | 1 | 0 | 0 |
| AD Wightman | eGFR | | | rs769449 | rs4420638 | 0.00E+00 | 0.00E+00 | 0.00E+00 | 1.00E+00 | 0.00E+00 | 1 | 0 | 0 |
| AD Wightman | eGFR | | | rs34404554 | rs4420638 | 0.00E+00 | 0.00E+00 | 0.00E+00 | 1.00E+00 | 0.00E+00 | 1 | 0 | 0 |
| AD Wightman | eGFR | | | rs6857 | rs4420638 | 0.00E+00 | 0.00E+00 | 0.00E+00 | 1.00E+00 | 0.00E+00 | 1 | 0 | 0 |
| AD Wightman | eGFR | | | rs2075650 | rs4420638 | 0.00E+00 | 0.00E+00 | 0.00E+00 | 1.00E+00 | 0.00E+00 | 1 | 0 | 0 |
| AD Wightman | eGFR | | | rs11556505 | rs4420638 | 0.00E+00 | 0.00E+00 | 0.00E+00 | 1.00E+00 | 0.00E+00 | 1 | 0 | 0 |
| AD Wightma | eGFR | | | rs429358 | rs283811 | 0.00E+00 | 0.00E+00 | 0.00E+00 | 1.00E+00 | 0.00E+00 | 1 | 0 | 0 |

| | | | coloc SUSIE method | | | | | | | | | | |
|---|---|---|---|---|---|---|---|---|---|---|---|---|---|
| **Trait1** | **Trait 2** | **cFDRlocusnum** | **nsnps 2** | **hit1** | **hit2** | **PP.H0.abf** | **PP.H1.abf** | **PP.H2.abf** | **PP.H3.abf** | **PP.H4.abf** | **PP.H3+PP.H4≥0.8** | **PP.H3/PP.H4≥3** | **PP.H4/PP.H3≥5** |
| n | | | | | | | | | | | | | |
| AD Wightman | eGFR | | | rs769449 | rs283811 | 0.00E+00 | 0.00E+00 | 0.00E+00 | 1.00E+00 | 0.00E+00 | 1 | 0 | 0 |
| AD Wightman | eGFR | | | rs34404554 | rs283811 | 0.00E+00 | 0.00E+00 | 0.00E+00 | 1.00E+00 | 0.00E+00 | 1 | 0 | 0 |
| AD Wightman | eGFR | | | rs6857 | rs283811 | 0.00E+00 | 0.00E+00 | 0.00E+00 | 1.00E+00 | 0.00E+00 | 1 | 0 | 0 |
| AD Wightman | eGFR | | | rs2075650 | rs283811 | 0.00E+00 | 0.00E+00 | 0.00E+00 | 1.00E+00 | 0.00E+00 | 1 | 0 | 0 |
| AD Wightman | eGFR | | | rs11556505 | rs283811 | 0.00E+00 | 0.00E+00 | 0.00E+00 | 1.00E+00 | 0.00E+00 | 1 | 0 | 0 |
| AD Wightman | eGFR | | | rs429358 | rs283815 | 0.00E+00 | 0.00E+00 | 0.00E+00 | 1.00E+00 | 0.00E+00 | 1 | 0 | 0 |
| AD Wightman | eGFR | | | rs769449 | rs283815 | 0.00E+00 | 0.00E+00 | 0.00E+00 | 1.00E+00 | 0.00E+00 | 1 | 0 | 0 |
| AD Wightman | eGFR | | | rs34404554 | rs283815 | 0.00E+00 | 0.00E+00 | 0.00E+00 | 1.00E+00 | 0.00E+00 | 1 | 0 | 0 |
| AD Wightman | eGFR | | | rs6857 | rs283815 | 0.00E+00 | 0.00E+00 | 0.00E+00 | 1.00E+00 | 0.00E+00 | 1 | 0 | 0 |
| AD Wightman | eGFR | | | rs2075650 | rs283815 | 0.00E+00 | 0.00E+00 | 0.00E+00 | 1.00E+00 | 0.00E+00 | 1 | 0 | 0 |
| AD Wightman | eGFR | | | rs11556505 | rs283815 | 0.00E+00 | 0.00E+00 | 0.00E+00 | 1.00E+00 | 0.00E+00 | 1 | 0 | 0 |
| AD Wightman | eGFR | | | rs429358 | rs157592 | 0.00E+00 | 1.09E-152 | 0.00E+00 | 1.00E+00 | 5.86E-60 | 1 | 0 | 0 |
| AD Wightman | eGFR | | | rs769449 | rs157592 | 0.00E+00 | 1.09E-152 | 0.00E+00 | 1.00E+00 | 1.78E-72 | 1 | 0 | 0 |

| | | | coloc SUSIE method | | | | | | | | | | |
|---|---|---|---|---|---|---|---|---|---|---|---|---|---|
| **Trait1** | **Trait 2** | **cFDRlocusnum** | **nsnps 2** | **hit1** | **hit2** | **PP.H0.abf** | **PP.H1.abf** | **PP.H2.abf** | **PP.H3.abf** | **PP.H4.abf** | **PP.H3+PP.H4≥0.8** | **PP.H3/PP.H4≥3** | **PP.H4/PP.H3≥5** |
| AD Wightman | eGFR | | | rs34404554 | rs157592 | 0.00E+00 | 1.09E-152 | 0.00E+00 | 1.00E+00 | 1.67E-101 | 1 | 0 | 0 |
| AD Wightman | eGFR | | | rs6857 | rs157592 | 0.00E+00 | 1.09E-152 | 0.00E+00 | 1.00E+00 | 8.48E-84 | 1 | 0 | 0 |
| AD Wightman | eGFR | | | rs2075650 | rs157592 | 0.00E+00 | 1.09E-152 | 0.00E+00 | 1.00E+00 | 2.39E-101 | 1 | 0 | 0 |
| AD Wightman | eGFR | | | rs11556505 | rs157592 | 0.00E+00 | 1.09E-152 | 0.00E+00 | 1.00E+00 | 9.89E-102 | 1 | 0 | 0 |
| AD Wightman | eGFR | | | rs429358 | rs66807996 | 0.00E+00 | 1.20E-05 | 0.00E+00 | 1.00E+00 | 1.11E-07 | 1 | 0 | 0 |
| AD Wightman | eGFR | | | rs769449 | rs66807996 | 0.00E+00 | 1.20E-05 | 0.00E+00 | 1.00E+00 | 9.67E-08 | 1 | 0 | 0 |
| AD Wightman | eGFR | | | rs34404554 | rs66807996 | 0.00E+00 | 1.20E-05 | 0.00E+00 | 1.00E+00 | 8.92E-08 | 1 | 0 | 0 |
| AD Wightman | eGFR | | | rs6857 | rs66807996 | 0.00E+00 | 1.20E-05 | 0.00E+00 | 1.00E+00 | 8.89E-08 | 1 | 0 | 0 |
| AD Wightman | eGFR | | | rs2075650 | rs66807996 | 0.00E+00 | 1.20E-05 | 0.00E+00 | 1.00E+00 | 8.98E-08 | 1 | 0 | 0 |
| AD Wightman | eGFR | | | rs11556505 | rs66807996 | 0.00E+00 | 1.20E-05 | 0.00E+00 | 1.00E+00 | 8.88E-08 | 1 | 0 | 0 |
| AD Wightman | eGFR | | | rs429358 | rs34342646 | 0.00E+00 | 2.74E-21 | 0.00E+00 | 1.00E+00 | 4.70E-10 | 1 | 0 | 0 |
| AD Wightman | eGFR | | | rs769449 | rs34342646 | 0.00E+00 | 2.74E-21 | 0.00E+00 | 1.00E+00 | 3.32E-10 | 1 | 0 | 0 |
| AD Wightman | eGFR | | | rs34404554 | rs34342646 | 0.00E+00 | 2.70E-21 | 0.00E+00 | 9.86E-01 | 1.42E-02 | 1 | 0 | 0 |
| AD | eGFR | | | rs6857 | rs34342646 | 0.00E+00 | 2.74E-21 | 0.00E+00 | 1.00E+00 | 2.48E-06 | 1 | 0 | 0 |

| | | | coloc SUSIE method | | | | | | | | | | |
|---|---|---|---|---|---|---|---|---|---|---|---|---|---|
| **Trait1** | **Trait 2** | **cFDRlocusnum** | **nsnps 2** | **hit1** | **hit2** | **PP.H0.abf** | **PP.H1.abf** | **PP.H2.abf** | **PP.H3.abf** | **PP.H4.abf** | **PP.H3+PP.H4≥0.8** | **PP.H3/PP.H4≥3** | **PP.H4/PP.H3≥5** |
| Wightman | | | | | | | | | | | | | |
| AD Wightman | eGFR | | | rs2075650 | rs34342646 | 0.00E+00 | 2.71E-21 | 0.00E+00 | 9.90E-01 | 1.01E-02 | 1 | 0 | 0 |
| AD Wightman | eGFR | | | rs11556505 | rs34342646 | 0.00E+00 | 2.68E-21 | 0.00E+00 | 9.81E-01 | 1.94E-02 | 1 | 0 | 0 |
| AD Wightman | eGFR | | | rs429358 | rs375972689 | 0.00E+00 | 5.86E-02 | 0.00E+00 | 9.40E-01 | 1.22E-03 | 1 | 0 | 0 |
| AD Wightman | eGFR | | | rs769449 | rs375972689 | 0.00E+00 | 5.86E-02 | 0.00E+00 | 9.41E-01 | 8.34E-04 | 1 | 0 | 0 |
| AD Wightman | eGFR | | | rs34404554 | rs375972689 | 0.00E+00 | 5.86E-02 | 0.00E+00 | 9.41E-01 | 6.33E-04 | 1 | 0 | 0 |
| AD Wightman | eGFR | | | rs6857 | rs375972689 | 0.00E+00 | 5.86E-02 | 0.00E+00 | 9.41E-01 | 6.40E-04 | 1 | 0 | 0 |
| AD Wightman | eGFR | | | rs2075650 | rs375972689 | 0.00E+00 | 5.86E-02 | 0.00E+00 | 9.41E-01 | 6.48E-04 | 1 | 0 | 0 |
| AD Wightman | eGFR | | | rs11556505 | rs375972689 | 0.00E+00 | 5.86E-02 | 0.00E+00 | 9.41E-01 | 6.24E-04 | 1 | 0 | 0 |
| AD Wightman | eGFR | 44b | 3425 | rs429358 | rs814573 | 0.00E+00 | 4.94E-324 | 0.00E+00 | 1.00E+00 | 3.22E-98 | 1 | 0 | 0 |
| AD Wightman | eGFR | | | rs769449 | rs814573 | 0.00E+00 | 4.94E-324 | 0.00E+00 | 1.00E+00 | 1.84E-134 | 1 | 0 | 0 |
| AD Wightman | eGFR | | | rs34404554 | rs814573 | 0.00E+00 | 4.94E-324 | 0.00E+00 | 1.00E+00 | 1.56E-197 | 1 | 0 | 0 |
| AD Wightman | eGFR | | | rs6857 | rs814573 | 0.00E+00 | 4.94E-324 | 0.00E+00 | 1.00E+00 | 8.43E-160 | 1 | 0 | 0 |
| AD | eGFR | | | rs2075650 | rs814573 | 0.00E+00 | 4.94E- | 0.00E+00 | 1.00E+00 | 2.16E- | 1 | 0 | 0 |

| | | | coloc SUSIE method | | | | | | | | | | |
|---|---|---|---|---|---|---|---|---|---|---|---|---|---|
| **Trait1** | **Trait 2** | **cFDRlocusnum** | **nsnps 2** | **hit1** | **hit2** | **PP.H0.abf** | **PP.H1.abf** | **PP.H2.abf** | **PP.H3.abf** | **PP.H4.abf** | **PP.H3+PP.H4≥0.8** | **PP.H3/PP.H4≥3** | **PP.H4/PP.H3≥5** |
| Wightman | | | | | | | 324 | | | 197 | | | |
| AD Wightman | eGFR | | | rs11556505 | rs814573 | 0.00E+00 | 4.94E-324 | 0.00E+00 | 1.00E+00 | 6.85E-198 | 1 | 0 | 0 |
| AD Wightman | eGFR | | | rs429358 | rs4420638 | 0.00E+00 | 0.00E+00 | 0.00E+00 | 1.00E+00 | 3.45E-256 | 1 | 0 | 0 |
| AD Wightman | eGFR | | | rs769449 | rs4420638 | 0.00E+00 | 0.00E+00 | 0.00E+00 | 1.00E+00 | 0.00E+00 | 1 | 0 | 0 |
| AD Wightman | eGFR | | | rs34404554 | rs4420638 | 0.00E+00 | 0.00E+00 | 0.00E+00 | 1.00E+00 | 0.00E+00 | 1 | 0 | 0 |
| AD Wightman | eGFR | | | rs6857 | rs4420638 | 0.00E+00 | 0.00E+00 | 0.00E+00 | 1.00E+00 | 0.00E+00 | 1 | 0 | 0 |
| AD Wightman | eGFR | | | rs2075650 | rs4420638 | 0.00E+00 | 0.00E+00 | 0.00E+00 | 1.00E+00 | 0.00E+00 | 1 | 0 | 0 |
| AD Wightman | eGFR | | | rs11556505 | rs4420638 | 0.00E+00 | 0.00E+00 | 0.00E+00 | 1.00E+00 | 0.00E+00 | 1 | 0 | 0 |
| AD Wightman | eGFR | | | rs429358 | rs283811 | 0.00E+00 | 0.00E+00 | 0.00E+00 | 1.00E+00 | 0.00E+00 | 1 | 0 | 0 |
| AD Wightman | eGFR | | | rs769449 | rs283811 | 0.00E+00 | 0.00E+00 | 0.00E+00 | 1.00E+00 | 0.00E+00 | 1 | 0 | 0 |
| AD Wightman | eGFR | | | rs34404554 | rs283811 | 0.00E+00 | 0.00E+00 | 0.00E+00 | 1.00E+00 | 0.00E+00 | 1 | 0 | 0 |
| AD Wightman | eGFR | | | rs6857 | rs283811 | 0.00E+00 | 0.00E+00 | 0.00E+00 | 1.00E+00 | 0.00E+00 | 1 | 0 | 0 |
| AD Wightman | eGFR | | | rs2075650 | rs283811 | 0.00E+00 | 0.00E+00 | 0.00E+00 | 1.00E+00 | 0.00E+00 | 1 | 0 | 0 |
| AD Wightma | eGFR | | | rs11556505 | rs283811 | 0.00E+00 | 0.00E+00 | 0.00E+00 | 1.00E+00 | 0.00E+00 | 1 | 0 | 0 |

| | | | coloc SUSIE method | | | | | | | | | | |
|---|---|---|---|---|---|---|---|---|---|---|---|---|---|
| **Trait1** | **Trait 2** | **cFDRlocusnum** | **nsnps 2** | **hit1** | **hit2** | **PP.H0.abf** | **PP.H1.abf** | **PP.H2.abf** | **PP.H3.abf** | **PP.H4.abf** | **PP.H3+PP.H4≥0.8** | **PP.H3/PP.H4≥3** | **PP.H4/PP.H3≥5** |
| n | | | | | | | | | | | | | |
| AD Wightman | eGFR | | | rs429358 | rs283815 | 0.00E+00 | 0.00E+00 | 0.00E+00 | 1.00E+00 | 0.00E+00 | 1 | 0 | 0 |
| AD Wightman | eGFR | | | rs769449 | rs283815 | 0.00E+00 | 0.00E+00 | 0.00E+00 | 1.00E+00 | 0.00E+00 | 1 | 0 | 0 |
| AD Wightman | eGFR | | | rs34404554 | rs283815 | 0.00E+00 | 0.00E+00 | 0.00E+00 | 1.00E+00 | 0.00E+00 | 1 | 0 | 0 |
| AD Wightman | eGFR | | | rs6857 | rs283815 | 0.00E+00 | 0.00E+00 | 0.00E+00 | 1.00E+00 | 0.00E+00 | 1 | 0 | 0 |
| AD Wightman | eGFR | | | rs2075650 | rs283815 | 0.00E+00 | 0.00E+00 | 0.00E+00 | 1.00E+00 | 0.00E+00 | 1 | 0 | 0 |
| AD Wightman | eGFR | | | rs11556505 | rs283815 | 0.00E+00 | 0.00E+00 | 0.00E+00 | 1.00E+00 | 0.00E+00 | 1 | 0 | 0 |
| AD Wightman | eGFR | | | rs429358 | rs157592 | 0.00E+00 | 1.08E-152 | 0.00E+00 | 1.00E+00 | 5.84E-60 | 1 | 0 | 0 |
| AD Wightman | eGFR | | | rs769449 | rs157592 | 0.00E+00 | 1.08E-152 | 0.00E+00 | 1.00E+00 | 1.76E-72 | 1 | 0 | 0 |
| AD Wightman | eGFR | | | rs34404554 | rs157592 | 0.00E+00 | 1.08E-152 | 0.00E+00 | 1.00E+00 | 1.66E-101 | 1 | 0 | 0 |
| AD Wightman | eGFR | | | rs6857 | rs157592 | 0.00E+00 | 1.08E-152 | 0.00E+00 | 1.00E+00 | 8.45E-84 | 1 | 0 | 0 |
| AD Wightman | eGFR | | | rs2075650 | rs157592 | 0.00E+00 | 1.08E-152 | 0.00E+00 | 1.00E+00 | 2.37E-101 | 1 | 0 | 0 |
| AD Wightman | eGFR | | | rs11556505 | rs157592 | 0.00E+00 | 1.08E-152 | 0.00E+00 | 1.00E+00 | 9.80E-102 | 1 | 0 | 0 |
| AD Wightman | eGFR | | | rs429358 | rs66807996 | 0.00E+00 | 1.19E-05 | 0.00E+00 | 1.00E+00 | 1.11E-07 | 1 | 0 | 0 |

| | | | coloc SUSIE method | | | | | | | | | | |
|---|---|---|---|---|---|---|---|---|---|---|---|---|---|
| **Trait1** | **Trait 2** | **cFDRlocusnum** | **nsnps 2** | **hit1** | **hit2** | **PP.H0.abf** | **PP.H1.abf** | **PP.H2.abf** | **PP.H3.abf** | **PP.H4.abf** | **PP.H3+PP.H4≥0.8** | **PP.H3/PP.H4≥3** | **PP.H4/PP.H3≥5** |
| AD Wightman | eGFR | | | rs769449 | rs66807996 | 0.00E+00 | 1.19E-05 | 0.00E+00 | 1.00E+00 | 9.64E-08 | 1 | 0 | 0 |
| AD Wightman | eGFR | | | rs34404554 | rs66807996 | 0.00E+00 | 1.19E-05 | 0.00E+00 | 1.00E+00 | 8.90E-08 | 1 | 0 | 0 |
| AD Wightman | eGFR | | | rs6857 | rs66807996 | 0.00E+00 | 1.19E-05 | 0.00E+00 | 1.00E+00 | 8.87E-08 | 1 | 0 | 0 |
| AD Wightman | eGFR | | | rs2075650 | rs66807996 | 0.00E+00 | 1.19E-05 | 0.00E+00 | 1.00E+00 | 8.96E-08 | 1 | 0 | 0 |
| AD Wightman | eGFR | | | rs11556505 | rs66807996 | 0.00E+00 | 1.19E-05 | 0.00E+00 | 1.00E+00 | 8.86E-08 | 1 | 0 | 0 |
| AD Wightman | eGFR | | | rs429358 | rs34342646 | 0.00E+00 | 2.69E-21 | 0.00E+00 | 1.00E+00 | 4.66E-10 | 1 | 0 | 0 |
| AD Wightman | eGFR | | | rs769449 | rs34342646 | 0.00E+00 | 2.69E-21 | 0.00E+00 | 1.00E+00 | 3.30E-10 | 1 | 0 | 0 |
| AD Wightman | eGFR | | | rs34404554 | rs34342646 | 0.00E+00 | 2.65E-21 | 0.00E+00 | 9.86E-01 | 1.42E-02 | 1 | 0 | 0 |
| AD Wightman | eGFR | | | rs6857 | rs34342646 | 0.00E+00 | 2.69E-21 | 0.00E+00 | 1.00E+00 | 2.46E-06 | 1 | 0 | 0 |
| AD Wightman | eGFR | | | rs2075650 | rs34342646 | 0.00E+00 | 2.66E-21 | 0.00E+00 | 9.90E-01 | 1.01E-02 | 1 | 0 | 0 |
| AD Wightman | eGFR | | | rs11556505 | rs34342646 | 0.00E+00 | 2.64E-21 | 0.00E+00 | 9.81E-01 | 1.94E-02 | 1 | 0 | 0 |
| AD Wightman | eGFR | | | rs429358 | rs375972689 | 0.00E+00 | 5.87E-02 | 0.00E+00 | 9.40E-01 | 1.22E-03 | 1 | 0 | 0 |
| AD Wightman | eGFR | | | rs769449 | rs375972689 | 0.00E+00 | 5.87E-02 | 0.00E+00 | 9.40E-01 | 8.34E-04 | 1 | 0 | 0 |
| AD | eGFR | | | rs34404554 | rs37597268 | 0.00E+00 | 5.87E-02 | 0.00E+00 | 9.41E-01 | 6.34E-04 | 1 | 0 | 0 |

| | | | coloc SUSIE method | | | | | | | | | | |
|---|---|---|---|---|---|---|---|---|---|---|---|---|---|
| **Trait1** | **Trait 2** | **cFDRlocusnum** | **nsnps 2** | **hit1** | **hit2** | **PP.H0.abf** | **PP.H1.abf** | **PP.H2.abf** | **PP.H3.abf** | **PP.H4.abf** | **PP.H3+PP.H4≥0.8** | **PP.H3/PP.H4≥3** | **PP.H4/PP.H3≥5** |
| Wightman | | | | | 9 | | | | | | | | |
| AD Wightman | eGFR | | | rs6857 | rs375972689 | 0.00E+00 | 5.87E-02 | 0.00E+00 | 9.41E-01 | 6.41E-04 | 1 | 0 | 0 |
| AD Wightman | eGFR | | | rs2075650 | rs375972689 | 0.00E+00 | 5.87E-02 | 0.00E+00 | 9.41E-01 | 6.48E-04 | 1 | 0 | 0 |
| AD Wightman | eGFR | | | rs11556505 | rs375972689 | 0.00E+00 | 5.87E-02 | 0.00E+00 | 9.41E-01 | 6.24E-04 | 1 | 0 | 0 |
| AD Wightman | eGFR | 44c | 3406 | rs429358 | rs814573 | 0.00E+00 | 4.94E-324 | 0.00E+00 | 1.00E+00 | 3.21E-98 | 1 | 0 | 0 |
| AD Wightman | eGFR | | | rs769449 | rs814573 | 0.00E+00 | 4.94E-324 | 0.00E+00 | 1.00E+00 | 1.84E-134 | 1 | 0 | 0 |
| AD Wightman | eGFR | | | rs34404554 | rs814573 | 0.00E+00 | 4.94E-324 | 0.00E+00 | 1.00E+00 | 1.55E-197 | 1 | 0 | 0 |
| AD Wightman | eGFR | | | rs6857 | rs814573 | 0.00E+00 | 4.94E-324 | 0.00E+00 | 1.00E+00 | 8.41E-160 | 1 | 0 | 0 |
| AD Wightman | eGFR | | | rs2075650 | rs814573 | 0.00E+00 | 4.94E-324 | 0.00E+00 | 1.00E+00 | 2.15E-197 | 1 | 0 | 0 |
| AD Wightman | eGFR | | | rs11556505 | rs814573 | 0.00E+00 | 4.94E-324 | 0.00E+00 | 1.00E+00 | 6.82E-198 | 1 | 0 | 0 |
| AD Wightman | eGFR | | | rs429358 | rs4420638 | 0.00E+00 | 0.00E+00 | 0.00E+00 | 1.00E+00 | 3.46E-256 | 1 | 0 | 0 |
| AD Wightman | eGFR | | | rs769449 | rs4420638 | 0.00E+00 | 0.00E+00 | 0.00E+00 | 1.00E+00 | 0.00E+00 | 1 | 0 | 0 |
| AD Wightman | eGFR | | | rs34404554 | rs4420638 | 0.00E+00 | 0.00E+00 | 0.00E+00 | 1.00E+00 | 0.00E+00 | 1 | 0 | 0 |
| AD | eGFR | | | rs6857 | rs4420638 | 0.00E+00 | 0.00E+00 | 0.00E+00 | 1.00E+00 | 0.00E+00 | 1 | 0 | 0 |

| | | | coloc SUSIE method | | | | | | | | | | |
|---|---|---|---|---|---|---|---|---|---|---|---|---|---|
| **Trait1** | **Trait 2** | **cFDRlocusnum** | **nsnps 2** | **hit1** | **hit2** | **PP.H0.abf** | **PP.H1.abf** | **PP.H2.abf** | **PP.H3.abf** | **PP.H4.abf** | **PP.H3+PP.H4≥0.8** | **PP.H3/PP.H4≥3** | **PP.H4/PP.H3≥5** |
| Wightman | | | | | | | | | | | | | |
| AD Wightman | eGFR | | | rs2075650 | rs4420638 | 0.00E+00 | 0.00E+00 | 0.00E+00 | 1.00E+00 | 0.00E+00 | 1 | 0 | 0 |
| AD Wightman | eGFR | | | rs11556505 | rs4420638 | 0.00E+00 | 0.00E+00 | 0.00E+00 | 1.00E+00 | 0.00E+00 | 1 | 0 | 0 |
| AD Wightman | eGFR | | | rs429358 | rs283811 | 0.00E+00 | 0.00E+00 | 0.00E+00 | 1.00E+00 | 0.00E+00 | 1 | 0 | 0 |
| AD Wightman | eGFR | | | rs769449 | rs283811 | 0.00E+00 | 0.00E+00 | 0.00E+00 | 1.00E+00 | 0.00E+00 | 1 | 0 | 0 |
| AD Wightman | eGFR | | | rs34404554 | rs283811 | 0.00E+00 | 0.00E+00 | 0.00E+00 | 1.00E+00 | 0.00E+00 | 1 | 0 | 0 |
| AD Wightman | eGFR | | | rs6857 | rs283811 | 0.00E+00 | 0.00E+00 | 0.00E+00 | 1.00E+00 | 0.00E+00 | 1 | 0 | 0 |
| AD Wightman | eGFR | | | rs2075650 | rs283811 | 0.00E+00 | 0.00E+00 | 0.00E+00 | 1.00E+00 | 0.00E+00 | 1 | 0 | 0 |
| AD Wightman | eGFR | | | rs11556505 | rs283811 | 0.00E+00 | 0.00E+00 | 0.00E+00 | 1.00E+00 | 0.00E+00 | 1 | 0 | 0 |
| AD Wightman | eGFR | | | rs429358 | rs283815 | 0.00E+00 | 0.00E+00 | 0.00E+00 | 1.00E+00 | 0.00E+00 | 1 | 0 | 0 |
| AD Wightman | eGFR | | | rs769449 | rs283815 | 0.00E+00 | 0.00E+00 | 0.00E+00 | 1.00E+00 | 0.00E+00 | 1 | 0 | 0 |
| AD Wightman | eGFR | | | rs34404554 | rs283815 | 0.00E+00 | 0.00E+00 | 0.00E+00 | 1.00E+00 | 0.00E+00 | 1 | 0 | 0 |
| AD Wightman | eGFR | | | rs6857 | rs283815 | 0.00E+00 | 0.00E+00 | 0.00E+00 | 1.00E+00 | 0.00E+00 | 1 | 0 | 0 |
| AD Wightma | eGFR | | | rs2075650 | rs283815 | 0.00E+00 | 0.00E+00 | 0.00E+00 | 1.00E+00 | 0.00E+00 | 1 | 0 | 0 |

| | | | coloc SUSIE method | | | | | | | | | | |
|---|---|---|---|---|---|---|---|---|---|---|---|---|---|
| Trait1 | Trait2 | cFDRlocusnum | nsnps2 | hit1 | hit2 | PP.H0.abf | PP.H1.abf | PP.H2.abf | PP.H3.abf | PP.H4.abf | PP.H3+PP.H4≥0.8 | PP.H3/PP.H4≥3 | PP.H4/PP.H3≥5 |
| n | | | | | | | | | | | | | |
| AD Wightman | eGFR | | | rs11556505 | rs283815 | 0.00E+00 | 0.00E+00 | 0.00E+00 | 1.00E+00 | 0.00E+00 | 1 | 0 | 0 |
| AD Wightman | eGFR | | | rs429358 | rs157592 | 0.00E+00 | 1.08E-152 | 0.00E+00 | 1.00E+00 | 5.84E-60 | 1 | 0 | 0 |
| AD Wightman | eGFR | | | rs769449 | rs157592 | 0.00E+00 | 1.08E-152 | 0.00E+00 | 1.00E+00 | 1.76E-72 | 1 | 0 | 0 |
| AD Wightman | eGFR | | | rs34404554 | rs157592 | 0.00E+00 | 1.08E-152 | 0.00E+00 | 1.00E+00 | 1.65E-101 | 1 | 0 | 0 |
| AD Wightman | eGFR | | | rs6857 | rs157592 | 0.00E+00 | 1.08E-152 | 0.00E+00 | 1.00E+00 | 8.45E-84 | 1 | 0 | 0 |
| AD Wightman | eGFR | | | rs2075650 | rs157592 | 0.00E+00 | 1.08E-152 | 0.00E+00 | 1.00E+00 | 2.37E-101 | 1 | 0 | 0 |
| AD Wightman | eGFR | | | rs11556505 | rs157592 | 0.00E+00 | 1.08E-152 | 0.00E+00 | 1.00E+00 | 9.79E-102 | 1 | 0 | 0 |
| AD Wightman | eGFR | | | rs429358 | rs66807996 | 0.00E+00 | 1.19E-05 | 0.00E+00 | 1.00E+00 | 1.11E-07 | 1 | 0 | 0 |
| AD Wightman | eGFR | | | rs769449 | rs66807996 | 0.00E+00 | 1.19E-05 | 0.00E+00 | 1.00E+00 | 9.63E-08 | 1 | 0 | 0 |
| AD Wightman | eGFR | | | rs34404554 | rs66807996 | 0.00E+00 | 1.19E-05 | 0.00E+00 | 1.00E+00 | 8.89E-08 | 1 | 0 | 0 |
| AD Wightman | eGFR | | | rs6857 | rs66807996 | 0.00E+00 | 1.19E-05 | 0.00E+00 | 1.00E+00 | 8.86E-08 | 1 | 0 | 0 |
| AD Wightman | eGFR | | | rs2075650 | rs66807996 | 0.00E+00 | 1.19E-05 | 0.00E+00 | 1.00E+00 | 8.96E-08 | 1 | 0 | 0 |
| AD Wightman | eGFR | | | rs11556505 | rs66807996 | 0.00E+00 | 1.19E-05 | 0.00E+00 | 1.00E+00 | 8.85E-08 | 1 | 0 | 0 |

| | | | coloc SUSIE method | | | | | | | | | | |
|---|---|---|---|---|---|---|---|---|---|---|---|---|---|
| **Trait1** | **Trait 2** | **cFDRlocusnum** | **nsnps 2** | **hit1** | **hit2** | **PP.H0.abf** | **PP.H1.abf** | **PP.H2.abf** | **PP.H3.abf** | **PP.H4.abf** | **PP.H3+PP.H4≥0.8** | **PP.H3/PP.H4≥3** | **PP.H4/PP.H3≥5** |
| AD Wightman | eGFR | | | rs429358 | rs34342646 | 0.00E+00 | 2.68E-21 | 0.00E+00 | 1.00E+00 | 4.65E-10 | 1 | 0 | 0 |
| AD Wightman | eGFR | | | rs769449 | rs34342646 | 0.00E+00 | 2.68E-21 | 0.00E+00 | 1.00E+00 | 3.30E-10 | 1 | 0 | 0 |
| AD Wightman | eGFR | | | rs34404554 | rs34342646 | 0.00E+00 | 2.64E-21 | 0.00E+00 | 9.86E-01 | 1.42E-02 | 1 | 0 | 0 |
| AD Wightman | eGFR | | | rs6857 | rs34342646 | 0.00E+00 | 2.68E-21 | 0.00E+00 | 1.00E+00 | 2.46E-06 | 1 | 0 | 0 |
| AD Wightman | eGFR | | | rs2075650 | rs34342646 | 0.00E+00 | 2.65E-21 | 0.00E+00 | 9.90E-01 | 1.01E-02 | 1 | 0 | 0 |
| AD Wightman | eGFR | | | rs11556505 | rs34342646 | 0.00E+00 | 2.62E-21 | 0.00E+00 | 9.81E-01 | 1.94E-02 | 1 | 0 | 0 |
| AD Wightman | eGFR | | | rs429358 | rs375972689 | 0.00E+00 | 5.86E-02 | 0.00E+00 | 9.40E-01 | 1.22E-03 | 1 | 0 | 0 |
| AD Wightman | eGFR | | | rs769449 | rs375972689 | 0.00E+00 | 5.87E-02 | 0.00E+00 | 9.41E-01 | 8.34E-04 | 1 | 0 | 0 |
| AD Wightman | eGFR | | | rs34404554 | rs375972689 | 0.00E+00 | 5.87E-02 | 0.00E+00 | 9.41E-01 | 6.34E-04 | 1 | 0 | 0 |
| AD Wightman | eGFR | | | rs6857 | rs375972689 | 0.00E+00 | 5.87E-02 | 0.00E+00 | 9.41E-01 | 6.40E-04 | 1 | 0 | 0 |
| AD Wightman | eGFR | | | rs2075650 | rs375972689 | 0.00E+00 | 5.87E-02 | 0.00E+00 | 9.41E-01 | 6.48E-04 | 1 | 0 | 0 |
| AD Wightman | eGFR | | | rs11556505 | rs375972689 | 0.00E+00 | 5.87E-02 | 0.00E+00 | 9.41E-01 | 6.24E-04 | 1 | 0 | 0 |
| AD Wightman | eGFR | 44d | 3399 | rs429358 | rs814573 | 0.00E+00 | 4.94E-324 | 0.00E+00 | 1.00E+00 | 3.22E-98 | 1 | 0 | 0 |

| | | | coloc SUSIE method | | | | | | | | | | |
|---|---|---|---|---|---|---|---|---|---|---|---|---|---|
| **Trait1** | **Trait 2** | **cFDRlocusnum** | **nsnps 2** | **hit1** | **hit2** | **PP.H0.abf** | **PP.H1.abf** | **PP.H2.abf** | **PP.H3.abf** | **PP.H4.abf** | **PP.H3+PP.H4≥0.8** | **PP.H3/PP.H4≥3** | **PP.H4/PP.H3≥5** |
| AD Wightman | eGFR | | | rs769449 | rs814573 | 0.00E+00 | 4.94E-324 | 0.00E+00 | 1.00E+00 | 1.84E-134 | 1 | 0 | 0 |
| AD Wightman | eGFR | | | rs34404554 | rs814573 | 0.00E+00 | 4.94E-324 | 0.00E+00 | 1.00E+00 | 1.55E-197 | 1 | 0 | 0 |
| AD Wightman | eGFR | | | rs6857 | rs814573 | 0.00E+00 | 4.94E-324 | 0.00E+00 | 1.00E+00 | 8.46E-160 | 1 | 0 | 0 |
| AD Wightman | eGFR | | | rs2075650 | rs814573 | 0.00E+00 | 4.94E-324 | 0.00E+00 | 1.00E+00 | 2.15E-197 | 1 | 0 | 0 |
| AD Wightman | eGFR | | | rs11556505 | rs814573 | 0.00E+00 | 4.94E-324 | 0.00E+00 | 1.00E+00 | 6.83E-198 | 1 | 0 | 0 |
| AD Wightman | eGFR | | | rs429358 | rs4420638 | 0.00E+00 | 0.00E+00 | 0.00E+00 | 1.00E+00 | 3.48E-256 | 1 | 0 | 0 |
| AD Wightman | eGFR | | | rs769449 | rs4420638 | 0.00E+00 | 0.00E+00 | 0.00E+00 | 1.00E+00 | 0.00E+00 | 1 | 0 | 0 |
| AD Wightman | eGFR | | | rs34404554 | rs4420638 | 0.00E+00 | 0.00E+00 | 0.00E+00 | 1.00E+00 | 0.00E+00 | 1 | 0 | 0 |
| AD Wightman | eGFR | | | rs6857 | rs4420638 | 0.00E+00 | 0.00E+00 | 0.00E+00 | 1.00E+00 | 0.00E+00 | 1 | 0 | 0 |
| AD Wightman | eGFR | | | rs2075650 | rs4420638 | 0.00E+00 | 0.00E+00 | 0.00E+00 | 1.00E+00 | 0.00E+00 | 1 | 0 | 0 |
| AD Wightman | eGFR | | | rs11556505 | rs4420638 | 0.00E+00 | 0.00E+00 | 0.00E+00 | 1.00E+00 | 0.00E+00 | 1 | 0 | 0 |
| AD Wightman | eGFR | | | rs429358 | rs283811 | 0.00E+00 | 0.00E+00 | 0.00E+00 | 1.00E+00 | 0.00E+00 | 1 | 0 | 0 |
| AD Wightman | eGFR | | | rs769449 | rs283811 | 0.00E+00 | 0.00E+00 | 0.00E+00 | 1.00E+00 | 0.00E+00 | 1 | 0 | 0 |
| AD | eGFR | | | rs34404554 | rs283811 | 0.00E+00 | 0.00E+00 | 0.00E+00 | 1.00E+00 | 0.00E+00 | 1 | 0 | 0 |

| | | | coloc SUSIE method | | | | | | | | | | |
|---|---|---|---|---|---|---|---|---|---|---|---|---|---|
| Trait1 | Trait 2 | cFDRlocusnum | nsnps 2 | hit1 | hit2 | PP.H0.abf | PP.H1.abf | PP.H2.abf | PP.H3.abf | PP.H4.abf | PP.H3+PP.H4≥0.8 | PP.H3/PP.H4≥3 | PP.H4/PP.H3≥5 |
| Wightman | | | | | | | | | | | | | |
| AD Wightman | eGFR | | | rs6857 | rs283811 | 0.00E+00 | 0.00E+00 | 0.00E+00 | 1.00E+00 | 0.00E+00 | 1 | 0 | 0 |
| AD Wightman | eGFR | | | rs2075650 | rs283811 | 0.00E+00 | 0.00E+00 | 0.00E+00 | 1.00E+00 | 0.00E+00 | 1 | 0 | 0 |
| AD Wightman | eGFR | | | rs11556505 | rs283811 | 0.00E+00 | 0.00E+00 | 0.00E+00 | 1.00E+00 | 0.00E+00 | 1 | 0 | 0 |
| AD Wightman | eGFR | | | rs429358 | rs283815 | 0.00E+00 | 0.00E+00 | 0.00E+00 | 1.00E+00 | 0.00E+00 | 1 | 0 | 0 |
| AD Wightman | eGFR | | | rs769449 | rs283815 | 0.00E+00 | 0.00E+00 | 0.00E+00 | 1.00E+00 | 0.00E+00 | 1 | 0 | 0 |
| AD Wightman | eGFR | | | rs34404554 | rs283815 | 0.00E+00 | 0.00E+00 | 0.00E+00 | 1.00E+00 | 0.00E+00 | 1 | 0 | 0 |
| AD Wightman | eGFR | | | rs6857 | rs283815 | 0.00E+00 | 0.00E+00 | 0.00E+00 | 1.00E+00 | 0.00E+00 | 1 | 0 | 0 |
| AD Wightman | eGFR | | | rs2075650 | rs283815 | 0.00E+00 | 0.00E+00 | 0.00E+00 | 1.00E+00 | 0.00E+00 | 1 | 0 | 0 |
| AD Wightman | eGFR | | | rs11556505 | rs283815 | 0.00E+00 | 0.00E+00 | 0.00E+00 | 1.00E+00 | 0.00E+00 | 1 | 0 | 0 |
| AD Wightman | eGFR | | | rs429358 | rs157592 | 0.00E+00 | 1.09E-152 | 0.00E+00 | 1.00E+00 | 5.88E-60 | 1 | 0 | 0 |
| AD Wightman | eGFR | | | rs769449 | rs157592 | 0.00E+00 | 1.09E-152 | 0.00E+00 | 1.00E+00 | 1.77E-72 | 1 | 0 | 0 |
| AD Wightman | eGFR | | | rs34404554 | rs157592 | 0.00E+00 | 1.09E-152 | 0.00E+00 | 1.00E+00 | 1.67E-101 | 1 | 0 | 0 |
| AD Wightma | eGFR | | | rs6857 | rs157592 | 0.00E+00 | 1.09E-152 | 0.00E+00 | 1.00E+00 | 8.52E-84 | 1 | 0 | 0 |

| | | | coloc SUSIE method | | | | | | | | | | |
|---|---|---|---|---|---|---|---|---|---|---|---|---|---|
| **Trait1** | **Trait 2** | **cFDRlocusnum** | **nsnps 2** | **hit1** | **hit2** | **PP.H0.abf** | **PP.H1.abf** | **PP.H2.abf** | **PP.H3.abf** | **PP.H4.abf** | **PP.H3+PP.H4≥0.8** | **PP.H3/PP.H4≥3** | **PP.H4/PP.H3≥5** |
| n | | | | | | | | | | | | | |
| AD Wightman | eGFR | | | rs2075650 | rs157592 | 0.00E+00 | 1.09E-152 | 0.00E+00 | 1.00E+00 | 2.38E-101 | 1 | 0 | 0 |
| AD Wightman | eGFR | | | rs11556505 | rs157592 | 0.00E+00 | 1.09E-152 | 0.00E+00 | 1.00E+00 | 9.86E-102 | 1 | 0 | 0 |
| AD Wightman | eGFR | | | rs429358 | rs66807996 | 0.00E+00 | 1.19E-05 | 0.00E+00 | 1.00E+00 | 1.11E-07 | 1 | 0 | 0 |
| AD Wightman | eGFR | | | rs769449 | rs66807996 | 0.00E+00 | 1.19E-05 | 0.00E+00 | 1.00E+00 | 9.62E-08 | 1 | 0 | 0 |
| AD Wightman | eGFR | | | rs34404554 | rs66807996 | 0.00E+00 | 1.19E-05 | 0.00E+00 | 1.00E+00 | 8.87E-08 | 1 | 0 | 0 |
| AD Wightman | eGFR | | | rs6857 | rs66807996 | 0.00E+00 | 1.19E-05 | 0.00E+00 | 1.00E+00 | 8.85E-08 | 1 | 0 | 0 |
| AD Wightman | eGFR | | | rs2075650 | rs66807996 | 0.00E+00 | 1.19E-05 | 0.00E+00 | 1.00E+00 | 8.94E-08 | 1 | 0 | 0 |
| AD Wightman | eGFR | | | rs11556505 | rs66807996 | 0.00E+00 | 1.19E-05 | 0.00E+00 | 1.00E+00 | 8.84E-08 | 1 | 0 | 0 |
| AD Wightman | eGFR | | | rs429358 | rs34342646 | 0.00E+00 | 2.65E-21 | 0.00E+00 | 1.00E+00 | 4.62E-10 | 1 | 0 | 0 |
| AD Wightman | eGFR | | | rs769449 | rs34342646 | 0.00E+00 | 2.65E-21 | 0.00E+00 | 1.00E+00 | 3.28E-10 | 1 | 0 | 0 |
| AD Wightman | eGFR | | | rs34404554 | rs34342646 | 0.00E+00 | 2.61E-21 | 0.00E+00 | 9.86E-01 | 1.42E-02 | 1 | 0 | 0 |
| AD Wightman | eGFR | | | rs6857 | rs34342646 | 0.00E+00 | 2.65E-21 | 0.00E+00 | 1.00E+00 | 2.45E-06 | 1 | 0 | 0 |
| AD Wightman | eGFR | | | rs2075650 | rs34342646 | 0.00E+00 | 2.62E-21 | 0.00E+00 | 9.90E-01 | 1.01E-02 | 1 | 0 | 0 |

| | | | coloc SUSIE method | | | | | | | | | | |
|---|---|---|---|---|---|---|---|---|---|---|---|---|---|
| **Trait1** | **Trait 2** | **cFDRlocusnum** | **nsnps 2** | **hit1** | **hit2** | **PP.H0.abf** | **PP.H1.abf** | **PP.H2.abf** | **PP.H3.abf** | **PP.H4.abf** | **PP.H3+PP.H4≥0.8** | **PP.H3/PP.H4≥3** | **PP.H4/PP.H3≥5** |
| AD Wightman | eGFR | | | rs11556505 | rs34342646 | 0.00E+00 | 2.60E-21 | 0.00E+00 | 9.81E-01 | 1.94E-02 | 1 | 0 | 0 |
| AD Wightman | eGFR | | | rs429358 | rs375972689 | 0.00E+00 | 5.86E-02 | 0.00E+00 | 9.40E-01 | 1.22E-03 | 1 | 0 | 0 |
| AD Wightman | eGFR | | | rs769449 | rs375972689 | 0.00E+00 | 5.86E-02 | 0.00E+00 | 9.41E-01 | 8.34E-04 | 1 | 0 | 0 |
| AD Wightman | eGFR | | | rs34404554 | rs375972689 | 0.00E+00 | 5.87E-02 | 0.00E+00 | 9.41E-01 | 6.33E-04 | 1 | 0 | 0 |
| AD Wightman | eGFR | | | rs6857 | rs375972689 | 0.00E+00 | 5.87E-02 | 0.00E+00 | 9.41E-01 | 6.40E-04 | 1 | 0 | 0 |
| AD Wightman | eGFR | | | rs2075650 | rs375972689 | 0.00E+00 | 5.87E-02 | 0.00E+00 | 9.41E-01 | 6.48E-04 | 1 | 0 | 0 |
| AD Wightman | eGFR | | | rs11556505 | rs375972689 | 0.00E+00 | 5.87E-02 | 0.00E+00 | 9.41E-01 | 6.24E-04 | 1 | 0 | 0 |
| AD Wightman | eGFR | 47 | 3088 | rs346771 | rs1800437 | 0.00E+00 | 3.82E-06 | 0.00E+00 | 1.00E+00 | 3.89E-08 | 1 | 0 | 0 |
| AD Wightman | eGFR | | | rs346773 | rs1800437 | 0.00E+00 | 3.82E-06 | 0.00E+00 | 1.00E+00 | 3.65E-08 | 1 | 0 | 0 |
| AD Wightman | eGFR | 49 | 3379 | rs11878908 | rs60068692 | 0.00E+00 | 8.49E-04 | 0.00E+00 | 9.99E-01 | 7.19E-06 | 1 | 0 | 0 |
| AD Wightman | eGFR | | | rs2287921 | rs60068692 | 0.00E+00 | 8.49E-04 | 0.00E+00 | 9.99E-01 | 7.17E-06 | 1 | 0 | 0 |
| AD Wightman | eGFR | | | rs2287922 | rs60068692 | 0.00E+00 | 8.49E-04 | 0.00E+00 | 9.99E-01 | 7.15E-06 | 1 | 0 | 0 |
| AD Wightma | eGFR | | | rs11878908 | rs187500064 | 0.00E+00 | 1.04E-29 | 0.00E+00 | 1.00E+00 | 6.97E-32 | 1 | 0 | 0 |

| | | | coloc SUSIE method | | | | | | | | | | |
|---|---|---|---|---|---|---|---|---|---|---|---|---|---|
| **Trait1** | **Trait 2** | **cFDRlocusnum** | **nsnps 2** | **hit1** | **hit2** | **PP.H0.abf** | **PP.H1.abf** | **PP.H2.abf** | **PP.H3.abf** | **PP.H4.abf** | **PP.H3+PP.H4≥0.8** | **PP.H3/PP.H4≥3** | **PP.H4/PP.H3≥5** |
| n | | | | | | | | | | | | | |
| AD Wightman | eGFR | | | rs2287921 | rs187500064 | 0.00E+00 | 1.04E-29 | 0.00E+00 | 1.00E+00 | 6.89E-32 | 1 | 0 | 0 |
| AD Wightman | eGFR | | | rs2287922 | rs187500064 | 0.00E+00 | 1.04E-29 | 0.00E+00 | 1.00E+00 | 6.68E-32 | 1 | 0 | 0 |
| AD Wightman | eGFR | | | rs11878908 | rs143462803 | 0.00E+00 | 3.88E-24 | 0.00E+00 | 1.00E+00 | 1.38E-25 | 1 | 0 | 0 |
| AD Wightman | eGFR | | | rs2287921 | rs143462803 | 0.00E+00 | 3.88E-24 | 0.00E+00 | 1.00E+00 | 1.33E-25 | 1 | 0 | 0 |
| AD Wightman | eGFR | | | rs2287922 | rs143462803 | 0.00E+00 | 3.88E-24 | 0.00E+00 | 1.00E+00 | 1.31E-25 | 1 | 0 | 0 |
| AD Wightman | eGFR | | | rs11878908 | rs12461216 | 0.00E+00 | 5.02E-04 | 0.00E+00 | 9.99E-01 | 2.52E-05 | 1 | 0 | 0 |
| AD Wightman | eGFR | | | rs2287921 | rs12461216 | 0.00E+00 | 5.02E-04 | 0.00E+00 | 9.99E-01 | 2.45E-05 | 1 | 0 | 0 |
| AD Wightman | eGFR | | | rs2287922 | rs12461216 | 0.00E+00 | 5.02E-04 | 0.00E+00 | 9.99E-01 | 2.34E-05 | 1 | 0 | 0 |
| AD Wightman | eGFR | | | rs11878908 | rs281380 | 0.00E+00 | 1.44E-08 | 0.00E+00 | 1.00E+00 | 2.06E-08 | 1 | 0 | 0 |
| AD Wightman | eGFR | | | rs2287921 | rs281380 | 0.00E+00 | 1.44E-08 | 0.00E+00 | 1.00E+00 | 2.10E-08 | 1 | 0 | 0 |
| AD Wightman | eGFR | | | rs2287922 | rs281380 | 0.00E+00 | 1.44E-08 | 0.00E+00 | 1.00E+00 | 2.19E-08 | 1 | 0 | 0 |
| AD Wightman | eGFR | 53 | 1877 | rs4820426 | rs73169043 | 0.00E+00 | 0.00E+00 | 0.00E+00 | 1.00E+00 | 0.00E+00 | 1 | 0 | 0 |
| AD Wightma | eGFR | | | rs4820423 | rs73169043 | 0.00E+00 | 0.00E+00 | 0.00E+00 | 1.00E+00 | 0.00E+00 | 1 | 0 | 0 |

| | | | coloc SUSIE method | | | | | | | | | | |
|---|---|---|---|---|---|---|---|---|---|---|---|---|---|
| **Trait1** | **Trait2** | **cFDRlocusnum** | **nsnps2** | **hit1** | **hit2** | **PP.H0.abf** | **PP.H1.abf** | **PP.H2.abf** | **PP.H3.abf** | **PP.H4.abf** | **PP.H3+PP.H4≥0.8** | **PP.H3/PP.H4≥3** | **PP.H4/PP.H3≥5** |
| n | | | | | | | | | | | | | |
| AD Wightman | eGFR | | | rs9611467 | rs73169043 | 0.00E+00 | 0.00E+00 | 0.00E+00 | 1.00E+00 | 0.00E+00 | 1 | 0 | 0 |
| AD Wightman | eGFR | | | rs9611466 | rs73169043 | 0.00E+00 | 0.00E+00 | 0.00E+00 | 1.00E+00 | 0.00E+00 | 1 | 0 | 0 |
| AD Wightman | eGFR | | | rs9611465 | rs73169043 | 0.00E+00 | 0.00E+00 | 0.00E+00 | 1.00E+00 | 0.00E+00 | 1 | 0 | 0 |
| AD Wightman | eGFR | | | rs12484477 | rs73169043 | 0.00E+00 | 0.00E+00 | 0.00E+00 | 1.00E+00 | 0.00E+00 | 1 | 0 | 0 |
| AD Wightman | eGFR | | | rs9611459 | rs73169043 | 0.00E+00 | 0.00E+00 | 0.00E+00 | 1.00E+00 | 0.00E+00 | 1 | 0 | 0 |
| AD Wightman | eGFR | | | rs75723348 | rs73169043 | 0.00E+00 | 0.00E+00 | 0.00E+00 | 1.00E+00 | 0.00E+00 | 1 | 0 | 0 |
| AD Wightman | eGFR | | | rs4820426 | rs116973859 | 0.00E+00 | 0.00E+00 | 0.00E+00 | 1.00E+00 | 0.00E+00 | 1 | 0 | 0 |
| AD Wightman | eGFR | | | rs4820423 | rs116973859 | 0.00E+00 | 0.00E+00 | 0.00E+00 | 1.00E+00 | 0.00E+00 | 1 | 0 | 0 |
| AD Wightman | eGFR | | | rs9611467 | rs116973859 | 0.00E+00 | 0.00E+00 | 0.00E+00 | 1.00E+00 | 0.00E+00 | 1 | 0 | 0 |
| AD Wightman | eGFR | | | rs9611466 | rs116973859 | 0.00E+00 | 0.00E+00 | 0.00E+00 | 1.00E+00 | 0.00E+00 | 1 | 0 | 0 |
| AD Wightman | eGFR | | | rs9611465 | rs116973859 | 0.00E+00 | 0.00E+00 | 0.00E+00 | 1.00E+00 | 0.00E+00 | 1 | 0 | 0 |
| AD Wightman | eGFR | | | rs12484477 | rs116973859 | 0.00E+00 | 0.00E+00 | 0.00E+00 | 1.00E+00 | 0.00E+00 | 1 | 0 | 0 |
| AD Wightman | eGFR | | | rs9611459 | rs116973859 | 0.00E+00 | 0.00E+00 | 0.00E+00 | 1.00E+00 | 0.00E+00 | 1 | 0 | 0 |

| | | | coloc SUSIE method | | | | | | | | | | |
|---|---|---|---|---|---|---|---|---|---|---|---|---|---|
| **Trait1** | **Trait 2** | **cFDRlocusnum** | **nsnps 2** | **hit1** | **hit2** | **PP.H0.abf** | **PP.H1.abf** | **PP.H2.abf** | **PP.H3.abf** | **PP.H4.abf** | **PP.H3+PP.H4≥0.8** | **PP.H3/PP.H4≥3** | **PP.H4/PP.H3≥5** |
| AD Wightman | eGFR | | | rs75723348 | rs116973859 | 0.00E+00 | 0.00E+00 | 0.00E+00 | 1.00E+00 | 0.00E+00 | 1 | 0 | 0 |

**Supplementary Table S28.** Summary of colocalization analyses of GWAS Summary Statistics with AD and eGFR using Sum of Single Effects (SuSiE) framework method of the AD EUR data summary. This concise summary table synthesizes colocalization findings across all European ancestry analyses, providing high-level statistics for rapid interpretation. For each combination of traits and AD dataset (Kunkle and Wightman, both corrected and uncorrected), the table reports: the total number of conjFDR loci that were tested for colocalization, the number of loci with any colocalized variants (PP.H3 or PP.H4 > threshold), the number specifically showing evidence for shared causal variants (high PP.H4), and percentages representing the proportion of tested loci showing colocalization, the proportion with distinct causal variants, and the proportion with shared causal variants among colocalized loci. Note: 0 indicates "No", and 1 indicates "Yes".

| Trait 1 | Trait 2 | Total conjFDR Loci Tested | # Loci with Colocalized Variants | # Loci with Suggesting Shared Causal Variant | Total Tested Loci that are Colocalized (%) | Colocalized and Distinct Causal Loci (%) | Colocalized and Shared Causal Loci (%) |
|---|---|---|---|---|---|---|---|
| eGFR | AD Kunkle | 17 | 7 | 6 | 41.18 | 14.29 | 85.71 |
| eGFR | AD Kunkle Corrected | 16 | 7 | 6 | 43.75 | 14.29 | 85.71 |
| eGFR | AD Wightman | 57 | 35 | 4 | 61.40 | 88.57 | 11.43 |
| eGFR | AD Wightman Corrected | 58 | 14 | 9 | 24.14 | 35.71 | 64.29 |

**Supplementary Table S29. Cis-Mendelian Randomization Results in European Ancestry (Kunkle Discovery).**
This table presents results from cis-Mendelian randomization (cis-MR) analysis, which uses genetic variants within specific genomic loci as instrumental variables to test for causal relationships between AD and eGFR. Unlike trans-MR which uses genome-wide variants, cis-MR focuses on local genetic architecture to assess locus-specific causal effects. For each pleiotropic locus identified in conjFDR analysis, bidirectional cis-MR was performed: forward analysis (eGFR as exposure, AD as outcome) and reverse analysis (AD as exposure, eGFR as outcome). The table reports: locus identifiers, ancestry, analysis direction, causal effect estimates (theta) with standard errors, z-scores, p-values, and 95% confidence intervals. Statistically significant causal effects suggest that genetic variation in kidney function causally influences AD risk, or vice versa, within those specific pleiotropic genomic regions identified in conjFDR.

| Locus Num | Locus | Ancestry | Analysis Type | Theta (Causal Estimate) | Theta SE | Z-Score | P-Value | 95% CI Lower | 95% CI Upper | N Input SNPs | N Informative SNPs | N Horizontal Pleiotropic SNPs | FDR P-Value | FDR Significant |
|---|---|---|---|---|---|---|---|---|---|---|---|---|---|---|
| 1 | Chr2:55.2Mb | EUR | Forward MR | -0.387 | 0.971 | -0.398 | 6.906E-01 | -2.290 | 1.517 | 84 | 4 | 3 | 7.620E-01 | FALSE |
| 1 | Chr2:55.2Mb | EUR | Reverse MR | 0.065 | 0.056 | 1.170 | 2.418E-01 | -0.044 | 0.174 | 91 | 3 | 12 | 4.338E-01 | FALSE |
| 10 | Chr19:45.3Mb_10 | EUR | Forward MR | -1.846 | 1.969 | -0.938 | 3.485E-01 | -5.704 | 2.013 | 259 | 4 | 118 | 5.310E-01 | FALSE |
| 10 | Chr19:45.3Mb_10 | EUR | Reverse MR | -0.023 | 0.004 | -5.321 | 1.030E-07 | -0.031 | -0.014 | 281 | 5 | 4 | 4.400E-07 | TRUE |
| 11 | Chr19:45.3Mb_11 | EUR | Forward MR | 1.670 | 2.224 | 0.751 | 4.527E-01 | -2.689 | 6.030 | 258 | 4 | 114 | 6.036E-01 | FALSE |
| 11 | Chr19:45.3Mb_11 | EUR | Reverse MR | -0.023 | 0.004 | -5.316 | 1.060E-07 | -0.031 | -0.014 | 280 | 5 | 4 | 4.400E-07 | TRUE |
| 12 | Chr19:45.4Mb_12 | EUR | Forward MR | -0.566 | 2.157 | -0.262 | 7.930E-01 | -4.795 | 3.662 | 263 | 4 | 115 | 8.186E-01 | FALSE |
| 12 | Chr19:45.4Mb_12 | EUR | Reverse MR | -0.023 | 0.004 | -5.347 | 8.960E-08 | -0.031 | -0.015 | 285 | 5 | 4 | 4.400E-07 | TRUE |
| 13a | Chr19:45.4Mb_13a | EUR | Forward MR | -1.094 | 2.142 | -0.511 | 6.096E-01 | -5.291 | 3.104 | 265 | 5 | 117 | 7.225E-01 | FALSE |
| 13a | Chr19:45.4Mb_13a | EUR | Reverse MR | -0.023 | 0.004 | -5.309 | 1.100E-07 | -0.031 | -0.014 | 288 | 5 | 5 | 4.400E-07 | TRUE |
| 13b | Chr19:45.4Mb_13b | EUR | Forward MR | -1.094 | 2.142 | -0.511 | 6.096E-01 | -5.291 | 3.104 | 265 | 5 | 117 | 7.225E-01 | FALSE |
| 13b | Chr19:45.4Mb_13b | EUR | Reverse MR | -0.023 | 0.004 | -5.309 | 1.100E-07 | -0.031 | -0.014 | 288 | 5 | 5 | 4.400E-07 | TRUE |
| 13c | Chr19:45.4Mb_13c | EUR | Forward MR | -1.094 | 2.142 | -0.511 | 6.096E-01 | -5.291 | 3.104 | 265 | 5 | 117 | 7.225E-01 | FALSE |
| 13c | Chr19:45.4Mb_13c | EUR | Reverse MR | -0.023 | 0.004 | -5.309 | 1.100E-07 | -0.031 | -0.014 | 288 | 5 | 5 | 4.400E-07 | TRUE |
| 16 | Chr19:46.3Mb | EUR | Forward MR | 1.957 | 0.573 | 3.415 | 6.376E-04 | 0.834 | 3.081 | 42 | 9 | 10 | 2.024E-03 | TRUE |
| 16 | Chr19:46.3Mb | EUR | Reverse MR | -0.090 | 0.080 | -1.132 | 2.576E-01 | -0.247 | 0.066 | 44 | 6 | 8 | 4.338E-01 | FALSE |
| 2 | Chr6:47.7Mb | EUR | Forward MR | 2.539 | 0.189 | 13.451 | 3.040E-41 | 2.169 | 2.909 | 21 | 1 | 6 | 4.864E-40 | TRUE |
| 2 | Chr6:47.7Mb | EUR | Reverse MR | 0.038 | 0.033 | 1.145 | 2.522E-01 | -0.027 | 0.102 | 26 | 15 | 2 | 4.338E-01 | FALSE |
| 3 | Chr8:96Mb | EUR | Forward MR | -0.552 | 0.680 | -0.812 | 4.168E-01 | -1.885 | 0.781 | 22 | 2 | 3 | 5.799E-01 | FALSE |
| 3 | Chr8:96Mb | EUR | Reverse MR | -0.067 | 0.063 | -1.077 | 2.814E-01 | -0.190 | 0.055 | 23 | 19 | 2 | 4.502E-01 | FALSE |
| 4 | Chr10:82.3Mb | EUR | Forward MR | -2.277 | 0.751 | -3.033 | 2.423E-03 | -3.749 | -0.806 | 57 | 7 | 8 | 5.964E-03 | TRUE |
| 4 | Chr10:82.3Mb | EUR | Reverse MR | 0.029 | 0.071 | 0.412 | 6.804E-01 | -0.109 | 0.168 | 64 | 9 | 11 | 7.620E-01 | FALSE |
| 5 | Chr11:47.4Mb | EUR | Forward MR | 2.849 | 0.840 | 3.391 | 6.958E-04 | 1.202 | 4.496 | 53 | 7 | 6 | 2.024E-03 | TRUE |
| 5 | Chr11:47.4Mb | EUR | Reverse MR | 0.144 | 0.058 | 2.469 | 1.354E-02 | 0.030 | 0.258 | 21 | 9 | 6 | 2.888E-02 | TRUE |
| 6 | Chr11:85.7Mb | EUR | Forward MR | 0.711 | 0.206 | 3.448 | 5.654E-04 | 0.307 | 1.115 | 40 | 15 | 3 | 2.010E-03 | TRUE |
| 6 | Chr11:85.7Mb | EUR | Reverse MR | -0.082 | 0.055 | -1.482 | 1.383E-01 | -0.190 | 0.026 | 43 | 5 | 9 | 2.767E-01 | FALSE |
| 7 | Chr15:82.4Mb | EUR | Forward MR | -0.972 | 0.360 | -2.702 | 6.902E-03 | -1.677 | -0.267 | 26 | 11 | 5 | 1.578E-02 | TRUE |
| 7 | Chr15:82.4Mb | EUR | Reverse MR | -0.024 | 0.069 | -0.353 | 7.240E-01 | -0.159 | 0.110 | 26 | 8 | 5 | 7.722E-01 | FALSE |
| 8 | Chr16:20.5Mb | EUR | Forward MR | 0.128 | 0.147 | 0.870 | 3.844E-01 | -0.160 | 0.415 | 115 | 6 | 5 | 5.591E-01 | FALSE |
| 8 | Chr16:20.5Mb | EUR | Reverse MR | -0.005 | 0.056 | -0.088 | 9.300E-01 | -0.115 | 0.105 | 124 | 1 | 20 | 9.300E-01 | FALSE |
| 9 | Chr17:5.1Mb | EUR | Forward MR | 2.922 | 0.097 | 30.224 | 1.130E-200 | 2.733 | 3.112 | 27 | 9 | 2 | 3.616E-199 | TRUE |
| 9 | Chr17:5.1Mb | EUR | Reverse MR | 0.208 | 0.065 | 3.197 | 1.387E-03 | 0.080 | 0.335 | 15 | 8 | 10 | 3.698E-03 | TRUE |

**Supplementary Table S30. Instrumental Variables for Forward Cis-MR Analysis (eGFR to AD).**
This table provides detailed information about the instrumental variables (genetic variants) used in forward cis-MR analysis (eGFR as exposure, AD as outcome) for each locus. Understanding which variants serve as instruments is critical for interpreting causal estimates and assessing the validity of MR assumptions. For each locus, the table reports: locus number and label, the complete list of input SNPs considered for the analysis, SNPs identified as informative instruments after SuSiE fine-mapping, SNPs excluded due to pleiotropy concerns (showing independent effects on the outcome), and the top SNPs with their posterior inclusion probabilities (PIPs) from fine-mapping. High-PIP SNPs represent the most likely causal variants for the exposure (eGFR) within each locus. The identification and exclusion of pleiotropic SNPs is essential for satisfying the MR exclusion restriction assumption, which requires that instruments affect the outcome only through the exposure.

| locus_num | locus | input_snps | informative_snps | pleiotropy_snps | top_snps_pip |
|---|---|---|---|---|---|
| 1 | Chr2:55.2Mb | rs17045898;rs10201566;rs59531970;rs114437785;rs4426561;rs4024576;rs4671934;rs139103761;rs115486808;rs4362595;rs35028480;rs76279463;rs143435618;rs72804692;rs189992573;rs56381959;rs6545423;rs150747574;rs75475627;rs10205182;rs10205410;rs62134681;rs77456006;rs79254608;rs6726004;rs6741053;rs12624153;rs2941580;rs6760298;rs55827937;rs2271322;rs28710338;rs2971884;rs2941589;rs354229;rs181347;rs12713275;rs354211;rs35006042;rs354243;rs72913519;rs78694233;rs143216688;rs1510613;rs17046269;rs1510614;rs35880160;rs11125546;rs165048;rs165050;rs2063391;rs164943;rs62136882;rs165052;rs165053;rs17046344;rs13416919;rs2578714;rs34170608;rs2567976;rs2631839;rs12467761;rs2631842;rs12713277;rs12713281;rs17046442;rs2163618;rs6545453;rs13408207;rs13430119;rs6744548;rs75010292;rs4625954;rs6725187;rs6745543;rs3821111;rs17347222;rs78539852;rs2920843;rs35950075;rs72799507;rs1134722;rs2589073;rs6721972 | rs4426561;rs4024576;rs2271322;rs354211 | rs75010292;rs17347222;rs78539852 | rs354211:0.9994;rs2271322:0.9785;rs4426561:0.9254;rs4024576:0.2297;rs150747574:0.0809 |
| 10 | Chr19:45.3Mb_10 | rs76602578;rs1836280;rs2722683;rs80257887;rs75161053;rs139140823;rs62118204;rs1727760;rs12459712;rs1727762;rs74172479;rs846881;rs846876;rs16979292;rs846892;rs11882134;rs112027066;rs11671606;rs62119270;rs58967105;rs7260482;rs714948;rs111780744;rs2289494;rs62119328;rs2289495;rs150735888;rs2965156;rs11879980;rs111358663;rs151330717;rs62120566;rs2965162;rs73035597;rs2965163;rs12150984;rs2965164;rs2919848;rs143934888;rs62120575;rs111278137;rs79745266;rs4516335;rs11881756;rs79638902;rs73037426;rs150820726;rs7254776;rs186110295;rs8108110;rs80307900;rs74607435;rs77775146;rs73048230;rs2927437;rs192016015;rs12459810;rs2965169;rs8100239;rs10401176;rs8103315;rs111677968;rs62117206;rs2006134;rs145414981;rs139326841;rs35106910;rs112450640;rs139425456;rs2889414;rs10419669;rs148933445;rs2967668;rs139246905;rs28399596;rs140824606;rs73048258;rs28399649;rs28399607;rs2968180;rs117737673;rs1135062;rs28399635;rs28399663;rs28399637;rs7026;rs28399664;rs78986976;rs7343130;rs4803760;rs2972555;rs56394238;rs10407439;rs1466435;rs55840414;rs28615360;rs111371860;rs77241309;rs41289512;rs1871046;rs12974942;rs57537848;rs2972566;rs2972558;rs10421035;rs7249933;rs10410835;rs56317818;rs12463239;rs440277;rs365653;rs12978931;rs394353;rs112283362;rs3112439;rs112422902;rs41290100;rs41290102;rs8104483;rs387465;rs11879589;rs71352236;rs393584;rs2075642;rs41290108;rs73050293;rs11668861;rs6859;rs41290120;rs11673139;rs3852861;rs148303016;rs11083749;rs406315;rs113743631;rs12972156;rs283810;rs283814;rs76692773;rs157580;rs73936968;rs77301115;rs1160983;rs11668327;rs35568738;rs117264457;rs405697;rs117843462;rs7259620;rs449647;rs769446;rs429358;rs7412;rs75627662;rs72654472;rs72654437;rs59325138;rs72654445;rs12691088;rs3925681;rs12721046;rs1064725;rs144311893;rs157595;rs75654248;rs141622900;rs4803771;rs71352239;rs60049679;rs11878790;rs113345881;rs8106813;rs73045691;rs76214972;rs12721109;rs1132899;rs117316645;rs143674704;rs | rs1836280;rs8104483;rs1160983;rs429358 | rs76602578;rs75161053;rs74172479;rs846892;rs11882134;rs112027066;rs2289494;rs62119328;rs2965156;rs111358663;rs2965162;rs12150984;rs73048230;rs192016015;rs8103315;rs111677968;rs2006134;rs35106910;rs139425456;rs2889414;rs2967668;rs28399596;rs140824606;rs28399607;rs2968180;rs28399635;rs28399663;rs28399637;rs78986976;rs7343130;rs4803760;rs2972555;rs10407439;rs55840414;rs1871046;rs57537848;rs2972566;rs2972558;rs12463239;rs394353;rs112283362;rs112422902;rs8104483;rs387465;rs11879589;rs393584;rs2075642;rs73050293;rs41290120;rs11673139;rs3852861;rs11083749;rs12972156;rs283810;rs283814;rs76692773;rs157580;rs73936968;rs1160983;rs11668327;rs35568738;rs405697;rs117843462;rs7259620;rs449647;rs429358;rs7412;rs75627662;rs72654437;rs59325138;rs72654445;rs12691088;rs3925681;rs12721046;rs1064725;rs144311893;rs157595;rs75654248;rs141622900;rs4803771;rs71352239;rs60049679;rs11878790;rs113345881;rs73045691;rs12721109;rs1132899;rs143674704;rs204906;rs10775543;rs204478;rs35194062;rs28809016;rs35194383;rs1971786;rs34583389;rs1076986;rs34293391;rs7259034;rs149013045;rs79067230;rs10413253;rs75463276;rs80182863;rs10421247;rs10416371;rs149553138;rs76971643;rs150685845;rs8113514;rs12977719;rs117783785;rs631920;rs4802250;rs10417555;rs12461144;rs10407278;rs73036542 | rs429358:1;rs8104483:0.9246;rs1160983:0.4918;rs1836280:0.2189;rs394353:0.0787 |

| locus_num | locus | input_snps | informative_snps | pleiotropy_snps | top_snps_pip |
|---|---|---|---|---|---|
| | | 75178253;rs204906;rs138692741;rs117198034;rs10775543;rs204478;rs193143021;rs741767;rs117261169;rs2376866;rs35194062;rs28809016;rs35194383;rs116518981;rs35912475;rs4803791;rs62117224;rs1971786;rs34583389;rs1076986;rs34293391;rs139126319;rs10856;rs1968449;rs112481437;rs147188206;rs7259034;rs112394906;rs60239918;rs8100183;rs6509181;rs149013045;rs138625214;rs12461065;rs79067230;rs11083760;rs10413253;rs79322367;rs17643262;rs142883151;rs75463276;rs116863911;rs78073763;rs80182863;rs10421247;rs10416371;rs149553138;rs76971643;rs150685845;rs4580299;rs76133116;rs116958755;rs75858218;rs12460985;rs62118471;rs12461027;rs111243475;rs8113514;rs12977719;rs117783785;rs631920;rs2627642;rs620807;rs8105802;rs4802250;rs182645183;rs10417555;rs10405928;rs11667509;rs10415392;rs12461144;rs10407278;rs346773;rs10402739;rs62118504;rs75727214;rs346748;rs11673093;rs77740243;rs73036542;rs1872259 | | | |
| 11 | Chr19:45.3Mb_11 | rs1836280;rs2722683;rs80257887;rs75161053;rs139140823;rs62118204;rs1727760;rs12459712;rs1727762;rs74172479;rs846881;rs846876;rs16979292;rs846892;rs11882134;rs112027066;rs11671606;rs62119270;rs58967105;rs7260482;rs714948;rs111780744;rs2289494;rs62119328;rs2289495;rs150735888;rs2965156;rs11879980;rs111358663;rs151330717;rs62120566;rs2965162;rs73035597;rs2965163;rs12150984;rs2965164;rs2919848;rs143934888;rs62120575;rs111278137;rs79745266;rs4516335;rs11881756;rs79638902;rs73037426;rs150820726;rs7254776;rs186110295;rs8108110;rs80307900;rs74607435;rs77775146;rs73048230;rs2927437;rs192016015;rs12459810;rs2965169;rs8100239;rs10401176;rs8103315;rs111677968;rs62117206;rs2006134;rs145414981;rs139326841;rs35106910;rs112450640;rs139425456;rs2889414;rs10419669;rs148933445;rs2967668;rs139246905;rs28399596;rs140824606;rs73048258;rs28399649;rs28399607;rs2968180;rs117737673;rs1135062;rs28399635;rs28399663;rs28399637;rs7026;rs28399664;rs78986976;rs7343130;rs4803760;rs2972555;rs56394238;rs10407439;rs1466435;rs55840414;rs28615360;rs111371860;rs77241309;rs41289512;rs1871046;rs12974942;rs57537848;rs2972566;rs2972558;rs10421035;rs7249933;rs10410835;rs56317818;rs12463239;rs440277;rs365653;rs12978931;rs394353;rs112283362;rs3112439;rs112422902;rs41290100;rs41290102;rs8104483;rs387465;rs11879589;rs71352236;rs393584;rs2075642;rs41290108;rs73050293;rs11668861;rs6859;rs41290120;rs11673139;rs3852861;rs148303016;rs11083749;rs406315;rs113743631;rs12972156;rs283810;rs283814;rs76692773;rs157580;rs73936968;rs77301115;rs1160983;rs11668327;rs35568738;rs117264457;rs405697;rs117843462;rs7259620;rs449647;rs769446;rs429358;rs7412;rs75627662;rs72654472;rs72654437;rs59325138;rs72654445;rs12691088;rs3925681;rs12721046;rs1064725;rs144311893;rs157595;rs75654248;rs141622900;rs4803771;rs71352239;rs60049679;rs11878790;rs113345881;rs8106813;rs73045691;rs76214972;rs12721109;rs1132899;rs117316645;rs143674704;rs75178253;rs204906;rs138692741;rs117198034;rs10775543;rs204478;rs193143021;rs741767;rs117261169;rs2376866;rs35194062;rs28809016;rs35194383;rs116518981;rs35912475;rs4803791;rs62117224;rs1971786;rs34583389;rs1076986;rs34293391;rs139126319;rs10856;rs1968449;rs112481437;rs147188206;rs7259034;rs112394906;rs60239918;rs8100183;rs6509181;rs149013045;rs138625214;rs12461065;rs79067230;rs11083760;rs10413253;rs79322367;rs17643262;rs142883151;rs75463276;rs116863911;rs78073763;rs80182863;rs10421247;rs10416371;rs149553138;rs76971643;rs150685845;rs4580299;rs76133116;rs116958755;rs75858218;rs12460985;rs62118471;rs12461027;rs111243475;rs8113514;rs12977719;rs117783785;rs631920;rs2627642;rs620807;rs8105802;rs4802250;rs182645183;rs10417555;rs10405928;rs11667509;rs10415392;rs12461144;rs10407278;rs346773;rs10402739;rs62118504;rs75727214;rs346748;rs11673093;rs77740243;rs73036542;rs345421 | rs1836280;rs8104483;rs1160983;rs429358 | rs75161053;rs74172479;rs846892;rs11882134;rs112027066;rs2289494;rs62119328;rs2965156;rs2965162;rs12150984;rs80307900;rs73048230;rs192016015;rs8103315;rs111677968;rs2006134;rs35106910;rs139425456;rs2889414;rs2967668;rs28399596;rs28399607;rs2968180;rs28399635;rs28399663;rs28399637;rs7026;rs78986976;rs7343130;rs4803760;rs2972555;rs10407439;rs1871046;rs57537848;rs2972566;rs2972558;rs12463239;rs112283362;rs3112439;rs112422902;rs41290100;rs8104483;rs387465;rs11879589;rs73050293;rs41290120;rs3852861;rs11083749;rs12972156;rs283810;rs283814;rs76692773;rs157580;rs73936968;rs1160983;rs11668327;rs35568738;rs405697;rs117843462;rs7259620;rs449647;rs429358;rs7412;rs75627662;rs72654437;rs59325138;rs72654445;rs12691088;rs3925681;rs12721046;rs1064725;rs144311893;rs157595;rs75654248;rs141622900;rs4803771;rs71352239;rs60049679;rs11878790;rs113345881;rs73045691;rs12721109;rs1132899;rs143674704;rs204906;rs10775543;rs204478;rs35194062;rs28809016;rs35194383;rs1971786;rs34583389;rs1076986;rs34293391;rs7259034;rs149013045;rs138625214;rs79067230;rs10413253;rs75463276;rs80182863;rs10421247;rs10416371;rs149553138;rs76971643;rs150685845;rs8113514;rs12977719;rs117783785;rs631920;rs10417555;rs12461144;rs10407278;rs73036542 | rs429358:1;rs8104483:0.9246;rs1160983:0.4925;rs1836280:0.2192;rs394353:0.0787 |
| 12 | Chr19: | rs1836280;rs2722683;rs80257887;rs75161053;rs139140823;rs62118204;rs1727760;rs12459712;rs1727762;rs74172479;rs846881;rs846876;rs16979292;rs846892;rs11882134;rs112027066;rs11671606;rs6 | rs1836280;rs8104483;rs1160983;rs429358 | rs75161053;rs74172479;rs846892;rs11882134;rs112027066;rs2289494;rs62119328;rs2965156;rs2965162;rs12150984;rs74607435;rs73048230;rs192016015;rs8103315;rs111677968;rs2006134;rs3510691 | rs429358:1;rs8104483:0.9249;rs1160983:0.4895;rs18362 |

| locus_num | locus | input_snps | informative_snps | pleiotropy_snps | top_snps_pip |
|---|---|---|---|---|---|
| | 45.4Mb_12 | 2119270;rs58967105;rs7260482;rs714948;rs111780744;rs2289494;rs62119328;rs2289495;rs150735888;rs2965156;rs11879980;rs111358663;rs151330717;rs62120566;rs2965162;rs73035597;rs2965163;rs12150984;rs2965164;rs2919848;rs143934888;rs62120575;rs111278137;rs79745266;rs4516335;rs11881756;rs79638902;rs73037426;rs150820726;rs7254776;rs186110295;rs8108110;rs80307900;rs74607435;rs77775146;rs73048230;rs2927437;rs192016015;rs12459810;rs2965169;rs8100239;rs10401176;rs8103315;rs111677968;rs62117206;rs2006134;rs145414981;rs139326841;rs35106910;rs112450640;rs139425456;rs2889414;rs10419669;rs148933445;rs2967668;rs139246905;rs28399596;rs140824606;rs73048258;rs28399649;rs28399607;rs2968180;rs117737673;rs1135062;rs28399635;rs28399663;rs28399637;rs7026;rs28399664;rs78986976;rs7343130;rs4803760;rs2972555;rs56394238;rs10407439;rs1466435;rs55840414;rs28615360;rs111371860;rs77241309;rs41289512;rs1871046;rs12974942;rs57537848;rs2972566;rs2972558;rs10421035;rs7249933;rs10410835;rs56317818;rs12463239;rs440277;rs365653;rs12978931;rs394353;rs112283362;rs3112439;rs112422902;rs41290100;rs41290102;rs8104483;rs387465;rs11879589;rs71352236;rs393584;rs2075642;rs41290108;rs73050293;rs11668861;rs6859;rs41290120;rs11673139;rs3852861;rs148303016;rs11083749;rs406315;rs113743631;rs12972156;rs283810;rs283814;rs76692773;rs157580;rs73936968;rs77301115;rs1160983;rs11668327;rs35568738;rs117264457;rs405697;rs117843462;rs7259620;rs449647;rs769446;rs429358;rs7412;rs75627662;rs72654472;rs72654437;rs59325138;rs72654445;rs12691088;rs3925681;rs12721046;rs1064725;rs144311893;rs157595;rs75654248;rs141622900;rs4803771;rs71352239;rs60049679;rs11878790;rs113345881;rs8106813;rs73045691;rs76214972;rs12721109;rs1132899;rs117316645;rs143674704;rs75178253;rs204906;rs138692741;rs117198034;rs10775543;rs204478;rs193143021;rs741767;rs117261169;rs2376866;rs35194062;rs28809016;rs35194383;rs116518981;rs35912475;rs4803791;rs62117224;rs1971786;rs34583389;rs1076986;rs34293391;rs139126319;rs10856;rs1968449;rs112481437;rs147188206;rs7259034;rs112394906;rs60239918;rs8100183;rs6509181;rs149013045;rs138625214;rs12461065;rs79067230;rs11083760;rs10413253;rs79322367;rs17643262;rs142883151;rs75463276;rs116863911;rs78073763;rs80182863;rs10421247;rs10416371;rs149553138;rs76971643;rs150685845;rs4580299;rs76133116;rs116958755;rs75858218;rs12460985;rs62118471;rs12461027;rs111243475;rs8113514;rs12977719;rs117783785;rs631920;rs2627642;rs620807;rs8105802;rs4802250;rs182645183;rs10417555;rs10405928;rs11667509;rs10415392;rs12461144;rs10407278;rs346773;rs10402739;rs62118504;rs75727214;rs346748;rs11673093;rs77740243;rs73036542;rs345421;rs344793;rs144645090;rs12983944;rs344818;rs12973593 | | 0;rs139425456;rs2889414;rs2967668;rs28399596;rs140824606;rs28399607;rs2968180;rs28399635;rs28399663;rs28399637;rs7026;rs78986976;rs7343130;rs4803760;rs2972555;rs10407439;rs1871046;rs57537848;rs2972566;rs2972558;rs12463239;rs112283362;rs112422902;rs8104483;rs387465;rs11879589;rs73050293;rs41290120;rs3852861;rs11083749;rs12972156;rs283810;rs283814;rs76692773;rs157580;rs73936968;rs1160983;rs11668327;rs35568738;rs405697;rs117843462;rs7259620;rs449647;rs429358;rs7412;rs75627662;rs72654437;rs59325138;rs72654445;rs12691088;rs3925681;rs12721046;rs1064725;rs144311893;rs157595;rs75654248;rs141622900;rs4803771;rs71352239;rs60049679;rs11878790;rs113345881;rs73045691;rs12721109;rs1132899;rs143674704;rs204906;rs10775543;rs204478;rs35194062;rs28809016;rs35194383;rs1971786;rs34583389;rs1076986;rs34293391;rs1968449;rs7259034;rs149013045;rs79067230;rs10413253;rs75463276;rs80182863;rs10421247;rs149553138;rs76971643;rs150685845;rs8113514;rs12977719;rs117783785;rs631920;rs4802250;rs10417555;rs12461144;rs10407278;rs73036542;rs144645090;rs12983944 | 80:0.2179;rs394353:0.0781 |
| 13a | Chr19:45.4Mb_13a | rs2722683;rs80257887;rs75161053;rs139140823;rs62118204;rs1727760;rs12459712;rs1727762;rs74172479;rs846881;rs846876;rs16979292;rs846892;rs11882134;rs112027066;rs11671606;rs62119270;rs58967105;rs7260482;rs714948;rs111780744;rs2289494;rs62119328;rs2289495;rs150735888;rs2965156;rs11879980;rs111358663;rs151330717;rs62120566;rs2965162;rs73035597;rs2965163;rs12150984;rs2965164;rs2919848;rs143934888;rs62120575;rs111278137;rs79745266;rs4516335;rs11881756;rs79638902;rs73037426;rs150820726;rs7254776;rs186110295;rs8108110;rs80307900;rs74607435;rs77775146;rs73048230;rs2927437;rs192016015;rs12459810;rs2965169;rs8100239;rs10401176;rs8103315;rs111677968;rs62117206;rs2006134;rs145414981;rs139326841;rs35106910;rs112450640;rs139425456;rs2889414;rs10419669;rs148933445;rs2967668;rs139246905;rs28399596;rs140824606;rs73048258;rs28399649;rs28399607;rs2968180;rs117737673;rs1135062;rs28399635;rs28399663;rs28399637;rs7026;rs28399664;rs78986976;rs7343130;rs4803760;rs2972555;rs56394238;rs10407439;rs1466435;rs55840414;rs28615360;rs111371860;rs77241309;rs41289512;rs1871046;rs12974942;rs57537848;rs2972566;rs2972558;rs10421035;rs7249933;rs10410835;rs56317818;rs12463239;rs440277;rs365653;rs1 | rs8104483;rs1160983;rs429358;rs3916898;rs34631405 | rs75161053;rs74172479;rs846892;rs11882134;rs112027066;rs2289494;rs62119328;rs2965156;rs2965162;rs12150984;rs74607435;rs73048230;rs192016015;rs8103315;rs111677968;rs2006134;rs35106910;rs139425456;rs2889414;rs2967668;rs28399596;rs140824606;rs28399607;rs2968180;rs117737673;rs28399635;rs28399663;rs28399637;rs7026;rs78986976;rs7343130;rs4803760;rs2972555;rs10407439;rs1871046;rs57537848;rs2972566;rs2972558;rs12463239;rs112283362;rs112422902;rs8104483;rs387465;rs11879589;rs73050293;rs41290120;rs3852861;rs11083749;rs12972156;rs283810;rs283814;rs76692773;rs157580;rs73936968;rs1160983;rs11668327;rs35568738;rs405697;rs117843462;rs7259620;rs449647;rs429358;rs7412;rs75627662;rs72654437;rs59325138;rs72654445;rs12691088;rs3925681;rs12721046;rs1064725;rs144311893;rs157595;rs75654248;rs141622900;rs4803771;rs71352239;rs60049679;rs11878790;rs113345881;rs73045691;rs12721109;rs1132899;rs143674704;rs204906;rs10775543;rs204478;rs35194062;rs28809016;rs35194383;rs1971786;rs34583389;rs1076986;rs34293391;rs1968449;rs7259034;rs149013045;rs79067230;rs10413253;rs75463276;rs80182863;rs10421247;rs149553138;rs76971643;rs150685845;rs8113514;rs12977719;rs117783785;rs631920;rs4802250;rs10417555;r | rs429358:1;rs8104483:0.9305;rs1160983:0.5042;rs34631405:0.1332;rs3916898:0.119 |

| locus_num | locus | input_snps | informative_snps | pleiotropy_snps | top_snps_pip |
| --- | --- | --- | --- | --- | --- |
| | | 2978931;rs394353;rs112283362;rs3112439;rs112422902;rs41290100;rs41290102;rs8104483;rs387465;rs11879589;rs71352236;rs393584;rs2075642;rs41290108;rs73050293;rs11668861;rs6859;rs41290120;rs11673139;rs3852861;rs148303016;rs11083749;rs406315;rs113743631;rs12972156;rs283810;rs283814;rs76692773;rs157580;rs73936968;rs77301115;rs1160983;rs11668327;rs35568738;rs117264457;rs405697;rs117843462;rs7259620;rs449647;rs769446;rs429358;rs7412;rs75627662;rs72654472;rs72654437;rs59325138;rs72654445;rs12691088;rs3925681;rs12721046;rs1064725;rs144311893;rs157595;rs75654248;rs141622900;rs4803771;rs71352239;rs60049679;rs11878790;rs113345881;rs8106813;rs73045691;rs76214972;rs12721109;rs1132899;rs117316645;rs143674704;rs75178253;rs204906;rs138692741;rs117198034;rs10775543;rs204478;rs193143021;rs741767;rs117261169;rs2376866;rs35194062;rs28809016;rs35194383;rs116518981;rs35912475;rs4803791;rs62117224;rs1971786;rs34583389;rs1076986;rs34293391;rs139126319;rs10856;rs1968449;rs112481437;rs147188206;rs7259034;rs112394906;rs60239918;rs8100183;rs6509181;rs149013045;rs138625214;rs12461065;rs79067230;rs11083760;rs10413253;rs79322367;rs17643262;rs142883151;rs75463276;rs116863911;rs78073763;rs80182863;rs10421247;rs10416371;rs149553138;rs76971643;rs150685845;rs4580299;rs76133116;rs116958755;rs75858218;rs12460985;rs62118471;rs12461027;rs111243475;rs8113514;rs12977719;rs117783785;rs631920;rs2627642;rs620807;rs8105802;rs4802250;rs182645183;rs10417555;rs10405928;rs11667509;rs10415392;rs12461144;rs10407278;rs346773;rs10402739;rs62118504;rs75727214;rs346748;rs11673093;rs77740243;rs73036542;rs345421;rs344793;rs144645090;rs12983944;rs344818;rs12973593;rs3916898;rs8109829;rs34631405 | | s12461144;rs10407278;rs73036542;rs144645090;rs12983944;rs3916898 | |
| 13b | Chr19:45.4Mb_13b | rs2722683;rs80257887;rs75161053;rs139140823;rs62118204;rs1727760;rs12459712;rs1727762;rs74172479;rs846881;rs846876;rs16979292;rs846892;rs11882134;rs112027066;rs11671606;rs62119270;rs58967105;rs7260482;rs714948;rs111780744;rs2289494;rs62119328;rs2289495;rs150735888;rs2965156;rs11879980;rs111358663;rs151330717;rs62120566;rs2965162;rs73035597;rs2965163;rs12150984;rs2965164;rs2919848;rs143934888;rs62120575;rs111278137;rs79745266;rs4516335;rs11881756;rs79638902;rs73037426;rs150820726;rs7254776;rs186110295;rs8108110;rs80307900;rs74607435;rs77775146;rs73048230;rs2927437;rs192016015;rs12459810;rs2965169;rs8100239;rs10401176;rs8103315;rs111677968;rs62117206;rs2006134;rs145414981;rs139326841;rs35106910;rs112450640;rs139425456;rs2889414;rs10419669;rs148933445;rs2967668;rs139246905;rs28399596;rs140824606;rs73048258;rs28399649;rs28399607;rs2968180;rs117737673;rs1135062;rs28399635;rs28399663;rs28399637;rs7026;rs28399664;rs78986976;rs7343130;rs4803760;rs2972555;rs56394238;rs10407439;rs1466435;rs55840414;rs28615360;rs111371860;rs77241309;rs41289512;rs1871046;rs12974942;rs57537848;rs2972566;rs2972558;rs10421035;rs7249933;rs10410835;rs56317818;rs12463239;rs440277;rs365653;rs12978931;rs394353;rs112283362;rs3112439;rs112422902;rs41290100;rs41290102;rs8104483;rs387465;rs11879589;rs71352236;rs393584;rs2075642;rs41290108;rs73050293;rs11668861;rs6859;rs41290120;rs11673139;rs3852861;rs148303016;rs11083749;rs406315;rs113743631;rs12972156;rs283810;rs283814;rs76692773;rs157580;rs73936968;rs77301115;rs1160983;rs11668327;rs35568738;rs117264457;rs405697;rs117843462;rs7259620;rs449647;rs769446;rs429358;rs7412;rs75627662;rs72654472;rs72654437;rs59325138;rs72654445;rs12691088;rs3925681;rs12721046;rs1064725;rs144311893;rs157595;rs75654248;rs141622900;rs4803771;rs71352239;rs60049679;rs11878790;rs113345881;rs8106813;rs73045691;rs76214972;rs12721109;rs1132899;rs117316645;rs143674704;rs75178253;rs204906;rs138692741;rs117198034;rs10775543;rs204478;rs193143021;rs741767;rs117261169;rs2376866;rs35194062;rs28809016;rs35194383;rs116518981;rs35912475;rs4803791;rs62117224;rs1971786;rs34583389;rs1076986;rs34293391;rs139126319;rs10856;rs1968449;rs112481437;rs147188206;rs7259034;rs11 | rs8104483;rs1160983;rs429358;rs3916898;rs34631405 | rs75161053;rs74172479;rs846892;rs11882134;rs112027066;rs2289494;rs62119328;rs2965156;rs2965162;rs12150984;rs74607435;rs73048230;rs192016015;rs8103315;rs111677968;rs2006134;rs35106910;rs139425456;rs2889414;rs2967668;rs28399596;rs140824606;rs28399607;rs2968180;rs117737673;rs28399635;rs28399663;rs28399637;rs7026;rs78986976;rs7343130;rs4803760;rs2972555;rs10407439;rs1871046;rs57537848;rs2972566;rs2972558;rs12463239;rs112283362;rs112422902;rs8104483;rs387465;rs11879589;rs73050293;rs41290120;rs3852861;rs11083749;rs12972156;rs283810;rs283814;rs76692773;rs157580;rs73936968;rs1160983;rs11668327;rs35568738;rs405697;rs117843462;rs7259620;rs449647;rs429358;rs7412;rs75627662;rs72654437;rs59325138;rs72654445;rs12691088;rs3925681;rs12721046;rs1064725;rs144311893;rs157595;rs75654248;rs141622900;rs4803771;rs71352239;rs60049679;rs11878790;rs113345881;rs73045691;rs12721109;rs1132899;rs143674704;rs204906;rs10775543;rs204478;rs35194062;rs28809016;rs35194383;rs1971786;rs34583389;rs1076986;rs34293391;rs1968449;rs7259034;rs149013045;rs79067230;rs10413253;rs75463276;rs80182863;rs10421247;rs149553138;rs76971643;rs150685845;rs8113514;rs12977719;rs117783785;rs631920;rs4802250;rs10417555;rs12461144;rs10407278;rs73036542;rs144645090;rs12983944;rs3916898 | rs429358:1;rs8104483:0.9305;rs1160983:0.5042;rs34631405:0.1332;rs3916898:0.119 |

| locus_num | locus | input_snps | informative_snps | pleiotropy_snps | top_snps_pip |
|---|---|---|---|---|---|
| | | 2394906;rs60239918;rs8100183;rs6509181;rs149013045;rs138625214;rs12461065;rs79067230;rs11083760;rs10413253;rs79322367;rs17643262;rs142883151;rs75463276;rs116863911;rs78073763;rs80182863;rs10421247;rs10416371;rs149553138;rs76971643;rs150685845;rs4580299;rs76133116;rs116958755;rs75858218;rs12460985;rs62118471;rs12461027;rs111243475;rs8113514;rs12977719;rs117783785;rs631920;rs2627642;rs620807;rs8105802;rs4802250;rs182645183;rs10417555;rs10405928;rs11667509;rs10415392;rs12461144;rs10407278;rs346773;rs10402739;rs62118504;rs75727214;rs346748;rs11673093;rs77740243;rs73036542;rs345421;rs344793;rs144645090;rs12983944;rs344818;rs12973593;rs3916898;rs8109829;rs34631405 | | | |
| 13c | Chr19:45.4Mb_13c | rs2722683;rs80257887;rs75161053;rs139140823;rs62118204;rs1727760;rs12459712;rs1727762;rs74172479;rs846881;rs846876;rs16979292;rs846892;rs11882134;rs112027066;rs11671606;rs62119270;rs58967105;rs7260482;rs714948;rs111780744;rs2289494;rs62119328;rs2289495;rs150735888;rs2965156;rs11879980;rs111358663;rs151330717;rs62120566;rs2965162;rs73035597;rs2965163;rs12150984;rs2965164;rs2919848;rs143934888;rs62120575;rs111278137;rs79745266;rs4516335;rs11881756;rs79638902;rs73037426;rs150820726;rs7254776;rs186110295;rs8108110;rs80307900;rs74607435;rs77775146;rs73048230;rs2927437;rs192016015;rs12459810;rs2965169;rs8100239;rs10401176;rs8103315;rs111677968;rs62117206;rs2006134;rs145414981;rs139326841;rs35106910;rs112450640;rs139425456;rs2889414;rs10419669;rs148933445;rs2967668;rs139246905;rs28399596;rs140824606;rs73048258;rs28399649;rs28399607;rs2968180;rs117737673;rs1135062;rs28399635;rs28399663;rs28399637;rs7026;rs28399664;rs78986976;rs7343130;rs4803760;rs2972555;rs56394238;rs10407439;rs1466435;rs55840414;rs28615360;rs111371860;rs77241309;rs41289512;rs1871046;rs12974942;rs57537848;rs2972566;rs2972558;rs10421035;rs7249933;rs10410835;rs56317818;rs12463239;rs440277;rs365653;rs12978931;rs394353;rs112283362;rs3112439;rs112422902;rs41290100;rs41290102;rs8104483;rs387465;rs11879589;rs71352236;rs393584;rs2075642;rs41290108;rs73050293;rs11668861;rs6859;rs41290120;rs11673139;rs3852861;rs148303016;rs11083749;rs406315;rs113743631;rs12972156;rs283810;rs283814;rs76692773;rs157580;rs73936968;rs77301115;rs1160983;rs11668327;rs35568738;rs117264457;rs405697;rs117843462;rs7259620;rs449647;rs769446;rs429358;rs7412;rs75627662;rs72654472;rs72654437;rs59325138;rs72654445;rs12691088;rs3925681;rs12721046;rs1064725;rs144311893;rs157595;rs75654248;rs141622900;rs4803771;rs71352239;rs60049679;rs11878790;rs113345881;rs8106813;rs73045691;rs76214972;rs12721109;rs1132899;rs117316645;rs143674704;rs75178253;rs204906;rs138692741;rs117198034;rs10775543;rs204478;rs193143021;rs741767;rs117261169;rs2376866;rs35194062;rs28809016;rs35194383;rs116518981;rs35912475;rs4803791;rs62117224;rs1971786;rs34583389;rs1076986;rs34293391;rs139126319;rs10856;rs1968449;rs112481437;rs147188206;rs7259034;rs112394906;rs60239918;rs8100183;rs6509181;rs149013045;rs138625214;rs12461065;rs79067230;rs11083760;rs10413253;rs79322367;rs17643262;rs142883151;rs75463276;rs116863911;rs78073763;rs80182863;rs10421247;rs10416371;rs149553138;rs76971643;rs150685845;rs4580299;rs76133116;rs116958755;rs75858218;rs12460985;rs62118471;rs12461027;rs111243475;rs8113514;rs12977719;rs117783785;rs631920;rs2627642;rs620807;rs8105802;rs4802250;rs182645183;rs10417555;rs10405928;rs11667509;rs10415392;rs12461144;rs10407278;rs346773;rs10402739;rs62118504;rs75727214;rs346748;rs11673093;rs77740243;rs73036542;rs345421;rs344793;rs144645090;rs12983944;rs344818;rs12973593;rs3916898;rs8109829;rs34631405 | rs8104483;rs1160983;rs429358;rs3916898;rs34631405 | rs75161053;rs74172479;rs846892;rs11882134;rs112027066;rs2289494;rs62119328;rs2965156;rs2965162;rs12150984;rs74607435;rs73048230;rs192016015;rs8103315;rs111677968;rs2006134;rs35106910;rs139425456;rs2889414;rs2967668;rs28399596;rs140824606;rs28399607;rs2968180;rs117737673;rs28399635;rs28399663;rs28399637;rs7026;rs78986976;rs7343130;rs4803760;rs2972555;rs10407439;rs1871046;rs57537848;rs2972566;rs2972558;rs12463239;rs112283362;rs112422902;rs8104483;rs387465;rs11879589;rs73050293;rs41290120;rs3852861;rs11083749;rs12972156;rs283810;rs283814;rs76692773;rs157580;rs73936968;rs1160983;rs11668327;rs35568738;rs405697;rs117843462;rs7259620;rs449647;rs429358;rs7412;rs75627662;rs72654437;rs59325138;rs72654445;rs12691088;rs3925681;rs12721046;rs1064725;rs144311893;rs157595;rs75654248;rs141622900;rs4803771;rs71352239;rs60049679;rs11878790;rs113345881;rs73045691;rs12721109;rs1132899;rs143674704;rs204906;rs10775543;rs204478;rs35194062;rs28809016;rs35194383;rs1971786;rs34583389;rs1076986;rs34293391;rs1968449;rs7259034;rs149013045;rs79067230;rs10413253;rs75463276;rs80182863;rs10421247;rs149553138;rs76971643;rs150685845;rs8113514;rs12977719;rs117783785;rs631920;rs4802250;rs10417555;rs12461144;rs10407278;rs73036542;rs144645090;rs12983944;rs3916898 | rs429358:1;rs8104483:0.9305;rs1160983:0.5042;rs34631405:0.1332;rs3916898:0.119 |
| 16 | Chr19:46. | rs3916898;rs8109829;rs34631405;rs117089239;rs10415949;rs2276471;rs59702211;rs73048409;rs62109650;rs11673516;rs147555396;rs145627316;rs7254282;rs4803836;rs35650950;rs4803840;rs12974310;rs62111724;rs35248551;rs34089191;rs2238689;rs1800437;rs1964272;rs34399924;rs11673709;rs7 | rs3916898;rs34631405;rs2276471;rs4803840;rs78784145;rs1055220;rs11545326;rs10415615;rs12609577 | rs8109829;rs117089239;rs59702211;rs73048409;rs147555396;rs35650950;rs35248551;rs17878252;rs75987583;rs112803677 | rs1055220:0.9994;rs10415615:0.8208;rs11545326:0.6534;rs4803840:0.3613;rs2276471 |

| locus_num | locus | input_snps | informative_snps | pleiotropy_snps | top_snps_pip |
|---|---|---|---|---|---|
| | 3Mb | 8784145;rs11083780;rs8100587;rs1055220;rs4803850;rs17878252;rs75987583;rs11083781;rs2070737;rs11545326;rs10520390;rs117379371;rs16980051;rs61703905;rs112803677;rs10415615;rs12609577 | | | :0.3069 |
| 2 | Chr6:47.7Mb | rs7756926;rs12198711;rs9381546;rs4368808;rs6458567;rs6934355;rs1931838;rs13207334;rs9381558;rs7745848;rs10948358;rs12197146;rs72866651;rs13200221;rs1385742;rs9395288;rs143625647;rs7740538;rs7763746;rs9381607;rs116523017 | rs7763746 | rs6934355;rs9381558;rs7745848;rs72866651;rs1385742;rs116523017 | rs7763746:0.9054;rs7740538:0.0313;rs9381607:0.0284;rs1385742:0.0044;rs7745848:0.0033 |
| 3 | Chr8:96Mb | rs16916899;rs17716607;rs7826830;rs3764239;rs17717431;rs7462000;rs67763258;rs12546825;rs6986659;rs7817845;rs4735327;rs35106232;rs113930555;rs62523075;rs2467673;rs4735334;rs896855;rs579798;rs574183;rs72679210;rs13276936;rs13281402 | rs17717431;rs4735334 | rs72679210;rs13276936;rs13281402 | rs4735334:0.9672;rs17717431:0.8526;rs67763258:0.0807;rs7826830:0.0436;rs6986659:0.0136 |
| 4 | Chr10:82.3Mb | rs12244122;rs143290766;rs10887356;rs3762097;rs2819874;rs2819872;rs7071579;rs3851053;rs11201989;rs3122586;rs2789642;rs11596730;rs7076554;rs2819939;rs139414179;rs2788298;rs2437360;rs3827874;rs2259619;rs11592611;rs35213726;rs998766;rs7904369;rs1985908;rs4934028;rs60204118;rs2342812;rs117407472;rs143396142;rs10736339;rs72815310;rs7099331;rs61859191;rs11202666;rs10736343;rs145663320;rs6585994;rs7907770;rs3195900;rs7087356;rs4934169;rs17680430;rs4933397;rs7080169;rs10788639;rs10749604;rs4933402;rs4934284;rs873257;rs7096909;rs17681121;rs9665686;rs7097741;rs71483311;rs6586106;rs71501966;rs2138286 | rs35213726;rs143396142;rs10736343;rs3195900;rs7087356;rs7096909;rs17681121 | rs12244122;rs143290766;rs2788298;rs2437360;rs11592611;rs7096909;rs9665686;rs71501966 | rs143396142:0.9979;rs7087356:0.4985;rs3195900:0.4381;rs7096909:0.1771;rs17681121:0.1657 |
| 5 | Chr11:47.4Mb | rs35959877;rs4752817;rs71474198;rs4494268;rs71474199;rs7101470;rs34942378;rs4579897;rs139240747;rs7117404;rs1055447;rs11039130;rs1685404;rs7947269;rs3781620;rs35677603;rs10838681;rs11039149;rs142289341;rs41299181;rs3816725;rs147206042;rs11570115;rs2596407;rs10769254;rs3729986;rs67472071;rs55957907;rs11039216;rs10838702;rs78529890;rs2293576;rs71475912;rs3824867;rs74385561;rs12361415;rs11603587;rs67132220;rs10838724;rs12287076;rs79754936;rs150268487;rs12286721;rs10769288;rs1017730;rs112318300;rs11039359;rs55743056;rs11819979;rs67019430;rs117399779;rs61895765;rs7942031 | rs71474198;rs139240747;rs3781620;rs10838702;rs74385561;rs150268487;rs67019430 | rs35959877;rs71474198;rs7101470;rs78529890;rs10838724;rs7942031 | rs10838702:0.9999;rs71474198:0.2473;rs3781620:0.2016;rs67019430:0.1726;rs139240747:0.1694;rs74385561:0.1506;rs150268487:0.1121 |
| 6 | Chr11:85.7Mb | rs145403697;rs77020006;rs623120;rs10898403;rs11234419;rs1445499;rs518691;rs145113481;rs11234462;rs7949837;rs56395100;rs11825598;rs598561;rs579406;rs11234493;rs12795381;rs12273113;rs534128;rs12294130;rs17745409;rs521711;rs149332738;rs7938634;rs56369637;rs10751134;rs10160337;rs3851179;rs7113976;rs7129687;rs61904266;rs55808779;rs3901732;rs11602811;rs10501612;rs12799380;rs12293081;rs2291813;rs10898454;rs7924472;rs291243 | rs145403697;rs77020006;rs623120;rs11234419;rs518691;rs11234462;rs7949837;rs56395100;rs11234493;rs12294130;rs10160337;rs10501612;rs12293081;rs2291813;rs7924472 | rs145113481;rs56369637;rs3851179 | rs145403697:0.7976;rs12293081:0.5537;rs11234419:0.2992;rs10501612:0.2766;rs56395100:0.2751 |
| 7 | Chr15:82.4Mb | rs117059514;rs6495618;rs8032120;rs79592067;rs12910666;rs62037263;rs28723034;rs113553821;rs146586218;rs877888;rs80260384;rs2665103;rs148113593;rs1501371;rs16973451;rs111491984;rs72749575;rs112500920;rs118142365;rs56042848;rs2047679;rs75810422;rs8028284;rs16973456;rs11854289;rs11630197 | rs117059514;rs62037263;rs80260384;rs148113593;rs16973451;rs111491984;rs112500920;rs56042848;rs8028284;rs16973456;rs11630197 | rs6495618;rs12910666;rs877888;rs1501371;rs75810422 | rs112500920:0.9696;rs148113593:0.7036;rs16973456:0.4604;rs62037263:0.346;rs111491984:0.2861 |
| 8 | Chr16:20.5Mb | rs9927753;rs35524209;rs77711368;rs80251477;rs12925140;rs34613945;rs148209904;rs145309364;rs9922273;rs139877448;rs78951139;rs111290919;rs118149895;rs113945953;rs78869901;rs35525813;rs142517943;rs76471758;rs34561880;rs35328637;rs4780882;rs12444268;rs8060932;rs111699931;rs72776659;rs6497474;rs111624876;rs9923532;rs55772253;rs7203451;rs7193058;rs74936567;rs36060036;rs7189301;rs72776665;rs118190679;rs4500734;rs75044573;rs76621572;rs7186720;rs77924615;rs146911540;rs12598706;rs11864909;rs12920708;rs12446492;rs7195740;rs7185389;rs11649245;rs117795282;rs8060250;rs62034982;rs13337272;rs9939588;rs28562514;rs12920537;rs77322809;rs142481473;rs11643963;rs8059661;rs150994345;rs117080231;rs12931877;rs76238628;rs34927101;rs649748 | rs74936567;rs36060036;rs4500734;rs77924615;rs12598706;rs12931877 | rs148209904;rs9939588;rs2158031;rs117716220;rs28868590 | rs36060036:1;rs77924615:1;rs12598706:1;rs12931877:0.9935;rs74936567:0.8062 |

| locus_num | locus | input_snps | informative_snps | pleiotropy_snps | top_snps_pip |
|---|---|---|---|---|---|
| | | 8;rs111945505;rs12448840;rs1354576;rs9302387;rs117990195;rs2221448;rs140836105;rs1634306;rs2188760;rs72778610;rs2158031;rs77291768;rs12324972;rs35984880;rs7187002;rs140845299;rs150139789;rs115812285;rs75692301;rs234993;rs12929314;rs16970500;rs433598;rs163257;rs11641336;rs113298888;rs12922077;rs141342711;rs117716220;rs28407583;rs11647875;rs10852248;rs117745671;rs5713;rs140059522;rs72778668;rs146046980;rs113213900;rs11642550;rs2285835;rs111494588;rs9921200;rs4783512;rs1267463;rs111171;rs411045;rs28868590;rs1389146;rs56962160 | | | |
| 9 | Chr17:5.1Mb | rs74476245;rs75364523;rs9893881;rs8066138;rs140850742;rs2302316;rs8834;rs3514;rs55735639;rs112976693;rs191971629;rs2243086;rs238231;rs8071428;rs3744704;rs8067699;rs8071989;rs62076287;rs61740308;rs112061257;rs113260531;rs4790763;rs76419842;rs11650141;rs2309383;rs55910656;rs151195310 | rs9893881;rs140850742;rs2302316;rs3514;rs55735639;rs238231;rs8071989;rs61740308;rs11650141 | rs55735639;rs151195310 | rs8071989:0.795;rs2302316:0.6756;rs140850742:0.3075;rs55735639:0.2882;rs238231:0.235 |

**Supplementary Table S31. Instrumental Variables for Reverse Cis-MR Analysis (AD to eGFR).**
This table mirrors Table S30 but for the reverse cis-MR analysis, where AD is the exposure and eGFR is the outcome. The table provides complete transparency about instrumental variable selection for assessing whether genetic liability for AD causally influences kidney function. For each locus, comprehensive information includes: locus identifiers, all input SNPs, informative SNPs identified through fine-mapping as the primary instruments for AD, SNPs excluded due to pleiotropy (independent effects on eGFR), and top SNPs with posterior inclusion probabilities. Notably, the sets of informative and pleiotropic SNPs often differ between forward and reverse analyses, reflecting the distinct genetic architectures of AD and eGFR within each locus.

| locus_num | analysis_type | locus | input_snps | informative_snps | pleiotropy_snps | top_snps_pip |
|---|---|---|---|---|---|---|
| 1 | reverse | Chr2:55.2Mb | rs17045898;rs10201566;rs59531970;rs114437785;rs4426561;rs4024576;rs4671934;rs139103761;rs115486808;rs4362595;rs35028480;rs76279463;rs143435618;rs72804692;rs6746016;rs189992573;rs56381959;rs6545423;rs150747574;rs75475627;rs10205182;rs10205410;rs62134681;rs77456006;rs79254608;rs6741053;rs12624153;rs61732872;rs2941580;rs6760298;rs55827937;rs2271322;rs28710338;rs2971884;rs2941589;rs354229;rs181347;rs12713275;rs354211;rs35006042;rs354190;rs354243;rs72913519;rs78694233;rs143216688;rs58175244;rs1510613;rs1510614;rs2631835;rs35880160;rs6719425;rs2631832;rs165048;rs165050;rs2063391;rs164943;rs62136882;rs165052;rs165053;rs17046344;rs13416919;rs2578714;rs34170608;rs2567976;rs2631839;rs12467761;rs2631842;rs2244568;rs10205156;rs1438814;rs12713281;rs72482058;rs17046442;rs2163618;rs6545453;rs13408207;rs13430119;rs6744548;rs75010292;rs4625954;rs6725187;rs6745543;rs3821111;rs17347222;rs78539852;rs2920843;rs35950075;rs72799507;rs1134722;rs2589073;rs6721972 | rs75010292;rs17347222;rs78539852 | rs4426561;rs4024576;rs150747574;rs62134681;rs6741053;rs2271322;rs17046442;rs2163618;rs6745543;rs2920843;rs35950075;rs2589073 | rs75010292:0.7908;rs17347222:0.6543;rs78539852:0.2171;rs6744548:0.093;rs6725187:0.0397 |
| 10 | reverse | Chr19:45.3Mb_10 | rs76602578;rs1836280;rs2722683;rs80257887;rs75161053;rs62116959;rs139140823;rs62118204;rs1727760;rs12459712;rs1727762;rs74172479;rs846881;rs846876;rs16979292;rs846892;rs11882134;rs112027066;rs11671606;rs62119270;rs58967105;rs7260482;rs714948;rs111780744;rs2289494;rs62119328;rs2289495;rs150735888;rs2965156;rs11879980;rs111358663;rs151330717;rs62120566;rs2965162;rs73035597;rs2965163;rs12150984;rs2965164;rs2919848;rs143934888;rs62120575;rs111278137;rs79745266;rs4516335;rs11881756;rs79638902;rs73037426;rs4803743;rs2927487;rs150820726;rs7254776;rs10422350;rs186110295;rs8108110;rs80307900;rs74607435;rs77775146;rs73048230;rs2927437;rs192016015;rs12459810;rs2965169;rs8100239;rs10401176;rs8103315;rs111677968;rs62117206;rs2006134;rs111897780;rs145414981;rs139326841;rs35106910;rs112450640;rs139425456;rs2889414;rs10419669;rs148933445;rs2967668;rs139246905;r | rs429358;rs59325138;rs12691088;rs157595;rs141622900 | rs1836280;rs394353;rs8104483;rs76971643 | rs429358:1;rs59325138:1;rs12691088:1;rs157595:1;rs141622900:1 |

| locus_num | analysis_type | locus | input_snps | informative_snps | pleiotropy_snps | top_snps_pip |
|---|---|---|---|---|---|---|
| | | | s28399596;rs140824606;rs73048258;rs28399649;rs28399607;rs2968180;rs117737673;rs1135062;rs28399635;rs28399663;rs28399637;rs7026;rs28399664;rs78986976;rs7343130;rs4802238;rs4803760;rs2972555;rs56394238;rs10407439;rs1466435;rs55840414;rs28615360;rs111371860;rs77241309;rs41289512;rs1871046;rs12974942;rs57537848;rs2972566;rs2972558;rs10421035;rs7249933;rs10410835;rs56317818;rs12463239;rs8112526;rs440277;rs365653;rs12978931;rs394353;rs112283362;rs3112439;rs112422902;rs41290100;rs41290102;rs8104483;rs387465;rs11879589;rs71352236;rs393584;rs2075642;rs41290108;rs73050293;rs11668861;rs6859;rs41290120;rs11673139;rs3852861;rs148303016;rs11083749;rs406315;rs113743631;rs3745150;rs12972156;rs283810;rs283814;rs76692773;rs157580;rs73936968;rs77301115;rs1160983;rs11668327;rs35568738;rs117264457;rs405697;rs117843462;rs7259620;rs449647;rs769446;rs440446;rs429358;rs7412;rs75627662;rs72654472;rs72654437;rs59325138;rs72654445;rs12691088;rs3925681;rs12721046;rs1064725;rs144311893;rs157595;rs75654248;rs141622900;rs4803771;rs4803772;rs71352239;rs60049679;rs11878790;rs113345881;rs8106813;rs73045691;rs34810028;rs12977604;rs111809714;rs76214972;rs12721109;rs1132899;rs5167;rs117316645;rs143674704;rs75178253;rs204906;rs138692741;rs117198034;rs10775543;rs204482;rs146390218;rs204478;rs193143021;rs741767;rs117261169;rs35080293;rs2376866;rs35194062;rs28809016;rs35194383;rs116518981;rs35912475;rs4803791;rs62117224;rs1971786;rs34583389;rs1076986;rs34293391;rs139126319;rs10856;rs34827707;rs1968449;rs112481437;rs147188206;rs7259034;rs112394906;rs11083759;rs60239918;rs8100183;rs10426398;rs9653111;rs149013045;rs138625214;rs12461065;rs79067230;rs11083760;rs10413253;rs79322367;rs17643262;rs142883151;rs75463276;rs116863911;rs78073763;rs80182863;rs10421247;rs10416371;rs149553138;rs76971643;rs150685845;rs78979751;rs4580299;rs76133116;rs59839536;rs116958755;rs12460985;rs62118471;rs75348771;rs111243475;rs8113514;rs12977719;rs117783785;rs631920;rs2627642;rs620807;rs8105802;rs4802250;rs182645183;rs10417555;rs10405928;rs11667509;rs10415392;rs12461144;rs10407278;rs28607628;rs346773;rs10402739;rs62118504;rs75727214;rs346748;rs11673093;rs77740243;rs73036542;rs1872259 | | | |
| 11 | reverse | Chr19:45.3Mb_11 | rs1836280;rs2722683;rs80257887;rs75161053;rs62116959;rs139140823;rs62118204;rs1727760;rs12459712;rs1727762;rs74172479;rs846881;rs846876;rs16979292;rs846892;rs11882134;rs112027066;rs11671606;rs62119270;rs58967105;rs7260482;rs714948;rs111780744;rs2289494;rs62119328;rs2289495;rs150735888;rs2965156;rs11879980;rs111358663;rs151330717;rs62120566;rs2965162;rs73035597;rs2965163;rs12150984;rs2965164;rs2919848;rs143934888;rs62120575;rs111278137;rs79745266;rs4516335;rs11881756;rs79638902;rs73037426;rs4803743;rs2927487;rs150820726;rs7254776;rs10422350;rs186110295;rs8108110;rs80307900;rs74607435;rs77775146;rs73048230;rs2927437;rs192016015;rs12459810;rs2965169;rs8100239;rs10401176;rs8103315;rs111677968;rs62117206;rs2006134;rs111897780;rs145414981;rs139326841;rs35106910;rs112450640;rs139425456;rs2889414;rs10419669;rs148933445;rs2967668;rs139246905;rs28399596;rs140824606;rs73048258;rs28399649;rs28399607;rs2968180;rs117737673;rs1135062;rs28399635;rs28399663;rs28399637;rs7026;rs28399664;rs78986976;rs7343130;rs4802238;rs4803760;rs2972555;rs56394238;rs10407439;rs1466435;rs55840414;rs28615360;rs111371860;rs77241309;rs41289512;rs1871046;rs12974942;rs57537848;rs2972566;rs2972558;rs10421035;rs7249933;rs10410835;rs56317818;rs12463239;rs8112526;rs440277;rs365653;rs12978931;rs394353;rs112283362;rs3112439;rs112422902;rs41290100;rs41290102;rs8104483;rs387465;rs11879589;rs71352236;rs393584;rs2075642;rs41290108;rs73050293;rs11668861;rs6859;rs41290120;rs11673139;rs3852861;rs148303016;rs11083749;rs406315;rs113743631;rs3745150;rs12972156;rs283810;rs283814;rs76692773;rs157580;rs73936968;rs77301115;rs1160983;rs11668327;rs35568738;rs117264457;rs405697;rs117843462;rs7259620;rs449647;rs769446;rs440446;rs429358;rs7412;rs75627662;rs72654472;rs72654437;rs59325138;rs72654445;rs12691088;rs3925681;rs12721046;rs1064725;rs144311893;rs157595;rs75654248;rs141622900;rs4803771;rs4803772;rs71352239;rs60049679;rs11878790;rs113345881;rs8106813;rs73045691;rs34810028;rs12977604;rs111809714;rs76214972;rs12721109;rs1132899;rs5167;rs117316645;rs143674704;rs75178253;rs204906;rs138692741;rs117198034;rs10775543;rs204482;rs146390218;rs204478;rs193143021;rs741767;rs117261169;rs35080293;rs2376866;rs35194062;rs28809016;rs35194383;rs116518981;rs35912475;rs4803791;rs62117224;rs1971786;rs34583389;rs1076986;rs34293391;rs139126319;rs10856;rs34827707;rs1968449;rs112481437;rs147188206;rs7259034;rs112394906;rs11083759;rs60239918;rs8100183;rs10426398;rs9653111;rs149013045;rs138625214;rs12461065;rs79067230;rs11083760;rs10413253;rs79322367;rs17643262;rs142883151;rs75463276;rs116863911;rs78073763;rs80182 | rs429358;rs59325138;rs12691088;rs157595;rs141622900 | rs1836280;rs394353;rs8104483;rs76971643 | rs429358:1;rs59325138:1;rs12691088:1;rs157595:1;rs141622900:1 |

| locus_num | analysis_type | locus | input_snps | informative_snps | pleiotropy_snps | top_snps_pip |
|---|---|---|---|---|---|---|
| | | | 863;rs10421247;rs10416371;rs149553138;rs76971643;rs150685845;rs78979751;rs4580299;rs76133116;rs59839536;rs116958755;rs12460985;rs62118471;rs75348771;rs111243475;rs8113514;rs12977719;rs117783785;rs631920;rs2627642;rs620807;rs8105802;rs4802250;rs182645183;rs10417555;rs10405928;rs11667509;rs10415392;rs12461144;rs10407278;rs28607628;rs346773;rs10402739;rs62118504;rs75727214;rs346748;rs11673093;rs77740243;rs73036542;rs345421 | | | |
| 12 | reverse | Chr19:45.4Mb_12 | rs1836280;rs2722683;rs80257887;rs75161053;rs62116959;rs139140823;rs62118204;rs1727760;rs12459712;rs1727762;rs74172479;rs846881;rs846876;rs16979292;rs846892;rs11882134;rs112027066;rs11671606;rs62119270;rs58967105;rs7260482;rs714948;rs111780744;rs2289494;rs62119328;rs2289495;rs150735888;rs2965156;rs11879980;rs111358663;rs151330717;rs62120566;rs2965162;rs73035597;rs2965163;rs12150984;rs2965164;rs2919848;rs143934888;rs62120575;rs111278137;rs79745266;rs4516335;rs11881756;rs79638902;rs73037426;rs4803743;rs2927487;rs150820726;rs7254776;rs10422350;rs186110295;rs8108110;rs80307900;rs74607435;rs77775146;rs73048230;rs2927437;rs192016015;rs12459810;rs2965169;rs8100239;rs10401176;rs8103315;rs111677968;rs62117206;rs2006134;rs111897780;rs145414981;rs139326841;rs35106910;rs112450640;rs139425456;rs2889414;rs10419669;rs148933445;rs2967668;rs139246905;rs28399596;rs140824606;rs73048258;rs28399649;rs28399607;rs2968180;rs117737673;rs1135062;rs28399635;rs28399663;rs28399637;rs7026;rs28399664;rs78986976;rs7343130;rs4802238;rs4803760;rs2972555;rs56394238;rs10407439;rs1466435;rs55840414;rs28615360;rs111371860;rs77241309;rs41289512;rs1871046;rs12974942;rs57537848;rs2972566;rs2972558;rs10421035;rs7249933;rs10410835;rs56317818;rs12463239;rs8112526;rs440277;rs365653;rs12978931;rs394353;rs112283362;rs3112439;rs112422902;rs41290100;rs41290102;rs8104483;rs387465;rs11879589;rs71352236;rs393584;rs2075642;rs41290108;rs73050293;rs11668861;rs6859;rs41290120;rs11673139;rs3852861;rs148303016;rs11083749;rs406315;rs113743631;rs3745150;rs12972156;rs283810;rs283814;rs76692773;rs157580;rs73936968;rs77301115;rs1160983;rs11668327;rs35568738;rs117264457;rs405697;rs117843462;rs7259620;rs449647;rs769446;rs440446;rs429358;rs7412;rs75627662;rs72654472;rs72654437;rs59325138;rs72654445;rs12691088;rs3925681;rs12721046;rs1064725;rs144311893;rs157595;rs75654248;rs141622900;rs4803771;rs4803772;rs71352239;rs60049679;rs11878790;rs113345881;rs8106813;rs73045691;rs34810028;rs12977604;rs111809714;rs76214972;rs12721109;rs1132899;rs5167;rs117316645;rs143674704;rs75178253;rs204906;rs138692741;rs117198034;rs10775543;rs204482;rs146390218;rs204478;rs193143021;rs741767;rs117261169;rs35080293;rs2376866;rs35194062;rs28809016;rs35194383;rs116518981;rs35912475;rs4803791;rs62117224;rs1971786;rs34583389;rs1076986;rs34293391;rs139126319;rs10856;rs34827707;rs1968449;rs112481437;rs147188206;rs7259034;rs112394906;rs11083759;rs60239918;rs8100183;rs10426398;rs9653111;rs149013045;rs138625214;rs12461065;rs79067230;rs11083760;rs10413253;rs79322367;rs17643262;rs142883151;rs75463276;rs116863911;rs78073763;rs80182863;rs10421247;rs10416371;rs149553138;rs76971643;rs150685845;rs78979751;rs4580299;rs76133116;rs59839536;rs116958755;rs12460985;rs62118471;rs75348771;rs111243475;rs8113514;rs12977719;rs117783785;rs631920;rs2627642;rs620807;rs8105802;rs4802250;rs182645183;rs10417555;rs10405928;rs11667509;rs10415392;rs12461144;rs10407278;rs28607628;rs346773;rs10402739;rs62118504;rs75727214;rs346748;rs11673093;rs77740243;rs73036542;rs345421;rs344793;rs144645090;rs12983944;rs344818;rs12973593 | rs429358;rs59325138;rs12691088;rs157595;rs141622900 | rs1836280;rs394353;rs8104483;rs76971643 | rs429358:1;rs59325138:1;rs12691088:1;rs157595:1;rs141622900:1 |
| 13a | reverse | Chr19:45.4Mb_13a | rs2722683;rs80257887;rs75161053;rs62116959;rs139140823;rs62118204;rs1727760;rs12459712;rs1727762;rs74172479;rs846881;rs846876;rs16979292;rs846892;rs11882134;rs112027066;rs11671606;rs62119270;rs58967105;rs7260482;rs714948;rs111780744;rs2289494;rs62119328;rs2289495;rs150735888;rs2965156;rs11879980;rs111358663;rs151330717;rs62120566;rs2965162;rs73035597;rs2965163;rs12150984;rs2965164;rs2919848;rs143934888;rs62120575;rs111278137;rs79745266;rs4516335;rs11881756;rs79638902;rs73037426;rs4803743;rs2927487;rs150820726;rs7254776;rs10422350;rs186110295;rs8108110;rs80307900;rs74607435;rs77775146;rs73048230;rs2927437;rs192016015;rs12459810;rs2965169;rs8100239;rs10401176;rs8103315;rs111677968;rs62117206;rs2006134;rs111897780;rs145414981;rs139326841;rs35106910;rs112450640;rs139425456;rs2889414;rs10419669;rs148933445;rs2967668;rs139246905;rs28399596;rs140824606;rs73048258;rs28399649;rs28399607;rs2968180;rs117737673;rs1135062;rs28399635;rs28399663;rs28399637;rs7026;rs28399664;rs78986976;rs7343130;rs4802238;rs4803760;rs2972555;rs56394238;rs10407439;rs1466435;rs55840414;rs28615360;rs111371860;rs77241309;rs41289512;rs1871046;rs12974942;rs57537848;rs2972566;rs2972558;rs10421035;rs7249933;rs10410835;rs56317818;rs12463239;rs8112526;rs440277;rs365653;rs12978931;rs394353;rs112283362;rs311243 | rs429358;rs59325138;rs12691088;rs157595;rs141622900 | rs394353;rs8104483;rs76971643;rs3916898;rs34631405 | rs429358:1;rs59325138:1;rs12691088:1;rs157595:1;rs141622900:1 |

| locus_num | analysis_type | locus | input_snps | informative_snps | pleiotropy_snps | top_snps_pip |
|---|---|---|---|---|---|---|
| | | | 9;rs112422902;rs41290100;rs41290102;rs8104483;rs387465;rs11879589;rs71352236;rs393584;rs2075642;rs41290108;rs73050293;rs11668861;rs6859;rs41290120;rs11673139;rs3852861;rs148303016;rs11083749;rs406315;rs113743631;rs3745150;rs12972156;rs283810;rs283814;rs76692773;rs157580;rs73936968;rs77301115;rs1160983;rs11668327;rs35568738;rs117264457;rs405697;rs117843462;rs7259620;rs449647;rs769446;rs440446;rs429358;rs7412;rs75627662;rs72654472;rs72654437;rs59325138;rs72654445;rs12691088;rs3925681;rs12721046;rs1064725;rs144311893;rs157595;rs75654248;rs141622900;rs4803771;rs4803772;rs71352239;rs60049679;rs11878790;rs113345881;rs8106813;rs73045691;rs34810028;rs12977604;rs111809714;rs76214972;rs12721109;rs1132899;rs5167;rs117316645;rs143674704;rs75178253;rs204906;rs138692741;rs117198034;rs10775543;rs204482;rs146390218;rs204478;rs193143021;rs741767;rs117261169;rs35080293;rs2376866;rs35194062;rs28809016;rs35194383;rs116518981;rs35912475;rs4803791;rs62117224;rs1971786;rs34583389;rs1076986;rs34293391;rs139126319;rs10856;rs34827707;rs1968449;rs112481437;rs147188206;rs7259034;rs112394906;rs11083759;rs60239918;rs8100183;rs10426398;rs9653111;rs149013045;rs138625214;rs12461065;rs79067230;rs11083760;rs10413253;rs79322367;rs17643262;rs142883151;rs75463276;rs116863911;rs78073763;rs80182863;rs10421247;rs10416371;rs149553138;rs76971643;rs150685845;rs78979751;rs4580299;rs76133116;rs59839536;rs116958755;rs12460985;rs62118471;rs75348771;rs111243475;rs8113514;rs12977719;rs117783785;rs631920;rs2627642;rs620807;rs8105802;rs4802250;rs182645183;rs10417555;rs10405928;rs11667509;rs10415392;rs12461144;rs10407278;rs28607628;rs346773;rs10402739;rs62118504;rs75727214;rs346748;rs11673093;rs77740243;rs73036542;rs345421;rs344793;rs144645090;rs12983944;rs344818;rs12973593;rs3916898;rs1799786;rs8109829;rs34631405 | | | |
| 13b | reverse | Chr19:45.4Mb_13b | rs2722683;rs80257887;rs75161053;rs62116959;rs139140823;rs62118204;rs1727760;rs12459712;rs1727762;rs74172479;rs846881;rs846876;rs16979292;rs846892;rs11882134;rs112027066;rs11671606;rs62119270;rs58967105;rs7260482;rs714948;rs111780744;rs2289494;rs62119328;rs2289495;rs150735888;rs2965156;rs11879980;rs111358663;rs151330717;rs62120566;rs2965162;rs73035597;rs2965163;rs12150984;rs2965164;rs2919848;rs143934888;rs62120575;rs111278137;rs79745266;rs4516335;rs11881756;rs79638902;rs73037426;rs4803743;rs2927487;rs150820726;rs7254776;rs10422350;rs186110295;rs8108110;rs80307900;rs74607435;rs77775146;rs73048230;rs2927437;rs192016015;rs12459810;rs2965169;rs8100239;rs10401176;rs8103315;rs111677968;rs62117206;rs2006134;rs111897780;rs145414981;rs139326841;rs35106910;rs112450640;rs139425456;rs2889414;rs10419669;rs148933445;rs2967668;rs139246905;rs28399596;rs140824606;rs73048258;rs28399649;rs28399607;rs2968180;rs117737673;rs1135062;rs28399635;rs28399663;rs28399637;rs7026;rs28399664;rs78986976;rs7343130;rs4802238;rs4803760;rs2972555;rs56394238;rs10407439;rs1466435;rs55840414;rs28615360;rs111371860;rs77241309;rs41289512;rs1871046;rs12974942;rs57537848;rs2972566;rs2972558;rs10421035;rs7249933;rs10410835;rs56317818;rs12463239;rs8112526;rs440277;rs365653;rs12978931;rs394353;rs112283362;rs3112439;rs112422902;rs41290100;rs41290102;rs8104483;rs387465;rs11879589;rs71352236;rs393584;rs2075642;rs41290108;rs73050293;rs11668861;rs6859;rs41290120;rs11673139;rs3852861;rs148303016;rs11083749;rs406315;rs113743631;rs3745150;rs12972156;rs283810;rs283814;rs76692773;rs157580;rs73936968;rs77301115;rs1160983;rs11668327;rs35568738;rs117264457;rs405697;rs117843462;rs7259620;rs449647;rs769446;rs440446;rs429358;rs7412;rs75627662;rs72654472;rs72654437;rs59325138;rs72654445;rs12691088;rs3925681;rs12721046;rs1064725;rs144311893;rs157595;rs75654248;rs141622900;rs4803771;rs4803772;rs71352239;rs60049679;rs11878790;rs113345881;rs8106813;rs73045691;rs34810028;rs12977604;rs111809714;rs76214972;rs12721109;rs1132899;rs5167;rs117316645;rs143674704;rs75178253;rs204906;rs138692741;rs117198034;rs10775543;rs204482;rs146390218;rs204478;rs193143021;rs741767;rs117261169;rs35080293;rs2376866;rs35194062;rs28809016;rs35194383;rs116518981;rs35912475;rs4803791;rs62117224;rs1971786;rs34583389;rs1076986;rs34293391;rs139126319;rs10856;rs34827707;rs1968449;rs112481437;rs147188206;rs7259034;rs112394906;rs11083759;rs60239918;rs8100183;rs10426398;rs9653111;rs149013045;rs138625214;rs12461065;rs79067230;rs11083760;rs10413253;rs79322367;rs17643262;rs142883151;rs75463276;rs116863911;rs78073763;rs80182863;rs10421247;rs10416371;rs149553138;rs76971643;rs150685845;rs78979751;rs4580299;rs76133116;rs59839536;rs116958755;rs12460985;rs62118471;rs75348771;rs111243475;rs8113514;rs12977719;rs117783785;rs631920;rs2627642;rs620807;rs8105802;rs4802250;rs182645183;rs10417555;rs10405928;rs11667509;rs10415392;rs12461144;rs10407278;rs28607628;rs346773;rs10402739;rs62118504;rs75727214;rs346748;rs11673093;rs77740243;rs73036542;rs345421;rs344793;rs14464 | rs429358;rs59325138;rs12691088;rs157595;rs141622900 | rs394353;rs8104483;rs76971643;rs3916898;rs34631405 | rs429358:1;rs59325138:1;rs12691088:1;rs157595:1;rs141622900:1 |

| locus_num | analysis_type | locus | input_snps | informative_snps | pleiotropy_snps | top_snps_pip |
|---|---|---|---|---|---|---|
| | | | 5090;rs12983944;rs344818;rs12973593;rs3916898;rs1799786;rs8109829;rs34631405 | | | |
| 13c | reverse | Chr 19:45.4 Mb_13c | rs2722683;rs80257887;rs75161053;rs62116959;rs139140823;rs62118204;rs1727760;rs12459712;rs1727762;rs74172479;rs846881;rs846876;rs16979292;rs846892;rs11882134;rs112027066;rs11671606;rs62119270;rs58967105;rs7260482;rs714948;rs111780744;rs2289494;rs62119328;rs2289495;rs150735888;rs2965156;rs11879980;rs111358663;rs151330717;rs62120566;rs2965162;rs73035597;rs2965163;rs12150984;rs2965164;rs2919848;rs143934888;rs62120575;rs111278137;rs79745266;rs4516335;rs11881756;rs79638902;rs73037426;rs4803743;rs2927487;rs150820726;rs7254776;rs10422350;rs186110295;rs8108110;rs80307900;rs74607435;rs77775146;rs73048230;rs2927437;rs192016015;rs12459810;rs2965169;rs8100239;rs10401176;rs8103315;rs111677968;rs62117206;rs2006134;rs111897780;rs145414981;rs139326841;rs35106910;rs112450640;rs139425456;rs2889414;rs10419669;rs148933445;rs2967668;rs139246905;rs28399596;rs140824606;rs73048258;rs28399649;rs28399607;rs2968180;rs117737673;rs1135062;rs28399635;rs28399663;rs28399637;rs7026;rs28399664;rs78986976;rs7343130;rs4802238;rs4803760;rs2972555;rs56394238;rs10407439;rs1466435;rs55840414;rs28615360;rs111371860;rs77241309;rs41289512;rs1871046;rs12974942;rs57537848;rs2972566;rs2972558;rs10421035;rs7249933;rs10410835;rs56317818;rs12463239;rs8112526;rs440277;rs365653;rs12978931;rs394353;rs112283362;rs3112439;rs112422902;rs41290100;rs41290102;rs8104483;rs387465;rs11879589;rs71352236;rs393584;rs2075642;rs41290108;rs73050293;rs11668861;rs6859;rs41290120;rs11673139;rs3852861;rs148303016;rs11083749;rs406315;rs113743631;rs3745150;rs12972156;rs283810;rs283814;rs76692773;rs157580;rs73936968;rs77301115;rs1160983;rs11668327;rs35568738;rs117264457;rs405697;rs117843462;rs7259620;rs449647;rs769446;rs440446;rs429358;rs7412;rs75627662;rs72654472;rs72654437;rs59325138;rs72654445;rs12691088;rs3925681;rs12721046;rs1064725;rs144311893;rs157595;rs75654248;rs141622900;rs4803771;rs4803772;rs71352239;rs60049679;rs11878790;rs113345881;rs8106813;rs73045691;rs34810028;rs12977604;rs111809714;rs76214972;rs12721109;rs1132899;rs5167;rs117316645;rs143674704;rs75178253;rs204906;rs138692741;rs117198034;rs10775543;rs204482;rs146390218;rs204478;rs193143021;rs741767;rs117261169;rs35080293;rs2376866;rs35194062;rs28809016;rs35194383;rs116518981;rs35912475;rs4803791;rs62117224;rs1971786;rs34583389;rs1076986;rs34293391;rs139126319;rs10856;rs34827707;rs1968449;rs112481437;rs147188206;rs7259034;rs112394906;rs11083759;rs60239918;rs8100183;rs10426398;rs9653111;rs149013045;rs138625214;rs12461065;rs79067230;rs11083760;rs10413253;rs79322367;rs17643262;rs142883151;rs75463276;rs116863911;rs78073763;rs80182863;rs10421247;rs10416371;rs149553138;rs76971643;rs150685845;rs78979751;rs4580299;rs76133116;rs59839536;rs116958755;rs12460985;rs62118471;rs75348771;rs111243475;rs8113514;rs12977719;rs117783785;rs631920;rs2627642;rs620807;rs8105802;rs4802250;rs182645183;rs10417555;rs10405928;rs11667509;rs10415392;rs12461144;rs10407278;rs28607628;rs346773;rs10402739;rs62118504;rs75727214;rs346748;rs11673093;rs77740243;rs73036542;rs345421;rs344793;rs144645090;rs12983944;rs344818;rs12973593;rs3916898;rs1799786;rs8109829;rs34631405 | rs429358;rs59325138;rs12691088;rs157595;rs141622900 | rs394353;rs8104483;rs76971643;rs3916898;rs34631405 | rs429358:1;rs59325138:1;rs12691088:1;rs157595:1;rs141622900:1 |
| 16 | reverse | Chr 19:46.3 Mb | rs3916898;rs1799786;rs8109829;rs34631405;rs117089239;rs10415949;rs2276471;rs59702211;rs73048409;rs62109650;rs11673516;rs147555396;rs145627316;rs7254282;rs4803836;rs35650950;rs4803840;rs12974310;rs62111724;rs35248551;rs34089191;rs2238689;rs1800437;rs1964272;rs34399924;rs11673709;rs111713189;rs78784145;rs11083780;rs8100587;rs1055220;rs4803850;rs17878252;rs75987583;rs11083781;rs2070737;rs11545326;rs10520390;rs117379371;rs16980051;rs61703905;rs112803677;rs10415615;rs12609577 | rs117089239;rs59702211;rs73048409;rs147555396;rs17878252;rs112803677 | rs3916898;rs34631405;rs145627316;rs7254282;rs4803840;rs1055220;rs11545326;rs10415615 | rs59702211:0.9893;rs147555396:0.9185;rs112803677:0.8832;rs17878252:0.7161;rs117089239:0.3301 |
| 2 | reverse | Chr 6:47.7Mb | rs7756926;rs12197270;rs6925189;rs9381546;rs4368808;rs6458566;rs6458567;rs6934355;rs1931838;rs13207334;rs9381558;rs7745848;rs12197146;rs10948359;rs72866651;rs9349405;rs13200221;rs12195738;rs1385742;rs143625647;rs9473147;rs7740538;rs7763746;rs2396831;rs9381607;rs116523017 | rs12197270;rs9381546;rs4368808;rs6458567;rs6934355;rs13207334;rs7745848;rs12197146;rs10948359;rs72866651;rs12195738;rs1385742;rs9473147;rs7740538;rs116523017 | rs7763746;rs2396831 | rs116523017:0.8899;rs7745848:0.849;rs1385742:0.2372;rs12195738:0.2303;rs6934355:0.2225 |
| 3 | reverse | Chr 8:96 Mb | rs16916899;rs17716607;rs7826830;rs3764239;rs17717431;rs7462000;rs67763258;rs13262255;rs12546825;rs6986659;rs7817845;rs4735327;rs35106232;rs113930555;rs62523075;rs2467673;rs4735334;rs896855;rs579798;rs574183;rs72679210;rs13276936;rs13281402 | rs16916899;rs17716607;rs7826830;rs7462000;rs67763258;rs13262255;rs12546825;rs6986659;rs4735327;rs35106232;rs113930555;rs62523075;rs2467673;rs896855;rs579798;rs574183;rs72679210;rs13276936;rs13281402 | rs17717431;rs4735334 | rs13276936:0.8769;rs13281402:0.8405;rs72679210:0.2863;rs113930555:0.285;rs574183:0.1918 |
| 4 | rev | Chr | rs12244122;rs143290766;rs10887356;rs2789697;rs2789655;rs2819872;rs7071579;rs3748242;rs1079242;rs3851053;rs11 | rs12244122;rs143290766;rs139414179;rs4934028;rs70 | rs35213726;rs60204118;rs117407472;rs143396142;rs72815 | rs71501966:0.8804;rs493 |

| locus_num | analysis_type | locus | input_snps | informative_snps | pleiotropy_snps | top_snps_pip |
|---|---|---|---|---|---|---|
| | erse | 10:82.3 Mb | 201989;rs3122586;rs12770550;rs2789642;rs7076554;rs7094956;rs2819939;rs139414179;rs56236678;rs2788298;rs2437360;rs3827874;rs2789643;rs2259619;rs11592611;rs35213726;rs998766;rs7904369;rs1985908;rs4934028;rs60204118;rs2342812;rs117407472;rs143396142;rs10736339;rs72815310;rs7099331;rs61859191;rs11202666;rs10736343;rs145663320;rs11202759;rs6585994;rs7907770;rs3195900;rs7087356;rs4934169;rs17680430;rs11203034;rs7080169;rs10788639;rs4933402;rs6586030;rs4934284;rs873257;rs7096909;rs17681121;rs9665686;rs7097741;rs71483311;rs6586106;rs71501966;rs2138286;rs1913752 | 80169;rs6586030;rs7096909;rs9665686;rs71501966 | 310;rs3195900;rs6586030;rs7096909;rs17681121;rs9665686;rs7097741 | 4028:0.3698;rs143290766:0.2524;rs7096909:0.2233;rs6586030:0.1845 |
| 5 | reverse | Chr 11:47.4 Mb | rs898604;rs71474198;rs7101470;rs139240747;rs11039130;rs1685404;rs10838681;rs142289341;rs147206042;rs3729986;rs67472071;rs55957907;rs10838702;rs78529890;rs74385561;rs67132220;rs150268487;rs10769288;rs112318300;rs117399779;rs61895765 | rs71474198;rs7101470;rs11039130;rs1685404;rs147206042;rs67472071;rs78529890;rs67132220;rs61895765 | rs71474198;rs139240747;rs10838681;rs10838702;rs74385561;rs150268487 | rs67472071:0.998;rs7101470:0.8073;rs71474198:0.274;rs11039130:0.2119;rs78529890:0.2102 |
| 6 | reverse | Chr 11:85.7 Mb | rs145403697;rs77020006;rs623120;rs10898403;rs11234419;rs1445499;rs518691;rs145113481;rs11234462;rs7949837;rs56395100;rs11825598;rs598561;rs579406;rs11234493;rs12795381;rs12273113;rs534128;rs787928;rs12294130;rs629343;rs17745409;rs149332738;rs646260;rs7938634;rs56369637;rs10751134;rs6592272;rs10160337;rs3851179;rs7113976;rs7129687;rs61904266;rs55808779;rs3901732;rs11602811;rs10501612;rs12799380;rs12293081;rs2291813;rs10898454;rs7924472;rs291243 | rs145113481;rs149332738;rs56369637;rs3851179;rs12799380 | rs145403697;rs77020006;rs518691;rs12795381;rs10501612;rs12293081;rs2291813;rs10898454;rs7924472 | rs3851179:0.9772;rs145113481:0.3235;rs149332738:0.2104;rs56369637:0.1179;rs12799380:0.1104 |
| 7 | reverse | Chr 15:82.4 Mb | rs117059514;rs6495618;rs8032120;rs79592067;rs12910666;rs62037263;rs28723034;rs113553821;rs146586218;rs877888;rs80260384;rs2665103;rs148113593;rs1501371;rs16973451;rs111491984;rs72749575;rs112500920;rs118142365;rs56042848;rs2047679;rs75810422;rs8028284;rs16973456;rs11854289;rs11630197 | rs6495618;rs8032120;rs12910666;rs877888;rs2665103;rs1501371;rs111491984;rs75810422 | rs62037263;rs148113593;rs111491984;rs112500920;rs16973456 | rs1501371:0.5082;rs2665103:0.4257;rs8032120:0.3434;rs6495618:0.336;rs12910666:0.2944 |
| 8 | reverse | Chr 16:20.5 Mb | rs9927753;rs35524209;rs77711368;rs80251477;rs12925140;rs34613945;rs148209904;rs145309364;rs9922273;rs139877448;rs78951139;rs111290919;rs118149895;rs113945953;rs78869901;rs35525813;rs142517943;rs76471758;rs34561880;rs35328637;rs34230407;rs12444268;rs8060932;rs111699931;rs72776659;rs6497474;rs111624876;rs71384446;rs9923532;rs55772253;rs7203451;rs7193058;rs74936567;rs36060036;rs7189301;rs72776665;rs118190679;rs4500734;rs75044573;rs11865589;rs76621572;rs7186720;rs77924615;rs146911540;rs12598706;rs11864909;rs12920708;rs12446492;rs7195740;rs7185389;rs11649245;rs117795282;rs8060250;rs62034982;rs13337272;rs9939588;rs28562514;rs12920537;rs77322809;rs142481473;rs11643963;rs8059661;rs150994345;rs28421037;rs117080231;rs12931877;rs150216898;rs12924372;rs76238628;rs34927101;rs6497488;rs111945505;rs12448840;rs1354576;rs9302387;rs117990195;rs2221448;rs140836105;rs1634306;rs2188760;rs72778610;rs2158031;rs77291768;rs12324972;rs35984880;rs7187002;rs140845299;rs150139789;rs115812285;rs75692301;rs234993;rs12929314;rs16970500;rs433598;rs111590267;rs163257;rs11641336;rs113298888;rs12922077;rs141342711;rs117716220;rs79211750;rs28407583;rs11647875;rs10852248;rs117745671;rs5713;rs140059522;rs72778668;rs146046980;rs113213900;rs147868418;rs11642550;rs2285835;rs111494588;rs9921200;rs4783512;rs1267463;rs111171;rs428863;rs411045;rs28868590;rs1389146;rs56962160 | rs2158031 | rs35524209;rs80251477;rs145309364;rs113945953;rs6497474;rs74936567;rs36060036;rs7189301;rs7186720;rs77924615;rs9939588;rs28562514;rs12920537;rs77322809;rs12931877;rs2221448;rs150139789;rs10852248;rs28868590;rs56962160 | rs2158031:0.7081;rs12931877:0.0584;rs117716220:0.057;rs28868590:0.0364;rs147868418:0.0211 |
| 9 | reverse | Chr 17:5.1Mb | rs74476245;rs75364523;rs9893881;rs140850742;rs2302316;rs55735639;rs112976693;rs8071428;rs8071989;rs112061257;rs76419842;rs75367124;rs11650141;rs2309383;rs151195310 | rs74476245;rs75364523;rs55735639;rs112976693;rs8071989;rs112061257;rs75367124;rs151195310 | rs74476245;rs9893881;rs140850742;rs2302316;rs8071428;rs8071989;rs112061257;rs11650141;rs2309383;rs151195310 | rs55735639:0.8964;rs151195310:0.6795;rs112061257:0.2408;rs74476245:0.2391;rs75367124:0.2158 |

**Supplementary Table S32. Instrumental Variable Overlap Between Forward and Reverse Cis-MR Analyses.** This summary table quantifies the overlap of instrumental variables between forward and reverse cis-MR analyses at each locus, which is critical for interpreting bidirectional causal relationships and assessing the potential for bias due to weak instruments or horizontal pleiotropy. For each locus, the table reports: the number of input SNPs considered in forward and reverse analyses, the number of overlapping input SNPs, the number of informative instrumental variables identified for each direction, the overlap in informative SNPs between directions, the number of SNPs flagged for pleiotropy in each direction, and the overlap in pleiotropic SNPs. High overlap in informative SNPs suggests that the same genetic variants influence both traits, supporting biological pleiotropy. High overlap in pleiotropic SNPs indicates that horizontal pleiotropy may be present, potentially violating MR assumptions and suggesting that causal estimates should be interpreted with caution. Minimal overlap in instrumental variables between forward and reverse directions provides stronger evidence for valid causal inference in both directions.

| Locus Num | Locus | N Input Forward | N Input Reverse | Overlap Input | N Informative Forward | N Informative Reverse | Overlap Informative | N Pleiotropy Forward | N Pleiotropy Reverse | Overlap Pleiotropy | Overlap Informative SNPs | Overlap Pleiotropy SNPs |
|---|---|---|---|---|---|---|---|---|---|---|---|---|
| 1 | Chr2:55.2Mb | 84 | 91 | 80 | 4 | 3 | 0 | 3 | 12 | 0 | | |
| 10 | Chr19:45.3Mb_10 | 259 | 281 | 256 | 4 | 5 | 1 | 118 | 4 | 3 | rs429358 | rs394353;rs76971643;rs8104483 |
| 11 | Chr19:45.3Mb_11 | 258 | 280 | 255 | 4 | 5 | 1 | 114 | 4 | 2 | rs429358 | rs76971643;rs8104483 |
| 12 | Chr19:45.4Mb_12 | 263 | 285 | 260 | 4 | 5 | 1 | 115 | 4 | 2 | rs429358 | rs76971643;rs8104483 |
| 13a | Chr19:45.4Mb_13a | 265 | 288 | 262 | 5 | 5 | 1 | 117 | 5 | 3 | rs429358 | rs3916898;rs76971643;rs8104483 |
| 13b | Chr19:45.4Mb_13b | 265 | 288 | 262 | 5 | 5 | 1 | 117 | 5 | 3 | rs429358 | rs3916898;rs76971643;rs8104483 |
| 13c | Chr19:45.4Mb_13c | 265 | 288 | 262 | 5 | 5 | 1 | 117 | 5 | 3 | rs429358 | rs3916898;rs76971643;rs8104483 |
| 16 | Chr19:46.3Mb | 42 | 44 | 42 | 9 | 6 | 0 | 10 | 8 | 0 | | |
| 2 | Chr6:47.7Mb | 21 | 26 | 18 | 1 | 15 | 0 | 6 | 2 | 0 | | |
| 3 | Chr8:96Mb | 22 | 23 | 22 | 2 | 19 | 0 | 3 | 2 | 0 | | |
| 4 | Chr10:82.3Mb | 57 | 64 | 52 | 7 | 9 | 1 | 8 | 11 | 2 | rs7096909 | rs7096909;rs9665686 |
| 5 | Chr11:47.4Mb | 53 | 21 | 20 | 7 | 9 | 1 | 6 | 6 | 1 | rs71474198 | rs71474198 |
| 6 | Chr11:85.7Mb | 40 | 43 | 39 | 15 | 5 | 0 | 3 | 9 | 0 | | |
| 7 | Chr15:82.4Mb | 26 | 26 | 26 | 11 | 8 | 1 | 5 | 5 | 0 | rs111491984 | |
| 8 | Chr16:20.5Mb | 115 | 124 | 114 | 6 | 1 | 0 | 5 | 20 | 2 | rs28868590;rs9939588 | |
| 9 | Chr17:5.1Mb | 27 | 15 | 14 | 9 | 8 | 2 | 2 | 10 | 1 | rs55735639;rs8071989 | rs151195310 |
